# Bibliometric analysis of emerging trend and top 100 most cited articles in primary central nervous system lymphoma

**DOI:** 10.1101/2024.07.16.24310545

**Authors:** Yibo Geng, Yiqi Liu, Yang Wang, Xiong Li

## Abstract

**Background:** Over the past thirty years, numerous studies on primary central nervous system lymphoma (PCNSL) have been conducted, encompassing epidemiology, diagnosis, treatment, and mechanisms. However, no bibliometric analysis has been performed in this field. Therefore, we conducted a bibliometric analysis to elucidate the evolution and current status of PCNSL research and identify the most-cited articles across various disciplines.

**Material and Methods:** Literature published between January 1992 and June 2024 in the Web of Science Core Collection (WoSCC) was extracted and classified. Three platforms (R-bibliometrix, VOSviewer, and Citespace) were used for the bibliometric analysis. Negative binomial regression was employed to establish a model for the risk ratio of citation.

**Results:** A total of 1,798 publications were included in the analysis. The annual number of publications has shown a steady increase over time. The United States emerged as the leading country in terms of both the number of publications and total citations. Among individual researchers, Hoang-Xuan Khe from Sorbonne University was the most prolific in terms of publication count, while Lisa M. DeAngelis from Memorial Sloan Kettering Cancer Center received the highest number of citations. The study titled “Report of an International Workshop to Standardize Baseline Evaluation and Response Criteria for Primary CNS Lymphoma” was identified as the most cited work in this field. Keyword analysis indicated that immune therapy and non-invasive diagnosis are the primary focal points of current research. Among the top 100 most-cited articles, interventional studies demonstrated a higher citation ratio (RR = 1.357, 95% CI 1.044-1.764, P=0.023), whereas chemoradiotherapy studies exhibited a lower citation ratio (RR = 0.763, 95% CI 0.603-0.965, P=0.024).

**Conclusion:** Research hotspots and trends in PCNSL were identified and explored using bibliometric and visual methods. The highly cited articles across multiple disciplines related to PCNSL may help generate high-quality research ideas, particularly in the fields of immunotherapy and non-invasive diagnosis.

## Background

Primary central nervous system lymphoma (PCNSL) is an extranodal non-Hodgkin lymphoma that affects the brain, spine, and cerebrospinal fluid (CSF)^1^. This rare cancer has an annual incidence of 0.4 per 100,000 population and accounts for 4% to 6% of all extranodal lymphomas and 4% of newly diagnosed malignant brain tumors^2^. Immunosuppressed patients, such as those with HIV/AIDS or post-transplant patients, are at an increased risk for PCNSL, which is often associated with the Epstein-Barr virus^3^.

Bibliometric analysis provides a citation-based assessment of the significance and impact of individual articles within their respective fields. Officially defined and introduced in 1962^4^, this method consolidates evidence-based studies and examines trends in medical practice. To identify research hotspots and the citation ratios of various variables, we combined bibliometric analysis with negative binomial regression analysis (NBRA)^5^. In our study, we analyzed 1,798 documents related to PCNSL, summarizing the number of publications, citations, publication journals, and notable scientists. We also identified the top 100 most-cited documents and analyzed the variables influencing their citations.

## Material and Methods

### Search strategy

A comprehensive search was conducted in the Web of Science Core Collection (WoSCC) database, covering publications from January 1, 1992, to June 4, 2024. The search utilized the following keyword strategy: TS = [“PCNSL” OR “primary*central*nervous*system*lymphoma” OR “primary*CNS*lymphoma”]. No limitations were applied regarding the year of publication or language. This search yielded a total of 2,217 relevant documents. After screening for articles and reviews, 1,798 documents remained for analysis (Figure 1). The raw data is available in Supplementary File 1.

**Figure 1.**
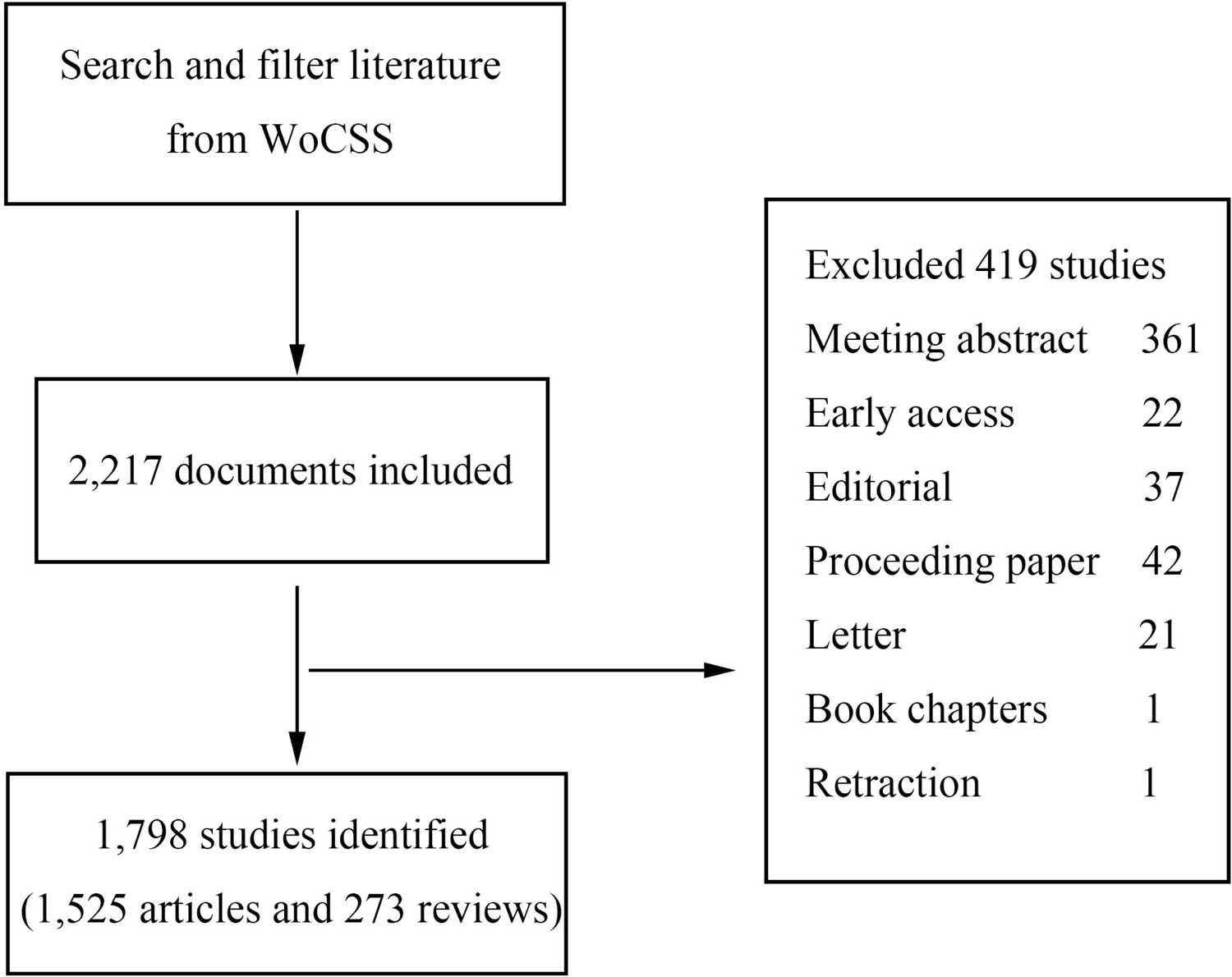
Flowchart of data filtration processing and excluding publications.

For the analysis of the top 100 most-cited papers, the 1,798 documents were sorted in descending order based on citation counts as defined by WoSCC. Two authors, Yibo Geng and Yiqi Liu, independently screened the articles. Any discrepancies were resolved through discussion.

### Data analysis

The analysis and visualization of bibliometric data were conducted using a suite of specialized software tools, including the R Bibliometrix package (R, Version 4.3.2), VOSviewer (Version 1.6.20), Citespace (Version 6.2.R7), and Microsoft Office Excel 2016^6,7^. This analysis encompassed PCNSL-related publications, institutions, journals, authors and their countries, keywords, citations, and other relevant indicators to comprehensively evaluate the quality and impact of the publications.

### Documents classification

We classified the study designs into reviews, laboratory studies, bioinformatic analyses, observational studies, and interventional studies. All papers were further grouped according to the most general topics: chemoradiotherapy, cognition, diagnosis, epidemiology, genetic alteration, guidelines, imaging, knowledge, and risk factors.

### Statistics

Negative binomial regression analysis was employed to determine associations between the total number of citations (WoSCC) and various factors, including study design, topic, region, open access status, and publication year, using SPSS software (Version 27.0.1.0, IBM Corp). Study design was categorized into three groups: interventional studies, observational studies, and others (including reviews, bioinformatic analyses, and laboratory studies). Topics were similarly categorized into three groups: chemoradiotherapy, knowledge, and others (encompassing cognition, diagnosis, epidemiology, genetic alterations, guidelines, imaging, and risk factors). The results were presented as rate ratios (RR) with 95% confidence intervals (CI), and a P-value of less than 0.05 was considered statistically significant.

## Results

### Trends and annual publications

The volume of global publications in the field of PCNSL exhibited consistent growth, rising from 7 publications in 1992 to a peak of 157 publications in 2023 (Figure 2A). The overall trend shows a smooth upward trajectory, with a significant increase beginning in 2018 (Figure 2B).

**Figure 2.**
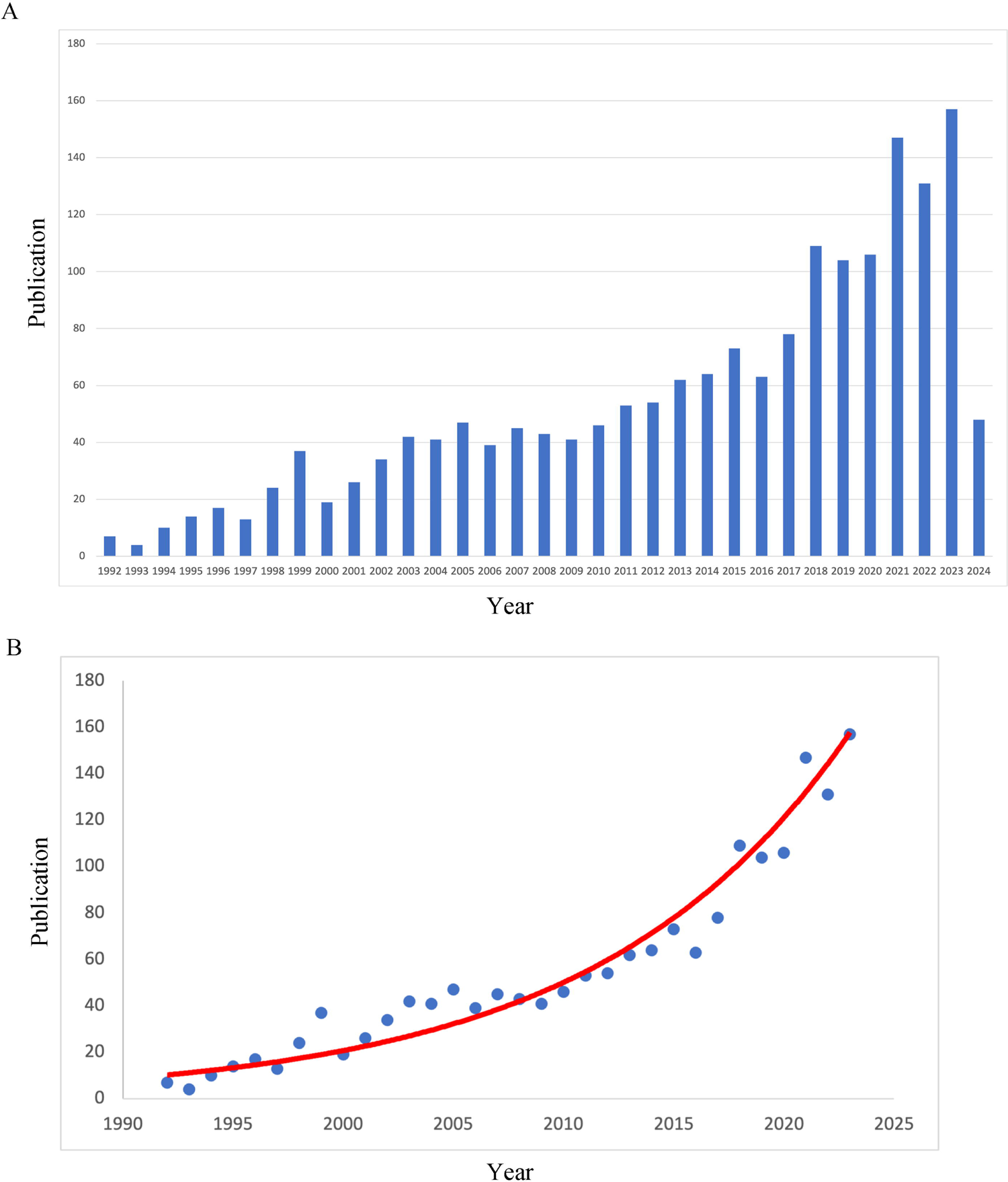
Analysis of publication trend in PCNSL. (A) Annual global publications between 1992 to 2024. (B) The publication consistently increased suggested by time curve (The data of incomplete 2024 year was excluded).

### Countries and organizations

A total of 1,798 articles originated from 62 different countries and regions. The frequency of scientific production by country is shown in Figure 3A. The United States led with 395 papers (22.0% of the total), followed by China with 310 papers (17.2%), Germany with 225 papers (12.5%), Japan with 203 papers (11.3%), and France with 104 papers (5.8%) (Figure 3B). In terms of citations, the United States amassed the highest number, with 18,909 citations. Germany followed with 7,613 citations, Japan with 3,525 citations, and France with 3,502 citations (Figure 3C). An analysis of total link strength among 43 countries revealed the top five nations as the United States (211 times), Germany (189 times), Italy (131 times), Switzerland (117 times), and France (108 times) (Figure 4A).

**Figure 3.**
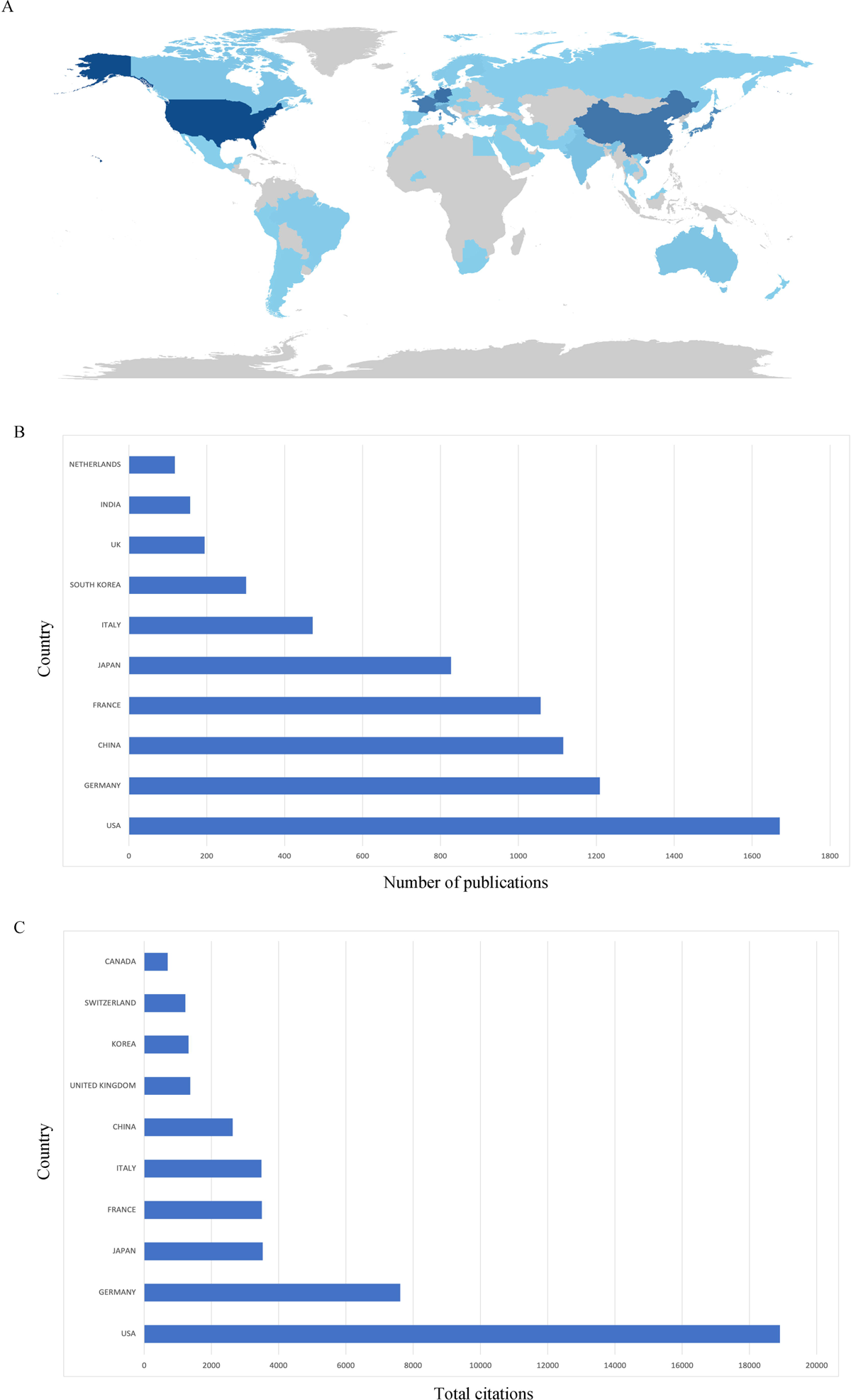
Global countries investigating PCNSL. (A) The heatmap of global publication distribution. (B) Publications of top ten countries. (C) Citations of top ten countries.

**Figure 4.**
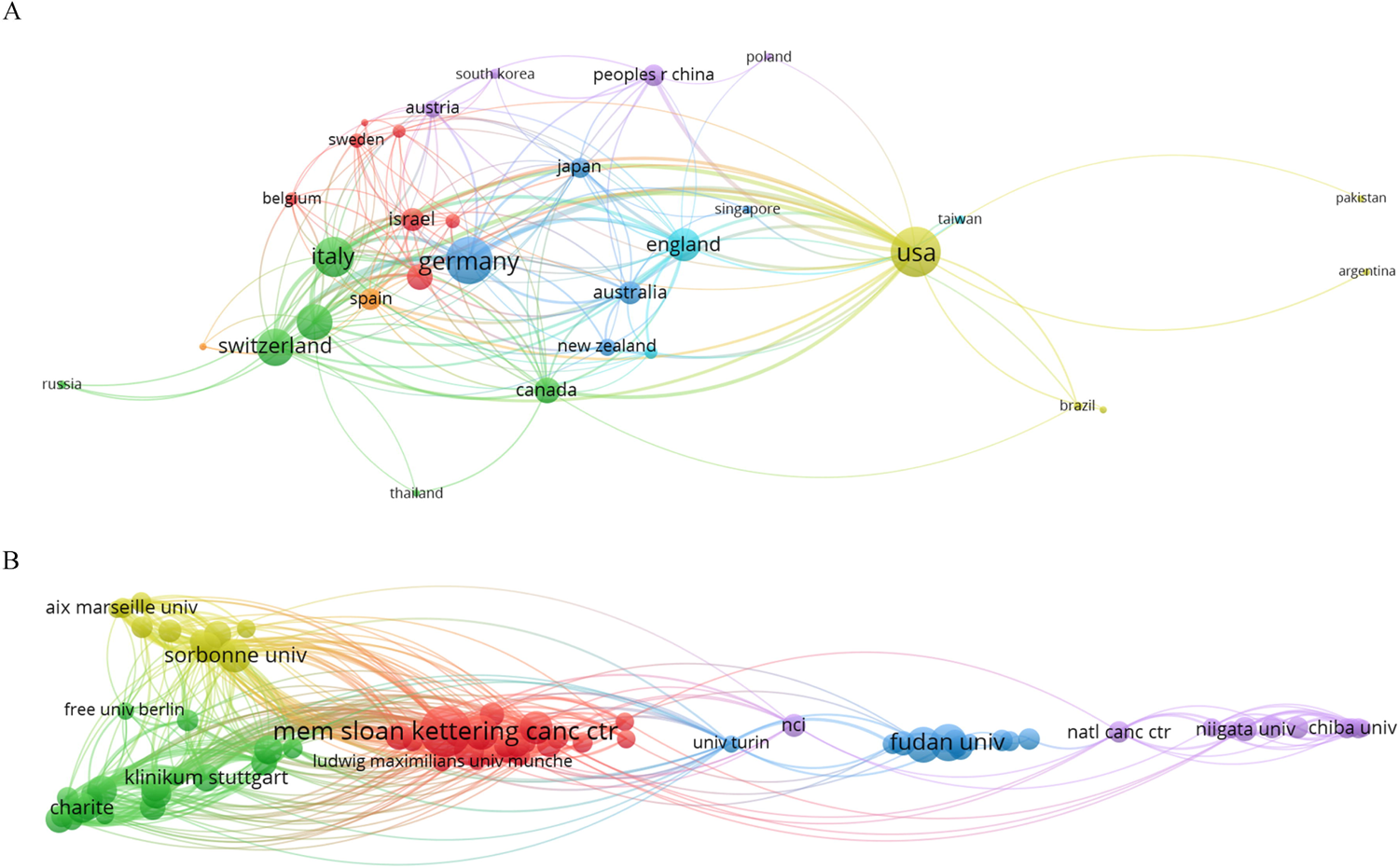
Total link strength analysis of countries and organizations. A network map revealing correlation between countries (A) and organizations (B).

Regarding organizational contributions, a total of 2,354 institutions participated in brainstem tumor research. The Memorial Sloan Kettering Cancer Center had the most substantial impact, with 100 records (5.6% of all articles), followed by Fudan University (51 records, 2.8%), Capital Medical University (45 records, 2.5%), Mayo Clinic (44 records, 2.5%), and Sorbonne University (39 records, 2.2%). The assessment of total link strength among institutions covered 79 institutions, which were summarized into five clusters by region: America (red), France (yellow), Germany (green), China (blue), and Japan (purple) (Figure 4B). Among the top institutions in terms of link strength were Memorial Sloan Kettering Cancer Center (131), Hôpital Universitaire Pitié-Salpêtrière (118), Sorbonne University (117), Massachusetts General Hospital (86), and Institut Curie (78) (Figure 4B).

### Sources of publications

The top 10 journals from a pool of 471 journals with the highest number of PCNSL-related publications are summarized in Table 1. The Journal of Neuro-Oncology led with 143 records (8.0% of all articles), followed by Leukemia & Lymphoma (48 records, 2.7%), Neuro-Oncology (43 records, 2.4%), Cancers (37 records, 2.1%), and Neurology (37 records, 2.1%). Co-citation analysis among journals, illustrated in Figure 5, showed a pronounced multi-center trend, with active citation relationships among the Journal of Clinical Oncology, Journal of Neuro-Oncology, Blood, and Neuro-Oncology. The top five journals with the highest number of cited brainstem tumor-related publications were Journal of Clinical Oncology (6,730 citations), Journal of Neuro-Oncology (4,009 citations), Blood (2,829 citations), Neurology (2,748 citations), and Cancer (2,064 citations). Notably, the Journal of Clinical Oncology, despite not being the top in publication count, had a significantly higher citation count per paper (240 citations/paper), indicating its high impact and limited publication frequency.

**Figure 5.**
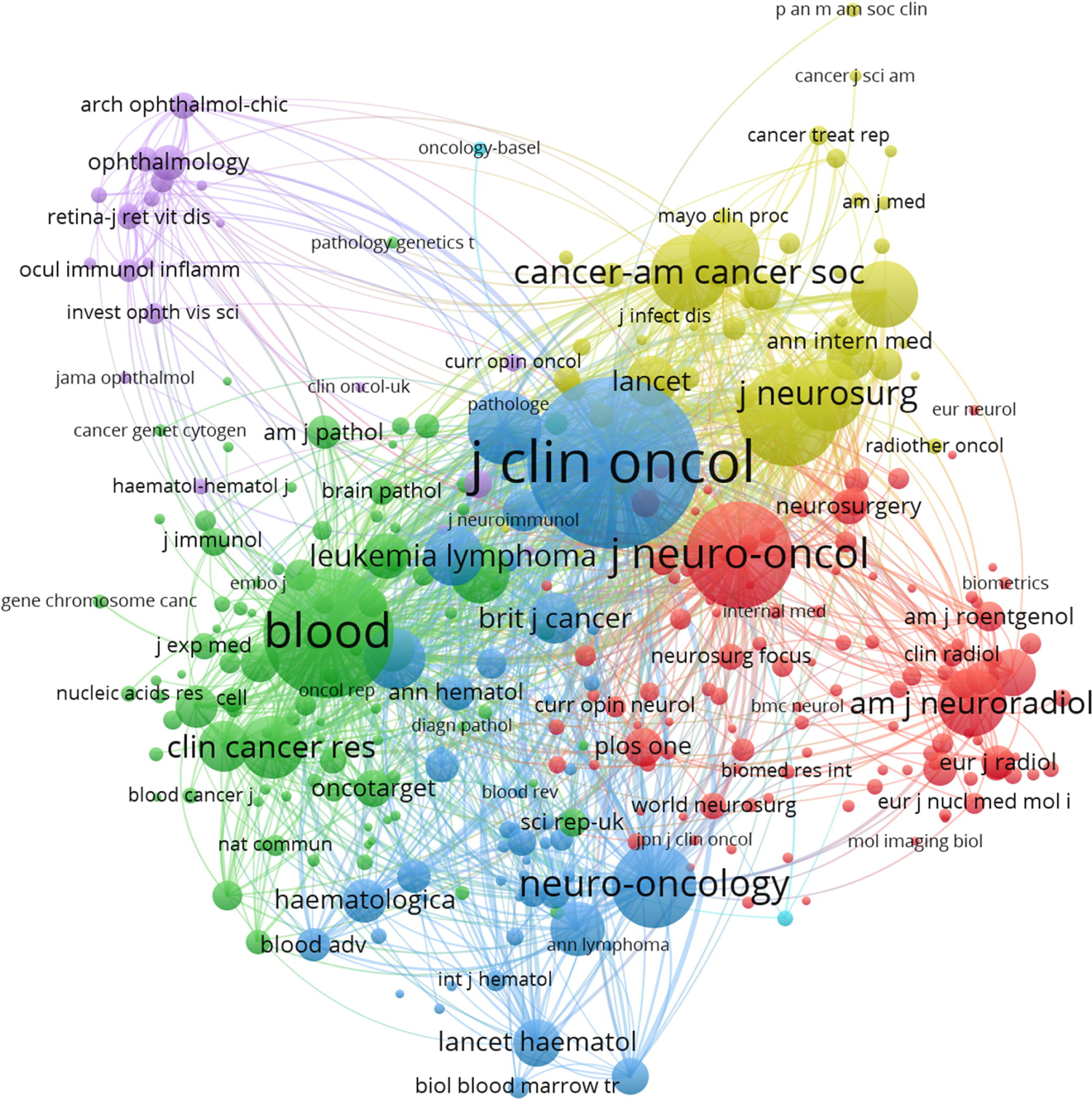
Cluster visualization for journals co-citation network.

**Table 1.** Top 10 journals that have the outmost number of publications and cited journals have the largest number of citations.

| Journals | Number of publications | Impact Factor (partition) | Cited journals | Number of citations | Impact Factor (partition) |
| --- | --- | --- | --- | --- | --- |
| Journal of neuro-oncology | 143 | 3.2 (Q2) | Journal of clinical oncology | 6326 | 42.1 (Q1) |
| Leukemia & lymphoma | 48 | 2.2 (Q3) | Blood | 3711 | 21.0 (Q1) |
| Neuro-oncology | 43 | 16.4 (Q1) | Journal of neuro-oncology | 2502 | 3.2 (Q2) |
| Cancers | 37 | 4.5 (Q1) | Neurology | 2237 | 7.7 (Q1) |
| Neurology | 37 | 7.7 (Q1) | Neuro-oncology | 1613 | 16.4 (Q1) |

### Authors and co-cited authors

A total of 10,205 authors have contributed articles related to PCNSL. Hoang-Xuan Khe from Sorbonne University was the most prolific author with 45 articles (Figure 6A, 6B). He was followed by Carole Soussain from Institut Curie with 39 publications, Andrés J. M. Ferreri from IRCCS San Raffaele Scientific Institute with 37, and Caroline Houillier from Sorbonne Université with 34. In terms of citations, LM DeAngelis led with 4,072 citations, followed by Lauren E. Abrey with 2,929 citations, Lisa M. DeAngelis with 2,485 citations, Andrés J. M. Ferreri with 2,276, and Hoang-Xuan Khe with 1,827 citations (Figure 6C).

**Figure 6.**
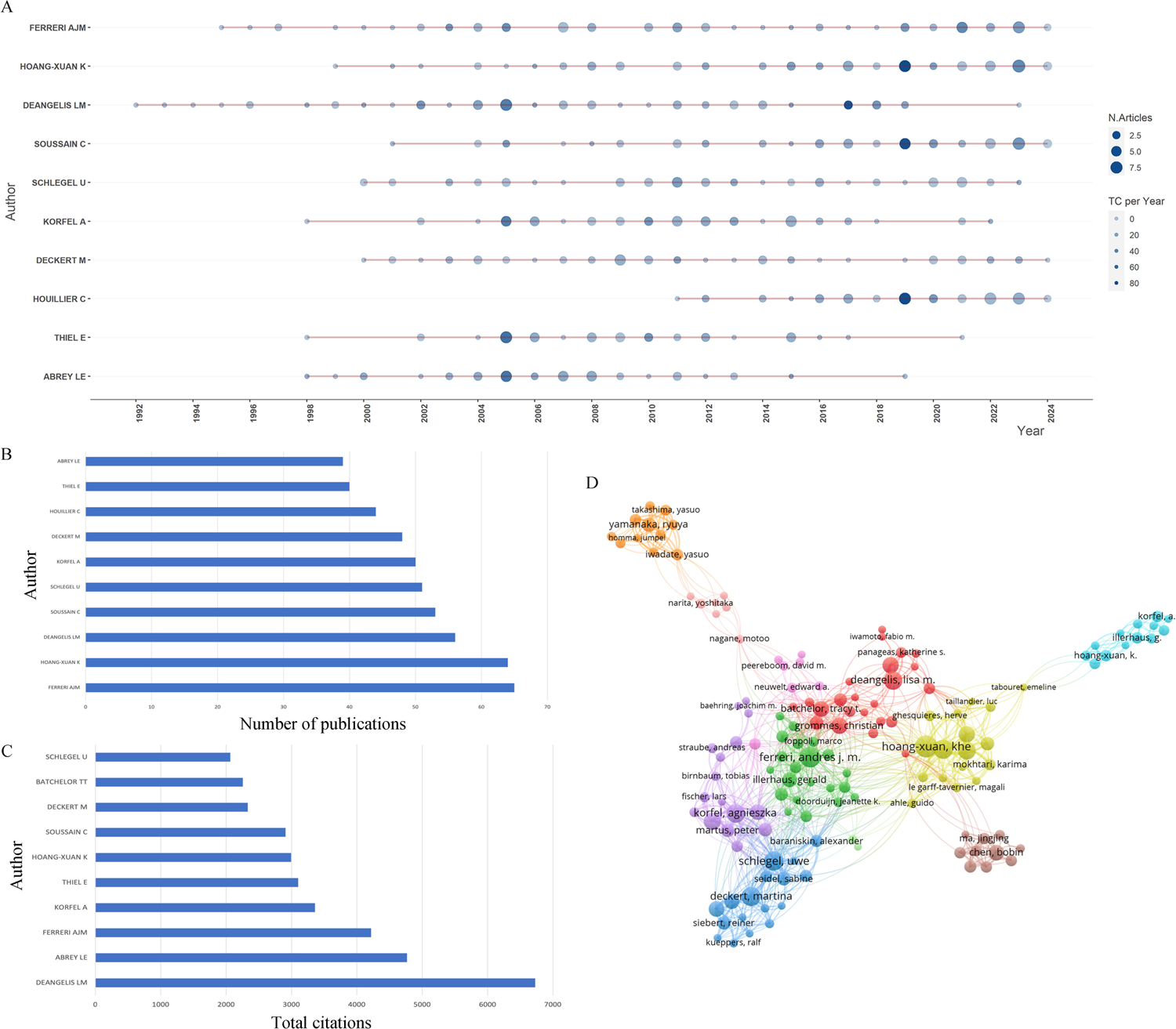
Authors analysis. (A) Top ten authors’ production over time. Node size indicated annual number of publications, and node color indicated annual total citation. (B) The top ten productive authors. (C) The top ten citation authors. (D) Co-occurrence analysis of 144 authors.

The h-index, which measures both the productivity and impact of researchers, identified the top five authors as LM DeAngelis (39), Andrés J. M. Ferreri (36), Lauren E. Abrey (32), Agnieszka Korfel (29), and Martina Deckert and Eckhard Thiel (both with 27) (Supplementary Figure 1). Co-author analysis revealed several prominent research groups led by Hoang-Xuan Khe, Andrés J. M. Ferreri, Agnieszka Korfel, Martina Deckert, Lisa M. DeAngelis, Ryuya Yamanaka, and Bobin Chen (Figure 6D).

### The hotspots and frontiers

Keywords summarize research topics and core content. Keyword co-occurrence analysis helps understand the distribution and development of research hotspots. Excluding keywords directly associated with PCNSL, such as “PCNSL”, “primary CNS lymphoma”, “lymphoma”, “central nervous system”, “non-Hodgkins lymphoma”, and “nervous system lymphoma”, the most frequent keywords were chemotherapy (393), survival (333), methotrexate (329), and radiotherapy (266) (Figure 7A). These keywords were summarized into 14 clusters, including #0 prognosis, #1 neurotoxicity, #2 rituximab, #3 glioblastoma, #4 clinical presentation, #5 acquired immune deficiency syndrome, #6 myd88, #7 intraocular lymphoma, #8 HAART, #9 acquired immunodeficiency syndrome, #10 bcl-2, #11 Epstein-Barr virus genome, #12 perivascular reticulin network, #13 natural rubber, and #14 radiotherapy (Figure 7B). Over time, the most frequent keywords, such as methotrexate, chemotherapy, radiotherapy, survival, and diagnosis, were active around 2010 (Figure 7C). Recent trends in PCNSL research have shifted from chemoradiotherapy and risk factors to immunotherapy and non-invasive diagnosis. However, no novel keywords have emerged in the central position of the occurrence network, indicating that these hotspots still require further investigation.

**Figure 7.**
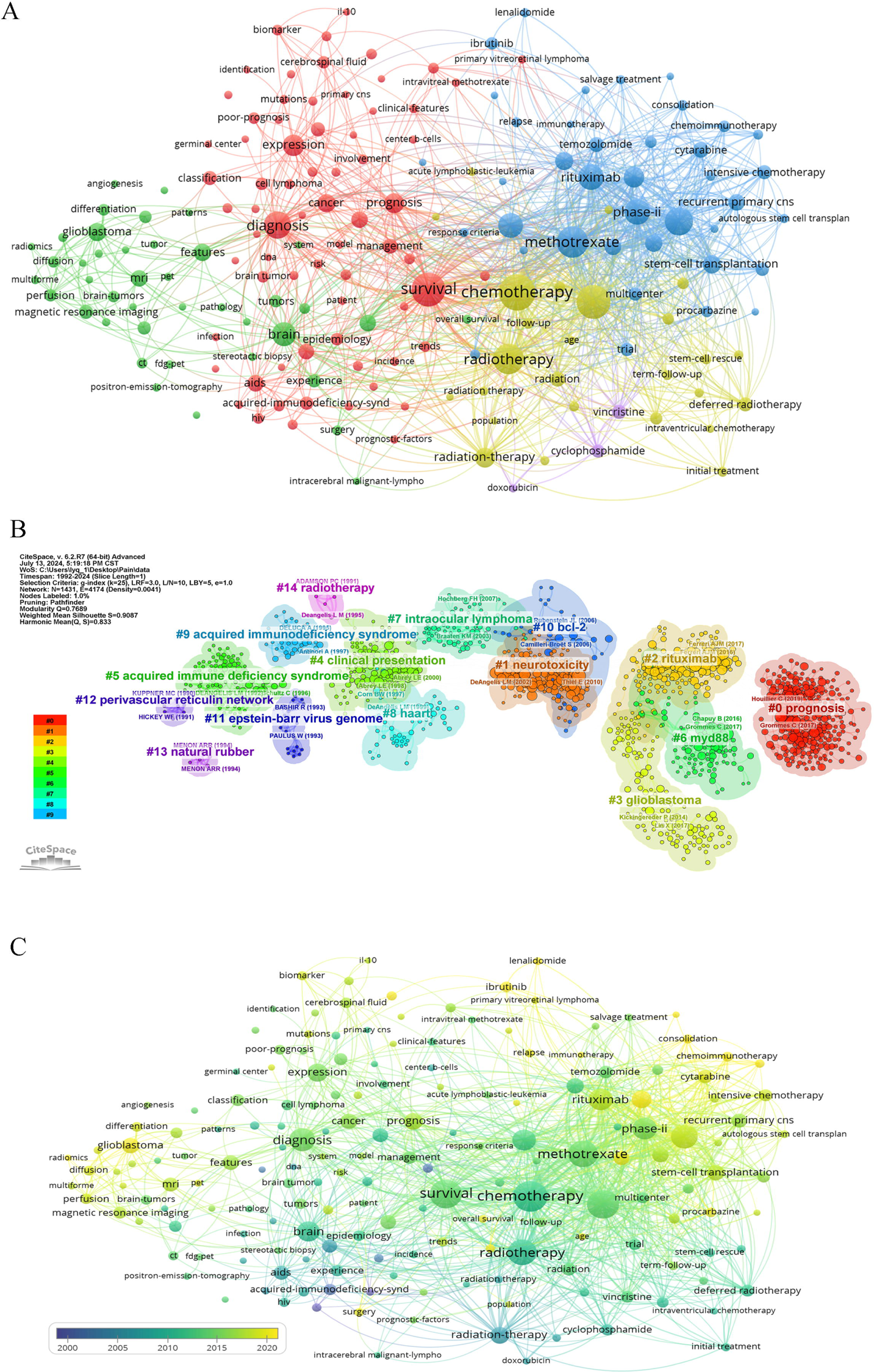
Keyword analysis (1992–2024). (A) Cluster visualization of keywords. (B) A total of 14 clusters are distinguished by different colors. Cluster #0 is the largest, followed by cluster #1, and so on. A cluster label was assigned by CiteSpace based on the terms that appear most frequently in relevant articles. (C) Timeline visualization of keywords.

### Top 100 cited Document citation analysis

We organized the 1,798 documents in descending order of citations, identifying the top 100 most-cited papers (Supplementary Table 1). These articles garnered a total of 19,011 citations, with thirteen receiving more than 300 citations. The most-cited article was an evaluation guideline published in the Journal of Clinical Oncology in 2005, with 593 citations. The article with the highest citation density (48.6) among the top 100 revealed genetic features of PCNSL and was published in Blood in 2016. The United States published 52% of the high-cited articles, accounting for 62.1% of the citations. Only 3% of the highly-cited articles were from outside America or Europe (Table 2).

**Table 2.** Country and their citations in the top 100 citation papers.

| Country | Papers | Citations |
| --- | --- | --- |
| USA | 52 | 10796 |
| Germany | 18 | 3182 |
| France | 13 | 2117 |
| Italy | 6 | 1288 |
| UK | 3 | 408 |
| Switzerland | 2 | 284 |
| Japan | 2 | 279 |
| Netherlands | 2 | 261 |
| Norway | 1 | 265 |
| Canada | 1 | 131 |

Interventional studies accounted for 34% of publications and 7,193 citations, followed by observational studies (30% of articles and 5,378 citations). The oldest and most frequently cited topic was chemoradiotherapy, which also accumulated the highest number of citations (42% of articles and 8,249 citations). Other identified topics included genetic alterations (12%), knowledge (12%), risk factors (11%), imaging (7%), diagnosis (6%), and epidemiology (6%) (Table 3).

**Table 3.** Topic of interest and study design in top 100 citation papers.

| Variables | Papers | Citations |
| --- | --- | --- |
| Study design |  |  |
| Interventional study | 34 | 7193 |
| Observational study | 30 | 5378 |
| Laboratorial | 7 | 1388 |
| Review | 18 | 3379 |
| Bioinformatic analysis | 11 | 1673 |
| Topic of interest |  |  |
| Chemoradiotherapy | 42 | 8249 |
| Cognition | 4 | 511 |
| Diagnosis | 6 | 948 |
| Epidemiology | 6 | 870 |
| Genetic alteration | 12 | 1904 |
| Guideline | 1 | 593 |
| Imaging | 7 | 1258 |
| Knowledge | 12 | 1796 |
| Risk factor | 11 | 2882 |

The regression model indicated that interventional studies (RR = 1.357; 95% CI, 1.044-1.764, P=0.023) had a higher citation ratio, while chemoradiotherapy (RR = 0.763; 95% CI, 0.603-0.965, P=0.024) had a lower citation ratio. Additionally, papers authored by researchers from the United States showed a tendency towards a higher citation ratio (RR = 1.222; 95% CI, 0.999-1.494, P=0.051) (Table 4).

**Table 4.** Negative Binomial Regression Analysis between total number of citations and bibliometric data.

| Variables | n (%) | Risk ratio (95% CI) | P |
| --- | --- | --- | --- |
| Study design |  |  |  |
| Interventional study | 34 | 1.357 (1.044-1.764) | 0.023 |
| Observational study | 30 | 0.923 (0.719-1.185) | 0.53 |
| Others | 36 | 1 |  |
| Topic of interest |  |  |  |
| Treatment | 42 | 0.763 (0.603-0.965) | 0.024 |
| Knowledge | 12 | 0.872 (0.688-1.105) | 0.258 |
| Others | 46 | 1 |  |
| Worldwide distribution |  |  |  |
| America | 52 | 1.222 (0.999-1.494) | 0.051 |
| Europe & Others | 48 | 1 |  |
| Open access |  |  |  |
| No | 50 | 1.066 (0.860-1.322) | 0.56 |
| Yes | 50 | 1 |  |
| Publication period |  |  |  |
| 1993-2006 | 52 | 1.125 (0.918-1.378) | 0.257 |
| 2007-2020 | 48 | 1 |  |

## Discussion

To our knowledge, this is the first bibliometric study to evaluate the articles published on PCNSL and analyze the contributions of various factors to citation counts. As evidenced by our findings, the annual publication rate has shown consistent growth. Prior to 2018, the number of publications never exceeded 80 per year; however, in the last five years, this number has doubled, suggesting that this rare intracerebral tumor has garnered increasing attention from the scientific community. Our analysis of national contributions indicates that the USA holds the greatest influence in this field, leading in both the number of publications and citations. Notably, the gap between the USA and other countries is smaller compared to other fields of research^8,9^. European countries, particularly Germany, France, and Italy, also make significant contributions, which reflect their early initiation of high-quality clinical trials and extensive research history. China, ranked third in publications and sixth in citations, still lacks high-quality interventional studies focusing on diagnosis, treatment, and prognosis. Interestingly, two of the top five institutions in terms of publications, Fudan University and Capital Medical University, are located in China, indicating that PCNSL research in China is concentrated in major neurosurgical centers. While the volume of research in these centers surpasses that of individual centers in the USA or Europe, the quality of research in China needs improvement. Thus, we can conclude that the USA maintains a leading position, with both the USA and Europe conducting multicenter, high-quality research, while China’s research is concentrated in a few neurological centers.

Regarding author contributions, notable scientists such as Deangelis LM, Hoang Xuan K, and Ferreri AJM have made significant impacts. Deangelis LM from Memorial Sloan Kettering Cancer Center established the first-line treatment for primary and recurrent PCNSL using high-dose methotrexate through prospective interventional studies, with a publication period spanning over 30 years^10^. Hoang Xuan Khe from Sorbonne University identified typical mutations in PCNSL and explored the effects of novel targeted and precision drugs through randomized clinical trials in France^11^. Ferreri AJM from IRCCS San Raffaele Scientific Institute conducted several clinical trials comparing different chemotherapy regimens and is currently focused on stem-cell transplantation^12^.

Methotrexate (MTX) remains the cornerstone of treatment, often combined with radiotherapy, and clinical trials involving novel immune and pathway inhibitors are central to the field^13^. Despite significant advances, PCNSL diagnosis remains challenging before histopathological confirmation, as it can easily be confused with glioma, metastasis, tumefactive demyelinating lesions, or infections based on MRI findings^14,15^. Researchers are paying significant attention to non-invasive diagnostic methods such as MRS^16^, amide proton transfer-weighted imaging^17^, PET^18^, and CSF analysis. Yamaguchi et al. demonstrated that PET-MRI could diagnose PCNSL with good accuracy^19^. Recent studies on CSF have focused on flow cytometry, cytomorphology, interleukins-6/10, and miRNA-21/92 for diagnosing meningeal involvement from lymphoid neoplasms^20–23^, as well as miR-222 in blood^24^. Liquid biopsy, which involves analyzing tumor-derived DNA from blood specimens, offers a non-invasive alternative to tissue biopsies^25^. MYD88 mutations detected in cell-free DNA fragments in the bloodstream have been reported as sensitive and specific biomarkers for differentiating PCNSL from other CNS cancers, with the L265P variant associated with poor prognosis, particularly in elderly patients^26,27^.

NBRA is a statistical modeling technique used to model over-dispersed count outcome variables and examine the relationship between a dependent variable representing counts or frequencies and one or more independent variables^28^. In our study, we analyzed variables influencing citations among the top 100 cited documents using NBRA. The results indicated that interventional studies published in the USA tend to receive more citations, while treatment studies tend to receive fewer citations. This finding highlights the hotspots in study type, topic, and region within the PCNSL field.

While our study provides valuable insights, it has certain limitations. Firstly, our analysis relied exclusively on data from the WoSCC database. A more comprehensive perspective could have been achieved by incorporating additional databases such as Scopus and PubMed, capturing a broader spectrum of research output. Secondly, our study may underrepresent cutting-edge research and emerging technologies, as these may not have accumulated substantial citation counts at the time of our analysis. Lastly, our focus on the top 100 cited documents may not fully reflect the comprehensive influence of the analyzed variables on the total citation landscape.

## Conclusion

Overall, this bibliometric analysis elucidates the evolution, current status, and influential factors in PCNSL research, providing a foundation for future investigations and collaborations to advance the understanding and treatment of PCNSL.

## Data Availability

All data produced in the present work are contained in the manuscript

## Acknowledgements

All authors contributed to the study conception and design. Study design, data collection and analysis were performed by Yibo Geng, Yiqi Liu and Yang Wang. The first draft of the manuscript was written by Yibo Geng and supervised by Xiong Li. All authors read and approved the final manuscript.

## Funding

This work was supported by the Beijing Municipal Natural Science Youth Foundation (No.7244344), Beijing Chao-yang Hospital Golden Seed foundation (No. CYJZ202201) and the inner-hospital foundation of Beijing Chao-yang Hospital.

## Notes

### Competing Interest Statement

The authors have declared no competing interest.

