## Supplementary material for "Bibliometric analysis of emerging trend and top 100 most cited articles in primary central nervous system lymphoma": Supple File 1: savedrecs (1).txt

FN Clarivate Analytics Web of Science
VR 1.0
PT J
AU Paul, RK
   Pillai, CKS
AF Paul, RK
   Pillai, CKS
TI Melt/solution processable polyaniline with functionalized phosphate
   ester dopants and its thermoplastic blends
SO JOURNAL OF APPLIED POLYMER SCIENCE
LA English
DT Article
DE polyaniline; doping studies; phosphate ester dopants; melt/solution
   processibility; thermoplastic blends
ID ELECTRICALLY CONDUCTIVE POLYANILINE; COUNTERION INDUCED PROCESSIBILITY;
   POLY(ALKYLENE PHOSPHATES); POLYMERS; METHACRYLATE); PROTONATION;
   MORPHOLOGY; COMPOSITES; ACIDS
AB Polyaniline (PANI) was doped with five novel dopants, 3-pentadecylphenylphosphoric acid (PDPPA), pentadecylphenyl(bis)phosphoric acid [PDP(bis)PAI, monocardanylphosphoric acid (MCPA), dicardanylphosphoric acid (DCPA), and phosphorylated cashew nut shell liquid prepolymer (PCNSL) and the doping behavior was studied. All dopants were synthesized from inexpensive naturally existing monomers [obtained from cashew nut shell liquid (CNSL)] having a long hydrophobic hydrocarbon side chain in the meta position of the aromatic ring. These dopants can act as plasticizing cum protonating agents for PANI so that free-standing films of PANI could be prepared by both thermal processing and solution processing techniques. Protonatiun was performed either by mechanical mixing of emeraldine base and the dopant or by an in situ doping emulsion polymerization route using xylene or chloroform as the solvent. Further, conductive flexible blends of the protonated PANl with poly(vinyl chloride) (PVC) were also prepared and studied for their conductivity and related properties. The PANI-PDPPA complex obtained by the in situ doping emulsion polymerization route exhibited an exceptionally high degree of crystalline order and orientation. A maximum conductivity value of 1.8 S cm(-1) was obtained for a PANI-PDPPA film hot-pressed at 120 degreesC. On the other hand, dopants based on cardanol having an unsaturated side chain gave only lower values. This was understood to be due the capability of the saturated analog to contribute to the ordered arrangement of PANI, thus improving the crystallinity. The conductivity values further decreased when bulky/oligomeric dopants such as PCNSL were used. The thermoplastic blends with PVC exhibited an exceptionally low-level percolation threshold because of the plasticizing nature of the dopants, The doped polymers and blends were characterized by FTIR and W-visible spectroscopic methods, four-probe conductivity measurements, XRD, SEM, TGA, and DSC. (C) 2001 John Wiley & Sons, Inc.
C1 CSIR, Reg Res Lab, Trivandrum 695019, Kerala, India.
C3 Council of Scientific & Industrial Research (CSIR) - India; CSIR -
   National Institute Interdisciplinary Science & Technology (NIIST)
RP Pillai, CKS (corresponding author), CSIR, Reg Res Lab, Trivandrum 695019, Kerala, India.
CR ANGELOPOULOS M, 1997, HDB CONDUCTIVE POLYM
   ANTONY R, 1993, J APPL POLYM SCI, V49, P2129, DOI 10.1002/app.1993.070491209
   ASTURIAS GE, 1989, SYNTHETIC MET, V29, P157
   BALLAUFF M, 1987, J POLYM SCI POL PHYS, V25, P739, DOI 10.1002/polb.1987.090250404
   Banerjee P, 1995, SYNTHETIC MET, V74, P257, DOI 10.1016/0379-6779(95)03370-Y
   Bredas J.L., 1991, Conjugated Polymers
   CAO Y, 1993, POLYMER, V34, P3139, DOI 10.1016/0032-3861(93)90381-J
   CAO Y, 1992, SYNTHETIC MET, V48, P91, DOI 10.1016/0379-6779(92)90053-L
   CAO Y, 1992, Patent No. 2222911
   CHAN HSO, 1994, MACROMOLECULES, V27, P2159, DOI 10.1021/ma00086a027
   CHIANG CK, 1977, PHYS REV LETT, V39, P1098, DOI 10.1103/PhysRevLett.39.1098
   CHIANG JC, 1986, SYNTHETIC MET, V13, P193, DOI 10.1016/0379-6779(86)90070-6
   CONN C, 1995, ADV MATER, V7, P790, DOI 10.1002/adma.19950070905
   COYHM, 1995, J AM CHEM SOC, V117, P33
   DAPRANO G, 1995, CHEM MATER, V7, P33, DOI 10.1021/cm00049a008
   FROMMER JE, 1988, ENCY POLYM SCI ENG, V5
   GREENBERG R, 1985, STANDARD METHODS EXA, P437
   IKKALA OT, 1995, SYNTHETIC MET, V69, P97, DOI 10.1016/0379-6779(94)02376-A
   JARVINEN H, 1995, Patent No. 07258409
   JARVINEN H, 1995, Patent No. 668594
   KARHU E, 1998, Patent No. 101075
   KARHU E, 1995, Patent No. 9528716
   KARNA T, 1999, Patent No. 5928565
   KulszewiczBajer I, 1996, POLYMER, V37, P25, DOI 10.1016/0032-3861(96)81596-5
   KULSZEWICZBAJER I, 1995, MACROMOLECULES, V28, P610, DOI 10.1021/ma00106a028
   LAASKA J, 1995, J POLYM SCI POL CHEM, V33, P1437
   LAASKA J, 1995, SYNTHETIC MET, V69, P113
   LEVON K, 1996, POLYM PREPR, V37, P60
   MACDIARMID AG, 1991, SCIENCE AND APPLICATIONS OF CONDUCTING POLYMERS, P117
   OKA O, Patent No. 95239207
   OKA O, 1991, Patent No. 9119356509
   Ong CH, 1997, POLYMER, V38, P1065, DOI 10.1016/S0032-3861(96)00618-0
   OSTERHOLM JE, 1993, SYNTHETIC MET, V55, P1034, DOI 10.1016/0379-6779(93)90195-3
   Paul RK, 1999, SYNTHETIC MET, V104, P189, DOI 10.1016/S0379-6779(99)00063-6
   PILLAI KS, 1988, Patent No. 176069
   Pron A, 1996, SYNTHETIC MET, V80, P191, DOI 10.1016/S0379-6779(96)03701-0
   PRON A, 1993, POLYMER, V34, P4235, DOI 10.1016/0032-3861(93)90182-A
   Roichman Y, 1999, J MACROMOL SCI PHYS, VB38, P145, DOI 10.1080/00222349908248111
   RUCKENSTEIN E, 1993, SYNTHETIC MET, V53, P283, DOI 10.1016/0379-6779(93)91097-L
   Singh R, 1997, POLYMER, V38, P4897, DOI 10.1016/S0032-3861(97)00013-X
   Skotheim TA, 1989, HDB CONDUCTING POLYM, V11
   Skotheim TA, 1989, HDB CONDUCTING POLYM, V1
   SUBRAMANIAM K, 1992, SOLID STATE COMMUN, V97, P235
   SUBRAMANIAM K, 1993, J POLYM SCI PP, V31, P25
   TERLEMEZYAN L, 1995, POLYM J, V27, P867
   TRAORE MK, 1991, SYNTHETIC MET, V40, P137, DOI 10.1016/0379-6779(91)91770-B
   Vikki T, 1996, MACROMOLECULES, V29, P2945, DOI 10.1021/ma951555v
   WAN P, 1999, Patent No. 9941756
   WAN P, 1999, Patent No. 5908898
   WANG ZH, 1991, PHYS REV LETT, V66, P1745, DOI 10.1103/PhysRevLett.66.1745
   WEI Y, 1989, J PHYS CHEM-US, V93, P495, DOI 10.1021/j100338a095
   WESSLING B, 1997, HDB CONDUCTIVE POLYM
   WESSLING B, 1999, HDB NANOSTRUCTURED M, V5, P501
   YANG CY, 1993, SYNTHETIC MET, V53, P293, DOI 10.1016/0379-6779(93)91098-M
   YANG SY, 1993, SYNTHETIC MET, V59, P1, DOI 10.1016/0379-6779(93)91152-R
   ZHENG W, 1995, J POLYM SCI PP, V33, P89
   ZHENG WY, 1994, MACROMOLECULES, V27, P7754, DOI 10.1021/ma00104a034
   FARADAY DISCUSS CHEM
NR 58
TC 28
Z9 30
U1 0
U2 15
PU JOHN WILEY & SONS INC
PI NEW YORK
PA 605 THIRD AVE, NEW YORK, NY 10158-0012 USA
SN 0021-8995
J9 J APPL POLYM SCI
JI J. Appl. Polym. Sci.
PD MAY 31
PY 2001
VL 80
IS 9
BP 1354
EP 1367
DI 10.1002/app.1224
PG 14
WC Polymer Science
WE Science Citation Index Expanded (SCI-EXPANDED)
SC Polymer Science
GA 415XY
UT WOS:000167749900003
DA 2024-06-03
ER

PT J
AU Krogh-Jensen, M
   Johansen, P
   D'Amore, F
AF Krogh-Jensen, M
   Johansen, P
   D'Amore, F
TI Primary central nervous system lymphomas in immunocompetent individuals:
   Histology, Epstein-Barr virus genome, Ki-67 proliferation index, p53 and
   bcl-2 gene expression
SO LEUKEMIA & LYMPHOMA
LA English
DT Article
DE lymphoma; non-Hodgkin's; primary central nervous system lymphoma;
   Epstein-Barr virus genome; protein bcl-2; protein p53; proliferation
   index
ID NON-HODGKINS-LYMPHOMAS; PRIMARY BRAIN LYMPHOMA; IMMUNODEFICIENCY
   SYNDROME; PROTEIN EXPRESSION; PROGNOSTIC FACTORS; CLINICOPATHOLOGICAL
   FEATURES; CLINICAL-SIGNIFICANCE; NUCLEAR ANTIGEN; CELL LYMPHOMA; AIDS
AB Epstein-Barr virus (EBV) has been detected in the large majority of HN-related primary central nervous system lymphomas (PCNSL) suggesting a pathogenetic role of the virus. Unlike HIV-related PCNSL, conflicting data exist with regard to the presence of EBV in non immunodeficiency-related (sporadic) PCNSL: For this reason, a population based material of 41 sporadic PCNSL was analysed for the presence of EBV genome (EBER, BHLF) using RNA in situ hybridisation (RISH). Furthermore, the expression of the gene products of the bcl-2 oncogene and the p53 tumor suppressor gene and the tumor growth fraction reactive with the monoclonal antibody Ki-67 have been evaluated. All cases but two were EBV genome negative. In the two positive cases less than 5% of tumor cells showed EBER positivity. In contrast, more than 75% of cells morphologically belonging to the tumor-cell population stained positively for EBER in two cases of HIV related PCNSL. Immunostaining for the bcl-2 oncoprotein was positive in 28 (72%) of 39 cases examined. In most cases more than 75% of tumor cells showed cytoplasmic expression. Of 37 cases investigated for p53 expression, 21 (57%) stained positively. However, in the large majority of positive cases less than 10% of the neoplastic cells stained. The percentage of Ki-67 positive cells ranged between 10% and 80% with a mean of 50%. The expression of the p53 and bcl-2 oncoproteins and the growth fraction did not have any prognostic impact.
   We conclude that the EBV genome is rarely detected in sporadic PCNSL, indicating that a pathogenetic role of EBV is unlikely. Like extracerebral B-cell lymphomas a large fraction of PCNSL expresses the p53 and bcl-2 oncoproteins, a feature, however, which does not seem to have prognostic implications.
C1 Aalborg Hosp, Dept Haematol, Aalborg, Denmark.
C3 Aalborg University; Aalborg University Hospital
RP Krogh-Jensen, M (corresponding author), Copenhagen Univ Hosp, Rigshosp, Dept Haematol L, DK-2100 Copenhagen O, Denmark.
CR AUPERIN I, 1994, NEUROPATH APPL NEURO, V20, P243, DOI 10.1111/j.1365-2990.1994.tb00966.x
   Bessell EM, 1996, J CLIN ONCOL, V14, P945, DOI 10.1200/JCO.1996.14.3.945
   CAMILLERIBROET S, 1995, BLOOD, V86, P432, DOI 10.1182/blood.V86.2.432.bloodjournal862432
   CHANG KL, 1993, MODERN PATHOL, V6, P427
   CRAIG FE, 1993, AM J CLIN PATHOL, V99, P265, DOI 10.1093/ajcp/99.3.265
   DAMORE F, 1991, EUR J CANCER, V27, P1201, DOI 10.1016/0277-5379(91)90081-N
   dAmore F, 1996, BLOOD, V87, P1045, DOI 10.1182/blood.V87.3.1045.bloodjournal8731045
   DEANGELIS LM, 1992, CANCER, V70, P1607, DOI 10.1002/1097-0142(19920915)70:6<1607::AID-CNCR2820700628>3.0.CO;2-J
   EBY NL, 1988, CANCER-AM CANCER SOC, V62, P2461, DOI 10.1002/1097-0142(19881201)62:11<2461::AID-CNCR2820621135>3.0.CO;2-M
   Ferreri AJM, 1995, CANCER TREAT REV, V21, P415, DOI 10.1016/0305-7372(95)90028-4
   GEDDES JF, 1992, J CLIN PATHOL, V45, P587, DOI 10.1136/jcp.45.7.587
   GILLIGAN K, 1990, P NATL ACAD SCI USA, V87, P8790, DOI 10.1073/pnas.87.22.8790
   GLICKMAN JN, 1988, J VIROL, V62, P90
   GULLEY ML, 1992, CANCER, V69, P1600, DOI 10.1002/1097-0142(19920315)69:6+<1600::AID-CNCR2820691316>3.0.CO;2-#
   HALL PA, 1988, J PATHOL, V154, P223, DOI 10.1002/path.1711540305
   HAMILTONDUTOIT SJ, 1993, BLOOD, V82, P619
   HARRIS CC, 1993, NEW ENGL J MED, V329, P1318, DOI 10.1056/NEJM199310283291807
   HENDERSON S, 1991, CELL, V65, P1107, DOI 10.1016/0092-8674(91)90007-L
   Hermine O, 1996, BLOOD, V87, P265, DOI 10.1182/blood.V87.1.265.bloodjournal871265
   Hockenbery D M, 1992, Semin Immunol, V4, P413
   IRONSIDE JW, 1992, CLIN NEUROPATHOL, V11, P210
   JELLINGER KA, 1992, J CANCER RES CLIN, V119, P7, DOI 10.1007/BF01209483
   KARP JE, 1992, LEUKEMIA LYMPHOMA, V8, P167, DOI 10.3109/10428199209054903
   Knutson G., 1995, Modern Pathology, V8, p166A
   KORKOLOPOULOU P, 1994, J CLIN PATHOL, V47, P9, DOI 10.1136/jcp.47.1.9
   Kramer MHH, 1996, J CLIN ONCOL, V14, P2131, DOI 10.1200/JCO.1996.14.7.2131
   KROGHJENSEN M, 1995, LEUKEMIA LYMPHOMA, V19, P223
   LING SM, 1994, CANCER-AM CANCER SOC, V73, P2570, DOI 10.1002/1097-0142(19940515)73:10<2570::AID-CNCR2820731019>3.0.CO;2-1
   MURPHY JK, 1990, J CLIN PATHOL, V43, P220, DOI 10.1136/jcp.43.3.220
   NIEDOBITEK G, 1992, BLOOD, V79, P2520
   ORAZI A, 1993, MODERN PATHOL, V6, P521
   PIRIS MA, 1994, BRIT J CANCER, V69, P337, DOI 10.1038/bjc.1994.61
   PIRIS MA, 1995, LEUKEMIA LYMPHOMA, V17, P35, DOI 10.3109/10428199509051701
   POLLACK IF, 1989, CANCER-AM CANCER SOC, V63, P939, DOI 10.1002/1097-0142(19890301)63:5<939::AID-CNCR2820630526>3.0.CO;2-V
   ROUAH E, 1990, HUM PATHOL, V21, P545, DOI 10.1016/0046-8177(90)90012-T
   ROUBY SE, 1993, BLOOD, V82, P3542
   SANDER CA, 1992, AM J SURG PATHOL, V16, P1170, DOI 10.1097/00000478-199212000-00004
   SHIRAMIZU B, 1991, BLOOD, V77, P1516
   SOINI Y, 1992, J CLIN PATHOL, V45, P1011, DOI 10.1136/jcp.45.11.1011
   SZEKELY L, 1993, P NATL ACAD SCI USA, V90, P5455, DOI 10.1073/pnas.90.12.5455
   TANG SC, 1994, J CLIN ONCOL, V12, P149, DOI 10.1200/JCO.1994.12.1.149
   WEISS LM, 1992, BLOOD, V79, P1789
   Wilson WH, 1997, BLOOD, V89, P601, DOI 10.1182/blood.V89.2.601
NR 43
TC 28
Z9 29
U1 0
U2 0
PU HARWOOD ACAD PUBL GMBH
PI READING
PA C/O STBS LTD, PO BOX 90, READING, BERKS, ENGLAND RG1 8JL
SN 1042-8194
J9 LEUKEMIA LYMPHOMA
JI Leuk. Lymphoma
PD JUN
PY 1998
VL 30
IS 1-2
BP 131
EP +
DI 10.3109/10428199809050936
PG 15
WC Oncology; Hematology
WE Science Citation Index Expanded (SCI-EXPANDED)
SC Oncology; Hematology
GA ZX229
UT WOS:000074493000013
PM 9669683
DA 2024-06-03
ER

PT J
AU Bashir, R
   Chamberlain, M
   Ruby, E
   Hochberg, FH
AF Bashir, R
   Chamberlain, M
   Ruby, E
   Hochberg, FH
TI T-cell infiltration of primary CNS lymphoma
SO NEUROLOGY
LA English
DT Article; Proceedings Paper
CT 47th Annual Meeting of the American-Academy-of-Neurology
CY MAY 11, 1995
CL SEATTLE, WA
SP Amer Acad Neurol
ID NERVOUS-SYSTEM LYMPHOMA; EPSTEIN-BARR-VIRUS; IMMUNE-DEFICIENCY-SYNDROME;
   INSITU HYBRIDIZATION; ADHESION MOLECULE-1; BRAIN; EXPRESSION; AIDS;
   BINDING; DOMAIN
AB We stained 13 primary CNS lymphomas (PCNSLs) (six from patients with AIDS, seven from immunocompetent patients) with a panel of antibodies to T cells (pan T cell [CD3], T helper cell [CD4], T suppressor cell [CD8], delta/Delta cell [CD4(-)8(-)]), B cells (CD20), hematopoietic cells (T200), and NK cell (CD56). We estimated the percentage of tumor cells staining with each antibody. All tumors were B-cell lymphomas. The non-AIDS tumors showed a significant infiltration with CD3(+) cells (mean of 10.82% of total cells). The AIDS patients' tumors showed a smaller percentage of CD3(+) infiltrating cells (mean, 4.88% of total cells) (p < 0.01). CD4(+) cells were 9.11% of the total hematopoietic cells in the non-AIDS patients and 3.13% in AIDS patients (p 0.01). AIDS patients showed some CD8(+) cells (0.3%), which was significantly higher than in immunocompetent patients (0%) (p < 0.05). Very few tumor cells stained with the NK cell and delta Delta cell markers. Both immunocompetent and AIDS patients with PCNSL exhibit significant CD3(+) and CD4(+) cell infiltration of their tumors; this infiltration is significantly lower in AIDS patients. AIDS patients show a minor CD8(+) cell infiltration of their tumors. These results on PCNSL are different from systemic lymphomas, which show a higher CD4 and CD8 cell infiltration, and may offer insights into the more aggressive nature of AIDS-related PCNSL.
C1 UNIV CALIF SAN DIEGO, MED CTR, SAN DIEGO, CA 92103 USA.
   UNIV NEBRASKA, MED CTR, DEPT PREVENT & SOCIETAL MED, OMAHA, NE USA.
   MASSACHUSETTS GEN HOSP, DEPT NEUROL, BOSTON, MA 02114 USA.
C3 University of California System; University of California San Diego;
   University of Nebraska System; University of Nebraska Medical Center;
   Harvard University; Massachusetts General Hospital
RP UNIV NEBRASKA, MED CTR, DIV NEUROL, DEPT INTERNAL MED, 600 S 42ND ST, OMAHA, NE 68198 USA.
CR BASHIR R, 1993, NEUROLOGY, V43, P2358, DOI 10.1212/WNL.43.11.2358
   BASHIR R, 1992, J NEURO-ONCOL, V12, P103
   BASHIR R, 1989, J NEURO-ONCOL, V7, P249
   BASHIR R, 1994, J NEURO-ONCOL, V20, P47, DOI 10.1007/BF01057960
   BASHIR RM, 1989, NEUROLOGY, V39, P813, DOI 10.1212/WNL.39.6.813
   CARLOS TM, 1994, BLOOD, V84, P2068
   COLTON T, 1974, STAT MED, P221
   DEANGELIS LM, 1992, CANCER, V70, P1607, DOI 10.1002/1097-0142(19920915)70:6<1607::AID-CNCR2820700628>3.0.CO;2-J
   DIAMOND MS, 1991, CELL, V65, P961, DOI 10.1016/0092-8674(91)90548-D
   FAUCI AS, 1994, TXB AIDS MED, P55
   FINE HA, 1993, ANN INTERN MED, V119, P1093, DOI 10.7326/0003-4819-119-11-199312010-00007
   FROHMAN EM, 1989, J CLIN IMMUNOL, V9, P1, DOI 10.1007/BF00917121
   GOLDSTEIN GW, 1983, ANN NEUROL, V14, P389, DOI 10.1002/ana.410140402
   HAAS W, 1993, ANNU REV IMMUNOL, V11, P637, DOI 10.1146/annurev.immunol.11.1.637
   HOCHBERG FH, 1988, J NEUROSURG, V68, P835, DOI 10.3171/jns.1988.68.6.0835
   HUYNH HK, 1993, AM J PATHOL, V142, P1265
   KARRE K, 1995, SCIENCE, V267, P978, DOI 10.1126/science.7863341
   KUBY J, 1994, IMMUNOLOGY, P271
   LEVINE AM, 1992, BLOOD, V80, P8
   LIST AF, 1993, LEUKEMIA, V7, P398
   LJUNGGREN HG, 1988, J EXP MED, V167, P730, DOI 10.1084/jem.167.2.730
   MCCUE MP, 1993, NEUROLOGY, V43, P377, DOI 10.1212/WNL.43.2.377
   MEDAWAR PB, 1948, BRIT J EXP PATHOL, V29, P58
   NAKHLEH RE, 1991, CANCER, V67, P444, DOI 10.1002/1097-0142(19910115)67:2<444::AID-CNCR2820670221>3.0.CO;2-W
   OSBORN L, 1992, J EXP MED, V176, P99, DOI 10.1084/jem.176.1.99
   REESE TS, 1967, J CELL BIOL, V34, P207, DOI 10.1083/jcb.34.1.207
   ROSENBERG NL, 1986, ANN NEUROL, V20, P98, DOI 10.1002/ana.410200118
   SASSEVILLE VG, 1992, AM J PATHOL, V141, P1021
   SO YT, 1986, ANN NEUROL, V20, P566, DOI 10.1002/ana.410200503
   WEKERLE H, 1986, TRENDS NEUROSCI, V9, P271, DOI 10.1016/0166-2236(86)90077-9
   WEKERLE H, 1984, LAB INVEST, V51, P199
NR 31
TC 28
Z9 30
U1 0
U2 1
PU LIPPINCOTT WILLIAMS & WILKINS
PI PHILADELPHIA
PA TWO COMMERCE SQ, 2001 MARKET ST, PHILADELPHIA, PA 19103 USA
SN 0028-3878
EI 1526-632X
J9 NEUROLOGY
JI Neurology
PD FEB
PY 1996
VL 46
IS 2
BP 440
EP 444
DI 10.1212/WNL.46.2.440
PG 5
WC Clinical Neurology
WE Science Citation Index Expanded (SCI-EXPANDED); Conference Proceedings Citation Index - Science (CPCI-S)
SC Neurosciences & Neurology
GA TZ712
UT WOS:A1996TZ71200025
PM 8614509
DA 2024-06-03
ER

PT J
AU Cook, MR
   Dorris, CS
   Makambi, KH
   Luo, YT
   Munshi, PN
   Donato, M
   Rowley, S
   Saad, A
   Goy, A
   Dunleavy, K
   Ali, A
AF Cook, Michael R.
   Dorris, C. Scott
   Makambi, Kepher H.
   Luo, Yutong
   Munshi, Pashna N.
   Donato, Michelle
   Rowley, Scott
   Saad, Ayman
   Goy, Andre
   Dunleavy, Kieron
   Ali, Alaa
TI Toxicity and efficacy of CAR T-cell therapy in primary and secondary CNS
   lymphoma: a meta-analysis of 128 patients
SO BLOOD ADVANCES
LA English
DT Review
ID NERVOUS-SYSTEM LYMPHOMA; TISAGENLECLEUCEL; MULTICENTER; RELAPSE
AB Relapsed/refractory primary central nervous system lymphoma (PCNSL) and secondary central nervous system lymphoma (SCNSL) are associated with short survival and represent an unmet need, requiring novel effective strategies. Anti-CD19 chimeric antigen receptor (CAR) T cells, effective in systemic large B-cell lymphoma (LBCL), have shown responses in PCNSL and SCNSL in early reports, but with limited sample size. We, therefore, performed a comprehensive systematic review and meta-analysis of all published data describing CAR T-cell use in PCNSL and SCNSL. This identified 128 patients with PCNSL (30) and SCNSL (98). Our primary objectives were to evaluate CAR T-cell specific toxicity (immune effector cellassociated neurotoxicity syndrome [ICANS] and cytokine release syndrome [CRS]) as well as response rates in these 2 populations. Seventy percent of patients with PCNSL had CRS of any grade (13% grade 3-4) and 53% had ICANS of any grade (18% grade 3-4). Comparatively, 72% of the SCNSL cohort experienced CRS of any grade (11% grade 3-4) and 48% had ICANS of any grade (26% grade 3-4). Of the patients with PCNSL, 56% achieved a complete remission (CR) with 37% remaining in remission at 6 months. Similarly, 47% of patients with SCNSL had a CR, with 37% in remission at 6 months. In a large meta-analysis of central nervous system (CNS) lymphomas, toxicity of anti-CD19-CAR T-cell therapy was similar to that of registrational studies in systemic LBCL with no increased signal of neurotoxicity observed. Encouraging efficacy was demonstrated in patients with CNS lymphoma with no discernible differences between PCNSL and SCNSL.
C1 [Cook, Michael R.; Munshi, Pashna N.; Rowley, Scott; Dunleavy, Kieron; Ali, Alaa] Lombardi Comprehens Canc Ctr, Washington, DC USA.
   [Cook, Michael R.; Munshi, Pashna N.; Rowley, Scott; Dunleavy, Kieron; Ali, Alaa] MedStar Georgetown Univ Hosp, Washington, DC USA.
   [Dorris, C. Scott] Georgetown Univ, Dahlgren Mem Lib, Washington, DC USA.
   [Makambi, Kepher H.; Luo, Yutong] Georgetown Univ, Dept Biostat Bioinformat & Biomath, Washington, DC USA.
   [Donato, Michelle; Goy, Andre] Hackensack Univ, John Theurer Canc Ctr, Med Ctr, Hackensack, NJ USA.
   [Saad, Ayman] Ohio State Univ, Div Hematol, Columbus, OH USA.
C3 Georgetown University; Georgetown University; Georgetown University;
   Hackensack University Medical Center; University System of Ohio; Ohio
   State University
RP Ali, A (corresponding author), Georgetown Univ, Stem Cell Transplant & Cellular Immunotherapy Pro, 3800 Reservoir Rd NW, Washington, DC 20007 USA.
EM
RI Saad, Ayman/JOZ-2117-2023; Dorris, C. Scott/J-8932-2018
OI Saad, Ayman/0000-0003-0003-0130; Cook, Michael/0000-0002-6171-9647; Ali,
   Alaa/0000-0003-1813-515X; Dorris, C. Scott/0000-0002-4664-6793; Rowley,
   Scott/0000-0002-3299-835X; Goy, Andre/0000-0001-5125-6522
CR Abbasi A, 2020, J HEMATOL ONCOL, V13, DOI 10.1186/s13045-019-0838-y
   Abramson JS, 2020, LANCET, V396, P839, DOI 10.1016/S0140-6736(20)31366-0
   Abramson JS, 2017, NEW ENGL J MED, V377, P783, DOI 10.1056/NEJMc1704610
   Ahmed G, 2021, BLOOD ADV, V5, P5626, DOI 10.1182/bloodadvances.2021005292
   Alcantara M, 2022, BLOOD, V139, P792, DOI 10.1182/blood.2021012932
   Bennani NN, 2019, BLOOD, V134, DOI 10.1182/blood-2019-129097
   Bishop MR, 2019, BLOOD ADV, V3, P2230, DOI 10.1182/bloodadvances.2019000151
   Chihara D, 2021, CL LYMPH MYELOM LEUK, V21, P73, DOI 10.1016/j.clml.2020.10.015
   DERSIMONIAN R, 1986, CONTROL CLIN TRIALS, V7, P177, DOI 10.1016/0197-2456(86)90046-2
   Foundation TR, R: a language and environment for statistical computing
   Frigault MJ, 2022, BLOOD, V139, P2306, DOI 10.1182/blood.2021014738
   Frigault MJ, 2019, BLOOD, V134, P860, DOI 10.1182/blood.2019001694
   Ghafouri S, 2021, BONE MARROW TRANSPL, V56, P974, DOI 10.1038/s41409-020-01099-4
   Green K, 2022, Central nervous system lymphoma
   Grupp SA, 2013, NEW ENGL J MED, V368, P1509, DOI 10.1056/NEJMoa1215134
   Higgins JPT, 2002, STAT MED, V21, P1539, DOI 10.1002/sim.1186
   Holtzman NG, 2021, NEURO-ONCOLOGY, V23, P112, DOI 10.1093/neuonc/noaa183
   Jahnke K, 2006, J NEURO-ONCOL, V80, P159, DOI 10.1007/s11060-006-9165-6
   Jin LC, 2019, NAT COMMUN, V10, DOI 10.1038/s41467-019-11869-4
   Karschnia P, 2022, NEUROLOGY, V98, P884, DOI 10.1212/WNL.0000000000200608
   Keu KV, 2017, SCI TRANSL MED, V9, DOI 10.1126/scitranslmed.aag2196
   Li TJ, 2020, FRONT ONCOL, V10, DOI 10.3389/fonc.2020.01306
   Liu R, 2022, HUM GENE THER, V33, P318, DOI 10.1089/hum.2021.249
   Locke FL, 2019, LANCET ONCOL, V20, P31, DOI 10.1016/S1470-2045(18)30864-7
   Nastoupil LJ, 2020, J CLIN ONCOL, V38, P3119, DOI 10.1200/JCO.19.02104
   Neelapu SS, 2017, NEW ENGL J MED, V377, P2531, DOI 10.1056/NEJMoa1707447
   Ouzzani M, 2016, SYST REV-LONDON, V5, DOI 10.1186/s13643-016-0384-4
   Page MJ, 2021, BMJ-BRIT MED J, V372, DOI [10.1136/bmj.n71, 10.1016/j.ijsu.2021.105906, 10.1136/bmj.n160]
   Parker KR, 2020, CELL, V183, P126, DOI 10.1016/j.cell.2020.08.022
   Roddie C, 2022, RELAPSEDREFRACTORY P, P1460
   Sanderson R, 2021, BLOOD, V138, DOI 10.1182/blood-2021-149501VVV
   Schuster SJ, 2021, LANCET ONCOL, V22, P1403, DOI 10.1016/S1470-2045(21)00375-2
   Schuster SJ, 2019, NEW ENGL J MED, V380, P45, DOI 10.1056/NEJMoa1804980
   Siddiqi T, 2021, BLOOD ADV, V5, P4059, DOI 10.1182/bloodadvances.2020004106
   Wu JY, 2021, BLOOD CANCER J, V11, DOI 10.1038/s41408-021-00523-2
   Zinzani PL, 1999, LEUKEMIA LYMPHOMA, V32, P571, DOI 10.3109/10428199909058415
NR 36
TC 27
Z9 27
U1 5
U2 11
PU ELSEVIER
PI AMSTERDAM
PA RADARWEG 29, 1043 NX AMSTERDAM, NETHERLANDS
SN 2473-9529
EI 2473-9537
J9 BLOOD ADV
JI Blood Adv.
PD JAN 10
PY 2023
VL 7
IS 1
BP 32
EP 39
DI 10.1182/bloodadvances.2022008525
EA JAN 2023
PG 8
WC Hematology
WE Science Citation Index Expanded (SCI-EXPANDED)
SC Hematology
GA 8F4SO
UT WOS:000919655200001
PM 36260735
OA gold, Green Published
HC Y
HP N
DA 2024-06-03
ER

PT J
AU Takashima, Y
   Hayano, A
   Yamanaka, R
AF Takashima, Yasuo
   Hayano, Azusa
   Yamanaka, Ryuya
TI Metabolome Analysis Reveals Excessive Glycolysis via PI3K/AKT/mTOR and
   RAS/MAPK Signaling in Methotrexate-Resistant Primary CNS
   Lymphoma-Derived Cells
SO CLINICAL CANCER RESEARCH
LA English
DT Article
ID CENTRAL-NERVOUS-SYSTEM; EXPRESSION; PLASMA; TUMORS; LINES
AB Purpose: Metabolome analysis is an emerging method that provides insight into intracellular and physiologic responses. Methotrexate (MTX) is an antifolate that suppresses DNA syntheses by inhibiting dihydrofolate reductase. High-dose methotrexate treatment with deferred radiotherapy is a standard protocol in primary central nervous system lymphoma (PCNSL) treatments. However, most cases come to relapse-acquired resistance, in which the role of metabolic pathways is largely unknown.
   Experimental Design: Metabolome analysis in methotrexate-resistant PCNSL-derived cells (designated as TK-MTX and HKBML-MTX) was performed to detect alternative metabolites and pathways.
   Results: The metabolomic analyses using capillary electrophoresis-time-of-flight mass spectrometry detected 188 and 169 peaks in TK- and HKBML-derived cells, respectively, including suppression of central carbon metabolism, lipid metabolism, nucleic acid metabolism, urea cycle, branched chain and aromatic amino acids, and coenzyme metabolism. Particularly, whole suppressive metabolic pathways were demonstrated in TK-MTX, whereas HKBML-MTX indicated partially enhanced pathways of the urea cycle, amino acid metabolism, and coenzyme metabolism. Reciprocally detected metabolites for glycolysis, including induced glucose and reduced glycogen, and induced lactate and reduced pyruvate, in addition to increased lactate dehydrogenase activity, which is involved in Warburg effect. Thereby, ATP was increased in both methotrexate-resistant PCNSL-derived cells. Furthermore, we specifically found that PI3K/AKT/mTOR and RAS/MAPK signaling pathways were activated in TK-MTX but not in HKBML-MTX by growth rate with inhibitors and gene expression analysis, suggestive of cell type-specific methotrexate-resistant metabolic pathways.
   Conclusions: These results can help us understand targeted therapies with selective anticancer drugs in recurrent CNS lymphoma-acquired resistance against methotrexate.
C1 [Takashima, Yasuo; Hayano, Azusa; Yamanaka, Ryuya] Kyoto Prefectural Univ Med, Grad Sch Med Sci, Lab Mol Target Therapy Canc, Kyoto, Japan.
C3 Kyoto Prefectural University of Medicine
RP Yamanaka, R (corresponding author), Kyoto Prefectural Univ Med, Kamigyo Ku, 465 Kajii Cho, Kyoto 6028566, Japan.
EM
RI Yamanaka, Ryuya/AFL-0782-2022
OI Yamanaka, Ryuya/0000-0003-2941-7996; Takashima,
   Yasuo/0000-0001-8876-6673
FU JSPS KAKENHI [16H05441, 18K09001]; Grants-in-Aid for Scientific Research
   [18K09001, 16H05441] Funding Source: KAKEN
FX This work was supported by JSPS KAKENHI (grant numbers 16H05441, to R.
   Yamanaka; 18K09001, to Y. Takashima). The authors thank Dr. Junya Fukai
   (Wakayama Medical University), Dr. Yasuo Iwadate (Chiba University), Dr.
   Koji Kajiwara (Yamaguchi University), and Dr. Hiroaki Hondoh (Toyama
   Prefectural Central Hospital) for providing PCNSL samples and clinical
   data.
CR Cairns RA, 2011, NAT REV CANCER, V11, P85, DOI 10.1038/nrc2981
   CHABNER BA, 1985, J CLIN INVEST, V76, P907, DOI 10.1172/JCI112088
   Ducker GS, 2017, P NATL ACAD SCI USA, V114, P11404, DOI 10.1073/pnas.1706617114
   Garcia-Manteiga JM, 2011, J PROTEOME RES, V10, P4165, DOI 10.1021/pr200328f
   Goecke IA, 2007, MOL IMMUNOL, V44, P2115, DOI 10.1016/j.molimm.2006.07.303
   Hayano A, 2019, INT J CLIN ONCOL, V24, P1020, DOI 10.1007/s10147-019-01451-9
   Hu J, 2013, NAT BIOTECHNOL, V31, P522, DOI 10.1038/nbt.2530
   Iwadate Y, 2014, J NEURO-ONCOL, V117, P261, DOI 10.1007/s11060-014-1379-4
   Jiang GT, 2018, J CHROMATOGR B, V1077, P71, DOI 10.1016/j.jchromb.2018.01.034
   Kasenda B, 2015, ANN ONCOL, V26, P1305, DOI 10.1093/annonc/mdv076
   Kawaguchi A, 2012, CLIN CANCER RES, V18, P5672, DOI 10.1158/1078-0432.CCR-12-0596
   Koppenol WH, 2011, NAT REV CANCER, V11, P325, DOI 10.1038/nrc3038
   Le YC, 2018, J MASS SPECTROM, V53, P172, DOI 10.1002/jms.4048
   Louis D.N., 2016, WHO CLASSIFICATION T, V1
   Louis DN, 2016, ACTA NEUROPATHOL, V131, P803, DOI 10.1007/s00401-016-1545-1
   McDonnell SRP, 2013, BLOOD, V122, P958, DOI 10.1182/blood-2013-01-482026
   Metallo CM, 2012, NATURE, V481, P380, DOI 10.1038/nature10602
   Pera B, 2018, EBIOMEDICINE, V28, P80, DOI 10.1016/j.ebiom.2018.01.014
   Reni M, 1999, BRIT J CANCER, V79, P530, DOI 10.1038/sj.bjc.6690083
   Ricard D, 2012, LANCET, V379, P1984, DOI 10.1016/S0140-6736(11)61346-9
   Rushworth D, 2015, J BIOL CHEM, V290, P22970, DOI 10.1074/jbc.C115.671123
   Schwarzfischer P, 2017, J PROTEOME RES, V16, P1105, DOI 10.1021/acs.jproteome.6b00164
   Shah HB, 2015, METHODS MOL BIOL, V1312, P419, DOI 10.1007/978-1-4939-2694-7_42
   Stenson M, 2016, LEUKEMIA LYMPHOMA, V57, P1814, DOI 10.3109/10428194.2016.1140164
   Takashima Y, 2019, BMC CANCER, V19, DOI 10.1186/s12885-019-6129-8
   Takashima Y, 2019, SCI REP-UK, V9, DOI 10.1038/s41598-019-46473-5
   Takashima Y, 2018, CANCER SCI, V109, P3543, DOI 10.1111/cas.13798
   Takashima Yasuo, 2018, Oncotarget, V9, P27471, DOI 10.18632/oncotarget.25463
   Takashima Yasuo, 2018, Oncotarget, V9, P19065, DOI 10.18632/oncotarget.24897
   Takashima Y, 2016, HEPATOLOGY, V64, P245, DOI 10.1002/hep.28548
   Vander Heiden MG, 2013, J CLIN INVEST, V123, P3648, DOI 10.1172/JCI72391
   WARBURG O, 1956, SCIENCE, V123, P309, DOI 10.1126/science.123.3191.309
   Ward PS, 2012, CANCER CELL, V21, P297, DOI 10.1016/j.ccr.2012.02.014
   Yamanaka R., 2016, PRIMARY CENTRAL NERV, P175
   Yang FM, 2017, PLOS ONE, V12, DOI 10.1371/journal.pone.0170896
   Yoo BC, 2010, BMC CANCER, V10, DOI 10.1186/1471-2407-10-55
   Yun JY, 2009, SCIENCE, V325, P1555, DOI 10.1126/science.1174229
   Zhang Jianqing, 2016, Oncoscience, V3, P31
   Zhou QY, 2014, ASIAN PAC J CANCER P, V15, P5417, DOI 10.7314/APJCP.2014.15.13.5417
NR 39
TC 27
Z9 27
U1 4
U2 20
PU AMER ASSOC CANCER RESEARCH
PI PHILADELPHIA
PA 615 CHESTNUT ST, 17TH FLOOR, PHILADELPHIA, PA 19106-4404 USA
SN 1078-0432
EI 1557-3265
J9 CLIN CANCER RES
JI Clin. Cancer Res.
PD JUN 1
PY 2020
VL 26
IS 11
BP 2754
EP 2766
DI 10.1158/1078-0432.CCR-18-3851
PG 13
WC Oncology
WE Science Citation Index Expanded (SCI-EXPANDED)
SC Oncology
GA LU6GQ
UT WOS:000537852100034
PM 32108030
DA 2024-06-03
ER

PT J
AU Cho, I
   Lee, H
   Yoon, SE
   Ryu, KJ
   Ko, YH
   Kim, WS
   Kim, SJ
AF Cho, Inju
   Lee, Hansang
   Yoon, Sang Eun
   Ryu, Kyung Ju
   Ko, Young Hyeh
   Kim, Won Seog
   Kim, Seok Jin
TI Serum levels of soluble programmed death-ligand 1 (sPD-L1) in patients
   with primary central nervous system diffuse large B-cell lymphoma
SO BMC CANCER
LA English
DT Article
DE Soluble PD-L1; PD-1; Primary central nervous system lymphoma
ID PRIMARY CNS LYMPHOMA; HIGH-DOSE METHOTREXATE; IMMUNOCOMPETENT PATIENTS;
   PD-L1 IMMUNOHISTOCHEMISTRY; PDL1 EXPRESSION; SURVIVAL; CHEMOTHERAPY;
   RADIOTHERAPY; ASSOCIATION; RADIATION
AB Background The interaction of programmed death-1 protein (PD-1) and programmed death-1 ligand (PD-L1) produces immunosuppressive activity, protecting tumor cells from anti-tumor immunity and possibly releasing soluble PD-L1 (sPD-L1) from PD-L1 expressing tumor cells. Therefore, we measured serum levels of sPD-L1 in patients with primary central nervous system lymphoma (PCNSL) and explored its clinical implications. Methods Sixty-eight patients with newly diagnosed PCNSL had diffuse large B-cell lymphoma and were treated with high-dose methotrexate-containing chemotherapy. The measurement of sPD-L1 and cytokines was performed using serum samples archived at diagnosis, and the tissue expression of PD-L1 was also analyzed from archived paraffin-embedded tissue blocks. Disease relapse, progression-free survival (PFS), and overall survival (OS) were analyzed according to the extent of sPD-L1 in serum and PD-L1 in tissue. Results The median level of serum sPD-L1 (0.429 ng/mL) was higher than in healthy control patients (0.364 ng/mL). The occurrence of relapse was more frequent in the high sPD-L1 (78%) than the low sPD-L1 group (50%), though the groups did not have different clinical or pathological characteristics at diagnosis. As a result, the OS and PFS for the high sPD-L1 group were significantly lower than those in the low group. PD-L1-positive tumor cells were found in 35 patients (67%), and the extent of PD-L1-postive tumor cells was positively associated with serum sPD-L1 levels (r = 0.299, P = 0.031). Among the 34 cytokines analyzed, only the serum level of IL-7 correlated with the serum level of sPD-L1 (r = 0.521, P < 0.001). Conclusions Serum levels of sPD-L1 could reflect the expression of PD-L1 in PCNSL tumor cells and predict patient survival outcomes. Therefore, sPD-L1 in serum could be a feasible biomarker for determining a risk-adapted treatment strategy for PCNSL patients.
C1 [Cho, Inju] Catholic Univ Korea, Yeouido St Marys Hosp, Dept Pathol, Seoul, South Korea.
   [Lee, Hansang] Dankook Univ, Dept Internal Med, Coll Med, Cheonan, South Korea.
   [Yoon, Sang Eun; Kim, Won Seog; Kim, Seok Jin] Sungkyunkwan Univ, Samsung Med Ctr, Dept Med, Div Hematol & Oncol,Sch Med, 81 Irwon Ro, Seoul 06351, South Korea.
   [Ryu, Kyung Ju; Kim, Won Seog; Kim, Seok Jin] Sungkyunkwan Univ, Samsung Adv Inst Hlth Sci & Technol, Dept Hlth Sci & Technol, Seoul, South Korea.
   [Ko, Young Hyeh] Sungkyunkwan Univ, Samsung Med Ctr, Dept Pathol & Translat Genom, Coll Med, Seoul, South Korea.
C3 Catholic University of Korea; Dankook University; Sungkyunkwan
   University (SKKU); Samsung Medical Center; Sungkyunkwan University
   (SKKU); Samsung Medical Center; Sungkyunkwan University (SKKU); Samsung
   Medical Center
RP Kim, SJ (corresponding author), Sungkyunkwan Univ, Samsung Med Ctr, Dept Med, Div Hematol & Oncol,Sch Med, 81 Irwon Ro, Seoul 06351, South Korea.; Kim, SJ (corresponding author), Sungkyunkwan Univ, Samsung Adv Inst Hlth Sci & Technol, Dept Hlth Sci & Technol, Seoul, South Korea.
EM
FU Basic Science Research Program through the National Research Foundation
   of Korea (NRF) - Ministry of Education, Science, and Technology
   [NRF-2017R1A2B4005136]
FX This study was supported by a grant from the Basic Science Research
   Program through the National Research Foundation of Korea (NRF), which
   is funded by the Ministry of Education, Science, and Technology
   (NRF-2017R1A2B4005136). The experiments of this study including
   immunohistochemistry and cytokine measurements were funded by the
   aforementioned grant. However, there was no role of the funding body in
   the design of the study and collection, analysis, and interpretation of
   data and in writing the manuscript.
CR Abrey LE, 2005, J CLIN ONCOL, V23, P5034, DOI 10.1200/JCO.2005.13.524
   Berghoff AS, 2014, CLIN NEUROPATHOL, V33, P42, DOI 10.5414/NP300698
   Camilleri-Broet S, 1998, AM J CLIN PATHOL, V110, P607, DOI 10.1093/ajcp/110.5.607
   Chapuy B, 2016, BLOOD, V127, P869, DOI 10.1182/blood-2015-10-673236
   Chen YJ, 2011, CYTOKINE, V56, P231, DOI 10.1016/j.cyto.2011.06.004
   Cho H, 2017, ONCOTARGET, V8, P87317, DOI 10.18632/oncotarget.20264
   Choi MK, 2013, INT J HEMATOL, V98, P346, DOI 10.1007/s12185-013-1403-z
   da Silva PB, 2018, HEMATOL ONCOL, V36, P709, DOI 10.1002/hon.2542
   DeAngelis LM, 2002, J CLIN ONCOL, V20, P4643, DOI 10.1200/JCO.2002.11.013
   Ding Y, 2017, SCAND J IMMUNOL, V86, P361, DOI 10.1111/sji.12596
   Durum SK, 1998, J EXP MED, V188, P2233, DOI 10.1084/jem.188.12.2233
   Ferreri AJM, 2009, LANCET, V374, P1512, DOI 10.1016/S0140-6736(09)61416-1
   Four M, 2017, HEMATOL ONCOL, V35, P487, DOI 10.1002/hon.2375
   Frigola X, 2011, CLIN CANCER RES, V17, P1915, DOI 10.1158/1078-0432.CCR-10-0250
   Gerstner ER, 2010, ARCH NEUROL-CHICAGO, V67, P291, DOI 10.1001/archneurol.2010.3
   Grommes C, 2017, J CLIN ONCOL, V35, P2410, DOI 10.1200/JCO.2017.72.7602
   Ha H, 2016, ONCOTARGET, V7, P76604, DOI 10.18632/oncotarget.12810
   Hayano A, 2017, ANTICANCER RES, V37, P5655, DOI 10.21873/anticanres.12001
   Hoang-Xuan K, 2015, LANCET ONCOL, V16, pE322, DOI 10.1016/S1470-2045(15)00076-5
   Hong JY, 2017, HEMATOL ONCOL, V35, P480, DOI 10.1002/hon.2374
   Huang SY, 2016, ONCOTARGET, V7, P62490, DOI 10.18632/oncotarget.11519
   Hude I, 2017, HAEMATOLOGICA, V102, P30, DOI 10.3324/haematol.2016.150656
   Kim Jeong Eun, 2012, Korean J Hematol, V47, P60, DOI 10.5045/kjh.2012.47.1.60
   Kim SJ, 2015, J HEMATOL ONCOL, V8, DOI 10.1186/s13045-015-0142-4
   Kinter AL, 2008, J IMMUNOL, V181, P6738, DOI 10.4049/jimmunol.181.10.6738
   Kiyasu J, 2015, BLOOD, V126, P2193, DOI 10.1182/blood-2015-02-629600
   Korfel A, 2013, NAT REV NEUROL, V9, P317, DOI 10.1038/nrneurol.2013.83
   Langner-Lemercier S, 2016, NEURO-ONCOLOGY, V18, P1297, DOI 10.1093/neuonc/now033
   Lipson EJ, 2013, CANCER IMMUNOL RES, V1, P54, DOI 10.1158/2326-6066.CIR-13-0034
   Menter T, 2016, HUM PATHOL, V54, P17, DOI 10.1016/j.humpath.2016.03.005
   Juárez-Salcedo LM, 2017, CRIT REV ONCOL HEMAT, V113, P52, DOI 10.1016/j.critrevonc.2017.02.027
   Nayak L, 2017, BLOOD, V129, P3071, DOI 10.1182/blood-2017-01-764209
   Nguyen PL, 2005, J CLIN ONCOL, V23, P1507, DOI 10.1200/JCO.2005.01.161
   Okuma Y, 2017, LUNG CANCER, V104, P1, DOI 10.1016/j.lungcan.2016.11.023
   Phillips T, 2015, APPL IMMUNOHISTO M M, V23, P541, DOI 10.1097/PAI.0000000000000256
   Poortmans PMP, 2003, J CLIN ONCOL, V21, P4483, DOI 10.1200/JCO.2003.03.108
   Rossille D, 2017, LEUKEMIA, V31, P988, DOI 10.1038/leu.2016.385
   Scheel AH, 2016, MODERN PATHOL, V29, P1165, DOI 10.1038/modpathol.2016.117
   Soussain C, 2008, J CLIN ONCOL, V26, P2512, DOI 10.1200/JCO.2007.13.5533
   Sugita Y, 2018, NEUROPATHOLOGY, V38, P125, DOI 10.1111/neup.12435
   Swerdlow S. H, 2017, WHO CLASSIFICATION T
   Thiel E, 2010, LANCET ONCOL, V11, P1036, DOI 10.1016/S1470-2045(10)70229-1
   Wang H, 2016, ONCOTARGET, V7, P33035, DOI 10.18632/oncotarget.8847
   Wang L, 2015, ONCOTARGET, V6, P41228, DOI 10.18632/oncotarget.5682
   Yi JH, 2019, CYTOKINE, V113, P371, DOI 10.1016/j.cyto.2018.10.009
   Zou WP, 2008, NAT REV IMMUNOL, V8, P467, DOI 10.1038/nri2326
NR 46
TC 27
Z9 27
U1 0
U2 6
PU BMC
PI LONDON
PA CAMPUS, 4 CRINAN ST, LONDON N1 9XW, ENGLAND
EI 1471-2407
J9 BMC CANCER
JI BMC Cancer
PD FEB 13
PY 2020
VL 20
IS 1
AR 120
DI 10.1186/s12885-020-6612-2
PG 11
WC Oncology
WE Science Citation Index Expanded (SCI-EXPANDED)
SC Oncology
GA KO6DL
UT WOS:000515638800001
PM 32054467
OA Green Published, gold, Green Submitted
DA 2024-06-03
ER

PT J
AU Watanabe, J
   Natsumeda, M
   Okada, M
   Kobayashi, D
   Kanemaru, Y
   Tsukamoto, Y
   Oishi, M
   Kakita, A
   Fujii, Y
AF Watanabe, Jun
   Natsumeda, Manabu
   Okada, Masayasu
   Kobayashi, Daiki
   Kanemaru, Yu
   Tsukamoto, Yoshihiro
   Oishi, Makoto
   Kakita, Akiyoshi
   Fujii, Yukihiko
TI High Detection Rate of <i>MYD88</i> Mutations in Cerebrospinal Fluid
   From Patients With CNS Lymphomas
SO JCO PRECISION ONCOLOGY
LA English
DT Article
ID CENTRAL-NERVOUS-SYSTEM; B-CELL LYMPHOMA; FETAL DNA; MATERNAL PLASMA;
   IBRUTINIB; METHYLATION; CLEARANCE; DISEASE; TUMORS; BRAIN
AB PURPOSE Biopsy is the gold standard for the diagnosis of primary CNS lymphoma (PCNSL). However, surgical biopsy has problems of morbidity related to hemorrhagic complications and false-negative findings, so safer and more reliable diagnostic methods are required. The aim of this study is to detect the MYD88 mutation, an important driver mutation, in the cerebrospinal fluid (CSF) of patients with CNS lymphoma.
   PATIENTS AND METHODS Twenty-six patients with CNS lymphoma (20 primary CNS lymphoma and six CNS relapse from systemic lymphoma) were studied. We extracted cell-free DNA (cfDNA) from CSF by lumbar puncture. cfDNA was extracted from 1 mL of CSF, and Sanger sequencing and droplet digital polymerase chain reaction (ddPCR) were performed. Furthermore, we performed DNA sequencing of MYD88 in 21 cases with available surgically obtained formalin-fixed paraffin-embedded (FFPE) tissue and compared the results.
   RESULTS The median cfDNA amount extracted from 1 mL CSF was 219 ng/mL (25th to 75th percentile, 129 to 333 ng/mL). MYD88 mutations were detected from CSF in 76.9% (20 of 26 cases), and L265P in exon 5 was the most frequent mutation in 19 out of 20 (95.0%) cases. S219C in exon 3 was detected in one case. In four patients, MYD88 mutation was confirmed by ddPCR but not by Sanger sequencing. In all 21 cases with sufficient FFPE tissue for DNA analysis, the detection of MYD88 mutation from cfDNA was consistent with those of tumor-derived DNA from FFPE tissue.
   CONCLUSION This pilot study provided evidence that the somatic driver mutation MYD88 can be reliably detected by combination of Sanger sequencing and ddPCR in the cfDNA taken from 1 mL of CSF in patients with CNS lymphomas. (C) 2019 by American Society of Clinical Oncology
C1 [Watanabe, Jun; Natsumeda, Manabu; Okada, Masayasu; Kobayashi, Daiki; Kanemaru, Yu; Tsukamoto, Yoshihiro; Oishi, Makoto; Kakita, Akiyoshi; Fujii, Yukihiko] Niigata Univ, Niigata, Japan.
C3 Niigata University
RP Natsumeda, M (corresponding author), Niigata Univ, Brain Res Inst, Dept Neurosurg, Chuo Ku, 1-757 Asahimachi, Niigata 9518585, Japan.
EM
RI Natsumeda, Manabu/Z-2072-2019; Okada, MASAYASU/HJI-9434-2023
OI Natsumeda, Manabu/0000-0003-3098-8323; 
FU Japan Society for Promotion of Science [17K16632, 17K17735, 17K17739];
   2016 Niigata University Interdisciplinary Research (U-go) grant;
   Grants-in-Aid for Scientific Research [17K17735, 17K16632, 17K17739]
   Funding Source: KAKEN
FX Supported in part by Japan Society for Promotion of Science Grants No.
   17K16632 (M.N.), 17K17735 (Y.T.), and 17K17739 (M.O.); and 2016 Niigata
   University Interdisciplinary Research (U-go) grant (J.W. and M.N.).
CR Abrey LE, 2005, J CLIN ONCOL, V23, P5034, DOI 10.1200/JCO.2005.13.524
   Aoki H, 2013, NEUROL MED-CHIR, V53, P797, DOI 10.2176/nmc.oa2013-0195
   Behin A, 2003, LANCET, V361, P323, DOI 10.1016/S0140-6736(03)12328-8
   Braggio E, 2015, CLIN CANCER RES, V21, P3986, DOI 10.1158/1078-0432.CCR-14-2116
   Bruno A, 2014, ONCOTARGET, V5, P5065, DOI 10.18632/oncotarget.2080
   Citterio G, 2017, CRIT REV ONCOL HEMAT, V113, P97, DOI 10.1016/j.critrevonc.2017.03.019
   De Mattos-Arruda L, 2015, NAT COMMUN, V6, DOI 10.1038/ncomms9839
   Fernando MR, 2010, PRENATAL DIAG, V30, P418, DOI 10.1002/pd.2484
   Fontanilles M, 2017, ONCOTARGET, V8, P48157, DOI 10.18632/oncotarget.18325
   Genovese G, 2014, NEW ENGL J MED, V371, P2477, DOI 10.1056/NEJMoa1409405
   Gonzalez-Aguilar A, 2012, CLIN CANCER RES, V18, P5203, DOI 10.1158/1078-0432.CCR-12-0845
   Grommes C, 2017, CANCER DISCOV, V7, P1018, DOI 10.1158/2159-8290.CD-17-0613
   Hans CP, 2004, BLOOD, V103, P275, DOI 10.1182/blood-2003-05-1545
   Hattori K, 2018, CANCER SCI, V109, P225, DOI 10.1111/cas.13450
   Hattori K, 2017, BRIT J HAEMATOL, V177, P492, DOI 10.1111/bjh.14080
   Hiemcke-Jiwa LS, 2018, HEMATOL ONCOL, V36, P429, DOI 10.1002/hon.2489
   Kreher S, 2015, NEURO-ONCOLOGY, V17, P1016, DOI 10.1093/neuonc/nov046
   Lionakis MS, 2017, CANCER CELL, V31, P833, DOI 10.1016/j.ccell.2017.04.012
   Lo YMD, 1999, AM J HUM GENET, V64, P218, DOI 10.1086/302205
   MILLER DC, 1994, CANCER-AM CANCER SOC, V74, P1383, DOI 10.1002/1097-0142(19940815)74:4<1383::AID-CNCR2820740432>3.0.CO;2-1
   Nakamura T, 2016, NEUROPATH APPL NEURO, V42, P279, DOI 10.1111/nan.12259
   Nakamura T, 2017, ACTA NEUROPATHOL, V133, P321, DOI 10.1007/s00401-016-1664-8
   Natsumeda M, 2011, NEUROPATHOLOGY, V31, P486, DOI 10.1111/j.1440-1789.2010.01197.x
   Ngo VN, 2011, NATURE, V470, P115, DOI 10.1038/nature09671
   Rossi D, 2017, BLOOD, V129, P1947, DOI 10.1182/blood-2016-05-719641
   Scherer F, 2016, SCI TRANSL MED, V8, DOI 10.1126/scitranslmed.aai8545
   Schroers R, 2010, EUR J HAEMATOL, V85, P520, DOI 10.1111/j.1600-0609.2010.01516.x
   Schroers R, 2010, EUR J HAEMATOL, V85, P236, DOI 10.1111/j.1600-0609.2010.01475.x
   Schwarzenbach H, 2011, NAT REV CANCER, V11, P426, DOI 10.1038/nrc3066
   Sorber L, 2017, J MOL DIAGN, V19, P162, DOI 10.1016/j.jmoldx.2016.09.009
   Takano S, 2018, WORLD NEUROSURG, V112, pE69, DOI 10.1016/j.wneu.2017.12.028
   Wang YX, 2015, P NATL ACAD SCI USA, V112, P9704, DOI 10.1073/pnas.1511694112
   Wilson WH, 2015, NAT MED, V21, P922, DOI 10.1038/nm.3884
   Yamada S, 2015, LEUKEMIA LYMPHOMA, V56, P2141, DOI 10.3109/10428194.2014.979413
   Yu SCY, 2013, CLIN CHEM, V59, P1228, DOI 10.1373/clinchem.2013.203679
   Yu XF, 2018, CANCER RES, V78, P2457, DOI 10.1158/0008-5472.CAN-18-0215
   Zheng M, 2017, NEUROPATHOLOGY, V37, P509, DOI 10.1111/neup.12405
   Zorofchian S, 2018, FRONT ONCOL, V8, DOI 10.3389/fonc.2018.00382
NR 38
TC 27
Z9 27
U1 3
U2 3
PU AMER SOC CLINICAL ONCOLOGY
PI ALEXANDRIA
PA 2318 MILL ROAD, STE 800, ALEXANDRIA, VA 22314 USA
EI 2473-4284
J9 JCO PRECIS ONCOL
JI JCO Precis. Oncol.
PD APR 18
PY 2019
VL 3
DI 10.1200/PO.18.00308
PG 13
WC Oncology
WE Science Citation Index Expanded (SCI-EXPANDED)
SC Oncology
GA JI8WX
UT WOS:000493745100001
PM 35100686
DA 2024-06-03
ER

PT J
AU Mei, SH
   Li, XG
   Jiang, XY
   Yu, KF
   Lin, S
   Zhao, ZG
AF Mei, Shenghui
   Li, Xingang
   Jiang, Xueyun
   Yu, Kefu
   Lin, Song
   Zhao, Zhigang
TI Population Pharmacokinetics of High-Dose Methotrexate in Patients With
   Primary Central Nervous System Lymphoma
SO JOURNAL OF PHARMACEUTICAL SCIENCES
LA English
DT Article
DE methotrexate; primary central nervous system lymphoma; population
   pharmacokinetic model; nonlinear mixed-effects modeling; serum
   creatinine; body surface area; age; personalized medicine
ID ACUTE LYMPHOBLASTIC-LEUKEMIA; RHEUMATOID-ARTHRITIS; PEDIATRIC-PATIENTS;
   OSTEOSARCOMA; MODEL; LEVETIRACETAM; DETERMINANTS; LIPOPROTEINS;
   MALIGNANCIES; POLYMORPHISM
AB The intra- and inter-individual variances of methotrexate (MTX) pharmacokinetics are extremely large, and the pharmacokinetic property of MTX in patients with primary central nervous system lymphoma (PCNSL) is unestablished. A total of 701 MTX plasma concentrations from 98 patients with PCNSL under high-dose MTX therapy were used to develop the population pharmacokinetic (popPK) model of MTX by using the nonlinear mixed-effects modeling method. A 2-compartment model was employed to describe the pharmacokinetic property of MTX. In the final popPK model, inclusion of serum creatinine and body surface area significantly reduced objective function value for clearance over the base model (p < 0.001), and inclusion of age significantly reduced objective function value for distribution volume of central compartment (V-c) over the base model (p < 0.001). In the final popPK model, the inter-individual clearance = 6.67 x (SCR/68.1)(-0.48) x (BSA/1.75)(1.17); V-c = 24.46 x (age/57.16)(0.81). The precision of all parameters was acceptable (relative standard error < 28.61%). Bootstrap and visual predictive check results indicated that the final popPK model was stable with acceptable predictive ability. The popPK model may be useful for personalized medication in PCNSL patients under high-dose MTX therapy. Further studies are warranted to confirm the results. (C) 2018 American Pharmacists Association (R). Published by Elsevier Inc. All rights reserved.
C1 [Mei, Shenghui; Li, Xingang; Yu, Kefu; Zhao, Zhigang] Capital Med Univ, Beijing Tiantan Hosp, Dept Pharm, Beijing 100050, Peoples R China.
   [Mei, Shenghui; Li, Xingang; Zhao, Zhigang] Capital Med Univ, Coll Pharmaceut Sci, Dept Clin Pharmacol, Beijing 100069, Peoples R China.
   [Jiang, Xueyun] Capital Med Univ, Coll Pharmaceut Sci, Beijing 100069, Peoples R China.
   [Lin, Song] Capital Med Univ, Beijing Tiantan Hosp, Dept Neurosurg, 6 Tiantan Xili, Beijing 100050, Peoples R China.
C3 Capital Medical University; Capital Medical University; Capital Medical
   University; Capital Medical University
RP Zhao, ZG (corresponding author), Capital Med Univ, Beijing Tiantan Hosp, Dept Pharm, Beijing 100050, Peoples R China.; Zhao, ZG (corresponding author), Capital Med Univ, Coll Pharmaceut Sci, Dept Clin Pharmacol, Beijing 100069, Peoples R China.; Lin, S (corresponding author), Capital Med Univ, Beijing Tiantan Hosp, Dept Neurosurg, 6 Tiantan Xili, Beijing 100050, Peoples R China.
EM
RI Mei, Shenghui/X-3037-2019; Li, Xingang/L-2443-2019
OI Li, Xingang/0000-0001-6726-9571
FU Capital Medical University [16JL13]
FX S.M. and X.J. were supported by the Capital Medical University (No.
   16JL13).
CR Aumente D, 2006, CLIN PHARMACOKINET, V45, P1227, DOI 10.2165/00003088-200645120-00007
   Bain E, 2014, ANN PHARMACOTHER, V48, P292, DOI 10.1177/1060028013511951
   Bauters TGM, 2008, PHARM WORLD SCI, V30, P316, DOI 10.1007/s11096-008-9204-9
   Charles-Schoeman C, 2017, ARTHRITIS RHEUMATOL, V69, P46, DOI 10.1002/art.39833
   Cheng Q, 2004, PHARMACOGENETICS, V14, P557, DOI 10.1097/01.fpc.0000114761.78957.7e
   Colom H, 2009, THER DRUG MONIT, V31, P76, DOI 10.1097/FTD.0b013e3181945624
   den Boer E, 2014, J RHEUMATOL, V41, P2167, DOI 10.3899/jrheum.131290
   Desoky ESE., 2013, PHARM PHARMACOL-RUSS, V4, P139, DOI [10.4236/pp.2013.42020, DOI 10.4236/PP.2013.42020]
   Ding JJ, 2015, CLIN PHARMACOKINET, V54, P305, DOI 10.1007/s40262-014-0212-8
   Dupuis C, 2008, ANTI-CANCER DRUG, V19, P267, DOI 10.1097/CAD.0b013e3282f21376
   Ette EL, 2003, J CLIN PHARMACOL, V43, P610, DOI 10.1177/0091270003253624
   Faltaos DW, 2006, CANCER CHEMOTH PHARM, V58, P626, DOI 10.1007/s00280-006-0202-0
   Fischer M, 2017, CLIN PHARM DRUG DEV, V6, P240, DOI 10.1002/cpdd.279
   Fuksa L, 2010, TOXICOLOGY, V267, P165, DOI 10.1016/j.tox.2009.11.010
   Fukuhara K, 2008, J CLIN PHARM THER, V33, P677, DOI 10.1111/j.1365-2710.2008.00966.x
   FURST DE, 1990, J PHARM SCI-US, V79, P782, DOI 10.1002/jps.2600790907
   Hospira, 2011, LAB METH INJ
   Joerger M, 2012, LEUKEMIA LYMPHOMA, V53, P1867, DOI 10.3109/10428194.2012.676177
   Joerger M, 2010, BRIT J CANCER, V102, P673, DOI 10.1038/sj.bjc.6605559
   Johansson AM, 2011, THER DRUG MONIT, V33, P711, DOI 10.1097/FTD.0b013e31823615e1
   Kasenda B, 2012, ANN HEMATOL, V91, P1257, DOI 10.1007/s00277-012-1441-2
   Khalil HA, 2016, LIPIDS HEALTH DIS, V15, DOI 10.1186/s12944-016-0318-0
   Kim IW, 2012, CLIN THER, V34, P1816, DOI 10.1016/j.clinthera.2012.06.022
   Li X, 2017, EUR J DRUG METAB PHA
   McBride Ali, 2012, J Pharm Pract, V25, P477, DOI 10.1177/0897190012442717
   Mei SH, 2017, ANAL SCI, V33, P665, DOI 10.2116/analsci.33.665
   Min Y, 2009, BIOPHARM DRUG DISPOS, V30, P437, DOI 10.1002/bdd.678
   Molnar P P, 1999, Neuro Oncol, V1, P89, DOI 10.1093/neuonc/1.2.89
   Nader A, 2017, EUR J DRUG METAB PH, V42, P221, DOI 10.1007/s13318-016-0338-1
   Nagulu M, 2010, ASIAN PAC J CANCER P, V11, P403
   Reeves D, 2016, ANN PHARMACOTHER, V50, P1016, DOI 10.1177/1060028016661572
   Ronda N, 2015, ARTHRITIS RHEUMATOL, V67, P1155, DOI 10.1002/art.39039
   Santucci R, 2010, ANTICANCER RES, V30, P963
   SCHWARTZ GJ, 1985, J PEDIATR-US, V106, P522, DOI 10.1016/S0022-3476(85)80697-1
   Stamp LK, 2009, ARTHRITIS RHEUM, V60, P2248, DOI 10.1002/art.24653
   Sun XF, 2017, ONCOTARGET, V8, P49156, DOI 10.18632/oncotarget.17101
   Trevin LR, 2009, J CLIN ONCOL, V27, P5972, DOI 10.1200/JCO.2008.20.4156
   Wasan KM, 2008, NAT REV DRUG DISCOV, V7, P84, DOI 10.1038/nrd2353
   Zhang W, 2015, CHINESE MED J-PEKING, V128, P111, DOI 10.4103/0366-6999.147829
NR 39
TC 27
Z9 27
U1 0
U2 12
PU WILEY
PI HOBOKEN
PA 111 RIVER ST, HOBOKEN 07030-5774, NJ USA
SN 0022-3549
EI 1520-6017
J9 J PHARM SCI-US
JI J. Pharm. Sci.
PD MAY
PY 2018
VL 107
IS 5
BP 1454
EP 1460
DI 10.1016/j.xphs.2018.01.004
PG 7
WC Chemistry, Medicinal; Chemistry, Multidisciplinary; Pharmacology &
   Pharmacy
WE Science Citation Index Expanded (SCI-EXPANDED)
SC Pharmacology & Pharmacy; Chemistry
GA GD6CT
UT WOS:000430597100029
PM 29331383
DA 2024-06-03
ER

PT J
AU Zalewski, NL
   Tobin, WO
AF Zalewski, Nicholas L.
   Tobin, W. Oliver
TI CLIPPERS
SO CURRENT NEUROLOGY AND NEUROSCIENCE REPORTS
LA English
DT Review
DE CLIPPERS; Brainstem encephalitis; Encephalomyelitis; Vasculitis;
   Lymphoma; PCNSL
ID CHRONIC LYMPHOCYTIC INFLAMMATION; CENTRAL-NERVOUS-SYSTEM; B-CELL
   LYMPHOMA; STEROIDS CLIPPERS; MULTIPLE-SCLEROSIS; CNS LYMPHOMA;
   BRAIN-STEM; PATIENT; DIAGNOSIS; BIOPSY
AB Purpose of Review Chronic lymphocytic inflammation with pontine perivascular enhancement responsive to steroids (CLIPPERS) is a recently described treatable, inflammatory, brainstem predominant encephalomyelitis. The diagnosis of CLIPPERS is challenging without a specific biomarker, and thus it is important to consider if both the clinical and radiographic features are consistent with the diagnosis, or rather a disease mimicker.
   Recent Findings Many patients with CLIPPERS-like lesions have been described in the literature with follow-up revealing a range of alternative diagnoses, such as malignancies, vasculitis, and other specific inflammatory diseases. As a result, some have proposed that CLIPPERS might represent a premalignancy state or simply an initial clinical syndrome of a variety of possible etiologies.
   Summary We describe the typical clinical, radiographic, and pathological features of CLIPPERS and emphasize consideration for alternative diagnoses when findings are not classic. A recommended diagnostic evaluation and initial treatment plan is provided.
C1 [Zalewski, Nicholas L.; Tobin, W. Oliver] Mayo Clin, Dept Neurol, Rochester, MN 55905 USA.
   [Tobin, W. Oliver] Mayo Clin, Coll Med, Rochester, MN 55905 USA.
C3 Mayo Clinic; Mayo Clinic
RP Tobin, WO (corresponding author), Mayo Clin, Dept Neurol, Rochester, MN 55905 USA.
EM
RI Tobin, William Oliver/W-7553-2019; Tobin, William Oliver/HII-8589-2022
OI Tobin, William Oliver/0000-0002-8141-2394; Tobin, William
   Oliver/0000-0002-8141-2394
CR Alsherbini K, 2014, JAMA NEUROL, V71, P110, DOI 10.1001/jamaneurol.2013.1578
   Binnahil M, 2016, CAN J NEUROL SCI, V43, P721, DOI 10.1017/cjn.2016.255
   Biotti Damien, 2011, Pract Neurol, V11, P349, DOI 10.1136/practneurol-2011-000043
   Blaabjerg M, 2016, NEUROL-NEUROIMMUNOL, V3, DOI 10.1212/NXI.0000000000000226
   De Graaff HJ, 2013, JAMA NEUROL, V70, P915, DOI 10.1001/jamaneurol.2013.2016
   Dudesek A, 2014, CLIN EXP IMMUNOL, V175, P385, DOI 10.1111/cei.12204
   Fang BY, 2016, JAMA NEUROL, V73, P1297, DOI 10.1001/jamaneurol.2016.2549
   Ferreira RM, 2013, J NEUROL SCI, V327, P61, DOI 10.1016/j.jns.2013.01.032
   Gabilondo I, 2011, J NEUROL, V258, P2090, DOI 10.1007/s00415-011-6068-z
   Gul M, 2015, JCR-J CLIN RHEUMATOL, V21, P144, DOI 10.1097/RHU.0000000000000238
   Hillesheim PB, 2012, ARCH PATHOL LAB MED, V136, P681, DOI 10.5858/arpa.2011-0428-CR
   Jones JL, 2011, BRAIN, V134, pE187, DOI 10.1093/brain/awr134
   Kastrup O, 2011, J NEUROL, V258, P2140, DOI 10.1007/s00415-011-6071-4
   Keegan BM, 2012, ARCH NEUROL-CHICAGO, V69, P819, DOI 10.1001/archneurol.2012.1015
   Kira J, 2012, J NEUROL NEUROSUR PS, V83, P2, DOI 10.1136/jnnp-2011-301626
   Kleinschmidt-DeMasters BK, 2014, J NEUROPATH EXP NEUR, V73, P262, DOI 10.1097/NEN.0000000000000050
   Limousin N, 2012, J NEURO-ONCOL, V107, P223, DOI 10.1007/s11060-011-0724-0
   Lin AW, 2014, CAN J NEUROL SCI, V41, P528, DOI 10.1017/S0317167100018631
   Mashima K, 2015, INT J HEMATOL, V102, P709, DOI 10.1007/s12185-015-1850-9
   Nakamura R, 2016, J NEUROL SCI, V364, P6, DOI 10.1016/j.jns.2016.02.057
   Ortega MR, 2012, NEUROLOGY, V79, P715, DOI 10.1212/WNL.0b013e3182648b77
   Pittock SJ, 2010, BRAIN, V133, P2626, DOI 10.1093/brain/awq164
   Porter AB, 2008, ANN NEUROL, V63, P662, DOI 10.1002/ana.21366
   Simon NG, 2012, J NEUROL NEUROSUR PS, V83, P15, DOI 10.1136/jnnp-2011-301054
   Smith A, 2014, J CUTAN PATHOL, V41, P657, DOI 10.1111/cup.12344
   Symmonds M, 2015, NEUROLOGY, V84, P1177, DOI 10.1212/WNL.0000000000001370
   Taieb G, 2016, NEURORADIOLOGY, V58, P221, DOI 10.1007/s00234-015-1629-y
   Taieb G, 2014, J CUTAN PATHOL, V41, P759, DOI 10.1111/cup.12367
   Taieb G, 2014, J NEUROL SCI, V344, P224, DOI 10.1016/j.jns.2014.06.015
   Taieb G, 2013, JAMA NEUROL, V70, P1200, DOI 10.1001/jamaneurol.2013.3652
   Taieb G, 2012, ARCH NEUROL-CHICAGO, V69, P847, DOI 10.1001/archneurol.2012.122
   Tan BL, 2015, NEUROL-NEUROIMMUNOL, V2, DOI 10.1212/NXI.0000000000000056
   Tobin WO, 2015, WORLD NEUROSURG, V84, P1973, DOI 10.1016/j.wneu.2015.08.062
   Tohge R, 2012, INTERNAL MED, V51, P1115, DOI 10.2169/internalmedicine.51.7040
   Wang L, 2015, J NEUROL SCI, V348, P277, DOI 10.1016/j.jns.2014.11.022
   Wang XL, 2017, J NEUROIMMUNOL, V305, P68, DOI 10.1016/j.jneuroim.2017.01.014
   Weng Ching-Fu, 2015, J Med Case Rep, V9, P266, DOI 10.1186/s13256-015-0750-1
   Zhang YX, 2016, NEUROLOGY, V86, P103, DOI 10.1212/WNL.0000000000002212
NR 38
TC 27
Z9 29
U1 1
U2 11
PU SPRINGER
PI NEW YORK
PA 233 SPRING ST, NEW YORK, NY 10013 USA
SN 1528-4042
EI 1534-6293
J9 CURR NEUROL NEUROSCI
JI Curr. Neurol. Neurosci. Rep.
PD SEP
PY 2017
VL 17
IS 9
AR 65
DI 10.1007/s11910-017-0773-7
PG 7
WC Clinical Neurology; Neurosciences
WE Science Citation Index Expanded (SCI-EXPANDED)
SC Neurosciences & Neurology
GA FH4TT
UT WOS:000411155700001
PM 28726127
DA 2024-06-03
ER

PT J
AU Zeremski, V
   Koehler, M
   Fischer, T
   Schalk, E
AF Zeremski, Vanja
   Koehler, Michael
   Fischer, Thomas
   Schalk, Enrico
TI Characteristics and outcome of patients with primary CNS lymphoma in a
   "real-life" setting compared to a clinical trial
SO ANNALS OF HEMATOLOGY
LA English
DT Article
DE PCNSL; Methotrexate; G-PCNSL-SG-1; Real-life; Clinical trial
ID CENTRAL-NERVOUS-SYSTEM; HIGH-DOSE METHOTREXATE; WHOLE-BRAIN
   RADIOTHERAPY; ELDERLY-PATIENTS; FOLLOW-UP; RITUXIMAB; CYTARABINE;
   SURVIVAL; G-PCNSL-SG-1; CHEMOTHERAPY
AB We aimed to compare the characteristics and outcome of patients treated within the multi-centre German Primary CNS Lymphoma Study Group 1 trial (G-PCNSL-SG-1; TRIAL group) and patients treated outside this clinical trial ("real-life" setting, R-LIFE group). Therefore, we conducted a retrospective single-centre study in order to analyse all patients with newly diagnosed primary CNS lymphoma (PCNSL) treated consecutively in our institution between November 2000 and June 2015. Altogether, 86 patients were analysed (median 68 years). Twenty patients were treated within (TRIAL) and 66 patients outside the clinical trial (R-LIFE), respectively. The majority (n = 75; 87 %) received high-dose methotrexate as the first-line treatment. Thirty-eight of 66 patients (57.6 %) responded to the first-line therapy. The R-LIFE patients were older (median age 70 vs. 62 years; p = 0.005) and had more frequently a worse performance status (ECOG score 2-4: 59.1 vs. 20.0 %; p = 0.004; median Karnofsky index 70 vs. 80 %; p = 0.003) and less frequently a low prognostic score (IELSG score 0-1: 19.7 vs. 45.0 %; p = 0.038), than the TRIAL patients. Median overall survial (OS) was shorter for the R-LIFE patients (9.3 months [95 % CI 1.9-16.7] vs. 33.4 months [95 % CI 17.6-49.2]; p = 0.065). Median progression-free survival (PFS) was significantly inferior for the R-LIFE patients (3.4 months [95 % CI 2.4-4.4] vs. 24.8 months [95 % CI 4.6-45.0]; p = 0.037). Our data indicate that the outcome of PCNSL patients treated outside, but about analogous to the G-PCNSL-SG-1 trial, was poor. This is likely explained by more unfavourable prognostic factors in patients being treated off trial.
C1 [Zeremski, Vanja; Koehler, Michael; Fischer, Thomas; Schalk, Enrico] Univ Magdeburg, Med Ctr, Dept Haematol & Oncol, Leipziger Str 44, D-39120 Magdeburg, Germany.
C3 Otto von Guericke University
RP Schalk, E (corresponding author), Univ Magdeburg, Med Ctr, Dept Haematol & Oncol, Leipziger Str 44, D-39120 Magdeburg, Germany.
EM
RI Schalk, Enrico/X-9283-2019
OI Schalk, Enrico/0000-0003-1892-5098
CR Abrey LE, 2006, J CLIN ONCOL, V24, P5711, DOI 10.1200/JCO.2006.08.2941
   Abrey LE, 2005, J CLIN ONCOL, V23, P5034, DOI 10.1200/JCO.2005.13.524
   [Anonymous], JAMA
   Batchelor T, 2003, J CLIN ONCOL, V21, P1044, DOI 10.1200/JCO.2003.03.036
   CHARLSON ME, 1987, J CHRON DIS, V40, P373, DOI 10.1016/0021-9681(87)90171-8
   Dalia S, 2014, INT J HEMATOL, V99, P450, DOI 10.1007/s12185-014-1540-z
   Ferreri AJM, 2003, J CLIN ONCOL, V21, P266, DOI 10.1200/JCO.2003.09.139
   Ferreri AJM, 2009, LANCET, V374, P1512, DOI 10.1016/S0140-6736(09)61416-1
   Gregory G, 2013, NEURO-ONCOLOGY, V15, P1068, DOI 10.1093/neuonc/not032
   Herrlinger U, 2005, ANN NEUROL, V57, P843, DOI 10.1002/ana.20495
   Herrlinger U, 2002, ANN NEUROL, V51, P247, DOI 10.1002/ana.10102
   Holdhoff M, 2014, NEUROLOGY, V83, P235, DOI 10.1212/WNL.0000000000000593
   Illerhaus G, 2015, ONCOL RES TREAT, V38, P177
   KAPLAN EL, 1958, J AM STAT ASSOC, V53, P457, DOI 10.2307/2281868
   Kasenda B, 2015, ANN ONCOL, V26, P1305, DOI 10.1093/annonc/mdv076
   Korfel A, 2015, NEUROLOGY, V84, P1242, DOI 10.1212/WNL.0000000000001395
   Madle M, 2015, ANN HEMATOL, V94, P1853, DOI 10.1007/s00277-015-2470-4
   O'Brien PC, 2006, INT J RADIAT ONCOL, V64, P408, DOI 10.1016/j.ijrobp.2005.07.958
   Omuro A, 2015, LANCET HAEMATOL, V2, pE251, DOI 10.1016/S2352-3026(15)00074-5
   Omuro A, 2015, BLOOD, V125, P1403, DOI 10.1182/blood-2014-10-604561
   Phillips EH, 2014, CURR HEMATOL MALIG R, V9, P243, DOI 10.1007/s11899-014-0217-2
   Roth P, 2012, NEUROLOGY, V79, P890, DOI 10.1212/WNL.0b013e318266fcb2
   Rubenstein J, 2008, LEUKEMIA LYMPHOMA, V49, P43, DOI 10.1080/10428190802311441
   Rubenstein JL, 2013, J CLIN ONCOL, V31, P3061, DOI 10.1200/JCO.2012.46.9957
   SHUSTER JJ, 1991, J CLIN ONCOL, V9, P191, DOI 10.1200/JCO.1991.9.1.191
   Thiel E, 2010, LANCET ONCOL, V11, P1036, DOI 10.1016/S1470-2045(10)70229-1
   Unger JM, 2014, JNCI-J NATL CANCER I, V106, DOI 10.1093/jnci/dju002
   Villano JL, 2011, BRIT J CANCER, V105, P1414, DOI 10.1038/bjc.2011.357
   Wang XX, 2014, LEUKEMIA LYMPHOMA, V55, P2497, DOI 10.3109/10428194.2014.889823
   Zeremski V, 2015, ONCOL RES TREAT, V38, P155
NR 30
TC 27
Z9 29
U1 0
U2 1
PU SPRINGER
PI NEW YORK
PA 233 SPRING ST, NEW YORK, NY 10013 USA
SN 0939-5555
EI 1432-0584
J9 ANN HEMATOL
JI Ann. Hematol.
PD APR
PY 2016
VL 95
IS 5
BP 793
EP 799
DI 10.1007/s00277-016-2602-5
PG 7
WC Hematology
WE Science Citation Index Expanded (SCI-EXPANDED)
SC Hematology
GA DI1ZS
UT WOS:000373296100014
PM 26801791
DA 2024-06-03
ER

PT J
AU Yamaguchi, S
   Hirata, K
   Kobayashi, H
   Shiga, T
   Manabe, O
   Kobayashi, K
   Motegi, H
   Terasaka, S
   Houkin, K
AF Yamaguchi, Shigeru
   Hirata, Kenji
   Kobayashi, Hiroyuki
   Shiga, Tohru
   Manabe, Osamu
   Kobayashi, Kentaro
   Motegi, Hiroaki
   Terasaka, Shunsuke
   Houkin, Kiyohiro
TI The diagnostic role of <SUP>18</SUP>F-FDG PET for primary central
   nervous system lymphoma
SO ANNALS OF NUCLEAR MEDICINE
LA English
DT Article
DE Brain tumor; Corticosteroid; F-18-FDG PET; Primary central nervous
   system lymphoma; ROC curve
ID B-CELL LYMPHOMA; EXPRESSION; MRI
AB F-18-FDG PET has become one of the most important methods for studying malignant lymphoma, but its diagnostic role for primary central nervous system lymphoma (PCNSL) has not been established. The aim of this study was to determine the appropriate cut-off values of FDG uptake and to investigate how corticosteroid administration influences PCNSL.
   We retrospectively reviewed 82 patients with contrast-enhanced brain tumors who underwent an FDG PET scan at onset, including 19 PCNSLs. FDG uptake of the lesion was assessed by the maximum standardized uptake value (SUVmax) and the ratio of tumor to normal contralateral cortex activity (T/N ratio). Receiver operating characteristic (ROC) curves were generated from the SUVmax and T/N ratios. To investigate the influence of corticosteroid application before a FDG PET scan, we evaluated the association between the FDG uptake of the lesion and the cumulative dose of corticosteroid administration on 13 PCNSL patients who had received steroid treatment before an FDG PET examination.
   The mean FDG SUVmax and T/N ratio of PCNSLs were 22.6 and 2.79, respectively, and these values were significantly higher than those of the other malignant brain tumors. ROC analysis indicated that the evaluation of FDG uptake using the T/N ratio was more reliable than the SUVmax with respect to the differential diagnosis. When PCNSL patients went without steroid application before FDG PET, the accuracy of the T/N ratio with a cut-off point of 2.0 was 91.1 %, the sensitivity was 94.7 %, and the specificity was 87.3 %. Although there are no significant differences in the FDG T/N ratio for PCNSL patients with or without steroid treatment, a negative correlation was found between the T/N ratio and cumulative dose of corticosteroid before PET study (r = -0.71, p = 0.032).
   We concluded that the T/N ratio was superior to SUVmax for FDG uptake assessment as for distinguishing PCNSLs from other malignant brain tumors; the appropriate T/N ratio cut-off point was 2.0. In addition, FDG uptake could be influenced by cumulative doses of corticosteroid before a PET scan, and thus this fact should be taken into consideration when evaluating FDG PET for PCNSL diagnosis.
C1 [Yamaguchi, Shigeru; Kobayashi, Hiroyuki; Motegi, Hiroaki; Terasaka, Shunsuke; Houkin, Kiyohiro] Hokkaido Univ, Grad Sch Med, Dept Neurosurg, Kita Ku, Sapporo, Hokkaido 0608638, Japan.
   [Hirata, Kenji; Shiga, Tohru; Manabe, Osamu; Kobayashi, Kentaro] Hokkaido Univ, Grad Sch Med, Dept Nucl Med, Sapporo, Hokkaido 0608638, Japan.
C3 Hokkaido University; Hokkaido University
RP Terasaka, S (corresponding author), Hokkaido Univ, Grad Sch Med, Dept Neurosurg, Kita Ku, North 15 West 7, Sapporo, Hokkaido 0608638, Japan.
EM
RI 真鍋, 治/JDC-9806-2023; Yamaguchi, Shigeru/G-5419-2012
OI Manabe, Osamu/0000-0001-8518-8441; mao mu, yang
   huang/0000-0001-6153-5461
CR Baraniskin A, 2012, BRIT J HAEMATOL, V156, P421, DOI 10.1111/j.1365-2141.2011.08928.x
   Erdag N, 2001, AM J ROENTGENOL, V176, P1319, DOI 10.2214/ajr.176.5.1761319
   Ferreri AJM, 2011, BLOOD, V118, P510, DOI 10.1182/blood-2011-03-321349
   Hirose Y, 2014, LEUKEMIA LYMPHOMA, V55, P520, DOI 10.3109/10428194.2013.807509
   Hustinx R, 1999, EUR J NUCL MED, V26, P1501, DOI 10.1007/s002590050487
   Karantanis D, 2007, NUCL MED COMMUN, V28, P834, DOI 10.1097/MNM.0b013e328264ae7f
   Kawai N, 2010, J NEURO-ONCOL, V100, P225, DOI 10.1007/s11060-010-0182-0
   Koga H, 2003, ANN NUCL MED, V17, P327, DOI 10.1007/BF02988530
   Kosaka N, 2008, AM J ROENTGENOL, V190, pW365, DOI 10.2214/AJR.07.2660
   Küker W, 2005, J NEURO-ONCOL, V72, P169, DOI 10.1007/s11060-004-3390-7
   Kuratsu Jun-ichi, 2001, International Journal of Clinical Oncology, V6, P183
   Makino K, 2006, SURG NEUROL, V66, P503, DOI 10.1016/j.surneu.2006.05.055
   Makino K, 2011, ANN NUCL MED, V25, P432, DOI 10.1007/s12149-011-0483-1
   Mohile Nimish A, 2008, Clin Adv Hematol Oncol, V6, P818
   Nakamura H, 2011, INT J CLIN ONCOL, V16, P314, DOI 10.1007/s10147-010-0178-y
   Ngeow JYY, 2009, ANN ONCOL, V20, P1543, DOI 10.1093/annonc/mdp030
   Palmedo H, 2006, EUR J NUCL MED MOL I, V33, P164, DOI 10.1007/s00259-005-1917-6
   Roelcke U, 1999, J NEURO-ONCOL, V43, P231, DOI 10.1023/A:1006202402010
   ROSENFELD SS, 1992, J NUCL MED, V33, P532
   Shim HK, 2009, NUCL MED BIOL, V36, P191, DOI 10.1016/j.nucmedbio.2008.11.009
   Watanabe Y, 2013, INT J HEMATOL, V97, P43, DOI 10.1007/s12185-012-1225-4
   Zacharia TT, 2008, J NEUROIMAGING, V18, P411, DOI 10.1111/j.1552-6569.2007.00231.x
NR 22
TC 27
Z9 30
U1 0
U2 8
PU SPRINGER
PI NEW YORK
PA 233 SPRING ST, NEW YORK, NY 10013 USA
SN 0914-7187
EI 1864-6433
J9 ANN NUCL MED
JI Ann. Nucl. Med.
PD AUG
PY 2014
VL 28
IS 7
BP 603
EP 609
DI 10.1007/s12149-014-0851-8
PG 7
WC Radiology, Nuclear Medicine & Medical Imaging
WE Science Citation Index Expanded (SCI-EXPANDED)
SC Radiology, Nuclear Medicine & Medical Imaging
GA AN5VV
UT WOS:000340660700001
PM 24743915
OA Green Submitted
DA 2024-06-03
ER

PT J
AU Joerger, M
   Ferreri, AJM
   Krähenbühl, S
   Schellens, JHM
   Cerny, T
   Zucca, E
   Huitema, ADR
AF Joerger, Markus
   Ferreri, Andres J. M.
   Kraehenbuehl, Stephan
   Schellens, Jan H. M.
   Cerny, Thomas
   Zucca, Emanuele
   Huitema, Alwin D. R.
TI Dosing algorithm to target a predefined AUC in patients with primary
   central nervous system lymphoma receiving high dose methotrexate
SO BRITISH JOURNAL OF CLINICAL PHARMACOLOGY
LA English
DT Article
DE Bayesian estimate; CNS lymphoma; high dose chemotherapy; methotrexate;
   therapeutic drug monitoring
ID PRIMARY CNS LYMPHOMA; UNDER-THE-CURVE; COMBINATION CHEMOTHERAPY;
   LEUCOVORIN RESCUE; CANCER-PATIENTS; RADIOTHERAPY; PHARMACOLOGY; THERAPY;
   TRIAL; PHARMACOKINETICS
AB AIM
   There is no consensus regarding optimal dosing of high dose methotrexate (HDMTX) in patients with primary CNS lymphoma. Our aim was to develop a convenient dosing algorithm to target AUCMTX in the range between 1000 and 1100 mmol l(-1) h.
   METHODS
   A population covariate model from a pooled dataset of 131 patients receiving HDMTX was used to simulate concentration-time curves of 10 000 patients and test the efficacy of a dosing algorithm based on 24 h MTX plasma concentrations to target the prespecified AUC(MTX). These data simulations included interindividual, interoccasion and residual unidentified variability. Patients received a total of four simulated cycles of HDMTX and adjusted MTX dosages were given for cycles two to four.
   RESULTS
   The dosing algorithm proposes MTX dose adaptations ranging from + 75% in patients with MTX C24 < 0.5 mmol ll(-1) up to - 35% in patients with MTX C(24) > 12 mmol l(-1). The proposed dosing algorithm resulted in a marked improvement of the proportion of patients within the AUCMTX target between 1000 and 1100 mmol l(-1) h (11% with standard MTX dose, 35% with the adjusted dose) and a marked reduction of the interindividual variability of MTX exposure.
   CONCLUSIONS
   A simple and practical dosing algorithm for HDMTX has been developed based on MTX 24 h plasma concentrations, and its potential efficacy in improving the proportion of patients within a prespecified target AUCMTX and reducing the interindividual variability of MTX exposure has been shown by data simulations. The clinical benefit of this dosing algorithm should be assessed in patients with primary central nervous system lymphoma (PCNSL).
C1 [Joerger, Markus; Cerny, Thomas] Cantonal Hosp, Dept Hematol & Oncol, Gallen, Switzerland.
   [Ferreri, Andres J. M.] Ist Sci San Raffaele, Dept Oncohematol, Unit Lymphoid Malignancies, I-20132 Milan, Italy.
   [Kraehenbuehl, Stephan] Univ Hosp, Div Clin Pharmacol & Toxicol, CH-4056 Basel, Switzerland.
   [Schellens, Jan H. M.] Netherlands Canc Inst, Antoni van Leeuwenhoek Hosp, Dept Med Oncol, NL-1066 CX Amsterdam, Netherlands.
   [Schellens, Jan H. M.] Univ Utrecht, Fac Pharmaceut Sci, Dept Biomed Anal, Div Drug Toxicol,FAFC Wentgebouw, NL-3584 CA Utrecht, Netherlands.
   [Zucca, Emanuele] Oncol Inst So Switzerland, CH-6500 Bellinzona, Switzerland.
   [Huitema, Alwin D. R.] Netherlands Canc Inst, Slotervaart Hosp, Dept Pharm & Pharmacol, NL-1066 CX Amsterdam, Netherlands.
C3 Vita-Salute San Raffaele University; IRCCS Ospedale San Raffaele;
   Netherlands Cancer Institute; Utrecht University; Institute of Oncology
   Research (IOR); Slotervaart Hospital; Netherlands Cancer Institute
RP Joerger, M (corresponding author), Cantonal Hosp, Dept Hematol & Oncol, Rorschacherstr 95,9007 St, Gallen, Switzerland.
EM
RI Krähenbühl, Stephan/Z-4744-2019; Ferreri, Andrés J. M./A-6662-2013
OI Zucca, Emanuele/0000-0002-5522-6109
CR Abrey LE, 2010, NAT REV CLIN ONCOL, V7, P306, DOI 10.1038/nrclinonc.2010.66
   Abrey LE, 2000, J CLIN ONCOL, V18, P3144, DOI 10.1200/JCO.2000.18.17.3144
   Batchelor T, 2003, J CLIN ONCOL, V21, P1044, DOI 10.1200/JCO.2003.03.036
   Beal S., 1988, NONMEM User Guides
   Blasco H, 2010, BRIT J CLIN PHARMACO, V70, P367, DOI 10.1111/j.1365-2125.2010.03712.x
   Bourguignon L, 2010, FUND CLIN PHARMACOL, V24, P109, DOI 10.1111/j.1472-8206.2009.00732.x
   Bustad A, 2006, CLIN PHARMACOKINET, V45, P365, DOI 10.2165/00003088-200645040-00003
   Calderoni A, 2002, J NEURO-ONCOL, V59, P227, DOI 10.1023/A:1019993018162
   DeAngelis LM, 2002, J CLIN ONCOL, V20, P4643, DOI 10.1200/JCO.2002.11.013
   Dombrowsky E, 2011, THER DRUG MONIT, V33, P99, DOI 10.1097/FTD.0b013e318203b41e
   Ferreri AJM, 2002, NEUROLOGY, V58, P1513, DOI 10.1212/WNL.58.10.1513
   Ferreri AJM, 2004, BRIT J CANCER, V90, P353, DOI 10.1038/sj.bjc.6601472
   Ferreri AJM, 2009, LANCET, V374, P1512, DOI 10.1016/S0140-6736(09)61416-1
   GOORIN A, 1995, MED PEDIATR ONCOL, V24, P362, DOI 10.1002/mpo.2950240605
   Herrlinger U, 2005, ANN NEUROL, V57, P843, DOI 10.1002/ana.20495
   Herrlinger U, 2002, ANN NEUROL, V51, P247, DOI 10.1002/ana.10102
   Hiraga S, 1999, J NEUROSURG, V91, P221, DOI 10.3171/jns.1999.91.2.0221
   ILIADIS A, 1985, J PHARMACOKINET BIOP, V13, P101, DOI 10.1007/BF01073659
   ISACOFF WH, 1977, CANCER TREAT REP, V61, P1665
   Jelliffe R, 2000, THER DRUG MONIT, V22, P354, DOI 10.1097/00007691-200006000-00019
   Joerger M, 2007, BRIT J CLIN PHARMACO, V64, P622, DOI 10.1111/j.1365-2125.2007.02956.x
   Joerger M, 2006, BRIT J CLIN PHARMACO, V62, P71, DOI 10.1111/j.1365-2125.2005.02513.x
   Joerger M, 2010, BRIT J CANCER, V102, P673, DOI 10.1038/sj.bjc.6605559
   JOLIVET J, 1983, NEW ENGL J MED, V309, P1094, DOI 10.1056/NEJM198311033091805
   LEVITT M, 1973, CANCER RES, V33, P1729
   Li J, 2002, CANCER CHEMOTH PHARM, V50, P373, DOI 10.1007/s00280-002-0512-9
   MALLET A, 1986, BIOMETRIKA, V73, P645
   Marcus R., 2009, GUIDELINES DIAGNOSIS
   Morris PG, 2009, LANCET NEUROL, V8, P581, DOI 10.1016/S1474-4422(09)70091-2
   Plotkin SR, 2004, CLIN CANCER RES, V10, P5643, DOI 10.1158/1078-0432.CCR-04-0159
   Poortmans PMP, 2003, J CLIN ONCOL, V21, P4483, DOI 10.1200/JCO.2003.03.108
   SAETER G, 1991, J CLIN ONCOL, V9, P1766, DOI 10.1200/JCO.1991.9.10.1766
   SCHORNAGEL JH, 1983, CANCER TREAT REV, V10, P53, DOI 10.1016/S0305-7372(83)80032-2
   STEWART AL, 1985, CANCER CHEMOTH PHARM, V14, P165, DOI 10.1007/BF00434358
   Widemann BC, 1997, J CLIN ONCOL, V15, P2125, DOI 10.1200/JCO.1997.15.5.2125
NR 35
TC 27
Z9 28
U1 0
U2 1
PU WILEY-BLACKWELL
PI MALDEN
PA COMMERCE PLACE, 350 MAIN ST, MALDEN 02148, MA USA
SN 0306-5251
J9 BRIT J CLIN PHARMACO
JI Br. J. Clin. Pharmacol.
PD FEB
PY 2012
VL 73
IS 2
BP 240
EP 247
DI 10.1111/j.1365-2125.2011.04084.x
PG 8
WC Pharmacology & Pharmacy
WE Science Citation Index Expanded (SCI-EXPANDED)
SC Pharmacology & Pharmacy
GA 873IU
UT WOS:000298876300011
PM 21838788
OA Green Published
DA 2024-06-03
ER

PT J
AU Thapa, DR
   Li, XM
   Jamieson, BD
   Martínez-Maza, O
AF Thapa, Dharma R.
   Li, Xinmin
   Jamieson, Beth D.
   Martinez-Maza, Otoniel
TI Overexpression of MicroRNAs from the miR-17-92 Paralog Clusters in
   AIDS-Related Non-Hodgkin's Lymphomas
SO PLOS ONE
LA English
DT Article
ID DOWN-REGULATION; P53 PROTEIN; EXPRESSION; GENE; AMPLIFICATION;
   POLYCISTRON; MIR-155; RNA; IDENTIFICATION; DELETION
AB Background: Individuals infected by HIV are at an increased risk for developing non-Hodgkin's lymphomas (AIDS-NHL). In the highly active antiretroviral therapy (HAART) era, there has been a significant decline in the incidence of AIDS-associated primary central nervous system lymphoma (PCNSL). However, only a modest decrease in incidence has been reported for other AIDS-NHL subtypes. Thus, AIDS-NHLs remain a significant cause of morbidity and mortality in HIV infected individuals. Recently, much attention has been directed toward the role of miRNAs in cancer, including NHL. Several miRNAs, including those encoded by the miR-17-92 polycistron, have been shown to play significant roles in B cell tumorigenesis. However, the role of miRNAs in NHL in the setting of HIV infection has not been defined.
   Methodology/Principal Findings: We used quantitative realtime PCR to assess the expression of miRNAs from three different paralog clusters, miR-17-92, miR-106a-363, and miR-106b-25 in 24 cases of AIDS-NHLs representing four tumor types, Burkitt's lymphoma (BL, n = 6), diffuse large B-cell lymphoma (DLBCL, n = 8), primary central nervous system lymphoma (PCNSL, n = 5), and primary effusion lymphoma (PEL, n = 5). We also used microarray analysis to identify a differentiation specific miRNA signature of naive, germinal center, and memory B cell subsets from tonsils (n = 4). miRNAs from the miR-17-92 paralog clusters were upregulated by B cells, specifically during the GC differentiation stage. We also found overexpression of these miRNA clusters in all four AIDS-NHL subtypes. Finally, we also show that select miRNAs from these clusters (miR-17, miR-106a, and miR-106b) inhibited p21 in AIDS-BL and DLBCL cases, thus providing a mechanistic role for these miRNAs in AIDS-NHL pathogenesis.
   Conclusion: Dysregulation of miR-17-92 paralog clusters is a common feature of AIDS-associated NHLs.
C1 [Thapa, Dharma R.; Martinez-Maza, Otoniel] Univ Calif Los Angeles, David Geffen Sch Med, Dept Microbiol Immunol & Mol Genet, Los Angeles, CA 90095 USA.
   [Li, Xinmin] Univ Calif Los Angeles, David Geffen Sch Med, Dept Pathol & Lab Med, Los Angeles, CA 90095 USA.
   [Jamieson, Beth D.] Univ Calif Los Angeles, David Geffen Sch Med, Dept Med, Los Angeles, CA 90095 USA.
   [Martinez-Maza, Otoniel] Univ Calif Los Angeles, David Geffen Sch Med, Dept Obstet & Gynecol, Los Angeles, CA 90095 USA.
   [Martinez-Maza, Otoniel] Univ Calif Los Angeles, Sch Publ Hlth, Dept Epidemiol, Los Angeles, CA 90024 USA.
   [Martinez-Maza, Otoniel] Univ Calif Los Angeles, UCLA AIDS Inst, Los Angeles, CA USA.
   [Martinez-Maza, Otoniel] Univ Calif Los Angeles, Jonsson Comprehens Canc Ctr, Los Angeles, CA 90024 USA.
C3 University of California System; University of California Los Angeles;
   University of California Los Angeles Medical Center; David Geffen School
   of Medicine at UCLA; University of California System; University of
   California Los Angeles; University of California Los Angeles Medical
   Center; David Geffen School of Medicine at UCLA; University of
   California System; University of California Los Angeles; University of
   California Los Angeles Medical Center; David Geffen School of Medicine
   at UCLA; University of California System; University of California Los
   Angeles; University of California Los Angeles Medical Center; David
   Geffen School of Medicine at UCLA; University of California System;
   University of California Los Angeles; University of California System;
   University of California Los Angeles; UCLA Jonsson Comprehensive Cancer
   Center; University of California System; University of California Los
   Angeles
RP Thapa, DR (corresponding author), Univ Calif Los Angeles, David Geffen Sch Med, Dept Microbiol Immunol & Mol Genet, Los Angeles, CA 90095 USA.
EM
RI Thapa, Dharma R/C-2854-2012; Martinez-Maza, Otoniel/B-2667-2009
OI Martinez-Maza, Otoniel/0000-0003-1364-0675
FU California HIV Research Program (CHRP) [D08-LA-311]; National Institutes
   of Health [R01-CA57152, NCI U01-CA096230, P30-AI28697]; NCI
   [P30-AI28697, U01-AI35040, P30-CA016042]; James B. Pendleton Charitable
   Trust; McCarthy Family Foundation
FX DRT received fellowship support from the California HIV Research Program
   (CHRP) (D08-LA-311). These studies were supported by grants from the
   National Institutes of Health (R01-CA57152, and pilot project
   supplemental funds from the NCI to U01-AI35040, P30-AI28697 and
   P30-CA016042). Lymphoma specimens were provided by the AIDS and Cancer
   Specimen Resource (ACSR), which is supported by the NIH (NCI
   U01-CA096230). UCLA AIDS Institute Center for AIDS Research (CFAR) core
   facilities were used in this study and were supported by an NIH grant
   (P30-AI28697). This work was carried out in the facilities of the UCLA
   AIDS Institute, which were supported, in part, by funds from the James
   B. Pendleton Charitable Trust and the McCarthy Family Foundation. No
   additional external funding was received for this study. The funders had
   no role in study design, data collection, and analysis, decision to
   publish, or preparation of the manuscript.
CR BALLERINI P, 1993, BLOOD, V81, P166
   Baraniskin A, 2011, BLOOD
   Bartel DP, 2004, CELL, V116, P281, DOI 10.1016/S0092-8674(04)00045-5
   Calin G.A., 2006, Nature Rev, P847
   Chen RW, 2008, BLOOD, V112, P822, DOI 10.1182/blood-2008-03-142182
   Cimmino A, 2005, P NATL ACAD SCI USA, V102, P13944, DOI 10.1073/pnas.0506654102
   Costinean S, 2006, P NATL ACAD SCI USA, V103, P7024, DOI 10.1073/pnas.0602266103
   Crum-Cianflone N, 2009, AIDS, V23, P41, DOI 10.1097/QAD.0b013e328317cc2d
   Eis PS, 2005, P NATL ACAD SCI USA, V102, P3627, DOI 10.1073/pnas.0500613102
   Gaidano G, 1998, AM J PATHOL, V152, P623
   Goedert JJ, 1998, LANCET, V351, P1833, DOI 10.1016/S0140-6736(97)09028-4
   Gottwein E, 2007, NATURE, V450, P1096, DOI 10.1038/nature05992
   Gottwein E, 2010, J VIROL, V84, P5229, DOI 10.1128/JVI.00202-10
   Griffiths-Jones S, 2006, NUCLEIC ACIDS RES, V34, pD140, DOI 10.1093/nar/gkj112
   He L, 2005, NATURE, V435, P828, DOI 10.1038/nature03552
   Inomata M, 2009, BLOOD, V113, P396, DOI 10.1182/blood-2008-07-163907
   Ivanovska I, 2008, MOL CELL BIOL, V28, P2167, DOI 10.1128/MCB.01977-07
   Jiang JM, 2006, GENE CHROMOSOME CANC, V45, P103, DOI 10.1002/gcc.20264
   Kan T, 2009, GASTROENTEROLOGY, V136, P1689, DOI 10.1053/j.gastro.2009.02.002
   Kim YK, 2009, NUCLEIC ACIDS RES, V37, P1672, DOI 10.1093/nar/gkp002
   Kirk O, 2001, BLOOD, V98, P3406, DOI 10.1182/blood.V98.12.3406
   Klein U, 2003, P NATL ACAD SCI USA, V100, P2639, DOI 10.1073/pnas.0437996100
   Kluiver J, 2006, GENE CHROMOSOME CANC, V45, P147, DOI 10.1002/gcc.20273
   Kluiver J, 2005, J PATHOL, V207, P243, DOI 10.1002/path.1825
   Lawrie CH, 2008, BRIT J HAEMATOL, V141, P672, DOI 10.1111/j.1365-2141.2008.07077.x
   Lewis BP, 2005, CELL, V120, P15, DOI 10.1016/j.cell.2004.12.035
   Li C, 2009, BLOOD, V113, P6681, DOI 10.1182/blood-2009-01-202028
   Lin J, 2011, LEUKEMIA, V25, P145, DOI 10.1038/leu.2010.230
   Malumbres R, 2009, BLOOD, V113, P3754, DOI 10.1182/blood-2008-10-184077
   Mao X, 2002, GENE CHROMOSOME CANC, V35, P144, DOI 10.1002/gcc.10104
   Martin A, 1998, BRIT J HAEMATOL, V101, P311, DOI 10.1046/j.1365-2141.1998.00692.x
   Mi SL, 2010, P NATL ACAD SCI USA, V107, P3710, DOI 10.1073/pnas.0914900107
   Monni O, 1998, GENE CHROMOSOME CANC, V21, P298, DOI 10.1002/(SICI)1098-2264(199804)21:4<298::AID-GCC3>3.0.CO;2-U
   Mu P, 2009, GENE DEV, V23, P2806, DOI 10.1101/gad.1872909
   Neat MJ, 2001, GENE CHROMOSOME CANC, V32, P236, DOI 10.1002/gcc.1187
   Nie K, 2008, AM J PATHOL, V173, P242, DOI 10.2353/ajpath.2008.080009
   O'Donnell KA, 2005, NATURE, V435, P839, DOI 10.1038/nature03677
   O'Hara AJ, 2008, BLOOD, V111, P2347, DOI 10.1182/blood-2007-08-104463
   O'Hara AJ, 2009, BLOOD, V113, P5938, DOI 10.1182/blood-2008-09-179168
   Olive V, 2009, GENE DEV, V23, P2839, DOI 10.1101/gad.1861409
   Ota A, 2004, CANCER RES, V64, P3087, DOI 10.1158/0008-5472.CAN-03-3773
   PASCUAL V, 1994, J EXP MED, V180, P329, DOI 10.1084/jem.180.1.329
   Pickering MT, 2009, ONCOGENE, V28, P140, DOI 10.1038/onc.2008.372
   Rao PH, 1998, BLOOD, V92, P234, DOI 10.1182/blood.V92.1.234.413k22_234_240
   Raphael M., 2001, WHO CLASSIFICATION T, P260
   Saeed AI, 2003, BIOTECHNIQUES, V34, P374, DOI 10.2144/03342mt01
   Skalsky RL, 2007, J VIROL, V81, P12836, DOI 10.1128/JVI.01804-07
   Tam W, 1997, MOL CELL BIOL, V17, P1490, DOI 10.1128/MCB.17.3.1490
   TAN LP, 2009, LAB INVEST, P1
   Tanzer A, 2004, J MOL BIOL, V339, P327, DOI 10.1016/j.jmb.2004.03.065
   van den Berg A, 2003, GENE CHROMOSOME CANC, V37, P20, DOI 10.1002/gcc.10186
   Ventura A, 2008, CELL, V132, P875, DOI 10.1016/j.cell.2008.02.019
   Venturini L, 2007, BLOOD, V109, P4399, DOI 10.1182/blood-2006-09-045104
   VILLUENDAS R, 1993, BLOOD, V82, P3151
   Wong P, 2010, CANCER RES, V70, P3833, DOI 10.1158/0008-5472.CAN-09-3268
   Woods K, 2007, J BIOL CHEM, V282, P2130, DOI 10.1074/jbc.C600252200
   Xia T, 2008, CANCER RES, V68, P1436, DOI 10.1158/0008-5472.CAN-07-5126
   Xiao CC, 2008, NAT IMMUNOL, V9, P405, DOI 10.1038/ni1575
   Yan HL, 2009, EMBO J, V28, P2719, DOI 10.1038/emboj.2009.214
   Yu ZR, 2008, J CELL BIOL, V182, P509, DOI 10.1083/jcb.200801079
   Zhang J, 2009, BLOOD, V113, P4586, DOI 10.1182/blood-2008-09-178186
NR 61
TC 27
Z9 33
U1 0
U2 4
PU PUBLIC LIBRARY SCIENCE
PI SAN FRANCISCO
PA 1160 BATTERY STREET, STE 100, SAN FRANCISCO, CA 94111 USA
SN 1932-6203
J9 PLOS ONE
JI PLoS One
PD JUN 16
PY 2011
VL 6
IS 6
AR e20781
DI 10.1371/journal.pone.0020781
PG 11
WC Multidisciplinary Sciences
WE Science Citation Index Expanded (SCI-EXPANDED)
SC Science & Technology - Other Topics
GA 778UW
UT WOS:000291734100021
PM 21698185
OA Green Published, Green Submitted, gold
DA 2024-06-03
ER

PT J
AU Jahnke, K
   Thiel, E
   Bechrakis, NE
   Willerding, G
   Kraemer, DF
   Fischer, L
   Korfel, A
AF Jahnke, Kristoph
   Thiel, Eckhard
   Bechrakis, Nikolaos E.
   Willerding, Gregor
   Kraemer, Dale F.
   Fischer, Lars
   Korfel, Agnieszka
TI Ifosfamide or trofosfamide in patients with intraocular lymphoma
SO JOURNAL OF NEURO-ONCOLOGY
LA English
DT Article
DE Intraocular lymphoma; Ifosfamide; Primary CNS lymphoma; Trofosfamide
ID NERVOUS-SYSTEM LYMPHOMA; PRIMARY CNS LYMPHOMA; COLLABORATIVE
   GROUP-REPORT; RETICULUM-CELL SARCOMA; DEFERRED RADIOTHERAPY;
   METHOTREXATE; INVOLVEMENT; MANAGEMENT; DIAGNOSIS
AB We evaluated response, progression-free survival (PFS), overall survival (OS), relapse patterns and long-term toxicity of intraocular lymphoma (IOL) patients treated with ifosfamide (IFO) or trofosfamide (TRO). In a prospective single center study, IFO or TRO were given to 10 patients with IOL. The median patient age was 73 (range 46-83) years. Six patients were pretreated with up to four treatment regimens for ocular or cerebral lymphoma, and four were therapy-naive. All patients responded, including nine complete remissions and one partial remission, with a median PFS of 18 (7-36) months. Seven patients relapsed: five in the eye and two in the brain. Median OS was 32 (7-37+) months. No long-term toxicity was observed in patients treated with IFO or TRO alone. IFO or TRO were active and well tolerated in this study. Thus, they may represent suitable combination partners for other cytostatics used for PCNSL and IOL treatment.
C1 [Jahnke, Kristoph; Thiel, Eckhard; Fischer, Lars; Korfel, Agnieszka] Charite Univ Med Berlin, Dept Hematol & Oncol, D-12200 Berlin, Germany.
   [Bechrakis, Nikolaos E.; Willerding, Gregor] Charite Univ Med Berlin, Dept Ophthalmol, D-12200 Berlin, Germany.
   [Kraemer, Dale F.] Oregon Hlth & Sci Univ, Dept Med Informat & Clin Epidemiol, Portland, OR 97239 USA.
   [Kraemer, Dale F.] Oregon Hlth & Sci Univ, Dept Publ Hlth & Prevent Med, Portland, OR 97239 USA.
   [Kraemer, Dale F.] Oregon State Univ, Dept Pharm Practice, Corvallis, OR 97331 USA.
C3 Free University of Berlin; Humboldt University of Berlin; Charite
   Universitatsmedizin Berlin; Free University of Berlin; Humboldt
   University of Berlin; Charite Universitatsmedizin Berlin; Oregon Health
   & Science University; Oregon Health & Science University; Oregon State
   University
RP Jahnke, K (corresponding author), Charite Univ Med Berlin, Dept Hematol & Oncol, Campus Benjamin Franklin,Hindenburgdamm 30, D-12200 Berlin, Germany.
EM
CR Abrey LE, 2005, J CLIN ONCOL, V23, P5034, DOI 10.1200/JCO.2005.13.524
   Batchelor T, 2003, J CLIN ONCOL, V21, P1044, DOI 10.1200/JCO.2003.03.036
   Batchelor TT, 2003, CLIN CANCER RES, V9, P711
   Buggage RR, 2001, CURR OPIN ONCOL, V13, P137, DOI 10.1097/00001622-200105000-00001
   CHAR DH, 1988, OPHTHALMOLOGY, V95, P625
   Ferreri AJM, 2002, ANN ONCOL, V13, P531, DOI 10.1093/annonc/mdf080
   Ferreri AJM, 2003, J CLIN ONCOL, V21, P266, DOI 10.1200/JCO.2003.09.139
   FINE HA, 1993, ANN INTERN MED, V119, P1093, DOI 10.7326/0003-4819-119-11-199312010-00007
   Grimm SA, 2008, NEUROLOGY, V71, P1355, DOI 10.1212/01.wnl.0000327672.04729.8c
   Grimm SA, 2007, ANN ONCOL, V18, P1851, DOI 10.1093/annonc/mdm340
   Hormigo A, 2004, BRIT J HAEMATOL, V126, P202, DOI 10.1111/j.1365-2141.2004.05028.x
   Jahnke K, 2006, GRAEF ARCH CLIN EXP, V244, P663, DOI 10.1007/s00417-005-0138-9
   Jahnke K, 2005, ANN ONCOL, V16, P1974, DOI 10.1093/annonc/mdi409
   KAPLAN EL, 1958, J AM STAT ASSOC, V53, P457, DOI 10.2307/2281868
   Kitzmann AS, 2007, EYE, V21, P1524, DOI 10.1038/sj.eye.6702804
   Pels H, 2003, J CLIN ONCOL, V21, P4489, DOI 10.1200/JCO.2003.04.056
   PETERSON K, 1993, CANCER, V72, P843, DOI 10.1002/1097-0142(19930801)72:3<843::AID-CNCR2820720333>3.0.CO;2-#
   ROCKWOOD EJ, 1984, J NEUROSURG, V61, P369, DOI 10.3171/jns.1984.61.2.0369
   Rodeghiero F, 2003, ONCOLOGY-BASEL, V65, P85, DOI 10.1159/000073367
   Smith JR, 2002, OPHTHALMOLOGY, V109, P1709, DOI 10.1016/S0161-6420(02)01125-9
   Sneller V, 2003, ANN ONCOL, V14, P1, DOI 10.1093/annonc/mdg701
   Wagner A, 1997, ANTI-CANCER DRUG, V8, P419, DOI 10.1097/00001813-199706000-00002
   Yule SM, 1997, CLIN CANCER RES, V3, P1985
NR 23
TC 27
Z9 29
U1 0
U2 0
PU SPRINGER
PI NEW YORK
PA 233 SPRING ST, NEW YORK, NY 10013 USA
SN 0167-594X
EI 1573-7373
J9 J NEURO-ONCOL
JI J. Neuro-Oncol.
PD JUN
PY 2009
VL 93
IS 2
BP 213
EP 217
DI 10.1007/s11060-008-9761-8
PG 5
WC Oncology; Clinical Neurology
WE Science Citation Index Expanded (SCI-EXPANDED)
SC Oncology; Neurosciences & Neurology
GA 446SP
UT WOS:000266141700006
PM 19099202
DA 2024-06-03
ER

PT J
AU Ng, S
   Butzkueven, H
   Kalnins, R
   Rowe, C
AF Ng, Steven
   Butzkueven, Helmut
   Kalnins, Renate
   Rowe, Christopher
TI Prolonged interval between sentinel pseudotumoral demyelination and
   development of primary CNS lymphoma
SO JOURNAL OF CLINICAL NEUROSCIENCE
LA English
DT Article
DE PCNSL; lymphoma; demyelination; pseudotumoral; sentinel; post-partum
ID POSITRON-EMISSION-TOMOGRAPHY; NERVOUS-SYSTEM LYMPHOMA;
   MULTIPLE-SCLEROSIS; PREGNANCY
AB Primary central nervous system lymphoma (PCNSL) can be associated with preceding demyelinating pseudotumoral brain lesions. The 'sentinel' demyelinating lesions recede spontaneously or with corticosteroid, and are followed by development of PCNSL typically within 12 months. This report describes a 29 year-old post-partum woman who developed PCNSL 4 years after a biopsy-proven pseudotumoral demyelinating episode. She presented with focal seizures in February 2005. She subsequently developed hemiparesis and raised intracranial pressure. MRI showed two contrast enhancing lesions in the right frontal lobe, which were hypermetabolic on F-18-FDG PET. A provisional diagnosis of tumefactive multiple sclerosis was made. Symptoms recurred despite multiple courses of high dose corticosteroid. Brain biopsy confirmed large B-cell non-Hodgkin's lymphoma. This patient illustrates the importance of considering PCNSL in patients presenting with a space-occupying lesion, even with previously confirmed demyelination, and that the interval between the two events may be several years. (C) 2006 Elsevier Ltd. All rights reserved.
C1 Austin Hlth, Dept Nucl Med, Melbourne, Vic 3084, Australia.
   Austin Hlth, Ctr PET, Melbourne, Vic 3084, Australia.
   Royal Melbourne Hosp, Dept Neurol, Melbourne, Vic 3050, Australia.
   Austin Hlth, Dept Anat Pathol, Melbourne, Vic 3084, Australia.
C3 Florey Institute of Neuroscience & Mental Health; Austin Research
   Institute; Austin Research Institute; Florey Institute of Neuroscience &
   Mental Health; Royal Melbourne Hospital; Austin Research Institute;
   Florey Institute of Neuroscience & Mental Health
RP Ng, S (corresponding author), Austin Hlth, Dept Nucl Med, 145 Studley Rd, Melbourne, Vic 3084, Australia.
EM
RI Rowe, Christopher/AAX-8748-2021
OI Butzkueven, Helmut/0000-0003-3940-8727
CR Alarcia R, 2000, REV NEUROLOGIA, V31, P955, DOI 10.33588/rn.3110.2000363
   Alderson L, 1996, J NEUROL NEUROSUR PS, V60, P102, DOI 10.1136/jnnp.60.1.102
   Bakshi R, 1998, J NEUROIMAGING, V8, P228, DOI 10.1111/jon199884228
   Confavreux C, 1998, NEW ENGL J MED, V339, P285, DOI 10.1056/NEJM199807303390501
   DEANGELIS LM, 1990, J NEURO-ONCOL, V9, P177, DOI 10.1007/BF02427840
   Ding Xinmin, 2003, Sichuan Da Xue Xue Bao Yi Xue Ban, V34, P327
   Heckmann JG, 2000, NERVENARZT, V71, P305, DOI 10.1007/s001150050562
   IOACHIM HL, 1985, ARCH PATHOL LAB MED, V109, P803
   ODoherty MJ, 1997, J NUCL MED, V38, P1575
   Okamoto Kouichirou, 2004, Radiat Med, V22, P63
   ROSENFELD SS, 1992, J NUCL MED, V33, P532
   Schiepers C, 1997, MULT SCLER, V3, P8, DOI 10.1177/135245859700300102
NR 12
TC 27
Z9 31
U1 0
U2 2
PU ELSEVIER SCI LTD
PI OXFORD
PA THE BOULEVARD, LANGFORD LANE, KIDLINGTON, OXFORD OX5 1GB, OXON, ENGLAND
SN 0967-5868
EI 1532-2653
J9 J CLIN NEUROSCI
JI J. Clin. Neurosci.
PD NOV
PY 2007
VL 14
IS 11
BP 1126
EP 1129
DI 10.1016/j.jocn.2006.05.003
PG 4
WC Clinical Neurology; Neurosciences
WE Science Citation Index Expanded (SCI-EXPANDED)
SC Neurosciences & Neurology
GA 236CW
UT WOS:000251281600017
PM 17890092
DA 2024-06-03
ER

PT J
AU Ferreri, AJM
   Reni, M
AF Ferreri, Andres J. M.
   Reni, Michele
TI Primary central nervous system lymphoma
SO CRITICAL REVIEWS IN ONCOLOGY HEMATOLOGY
LA English
DT Review
DE PCNSL; radiotherapy; cerebrospinal fluid; methotrexate; ternozolornide;
   rituximab
ID PRIMARY-CNS-LYMPHOMA; NON-HODGKINS-LYMPHOMA; HIGH-DOSE METHOTREXATE;
   STEM-CELL TRANSPLANTATION; PRIMARY BRAIN LYMPHOMAS; IMMUNOCOMPETENT
   PATIENTS; RADIATION-THERAPY; PHASE-II; DEFERRED RADIOTHERAPY; INITIAL
   TREATMENT
AB Primary central nervous system lymphomas (PCNSL) are aggressive malignancies that arise in distinct anatomical sites, which display unique structural, biological and immunological conditions. So far, despite recent therapeutic advances, these malignancies exhibit one of the worst prognoses among all non-Hodgkin lymphomas (NHL). For a long time, radiotherapy (RT) has been the standard treatment, producing a response rate of 60-65% and a notable neurological improvement in most cases. However, relapse usually occurred within a few months after RT, with a median survival of 14 months and a 5-year survival of approximately 15-24%. Although the introduction of systemic chemotherapy has consistently improved survival, the prognosis of PCNSL is still dismal, with high rates of local relapse and consequent death. Defining the optimum therapeutic management is difficult because of potential selection biases in large retrospective reviews and the limited number of prospective studies. Although studies published on PCNSL are increasing, several therapeutic questions still remain unanswered after a decade of research. (c) 2007 Elsevier Ireland Ltd. All rights reserved.
C1 Ist Sci San Raffaele, Med Oncol Unit, I-20132 Milan, Italy.
C3 Vita-Salute San Raffaele University; IRCCS Ospedale San Raffaele
RP Ferreri, AJM (corresponding author), Ist Sci San Raffaele, Med Oncol Unit, I-20132 Milan, Italy.
EM
RI Ferreri, Andrés J. M./A-6662-2013; tonekaboni, seyed Hassan/K-1575-2019;
   Reni, Michele/J-8296-2016
OI Reni, Michele/0000-0002-6463-0293
CR Abrey LE, 2003, J CLIN ONCOL, V21, P4151, DOI 10.1200/JCO.2003.05.024
   Abrey LE, 2005, J CLIN ONCOL, V23, P5034, DOI 10.1200/JCO.2005.13.524
   Abrey LE, 1998, J CLIN ONCOL, V16, P859, DOI 10.1200/JCO.1998.16.3.859
   Abrey LE, 2000, J CLIN ONCOL, V18, P3144, DOI 10.1200/JCO.2000.18.17.3144
   BASHIR R, 1994, J NEURO-ONCOL, V20, P47, DOI 10.1007/BF01057960
   Batchelor T, 2003, J CLIN ONCOL, V21, P1044, DOI 10.1200/JCO.2003.03.036
   Batchelor TT, 2003, CLIN CANCER RES, V9, P711
   BERGMANN M, 1994, CLIN NEUROL NEUROSUR, V96, P236, DOI 10.1016/0303-8467(94)90075-2
   Bessell EM, 1996, J CLIN ONCOL, V14, P945, DOI 10.1200/JCO.1996.14.3.945
   Blay JY, 1998, J CLIN ONCOL, V16, P864, DOI 10.1200/JCO.1998.16.3.864
   BLAY JY, 1995, BLOOD, V86, P2922
   Blumenthal DT, 1999, NEUROLOGY, V52, P1648, DOI 10.1212/WNL.52.8.1648
   Boiardi A, 1999, J NEUROL, V246, P31, DOI 10.1007/s004150050302
   Brada M, 1998, INT J RADIAT ONCOL, V40, P1157, DOI 10.1016/S0360-3016(98)00002-9
   Bromberg JEC, 2002, NEUROLOGY, V59, P762, DOI 10.1212/WNL.59.5.762
   Camilleri-Broet S, 1998, AM J CLIN PATHOL, V110, P607, DOI 10.1093/ajcp/110.5.607
   Camilleri-Broët S, 2006, BLOOD, V107, P190, DOI 10.1182/blood-2005-03-1024
   CARBONE PP, 1971, CANCER RES, V31, P1860
   Cheng AL, 1998, CANCER, V82, P1946, DOI 10.1002/(SICI)1097-0142(19980515)82:10<1946::AID-CNCR19>3.0.CO;2-T
   Cher L, 1996, NEUROLOGY, V46, P1757, DOI 10.1212/WNL.46.6.1757
   Colombat P, 2006, BONE MARROW TRANSPL, V38, P417, DOI 10.1038/sj.bmt.1705452
   Dabaja BS, 2003, CANCER, V98, P1021, DOI 10.1002/cncr.11627
   DeAngelis L M, 1994, Recent Results Cancer Res, V135, P155
   DEANGELIS LM, 1992, J CLIN ONCOL, V10, P635, DOI 10.1200/JCO.1992.10.4.635
   DEANGELIS LM, 1991, CANCER-AM CANCER SOC, V67, P1431, DOI 10.1002/1097-0142(19910301)67:5<1431::AID-CNCR2820670527>3.0.CO;2-K
   Doolittle ND, 2000, CANCER, V88, P637, DOI 10.1002/(SICI)1097-0142(20000201)88:3<637::AID-CNCR22>3.0.CO;2-Y
   EBY NL, 1988, CANCER-AM CANCER SOC, V62, P2461, DOI 10.1002/1097-0142(19881201)62:11<2461::AID-CNCR2820621135>3.0.CO;2-M
   Enting RH, 2004, NEUROLOGY, V63, P901, DOI 10.1212/01.WNL.0000137050.43114.42
   Ferreri AJM, 1995, CANCER TREAT REV, V21, P415, DOI 10.1016/0305-7372(95)90028-4
   Ferreri AJM, 1996, CANCER, V77, P827, DOI 10.1002/(SICI)1097-0142(19960301)77:5<827::AID-CNCR4>3.0.CO;2-B
   Ferreri AJM, 2003, J CLIN ONCOL, V21, P266, DOI 10.1200/JCO.2003.09.139
   Ferreri AJM, 2002, NEUROLOGY, V58, P1513, DOI 10.1212/WNL.58.10.1513
   FINE HA, 1993, ANN INTERN MED, V119, P1093, DOI 10.7326/0003-4819-119-11-199312010-00007
   Fischer L, 2004, NEUROLOGY, V62, P1885, DOI 10.1212/01.WNL.0000125334.16420.D4
   Freilich RJ, 1996, NEUROLOGY, V46, P435, DOI 10.1212/WNL.46.2.435
   GALETTO G, 1993, JAMA-J AM MED ASSOC, V269, P92, DOI 10.1001/jama.269.1.92
   GLASS J, 1994, J NEUROSURG, V81, P188, DOI 10.3171/jns.1994.81.2.0188
   Glick JH., 1995, P AN M AM SOC CLIN, V14, P391
   GREIG NH, 1990, JNCI-J NATL CANCER I, V82, P1621, DOI 10.1093/jnci/82.20.1621
   Guha-Thakurta N, 1999, J NEURO-ONCOL, V43, P259, DOI 10.1023/A:1006210703827
   HACKER SM, 1992, TRANSPLANTATION, V53, P691, DOI 10.1097/00007890-199203000-00042
   HAYAKAWA T, 1994, J NEURO-ONCOL, V19, P197, DOI 10.1007/BF01053274
   HENRY JM, 1974, CANCER, V34, P1293, DOI 10.1002/1097-0142(197410)34:4<1293::AID-CNCR2820340441>3.0.CO;2-P
   Herrlinger U, 2005, ANN NEUROL, V57, P843, DOI 10.1002/ana.20495
   Herrlinger U, 2002, ANN NEUROL, V51, P247, DOI 10.1002/ana.10102
   HOCHBERG FH, 1988, J NEUROSURG, V68, P835, DOI 10.3171/jns.1988.68.6.0835
   Hu XW, 1996, CHINESE MED J-PEKING, V109, P414
   Illerhaus G, 2006, J CLIN ONCOL, V24, P3865, DOI 10.1200/JCO.2006.06.2117
   Jahnke K, 2006, J CLIN ONCOL, V24, P4754, DOI 10.1200/JCO.2006.06.7165
   JELLINGER KA, 1995, J NEURO-ONCOL, V24, P33, DOI 10.1007/BF01052655
   JELLINGER KA, 1992, J CANCER RES CLIN, V119, P7, DOI 10.1007/BF01209483
   Kadan-Lottick NS, 2002, CANCER, V95, P193, DOI 10.1002/cncr.10643
   Khalfallah S, 1996, BONE MARROW TRANSPL, V18, P1021
   KRANZ BR, 1992, LANCET, V340, P727, DOI 10.1016/0140-6736(92)92263-F
   LACHANCE DH, 1991, NEUROLOGY, V41, P95, DOI 10.1212/WNL.41.1.95
   Laperriere NJ, 1997, RADIOTHER ONCOL, V43, P247, DOI 10.1016/S0167-8140(97)00074-1
   Levy-Clarke GA, 2005, HEMATOL ONCOL CLIN N, V19, P739, DOI 10.1016/j.hoc.2005.05.011
   LOEFFLER JS, 1985, J CLIN ONCOL, V3, P490, DOI 10.1200/JCO.1985.3.4.490
   Marshall AG, 1998, J NEUROL SCI, V158, P226, DOI 10.1016/S0022-510X(98)00111-7
   Montemurro M, 2005, BLOOD, V106, p934A
   NELSON DF, 1992, INT J RADIAT ONCOL, V23, P9, DOI 10.1016/0360-3016(92)90538-S
   Ng VL, 1998, IMMUNOL REV, V162, P293, DOI 10.1111/j.1600-065X.1998.tb01449.x
   Nguyen PL, 2005, J CLIN ONCOL, V23, P1507, DOI 10.1200/JCO.2005.01.161
   OBrien PC, 1996, MED J AUSTRALIA, V165, P424
   ONEILL BP, 1995, J NEURO-ONCOL, V25, P67, DOI 10.1007/BF01054724
   ONEILL BP, 1993, 4 INT C ANT CANC CHE, P119
   Palmedo H, 2006, EUR J NUCL MED MOL I, V33, P164, DOI 10.1007/s00259-005-1917-6
   Panageas KS, 2005, CANCER-AM CANCER SOC, V104, P2466, DOI 10.1002/cncr.21481
   Plotkin SR, 2004, CLIN CANCER RES, V10, P5643, DOI 10.1158/1078-0432.CCR-04-0159
   Raizer JJ, 2005, J NEURO-ONCOL, V71, P173, DOI 10.1007/s11060-004-1360-8
   Reni M, 1997, J NEURO-ONCOL, V32, P135, DOI 10.1023/A:1005781815202
   Reni M, 2004, EUR J CANCER, V40, P1682, DOI 10.1016/j.ejca.2004.03.008
   Reni M, 1999, BRIT J CANCER, V79, P530, DOI 10.1038/sj.bjc.6690083
   Reni M, 1997, ANN ONCOL, V8, P227, DOI 10.1023/A:1008201717089
   Reni M, 2001, INT J RADIAT ONCOL, V51, P419, DOI 10.1016/S0360-3016(01)01639-X
   Sandor V, 1998, J CLIN ONCOL, V16, P3000, DOI 10.1200/JCO.1998.16.9.3000
   SCHAUMBURG HH, 1972, BRAIN, V95, P199, DOI 10.1093/brain/95.2.199
   Schlegel U, 2001, J NEUROL NEUROSUR PS, V71, P118, DOI 10.1136/jnnp.71.1.118
   Schultz C, 1996, J CLIN ONCOL, V14, P556, DOI 10.1200/JCO.1996.14.2.556
   Schulz H, 2004, HAEMATOLOGICA, V89, P753
   Shenkier TN, 2005, J CLIN ONCOL, V23, P2233, DOI 10.1200/JCO.2005.07.109
   SHIBATA S, 1989, NEUROSURGERY, V25, P14, DOI 10.1227/00006123-198907000-00003
   SOCIE G, 1990, CANCER-AM CANCER SOC, V65, P322, DOI 10.1002/1097-0142(19900115)65:2<322::AID-CNCR2820650223>3.0.CO;2-D
   Soussain C, 2001, J CLIN ONCOL, V19, P742, DOI 10.1200/JCO.2001.19.3.742
   STEWART DA, 2004, BLOOD, V104, P901
   Tyson RM, 2003, LEUKEMIA LYMPHOMA, V44, P627, DOI 10.1080/1042819021000055057
   Varadi G, 1999, LEUKEMIA LYMPHOMA, V34, P185, DOI 10.3109/10428199909083396
   Weigel R, 2004, CLIN NEUROL NEUROSUR, V106, P82, DOI 10.1016/j.clineuro.2003.09.005
NR 88
TC 27
Z9 35
U1 0
U2 2
PU ELSEVIER SCIENCE INC
PI NEW YORK
PA STE 800, 230 PARK AVE, NEW YORK, NY 10169 USA
SN 1040-8428
EI 1879-0461
J9 CRIT REV ONCOL HEMAT
JI Crit. Rev. Oncol./Hematol.
PD SEP
PY 2007
VL 63
IS 3
BP 257
EP 268
DI 10.1016/j.critrevonc.2007.04.012
PG 12
WC Oncology; Hematology
WE Science Citation Index Expanded (SCI-EXPANDED)
SC Oncology; Hematology
GA 208AL
UT WOS:000249292100007
PM 17590348
DA 2024-06-03
ER

PT J
AU Abla, O
   Sandlund, JT
   Sung, L
   Brock, P
   Corbett, R
   Kirov, I
   Griffin, TC
   Blaser, S
   Weitzman, S
AF Abla, Oussama
   Sandlund, John T.
   Sung, Lillian
   Brock, Penelope
   Corbett, Rob
   Kirov, Ivan
   Griffin, Timothy C.
   Blaser, Susan
   Weitzman, Sheila
TI A case series of pediatric primary central nervous system lymphoma:
   Favorable outcome without cranial irradiation
SO PEDIATRIC BLOOD & CANCER
LA English
DT Article
DE children; lymphoma; primary CNS
ID PRIMARY CNS LYMPHOMA; ACUTE LYMPHOBLASTIC-LEUKEMIA; LONG-TERM REMISSION;
   DEFERRED RADIOTHERAPY; FOLLOW-UP; PHASE-II; CHEMOTHERAPY; METHOTREXATE;
   THERAPY; MULTICENTER
AB Purpose. To describe the outcome of childhood primary central nervous system lymphoma (PCNSL) treated with chemotherapy alone or with chemotherapy plus cranial radiotherapy (CRT).
   Methods. Retrospective chart review of children with PCNSL at six tertiary care pediatric centers.
   Results. Eight immunocompetent and four immunocompromised children were included. Ten children received chemotherapy alone without CRT, with most receiving high-dose methotrexate and high-dose cytarabine. Five year event-free survival (EFS) in this group was 70.0 +/- 14.5%. Two children received chemotherapy plus CRT; one relapsed and died while the other is alive in remission. Three children died, including two from relapsed disease. The other child with human immunodeficiency virus infection died of an opportunistic infection while in remission following chemotherapy alone.
   Conclusions. Most children with PCNSL can achieve long-term remissions with chemotherapy alone without CRT. Multi-center prospective studies are needed to confirm or refute these results in a larger number of patients.
C1 Hosp Sick Children, Div Hematol Oncol, Toronto, ON M5G 1X8, Canada.
   Hosp Sick Children, Div Radiol, Toronto, ON M5G 1X8, Canada.
   St Jude Childrens Res Hosp, Memphis, TN 38105 USA.
   Great Ormond St Hosp Sick Children, London WC1N 3JH, England.
   S Isl Child Canc Serv, Christchurch, New Zealand.
   Childrens Hosp Orange Cty, Orange, CA USA.
   Cook Childrens Med Ctr, Ft Worth, TX USA.
C3 University of Toronto; Hospital for Sick Children (SickKids); University
   of Toronto; Hospital for Sick Children (SickKids); St Jude Children's
   Research Hospital; University of London; University College London;
   Great Ormond Street Hospital for Children NHS Foundation Trust;
   Childrens Hospital of Orange County; Cook Children's Medical Center
RP Abla, O (corresponding author), Hosp Sick Children, Div Hematol Oncol, 555 Univ Ave, Toronto, ON M5G 1X8, Canada.
EM
FU NCI NIH HHS [CA 21765] Funding Source: Medline
CR Abrey LE, 2000, J CLIN ONCOL, V18, P3144, DOI 10.1200/JCO.2000.18.17.3144
   Batchelor T, 2003, J CLIN ONCOL, V21, P1044, DOI 10.1200/JCO.2003.03.036
   Bessell EM, 2004, INT J RADIAT ONCOL, V59, P501, DOI 10.1016/j.ijrobp.2003.11.001
   Blay JY, 2000, ANN ONCOL, V11, P39, DOI 10.1023/A:1008364612406
   CAIRO MS, 2003, P AN M AM SOC CLIN, V22, P796
   Cheng G, 2003, BONE MARROW TRANSPL, V31, P679, DOI 10.1038/sj.bmt.1703917
   COHEN IJ, 1986, CANCER-AM CANCER SOC, V57, P6, DOI 10.1002/1097-0142(19860101)57:1<6::AID-CNCR2820570104>3.0.CO;2-B
   DeAngelis LM, 2002, J CLIN ONCOL, V20, P4643, DOI 10.1200/JCO.2002.11.013
   Ferreri AJM, 2000, ANN ONCOL, V11, P927, DOI 10.1023/A:1008376412784
   Ferreri AJM, 2002, NEUROLOGY, V58, P1513, DOI 10.1212/WNL.58.10.1513
   Ferreri AJM, 2003, J CLIN ONCOL, V21, P1649, DOI 10.1200/JCO.2003.99.053
   Guha-Thakurta N, 1999, J NEURO-ONCOL, V43, P259, DOI 10.1023/A:1006210703827
   Herrlinger U, 2002, ANN NEUROL, V51, P247, DOI 10.1002/ana.10102
   Khalfallah S, 1996, BONE MARROW TRANSPL, V18, P1021
   Khan RB, 2002, J NEURO-ONCOL, V58, P175, DOI 10.1023/A:1016077907952
   KIROV I, 2003, J PEDIAT HEMATOL ONC, V25, pS11
   Kraemer DF, 2001, NEUROSURGERY, V48, P1033, DOI 10.1097/00006123-200105000-00013
   Löning L, 2000, BLOOD, V95, P2770, DOI 10.1182/blood.V95.9.2770.009k16_2770_2775
   McAllister LD, 2000, NEUROSURGERY, V46, P51, DOI 10.1093/neurosurgery/46.1.51
   Newell ME, 2004, CANCER-AM CANCER SOC, V100, P2627, DOI 10.1002/cncr.20300
   O'Brien P, 2000, J CLIN ONCOL, V18, P519, DOI 10.1200/JCO.2000.18.3.519
   Pels H, 2003, J CLIN ONCOL, V21, P4489, DOI 10.1200/JCO.2003.04.056
   Pirotte B, 1997, J NEURO-ONCOL, V32, P63, DOI 10.1023/A:1005733416571
   Roychowdhury S, 2003, CANCER RES, V63, P965
   SAID JA, 1989, J CONSULT CLIN PSYCH, V57, P251, DOI 10.1037/0022-006X.57.2.251
   Schabet M, 1999, J NEURO-ONCOL, V43, P199, DOI 10.1023/A:1006290032052
   SLEVIN ML, 1982, MED PEDIATR ONCOL, P157, DOI 10.1002/mpo.2950100715
   Soussain C, 2001, J CLIN ONCOL, V19, P742, DOI 10.1200/JCO.2001.19.3.742
   Watanabe T, 2003, J NEURO-ONCOL, V63, P87, DOI 10.1023/A:1023760824739
NR 29
TC 27
Z9 30
U1 1
U2 5
PU WILEY
PI HOBOKEN
PA 111 RIVER ST, HOBOKEN 07030-5774, NJ USA
SN 1545-5009
EI 1545-5017
J9 PEDIATR BLOOD CANCER
JI Pediatr. Blood Cancer
PD DEC
PY 2006
VL 47
IS 7
BP 880
EP 885
DI 10.1002/pbc.20736
PG 6
WC Oncology; Hematology; Pediatrics
WE Science Citation Index Expanded (SCI-EXPANDED)
SC Oncology; Hematology; Pediatrics
GA 102IH
UT WOS:000241803700004
PM 16365864
DA 2024-06-03
ER

PT J
AU Sarkar, C
   Sharma, MC
   Deb, P
   Singh, R
   Santosh, V
   Shankar, SK
AF Sarkar, C
   Sharma, MC
   Deb, P
   Singh, R
   Santosh, V
   Shankar, SK
TI Primary central nervous system lymphoma - a hospital based study of
   incidence and clinicopathological features from India (1980-2003)
SO JOURNAL OF NEURO-ONCOLOGY
LA English
DT Article; Proceedings Paper
CT 14th International Congress of Neuropathology
CY SEP 03-06, 2000
CL BIRMINGHAM, ENGLAND
DE age; immunocompetent; immunocompromised; incidence; India; PCNSL
ID BRAIN LYMPHOMA; CNS-LYMPHOMA; EPIDEMIOLOGY; AIDS; ERA; SURVIVAL;
   PROFILE; TRENDS
AB Over the last two decades, an increase in the incidence of PCNSL cases has been reported in the West, both among immunosuppressed and immunocompetent patients. The present study was undertaken to assess the trend of incidence of PCNSL cases in India. To the best of our knowledge, only a single such report is available from India.
   All biopsy proven PCNSL cases obtained from the Neurosurgical databases of two large referral hospitals, one in Northern India (AIIMS, New Delhi) and another in Southern India (NIMHANS, Bangalore) from the period 1980 to 2003, were reviewed. Immunophenotyping was done and relevant clinical details collected. Appropriate statistical analysis was done to assess any change in trend of incidence or age at presentation.
   PCNSL cases constituted 0.95% and 0.92% of the total intracranial neoplasms at AIIMS and at NIMHANS, respectively. The mean age for cases diagnosed at AIIMS was 44.35 years, while that for NIMHANS was 39.51 years. Statistical analysis to evaluate any change in trend either of incidence or of age at presentation, over the study period, did not reveal any significant change. All the cases occurred in immunocompetent patients, except one case of HIV positive at NIMHANS, and one case of renal transplant at AIIMS. Frontal lobe was the most common site of involvement. Majority of the cases were diffuse, high grade, large cell lymphoma, B-cell immunophenotype.
   Thus, this multicentric hospital based study did not reveal any increase in incidence of PCNSL cases in India over the past 24 years. Further, in contrast to the West, majority of the cases in this Indian study were immunocompetent and a decade younger than in the West. The association of PCNSL with HIV/AIDS has been low in India, possibly due to early death in AIDS on account of opportunistic infections.
C1 All India Inst Med Sci, Dept Pathol, New Delhi 110029, India.
   All India Inst Med Sci, Dept Biostat, New Delhi 110029, India.
   NIMHANS, Dept Neuropathol, Bangalore, Karnataka, India.
C3 All India Institute of Medical Sciences (AIIMS) New Delhi; All India
   Institute of Medical Sciences (AIIMS) New Delhi; National Institute of
   Mental Health & Neurosciences - India
RP All India Inst Med Sci, Dept Pathol, Ansari Nagar, New Delhi 110029, India.
EM
CR Basso U, 2002, EUR J CANCER, V38, P1298, DOI 10.1016/S0959-8049(02)00031-X
   Bataille B, 2000, J NEUROSURG, V92, P261, DOI 10.3171/jns.2000.92.2.0261
   Batara JF, 2003, CURR OPIN NEUROL, V16, P671, DOI 10.1097/00019052-200312000-00005
   Chow KU, 2001, LEUKEMIA LYMPHOMA, V41, P105, DOI 10.3109/10428190109057959
   Cote TR, 1996, J NATL CANCER I, V88, P675, DOI 10.1093/jnci/88.10.675
   EBY NL, 1988, CANCER-AM CANCER SOC, V62, P2461, DOI 10.1002/1097-0142(19881201)62:11<2461::AID-CNCR2820621135>3.0.CO;2-M
   FINE HA, 1993, ANN INTERN MED, V119, P1093, DOI 10.7326/0003-4819-119-11-199312010-00007
   Forsyth PA, 1996, HEMATOL ONCOL CLIN N, V10, P1125, DOI 10.1016/S0889-8588(05)70388-9
   Gates AE, 2002, ONCOLOGY-NY, V16, P657
   Hao D, 1999, ANN ONCOL, V10, P65, DOI 10.1023/A:1008308313914
   HAYAKAWA T, 1994, J NEURO-ONCOL, V19, P197, DOI 10.1007/BF01053274
   Ironside JW, 2002, DIAGNOSTIC PATHOLOGY, P291
   JELLINGER KA, 1992, J CANCER RES CLIN, V119, P7, DOI 10.1007/BF01209483
   JONCAS JH, 1990, J CLIN ONCOL, V8, P378, DOI 10.1200/JCO.1990.8.3.378
   Kadan-Lottick NS, 2002, CANCER, V95, P193, DOI 10.1002/cncr.10643
   Kleihues P., 2000, Pathology and Genetics of Tumours of the Nervous System, P29
   KLEIHUES PDR, 2000, WHO CLASSIFICATION T, P27
   KROGHJENSEN M, 1995, LEUKEMIA LYMPHOMA, V19, P223
   Lanjewar DN, 1998, AIDS, V12, P309, DOI 10.1097/00002030-199803000-00009
   Launay O, 2003, B CANCER, V90, P387
   Martinez AJ, 1998, PATHOL RES PRACT, V194, P473, DOI 10.1016/S0344-0338(98)80116-7
   Nakamura M, 2004, HISTOL HISTOPATHOL, V19, P211, DOI 10.14670/HH-19.211
   Olson JE, 2002, CANCER, V95, P1504, DOI 10.1002/cncr.10851
   Powari M, 2002, SURG NEUROL, V57, P113, DOI 10.1016/S0090-3019(01)00675-9
   Rao CR, 2003, NEUROL INDIA, V51, P237
   Sacktor N, 2002, J NEUROVIROL, V8, P115, DOI 10.1080/13550280290101094
   Satishchandra P, 2000, INDIAN J MED RES, V111, P14
   Slowik F, 1975, ACTA NEUROPATH BER S, V6, P95, DOI DOI 10.1007/978-3-662-08456-4_16
   Yau YH, 1996, LANCET, V348, P890, DOI 10.1016/S0140-6736(05)64759-9
NR 29
TC 27
Z9 35
U1 0
U2 1
PU SPRINGER
PI NEW YORK
PA ONE NEW YORK PLAZA, SUITE 4600, NEW YORK, NY, UNITED STATES
SN 0167-594X
EI 1573-7373
J9 J NEURO-ONCOL
JI J. Neuro-Oncol.
PD JAN
PY 2005
VL 71
IS 2
BP 199
EP 204
DI 10.1007/s11060-004-1385-z
PG 6
WC Oncology; Clinical Neurology
WE Science Citation Index Expanded (SCI-EXPANDED); Conference Proceedings Citation Index - Science (CPCI-S)
SC Oncology; Neurosciences & Neurology
GA 896NI
UT WOS:000226940400019
PM 15690139
DA 2024-06-03
ER

PT J
AU Shi, XX
   Miao, WM
   Pang, DW
   Wu, JS
   Tong, QS
   Li, JX
   Luo, JQ
   Li, WY
   Du, JZ
   Wang, J
AF Shi, Xiao-Xiao
   Miao, Wei-Min
   Pang, Di-Wen
   Wu, Jia-Si
   Tong, Qi-Song
   Li, Jia-Xian
   Luo, Jia-Qi
   Li, Wen-Yu
   Du, Jin-Zhi
   Wang, Jun
TI Angiopep-2 conjugated nanoparticles loaded with doxorubicin for the
   treatment of primary central nervous system lymphoma
SO BIOMATERIALS SCIENCE
LA English
DT Article
ID BRAIN; DELIVERY; TRANSFERRIN; STRATEGIES; DIAGNOSIS
AB Primary central nervous system lymphoma (PCNSL) is a rare brain tumor. Its therapeutic efficacy is much lower than that of traditional lymphoma, largely due to the presence of the blood-brain barrier (BBB), which hinders the effective drug delivery and deposition on the disease site. Angiopep-2 (ANG) can target low-density lipoprotein receptor-related protein (LRP) on the surface of brain capillary endothelial cells (BCECs) and exhibits high BBB transport capability. In this study, we designed an ANG conjugated poly(ethylene glycol)-b-poly(epsilon-caprolactone) (PEG-b-PCL) (APP) nanoparticle to deliver doxorubicin (DOX) for the treatment of PCNSL. Our data indicated that the targeted APP nanoparticles showed significantly increased cellular uptake by BCECs compared with the control nanoparticles. In the intracranial SU-DHL-2-LUC lymphoma xenograft mice model, APP enhanced drug deposition in tumor tissues, and DOX-loaded APP (APP@DOX) exhibited a better therapeutic effect than free DOX and nontargeted PP@DOX, which significantly prolonged the survival time of mice.
C1 [Shi, Xiao-Xiao; Pang, Di-Wen; Li, Jia-Xian; Du, Jin-Zhi; Wang, Jun] South China Univ Technol, Guangzhou Peoples Hosp 1, Guangzhou 510006, Peoples R China.
   [Shi, Xiao-Xiao; Pang, Di-Wen; Li, Jia-Xian; Du, Jin-Zhi; Wang, Jun] South China Univ Technol, Inst Life Sci, Sch Med, Guangzhou 510006, Peoples R China.
   [Miao, Wei-Min] South China Univ Technol, Sch Biol & Biol Engn, Guangzhou Int Campus, Guangzhou 510006, Peoples R China.
   [Wu, Jia-Si; Tong, Qi-Song; Luo, Jia-Qi; Wang, Jun] South China Univ Technol, Sch Biomed Sci & Engn, Guangzhou Int Campus, Guangzhou 510006, Peoples R China.
   [Li, Wen-Yu] Guangdong Prov Peoples Hosp, Lymphoma Div, Guangzhou 510080, Peoples R China.
   [Du, Jin-Zhi] South China Univ Technol, Natl Engn Res Ctr Tissue Restorat & Reconstruct, Guangzhou 510006, Peoples R China.
   [Du, Jin-Zhi] Guangzhou Regenerat Med & Hlth Guangdong Lab, Guangzhou 510005, Peoples R China.
   [Wang, Jun] South China Univ Technol, Key Lab Biomed Mat & Engn, Minist Educ, Guangzhou 510006, Peoples R China.
   [Wang, Jun] South China Univ Technol, Key Lab Biomed Engn Guangdong Prov, Guangzhou 510006, Peoples R China.
   [Wang, Jun] South China Univ Technol, Innovat Ctr Tissue Restorat & Reconstruct, Guangzhou 510006, Peoples R China.
C3 South China University of Technology; South China University of
   Technology; South China University of Technology; South China University
   of Technology; Guangdong Academy of Medical Sciences & Guangdong General
   Hospital; South China University of Technology; Bioland Lab; South China
   University of Technology; South China University of Technology; South
   China University of Technology
RP Du, JZ (corresponding author), South China Univ Technol, Guangzhou Peoples Hosp 1, Guangzhou 510006, Peoples R China.; Du, JZ (corresponding author), South China Univ Technol, Inst Life Sci, Sch Med, Guangzhou 510006, Peoples R China.; Du, JZ (corresponding author), South China Univ Technol, Natl Engn Res Ctr Tissue Restorat & Reconstruct, Guangzhou 510006, Peoples R China.; Du, JZ (corresponding author), Guangzhou Regenerat Med & Hlth Guangdong Lab, Guangzhou 510005, Peoples R China.
EM
RI wei, xiao/ISB-6027-2023; Wang, Jun/F-9033-2011; Jiang,
   Tao/IWM-7503-2023; jiang, lei/IWE-1124-2023; zhang, lin/IZQ-4870-2023;
   Shi, Xiaoxiao/D-6554-2014
OI Wang, Jun/0000-0001-9957-9208; 
FU National Key R&D Program of China [2017YFA0205600]; National Natural
   Science Foundation of China [31771091, 51922043]; Guangdong Natural
   Science Funds for Distinguished Young Scholar [2017A030306018]; Program
   for Guangdong Introducing Innovative and Entrepreneurial Teams
   [2017ZT07S054]; Guangdong Provincial Pearl River Talents Program
   [2017GC010713, 2017GC010304]; Outstanding Scholar Program of Guangzhou
   Regenerative Medicine and Health Guangdong Laboratory
   [2018GZR110102001]; Fundamental Research Funds for Central Universities
FX This work was supported by the National Key R&D Program of China
   (2017YFA0205600), the National Natural Science Foundation of China
   (31771091, and 51922043), the Guangdong Natural Science Funds for
   Distinguished Young Scholar (2017A030306018), the Program for Guangdong
   Introducing Innovative and Entrepreneurial Teams (2017ZT07S054), the
   Guangdong Provincial Pearl River Talents Program (2017GC010713,
   2017GC010304), the Outstanding Scholar Program of Guangzhou Regenerative
   Medicine and Health Guangdong Laboratory (2018GZR110102001), and the
   Fundamental Research Funds for Central Universities. We are very
   grateful to Prof. Peng Li (Guangzhou Institute of Biomedicine and
   Health, Chinese Academy of Sciences) for giving us the lymphoma cell
   lines (SU-DHL-2 and SU-DHL-2-Luc).
CR Abbott NJ, 2006, NAT REV NEUROSCI, V7, P41, DOI 10.1038/nrn1824
   Anraku Y, 2017, NAT COMMUN, V8, DOI 10.1038/s41467-017-00952-3
   BARAKAT M, 2019, J CLIN ONCOL S, V37
   Batchelor TT, 2019, HEMATOL ONCOL, V37, P15, DOI 10.1002/hon.2598
   Bentivoglio M, 2014, TRENDS NEUROSCI, V37, P325, DOI 10.1016/j.tins.2014.03.007
   Carvalho SM, 2019, BIOMATER SCI-UK, V7, P2102, DOI 10.1039/c8bm01528g
   Chen Y, 2012, ADV DRUG DELIVER REV, V64, P640, DOI 10.1016/j.addr.2011.11.010
   Clark AJ, 2015, P NATL ACAD SCI USA, V112, P12486, DOI 10.1073/pnas.1517048112
   Deckert M, 2011, LEUKEMIA, V25, P1797, DOI 10.1038/leu.2011.169
   Faivre G., 2019, CLIN LYMPHOMA MYELOM, V10, P2152
   Furtado D, 2018, ADV MATER, V30, DOI 10.1002/adma.201801362
   Hoffman S, 2006, NEURO-ONCOLOGY, V8, P27, DOI 10.1215/S1522851705000323
   Illerhaus G, 2018, BLOOD, V132, P681, DOI 10.1182/blood-2018-01-791558
   Korfel A, 2013, NAT REV NEUROL, V9, P317, DOI 10.1038/nrneurol.2013.83
   Lee JY, 2019, EUR RADIOL, V29, P5539, DOI 10.1007/s00330-019-06097-9
   Li SY, 2018, PATHOLOGY, V50, P74, DOI 10.1016/j.pathol.2017.09.006
   Pardridge WM, 2001, DRUG DISCOV TODAY, V6, P381, DOI 10.1016/S1359-6446(01)01721-4
   Ruan SB, 2018, ADV FUNCT MATER, V28, DOI 10.1002/adfm.201802227
   Rubenstein JL, 2013, BLOOD, V122, P2318, DOI 10.1182/blood-2013-06-453084
   Saunders NR, 2014, FRONT NEUROSCI-SWITZ, V8, DOI 10.3389/fnins.2014.00404
   Schlegel U, 2018, CURR OPIN NEUROL, V31, P733, DOI 10.1097/WCO.0000000000000619
   Sethi TK, 2019, LEUKEMIA LYMPHOMA, V60, P6, DOI 10.1080/10428194.2018.1466296
   Shuai XT, 2003, MACROMOLECULES, V36, P5751, DOI 10.1021/ma034390w
   Sun CM, 2017, NANOMED-NANOTECHNOL, V13, P2605, DOI 10.1016/j.nano.2017.07.009
   Tang W, 2019, CHEM SOC REV, V48, P2967, DOI 10.1039/c8cs00805a
   Tsou YH, 2017, SMALL, V13, DOI 10.1002/smll.201701921
   Vasudevan SM, 2019, BIOMATER SCI-UK, V7, P4017, DOI 10.1039/c9bm00955h
   Wang R, 2015, J NEURO-ONCOL, V122, P35, DOI 10.1007/s11060-014-1690-0
   Wang Y, 2013, J AM CHEM SOC, V135, P4799, DOI 10.1021/ja312221g
   Washington KE, 2018, ACS BIOMATER SCI ENG, V4, P997, DOI 10.1021/acsbiomaterials.7b00972
   Wu D, 2019, ADV MATER, V31, DOI 10.1002/adma.201807557
   Xin HL, 2012, BIOMATERIALS, V33, P1673, DOI 10.1016/j.biomaterials.2011.11.018
   Xin HL, 2011, BIOMATERIALS, V32, P4293, DOI 10.1016/j.biomaterials.2011.02.044
   Xuan K. H., 2015, LANCET ONCOL, V16, P322
   Yang XC, 2019, SAUDI J BIOL SCI, V26, P595, DOI 10.1016/j.sjbs.2018.11.004
   Zhang PC, 2012, J CONTROL RELEASE, V159, P429, DOI 10.1016/j.jconrel.2012.01.031
   Zhang TT, 2016, BIOMATER SCI-UK, V4, P219, DOI 10.1039/c5bm00383k
   Zhao PF, 2018, CHEM SCI, V9, P2674, DOI 10.1039/c7sc04853j
   Zheng M, 2019, ADV MATER, V31, DOI 10.1002/adma.201903277
NR 39
TC 26
Z9 28
U1 1
U2 60
PU ROYAL SOC CHEMISTRY
PI CAMBRIDGE
PA THOMAS GRAHAM HOUSE, SCIENCE PARK, MILTON RD, CAMBRIDGE CB4 0WF, CAMBS,
   ENGLAND
SN 2047-4830
EI 2047-4849
J9 BIOMATER SCI-UK
JI Biomater. Sci.
PD MAR 7
PY 2020
VL 8
IS 5
BP 1290
EP 1297
DI 10.1039/c9bm01750j
PG 8
WC Materials Science, Biomaterials
WE Science Citation Index Expanded (SCI-EXPANDED)
SC Materials Science
GA KU2SR
UT WOS:000519558700005
PM 31899467
DA 2024-06-03
ER

PT J
AU Takashima, Y
   Kawaguchi, A
   Sato, R
   Yoshida, K
   Hayano, A
   Homma, J
   Fukai, J
   Iwadate, Y
   Kajiwara, K
   Ishizawa, S
   Hondoh, H
   Nakano, M
   Ogawa, S
   Tashiro, K
   Yamanaka, R
AF Takashima, Yasuo
   Kawaguchi, Atsushi
   Sato, Ryuichi
   Yoshida, Kenichi
   Hayano, Azusa
   Homma, Jumpei
   Fukai, Junya
   Iwadate, Yasuo
   Kajiwara, Koji
   Ishizawa, Shin
   Hondoh, Hiroaki
   Nakano, Masakazu
   Ogawa, Seishi
   Tashiro, Kei
   Yamanaka, Ryuya
TI Differential expression of individual transcript variants of PD-1 and
   PD-L2 genes on Th-1/Th-2 status is guaranteed for prognosis prediction
   in PCNSL
SO SCIENTIFIC REPORTS
LA English
DT Article
ID CENTRAL-NERVOUS-SYSTEM; LIQUID BIOPSIES; RISK SCORE; LYMPHOMAS;
   BLOCKADE; CELLS; RESISTANCE; MUTATIONS; NIVOLUMAB; LIGANDS
AB In current molecular medicine, next-generation sequencing (NGS) for transcript variant detection and multivariable analyses are valid methods for evaluating gene expression, cancer mechanisms, and prognoses of patients. We conducted RNA-sequencing on samples from patients with primary central nervous system lymphoma (PCNSL) using NGS and performed multivariable analysis on gene expression data and correlations focused on Th-1/Th-2 helper T cell balance and immune checkpoint to identify diagnosis/prognosis markers and cancer immune pathways in PCNSL. We selected 84 transcript variants to limit the analysis range for Th-1/Th-2 balance and stimulatory and inhibitory checkpoints in 31 PCNSLs. Of these, 21 highly-expressed transcript variants were composed of the formulas for prognoses based on Th-1/Th-2 status and checkpoint activities. Using formulas, Th-1(low), Th-2(high), and stimulatory checkpoint(high) resulted in poor prognoses. Further, Th-1(high)Th-2(low) was associated with good prognoses. On the other hand, CD40-001(high) and CD70-001(high) as stimulatory genes, and LAG3-001(high), PDCD1 (PD-1)-001/002/003(high), and PDCD1LG2 (PD-L2)-201(low) as inhibitory genes were associated with poor prognoses. Interestingly, Th-1(high)Th-2(low) and Th-1(low)Th-2(high) were correlated with stimulatory checkpoint(low) as CD70-001(low) and inhibitory checkpointlow as HAVCR2 (TIM-3)-001(low) and PDCD1LG2-001/201(low), respectively. Focused on the inhibitory checkpoint, specific variants of CD274 (PD-L1)-001 and PDCD1-002 served severe hazard ratios. In particular, PDCD1-002(high) by a cut off score was associated with poor prognoses, in addition to PDCD1-001/003(high), PDCD1LG2-201(low), and LAG3-001(high). These results mainly suggest that expression of transcript variants of PDCD1 and PDCD1LG2 on the Th-1/Th-2 balance enable prognostic prediction in PCNSL. This study provides insights for development of molecular target therapies and identification of diagnosis/prognosis markers in PCNSL.
C1 [Takashima, Yasuo; Hayano, Azusa; Yamanaka, Ryuya] Kyoto Prefectural Univ Med, Grad Sch Med Sci, Lab Mol Target Therapy Canc, Kyoto, Japan.
   [Kawaguchi, Atsushi] Saga Univ, Ctr Comprehens Community Med, Fac Med, Saga, Japan.
   [Sato, Ryuichi; Nakano, Masakazu; Tashiro, Kei] Kyoto Prefectural Univ Med, Grad Sch Med Sci, Dept Genom Med Sci, Kyoto, Japan.
   [Yoshida, Kenichi; Ogawa, Seishi] Kyoto Univ, Grad Sch Med, Dept Pathol & Tumor Biol, Kyoto, Japan.
   [Homma, Jumpei; Hondoh, Hiroaki] Toyama Prefectural Cent Hosp, Dept Neurosurg, Toyama, Japan.
   [Fukai, Junya] Wakayama Med Univ, Dept Neurol Surg, Sch Med, Wakayama, Japan.
   [Iwadate, Yasuo] Chiba Univ, Grad Sch Med Sci, Dept Neurosurg, Chiba, Japan.
   [Kajiwara, Koji] Yamaguchi Univ, Grad Sch Med Sci, Dept Neurosurg, Ube, Yamaguchi, Japan.
   [Ishizawa, Shin] Toyama Prefectural Cent Hosp, Dept Pathol, Toyama, Japan.
C3 Kyoto Prefectural University of Medicine; Saga University; Kyoto
   Prefectural University of Medicine; Kyoto University; Wakayama Medical
   University; Chiba University; Yamaguchi University
RP Yamanaka, R (corresponding author), Kyoto Prefectural Univ Med, Grad Sch Med Sci, Lab Mol Target Therapy Canc, Kyoto, Japan.
EM
RI Ogawa, Seishi/AAE-7088-2019; Yamanaka, Ryuya/AFL-0782-2022
OI Yamanaka, Ryuya/0000-0003-2941-7996; Takashima,
   Yasuo/0000-0001-8876-6673
FU MEXT KAKENHI [16H05441, 18K09001]; Grants-in-Aid for Scientific Research
   [18K09001, 16H05441] Funding Source: KAKEN
FX This study was supported by the MEXT KAKENHI Grant Numbers 16H05441 and
   18K09001.
CR Andrews LP, 2017, IMMUNOL REV, V276, P80, DOI 10.1111/imr.12519
   Ansell SM, 2015, NEW ENGL J MED, V372, P311, DOI 10.1056/NEJMoa1411087
   Ansell SM, 2009, CLIN CANCER RES, V15, P6446, DOI 10.1158/1078-0432.CCR-09-1339
   Armand P, 2013, J CLIN ONCOL, V31, P4199, DOI 10.1200/JCO.2012.48.3685
   Batlevi CL, 2016, NAT REV CLIN ONCOL, V13, P25, DOI 10.1038/nrclinonc.2015.187
   Berghoff AS, 2014, CLIN NEUROPATHOL, V33, P42, DOI 10.5414/NP300698
   Burger JA, 2009, BLOOD, V114, P3367, DOI 10.1182/blood-2009-06-225326
   Cani AK, 2017, ONCOTARGET, V8, P7989, DOI 10.18632/oncotarget.14008
   Chandraratna RAS, 2016, AM J TRANSL RES, V8, P1016
   Chapuy B, 2016, BLOOD, V127, P869, DOI 10.1182/blood-2015-10-673236
   Cho H, 2017, ONCOTARGET, V8, P87317, DOI 10.18632/oncotarget.20264
   Cui CG, 2019, CLIN EXP PHARMACOL P, V46, P3, DOI 10.1111/1440-1681.13028
   Fontanilles M, 2017, ONCOTARGET, V8, P48157, DOI 10.18632/oncotarget.18325
   Friedman J, 2008, BIOSTATISTICS, V9, P432, DOI 10.1093/biostatistics/kxm045
   Goecke IA, 2007, MOL IMMUNOL, V44, P2115, DOI 10.1016/j.molimm.2006.07.303
   Hayano A, 2017, ANTICANCER RES, V37, P5655, DOI 10.21873/anticanres.12001
   Hiemcke-Jiwa LS, 2018, HEMATOL ONCOL, V36, P429, DOI 10.1002/hon.2489
   Illerhaus G, 2018, BLOOD, V132, P681, DOI 10.1182/blood-2018-01-791558
   Jenkins RW, 2018, BRIT J CANCER, V118, P9, DOI 10.1038/bjc.2017.434
   Kawaguchi A, 2013, CANCER SCI, V104, P1205, DOI 10.1111/cas.12214
   Kawaguchi A, 2012, CLIN CANCER RES, V18, P5672, DOI 10.1158/1078-0432.CCR-12-0596
   Kim HD, 2018, GASTROENTEROLOGY, V155, P1936, DOI 10.1053/j.gastro.2018.08.030
   Kumar D, 2018, FRONT ONCOL, V8, DOI 10.3389/fonc.2018.00288
   Louis DN, 2016, ACTA NEUROPATHOL, V131, P803, DOI 10.1007/s00401-016-1545-1
   Louis DN OH., 2016, World Health Organization classification of tumours of the Central Nervous System Revised
   Ma C, 2017, J CLIN EXP HEMATOP, V57, P21, DOI 10.3960/jslrt.17006
   Meinshausen N, 2006, ANN STAT, V34, P1436, DOI 10.1214/009053606000000281
   Miyasato Y, 2018, J CLIN EXP HEMATOP, V58, P95, DOI 10.3960/jslrt.18001
   Moreira A, 2019, INT J CANCER, V144, P1147, DOI 10.1002/ijc.31763
   Nayak L, 2017, BLOOD, V129, P3071, DOI 10.1182/blood-2017-01-764209
   Panda A, 2018, JCI INSIGHT, V3, DOI 10.1172/jci.insight.121522
   Parsons E, 2016, PLOS ONE, V11, DOI 10.1371/journal.pone.0167841
   Ricard D, 2012, LANCET, V379, P1984, DOI 10.1016/S0140-6736(11)61346-9
   Rodig SJ, 2018, SCI TRANSL MED, V10, DOI 10.1126/scitranslmed.aar3342
   Rushworth D, 2015, J BIOL CHEM, V290, P22970, DOI 10.1074/jbc.C115.671123
   Sato Y, 2013, NAT GENET, V45, P860, DOI 10.1038/ng.2699
   Sharma P, 2017, CELL, V168, P707, DOI 10.1016/j.cell.2017.01.017
   Shu CC, 2017, RESP RES, V18, DOI 10.1186/s12931-017-0580-3
   Sui HS, 2018, J IMMUNOL RES, V2018, DOI 10.1155/2018/6984948
   Takashima Y, 2018, CANCER SCI, V109, P3543, DOI 10.1111/cas.13798
   Takashima Yasuo, 2018, Oncotarget, V9, P27471, DOI 10.18632/oncotarget.25463
   Takashima Yasuo, 2018, Oncotarget, V9, P19065, DOI 10.18632/oncotarget.24897
   Terziev D, 2018, EUR J HAEMATOL, V101, P115, DOI 10.1111/ejh.13072
   Tsukamoto T, 2017, SCI REP-UK, V7, DOI 10.1038/s41598-017-14150-0
   Wang RYR, 2011, PLOS ONE, V6, DOI 10.1371/journal.pone.0022060
   Wang ZJ, 2018, CANCER RES, V78, P6486, DOI 10.1158/0008-5472.CAN-18-1814
   Xu XD, 2005, CELL, V120, P59, DOI 10.1016/j.cell.2004.11.036
   Yamanaka R., 2016, PRIMARY CENTRAL NERV, P175
   Zou WP, 2006, NAT REV IMMUNOL, V6, P295, DOI 10.1038/nri1806
   Zuo MX, 2018, LEUKEMIA RES, V70, P56, DOI 10.1016/j.leukres.2018.05.008
NR 50
TC 26
Z9 27
U1 0
U2 12
PU NATURE PORTFOLIO
PI BERLIN
PA HEIDELBERGER PLATZ 3, BERLIN, 14197, GERMANY
SN 2045-2322
J9 SCI REP-UK
JI Sci Rep
PD JUL 10
PY 2019
VL 9
AR 10004
DI 10.1038/s41598-019-46473-5
PG 13
WC Multidisciplinary Sciences
WE Science Citation Index Expanded (SCI-EXPANDED)
SC Science & Technology - Other Topics
GA IH8CH
UT WOS:000474731800005
PM 31292525
OA Green Published, gold
DA 2024-06-03
ER

PT J
AU Cheng, G
   Zhang, JN
AF Cheng, Gang
   Zhang, Jianning
TI Imaging features (CT, MRI, MRS, and PET/CT) of primary central nervous
   system lymphoma in immunocompetent patients
SO NEUROLOGICAL SCIENCES
LA English
DT Article
DE Primary central nervous system lymphoma; Non-Hodgkin lymphomas; Imaging
ID PRIMARY CNS LYMPHOMA; MAGNETIC-RESONANCE-SPECTROSCOPY; AIDS; NORWAY
AB Background Because of the low incidence of primary central nervous system lymphoma (PCNSL) in non-HIV individuals and because of the lack of specific clinical manifestations and auxiliary examinations, the disease is easily missed or misdiagnosed.
   Objective To analyze the imaging features of PCNSL in non-HIV patients.
   Methods This was a retrospective study of patients with PCNSL treated between January 2001 and December 2011 at the Naval General Hospital (Beijing, China). All included patients were pathologically diagnosed with PCNSL. Specimens were obtained by stereotactic biopsy and diagnosed by pathological examination. Serological panel had to be negative for HIV.
   Results Out of the 118 patients, 73 (61.9%) were male and 45 (38.1%) were female. Median age was 54 (range 11-83) years. All patients had B cell lymphoma. The lesions showed slightly hyperintense shadows on computed tomography (CT) images, and mostly hyperintense T1 and iso- or hyperintense T2 signals on magnetic resonance imaging (MRI). Most lesions showed patchy enhancement after enhanced scanning, and some had the characteristic "butterfly sign" on enhanced MRI. The magnetic resonance spectroscopy of PCNSL manifested as increased Cho peak, moderately decreased NAA peak, and slightly decreased Cr peak. Positron emission computed tomography indicated high metabolism of F-18-FDG in PCNSL lesions.
   Conclusion MRI is important in the diagnosis of PCNSL. Understanding the imaging features of PCNSL will help improve its diagnosis in clinics.
C1 [Cheng, Gang; Zhang, Jianning] Navy Gen Hosp, Inst Neurosurg, Beijing 100048, Peoples R China.
C3 Sixth Medical Center of Chinese PLA General Hospital
RP Zhang, JN (corresponding author), Navy Gen Hosp, Inst Neurosurg, Beijing 100048, Peoples R China.
EM
FU National Natural Science Foundation [81572899]
FX This study was supported by the National Natural Science Foundation (No.
   81572899).
CR Algazi AP, 2009, NEUROTHERAPEUTICS, V6, P587, DOI 10.1016/j.nurt.2009.04.013
   Barajas RF, 2010, AM J NEURORADIOL, V31, P60, DOI 10.3174/ajnr.A1750
   Berti V, 2014, PET CLIN, V9, P129, DOI 10.1016/j.cpet.2013.10.006
   BIZZI A, 1995, RADIOLOGY, V194, P271, DOI 10.1148/radiology.194.1.7997566
   Coulon A, 2002, EUR RADIOL, V12, P329, DOI 10.1007/s003300101037
   Eichler April F, 2006, Neurosurg Focus, V21, pE15
   Ferreri AJM, 2003, J CLIN ONCOL, V21, P2407, DOI 10.1200/JCO.2003.01.135
   Fraser E, 2015, CHIN CLIN ONCOL, V4, DOI 10.3978/j.issn.2304-3865.2015.02.01
   González-Aguilar A, 2011, CURR OPIN ONCOL, V23, P648, DOI 10.1097/CCO.0b013e32834b6adc
   Guo AC, 2002, RADIOLOGY, V224, P177, DOI 10.1148/radiol.2241010637
   Haldorsen IS, 2011, AM J NEURORADIOL, V32, P984, DOI 10.3174/ajnr.A2171
   Haldorsen IS, 2009, AM J NEURORADIOL, V30, P744, DOI 10.3174/ajnr.A1447
   Haldorsen IS, 2007, CANCER, V110, P1803, DOI 10.1002/cncr.22989
   Harting I, 2003, NEUROSCI LETT, V342, P163, DOI 10.1016/S0304-3940(03)00272-6
   Hochberg FH, 2007, NAT CLIN PRACT NEURO, V3, P24, DOI 10.1038/ncpneuro0395
   Horger M, 2009, AM J ROENTGENOL, V193, P1384, DOI 10.2214/AJR.09.2486
   Karantanis D, 2007, NUCL MED COMMUN, V28, P834, DOI 10.1097/MNM.0b013e328264ae7f
   Kasamon YL, 2005, HEMATOL ONCOL CLIN N, V19, P665, DOI 10.1016/j.hoc.2005.05.004
   Kawai N, 2010, ANN NUCL MED, V24, P335, DOI 10.1007/s12149-010-0356-z
   Küker W, 2005, J NEURO-ONCOL, V72, P169, DOI 10.1007/s11060-004-3390-7
   Latta S, 2010, CURR ONCOL, V17, P63
   Liao WH, 2009, ACTA RADIOL, V50, P217, DOI 10.1080/02841850802616752
   Mansour A, 2014, CANC IMAGING, V14, P1
   Omuro AMP, 2005, ARCH NEUROL-CHICAGO, V62, P1595, DOI 10.1001/archneur.62.10.1595
   Palmedo H, 2006, EUR J NUCL MED MOL I, V33, P164, DOI 10.1007/s00259-005-1917-6
   Qian LR, 2016, ONCOTARGET, V7, P25783, DOI 10.18632/oncotarget.8370
   Raizer JJ, 2005, J NEURO-ONCOL, V71, P173, DOI 10.1007/s11060-004-1360-8
   Schramm P, 2010, EUR RADIOL, V20, P2482, DOI 10.1007/s00330-010-1817-4
   Soares DP, 2009, CLIN RADIOL, V64, P12, DOI 10.1016/j.crad.2008.07.002
   Tang YZ, 2011, CLIN RADIOL, V66, P768, DOI 10.1016/j.crad.2011.03.006
   Verma A, 2016, BBA CLIN, V5, P170, DOI 10.1016/j.bbacli.2016.04.002
   White NS, 2014, CANCER RES, V74, P4638, DOI 10.1158/0008-5472.CAN-13-3534
   Zacharia TT, 2008, J NEUROIMAGING, V18, P411, DOI 10.1111/j.1552-6569.2007.00231.x
   Zhang D, 2010, KOREAN J RADIOL, V11, P269, DOI 10.3348/kjr.2010.11.3.269
   Zou YR, 2017, ONCOTARGET, V8, P41518, DOI 10.18632/oncotarget.17456
NR 35
TC 26
Z9 30
U1 0
U2 9
PU SPRINGER-VERLAG ITALIA SRL
PI MILAN
PA VIA DECEMBRIO, 28, MILAN, 20137, ITALY
SN 1590-1874
EI 1590-3478
J9 NEUROL SCI
JI Neurol. Sci.
PD MAR
PY 2019
VL 40
IS 3
BP 535
EP 542
DI 10.1007/s10072-018-3669-7
PG 8
WC Clinical Neurology; Neurosciences
WE Science Citation Index Expanded (SCI-EXPANDED)
SC Neurosciences & Neurology
GA HQ6EO
UT WOS:000462505300013
PM 30580380
OA Green Published, hybrid
DA 2024-06-03
ER

PT J
AU Zorofchian, S
   Lu, GR
   Zhu, JJ
   Duose, DY
   Windham, J
   Esquenazi, Y
   Ballester, LY
AF Zorofchian, Soheil
   Lu, Guangrong
   Zhu, Jay-Jiguang
   Duose, Dzifa Y.
   Windham, Justin
   Esquenazi, Yoshua
   Ballester, Leomar Y.
TI Detection of the <i>MYD88</i> p.L265P Mutation in the CSF of a Patient
   With Secondary Central Nervous System Lymphoma
SO FRONTIERS IN ONCOLOGY
LA English
DT Article
DE MYD88; central nervous system lymphoma; liquid biopsy; cerebrospinal
   fluid; digital droplet PCR; CSF
ID CLINICAL-SIGNIFICANCE; PRIMARY BRAIN; CANCER; DISEASE; TUMORS
AB Primary Central Nervous System Lymphoma (PCNSL) and Metastatic (or Secondary) Central Nervous System Lymphoma (MCNSL) are rare central nervous system (CNS) malignancies that exhibit aggressive clinical behavior and have a poor prognosis. The majority of CNS lymphomas are histologically classified as diffuse large-B cell lymphoma (DLBCL). DLBCL harbors a high frequency of mutations in MYD88 and CD79b. The MYD88 p.L265P mutation occurs at high frequency in CNS lymphoma and is extremely rare in non-hematologic malignancies. Currently, brain biopsy is considered the gold standard for CNS lymphoma diagnosis. However, brain biopsy is invasive, carries a risk of complications, and can delay initiation of systemic therapy. Circulating tumor DNA (ctDNA) in the cerebrospinal fluid (CSF) can be utilized to detect tumor-derived mutations. Testing of CSF-ctDNA is a minimally-invasive methodology that can be used to assess the genomic alterations present in CNS malignancies. We present a case of an 82-year-old man with a history of testicular lymphoma who presented with speech difficulty and a multifocal enhancing left inferior frontal mass. Analysis for both CSF-cytology and flow cytometry did not show evidence of neoplastic cells. A brain biopsy was performed and microscopic examination showed DLBCL. We isolated CSF-ctDNA and used droplet digital PCR (ddPCR) to detect the most common lymphoma-associated mutations in MYD88, L265P, and V217F. In conjunction, we evaluated the patient-matched CNS lymphoma tissue for MYD88 mutations. We detected the MYD88 p.L265P mutation in formalin fixed paraffin embedded (FFPE) tissue from the brain biopsy and the CSF-ctDNA. In contrast, both the tumor tissue and the CSF ctDNA were negative for the MYD88 p.V217F mutation. This study shows that testing CSF ctDNA for MYD88 mutations is a potentially minimally-invasive approach to diagnosing patients with suspected CNS lymphomas.
C1 [Zorofchian, Soheil; Ballester, Leomar Y.] Univ Texas Hlth Sci Ctr Houston, Dept Pathol & Lab Med, Houston, TX 77030 USA.
   [Lu, Guangrong; Zhu, Jay-Jiguang; Esquenazi, Yoshua; Ballester, Leomar Y.] Univ Texas Hlth Sci Ctr Houston, Vivian L Smith Dept Neurosurg, Houston, TX 77030 USA.
   [Duose, Dzifa Y.; Windham, Justin] UT MD Anderson Canc Ctr, Dept Translat Mol Pathol, Houston, TX USA.
C3 University of Texas System; University of Texas Health Science Center
   Houston; University of Texas System; University of Texas Health Science
   Center Houston; University of Texas System; UTMD Anderson Cancer Center
RP Ballester, LY (corresponding author), Univ Texas Hlth Sci Ctr Houston, Dept Pathol & Lab Med, Houston, TX 77030 USA.; Esquenazi, Y; Ballester, LY (corresponding author), Univ Texas Hlth Sci Ctr Houston, Vivian L Smith Dept Neurosurg, Houston, TX 77030 USA.
EM
RI Esquenazi, Yoshua/AAG-6576-2021
OI Duose, Dzifa/0000-0002-6763-9736
CR Ballester LY, 2018, J NEUROPATH EXP NEUR, V77, P628, DOI 10.1093/jnen/nly046
   Behin A, 2003, LANCET, V361, P323, DOI 10.1016/S0140-6736(03)12328-8
   Braggio E, 2015, CLIN CANCER RES, V21, P3986, DOI 10.1158/1078-0432.CCR-14-2116
   Chamberlain MC, 2009, SEMIN ONCOL, V36, pS35, DOI 10.1053/j.seminoncol.2009.05.005
   Forbes SA, 2017, NUCLEIC ACIDS RES, V45, pD777, DOI 10.1093/nar/gkw1121
   Glantz MJ, 1998, CANCER, V82, P733, DOI 10.1002/(SICI)1097-0142(19980215)82:4<733::AID-CNCR17>3.0.CO;2-Z
   Haldorsen IS, 2007, CANCER, V110, P1803, DOI 10.1002/cncr.22989
   Hattori K, 2018, CANCER SCI, V109, P225, DOI 10.1111/cas.13450
   Hiemcke-Jiwa LS, 2018, HEMATOL ONCOL, V36, P429, DOI 10.1002/hon.2489
   Jeelall YS, 2011, IMMUNOL CELL BIOL, V89, P659, DOI 10.1038/icb.2011.31
   Kickingereder P, 2014, RADIOLOGY, V272, P843, DOI 10.1148/radiol.14132740
   Korfel A, 2013, NAT REV NEUROL, V9, P317, DOI 10.1038/nrneurol.2013.83
   Kraan W, 2013, BLOOD CANCER J, V3, DOI 10.1038/bcj.2013.28
   Lee JH, 2017, SCI REP-UK, V7, DOI 10.1038/s41598-017-01998-5
   Mehrotra Meenakshi, 2018, Oncotarget, V9, P10259, DOI 10.18632/oncotarget.21982
   Mohile NA, 2007, SEMIN RADIAT ONCOL, V17, P223, DOI 10.1016/j.semradonc.2007.02.008
   Nayak L, 2013, CURR TREAT OPTION ON, V14, P539, DOI 10.1007/s11864-013-0252-6
   Ostrom QT, 2017, NEURO-ONCOLOGY, V19, pV1, DOI 10.1093/neuonc/nox158
   Shalaby T., 2016, JCMT, V2016, P176, DOI DOI 10.20517/2394-4722.2016.12
   van der Sanden GAC, 2002, CANCER, V94, P1548, DOI 10.1002/cncr.10357
   Varettoni M, 2013, BLOOD, V121, P2522, DOI 10.1182/blood-2012-09-457101
   Vater I, 2015, LEUKEMIA, V29, P677, DOI 10.1038/leu.2014.264
   Weston CL, 2011, FLUIDS BARRIERS CNS, V8, DOI 10.1186/2045-8118-8-14
   Wong ET, 2005, CLIN CANCER RES, V11, p7151S, DOI 10.1158/1078-0432.CCR-1004-0002
NR 24
TC 26
Z9 26
U1 4
U2 10
PU FRONTIERS MEDIA SA
PI LAUSANNE
PA AVENUE DU TRIBUNAL FEDERAL 34, LAUSANNE, CH-1015, SWITZERLAND
SN 2234-943X
J9 FRONT ONCOL
JI Front. Oncol.
PD SEP 20
PY 2018
VL 8
AR 382
DI 10.3389/fonc.2018.00382
PG 6
WC Oncology
WE Science Citation Index Expanded (SCI-EXPANDED)
SC Oncology
GA GU2FX
UT WOS:000445085900001
PM 30294590
OA gold, Green Published
DA 2024-06-03
ER

PT J
AU Strehlow, F
   Bauer, S
   Martus, P
   Weller, M
   Roth, P
   Schlegel, U
   Seidel, S
   Scheibenbogen, C
   Korfel, A
   Kreher, S
AF Strehlow, Felicitas
   Bauer, Sandra
   Martus, Peter
   Weller, Michael
   Roth, Patrick
   Schlegel, Uwe
   Seidel, Sabine
   Scheibenbogen, Carmen
   Korfel, Agnieszka
   Kreher, Stephan
TI Osteopontin in cerebrospinal fluid as diagnostic biomarker for central
   nervous system lymphoma
SO JOURNAL OF NEURO-ONCOLOGY
LA English
DT Article
DE Osteopontin; Cerebrospinal fluid; PCNSL; SCNSL
ID PRIMARY CNS LYMPHOMA; NON-HODGKINS-LYMPHOMA; MULTIPLE-SCLEROSIS; SERUM
   OSTEOPONTIN; RECURRENCE; LEVEL; CSF; INTERLEUKIN-10; INVOLVEMENT;
   EXPRESSION
AB Central nervous system lymphoma (CNSL) is diagnostically challenging. The identification of reliable and easy to measure biomarkers is desirable to facilitate diagnosis. Here, we evaluated the value of cerebrospinal fluid (CSF) osteopontin (OPN) as a diagnostic biomarker for CNSL. OPN concentrations in CSF from 37 patients with CNSL (29 with primary CNSL and 8 with secondary CNS involvement of systemic lymphoma) and 36 controls [6 patients with inflammatory CNS disease other than multiple sclerosis (MS), 8 with MS, 9 with glioblastoma (GBM) and 13 healthy controls] were determined using an enzyme-linked immunosorbent assay. Non-parametric tests and receiver operating characteristic (ROC) curves were performed for determination of diagnostic accuracy. Median CSF OPN level in all CNSL patients was 620 ng/mL and higher than in patients with inflammatory CNS disease (356 ng/mL); P < .05, MS (163 ng/mL); P < .01, GBM (41 ng/mL); P < .01, or healthy controls (319 ng/mL); P < .01. The area under the ROC curve was 0.865 [95 % confidence interval (CI) 0.745-0.985] for differentiating CNSL and patients with inflammatory CNS disease; 0.956 (95 % CI 0.898-1.000) for CNSL and MS patients; 0.988 (95 % CI 0.964-1.000) for CNSL and GBM patients, and 0.915 (95 % CI 0.834-0.996) for CNSL patients and healthy controls. In multivariate analysis, high CSF OPN level was associated with shorter progression-free (HR 1.61, 95 % CI 1.13-2.31; P = .009) and overall survival (HR 1.52, 95 % CI 1.04-2.21; P = .029). CSF OPN is a potential biomarker in CNSL.
C1 [Strehlow, Felicitas; Korfel, Agnieszka; Kreher, Stephan] Charite, Dept Hematol Oncol & Tumour Immunol, Campus Benjamin Franklin,Hindenburgdamm 30, D-12200 Berlin, Germany.
   [Bauer, Sandra; Scheibenbogen, Carmen] Charite, Dept Med Immunol, Berlin, Germany.
   [Martus, Peter] Univ Tubingen, Inst Clin Epidemiol & Appl Biostat, Tubingen, Germany.
   [Weller, Michael; Roth, Patrick] Univ Zurich Hosp, Dept Neurol, Zurich, Switzerland.
   [Schlegel, Uwe; Seidel, Sabine] Univ Hosp Bochum, Dept Neurol, Knappschaftskrankenhaus, Bochum, Germany.
C3 Free University of Berlin; Humboldt University of Berlin; Charite
   Universitatsmedizin Berlin; Free University of Berlin; Humboldt
   University of Berlin; Charite Universitatsmedizin Berlin; Eberhard Karls
   University of Tubingen; University of Zurich; University Zurich
   Hospital; Ruhr University Bochum
RP Kreher, S (corresponding author), Charite, Dept Hematol Oncol & Tumour Immunol, Campus Benjamin Franklin,Hindenburgdamm 30, D-12200 Berlin, Germany.
EM
RI McDermott, Michael Francis/GZM-5330-2022; Roth, Patrick/R-6671-2018;
   Seidel, Sabine/HMW-0156-2023
OI Scheibenbogen, Carmen/0000-0002-8554-9638; Bauer,
   Sandra/0000-0002-3497-4730; Weller, Michael/0000-0002-1748-174X
CR Baraniskin A, 2012, J NEURO-ONCOL, V109, P239, DOI 10.1007/s11060-012-0908-2
   Baraniskin A, 2012, BRIT J HAEMATOL, V156, P421, DOI 10.1111/j.1365-2141.2011.08928.x
   Baraniskin A, 2011, BLOOD, V117, P3140, DOI 10.1182/blood-2010-09-308684
   Bessell EM, 1996, J CLIN ONCOL, V14, P945, DOI 10.1200/JCO.1996.14.3.945
   Boehme V, 2007, ANN ONCOL, V18, P149, DOI 10.1093/annonc/mdl327
   Börnsen L, 2011, MULT SCLER J, V17, P32, DOI 10.1177/1352458510382247
   Braitch M, 2008, ARCH NEUROL-CHICAGO, V65, P633, DOI 10.1001/archneur.65.5.633
   Bromberg JE, 2013, HAEMATOLOGICA, V98, P808, DOI 10.3324/haematol.2012.070839
   Brown A, 2012, TRANSL NEUROSCI, V3, P288, DOI 10.2478/s13380-012-0028-7
   Chowdhury SA, 2008, ARCH NEUROL-CHICAGO, V65, P232, DOI 10.1001/archneurol.2007.33
   Dai JX, 2010, CANCER IMMUNOL IMMUN, V59, P355, DOI 10.1007/s00262-009-0754-z
   Deckert M, 2011, LEUKEMIA, V25, P1797, DOI 10.1038/leu.2011.169
   Edwards LJ, 2013, AM J CLIN EXP IMMUNO, V2, P234
   Fischer L, 2009, CLIN CANCER RES, V15, P5968, DOI 10.1158/1078-0432.CCR-09-0108
   Gleissner B, 2002, NEUROLOGY, V58, P390, DOI 10.1212/WNL.58.3.390
   Han SS, 2013, TUBERC RESP DIS, V75, P104, DOI 10.4046/trd.2013.75.3.104
   Han YP, 2014, J NEURO-ONCOL, V119, P343, DOI 10.1007/s11060-014-1484-4
   Hollender A, 2002, ANN ONCOL, V13, P1099, DOI 10.1093/annonc/mdf175
   Incesoy-Özdemir S, 2013, TURKISH J PEDIATR, V55, P42
   Jiang LY, 2010, INT J CLIN EXP PATHO, V3, P763
   Korfel A, 2012, ANN ONCOL, V23, P2374, DOI 10.1093/annonc/mdr627
   Korfel A, 2013, NAT REV NEUROL, V9, P317, DOI 10.1038/nrneurol.2013.83
   Li Y, 2012, MED ONCOL, V29, P1429, DOI 10.1007/s12032-011-0012-z
   Megaptche AP, 2014, IMMUNOL CELL BIOL, V92, P709, DOI 10.1038/icb.2014.47
   Poruk KE, 2013, PANCREAS, V42, P193, DOI 10.1097/MPA.0b013e31825e354d
   Rubenstein JL, 2013, BLOOD, V121, P4740, DOI 10.1182/blood-2013-01-476333
   Rubenstein JL, 2009, BLOOD, V113, P266, DOI 10.1182/blood-2008-04-152835
   Rud AK, 2013, BMC CANCER, V13, DOI 10.1186/1471-2407-13-540
   Sasayama T, 2012, NEURO-ONCOLOGY, V14, P368, DOI 10.1093/neuonc/nor203
   Schroers R, 2010, EUR J HAEMATOL, V85, P520, DOI 10.1111/j.1600-0609.2010.01516.x
   Shevde LA, 2014, MATRIX BIOL, V37, P131, DOI 10.1016/j.matbio.2014.03.001
   Szalardy L, 2013, J NEUROL SCI, V331, P38, DOI 10.1016/j.jns.2013.04.024
   Tun HW, 2008, BLOOD, V111, P3200, DOI 10.1182/blood-2007-10-119099
   Viaccoz A, 2015, NEURO-ONCOLOGY, V17, P1497, DOI 10.1093/neuonc/nov092
   Weller M, 2012, NEURO-ONCOLOGY, V14, P1481, DOI 10.1093/neuonc/nos159
   Wen SR, 2012, J NEUROIMMUNOL, V244, P94, DOI 10.1016/j.jneuroim.2011.12.004
   Yamaguchi Y, 2013, J BIOL CHEM, V288, P3097, DOI 10.1074/jbc.M112.362954
   Yuan J, 2013, HUM PATHOL, V44, P606, DOI 10.1016/j.humpath.2012.07.007
   Zhou C, 2013, ANN SURG ONCOL, V20, P929, DOI 10.1245/s10434-012-2749-9
NR 39
TC 26
Z9 27
U1 1
U2 9
PU SPRINGER
PI NEW YORK
PA 233 SPRING ST, NEW YORK, NY 10013 USA
SN 0167-594X
EI 1573-7373
J9 J NEURO-ONCOL
JI J. Neuro-Oncol.
PD AUG
PY 2016
VL 129
IS 1
BP 165
EP 171
DI 10.1007/s11060-016-2162-5
PG 7
WC Oncology; Clinical Neurology
WE Science Citation Index Expanded (SCI-EXPANDED)
SC Oncology; Neurosciences & Neurology
GA DT0DE
UT WOS:000381152000019
PM 27294357
OA Green Accepted
DA 2024-06-03
ER

PT J
AU Bathla, G
   Hegde, A
AF Bathla, G.
   Hegde, A.
TI Lymphomatous involvement of the central nervous system
SO CLINICAL RADIOLOGY
LA English
DT Review
ID PRIMARY CNS LYMPHOMA; MRI FINDINGS; CT; AIDS
AB Lymphoma may arise within the central nervous system (CNS), known as primary CNS lymphoma (PCNSL) or may involve the CNS secondary to systemic disease. Clinical features are non-specific. A provisional diagnosis of PCNSL can be blade on imaging, potentially changing the management algorithm from neurosurgical resection to biopsy. PCNSL in immunocompetent patients generally presents late, is solid, is bright on diffusion weighted imaging and shows uniform enhancement. Contiguity with a cerebrospinal fluid (CSF) surface and perivascular enhancement are useful clues. Immunocompromised patients, on the other hand, present earlier and often have multiple, necrotic, haemorrhagic lesions with irregular or rim enhancement. Secondary CNS involvement predominantly affects the leptomeninges. This review illustrates the varied imaging features of CNS lymphoma, atypical presentations, and differential diagnoses. (C) 2016 Published by Elsevier Ltd on behalf of The Royal College of Radiologists.
C1 [Bathla, G.] Univ Iowa Hosp & Clin, Dept Radiol, 200 Hawkins Dr, Iowa City, IA 52242 USA.
   [Hegde, A.] Raffles Hosp, Dept Radiol, 585 North Bridge Dr, Singapore 188770, Singapore.
C3 University of Iowa; Raffles Hospital
RP Hegde, A (corresponding author), Raffles Hosp, Dept Radiol, 585 North Bridge Dr, Singapore 188770, Singapore.
EM
OI BATHLA, GIRISH/0000-0003-3170-242X
CR Adachi K, 2013, J NIPPON MED SCH, V80, P174, DOI 10.1272/jnms.80.174
   CHANG L, 1995, RADIOLOGY, V197, P525, DOI 10.1148/radiology.197.2.7480706
   CHINN RJS, 1995, RADIOLOGY, V197, P649, DOI 10.1148/radiology.197.3.7480733
   Coulon A, 2002, EUR RADIOL, V12, P329, DOI 10.1007/s003300101037
   Erdag N, 2001, AM J ROENTGENOL, V176, P1319, DOI 10.2214/ajr.176.5.1761319
   Haldorsen IS, 2011, AM J NEURORADIOL, V32, P984, DOI 10.3174/ajnr.A2171
   Haldorsen IS, 2007, CANCER, V110, P1803, DOI 10.1002/cncr.22989
   Iwamoto FM, 2006, NEUROLOGY, V66, P1763, DOI 10.1212/01.wnl.0000218284.23872.eb
   Jayaraman MV, 2009, MAGNETIC RESONANCE I, P525
   Küker W, 2005, J NEURO-ONCOL, V72, P169, DOI 10.1007/s11060-004-3390-7
   Mansour A, 2014, CANCER IMAGING, V14, DOI 10.1186/1470-7330-14-22
   OGAWA T, 1994, RADIOGRAPHICS, V14, P101, DOI 10.1148/radiographics.14.1.8128041
   Opelz G, 2004, AM J TRANSPLANT, V4, P222, DOI 10.1046/j.1600-6143.2003.00325.x
   Partovi S, 2014, BRIT J RADIOL, V87, DOI 10.1259/bjr.20130684
   Phan TG, 2000, NEURO-ONCOLOGY, V2, P229, DOI 10.1093/neuonc/2.4.229
   Raz E, 2011, J NEUROIMAGING, V21, pe183, DOI 10.1111/j.1552-6569.2010.00477.x
   REMICK SC, 1990, MEDICINE, V69, P345, DOI 10.1097/00005792-199011000-00003
   Ruiz A, 1997, NEUROIMAG CLIN N AM, V7, P281
   Slone HW, 2005, AM J ROENTGENOL, V184, P1679, DOI 10.2214/ajr.184.5.01841679
   Tang YZ, 2011, CLIN RADIOL, V66, P768, DOI 10.1016/j.crad.2011.03.006
   Taylor JW, 2013, NEUROLOGY, V81, P1690, DOI 10.1212/01.wnl.0000435302.02895.f3
   Thomas AG, 2011, AM J ROENTGENOL, V197, P350, DOI 10.2214/AJR.10.7266
   Thurnher MM, 2001, NEURORADIOLOGY, V43, P29, DOI 10.1007/s002340000480
   van der Sanden GAC, 2002, CANCER, V94, P1548, DOI 10.1002/cncr.10357
   Williams RL, 1998, AM J NEURORADIOL, V19, P427
   Yap KK, 2012, J MED IMAG RADIAT ON, V56, P295
   Zhang D, 2010, KOREAN J RADIOL, V11, P269, DOI 10.3348/kjr.2010.11.3.269
NR 27
TC 26
Z9 30
U1 0
U2 10
PU W B SAUNDERS CO LTD
PI LONDON
PA 32 JAMESTOWN RD, LONDON NW1 7BY, ENGLAND
SN 0009-9260
EI 1365-229X
J9 CLIN RADIOL
JI Clin. Radiol.
PD JUN
PY 2016
VL 71
IS 6
BP 602
EP 609
DI 10.1016/j.crad.2016.02.006
PG 8
WC Radiology, Nuclear Medicine & Medical Imaging
WE Science Citation Index Expanded (SCI-EXPANDED)
SC Radiology, Nuclear Medicine & Medical Imaging
GA DM9PA
UT WOS:000376697000017
PM 27038652
DA 2024-06-03
ER

PT J
AU Jang, JE
   Kim, YR
   Kim, SJ
   Cho, H
   Chung, H
   Lee, JY
   Park, H
   Kim, Y
   Cheong, JW
   Min, YH
   Kim, JS
AF Jang, Ji Eun
   Kim, Yu Ri
   Kim, Soo-Jeong
   Cho, Hyunsoo
   Chung, Haerim
   Lee, Jung Yeon
   Park, Hyunsung
   Kim, Yundeok
   Cheong, June-Won
   Min, Yoo Hong
   Kim, Jin Seok
TI A new prognostic model using absolute lymphocyte count in patients with
   primary central nervous system lymphoma
SO EUROPEAN JOURNAL OF CANCER
LA English
DT Article
DE Primary central nervous system lymphoma; Prognosis; Lymphopenia
ID B-CELL-LYMPHOMA; PRIMARY CNS LYMPHOMA; EPSTEIN-BARR-VIRUS; PREDICTS
   SURVIVAL; HODGKINS-LYMPHOMA; MARKER; CHEMOTHERAPY; EXPRESSION;
   CARCINOMA; THERAPY
AB Purpose: Primary central nervous system lymphoma (PCNSL) is an aggressive and rare extranodal non-Hodgkin lymphoma (NHL). Absolute lymphocyte count (ALC) has been suggested to have a prognostic value in several subtypes of NHL. We evaluated the prognostic significance of clinical factors, including ALC, in patients with PCNSL to develop a new prognostic model.
   Methods: We analysed prognostic factors, including ALC, at diagnosis in 81 PCNSL patients receiving high-dose methotrexate-based therapy.
   Results: The median ALC at diagnosis was 1210 x 10(6)/L (range, 210-3610), with lymphopenia (<= 875 x 10(6)/L) being detected in 27 (33.3%) patients. In the multivariate analysis, Eastern Cooperative Oncology Group performance status (ECOG PS) > 1 (hazard ratio [HR] 3.18, P = 0.003), age > 50 years (HR 4.23, P = 0.012), and lymphopenia at diagnosis (HR 2.83, P = 0.008) remained independent prognostic factors for low overall survival (OS). Lymphopenia was also a significant prognostic factor for progression-free survival (HR 3.17, P = 0.001). By means of a new three-factor prognostic model using ECOG PS > 1, age > 50 years, and presence of lymphopenia, with 1 point assigned to each factor, we successfully classified the patients into three risk groups: low (0 and 1), intermediate (2), and high (3). The 5-year OS rates of the patients in the low-, intermediate-, and high-risk groups were 74.3%, 21.7%, and 12.5%, respectively (P < 0.001).
   Conclusions: Low ALC is a useful indicator of poor prognosis in patients with PCNSL. The proposed three-factor model should be validated in large-scale studies. (C) 2016 Elsevier Ltd. All rights reserved.
C1 [Jang, Ji Eun; Kim, Yu Ri; Kim, Soo-Jeong; Cho, Hyunsoo; Chung, Haerim; Lee, Jung Yeon; Park, Hyunsung; Kim, Yundeok; Cheong, June-Won; Min, Yoo Hong; Kim, Jin Seok] Yonsei Univ, Coll Med, Dept Internal Med, Div Hematol, 50-1 Yonsei Ro, Seoul 120752, South Korea.
C3 Yonsei University; Yonsei University Health System
RP Kim, JS (corresponding author), Yonsei Univ, Coll Med, Severance Hosp, Div Hematol,Dept Internal Med, 50-1 Yonsei Ro, Seoul 120752, South Korea.
EM
RI KIM, JIN SEOK/I-6927-2019; Cho, Hyunsoo/JLR-6939-2023; Kim,
   Ji-Soo/D-8744-2012; Kim, Soo-Jeong/F-1024-2017
OI KIM, JIN SEOK/0000-0001-8986-8436; Kim, Ji-Soo/0000-0002-1508-2024;
   Chung, Haerim/0000-0002-7926-9285; Jang, Ji Eun/0000-0001-8832-1412;
   min, yoo hong/0000-0001-8542-9583; Kim, YUNDEOK/0000-0002-5336-7936;
   Kim, Soo-Jeong/0000-0001-8859-3573; CHEONG,
   JUNE-WON/0000-0002-1744-0921; Cho, Hyunsoo/0000-0003-2651-6403
FU faculty research grant of Yonsei University College of Medicine
   [6-2015-0029]
FX This study was supported by a faculty research grant of Yonsei
   University College of Medicine for 2015 (6-2015-0029).
CR Abrey LE, 2006, J CLIN ONCOL, V24, P5711, DOI 10.1200/JCO.2006.08.2941
   Abrey LE, 2005, J CLIN ONCOL, V23, P5034, DOI 10.1200/JCO.2005.13.524
   Batchelor T, 2006, J CLIN ONCOL, V24, P1281, DOI 10.1200/JCO.2005.04.8819
   Bessell EM, 2004, INT J RADIAT ONCOL, V59, P501, DOI 10.1016/j.ijrobp.2003.11.001
   Burnham KP, 2011, BEHAV ECOL SOCIOBIOL, V65, P23, DOI 10.1007/s00265-010-1029-6
   Cox MC, 2008, BRIT J HAEMATOL, V141, P265, DOI 10.1111/j.1365-2141.2008.07028.x
   Cox MC, 2008, LEUKEMIA LYMPHOMA, V49, P1745, DOI 10.1080/10428190802226425
   Deckert M, 2011, LEUKEMIA, V25, P1797, DOI 10.1038/leu.2011.169
   Dürr C, 2010, CANCER RES, V70, P10170, DOI 10.1158/0008-5472.CAN-10-1943
   Ferreri AJM, 2003, J CLIN ONCOL, V21, P266, DOI 10.1200/JCO.2003.09.139
   Ferreri AJM, 2011, ONCOLOGIST, V16, P336, DOI 10.1634/theoncologist.2010-0361
   Ferreri AJM, 2009, LANCET, V374, P1512, DOI 10.1016/S0140-6736(09)61416-1
   Glatting G, 2007, MED PHYS, V34, P4285, DOI 10.1118/1.2794176
   Hans CP, 2004, BLOOD, V103, P275, DOI 10.1182/blood-2003-05-1545
   Hsu CY, 2010, CANCER-AM CANCER SOC, V116, P3006, DOI 10.1002/cncr.25044
   Ishigami S, 2000, CANCER, V88, P577, DOI 10.1002/(SICI)1097-0142(20000201)88:3<577::AID-CNCR13>3.0.CO;2-V
   Keegan THM, 2005, J CLIN ONCOL, V23, P7604, DOI 10.1200/JCO.2005.02.6310
   Kim Jeong Eun, 2012, Korean J Hematol, V47, P60, DOI 10.5045/kjh.2012.47.1.60
   Kim SH, 2007, ARCH PATHOL LAB MED, V131, P457
   Kim YR, 2014, ANN HEMATOL, V93, P211, DOI 10.1007/s00277-013-1853-7
   Kreher S, 2015, NEURO-ONCOLOGY, V17, P1016, DOI 10.1093/neuonc/nov046
   Oki Y, 2008, EUR J HAEMATOL, V81, P448, DOI 10.1111/j.1600-0609.2008.01129.x
   Ponzoni M, 2007, BRIT J HAEMATOL, V138, P316, DOI 10.1111/j.1365-2141.2007.06661.x
   Roth P, 2015, EUR J CANCER, V51, P382, DOI 10.1016/j.ejca.2014.10.028
   Roychowdhury S, 2003, CANCER RES, V63, P965
   Siddiqui M, 2006, BRIT J HAEMATOL, V134, P596, DOI 10.1111/j.1365-2141.2006.06232.x
   Song MK, 2010, ANN ONCOL, V21, P140, DOI 10.1093/annonc/mdp505
   Suzuki R, 2011, BLOOD, V118, P6018, DOI 10.1182/blood-2011-05-354142
   Talaulikar D, 2008, LEUKEMIA LYMPHOMA, V49, P959, DOI 10.1080/10428190801959026
   Utsuki S, 2011, BRAIN TUMOR PATHOL, V28, P145, DOI 10.1007/s10014-011-0020-x
   Venetz D, 2010, INT J CANCER, V127, P2300, DOI 10.1002/ijc.25236
   Wilcox RA, 2011, LEUKEMIA, V25, P1502, DOI 10.1038/leu.2011.112
   Zou WP, 2005, NAT REV CANCER, V5, P263, DOI 10.1038/nrc1586
NR 33
TC 26
Z9 28
U1 0
U2 2
PU ELSEVIER SCI LTD
PI OXFORD
PA THE BOULEVARD, LANGFORD LANE, KIDLINGTON, OXFORD OX5 1GB, OXON, ENGLAND
SN 0959-8049
EI 1879-0852
J9 EUR J CANCER
JI Eur. J. Cancer
PD APR
PY 2016
VL 57
BP 127
EP 135
DI 10.1016/j.ejca.2016.01.016
PG 9
WC Oncology
WE Science Citation Index Expanded (SCI-EXPANDED)
SC Oncology
GA DG0UD
UT WOS:000371780300015
PM 26918738
DA 2024-06-03
ER

PT J
AU Patrick, LB
   Mohile, NA
AF Patrick, Lauren B.
   Mohile, Nimish A.
TI Advances in Primary Central Nervous System Lymphoma
SO CURRENT ONCOLOGY REPORTS
LA English
DT Review
DE Primary central nervous system lymphoma; Diffuse large B cell lymphoma;
   Ocular lymphoma; Leptomeningeal metastases; Bone marrow transplant;
   Leukoencephalopathy; High-dose methotrexate
ID PRIMARY CNS LYMPHOMA; WHOLE-BRAIN RADIOTHERAPY; HIGH-DOSE METHOTREXATE;
   TERM-FOLLOW-UP; 1ST-LINE TREATMENT; DEFERRED RADIOTHERAPY; CELL
   TRANSPLANTATION; GENE REARRANGEMENT; PHASE-II; CHEMOTHERAPY
AB Primary central nervous system lymphoma (PCNSL) is a rare form of non-Hodgkin lymphoma that is limited to the CNS. Although novel imaging techniques aid in discriminating lymphoma from other brain tumors, definitive diagnosis requires brain biopsy, vitreoretinal biopsy, or cerebrospinal fluid analysis. Survival rates in clinical studies have improved over the past 20 years due to the addition of high-dose methotrexate-based chemotherapy regimens to whole-brain radiotherapy. Long-term survival, however, is complicated by clinically devastating delayed neurotoxicity. Newer regimens are attempting to reduce or eliminate radiotherapy from first-line treatment with chemotherapy dose intensification. Significant advances have also been made in the fields of pathobiology and treatment, with more targeted treatments on the horizon. The rarity of the disease makes conducting of prospective clinical trials challenging, requiring collaborative efforts between institutions. This review highlights recent advances in the biology, detection, and treatment of PCNSL in immunocompetent patients.
C1 [Patrick, Lauren B.; Mohile, Nimish A.] Univ Rochester, Med Ctr, Dept Neurol, Rochester, NY 14642 USA.
   [Mohile, Nimish A.] Univ Rochester, Med Ctr, Dept Neurol, Rochester, NY 14642 USA.
C3 University of Rochester; University of Rochester
RP Mohile, NA (corresponding author), Univ Rochester, Med Ctr, Dept Neurol, 601 Elmwood Ave,Box 356, Rochester, NY 14642 USA.
EM
CR Abrey LE, 2006, J CLIN ONCOL, V24, P5711, DOI 10.1200/JCO.2006.08.2941
   Abrey LE, 1998, J CLIN ONCOL, V16, P859, DOI 10.1200/JCO.1998.16.3.859
   Abrey LE, 2000, J CLIN ONCOL, V18, P3144, DOI 10.1200/JCO.2000.18.17.3144
   Angelov L, 2009, J CLIN ONCOL, V27, P3503, DOI 10.1200/JCO.2008.19.3789
   Baehring JM, 2006, J NEUROL SCI, V247, P208, DOI 10.1016/j.jns.2006.05.044
   Baraniskin A, 2011, BLOOD, V117, P3140, DOI 10.1182/blood-2010-09-308684
   Batchelor T, 2003, J CLIN ONCOL, V21, P1044, DOI 10.1200/JCO.2003.03.036
   Bellinzona M, 2005, EJSO, V31, P100, DOI 10.1016/j.ejso.2004.10.002
   Bierman PJ, 2014, ONCOLOGY-NY, V28, P632
   Braggio E, 2015, CLIN CANC RES
   Bühring U, 2001, NEUROLOGY, V57, P393, DOI 10.1212/WNL.57.3.393
   Camilleri-Broët S, 2006, BLOOD, V107, P190, DOI 10.1182/blood-2005-03-1024
   Chamberlain MC, 2010, NEURO-ONCOLOGY, V12, P736, DOI 10.1093/neuonc/noq011
   CONSTINE LS, 1988, INT J RADIAT ONCOL, V15, P319, DOI 10.1016/S0360-3016(98)90011-6
   Correa DD, 2004, NEUROLOGY, V62, P548, DOI 10.1212/01.WNL.0000109673.75316.D8
   DEANGELIS LM, 1992, J CLIN ONCOL, V10, P635, DOI 10.1200/JCO.1992.10.4.635
   Dolecek TA, 2012, NEURO-ONCOLOGY, V14, pv1, DOI 10.1093/neuonc/nos218
   Ekstein D, 2006, J NEUROL SCI, V247, P39, DOI 10.1016/j.jns.2006.03.012
   Enting RH, 2004, NEUROLOGY, V63, P901, DOI 10.1212/01.WNL.0000137050.43114.42
   Ferreri AJ, 2015, 13 INT C MAL LYMPH J
   Ferreri AJM, 2003, J CLIN ONCOL, V21, P266, DOI 10.1200/JCO.2003.09.139
   Ferreri AJM, 2009, LANCET, V374, P1512, DOI 10.1016/S0140-6736(09)61416-1
   Fischer L, 2008, NEUROLOGY, V71, P1102, DOI 10.1212/01.wnl.0000326958.52546.f5
   Fischer L, 2011, NEURO-ONCOLOGY, V13, P1090, DOI 10.1093/neuonc/nor107
   Gavrilovic IT, 2006, J CLIN ONCOL, V24, P4570, DOI 10.1200/JCO.2006.06.6910
   Gonzalez-Aguilar A, 2012, CLIN CANCER RES, V18, P5203, DOI 10.1158/1078-0432.CCR-12-0845
   Hirono S, 2015, J NEURO-ONCOL, V123, P237, DOI 10.1007/s11060-015-1786-1
   Hoang-Xuan K, 2015, LANCET ONCOL, V16, pE322, DOI 10.1016/S1470-2045(15)00076-5
   Hottinger AF, 2007, NEUROLOGY, V69, P1178, DOI 10.1212/01.wnl.0000276986.19602.c1
   Houillier C, 2015, NEUROLOGY, V84, P325, DOI 10.1212/WNL.0000000000001158
   Illerhaus G, 2006, J CLIN ONCOL, V24, P3865, DOI 10.1200/JCO.2006.06.2117
   Jahnke K, 2006, J CLIN ONCOL, V24, P4754, DOI 10.1200/JCO.2006.06.7165
   Kasenda B, 2012, ANN ONCOL, V23, P2670, DOI 10.1093/annonc/mds059
   Khan RB, 2002, J NEURO-ONCOL, V58, P175, DOI 10.1023/A:1016077907952
   LACHANCE DH, 1994, NEUROLOGY, V44, P1721, DOI 10.1212/WNL.44.9.1721
   Lai R, 2002, NEUROLOGY, V59, P1557, DOI 10.1212/01.WNL.0000034256.20173.EA
   Lai R, 2004, NEUROLOGY, V62, P451, DOI 10.1212/01.WNL.0000106941.51340.A2
   LEIBEL SA, 1987, J NEUROSURG, V66, P1, DOI 10.3171/jns.1987.66.1.0001
   Mappa S, 2013, HEMATOL ONCOL, V31, P143, DOI 10.1002/hon.2037
   Milpied N, 2004, NEW ENGL J MED, V350, P1287, DOI 10.1056/NEJMoa031770
   Mohile NA, 2008, NEURO-ONCOLOGY, V10, P223, DOI 10.1215/15228517-2007-061
   Montesinos-Rongen M, 2011, ACTA NEUROPATHOL, V122, P791, DOI 10.1007/s00401-011-0891-2
   Morris PG, 2013, J CLIN ONCOL, V31, P3971, DOI 10.1200/JCO.2013.50.4910
   Nayak L, 2013, LEUKEMIA LYMPHOMA, V54, P58, DOI 10.3109/10428194.2012.698736
   Olson JE, 2002, CANCER, V95, P1504, DOI 10.1002/cncr.10851
   Omuro AMP, 2005, ARCH NEUROL-CHICAGO, V62, P1595, DOI 10.1001/archneur.62.10.1595
   Omuro A, 2015, BLOOD, V125, P1403, DOI 10.1182/blood-2014-10-604561
   Panageas KS, 2005, CANCER-AM CANCER SOC, V104, P2466, DOI 10.1002/cncr.21481
   Pels H, 2009, J NEURO-ONCOL, V91, P299, DOI 10.1007/s11060-008-9712-4
   Pentsova E, 2014, J NEURO-ONCOL, V117, P161, DOI 10.1007/s11060-014-1370-0
   Reni M, 2001, ANN HEMATOL, V80, pB113
   Rubenstein J, 2008, LEUKEMIA LYMPHOMA, V49, P43, DOI 10.1080/10428190802311441
   Rubenstein JL, 2013, J CLIN ONCOL, V31, P3061, DOI 10.1200/JCO.2012.46.9957
   Rubenstein JL, 2013, BLOOD, V121, P4740, DOI 10.1182/blood-2013-01-476333
   Rubenstein JL, 2006, BLOOD, V107, P3716, DOI 10.1182/blood-2005-03-0897
   Shah GD, 2007, J CLIN ONCOL, V25, P4730, DOI 10.1200/JCO.2007.12.5062
   SHELINE GE, 1980, INT J RADIAT ONCOL, V6, P1215, DOI 10.1016/0360-3016(80)90175-3
   Shibamoto Y, 2003, CANCER-AM CANCER SOC, V97, P128, DOI 10.1002/cncr.11035
   Thiel E, 2010, LANCET ONCOL, V11, P1036, DOI 10.1016/S1470-2045(10)70229-1
   Toh CH, 2013, AM J NEURORADIOL, V34, P1145, DOI 10.3174/ajnr.A3383
   Weller M, 1999, J NEURO-ONCOL, V43, P237, DOI 10.1023/A:1006254518848
   Wong SF, 2012, J CLIN NEUROSCI, V19, P1501, DOI 10.1016/j.jocn.2012.04.001
NR 62
TC 26
Z9 31
U1 0
U2 18
PU SPRINGER
PI NEW YORK
PA 233 SPRING ST, NEW YORK, NY 10013 USA
SN 1523-3790
EI 1534-6269
J9 CURR ONCOL REP
JI Curr. Oncol. Rep.
PD DEC
PY 2015
VL 17
IS 12
AR 60
DI 10.1007/s11912-015-0483-8
PG 8
WC Oncology
WE Science Citation Index Expanded (SCI-EXPANDED)
SC Oncology
GA CZ0PG
UT WOS:000366807600008
PM 26475775
DA 2024-06-03
ER

PT J
AU Thapa, DR
   Hussain, SK
   Tran, WC
   D'souza, G
   Bream, JH
   Achenback, CJ
   Ayyavoo, V
   Detels, R
   Martínez-Maza, O
AF Thapa, Dharma R.
   Hussain, Shehnaz K.
   Wen-Ching Tran
   D'souza, Gypsyamber
   Bream, Jay H.
   Achenback, Chad J.
   Ayyavoo, Velpandi
   Detels, Roger
   Martinez-Maza, Otoniel
TI Serum MicroRNAs in HIV-Infected Individuals as Pre-Diagnosis Biomarkers
   for AIDS-NHL
SO JAIDS-JOURNAL OF ACQUIRED IMMUNE DEFICIENCY SYNDROMES
LA English
DT Article
DE HIV; lymphoma; miR-21; miR-223; miR-122; miR-222
ID HUMAN-IMMUNODEFICIENCY-VIRUS; B-CELL LYMPHOMA; NON-HODGKINS-LYMPHOMA;
   ACTIVE ANTIRETROVIRAL THERAPY; CIRCULATING MICRORNAS;
   HEPATOCELLULAR-CARCINOMA; GERMINAL CENTER; ELEVATED LEVELS; EXPRESSION;
   RISK
AB Objective:To determine if changes in levels of serum microRNAs (miRNAs) were seen preceding the diagnosis of AIDS-related non-Hodgkin lymphoma (AIDS-NHL).Design:Serum miRNA levels were compared in 3 subject groups from the Multicenter AIDS Cohort Study: HIV-negative men (n = 43), HIV-positive men who did not develop NHL (n = 45), and HIV-positive men before AIDS-NHL diagnosis (n = 62, median time before diagnosis, 8.8 months).Methods:A total of 175 serum-enriched miRNAs were initially screened to identify differentially expressed miRNAs among these groups and the results validated by quantitative polymerase chain reaction. Receiver-operating characteristic analysis was then performed to assess biomarker utility.Results:Higher levels of miR-21 and miR-122, and a lower level of miR-223, were able to discriminate HIV-infected from the HIV-uninfected groups, suggesting that these miRNAs are biomarkers for HIV infection but are not AIDS-NHL specific. Among the HIV-infected groups, a higher level of miR-222 was able to discriminate diffuse large B-cell lymphoma (DLBCL) and primary central nervous system lymphoma (PCNSL) subjects from HIV-infected subjects who did not develop NHL, with area under the receiver-operating characteristic curve of 0.777 and 0.792, respectively. At miR-222 cutoff values of 0.105 for DLBCL and 0.109 for PCNSL, the sensitivity and specificity were 75% and 77%, and 80% and 82%, respectively.Conclusions:Altered serum levels of miR-21, miR-122, and miR-223 are seen in HIV-infected individuals. Higher serum level of miR-222 has clear potential as a serum biomarker for earlier detection of DLBCL and PCNSL among HIV-infected individuals.
C1 [Thapa, Dharma R.; Martinez-Maza, Otoniel] Univ Calif Los Angeles, AIDS Inst, Los Angeles, CA 90095 USA.
   [Thapa, Dharma R.; Martinez-Maza, Otoniel] Univ Calif Los Angeles, David Geffen Sch Med, Dept Obstet & Gynecol, Los Angeles, CA 90095 USA.
   [Thapa, Dharma R.; Wen-Ching Tran; Martinez-Maza, Otoniel] Univ Calif Los Angeles, David Geffen Sch Med, Dept Microbiol Immunol & Mol Genet, Los Angeles, CA 90095 USA.
   [Hussain, Shehnaz K.; Martinez-Maza, Otoniel] Univ Calif Los Angeles, Jonsson Comprehens Canc Ctr, Los Angeles, CA 90095 USA.
   [Hussain, Shehnaz K.; Detels, Roger; Martinez-Maza, Otoniel] Univ Calif Los Angeles, Fielding Sch Publ Hlth, Dept Epidemiol, Los Angeles, CA 90095 USA.
   [D'souza, Gypsyamber] Johns Hopkins Bloomberg Sch Publ Hlth, Dept Epidemiol, Baltimore, MD USA.
   [Bream, Jay H.] Johns Hopkins Bloomberg Sch Publ Hlth, Dept Mol Microbiol & Immunol, Baltimore, MD USA.
   [Achenback, Chad J.] Northwestern Univ, Dept Med, Div Infect Dis, Feinberg Sch Med, Chicago, IL 60611 USA.
   [Ayyavoo, Velpandi] Univ Pittsburgh, Grad Sch Publ Hlth, Dept Infect Dis & Microbiol, Pittsburgh, PA 15261 USA.
C3 University of California System; University of California Los Angeles;
   University of California System; University of California Los Angeles;
   University of California Los Angeles Medical Center; David Geffen School
   of Medicine at UCLA; University of California System; University of
   California Los Angeles; University of California Los Angeles Medical
   Center; David Geffen School of Medicine at UCLA; University of
   California System; University of California Los Angeles; UCLA Jonsson
   Comprehensive Cancer Center; University of California System; University
   of California Los Angeles; Johns Hopkins University; Johns Hopkins
   Bloomberg School of Public Health; Johns Hopkins University; Johns
   Hopkins Bloomberg School of Public Health; Northwestern University;
   Feinberg School of Medicine; Pennsylvania Commonwealth System of Higher
   Education (PCSHE); University of Pittsburgh
RP Martínez-Maza, O (corresponding author), Univ Calif Los Angeles, AIDS Inst, 173 BSRB,615 Charles E Young Dr South, Los Angeles, CA 90095 USA.
EM
RI Thapa, Dharma R/C-2854-2012; Martinez-Maza, Otoniel/B-2667-2009;
   Achenbach, Chad/AAQ-4035-2020
OI Martinez-Maza, Otoniel/0000-0003-1364-0675; Achenbach,
   Chad/0000-0003-4847-7249; Bream, Jay/0000-0002-9149-7643
FU Margaret E. Early Trust [B-MEEMR 20113120]; National Institutes of
   Health [R01-CA-168482, R01-CA-057152]; National Institutes of Health
   (NCI) [U01-AI-035040, K07-CA-140360]; National Cancer Institute
   [UM1-CA-181255]
FX Supported by grants from the Margaret E. Early Trust (B-MEEMR 20113120)
   and the National Institutes of Health (R01-CA-168482, R01-CA-057152, NCI
   supplements to U01-AI-035040, and K07-CA-140360 to S. K. H.). The
   Multicenter AIDS Cohort Study includes the following: Baltimore-The
   Johns Hopkins University Bloomberg School of Public Health: Joseph B.
   Margolick (PI), Barbara Crain, Adrian Dobs, Homayoon Farzadegan, Joel
   Gallant, Lisette Johnson-Hill, Michael W. Plankey, Ned Sacktor, Ola
   Selnes, James Shepard, and Chloe Thio. Chicago-Feinberg School of
   Medicine, Northwestern University, and Cook County Bureau of Health
   Services: Steven M. Wolinsky (PI), John P. Phair, Sheila Badri, Maurice
   O'Gorman, David Ostrow, Frank Palella, and Ann Ragin. Los
   Angeles-University of California, University of California, Los Angeles,
   Schools of Public Health and Medicine: R. D. (PI), O.M-M. (Co-PI), Aaron
   Aronow, Robert Bolan, Elizabeth Breen, Anthony Butch, Beth Jamieson,
   Eric N. Miller, John Oishi, Harry Vinters, Dorothy Wiley, Mallory Witt,
   Otto Yang, Stephen Young, and Zuo Feng Zhang. Pittsburgh-University of
   Pittsburgh, Graduate School of Public Health: Charles R. Rinaldo (PI),
   Lawrence A. Kingsley (Co-PI), James T. Becker, Ross D. Cranston, Jeremy
   J. Martinson, John W. Mellors, Anthony J. Silvestre, and Ronald D.
   Stall. Data Coordinating Center-The Johns Hopkins University Bloomberg
   School of Public Health: Lisa P. Jacobson (PI), Alvaro Munoz (Co-PI),
   Alison, Abraham, Keri Althoff, Christopher Cox, G.D., Priya Duggal,
   Elizabeth Golub, Janet Schollenberger, Eric C. Seaberg, Sol Su, and
   Pamela Surkan. NIH-National Institute of Allergy and Infectious
   Diseases: Robin E. Huebner; National Cancer Institute: Geraldina
   Dominguez. UO1-AI-35042, UM1-AI-35043, UO1-AI-35039, UO1-AI-35040,
   UO1-AI-35041, and UL1-RR025005 (NCATS, National Center for Advancing
   Translational Sciences). Website located at http://www.statepi.
   jhsph.edu/macs/macs.html. Specimens were provided by the AIDS and Cancer
   Specimen Resource, funded by the National Cancer Institute
   (UM1-CA-181255).
CR Ahluwalia JK, 2008, RETROVIROLOGY, V5, DOI 10.1186/1742-4690-5-117
   Alencar AJ, 2011, CLIN CANCER RES, V17, P4125, DOI 10.1158/1078-0432.CCR-11-0224
   Allegra A, 2012, INT J ONCOL, V41, P1897, DOI 10.3892/ijo.2012.1647
   Asslaber D, 2010, BLOOD, V115, P4191, DOI 10.1182/blood-2009-07-234823
   Baraniskin A, 2011, BLOOD, V117, P3140, DOI 10.1182/blood-2010-09-308684
   Bartel DP, 2004, CELL, V116, P281, DOI 10.1016/S0092-8674(04)00045-5
   BERAL V, 1991, LANCET, V337, P805, DOI 10.1016/0140-6736(91)92513-2
   Bignami F, 2012, BLOOD, V119, P6259, DOI 10.1182/blood-2011-09-379503
   Bohlius J, 2009, ANTIVIR THER, V14, P1065, DOI 10.3851/IMP1462
   Bonnet F, 2004, CANCER-AM CANCER SOC, V101, P317, DOI 10.1002/cncr.20354
   Breen EC, 2005, AIDS, V19, P1711, DOI 10.1097/01.aids.0000184924.04983.7c
   Breen EC, 2011, CANCER EPIDEM BIOMAR, V20, P1303, DOI 10.1158/1055-9965.EPI-11-0037
   Calin GA, 2005, NEW ENGL J MED, V353, P1793, DOI 10.1056/NEJMoa050995
   Calin GA, 2004, P NATL ACAD SCI USA, V101, P11755, DOI 10.1073/pnas.0404432101
   Cote TR, 1997, INT J CANCER, V73, P645, DOI 10.1002/(SICI)1097-0215(19971127)73:5<645::AID-IJC6>3.0.CO;2-X
   Culpin RE, 2010, INT J ONCOL, V37, P367, DOI 10.3892/ijo_00000685
   De Roos AJ, 2012, CANCER RES, V72, P4733, DOI 10.1158/0008-5472.CAN-12-1639
   Di Lisio L, 2012, BLOOD CANCER J, V2, DOI 10.1038/bcj.2012.1
   Di Lisio L, 2012, BLOOD, V120, P1782, DOI 10.1182/blood-2012-05-402784
   Diamond C, 2006, CANCER, V106, P128, DOI 10.1002/cncr.21562
   Engels EA, 2008, INT J CANCER, V123, P187, DOI 10.1002/ijc.23487
   Epeldegui M, 2007, AIDS, V21, P2265, DOI 10.1097/QAD.0b013e3282ef9f59
   Fang C, 2012, ANN HEMATOL, V91, P553, DOI 10.1007/s00277-011-1350-9
   Fassina A, 2012, LAB INVEST, V92, P1574, DOI 10.1038/labinvest.2012.129
   Ferracin M, 2010, MOL CANCER, V9, DOI 10.1186/1476-4598-9-123
   Fischer L, 2011, NEURO-ONCOLOGY, V13, P1090, DOI 10.1093/neuonc/nor107
   Fu XN, 2011, EUR J CLIN INVEST, V41, P1245, DOI 10.1111/j.1365-2362.2011.02535.x
   Gupta A, 2011, PLOS ONE, V6, DOI [10.1371/journal.pone.0022730, 10.1371/journal.pone.0028691]
   Houzet L, 2008, RETROVIROLOGY, V5, DOI 10.1186/1742-4690-5-118
   Huang JL, 2007, NAT MED, V13, P1241, DOI 10.1038/nm1639
   Jima DD, 2010, BLOOD, V116, pE118, DOI 10.1182/blood-2010-05-285403
   KASLOW RA, 1987, AM J EPIDEMIOL, V126, P310, DOI 10.1093/aje/126.2.310
   Kosaka N, 2010, CANCER SCI, V101, P2087, DOI 10.1111/j.1349-7006.2010.01650.x
   Lagos-Quintana M, 2002, CURR BIOL, V12, P735, DOI 10.1016/S0960-9822(02)00809-6
   Lawrie CH, 2008, BRIT J HAEMATOL, V141, P672, DOI 10.1111/j.1365-2141.2008.07077.x
   Lawrie CH, 2007, INT J CANCER, V121, P1156, DOI 10.1002/ijc.22800
   Lawrie CH, 2009, J CELL MOL MED, V13, P1248, DOI 10.1111/j.1582-4934.2008.00628.x
   Lenze D, 2011, LEUKEMIA, V25, P1869, DOI 10.1038/leu.2011.156
   Levine AM, 2006, ARCH INTERN MED, V166, P405, DOI 10.1001/.405
   Li C, 2009, BLOOD, V113, P6681, DOI 10.1182/blood-2009-01-202028
   Long JL, 2008, AIDS, V22, P489, DOI 10.1097/QAD.0b013e3282f47082
   Malumbres R, 2009, BLOOD, V113, P3754, DOI 10.1182/blood-2008-10-184077
   Monforte AD, 2005, ARCH INTERN MED, V165, P416
   Montes-Moreno S, 2011, BLOOD, V118, P1034, DOI 10.1182/blood-2010-11-321554
   Moussay E, 2011, P NATL ACAD SCI USA, V108, P6573, DOI 10.1073/pnas.1019557108
   Navarro A, 2008, BLOOD, V111, P2825, DOI 10.1182/blood-2007-06-096784
   O'Hara AJ, 2008, BLOOD, V111, P2347, DOI 10.1182/blood-2007-08-104463
   Palella FJ, 2006, JAIDS-J ACQ IMM DEF, V43, P27, DOI 10.1097/01.qai.0000233310.90484.16
   PLUDA JM, 1993, J CLIN ONCOL, V11, P1099, DOI 10.1200/JCO.1993.11.6.1099
   Price JC, 2010, CLIN GASTROENTEROL H, V8, P1002, DOI 10.1016/j.cgh.2010.08.024
   Pritchard CC, 2012, CANCER PREV RES, V5, P492, DOI 10.1158/1940-6207.CAPR-11-0370
   Qu KZ, 2011, J CLIN GASTROENTEROL, V45, P355, DOI 10.1097/MCG.0b013e3181f18ac2
   Raphael M., 2001, WHO CLASSIFICATION T, P260
   Robertus JL, 2010, BRIT J HAEMATOL, V149, P896, DOI 10.1111/j.1365-2141.2010.08111.x
   Roehle A, 2008, BRIT J HAEMATOL, V142, P732, DOI 10.1111/j.1365-2141.2008.07237.x
   Rossi S, 2010, BLOOD, V116, P945, DOI 10.1182/blood-2010-01-263889
   Saeed AI, 2003, BIOTECHNIQUES, V34, P374, DOI 10.2144/03342mt01
   Seaberg EC, 2010, CANCER-AM CANCER SOC, V116, P5507, DOI 10.1002/cncr.25530
   Silverberg MJ, 2011, CANCER EPIDEM BIOMAR, V20, P2551, DOI 10.1158/1055-9965.EPI-11-0777
   Stamatopoulos B, 2009, BLOOD, V113, P5237, DOI 10.1182/blood-2008-11-189407
   Thapa DR, 2012, AIDS, V26, P1177, DOI 10.1097/QAD.0b013e3283543e0e
   Thapa DR, 2011, PLOS ONE, V6, DOI 10.1371/journal.pone.0020781
   Trebicka J, 2013, J HEPATOL, V58, P234, DOI 10.1016/j.jhep.2012.10.015
   Triboulet R, 2007, SCIENCE, V315, P1579, DOI 10.1126/science.1136319
   Visone R, 2011, BLOOD, V118, P3072, DOI 10.1182/blood-2011-01-333484
   Wang B, 2012, J CANCER RES CLIN, V138, P1659, DOI 10.1007/s00432-012-1244-9
   Wang JF, 2010, BIOCHEM BIOPH RES CO, V394, P184, DOI 10.1016/j.bbrc.2010.02.145
   Wang K, 2009, P NATL ACAD SCI USA, V106, P4402, DOI 10.1073/pnas.0813371106
   Wang X, 2009, BLOOD, V113, P671, DOI 10.1182/blood-2008-09-175000
   Witwer KW, 2012, RETROVIROLOGY, V9, DOI 10.1186/1742-4690-9-5
   Witwer KW, 2011, AIDS, V25, P2057, DOI 10.1097/QAD.0b013e32834b95bf
   Xu JA, 2011, MOL CARCINOGEN, V50, P136, DOI 10.1002/mc.20712
   Xu J, 2012, LIVER INT, V32, P752, DOI 10.1111/j.1478-3231.2011.02750.x
   Yeung ML, 2005, RETROVIROLOGY, V2, DOI 10.1186/1742-4690-2-81
   Zhang J, 2009, BLOOD, V113, P4586, DOI 10.1182/blood-2008-09-178186
   ZON LI, 1987, BRIT J HAEMATOL, V66, P251, DOI 10.1111/j.1365-2141.1987.tb01307.x
NR 76
TC 26
Z9 30
U1 0
U2 5
PU LIPPINCOTT WILLIAMS & WILKINS
PI PHILADELPHIA
PA TWO COMMERCE SQ, 2001 MARKET ST, PHILADELPHIA, PA 19103 USA
SN 1525-4135
EI 1077-9450
J9 JAIDS-J ACQ IMM DEF
JI JAIDS
PD JUN 1
PY 2014
VL 66
IS 2
BP 229
EP 237
DI 10.1097/QAI.0000000000000146
PG 9
WC Immunology; Infectious Diseases
WE Science Citation Index Expanded (SCI-EXPANDED)
SC Immunology; Infectious Diseases
GA AJ4YJ
UT WOS:000337685800027
PM 24675587
OA Green Accepted, Bronze, Green Published
DA 2024-06-03
ER

PT J
AU Brastianos, PK
   Batchelor, TT
AF Brastianos, Priscilla K.
   Batchelor, Tracy T.
TI Primary Central Nervous System Lymphoma Overview of Current Treatment
   Strategies
SO HEMATOLOGY-ONCOLOGY CLINICS OF NORTH AMERICA
LA English
DT Article
DE Primary central nervous system lymphoma; Treatment; Chemotherapy;
   Radiation
ID PRIMARY CNS LYMPHOMA; HIGH-DOSE METHOTREXATE; NON-HODGKINS-LYMPHOMA;
   WHOLE-BRAIN RADIOTHERAPY; STEM-CELL TRANSPLANTATION; RECURRENT PRIMARY
   CNS; MULTICENTER PHASE-II; RADIATION-THERAPY; INITIAL TREATMENT;
   INTRAVENTRICULAR CHEMOTHERAPY
AB Primary central nervous system lymphoma (PCNSL) is an uncommon and aggressive variant of non-Hodgkin lymphoma that involves the brain, eyes, leptomeninges, or spinal cord. Therapeutic progress has been modest, and our understanding of the molecular mechanisms that drive this disease is limited. Clinicians treating PCNSL face a challenge to balance the need to administer aggressive regimens to achieve a cure with the risks of delayed neurotoxicity after treatment. The standard treatment is a methotrexate-containing chemotherapy regimen. The timing and dose of whole-brain radiation therapy is controversial, given the significant risks of late neurotoxic effects, particularly in elderly patients.
C1 [Brastianos, Priscilla K.; Batchelor, Tracy T.] Harvard Univ, Massachusetts Gen Hosp Canc Ctr, Stephen E & Catherine Pappas Ctr Neurooncol, Div Hematol & Oncol,Dept Neurol,Med Sch, Boston, MA 02114 USA.
   [Brastianos, Priscilla K.] Harvard Univ, Dept Med Oncol, Dana Farber Brigham & Womens Canc Ctr, Sch Med, Boston, MA 02115 USA.
   [Batchelor, Tracy T.] Harvard Univ, Dept Radiat Oncol, Massachusetts Gen Hosp, Sch Med, Boston, MA 02114 USA.
C3 Harvard University; Harvard Medical School; Massachusetts General
   Hospital; Harvard University; Harvard Medical School; Dana-Farber Cancer
   Institute; Harvard University; Massachusetts General Hospital; Harvard
   Medical School
RP Brastianos, PK (corresponding author), Harvard Univ, Massachusetts Gen Hosp Canc Ctr, Stephen E & Catherine Pappas Ctr Neurooncol, Div Hematol & Oncol,Dept Neurol,Med Sch, 55 Fruit St,Yawkey 9 E, Boston, MA 02114 USA.
EM
OI Brastianos, Priscilla/0000-0003-4470-8425
CR Abrey LE, 2006, J CLIN ONCOL, V24, P5711, DOI 10.1200/JCO.2006.08.2941
   Abrey LE, 2003, J CLIN ONCOL, V21, P4151, DOI 10.1200/JCO.2003.05.024
   Abrey LE, 2005, J CLIN ONCOL, V23, P5034, DOI 10.1200/JCO.2005.13.524
   Abrey LE, 1998, J CLIN ONCOL, V16, P859, DOI 10.1200/JCO.1998.16.3.859
   Abrey LE, 2000, J CLIN ONCOL, V18, P3144, DOI 10.1200/JCO.2000.18.17.3144
   BALMACEDA C, 1995, ANN NEUROL, V38, P202, DOI 10.1002/ana.410380212
   Bataille B, 2000, J NEUROSURG, V92, P261, DOI 10.3171/jns.2000.92.2.0261
   Batchelor T, 2006, J CLIN ONCOL, V24, P1281, DOI 10.1200/JCO.2005.04.8819
   Batchelor T, 2003, J CLIN ONCOL, V21, P1044, DOI 10.1200/JCO.2003.03.036
   Batchelor TT, 2011, NEUROLOGY, V76, P929, DOI 10.1212/WNL.0b013e31820f2d94
   Bellinzona M, 2005, EJSO, V31, P100, DOI 10.1016/j.ejso.2004.10.002
   BERRY MP, 1981, INT J RADIAT ONCOL, V7, P55, DOI 10.1016/0360-3016(81)90060-2
   BLAY JY, 1995, BLOOD, V86, P2922
   Braaten KM, 2003, CLIN CANCER RES, V9, P1063
   Brevet M, 2005, EUR J HAEMATOL, V75, P288, DOI 10.1111/j.1600-0609.2005.00508.x
   CBTRUS, 2011, PRIM BRAIN CENTR NER
   Colombat P, 2006, BONE MARROW TRANSPL, V38, P417, DOI 10.1038/sj.bmt.1705452
   Correa DD, 2007, ANN ONCOL, V18, P1145, DOI 10.1093/annonc/mdl464
   DEANGELIS LM, 1992, J CLIN ONCOL, V10, P635, DOI 10.1200/JCO.1992.10.4.635
   DeAngelis LM, 2002, J CLIN ONCOL, V20, P4643, DOI 10.1200/JCO.2002.11.013
   Enting RH, 2004, NEUROLOGY, V63, P901, DOI 10.1212/01.WNL.0000137050.43114.42
   Ferreri AJM, 1995, CANCER TREAT REV, V21, P415, DOI 10.1016/0305-7372(95)90028-4
   Ferreri AJM, 1996, CANCER, V77, P827, DOI 10.1002/(SICI)1097-0142(19960301)77:5<827::AID-CNCR4>3.0.CO;2-B
   Ferreri AJM, 2006, NEUROLOGY, V66, P1435, DOI 10.1212/01.wnl.0000210464.94122.e1
   Ferreri AJM, 2001, ONCOLOGY-BASEL, V60, P134, DOI 10.1159/000055310
   Ferreri AJM, 2003, J CLIN ONCOL, V21, P266, DOI 10.1200/JCO.2003.09.139
   Ferreri AJM, 2002, NEUROLOGY, V58, P1513, DOI 10.1212/WNL.58.10.1513
   Ferreri AJM, 2007, CRIT REV ONCOL HEMAT, V63, P257, DOI 10.1016/j.critrevonc.2007.04.012
   Ferreri AJM, 2011, BLOOD, V118, P510, DOI 10.1182/blood-2011-03-321349
   Ferreri AJM, 2009, LANCET, V374, P1512, DOI 10.1016/S0140-6736(09)61416-1
   Filley CM, 2001, NEW ENGL J MED, V345, P425, DOI 10.1056/NEJM200108093450606
   FINE HA, 1993, ANN INTERN MED, V119, P1093, DOI 10.7326/0003-4819-119-11-199312010-00007
   Fischer L, 2006, ANN ONCOL, V17, P1141, DOI 10.1093/annonc/mdl070
   Fischer L, 2006, HAEMATOLOGICA, V91, P429
   Flanagan EP, 2011, NEUROLOGY, V77, P784, DOI 10.1212/WNL.0b013e31822b00b9
   Freilich RJ, 1996, NEUROLOGY, V46, P435, DOI 10.1212/WNL.46.2.435
   Fritsch K, 2011, ANN ONCOL, V22, P2080, DOI 10.1093/annonc/mdq712
   Gavrilovic IT, 2006, J CLIN ONCOL, V24, P4570, DOI 10.1200/JCO.2006.06.6910
   Gerstner ER, 2008, NEUROLOGY, V70, P401, DOI 10.1212/01.wnl.0000300671.37279.0e
   Gerstner ER, 2010, ARCH NEUROL-CHICAGO, V67, P291, DOI 10.1001/archneurol.2010.3
   Glass J, 1996, J NEURO-ONCOL, V30, P257
   GLASS J, 1994, J NEUROSURG, V81, P188, DOI 10.3171/jns.1994.81.2.0188
   Guha-Thakurta N, 1999, J NEURO-ONCOL, V43, P259, DOI 10.1023/A:1006210703827
   Herrlinger U, 2000, NEUROLOGY, V54, P1707, DOI 10.1212/WNL.54.8.1707
   Herrlinger U, 2005, ANN NEUROL, V57, P843, DOI 10.1002/ana.20495
   Herrlinger U, 2002, ANN NEUROL, V51, P247, DOI 10.1002/ana.10102
   Hiraga S, 1999, J NEUROSURG, V91, P221, DOI 10.3171/jns.1999.91.2.0221
   Hoang-Xuan K, 2003, J CLIN ONCOL, V21, P2726, DOI 10.1200/JCO.2003.11.036
   Hottinger AF, 2007, NEUROLOGY, V69, P1178, DOI 10.1212/01.wnl.0000276986.19602.c1
   Illerhaus G, 2009, ANN ONCOL, V20, P319, DOI 10.1093/annonc/mdn628
   Illerhaus G, 2007, HAEMATOLOGICA, V93, P147, DOI 10.3324/haematol.11771
   Illerhaus G, 2006, J CLIN ONCOL, V24, P3865, DOI 10.1200/JCO.2006.06.2117
   Kadan-Lottick NS, 2002, CANCER, V95, P193, DOI 10.1002/cncr.10643
   Khan RB, 2002, J NEURO-ONCOL, V58, P175, DOI 10.1023/A:1016077907952
   Küker W, 2005, J NEURO-ONCOL, V72, P169, DOI 10.1007/s11060-004-3390-7
   Mathew BS, 2006, CANCER-AM CANCER SOC, V106, P383, DOI 10.1002/cncr.21583
   Mead GM, 2000, CANCER, V89, P1359, DOI 10.1002/1097-0142(20000915)89:6<1359::AID-CNCR21>3.0.CO;2-9
   MILLER DC, 1994, CANCER-AM CANCER SOC, V74, P1383, DOI 10.1002/1097-0142(19940815)74:4<1383::AID-CNCR2820740432>3.0.CO;2-1
   Montemurro M, 2007, ANN ONCOL, V18, P665, DOI 10.1093/annonc/mdl458
   Montesinos-Rongen M, 2004, BLOOD, V103, P1869, DOI 10.1182/blood-2003-05-1465
   Montesinos-Rongen M, 2003, BRAIN PATHOL, V13, P534
   NELSON DF, 1992, INT J RADIAT ONCOL, V23, P9, DOI 10.1016/0360-3016(92)90538-S
   Ney DE, 2010, CANCER-AM CANCER SOC, V116, P4605, DOI 10.1002/cncr.25363
   Nguyen PL, 2005, J CLIN ONCOL, V23, P1507, DOI 10.1200/JCO.2005.01.161
   O'Brien P, 2000, J CLIN ONCOL, V18, P519, DOI 10.1200/JCO.2000.18.3.519
   OBBENS EAMT, 1985, NEUROLOGY, V35, P1274, DOI 10.1212/WNL.35.9.1274
   Omuro AMP, 2007, J NEURO-ONCOL, V85, P207, DOI 10.1007/s11060-007-9397-0
   ONEILL BP, 1995, J NEURO-ONCOL, V25, P67, DOI 10.1007/BF01054724
   Ostermann K, 2011, J NEURO-ONCOL, V103, P635, DOI 10.1007/s11060-010-0435-y
   Panageas KS, 2007, CANCER, V110, P1338, DOI 10.1002/cncr.22907
   Pels H, 2003, J CLIN ONCOL, V21, P4489, DOI 10.1200/JCO.2003.04.056
   Pels H, 2006, NEURO-ONCOLOGY, V8, P293
   Pels H, 2009, J NEURO-ONCOL, V91, P299, DOI 10.1007/s11060-008-9712-4
   Plotkin SR, 2004, CLIN CANCER RES, V10, P5643, DOI 10.1158/1078-0432.CCR-04-0159
   Poortmans PMP, 2003, J CLIN ONCOL, V21, P4483, DOI 10.1200/JCO.2003.03.108
   Reni M, 2007, BRIT J CANCER, V96, P864, DOI 10.1038/sj.bjc.6603660
   Reni M, 1997, ANN ONCOL, V8, P227, DOI 10.1023/A:1008201717089
   Rickert CH, 1999, AM J PATHOL, V155, P1445, DOI 10.1016/S0002-9440(10)65458-2
   Rubenstein JL, 2007, J CLIN ONCOL, V25, P1350, DOI 10.1200/JCO.2006.09.7311
   Rubenstein JL, 2010, BLOOD, V116, P335, DOI 10.1182/blood.V116.21.763.763
   Rubenstein JL, 2009, BLOOD, V113, P266, DOI 10.1182/blood-2008-04-152835
   Rubenstein JL, 2006, BLOOD, V107, P3716, DOI 10.1182/blood-2005-03-0897
   Sandor V, 1998, J CLIN ONCOL, V16, P3000, DOI 10.1200/JCO.1998.16.9.3000
   Schabet M, 1999, J NEURO-ONCOL, V43, P199, DOI 10.1023/A:1006290032052
   Schlegel U, 2001, J NEUROL NEUROSUR PS, V71, P118, DOI 10.1136/jnnp.71.1.118
   Shah GD, 2007, J CLIN ONCOL, V25, P4730, DOI 10.1200/JCO.2007.12.5062
   SHAPIRO WR, 1975, NEW ENGL J MED, V293, P161, DOI 10.1056/NEJM197507242930402
   Shibamoto Y, 2005, INT J RADIAT ONCOL, V62, P809, DOI 10.1016/j.ijrobp.2004.12.043
   Soussain C, 2008, J CLIN ONCOL, V26, P2512, DOI 10.1200/JCO.2007.13.5533
   Thiel E, 2010, LANCET ONCOL, V11, P1036, DOI 10.1016/S1470-2045(10)70229-1
   Tun HW, 2008, BLOOD, V111, P3200, DOI 10.1182/blood-2007-10-119099
   Zhu JJ, 2009, NEURO-ONCOLOGY, V11, P211, DOI 10.1215/15228517-2008-067
NR 92
TC 26
Z9 29
U1 0
U2 6
PU W B SAUNDERS CO-ELSEVIER INC
PI PHILADELPHIA
PA 1600 JOHN F KENNEDY BOULEVARD, STE 1800, PHILADELPHIA, PA 19103-2899 USA
SN 0889-8588
EI 1558-1977
J9 HEMATOL ONCOL CLIN N
JI Hematol. Oncol. Clin. North Am.
PD AUG
PY 2012
VL 26
IS 4
BP 897
EP +
DI 10.1016/j.hoc.2012.05.003
PG 21
WC Oncology; Hematology
WE Science Citation Index Expanded (SCI-EXPANDED)
SC Oncology; Hematology
GA 989ZW
UT WOS:000307601800011
PM 22794289
DA 2024-06-03
ER

PT J
AU Schuurmans, M
   Bromberg, JEC
   Doorduijn, J
   Poortmans, P
   Taphoorn, MJB
   Seute, T
   Enting, R
   van Imhoff, G
   van Norden, Y
   van den Bent, MJ
AF Schuurmans, Mascha
   Bromberg, Jacoline E. C.
   Doorduijn, Jeanette
   Poortmans, Philip
   Taphoorn, Martin J. B.
   Seute, Tatjana
   Enting, Roelien
   van Imhoff, Gustaaf
   van Norden, Yvette
   van den Bent, Martin J.
TI Primary central nervous system lymphoma in the elderly: a multicentre
   retrospective analysis
SO BRITISH JOURNAL OF HAEMATOLOGY
LA English
DT Article
DE central nervous system lymphoma; radiotherapy; chemotherapy; age;
   performance status
ID HIGH-DOSE METHOTREXATE; PRIMARY CNS LYMPHOMA; WHOLE-BRAIN RADIOTHERAPY;
   PHASE-II; EUROPEAN ORGANIZATION; CHEMOTHERAPY; RADIATION; SURVIVAL;
   TOXICITY; SALVAGE
AB Elderly patients with primary central nervous ystem lymphoma (PCNSL) do not tolerate treatment with combined radio-chemotherapy well because of leuco-encephalopathy; they are usually treated initially with chemotherapy or radiotherapy alone. Little is known about the efficacy and toxicity of these treatments outside clinical studies. This study was a retrospective analysis of all patients aged 60 years or over who were admitted with PCNSL to one of five Dutch centers between 1998 and 2007. A total of 74 patients were identified. Twenty-nine were treated with radiotherapy only (Group A), in 36 the intended treatment was chemotherapy alone (Group B), and nine were planned to receive chemotherapy followed by radiotherapy (Group C). Median overall survival was 20 months; 4 months in patients with a Karnofsky performance status (KPS) < 70, 25 months in patients with a KPS >= 70 (P < 0 center dot 001). Treatment modality was not an independent prognostic factor. Forty patients were treated with methotrexate 3 g/m2: there were two toxic deaths. Ten patients discontinued chemotherapy because of toxicity. Delayed encephalopathy was reported in 10 patients. In conclusion, community hospitals still frequently utilize whole brain radiotherapy in elderly PCNSL patients, though a majority tolerates chemotherapy well. Performance status was the most important variable determining prognosis. Short and long term toxicities must be weighed against possible clinical benefits of each treatment, making treatment decisions a highly individualized process.
C1 [Schuurmans, Mascha; Bromberg, Jacoline E. C.; van den Bent, Martin J.] Erasmus MC, Daniel den Hoed Canc Ctr, Dept Neurooncol, Rotterdam, Netherlands.
   [Doorduijn, Jeanette] Erasmus MC, Daniel den Hoed Canc Ctr, Dept Haematol, Rotterdam, Netherlands.
   [Poortmans, Philip] Bernard Verbeeten Inst Radiotherapy, Rotterdam, Netherlands.
   [Taphoorn, Martin J. B.] MC Haaglanden, Dept Neurol, Rotterdam, Netherlands.
   [Seute, Tatjana] Univ Utrecht, Med Ctr, Dept Neurol, Rotterdam, Netherlands.
   [Enting, Roelien] Univ Groningen, Dept Neurol, Med Ctr, Rotterdam, Netherlands.
   [van Imhoff, Gustaaf] Univ Groningen, Dept Haematol, Med Ctr, Rotterdam, Netherlands.
   [van Norden, Yvette] Erasmus MC, Daniel den Hoed Canc Ctr, Dept Trials & Stat, Rotterdam, Netherlands.
C3 Erasmus University Rotterdam; Erasmus MC; Erasmus MC Cancer Institute;
   Erasmus University Rotterdam; Erasmus MC; Erasmus MC Cancer Institute;
   Instituut Verbeeten; Utrecht University; University of Groningen;
   University of Groningen; Erasmus University Rotterdam; Erasmus MC;
   Erasmus MC Cancer Institute
RP Bromberg, JEC (corresponding author), Erasmus Univ, Daniel den Hoed Canc Ctr, Dept Neurooncol, Med Ctr, POB 5201, NL-3008 AE Rotterdam, Netherlands.
EM
RI van den Bent, Martin J/JPK-6126-2023; Poortmans, P.M.P./L-4581-2015
OI van den Bent, Martin J/0000-0001-5710-5127; Doorduijn,
   Jeanette/0000-0002-1014-0918; Bromberg, Jacoline/0000-0002-3830-7142
CR Abrey LE, 2006, J CLIN ONCOL, V24, P5711, DOI 10.1200/JCO.2006.08.2941
   Batchelor T, 2006, J CLIN ONCOL, V24, P1281, DOI 10.1200/JCO.2005.04.8819
   Bessell EM, 2007, EUR J CANCER, V43, P1141, DOI 10.1016/j.ejca.2006.12.011
   Blay JY, 1998, J CLIN ONCOL, V16, P864, DOI 10.1200/JCO.1998.16.3.864
   DeAngelis LM, 2002, J CLIN ONCOL, V20, P4643, DOI 10.1200/JCO.2002.11.013
   Ekenel M, 2008, CANCER-AM CANCER SOC, V113, P1025, DOI 10.1002/cncr.23670
   Ferreri AJM, 2003, J CLIN ONCOL, V21, P266, DOI 10.1200/JCO.2003.09.139
   Hoang-Xuan K, 2003, J CLIN ONCOL, V21, P2726, DOI 10.1200/JCO.2003.11.036
   Hottinger AF, 2007, NEUROLOGY, V69, P1178, DOI 10.1212/01.wnl.0000276986.19602.c1
   Illerhaus G, 2009, ANN ONCOL, V20, P319, DOI 10.1093/annonc/mdn628
   Jahnke K, 2005, ANN ONCOL, V16, P445, DOI 10.1093/annonc/mdi075
   Laack NN, 2006, INT J RADIAT ONCOL, V65, P1429, DOI 10.1016/j.ijrobp.2006.03.061
   NELSON DF, 1992, INT J RADIAT ONCOL, V23, P9, DOI 10.1016/0360-3016(92)90538-S
   Ng S, 2000, NEURO-ONCOLOGY, V2, P40, DOI 10.1093/neuonc/2.1.40
   Nguyen PL, 2005, J CLIN ONCOL, V23, P1507, DOI 10.1200/JCO.2005.01.161
   Omuro AMP, 2005, ARCH NEUROL-CHICAGO, V62, P1595, DOI 10.1001/archneur.62.10.1595
   Omuro AMP, 2007, J NEURO-ONCOL, V85, P207, DOI 10.1007/s11060-007-9397-0
   Panageas KS, 2007, CANCER, V110, P1338, DOI 10.1002/cncr.22907
   Pels H, 2003, J CLIN ONCOL, V21, P4489, DOI 10.1200/JCO.2003.04.056
   Poortmans PMP, 2003, J CLIN ONCOL, V21, P4483, DOI 10.1200/JCO.2003.03.108
   Shibamoto Y, 2005, INT J RADIAT ONCOL, V62, P809, DOI 10.1016/j.ijrobp.2004.12.043
   Wassenberg MWM, 2001, J NEURO-ONCOL, V52, P73, DOI 10.1023/A:1010676807228
NR 22
TC 26
Z9 26
U1 0
U2 3
PU WILEY-BLACKWELL
PI MALDEN
PA COMMERCE PLACE, 350 MAIN ST, MALDEN 02148, MA USA
SN 0007-1048
J9 BRIT J HAEMATOL
JI Br. J. Haematol.
PD OCT
PY 2010
VL 151
IS 2
BP 179
EP 184
DI 10.1111/j.1365-2141.2010.08328.x
PG 6
WC Hematology
WE Science Citation Index Expanded (SCI-EXPANDED)
SC Hematology
GA 656XN
UT WOS:000282374500007
PM 20738305
OA Green Submitted, Bronze
DA 2024-06-03
ER

PT J
AU Hardwig, PW
   Pulido, JS
   Bakri, SJ
AF Hardwig, Paul W.
   Pulido, Jose S.
   Bakri, Sophie J.
TI The safety of intraocular methotrexate in silicone-filled eyes
SO RETINA-THE JOURNAL OF RETINAL AND VITREOUS DISEASES
LA English
DT Article
DE intravitreal methotrexate; silicone oil; proliferative
   vitreoretinopathy; proliferative diabetic retinopathy; uveitis
ID NERVOUS-SYSTEM LYMPHOMA; INTRAVITREAL METHOTREXATE; CLEARANCE;
   DEXAMETHASONE; FLUOROURACIL; CHEMOTHERAPY; GENTAMICIN; INJECTION;
   TOXICITY; DRUGS
AB Background: Intraocular methotrexate has been safely used in eyes with primary CNS lymphoma (PCNSL), and in eyes with uveitis and proliferative diabetic retinopathy. Dosing in silicone-filled eyes was reduced from a standard 400 mu g intravitreal injection due to concerns of toxicity. The present study reports the visual results of non-PCNSL, silicone-filled eyes treated with intravitreal methotrexate using cumulative dosages ranging from 200 mu g to 1,200 mu g.
   Method: In this retrospective case series, all patients with silicone-filled eyes who received intraocular methotrexate were included. Patients were observed with serial ophthalmic examinations. Best-corrected visual acuity was measured by Snellen acuity. Pretreatment acuities were compared to those obtained at last follow-up.
   Results: The cohort included 12 patients (13 eyes) with disease other than PCNSL. The cumulative dose of intraocular methotrexate in any one patient ranged from 200 mu g to 1,200 mu g. Mean follow-up was 9 months (median, 10 months; range, 2 weeks to 16 months). Best-corrected vision at last follow-up was either stable or improved from pretreatment acuity in 12 of 13 eyes.
   Conclusion: Preservation of acuity in 12 of 13 study eyes suggests that intravitreal methotrexate in a cumulative dose of up to 1,200 mu g is safe in silicone-filled eyes.
C1 [Hardwig, Paul W.; Pulido, Jose S.; Bakri, Sophie J.] Mayo Clin, Dept Ophthalmol, Rochester, MN 55905 USA.
C3 Mayo Clinic
RP Hardwig, PW (corresponding author), Mayo Clin, Dept Ophthalmol, W-7, Rochester, MN 55905 USA.
EM
CR BARRADA A, 1983, OPHTHALMIC SURG LAS, V14, P845
   CASE JL, 1985, OPHTHALMIC SURG LAS, V16, P378
   COBO LM, 1981, AM J OPHTHALMOL, V92, P59, DOI 10.1016/S0002-9394(14)75908-2
   de Smet MD, 1999, BRIT J OPHTHALMOL, V83, P448, DOI 10.1136/bjo.83.4.448
   Deng SX, 2006, INVEST OPHTH VIS SCI, V47, P1516, DOI 10.1167/iovs.05-0880
   FISCELLA R, 1985, OPHTHALMIC SURG LAS, V16, P247
   Fishburne BC, 1997, ARCH OPHTHALMOL-CHIC, V115, P1152, DOI 10.1001/archopht.1997.01100160322009
   Hardwig PW, 2006, AM J OPHTHALMOL, V142, P883, DOI 10.1016/j.ajo.2006.06.002
   Holladay JT, 2004, J CATARACT REFR SURG, V30, P287, DOI 10.1016/j.jcrs.2004.01.014
   JARUS G, 1985, OPHTHALMOLOGY, V92, P91
   KANE A, 1981, INVEST OPHTH VIS SCI, V20, P593
   Samson CM, 2001, OPHTHALMOLOGY, V108, P1134, DOI 10.1016/S0161-6420(01)00576-0
   SCHINDLER RH, 1982, AM J OPHTHALMOL, V93, P415, DOI 10.1016/0002-9394(82)90130-1
   Smith JR, 2002, OPHTHALMOLOGY, V109, P1709, DOI 10.1016/S0161-6420(02)01125-9
   *US STAT PHARM CON, 2006, PHARM COMP STER PREP, V1, P334
   Velez G, 2001, ARCH OPHTHALMOL-CHIC, V119, P1518
NR 16
TC 26
Z9 28
U1 0
U2 3
PU LIPPINCOTT WILLIAMS & WILKINS
PI PHILADELPHIA
PA 530 WALNUT ST, PHILADELPHIA, PA 19106-3621 USA
SN 0275-004X
J9 RETINA-J RET VIT DIS
JI Retin.-J. Retin. Vitr. Dis.
PD SEP
PY 2008
VL 28
IS 8
BP 1082
EP 1086
DI 10.1097/IAE.0b013e3181754231
PG 5
WC Ophthalmology
WE Science Citation Index Expanded (SCI-EXPANDED)
SC Ophthalmology
GA 350CC
UT WOS:000259329100008
PM 18779714
DA 2024-06-03
ER

PT J
AU Kanai, R
   Shibuya, M
   Hata, T
   Hori, M
   Hirabayashi, K
   Terada, T
   Fujii, K
AF Kanai, Ryuichi
   Shibuya, Makoto
   Hata, Takashi
   Hori, Makoto
   Hirabayashi, Kenichi
   Terada, Tadashi
   Fujii, Koji
TI A case of 'lymphomatosis cerebri' diagnosed in an early phase and
   treated by whole brain radiation: case report and literature review
SO JOURNAL OF NEURO-ONCOLOGY
LA English
DT Article
DE leukoencephalopathy; lymphomatosis cerebri; magnetic resonance imaging;
   primary CNS lymphoma; white matter dementia
ID CENTRAL-NERVOUS-SYSTEM; RAPIDLY PROGRESSIVE DEMENTIA; MRI FEATURES; CNS
   LYMPHOMA; AIDS; CT
AB 'Lymphomatosis cerebri' (LC) is a term indicating a diffusely infiltrating form of primary central nervous system lymphoma (PCNSL) without evidence of a mass lesion. Not infrequently, diagnostic confusion is caused by its presentation on cranial magnetic resonance images (MRI), which is characterized by diffuse leukoencephalopathy without contrast enhancement. In this report, we describe a 53-year-old, immunocompetent man who had an insidiously progressive dementia and right weakness. On serial MRI in 4 months duration, diffuse white matter lesions without contrast enhancement gradually progressed, which was clinically consistent with his worsening condition. Biopsy specimen demonstrated nondestructive, diffusely infiltrating, large B-cell lymphoma, diagnosing LC. After the biopsy, corticosteroids were initiated, which dramatically alleviated his symptoms. Afterwards, he was treated by whole brain irradiation (total 36Gy) and discharged without noticeable deficits. Diagnosis of LC requires additional examinations generally not performed in the other white matter disorders. In suspected cases, biopsy should be performed to avoid deferring adequate cytostatic treatment.
C1 Shizuoka City Shimizu Hosp, Dept Neurosurg, Shizuoka, Japan.
   Tokyo Med Univ, Hachioji Med Ctr, Dept Pathol, Tokyo, Japan.
   Shizuoka City Shimizu Hosp, Dept Neurol, Shizuoka, Japan.
   Tokai Univ, Sch Med, Dept Pathol, Kanagawa 2591100, Japan.
   Shizuoka City Shimizu Hosp, Dept Pathol, Shizuoka, Japan.
C3 Tokyo Medical University; Tokai University
RP Kanai, R (corresponding author), Shizuoka City Shimizu Hosp, Dept Neurosurg, Shizuoka, Japan.
EM
RI Hirabayashi, Kenichi/GYA-2785-2022
OI Hirabayashi, Kenichi/0000-0002-9516-1792
CR Bakshi R, 1999, DEMENT GERIATR COGN, V10, P152, DOI 10.1159/000017116
   Bühring U, 2001, NEUROLOGY, V57, P393, DOI 10.1212/WNL.57.3.393
   Carlson BA, 1996, AM J NEURORADIOL, V17, P1695
   DeAngelis L M, 1994, Recent Results Cancer Res, V135, P155
   DEANGELIS LM, 1993, ANN NEUROL, V33, P308, DOI 10.1002/ana.410330313
   Furusawa T, 1998, Radiat Med, V16, P137
   Kleihues PCW., 2000, WORLD HLTH ORG CLASS
   Küker W, 2005, J NEURO-ONCOL, V72, P169, DOI 10.1007/s11060-004-3390-7
   Lewerenz J, 2007, J NEUROL NEUROSUR PS, V78, P777, DOI 10.1136/jnnp.2006.106385
   Matsumoto Katsumi, 1995, Neurologia Medico-Chirurgica, V35, P360, DOI 10.2176/nmc.35.360
   Partap S, 2006, J NEURO-ONCOL, V80, P305, DOI 10.1007/s11060-006-9192-3
   POON T, 1989, J COMPUT ASSIST TOMO, V13, P6, DOI 10.1097/00004728-198901000-00002
   Rollins KE, 2005, HUM PATHOL, V36, P282, DOI 10.1016/j.humpath.2005.01.014
   SCHWAIGHOFER BW, 1989, AM J NEURORADIOL, V10, P725
   Terae S, 1996, NEURORADIOLOGY, V38, P34, DOI 10.1007/BF00593213
   Thurnher MM, 2001, NEURORADIOLOGY, V43, P29, DOI 10.1007/s002340000480
   VAQUERO J, 1984, J NEUROSURG, V60, P174, DOI 10.3171/jns.1984.60.1.0174
NR 17
TC 26
Z9 30
U1 0
U2 4
PU SPRINGER
PI NEW YORK
PA 233 SPRING STREET, NEW YORK, NY 10013 USA
SN 0167-594X
J9 J NEURO-ONCOL
JI J. Neuro-Oncol.
PD JAN
PY 2008
VL 86
IS 1
BP 83
EP 88
DI 10.1007/s11060-007-9437-9
PG 6
WC Oncology; Clinical Neurology
WE Science Citation Index Expanded (SCI-EXPANDED)
SC Oncology; Neurosciences & Neurology
GA 238ZS
UT WOS:000251487800012
PM 17611716
DA 2024-06-03
ER

PT J
AU Green, MR
   Chowdhary, S
   Lombardi, KM
   Chalmers, LM
   Chamberlain, M
AF Green, Myke R.
   Chowdhary, Sajeel
   Lombardi, Kristina M.
   Chalmers, Lisa M.
   Chamberlain, Marc
TI Clinical utility and pharmacology of high-dose methotrexate in the
   treatment of primary CNS lymphoma
SO EXPERT REVIEW OF NEUROTHERAPEUTICS
LA English
DT Review
DE high-dose methotrexate; non-Hodgkin's lymphoma; primary CNS lymphoma
ID NERVOUS-SYSTEM LYMPHOMA; NON-HODGKINS-LYMPHOMA; LONG-TERM SURVIVAL;
   PRIMARY CEREBRAL LYMPHOMA; MALIGNANT BRAIN-TUMORS; POPULATION-BASED
   DATA; STEM-CELL RESCUE; DIHYDROFOLATE-REDUCTASE; INTRAVENTRICULAR
   CHEMOTHERAPY; CLINICOPATHOLOGICAL FEATURES
AB Primary CNS non-Hodgkin lymphoma (PCNSL) has been shown to be increasing in incidence. This appears to be a consequence of the increasing population of those older than 65 years of age in whom PCNSL occurs most often. PCNSL often has a favorable response to treatment and aggressive management may result in extended survival and, in a proportion of patients less than 65 years of age, cure. The majority of neuro-oncologist's advocate utilizing high-dose methotrexate (HD-MTX) as a platform for the chemotherapy treatment of these neoplasms. In this review, the literature regarding HDHMTX as a treatment for PCNSL is summarized as are the pharmacological principles of HD-MTX.
C1 [Green, Myke R.; Chowdhary, Sajeel; Lombardi, Kristina M.; Chalmers, Lisa M.; Chamberlain, Marc] Univ S Florida, H Lee Moffitt Canc Ctr & Res Inst, Dept Interdisciplinary Oncol, Tampa, FL 33611 USA.
C3 H Lee Moffitt Cancer Center & Research Institute; State University
   System of Florida; University of South Florida
RP Chamberlain, M (corresponding author), Univ S Florida, H Lee Moffitt Canc Ctr & Res Inst, Dept Interdisciplinary Oncol, 12902 Magnolia Dr, Tampa, FL 33611 USA.
EM;
  ;
  
CR Abrey LE, 2003, J CLIN ONCOL, V21, P4151, DOI 10.1200/JCO.2003.05.024
   Abrey LE, 1998, J CLIN ONCOL, V16, P859, DOI 10.1200/JCO.1998.16.3.859
   Abrey LE, 2000, J CLIN ONCOL, V18, P3144, DOI 10.1200/JCO.2000.18.17.3144
   Ahsan H, 1995, INT J EPIDEMIOL, V24, P1078, DOI 10.1093/ije/24.6.1078
   Alkins SA, 1996, CANCER, V77, P2123, DOI 10.1002/(SICI)1097-0142(19960515)77:10<2123::AID-CNCR24>3.0.CO;2-W
   ALLEGRA CJ, 1985, NEW ENGL J MED, V313, P184
   ALLEGRA CJ, 1985, J BIOL CHEM, V260, P9720
   [Anonymous], 2006, MICR HEALTHC SER
   ANTONY AC, 1985, J BIOL CHEM, V260, P4911
   Batchelor T, 2003, J CLIN ONCOL, V21, P1044, DOI 10.1200/JCO.2003.03.036
   Batchelor TT, 2003, CLIN CANCER RES, V9, P711
   BERTINO JR, 1990, J CLIN ONCOL, V8, P193, DOI 10.1200/JCO.1990.8.2.193
   Bessell EM, 2004, INT J RADIAT ONCOL, V59, P501, DOI 10.1016/j.ijrobp.2003.11.001
   Bessell EM, 2002, J CLIN ONCOL, V20, P231, DOI 10.1200/JCO.20.1.231
   Bessell EM, 1996, J CLIN ONCOL, V14, P945, DOI 10.1200/JCO.1996.14.3.945
   Blay JY, 1997, B CANCER, V84, P976
   BLEYER WA, 1977, CANCER TREAT REV, V4, P87, DOI 10.1016/S0305-7372(77)80007-8
   BLEYER WA, 1989, CANCER-AM CANCER SOC, V63, P995
   BRADA M, 1990, INT J RADIAT ONCOL, V18, P787, DOI 10.1016/0360-3016(90)90398-4
   Brada M, 1998, INT J RADIAT ONCOL, V40, P1157, DOI 10.1016/S0360-3016(98)00002-9
   BREITHAUPT H, 1982, CANCER TREAT REP, V66, P1733
   Bromberg JEC, 2002, NEUROLOGY, V59, P762, DOI 10.1212/WNL.59.5.762
   CALVERT AH, 1977, CANCER TREAT REP, V61, P1647
   Calvert H, 2002, SEMIN ONCOL, V29, P3, DOI 10.1016/S0093-7754(02)70209-1
   Campbell C, 2003, CURR ONCOL, V10, P102
   CHABNER BA, 1973, J CLIN INVEST, V52, P1804, DOI 10.1172/JCI107362
   CHABNER BA, 1985, J CLIN INVEST, V76, P907, DOI 10.1172/JCI112088
   CHAMBERLAIN MC, 1990, ARCH NEUROL-CHICAGO, V47, P1113, DOI 10.1001/archneur.1990.00530100081017
   Cher L, 1996, NEUROLOGY, V46, P1757, DOI 10.1212/WNL.46.6.1757
   Chu E., 1996, CANC CHEMOTHERAPY BI, P109
   CHUNGI VS, 1978, J PHARM SCI-US, V67, P560, DOI 10.1002/jps.2600670434
   CORDER MP, 1976, CANCER, V37, P1660, DOI 10.1002/1097-0142(197604)37:4<1660::AID-CNCR2820370407>3.0.CO;2-J
   Corn BW, 1997, CANCER, V79, P2409
   Correa DD, 2004, NEUROLOGY, V62, P548, DOI 10.1212/01.WNL.0000109673.75316.D8
   CREAVEN PJ, 1976, BRIT J CANCER, V34, P571, DOI 10.1038/bjc.1976.213
   Dahlborg SA, 1998, CANCER J, V4, P110
   DeAngelis LM, 2004, SEMIN ONCOL, V31, P684, DOI 10.1053/j.seminoncol.2004.07.011
   DeAngelis LM, 2003, J CLIN ONCOL, V21, P4471, DOI 10.1200/JCO.2003.08.900
   DEANGELIS LM, 1992, J CLIN ONCOL, V10, P635, DOI 10.1200/JCO.1992.10.4.635
   DeAngelis LM, 2002, J CLIN ONCOL, V20, P4643, DOI 10.1200/JCO.2002.11.013
   DeVita VT, 2001, CANC PRINCIPLES PRAC, P391
   DIDDENS H, 1984, CANCER TREAT REV, V11, P37, DOI 10.1016/0305-7372(84)90041-0
   DOMIN BA, 1983, CANCER RES, V43, P2155
   DOMIN BA, 1982, MOL PHARMACOL, V21, P478
   Dooley MJ, 2004, BRIT J CANCER, V90, P991, DOI 10.1038/sj.bjc.6601641
   Doolittle N D, 1998, J Neurosci Nurs, V30, P81
   Dorr RT, 1994, CANC CHEMOTHERAPY HD, P698
   DOYLE LA, 1983, ANN INTERN MED, V98, P611, DOI 10.7326/0003-4819-98-5-611
   DRINKA PJ, 1989, J AM GERIATR SOC, V37, P820, DOI 10.1111/j.1532-5415.1989.tb02250.x
   EVANS WE, 1978, CLIN PHARMACOL THER, V23, P68
   EVANS WE, 1979, CANCER CHEMOTH PHARM, V3, P161
   FABRE I, 1984, CANCER RES, V44, P3190
   Ferracini R., 1997, Pathologica (Genoa), V89, P146
   Ferreri AJ, 1996, BRIT J CANCER, V90, P353
   Ferreri AJM, 2000, ANN ONCOL, V11, P927, DOI 10.1023/A:1008376412784
   Ferreri AJM, 2003, J CLIN ONCOL, V21, P2407, DOI 10.1200/JCO.2003.01.135
   Ferreri AJM, 2002, NEUROLOGY, V58, P1513, DOI 10.1212/WNL.58.10.1513
   Ferreri AJM, 2004, BRIT J HAEMATOL, V126, P657, DOI 10.1111/j.1365-2141.2004.05109.x
   Fine HA, 2002, J CLIN ONCOL, V20, P4615, DOI 10.1200/JCO.2002.20.24.4615
   FINE HA, 1993, ANN INTERN MED, V119, P1093, DOI 10.7326/0003-4819-119-11-199312010-00007
   FLEMING RA, 1991, CANCER, V68, P1247, DOI 10.1002/1097-0142(19910915)68:6<1247::AID-CNCR2820680611>3.0.CO;2-O
   Fliessbach K, 2005, NEUROLOGY, V64, P1184, DOI 10.1212/01.WNL.0000156350.49336.E2
   Fliessbach K, 2003, ARCH NEUROL-CHICAGO, V60, P563, DOI 10.1001/archneur.60.4.563
   FREEMAN C, 1972, CANCER, V29, P252, DOI 10.1002/1097-0142(197201)29:1<252::AID-CNCR2820290138>3.0.CO;2-#
   Freilich RJ, 1996, NEUROLOGY, V46, P435, DOI 10.1212/WNL.46.2.435
   FRY DW, 1983, CANCER RES, V43, P1087
   GABBAI AA, 1989, J NEUROSURG, V70, P190, DOI 10.3171/jns.1989.70.2.0190
   GALIVAN J, 1987, ADV ENZYME REGUL, V26, P147, DOI 10.1016/0065-2571(87)90011-2
   GLASS J, 1994, J NEUROSURG, V81, P188, DOI 10.3171/jns.1994.81.2.0188
   GOLDMAN ID, 1985, PHARMACOL THERAPEUT, V28, P77, DOI 10.1016/0163-7258(85)90083-X
   GOLDMAN ID, 1971, BIOCHIM BIOPHYS ACTA, V233, P624, DOI 10.1016/0005-2736(71)90162-3
   Gorlick R, 1996, NEW ENGL J MED, V335, P1041, DOI 10.1056/NEJM199610033351408
   GOULIAN M, 1980, P NATL ACAD SCI-BIOL, V77, P1956, DOI 10.1073/pnas.77.4.1956
   HENDERSON ES, 1965, CANCER RES, V25, P1018
   Herrlinger U, 1998, ACTA NEUROL SCAND, V97, P257
   Herrlinger U, 2002, ANN NEUROL, V51, P247, DOI 10.1002/ana.10102
   Herrlinger U, 2001, CANCER, V91, P130, DOI 10.1002/1097-0142(20010101)91:1<130::AID-CNCR17>3.0.CO;2-8
   Hesketh PJ, 1997, J CLIN ONCOL, V15, P103, DOI 10.1200/JCO.1997.15.1.103
   Hoang-Xuan K, 2003, J CLIN ONCOL, V21, P2726, DOI 10.1200/JCO.2003.11.036
   HOCHBERG FH, 1988, J NEUROSURG, V68, P835, DOI 10.3171/jns.1988.68.6.0835
   ISACOFF WH, 1978, CANCER TREAT REP, V62, P1295
   JACOBS SA, 1976, J CLIN INVEST, V57, P534, DOI 10.1172/JCI108308
   Jahnke K, 2005, ANN ONCOL, V16, P445, DOI 10.1093/annonc/mdi075
   Jahnke K, 2003, BRIT J HAEMATOL, V128, P616
   JELLIFFE RW, 1973, ANN INTERN MED, V79, P604, DOI 10.7326/0003-4819-79-4-604
   JELLIFFE RW, 1971, LANCET, V2, P710
   JOLIVET J, 1983, J CLIN INVEST, V72, P773, DOI 10.1172/JCI111048
   KAMEN BA, 1983, BIOCHEM PHARMACOL, V32, P1837, DOI 10.1016/0006-2952(83)90047-3
   KAMEN BA, 1986, P NATL ACAD SCI USA, V83, P5983, DOI 10.1073/pnas.83.16.5983
   KAMEN BA, 1988, J BIOL CHEM, V263, P13602
   Kepka L, 1998, LEUKEMIA LYMPHOMA, V29, P205, DOI 10.3109/10428199809058397
   KOROSSY KS, 1981, ARCH DERMATOL, V117, P310, DOI 10.1001/archderm.117.5.310
   KROGHJENSEN M, 1995, LEUKEMIA LYMPHOMA, V19, P223
   KROGHJENSEN M, 1994, ANN ONCOL, V5, P349, DOI 10.1093/oxfordjournals.annonc.a058839
   KUMAR P, 1989, BIOCHEM PHARMACOL, V38, P541, DOI 10.1016/0006-2952(89)90397-3
   Linnebank M, 2005, NEUROLOGY, V64, P912, DOI 10.1212/01.WNL.0000152840.26156.74
   Marx GM, 2004, ANN ONCOL, V15, P291, DOI 10.1093/annonc/mdh079
   MATHERLY LH, 1987, PHARMACOL THERAPEUT, V35, P27, DOI 10.1016/0163-7258(87)90104-5
   McGuire J.J., 1984, FOLATES PTERINS, V1, P135
   Mead GM, 2000, CANCER, V89, P1359, DOI 10.1002/1097-0142(20000915)89:6<1359::AID-CNCR21>3.0.CO;2-9
   MILLER DC, 1994, CANCER-AM CANCER SOC, V74, P1383, DOI 10.1002/1097-0142(19940815)74:4<1383::AID-CNCR2820740432>3.0.CO;2-1
   MOLLER H, 1969, J INVEST DERMATOL, V52, P437
   MONJANEL S, 1979, CANCER CHEMOTH PHARM, V3, P189
   Morikawa N, 1997, ANN PHARMACOTHER, V31, P1153, DOI 10.1177/106002809703101007
   NCI Special Exception Protocol E, 2004, NCI SPEC EXC PROT E, P1
   NELSON DF, 1992, INT J RADIAT ONCOL, V23, P9, DOI 10.1016/0360-3016(92)90538-S
   NEUWELT EA, 1991, J CLIN ONCOL, V9, P1580, DOI 10.1200/JCO.1991.9.9.1580
   Nguyen PL, 2005, J CLIN ONCOL, V23, P1507, DOI 10.1200/JCO.2005.01.161
   O'Brien PC, 2005, INT J RAD ONCOL BIOL, V27
   OBrien PC, 1996, MED J AUSTRALIA, V165, P424
   Olson JE, 2002, CANCER, V95, P1504, DOI 10.1002/cncr.10851
   Omuro AMP, 2005, NEUROLOGY, V64, P69, DOI 10.1212/01.WNL.0000148641.98241.5E
   Omuro AMP, 2005, ARCH NEUROL-CHICAGO, V62, P1595, DOI 10.1001/archneur.62.10.1595
   ONEILL BP, 1995, INT J RADIAT ONCOL, V33, P663, DOI 10.1016/0360-3016(95)00207-F
   Park Deric Minwoo, 2002, Expert Opin Pharmacother, V3, P39, DOI 10.1517/14656566.3.1.39
   Pech IV, 1998, ONCOLOGY-NY, V12, P537
   PERRY MC, 1982, SEMIN ONCOL, V9, P65
   Perry MC, 1984, TOXICITY CHEMOTHERAP, P325
   Plotkin SR, 2001, LANCET ONCOL, V2, P455
   Poortmans PMP, 2003, J CLIN ONCOL, V21, P4483, DOI 10.1200/JCO.2003.03.108
   Proudfoot AT, 2004, J TOXICOL-CLIN TOXIC, V42, P1, DOI 10.1081/CLT-120028740
   Qin DX, 1997, AM J CLIN ONCOL-CANC, V20, P263, DOI 10.1097/00000421-199706000-00011
   RICE GC, 1987, P NATL ACAD SCI USA, V84, P9261, DOI 10.1073/pnas.84.24.9261
   ROSENTHAL MA, 1993, AUST NZ J SURG, V63, P30, DOI 10.1111/j.1445-2197.1993.tb00029.x
   Schabet M, 1999, J NEURO-ONCOL, V43, P199, DOI 10.1023/A:1006290032052
   Schlegel U, 2001, J NEUROL NEUROSUR PS, V71, P118, DOI 10.1136/jnnp.71.1.118
   Schultz C, 1996, J CLIN ONCOL, V14, P556, DOI 10.1200/JCO.1996.14.2.556
   Schwartz JI, 2001, J CLIN PHARMACOL, V41, P1120, DOI 10.1177/00912700122012616
   Shenkier TN, 2005, CANCER, V103, P1008, DOI 10.1002/cncr.20868
   Shibamoto Y, 1990, CANCER, V65, P1907
   Shinozaki T, 2000, MED PEDIATR ONCOL, V34, P226, DOI 10.1002/(SICI)1096-911X(200003)34:3<226::AID-MPO15>3.0.CO;2-#
   SHOLAR PW, 1988, BIOCHEM PHARMACOL, V37, P3531, DOI 10.1016/0006-2952(88)90709-5
   SIROTNAK FM, 1979, BIOCHEM PHARMACOL, V28, P2993, DOI 10.1016/0006-2952(79)90599-9
   Sonstein W, 1998, MED ONCOL, V15, P61, DOI 10.1007/BF02787347
   Soussain C, 2004, B CANCER, V91, P189
   STRAW JA, 1984, CANCER RES, V44, P3114
   VALERINO DM, 1972, BIOCHEM PHARMACOL, V21, P821, DOI 10.1016/0006-2952(72)90125-6
   Wall SM, 1996, AM J KIDNEY DIS, V28, P846, DOI 10.1016/S0272-6386(96)90384-4
   WAN SH, 1974, CANCER RES, V34, P3487
   WHITE JC, 1979, J BIOL CHEM, V254, P889
   WIDEMANN BC, 1995, CANCER, V76, P521, DOI 10.1002/1097-0142(19950801)76:3<521::AID-CNCR2820760325>3.0.CO;2-M
   Yang CH, 2000, INT J DERMATOL, V39, P621, DOI 10.1046/j.1365-4362.2000.00022-3.x
   Zhao RB, 2003, ONCOGENE, V22, P7431, DOI 10.1038/sj.onc.1206946
NR 143
TC 26
Z9 28
U1 0
U2 7
PU TAYLOR & FRANCIS LTD
PI ABINGDON
PA 2-4 PARK SQUARE, MILTON PARK, ABINGDON OR14 4RN, OXON, ENGLAND
SN 1473-7175
EI 1744-8360
J9 EXPERT REV NEUROTHER
JI Expert Rev. Neurother.
PD MAY
PY 2006
VL 6
IS 5
BP 635
EP 652
DI 10.1586/14737175.6.5.635
PG 18
WC Clinical Neurology; Pharmacology & Pharmacy
WE Science Citation Index Expanded (SCI-EXPANDED)
SC Neurosciences & Neurology; Pharmacology & Pharmacy
GA V28TC
UT WOS:000208702100008
PM 16734512
DA 2024-06-03
ER

PT J
AU Tachikawa, N
   Goto, M
   Hoshino, Y
   Gatanaga, H
   Yasuoka, A
   Wakabayashi, T
   Katano, H
   Kimura, S
   Oka, S
   Iwamoto, A
AF Tachikawa, N
   Goto, M
   Hoshino, Y
   Gatanaga, H
   Yasuoka, A
   Wakabayashi, T
   Katano, H
   Kimura, S
   Oka, S
   Iwamoto, A
TI Detection of <i>Toxoplasma gondii</i>, Epstein-Barr virus, and JC virus
   DNAs in the cerebrospinal fluid in acquired immunodeficiency syndrome
   patients with focal central nervous system complications
SO INTERNAL MEDICINE
LA English
DT Article
DE brain infection; polymerase chain reaction (PCR); toxoplasmic
   encephalitis; progressive multifocal leukoencephalopathy; primary
   central nervous system (CNS) lymphoma; diagnosis
ID POLYMERASE CHAIN-REACTION; AIDS PATIENTS; CLINICAL-APPLICATION;
   STEREOTAXIC BIOPSY; ANIMAL-MODELS; LESIONS; ENCEPHALITIS; LYMPHOMA;
   NEUROPATHOLOGY; HYBRIDIZATION
AB Object Toxoplasmic encephalitis (TE), primary central nervous system lymphoma (PCNSL) and progressive multifocal leukoencephalopathy (PML) are major central nervous system (CNS) diseases in patients with acquired immunodefieiency syndrome (AIDS). We assessed the diagnostic value of polymerase chain reaction (PCR) in the detection of DNAs of Toxoplasma gondii (T. gondii), Epstein-Barr virus (EBV) and JC virus (JCV) in the cerebrospinal fluid (CSF). Methods We compared the PCR results with those of pathological findings at autopsy. Patients or Materials The present study included 23 autopsies representing those in whom CSF samples were obtained before death while the patient was hospitalized or at autopsy. Results The threshold levels for PCR detection were 4 tachyzoites of T. gondii, 5-15 genomes of EBV and 10 genomes of JCV. We identified T. gondii DNA in 4 out of 5 autopsy-defined cases of TE, EBV DNA in 5 out of 5 cases with PCNSL, and JCV DNA in 2 out of 2 cases with PML. The specificity of PCR was 100% in TE, 78% in PCNSL, and 100% in PML. Conclusion Although the number of cases was relatively small in this study PCR correctly identified T. gondii DNA in those cases in which PML or PCNSL was the sole clinical diagnosis. Our results indicate that PCR examination of CSF is a clinically useful tool for the diagnosis of focal brain lesions in patients with AIDS.
C1 Int Med Ctr Japan, AIDS Clin Ctr, Shinjuku Ku, Tokyo 1628655, Japan.
C3 National Center for Global Health & Medicine - Japan
RP Tachikawa, N (corresponding author), Int Med Ctr Japan, AIDS Clin Ctr, Shinjuku Ku, 1-21-1 Toyama, Tokyo 1628655, Japan.
RI Hoshino, Yoshihiko/B-5159-2019; Hoshino, Yoshihiko/JPX-0557-2023;
   HOSHINO, Yoshihiko/A-2461-2008
OI Hoshino, Yoshihiko/0000-0002-3132-0715; 
CR BIRX DL, 1986, NEW ENGL J MED, V314, P874, DOI 10.1056/NEJM198604033141403
   CHAPPELL ET, 1992, NEUROSURGERY, V30, P825, DOI 10.1227/00006123-199206000-00001
   CINQUE P, 1993, LANCET, V342, P398, DOI 10.1016/0140-6736(93)92814-A
   CIRICILLO SF, 1990, J NEUROSURG, V73, P720, DOI 10.3171/jns.1990.73.5.0720
   CIRICILLO SF, 1991, J NEUROSURG, V74, P1029
   COHN JA, 1989, AM J MED, V86, P521, DOI 10.1016/0002-9343(89)90378-1
   Council of State and Territorial Epidemiologists, 1987, MMWR MORB MORTAL S1, V36, p1S
   Davies J, 1997, AIDS, V11, P1145, DOI 10.1097/00002030-199709000-00010
   Dubois V, 1996, AIDS, V10, P353, DOI 10.1097/00002030-199604000-00001
   FEIDEN W, 1993, ACTA NEUROL SCAND, V87, P228
   GRAY F, 1989, J NEUROL, V236, P273, DOI 10.1007/BF00314455
   HAMILTONDUTOIT SJ, 1991, AM J PATHOL, V138, P149
   HISHIMA T, 1998, PATHOLOGY CLIN MED, V16, P826
   HOWLEY PM, 1980, J VIROL, V36, P878, DOI 10.1128/JVI.36.3.878-882.1980
   Katano H, 1996, PATHOL INT, V46, P568, DOI 10.1111/j.1440-1827.1996.tb03655.x
   KIKUCHI Y, 1992, INTERNAL MED, V31, P1016, DOI 10.2169/internalmedicine.31.1016
   KITADA K, 1991, J CLIN MICROBIOL, V29, P1985, DOI 10.1128/JCM.29.9.1985-1990.1991
   KOVACH NL, 1996, HIV MANUAL GUIDE DIA
   LUFT BJ, 1993, NEW ENGL J MED, V329, P995, DOI 10.1056/NEJM199309303291403
   MACMAHON EME, 1991, LANCET, V338, P969, DOI 10.1016/0140-6736(91)91837-K
   MARTINEZ AJ, 1995, PATHOL RES PRACT, V191, P427, DOI 10.1016/S0344-0338(11)80730-2
   MOSKOWITZ LB, 1984, ARCH PATHOL LAB MED, V108, P867
   OLSEN WL, 1988, RADIOLOGY, V169, P445, DOI 10.1148/radiology.169.2.3174991
   Pelloux H, 1997, AIDS, V11, P1888, DOI 10.1097/00002030-199715000-00018
   PORTER SB, 1992, NEW ENGL J MED, V327, P1643, DOI 10.1056/NEJM199212033272306
   Rosenblum M L, 1988, Clin Neurosurg, V34, P419
   SAEKI K, 1993, DIAGN MOL PATHOL, V2, P108, DOI 10.1097/00019606-199300020-00017
   SCHOONDERMARKVANDEVEN E, 1993, CLIN INFECT DIS, V16, P661, DOI 10.1093/clind/16.5.661
   WEBER T, 1994, AIDS, V8, P49, DOI 10.1097/00002030-199401000-00008
   ZIMMER C, 1992, J NEUROL, V239, P394
NR 30
TC 26
Z9 26
U1 0
U2 4
PU JAPAN SOC INTERNAL MEDICINE
PI TOKYO
PA 34-3 3-CHOME HONGO BUNKYO-KU, TOKYO, 113, JAPAN
SN 0918-2918
J9 INTERNAL MED
JI Intern. Med.
PD JUL
PY 1999
VL 38
IS 7
BP 556
EP 562
DI 10.2169/internalmedicine.38.556
PG 7
WC Medicine, General & Internal
WE Science Citation Index Expanded (SCI-EXPANDED)
SC General & Internal Medicine
GA 215EV
UT WOS:000081369700008
PM 10435361
OA Bronze
DA 2024-06-03
ER

PT J
AU Khalfallah, S
   Stamatoullas, A
   Fruchart, C
   Proust, F
   Delangre, T
   Tilly, H
AF Khalfallah, S
   Stamatoullas, A
   Fruchart, C
   Proust, F
   Delangre, T
   Tilly, H
TI Durable remission of a relapsing primary central nervous system lymphoma
   after autologous bone marrow transplantation
SO BONE MARROW TRANSPLANTATION
LA English
DT Article
DE autologous bone marrow transplantation; non-Hodgkin's lymphoma; central
   nervous system lymphoma
ID HIGH-DOSE CHEMOTHERAPY; NON-HODGKINS-LYMPHOMA; MALIGNANT-LYMPHOMA;
   RADIOTHERAPY; INVOLVEMENT; PATTERNS; FAILURE; BRAIN
AB A 30-year-old patient with a relapsing primary central nervous system lymphoma (PCNSL), was successfully treated with salvage chemotherapy and high-dose therapy including drugs that cross the blood-brain barrier followed by ABMT. Cerebrospinal irradiation was administered after hematological recovery, Six years after transplantation the patient is alive without evidence of recurrent disease with a good neuropsychological status, This result could justify further studies of aggressive management of PCNSL in relapse.
C1 CTR HENRI BECQUEREL,DEPT HEMATOL,F-76038 ROUEN,FRANCE.
   CHU CHARLES NICOLLE,FEDERAT NEUROL,ROUEN,FRANCE.
C3 UNICANCER; Centre Henri Becquerel; Universite de Rouen Normandie
CR Alvarnas J. C., 1995, Blood, V86, p208A
   BLAY JY, 1995, BLOOD, V86, P2922
   BRADA M, 1990, INT J RADIAT ONCOL, V18, P787, DOI 10.1016/0360-3016(90)90398-4
   DEANGELIS LM, 1992, J CLIN ONCOL, V10, P635, DOI 10.1200/JCO.1992.10.4.635
   ERVIN T, 1980, CANCER, V45, P1556, DOI 10.1002/1097-0142(19800401)45:7<1556::AID-CNCR2820450707>3.0.CO;2-B
   Ferreri AJM, 1995, CANCER TREAT REV, V21, P415, DOI 10.1016/0305-7372(95)90028-4
   FRICK JC, 1986, ARCH INTERN MED, V146, P791, DOI 10.1001/archinte.146.4.791
   ISHII E, 1989, AM J CLIN ONCOL-CANC, V12, P132, DOI 10.1097/00000421-198904000-00009
   LOEFFLER JS, 1985, J CLIN ONCOL, V3, P490, DOI 10.1200/JCO.1985.3.4.490
   MICHALSKI JM, 1990, RADIOLOGY, V176, P855, DOI 10.1148/radiology.176.3.2389047
   PHILIP T, 1995, NEW ENGL J MED, V333, P1540, DOI 10.1056/NEJM199512073332305
   POLLACK IF, 1989, CANCER-AM CANCER SOC, V63, P939, DOI 10.1002/1097-0142(19890301)63:5<939::AID-CNCR2820630526>3.0.CO;2-V
   SOCIE G, 1990, CANCER-AM CANCER SOC, V65, P322, DOI 10.1002/1097-0142(19900115)65:2<322::AID-CNCR2820650223>3.0.CO;2-D
   VANDERWALL E, 1995, CANCER TREAT REV, V21, P105, DOI 10.1016/0305-7372(95)90023-3
   WATNE K, 1992, ACTA ONCOL, V31, P545, DOI 10.3109/02841869209088304
NR 15
TC 26
Z9 26
U1 0
U2 0
PU STOCKTON PRESS
PI BASINGSTOKE
PA HOUNDMILLS, BASINGSTOKE, HAMPSHIRE, ENGLAND RG21 6XS
SN 0268-3369
J9 BONE MARROW TRANSPL
JI Bone Marrow Transplant.
PD NOV
PY 1996
VL 18
IS 5
BP 1021
EP 1023
PG 3
WC Biophysics; Oncology; Hematology; Immunology; Transplantation
WE Science Citation Index Expanded (SCI-EXPANDED)
SC Biophysics; Oncology; Hematology; Immunology; Transplantation
GA VT498
UT WOS:A1996VT49800029
PM 8932861
DA 2024-06-03
ER

PT J
AU Chen, FL
   Pang, DW
   Guo, HG
   Ou, QX
   Wu, X
   Jiang, XM
   Wei, XJ
   Liu, SC
   Huang, L
   Liang, ZL
   Zhou, D
   Li, WY
AF Chen, Feili
   Pang, Diwen
   Guo, Hanguo
   Ou, Qiuxiang
   Wu, Xue
   Jiang, Xinmiao
   Wei, Xiaojuan
   Liu, Sichu
   Huang, Ling
   Liang, Zhanli
   Zhou, Dong
   Li, Wenyu
TI Clinical outcomes of newly diagnosed primary CNS lymphoma treated with
   ibrutinib-based combination therapy: A real-world experience of
   off-label ibrutinib use
SO CANCER MEDICINE
LA English
DT Article
ID CENTRAL-NERVOUS-SYSTEM; TYROSINE KINASE; PHASE-II; LENALIDOMIDE
AB Ibrutinib-based combination therapy with high-dose methotrexate (HD-MTX) has recently shown clinical activity against relapse/refractory (R/R) primary central nervous system lymphoma (PCNSL). Herein, we report our real-world experience of treating 11 newly diagnosed PCNSL patients with the ibrutinib/MTX combination. HD-MTX was given at 3.5 g/m(2)every 2-week for eight doses. Ibrutinib was held upon HD-MTX infusion until clearance and was administered daily post-induction until disease progression, intolerable toxicity, or death. Nine out of 11 patients completed the induction phase and received ibrutinib as maintenance therapy. An objective response rate (ORR) of 82% (9/11) was observed including complete response (64%) and partial response (18%). The median progression-free survival (PFS) was 7.4 months while the median overall survival (OS) was not reached. The ibrutinib/MTX combination was well tolerated in these treatment-naive PCNSL patients with an acceptable safety profile. Moreover, the longitudinal analysis of cerebrospinal fluid (CSF) circulating tumor DNA (ctDNA) revealed that CSF ctDNA detection was closely associated with tumor response, and sustained tumor responses correlated with the clearance of ctDNA from the CSF. In sum, our data not only demonstrated the clinical benefit of the ibrutinib and HD-MTX combination regimen in treating newly diagnosed PCNSL patients in a real-world setting, but also highlighted the significance of liquid biopsy including CSF ctDNA in tracing tumor burden and assessing treatment response.
C1 [Chen, Feili; Pang, Diwen; Guo, Hanguo; Wu, Xue; Jiang, Xinmiao; Wei, Xiaojuan; Liu, Sichu; Huang, Ling; Liang, Zhanli; Li, Wenyu] South China Univ Technol, Sch Med, Guangdong Acad Med Sci, Guangdong Prov Peoples Hosp,Lymphoma Div, Guangzhou, Peoples R China.
   [Ou, Qiuxiang] Geneseeq Technol Inc, Translat Med Res Inst, Toronto, ON, Canada.
   [Zhou, Dong] Guangdong Acad Med Sci, Guangdong Prov Peoples Hosp, Dept Neurosurg, Guangzhou, Peoples R China.
C3 Southern Medical University - China; South China University of
   Technology; Guangdong Academy of Medical Sciences & Guangdong General
   Hospital; Geneseeq Technology Inc.; Guangdong Academy of Medical
   Sciences & Guangdong General Hospital; Southern Medical University -
   China
RP Li, WY (corresponding author), South China Univ Technol, Sch Med, Guangdong Acad Med Sci, Guangdong Prov Peoples Hosp,Lymphoma Div, Guangzhou, Peoples R China.; Zhou, D (corresponding author), Guangdong Acad Med Sci, Guangdong Prov Peoples Hosp, Dept Neurosurg, Guangzhou, Peoples R China.
EM
RI wei, xiao/ISB-6027-2023; Liu, Donghua/KEJ-1974-2024; Jiang,
   Tao/IWM-7503-2023; jiang, lei/IWE-1124-2023; zhang, lin/IZQ-4870-2023;
   li, fang/KDO-8841-2024
OI Liu, Donghua/0000-0002-5830-9540; Ou, Qiuxiang/0000-0002-2961-2057
FU Science and Technology Planning Project Foundation of Guangdong
   Province, China [2016A050502028]
FX This study was supported by the Science and Technology Planning Project
   Foundation of Guangdong Province, China (No. 2016A050502028).
CR Batchelor TT, 2016, HEMATOL-AM SOC HEMAT, P379, DOI 10.1182/asheducation-2016.1.379
   Braggio E, 2015, CLIN CANCER RES, V21, P3986, DOI 10.1158/1078-0432.CCR-14-2116
   Bromberg JEC, 2019, LANCET ONCOL, V20, P216, DOI 10.1016/S1470-2045(18)30747-2
   Challa-Malladi M, 2011, CANCER CELL, V20, P728, DOI 10.1016/j.ccr.2011.11.006
   Chukwueke UN, 2019, HEMATOL ONCOL CLIN N, V33, P597, DOI 10.1016/j.hoc.2019.03.008
   Ferreri AJM, 2016, BLOOD, V127, P1642, DOI 10.1182/blood-2015-10-636340
   Ferreri AJM, 2009, LANCET, V374, P1512, DOI 10.1016/S0140-6736(09)61416-1
   Ferreri AJM, 2017, HEMATOL-AM SOC HEMAT, P565, DOI 10.1182/asheducation-2017.1.565
   Ghesquieres H, 2019, ANN ONCOL, V30, P621, DOI 10.1093/annonc/mdz032
   Goy A, 2019, BLOOD, V134, P1024, DOI 10.1182/blood.2018891598
   Grommes C, 2019, BLOOD, V133, P436, DOI 10.1182/blood-2018-09-875732
   Grommes C, 2017, CANCER DISCOV, V7, P1018, DOI 10.1158/2159-8290.CD-17-0613
   Hilal T, 2019, LEUKEMIA LYMPHOMA, V60, P3581, DOI 10.1080/10428194.2019.1633637
   Jeelall YS, 2011, IMMUNOL CELL BIOL, V89, P659, DOI 10.1038/icb.2011.31
   Kasenda B, 2015, ANN ONCOL, V26, P1305, DOI 10.1093/annonc/mdv076
   Langner-Lemercier S, 2016, NEURO-ONCOLOGY, V18, P1297, DOI 10.1093/neuonc/now033
   Lionakis MS, 2017, CANCER CELL, V31, P833, DOI 10.1016/j.ccell.2017.04.012
   Mao CL, 2019, WORLD NEUROSURG, V123, pE15, DOI 10.1016/j.wneu.2018.10.034
   Mendez Joe S, 2018, Am Soc Clin Oncol Educ Book, V38, P604, DOI 10.1200/EDBK_200829
   Montesinos-Rongen M, 2010, ACTA NEUROPATHOL, V120, P529, DOI 10.1007/s00401-010-0709-7
   Morabito F, 2019, EXPERT OPIN PHARMACO, V20, P487, DOI 10.1080/14656566.2018.1561865
   Nakamura T, 2016, NEUROPATH APPL NEURO, V42, P279, DOI 10.1111/nan.12259
   Nayak L, 2017, BLOOD, V129, P3071, DOI 10.1182/blood-2017-01-764209
   Ngo VN, 2011, NATURE, V470, P115, DOI 10.1038/nature09671
   Nijland M, 2018, CANCERS, V10, DOI 10.3390/cancers10110459
   Rubenstein JL, 2018, BLOOD ADV, V2, P1595, DOI 10.1182/bloodadvances.2017014845
   Schmitz R, 2018, NEW ENGL J MED, V378, P1396, DOI 10.1056/NEJMoa1801445
   Shu YQ, 2017, SCI REP-UK, V7, DOI 10.1038/s41598-017-00520-1
   Soussain C, 2019, EUR J CANCER, V117, P121, DOI 10.1016/j.ejca.2019.05.024
   Steidl C, 2011, NATURE, V471, P377, DOI 10.1038/nature09754
   Wilson WH, 2015, NAT MED, V21, P922, DOI 10.1038/nm.3884
   Woyach JA, 2014, NEW ENGL J MED, V370, P2286, DOI 10.1056/NEJMoa1400029
   Yang YB, 2012, CANCER CELL, V21, P723, DOI 10.1016/j.ccr.2012.05.024
   Yang Z, 2018, CLIN CANCER RES, V24, P3097, DOI 10.1158/1078-0432.CCR-17-2310
   Young RM, 2015, SEMIN HEMATOL, V52, P77, DOI 10.1053/j.seminhematol.2015.01.008
NR 35
TC 25
Z9 25
U1 2
U2 10
PU WILEY
PI HOBOKEN
PA 111 RIVER ST, HOBOKEN 07030-5774, NJ USA
SN 2045-7634
J9 CANCER MED-US
JI Cancer Med.
PD NOV
PY 2020
VL 9
IS 22
BP 8676
EP 8684
DI 10.1002/cam4.3499
EA OCT 2020
PG 9
WC Oncology
WE Science Citation Index Expanded (SCI-EXPANDED)
SC Oncology
GA OR0QH
UT WOS:000578039700001
PM 33068336
OA Green Published, gold
DA 2024-06-03
ER

PT J
AU Martinez-Calle, N
   Poynton, E
   Alchawaf, A
   Kassam, S
   Horan, M
   Rafferty, M
   Kelsey, P
   Scott, G
   Culligan, DJ
   Buckley, H
   Lim, YY
   Ngu, L
   McCulloch, R
   Rowntree, C
   Wright, J
   McKay, P
   Fourali, S
   Eyre, TA
   Smith, J
   Osborne, W
   Yallop, D
   Linton, K
   Fox, CP
   Cwynarski, K
AF Martinez-Calle, Nicolas
   Poynton, Edward
   Alchawaf, Alia
   Kassam, Shireen
   Horan, Matthew
   Rafferty, Mark
   Kelsey, Phillipa
   Scott, Gemma
   Culligan, Dominic J.
   Buckley, Hannah
   Lim, Yeong Y.
   Ngu, Loretta
   McCulloch, Rory
   Rowntree, Clare
   Wright, Josh
   McKay, Pamela
   Fourali, Samih
   Eyre, Toby A.
   Smith, Jeffrey
   Osborne, Wendy
   Yallop, Deborah
   Linton, Kim
   Fox, Christopher P.
   Cwynarski, Kate
TI Outcomes of older patients with primary central nervous system lymphoma
   treated in routine clinical practice in the UK: methotrexate dose
   intensity correlates with response and survival
SO BRITISH JOURNAL OF HAEMATOLOGY
LA English
DT Article
DE elderly; PCNSL; DLBCL; real world; high-dose therapy; Mthotrexate
ID PRIMARY CNS LYMPHOMA; INTERNATIONAL EXTRANODAL LYMPHOMA; STEM-CELL
   TRANSPLANTATION; WHOLE-BRAIN RADIOTHERAPY; ELDERLY-PATIENTS; CYTARABINE;
   RITUXIMAB; CHEMOIMMUNOTHERAPY; RANDOMIZATION; CHEMOTHERAPY
AB Data on older patients with primary central nervous system lymphoma (PCNSL) are scarce. Comorbidities and performance status frequently compromise outcomes in this group. Medical records for consecutive patients >= 65 years (n = 244) with PCNSL diagnosed 2012-2017 from 14 UK centres were retrospectively reviewed. Of these 192 patients received methotrexate (MTX)-based treatment. Patients were categorised based on clinician's treatment choice into 'palliative' (n = 52), 'less intensive: MTX +/- rituximab +/- alkylators' (n = 74) and 'intensive: MTX/cytarabine combinations' (n = 118) groups. Complete remission (CR) rate, two-year progression-free survival (PFS) and overall survival (OS) rates were 49%, 11% and 24% for the less intensive and 69%, 40% and 50% for the intensive groups. Treatment-related mortality (TRM) was 6 center dot 8% for MTX-treated patients. Median MTX cumulative dose was 8 center dot 8 g/m(2) (range 1 center dot 5-21) over a median of three cycles. Higher relative dose intensity of MTX (MTX-RDI) was associated with improved PFS and OS in both groups adjusting for age, Eastern cooperative oncology group (ECOG) score and baseline parameters. Two-year PFS and OS for patients receiving four or more induction cycles followed by consolidation (n = 36) were 65% and 70% respectively. Older patients completing MTX-based induction and consolidation had clinical outcomes similar to those in younger cohorts. These retrospective data suggest that maximising MTX-RDI and delivering consolidation in a subgroup of older patients may improve clinical outcomes.
C1 [Martinez-Calle, Nicolas; Fox, Christopher P.] Nottingham Univ Hosp NHS Trust, Dept Clin Haematol, City Hosp Campus,Hucknall Rd, Nottingham NG5 1PB, England.
   [Poynton, Edward; Cwynarski, Kate] Univ Coll London Hosp NHS Fdn Trust, London, England.
   [Alchawaf, Alia; Linton, Kim] Christie NHS Fdn Trust, Manchester, Lancs, England.
   [Kassam, Shireen; Yallop, Deborah] Kings Coll Hosp NHS Fdn Trust, London, England.
   [Horan, Matthew; Osborne, Wendy] Newcastle Upon Tyne Hosp NHS Trust, Newcastle, NSW, Australia.
   [Rafferty, Mark; McKay, Pamela] Beatson West Scotland Canc Ctr, Dept Haematol, Glasgow, Lanark, Scotland.
   [Kelsey, Phillipa; Wright, Josh] Sheffield Teaching Hosp NHS Trust, Sheffield, S Yorkshire, England.
   [Scott, Gemma; Rowntree, Clare] Univ Hosp Wales, Cardiff, Wales.
   [Culligan, Dominic J.] Aberdeen Royal Infirm, Aberdeen, Scotland.
   [Buckley, Hannah] Cambridge Univ Hosp NHS Trust, Cambridge, England.
   [Lim, Yeong Y.; Smith, Jeffrey] Aintree Univ Hosp NHS Fdn Trust, Liverpool, Merseyside, England.
   [Ngu, Loretta] Royal Devon & Exeter NHS Fdn Trust, Exeter, Devon, England.
   [McCulloch, Rory] Univ Hosp Plymouth NHS Trust, Plymouth, Devon, England.
   [Fourali, Samih; Eyre, Toby A.] Oxford Univ Hosp NHS Fdn Trust, Dept Haematol, Oxford, England.
C3 Nottingham University Hospital NHS Trust; Nottingham City Hospital;
   University of London; University College London; University College
   London Hospitals NHS Foundation Trust; Christie NHS Foundation Trust;
   King's College Hospital NHS Foundation Trust; Beatson Oncology Centre;
   University of Sheffield; Cardiff University; University of Aberdeen;
   Aintree University Hospitals NHS Foundation Trust; University of Exeter;
   Oxford University Hospitals NHS Foundation Trust
RP Martinez-Calle, N (corresponding author), Nottingham Univ Hosp NHS Trust, Dept Clin Haematol, City Hosp Campus,Hucknall Rd, Nottingham NG5 1PB, England.
EM
RI Martinez-Calle, Nicolas/ABE-4394-2020; Linton, Kim M/O-9903-2015
OI Martinez-Calle, Nicolas/0000-0002-5184-9464; Lim, Yeong
   Jer/0000-0002-9022-2904; Fox, Christopher Paul/0000-0002-6322-9254
CR Abrey LE, 2005, J CLIN ONCOL, V23, P5034, DOI 10.1200/JCO.2005.13.524
   Bromberg J, 2017, BLOOD, V130
   Bromberg JEC, 2019, LANCET ONCOL, V20, P216, DOI 10.1016/S1470-2045(18)30747-2
   Farhi J, 2018, ANN HEMATOL, V97, P2391, DOI 10.1007/s00277-018-3468-5
   Ferreri AJM, 2017, LANCET HAEMATOL, V4, pE510, DOI 10.1016/S2352-3026(17)30174-6
   Ferreri AJM, 2016, LANCET HAEMATOL, V3, pE217, DOI 10.1016/S2352-3026(16)00036-3
   Fox CP, 2019, BRIT J HAEMATOL, V184, P348, DOI 10.1111/bjh.15661
   Fritsch K, 2017, LEUKEMIA, V31, P846, DOI 10.1038/leu.2016.334
   GHESQUIERES H, 2016, BLOOD, V128
   Gittleman HR, 2015, CANCER-AM CANCER SOC, V121, P102, DOI 10.1002/cncr.29015
   Grommes C., 2018, Blood, V132, P2965, DOI [10.1182/blood-2018-99-118538, DOI 10.1182/BLOOD-2018-99-118538]
   Grommes C, 2016, BLOOD, V128, DOI 10.1182/blood.V128.22.783.783
   Houillier C, 2019, J CLIN ONCOL, V37, P823, DOI 10.1200/JCO.18.00306
   Kasenda B, 2015, ANN ONCOL, V26, P1305, DOI 10.1093/annonc/mdv076
   Lee W, 2019, HEMATOL ONCOL, V37, P459, DOI DOI 10.1002/HON.136_2631
   Madle M, 2015, ANN HEMATOL, V94, P1853, DOI 10.1007/s00277-015-2470-4
   Mendez JS, 2018, NEURO-ONCOLOGY, V20, P687, DOI 10.1093/neuonc/nox187
   Morris PG, 2013, J CLIN ONCOL, V31, P3971, DOI 10.1200/JCO.2013.50.4910
   Nayak L, 2013, CURR TREAT OPTION ON, V14, P539, DOI 10.1007/s11864-013-0252-6
   Ney DE, 2010, CANCER-AM CANCER SOC, V116, P4605, DOI 10.1002/cncr.25363
   Omuro A, 2015, LANCET HAEMATOL, V2, pE251, DOI 10.1016/S2352-3026(15)00074-5
   Omuro A, 2015, BLOOD, V125, P1403, DOI 10.1182/blood-2014-10-604561
   Rigau V, 2011, BRAIN PATHOL, V21, P633, DOI 10.1111/j.1750-3639.2011.00491.x
   Roth P, 2014, CURR OPIN NEUROL, V27, P697, DOI 10.1097/WCO.0000000000000145
   Rubenstein JL, 2013, J CLIN ONCOL, V31, P3061, DOI 10.1200/JCO.2012.46.9957
   Schmitt AM, 2019, HEMATOL ONCOL, V37, P548, DOI 10.1002/hon.2666
   Schorb E, 2019, HEMATOL ONCOL, V37, P271, DOI [10.1002/hon.81_2630, DOI 10.1002/HON.81_2630]
   Schorb E, 2019, BMC CANCER, V19, DOI 10.1186/s12885-019-5473-z
   Schuurmans M, 2010, BRIT J HAEMATOL, V151, P179, DOI 10.1111/j.1365-2141.2010.08328.x
   Siegal T, 2019, ACTA HAEMATOL-BASEL, V141, P138, DOI 10.1159/000495284
   Soussain C, 2017, HEMATOL ONCOL, V35, P72
NR 31
TC 25
Z9 25
U1 1
U2 2
PU WILEY
PI HOBOKEN
PA 111 RIVER ST, HOBOKEN 07030-5774, NJ USA
SN 0007-1048
EI 1365-2141
J9 BRIT J HAEMATOL
JI Br. J. Haematol.
PD AUG
PY 2020
VL 190
IS 3
BP 394
EP 404
DI 10.1111/bjh.16592
EA MAR 2020
PG 11
WC Hematology
WE Science Citation Index Expanded (SCI-EXPANDED)
SC Hematology
GA MT0RK
UT WOS:000522491900001
PM 32232989
OA Bronze
DA 2024-06-03
ER

PT J
AU Liu, CJ
   Lin, SY
   Yang, CF
   Yeh, CM
   Kuan, AS
   Wang, HY
   Tsai, CK
   Gau, JP
   Hsiao, LT
   Chen, PM
   Liu, YC
   Hong, YC
   Ko, PS
   Liu, JH
   Lin, CH
AF Liu, Chia-Jen
   Lin, Shinn-Yn
   Yang, Ching-Fen
   Yeh, Chiu-Mei
   Kuan, Ai-Seon
   Wang, Hao-Yuan
   Tsai, Chun-Kuang
   Gau, Jyh-Pyng
   Hsiao, Liang-Tsai
   Chen, Po-Min
   Liu, Yao-Chung
   Hong, Ying-Chung
   Ko, Po-Shen
   Liu, Jin-Hwang
   Lin, Chia-Hsin
TI A new prognostic score for disease progression and mortality in patients
   with newly diagnosed primary CNS lymphoma
SO CANCER MEDICINE
LA English
DT Article
DE epidemiology; external validation; primary CNS lymphoma; prognostic
   model
ID NERVOUS-SYSTEM LYMPHOMA; B-CELL LYMPHOMA; INTERNATIONAL EXTRANODAL
   LYMPHOMA; WHOLE-BRAIN RADIOTHERAPY; HIGH-DOSE CHEMOTHERAPY;
   ELDERLY-PATIENTS; SURVIVAL; METHOTREXATE; TRENDS; AGE
AB Background Although various prognostic models for primary central nervous system lymphoma (PCNSL) have been developed, there is no consensus regarding the optimal prognostic index. We aimed to evaluate potential prognostic factors and construct a novel predictive model for PCNSL patients. Methods We enrolled newly diagnosed PCNSL patients between 2003 and 2015. The primary endpoint was progression-free survival (PFS), and the secondary endpoint was overall survival (OS). The prognostic factors identified using multivariate Cox proportional hazards models were used to develop a predictive model. We subsequently validated the prognostic model in an independent cohort. We also evaluated the validity of the existing scores: the International Extranodal Lymphoma Study Group (IELSG), the Nottingham/Barcelona (NB), and the Memorial Sloan-Kettering Cancer Center models (MSKCC). Results We identified 101 patients with newly diagnosed PCNSL at our center. Multivariate analysis showed that age >= 80, deep brain lesions, and ECOG >= 2 were independent risk factors of PFS. Assigning one point for each factor, we constructed a novel prognostic model, the Taipei Score, with four distinct risk groups (0-3 points). The performances of the Taipei Score in discriminating both PFS and OS in the training cohort were significant, and the score was validated in the external validation cohort. The IELSG, NB and MSKCC models had insufficient discriminative ability for either PFS or OS in both cohorts. Conclusion The Taipei Score is a simple model that discriminates PFS and OS for PCNSL patients. The score may offer disease risk stratification and facilitate clinical decision-making.
C1 [Liu, Chia-Jen; Yeh, Chiu-Mei; Wang, Hao-Yuan; Tsai, Chun-Kuang; Gau, Jyh-Pyng; Hsiao, Liang-Tsai; Chen, Po-Min; Liu, Yao-Chung; Ko, Po-Shen; Liu, Jin-Hwang; Lin, Chia-Hsin] Taipei Vet Gen Hosp, Dept Med, Div Hematol & Oncol, Taipei, Taiwan.
   [Liu, Chia-Jen; Yeh, Chiu-Mei] Natl Yang Ming Univ, Inst Publ Hlth, Taipei, Taiwan.
   [Lin, Shinn-Yn; Lin, Chia-Hsin] Linkou Chang Gung Mem Hosp, Dept Radiat Oncol, Med Ctr, Taoyuan, Taiwan.
   [Lin, Shinn-Yn] Chang Gung Univ, Coll Med, Dept Med Imaging & Radiol Sci, Taoyuan, Taiwan.
   [Yang, Ching-Fen] Taipei Vet Gen Hosp, Dept Pathol & Lab Med, Taipei, Taiwan.
   [Kuan, Ai-Seon] Univ Oxford, Nuffield Dept Populat Hlth, Canc Epidemiol Unit, Oxford, England.
   [Gau, Jyh-Pyng; Hsiao, Liang-Tsai; Chen, Po-Min; Liu, Yao-Chung; Hong, Ying-Chung; Ko, Po-Shen; Lin, Chia-Hsin] Natl Yang Ming Univ, Sch Med, Taipei, Taiwan.
   [Hong, Ying-Chung; Liu, Jin-Hwang] Kaohsiung Vet Gen Hosp, Div Hematol & Oncol, Kaohsiung, Taiwan.
   [Liu, Jin-Hwang] Natl Yang Ming Univ, Inst Biopharmaceut Sci, Taipei, Taiwan.
   [Lin, Chia-Hsin] Natl Yang Ming Univ, Chong Hin Loon Mem Canc & Biotherapy Res Ctr, Taipei, Taiwan.
C3 Taipei Veterans General Hospital; National Yang Ming Chiao Tung
   University; Chang Gung Memorial Hospital; Chang Gung University; Taipei
   Veterans General Hospital; University of Oxford; National Yang Ming
   Chiao Tung University; Kaohsiung Veterans General Hospital; National
   Yang Ming Chiao Tung University; National Yang Ming Chiao Tung
   University
RP Lin, CH (corresponding author), Taipei Vet Gen Hosp, Dept Med, Div Hematol & Oncol, Taipei, Taiwan.
EM
RI wang, hao/HSE-7975-2023; wang, haoyuan/GWR-3322-2022; Lin,
   Celine/KFB-7293-2024; Han, Liang/KFR-6745-2024
OI Liu, Jin-Hwang/0000-0001-8201-5529; Liu, Chia-Jen/0000-0001-7945-9292
FU Taipei Veterans General Hospital [V105B-016, V105E10-002-MY2-1];
   Ministry of Science and Technology [MOST 104-2314-B-075-085-MY2, MOST
   105-2314-B-418-003-MY3]; Taiwan Clinical Oncology Research Foundation;
   Szu-Yuan Research Foundation of Internal Medicine; Chong Hin Loon
   Memorial Cancer and Biotherapy Research Center
FX Taipei Veterans General Hospital, Grant/Award Number: V105B-016 and
   V105E10-002-MY2-1; Ministry of Science and Technology, Grant/Award
   Number: MOST 104-2314-B-075-085-MY2 and MOST 105-2314-B-418-003-MY3;
   Taiwan Clinical Oncology Research Foundation; Szu-Yuan Research
   Foundation of Internal Medicine; Chong Hin Loon Memorial Cancer and
   Biotherapy Research Center
CR Abrey LE, 2006, J CLIN ONCOL, V24, P5711, DOI 10.1200/JCO.2006.08.2941
   Abrey LE, 2009, CURR OPIN NEUROL, V22, P675, DOI 10.1097/WCO.0b013e328332533b
   Bessell EM, 2004, INT J RADIAT ONCOL, V59, P501, DOI 10.1016/j.ijrobp.2003.11.001
   Bromberg JEC, 2019, LANCET ONCOL, V20, P216, DOI 10.1016/S1470-2045(18)30747-2
   Corry J, 1998, INT J RADIAT ONCOL, V41, P615, DOI 10.1016/S0360-3016(97)00571-3
   Ferreri AJM, 2003, J CLIN ONCOL, V21, P266, DOI 10.1200/JCO.2003.09.139
   Ferreri AJM, 2002, NEUROLOGY, V58, P1513, DOI 10.1212/WNL.58.10.1513
   Ferreri AJM, 2016, LANCET HAEMATOL, V3, pE217, DOI 10.1016/S2352-3026(16)00036-3
   Ferreri AJM, 2009, LANCET, V374, P1512, DOI 10.1016/S0140-6736(09)61416-1
   Fraser E, 2015, CHIN CLIN ONCOL, V4, DOI 10.3978/j.issn.2304-3865.2015.02.01
   Ghesquières H, 2010, ANN ONCOL, V21, P842, DOI 10.1093/annonc/mdp529
   Grommes C, 2017, J CLIN ONCOL, V35, P2410, DOI 10.1200/JCO.2017.72.7602
   Harrell FE, 1996, STAT MED, V15, P361, DOI 10.1002/(SICI)1097-0258(19960229)15:4<361::AID-SIM168>3.0.CO;2-4
   Hoang-Xuan K, 2015, LANCET ONCOL, V16, pE322, DOI 10.1016/S1470-2045(15)00076-5
   Jahnke K, 2006, J NEURO-ONCOL, V80, P159, DOI 10.1007/s11060-006-9165-6
   Jahr G, 2018, BRAIN BEHAV, V8, DOI 10.1002/brb3.928
   Jang JE, 2016, EUR J CANCER, V57, P127, DOI 10.1016/j.ejca.2016.01.016
   Kasenda B, 2015, ANN ONCOL, V26, P1305, DOI 10.1093/annonc/mdv076
   Lanic H, 2014, LEUKEMIA LYMPHOMA, V55, P817, DOI 10.3109/10428194.2013.816421
   Le M, 2019, ONCOLOGIST, V24, pE898, DOI 10.1634/theoncologist.2018-0629
   Lin CH, 2019, J CANCER, V10, P3958, DOI 10.7150/jca.32467
   Löw S, 2018, THER ADV NEUROL DISO, V11, DOI 10.1177/1756286418793562
   Mantripragada K, 2015, BLOOD, V126
   Melchardt T, 2015, J NATL COMPR CANC NE, V13, P1501, DOI 10.6004/jnccn.2015.0178
   Mendez JS, 2018, NEURO-ONCOLOGY, V20, P687, DOI 10.1093/neuonc/nox187
   Morris PG, 2013, J CLIN ONCOL, V31, P3971, DOI 10.1200/JCO.2013.50.4910
   Norden AD, 2011, J NEURO-ONCOL, V101, P487, DOI 10.1007/s11060-010-0269-7
   Omuro A, 2015, BLOOD, V125, P1403, DOI 10.1182/blood-2014-10-604561
   Panageas KS, 2005, CANCER-AM CANCER SOC, V104, P2466, DOI 10.1002/cncr.21481
   Rubenstein JL, 2013, J CLIN ONCOL, V31, P3061, DOI 10.1200/JCO.2012.46.9957
   Schorb E, 2013, HAEMATOLOGICA, V98, P765, DOI 10.3324/haematol.2012.076075
   Shiels MS, 2016, BRIT J HAEMATOL, V174, P417, DOI 10.1111/bjh.14073
   Sinicrope K, 2018, NEUROL CLIN, V36, P517, DOI 10.1016/j.ncl.2018.04.008
   Swerdlow SH, 2016, BLOOD, V127, P2375, DOI 10.1182/blood-2016-01-643569
   Thiel E, 2010, LANCET ONCOL, V11, P1036, DOI 10.1016/S1470-2045(10)70229-1
   Troppan KT, 2014, BRIT J CANCER, V111, P55, DOI 10.1038/bjc.2014.277
   Villano JL, 2011, BRIT J CANCER, V105, P1414, DOI 10.1038/bjc.2011.357
   Welch MR, 2012, NEURO-ONCOLOGY, V14, P1304, DOI 10.1093/neuonc/nos207
   Wieduwilt MJ, 2012, CLIN CANCER RES, V18, P1146, DOI 10.1158/1078-0432.CCR-11-0625
NR 39
TC 25
Z9 26
U1 0
U2 2
PU WILEY
PI HOBOKEN
PA 111 RIVER ST, HOBOKEN 07030-5774, NJ USA
SN 2045-7634
J9 CANCER MED-US
JI Cancer Med.
PD MAR
PY 2020
VL 9
IS 6
BP 2134
EP 2145
DI 10.1002/cam4.2872
EA FEB 2020
PG 12
WC Oncology
WE Science Citation Index Expanded (SCI-EXPANDED)
SC Oncology
GA KT4WK
UT WOS:000510507200001
PM 32011103
OA gold, Green Published
DA 2024-06-03
ER

PT J
AU Saini, J
   Gupta, PK
   Awasthi, A
   Pandey, CM
   Singh, A
   Patir, R
   Ahlawat, S
   Sadashiva, N
   Mahadevan, A
   Gupta, RK
AF Saini, J.
   Gupta, P. Kumar
   Awasthi, A.
   Pandey, C. M.
   Singh, A.
   Patir, R.
   Ahlawat, S.
   Sadashiva, N.
   Mahadevan, A.
   Gupta, R. Kumar
TI Multiparametric imaging-based differentiation of lymphoma and
   glioblastoma: using T1-perfusion, diffusion, and susceptibility-weighted
   MRI
SO CLINICAL RADIOLOGY
LA English
DT Article
ID NERVOUS-SYSTEM LYMPHOMA; PRIMARY CNS LYMPHOMA; CONTRAST-ENHANCED-MRI;
   POSITRON-EMISSION-TOMOGRAPHY; CEREBRAL BLOOD-VOLUME; EPSTEIN-BARR-VIRUS;
   DCE-MRI; IMMUNOCOMPETENT PATIENTS; DIAGNOSTIC-ACCURACY; PERFUSION
AB AIM: To compare the diagnostic performance of T1 perfusion magnetic resonance imaging (MRI), diffusion-weighted imaging (DWI), and susceptibility-weighted imaging (SWI) for differentiating primary central nervous system lymphoma (PCNSL) and glioblastoma (GBM).
   MATERIALS AND METHODS: This retrospective study comprised a cohort of 70 patients with glioblastoma and 30 patients with PCNSL. T1 perfusion MRI-derived rCBV_corr (leakage corrected relative cerebral blood volume), apparent diffusion coefficient (ADC) derived from DWI, and intratumoural susceptibility signals intensity (ITSS) measured on SWI were evaluated in these 100 patients. The Mann-Whitney U-test was used for pairwise comparison between groups. The diagnostic performance for differentiating PCNSL from glioblastoma was evaluated by using univariate and multivariable logistic regression analyses and receiver operating characteristic (ROC) analysis.
   RESULTS: Minimum ADC, maximum rCBVs_corr, k(ep) (back flux exchange rate), and ITSS scores were significantly lower in patients with PCNSL than in those with glioblastoma (p<0.05). On ROC analysis, ITSS showed the best discrimination ability for differentiation of GBM and PCNSL with an area under the ROC curve (AUC) of 0.80. rCBV_corr and ADC showed AUCs of 0.68 and 0.63, respectively. Multiparametric assessment using ADC, rCBV_corr, k(ep), and ITSS scores significantly increased the diagnostic ability for differentiating PCNSL from GBM as compared to mean ADC, mean rCBV_corr, and ITSS alone or a combination of these parameters. The multiparametric model could correctly discriminate 84% of tumours with a sensitivity and specificity of 90% and 70% with an AUC of 0.92.
   CONCLUSION: Multiparametric MRI evaluation using DWI, T1 perfusion MRI, and SWI enabled reliable differentiation of PCNSL and GBM in the majority patients, and these results support an integration of advanced MRI techniques for the diagnostic work-up of patients with these tumours. (C) 2018 The Royal College of Radiologists. Published by Elsevier Ltd. All rights reserved.
C1 [Saini, J.] Natl Inst Mental Hlth & Neurosci, Neuroimaging & Intervent Radiol, Bangalore, Karnataka, India.
   [Gupta, P. Kumar; Gupta, R. Kumar] Fortis Mem Res Inst, Dept Radiol, Gurugram, India.
   [Gupta, P. Kumar; Gupta, R. Kumar] Fortis Mem Res Inst, Dept Imaging, Gurugram, India.
   [Awasthi, A.; Pandey, C. M.] Sanjay Gandhi Post Grad Inst Med Sci, Biostat, Lucknow, Uttar Pradesh, India.
   [Singh, A.] Indian Inst Technol Delhi, Ctr Biomed Engn, Delhi, India.
   [Patir, R.] Fortis Mem Res Inst, Dept Neurosurg, Gurugram, India.
   [Ahlawat, S.] Fortis Mem Res Inst, SRL Diagnost, Gurugram, India.
   [Sadashiva, N.] Natl Inst Mental Hlth & Neurosci, Dept Neurosurg, Bangalore, Karnataka, India.
   [Mahadevan, A.] Natl Inst Mental Hlth & Neurosci, Dept Neuropathol, Bangalore, Karnataka, India.
C3 National Institute of Mental Health & Neurosciences - India; Sanjay
   Gandhi Postgraduate Institute of Medical Sciences; Indian Institute of
   Technology System (IIT System); Indian Institute of Technology (IIT) -
   Delhi; National Institute of Mental Health & Neurosciences - India;
   National Institute of Mental Health & Neurosciences - India
RP Gupta, RK (corresponding author), Dept Radiol, Gurgaon 122002, Haryana, India.; Gupta, RK (corresponding author), Imaging Fortis Mem Res Inst, Gurgaon 122002, Haryana, India.
EM
RI Sadashiva, Nishanth/J-9587-2015; Awasthi, Ashish/Y-5915-2019
OI Awasthi, Ashish/0000-0002-9308-9782; GUPTA, PRADEEP
   KUMAR/0000-0002-4036-3899; Singh, Anup/0000-0001-6744-8326
CR Abe T, 2015, MAGN RESON MED SCI, V14, P313, DOI 10.2463/mrms.2014-0083
   Alifieris C, 2015, PHARMACOL THERAPEUT, V152, P63, DOI 10.1016/j.pharmthera.2015.05.005
   Awasthi R, 2012, NEURORADIOLOGY, V54, P205, DOI 10.1007/s00234-011-0874-y
   Barajas RF, 2015, CLIN RADIOL, V70, P1087, DOI 10.1016/j.crad.2015.03.006
   Blasel S, 2013, J NEURO-ONCOL, V114, P127, DOI 10.1007/s11060-013-1161-z
   Boxerman JL, 2006, AM J NEURORADIOL, V27, P859
   Choi YS, 2017, EUR RADIOL, V27, P1344, DOI 10.1007/s00330-016-4484-2
   Furtner J, 2014, EUR J RADIOL, V83, P806, DOI 10.1016/j.ejrad.2014.01.017
   Guo AC, 2002, RADIOLOGY, V224, P177, DOI 10.1148/radiol.2241010637
   Haldorsen IS, 2011, AM J NEURORADIOL, V32, P984, DOI 10.3174/ajnr.A2171
   Hoang-Xuan K, 2015, LANCET ONCOL, V16, pE322, DOI 10.1016/S1470-2045(15)00076-5
   Horger M, 2009, AM J ROENTGENOL, V193, P1384, DOI 10.2214/AJR.09.2486
   de la Peña MDJ, 2017, CURR PROBL DIAGN RAD, V46, P136, DOI 10.1067/j.cpradiol.2016.04.004
   Kickingereder P, 2014, AM J NEURORADIOL, V35, P1503, DOI 10.3174/ajnr.A3915
   Kickingereder P, 2014, RADIOLOGY, V272, P843, DOI 10.1148/radiol.14132740
   Kim EY, 2005, ACTA RADIOL, V46, P187, DOI 10.1080/02841850510022714
   Kim HS, 2009, AM J NEURORADIOL, V30, P1574, DOI 10.3174/ajnr.A1635
   Kim IY, 2008, SURG NEUROL, V70, P308, DOI 10.1016/j.surneu.2007.04.009
   Lee IH, 2010, EUR J RADIOL, V76, P48, DOI 10.1016/j.ejrad.2009.05.013
   Liao WH, 2009, ACTA RADIOL, V50, P217, DOI 10.1080/02841850802616752
   Lin X, 2017, AM J NEURORADIOL, V38, P485, DOI 10.3174/ajnr.A5023
   Lu SS, 2017, J COMPUT ASSIST TOMO, V41, P898, DOI 10.1097/RCT.0000000000000622
   Makino K, 2011, ANN NUCL MED, V25, P432, DOI 10.1007/s12149-011-0483-1
   Malikova H, 2016, CANCER IMAGING, V16, DOI 10.1186/s40644-016-0098-9
   Murayama K, 2018, MAGN RESON MED SCI, V17, P42, DOI 10.2463/mrms.mp.2016-0113
   Nakajima S, 2015, CLIN RADIOL, V70, P1393, DOI 10.1016/j.crad.2015.08.004
   Nakajima S, 2015, CLIN IMAG, V39, P390, DOI 10.1016/j.clinimag.2014.12.002
   Park MJ, 2009, AM J NEURORADIOL, V30, P1402, DOI 10.3174/ajnr.A1593
   Peters S, 2012, ROFO-FORTSCHR RONTG, V184, P713, DOI 10.1055/s-0032-1312862
   Radbruch A, 2013, EUR J RADIOL, V82, P552, DOI 10.1016/j.ejrad.2012.11.002
   Rubenstein J, 2002, J NEURO-ONCOL, V58, P53, DOI 10.1023/A:1015887312455
   Sahoo P, 2017, MAGN RESON IMAGING, V44, P32, DOI 10.1016/j.mri.2017.08.003
   Sahoo P, 2013, J MAGN RESON IMAGING, V38, P677, DOI 10.1002/jmri.24021
   Server A, 2009, ACTA RADIOL, V50, P682, DOI 10.1080/02841850902933123
   Sharma MC, 2016, INDIAN J MED RES, V143, P605, DOI 10.4103/0971-5916.187109
   Singh A, 2007, J MAGN RESON IMAGING, V26, P871, DOI 10.1002/jmri.21080
   Singh A, 2009, J MAGN RESON IMAGING, V29, P166, DOI 10.1002/jmri.21624
   Suh CH, 2014, RADIOLOGY, V272, P504, DOI 10.1148/radiol.14131895
   Takeuchi H, 2007, J NEURO-ONCOL, V84, P141, DOI 10.1007/s11060-007-9363-x
   Tandon A, 2009, NEUROL INDIA, V57, P36, DOI 10.4103/0028-3886.48813
   Toh CH, 2013, AM J NEURORADIOL, V34, P1145, DOI 10.3174/ajnr.A3383
   Wang S, 2011, AM J NEURORADIOL, V32, P507, DOI 10.3174/ajnr.A2333
   Xing Z, 2014, CLIN NEURORADIOL, V24, P329, DOI 10.1007/s00062-013-0255-5
   Yamashita K, 2013, NEURORADIOLOGY, V55, P135, DOI 10.1007/s00234-012-1089-6
   Zacharia TT, 2008, J NEUROIMAGING, V18, P411, DOI 10.1111/j.1552-6569.2007.00231.x
   Zaharchuk G, 2007, AM J NEURORADIOL, V28, P1850, DOI 10.3174/ajnr.A0831
NR 46
TC 25
Z9 26
U1 0
U2 8
PU W B SAUNDERS CO LTD
PI LONDON
PA 32 JAMESTOWN RD, LONDON NW1 7BY, ENGLAND
SN 0009-9260
EI 1365-229X
J9 CLIN RADIOL
JI Clin. Radiol.
PD NOV
PY 2018
VL 73
IS 11
AR 986.e7
DI 10.1016/j.crad.2018.07.107
PG 9
WC Radiology, Nuclear Medicine & Medical Imaging
WE Science Citation Index Expanded (SCI-EXPANDED)
SC Radiology, Nuclear Medicine & Medical Imaging
GA GV9LG
UT WOS:000446479000024
PM 30197047
DA 2024-06-03
ER

PT J
AU Cho, H
   Kim, SH
   Kim, SJ
   Chang, JH
   Yang, WI
   Suh, CO
   Kim, YR
   Jang, JE
   Cheong, JW
   Min, YH
   Kim, JS
AF Cho, Hyunsoo
   Kim, Se Hoon
   Kim, Soo-Jeong
   Chang, Jong Hee
   Yang, Woo-Ick
   Suh, Chang-Ok
   Kim, Yu Ri
   Jang, Ji Eun
   Cheong, June-Won
   Min, Yoo Hong
   Kim, Jin Seok
TI Programmed cell death 1 expression is associated with inferior survival
   in patients with primary central nervous system lymphoma
SO ONCOTARGET
LA English
DT Article
DE primary central nervous system lymphoma; programmed cell death 1;
   programmed cell death-ligand 1; programmed cell death-ligand 2;
   prognosis
ID PD-1 BLOCKADE; NIVOLUMAB
AB Programmed cell death 1 (PD-1) and its ligands PD-L1/PD-L2 have been shown to mediate immune evasion in various cancers, but their prognostic implications in patients with primary central nervous system lymphoma (PCNSL) are poorly understood. Therefore, we analyzed 76 PCNSL patients at initial diagnosis who were treated homogenously with high-dose methotrexate-based chemotherapy, and evaluated the prognostic roles of high immunohistochemical PD-1, PD-L1, and PD-L2 expression. The cut-off values for high PD-1 (>= 70 cells/high power field [HPF]), PD-L1 (>= 100 cells/HPF), and PD-L2 (>= 100 cells/HPF) were determined by the area under the receiver operating characteristic curve. Expression of PD-1, PD-L1, and PD-L2 was high in 7.9%, 13.2%, and 42.1% patients, respectively. High PD-1, (P = 0.007) and Memorial Sloan Kettering Cancer Center (MSKCC) prognostic scoring (P = 0.019) were independently associated with inferior overall survival on multivariate analysis. High PD-1 also remained an independent prognostic factor for inferior progression-free survival (P = 0.028), as did MSKCC prognostic scoring (P = 0.041) on multivariate analysis. However, there were no differences in survival according to the expression levels of PD-L1/PD-L2 in PCNSL tumor microenvironment. Our results suggest that PD-1 may be considered a biomarker and potential therapeutic target in PCNSL.
C1 [Cho, Hyunsoo; Kim, Soo-Jeong; Kim, Yu Ri; Jang, Ji Eun; Cheong, June-Won; Min, Yoo Hong; Kim, Jin Seok] Yonsei Univ, Coll Med, Severance Hosp, Div Hematol,Dept Internal Med, Seoul 03722, South Korea.
   [Cho, Hyunsoo] Korea Adv Inst Sci & Technol, Grad Sch Med Sci & Engn, Daejeon 34141, South Korea.
   [Kim, Se Hoon; Yang, Woo-Ick] Yonsei Univ, Coll Med, Severance Hosp, Dept Pathol, Seoul 03722, South Korea.
   [Chang, Jong Hee] Yonsei Univ, Coll Med, Severance Hosp, Dept Neurosurg, Seoul 03722, South Korea.
   [Suh, Chang-Ok] Yonsei Univ, Coll Med, Severance Hosp, Dept Radiat Oncol, Seoul 03722, South Korea.
C3 Yonsei University; Yonsei University Health System; Korea Advanced
   Institute of Science & Technology (KAIST); Yonsei University; Yonsei
   University Health System; Yonsei University; Yonsei University Health
   System; Yonsei University; Yonsei University Health System
RP Kim, JS (corresponding author), Yonsei Univ, Coll Med, Severance Hosp, Div Hematol,Dept Internal Med, Seoul 03722, South Korea.
EM
RI Kim, Hyoungil/M-7288-2019; Kim, Tae HWan/ABC-5406-2021; Kim,
   Ji-Soo/D-8744-2012; Cho, Hyunsoo/JLR-6939-2023; Kim,
   Soo-Jeong/F-1024-2017
OI Kim, Hyoungil/0000-0002-6134-4523; Kim, Tae HWan/0000-0002-5585-6522;
   Kim, Ji-Soo/0000-0002-1508-2024; Kim, Se Hoon/0000-0001-7516-7372; Cho,
   Hyunsoo/0000-0003-2651-6403; Jang, Ji Eun/0000-0001-8832-1412; CHEONG,
   JUNE-WON/0000-0002-1744-0921; min, yoo hong/0000-0001-8542-9583; Chang,
   Jong Hee/0000-0003-1509-9800; KIM, JIN SEOK/0000-0001-8986-8436; Kim,
   Soo-Jeong/0000-0001-8859-3573
FU faculty research grant, Yonsei University College of Medicine
   [62015-0029]
FX This work was supported by a faculty research grant, Yonsei University
   College of Medicine, 2015 (62015-0029).
CR Abrey LE, 2006, J CLIN ONCOL, V24, P5711, DOI 10.1200/JCO.2006.08.2941
   Abrey LE, 2005, J CLIN ONCOL, V23, P5034, DOI 10.1200/JCO.2005.13.524
   Ansell SM, 2015, NEW ENGL J MED, V372, P311, DOI 10.1056/NEJMoa1411087
   Berghoff AS, 2015, NEURO-ONCOLOGY, V17, P1064, DOI 10.1093/neuonc/nou307
   Berghoff AS, 2014, CLIN NEUROPATHOL, V33, P42, DOI 10.5414/NP300698
   Braaten KM, 2003, CLIN CANCER RES, V9, P1063
   Chapuy B, 2016, BLOOD, V127, P869, DOI 10.1182/blood-2015-10-673236
   Cho H, 2017, ANN HEMATOL, V96, P1163, DOI 10.1007/s00277-017-3014-x
   Cho H, 2016, BRIT J HAEMATOL, V174, P444, DOI 10.1111/bjh.14069
   Deckert M, 2011, LEUKEMIA, V25, P1797, DOI 10.1038/leu.2011.169
   Ferreri AJM, 2003, J CLIN ONCOL, V21, P266, DOI 10.1200/JCO.2003.09.139
   Four M, 2016, HEMATOL ONCOL, DOI 10.100/hon.2375
   Lopez-Girona A, 2011, BRIT J HAEMATOL, V154, P325, DOI 10.1111/j.1365-2141.2011.08689.x
   Myklebust JH, 2013, BLOOD, V121, P1367, DOI 10.1182/blood-2012-04-421826
   Nayak L, 2017, BLOOD, V129, P3071, DOI 10.1182/blood-2017-01-764209
   Paydas S, 2015, ANN HEMATOL, V94, P1545, DOI 10.1007/s00277-015-2403-2
   Ponzoni M, 2007, BRIT J HAEMATOL, V138, P316, DOI 10.1111/j.1365-2141.2007.06661.x
   Taube JM, 2014, CLIN CANCER RES, V20, P5064, DOI 10.1158/1078-0432.CCR-13-3271
   Topalian SL, 2016, NAT REV CANCER, V16, P275, DOI 10.1038/nrc.2016.36
   Tun HW, 2008, BLOOD, V111, P3200, DOI 10.1182/blood-2007-10-119099
   Wang CC, 2014, BRIT J HAEMATOL, V166, P311, DOI 10.1111/bjh.12938
NR 21
TC 25
Z9 27
U1 1
U2 2
PU IMPACT JOURNALS LLC
PI ORCHARD PARK
PA 6666 E QUAKER ST, STE 1, ORCHARD PARK, NY 14127 USA
EI 1949-2553
J9 ONCOTARGET
JI Oncotarget
PD OCT 20
PY 2017
VL 8
IS 50
BP 87317
EP 87328
DI 10.18632/oncotarget.20264
PG 12
WC Oncology; Cell Biology
WE Science Citation Index Expanded (SCI-EXPANDED)
SC Oncology; Cell Biology
GA FK2VP
UT WOS:000413341400027
PM 29152083
OA Green Submitted, gold, Green Published
DA 2024-06-03
ER

PT J
AU Kerbauy, MN
   Moraes, FY
   Lok, BH
   Ma, J
   Kerbauy, LN
   Spratt, DE
   Santos, FPS
   Perini, GF
   Berlin, A
   Chung, C
   Hamerschlak, N
   Yahalom, J
AF Kerbauy, Mariana N.
   Moraes, Fabio Y.
   Lok, Benjamin H.
   Ma, Jennifer
   Kerbauy, Lucila N.
   Spratt, Daniel E.
   Santos, Fabio P. S.
   Perini, Guilherme F.
   Berlin, Alejandro
   Chung, Caroline
   Hamerschlak, Nelson
   Yahalom, Joachim
TI Challenges and opportunities in primary CNS lymphoma: A systematic
   review
SO RADIOTHERAPY AND ONCOLOGY
LA English
DT Review
DE Non-Hodgkin lymphoma; Lymphoma Central Nervous System; Neoplasms; Drug
   therapy; Stem cell transplantation; Hematology; Radiotherapy
ID CENTRAL-NERVOUS-SYSTEM; HIGH-DOSE METHOTREXATE; STEM-CELL
   TRANSPLANTATION; WHOLE-BRAIN RADIOTHERAPY; RECURRENT PRIMARY CNS;
   MULTICENTER PHASE-II; TERM-FOLLOW-UP; RADIATION-THERAPY;
   ELDERLY-PATIENTS; IMMUNOCOMPETENT PATIENTS
AB Background: Historically, high-dose methotrexate (HD-MTX) plus consolidation chemotherapy and/or whole brain radiotherapy (WBRT) has been the gold standard on Primary Central Nervous System Lymphoma (PCNSL) management. We sought to examine and summarize the data, on clinical trial (CT) setting, investigating multi-modality treatment to PCNSL. Methods: We performed a systematic review of electronic databases (Medline, EMBASE, Cochrane Database and clinicaltrials.gov) and a manual search to identify original PCNSL phase 2 and phase 3 CT from the last 10 years. After a 4 stage Prisma based selection process, 32 published (3 Randomized CT and 29 phases 2 CT) studies ultimately were selected for review. Four ongoing clinical trials found on clinicaltrial.gov were reviewed. Two investigators reviewed titles, abstracts, and articles independently. Two investigators abstracted data sequentially and evaluated each study independently. Findings: Treatment of PCNSL requires a multidisciplinary approach. HD-MTX represents the most accepted standard of care induction therapy for newly diagnosed PCNSL. When HD-MTX is given with WBRT for consolidation delayed neurotoxicity can be an important complication, particularly in elderly patients. Studies have suggested that WBRT may be deferred until relapse without compromising survival and deferring WBRT may be the best approach in elderly patients. Results from dose-reduced WBRT and consolidative HD-Ara-C are encouraging. High-dose chemotherapy in combination with autologous stem cell transplantation (HDC-ASCT) as chemotherapy alone has emerged as an important consolidative treatment for selected population. The optimal salvage therapy is still to be defined. Conclusion: WBRT for consolidation is a well-studied modality; however emerging options to selected population such as HDC-ASCf, dose-reduced WBRT or chemotherapy alone are associated with similar survival outcome and less neurotoxicity in selected series. Ongoing and future clinical trials will better define the best approach on this rare disease. (C) 2017 Elsevier B.V. All rights reserved. Radiotherapy and Oncology 122 (2017) 352-361
C1 [Kerbauy, Mariana N.; Kerbauy, Lucila N.; Santos, Fabio P. S.; Perini, Guilherme F.; Hamerschlak, Nelson] Hosp Israelita Albert Einstein, Dept Hematol, Sao Paulo, Brazil.
   [Moraes, Fabio Y.] Hosp Sirio Libanes, Dept Radiat Oncol, Sao Paulo, Brazil.
   [Moraes, Fabio Y.; Berlin, Alejandro] Univ Toronto, Dept Radiat Oncol, Toronto, ON, Canada.
   [Moraes, Fabio Y.; Berlin, Alejandro] Univ Hlth Network, Radiat Med Program, Princess Margaret Canc Ctr, OPG Bldg-7-215-700, Toronto, ON M5G 1Z5, Canada.
   [Lok, Benjamin H.; Ma, Jennifer; Yahalom, Joachim] Mem Sloan Kettering Canc Ctr, Dept Radiat Oncol, New York, NY USA.
   [Spratt, Daniel E.] Univ Michigan, Dept Radiat Oncol, Med Ctr, Ann Arbor, MI USA.
   [Chung, Caroline] Univ Texas MD Anderson Canc Ctr, Dept Radiat Oncol, Houston, TX USA.
C3 Hospital Israelita Albert Einstein; Hospital Sirio Libanes; University
   of Toronto; University of Toronto; University Health Network Toronto;
   Princess Margaret Cancer Centre; Memorial Sloan Kettering Cancer Center;
   University of Michigan System; University of Michigan; University of
   Texas System; UTMD Anderson Cancer Center
RP Moraes, FY (corresponding author), Univ Hlth Network, Radiat Med Program, Princess Margaret Canc Ctr, OPG Bldg-7-215-700, Toronto, ON M5G 1Z5, Canada.
EM
RI Kerbauy, Mariana/AAW-6720-2020; Lok, Benjamin/AAC-9212-2019; Spratt,
   Daniel/AAM-4612-2020; Moraes, Fabio Ynoe/AAE-9645-2019; Hamerschlak,
   Nelson/M-8060-2015; Chung, Caroline/AAM-5504-2021
OI Kerbauy, Mariana/0000-0002-4441-9212; Lok, Benjamin/0000-0002-3675-8847;
   Berlin, Alejandro/0000-0002-1880-6905; Ma, Jennifer/0000-0001-8133-2282;
   Chung, Caroline/0000-0002-9662-1519; Spratt, Daniel/0000-0002-5973-4741;
   Ynoe Moraes, Fabio/0000-0003-3407-9215
CR Abrey LE, 2006, J CLIN ONCOL, V24, P5711, DOI 10.1200/JCO.2006.08.2941
   Abrey LE, 2003, J CLIN ONCOL, V21, P4151, DOI 10.1200/JCO.2003.05.024
   Abrey LE, 2005, J CLIN ONCOL, V23, P5034, DOI 10.1200/JCO.2005.13.524
   Abrey LE, 2000, J CLIN ONCOL, V18, P3144, DOI 10.1200/JCO.2000.18.17.3144
   Alimohamed N, 2012, LEUKEMIA LYMPHOMA, V53, P862, DOI 10.3109/10428194.2011.633250
   [Anonymous], 2008, WHO Classification of Tumors of Haematopoietic and Lymphoid Tissues
   [Anonymous], HAEMATOL ONCOL
   [Anonymous], J CLIN ONCOL
   [Anonymous], ASCO M
   [Anonymous], BLOOD
   Bataille B, 2000, J NEUROSURG, V92, P261, DOI 10.3171/jns.2000.92.2.0261
   Batchelor T, 2003, J CLIN ONCOL, V21, P1044, DOI 10.1200/JCO.2003.03.036
   Bessell EM, 2001, INT J RADIAT ONCOL, V50, P457, DOI 10.1016/S0360-3016(01)01451-1
   Braaten KM, 2003, CLIN CANCER RES, V9, P1063
   Camilleri-Broet S, 1998, AM J CLIN PATHOL, V110, P607, DOI 10.1093/ajcp/110.5.607
   Camilleri-Broët S, 2006, BLOOD, V107, P190, DOI 10.1182/blood-2005-03-1024
   Chen CC, 2010, RADIOSURG, V7, P276, DOI 10.1159/000288739
   Chen YB, 2015, CANCER-AM CANCER SOC, V121, P226, DOI 10.1002/cncr.29023
   Cheng AL, 1998, CANCER, V82, P1946, DOI 10.1002/(SICI)1097-0142(19980515)82:10<1946::AID-CNCR19>3.0.CO;2-T
   Cheng G, 2003, BONE MARROW TRANSPL, V31, P679, DOI 10.1038/sj.bmt.1703917
   Cher L, 1996, NEUROLOGY, V46, P1757, DOI 10.1212/WNL.46.6.1757
   Colombat P, 2006, BONE MARROW TRANSPL, V38, P417, DOI 10.1038/sj.bmt.1705452
   Correa DD, 2007, ANN ONCOL, V18, P1145, DOI 10.1093/annonc/mdl464
   Courts C, 2007, J NEUROPATH EXP NEUR, V66, P230, DOI 10.1097/01.jnen.0000248553.45456.96
   DeAngelis LM, 2002, J CLIN ONCOL, V20, P4643, DOI 10.1200/JCO.2002.11.013
   Ekenel M, 2008, CANCER-AM CANCER SOC, V113, P1025, DOI 10.1002/cncr.23670
   Ferreri AJM, 2003, J CLIN ONCOL, V21, P266, DOI 10.1200/JCO.2003.09.139
   Ferreri AJM, 2002, NEUROLOGY, V58, P1513, DOI 10.1212/WNL.58.10.1513
   Ferreri AJM, 2009, LANCET, V374, P1512, DOI 10.1016/S0140-6736(09)61416-1
   Fisher B, 2005, J NEURO-ONCOL, V74, P201, DOI 10.1007/s11060-004-6596-9
   Flanagan EP, 2011, NEUROLOGY, V77, P784, DOI 10.1212/WNL.0b013e31822b00b9
   Freilich RJ, 1996, NEUROLOGY, V46, P435, DOI 10.1212/WNL.46.2.435
   Fritsch K, 2011, ANN ONCOL, V22, P2080, DOI 10.1093/annonc/mdq712
   Fukumura K, 2016, ACTA NEUROPATHOL, V131, P865, DOI 10.1007/s00401-016-1536-2
   Gavrilovic IT, 2006, J CLIN ONCOL, V24, P4570, DOI 10.1200/JCO.2006.06.6910
   Guha-Thakurta N, 1999, J NEURO-ONCOL, V43, P259, DOI 10.1023/A:1006210703827
   Herrlinger U, 2002, ANN NEUROL, V51, P247, DOI 10.1002/ana.10102
   Hoang-Xuan K, 2003, J CLIN ONCOL, V21, P2726, DOI 10.1200/JCO.2003.11.036
   Hoang-Xuan K, 2015, LANCET ONCOL, V16, pE322, DOI 10.1016/S1470-2045(15)00076-5
   Hottinger AF, 2007, NEUROLOGY, V69, P1178, DOI 10.1212/01.wnl.0000276986.19602.c1
   Illerhaus G, 2009, ANN ONCOL, V20, P319, DOI 10.1093/annonc/mdn628
   Illerhaus G, 2007, HAEMATOLOGICA, V93, P147, DOI 10.3324/haematol.11771
   Illerhaus G, 2006, J CLIN ONCOL, V24, P3865, DOI 10.1200/JCO.2006.06.2117
   Jahnke K, 2005, ANN ONCOL, V16, P445, DOI 10.1093/annonc/mdi075
   Joerger M, 2012, LEUKEMIA LYMPHOMA, V53, P1867, DOI 10.3109/10428194.2012.676177
   Josephson SA, 2007, J NEUROSURG, V106, P72, DOI 10.3171/jns.2007.106.1.72
   Kasenda B, 2015, ANN ONCOL, V26, P1305, DOI 10.1093/annonc/mdv076
   Khan RB, 2002, J NEURO-ONCOL, V58, P175, DOI 10.1023/A:1016077907952
   Kiefer T, 2012, ANN ONCOL, V23, P1809, DOI 10.1093/annonc/mdr553
   Korfel A, 2013, NAT REV NEUROL, V9, P317, DOI 10.1038/nrneurol.2013.83
   Mappa S, 2013, HEMATOL ONCOL, V31, P143, DOI 10.1002/hon.2037
   Milgrom SA, 2015, LEUKEMIA LYMPHOMA, V56, P1197, DOI 10.3109/10428194.2014.961014
   Moher D, 2009, BMJ-BRIT MED J, V339, DOI [10.1136/bmj.b2535, 10.1186/2046-4053-4-1, 10.1136/bmj.b2700, 10.1016/j.ijsu.2010.07.299, 10.1371/journal.pmed.1000097, 10.1016/j.ijsu.2010.02.007, 10.1136/bmj.i4086]
   Montemurro M, 2007, ANN ONCOL, V18, P665, DOI 10.1093/annonc/mdl458
   Montesinos-Rongen M, 2008, LEUKEMIA, V22, P400, DOI 10.1038/sj.leu.2405019
   Montesinos-Rongen M, 2009, BLOOD, V113, P7, DOI 10.1182/blood-2008-04-149005
   Nabors LB, 2014, J NATL COMPR CANC NE, V12, P1517, DOI 10.6004/jnccn.2014.0151
   NELSON DF, 1992, INT J RADIAT ONCOL, V23, P9, DOI 10.1016/0360-3016(92)90538-S
   Nguyen PL, 2005, J CLIN ONCOL, V23, P1507, DOI 10.1200/JCO.2005.01.161
   O'Neill BP, 2013, AM J HEMATOL, V88, P997, DOI 10.1002/ajh.23551
   Omuro A, 2015, LANCET HAEMATOL, V2, pE251, DOI 10.1016/S2352-3026(15)00074-5
   Omuro A, 2015, BLOOD, V125, P1403, DOI 10.1182/blood-2014-10-604561
   Pels H, 2003, J CLIN ONCOL, V21, P4489, DOI 10.1200/JCO.2003.04.056
   Pels H, 2009, J NEURO-ONCOL, V91, P299, DOI 10.1007/s11060-008-9712-4
   Perini GF, 2013, ARQ NEURO-PSIQUIAT, V71, P254, DOI 10.1590/0004-282X20130011
   Poortmans PMP, 2003, J CLIN ONCOL, V21, P4483, DOI 10.1200/JCO.2003.03.108
   Pulczynski EJ, 2015, HAEMATOLOGICA, V100, P534, DOI 10.3324/haematol.2014.108472
   Roth P, 2014, CURR OPIN NEUROL, V27, P697, DOI 10.1097/WCO.0000000000000145
   Rubenstein JL, 2013, J CLIN ONCOL, V31, P3061, DOI 10.1200/JCO.2012.46.9957
   Sakamoto M, 2006, J NEURO-ONCOL, V77, P53, DOI 10.1007/s11060-005-7698-8
   Sandor V, 1998, J CLIN ONCOL, V16, P3000, DOI 10.1200/JCO.1998.16.9.3000
   Schabet M, 1999, J NEURO-ONCOL, V43, P199, DOI 10.1023/A:1006290032052
   Schorb E, 2013, HAEMATOLOGICA, V98, P765, DOI 10.3324/haematol.2012.076075
   Schultz C, 1996, J CLIN ONCOL, V14, P556, DOI 10.1200/JCO.1996.14.2.556
   Shah GD, 2007, J CLIN ONCOL, V25, P4730, DOI 10.1200/JCO.2007.12.5062
   Shenkier TN, 2005, J CLIN ONCOL, V23, P2233, DOI 10.1200/JCO.2005.07.109
   Soussain C, 2001, J CLIN ONCOL, V19, P742, DOI 10.1200/JCO.2001.19.3.742
   Soussain C, 2008, J CLIN ONCOL, V26, P2512, DOI 10.1200/JCO.2007.13.5533
   Thiel E, 2010, LANCET ONCOL, V11, P1036, DOI 10.1016/S1470-2045(10)70229-1
   Villano JL, 2011, BRIT J CANCER, V105, P1414, DOI 10.1038/bjc.2011.357
   Wieduwilt MJ, 2012, CLIN CANCER RES, V18, P1146, DOI 10.1158/1078-0432.CCR-11-0625
   Yahalom J, 2015, INT J RADIAT ONCOL, V92, P11, DOI 10.1016/j.ijrobp.2015.01.009
NR 82
TC 25
Z9 28
U1 0
U2 12
PU ELSEVIER IRELAND LTD
PI CLARE
PA ELSEVIER HOUSE, BROOKVALE PLAZA, EAST PARK SHANNON, CO, CLARE, 00000,
   IRELAND
SN 0167-8140
EI 1879-0887
J9 RADIOTHER ONCOL
JI Radiother. Oncol.
PD MAR
PY 2017
VL 122
IS 3
BP 352
EP 361
DI 10.1016/j.radonc.2016.12.033
PG 10
WC Oncology; Radiology, Nuclear Medicine & Medical Imaging
WE Science Citation Index Expanded (SCI-EXPANDED)
SC Oncology; Radiology, Nuclear Medicine & Medical Imaging
GA EP9KC
UT WOS:000397691500005
PM 28104300
DA 2024-06-03
ER

PT J
AU Enblad, G
   Martinsson, G
   Baecklund, E
   Hesselager, G
   Sundström, C
   Amini, RM
   Hagberg, H
AF Enblad, Gunilla
   Martinsson, Gustaf
   Baecklund, Eva
   Hesselager, Goran
   Sundstrom, Christer
   Amini, Rosie-Marie
   Hagberg, Hans
TI Population-based experience on primary central nervous system lymphoma
   2000-2012: the incidence is increasing
SO ACTA ONCOLOGICA
LA English
DT Article
ID PRIMARY CNS LYMPHOMA; HIGH-DOSE METHOTREXATE; INFLAMMATORY DISORDERS;
   SINGLE-INSTITUTION; CHEMOTHERAPY; TEMOZOLOMIDE; AUTOIMMUNE; SURVIVAL;
   TRENDS
AB Background: Primary central nervous system lymphomas (PCNSL) are rare lymphomas with a poor prognosis. Recently, an increased incidence has been reported. The present study is a population-based study of all patients with PCNSL in the Uppsala/orebro region of middle Sweden.Patients and methods: All patients diagnosed with a PCNSL at Uppsala University Hospital 2000-2012 were identified. Altogether, 96 patients (50 women and 46 men) were included. The median age at diagnosis was 66 years (17-95).Results: There was a statistically significant increase in age-standardized incidence during the study period, 30 patients were diagnosed in the first half and 66 in the second half of the period. No patient had an HIV-infection. Two patients had undergone kidney transplantation and were treated with immunosuppressive drugs. A high proportion of the patients, 29%, had a history of an autoimmune or inflammatory disease. The prognosis was poor with a median survival of only four months. In the 70 (73%) patients treated with curative intention the median survival was 12 months. Patients treated with high-dose methotrexate, radiotherapy and/or temozolomide appeared to have a better survival. There was no improvement in survival during the study period or after the introduction of rituximab. There also was no difference in any of the analyzed variables that could explain the increased incidence.Conclusion: In this population-based study we could confirm the previously described increased incidence of PCNSL. The prognosis remains poor despite the inclusion of treatment with rituximab during the study period. A high proportion of the patients had a history of an autoimmune or inflammatory disease not previously described but there was no increase during the study period.
C1 [Enblad, Gunilla; Martinsson, Gustaf; Hagberg, Hans] Uppsala Univ, Dept Immunol Genet & Pathol, Expt & Clin Oncol, S-75185 Uppsala, Sweden.
   [Baecklund, Eva] Uppsala Univ, Dept Med Sci, Uppsala, Sweden.
   [Hesselager, Goran] Uppsala Univ, Dept Neurosci, Uppsala, Sweden.
   [Sundstrom, Christer; Amini, Rosie-Marie] Uppsala Univ, Dept Immunol Genet & Pathol, Uppsala, Sweden.
C3 Uppsala University; Uppsala University; Uppsala University; Uppsala
   University
RP Enblad, G (corresponding author), Uppsala Univ, Dept Immunol Genet & Pathol, Expt & Clin Oncol, S-75185 Uppsala, Sweden.
EM
FU Swedish Cancer Foundation; Lions research foundation
FX This work was supported by The Swedish Cancer Foundation and the Lions
   research foundation.
CR Abrey LE, 2006, J CLIN ONCOL, V24, P5711, DOI 10.1200/JCO.2006.08.2941
   Abrey LE, 2000, J CLIN ONCOL, V18, P3144, DOI 10.1200/JCO.2000.18.17.3144
   Aoki H, 2013, NEUROL MED-CHIR, V53, P797, DOI 10.2176/nmc.oa2013-0195
   Baecklund E, 2014, SEMIN CANCER BIOL, V24, P61, DOI 10.1016/j.semcancer.2013.12.001
   Bhat Neelima M, 2004, Hum Antibodies, V13, P63
   Birnbaum T, 2012, J NEURO-ONCOL, V109, P285, DOI 10.1007/s11060-012-0891-7
   Booman M, 2008, J PATHOL, V216, P209, DOI 10.1002/path.2399
   Camilleri-Broët S, 2006, BLOOD, V107, P190, DOI 10.1182/blood-2005-03-1024
   Campo E, 2011, BLOOD, V117, P5019, DOI 10.1182/blood-2011-01-293050
   DEANGELIS LM, 1990, NEUROLOGY, V40, P80, DOI 10.1212/WNL.40.1.80
   Doucet S, 2013, CURR TREAT OPTION ON, V14, P185, DOI 10.1007/s11864-013-0227-7
   Hellgren K, 2010, ANN RHEUM DIS, V69, P654, DOI 10.1136/ard.2008.096925
   Holdhoff M, 2014, NEUROLOGY, V83, P235, DOI 10.1212/WNL.0000000000000593
   Lu JQ, 2016, WORLD NEUROSURG, V88, DOI 10.1016/j.wneu.2015.11.099
   Mocikova H, 2016, LEUKEMIA LYMPHOMA, V57, P2777, DOI 10.3109/10428194.2016.1167203
   Montesinos-Rongen M, 2014, J NEUROPATH EXP NEUR, V73, P1116, DOI 10.1097/NEN.0000000000000133
   O'Neill BP, 2013, AM J HEMATOL, V88, P997, DOI 10.1002/ajh.23551
   Phillips EH, 2014, CURR HEMATOL MALIG R, V9, P243, DOI 10.1007/s11899-014-0217-2
   Pulczynski EJ, 2015, HAEMATOLOGICA, V100, P534, DOI 10.3324/haematol.2014.108472
   Sagoo MS, 2014, SURV OPHTHALMOL, V59, P503, DOI 10.1016/j.survophthal.2013.12.001
   Schorb E, 2016, BMC CANCER, V16, DOI 10.1186/s12885-016-2311-4
   Shiels MS, 2016, BRIT J HAEMATOL, V174, P417, DOI 10.1111/bjh.14073
   Smedby KE, 2008, J INTERN MED, V264, P514, DOI 10.1111/j.1365-2796.2008.02029.x
   Vanderpump MPJ, 2011, BRIT MED BULL, V99, P39, DOI 10.1093/bmb/ldr030
   Villano JL, 2011, BRIT J CANCER, V105, P1414, DOI 10.1038/bjc.2011.357
   Wang XX, 2014, LEUKEMIA LYMPHOMA, V55, P2497, DOI 10.3109/10428194.2014.889823
NR 26
TC 25
Z9 27
U1 0
U2 3
PU TAYLOR & FRANCIS LTD
PI ABINGDON
PA 2-4 PARK SQUARE, MILTON PARK, ABINGDON OR14 4RN, OXON, ENGLAND
SN 0284-186X
EI 1651-226X
J9 ACTA ONCOL
JI Acta Oncol.
PY 2017
VL 56
IS 4
BP 599
EP 607
DI 10.1080/0284186X.2016.1270465
PG 9
WC Oncology
WE Science Citation Index Expanded (SCI-EXPANDED)
SC Oncology
GA ES4JD
UT WOS:000399499600014
PM 28084866
OA Bronze
DA 2024-06-03
ER

PT J
AU Tarabay, A
   Cossu, G
   Berhouma, M
   Levivier, M
   Daniel, RT
   Messerer, M
AF Tarabay, A.
   Cossu, G.
   Berhouma, M.
   Levivier, M.
   Daniel, R. T.
   Messerer, M.
TI Primary pituitary lymphoma: an update of the literature
SO JOURNAL OF NEURO-ONCOLOGY
LA English
DT Review
DE Primary pituitary lymphoma; Primary central nervous system lymphoma;
   Sellar mass; Endoscopic endonasal surgery; Trans-sphenoidal surgery;
   Radiotherapy; Chemotherapy
ID PRIMARY CNS LYMPHOMA; NERVOUS-SYSTEM LYMPHOMA; HIGH-DOSE METHOTREXATE;
   WHOLE-BRAIN RADIOTHERAPY; PRIMARY SELLAR LYMPHOMA; B-CELL LYMPHOMA;
   STEREOTACTIC RADIOSURGERY; MALIGNANT-LYMPHOMA; 1ST-LINE TREATMENT;
   SURGERY
AB Primary intracranial lymphomas (Weller et al. in Neuro Oncol 14(12):1481-1484, 2012) are an emerging disease and an isolated localization in the pituitary gland i.e. primary pituitary lymphoma (PPL) represents a rare condition. We present an update of the most recent evidence for PPL through a systematic review of the literature. A systematic literature review was conducted using PubMed database up to October 2015. The population was defined as immunocompetent patients with a pathologically confirmed diagnosis of PPL. Patients' characteristics, clinical presentation, radiological features, pathology reports, adjuvant treatment and follow-up data were analyzed. We reported one case of PPL and included our data in this analysis. A total of 33 cases of PPL were identified, including ours. A slight not significant female prevalence was evident, with a mean age of 59 years at diagnosis. Visual troubles and headaches were the most common presenting symptoms. About 80 % of patients presented a cranial nerve (CN) deficit. The most frequently involved were the II and III CN. Anterior hypopituitarism was present in 70 % of cases and a diabetes insipidus in 36 % of cases. PPL was rarely limited to the sella and most often extended to the suprasellar and parasellar space. 70 % of cases underwent resection, 21 % a biopsy. A B-cell lymphoma was isolated in 82 % of cases, a T-cell lymphoma in 15 % and a NK/T cell lymphoma in one case. Overall mean survival rate was 14.4 months (95 % confidence interval 9.0-19.8 months) and there was no difference in terms of survival rates when patients were stratified according to the treatment they received. PPL is an emerging clinical entity. Literature data are too scarce to allow the definition of specific protocols of treatment and the management is based on the guidelines present for PCNSL. The role of surgery aiming at a complete resection of PPL should be reevaluated in wider studies including only this category of patients, to establish the real role of each therapeutic strategy.
C1 [Tarabay, A.; Cossu, G.; Levivier, M.; Daniel, R. T.; Messerer, M.] Univ Lausanne Hosp, Dept Neurosurg, Rue Bugnon 46, CH-1010 Lausanne, Switzerland.
   [Berhouma, M.] Hosp Civils Lyon, Pierre Wertheimer Neurol & Neurosurg Hosp, Dept Neurosurg B, Skull Base Surg Unit, Lyon, France.
C3 University of Lausanne; Centre Hospitalier Universitaire Vaudois (CHUV);
   CHU Lyon
RP Messerer, M (corresponding author), Univ Lausanne Hosp, Dept Neurosurg, Rue Bugnon 46, CH-1010 Lausanne, Switzerland.
EM
RI Berhouma, Moncef/AAF-4058-2020; Messerer, Mahmoud/AAZ-3527-2020; Cossu,
   Giulia/ABF-6396-2020
OI Levivier, Marc/0000-0002-6273-2595; Cossu, Giulia/0000-0003-0913-8965;
   Messerer, Mahmoud/0000-0003-4975-8237
CR Anila KR, 2012, INDIAN J PATHOL MICR, V55, P549, DOI 10.4103/0377-4929.107811
   Au WY, 2000, AM J HEMATOL, V63, P231, DOI 10.1002/(SICI)1096-8652(200004)63:4<231::AID-AJH14>3.0.CO;2-Z
   Bataille B, 2000, J NEUROSURG, V92, P261, DOI 10.3171/jns.2000.92.2.0261
   Bayraktar S, 2010, ACTA HAEMATOL-BASEL, V123, P121, DOI 10.1159/000272920
   Bellinzona M, 2005, EJSO, V31, P100, DOI 10.1016/j.ejso.2004.10.002
   Carrasco CA, 2012, PITUITARY, V15, P93, DOI 10.1007/s11102-010-0219-6
   Chen SM, 2008, J CLIN NEUROSCI, V15, P1148, DOI 10.1016/j.jocn.2007.08.021
   Clayton RN, 1999, BEST PRACT RES CL EN, V13, P451, DOI 10.1053/beem.1999.0036
   EBY NL, 1988, CANCER-AM CANCER SOC, V62, P2461, DOI 10.1002/1097-0142(19881201)62:11<2461::AID-CNCR2820621135>3.0.CO;2-M
   Famini P, 2011, J CLIN ENDOCR METAB, V96, P1633, DOI 10.1210/jc.2011-0168
   FINE HA, 1993, ANN INTERN MED, V119, P1093, DOI 10.7326/0003-4819-119-11-199312010-00007
   Freda PU, 1999, ENDOCRIN METAB CLIN, V28, P81, DOI 10.1016/S0889-8529(05)70058-X
   Gerstner ER, 2008, NEUROLOGY, V70, P401, DOI 10.1212/01.wnl.0000300671.37279.0e
   Giustina A, 2001, J CLIN ENDOCR METAB, V86, P4567, DOI 10.1210/jc.86.10.4567
   Hayasaka K, 2010, CLIN NUCL MED, V35, P205, DOI 10.1097/RLU.0b013e3181cc64a2
   Martinez JH, 2011, CASE REP ENDOCRINOL, V2011, DOI 10.1155/2011/941738
   Hirono S, 2015, J NEURO-ONCOL, V123, P237, DOI 10.1007/s11060-015-1786-1
   Huang YY, 2005, ENDOCR J, V52, P543, DOI 10.1507/endocrj.52.543
   Jahnke K, 2006, J NEURO-ONCOL, V80, P159, DOI 10.1007/s11060-006-9165-6
   Kasenda B, 2015, ANN ONCOL, V26, P1305, DOI 10.1093/annonc/mdv076
   Katz Bradley J, 2003, Endocr Pract, V9, P296
   Kaufmann TJ, 2002, AM J NEURORADIOL, V23, P364
   Kenai H, 2006, J NEUROSURG, V105, P133, DOI 10.3171/sup.2006.105.7.133
   Korfel A, 2015, NEUROLOGY, V84, P1242, DOI 10.1212/WNL.0000000000001395
   Kozakova D, 2008, Cas Lek Cesk, V147, P569
   Kuhn D, 1999, ACTA NEUROPATHOL, V97, P311, DOI 10.1007/s004010050990
   Kumar R, 2015, WORLD NEUROSURG, V84, P390, DOI 10.1016/j.wneu.2015.03.062
   Lai R, 2002, NEUROLOGY, V59, P1557, DOI 10.1212/01.WNL.0000034256.20173.EA
   Landman RE, 2001, J CLIN ENDOCR METAB, V86, P1470, DOI 10.1210/jc.86.4.1470
   Lee JH, 2002, AM J NEURORADIOL, V23, P838
   Li YX, 2012, J NEUROL, V259, P297, DOI 10.1007/s00415-011-6179-6
   Liu JK, 2007, J NEUROSURG, V107, P660, DOI 10.3171/JNS-07/09/0660
   Mathiasen R A, 2000, Pituitary, V2, P283, DOI 10.1023/A:1009969417380
   Messerer M, 2011, NEUROSURG FOCUS, V30, DOI 10.3171/2011.1.FOCUS10308
   Morris PG, 2013, J CLIN ONCOL, V31, P3971, DOI 10.1200/JCO.2013.50.4910
   Moshkin O, 2009, ENDOCR PATHOL, V20, P46, DOI 10.1007/s12022-009-9062-6
   ONEAL KD, 1991, MOL CELL ENDOCRINOL, V82, P127, DOI 10.1016/0303-7207(91)90023-L
   Papanastasiou L, 2012, HORM-INT J ENDOCRINO, V11, P488, DOI 10.14310/horm.2002.1382
   Popovic V, 1998, CLIN ENDOCRINOL, V49, P441, DOI 10.1046/j.1365-2265.1998.00536.x
   Rainsbury Pg, 2012, JRSM Short Rep, V3, P55, DOI 10.1258/shorts.2012.012067
   Romeike BFM, 2008, ACTA NEUROCHIR, V150, P833, DOI 10.1007/s00701-008-1585-y
   Rudnik A, 2007, POL ACAD SCI, V45, P144
   Samaratunga H, 1997, ENDOCR PATHOL, V8, P335, DOI 10.1007/BF02739936
   Shaw JAM, 1997, J ROY SOC MED, V90, P274, DOI 10.1177/014107689709000512
   Singh V P, 1993, Indian J Cancer, V30, P88
   SNIDER WD, 1983, NEW ENGL J MED, V308, P45
   Tamer Gonca, 2009, J Med Case Rep, V3, P9293, DOI 10.1186/1752-1947-3-9293
   Thiel E, 2010, LANCET ONCOL, V11, P1036, DOI 10.1016/S1470-2045(10)70229-1
   Weller M, 2012, NEURO-ONCOLOGY, V14, P1481, DOI 10.1093/neuonc/nos159
   Wiens AL, 2012, NEUROPATHOLOGY, V32, P668, DOI 10.1111/j.1440-1789.2012.01314.x
   Wilkie MD, 2012, BRIT J NEUROSURG, V26, P782, DOI 10.3109/02688697.2012.674578
   Wolfe SQ, 2009, PITUITARY, V12, P76, DOI 10.1007/s11102-008-0084-8
   Yang SH, 2009, J NEURO-ONCOL, V92, P65, DOI 10.1007/s11060-008-9736-9
NR 53
TC 25
Z9 26
U1 0
U2 9
PU SPRINGER
PI NEW YORK
PA 233 SPRING ST, NEW YORK, NY 10013 USA
SN 0167-594X
EI 1573-7373
J9 J NEURO-ONCOL
JI J. Neuro-Oncol.
PD DEC
PY 2016
VL 130
IS 3
BP 383
EP 395
DI 10.1007/s11060-016-2249-z
PG 13
WC Oncology; Clinical Neurology
WE Science Citation Index Expanded (SCI-EXPANDED)
SC Oncology; Neurosciences & Neurology
GA ED6FE
UT WOS:000388950700001
PM 27581598
DA 2024-06-03
ER

PT J
AU Huang, WY
   Wen, JB
   Wu, G
   Yin, B
   Li, JJ
   Geng, DY
AF Huang, W. -Y.
   Wen, J. -B.
   Wu, G.
   Yin, B.
   Li, J. -J.
   Geng, D. -Y.
TI Diffusion-Weighted Imaging for Predicting and Monitoring Primary Central
   Nervous System Lymphoma Treatment Response
SO AMERICAN JOURNAL OF NEURORADIOLOGY
LA English
DT Article
ID PRIMARY CNS LYMPHOMA; THERAPEUTIC RESPONSE; GRADE ASTROCYTOMAS;
   BREAST-CANCER; BRAIN-TUMORS; CONSOLIDATION; METHOTREXATE; COEFFICIENT;
   RITUXIMAB; BIOMARKER
AB BACKGROUND AND PURPOSE: Whether ADC value predicts the therapy response and outcomes of primary central system lymphoma remains controversial. This study assessed the minimum ADC correlated with treatment response in patients with primary central nervous system lymphoma undergoing methotrexate-based chemotherapy.
   MATERIALS AND METHODS: Thirty-five patients with primary central nervous system lymphoma underwent conventional MR imaging and DWI before chemotherapy and after 1 and 5 cycles of chemotherapy. Treatment response was determined according to the International PCNSL Collaborative Group criteria and was classified as a complete response, partial response, or progressive disease. Pretreatment minimum ADC, minimum ADC after 1 cycle, minimum ADC after 5 cycles, and change in minimum ADC were compared among the different response groups. The Pearson correlation test was calculated between these ADC parameters and tumor response.
   RESULTS: The pretreatment minimum ADC of the progressive disease group was lower than that of the complete response and partial response groups, but there was no significant difference among them. The minimum ADC after 1 cycle and minimum ADC after 5 cycles were statistically significantly higher than the pretreatment minimum ADC. A comparison among groups showed that minimum ADC after 1 cycle, minimum ADC after 5 cycles, minimum ADC change, and the percentage of minimum ADC change were all significantly different among the 3 groups. A significant positive correlation was observed between the percentage of minimum ADC after 1 cycle of chemotherapy and the size reduction percentage after 5 cycles of chemotherapy. The minimum ADC change and the percentage of minimum ADC change performed better in the differentiation of the final treatment response, specifically in complete response and partial response from progressive disease.
   CONCLUSIONS: The minimum ADC after 1 cycle and minimum ADC changes were better correlated with the treatment response than the pretreatment minimum ADC. Minimum ADC after early therapy may potentially to be used to predict and monitor the response of primary central nervous system lymphoma to chemotherapy.
C1 [Huang, W. -Y.; Li, J. -J.] Hainan Gen Hosp, Dept Radiol, Haikou, Hainan, Peoples R China.
   [Wu, G.] Hainan Gen Hosp, Dept Radiotherapy, Haikou, Hainan, Peoples R China.
   [Wen, J. -B.; Yin, B.; Geng, D. -Y.] Fudan Univ, Huashan Hosp, Dept Radiol, 12 Wulumuqi Rd, Shanghai, Peoples R China.
C3 Fudan University
RP Geng, DY (corresponding author), Fudan Univ, Huashan Hosp, Dept Radiol, 12 Wulumuqi Rd, Shanghai, Peoples R China.; Li, JJ (corresponding author), Hainan Gen Hosp, Dept Radiol, 19 Xiuhua Rd, Haikou, Hainan, Peoples R China.
EM
RI Cao, Xin/AAX-8367-2021
FU National Key Discipline Project; Hainan Provincial Key Foundation
   Project [ZDXM2015066]; National Natural Science Foundation of China
   [81471627]
FX This research was supported by funding from the National Key Discipline
   Project, the Hainan Provincial Key Foundation Project (ZDXM2015066), and
   the National Natural Science Foundation of China under grant No.
   81471627.
CR Abrey LE, 2005, J CLIN ONCOL, V23, P5034, DOI 10.1200/JCO.2005.13.524
   Africa Emilio, 2005, Rays, V30, P221
   Barajas RF, 2010, AM J NEURORADIOL, V31, P60, DOI 10.3174/ajnr.A1750
   Bhagavathi S, 2008, ARCH PATHOL LAB MED, V132, P1830, DOI 10.1043/1543-2165-132.11.1830
   Chenevert TL, 1997, CLIN CANCER RES, V3, P1457
   Chenevert TL, 2000, JNCI-J NATL CANCER I, V92, P2029, DOI 10.1093/jnci/92.24.2029
   Fujimoto H, 2014, BREAST CANCER-TOKYO, V21, P724, DOI 10.1007/s12282-013-0449-3
   Guo AC, 2002, RADIOLOGY, V224, P177, DOI 10.1148/radiol.2241010637
   Iwadate Y, 2014, J NEURO-ONCOL, V117, P261, DOI 10.1007/s11060-014-1379-4
   Kansara R, 2015, AM J HEMATOL, V90, P1149, DOI 10.1002/ajh.24204
   Lee EJ, 2008, AM J NEURORADIOL, V29, P1872, DOI 10.3174/ajnr.A1254
   Lee KC, 2007, CLIN CANCER RES, V13, P443, DOI 10.1158/1078-0432.CCR-06-1888
   Liu Y, 2009, CLIN RADIOL, V64, P1067, DOI 10.1016/j.crad.2009.07.010
   Moffat BA, 2005, P NATL ACAD SCI USA, V102, P5524, DOI 10.1073/pnas.0501532102
   Moffat BA, 2004, MAGN RESON MATER PHY, V17, P249, DOI 10.1007/s10334-004-0079-z
   Morris PG, 2013, J CLIN ONCOL, V31, P3971, DOI 10.1200/JCO.2013.50.4910
   Nakajima S, 2015, CLIN RADIOL, V70, P1393, DOI 10.1016/j.crad.2015.08.004
   PDQAdult Treatment Editorial Board, 2015, PRIMARYCNS LYMPH TRE
   Prasad SR, 2002, RADIOLOGY, V225, P416, DOI 10.1148/radiol.2252011604
   Server A, 2009, ACTA RADIOL, V50, P682, DOI 10.1080/02841850902933123
   Valles FE, 2013, AM J NEURORADIOL, V34, P35, DOI 10.3174/ajnr.A3165
   Wieduwilt MJ, 2012, CLIN CANCER RES, V18, P1146, DOI 10.1158/1078-0432.CCR-11-0625
   Wu XC, 2011, NMR BIOMED, V24, P1181, DOI 10.1002/nbm.1689
   Zacharia TT, 2008, J NEUROIMAGING, V18, P411, DOI 10.1111/j.1552-6569.2007.00231.x
NR 24
TC 25
Z9 28
U1 0
U2 9
PU AMER SOC NEURORADIOLOGY
PI DENVILLE
PA PO BOX 3000, DENVILLE, NJ 07834-9349 USA
SN 0195-6108
EI 1936-959X
J9 AM J NEURORADIOL
JI Am. J. Neuroradiol.
PD NOV
PY 2016
VL 37
IS 11
BP 2010
EP 2018
DI 10.3174/ajnr.A4867
PG 9
WC Clinical Neurology; Neuroimaging; Radiology, Nuclear Medicine & Medical
   Imaging
WE Science Citation Index Expanded (SCI-EXPANDED)
SC Neurosciences & Neurology; Radiology, Nuclear Medicine & Medical Imaging
GA EC2PI
UT WOS:000387965400016
PM 27390318
OA Green Published, hybrid
DA 2024-06-03
ER

PT J
AU Roth, P
   Khê, HX
AF Roth, Patrick
   Khe Hoang-Xuan
TI Challenges in the treatment of elderly patients with primary central
   nervous system lymphoma
SO CURRENT OPINION IN NEUROLOGY
LA English
DT Review
DE age; elderly; primary central nervous system lymphoma; treatment
ID PRIMARY CNS LYMPHOMA; HIGH-DOSE METHOTREXATE; WHOLE-BRAIN RADIOTHERAPY;
   OLDER; PROCARBAZINE; G-PCNSL-SG-1; CHEMOTHERAPY; RITUXIMAB; OUTCOMES
AB Purpose of review
   Approximately 50% of all patients with primary central nervous system lymphoma (PCNSL) are 60 years or older and may therefore be considered as elderly. Although the diagnostic work-up is basically the same in young and in elderly patients, therapeutic strategies vary considerably. Here, we review the characteristics of elderly PCNSL patients with a particular focus on advances in the optimization of treatment regimens.
   Recent findings
   Age has been repeatedly confirmed as a major therapy-independent negative prognostic factor. Benefit from treatment and the tolerability of tumor-specific therapy, particularly whole-brain radiotherapy, are significantly lower in the elderly patients. Still, for patients with newly diagnosed PCNSL, several studies emphasized the indisputable role of high-dose methotrexate as backbone for any therapy regimen also in elderly patients. However, the durability of responses to primary chemotherapy is significantly shorter than in young patients. Recent data from a randomized phase II study for elderly PCNSL patients suggest that the combination of high-dose methotrexate, procarbazine, vincristine and cytarabine is superior to methotrexate in combination with temozolomide.
   Summary
   Current efforts aim at treating elderly PCNSL patients within clinical trials that are specifically designed for this group of patients. Determining adapted consolidation and/or maintenance treatment to improve disease control in responding patients are the main challenges to be faced by future trials. Together with a better understanding of age-specific changes in the biology of PCNSL, this will pave the road for elderly tailored therapies.
C1 [Roth, Patrick] Univ Zurich Hosp, Dept Neurol, CH-8091 Zurich, Switzerland.
   [Roth, Patrick] Univ Zurich Hosp, Brain Tumor Ctr, CH-8091 Zurich, Switzerland.
   [Khe Hoang-Xuan] Grp Hosp Pitie Salpetriere, APHP, Dept Neurol, Paris, France.
   [Khe Hoang-Xuan] Univ Paris 06, Sorbonne Univ, UM 75, F-75252 Paris 05, France.
   [Khe Hoang-Xuan] ICM, Paris, France.
   [Khe Hoang-Xuan] LOC Network, Paris, France.
   [Khe Hoang-Xuan] IHU Paris, Paris, France.
C3 University of Zurich; University Zurich Hospital; University of Zurich;
   University Zurich Hospital; Assistance Publique Hopitaux Paris (APHP);
   Hopital Universitaire Ambroise-Pare - APHP; Hopital Universitaire
   Pitie-Salpetriere - APHP; Universite Paris Cite; Hopital Universitaire
   Hotel-Dieu - APHP; Sorbonne Universite; Sorbonne Universite
RP Roth, P (corresponding author), Univ Zurich Hosp, Dept Neurol, Frauenklin Str 26, CH-8091 Zurich, Switzerland.
EM
RI Roth, Patrick/R-6671-2018; Hoang-Xuan, Khe/O-7527-2017
OI Hoang Xuan, Khe/0000-0003-4072-1999
CR Abrey LE, 2006, J CLIN ONCOL, V24, P5711, DOI 10.1200/JCO.2006.08.2941
   Abrey LE, 2000, J CLIN ONCOL, V18, P3144, DOI 10.1200/JCO.2000.18.17.3144
   Bergner N, 2012, COCHRANE DB SYST REV, DOI 10.1002/14651858.CD009355.pub2
   Bruno A, 2014, ONCOTARGET, V5, P5065, DOI 10.18632/oncotarget.2080
   Correa DD, 2012, NEURO-ONCOLOGY, V14, P101, DOI 10.1093/neuonc/nor186
   Doolittle ND, 2013, NEUROLOGY, V81, P84, DOI 10.1212/WNL.0b013e318297eeba
   Ferreri AJM, 2003, J CLIN ONCOL, V21, P266, DOI 10.1200/JCO.2003.09.139
   Fritsch K, 2011, ANN ONCOL, V22, P2080, DOI 10.1093/annonc/mdq712
   Gonzalez-Aguilar A, 2012, CLIN CANCER RES, V18, P5203, DOI 10.1158/1078-0432.CCR-12-0845
   Hajjar Emily R, 2007, Am J Geriatr Pharmacother, V5, P345, DOI 10.1016/j.amjopharm.2007.12.002
   Hoang-Xuan K, 2003, J CLIN ONCOL, V21, P2726, DOI 10.1200/JCO.2003.11.036
   Houillier C, 2014, NEUROLOGY IN PRESS
   Illerhaus G, 2009, ANN ONCOL, V20, P319, DOI 10.1093/annonc/mdn628
   Jahnke K, 2005, ANN ONCOL, V16, P445, DOI 10.1093/annonc/mdi075
   Kasenda B, 2012, ANN HEMATOL, V91, P1257, DOI 10.1007/s00277-012-1441-2
   Korfel A, 2013, NAT REV NEUROL, V9, P317, DOI 10.1038/nrneurol.2013.83
   Kurzwelly D, 2010, J NEURO-ONCOL, V97, P389, DOI 10.1007/s11060-009-0032-0
   Lange M, 2014, CANCER TREAT REV, V40, P810, DOI 10.1016/j.ctrv.2014.03.003
   Lewis JH, 2003, J CLIN ONCOL, V21, P1383, DOI 10.1200/JCO.2003.08.010
   Morris PG, 2013, J CLIN ONCOL, V31, P3971, DOI 10.1200/JCO.2013.50.4910
   Nelson DF, 1999, J NEURO-ONCOL, V43, P241, DOI 10.1023/A:1006206602918
   Ney DE, 2010, CANCER-AM CANCER SOC, V116, P4605, DOI 10.1002/cncr.25363
   Ng S, 2000, NEURO-ONCOLOGY, V2, P40, DOI 10.1093/neuonc/2.1.40
   O'Neill BP, 2013, AM J HEMATOL, V88, P997, DOI 10.1002/ajh.23551
   Omuro AMP, 2007, J NEURO-ONCOL, V85, P207, DOI 10.1007/s11060-007-9397-0
   Omuro AMP, 2013, J CLIN ONCOL, V31
   Panageas KS, 2007, CANCER, V110, P1338, DOI 10.1002/cncr.22907
   Ramjaun A, 2013, J GERIATR ONCOL, V4, P271, DOI 10.1016/j.jgo.2013.04.002
   Roth P, 2014, CURR TREAT OPTION NE, V16, DOI 10.1007/s11940-013-0277-y
   Roth P, 2012, NEUROLOGY, V79, P890, DOI 10.1212/WNL.0b013e318266fcb2
   Rubenstein JL, 2013, J CLIN ONCOL, V31, P3061, DOI 10.1200/JCO.2012.46.9957
   Schuurmans M, 2010, BRIT J HAEMATOL, V151, P179, DOI 10.1111/j.1365-2141.2010.08328.x
   Thiel E, 2010, LANCET ONCOL, V11, P1036, DOI 10.1016/S1470-2045(10)70229-1
   Weiss B, 2008, NEUROTOXICOLOGY, V29, P891, DOI 10.1016/j.neuro.2008.03.009
   Welch MR, 2012, NEURO-ONCOLOGY, V14, P1304, DOI 10.1093/neuonc/nos207
   Weller M, 2012, NEURO-ONCOLOGY, V14, P1481, DOI 10.1093/neuonc/nos159
   Zhu JJ, 2009, NEURO-ONCOLOGY, V11, P211, DOI 10.1215/15228517-2008-067
NR 37
TC 25
Z9 25
U1 0
U2 12
PU LIPPINCOTT WILLIAMS & WILKINS
PI PHILADELPHIA
PA TWO COMMERCE SQ, 2001 MARKET ST, PHILADELPHIA, PA 19103 USA
SN 1350-7540
EI 1473-6551
J9 CURR OPIN NEUROL
JI Curr. Opin. Neurol.
PD DEC
PY 2014
VL 27
IS 6
BP 697
EP 701
DI 10.1097/WCO.0000000000000145
PG 5
WC Clinical Neurology; Neurosciences
WE Science Citation Index Expanded (SCI-EXPANDED)
SC Neurosciences & Neurology
GA AS8XG
UT WOS:000344527800012
PM 25313692
OA Green Published
DA 2024-06-03
ER

PT J
AU Manoj, N
   Arivazhagan, A
   Mahadevan, A
   Bhat, DI
   Arvinda, HR
   Devi, BI
   Sampath, S
   Chandramouli, BA
AF Manoj, N.
   Arivazhagan, A.
   Mahadevan, Anita
   Bhat, Dhananjaya I.
   Arvinda, H. R.
   Devi, B. Indira
   Sampath, S.
   Chandramouli, B. A.
TI Central nervous system lymphoma: Patterns of incidence in Indian
   population and effect of steroids on stereotactic biopsy yield
SO NEUROLOGY INDIA
LA English
DT Article
DE Central nervous system lymphoma; human immunodeficiency virus;
   incidence; stereotactic biopsy; steroids
ID CLINICOPATHOLOGICAL FEATURES; CLINICAL PRESENTATION; MALIGNANT-LYMPHOMA;
   CNS-LYMPHOMA; DIAGNOSIS; IMMUNOCOMPETENT; EPIDEMIOLOGY; EXPERIENCE;
   TUMORS
AB Background: Epidemiology of primary central nervous system lymphoma (PCNSL) world-wide shows an increase in incidence linked to human immunodeficiency virus (HIV) pandemic. Materials and Methods: This retrospective review of case records analyzed the trends of hospital-based incidence of PCNSL over two decades (1991-2010), relation to immune status and effect of steroids on yield of stereotactic biopsy (STB). Results: A total of 76 cases of PCNSL were diagnosed over a period of two decades. Incidence of lymphomas amongst all biopsied lesions showed a gradual increase from 0.18% at the beginning of study period to 0.41% at the end of study period. Only 8.6% (3 of 35 tested) of the PCNSL patients were positive for HIV. The mean age of patients with HIV infection (31.3 +/- 3.5 years) was significantly lower compared with those without HIV infection (44.7 +/- 10.9 years) (P = 0.033). Diagnosis was obtained by open biopsy in 32 patients (42.1%) and STB in 44 patients (57.9%). Open biopsy yielded a histological confirmation of PCNSL in all cases. Among those who underwent STB, the incidence of negative biopsy with short duration of steroids (<= 1 week) was 33.3% and increased to 57.1% with increasing duration of steroid treatment (> 1 week). Conclusions: This study documented an increase in hospital based incidence of PCNSL in our institute, independent of HIV association. Steroid intake administration for more than a week prior to biopsy adversely affected the yield of STB in PCNSL.
C1 [Manoj, N.; Arivazhagan, A.; Bhat, Dhananjaya I.; Devi, B. Indira; Sampath, S.] Natl Inst Mental Hlth & Neurosci, Dept Neurosurg, Bengaluru, Karnataka, India.
   [Mahadevan, Anita] Natl Inst Mental Hlth & Neurosci, Dept Neuropathol, Bengaluru, Karnataka, India.
   [Arvinda, H. R.] Natl Inst Mental Hlth & Neurosci, Dept Neuroimaging & Intervent Radiol, Bengaluru, Karnataka, India.
   [Chandramouli, B. A.] Vikram Hosp, Dept Neurosurg, Bengaluru, Karnataka, India.
C3 National Institute of Mental Health & Neurosciences - India; National
   Institute of Mental Health & Neurosciences - India; National Institute
   of Mental Health & Neurosciences - India
RP Arivazhagan, A (corresponding author), Natl Inst Mental Hlth & Neurosci, Dept Neurosurg, Bengaluru, Karnataka, India.
EM
CR Agarwal PA, 2009, NEUROL INDIA, V57, P756, DOI 10.4103/0028-3886.59472
   Aker FV, 2005, NEUROPATHOLOGY, V25, P207, DOI 10.1111/j.1440-1789.2005.00634.x
   Bataille B, 2000, J NEUROSURG, V92, P261, DOI 10.3171/jns.2000.92.2.0261
   Batchelor T, 2006, J CLIN ONCOL, V24, P1281, DOI 10.1200/JCO.2005.04.8819
   Bayraktar S, 2011, J NEURO-ONCOL, V101, P257, DOI 10.1007/s11060-010-0252-3
   BRAUS DF, 1992, J NEUROL, V239, P117, DOI 10.1007/BF00833908
   Choi YL, 2006, CLIN NEUROPATHOL, V25, P29
   Donaghy M., 2002, Pract Neurol, V2, P218
   Ferreira MP, 2006, SURG NEUROL S1, V65
   FINE HA, 1993, ANN INTERN MED, V119, P1093, DOI 10.7326/0003-4819-119-11-199312010-00007
   GEPPERT M, 1990, ACTA NEUROPATHOL, V80, P629, DOI 10.1007/BF00307631
   Haldorsen IS, 2008, BMC CANCER, V8, P225
   HAYAKAWA T, 1994, J NEURO-ONCOL, V19, P197, DOI 10.1007/BF01053274
   Heper AO, 2005, SURG NEUROL, V64, P82, DOI 10.1016/j.surneu.2005.07.055
   Herrlinger U, 1998, ACTA NEUROL SCAND, V97, P257
   Herrlinger U, 1999, J NEURO-ONCOL, V43, P219, DOI 10.1023/A:1006298201101
   Jaffe Elaine S, 2009, Hematology Am Soc Hematol Educ Program, P523, DOI 10.1182/asheducation-2009.1.523
   Kadoch Cigall, 2006, Neurosurg Focus, V21, pE1
   Kumari N, 2009, J POSTGRAD MED, V55, P247, DOI 10.4103/0022-3859.58926
   Lister A., 2002, Hematology Am Soc Hematol Educ Program, P283, DOI DOI 10.1182/ASHEDUCATION-2002.1.283
   Louis DN, 2007, ACTA NEUROPATHOL, V114, P547, DOI 10.1007/s00401-007-0278-6
   Makhdoomi R, 2011, TURK NEUROSURG, V21, P39
   Olson JE, 2002, CANCER, V95, P1504, DOI 10.1002/cncr.10851
   Pasricha S, 2011, INDIAN J CANCER, V48, P199, DOI 10.4103/0019-509X.82890
   Paul T. R., 2008, Indian Journal of Cancer, V45, P112
   Rao CR, 2003, NEUROL INDIA, V51, P237
   Sarkar C, 2005, J NEURO-ONCOL, V71, P199, DOI 10.1007/s11060-004-1385-z
   Schabet M, 1999, J NEURO-ONCOL, V43, P199, DOI 10.1023/A:1006290032052
   Tandon A, 2009, NEUROL INDIA, V57, P36, DOI 10.4103/0028-3886.48813
NR 29
TC 25
Z9 26
U1 0
U2 0
PU WOLTERS KLUWER MEDKNOW PUBLICATIONS
PI MUMBAI
PA WOLTERS KLUWER INDIA PVT LTD , A-202, 2ND FLR, QUBE, C T S  NO 1498A-2
   VILLAGE MAROL, ANDHERI EAST, MUMBAI, Maharashtra, INDIA
SN 0028-3886
EI 1998-4022
J9 NEUROL INDIA
JI Neurol. India
PD JAN-FEB
PY 2014
VL 62
IS 1
BP 19
EP 25
DI 10.4103/0028-3886.128272
PG 7
WC Neurosciences
WE Science Citation Index Expanded (SCI-EXPANDED)
SC Neurosciences & Neurology
GA AD7VY
UT WOS:000333476200005
PM 24608449
DA 2024-06-03
ER

PT J
AU Zhang, JP
   Lee, EQ
   Nayak, L
   Doherty, L
   Kesari, S
   Muzikansky, A
   Norden, AD
   Chen, HC
   Wen, PY
   Drappatz, J
AF Zhang, Jun-Ping
   Lee, Eudocia Q.
   Nayak, Lakshmi
   Doherty, Lisa
   Kesari, Santosh
   Muzikansky, Alona
   Norden, Andrew D.
   Chen, Huichao
   Wen, Patrick Y.
   Drappatz, Jan
TI Retrospective study of pemetrexed as salvage therapy for central nervous
   system lymphoma
SO JOURNAL OF NEURO-ONCOLOGY
LA English
DT Article
DE Pemetrexed; Primary central nervous system lymphoma; Secondary central
   nervous system lymphoma; Salvage chemotherapy
ID PRIMARY CNS LYMPHOMA; RECURRENT PRIMARY CNS; PHASE-II; TEMOZOLOMIDE;
   RITUXIMAB; CHEMOTHERAPY; COMBINATION; TOPOTECAN
AB There is currently no standard therapy for recurrent or chemotherapy-refractory central nervous system lymphoma (CNSL). Pemetrexed has been reported to have activity in patients with primary CNSL (PCNSL). The use of pemetrexed in secondary CNS lymphoma (SCNSL) has not previously been reported. Here we retrospectively review the outcomes and toxicities of standard and modified doses of pemetrexed as salvage therapy in 18 PCNSL and 12 SCNSL patients. The overall response rate for PCNSL patients was 64.7 %, all of whom achieved a complete response (CR). The median progression-free survival (PFS) was 5.8 months. For the SCNSL patients, RR was 58.3 % with 2 CR (16.7 %); the median PFS was 2.5 months. Grade a parts per thousand yen3 adverse events included leukopenia in 5 patients (16.7 %), neutropenia in 1 patient (3.3 %), and fatigue in 3 patients (10.0 %). 3 patients died while on treatment, 2 due to infections and 1 due to pulmonary embolism. Our results indicate that pemetrexed has activity as salvage therapy in recurrent PCNSL, even with modified dosing, but outcomes trend towards less favorable in SCNSL.
C1 [Zhang, Jun-Ping] Capital Med Univ, Beijing Sanbo Brain Hosp, Dept Neurooncol, Beijing, Peoples R China.
   [Lee, Eudocia Q.; Nayak, Lakshmi; Doherty, Lisa; Norden, Andrew D.; Wen, Patrick Y.] Dana Farber Brigham & Womens Canc Ctr, Ctr Neurooncol, Boston, MA USA.
   [Kesari, Santosh] Univ Calif San Diego, Ctr Translat Neurooncol, Dept Neurosci, La Jolla, CA USA.
   [Muzikansky, Alona] Massachusetts Gen Hosp, Dept Biostat, Boston, MA 02114 USA.
   [Chen, Huichao] Harvard Univ, Sch Publ Hlth, Dept Biostat, Bostan, MA USA.
   [Drappatz, Jan] Univ Pittsburgh, Med Ctr, Dept Neurol, Div Hematol & Oncol, Pittsburgh, PA 15232 USA.
   [Drappatz, Jan] Univ Pittsburgh, Med Ctr, Dept Med, Div Hematol & Oncol, Pittsburgh, PA 15232 USA.
C3 Capital Medical University; Harvard University; Dana-Farber Cancer
   Institute; University of California System; University of California San
   Diego; Harvard University; Massachusetts General Hospital; Harvard
   University; Harvard T.H. Chan School of Public Health; Pennsylvania
   Commonwealth System of Higher Education (PCSHE); University of
   Pittsburgh; Pennsylvania Commonwealth System of Higher Education
   (PCSHE); University of Pittsburgh
RP Drappatz, J (corresponding author), Univ Pittsburgh, Med Ctr, Dept Neurol, Div Hematol & Oncol, UPMC Canc Pavil,5150 Ctr Ave, Pittsburgh, PA 15232 USA.
EM
RI Wen, Patrick/AAE-3776-2022; Kesari, Santosh/E-8461-2013
FU Society for Neuro-Oncology International Outreach Fellowship; American
   Brain Tumor Association; EMD Serono
FX J-PZ was supported by the Society for Neuro-Oncology International
   Outreach Fellowship with funding from the American Brain Tumor
   Association and EMD Serono.
CR Arellano-Rodrigo E, 2003, EUR J HAEMATOL, V70, P219, DOI 10.1034/j.1600-0609.2003.00045.x
   Batchelor TT, 2011, NEUROLOGY, V76, P929, DOI 10.1212/WNL.0b013e31820f2d94
   Bernstein SH, 2009, J CLIN ONCOL, V27, P114, DOI 10.1200/JCO.2008.16.8021
   Enting RH, 2004, NEUROLOGY, V63, P901, DOI 10.1212/01.WNL.0000137050.43114.42
   Fischer L, 2006, ANN ONCOL, V17, P1141, DOI 10.1093/annonc/mdl070
   Gerstner ER, 2010, ARCH NEUROL-CHICAGO, V67, P291, DOI 10.1001/archneurol.2010.3
   Herrlinger U, 2000, NEUROLOGY, V54, P1707, DOI 10.1212/WNL.54.8.1707
   MACDONALD DR, 1990, J CLIN ONCOL, V8, P1277, DOI 10.1200/JCO.1990.8.7.1277
   Makino K, 2012, J NEURO-ONCOL, V106, P155, DOI 10.1007/s11060-011-0652-z
   Nayak L, 2012, LEUKEMIA LYMPHOMA, V19, P1501
   Nayak L, 2013, LEUKEMIA LYMPHOMA, V54, P58, DOI 10.3109/10428194.2012.698736
   Plotkin SR, 2004, CLIN CANCER RES, V10, P5643, DOI 10.1158/1078-0432.CCR-04-0159
   Raizer JJ, 2012, CANCER-AM CANCER SOC, V118, P3743, DOI 10.1002/cncr.26709
   Reni M, 2007, BRIT J CANCER, V96, P864, DOI 10.1038/sj.bjc.6603660
   Reni M, 2000, J NATL CANCER I, V92, P575, DOI 10.1093/jnci/92.7.575
   Soussain C, 2008, J CLIN ONCOL, V26, P2512, DOI 10.1200/JCO.2007.13.5533
   Voloschin AD, 2008, J NEURO-ONCOL, V86, P211, DOI 10.1007/s11060-007-9464-6
   Wong ET, 2005, NEUROLOGY, V64, P934, DOI 10.1212/WNL.64.5.934
   Wong SF, 2012, J CLIN NEUROSCI, V19, P1501, DOI 10.1016/j.jocn.2012.04.001
NR 19
TC 25
Z9 27
U1 0
U2 3
PU SPRINGER
PI NEW YORK
PA 233 SPRING ST, NEW YORK, NY 10013 USA
SN 0167-594X
J9 J NEURO-ONCOL
JI J. Neuro-Oncol.
PD OCT
PY 2013
VL 115
IS 1
BP 71
EP 77
DI 10.1007/s11060-013-1196-1
PG 7
WC Oncology; Clinical Neurology
WE Science Citation Index Expanded (SCI-EXPANDED)
SC Oncology; Neurosciences & Neurology
GA 227VP
UT WOS:000325143900009
PM 23828279
DA 2024-06-03
ER

PT J
AU Bessell, EM
   Dickinson, P
   Dickinson, S
   Salmon, J
AF Bessell, Eric M.
   Dickinson, Peter
   Dickinson, Stephen
   Salmon, Jennifer
TI Increasing age at diagnosis and worsening renal function in patients
   with primary central nervous system lymphoma
SO JOURNAL OF NEURO-ONCOLOGY
LA English
DT Article
DE Primary central nervous system lymphoma; Renal function; Age;
   Methotrexate
ID HIGH-DOSE METHOTREXATE; NON-HODGKINS-LYMPHOMA; PRIMARY CNS LYMPHOMA;
   RADIOTHERAPY; CHEMOTHERAPY; MODEL
AB High-dose Methotrexate (MTX) is the most useful cytotoxic drug used in the treatment of primary central nervous system lymphoma (PCNSL). Dose reduction should be made in patients with reduced renal function. We evaluated the age of patients over a period of 22 years and estimated their glomerular filtration rate. One hundred and two patients were treated in Nottingham University Hospitals (a regional centre for neurosurgery) during the period 1986-2008. Patients were treated either with the BVAM regimen (carmustine, vincristine, cytarabine, MTX) or with CHOD (cyclophosphamide, doxorubicin, vincristine and dexamethasone) given for one cycle prior to BVAM. The age at which patients were diagnosed with PCNSL increased during the period of the study. During the first half of the study period (1986-1997) the median age was 60.5 years, compared to a median age of 65 years during the second half of the study period (1998-2008) (P = 0.001). The estimated glomerular filtration rate decreased with age over 40 years in a similar way to the general population. The increasing age of patients with PCNSL and decreasing renal function limit the intensity of chemotherapy with MTX containing regimens.
C1 [Bessell, Eric M.; Dickinson, Peter; Salmon, Jennifer] City Hosp Nottingham, Dept Clin Oncol, Nottingham NG5 1PB, England.
   [Dickinson, Stephen] Royal Cornwall Hosp, Dept Renal Med, Truro TR1 3LJ, Cornwall, England.
C3 Nottingham University Hospital NHS Trust; Nottingham City Hospital;
   Royal Cornwall Hospital
RP Dickinson, P (corresponding author), City Hosp Nottingham, Dept Clin Oncol, Hucknall Rd, Nottingham NG5 1PB, England.
EM
OI Dickinson, Stephen/0000-0002-9067-6957
CR Abrey LE, 2006, J CLIN ONCOL, V24, P5711, DOI 10.1200/JCO.2006.08.2941
   [Anonymous], 1993, N ENGL J MED, V329, P987
   Batchelor T, 2003, J CLIN ONCOL, V21, P1044, DOI 10.1200/JCO.2003.03.036
   Bessell E M, 1991, Clin Oncol (R Coll Radiol), V3, P193, DOI 10.1016/S0936-6555(05)80738-1
   Bessell EM, 2004, INT J RADIAT ONCOL, V59, P501, DOI 10.1016/j.ijrobp.2003.11.001
   Bessell EM, 2007, EUR J CANCER, V43, P1141, DOI 10.1016/j.ejca.2006.12.011
   DEANGELIS LM, 1990, NEUROLOGY, V40, P80, DOI 10.1212/WNL.40.1.80
   FERRERI AJ, 2008, ANN ONCOL, V19
   Ferreri AJM, 2003, J CLIN ONCOL, V21, P266, DOI 10.1200/JCO.2003.09.139
   Grewal GS, 2005, NUCL MED COMMUN, V26, P61, DOI 10.1097/00006231-200501000-00010
   Herrlinger U, 2005, ANN NEUROL, V57, P843, DOI 10.1002/ana.20495
   Jahnke K, 2005, ANN ONCOL, V16, P445, DOI 10.1093/annonc/mdi075
   Kadan-Lottick NS, 2002, CANCER, V95, P193, DOI 10.1002/cncr.10643
   Olson JE, 2002, CANCER, V95, P1504, DOI 10.1002/cncr.10851
   Omuro Antonio M P, 2006, Neurosurg Focus, V21, pE12
NR 15
TC 25
Z9 26
U1 0
U2 1
PU SPRINGER
PI NEW YORK
PA 233 SPRING ST, NEW YORK, NY 10013 USA
SN 0167-594X
J9 J NEURO-ONCOL
JI J. Neuro-Oncol.
PD AUG
PY 2011
VL 104
IS 1
BP 191
EP 193
DI 10.1007/s11060-010-0457-5
PG 3
WC Oncology; Clinical Neurology
WE Science Citation Index Expanded (SCI-EXPANDED)
SC Oncology; Neurosciences & Neurology
GA 804IS
UT WOS:000293648100018
PM 21103909
DA 2024-06-03
ER

PT J
AU Ferreri, AJM
   Licata, G
   Foppoli, M
   Corazzelli, G
   Zucca, E
   Stelitano, C
   Zaja, F
   Fava, S
   Paolini, R
   Franzin, A
   Politi, LS
   Ponzoni, M
   Reni, M
AF Ferreri, Andres J. M.
   Licata, Giada
   Foppoli, Marco
   Corazzelli, Gaetano
   Zucca, Emanuele
   Stelitano, Caterina
   Zaja, Francesco
   Fava, Sergio
   Paolini, Rossella
   Franzin, Alberto
   Politi, Letterio S.
   Ponzoni, Maurilio
   Reni, Michele
TI Clinical Relevance of the Dose of Cytarabine in the Upfront Treatment of
   Primary CNS Lymphomas with Methotrexate-Cytarabine Combination
SO ONCOLOGIST
LA English
DT Article
DE Primary central nervous system lymphoma; Methotrexate; Cytarabine; IELSG
   prognostic score; Chemotherapy; Thiotepa
ID CYTOSINE-ARABINOSIDE; CEREBROSPINAL-FLUID; PHARMACOKINETICS;
   1-BETA-D-ARABINOFURANOSYLCYTOSINE; SYSTEM; PLASMA
AB Background. The combination of high doses of methotrexate (MTX) and cytarabine (araC) is the standard chemotherapy for patients with primary CNS lymphoma (PCNSL). The addition of an alkylating agent could improve MTX-araC efficacy because it is active against quiescent G0 cells and increases anti-metabolites cytotoxicity. A pilot experience with high doses of MTX, araC, and thiotepa (MAT regimen) was performed to investigate feasibility and efficacy of adding an alkylating agent. With respect to MTX-araC combination, araC dose was halved to minimize toxicity. Herein, we report tolerability, activity, and efficacy of MAT regimen and compare these results to those previously reported with MTX/ara-C combination.
   Methods. Twenty HIV-negative patients with PCNSL treated with MAT regimen and whole-brain irradiation and selected according to eligibility criteria of the International Extranodal Lymphoma Study Group (IELSG) #20 trial were analyzed.
   Results. Patient characteristics of MAT and MTX-araC series were similar. G4 hematologic toxicity was common after MAT chemotherapy, with dose reductions in 60% of patients, infections in 20%, G4 non-hematologic toxicity in 15%, and one (5%) toxic death. Response after chemotherapy was complete in four patients (clinical response rate, 20%; 95% confidence interval, 3%-37%) and partial in three (overall response rate, 35%; 95% confidence interval, 15%-55%). Fifteen patients experienced failure and 16 died (median follow-up, 26 months), with a 2-year overall survival of 24% +/- 9%.
   Conclusions. MAT and MTX-araC combinations showed similar tolerability, whereas araC dose reduction was associated with a remarkably lower efficacy, hiding any potential benefit of thiotepa. Four doses of araC 2 g/m(2) per course are recommended in patients with PCNSL. The Oncologist 2011;16:336-341
C1 [Ferreri, Andres J. M.; Licata, Giada; Foppoli, Marco; Ponzoni, Maurilio; Reni, Michele] Ist Sci San Raffaele, Dept Oncol, Med Oncol Unit, Unit Lymphoid Malignancies, I-20132 Milan, Italy.
   [Licata, Giada; Foppoli, Marco] Ist Sci San Raffaele, Dept Oncol, Unit Internal Med, I-20132 Milan, Italy.
   [Franzin, Alberto] Ist Sci San Raffaele, Dept Oncol, Neurosurg Unit, I-20132 Milan, Italy.
   [Politi, Letterio S.] Ist Sci San Raffaele, Dept Oncol, Neuroradiol Unit, I-20132 Milan, Italy.
   [Ponzoni, Maurilio] Ist Sci San Raffaele, Dept Oncol, Pathol Unit, I-20132 Milan, Italy.
   [Corazzelli, Gaetano] Ist Pascale, Div Hematol, Naples, Italy.
   [Zucca, Emanuele] Oncol Inst So Switzerland, Lymphoma Unit, Bellinzona, Switzerland.
   [Stelitano, Caterina] Azienda Osped Reggio, Div Hematol, Calabria, Italy.
   [Zaja, Francesco] Univ Udine, Clin Haematol, I-33100 Udine, Italy.
   [Fava, Sergio] Azienda Osped Legnano, Oncol Unit, Legnano, Italy.
   [Paolini, Rossella] Azienda Osped Rovigo, Dept Oncol, Oncohematol Unit, Rovigo, Italy.
C3 Vita-Salute San Raffaele University; IRCCS Ospedale San Raffaele;
   Vita-Salute San Raffaele University; IRCCS Ospedale San Raffaele;
   Vita-Salute San Raffaele University; IRCCS Ospedale San Raffaele;
   Vita-Salute San Raffaele University; IRCCS Ospedale San Raffaele;
   Vita-Salute San Raffaele University; IRCCS Ospedale San Raffaele; IRCCS
   Fondazione Pascale; Institute of Oncology Research (IOR); University of
   Udine; ULSS 5 Polesana; Ospedale di Rovigo
RP Ferreri, AJM (corresponding author), Ist Sci San Raffaele, Dept Oncol, Med Oncol Unit, Unit Lymphoid Malignancies, I-20132 Milan, Italy.
EM
RI Italiana Linfomi, Fondazione/AAW-6910-2021; Politi, Letterio
   Salvatore/AAB-1092-2021; Ferreri, Andrés J. M./A-6662-2013; Reni,
   Michele/J-8296-2016
OI Italiana Linfomi, Fondazione/0000-0002-5918-8824; Politi, Letterio
   Salvatore/0000-0002-6190-6688; PAOLINI, Rossella/0000-0001-7605-1531;
   Franzin, Alberto/0000-0002-4962-6413; Reni, Michele/0000-0002-6463-0293;
   Zaja, Francesco/0000-0003-3995-8817; Zucca, Emanuele/0000-0002-5522-6109
CR Ansell SM, 2010, NAT REV CLIN ONCOL, V7, P125, DOI 10.1038/nrclinonc.2010.9
   BREITHAUPT H, 1982, CANCER-AM CANCER SOC, V50, P1248, DOI 10.1002/1097-0142(19821001)50:7<1248::AID-CNCR2820500705>3.0.CO;2-5
   Cheson BD, 1999, J CLIN ONCOL, V17, P1244, DOI 10.1200/JCO.1999.17.4.1244
   DAMON LE, 1991, CANCER RES, V51, P4141
   Ferreri AJM, 2003, J CLIN ONCOL, V21, P266, DOI 10.1200/JCO.2003.09.139
   Ferreri AJM, 2009, LANCET, V374, P1512, DOI 10.1016/S0140-6736(09)61416-1
   HRYNIUK WM, 1990, J CLIN ONCOL, V8, P1935, DOI 10.1200/JCO.1990.8.12.1935
   O'Brien PC, 2009, LANCET, V374, P1477, DOI 10.1016/S0140-6736(09)61488-4
   SLEVIN ML, 1983, J CLIN ONCOL, V1, P546, DOI 10.1200/JCO.1983.1.9.546
   TAKASHIMA Y, 1987, J CLIN PHARMACOL, V27, P330, DOI 10.1002/j.1552-4604.1987.tb03025.x
   Trotti A, 2003, SEMIN RADIAT ONCOL, V13, P176, DOI 10.1016/S1053-4296(03)00031-6
NR 11
TC 25
Z9 27
U1 0
U2 1
PU ALPHAMED PRESS
PI DURHAM
PA 318 BLACKWELL ST, STE 260, DURHAM, NC 27701-2884 USA
SN 1083-7159
J9 ONCOLOGIST
JI Oncologist
PY 2011
VL 16
IS 3
BP 336
EP 341
DI 10.1634/theoncologist.2010-0361
PG 6
WC Oncology
WE Science Citation Index Expanded (SCI-EXPANDED)
SC Oncology
GA 737WU
UT WOS:000288601800009
PM 21346023
OA Green Published, Bronze
DA 2024-06-03
ER

PT J
AU Kawai, N
   Nishiyama, Y
   Miyake, K
   Tamiya, T
   Nagao, S
AF Kawai, N
   Nishiyama, Y
   Miyake, K
   Tamiya, T
   Nagao, S
TI Evaluation of tumor FDG transport and metabolism in primary central
   nervous system lymphoma using [<SUP>18</SUP>F]fluorodeoxyglucose (FDG)
   positron emission tomography (PET) kinetic analysis
SO ANNALS OF NUCLEAR MEDICINE
LA English
DT Article
DE glucose transport; glucose phosphorylation; fluorodeoxyglucose; PET
   (positron emission tomography); primary central nervous system lymphoma
ID FOLLOW-UP; BRAIN; CHEMOTHERAPY; DIAGNOSIS
AB Objective: Although F-18-fluorodeoxyglucose (FDG) positron emission tomography (PET) has been used as a promising tool to diagnose primary central nervous system lymphoma (PCNSL) because the tumor shows very high FDG accumulation, no data exist evaluating the extent of tumor FDG transport and metabolism. The aim of this study was to evaluate the feasibility of FDG-PET kinetic analysis in measurement of uptake parameters of FDG in the lymphoma tissues and in the assessment of treatment effects in patients with PCNSL. Methods: Dynamic FDG-PET examination was performed in 7 histologically proven PCNSL patients before and after methotrexate-based chemotherapy. Results: Before the chemotherapy, the highest CMRglc in the tumor for all 7 patients was 79.4 +/- 27.2 mu mol/100 g/min. This value was significantly higher than that observed in the normal cortex in 14 control patients (44.3 +/- 6.0 mu mol/100 g/min, p < 0.001). The phosphorylation (k(3)) activity was also significantly higher in the tumor (0.093 +/- 0.026 min(-1)) compared with the normal cortex (0.064 +/- 0.014 min(-1), p < 0.05). On the other hand, the transporter (K-I) activity in the tumor (0.079 +/- 0.016 ml/min) was similar to that observed in the normal cortex (0.082 +/- 0.012 ml/min). The chemotherapy significantly reduced the volume of the tumor in 6 of 7 patients and the highest CMRglc in the tumor examined 18.0 +/- 5.5 days after the chemotherapy (34.0 +/- 21.8 mu mol/ 100 g/min) was significantly lower than that observed before the chemotherapy (p < 0.01). This reduction in FDG uptake was concomitant with a significant reduction in both the K-I and k(3) values (p < 0.05). The reduction in the k(3) value after the chemotherapy was marked in 6 of 7 patients in whom the tumor responded to the first chemotherapy. Conclusions: Dynamic image acquisition can separate regional FDG uptake into FDG transport and phosphorylation activity in the lymphoma tissues. Tumor FDG uptake was significantly higher with accelerated phosphorylation activity compared with that observed in the normal cortex.
C1 Kagawa Univ, Sch Med, Dept Neurol Surg, Kagawa 7610793, Japan.
   Kagawa Univ, Sch Med, Dept Radiol, Kagawa 7610793, Japan.
C3 Kagawa University; Kagawa University
RP Kawai, N (corresponding author), Kagawa Univ, Sch Med, Dept Neurol Surg, 1750-1 Mikicho, Kagawa 7610793, Japan.
EM
CR Corn BW, 1997, CANCER, V79, P2409
   HERHOLZ K, 1992, J NEURO-ONCOL, V12, P159
   HOEKSTRA OS, 1993, J NUCL MED, V34, P1706
   Kim L, 2000, NEURORADIOLOGY, V42, P406, DOI 10.1007/s002340000301
   OGAWA T, 1994, RADIOGRAPHICS, V14, P101, DOI 10.1148/radiographics.14.1.8128041
   Roelcke U, 1999, J NEURO-ONCOL, V43, P231, DOI 10.1023/A:1006202402010
   Römer W, 1998, BLOOD, V91, P4464
   ROSENFELD SS, 1992, J NUCL MED, V33, P532
   Schaller B, 2004, NEUROBIOL DIS, V15, P437, DOI 10.1016/j.nbd.2003.11.023
   Spence AM, 1998, J NUCL MED, V39, P440
   VILLRINGER K, 1995, J COMPUT ASSIST TOMO, V19, P532, DOI 10.1097/00004728-199507000-00004
   WONG KP, 2001, PHISIOLOGICAL IMAGIN, P121
   Yamane T, 2004, J NUCL MED, V45, P1838
NR 13
TC 25
Z9 26
U1 0
U2 0
PU SPRINGER
PI NEW YORK
PA 233 SPRING ST, NEW YORK, NY 10013 USA
SN 0914-7187
EI 1864-6433
J9 ANN NUCL MED
JI Ann. Nucl. Med.
PD DEC
PY 2005
VL 19
IS 8
BP 685
EP 690
DI 10.1007/BF02985117
PG 6
WC Radiology, Nuclear Medicine & Medical Imaging
WE Science Citation Index Expanded (SCI-EXPANDED)
SC Radiology, Nuclear Medicine & Medical Imaging
GA 998HM
UT WOS:000234309300007
PM 16444994
DA 2024-06-03
ER

PT J
AU Pramstaller, PP
   Salerno, A
   Bhatia, KP
   Prugger, M
   Marsden, CD
AF Pramstaller, PP
   Salerno, A
   Bhatia, KP
   Prugger, M
   Marsden, CD
TI Primary central neurons system lymphoma presenting with a parkinsonian
   syndrome of pure akinesia
SO JOURNAL OF NEUROLOGY
LA English
DT Article
DE parkinsonism; akinesia; lymphoma
ID CENTRAL-NERVOUS-SYSTEM; GLOBUS-PALLIDUS; LESIONS; CLASSIFICATION;
   EXPERIENCE
AB The incidence of primary central nervous system lymphoma (PCNSL), once a ran tumour, has risen significantly in both immunocompetent and immunosuppressed patients. Although infiltration of the basal ganglia is not uncommon in PCNSL, extrapyramidal movement disorders an generally not recognised as a mode of clinical presentation of this type of cerebral tumour. We present the unusual case of a 75-year-old man who developed a parkinsonian syndrome of "pure akinesia" due to autopsy-confirmed PCNSL primarily involving the globus pallidus bilaterally. Parkinsonism due to bilateral pallidal lesions is known but rare, and such cases help in the understanding of basal ganglia function with regard to akinesia and freezing.
C1 Inst Neurol, Univ Dept Clin Neurol, London WC1N 3BG, England.
   Reg Gen Hosp, Dept Neurol, Bolzano, Italy.
   Reg Gen Hosp, Dept Pathol, Bolzano, Italy.
   Univ Dept Neurol, Innsbruck, Austria.
C3 University of London; University College London
RP Bhatia, KP (corresponding author), Inst Neurol, Univ Dept Clin Neurol, Queen Sq, London WC1N 3BG, England.
RI Pramstaller, Peter P/C-2357-2008
CR BHATIA KP, 1994, BRAIN, V117, P859, DOI 10.1093/brain/117.4.859
   CHAN JKC, 1994, HISTOPATHOLOGY, V25, P517, DOI 10.1111/j.1365-2559.1994.tb01371.x
   DEANGELIS LM, 1990, J NEURO-ONCOL, V9, P177, DOI 10.1007/BF02427840
   EBY NL, 1988, CANCER, V62, P2641
   FEVE AP, 1993, MOVEMENT DISORD, V8, P321, DOI 10.1002/mds.870080311
   FINE HA, 1993, ANN INTERN MED, V119, P1093, DOI 10.7326/0003-4819-119-11-199312010-00007
   FREILICH RJ, 1995, NEUROL CLIN, V13, P901, DOI 10.1016/S0733-8619(18)30024-0
   GRANT JW, 1992, BRAIN PATHOL, V2, P97, DOI 10.1111/j.1750-3639.1992.tb00677.x
   Haaxma R, 1995, MOVEMENT DISORD, V10, P761, DOI 10.1002/mds.870100610
   HELLE TL, 1984, J NEUROSURG, V60, P94, DOI 10.3171/jns.1984.60.1.0094
   HENRY JM, 1974, CANCER, V34, P1293, DOI 10.1002/1097-0142(197410)34:4<1293::AID-CNCR2820340441>3.0.CO;2-P
   HERMAN MD, 1993, J NEUROSURG, V78, P997, DOI 10.3171/jns.1993.78.6.0997
   IMAI H, 1993, ADV NEUROL, V60, P622
   Imai H, 1974, SHINKEI XENKYU SHIMP, V18, P787
   JACK CR, 1988, RADIOLOGY, V167, P211, DOI 10.1148/radiology.167.1.3279454
   LAPLANE D, 1993, BRAIN, V50, P873
   MATSUO H, 1991, J NEUROL NEUROSUR PS, V54, P397, DOI 10.1136/jnnp.54.5.397
   MILLER DC, 1994, CANCER-AM CANCER SOC, V74, P1383, DOI 10.1002/1097-0142(19940815)74:4<1383::AID-CNCR2820740432>3.0.CO;2-1
   MISUZAWA H, 1993, ADV NEUROL, V60, P618
   Narabayashi H., 1976, ADV PARKINSONISM, P335
   POEWE WH, 1988, MOVEMENT DISORD, V4, P320
   ROSENBERG SA, 1982, CANCER, V49, P2112
   STANSFELD AG, 1988, LANCET, V1, P292
   Suzuki T, 1984, No To Shinkei, V36, P689
   WEISSMAN DE, 1990, J NEURO-ONCOL, V9, P171, DOI 10.1007/BF02427839
NR 25
TC 25
Z9 27
U1 0
U2 1
PU DR DIETRICH STEINKOPFF VERLAG
PI DARMSTADT
PA PLATZ DER DEUTSCHEN EINHEIT 25, D-64293 DARMSTADT, GERMANY
SN 0340-5354
J9 J NEUROL
JI J. Neurol.
PD OCT
PY 1999
VL 246
IS 10
BP 934
EP 938
DI 10.1007/s004150050485
PG 5
WC Clinical Neurology
WE Science Citation Index Expanded (SCI-EXPANDED)
SC Neurosciences & Neurology
GA 252GU
UT WOS:000083494400010
PM 10552241
DA 2024-06-03
ER

PT J
AU Harada, K
   Kurisu, K
   Arita, K
   Sadatomo, T
   Tahara, H
   Tahara, E
   Ide, T
   Tahara, E
   Uozumi, T
AF Harada, K
   Kurisu, K
   Arita, K
   Sadatomo, T
   Tahara, H
   Tahara, E
   Ide, T
   Tahara, E
   Uozumi, T
TI Telomerase activity in central nervous system malignant lymphoma
SO CANCER
LA English
DT Article
DE telomerase; hTERT; hTERC; brain; malignant lymphoma
ID CATALYTIC SUBUNIT; HEPATOCELLULAR CARCINOMAS; INCREASING INCIDENCE;
   IMMORTAL CELLS; CNS-LYMPHOMA; BRAIN; CANCER; GENE; CHEMOTHERAPY;
   RADIOTHERAPY
AB BACKGROUND, Primary central nervous system malignant lymphoma (PCNSL) has been regarded as a rare neoplasm. Recently, however, its incidence has been rapidly increasing. Despite active clinical trials, its clinical and biologic features remain unknown and there has been no effective treatment or prognostic factor. The current study attempted to elucidate telomerase activity and expression of the telomere-related RNA of PCNSL as novel prognostic factors.
   METHODS. The subjects were 12 patients with histologically diagnosed PCNSL. All patients were treated with chemoradiotherapy consisting of whole-brain radiation, vincristine, etoposide, and prednisolone. Telomerase activity in the resected tumor was analyzed both qualitatively and quantitatively using the TRAP (telomeric amplification assay protocol) /TRAP-HPA (hybridization protection assay) method. The expression of telomere-related RNA (hTERT, hTERC, and TEP1) was analyzed by reverse transcriptase-polymerase chain reaction.
   RESULTS. Telomerase activity was detected in 10 of 12 patients with PCNSL, with an average activity of 148.9 relative light units. The average telomere length was 5.55 kb. The expression of hTERT correlated with telomerase activity, and there was a statistically significant correlation between telomerase activity and both the duration of survival and the interval to tumor progression.
   CONCLUSIONS. Telomerase activity and the expression of hTERT may be novel prognostic factors in patients with PCNSL. (C) 1999 American Cancer Society.
C1 Hiroshima Univ, Sch Med, Dept Neurosurg, Minami Ku, Hiroshima, Japan.
   Hiroshima Univ, Sch Med, Dept Cellular & Mol Biol, Hiroshima, Japan.
   Hiroshima Univ, Sch Med, Dept Pathol 1, Hiroshima, Japan.
   Hiroshima Prefectural Hosp, Hiroshima, Japan.
C3 Hiroshima University; Hiroshima University; Hiroshima University
RP Harada, K (corresponding author), Hiroshima Univ, Sch Med, Dept Neurosurg, Minami Ku, Kasumi 1-2-3, Hiroshima, Japan.
RI Kowaltowski, Alicia J/H-8698-2012; Arita, Ken/A-5451-2012
CR AMADORI S, 1984, J CLIN ONCOL, V2, P98, DOI 10.1200/JCO.1984.2.2.98
   AOZASA K, 1985, EUR J CANCER CLIN ON, V21, P487, DOI 10.1016/0277-5379(85)90042-2
   BRADA M, 1990, INT J RADIAT ONCOL, V18, P787, DOI 10.1016/0360-3016(90)90398-4
   CHADENEAU C, 1995, CANCER RES, V55, P2533
   COUNTER CM, 1992, EMBO J, V11, P1921, DOI 10.1002/j.1460-2075.1992.tb05245.x
   Counter CM, 1997, P NATL ACAD SCI USA, V94, P9202, DOI 10.1073/pnas.94.17.9202
   COX DR, 1972, J R STAT SOC B, V34, P187
   DEANGELIS LM, 1990, NEUROLOGY, V40, P80, DOI 10.1212/WNL.40.1.80
   EBY NL, 1988, CANCER-AM CANCER SOC, V62, P2461, DOI 10.1002/1097-0142(19881201)62:11<2461::AID-CNCR2820621135>3.0.CO;2-M
   FENG JL, 1995, SCIENCE, V269, P1236, DOI 10.1126/science.7544491
   FINE HA, 1993, ANN INTERN MED, V119, P1093, DOI 10.7326/0003-4819-119-11-199312010-00007
   HARADA K, IN PRESS NEUROL MED
   HARLEY CB, 1990, NATURE, V345, P458, DOI 10.1038/345458a0
   HARLEY CB, 1995, CURR OPIN GENET DEV, V5, P249, DOI 10.1016/0959-437X(95)80016-6
   HAYAKAWA T, 1994, J NEURO-ONCOL, V19, P197, DOI 10.1007/BF01053274
   Hirose M, 1997, J CANCER RES CLIN, V123, P337, DOI 10.1007/BF01438310
   HIYAMA E, 1995, NAT MED, V1, P249, DOI 10.1038/nm0395-249
   HOCHBERG FH, 1988, J NEUROSURG, V68, P835, DOI 10.3171/jns.1988.68.6.0835
   Jellinger K, 1975, Acta Neuropathol Suppl, VSuppl 6, P95
   KIM NW, 1994, SCIENCE, V266, P2011, DOI 10.1126/science.7605428
   Kleinschmidt-DeMasters BK, 1998, J CLIN PATHOL, V51, P284, DOI 10.1136/jcp.51.4.284
   Kubota Y, 1998, CANCER, V82, P176, DOI 10.1002/(SICI)1097-0142(19980101)82:1<176::AID-CNCR22>3.3.CO;2-N
   Kumanishi T, 1996, JPN J CANCER RES, V87, P691, DOI 10.1111/j.1349-7006.1996.tb00279.x
   LANGFORD LA, 1995, LANCET, V346, P1267, DOI 10.1016/S0140-6736(95)91865-5
   LETENDRE L, 1982, CANCER-AM CANCER SOC, V49, P939, DOI 10.1002/1097-0142(19820301)49:5<939::AID-CNCR2820490518>3.0.CO;2-1
   LITTMAN P, 1975, CANCER, V35, P1412, DOI 10.1002/1097-0142(197505)35:5<1412::AID-CNCR2820350526>3.0.CO;2-T
   LOEFFLER JS, 1985, J CLIN ONCOL, V3, P490, DOI 10.1200/JCO.1985.3.4.490
   MENDENHALL NP, 1983, CANCER-AM CANCER SOC, V52, P1993, DOI 10.1002/1097-0142(19831201)52:11<1993::AID-CNCR2820521104>3.0.CO;2-C
   Meyerson M, 1997, CELL, V90, P785, DOI 10.1016/S0092-8674(00)80538-3
   Nakamura TM, 1997, SCIENCE, V277, P955, DOI 10.1126/science.277.5328.955
   Nakayama JI, 1998, NAT GENET, V18, P65, DOI 10.1038/ng0198-65
   Nakayama JI, 1997, CELL, V88, P875, DOI 10.1016/S0092-8674(00)81933-9
   NOMURA K, 1993, BRAIN TUMOR REGISTRY, V8
   OSULLIVAN MG, 1991, LANCET, V338, P895, DOI 10.1016/0140-6736(91)91561-8
   REMICK SC, 1990, MEDICINE, V69, P345, DOI 10.1097/00005792-199011000-00003
   ROSENBERG SA, 1982, CANCER, V49, P2112
   Shay Jerry W., 1994, Molecular and Cellular Differentiation, V2, P1
   SHAY JW, 1991, BIOCHIM BIOPHYS ACTA, V1072, P1, DOI 10.1016/0304-419X(91)90003-4
   TAHARA H, 1995, CANCER RES, V55, P2734
   Tatematsu K, 1996, ONCOGENE, V13, P2265
   VOGELSTEIN B, 1993, TRENDS GENET, V9, P138, DOI 10.1016/0168-9525(93)90209-Z
   Zhang SJ, 1998, CANCER RES, V58, P1231
   Zimmerman H M, 1975, Acta Neuropathol Suppl, VSuppl 6, P69
NR 43
TC 25
Z9 27
U1 0
U2 1
PU WILEY-LISS
PI NEW YORK
PA DIV JOHN WILEY & SONS INC, 605 THIRD AVE, NEW YORK, NY 10158-0012 USA
SN 0008-543X
J9 CANCER
JI Cancer
PD SEP 15
PY 1999
VL 86
IS 6
BP 1050
EP 1055
DI 10.1002/(SICI)1097-0142(19990915)86:6<1050::AID-CNCR22>3.3.CO;2-B
PG 6
WC Oncology
WE Science Citation Index Expanded (SCI-EXPANDED)
SC Oncology
GA 236DL
UT WOS:000082582900022
PM 10491533
DA 2024-06-03
ER

PT J
AU Bonzheim, I
   Sander, P
   Salmerón-Villalobos, J
   Süsskind, D
   Szurman, P
   Gekeler, F
   Spitzer, MS
   Steinhilber, J
   Kohler, E
   Büssgen, M
   Schittenhelm, J
   Salaverria, I
   Campo, E
   Coupland, SE
   Quintanilla-Martinez, L
   Fend, F
AF Bonzheim, Irina
   Sander, Philip
   Salmeron-Villalobos, Julia
   Suesskind, Daniela
   Szurman, Peter
   Gekeler, Florian
   Spitzer, Martin S.
   Steinhilber, Julia
   Kohler, Esther
   Buessgen, Melanie
   Schittenhelm, Jens
   Salaverria, Itziar
   Campo, Elias
   Coupland, Sarah E.
   Quintanilla-Martinez, Leticia
   Fend, Falko
TI The molecular hallmarks of primary and secondary vitreoretinal lymphoma
SO BLOOD ADVANCES
LA English
DT Article
ID CENTRAL-NERVOUS-SYSTEM; PRIMARY INTRAOCULAR LYMPHOMA; MYD88 L265P
   MUTATION; B-CELL LYMPHOMA; IMMUNOGLOBULIN; PREVALENCE; TBL1XR1
AB Vitreoretinal lymphoma (VRL) is a rare subtype of diffuse large B-cell lymphoma (DLBCL) considered a variant of primary central nervous system lymphoma (PCNSL). The diagnosis of VRL requires examination of vitreous fluid, but cytologic differentiation from uveitis remains difficult. Because of its rarity and the difficulty in obtaining diagnostic material, little is known about the genetic profile of VRL. The purpose of our study was to investigate the mutational profile of a large series of primary and secondary VRL. Targeted next-generation sequencing using a custom panel containing the most frequent mutations in PCNSL was performed on 34 vitrectomy samples from 31 patients with VRL and negative controls with uveitis. In a subset of cases, genome-wide copy number alterations (CNAs) were assessed using the OncoScan platform. Mutations in MYD88 (74%), PIM1 (71%), CD79B (55%), IGLL5 (52%), TBL1XR1 (48%), ETV6 (45%), and 9p21/CDKN2A deletions (75%) were the most common alterations, with similar frequencies in primary (n = 16), synchronous (n = 3), or secondary (n = 12) VRL. This mutational spectrum is similar to MYD88(mut)/CD79B(mut) (MCD or cluster 5) DLBCL with activation of Toll-like and B-cell receptor pathways and CDKN2A loss, confirming their close relationship. OncoScan analysis demonstrated a high number of CNAs (mean 18.6 per case). Negative controls lacked mutations or CNAs. Using cell-free DNA of vitreous fluid supernatant, mutations present in cellular DNA were reliably detected in all cases examined. Mutational analysis is a highly sensitive and specific tool for the diagnosis of VRL and can also be applied successfully to cell-free DNA derived from the vitreous.
C1 [Bonzheim, Irina; Sander, Philip; Steinhilber, Julia; Kohler, Esther; Buessgen, Melanie; Quintanilla-Martinez, Leticia; Fend, Falko] Eberhard Karls Univ Tubingen, Inst Pathol & Neuropathol, Dept Gen & Mol Pathol & Pathol Anat, Tubingen, Germany.
   [Bonzheim, Irina; Sander, Philip; Suesskind, Daniela; Gekeler, Florian; Steinhilber, Julia; Kohler, Esther; Buessgen, Melanie; Schittenhelm, Jens; Quintanilla-Martinez, Leticia; Fend, Falko] Univ Hosp Tubingen, Comprehens Canc Ctr, Tubingen, Germany.
   [Salmeron-Villalobos, Julia; Salaverria, Itziar; Campo, Elias] Hosp Clin Barcelona, Inst Invest Biomed August Pi i Sunyer, Hematopathol Unit, Barcelona, Spain.
   [Salmeron-Villalobos, Julia; Salaverria, Itziar; Campo, Elias] Ctr Invest Biomed Red Oncol CIBERONC, Madrid, Spain.
   [Suesskind, Daniela; Gekeler, Florian] Eberhard Karls Univ Tubingen, Ctr Ophthalmol, Tubingen, Germany.
   [Szurman, Peter] Knappschaft Hosp Saar, Sulzbach Eye Clin, Sulzbach, Germany.
   [Gekeler, Florian] Klinikum Stuttgart, Dept Ophthalmol, Stuttgart, Germany.
   [Spitzer, Martin S.] Univ Med Ctr Hamburg Eppendorf UKE, Clin Ophthalmol, Hamburg, Germany.
   [Schittenhelm, Jens] Eberhard Karls Univ Tubingen, Inst Pathol & Neuropathol, Dept Neuropathol, Tubingen, Germany.
   [Coupland, Sarah E.] Univ Liverpool, Inst Syst Mol & Integrat Biol, Dept Mol & Clin Canc Med, Liverpool, Merseyside, England.
   [Quintanilla-Martinez, Leticia] Univ Tubingen, Image Guided & Funct Instructed Tumor Therapies, Cluster Excellence iFIT EXC 2180, Tubingen, Germany.
C3 Eberhard Karls University of Tubingen; Eberhard Karls University
   Hospital; Eberhard Karls University of Tubingen; Eberhard Karls
   University Hospital; Universitat de Barcelona; Hospital Clinic de
   Barcelona; IDIBAPS; CIBER - Centro de Investigacion Biomedica en Red;
   CIBERONC; Eberhard Karls University of Tubingen; Eberhard Karls
   University Hospital; Klinikum Stuttgart; University of Hamburg;
   University Medical Center Hamburg-Eppendorf; Eberhard Karls University
   of Tubingen; Eberhard Karls University Hospital; University of
   Liverpool; Eberhard Karls University of Tubingen
RP Bonzheim, I (corresponding author), Univ Hosp Tubingen, Inst Pathol & Neuropathol, Liebermeisterstr 8, D-72076 Tubingen, Germany.; Fend, F (corresponding author), Tubingen Univ Hosp, Inst Pathol & Neuropathol, Liebermeisterstr 8, D-72076 Tubingen, Germany.
EM
RI Salmeron-Villalobos, Julia/AAE-2161-2020; Campo, Elias/AAC-5593-2019
OI Campo, Elias/0000-0001-9850-9793; Salmeron Villalobos,
   Julia/0000-0002-7092-9160; Coupland, Sarah/0000-0002-1464-2069
FU Spanish Ministerio de Economia y Competitividad [RTI2018-094274-B-I00];
   Generalitat de Catalunya Suport Grups de Recerca [2017-SGR-1142]
FX The authors thank Sema Colak for excellent technical assistance. E.C. is
   supported by Spanish Ministerio de Economia y Competitividad
   (RTI2018-094274-B-I00) and Generalitat de Catalunya Suport Grups de
   Recerca (2017-SGR-1142) and is also an Academia Researcher of the
   "Institucio Catalana de Recerca i Estudis Avancats" (ICREA) of the
   Generalitat de Catalunya.
CR Bödör C, 2020, J NEUROPATH EXP NEUR, V79, P176, DOI 10.1093/jnen/nlz125
   Bonzheim I, 2015, BLOOD, V126, P76, DOI 10.1182/blood-2015-01-620518
   Braggio E, 2015, CLIN CANCER RES, V21, P3986, DOI 10.1158/1078-0432.CCR-14-2116
   Bruno A, 2018, NEURO-ONCOLOGY, V20, P1092, DOI 10.1093/neuonc/noy019
   Bruno A, 2014, ONCOTARGET, V5, P5065, DOI 10.18632/oncotarget.2080
   Camilleri-Broët S, 2006, BLOOD, V107, P190, DOI 10.1182/blood-2005-03-1024
   Cani AK, 2017, ONCOTARGET, V8, P7989, DOI 10.18632/oncotarget.14008
   Cassoux N, 2001, OPHTHALMOLOGY, V108, P426, DOI 10.1016/S0161-6420(00)00401-2
   Chapuy B, 2018, NAT MED, V24, P679, DOI 10.1038/s41591-018-0016-8
   Chapuy B, 2016, BLOOD, V127, P869, DOI 10.1182/blood-2015-10-673236
   Coupland SE, 2008, CLIN EXP OPHTHALMOL, V36, P564, DOI 10.1111/j.1442-9071.2008.01843.x
   Coupland SE, 2009, OCUL IMMUNOL INFLAMM, V17, P227, DOI 10.1080/09273940903168696
   Coupland SE, 2005, INVEST OPHTH VIS SCI, V46, P3507, DOI 10.1167/iovs.05-0401
   Fend F, 2016, BRIT J HAEMATOL, V173, P680, DOI 10.1111/bjh.14025
   Gonzalez-Aguilar A, 2012, CLIN CANCER RES, V18, P5203, DOI 10.1158/1078-0432.CCR-12-0845
   Grimm SA, 2007, ANN ONCOL, V18, P1851, DOI 10.1093/annonc/mdm340
   Hiemcke-Jiwa LS, 2018, JAMA OPHTHALMOL, V136, P1098, DOI 10.1001/jamaophthalmol.2018.2887
   Ho KG, 2019, CURR OPIN NEUROL, V32, P886, DOI 10.1097/WCO.0000000000000759
   Huang W, 2009, MOL CELL, V35, P48, DOI 10.1016/j.molcel.2009.05.023
   Karakawa A, 2018, BRIT J HAEMATOL, V183, P668, DOI 10.1111/bjh.15005
   Kraan W, 2014, LEUKEMIA, V28, P719, DOI 10.1038/leu.2013.348
   Kraan W, 2013, BLOOD CANCER J, V3, DOI 10.1038/bcj.2013.28
   Lee JN, 2020, HAEMATOLOGICA, V105, pE458, DOI 10.3324/haematol.2019.233783
   Montesinos-Rongen M, 2008, LEUKEMIA, V22, P400, DOI 10.1038/sj.leu.2405019
   Montesinos-Rongen M, 2004, BLOOD, V103, P1869, DOI 10.1182/blood-2003-05-1465
   Nakamura T, 2016, NEUROPATH APPL NEURO, V42, P279, DOI 10.1111/nan.12259
   Nayyar N, 2019, BLOOD ADV, V3, P375, DOI 10.1182/bloodadvances.2018027672
   Ou A, 2020, NEUROONCOL ADV, V2, DOI DOI 10.1093/NOAJNL/VDAA018
   Pulido Jose S, 2018, Int J Retina Vitreous, V4, P18, DOI 10.1186/s40942-018-0120-4
   Raja H, 2016, RETINA-J RET VIT DIS, V36, P624, DOI 10.1097/IAE.0000000000000996
   Rimelen V, 2019, ACTA NEUROPATHOL COM, V7, DOI 10.1186/s40478-019-0692-8
   Sagoo MS, 2014, SURV OPHTHALMOL, V59, P503, DOI 10.1016/j.survophthal.2013.12.001
   Salomao DR, 2013, JAMA OPHTHALMOL, V131, P1151, DOI 10.1001/jamaophthalmol.2013.334
   Schmitz R, 2018, NEW ENGL J MED, V378, P1396, DOI 10.1056/NEJMoa1801445
   Shi HM, 2021, OCUL IMMUNOL INFLAMM, V29, P448, DOI 10.1080/09273948.2019.1657903
   Sung CO, 2011, BLOOD, V117, P1291, DOI 10.1182/blood-2010-07-297861
   Swerdlow SH, 2017, WHO Classification of Tumours of Haematopoietic and Lymphoid Tissues, V2
   Tan WJ, 2019, BLOOD, V134, P709, DOI 10.1182/blood.2019000022
   Tijchon E, 2013, LEUKEMIA, V27, P541, DOI 10.1038/leu.2012.293
   van Dongen JJM, 2003, LEUKEMIA, V17, P2257, DOI 10.1038/sj.leu.2403202
   Venturutti L, 2020, BLOOD, V136, P2263, DOI 10.1182/blood.2020005857
   Venturutti L, 2020, CELL, V182, P297, DOI 10.1016/j.cell.2020.05.049
   Wang LD, 2014, CANCER SCI, V105, P592, DOI 10.1111/cas.12388
   Xu L, 2013, BLOOD, V121, P2051, DOI 10.1182/blood-2012-09-454355
   Yonese I, 2019, EUR J HAEMATOL, V102, P191, DOI 10.1111/ejh.13191
   Yuniati L, 2019, J CELL PHYSIOL, V234, P5379, DOI 10.1002/jcp.27407
NR 46
TC 24
Z9 24
U1 0
U2 7
PU ELSEVIER
PI AMSTERDAM
PA RADARWEG 29, 1043 NX AMSTERDAM, NETHERLANDS
SN 2473-9529
EI 2473-9537
J9 BLOOD ADV
JI Blood Adv.
PD MAR 8
PY 2022
VL 6
IS 5
BP 1598
EP 1607
DI 10.1182/bloodadvances.2021004212
EA MAR 2022
PG 10
WC Hematology
WE Science Citation Index Expanded (SCI-EXPANDED)
SC Hematology
GA 1I1WO
UT WOS:000797026400024
PM 34448823
OA Green Published, gold
DA 2024-06-03
ER

PT J
AU Yuan, Y
   Ding, TL
   Wang, S
   Chen, H
   Mao, Y
   Chen, T
AF Yuan, Yan
   Ding, Tianling
   Wang, Shu
   Chen, Hong
   Mao, Ying
   Chen, Tong
TI Current and emerging therapies for primary central nervous system
   lymphoma
SO BIOMARKER RESEARCH
LA English
DT Review
DE primary central nervous system lymphoma; diffuse large B-cell lymphoma;
   targeted therapy
ID PRIMARY CNS LYMPHOMA; WHOLE-BRAIN RADIOTHERAPY; BRUTON TYROSINE KINASE;
   HIGH-DOSE METHOTREXATE; TUMOR-NECROSIS-FACTOR; T-CELL THERAPY;
   CEREBROSPINAL-FLUID PHARMACOKINETICS; INTERNATIONAL EXTRANODAL LYMPHOMA;
   NON-HODGKIN-LYMPHOMA; QUALITY-OF-LIFE
AB Primary central nervous system (CNS) lymphoma (PCNSL) is a rare type of extranodal lymphoma exclusively involving the CNS at the onset, with diffuse large B-cell lymphoma (DLBCL) as the most common histological subtype. As PCNSL is a malignancy arising in an immune-privileged site, suboptimal delivery of systemic agents into tumor tissues results in poorer outcomes in PCNSL than in non-CNS DLBCLs. Commonly used regimens for PCNSL include high-dose methotrexate-based chemotherapy with rituximab for induction therapy and intensive chemotherapy followed by autologous hematopoietic stem cell transplantation or whole-brain radiotherapy for consolidation therapy. Targeted agents against the B-cell receptor signaling pathway, microenvironment immunomodulation and blood-brain barrier (BBB) permeabilization appear to be promising in treating refractory/relapsed patients. Chimeric antigen receptor-T cells (CAR-T cells) have been shown to penetrate the BBB as a potential tool to manipulate this disease entity while controlling CAR-T cell-related encephalopathy syndrome. Future approaches may stratify patients according to age, performance status, molecular biomarkers and cellular bioinformation. This review summarizes the current therapies and emerging agents in clinical development for PCNSL treatment.
C1 [Yuan, Yan; Ding, Tianling; Wang, Shu; Chen, Tong] Fudan Univ, Huashan Hosp, Dept Hematol, 12 Wulumuqi Middle Rd, Shanghai 200040, Peoples R China.
   [Chen, Hong] Fudan Univ, Huashan Hosp, Dept Pathol, 12 Wulumuqi Middle Rd, Shanghai 200040, Peoples R China.
   [Mao, Ying] Fudan Univ, Huashan Hosp, Dept Neurosurg, 12 Wulumuqi Middle Rd, Shanghai 200040, Peoples R China.
C3 Fudan University; Fudan University; Fudan University
RP Chen, T (corresponding author), Fudan Univ, Huashan Hosp, Dept Hematol, 12 Wulumuqi Middle Rd, Shanghai 200040, Peoples R China.; Mao, Y (corresponding author), Fudan Univ, Huashan Hosp, Dept Neurosurg, 12 Wulumuqi Middle Rd, Shanghai 200040, Peoples R China.
EM
RI Chen, Tong/HLV-7325-2023; liu, xq/JDW-2596-2023
OI Chen, Tong/0000-0001-7895-2077; Mao, Ying/0000-0001-8055-115X
FU National Natural & Science Foundation of China [81870081]; Program for
   Outstanding Medical Academic Leader of Shanghai [2019LJ05]; Shanghai
   Collaborative Innovation Program on Regenerative Medicine and Stem Cell
   Research [2019CXJQ01]; Translational Research Grant of theNational
   Clinical Research Center for Hematologic Diseases [2020ZKMB03]
FX This project was supported by the National Natural & Science Foundation
   of China (No.81870081), Program for Outstanding Medical Academic Leader
   of Shanghai (No.2019LJ05), Shanghai Collaborative Innovation Program on
   Regenerative Medicine and Stem Cell Research (No.2019CXJQ01) and
   Translational Research Grant of the National Clinical Research Center
   for Hematologic Diseases (No.2020ZKMB03) to Tong Chen.
CR Abbasi A, 2020, J HEMATOL ONCOL, V13, DOI 10.1186/s13045-019-0838-y
   Abramson JS, 2020, LANCET, V396, P839, DOI 10.1016/S0140-6736(20)31366-0
   Abrey LE, 2003, J CLIN ONCOL, V21, P4151, DOI 10.1200/JCO.2003.05.024
   Alvarnas JC, 2000, BIOL BLOOD MARROW TR, V6, P352, DOI 10.1016/S1083-8791(00)70060-7
   Argyropoulos KV, 2018, HEMATOL ONCOL CLIN N, V32, P853, DOI 10.1016/j.hoc.2018.05.012
   Armand P, 2015, BLOOD, V125, P3393, DOI 10.1182/blood-2015-02-567453
   Baker SD, 1996, CANCER CHEMOTH PHARM, V37, P195
   Barrantes-Freer A, 2018, BRAIN PATHOL, V28, P225, DOI 10.1111/bpa.12496
   Batchelor T, 2003, J CLIN ONCOL, V21, P1044, DOI 10.1200/JCO.2003.03.036
   Bercusson A, 2018, BLOOD, V132, P1985, DOI 10.1182/blood-2017-12-823393
   Bond DA, 2020, HEMATOL ONCOL CLIN N, V34, P903, DOI 10.1016/j.hoc.2020.06.007
   Bromberg JEC, 2019, LANCET ONCOL, V20, P216, DOI 10.1016/S1470-2045(18)30747-2
   Brorson K, 1997, J IMMUNOL, V159, P135
   Brown JR, 2018, BLOOD, V131, P379, DOI [10.1182/blood-2017-08-764712, 10.1182/blood-2018-02-832071]
   Brudno JN, 2016, BLOOD, V127, P3321, DOI 10.1182/blood-2016-04-703751
   Chamberlain MC, 2010, NEURO-ONCOLOGY, V12, P736, DOI 10.1093/neuonc/noq011
   Chamoun K, 2017, NEUROLOGY, V88, P101, DOI 10.1212/WNL.0000000000003420
   Chapuy B, 2018, NAT MED, V24, P679, DOI 10.1038/s41591-018-0016-8
   Chapuy B, 2016, BLOOD, V127, P869, DOI 10.1182/blood-2015-10-673236
   Chen FL, 2020, CANCER MED-US, V9, P8676, DOI 10.1002/cam4.3499
   Chen LY, 2020, TRANSL ONCOL, V13, DOI 10.1016/j.tranon.2020.100838
   Cho H, 2017, ONCOTARGET, V8, P87317, DOI 10.18632/oncotarget.20264
   Crippa L, 2008, CANCER RES, V68, P1154, DOI 10.1158/0008-5472.CAN-07-2085
   Csordas K, 2013, ANTI-CANCER DRUG, V24, P189, DOI 10.1097/CAD.0b013e32835b8662
   Curnis F, 2000, NAT BIOTECHNOL, V18, P1185, DOI 10.1038/81183
   Curnis F, 2002, CANCER RES, V62, P867
   DEANGELIS LM, 1992, CANCER CHEMOTH PHARM, V29, P173, DOI 10.1007/BF00686248
   DeAngelis LM, 2002, J CLIN ONCOL, V20, P4643, DOI 10.1200/JCO.2002.11.013
   Deckert M, 2011, LEUKEMIA, V25, P1797, DOI 10.1038/leu.2011.169
   Dong HD, 1999, NAT MED, V5, P1365
   Dubovsky JA, 2013, BLOOD, V122, P2539, DOI 10.1182/blood-2013-06-507947
   EGORIN MJ, 1982, CLIN PHARMACOL THER, V32, P122, DOI 10.1038/clpt.1982.135
   El-Khouly FE, 2017, FRONT ONCOL, V7, DOI 10.3389/fonc.2017.00254
   Ferreri AJM, 2020, BLOOD ADV, V4, P3648, DOI 10.1182/bloodadvances.2020002270
   Ferreri AJM, 2019, BLOOD, V134, P252, DOI 10.1182/blood.2019000633
   Ferreri AJM, 2017, LANCET HAEMATOL, V4, pE510, DOI 10.1016/S2352-3026(17)30174-6
   Ferreri AJM, 2016, LANCET HAEMATOL, V3, pE217, DOI 10.1016/S2352-3026(16)00036-3
   Ferreri AJM, 2016, BLOOD, V127, P1642, DOI 10.1182/blood-2015-10-636340
   Ferreri AJM, 2011, BLOOD, V118, P510, DOI 10.1182/blood-2011-03-321349
   Ferreri AJM, 2009, LANCET, V374, P1512, DOI 10.1016/S0140-6736(09)61416-1
   Freeman GJ, 2000, J EXP MED, V192, P1027, DOI 10.1084/jem.192.7.1027
   Furuse M, 2020, BMC CANCER, V20, DOI 10.1186/s12885-020-06755-y
   Gavrilovic IT, 2006, J CLIN ONCOL, V24, P4570, DOI 10.1200/JCO.2006.06.6910
   Ghesquieres H, 2019, ANN ONCOL, V30, P621, DOI 10.1093/annonc/mdz032
   GINSBERG S, 1981, CANCER TREAT REP, V65, P477
   Goldwirt L, 2018, CANCER CHEMOTH PHARM, V81, P783, DOI 10.1007/s00280-018-3546-3
   Goldwirt L, 2013, BIOMED CHROMATOGR, V27, P889, DOI 10.1002/bmc.2877
   Grommes C, 2019, BLOOD, V133, P436, DOI 10.1182/blood-2018-09-875732
   Grommes C, 2017, CANCER DISCOV, V7, P1018, DOI 10.1158/2159-8290.CD-17-0613
   Harjunpää A, 2001, LEUKEMIA LYMPHOMA, V42, P731, DOI 10.3109/10428190109099335
   HEIDEMAN RL, 1989, CANCER RES, V49, P736
   Herrlinger U, 2017, J CANCER RES CLIN, V143, P1815, DOI 10.1007/s00432-017-2423-5
   Hiemcke-Jiwa LS, 2018, CRIT REV ONCOL HEMAT, V127, P56, DOI 10.1016/j.critrevonc.2018.05.010
   Honigberg LA, 2010, P NATL ACAD SCI USA, V107, P13075, DOI 10.1073/pnas.1004594107
   Houillier C, 2019, J CLIN ONCOL, V37, P823, DOI 10.1200/JCO.18.00306
   Hu YX, 2016, J HEMATOL ONCOL, V9, DOI 10.1186/s13045-016-0299-5
   Illerhaus G, 2006, J CLIN ONCOL, V24, P3865, DOI 10.1200/JCO.2006.06.2117
   ISHIDA Y, 1992, EMBO J, V11, P3887, DOI 10.1002/j.1460-2075.1992.tb05481.x
   JACKSON DV, 1981, CANCER RES, V41, P1466
   Kang LQ, 2020, EXP HEMATOL ONCOL, V9, DOI 10.1186/s40164-020-00166-2
   Kasenda B, 2016, BLOOD, V128, P32, DOI 10.1182/blood-2016-01-650101
   Kepka L, 2018, CLIN TRANSL ONCOL, V20, P150, DOI 10.1007/s12094-017-1703-5
   Vu K, 2019, BRIT J HAEMATOL, V186, P180, DOI 10.1111/bjh.15787
   Kim S, 2019, ONCOIMMUNOLOGY, V8, DOI 10.1080/2162402X.2019.1626653
   Kochenderfer JN, 2013, NAT REV CLIN ONCOL, V10, P267, DOI 10.1038/nrclinonc.2013.46
   Kumthekar P, 2013, J NEURO-ONCOL, V112, P247, DOI 10.1007/s11060-013-1055-0
   Lapointe S, 2018, LANCET, V392, P432, DOI 10.1016/S0140-6736(18)30990-5
   Latchman Y, 2001, NAT IMMUNOL, V2, P261, DOI 10.1038/85330
   Li ZM, 2013, PLOS ONE, V8, DOI 10.1371/journal.pone.0071754
   Lionakis MS, 2017, CANCER CELL, V31, P833, DOI 10.1016/j.ccell.2017.04.012
   Mead GM, 2000, CANCER, V89, P1359, DOI 10.1002/1097-0142(20000915)89:6<1359::AID-CNCR21>3.0.CO;2-9
   Morris PG, 2013, J CLIN ONCOL, V31, P3971, DOI 10.1200/JCO.2013.50.4910
   Muscal JA, 2012, CANCER CHEMOTH PHARM, V69, P943, DOI 10.1007/s00280-011-1781-y
   Narita Y, 2021, NEURO-ONCOLOGY, V23, P122, DOI 10.1093/neuonc/noaa145
   Nayak L, 2019, BLOOD, V134, P221, DOI 10.1182/blood.2019001383
   Nayak L, 2017, BLOOD, V129, P3071, DOI 10.1182/blood-2017-01-764209
   Neelapu SS, 2018, NAT REV CLIN ONCOL, V15, P47, DOI 10.1038/nrclinonc.2017.148
   Norelli M, 2018, NAT MED, V24, P739, DOI 10.1038/s41591-018-0036-4
   Olson JE, 2002, CANCER, V95, P1504, DOI 10.1002/cncr.10851
   Omuro A, 2015, LANCET HAEMATOL, V2, pE251, DOI 10.1016/S2352-3026(15)00074-5
   Omuro A, 2015, BLOOD, V125, P1403, DOI 10.1182/blood-2014-10-604561
   Papich M.G., 2016, Saunders Handbook of Veterinary Drugs: Small and Large Animal, V4th ed., P110
   Parker KR, 2020, CELL, V183, P126, DOI 10.1016/j.cell.2020.08.022
   Petereit HF, 2009, MULT SCLER J, V15, P189, DOI 10.1177/1352458508098268
   Ponader S, 2012, BLOOD, V119, P1182, DOI 10.1182/blood-2011-10-386417
   Porter DL, 2011, NEW ENGL J MED, V365, P725, DOI 10.1056/NEJMoa1103849
   Portnow J, 2009, CLIN CANCER RES, V15, P7092, DOI 10.1158/1078-0432.CCR-09-1349
   Prica A, 2012, BRIT J HAEMATOL, V158, P600, DOI 10.1111/j.1365-2141.2012.09208.x
   Rubenstein JL, 2018, BLOOD ADV, V2, P1595, DOI 10.1182/bloodadvances.2017014845
   Rubenstein JL, 2013, J CLIN ONCOL, V31, P3061, DOI 10.1200/JCO.2012.46.9957
   Rubenstein JL, 2003, BLOOD, V101, P466, DOI 10.1182/blood-2002-06-1636
   Sacchi A, 2006, CLIN CANCER RES, V12, P175, DOI 10.1158/1078-0432.CCR-05-1147
   Sacchi A, 2004, CANCER RES, V64, P7150, DOI 10.1158/0008-5472.CAN-04-1445
   Sagoo MS, 2014, SURV OPHTHALMOL, V59, P503, DOI 10.1016/j.survophthal.2013.12.001
   Satterthwaite AB, 1998, J EXP MED, V188, P833, DOI 10.1084/jem.188.5.833
   Schmidt NW, 2006, J IMMUNOL, V177, P7203, DOI 10.4049/jimmunol.177.10.7203
   Schmitz N, 2015, BLOOD, V125, P1360, DOI 10.1182/blood-2015-01-622498
   Schmitz R, 2018, NEW ENGL J MED, V378, P1396, DOI 10.1056/NEJMoa1801445
   Shah GD, 2007, J CLIN ONCOL, V25, P4730, DOI 10.1200/JCO.2007.12.5062
   Shalabi H, 2018, J IMMUNOTHER, V41, P350, DOI 10.1097/CJI.0000000000000241
   Siegal T, 2019, ACTA HAEMATOL-BASEL, V141, P138, DOI 10.1159/000495284
   SLEVIN ML, 1983, J CLIN ONCOL, V1, P546, DOI 10.1200/JCO.1983.1.9.546
   Soussain C, 2019, EUR J CANCER, V117, P121, DOI 10.1016/j.ejca.2019.05.024
   Soussain C, 2008, J CLIN ONCOL, V26, P2512, DOI 10.1200/JCO.2007.13.5533
   STRONG JM, 1986, CANCER RES, V46, P6101
   Sugita Y, 2016, NEUROPATHOLOGY, V36, P313, DOI 10.1111/neup.12276
   Sukswai N, 2020, PATHOLOGY, V52, P53, DOI 10.1016/j.pathol.2019.08.013
   Swerdlow SH, 2016, BLOOD, V127, P2375, DOI 10.1182/blood-2016-01-643569
   T Low Justin, 2020, CNS Oncol, V9, pCNS51, DOI 10.2217/cns-2019-0022
   Taraseviciute A, 2018, CANCER DISCOV, V8, P750, DOI 10.1158/2159-8290.CD-17-1368
   Terziev D, 2018, EUR J HAEMATOL, V101, P115, DOI 10.1111/ejh.13072
   Tetef ML, 2000, CANCER CHEMOTH PHARM, V46, P19, DOI 10.1007/s002800000118
   Thiel E, 2010, LANCET ONCOL, V11, P1036, DOI 10.1016/S1470-2045(10)70229-1
   Tillman BF, 2018, EUR J HAEMATOL, V100, P325, DOI 10.1111/ejh.13020
   Torre M, 2018, J NEUROPATH EXP NEUR, V77, P877, DOI 10.1093/jnen/nly064
   Treon SP, 2015, NEW ENGL J MED, V372, P1430, DOI 10.1056/NEJMoa1501548
   Tu SF, 2019, FRONT ONCOL, V9, DOI 10.3389/fonc.2019.01350
   Tun HW, 2018, BLOOD, V132, P2240, DOI 10.1182/blood-2018-02-835496
   van Laarhoven HWM, 2006, INVEST NEW DRUG, V24, P27, DOI 10.1007/s10637-005-4540-2
   VASSAL G, 1989, CANCER CHEMOTH PHARM, V24, P386
   Vassilev A, 1999, J BIOL CHEM, V274, P1646, DOI 10.1074/jbc.274.3.1646
   Wang ML, 2013, NEW ENGL J MED, V369, P507, DOI 10.1056/NEJMoa1306220
   Wilson WH, 2015, NAT MED, V21, P922, DOI 10.1038/nm.3884
   Woyach JA, 2018, NEW ENGL J MED, V379, P2517, DOI 10.1056/NEJMoa1812836
   Wu JJ, 2018, J NEURO-ONCOL, V140, P427, DOI 10.1007/s11060-018-2970-x
   Wu JJ, 2016, J HEMATOL ONCOL, V9, DOI 10.1186/s13045-016-0313-y
   Wu L, 2008, CLIN CANCER RES, V14, P4650, DOI 10.1158/1078-0432.CCR-07-4405
   Yang YB, 2012, CANCER CELL, V21, P723, DOI 10.1016/j.ccr.2012.05.024
   Zhou QY, 2007, CLIN CANCER RES, V13, P4271, DOI 10.1158/1078-0432.CCR-07-0658
   Zhu YX, 2011, BLOOD, V118, P4771, DOI 10.1182/blood-2011-05-356063
   ZUCCHETTI M, 1991, ANN ONCOL, V2, P63, DOI 10.1093/oxfordjournals.annonc.a057826
NR 131
TC 24
Z9 25
U1 0
U2 16
PU BMC
PI LONDON
PA CAMPUS, 4 CRINAN ST, LONDON N1 9XW, ENGLAND
EI 2050-7771
J9 BIOMARK RES
JI Biomark. Res.
PD MAY 6
PY 2021
VL 9
IS 1
AR 32
DI 10.1186/s40364-021-00282-z
PG 15
WC Oncology; Medicine, Research & Experimental
WE Science Citation Index Expanded (SCI-EXPANDED)
SC Oncology; Research & Experimental Medicine
GA RY2LA
UT WOS:000647747600001
PM 33957995
OA Green Published, gold
DA 2024-06-03
ER

PT J
AU Chen, CY
   Zheng, AP
   Ou, XJ
   Wang, J
   Ma, XL
AF Chen, Chaoyue
   Zheng, Aiping
   Ou, Xuejin
   Wang, Jian
   Ma, Xuelei
TI Comparison of Radiomics-Based Machine-Learning Classifiers in Diagnosis
   of Glioblastoma From Primary Central Nervous System Lymphoma
SO FRONTIERS IN ONCOLOGY
LA English
DT Article
DE glioblastoma; primary central nervous system lymphoma; magnetic
   resonance imaging; radiomics; machine learning
ID DIFFERENTIATION; MRI; DIFFUSION; PERFUSION; BRAIN
AB Purpose:The purpose of the current study was to evaluate the ability of magnetic resonance (MR) radiomics-based machine-learning algorithms in differentiating glioblastoma (GBM) from primary central nervous system lymphoma (PCNSL). Method:One-hundred and thirty-eight patients were enrolled in this study. Radiomics features were extracted from contrast-enhanced MR images, and the machine-learning models were established using five selection methods (distance correlation, random forest, least absolute shrinkage and selection operator (LASSO), eXtreme gradient boosting (Xgboost), and Gradient Boosting Decision Tree) and three radiomics-based machine-learning classifiers [linear discriminant analysis (LDA), support vector machine (SVM), and logistic regression (LR)]. Sensitivity, specificity, accuracy, and areas under curves (AUC) of models were calculated, with which the performances of classifiers were evaluated and compared with each other. Result:Brilliant discriminative performance would be observed among all classifiers when combined with the suitable selection method. For LDA-based models, the optimal one was Distance Correlation + LDA with AUC of 0.978. For SVM-based models, Distance Correlation + SVM was the one with highest AUC of 0.959, while for LR-based models, the highest AUC was 0.966 established with LASSO + LR. Conclusion:Radiomics-based machine-learning algorithms potentially have promising performances in differentiating GBM from PCNSL.
C1 [Chen, Chaoyue; Ou, Xuejin; Ma, Xuelei] Sichuan Univ, West China Hosp, Collaborat Innovat Ctr Biotherapy, State Key Lab Biotherapy & Canc Ctr, Chengdu, Peoples R China.
   [Chen, Chaoyue] Sichuan Univ, West China Hosp, Dept Neurosurg, Chengdu, Peoples R China.
   [Zheng, Aiping; Ou, Xuejin] Sichuan Univ, West China Hosp, West China Sch Med, Chengdu, Peoples R China.
   [Wang, Jian] Nanjing Univ Sci & Technol, Sch Comp Sci, Nanjing, Peoples R China.
   [Ma, Xuelei] Sichuan Univ, Canc Ctr West China Hosp, Dept Biotherapy, Chengdu, Peoples R China.
C3 Sichuan University; Sichuan University; Sichuan University; Nanjing
   University of Science & Technology; Sichuan University
RP Ma, XL (corresponding author), Sichuan Univ, West China Hosp, Collaborat Innovat Ctr Biotherapy, State Key Lab Biotherapy & Canc Ctr, Chengdu, Peoples R China.; Ma, XL (corresponding author), Sichuan Univ, Canc Ctr West China Hosp, Dept Biotherapy, Chengdu, Peoples R China.
EM
CR Al-Okaili RN, 2007, RADIOLOGY, V243, P539, DOI 10.1148/radiol.2432060493
   Alcaide-Leon P, 2017, AM J NEURORADIOL, V38, P1145, DOI 10.3174/ajnr.A5173
   Castellano G, 2004, CLIN RADIOL, V59, P1061, DOI 10.1016/j.crad.2004.07.008
   Choi YS, 2017, EUR RADIOL, V27, P1344, DOI 10.1007/s00330-016-4484-2
   Ditmer A, 2018, J NEURO-ONCOL, V140, P583, DOI 10.1007/s11060-018-2984-4
   Dolecek TA, 2012, NEURO-ONCOLOGY, V14, pv1, DOI 10.1093/neuonc/nos218
   Guo AC, 2002, RADIOLOGY, V224, P177, DOI 10.1148/radiol.2241010637
   Kickingereder P, 2014, AM J NEURORADIOL, V35, P1503, DOI 10.3174/ajnr.A3915
   Kickingereder P, 2014, RADIOLOGY, V272, P843, DOI 10.1148/radiol.14132740
   Ko SY, 2019, HEAD NECK-J SCI SPEC, V41, P885, DOI 10.1002/hed.25415
   Koeller KK, 1997, RADIOGRAPHICS, V17, P1497, DOI 10.1148/radiographics.17.6.9397461
   Lee JH, 2019, EUR RADIOL, V29, P5452, DOI 10.1007/s00330-019-06098-8
   Li YM, 2018, EUR RADIOL, V28, P2960, DOI 10.1007/s00330-017-5267-0
   Malikova H, 2016, CANCER IMAGING, V16, DOI 10.1186/s40644-016-0098-9
   Nioche C, 2018, CANCER RES, V78, P4786, DOI 10.1158/0008-5472.CAN-18-0125
   Radbruch A, 2013, EUR J RADIOL, V82, P552, DOI 10.1016/j.ejrad.2012.11.002
   Scalco E, 2017, BRIT J RADIOL, V90, DOI 10.1259/bjr.20160642
   Schlegel Uwe, 2009, Ther Adv Neurol Disord, V2, P93, DOI 10.1177/1756285608101222
   Suh HB, 2018, EUR RADIOL, V28, P3832, DOI 10.1007/s00330-018-5368-4
   Toh CH, 2008, AM J NEURORADIOL, V29, P471, DOI 10.3174/ajnr.A0872
   Toh CH, 2013, AM J NEURORADIOL, V34, P1145, DOI 10.3174/ajnr.A3383
   van IJzendoorn DGP, 2019, PLOS COMPUT BIOL, V15, DOI 10.1371/journal.pcbi.1006826
   von Baumgarten L, 2018, DTSCH ARZTEBL INT, V115, P419, DOI 10.3238/arztebl.2018.0419
   Xie JY, 2019, FRONT GENET, V10, DOI 10.3389/fgene.2019.00080
   Zhou H, 2017, NEURO-ONCOLOGY, V19, P862, DOI 10.1093/neuonc/now256
NR 25
TC 24
Z9 28
U1 2
U2 26
PU FRONTIERS MEDIA SA
PI LAUSANNE
PA AVENUE DU TRIBUNAL FEDERAL 34, LAUSANNE, CH-1015, SWITZERLAND
SN 2234-943X
J9 FRONT ONCOL
JI Front. Oncol.
PD SEP 15
PY 2020
VL 10
AR 1151
DI 10.3389/fonc.2020.01151
PG 7
WC Oncology
WE Science Citation Index Expanded (SCI-EXPANDED)
SC Oncology
GA NY1RK
UT WOS:000576175300001
PM 33042784
OA Green Published, gold
DA 2024-06-03
ER

PT J
AU Velasco, R
   Mercadal, S
   Vidal, N
   Alañá, M
   Barcelo, MI
   Ibáñez-Juliá, MJ
   Bobillo, S
   Agud, RC
   Molina, EG
   Martínez, P
   Cacabelos, P
   Muntañola, A
   García-Catalán, G
   Sancho, JM
   Camro, I
   Lado, T
   Erro, ME
   Gómez-Vicente, L
   Salar, A
   Caballero, AC
   Solé-Rodríguez, M
   Pérez-Larraya, JG
   Huertas, N
   Estela, J
   Barón, M
   Barbero-Bordallo, N
   Encuentra, M
   Dlouhy, I
   Bruna, J
   Graus, F
AF Velasco, R.
   Mercadal, S.
   Vidal, N.
   Alana, M.
   Barcelo, M., I
   Ibanez-Julia, M. J.
   Bobillo, S.
   Caldu Agud, R.
   Garcia Molina, E.
   Martinez, P.
   Cacabelos, P.
   Muntanola, A.
   Garcia-Catalan, G.
   Sancho, J. M.
   Camro, I
   Lado, T.
   Erro, M. E.
   Gomez-Vicente, L.
   Salar, A.
   Caballero, A. C.
   Sole-Rodriguez, M.
   Gallego Perez-Larraya, J.
   Huertas, N.
   Estela, J.
   Baron, M.
   Barbero-Bordallo, N.
   Encuentra, M.
   Dlouhy, I
   Bruna, J.
   Graus, F.
CA GELTAMO & GENOSEN Grp
TI Diagnostic delay and outcome in immunocompetent patients with primary
   central nervous system lymphoma in Spain: a multicentric study
SO JOURNAL OF NEURO-ONCOLOGY
LA English
DT Article
DE Primary central nervous system lymphoma; Diagnostic delay; Steroids;
   Prognostic factors; Outcome
ID PRIMARY CNS LYMPHOMA; INTERNATIONAL EXTRANODAL LYMPHOMA; INCREASING
   INCIDENCE; UNITED-STATES; MRI FEATURES; LONG-TERM; SURVIVAL;
   GLIOBLASTOMA; EXPERIENCE; PATTERNS
AB Introduction To assess the management of immunocompetent patients with primary central nervous system lymphomas (PCNSL) in Spain. Methods Retrospective analysis of 327 immunocompetent patients with histologically confirmed PCNSL diagnosed between 2005 and 2014 in 27 Spanish hospitals. Results Median age was 64 years (range: 19-84; 33% >= 70 years), 54% were men, and 59% had a performance status (PS) >= 2 at diagnosis. Median delay to diagnosis was 47 days (IQR 24-81). Diagnostic delay > 47 days was associated with PS >= 2 (OR 1.99; 95% CI 1.13-3.50; p = 0.016) and treatment with corticosteroids (OR 2.47; 95% CI 1.14-5.40; p = 0.023), and it did not improve over the years. Patients treated with corticosteroids (62%) had a higher risk of additional biopsies (11.7% vs 4.0%, p = 0.04) but corticosteroids withdrawal before surgery did not reduce this risk and increased the diagnostic delay (64 vs 40 days, p = 0.04). Median overall survival (OS) was 8.9 months [95% CI 5.9-11.7] for the whole series, including 52 (16%) patients that were not treated, and 14.1 months (95%CI 7.7-20.5) for the 240 (73.4%) patients that received high-dose methotrexate (HD-MTX)-based chemotherapy. Median OS was shorter in patients >= 70 years (4.1 vs. 13.4 months; p < 0.0001). Multivariate analysis identified age >= 65 years, PS >= 2, no treatment, and cognitive/psychiatric symptoms at diagnosis as independent predictors of short survival. Conclusions Corticosteroids withdrawal before surgery does not decrease the risk of a negative biopsy but delays diagnosis. In this community-based study, only 73.4% of patients could receive HD-MTX-based chemotherapy and OS remains poor, particularly in elderly patients >= 70 years.
C1 [Velasco, R.; Vidal, N.; Bruna, J.] Hosp Univ Bellvitge ICO Duran i Reynals, Neurooncol Unit, IDIBELL, Barcelona, Spain.
   [Velasco, R.; Bruna, J.] Univ Autonoma Barcelona, Inst Neurosci, Dept Cell Biol Physiol & Immunol, Bellaterra, Spain.
   [Velasco, R.; Bruna, J.] Ctr Invest Biomed Red Enfermedades Neurodegenerat, Bellaterra, Spain.
   [Mercadal, S.; Encuentra, M.] Univ Barcelona, Dept Hematol, Catalan Inst Oncol, Hosp Duran i Reynals,IDIBELL, Barcelona, Spain.
   [Vidal, N.] Hosp Univ Bellvitge, Dept Pathol, ICO Duran i Reynals, IDIBELL,Unit Neurooncol, Barcelona, Spain.
   [Alana, M.] Complejo Asistencial Univ, Dept Neurol, Salamanca, Spain.
   [Barcelo, M., I] Hosp Univ Son Espases, Dept Neurol, Palma De Mallorca, Spain.
   [Ibanez-Julia, M. J.] Hosp Univ & Politecn La Fe, Dept Neurol, Valencia, Spain.
   [Bobillo, S.] Univ Hosp Vall dHebron, Vall dHebron Inst Oncol VHIO, Dept Hematol, Barcelona, Spain.
   [Caldu Agud, R.] Hosp Univ Miguel Servet, Dept Neurol, Zaragoza, Spain.
   [Garcia Molina, E.] Virgen Arrixaca Univ Hosp, Dept Neurol, Murcia, Spain.
   [Martinez, P.] Hosp Univ 12 Octubre, Dept Hematol, Madrid, Spain.
   [Cacabelos, P.] Hosp Clin Univ Santiago de Compostela, Dept Neurol, Santiago De Compostela, La Coruna, Spain.
   [Muntanola, A.] Hosp Univ Mutua Terrassa, Dept Hematol, Terrassa, Spain.
   [Garcia-Catalan, G.] Hosp Univ Marques de Valdecilla, Dept Neurosurg, Santander, Spain.
   [Sancho, J. M.] ICO ITC Hosp Germans Trias & Pujol, Dept Hematol, Badalona, Spain.
   [Camro, I] Hosp Univ Cruces, Dept Neurol, Vizcaya, Spain.
   [Lado, T.] Hosp Clin Univ Valladolid, Dept Hematol, Valladolid, Spain.
   [Erro, M. E.] Complejo Hosp Navarra, Navarra Inst Hlth Res IdiSNA, Neurol Dept, Pamplona, Spain.
   [Gomez-Vicente, L.] Hosp Univ Quironsalud Madrid, Dept Neurol, Madrid, Spain.
   [Salar, A.] Hosp del Mar, Dept Hematol, Barcelona, Spain.
   [Caballero, A. C.] Hosp Santa Creu & Sant Pau, Dept Hematol, Barcelona, Spain.
   [Sole-Rodriguez, M.] Hosp Univ Virgen del Rocio, Dept Hematol, Seville, Spain.
   [Gallego Perez-Larraya, J.] Inst Invest Sanitaria Navarra, Clin Univ Navarra, Dept Neurol, IdiSNA, Pamplona, Spain.
   [Huertas, N.] Hosp Univ Severo Ochoa, Dept Neurol, Madrid, Spain.
   [Estela, J.] Hosp Parc Tauli, Dept Neurol, Sabadell, Spain.
   [Baron, M.] Hosp Univ Fdn Alcorcon, Unit Neurol, Madrid, Spain.
   [Barbero-Bordallo, N.] Hosp Univ Rey Juan Carlos, Dept Neurol, HURJC HUIE HCV, Madrid, Spain.
   [Dlouhy, I; Graus, F.] Hosp Clin Barcelona, Dept Hematol & Neurol, Barcelona, Spain.
   [Velasco, R.] Hosp Univ Bellvitge, Neurooncol Unit, Dept Neurol, C Feixa Llarga S-N, Barcelona 08907, Spain.
   [Velasco, R.] ICO Hosp, C Feixa Llarga S-N, Barcelona 08907, Spain.
C3 Institut d'Investigacio Biomedica de Bellvitge (IDIBELL); Hospital Duran
   i Reynals; Autonomous University of Barcelona; CIBERNED; University of
   Barcelona; Institut d'Investigacio Biomedica de Bellvitge (IDIBELL);
   Hospital Duran i Reynals; Institut Catala d'Oncologia; Institut
   d'Investigacio Biomedica de Bellvitge (IDIBELL); Hospital Duran i
   Reynals; Bellvitge University Hospital; University of Barcelona;
   Hospital Universitari Son Espases; Hospital Universitari i Politecnic La
   Fe; Hospital Universitari Vall d'Hebron; Vall d'Hebron Institut
   d'Oncologia (VHIO); Miguel Servet University Hospital; Hospital Clinico
   Universitario Virgen de la Arrixaca; Hospital Universitario 12 de
   Octubre; Complexo Hospitalario Universitario de Santiago de Compostela;
   Hospital Universitario Mutua Terrassa; Hospital Universitario Marques de
   Valdecilla (HUMV); Hospital Universitario Cruces; Universidad de
   Valladolid; Servicio Navarro de Salud - Osasunbidea; quironsalud Group;
   Hospital del Mar Research Institute; Hospital del Mar; Hospital of Santa
   Creu i Sant Pau; Virgen del Rocio University Hospital; University of
   Navarra; Severo Ochoa University Hospital; Autonomous University of
   Barcelona; Parc Tauli Hospital Universitari; Alcorcon Foundation
   University Hospital; University of Barcelona; Hospital Clinic de
   Barcelona; Institut d'Investigacio Biomedica de Bellvitge (IDIBELL);
   Bellvitge University Hospital; University of Barcelona
RP Velasco, R (corresponding author), Hosp Univ Bellvitge ICO Duran i Reynals, Neurooncol Unit, IDIBELL, Barcelona, Spain.; Velasco, R (corresponding author), Univ Autonoma Barcelona, Inst Neurosci, Dept Cell Biol Physiol & Immunol, Bellaterra, Spain.; Velasco, R (corresponding author), Ctr Invest Biomed Red Enfermedades Neurodegenerat, Bellaterra, Spain.; Velasco, R (corresponding author), Hosp Univ Bellvitge, Neurooncol Unit, Dept Neurol, C Feixa Llarga S-N, Barcelona 08907, Spain.; Velasco, R (corresponding author), ICO Hosp, C Feixa Llarga S-N, Barcelona 08907, Spain.
EM
RI Sancho, Juan Manuel/AGB-5076-2022; Martín García-Sancho,
   Alejandro/B-8611-2018; Salar, Antonio/HJY-3106-2023; Caballero
   GonzÃ¡lez, Ana Carolina/JPY-0936-2023; Graus, Francesc/K-9904-2018
OI Sancho, Juan Manuel/0000-0001-7168-6538; Martín García-Sancho,
   Alejandro/0000-0001-6330-1028; CABALLERO GONZALEZ, ANA
   CAROLINA/0000-0002-4950-5259; Climent, Fina/0000-0002-4360-8388;
   Abrisqueta, Pau/0000-0001-9625-7422; Vidal, Noemi/0000-0002-2852-6667
FU ISCIII [PI1501303]; Fondo Europeo de Desarrollo Regional (FEDER)
FX This work was partially supported by Grant PI1501303 from ISCIII and
   Fondo Europeo de Desarrollo Regional (FEDER).
CR Abrey LE, 2006, J CLIN ONCOL, V24, P5711, DOI 10.1200/JCO.2006.08.2941
   Abrey LE, 1998, J CLIN ONCOL, V16, P859, DOI 10.1200/JCO.1998.16.3.859
   Bessell EM, 1996, J CLIN ONCOL, V14, P945, DOI 10.1200/JCO.1996.14.3.945
   Binnahil M, 2016, CAN J NEUROL SCI, V43, P721, DOI 10.1017/cjn.2016.255
   Brück W, 2013, PATHOLOGE, V34, P186, DOI 10.1007/s00292-013-1742-9
   Bühring U, 2001, NEUROLOGY, V57, P393, DOI 10.1212/WNL.57.3.393
   Burton EC, 2017, CUREUS J MED SCIENCE, V9, DOI 10.7759/cureus.1512
   Campo E, 2011, BLOOD, V117, P5019, DOI 10.1182/blood-2011-01-293050
   Cerqua R, 2016, NEUROL SCI, V37, P23, DOI 10.1007/s10072-015-2353-4
   Chang SM, 2005, JAMA-J AM MED ASSOC, V293, P557, DOI 10.1001/jama.293.5.557
   Correa DD, 2019, J NEURO-ONCOL, V144, P553, DOI 10.1007/s11060-019-03257-1
   DeAngelis LM, 1999, J NEUROL NEUROSUR PS, V66, P699, DOI 10.1136/jnnp.66.6.699
   Eloranta S, 2018, EUR J HAEMATOL, V100, P61, DOI 10.1111/ejh.12980
   Enblad G, 2017, ACTA ONCOL, V56, P599, DOI 10.1080/0284186X.2016.1270465
   Fabbro-Peray P, 2019, J NEURO-ONCOL, V142, P91, DOI 10.1007/s11060-018-03065-z
   Fallah J, 2016, BLOOD ADV, V1, P112, DOI 10.1182/bloodadvances.2016000927
   Ferreri AJM, 2003, J CLIN ONCOL, V21, P266, DOI 10.1200/JCO.2003.09.139
   Ferreri AJM, 2017, LANCET HAEMATOL, V4, pE510, DOI 10.1016/S2352-3026(17)30174-6
   Graus F, 2013, NEURO-ONCOLOGY, V15, P797, DOI 10.1093/neuonc/not013
   Grommes C, 2017, J CLIN ONCOL, V35, P2410, DOI 10.1200/JCO.2017.72.7602
   Haldorsen IS, 2009, AM J NEURORADIOL, V30, P744, DOI 10.3174/ajnr.A1447
   Haldorsen IS, 2007, CANCER, V110, P1803, DOI 10.1002/cncr.22989
   Haldorsen IS, 2005, ACTA ONCOL, V44, P728, DOI 10.1080/02841860500256272
   Hart A, 2014, NETH J MED, V72, P218
   Hoang-Xuan K, 2015, LANCET ONCOL, V16, pE322, DOI 10.1016/S1470-2045(15)00076-5
   Hoffman S, 2006, NEURO-ONCOLOGY, V8, P27, DOI 10.1215/S1522851705000323
   Houillier C, 2020, NEUROLOGY, V94, pE1027, DOI 10.1212/WNL.0000000000008900
   Houillier C, 2019, J CLIN ONCOL, V37, P823, DOI 10.1200/JCO.18.00306
   Izquierdo C, 2016, NEURO-ONCOLOGY, V18, P707, DOI 10.1093/neuonc/nov197
   Kasenda B, 2015, ANN ONCOL, V26, P1305, DOI 10.1093/annonc/mdv076
   Kiewe P, 2008, CANCER-AM CANCER SOC, V112, P1812, DOI 10.1002/cncr.23377
   Küker W, 2005, J NEURO-ONCOL, V72, P169, DOI 10.1007/s11060-004-3390-7
   Mendez JS, 2018, NEURO-ONCOLOGY, V20, P687, DOI 10.1093/neuonc/nox187
   Porter AB, 2008, ANN NEUROL, V63, P662, DOI 10.1002/ana.21366
   Shan YD, 2018, DIS MARKERS, V2018, DOI 10.1155/2018/7860494
   Siegal T, 2019, ACTA HAEMATOL-BASEL, V141, P138, DOI 10.1159/000495284
   van der Meulen M, 2017, LEUKEMIA, V31, P1822, DOI 10.1038/leu.2017.128
   van der Meulen M, 2018, LANCET ONCOL, V19, pE407, DOI 10.1016/S1470-2045(18)30356-5
   Weller M, 1999, J NEURO-ONCOL, V43, P237, DOI 10.1023/A:1006254518848
   Xie H, 2015, AM J CLIN ONCOL-CANC, V38, P140, DOI 10.1097/COC.0b013e31828f5a26
NR 40
TC 24
Z9 25
U1 0
U2 6
PU SPRINGER
PI NEW YORK
PA ONE NEW YORK PLAZA, SUITE 4600, NEW YORK, NY, UNITED STATES
SN 0167-594X
EI 1573-7373
J9 J NEURO-ONCOL
JI J. Neuro-Oncol.
PD JUL
PY 2020
VL 148
IS 3
BP 545
EP 554
DI 10.1007/s11060-020-03547-z
EA JUN 2020
PG 10
WC Oncology; Clinical Neurology
WE Science Citation Index Expanded (SCI-EXPANDED)
SC Oncology; Neurosciences & Neurology
GA ND8OJ
UT WOS:000539507000001
PM 32524392
DA 2024-06-03
ER

PT J
AU Yu, H
   Gao, B
   Liu, J
   Yu, YC
   Shiroishi, MS
   Huang, MM
   Yang, WX
   Guan, ZZ
AF Yu, Hui
   Gao, Bo
   Liu, Jing
   Yu, Yong-Cheng
   Shiroishi, Mark S.
   Huang, Ming-Ming
   Yang, Wen-Xiu
   Guan, Zhi-Zhong
TI Lymphomatosis cerebri: a rare variant of primary central nervous system
   lymphoma and MR imaging features
SO CANCER IMAGING
LA English
DT Article
DE Lymphomatosis cerebri; Primary central nervous system lymphoma; Magnetic
   resonance imaging; Diffusion weighted imaging; Magnetic resonance
   spectroscopy
ID PROGRESSIVE DEMENTIA; LEUKOENCEPHALOPATHY
AB Background: Lymphomatosis cerebri (LC) is a rare variant of primary central nervous system lymphoma (PCNSL), characterized by diffuse infiltration without the formation of a discrete mass. The diagnosis of LC is a challenge because the imaging findings are atypical for lymphoma. The purpose of present study is to investigate MRI characteristics and clinical features of LC and potentially facilitate an early and accurate diagnosis of this oftenmissed disease.
   Methods: Seven patients (average 44 years, 19-58 years) with LC proved basing on MRI and histology were retrospectively reviewed the clinical data and cerebral MR imaging findings.
   Results: The common presenting symptoms were cognitive decline, behavioral disturbance, gait disturbance. All patients had both deep and lobar lesion distribution, and two of them had infratentorial involvement. Lack of contrast enhancement and subtle patchy enhanced pattern were observed in two and three patients, respectively. The remaining two patients presented multiple patchy enhancement. Most of the lesions were slightly hyperintense to normal brain on DWI as well as hyperintense on ADC maps. Three patients presented a pattern of marked decrease of NAA/Cr, increase of Cho/Cr, and two of the three cases showed increased Lip/Cr and Lac/Cr on MRS.
   Conclusions: We conclude that diffuse bilateral lesions especially in deep and lobar region including white and gray matter, without enhancement or with patchy enhancement, marked decrease of NAA/Cr and increase of Cho/Cr, and increased Lip/Cr and Lac/Cr are suggestive of LC. Prompt recognition of these imaging patterns may lead to early diagnosis of LC and brain biopsy with improved prognosis.
C1 [Yu, Hui; Liu, Jing; Huang, Ming-Ming] Guizhou Med Univ, Affiliated Hosp, Dept Radiol, Guiyang 550004, Peoples R China.
   [Gao, Bo] Yantai Yuhuangding Hosp, Dept Radiol, Yantai 264000, Shandong, Peoples R China.
   [Yu, Yong-Cheng] Guizhou Med Univ, Dept Neurol, Affiliated Hosp 2, Kaili 556000, Peoples R China.
   [Shiroishi, Mark S.] Univ Southern Calif, Keck Sch Med, Dept Radiol, Los Angeles, CA USA.
   [Yang, Wen-Xiu; Guan, Zhi-Zhong] Guizhou Med Univ, Affiliated Hosp, Dept Pathol, Guiyang 550004, Peoples R China.
C3 Guizhou Medical University; Guizhou Medical University; University of
   Southern California; Guizhou Medical University
RP Guan, ZZ (corresponding author), Guizhou Med Univ, Affiliated Hosp, Dept Pathol, Guiyang 550004, Peoples R China.
EM
RI Yang, Wenxiu/GLR-5145-2022
FU Chinese National Natural Science Foundation [GR-81260173]; Guizhou
   Provincial Education Department [KY[2016] 012]; SC CTSI (NIH/NCRR/NCATS)
   [GR-KL2TR000131]; NIH [1 L30 CA209248-01]
FX Collection, interpretation of data and writing manuscript were supported
   by Chinese National Natural Science Foundation GR-81260173, Guizhou
   Provincial Education Department KY[2016] 012, SC CTSI (NIH/NCRR/NCATS)
   GR-KL2TR000131 and NIH 1 L30 CA209248-01.
CR Bakshi R, 1999, DEMENT GERIATR COGN, V10, P152, DOI 10.1159/000017116
   Courtois F, 2011, CASE REP NEUROL, V4, P181, DOI 10.1159/000343947
   Rocha Antonio José da, 2016, Rev. Bras. Hematol. Hemoter., V38, P44, DOI 10.1016/j.bjhh.2015.12.001
   Filley CM, 2001, NEW ENGL J MED, V345, P425, DOI 10.1056/NEJM200108093450606
   Haldorsen IS, 2011, AM J NEURORADIOL, V32, P984, DOI 10.3174/ajnr.A2171
   Hatanpaa KJ, 2015, JAMA NEUROL, V72, P1066, DOI 10.1001/jamaneurol.2015.1149
   Izquierdo C, 2016, NEURO-ONCOLOGY, V18, P707, DOI 10.1093/neuonc/nov197
   Kitai R, 2012, BRAIN TUMOR PATHOL, V29, P47, DOI 10.1007/s10014-011-0064-y
   Lai R, 2002, NEUROLOGY, V59, P1557, DOI 10.1212/01.WNL.0000034256.20173.EA
   Lee IH, 2010, EUR J RADIOL, V76, P48, DOI 10.1016/j.ejrad.2009.05.013
   Lewerenz J, 2007, J NEUROL NEUROSUR PS, V78, P777, DOI 10.1136/jnnp.2006.106385
   Lewerenz Jan, 2009, BMJ Case Rep, V2009, DOI 10.1136/bcr.08.2008.0752
   Mora P, 2014, EUR RADIOL, V24, P2895, DOI 10.1007/s00330-014-3308-5
   Phan TG, 2000, NEURO-ONCOLOGY, V2, P229, DOI 10.1093/neuonc/2.4.229
   Raizer JJ, 2005, J NEURO-ONCOL, V71, P173, DOI 10.1007/s11060-004-1360-8
   Raz E, 2011, J NEUROIMAGING, V21, pe183, DOI 10.1111/j.1552-6569.2010.00477.x
   Rollins KE, 2005, HUM PATHOL, V36, P282, DOI 10.1016/j.humpath.2005.01.014
   Samani A, 2015, J NEUROL NEUROSUR PS, V86, P238, DOI 10.1136/jnnp-2013-307327
   Sugino T, 2013, BRAIN TUMOR PATHOL, V30, P61, DOI 10.1007/s10014-012-0094-0
   Taillibert S, 2008, J NEURO-ONCOL, V86, P225, DOI 10.1007/s11060-007-9468-2
   Terae S, 1996, NEURORADIOLOGY, V38, P34, DOI 10.1007/BF00593213
   Toh CH, 2008, AM J NEURORADIOL, V29, P471, DOI 10.3174/ajnr.A0872
   Vital A, 2007, BRAIN PATHOL, V17, P117, DOI 10.1111/j.1750-3639.2007.00044_2.x
   Weaver JD, 2007, NEUROLOGIST, V13, P150, DOI 10.1097/01.nrl.0000254706.85609.95
   Zacharia TT, 2008, J NEUROIMAGING, V18, P411, DOI 10.1111/j.1552-6569.2007.00231.x
NR 25
TC 24
Z9 27
U1 1
U2 11
PU BMC
PI LONDON
PA CAMPUS, 4 CRINAN ST, LONDON N1 9XW, ENGLAND
SN 1470-7330
EI 1740-5025
J9 CANCER IMAGING
JI Cancer Imaging
PD OCT 5
PY 2017
VL 17
AR 26
DI 10.1186/s40644-017-0128-2
PG 9
WC Oncology; Radiology, Nuclear Medicine & Medical Imaging
WE Science Citation Index Expanded (SCI-EXPANDED)
SC Oncology; Radiology, Nuclear Medicine & Medical Imaging
GA FJ2IU
UT WOS:000412550200001
PM 28982392
OA Green Published, gold
DA 2024-06-03
ER

PT J
AU Cloney, MB
   Sonabend, AM
   Yun, J
   Yang, J
   Iwamoto, F
   Singh, S
   Bhagat, G
   Canoll, P
   Zanazzi, G
   Bruce, JN
   Sisti, M
   Sheth, S
   Connolly, ES
   McKhann, G
AF Cloney, Michael Brendan
   Sonabend, Adam M.
   Yun, Jonathan
   Yang, Jingyan
   Iwamoto, Fabio
   Singh, Suprit
   Bhagat, Govind
   Canoll, Peter
   Zanazzi, George
   Bruce, Jeffrey N.
   Sisti, Michael
   Sheth, Sameer
   Connolly, E. Sander
   McKhann, Guy
TI The safety of resection for primary central nervous system lymphoma: a
   single institution retrospective analysis
SO JOURNAL OF NEURO-ONCOLOGY
LA English
DT Article
DE CNS lymphoma; PCNSL; Central nervous; system lymphoma; Resection;
   Complications
ID PRIMARY CNS LYMPHOMA; INTRACEREBRAL MALIGNANT-LYMPHOMA; CHARLSON
   COMORBIDITY INDEX; PROVIDER CASELOAD; CLINICAL ARTICLE;
   ELDERLY-PATIENTS; UNITED-STATES; BRAIN-TUMORS; SURGERY; COMPLICATIONS
AB Surgical resection is not the standard of care for primary central nervous system lymphoma (PCNSL), as historical studies have demonstrated unfavorable complication rates and limited benefits. Some recent studies suggest that resection may provide a therapeutic benefit, yet the safety of these procedures has not been systematically investigated in the setting of modern neurosurgery. We examined the safety of surgical resection for PCNSL. We retrospectively analyzed all patients with PCNSL treated at Columbia University Medical Center between 2000 and 2015 to assess complications rates following biopsy or resection using the Glioma Outcomes Project system. We identified predictors of complications and selection for resection. Well-validated scales were used to quantify patients ' baseline clinical characteristics, including functional status, comorbid disease burden, and cardiac risk. The overall complication rate was 17.2% after resection, and 28.2% after biopsy. Cardiac risk (p = 0.047, OR 1.72 [1.01, 2.95]), and comorbid diagnoses (p = 0.004, OR 3.05 [1.42, 6.57]) predicted complications on multivariable regression. Patients who underwent resection had better KPS scores (median 70 v. 80, p = 0.0068,. 10 [0.0, 10.00]), and were less likely to have multiple (46.5% v. 27.6%, p = 0.030, OR 1.42 [1.05, 1.92]) or deep lesions (70.4% v. 39.7%, p = 0.001, OR 1.83 [1.26, 2.65]). Age (p = 0.048, OR 0.75 per 10-year increase [0.56, 1.00]) and deep lesions (p = 0.003, OR 0.29 [0.13, 0.65]) influenced selection for resection on multivariable regression. Surgical resection of PCNSL is safe for select patients, with complication rates comparable to rates for other intracranial neoplasms. Whether there is a clinical benefit to resection cannot be concluded.
C1 [Cloney, Michael Brendan; Sonabend, Adam M.; Yun, Jonathan; Singh, Suprit; Bruce, Jeffrey N.; Sisti, Michael; Sheth, Sameer; Connolly, E. Sander; McKhann, Guy] Columbia Univ, Coll Phys & Surg, Med Ctr, Dept Neurol Surg, 710 W 168th St,Room 426, New York, NY 10032 USA.
   [Yang, Jingyan] Columbia Univ, Mailman Sch Publ Hlth, Dept Epidemiol, New York, NY USA.
   [Iwamoto, Fabio] Columbia Univ, Coll Phys & Surg, Med Ctr, Dept Neurol, New York, NY USA.
   [Bhagat, Govind; Canoll, Peter; Zanazzi, George] Columbia Univ, Coll Phys & Surg, Med Ctr, Dept Pathol & Cell Biol, New York, NY USA.
C3 Columbia University; Columbia University; Columbia University; Columbia
   University
RP Sonabend, AM (corresponding author), Columbia Univ, Coll Phys & Surg, Med Ctr, Dept Neurol Surg, 710 W 168th St,Room 426, New York, NY 10032 USA.
EM
RI iwamoto, fabio/KJX-6609-2024; Bruce, Jeffrey N/AAC-5307-2022; Bhagat,
   Govind/AAE-9493-2021
OI Bhagat, Govind/0000-0001-6250-048X; Cloney, Michael/0000-0002-5085-3103
FU NIH [1DP5OD021356-01]; Senior Associate Provost for Faculty Diversity
   and Inclusion, Columbia University
FX NIH office of the Director, Award Number 1DP5OD021356-01, and Diversity
   Recruitment Award by the Office of Senior Associate Provost for Faculty
   Diversity and Inclusion, Columbia University.
CR Abrey LE, 2000, J CLIN ONCOL, V18, P3144, DOI 10.1200/JCO.2000.18.17.3144
   Barker FG, 2005, NEURO-ONCOLOGY, V7, P49, DOI 10.1215/S1152851704000146
   Bataille B, 2000, J NEUROSURG, V92, P261, DOI 10.3171/jns.2000.92.2.0261
   Bellinzona M, 2005, EJSO, V31, P100, DOI 10.1016/j.ejso.2004.10.002
   Benz RJ, 2001, CLIN ORTHOP RELAT R, P116
   Bierman PJ, 2014, ONCOLOGY, P28
   Birim Ö, 2003, EUR J CARDIO-THORAC, V23, P30, DOI 10.1016/S1010-7940(02)00721-2
   Brell M, 2000, ACTA NEUROCHIR, V142, P739, DOI 10.1007/s007010070088
   Burchenal J.H., 1949, EVALUATION CHEMOTHER
   Chang SM, 2003, J NEUROSURG, V98, P1175, DOI 10.3171/jns.2003.98.6.1175
   CHARLSON ME, 1987, J CHRON DIS, V40, P373, DOI 10.1016/0021-9681(87)90171-8
   Cheney FW, 2006, ANESTHESIOLOGY, V105, P1081, DOI 10.1097/00000542-200612000-00007
   Curry WT, 2005, J NEUROSURG, V102, P977, DOI 10.3171/jns.2005.102.6.0977
   D'Amico RS, 2015, WORLD NEUROSURG, V84, P913, DOI 10.1016/j.wneu.2015.05.072
   DEANGELIS LM, 1990, NEUROLOGY, V40, P80, DOI 10.1212/WNL.40.1.80
   Deyo RA, 2010, JAMA-J AM MED ASSOC, V303, P1259, DOI 10.1001/jama.2010.338
   Finks JF, 2011, NEW ENGL J MED, V364, P2128, DOI 10.1056/NEJMsa1010705
   FRANK G, 1985, SURG NEUROL, V23, P3, DOI 10.1016/0090-3019(85)90150-8
   GONZALEZ DG, 1983, CANCER-AM CANCER SOC, V51, P2048, DOI 10.1002/1097-0142(19830601)51:11<2048::AID-CNCR2820511115>3.0.CO;2-T
   Gumprecht HK, 1999, NEUROSURGERY, V44, P97, DOI 10.1097/00006123-199901000-00056
   HELLE TL, 1984, J NEUROSURG, V60, P94, DOI 10.3171/jns.1984.60.1.0094
   HENRY JM, 1974, CANCER, V34, P1293, DOI 10.1002/1097-0142(197410)34:4<1293::AID-CNCR2820340441>3.0.CO;2-P
   Hoang-Xuan K, 2015, LANCET ONCOL, V16, pE322, DOI 10.1016/S1470-2045(15)00076-5
   HOCHBERG FH, 1988, J NEUROSURG, V68, P835, DOI 10.3171/jns.1988.68.6.0835
   Hoover JM, 2013, J NEUROSURG, V118, P1224, DOI 10.3171/2013.2.JNS121731
   Iwamoto FM, 2009, CANCER-AM CANCER SOC, V115, P3758, DOI 10.1002/cncr.24413
   Lee TH, 1999, CIRCULATION, V100, P1043, DOI 10.1161/01.CIR.100.10.1043
   Malone HR, 2015, 2015 C NEUR SURG ANN
   Marcus R., 2009, GUIDELINES DIAGNOSIS
   MENDENHALL NP, 1983, CANCER-AM CANCER SOC, V52, P1993, DOI 10.1002/1097-0142(19831201)52:11<1993::AID-CNCR2820521104>3.0.CO;2-C
   MICHALSKI JM, 1990, RADIOLOGY, V176, P855, DOI 10.1148/radiology.176.3.2389047
   Moiyadi AV, 2012, J NEUROSCI RURAL PRA, V3, P28, DOI 10.4103/0976-3147.91927
   MURRAY K, 1986, J NEUROSURG, V65, P600, DOI 10.3171/jns.1986.65.5.0600
   Orringer D, 2012, J NEUROSURG, V117, P851, DOI 10.3171/2012.8.JNS12234
   Patil CG, 2007, CANCER, V110, P625, DOI 10.1002/cncr.22819
   Patil PG, 2005, NEUROSURGERY, V57, P753, DOI 10.1227/01.NEU.0000175729.79119.1d
   POLLACK IF, 1989, CANCER-AM CANCER SOC, V63, P939, DOI 10.1002/1097-0142(19890301)63:5<939::AID-CNCR2820630526>3.0.CO;2-V
   Sawaya R, 1998, NEUROSURGERY, V42, P1044, DOI 10.1097/00006123-199805000-00054
   Schebesch KM, 2015, CLIN NEUROL NEUROSUR, V139, P125, DOI 10.1016/j.clineuro.2015.09.015
   Siasios Ioannis, 2015, J Clin Med Res, V7, P1007, DOI 10.14740/jocmr2376w
   Singh B, 1997, LARYNGOSCOPE, V107, P1469, DOI 10.1097/00005537-199711000-00009
   Smith-Bindman R, 2012, JAMA-J AM MED ASSOC, V307, P2400, DOI 10.1001/jama.2012.5960
   Sonabend AM, 2016, NEUROSURGERY
   Talacchi A, 2010, J NEURO-ONCOL, V100, P417, DOI 10.1007/s11060-010-0193-x
   Tanaka S, 2013, J NEUROSURG, V118, P786, DOI 10.3171/2012.10.JNS112268
   TOMLINSON FH, 1995, J NEUROSURG, V82, P558, DOI 10.3171/jns.1995.82.4.0558
   Weller M, 2012, NEURO-ONCOLOGY, V14, P1481, DOI 10.1093/neuonc/nos159
   Zada G, 2011, J NEUROSURG, V114, P286, DOI 10.3171/2010.8.JNS10520
NR 48
TC 24
Z9 25
U1 0
U2 3
PU SPRINGER
PI NEW YORK
PA 233 SPRING ST, NEW YORK, NY 10013 USA
SN 0167-594X
EI 1573-7373
J9 J NEURO-ONCOL
JI J. Neuro-Oncol.
PD MAR
PY 2017
VL 132
IS 1
BP 189
EP 197
DI 10.1007/s11060-016-2358-8
PG 9
WC Oncology; Clinical Neurology
WE Science Citation Index Expanded (SCI-EXPANDED)
SC Oncology; Neurosciences & Neurology
GA EQ4MV
UT WOS:000398052800022
PM 28116650
OA Green Accepted
DA 2024-06-03
ER

PT J
AU Fallah, J
   Qunaj, L
   Olszewski, AJ
AF Fallah, Jaleh
   Qunaj, Lindor
   Olszewski, Adam J.
TI Therapy and outcomes of primary central nervous system lymphoma in the
   United States: analysis of the National Cancer Database
SO BLOOD ADVANCES
LA English
DT Article
ID PRIMARY CNS LYMPHOMA; HIGH-DOSE METHOTREXATE; INTERNATIONAL EXTRANODAL
   LYMPHOMA; STEM-CELL TRANSPLANTATION; WHOLE-BRAIN RADIOTHERAPY;
   NON-HODGKINS-LYMPHOMA; LARGE B-CELL; ELDERLY-PATIENTS; DATA-BASE;
   1ST-LINE TREATMENT
AB Although the role of radiation therapy and chemotherapy in primary central nervous system lymphoma (PCNSL) has evolved considerably over the past decade, the application of treatment modalities in the community has not been evaluated. We analyzed the use of chemotherapy, radiation therapy, and associated overall survival, among 9165 HIV-negative PCSNL cases reported to the US National Cancer Database in 2004-2013. During this time, the proportion of patients receiving chemotherapy significantly increased from 65.6% to 78.8% (P for trend <. 0001), whereas the proportion receiving radiation therapy decreased from 37.6% to 18.8% (P < .0001). Adjusting for the varying distribution of clinical and sociodemographic characteristics by type of treating facility, the risk of not receiving chemotherapy was significantly lower in academic/research cancer programs compared with community programs (adjusted relative risk, 0.69; 95% confidence interval [CI], 0.62-0.76; P < .0001). Furthermore, omission of chemotherapy was associated with increasing age, comorbidities, black race, and indicators of poor socioeconomic status. Overall survival at 3 years was 37.7% (95% CI, 36.6-38.8) and ranged from 14.1% for patients treated with radiation therapy alone to 51.8% for those who received multiagent chemotherapy. There was evidence of improved survival over time (P for trend =.0002). The disparities in application of chemotherapy for PCNSL underscore the need to provide access to expert management for this rare disease and improve safe delivery of systemic treatment in the community setting, where most older patients receive their care.
C1 [Fallah, Jaleh; Qunaj, Lindor; Olszewski, Adam J.] Brown Univ, Alpert Med Sch, Providence, RI 02912 USA.
   [Fallah, Jaleh] Mem Hosp Rhode Isl, Dept Med, Pawtucket, RI USA.
   [Olszewski, Adam J.] Rhode Isl Hosp, Div Hematol Oncol, George 353,593 Eddy St, Providence, RI 02903 USA.
C3 Brown University; Lifespan Health Rhode Island; Rhode Island Hospital
RP Olszewski, AJ (corresponding author), Rhode Isl Hosp, Div Hematol Oncol, George 353,593 Eddy St, Providence, RI 02903 USA.
EM
RI Fallah, Jaleh/AAD-3599-2020; Olszewski, Adam J/F-8449-2013; Fallah,
   Jaleh/AAB-7189-2020
OI Olszewski, Adam J/0000-0002-6472-6658; Fallah,
   Jaleh/0000-0001-7365-2104; Qunaj, Lindor/0000-0001-8461-8374
FU American Society of Hematology Scholar Award; American Cancer Society
   Research Scholar Award
FX A.J.O. is supported by the American Society of Hematology Scholar Award
   and the American Cancer Society Research Scholar Award.
CR Abrey LE, 2006, J CLIN ONCOL, V24, P5711, DOI 10.1200/JCO.2006.08.2941
   [Anonymous], NCCN clinical practice guidelines in Oncology: breast cancer
   Bilimoria KY, 2008, ANN SURG ONCOL, V15, P683, DOI 10.1245/s10434-007-9747-3
   Brem Steven S, 2005, J Natl Compr Canc Netw, V3, P644
   DeAngelis LM, 2002, J CLIN ONCOL, V20, P4643, DOI 10.1200/JCO.2002.11.013
   Deddens JA, 2008, OCCUP ENVIRON MED, V65, P501, DOI 10.1136/oem.2007.034777
   DEYO RA, 1992, J CLIN EPIDEMIOL, V45, P613, DOI 10.1016/0895-4356(92)90133-8
   Ferreri AJM, 2003, J CLIN ONCOL, V21, P266, DOI 10.1200/JCO.2003.09.139
   Ferreri AJM, 2016, LANCET HAEMATOL, V3, pE217, DOI 10.1016/S2352-3026(16)00036-3
   Ferreri AJM, 2016, BLOOD, V127, P1642, DOI 10.1182/blood-2015-10-636340
   Ferreri AJM, 2011, BLOOD, V118, P510, DOI 10.1182/blood-2011-03-321349
   Ferreri AJM, 2009, LANCET, V374, P1512, DOI 10.1016/S0140-6736(09)61416-1
   Giobbie-Hurder A, 2013, J CLIN ONCOL, V31, P2963, DOI 10.1200/JCO.2013.49.5283
   GLASS J, 1994, J NEUROSURG, V81, P188, DOI 10.3171/jns.1994.81.2.0188
   Hoang-Xuan K, 2015, LANCET ONCOL, V16, pE322, DOI 10.1016/S1470-2045(15)00076-5
   Illerhaus G, 2009, ANN ONCOL, V20, P319, DOI 10.1093/annonc/mdn628
   Illerhaus G, 2007, HAEMATOLOGICA, V93, P147, DOI 10.3324/haematol.11771
   Kansara R, 2015, AM J HEMATOL, V90, P1149, DOI 10.1002/ajh.24204
   Kasenda B, 2015, ANN ONCOL, V26, P1305, DOI 10.1093/annonc/mdv076
   Kasenda B, 2016, BLOOD, V128, P32, DOI 10.1182/blood-2016-01-650101
   Keegan THM, 2013, LEUKEMIA LYMPHOMA, V54, P743, DOI 10.3109/10428194.2012.727415
   Korfel A, 2013, NAT REV NEUROL, V9, P317, DOI 10.1038/nrneurol.2013.83
   Mead GM, 2000, CANCER, V89, P1359, DOI 10.1002/1097-0142(20000915)89:6<1359::AID-CNCR21>3.0.CO;2-9
   Morris PG, 2013, J CLIN ONCOL, V31, P3971, DOI 10.1200/JCO.2013.50.4910
   Nabhan C, 2012, CANCER-AM CANCER SOC, V118, P4842, DOI 10.1002/cncr.27513
   NELSON DF, 1992, INT J RADIAT ONCOL, V23, P9, DOI 10.1016/0360-3016(92)90538-S
   Ney DE, 2010, CANCER-AM CANCER SOC, V116, P4605, DOI 10.1002/cncr.25363
   O'Neill BP, 2013, AM J HEMATOL, V88, P997, DOI 10.1002/ajh.23551
   Olszewski AJ, 2016, CANCER-AM CANCER SOC, V122, P2689, DOI 10.1002/cncr.30112
   Olszewski AJ, 2015, LANCET HAEMATOL, V2, pE459, DOI 10.1016/S2352-3026(15)00188-X
   Olszewski AJ, 2015, J CLIN ONCOL, V33, P625, DOI 10.1200/JCO.2014.58.7543
   Omuro A, 2015, LANCET HAEMATOL, V2, pE251, DOI 10.1016/S2352-3026(15)00074-5
   Omuro A, 2015, BLOOD, V125, P1403, DOI 10.1182/blood-2014-10-604561
   Roth P, 2014, CURR OPIN NEUROL, V27, P697, DOI 10.1097/WCO.0000000000000145
   Rubenstein JL, 2013, J CLIN ONCOL, V31, P3061, DOI 10.1200/JCO.2012.46.9957
   Shiels MS, 2016, BRIT J HAEMATOL, V174, P417, DOI 10.1111/bjh.14073
   Thiel E, 2010, LANCET ONCOL, V11, P1036, DOI 10.1016/S1470-2045(10)70229-1
   Vardiman JW., 2008, WHO CLASSIFICATION T
   Villano JL, 2011, BRIT J CANCER, V105, P1414, DOI 10.1038/bjc.2011.357
   Warren JL, 2013, J CLIN ONCOL, V31, P1984, DOI 10.1200/JCO.2012.46.3323
   Welch MR, 2012, NEURO-ONCOLOGY, V14, P1304, DOI 10.1093/neuonc/nos207
   White IR, 2011, STAT MED, V30, P377, DOI 10.1002/sim.4067
NR 42
TC 24
Z9 24
U1 0
U2 2
PU AMER SOC HEMATOLOGY
PI WASHINGTON
PA 2021 L ST NW, SUITE 900, WASHINGTON, DC 20036 USA
SN 2473-9529
EI 2473-9537
J9 BLOOD ADV
JI Blood Adv.
PD DEC 13
PY 2016
VL 1
IS 2
BP 112
EP 121
DI 10.1182/bloodadvances.2016000927
PG 10
WC Hematology
WE Science Citation Index Expanded (SCI-EXPANDED)
SC Hematology
GA FG8AH
UT WOS:000410647200002
PM 29296804
OA Green Published, Bronze
DA 2024-06-03
ER

PT J
AU Chang, C
   Lin, CH
   Cheng, AL
   Medeiros, LJ
   Chang, KC
AF Chang, Chen
   Lin, Ching-Hung
   Cheng, Ann-Lii
   Medeiros, L. Jeffrey
   Chang, Kung-Chao
TI Primary central nervous system diffuse large B-cell lymphoma has poorer
   immune cell infiltration and prognosis than its peripheral counterpart
SO HISTOPATHOLOGY
LA English
DT Article
DE antitumour immunity; dendritic cells; diffuse large B-cell lymphoma;
   primary central nervous system; prognosis; T cells
ID CHRONIC LYMPHOCYTIC-LEUKEMIA; CYTOTOXIC T-LYMPHOCYTES; DENDRITIC CELLS;
   HIGH NUMBERS; EXPRESSION; SURVIVAL; PREDICTS; BRAIN; GENE;
   IMMUNOHISTOCHEMISTRY
AB AimsPrimary central nervous system (CNS) diffuse large B-cell lymphoma (PCNSL) is an ominous disease with a poor prognosis. The brain is an immune-privileged sanctuary, and this may contribute to an ineffective host immune response and thus a poorer outcome. The aim of this study was therefore to study the difference in the immune composition in PCNSL and non-CNS diffuse large B-cell lymphoma (DLBCL), and the role of the immune response in PCNSL prognosis.
   Methods and resultsThirty-two biopsy specimens of PCNSL and 30 specimens of low-stage non-CNS DLBCL from immunocompetent patients formed the study group. The density and distribution of immune cells, including dendritic cells (dendritic cell-specific lysosomal-associated membrane protein-positive and S100-positive), effector/memory T cells (CD45RO-positive), and cytotoxic T cells (granzyme B-positive), and the expression of HLA-DR by lymphoma cells, were evaluated immunohistochemically. PCNSL patients showed poorer overall survival (P=0.032). On comparison of the PCNSL and DLBCL biopsy specimens, the PCNSL cells showed less HLA-DR expression (P=0.003), and there were fewer S100-positive cells (P<0.01), and effector T cells (P= 0.026) infiltrating PCNSL than infiltrating DLBCL. For PCNSL patients, fewer cytotoxic T cells in the background constituted a poor prognostic factor (P=0.004). Intratumoral S100-positive cell infiltration was positively correlated with T-cell infiltration, and a T-cell rimming pattern.
   ConclusionsIn PCNSL, the baseline antitumour immune response is less as compared with non-CNS DLBCL, and this response may play a role in the poorer prognosis. Adjuvant dendritic cell and T-cell immunotherapy may further boost treatment responses in PCNSL patients.
C1 [Chang, Chen; Chang, Kung-Chao] Natl Cheng Kung Univ & Hosp, Coll Med, Dept Pathol, Tainan 704, Taiwan.
   [Lin, Ching-Hung; Cheng, Ann-Lii] Natl Taiwan Univ Hosp, Dept Oncol, Taipei, Taiwan.
   [Lin, Ching-Hung; Cheng, Ann-Lii] Natl Taiwan Univ Hosp, Dept Internal Med, Taipei 100, Taiwan.
   [Medeiros, L. Jeffrey] Univ Texas MD Anderson Canc Ctr, Dept Hematopathol, Houston, TX 77030 USA.
C3 National Cheng Kung University; National Cheng Kung University Hospital;
   National Taiwan University; National Taiwan University Hospital;
   National Taiwan University; National Taiwan University Hospital;
   University of Texas System; UTMD Anderson Cancer Center
RP Chang, KC (corresponding author), Natl Cheng Kung Univ & Hosp, Coll Med, Dept Pathol, 138 Sheng Li Rd, Tainan 704, Taiwan.
EM
RI Medeiros, L. Jeffrey/AAB-1018-2019
OI Medeiros, L. Jeffrey/0000-0001-6577-8006; Cheng,
   Ann-Lii/0000-0002-9152-6512; LIN, CHING-HUNG/0000-0003-2403-4056
FU Ministry of Science and Technology, Taiwan
   [MOST-103-2320-B-006-020-MY3]; National Cheng Kung University Hospital,
   Taiwan [NCKUH-10104006]
FX The authors thank Mi-Chia Ma (Associate Professor, Department of
   Statistics, National Cheng Kung University, Tainan, Taiwan) for advice
   on statistical interpretation. This study was supported by grants from
   the Ministry of Science and Technology, Taiwan
   (MOST-103-2320-B-006-020-MY3) and National Cheng Kung University
   Hospital, Taiwan (NCKUH-10104006) to K. C. Chang.
CR Alizadeh AA, 2000, NATURE, V403, P503, DOI 10.1038/35000501
   Ansell SM, 2001, J CLIN ONCOL, V19, P720, DOI 10.1200/JCO.2001.19.3.720
   Ansell SM, 2009, CLIN CANCER RES, V15, P6446, DOI 10.1158/1078-0432.CCR-09-1339
   Armand P, 2013, J CLIN ONCOL, V31, P4199, DOI 10.1200/JCO.2012.48.3685
   Banchereau J, 2000, ANNU REV IMMUNOL, V18, P767, DOI 10.1146/annurev.immunol.18.1.767
   BOHLEN H, 1993, BLOOD, V82, P1803
   Camilleri-Broët S, 2006, BLOOD, V107, P190, DOI 10.1182/blood-2005-03-1024
   Chang KC, 2007, CLIN CANCER RES, V13, P6666, DOI 10.1158/1078-0432.CCR-07-0504
   Choi WWL, 2009, CLIN CANCER RES, V15, P5494, DOI 10.1158/1078-0432.CCR-09-0113
   Colton CA, 2013, J NEUROIMMUNE PHARM, V8, P145, DOI 10.1007/s11481-012-9414-8
   COX DR, 1972, J R STAT SOC B, V34, P187
   D'Agostino PM, 2012, ACTA NEUROPATHOL, V124, P599, DOI 10.1007/s00401-012-1018-0
   Ferreri AJM, 2003, J CLIN ONCOL, V21, P266, DOI 10.1200/JCO.2003.09.139
   Ferreri AJM, 2011, BLOOD, V118, P510, DOI 10.1182/blood-2011-03-321349
   Fong L, 2000, ANNU REV IMMUNOL, V18, P245, DOI 10.1146/annurev.immunol.18.1.245
   Galand C, 2012, FRONT IMMUNOL, V3, DOI 10.3389/fimmu.2012.00098
   GEHAN EA, 1965, BIOMETRIKA, V52, P650, DOI 10.1093/biomet/52.3-4.650
   Giannini C, 2014, J NEUROPATH EXP NEUR, V73, P478, DOI 10.1097/NEN.0000000000000076
   Hans CP, 2004, BLOOD, V103, P275, DOI 10.1182/blood-2003-05-1545
   Hasselblom S, 2007, BRIT J HAEMATOL, V137, P364, DOI 10.1111/j.1365-2141.2007.06593.x
   Janeway Jr CA, 2005, IMMUNOBIOLOGY IMMUNE, P630
   Juszczynski P, 2008, ARCH IMMUNOL THER EX, V56, P245, DOI 10.1007/s00005-008-0028-z
   KAPLAN EL, 1958, J AM STAT ASSOC, V53, P457, DOI 10.2307/2281868
   Komohara Y, 2011, J CLIN EXP HEMATOP, V51, P93, DOI 10.3960/jslrt.51.93
   Lin CH, 2006, CLIN CANCER RES, V12, P1152, DOI 10.1158/1078-0432.CCR-05-1699
   López-Guillermo A, 2005, J CLIN ONCOL, V23, P2797, DOI 10.1200/JCO.2005.07.155
   McNutt M, 2013, SCIENCE, V342, P1417, DOI 10.1126/science.1249481
   Mellman I, 2011, NATURE, V480, P480, DOI 10.1038/nature10673
   Melzer P, 2001, EXP NEUROL, V168, P63, DOI 10.1006/exnr.2000.7554
   Meyer PN, 2011, J CLIN ONCOL, V29, P200, DOI 10.1200/JCO.2010.30.0368
   MILLER RA, 1989, NEW ENGL J MED, V321, P851, DOI 10.1056/NEJM198909283211302
   Monti S, 2005, BLOOD, V105, P1851, DOI 10.1182/blood-2004-07-2947
