## Supplementary material for "Bibliometric analysis of emerging trend and top 100 most cited articles in primary central nervous system lymphoma": Supple File 1: savedrecs (3).txt

FN Clarivate Analytics Web of Science
VR 1.0
PT J
AU Linnebank, M
   Moskau, S
   Kowoll, A
   Semmler, A
   Bangard, C
   Vogt-Schaden, M
   Egerer, G
   Schackert, G
   Reichmann, H
   Schmidt-Wolf, IGH
   Pels, H
   Schlegel, U
AF Linnebank, M.
   Moskau, S.
   Kowoll, A.
   Semmler, A.
   Bangard, C.
   Vogt-Schaden, M.
   Egerer, G.
   Schackert, G.
   Reichmann, H.
   Schmidt-Wolf, I. G. H.
   Pels, H.
   Schlegel, U.
TI Association of transcobalamin c. 776C&gt;G with overall survival in
   patients with primary central nervous system lymphoma
SO BRITISH JOURNAL OF CANCER
LA English
DT Article
DE primary central nervous system lymphoma; polymorphism; survival
ID WHITE-MATTER CHANGES; METHIONINE METABOLISM; CHEMOTHERAPY
AB BACKGROUND: Chemotherapy for primary central nervous system lymphoma (PCNSL) is based on methotrexate (MTX), which interferes with both nucleic acid synthesis and methionine metabolism. We have reported previously that genetic variants with influence on methionine metabolism are associated with MTX side effects, that is, the occurrence of white matter lesions as a sign of MTX neurotoxicity. Here, we investigated whether such variants are associated with MTX efficacy in terms of overall survival in MTX-treated PCNSL patients.
   METHODS: We analysed seven genetic variants influencing methionine metabolism in 68 PCNSL patients treated with systemic and facultative intraventricular MTX-based polychemotherapy (Bonn protocol).
   RESULTS: Median age at diagnosis was 59 years (range: 28-77), 32 patients were female. Younger age (Wald = 8.9; P = 0.003) and the wild-type C (CC) allele of the genotype transcobalamin c (Tc2). 776C>G (Wald = 6.7; P = 0.010) were associated with longer overall survival in a multivariate COX regression analysis.
   CONCLUSION: This observation suggests that the missense variant Tc2. 776C>G influences both neurotoxicity and efficacy of MTX in the Bonn PCNSL protocol. British Journal of Cancer (2012) 107, 1840-1843. doi:10.1038/bjc.2012.476 www.bjcancer.com Published online 25 October 2012 (C) 2012 Cancer Research UK
C1 [Linnebank, M.; Moskau, S.; Semmler, A.] Univ Hosp Bonn, Dept Neurol, Bonn, Germany.
   [Linnebank, M.; Semmler, A.] Univ Zurich Hosp, Dept Neurol, CH-8091 Zurich, Switzerland.
   [Kowoll, A.; Schlegel, U.] Univ Hosp Bochum, Dept Neurol, Bochum, Germany.
   [Bangard, C.] Univ Hosp Cologne, Dept Radiol, Cologne, Germany.
   [Vogt-Schaden, M.] Univ Heidelberg Hosp, Dept Neurol, Heidelberg, Germany.
   [Egerer, G.] Univ Heidelberg Hosp, Dept Med, Heidelberg, Germany.
   [Schackert, G.] Univ Hosp Dresden, Dept Neurosurg, Dresden, Germany.
   [Reichmann, H.] Univ Hosp Dresden, Dept Neurol, Dresden, Germany.
   [Schmidt-Wolf, I. G. H.] Univ Hosp Bonn, Dept Med 3, Bonn, Germany.
   [Pels, H.] Krankenhaus Barmherzige Bruder, Dept Neurol, Regensburg, Germany.
C3 University of Bonn; University of Zurich; University Zurich Hospital;
   Ruhr University Bochum; University of Cologne; Ruprecht Karls University
   Heidelberg; Ruprecht Karls University Heidelberg; Technische Universitat
   Dresden; Carl Gustav Carus University Hospital; Technische Universitat
   Dresden; Carl Gustav Carus University Hospital; University of Bonn
RP Linnebank, M (corresponding author), Univ Hosp Bonn, Dept Neurol, Bonn, Germany.
EM
FU Deutsche Krebshilfe [106262]
FX This study was supported by the 'Deutsche Krebshilfe', grant no. 106262
   (Linnebank M and Schlegel U).
CR Abrey LE, 2006, J CLIN ONCOL, V24, P5711, DOI 10.1200/JCO.2006.08.2941
   Abrey LE, 2005, J CLIN ONCOL, V23, P5034, DOI 10.1200/JCO.2005.13.524
   Ferreri AJM, 2003, J CLIN ONCOL, V21, P266, DOI 10.1200/JCO.2003.09.139
   FINKELSTEIN JD, 1990, J NUTR BIOCHEM, V1, P228, DOI 10.1016/0955-2863(90)90070-2
   Jahnke K, 2009, EXPERT REV NEUROTHER, V9, P1497, DOI 10.1586/ERN.09.100
   Linnebank M, 2005, NEUROLOGY, V64, P912, DOI 10.1212/01.WNL.0000152840.26156.74
   Linnebank M, 2007, J CHEMOTHERAPY, V19, P455, DOI 10.1179/joc.2007.19.4.455
   Linnebank M, 2009, NEURO-ONCOLOGY, V11, P2, DOI 10.1215/15228517-2008-082
   Pels H, 2003, J CLIN ONCOL, V21, P4489, DOI 10.1200/JCO.2003.04.056
   Pels H, 2010, NEURO-ONCOLOGY, V12, P720, DOI 10.1093/neuonc/noq010
   Pels H, 2009, J NEURO-ONCOL, V91, P299, DOI 10.1007/s11060-008-9712-4
   Semmler A, 2009, J NEUROL, V256, P1277, DOI 10.1007/s00415-009-5114-6
   Stanislawska-Sachadyn A, 2010, EUR J CLIN NUTR, V64, P1338, DOI 10.1038/ejcn.2010.157
   SURTEES R, 1991, LANCET, V338, P1550, DOI 10.1016/0140-6736(91)92373-A
   von Castel-Dunwoody KM, 2005, AM J CLIN NUTR, V81, P1436
NR 15
TC 2
Z9 2
U1 0
U2 5
PU NATURE PUBLISHING GROUP
PI LONDON
PA MACMILLAN BUILDING, 4 CRINAN ST, LONDON N1 9XW, ENGLAND
SN 0007-0920
EI 1532-1827
J9 BRIT J CANCER
JI Br. J. Cancer
PD NOV 20
PY 2012
VL 107
IS 11
BP 1840
EP 1843
DI 10.1038/bjc.2012.476
PG 4
WC Oncology
WE Science Citation Index Expanded (SCI-EXPANDED)
SC Oncology
GA 041RP
UT WOS:000311419200008
PM 23099805
OA hybrid, Green Published, Green Accepted
DA 2024-06-03
ER

PT J
AU Porto, L
   Hattingen, E
   Stuecher, A
   Herminghaus, S
   Lanfermann, H
   Pilatus, UPU
AF Porto, L.
   Hattingen, E.
   Stuecher, A.
   Herminghaus, S.
   Lanfermann, H.
   Pilatus, U. P. Ulrich
TI Cutoff Value of Choline Concentration Reliably Reveals High-Grade Brain
   Tumors among Other Contrast-Enhancing Brain Lesions
SO JOURNAL OF NEUROLOGICAL SURGERY PART A-CENTRAL EUROPEAN NEUROSURGERY
LA English
DT Article
DE choline; 1 H-MR spectroscopy; cerebral mass lesions; brain tumors;
   malignant
ID MAGNETIC-RESONANCE-SPECTROSCOPY; IN-VIVO; METABOLITE QUANTIFICATION;
   STEREOTACTIC BIOPSY; LOCALIZED PROTON; MR SPECTROSCOPY; H-1 MRS; NMR;
   GLIOMAS; DIFFERENTIATE
AB Background and Aim To evaluate whether there is a cutoff value for a metabolite concentration measured by 1 H MR spectroscopy (MRS), which can be used to differentiate malignant brain tumors (high-grade gliomas, primary CNS lymphomas [PCNSL] and metastases) from other contrast-enhancing lesions like low-grade gliomas and non-neoplastic lesions.
   Material and Methods 1 H MRS was performed in 252 consecutive patients with space-occupying brain lesions which were enhanced with application of a contrast agent. Concentrations of N-acetyl-aspartate, total creatine, choline containing metabolites (total choline, tCho), lipids, and lactate were evaluated from the contrast-enhancing part of the lesions and from the normal appearing brain tissue. Linear discriminant analysis was used to find the best predictor for malignant brain tumors. In addition, receiver operating characteristic analysis (ROC) was performed to determine a cutoff value for the best predictor in detecting malignant brain tumors with a specificity of >95%.
   Results All brain tumors and 20 out of 47 nonneoplastic lesions were examined histopathologically. The remaining 27 diagnoses were based on MR imaging, clinical findings, and follow-up. The final diagnosis was 134 high-grade gliomas (WHO grade III/IV), 36 metastases, 9 PCNSL, 8 low-grade gliomas (WHO grade I/II), 34 infections, 9 infarctions, 2 hematomas, and 2 vasculitides. 18 patients were excluded due to insufficient spectral quality. The tCho concentration was the best predictor to differentiate malignant brain tumors from enhancing low-grade gliomas or non-neoplastic lesions (F=26.6 [df: 25.833], p<0.0005). The ROC revealed that a cutoff tCho value, based on an increase of >= 40% compared to normal, yielded a specificity of 100% and a sensitivity of 89.4% to correctly diagnose a malignant brain tumor.
   Conclusion 1 H MRS reliably differentiates malignant brain tumors from other contrast-enhancing brain lesions. At least a 40% increase of tCho compared to normal brain tissue indicates a malignant tumor (WHO grade III/IV gliomas, PCNSL, metastases) with >90% specificity and sensitivity.
C1 [Porto, L.; Hattingen, E.; Stuecher, A.; Herminghaus, S.; Pilatus, U. P. Ulrich] Univ Frankfurt Klinikum, D-60528 Frankfurt, Germany.
   [Lanfermann, H.] Hannover Med Sch, D-30623 Hannover, Germany.
C3 Goethe University Frankfurt; Goethe University Frankfurt Hospital;
   Hannover Medical School
RP Porto, L (corresponding author), Univ Frankfurt Klinikum, Schleusenweg 2-16, D-60528 Frankfurt, Germany.
EM
RI Hattingen, Elke/D-7880-2015; Lanfermann, Heinrich/JCO-1877-2023; Porto,
   Luciana/AAF-6355-2020; Lanfermann, Heinrich/AAR-4069-2021
OI Porto, Luciana/0000-0003-0554-2493; Hattingen, Elke/0000-0002-8392-9004
CR Al-Okaili RN, 2007, RADIOLOGY, V243, P539, DOI 10.1148/radiol.2432060493
   Bitsch A, 1999, AM J NEURORADIOL, V20, P1619
   Blasel S, 2011, J NEUROL, V258, P113, DOI 10.1007/s00415-010-5703-4
   Bulakbasi N, 2003, AM J NEURORADIOL, V24, P225
   CHRISTIANSEN P, 1993, MAGN RESON IMAGING, V11, P107, DOI 10.1016/0730-725X(93)90418-D
   de Beer R, 1998, MAGN RESON IMAGING, V16, P1107, DOI 10.1016/S0730-725X(98)00119-2
   DeStefano N, 1995, ANN NEUROL, V38, P901, DOI 10.1002/ana.410380610
   ElDeredy W, 1997, NMR BIOMED, V10, P99, DOI 10.1002/(SICI)1099-1492(199705)10:3<99::AID-NBM461>3.0.CO;2-#
   EVANOCHKO WT, 1984, BIOCHIM BIOPHYS ACTA, V805, P104, DOI 10.1016/0167-4889(84)90042-9
   Galanaud D, 2003, J NEUROSURG, V98, P269, DOI 10.3171/jns.2003.98.2.0269
   GILLIES RJ, 1994, MAGNET RESON MED, V32, P310, DOI 10.1002/mrm.1910320306
   Hattingen E, 2010, NEUROL RES, V32, P593, DOI 10.1179/016164109X12478302362770
   Henriksen O, 1995, NMR BIOMED, V8, P139, DOI 10.1002/nbm.1940080402
   Hermann EJ, 2008, STEREOT FUNCT NEUROS, V86, P300, DOI 10.1159/000155232
   Herminghaus S, 2003, J NEUROSURG, V98, P74, DOI 10.3171/jns.2003.98.1.0074
   Hourani R, 2008, AM J NEURORADIOL, V29, P366, DOI 10.3174/ajnr.A0810
   Jackson JR, 1997, FASEB J, V11, P457, DOI 10.1096/fasebj.11.6.9194526
   Jansen JFA, 2006, RADIOLOGY, V240, P318, DOI 10.1148/radiol.2402050314
   Julià-Sapé M, 2006, J NEUROSURG, V105, P6, DOI 10.3171/jns.2006.105.1.6
   KAPLAN O, 1991, J BIOL CHEM, V266, P3688
   Keevil SF, 1998, MAGN RESON IMAGING, V16, P1093, DOI 10.1016/S0730-725X(98)00118-0
   Kleihues PCW., 2000, WORLD HLTH ORG CLASS
   Law M, 2003, AM J NEURORADIOL, V24, P1989
   Majós C, 2003, EUR RADIOL, V13, P582, DOI 10.1007/s00330-002-1547-3
   MICHAELIS T, 1993, RADIOLOGY, V187, P219, DOI 10.1148/radiology.187.1.8451417
   Möller-Hartmann W, 2002, NEURORADIOLOGY, V44, P371, DOI 10.1007/s00234-001-0760-0
   Muacevic A, 2003, NERVENARZT, V74, P350, DOI 10.1007/s00115-003-1487-4
   Panigrahy A, 2006, AM J NEURORADIOL, V27, P560
   Podo F, 1999, NMR BIOMED, V12, P413, DOI 10.1002/(SICI)1099-1492(199911)12:7<413::AID-NBM587>3.3.CO;2-L
   Podo F, 1996, ANTICANCER RES, V16, P1399
   POPTANI H, 1995, AM J NEURORADIOL, V16, P1593
   Senft C, 2009, NEUROSURGERY, V65, P908, DOI 10.1227/01.NEU.0000356982.82378.BA
   Stadlbauer A, 2006, RADIOLOGY, V238, P958, DOI 10.1148/radiol.2382041896
   SZE DY, 1990, BIOCHIM BIOPHYS ACTA, V1054, P198, DOI 10.1016/0167-4889(90)90241-5
   Tate AR, 1996, ANTICANCER RES, V16, P1575
   Tate AR, 1998, NMR BIOMED, V11, P177, DOI 10.1002/(SICI)1099-1492(199806/08)11:4/5<177::AID-NBM534>3.0.CO;2-U
   Vanhamme L, 1997, J MAGN RESON, V129, P35, DOI 10.1006/jmre.1997.1244
   White ML, 2005, AM J NEURORADIOL, V26, P784
   Yerli H, 2007, DIAGN INTERV RADIOL, V13, P3
NR 39
TC 2
Z9 2
U1 0
U2 2
PU THIEME MEDICAL PUBL INC
PI NEW YORK
PA 333 SEVENTH AVE, NEW YORK, NY 10001 USA
SN 2193-6315
EI 2193-6323
J9 J NEUROL SURG PART A
JI J. Neurol. Surg. Part A
PD MAY
PY 2012
VL 73
IS 3
BP 147
EP 152
DI 10.1055/s-0032-1313723
PG 6
WC Clinical Neurology; Surgery
WE Science Citation Index Expanded (SCI-EXPANDED)
SC Neurosciences & Neurology; Surgery
GA 175UO
UT WOS:000321259300004
PM 22190143
DA 2024-06-03
ER

PT J
AU Steinbeck, JA
   Stuplich, M
   Blasius, E
   Pels, H
   Glas, M
   Schlegel, U
   Herrlinger, U
AF Steinbeck, J. A.
   Stuplich, M.
   Blasius, E.
   Pels, H.
   Glas, M.
   Schlegel, U.
   Herrlinger, U.
TI Relapse of primary central nervous system lymphoma 13 years after
   high-dose methotrexate-based polychemotherapy
SO JOURNAL OF CLINICAL NEUROSCIENCE
LA English
DT Article
DE Methotrexate; MTX; Neurotoxicity; Polychemotherapy; Primary CNS
   lymphoma; Relapse
ID PRIMARY CNS LYMPHOMA; INTRAVENTRICULAR CHEMOTHERAPY; FEATURES
AB We report a now 74-year-old patient who was successfully treated with a methotrexate (MTX)-ssbased polychemotherapy protocol (Bonn protocol) for primary central nervous system lymphoma (PCNSL) in 1996. When presenting with an unusually late relapse after 13 years of tumor-free survival the diagnosis was made on the basis of clinical and radiological criteria. In the context of the very limited treatment options for recurrent PCNSL, it is reassuring that the re-application of high dose-MTX-based polychemotherapy, including intraventricular treatment, again succeeded in a sustained complete response with still low neurotoxicity. (C) 2011 Elsevier Ltd. All rights reserved.
C1 [Steinbeck, J. A.; Stuplich, M.; Blasius, E.; Glas, M.; Herrlinger, U.] Univ Bonn, Div Clin Neurooncol, Dept Neurol, Sch Med, D-53105 Bonn, Germany.
   [Steinbeck, J. A.] Univ Bonn, Inst Reconstruct Neurobiol, D-53105 Bonn, Germany.
   [Pels, H.; Schlegel, U.] Univ Bochum, Dept Neurol, Knappschaftskrankenhaus, Bochum, Germany.
C3 University of Bonn; University of Bonn; Ruhr University Bochum
RP Herrlinger, U (corresponding author), Univ Bonn, Div Clin Neurooncol, Dept Neurol, Sch Med, Sigmund Freud Str 25, D-53105 Bonn, Germany.
EM
CR Abrey LE, 1998, J CLIN ONCOL, V16, P859, DOI 10.1200/JCO.1998.16.3.859
   Batchelor T, 2006, J CLIN ONCOL, V24, P1281, DOI 10.1200/JCO.2005.04.8819
   de Jong D, 2003, BLOOD, V102, P324, DOI 10.1182/blood-2002-09-2822
   Enting RH, 2004, NEUROLOGY, V63, P901, DOI 10.1212/01.WNL.0000137050.43114.42
   Fliessbach K, 2003, ARCH NEUROL-CHICAGO, V60, P563, DOI 10.1001/archneur.60.4.563
   Herrlinger U, 2005, J NEUROL, V252, P1409, DOI 10.1007/s00415-005-0854-4
   Juergens A, 2010, ANN NEUROL, V67, P182, DOI 10.1002/ana.21824
   Küker W, 2005, J NEURO-ONCOL, V72, P169, DOI 10.1007/s11060-004-3390-7
   Pels H, 2003, J CLIN ONCOL, V21, P4489, DOI 10.1200/JCO.2003.04.056
NR 9
TC 2
Z9 3
U1 0
U2 1
PU ELSEVIER SCI LTD
PI OXFORD
PA THE BOULEVARD, LANGFORD LANE, KIDLINGTON, OXFORD OX5 1GB, OXON, ENGLAND
SN 0967-5868
J9 J CLIN NEUROSCI
JI J. Clin. Neurosci.
PD NOV
PY 2011
VL 18
IS 11
BP 1554
EP 1555
DI 10.1016/j.jocn.2011.04.003
PG 2
WC Clinical Neurology; Neurosciences
WE Science Citation Index Expanded (SCI-EXPANDED)
SC Neurosciences & Neurology
GA 839XM
UT WOS:000296402800032
PM 21868233
DA 2024-06-03
ER

PT J
AU Gonzalez-Aguilar, A
   Houillier, C
   Soussain, C
   Hoang-Xuan, K
AF Gonzalez-Aguilar, A.
   Houillier, C.
   Soussain, C.
   Hoang-Xuan, K.
TI Management of primary central nervous system lymphoma
SO REVUE NEUROLOGIQUE
LA French
DT Article
DE Lymphoma; Central nervous system; Immunocompetent; Steroids;
   Radiotherapy; Chemotherapy; Methotrexate; Neurotoxicity; Rituximab; Stem
   cells
ID PRIMARY CNS LYMPHOMA; HIGH-DOSE METHOTREXATE; WHOLE-BRAIN RADIOTHERAPY;
   NON-HODGKINS-LYMPHOMA; STEM-CELL RESCUE; TERM-FOLLOW-UP;
   MAGNETIC-RESONANCE-SPECTROSCOPY; RECURRENT PRIMARY CNS; MULTICENTER
   PHASE-II; TO-TREAT ANALYSIS
AB Primary CNS lymphoma is the malignant brain tumor whose prognosis has improved the most the two past decades. The majority of the patients achieve a complete remission with treatment and a substantial minority may hope to be cured. The treatment includes high-dose methotrexate polychemotherapy combined or not with whole brain radiotherapy (WBRT). Elderly patients who are exposed to a high risk of treatment induced neurotoxicity need a specific management avoiding or defering WBRT. In young patients, the main questions concerning the treatment are the role of consolidation WBRT and intensive chemotherapy with peripheral blood stem cell rescue in the initial treatment of PCNSL. As recently shown, national and international collaborative efforts make now possible randomized trials for this rare disease, which would contribute to better define the treatment strategy. New insights in PCNSL tumorigenesis would help to better understand the heterogeneity of outcome and to develop efficient targeted therapies. (C) 2011 Elsevier Masson SAS. All rights reserved.
C1 [Gonzalez-Aguilar, A.; Houillier, C.; Hoang-Xuan, K.] UPMC, Grp Hosp Pitie Salpetriere, AP HP, Serv Neurol Mazarin, F-75651 Paris 13, France.
   [Soussain, C.] Hop R Huguenin, Inst Curie, Dept Hematol, F-92210 St Cloud, France.
   [Houillier, C.; Soussain, C.; Hoang-Xuan, K.] Grp Hosp Pitie Salpetriere, Div Mazarin, Ctr Natl Reference Lymphomes Primitifs SNC Reseau, F-75651 Paris 13, France.
C3 Assistance Publique Hopitaux Paris (APHP); Hopital Universitaire
   Pitie-Salpetriere - APHP; Sorbonne Universite; UNICANCER; Universite
   PSL; Institut Curie; Rene Huguenin Hospital; Sorbonne Universite;
   Assistance Publique Hopitaux Paris (APHP); Hopital Universitaire
   Pitie-Salpetriere - APHP
RP Hoang-Xuan, K (corresponding author), UPMC, Grp Hosp Pitie Salpetriere, AP HP, Serv Neurol Mazarin, 47 Blvd Hop, F-75651 Paris 13, France.
EM
RI González, Alberto/IYJ-8593-2023; Hoang-Xuan, Khe/O-7527-2017
OI Gonzalez Aguilar, Alberto/0000-0001-8916-6258; Hoang Xuan,
   Khe/0000-0003-4072-1999; Houillier, Caroline/0000-0003-2931-7522
CR Abla O, 2011, CLIN CANCER RES, V17, P346, DOI 10.1158/1078-0432.CCR-10-1161
   Abrey LE, 2006, J CLIN ONCOL, V24, P5711, DOI 10.1200/JCO.2006.08.2941
   Abrey LE, 2003, J CLIN ONCOL, V21, P4151, DOI 10.1200/JCO.2003.05.024
   Abrey LE, 1998, J CLIN ONCOL, V16, P859, DOI 10.1200/JCO.1998.16.3.859
   Abrey LE, 2000, J CLIN ONCOL, V18, P3144, DOI 10.1200/JCO.2000.18.17.3144
   Alizadeh AA, 2000, NATURE, V403, P503, DOI 10.1038/35000501
   Angelov L, 2009, J CLIN ONCOL, V27, P3503, DOI 10.1200/JCO.2008.19.3789
   Batchelor T, 2003, J CLIN ONCOL, V21, P1044, DOI 10.1200/JCO.2003.03.036
   Batchelor TT, 2011, NEUROLOGY, V76, P929, DOI 10.1212/WNL.0b013e31820f2d94
   Bessell EM, 2004, INT J RADIAT ONCOL, V59, P501, DOI 10.1016/j.ijrobp.2003.11.001
   Bessell EM, 2002, J CLIN ONCOL, V20, P231, DOI 10.1200/JCO.20.1.231
   BLAY JY, 1995, BLOOD, V86, P2922
   Bohlius J, 2009, ANTIVIR THER, V14, P1065, DOI 10.3851/IMP1462
   Braaten KM, 2003, CLIN CANCER RES, V9, P1063
   Cady FM, 2008, J CLIN ONCOL, V26, P4814, DOI 10.1200/JCO.2008.16.1455
   Camilleri-Broet S, 1998, AM J CLIN PATHOL, V110, P607, DOI 10.1093/ajcp/110.5.607
   Camilleri-Broët S, 2006, BLOOD, V107, P190, DOI 10.1182/blood-2005-03-1024
   Cassoux N, 2007, INVEST OPHTH VIS SCI, V48, P3253, DOI 10.1167/iovs.06-0031
   CHAMBERLAIN MC, 2010, NEURO ONCOL
   Cheng G, 2003, BONE MARROW TRANSPL, V31, P679, DOI 10.1038/sj.bmt.1703917
   Cher L, 1996, NEUROLOGY, V46, P1757, DOI 10.1212/WNL.46.6.1757
   Colombat P, 2006, BONE MARROW TRANSPL, V38, P417, DOI 10.1038/sj.bmt.1705452
   COTE GM, 2011, BIOL BLOOD MARROW TR
   Coulon A, 2002, EUR RADIOL, V12, P329, DOI 10.1007/s003300101037
   Dahlborg S A, 1996, Cancer J Sci Am, V2, P166
   DeAngelis LM, 2011, NAT REV CLIN ONCOL, V8, P196, DOI 10.1038/nrclinonc.2011.18
   DEANGELIS LM, 1992, J CLIN ONCOL, V10, P635, DOI 10.1200/JCO.1992.10.4.635
   DeAngelis LM, 2002, J CLIN ONCOL, V20, P4643, DOI 10.1200/JCO.2002.11.013
   DEANGELIS LM, 1993, ANN NEUROL, V33, P308, DOI 10.1002/ana.410330313
   del Rio MS, 2009, ONCOLOGIST, V14, P526, DOI 10.1634/theoncologist.2008-0236
   Ekenel M, 2008, CANCER-AM CANCER SOC, V113, P1025, DOI 10.1002/cncr.23670
   Enting RH, 2004, NEUROLOGY, V63, P901, DOI 10.1212/01.WNL.0000137050.43114.42
   Ferreri AJM, 2003, J CLIN ONCOL, V21, P266, DOI 10.1200/JCO.2003.09.139
   Ferreri AJM, 2011, LANCET ONCOL, V12, P118, DOI 10.1016/S1470-2045(11)70018-3
   Ferreri AJM, 2009, LANCET, V374, P1512, DOI 10.1016/S0140-6736(09)61416-1
   Fliessbach K, 2005, NEUROLOGY, V64, P1184, DOI 10.1212/01.WNL.0000156350.49336.E2
   Freilich RJ, 1996, NEUROLOGY, V46, P435, DOI 10.1212/WNL.46.2.435
   FRITSCH K, 2011, ANN ONCOL
   Gavrilovic IT, 2006, J CLIN ONCOL, V24, P4570, DOI 10.1200/JCO.2006.06.6910
   Ghesquières H, 2010, ANN ONCOL, V21, P842, DOI 10.1093/annonc/mdp529
   Glass J, 1996, J NEURO-ONCOL, V30, P257
   Guha-Thakurta N, 1999, J NEURO-ONCOL, V43, P259, DOI 10.1023/A:1006210703827
   GUILLEVIN R, 2011, REV NEUROL, DOI DOI 10.1016/J.NEUR0L.2011.07.003
   Harting I, 2003, NEUROSCI LETT, V342, P163, DOI 10.1016/S0304-3940(03)00272-6
   Hartmann M, 2003, NEUROSCI LETT, V338, P119, DOI 10.1016/S0304-3940(02)01367-8
   Herrlinger U, 2005, ANN NEUROL, V57, P843, DOI 10.1002/ana.20495
   Hoang-Xuan K, 2003, J CLIN ONCOL, V21, P2726, DOI 10.1200/JCO.2003.11.036
   Hottinger AF, 2007, NEUROLOGY, V69, P1178, DOI 10.1212/01.wnl.0000276986.19602.c1
   Hottinger AF, 2011, ANN NEUROL, V70, P163, DOI 10.1002/ana.22360
   Illerhaus G, 2006, J CLIN ONCOL, V24, P3865, DOI 10.1200/JCO.2006.06.2117
   Jahnke K, 2009, NEURO-ONCOLOGY, V11, P503, DOI 10.1215/15228517-2008-119
   Juergens A, 2010, ANN NEUROL, V67, P182, DOI 10.1002/ana.21824
   Kadan-Lottick NS, 2002, CANCER, V95, P193, DOI 10.1002/cncr.10643
   Kanai R, 2008, J NEURO-ONCOL, V86, P83, DOI 10.1007/s11060-007-9437-9
   Khan RB, 2002, J NEURO-ONCOL, V58, P175, DOI 10.1023/A:1016077907952
   Lee IH, 2010, EUR J RADIOL, V76, P48, DOI 10.1016/j.ejrad.2009.05.013
   Levy O, 2008, CANCER, V112, P151, DOI 10.1002/cncr.23149
   Lin CH, 2006, CLIN CANCER RES, V12, P1152, DOI 10.1158/1078-0432.CCR-05-1699
   Mangla R, 2011, AM J NEURORADIOL, V32, P1004, DOI 10.3174/ajnr.A2441
   Mathew BS, 2006, CANCER-AM CANCER SOC, V106, P383, DOI 10.1002/cncr.21583
   McAllister LD, 2000, NEUROSURGERY, V46, P51, DOI 10.1093/neurosurgery/46.1.51
   Mohile NA, 2008, NEURO-ONCOLOGY, V10, P223, DOI 10.1215/15228517-2007-061
   Montemurro M, 2007, ANN ONCOL, V18, P665, DOI 10.1093/annonc/mdl458
   Montesinos-Rongen M, 2008, LEUKEMIA, V22, P400, DOI 10.1038/sj.leu.2405019
   Montesinos-Rongen M, 2009, BLOOD, V113, P7, DOI 10.1182/blood-2008-04-149005
   Nakamura M, 2003, CANCER RES, V63, P737
   Nayak L, 2011, NEURO-ONCOLOGY, V13, P525, DOI 10.1093/neuonc/nor014
   NELSON DF, 1992, INT J RADIAT ONCOL, V23, P9, DOI 10.1016/0360-3016(92)90538-S
   NEUWELT EA, 1991, J CLIN ONCOL, V9, P1580, DOI 10.1200/JCO.1991.9.9.1580
   Ney DE, 2010, CANCER-AM CANCER SOC, V116, P4605, DOI 10.1002/cncr.25363
   Nguyen PL, 2005, J CLIN ONCOL, V23, P1507, DOI 10.1200/JCO.2005.01.161
   O'Brien P, 2000, J CLIN ONCOL, V18, P519, DOI 10.1200/JCO.2000.18.3.519
   Olson JE, 2002, CANCER, V95, P1504, DOI 10.1002/cncr.10851
   Omuro A, 2011, J NEURO-ONCOL, V104, P323, DOI 10.1007/s11060-010-0497-x
   Omuro AMP, 2005, NEUROLOGY, V64, P69, DOI 10.1212/01.WNL.0000148641.98241.5E
   Omuro AMP, 2005, ARCH NEUROL-CHICAGO, V62, P1595, DOI 10.1001/archneur.62.10.1595
   Omuro AMP, 2007, J NEURO-ONCOL, V85, P207, DOI 10.1007/s11060-007-9397-0
   ONEILL BP, 1995, INT J RADIAT ONCOL, V33, P663, DOI 10.1016/0360-3016(95)00207-F
   Pels H, 2003, J CLIN ONCOL, V21, P4489, DOI 10.1200/JCO.2003.04.056
   PELS H, 2010, NEURO ONCOL
   Pels H, 2009, J NEURO-ONCOL, V91, P299, DOI 10.1007/s11060-008-9712-4
   Poortmans PMP, 2003, J CLIN ONCOL, V21, P4483, DOI 10.1200/JCO.2003.03.108
   Reni M, 2004, EUR J CANCER, V40, P1682, DOI 10.1016/j.ejca.2004.03.008
   Reni M, 1997, ANN ONCOL, V8, P227, DOI 10.1023/A:1008201717089
   Rubenstein JL, 2006, BLOOD, V107, P3716, DOI 10.1182/blood-2005-03-0897
   Sandor V, 1998, J CLIN ONCOL, V16, P3000, DOI 10.1200/JCO.1998.16.9.3000
   Schultz C, 1996, J CLIN ONCOL, V14, P556, DOI 10.1200/JCO.1996.14.2.556
   Schulz H, 2004, HAEMATOLOGICA, V89, P753
   Shah GD, 2007, J CLIN ONCOL, V25, P4730, DOI 10.1200/JCO.2007.12.5062
   Sierra Del Rio M, 2011, J NEUROONCOL
   SIOBHAN NG, 2000, NEURO-ONCOLOGY, V2, P40, DOI DOI 10.1215/15228517-2-1-40
   Soussain C, 2001, J CLIN ONCOL, V19, P742, DOI 10.1200/JCO.2001.19.3.742
   Soussain C, 2008, J CLIN ONCOL, V26, P2512, DOI 10.1200/JCO.2007.13.5533
   Soussain C, 2007, CLIN CANCER RES, V13, P2504, DOI 10.1158/1078-0432.CCR-06-2379
   Soussain C, 2009, CURR OPIN ONCOL, V21, P550, DOI 10.1097/CCO.0b013e3283310eb3
   Sung CO, 2011, BLOOD, V117, P1291, DOI 10.1182/blood-2010-07-297861
   Taillibert S, 2008, J NEURO-ONCOL, V86, P225, DOI 10.1007/s11060-007-9468-2
   Thiel E, 2010, LANCET ONCOL, V11, P1036, DOI 10.1016/S1470-2045(10)70229-1
   Tun HW, 2008, BLOOD, V111, P3200, DOI 10.1182/blood-2007-10-119099
   Zhu JJ, 2009, NEURO-ONCOLOGY, V11, P211, DOI 10.1215/15228517-2008-067
NR 100
TC 2
Z9 2
U1 0
U2 0
PU MASSON EDITEUR
PI MOULINEAUX CEDEX 9
PA 21 STREET CAMILLE DESMOULINS, ISSY, 92789 MOULINEAUX CEDEX 9, FRANCE
SN 0035-3787
EI 2213-0004
J9 REV NEUROL-FRANCE
JI Rev. Neurol.
PD OCT
PY 2011
VL 167
IS 10
BP 721
EP 728
DI 10.1016/j.neurol.2011.08.004
PG 8
WC Clinical Neurology
WE Science Citation Index Expanded (SCI-EXPANDED)
SC Neurosciences & Neurology
GA 835IO
UT WOS:000296030200013
PM 21906768
DA 2024-06-03
ER

PT J
AU Del Rio, MS
   Benouaich-Amiel, A
   Psimaras, D
   Dehais, C
   Hoang-Xuan, K
AF Del Rio, M. Sierra
   Benouaich-Amiel, A.
   Psimaras, D.
   Dehais, C.
   Hoang-Xuan, K.
TI Treatment of primary central nervous system lymphoma in the
   immunocompetent patient
SO REVUE NEUROLOGIQUE
LA French
DT Review
DE lymphoma; central nervous system; immunocompetent; steroids;
   radiotherapy; chemotherapy; methotrexate; neurotoxicity
ID PRIMARY CNS LYMPHOMA; HIGH-DOSE CHEMOTHERAPY; NON-HODGKINS-LYMPHOMA;
   METHOTREXATE-BASED CHEMOTHERAPY; MULTICENTER PHASE-II; LONG-TERM
   SURVIVAL; STEM-CELL RESCUE; RADIATION-THERAPY; DEFERRED RADIOTHERAPY;
   INITIAL TREATMENT
AB The incidence of primary CNS lymphoma (PCNSL), which has considerably increased these last years, remains stable in the immunocompetent population, while it is steadily decreasing in the immunosuppressed population. The addition of high dose methotrexate (MTX) based chemotherapy (CT) before whole brain radiotherapy (WBRT) has clearly improved the prognosis of PCNSL with a median survival of three to four years. About 30% of the patients may hope to have a long survival and to be cured. in the elderly (age over 60 years) CT alone (without RT) is recommended as initial treatment. This approach seems useful to avoid RT and to reduce the risk of delayed neurotoxicity due to the combined treatment. in the young population (age less than 60 years), intensive chemotherapy followed by hematopoietic stem cell rescue (ICH) appears as a promising approach in recurrent tumors and potentially as an alternative option to RT as consolidation treatment in newly diagnosed patients. A prospective trial will be activated in France soon randomizing ICH and RT in the initial treatment of PCNSL. (C) 2008 Publie par Elsevier Masson SAS.
C1 [Del Rio, M. Sierra; Benouaich-Amiel, A.; Psimaras, D.; Dehais, C.; Hoang-Xuan, K.] CHU Pitie Salpetriere, Serv Neurol Mazarin, F-75651 Paris, France.
C3 Assistance Publique Hopitaux Paris (APHP); Hopital Universitaire
   Pitie-Salpetriere - APHP; Sorbonne Universite
RP Hoang-Xuan, K (corresponding author), CHU Pitie Salpetriere, Serv Neurol Mazarin, 47 Blvd Hop, F-75651 Paris, France.
EM
RI Hoang-Xuan, Khe/O-7527-2017
OI psimaras, dimitri/0000-0002-6641-288X; Hoang Xuan,
   Khe/0000-0003-4072-1999
CR Abrey LE, 2003, J CLIN ONCOL, V21, P4151, DOI 10.1200/JCO.2003.05.024
   Abrey LE, 1998, J CLIN ONCOL, V16, P859, DOI 10.1200/JCO.1998.16.3.859
   Abrey LE, 2000, J CLIN ONCOL, V18, P3144, DOI 10.1200/JCO.2000.18.17.3144
   Batchelor T, 2003, J CLIN ONCOL, V21, P1044, DOI 10.1200/JCO.2003.03.036
   Bessell EM, 2004, INT J RADIAT ONCOL, V59, P501, DOI 10.1016/j.ijrobp.2003.11.001
   Bessell EM, 2002, J CLIN ONCOL, V20, P231, DOI 10.1200/JCO.20.1.231
   Bessell EM, 2007, EUR J CANCER, V43, P1141, DOI 10.1016/j.ejca.2006.12.011
   BLAY JY, 1995, BLOOD, V86, P2922
   Brada M, 1998, INT J RADIAT ONCOL, V40, P1157, DOI 10.1016/S0360-3016(98)00002-9
   Camilleri-Broët S, 2006, BLOOD, V107, P190, DOI 10.1182/blood-2005-03-1024
   Cheng G, 2003, BONE MARROW TRANSPL, V31, P679, DOI 10.1038/sj.bmt.1703917
   Cher L, 1996, NEUROLOGY, V46, P1757, DOI 10.1212/WNL.46.6.1757
   Colombat P, 2006, BONE MARROW TRANSPL, V38, P417, DOI 10.1038/sj.bmt.1705452
   Dahlborg S A, 1996, Cancer J Sci Am, V2, P166
   DEANGELIS LM, 1992, J CLIN ONCOL, V10, P635, DOI 10.1200/JCO.1992.10.4.635
   DeAngelis LM, 2002, J CLIN ONCOL, V20, P4643, DOI 10.1200/JCO.2002.11.013
   Freilich RJ, 1996, NEUROLOGY, V46, P435, DOI 10.1212/WNL.46.2.435
   GABBAI AA, 1989, J NEUROSURG, V70, P190, DOI 10.3171/jns.1989.70.2.0190
   Glass J, 1996, J NEURO-ONCOL, V30, P257
   GLASS J, 1994, J NEUROSURG, V81, P188, DOI 10.3171/jns.1994.81.2.0188
   Guha-Thakurta N, 1999, J NEURO-ONCOL, V43, P259, DOI 10.1023/A:1006210703827
   Herrlinger U, 2005, ANN NEUROL, V57, P843, DOI 10.1002/ana.20495
   Hoang-Xuan K, 2003, SEMIN ONCOL, V30, P53, DOI 10.1053/j.seminoncol.2003.11.032
   Hoang-Xuan K, 2004, CURR OPIN ONCOL, V16, P601, DOI 10.1097/01.cco.0000142927.88773.7e
   Hoang-Xuan K, 2003, J CLIN ONCOL, V21, P2726, DOI 10.1200/JCO.2003.11.036
   Hoang-Xuan K, 1998, NEUROLOGY, V50, pA353
   Illerhaus G, 2006, J CLIN ONCOL, V24, P3865, DOI 10.1200/JCO.2006.06.2117
   Kadan-Lottick NS, 2002, CANCER, V95, P193, DOI 10.1002/cncr.10643
   LOUIS E, 2005, ENCY MEDICOCHIRURGIC
   McAllister LD, 2000, NEUROSURGERY, V46, P51, DOI 10.1093/neurosurgery/46.1.51
   NELSON DF, 1992, INT J RADIAT ONCOL, V23, P9, DOI 10.1016/0360-3016(92)90538-S
   NEUWELT EA, 1991, J CLIN ONCOL, V9, P1580, DOI 10.1200/JCO.1991.9.9.1580
   O'Brien P, 2000, J CLIN ONCOL, V18, P519, DOI 10.1200/JCO.2000.18.3.519
   Omuro AMP, 2005, ARCH NEUROL-CHICAGO, V62, P1595, DOI 10.1001/archneur.62.10.1595
   Omuro AMP, 2007, J NEURO-ONCOL, V85, P207, DOI 10.1007/s11060-007-9397-0
   ONEILL BP, 1995, INT J RADIAT ONCOL, V33, P663, DOI 10.1016/0360-3016(95)00207-F
   Pels H, 2003, J CLIN ONCOL, V21, P4489, DOI 10.1200/JCO.2003.04.056
   Pirotte B, 1997, J NEURO-ONCOL, V32, P63, DOI 10.1023/A:1005733416571
   Poortmans PMP, 2003, J CLIN ONCOL, V21, P4483, DOI 10.1200/JCO.2003.03.108
   Reni M, 1997, ANN ONCOL, V8, P227, DOI 10.1023/A:1008201717089
   Sandor V, 1998, J CLIN ONCOL, V16, P3000, DOI 10.1200/JCO.1998.16.9.3000
   Schultz C, 1996, J CLIN ONCOL, V14, P556, DOI 10.1200/JCO.1996.14.2.556
   SIOBHAN NG, 2000, NEURO-ONCOLOGY, V2, P40, DOI DOI 10.1215/15228517-2-1-40
   Soussain C, 2001, J CLIN ONCOL, V19, P742, DOI 10.1200/JCO.2001.19.3.742
   SOUSSAIN C, 2008, J CLIN ONCOL
NR 45
TC 2
Z9 2
U1 0
U2 0
PU MASSON EDITEUR
PI MOULINEAUX CEDEX 9
PA 21 STREET CAMILLE DESMOULINS, ISSY, 92789 MOULINEAUX CEDEX 9, FRANCE
SN 0035-3787
EI 2213-0004
J9 REV NEUROL-FRANCE
JI Rev. Neurol.
PD JUN-JUL
PY 2008
VL 164
IS 6-7
BP 569
EP 574
DI 10.1016/j.neurol.2008.04.001
PG 6
WC Clinical Neurology
WE Science Citation Index Expanded (SCI-EXPANDED)
SC Neurosciences & Neurology
GA 326NS
UT WOS:000257666800010
PM 18565356
DA 2024-06-03
ER

PT J
AU Rujirojindakul, P
   Rohitoprakarn, M
   Kongkabpan, D
   Viboonjantra, P
   Lekhakula, A
AF Rujirojindakul, Pairaya
   Rohitoprakarn, Montanan
   Kongkabpan, Daolada
   Viboonjantra, Pongthep
   Lekhakula, Arnuparp
TI Incidence and radiographic findings of primary central nervous system
   lymphoma in immunocompetent patients in southern Thailand
SO TUMORI JOURNAL
LA English
DT Article
DE incidence; manifestation; CT; MRI; radiation; outcome
ID PRIMARY CNS LYMPHOMA; PRIMARY MALIGNANT-LYMPHOMA; PHASE-II;
   CHEMOTHERAPY; METHOTREXATE; RADIOTHERAPY; COMBINATION; RITUXIMAB;
   FEATURES; THERAPY
AB Aims and background. In the last two decades there have been conflicting reports concerning a rising incidence of primary central nervous system lymphoma (PCNSL) in immunocompetent patients. This study is being conducted to review the incidence and radiographic findings of PCNSL in a large tertiary care institution in southern Thailand.
   Patients. A review was carried out in Songklanagarind University Hospital of the clinical records, CT and MRI images of immunocompetent patients who had histologically proven PCNSL diagnosed between January 1993 and December 2004.
   Results. A total of 25 PCNSL patients were diagnosed over a 12-year period. The incidence trend was rising but not to a statistically significant extent. The median age at diagnosis was 57.4 years. Based on the Working Formulation classification, diffuse large cell was the most common subtype. All of the 20 patients with available immunohistochemical results had B-cell lymphomas. The radiographic findings in our series show that the majority of patients had a single lesion (60%) at a supratentorial location (50%). Most of the lesions were well circumscribed. Based on the density of the lesions before administration of a contrast medium, 56% of the lesions on CT images appeared hyperdense. On T1-weighted MRI images, the lesions of our patients were most frequently hypointense (67%) while the T2-weighted images were more commonly hyperintense (60%) and contrast enhancement was found in all lesions. There were 21 patients who received whole brain radiotherapy with doses ranging between 4.2 and 6.0 Gy. The median survival time was 14.4 months.
   Conclusions. This study indicates that there has been no significant increase in PCNSL cases over the last 12 years. The recognition of characteristic imaging features of PCNSL may facilitate a stereotactic procedure before steroid administration. In addition, we provide evidence that radiotherapy alone is insufficient in PCNSL.
C1 [Rujirojindakul, Pairaya; Kongkabpan, Daolada; Viboonjantra, Pongthep; Lekhakula, Arnuparp] Prince Songk1a Univ, Fac Med, Dept Med, Hat Yai 90110, Songkhla, Thailand.
   [Rohitoprakarn, Montanan] Prince Songk1a Univ, Fac Med, Dept Radiol, Hat Yai 90110, Songkhla, Thailand.
C3 Prince of Songkla University; Prince of Songkla University
RP Rujirojindakul, P (corresponding author), Prince Songk1a Univ, Fac Med, Dept Med, Hat Yai 90110, Songkhla, Thailand.
EM
OI Lekhakula, Arnuparp/0000-0002-0196-8107
CR Abrey LE, 1998, J CLIN ONCOL, V16, P859, DOI 10.1200/JCO.1998.16.3.859
   Abrey LE, 2000, J CLIN ONCOL, V18, P3144, DOI 10.1200/JCO.2000.18.17.3144
   BATABYAL AA, 2000, INT J ECOLOGY ENV SC, V26, P1
   Batchelor T, 2003, J CLIN ONCOL, V21, P1044, DOI 10.1200/JCO.2003.03.036
   Bellinzona M, 2005, EJSO, V31, P100, DOI 10.1016/j.ejso.2004.10.002
   Bühring U, 2001, NEUROLOGY, V57, P393, DOI 10.1212/WNL.57.3.393
   Coulon A, 2002, EUR RADIOL, V12, P329, DOI 10.1007/s003300101037
   DEANGELIS LM, 1990, NEUROLOGY, V40, P80, DOI 10.1212/WNL.40.1.80
   DeAngelis LM, 2002, J CLIN ONCOL, V20, P4643, DOI 10.1200/JCO.2002.11.013
   Enting RH, 2004, NEUROLOGY, V63, P901, DOI 10.1212/01.WNL.0000137050.43114.42
   Ferreri AJM, 2004, BRIT J CANCER, V90, P353, DOI 10.1038/sj.bjc.6601472
   FINE HA, 1993, ANN INTERN MED, V119, P1093, DOI 10.7326/0003-4819-119-11-199312010-00007
   Hao D, 1999, ANN ONCOL, V10, P65, DOI 10.1023/A:1008308313914
   HENRY JM, 1974, CANCER, V34, P1293, DOI 10.1002/1097-0142(197410)34:4<1293::AID-CNCR2820340441>3.0.CO;2-P
   Hoang-Xuan K, 2004, CURR OPIN ONCOL, V16, P601, DOI 10.1097/01.cco.0000142927.88773.7e
   Johnson BA, 1997, AM J NEURORADIOL, V18, P563
   Kadan-Lottick NS, 2002, CANCER, V95, P193, DOI 10.1002/cncr.10643
   Küker W, 2005, NEUROLOGY, V65, P1129, DOI 10.1212/01.wnl.0000178894.51436.54
   Maher EA, 1999, SEMIN ONCOL, V26, P346
   MILLER DC, 1994, CANCER-AM CANCER SOC, V74, P1383, DOI 10.1002/1097-0142(19940815)74:4<1383::AID-CNCR2820740432>3.0.CO;2-1
   MURRAY K, 1986, J NEUROSURG, V65, P600, DOI 10.3171/jns.1986.65.5.0600
   Olson JE, 2002, CANCER, V95, P1504, DOI 10.1002/cncr.10851
   Pels H, 2003, J CLIN ONCOL, V21, P4489, DOI 10.1200/JCO.2003.04.056
   Poortmans PMP, 2003, J CLIN ONCOL, V21, P4483, DOI 10.1200/JCO.2003.03.108
   Powari M, 2002, SURG NEUROL, V57, P113, DOI 10.1016/S0090-3019(01)00675-9
   Rubenstein JL, 2007, J CLIN ONCOL, V25, P1350, DOI 10.1200/JCO.2006.09.7311
   Takahashi W, 2005, REMOTE SENS ENVIRON, V97, P83, DOI 10.1016/j.rse.2005.04.019
   Ueda F, 1995, Radiat Med, V13, P51
   WATANABE M, 1992, NEURORADIOLOGY, V34, P36, DOI 10.1007/BF00588431
NR 29
TC 2
Z9 2
U1 0
U2 0
PU SAGE PUBLICATIONS LTD
PI LONDON
PA 1 OLIVERS YARD, 55 CITY ROAD, LONDON EC1Y 1SP, ENGLAND
SN 0300-8916
EI 2038-2529
J9 TUMORI J
JI Tumori J.
PD MAY-JUN
PY 2008
VL 94
IS 3
BP 304
EP 308
PG 5
WC Oncology
WE Science Citation Index Expanded (SCI-EXPANDED)
SC Oncology
GA 329AE
UT WOS:000257838500003
PM 18705395
DA 2024-06-03
ER

PT J
AU Günaydin, A
   Er, U
   Acuduman, A
   Sabuncuoglu, H
   Öztürk, E
AF Gunaydin, Ahmet
   Er, Uygur
   Acuduman, Ahmet
   Sabuncuoglu, Hakan
   Ozturk, Evrim
TI Diagnostic and surgical pitfalls of an unusual primary central nervous
   system lymphoma
SO TURKISH NEUROSURGERY
LA English
DT Article
DE primary CNS lymphoma; craniotomy; pathology; spontaneous remission;
   tumor regression
ID PRIMARY CNS LYMPHOMA; DURAL TAIL SIGN; CEREBRAL LYMPHOMA; TUMOR;
   LESIONS; BRAIN
AB BACKGROUND: Here we describe a case of PCNSL which was located peripherally as a single lesion and showed no evidence of pathological findings of lymphoma at the first biopsy.
   CASE DESCRIPTION: A 56-year-old man was admitted to the hospital with a talking disorder and attention deficit. The neurological examination was normal. There was a left temporoparietal, 24x20 mm. enhanced lesion on MRI. The patient underwent a craniotomy and the lesion was excised totally. The pathological examination revealed glial tissue which showed reactive astrocyte proliferations. A month later, an MRI was performed and a recurrent tumor was seen near the first lesion's location. The second operation was performed via the same craniotomy, the tumor was excised totally and the second pathological examination revealed diffuse large-cell, B-lymphoma. There were no pathological findings on abdominal, thoracic and bone marrow investigations.
   CONCLUSION: PCNSL may show various biological behaviors. Using steroids before the biopsy may lead to diagnostic and therapeutic failure.
C1 [Gunaydin, Ahmet; Acuduman, Ahmet] Etlik Specializat Hosp, Neurosurg Clin, Ankara, Turkey.
   [Er, Uygur] Yildirim Bayezit Training & Res Hosp, Neurosurg Clin 2, Ankara, Turkey.
   [Ozturk, Evrim] Yildirim Bayezit Training & Res Hosp, Pathol Clin, Ankara, Turkey.
   [Sabuncuoglu, Hakan] Ufuk Univ, Neurosurg Clin, Ankara, Turkey.
C3 Ankara Etlik Ihtisas Egitim ve Arastirma Hastanesi; Diskapi Yildirim
   Beyazit Training & Research Hospital; Diskapi Yildirim Beyazit Training
   & Research Hospital; Ufuk University
RP Er, U (corresponding author), 4 Sokak,22-7 Sogutozu, TR-06470 Ankara, Turkey.
EM
CR Al-Yamany M, 1999, J NEURO-ONCOL, V42, P151, DOI 10.1023/A:1006162615774
   Alderson L, 1996, J NEUROL NEUROSUR PS, V60, P102, DOI 10.1136/jnnp.60.1.102
   Bromberg JEC, 2002, NEUROLOGY, V59, P762, DOI 10.1212/WNL.59.5.762
   Cartmill M, 2000, BRIT J NEUROSURG, V14, P313
   Corry J, 1998, INT J RADIAT ONCOL, V41, P615, DOI 10.1016/S0360-3016(97)00571-3
   DeAngelis LM, 2003, J CLIN ONCOL, V21, P4471, DOI 10.1200/JCO.2003.08.900
   Guermazi A, 2005, CLIN RADIOL, V60, P171, DOI 10.1016/j.crad.2004.01.019
   Jaiswal A K, 2000, J Neurosurg Sci, V44, P145
   Jaiswal AK, 2004, J CLIN NEUROSCI, V11, P328, DOI 10.1016/S0967-5868(03)00110-3
   KAWAKAMI Y, 1985, J NEUROSURG, V62, P522, DOI 10.3171/jns.1985.62.4.0522
   Kawamura T, 2001, J CLIN NEUROSCI, V8, P574, DOI 10.1054/jocn.2000.0854
   KIKUCHI K, 1986, SURG NEUROL, V26, P291, DOI 10.1016/0090-3019(86)90165-5
   Pels H, 2004, NEUROLOGY, V63, P167, DOI 10.1212/01.WNL.0000132649.24618.8A
   Rokni-Yazdi H, 2006, EUR J RADIOL, V60, P42, DOI 10.1016/j.ejrad.2006.04.003
   RUSSELL DS, 1989, PATHOLOGY TUMOURS NE, P590
   SPILLANE JA, 1982, J NEUROL NEUROSUR PS, V45, P199, DOI 10.1136/jnnp.45.3.199
   UMUR AS, 2003, TURKISH NEUROSURGERY, V13, P122
   VAQUERO J, 1984, J NEUROSURG, V60, P174, DOI 10.3171/jns.1984.60.1.0174
NR 18
TC 2
Z9 2
U1 0
U2 0
PU TURKISH NEUROSURGICAL SOC
PI BAHCELIEVLER
PA TASKENT CADDESI 13-4, BAHCELIEVLER, ANKARA 06500, TURKEY
SN 1019-5149
J9 TURK NEUROSURG
JI Turk. Neurosurg.
PD APR
PY 2007
VL 17
IS 2
BP 129
EP 133
PG 5
WC Clinical Neurology; Surgery
WE Science Citation Index Expanded (SCI-EXPANDED)
SC Neurosciences & Neurology; Surgery
GA 314EQ
UT WOS:000256792700011
PM 17935030
DA 2024-06-03
ER

PT J
AU Moradi, A
   Tajedini, A
   Mehrabian, A
   Sadeghi, S
   Semnani, V
   Khodabakhshi, R
   Arefian, N
   Afrakhteh, M
   Keshvari, K
   Yavari, P
   Madani-Civi, M
AF Moradi, Afshin
   Tajedini, Aram
   Mehrabian, Abbasali
   Sadeghi, Sohrab
   Semnani, Vahid
   Khodabakhshi, Reza
   Arefian, Noormohammad
   Afrakhteh, Maryam
   Keshvari, Kayvan
   Yavari, Parvin
   Madani-Civi, Manouchehr
TI Clinicopathological features of primary central nervous system lymphoma
SO NEUROSCIENCES
LA English
DT Article
ID INTRACEREBRAL MALIGNANT-LYMPHOMA; CLINICAL PRESENTATION; BRAIN LYMPHOMA;
   CNS-LYMPHOMA; EPIDEMIOLOGY; TRENDS
AB Objectives: To investigate the anatomic location, immunologic, and clinicopathological features of patients with primary central nervous system lymphoma (PCNSL).
   Methods: From May 1993 to December 2004, at Shohada Hospital, Tehran, Iran, the clinical data of 110 PCNSL patients, including the age, sex, duration of symptoms, radiological findings, site of tumors, immune status, and history of immunocompromised state (such as organ transplantation, radiotherapy, steroid therapy or AIDS) were assessed.
   Results: The mean age of the patients with PCNSL was 47.02 +/- 15.8 years. There were 42 female and 68 male patients. One hundred and six cases (96.3%) were diagnosed as B-cell lymphoma. Most of the PCNSL in our study are unifocal. More than 70% of tumors were in a cerebral hemisphere and periventricular location, usually involving the corpus callosum or basal ganglia. No patients had been in immunocompromised states. Symptoms of increased intracranial pressure or changes in personality, vision, or motor function are most common. Seizures are seen in approximately 10% of patients. The number of PCNSL cases showed a gradual rise in incidence.
   Conclusion: The results of this single hospital 12-year survey of PCNSL are in agreement with data from other single institutions and regional surveys concerning clinical features. However, in contrast with the literature, most of our patients were immunocompetent. The age at diagnosis is also lower than in most reports.
C1 Shahid Beheshti Univ Med Sci, Dept Pathol, Shohada Hosp, Tehran, Iran.
   Shahid Beheshti Univ Med Sci, Dept Internal Med, Shohada Hosp, Tehran, Iran.
   Shahid Beheshti Univ Med Sci, Dept Neurosurg, Shohada Hosp, Tehran, Iran.
   Shahid Beheshti Univ Med Sci, Dept Oncol, Shohada Hosp, Tehran, Iran.
   Shahid Beheshti Univ Med Sci, Dept Anesthesiol, Shohada Hosp, Tehran, Iran.
   Shahid Beheshti Univ Med Sci, Dept Obstet & Gynecol, Shohada Hosp, Tehran, Iran.
   Shahid Beheshti Univ Med Sci, Dept Epidemiol, Shohada Hosp, Tehran, Iran.
   Univ Tehran Med Sci, Res Dept, THC Hosp, Tehran, Iran.
   Semnan Univ Med Sci, Dept Pathol, Semnan, Iran.
C3 Shahid Beheshti University Medical Sciences; Shahid Beheshti University
   Medical Sciences; Shahid Beheshti University Medical Sciences; Shahid
   Beheshti University Medical Sciences; Shahid Beheshti University Medical
   Sciences; Shahid Beheshti University Medical Sciences; Shahid Beheshti
   University Medical Sciences; Tehran University of Medical Sciences;
   Semnan University of Medical Sciences
RP Moradi, A (corresponding author), Shahid Beheshti Univ Med Sci, Dept Pathol, Shohada Hosp, POB 16765-3156, Tehran, Iran.
EM
RI afrakhteh, maryam/AAU-9989-2021; arefian, noormohammad/P-5160-2019;
   Sadeghi, Sohrab/ABC-2330-2020
CR Basso U, 2002, EUR J CANCER, V38, P1298, DOI 10.1016/S0959-8049(02)00031-X
   Bataille B, 2000, J NEUROSURG, V92, P261, DOI 10.3171/jns.2000.92.2.0261
   BRAUS DF, 1992, J NEUROL, V239, P117, DOI 10.1007/BF00833908
   Bromberg JEC, 2002, NEUROLOGY, V59, P762, DOI 10.1212/WNL.59.5.762
   Corn BW, 1997, CANCER, V79, P2409
   Cote TR, 1996, J NATL CANCER I, V88, P675, DOI 10.1093/jnci/88.10.675
   EBY NL, 1988, CANCER-AM CANCER SOC, V62, P2461, DOI 10.1002/1097-0142(19881201)62:11<2461::AID-CNCR2820621135>3.0.CO;2-M
   Ferreri AJM, 2003, J CLIN ONCOL, V21, P2407, DOI 10.1200/JCO.2003.01.135
   FINE HA, 1993, ANN INTERN MED, V119, P1093, DOI 10.7326/0003-4819-119-11-199312010-00007
   Gliemroth J, 2003, CLIN NEUROL NEUROSUR, V105, P78, DOI 10.1016/S0303-8467(02)00105-1
   HAYAKAWA T, 1994, J NEURO-ONCOL, V19, P197, DOI 10.1007/BF01053274
   HENRY JM, 1974, CANCER, V34, P1293, DOI 10.1002/1097-0142(197410)34:4<1293::AID-CNCR2820340441>3.0.CO;2-P
   Herrlinger U, 1999, J NEURO-ONCOL, V43, P219, DOI 10.1023/A:1006298201101
   HOCHBERG FH, 1988, J NEUROSURG, V68, P835, DOI 10.3171/jns.1988.68.6.0835
   LACHANCE DH, 1994, NEUROLOGY, V44, P1721, DOI 10.1212/WNL.44.9.1721
   LUTZ JM, 1994, BRIT J CANCER, V70, P716, DOI 10.1038/bjc.1994.381
   Martinez AJ, 1998, PATHOL RES PRACT, V194, P473, DOI 10.1016/S0344-0338(98)80116-7
   MILLER DC, 1994, CANCER-AM CANCER SOC, V74, P1383, DOI 10.1002/1097-0142(19940815)74:4<1383::AID-CNCR2820740432>3.0.CO;2-1
   Olson JE, 2002, CANCER, V95, P1504, DOI 10.1002/cncr.10851
   Schlegel U, 2000, J NEUROL SCI, V181, P1, DOI 10.1016/S0022-510X(00)00385-3
   TOMLINSON FH, 1995, J NEUROSURG, V82, P558, DOI 10.3171/jns.1995.82.4.0558
NR 21
TC 2
Z9 2
U1 0
U2 0
PU RIYADH ARMED FORCES HOSPITAL
PI RIYADH
PA PO BOX 7897, RIYADH, KSA 11159, SAUDI ARABIA
SN 1319-6138
J9 NEUROSCIENCES
JI Neurosciences
PD OCT
PY 2006
VL 11
IS 4
BP 284
EP 288
PG 5
WC Clinical Neurology
WE Science Citation Index Expanded (SCI-EXPANDED)
SC Neurosciences & Neurology
GA 131BV
UT WOS:000243843900010
PM 22266438
DA 2024-06-03
ER

PT J
AU Alécio-Mattei, T
   Alécio-Mattei, J
AF Alécio-Mattei, T
   Alécio-Mattei, J
TI Primary central nervous system lymphomas in immunocompetent patients
SO NEUROCIRUGIA
LA English
DT Article
DE brain lymphoma; immunocompetent; malignant lymphoma; PCNLS
ID B-CELL LYMPHOMA; SURVIVAL; CYCLOPHOSPHAMIDE; VINCRISTINE; DOXORUBICIN;
   CHEMOTHERAPY; RADIOTHERAPY; NEOPLASM; LESIONS; BIOPSY
AB Objectives. Primary central nervous system lymphoma (PCNSL) is a rare patology and is most often seen in immunodeficient patients. This article presents our casuistic of PCNSL in immunocompetent patients and make a literature review on this issue with focus on recent advances, investigations, and controversies in diagnosis and management of this patology.
   Matherial and methods. Nine patients operated in the last years in our clinics are analysed in relation to sex, age, time of symptoms, procedures and adjuvant treatment. Posteriorly the results are compared with those in the preview literature.
   Results. The age ranged from 44-68 years (middle of 66 years); 62,5% of the patients were female. The mean time of symptoms, when the diagnostic was made, was 3.2 months (range 1-6 months). The most common symptoms were hemiparesis (present in 75% of the patients) headache (37.5%) seizures (25%) and ataxia (25%). The most common localization was the parietal, frontal and temporal lobe surface with 25% of the patients for each localization. Five patients (50%) were treated with stereotactic biopsy, three with surgery (37.5%) and one (12.5%) with both of them. Five patients (62,5%) were submitted to pos-operative radiotherapy as adjuvant treatment. In relation to the hystology, the most common cell type was Difuse B Cells.
   Conclusions. This study demonstrated that complete surgical ressection followed by radiotherapy have shown good results. In opposition to the literature, the authors regards chemotherapy as a secondary line treatment and recommends its use only in some selected cases.
C1 Pinheiros Neurosurg Clin, Sao Paulo, Brazil.
   Inst Neurol, Curitiba, Parana, Brazil.
RP Rua Capote Valente 725 Ap 302, BR-05409002 Sao Paulo, Brazil.
OI Pires de Aguiar, Paulo Henrique/0000-0002-9059-4978
CR Altundag MK, 2000, J EXP CLIN CANC RES, V19, P249
   Bataille B, 2000, J NEUROSURG, V92, P261, DOI 10.3171/jns.2000.92.2.0261
   Bataille B, 1997, NEUROCHIRURGIE, V43, P385
   Bataille B, 1997, NEUROCHIRURGIE, V43, P372
   Bojanowski K, 2001, Neurol Neurochir Pol, V35, P91
   CHAMBERLAIN MC, 1992, J NEURO-ONCOL, V14, P271
   Chamberlain MC, 1998, ONCOL REP, V5, P521
   Cheuk W, 2001, AM J SURG PATHOL, V25, P1372, DOI 10.1097/00000478-200111000-00004
   Corn BW, 2000, INT J RADIAT ONCOL, V47, P299, DOI 10.1016/S0360-3016(00)00470-3
   Cuny E, 2002, ACTA NEUROCHIR, V144, P357, DOI 10.1007/s007010200049
   Cuny E, 1998, NEUROCHIRURGIE, V44, P19
   Delwail V, 1997, NEUROCHIRURGIE, V43, P380
   Fewings PE, 2002, CLIN NEUROPATHOL, V21, P243
   Gildenberg PL, 2000, CLIN INFECT DIS, V30, P491, DOI 10.1086/313685
   Gliemroth J, 2003, CLIN NEUROL NEUROSUR, V105, P78, DOI 10.1016/S0303-8467(02)00105-1
   Hayabuchi N, 1999, INT J RADIAT ONCOL, V44, P265, DOI 10.1016/S0360-3016(98)00564-1
   Kolasa P, 2002, FOLIA NEUROPATHOL, V40, P107
   Mead GM, 2000, CANCER, V89, P1359, DOI 10.1002/1097-0142(20000915)89:6<1359::AID-CNCR21>3.0.CO;2-9
   Monabati A, 2002, J NEUROSURG, V96, P1127, DOI 10.3171/jns.2002.96.6.1127
   Park Deric Minwoo, 2002, Expert Opin Pharmacother, V3, P39, DOI 10.1517/14656566.3.1.39
   Paueksakon P, 2003, HUM PATHOL, V34, P720, DOI 10.1016/S0046-8177(03)00192-8
   Sawin PD, 1998, SURG NEUROL, V49, P640, DOI 10.1016/S0090-3019(97)00435-7
   Shibamoto Y, 1999, J NEURO-ONCOL, V42, P161, DOI 10.1023/A:1006106530795
   SHIBAMOTO Y, 1990, CANCER-AM CANCER SOC, V65, P1907, DOI 10.1002/1097-0142(19900501)65:9<1907::AID-CNCR2820650906>3.0.CO;2-L
   Singhal D, 2002, CAN J NEUROL SCI, V29, P147, DOI 10.1017/S0317167100120918
   Sonstein W, 1998, MED ONCOL, V15, P61, DOI 10.1007/BF02787347
   Spinelli F, 2000, EUR J NUCL MED, V27, P46, DOI 10.1007/PL00006661
   Takeshita M, 1999, NEUROL MED-CHIR, V39, P452, DOI 10.2176/nmc.39.452
   Tseng SH, 2003, ACTA NEUROL SCAND, V108, P43, DOI 10.1034/j.1600-0404.2003.00080.x
   Uchino Masafumi, 2003, No To Shinkei, V55, P273
   Yang J, 1996, Zhonghua Wai Ke Za Zhi, V34, P102
   Yu H, 2001, Zhonghua Nei Ke Za Zhi, V40, P325
NR 32
TC 2
Z9 5
U1 0
U2 1
PU ELSEVIER ESPANA SLU
PI BARCELONA
PA AV JOSEP TARRADELLAS, 20-30, 1ERA PLANTA, BARCELONA, CP-08029, SPAIN
SN 1130-1473
EI 2340-6305
J9 NEUROCIRUGIA
JI Neurocirugia
PD FEB
PY 2006
VL 17
IS 1
BP 46
EP 53
PG 8
WC Neurosciences; Surgery
WE Science Citation Index Expanded (SCI-EXPANDED)
SC Neurosciences & Neurology; Surgery
GA 038HK
UT WOS:000237209700009
PM 16565780
DA 2024-06-03
ER

PT J
AU Park, S
   Leymarie, V
   Agbalika, F
   Galicier, L
   Oksenhendler, E
   Sigaux, F
   Noguera, ME
AF Park, S
   Leymarie, V
   Agbalika, F
   Galicier, L
   Oksenhendler, E
   Sigaux, F
   Noguera, ME
TI Varicella-Zoster viral meningitis mimicking lymphoma
SO LEUKEMIA & LYMPHOMA
LA English
DT Article
DE Varicella-Zoster Virus; meningitis; lymphoma; PCR
ID HUMAN-IMMUNODEFICIENCY-VIRUS; POLYMERASE CHAIN-REACTION;
   CENTRAL-NERVOUS-SYSTEM; CEREBROSPINAL-FLUID; INFECTED PATIENTS
AB We report the case of a 30-year-old HIV-infected man admitted for a meningeal syndrome and a zoster rash. The CSF had cytological features suggesting a primary CNS lymphoma (PCNSL). The large lymphoid cells had a fine chromatin with nucleoli, a basophilic cytoplasm with azurophilic granules and high mitotic activity. Several arguments demonstrated the viral origin of the meningitis: the large lymphoid cells were of T origin with no evidence of clonal TCR gamma gene rearrangement. The PCR was positive for Varicella-Zoster Virus (VZV) and EBV DNA. Clinical evolution was favorable under acyclovir. We should be cautious in the differential diagnosis between viral meningitis and PCNSL.
C1 Hop St Louis, Hematol Lab, F-75010 Paris, France.
   Hop St Louis, Serv Microbiol Virol, F-75010 Paris, France.
   Hop St Louis, Serv Immunohematol, F-75010 Paris, France.
C3 Universite Paris Cite; Assistance Publique Hopitaux Paris (APHP);
   Hopital Universitaire Saint-Louis - APHP; Universite Paris Cite;
   Assistance Publique Hopitaux Paris (APHP); Hopital Universitaire
   Saint-Louis - APHP; Universite Paris Cite; Assistance Publique Hopitaux
   Paris (APHP); Hopital Universitaire Saint-Louis - APHP
RP Park, S (corresponding author), Hop St Louis, Hematol Lab, 1 Av Claude Vellefaux, F-75010 Paris, France.
RI PARK, Sophie/M-6597-2014
OI PARK, Sophie/0000-0003-0514-5789
CR BERGSTROM T, 1996, SCAND J INFECT DIS S, V100, P41
   Burke DG, 1997, J INFECT DIS, V176, P1080, DOI 10.1086/516516
   Chretien F., 1997, Archives d'Anatomie et de Cytologie Pathologiques, V45, P142
   CINQUE P, 1993, LANCET, V342, P398, DOI 10.1016/0140-6736(93)92814-A
   Cinque P, 1996, AIDS, V10, P951, DOI 10.1097/00002030-199610090-00004
   DeMario MD, 1998, SEMIN ONCOL, V25, P492
   FORTSYTH PA, 1996, HEMAT ONCOL CLIN N A, V10, P1125
   GLESBY MJ, 1995, CLIN INFECT DIS, V21, P370, DOI 10.1093/clinids/21.2.370
   MACINTYRE EA, 1990, J CLIN INVEST, V86, P2125, DOI 10.1172/JCI114951
   Minjolle S, 1999, J CLIN MICROBIOL, V37, P950, DOI 10.1128/JCM.37.4.950-953.1999
   Nebuloni M, 1998, J NEUROVIROL, V4, P457, DOI 10.3109/13550289809114546
NR 11
TC 2
Z9 2
U1 0
U2 2
PU TAYLOR & FRANCIS LTD
PI ABINGDON
PA 4 PARK SQUARE, MILTON PARK, ABINGDON OX14 4RN, OXON, ENGLAND
SN 1042-8194
J9 LEUKEMIA LYMPHOMA
JI Leuk. Lymphoma
PD SEP
PY 2003
VL 44
IS 10
BP 1793
EP 1795
DI 10.1080/10428190310001606521
PG 3
WC Oncology; Hematology
WE Science Citation Index Expanded (SCI-EXPANDED)
SC Oncology; Hematology
GA 718GJ
UT WOS:000185136500022
PM 14692535
DA 2024-06-03
ER

PT J
AU Tewari, MK
   Singh, DP
   Pathak, A
   Khandelwal, N
   Radotra, BD
   Mathuriya, SN
   Gupta, SK
   Khosla, VK
   Sharma, BS
   Kak, VK
AF Tewari, MK
   Singh, DP
   Pathak, A
   Khandelwal, N
   Radotra, BD
   Mathuriya, SN
   Gupta, SK
   Khosla, VK
   Sharma, BS
   Kak, VK
TI Primary central nervous system lymphoma : Experience of 46 cases with
   review of literature
SO NEUROLOGY INDIA
LA English
DT Review
DE primary lymphoma; central nervous system; clinical spectrum; prognosis
ID PRIMARY CNS LYMPHOMA; NON-HODGKINS-LYMPHOMA; IMMUNODEFICIENCY-SYNDROME
   AIDS; PRIMARY MALIGNANT-LYMPHOMAS; COMBINED-MODALITY THERAPY;
   RETICULUM-CELL SARCOMA; CLINICAL PRESENTATION; CHEMOTHERAPY;
   RADIOTHERAPY; MANIFESTATIONS
AB Primary central nervous system lymphomas (PCNSL) constitutes only 1.0 to 1.5% of all brain tumors. Their incidence has gone up over tenfold in the last 25 years. Though, there has been an association of PCNSL with acquired immune deficiency syndrome (AIDS), yet the increased incidence of PCNSL appears to be real and unrelated to AIDS and organ transplantation. This increased incidence could be because of improvement in diagnostic technology and practice. The outcome remains gloomy despite surgical resection, radiotherapy and intensive adjuvant chemotherapy regimens, as majority of the patients succumb to the disease, with only 30-40% survival in patients under 70 years of age.
C1 Postgrad Inst Med Educ & Res, Dept Neurosurg, Chandigarh 160012, India.
   Postgrad Inst Med Educ & Res, Dept Radiodiagnosis & Imaging, Chandigarh 160012, India.
   Postgrad Inst Med Educ & Res, Dept Pathol, Chandigarh 160012, India.
   Postgrad Inst Med Educ & Res, Dept Radiodiag, Chandigarh 160012, India.
C3 Post Graduate Institute of Medical Education & Research (PGIMER),
   Chandigarh; Post Graduate Institute of Medical Education & Research
   (PGIMER), Chandigarh; Post Graduate Institute of Medical Education &
   Research (PGIMER), Chandigarh; Post Graduate Institute of Medical
   Education & Research (PGIMER), Chandigarh
EM
RI Gupta, Sunil Kumar/AAI-7143-2020
OI Gupta, Sunil Kumar/0000-0001-6810-6483
CR Abrey LE, 2000, J CLIN ONCOL, V18, P3144, DOI 10.1200/JCO.2000.18.17.3144
   ANDERS K, 1986, CLIN NEUROPATHOL, V5, P1
   Bataille B, 2000, J NEUROSURG, V92, P261, DOI 10.3171/jns.2000.92.2.0261
   Bessell EM, 1996, J CLIN ONCOL, V14, P945, DOI 10.1200/JCO.1996.14.3.945
   Boiardi A, 1999, J NEUROL, V246, P31, DOI 10.1007/s004150050302
   BRADA M, 1995, INT J RADIAT ONCOL, V33, P769, DOI 10.1016/0360-3016(95)02123-S
   Cartmill M, 2000, BRIT J NEUROSURG, V14, P313
   DEANGELIS LM, 1990, NEUROLOGY, V40, P80, DOI 10.1212/WNL.40.1.80
   DEANGELIS LM, 1992, J CLIN ONCOL, V10, P635, DOI 10.1200/JCO.1992.10.4.635
   DEANGELIS LM, 1991, NEUROLOGY, V41, P619, DOI 10.1212/WNL.41.5.619
   FINE HA, 1993, ANN INTERN MED, V119, P1093, DOI 10.7326/0003-4819-119-11-199312010-00007
   FORSYTH PA, 1994, NEUROLOGY, V44, P1473, DOI 10.1212/WNL.44.8.1473
   GABBAI AA, 1989, J NEUROSURG, V70, P190, DOI 10.3171/jns.1989.70.2.0190
   GILL PS, 1985, AM J MED, V78, P742, DOI 10.1016/0002-9343(85)90277-3
   GONZALEZ DG, 1983, CANCER-AM CANCER SOC, V51, P2048, DOI 10.1002/1097-0142(19830601)51:11<2048::AID-CNCR2820511115>3.0.CO;2-T
   HELLE TL, 1984, J NEUROSURG, V60, P94, DOI 10.3171/jns.1984.60.1.0094
   HENRY JM, 1974, CANCER, V34, P1293, DOI 10.1002/1097-0142(197410)34:4<1293::AID-CNCR2820340441>3.0.CO;2-P
   Herrlinger U, 1998, ACTA NEUROL SCAND, V97, P257
   HOCHBERG FH, 1988, J NEUROSURG, V68, P835, DOI 10.3171/jns.1988.68.6.0835
   JAFFE ES, 1974, NEW ENGL J MED, V290, P813, DOI 10.1056/NEJM197404112901501
   Jellinger K, 1975, Acta Neuropathol Suppl, VSuppl 6, P95
   JIDDANE M, 1986, J NEUROSURG, V65, P592, DOI 10.3171/jns.1986.65.5.0592
   KAWAKAMI Y, 1985, J NEUROSURG, V62, P522, DOI 10.3171/jns.1985.62.4.0522
   Kim DG, 1996, ACTA NEUROCHIR, V138, P280, DOI 10.1007/BF01411738
   Lal P, 2000, Indian J Cancer, V37, P32
   LEMANN W, 1985, J NEUROPATH EXP NEUR, V44, P349, DOI 10.1097/00005072-198505000-00135
   LEVY RM, 1985, J NEUROSURG, V62, P475, DOI 10.3171/jns.1985.62.4.0475
   LITTMAN P, 1975, CANCER, V35, P1412, DOI 10.1002/1097-0142(197505)35:5<1412::AID-CNCR2820350526>3.0.CO;2-T
   LUTZ JM, 1994, BRIT J CANCER, V70, P716, DOI 10.1038/bjc.1994.381
   Maher EA, 1999, SEMIN ONCOL, V26, P346
   MARSH WL, 1983, CANCER-AM CANCER SOC, V51, P1125, DOI 10.1002/1097-0142(19830315)51:6<1125::AID-CNCR2820510625>3.0.CO;2-U
   NEUWELT EA, 1986, CANCER-AM CANCER SOC, V58, P1609, DOI 10.1002/1097-0142(19861015)58:8<1609::AID-CNCR2820580805>3.0.CO;2-7
   OBrien PC, 1996, MED J AUSTRALIA, V165, P424
   ONEILL BP, 1995, INT J RADIAT ONCOL, V33, P663, DOI 10.1016/0360-3016(95)00207-F
   PERRY GS, 1980, J PEDIATR-US, V97, P72, DOI 10.1016/S0022-3476(80)80133-8
   ROCKWOOD EJ, 1984, J NEUROSURG, V61, P369, DOI 10.3171/jns.1984.61.2.0369
   Schlegel U, 2000, J NEUROL SCI, V181, P1, DOI 10.1016/S0022-510X(00)00385-3
   SCHLOM J, 1986, CANCER RES, V46, P3225
   SCHMITTGRAFF A, 1983, ACTA CYTOL, V27, P267
   TAYLOR CR, 1978, CANCER-AM CANCER SOC, V41, P2197, DOI 10.1002/1097-0142(197806)41:6<2197::AID-CNCR2820410619>3.0.CO;2-2
   WOODMAN R, 1985, MEDICINE, V64, P425, DOI 10.1097/00005792-198511000-00006
NR 41
TC 2
Z9 3
U1 0
U2 0
PU MEDKNOW PUBLICATIONS & MEDIA PVT LTD
PI MUMBAI
PA B-9, KANARA BUSINESS CENTRE, OFF LINK RD, GHAKTOPAR-E, MUMBAI, 400075,
   INDIA
SN 0028-3886
EI 1998-4022
J9 NEUROL INDIA
JI Neurol. India
PD DEC
PY 2002
VL 50
IS 4
BP 424
EP 429
PG 6
WC Neurosciences
WE Science Citation Index Expanded (SCI-EXPANDED)
SC Neurosciences & Neurology
GA 649KG
UT WOS:000181206900006
PM 12577089
DA 2024-06-03
ER

PT J
AU Hochberg, FH
   Tabatabai, G
AF Hochberg, FH
   Tabatabai, G
TI Therapy of PCNSL at the Massachusetts General Hospital with high dose
   methotrexate and deferred radiotherapy
SO ANNALS OF HEMATOLOGY
LA English
DT Article; Proceedings Paper
CT 1st International Symposium on Biology and Treatment of Aggressive
   Lymphomas (Lymphoma 2000)
CY SEP 23-27, 2000
CL SAARBRUCKEN, GERMANY
ID NERVOUS-SYSTEM LYMPHOMA; CHEMOTHERAPY
C1 Massachusetts Gen Hosp, Boston, MA 02114 USA.
C3 Harvard University; Massachusetts General Hospital
RP Hochberg, FH (corresponding author), Massachusetts Gen Hosp, Boston, MA 02114 USA.
RI ; Tabatabai, Ghazaleh/A-6365-2015
OI Tabatabai, Ghazaleh/0000-0002-3542-8782; Tabatabai,
   Ghazaleh/0000-0003-3702-923X
CR BATCHELOR TT, 2000, NABTT, P96
   Glass J, 1996, J NEURO-ONCOL, V30, P257
   GLASS J, 1994, J NEUROSURG, V81, P188, DOI 10.3171/jns.1994.81.2.0188
   Guha-Thakurta N, 1999, J NEURO-ONCOL, V43, P259, DOI 10.1023/A:1006210703827
   Soussain C, 1999, LEUKEMIA LYMPHOMA, V33, P543, DOI 10.3109/10428199909058458
NR 5
TC 2
Z9 2
U1 0
U2 1
PU SPRINGER-VERLAG
PI NEW YORK
PA 175 FIFTH AVE, NEW YORK, NY 10010 USA
SN 0939-5555
J9 ANN HEMATOL
JI Ann. Hematol.
PD NOV
PY 2001
VL 80
SU 3
BP B111
EP B112
PG 2
WC Hematology
WE Conference Proceedings Citation Index - Science (CPCI-S); Science Citation Index Expanded (SCI-EXPANDED)
SC Hematology
GA 500GR
UT WOS:000172618700027
PM 11757690
DA 2024-06-03
ER

PT J
AU García-Rayo, S
   Martínez-Monge, R
   Brugarolas, A
AF García-Rayo, S
   Martínez-Monge, R
   Brugarolas, A
TI Response to chemotherapy in cerebral primary lymphoma
SO REVISTA DE NEUROLOGIA
LA Spanish
DT Article
DE immunocompetent; intensive chemotherapy; multicentric; primary central
   nervous system lymphoma; radiosurgery; radiotherapy
ID CENTRAL NERVOUS-SYSTEM; PRIMARY MALIGNANT-LYMPHOMA; RADIATION; SURVIVAL;
   THERAPY; BRAIN
AB Introduction. Primary central nervous system lymphomas (PCNSL) are infrequent, rapidly growing and generally limited to the central nervous system (CNS). In recent years there has been a threefold increase in cases occurring in immunocompetent individuals. Conventional treatment with corticosteroids and radiotherapy is effective, but over 80% relapse in less than a year Chemotherapy has significantly improved the results of treatment. Clinical case. We present a case of PCNSL in an immunocompetent patient which was resistant to radiotherapy (RT), There was progression of the lymphoma three months after irradiation, and later multicentric progression one month after radiosurgery. The patient had complete radiological remission after systemic chemotherapy. The clinical improvement was obvious, since before chemotherapy was started the patient was in coma and had episodes of apnea. The therapeutic response obtained was consolidated by intensive chemotherapy and hemopoietic support. The condition is still in remission three years after completion of this treatment. Conclusions. There are no publications about the results of combined radiotherapy and chemotherapy in series of patients with PCNSL, since there are relatively few patients and many methods of treatment are tried [REV NEUROL 1998; 27: 992-4].
C1 Univ Navarra, Fac Med, Clin Univ, Dept Oncol, E-31080 Pamplona, Spain.
   Univ Navarra, Fac Med, Clin Univ, Dept Radioterapia, E-31080 Pamplona, Spain.
C3 University of Navarra; University of Navarra
RI Perez-Cornago, Aurora/C-1097-2016
OI Perez-Cornago, Aurora/0000-0002-5652-356X
CR BERRY MP, 1981, INT J RADIAT ONCOL, V7, P55, DOI 10.1016/0360-3016(81)90060-2
   DEANGELIS LM, 1992, J CLIN ONCOL, V10, P635, DOI 10.1200/JCO.1992.10.4.635
   DEANGELIS LM, 1997, CANC PRINCIPLES PRAC, P2236
   EBY NL, 1988, CANCER-AM CANCER SOC, V62, P2461, DOI 10.1002/1097-0142(19881201)62:11<2461::AID-CNCR2820621135>3.0.CO;2-M
   GONZALEZ DG, 1983, CANCER-AM CANCER SOC, V51, P2048, DOI 10.1002/1097-0142(19830601)51:11<2048::AID-CNCR2820511115>3.0.CO;2-T
   HENRY JM, 1974, CANCER, V34, P1293, DOI 10.1002/1097-0142(197410)34:4<1293::AID-CNCR2820340441>3.0.CO;2-P
   HOCHBERG FH, 1988, J NEUROSURG, V68, P835, DOI 10.3171/jns.1988.68.6.0835
   LOEFFLER JS, 1985, J CLIN ONCOL, V3, P490, DOI 10.1200/JCO.1985.3.4.490
   MURRAY K, 1986, J NEUROSURG, V65, P600, DOI 10.3171/jns.1986.65.5.0600
   ROSENBLUM ML, 1988, ANN NEUROL S, V23, P13
   SHIBAMOTO Y, 1990, CANCER-AM CANCER SOC, V65, P1907, DOI 10.1002/1097-0142(19900501)65:9<1907::AID-CNCR2820650906>3.0.CO;2-L
NR 11
TC 2
Z9 2
U1 0
U2 0
PU REVISTA DE NEUROLOGIA
PI BARCELONA
PA C/O CESAR VIGUERA, EDITOR, APDO 94121, 08080 BARCELONA, SPAIN
SN 0210-0010
J9 REV NEUROLOGIA
JI Rev. Neurologia
PD DEC
PY 1998
VL 27
IS 160
BP 992
EP 994
DI 10.33588/rn.27160.98199
PG 3
WC Clinical Neurology
WE Science Citation Index Expanded (SCI-EXPANDED)
SC Neurosciences & Neurology
GA 161FG
UT WOS:000078278100018
PM 9951022
DA 2024-06-03
ER

PT J
AU Schaller, C
   Kelly, PJ
AF Schaller, C
   Kelly, PJ
TI Primary central nervous system non-Hodgkin's lymphoma (PCNSL): Does age
   and histology at presentation affect outcome?
SO ZENTRALBLATT FUR NEUROCHIRURGIE
LA English
DT Article
DE primary central nervous system lymphoma; brain tumor; stereotactic
   biopsy; radiotherapy
ID IMMUNE-DEFICIENCY-SYNDROME; PRIMARY CNS LYMPHOMA; PRIMARY
   MALIGNANT-LYMPHOMA; T-CELL LYMPHOMA; INTRACRANIAL LYMPHOMA; SYNDROME
   AIDS; CHEMOTHERAPY; EXPERIENCE; DIAGNOSIS; RADIATION
AB Primary Non-Hodgkin's lymphoma of the central nervous system (PCNSL) was diagnosed by computer-assisted stereotactic biopsy in 27 non-AIDS patients (22 men, 5 women; mean age, 50.6 years, median age 53 years). Among the various histologies, fourteen patients had diffuse large cell lymphoma (DLC), five had diffuse, small, non-cleaved lymphoma (DSNC) and five had diffuse mixed cell lymphoma (DMC). Immunohistochemical analysis revealed 6 B-cell and 3 T-cell types. Multiple lesions occured in 11 patients. Most patients received radiation therapy and some received adjunctive and/or salvage chemotherapy. Several factors correlated with survival. median survival for the entire group was 356 days (0.975 years). However, Median survival was 1982 days (5.43 years) for the 8 patients who were less than 45 years old at diagnosis versus 283 days (0.77 years) for the 18 patients who were older (p = 0.03). One patient was lost for follow up. Four of the 5 patients who have survived for more than four years were less than 45 years of age; of these 2 had T-cell lymphoma. Only 3 of the 11 patients with multiple lesions died within the first year. A proportional hazards model was derived for the group and suggested that older age and uncleaved histology are associated with shorter survival. Treatment protocols should include stratification factors to allow comparison between groups with different characteristics and to account for age and/or histology dependent on differences in response.
C1 UNIV BONN,NEUROCHIRURG KLIN,D-53127 BONN,GERMANY.
   NYU,MED CTR,NEW YORK,NY 10016.
C3 University of Bonn; New York University
CR ARMITAGE JO, 1991, ANN ONCOL, P37
   BERRY MP, 1981, INT J RADIAT ONCOL, V7, P55, DOI 10.1016/0360-3016(81)90060-2
   CHAMBERLAIN MC, 1990, ARCH NEUROL-CHICAGO, V47, P1113, DOI 10.1001/archneur.1990.00530100081017
   DEANGELIS LM, 1990, NEUROLOGY, V40, P80, DOI 10.1212/WNL.40.1.80
   DESMISTRO A, 1990, AM J CLIN PATHOL, V94, P722
   EBY NL, 1988, CANCER-AM CANCER SOC, V62, P2461, DOI 10.1002/1097-0142(19881201)62:11<2461::AID-CNCR2820621135>3.0.CO;2-M
   EPSTEIN LG, 1988, PEDIATRICS, V82, P255
   GABBAI AA, 1989, J NEUROSURG, V70, P190, DOI 10.3171/jns.1989.70.2.0190
   GRANT JW, 1989, NEUROLOGY, V39, P1190
   HAYAKAWA T, 1994, J NEURO-ONCOL, V19, P197, DOI 10.1007/BF01053274
   HELLE TL, 1984, J NEUROSURG, V60, P94, DOI 10.3171/jns.1984.60.1.0094
   HOBSON DE, 1986, CAN J NEUROL SCI, V13, P55, DOI 10.1017/S0317167100035800
   HOCHBERG FH, 1988, J NEUROSURG, V68, P835, DOI 10.3171/jns.1988.68.6.0835
   INOUE M, 1990, NEUROSURGERY, V27, P148, DOI 10.1227/00006123-199007000-00025
   LACHANCE DH, 1991, NEUROLOGY, V41, P95, DOI 10.1212/WNL.41.1.95
   LANG W, 1989, ACTA NEUROPATHOL, V77, P379, DOI 10.1007/BF00687372
   LEVY RM, 1985, J NEUROSURG, V62, P475, DOI 10.3171/jns.1985.62.4.0475
   MILLER DC, 1994, CANCER-AM CANCER SOC, V74, P1383, DOI 10.1002/1097-0142(19940815)74:4<1383::AID-CNCR2820740432>3.0.CO;2-1
   MORGELLO S, 1989, NEUROLOGY, V39, P1190, DOI 10.1212/WNL.39.9.1190
   MURRAY K, 1986, J NEUROSURG, V65, P600, DOI 10.3171/jns.1986.65.5.0600
   NEUWELT EA, 1986, CANCER-AM CANCER SOC, V58, P1609, DOI 10.1002/1097-0142(19861015)58:8<1609::AID-CNCR2820580805>3.0.CO;2-7
   ONEILL BP, 1987, NEUROLOGY, V37, P1160, DOI 10.1212/WNL.37.7.1160
   Party MRCBTW, 1990, J NEUROONCOL, V9, P47
   POLLACK IF, 1990, AM J CLIN PATHOL, V94, P722
   REMICK SC, 1990, MEDICINE, V69, P345, DOI 10.1097/00005792-199011000-00003
   RUBIN M, 1987, CAN J NEUROL SCI, V14, P175
   SHIBAMOTO Y, 1990, CANCER-AM CANCER SOC, V65, P1907, DOI 10.1002/1097-0142(19900501)65:9<1907::AID-CNCR2820650906>3.0.CO;2-L
   SO YT, 1986, ANN NEUROL, V20, P566, DOI 10.1002/ana.410200503
   STRAUS DJ, 1983, CANCER, V51, P101, DOI 10.1002/1097-0142(19830101)51:1<101::AID-CNCR2820510121>3.0.CO;2-W
   SUGITA Y, 1988, SURG NEUROL, V30, P148, DOI 10.1016/0090-3019(88)90102-4
   TOMLINSON FH, 1995, J NEUROSURG, V82, P558, DOI 10.3171/jns.1995.82.4.0558
   VOSE JM, 1988, J CLIN ONCOL, V6, P1838, DOI 10.1200/JCO.1988.6.12.1838
NR 32
TC 2
Z9 2
U1 0
U2 0
PU JOHANN AMBROSIUS BARTH VERLAG
PI HEIDELBERG
PA IM WEIHER 10, D-69121 HEIDELBERG, GERMANY
SN 0044-4251
J9 ZBL NEUROCHIR
JI Zent.bl. Neurochir.
PY 1996
VL 57
IS 3
BP 156
EP 162
PG 7
WC Neurosciences; Surgery
WE Science Citation Index Expanded (SCI-EXPANDED)
SC Neurosciences & Neurology; Surgery
GA VD173
UT WOS:A1996VD17300006
PM 8794548
DA 2024-06-03
ER

PT J
AU FICKER, DM
   HAMMACK, JE
AF FICKER, DM
   HAMMACK, JE
TI PRIMARY CENTRAL-NERVOUS-SYSTEM LYMPHOMA PRESENTING AS AUTONOMIC
   DYSFUNCTION
SO JOURNAL OF NEURO-ONCOLOGY
LA English
DT Article
DE CENTRAL NERVOUS SYSTEM LYMPHOMA; DYSAUTONOMIA; BRAIN-STEM TUMOR
AB We describe a patient with primary central nervous system lymphoma (PCNSL) who presented with symptoms of subacute onset of dysautonomia. Autonomic testing indicated a peripheral autonomic neuropathy while magnetic resonance imaging revealed brainstem involvement. We propose that this patient's autonomic dysfunction could be the result of a paraneoplastic syndrome and PCNSL should be considered in the differential diagnosis of dysautonomia.
C1 MAYO CLIN,DEPT NEUROL,ROCHESTER,MN 55905.
C3 Mayo Clinic
CR HOCHBERG FH, 1988, J NEUROSURG, V68, P835, DOI 10.3171/jns.1988.68.6.0835
   HSU CY, 1984, NEUROLOGY, V34, P1137, DOI 10.1212/WNL.34.9.1137
   KHURANA RK, 1993, CLIN AUTONOMIC DISOR, P505
   LOW PA, 1993, J CLIN NEUROPHYSIOL, V10, P14, DOI 10.1097/00004691-199301000-00003
   VANLIESHOUT JJ, 1986, J NEUROL NEUROSUR PS, V48, P830
NR 5
TC 2
Z9 2
U1 0
U2 0
PU KLUWER ACADEMIC PUBL
PI DORDRECHT
PA SPUIBOULEVARD 50, PO BOX 17, 3300 AA DORDRECHT, NETHERLANDS
SN 0167-594X
J9 J NEURO-ONCOL
JI J. Neuro-Oncol.
PY 1995
VL 23
IS 3
BP 245
EP 248
DI 10.1007/BF01059956
PG 4
WC Oncology; Clinical Neurology
WE Science Citation Index Expanded (SCI-EXPANDED)
SC Oncology; Neurosciences & Neurology
GA RC909
UT WOS:A1995RC90900009
PM 7673987
DA 2024-06-03
ER

PT J
AU BISWAS, PK
   BALAGOPAL, N
   PILLAI, CKS
AF BISWAS, PK
   BALAGOPAL, N
   PILLAI, CKS
TI PHOSPHORYLATED CNSL PREPOLYMER AS A FOUNDRY-SAND CORE BINDER
SO INDIAN JOURNAL OF ENGINEERING AND MATERIALS SCIENCES
LA English
DT Article
AB The performance of phosphorylated cashew-nut shell liquid prepolymer (PCNSL) as a core binder in foundry applications is studied. Moulding properties such as strength and permeability and casting characteristics such as surface quality of castings, core-metal reactions, core collapsibility and anti-damping behaviour are analysed. PCNSL gave superior performance in all these factors when compared to other system of binders such as sodium silicate. Studies based on LR spectra and SEM on the nature of bonding between sand and PCNSL indicate involvement mostly of physical and mechanical interactions rather than chemical bonding. The formation (during casting) of higher content of char (due to the presence of phosphate group in the resin) with open porous structure as revealed by SEM is thought to give better collapsibility to the system.
C1 REG RES LAB, THIRUVANANTHAPURAM 695019, INDIA.
C3 Council of Scientific & Industrial Research (CSIR) - India; CSIR -
   National Institute Interdisciplinary Science & Technology (NIIST)
RI Nair, Balagopal N/A-7255-2008
OI Nair, Balagopal N/0000-0002-5128-3050
NR 0
TC 2
Z9 2
U1 0
U2 4
PU NATL INST SCIENCE COMMUNICATION-NISCAIR
PI NEW DELHI
PA DR K S KRISHNAN MARG, PUSA CAMPUS, NEW DELHI 110 012, INDIA
SN 0971-4588
EI 0975-1017
J9 INDIAN J ENG MATER S
JI Indian J. Eng. Mat. Sci.
PD APR
PY 1994
VL 1
IS 2
BP 99
EP 106
PG 8
WC Engineering, Multidisciplinary; Materials Science, Multidisciplinary
WE Science Citation Index Expanded (SCI-EXPANDED)
SC Engineering; Materials Science
GA PE726
UT WOS:A1994PE72600006
DA 2024-06-03
ER

PT J
AU Bathla, G
   Dhruba, DD
   Soni, N
   Liu, YA
   Larson, NB
   Kassmeyer, BA
   Mohan, S
   Roberts-Wolfe, D
   Rathore, S
   Le, NH
   Zhang, HH
   Sonka, M
   Priya, S
AF Bathla, Girish
   Dhruba, Durjoy Deb
   Soni, Neetu
   Liu, Yanan
   Larson, Nicholas B.
   Kassmeyer, Blake A.
   Mohan, Suyash
   Roberts-Wolfe, Douglas
   Rathore, Saima
   Le, Nam H.
   Zhang, Honghai
   Sonka, Milan
   Priya, Sarv
TI AI-based classification of three common malignant tumors in
   neuro-oncology: A multi-institutional comparison of machine learning and
   deep learning methods
SO JOURNAL OF NEURORADIOLOGY
LA English
DT Article
DE Arti ficial intelligence; Radiomics; Glioblastoma; Metastasis; CNS
   lymphoma
ID BRAIN; RADIOMICS; SYSTEM; CNN
AB Purpose: To determine if machine learning (ML) or deep learning (DL) pipelines perform better in AI -based three -class classi fication of glioblastoma (GBM), intracranial metastatic disease (IMD) and primary CNS lymphoma (PCNSL). Methodology: Retrospective analysis included 502 cases for training (208 GBM, 67 PCNSL and 227 IMD), with external validation on 86 cases (27:27:32). Multiparametric MRI images (T1W, T2W, FLAIR, DWI and T1 -CE) were co -registered, resampled, denoised and intensity normalized, followed by semiautomatic 3D segmentation of the enhancing tumor (ET) and peritumoral region (PTR). Model performance was assessed using several ML pipelines and 3D -convolutional neural networks (3D -CNN) using sequence speci fic masks, as well as combination of masks. All pipelines were trained and evaluated with 5 -fold nested cross -validation on internal data followed by external validation using multi -class AUC. Results: Two ML models achieved similar performance on test set, one using T2 -ET and T2-PTR masks (AUC: 0.885, 95% CI: [0.816, 0.935] and another using T1 -CE -ET and FLAIR-PTR mask (AUC: 0.878, CI: [0.804, 0.930]). The best performing DL models achieved an AUC of 0.854, (CI [0.774, 0.914]) on external data using T1 -CE -ET and T2-PTR masks, followed by model derived from T1 -CE -ET, ADC -ET and FLAIR-PTR masks (AUC: 0.851, CI [0.772, 0.909]). Conclusion: Both ML and DL derived pipelines achieved similar performance. T1 -CE mask was used in three of the top four overall models. Additionally, all four models had some mask derived from PTR, either T2WI or FLAIR. (c) 2023 Elsevier Masson SAS. All rights reserved.
C1 [Bathla, Girish; Soni, Neetu; Priya, Sarv] Univ Iowa Hosp & Clin, Dept Radiol, 200 Hawkins Dr, Iowa, IA 52242 USA.
   [Bathla, Girish] Mayo Clin, Dept Radiol, 200 1st St SW, Rochester, MN 55902 USA.
   [Dhruba, Durjoy Deb; Le, Nam H.; Zhang, Honghai; Sonka, Milan] Univ Iowa, Seamans Ctr Engn Arts & Sci 4016, Elect & Comp Engn, Iowa, IA 52242 USA.
   [Soni, Neetu] Univ Rochester, Med Ctr, Dept Imaging Sci, 601 Elmwood Ave,Box 648, Rochester, NY 14642 USA.
   [Liu, Yanan] Univ Iowa, Adv Pulm Physi Imaging Lab APPIL, 200 Hawkins Dr, Iowa, IA 52242 USA.
   [Larson, Nicholas B.; Kassmeyer, Blake A.] Mayo Clin, Dept Quantitat Hlth Sci, Div Clin Trials & Biostat, 200 1st St SW, Rochester, MN 55902 USA.
   [Mohan, Suyash; Roberts-Wolfe, Douglas] Univ Penn, Perelman Sch Med, Dept Radiol, 3400 Spruce St, Philadelphia, PA 19104 USA.
   [Rathore, Saima] Avid Radiopharmaceut, 3711 Market St, Philadelphia, PA 19104 USA.
C3 University of Iowa; Mayo Clinic; University of Iowa; University of
   Rochester; University of Iowa; Mayo Clinic; University of Pennsylvania;
   Avid Radiopharmaceuticals
RP Bathla, G (corresponding author), Mayo Clin, Dept Radiol, 200 1st St SW, Rochester, MN 55902 USA.
EM
RI Priya, Sarv/U-8494-2019; Dhruba, Durjoy Deb/JEO-9464-2023; Le, Nam
   Hoang/AFX-7331-2022
OI Priya, Sarv/0000-0003-2442-1902; Dhruba, Durjoy Deb/0000-0003-0813-8269;
   Le, Nam Hoang/0000-0003-2556-7612; Mohan, Suyash/0000-0002-4025-115X;
   BATHLA, GIRISH/0000-0003-3170-242X; Larson,
   Nicholas/0000-0002-3468-4215; Roberts-Wolfe, Doug/0000-0001-5456-490X;
   SONI, NEETU/0000-0003-2082-1634
FU ACS-Holden Cancer Center grant; NIH-NIBIB [R01-EB004640]
FX This work was supported, in part, ACS-Holden Cancer Center grant and by
   NIH-NIBIB grant R01-EB004640.
CR Afshar P, 2019 2019 IEEE INT C
   Bae S, 2020, SCI REP-UK, V10, DOI 10.1038/s41598-020-68980-6
   Bander ED, 2019, J NEURO-ONCOL, V141, P121, DOI 10.1007/s11060-018-03014-w
   Baro V, 2022, CURR ONCOL, V29, P3472, DOI 10.3390/curroncol29050280
   Bathla G, 2016, CLIN RADIOL, V71, P602, DOI 10.1016/j.crad.2016.02.006
   Bathla G, 2021, EUR RADIOL, V31, P8703, DOI 10.1007/s00330-021-07845-6
   Berghoff AS, 2016, ESMO OPEN, V1, DOI 10.1136/esmoopen-2015-000024
   Callovini GM, 2018, CLIN NEUROL NEUROSUR, V174, P101, DOI 10.1016/j.clineuro.2018.09.020
   Chakrabarty S, 2021, RADIOL-ARTIF INTELL, V3, DOI 10.1148/ryai.2021200301
   Chawla S, 2010, J COMPUT ASSIST TOMO, V34, P836, DOI 10.1097/RCT.0b013e3181ec554e
   Cheng J, 2015, PLOS ONE, V10, DOI 10.1371/journal.pone.0140381
   Fink Kathleen R, 2013, Surg Neurol Int, V4, pS209, DOI 10.4103/2152-7806.111298
   Fortin J, 2021, Harmonization of multi-site imaging data with ComBat, R Package Version 1.0.9
   Gao PY, 2022, JAMA NETW OPEN, V5, DOI 10.1001/jamanetworkopen.2022.25608
   Hand DJ, 2001, MACH LEARN, V45, P171, DOI 10.1023/A:1010920819831
   Joo B, 2022, J Neuroradiol., DOI [10.1016/j.neurad.2022.11.00110.1016/j.neurad.2022.11.001, DOI 10.1016/J.NEURAD.2022.11.00110.1016/J.NEURAD.2022.11.001]
   Lamba N, 2021, NEURO-ONCOLOGY, V23, P1447, DOI 10.1093/neuonc/noab101
   Lambin P, 2017, NAT REV CLIN ONCOL, V14, P749, DOI 10.1038/nrclinonc.2017.141
   Li YZ, 2022, EUR RADIOL, V32, P8039, DOI 10.1007/s00330-022-08828-x
   Ma JH, 2010, AM J NEURORADIOL, V31, P1699, DOI 10.3174/ajnr.A2161
   Neska-Matuszewska M, 2018, PLOS ONE, V13, DOI 10.1371/journal.pone.0191341
   Ostrom QT, 2019, NEURO-ONCOLOGY, V21, pV1, DOI 10.1093/neuonc/noz150
   Park JE, 2020, BMC CANCER, V20, DOI 10.1186/s12885-019-6504-5
   Priya S, 2021, SCI REP-UK, V11, DOI 10.1038/s41598-021-92155-6
   Priya S, 2021, CANCERS, V13, DOI 10.3390/cancers13112568
   Priya S, 2021, SCI REP-UK, V11, DOI 10.1038/s41598-021-90032-w
   Reza AW, 2023, APPL SCI-BASEL, V13, DOI 10.3390/app13010312
   Rudie JD, 2019, RADIOLOGY, V290, P607, DOI 10.1148/radiol.2018181928
   Sajjad M, 2019, J COMPUT SCI-NETH, V30, P174, DOI 10.1016/j.jocs.2018.12.003
   Shaver MM, 2019, CANCERS, V11, DOI 10.3390/cancers11060829
   Soni N, 2019, AM J NEURORADIOL, V40, P928, DOI 10.3174/ajnr.A6075
   Stadlbauer A, 2022, CANCERS, V14, DOI 10.3390/cancers14102363
   Steffan-Dewenter I., 2002, Ecology, V83, P1421, DOI 10.1890/0012-9658(2002)083[1421:SDEOLC]2.0.CO;2
   STONE M, 1974, J R STAT SOC B, V36, P111, DOI 10.1111/j.2517-6161.1974.tb00994.x
   Swinburne NC, 2019, ANN TRANSL MED, V7, DOI 10.21037/atm.2018.08.05
   Talo M, 2019, COMPUT MED IMAG GRAP, V78, DOI 10.1016/j.compmedimag.2019.101673
   Thomas RP, 2013, J NEURO-ONCOL, V112, P91, DOI 10.1007/s11060-012-1030-1
   van Griethuysen JJM, 2017, CANCER RES, V77, pE104, DOI 10.1158/0008-5472.CAN-17-0339
   Yin Y, 2010, IEEE T MED IMAGING, V29, P2023, DOI 10.1109/TMI.2010.2058861
   Zhang L, 2023, EUR J RADIOL, V158, DOI 10.1016/j.ejrad.2022.110639
NR 40
TC 1
Z9 1
U1 0
U2 2
PU MASSON EDITEUR
PI MOULINEAUX CEDEX 9
PA 21 STREET CAMILLE DESMOULINS, ISSY, 92789 MOULINEAUX CEDEX 9, FRANCE
SN 0150-9861
EI 1773-0406
J9 J NEURORADIOLOGY
JI J. Neuroradiol.
PD MAY
PY 2024
VL 51
IS 3
BP 258
EP 264
DI 10.1016/j.neurad.2023.08.007
PG 7
WC Clinical Neurology; Neuroimaging; Radiology, Nuclear Medicine & Medical
   Imaging
WE Science Citation Index Expanded (SCI-EXPANDED)
SC Neurosciences & Neurology; Radiology, Nuclear Medicine & Medical Imaging
GA QL0J3
UT WOS:001220908300002
PM 37652263
DA 2024-06-03
ER

PT J
AU Haran, A
   Even-Zohar, NG
   Haran, M
   Lebel, E
   Aumann, S
   Shaulov, A
   Gatt, M
   Nachmias, B
AF Haran, Arnon
   Even-Zohar, Noa Gross
   Haran, Michal
   Lebel, Eyal
   Aumann, Shlomzion
   Shaulov, Adir
   Gatt, Moshe
   Nachmias, Boaz
TI Impact of Folinic Acid Dosing on Efficacy and Toxicity of High-Dose
   Methotrexate in Central Nervous System Lymphoma
SO CLINICAL LYMPHOMA MYELOMA & LEUKEMIA
LA English
DT Article
DE CNS lymphoma; Progression-free survival; Retrospective
ID PRIMARY CNS LYMPHOMA; UNDER-THE-CURVE; INTERNATIONAL EXTRANODAL
   LYMPHOMA; LEUKEMIA; RESCUE; CYTARABINE; RITUXIMAB; SURVIVAL; THERAPY;
   FOLATE
AB High -dose methotrexate with folinic acid (FA) rescue is the preferred treatment for primary central nervous system lymphoma. However, FA dosing regimens vary widely, and the effect of different doses on treatment outcomes remains unknown. In this study of 36 PCNSL patients, higher per -treatment cumulative FA doses were linked to worse progression -free survival. Introduction: High -dose methotrexate (HDMTX)-based regimens are the treatment of choice in primar y central ner vous system lymphoma (PCNSL). Folinic acid (FA) rescue is used to mitigate the toxic effects of MTX on normal cells. However, the optimal dosing of FA in PCNSL remains uncertain. Methods: We analyzed the relationship between FA dosing and treatment efficacy and toxicity in a cohort of 36 PCNSL patients treated at our institute between the years 2014 and 2022. A combination of univariate and multivariate analyses using known prognostic factors were used to determine the association between FA dosing and treatment outcomes. Results: We found that higher per -treatment cumulative FA doses were associated with inferior progression -free survival (PFS), with a hazard ratio (HR) of 2.2 for each 100 mg/m2 increase in FA dose. We identified a threshold of 350 mg/m2 /treatment, above which there was a significant reduction in PFS. Notably, lower FA doses did not result in increased toxicity. Conclusion: Our findings suggest that optimizing FA dosing to avoid very high rescue doses may improve treatment outcomes in PCNSL patients receiving HDMTX. Further prospective studies are warranted to validate these findings.
C1 [Haran, Arnon; Even-Zohar, Noa Gross; Lebel, Eyal; Aumann, Shlomzion; Shaulov, Adir; Gatt, Moshe; Nachmias, Boaz] Hebrew Univ Jerusalem, Hadassah Med Ctr, Dept Hematol, POB 12000, IL-91120 Jerusalem, Israel.
   [Haran, Arnon; Even-Zohar, Noa Gross; Lebel, Eyal; Aumann, Shlomzion; Shaulov, Adir; Gatt, Moshe; Nachmias, Boaz] Hebrew Univ Jerusalem, Fac Med, POB 12000, IL-91120 Jerusalem, Israel.
   [Haran, Michal] Kaplan Med Ctr, Dept Hematol, Rehovot, Israel.
C3 Hebrew University of Jerusalem; Hadassah University Medical Center;
   Hebrew University of Jerusalem; Hebrew University of Jerusalem; Kaplan
   Medical Center
RP Haran, A (corresponding author), Hebrew Univ Jerusalem, Hadassah Med Ctr, Dept Hematol, POB 12000, IL-91120 Jerusalem, Israel.; Haran, A (corresponding author), Hebrew Univ Jerusalem, Fac Med, POB 12000, IL-91120 Jerusalem, Israel.
EM
OI Gatt, Moshe/0000-0002-9426-4482; Haran, Arnon/0000-0003-2414-3746
CR [Anonymous], COMMON TERMINOLOGY C
   Ariyoshi Nobuhiro, 2018, BMJ Case Rep, V2018, DOI 10.1136/bcr-2018-225482
   Bairey O, 2021, ACTA HAEMATOL-BASEL, V144, P389, DOI 10.1159/000511208
   BORSI JD, 1991, EUR J CANCER, V27, P1006, DOI 10.1016/0277-5379(91)90269-J
   Calimeriy T, 2021, ESMO OPEN, V6, DOI 10.1016/j.esmoop.2021.100213
   Chen T, 2022, J HEMATOL ONCOL, V15, DOI 10.1186/s13045-022-01356-7
   Cohen IJ, 2013, MED HYPOTHESES, V81, P942, DOI 10.1016/j.mehy.2013.08.027
   Cohen IJ, 2004, J PEDIAT HEMATOL ONC, V26, P156, DOI 10.1097/00043426-200403000-00004
   Cohen IJ, 2003, PEDIATR HEMAT ONCOL, V20, P579, DOI 10.1080/08880010390242898
   Cole BF, 2007, JAMA-J AM MED ASSOC, V297, P2351, DOI 10.1001/jama.297.21.2351
   Ferreri AJM, 2003, J CLIN ONCOL, V21, P266, DOI 10.1200/JCO.2003.09.139
   Ferreri AJM, 2004, BRIT J CANCER, V90, P353, DOI 10.1038/sj.bjc.6601472
   Ferreri AJM, 2016, LANCET HAEMATOL, V3, pE217, DOI 10.1016/S2352-3026(16)00036-3
   Fox CP, 2019, BRIT J HAEMATOL, V184, P348, DOI 10.1111/bjh.15661
   Glass J, 2016, J CLIN ONCOL, V34, P1620, DOI 10.1200/JCO.2015.64.8634
   Hansen MF, 2017, BRIT J CANCER, V116, P752, DOI 10.1038/bjc.2017.11
   Hoang-Xuan K, 2023, NEURO-ONCOLOGY, V25, P37, DOI 10.1093/neuonc/noac196
   Izraeli S, 2022, HAEMATOLOGICA, V107, P782, DOI 10.3324/haematol.2022.280670
   Joerger M, 2010, BRIT J CANCER, V102, P673, DOI 10.1038/sj.bjc.6605559
   Kim YI, 2018, AM J CLIN NUTR, V107, P139, DOI 10.1093/ajcn/nqx076
   Lebel E, 2022, LEUKEMIA LYMPHOMA, V63, P2102, DOI 10.1080/10428194.2022.2064996
   Lopez-Lopez E, 2020, J CLIN INVEST, V130, P6600, DOI 10.1172/JCI140797
   Mappa S, 2013, HEMATOL ONCOL, V31, P143, DOI 10.1002/hon.2037
   May J, 2014, LEUKEMIA LYMPHOMA, V55, P1345, DOI 10.3109/10428194.2013.840780
   Möricke A, 2008, BLOOD, V111, P4477, DOI 10.1182/blood-2007-09-112920
   Morris PG, 2013, J CLIN ONCOL, V31, P3971, DOI 10.1200/JCO.2013.50.4910
   Morris PG, 2011, LEUKEMIA LYMPHOMA, V52, P1891, DOI 10.3109/10428194.2011.585527
   Olivier G, 2014, AM J HEMATOL, V89, P1024, DOI 10.1002/ajh.23812
   Pels H, 2003, J CLIN ONCOL, V21, P4489, DOI 10.1200/JCO.2003.04.056
   Rubenstein JL, 2013, BLOOD, V122, P2318, DOI 10.1182/blood-2013-06-453084
   SIROTNAK FM, 1978, CANCER RES, V38, P345
   Skärby TVC, 2006, LEUKEMIA, V20, P1955, DOI 10.1038/sj.leu.2404404
   Song J, 2000, CANCER RES, V60, P3191
   Van Guelpen B, 2006, GUT, V55, DOI 10.1136/gut.2005.085480
   Villano JL, 2011, BRIT J CANCER, V105, P1414, DOI 10.1038/bjc.2011.357
   Villanueva G, 2023, CANCERS, V15, DOI 10.3390/cancers15051459
NR 36
TC 1
Z9 1
U1 0
U2 0
PU CIG MEDIA GROUP, LP
PI DALLAS
PA 3500 MAPLE AVENUE, STE 750, DALLAS, TX 75219-3931 USA
SN 2152-2650
EI 2152-2669
J9 CL LYMPH MYELOM LEUK
JI Clin. Lymphoma Myeloma Leuk.
PD MAR
PY 2024
VL 24
IS 3
DI 10.1016/j.clml.2023.10.012
EA FEB 2024
PG 8
WC Oncology; Hematology
WE Science Citation Index Expanded (SCI-EXPANDED)
SC Oncology; Hematology
GA MM2F5
UT WOS:001193965700001
PM 38008594
DA 2024-06-03
ER

PT J
AU Schorb, E
   Isbell, LK
   Kerkhoff, A
   Mathas, S
   Braulke, F
   Egerer, G
   Roeth, A
   Schliffke, S
   Borchmann, P
   Brunnberg, U
   Kroschinsky, F
   Moehle, R
   Rank, A
   Wellnitz, D
   Kasenda, B
   Pospiech, L
   Wendler, J
   Scherer, F
   Deckert, M
   Henkes, E
   von Gottberg, P
   Gmehlin, D
   Backenstrass, M
   Jensch, A
   Burger-Martin, E
   Grishina, O
   Fricker, H
   Malenica, N
   Orban, A
   Duyster, J
   Ihorst, G
   Finke, J
   Illerhaus, G
AF Schorb, Elisabeth
   Isbell, Lisa Kristina
   Kerkhoff, Andrea
   Mathas, Stephan
   Braulke, Friederike
   Egerer, Gerlinde
   Roeth, Alexander
   Schliffke, Simon
   Borchmann, Peter
   Brunnberg, Uta
   Kroschinsky, Frank
   Moehle, Robert
   Rank, Andreas
   Wellnitz, Dominique
   Kasenda, Benjamin
   Pospiech, Lisa
   Wendler, Julia
   Scherer, Florian
   Deckert, Martina
   Henkes, Elina
   von Gottberg, Philipp
   Gmehlin, Dennis
   Backenstrass, Matthias
   Jensch, Antje
   Burger-Martin, Elvira
   Grishina, Olga
   Fricker, Heidi
   Malenica, Natalie
   Orban, Andras
   Duyster, Justus
   Ihorst, Gabriele
   Finke, Juergen
   Illerhaus, Gerald
TI High-dose chemotherapy and autologous haematopoietic stem-cell
   transplantation in older, fit patients with primary diffuse large B-cell
   CNS lymphoma (MARTA): a single-arm, phase 2 trial
SO LANCET HAEMATOLOGY
LA English
DT Article
ID NERVOUS-SYSTEM LYMPHOMA; QUALITY-OF-LIFE; ELDERLY-PATIENTS; 1ST-LINE
   TREATMENT; METHOTREXATE; SURVIVAL; EORTC; AGE
AB Background Available treatments for older patients with primary diffuse large B-cell CNS lymphoma (PCNSL) offer progression -free survival of up to 16 months. We aimed to investigate an intensified treatment of high -dose chemotherapy and autologous haematopoietic stem-cell transplantation (HSCT) in older patients with PCNSL. Methods MARTA was a prospective, single -arm, phase 2 study done at 15 research hospitals in Germany. Patients aged 65 years or older with newly diagnosed, untreated PCNSL were enrolled if they had an Eastern Cooperative Oncology Group performance status of 0-2 and were fit for high -dose chemotherapy and autologous HSCT. Induction treatment consisted of two 21-day cycles of high -dose intravenous methotrexate 3 center dot 5 g/m2 (day 1), intravenous cytarabine 2 g/m2 twice daily (days 2 and 3), and intravenous rituximab 375 mg/m2 (days 0 and 4) followed by highdose chemotherapy with intravenous rituximab 375 mg/m2 (day -8), intravenous busulfan 3 center dot 2 mg/kg (days -7 and -6), and intravenous thiotepa 5 mg/kg (days -5 and -4) plus autologous HSCT. The primary endpoint was progression -free survival at 12 months in all patients who met eligibility criteria and started treatment. The study was registered with the German clinical trial registry, DRKS00011932, and recruitment is complete. Findings Between Nov 28, 2017, and Sept 16, 2020, 54 patients started induction treatment and 51 were included in the full analysis set. Median age was 71 years (IQR 68-75); 27 (53%) patients were female and 24 (47%) were male. At a median follow-up of 23 center dot 0 months (IQR 16 center dot 8-37 center dot 4), 23 (45%) of 51 patients progressed, relapsed, or died. 12-month progression -free survival was 58 center dot 8% (80% CI 48 center dot 9-68 center dot 2; 95% CI 44 center dot 1-70 center dot 9). During induction treatment, the most common grade 3-5 toxicities were thrombocytopenia and leukopenia (each in 52 [96%] of 54 patients). During highdose chemotherapy and autologous HSCT, the most common grade 3-5 toxicity was leukopenia (37 [100%] of 37 patients). Treatment-related deaths were reported in three (6%) of 54 patients, all due to infectious complications. Interpretation Although the primary efficacy threshold was not met, short induction followed by high -dose chemotherapy and autologous HSCT is active in selected older patients with PCNSL and could serve as a benchmark for comparative trials. Funding Else Kroner-Fresenius Foundation, Riemser Pharma, and Medical Center-University of Freiburg. Copyright (c) 2024 Elsevier Ltd. All rights reserved.
C1 [Schorb, Elisabeth; Isbell, Lisa Kristina; Scherer, Florian; Burger-Martin, Elvira; Fricker, Heidi; Malenica, Natalie; Orban, Andras; Duyster, Justus; Ihorst, Gabriele; Finke, Juergen] Univ Freiburg, Dept Med 1, Fac Med, Med Ctr, D-79106 Freiburg, Germany.
   [Burger-Martin, Elvira; Grishina, Olga; Ihorst, Gabriele] Univ Freiburg, Clin Trials Unit, Fac Med, Med Ctr, Freiburg, Germany.
   [Kerkhoff, Andrea] Univ Hosp Muenster, Med Klin A, Haematol & Oncol, Munster, Germany.
   [Mathas, Stephan] Charite Univ Med Berlin, Haematol Oncol & Canc Immunol, Berlin, Germany.
   [Mathas, Stephan] Free Univ Berlin, Berlin, Germany.
   [Mathas, Stephan] HumboldtUniv Berlin, Berlin, Germany.
   [Mathas, Stephan] Helmholtz Assoc, Biol Malignant Lymphomas, Max Delbruck Ctr Mol Med, Berlin, Germany.
   [Mathas, Stephan] Helmholtz Assoc, Expt & Clin Res Ctr, Max Delbruck Ctr Mol Med, Berlin, Germany.
   [Mathas, Stephan] Charite Univ Med Berlin, Berlin, Germany.
   [Braulke, Friederike] Univ Med Ctr Gottingen, Comprehens Canc Ctr, Gottingen, Germany.
   [Braulke, Friederike] Univ Med Ctr Gottingen, Dept Haematol & Med Oncol, Gottingen, Germany.
   [Egerer, Gerlinde] Heidelberg Univ, Dept Haematol & Oncol, Heidelberg, Germany.
   [Roeth, Alexander] Univ Duisburg Essen, Univ Hosp Essen, West German Canc Ctr, Dept Hematol & Stem Cell Transplantat, Essen, Germany.
   [Schliffke, Simon] Univ Canc Ctr Hamburg, Univ Med Ctr Hamburg Eppendorf, Dept Oncol & Haematol, BMT Sect Pneumol, Hamburg, Germany.
   [Borchmann, Peter] Univ Cologne, Dept Internal Med 1, Cologne, Germany.
   [Deckert, Martina] Univ Cologne, Inst Neuropathol, Cologne, Germany.
   [Deckert, Martina] Univ Cologne, Fac Med, Cologne, Germany.
   [Deckert, Martina] Univ Cologne, Univ Hosp Cologne, Cologne, Germany.
   [Brunnberg, Uta] Univ Hosp Frankfurt, Dept Internal Med Haematol & Oncol 2, Frankfurt, Germany.
   [Kroschinsky, Frank] Dresden Univ Hosp, Med Dept 1, Dresden, Germany.
   [Moehle, Robert] Univ Tubingen, Dept Haematol & Oncol, Tubingen, Germany.
   [Rank, Andreas] Augsburg Med Ctr, Dept Hematol & Oncol, Augsburg, Germany.
   [Wellnitz, Dominique] Univ Clin Schleswig Holstein, Dept Internal Med Haematol & Oncol 2, Campus Kiel, Kiel, Germany.
   [Kasenda, Benjamin] Univ Hosp Basel, Dept Med Oncol, Basel, Switzerland.
   [Pospiech, Lisa; Wendler, Julia; Illerhaus, Gerald] Klinikum Stuttgart, Dept Paediat Haematol Oncol & Palliat Care, Stuttgart, Germany.
   [Henkes, Elina; von Gottberg, Philipp] Klinikum Stuttgart, Clin Neuroradiol, Stuttgart, Germany.
   [Gmehlin, Dennis; Backenstrass, Matthias] Klinikum Stuttgart, Inst Clin Psychol, Stuttgart, Germany.
   [Jensch, Antje] Klinikum Stuttgart, Stuttgart Canc Ctr, Tumorzentrum Eva Mayr Stihl, Stuttgart, Germany.
C3 University of Freiburg; University of Freiburg; University of Munster;
   Free University of Berlin; Humboldt University of Berlin; Charite
   Universitatsmedizin Berlin; Free University of Berlin; Helmholtz
   Association; Max Delbruck Center for Molecular Medicine; Helmholtz
   Association; Max Delbruck Center for Molecular Medicine; Free University
   of Berlin; Humboldt University of Berlin; Charite Universitatsmedizin
   Berlin; University of Gottingen; University of Gottingen; Ruprecht Karls
   University Heidelberg; University of Duisburg Essen; University of
   Hamburg; University Medical Center Hamburg-Eppendorf; University of
   Cologne; University of Cologne; University of Cologne; University of
   Cologne; Goethe University Frankfurt; Goethe University Frankfurt
   Hospital; Technische Universitat Dresden; Carl Gustav Carus University
   Hospital; Eberhard Karls University of Tubingen; University of Kiel;
   Schleswig Holstein University Hospital; University of Basel; Klinikum
   Stuttgart; Klinikum Stuttgart; Klinikum Stuttgart; Klinikum Stuttgart
RP Schorb, E (corresponding author), Univ Freiburg, Dept Med 1, Fac Med, Med Ctr, D-79106 Freiburg, Germany.
EM
RI Wendler, Julia S/IYT-3112-2023
FU Else Kroner-Fresenius Foundation [2016_A133]; Medical Center-University
   of Freiburg and Riemser Pharma; Clinical Trials Unit Freiburg
FX The study was sponsored by the Else Kroener-Fresenius Foundation (grant
   number 2016_A133) . Additional research funding was provided by the
   Medical Center-University of Freiburg and Riemser Pharma (Esteve
   Pharmaceuticals since November, 2021) . The funding bodies played no
   role in analysing the data or writing the manuscript. Decision for
   publication was solely made by the academic investigators. The study had
   an independent data safety monitoring board. We are indebted to our
   patients and their families for their dedication. We are grateful for
   the excellent support provided by the Clinical Trials Unit Freiburg. We
   also acknowledge haematologists, oncologists, neuroradiologists,
   pathologists, and psychologists from the 15 participating centres for
   their continued scientific collaboration. During the preparation of this
   work, ES used DeepL for language editing. After using this tool, ES
   reviewed and edited the content as needed and takes full responsibility
   for the content of the publication.
CR AALEN O, 1978, ANN STAT, V6, P701, DOI 10.1214/aos/1176344247
   Abrey LE, 2005, J CLIN ONCOL, V23, P5034, DOI 10.1200/JCO.2005.13.524
   Correa DD, 2007, ANN ONCOL, V18, P1145, DOI 10.1093/annonc/mdl464
   Deckert M, 2021, Lymphomas. WHO Classification of tumours editorial board central nervous system tumours, P351
   Fayers P, 2002, EUR J CANCER, V38, pS125
   Ferreri AJM, 2017, LANCET HAEMATOL, V4, pE510, DOI 10.1016/S2352-3026(17)30174-6
   Ferreri AJM, 2016, BLOOD, V127, P1642, DOI 10.1182/blood-2015-10-636340
   Fritsch K, 2017, LEUKEMIA, V31, P846, DOI 10.1038/leu.2016.334
   Houillier C, 2020, NEUROLOGY, V94, pE1027, DOI 10.1212/WNL.0000000000008900
   Houillier C, 2019, J CLIN ONCOL, V37, P823, DOI 10.1200/JCO.18.00306
   Illerhaus G, 2016, LANCET HAEMATOL, V3, pE388, DOI 10.1016/S2352-3026(16)30050-3
   Isbell LK, 2023, BMC CANCER, V23, DOI 10.1186/s12885-023-11193-7
   Kasenda B, 2015, ANN ONCOL, V26, P1305, DOI 10.1093/annonc/mdv076
   Kassam S, 2017, BONE MARROW TRANSPL, V52, P1268, DOI 10.1038/bmt.2017.101
   Martinez-Calle N, 2022, BRIT J HAEMATOL, V196, P473, DOI 10.1111/bjh.17799
   Mendez JS, 2018, NEURO-ONCOLOGY, V20, P687, DOI 10.1093/neuonc/nox187
   Omuro A, 2015, LANCET HAEMATOL, V2, pE251, DOI 10.1016/S2352-3026(15)00074-5
   Schorb E, 2017, BONE MARROW TRANSPL, V52, P1113, DOI 10.1038/bmt.2017.23
   Schorb E, 2021, CANCERS, V13, DOI 10.3390/cancers13174268
   Schorb E, 2020, BLOOD ADV, V4, P3378, DOI 10.1182/bloodadvances.2020002064
   Schorb E, 2016, BMC CANCER, V16, DOI 10.1186/s12885-016-2311-4
   Taphoorn MJB, 2010, EUR J CANCER, V46, P1033, DOI 10.1016/j.ejca.2010.01.012
   van der Meulen M, 2021, HAEMATOLOGICA, V106, P597, DOI 10.3324/haematol.2020.247536
   Villano JL, 2011, BRIT J CANCER, V105, P1414, DOI 10.1038/bjc.2011.357
   Welch MR, 2012, NEURO-ONCOLOGY, V14, P1304, DOI 10.1093/neuonc/nos207
   Wendler J, 2022, BMC CANCER, V22, DOI 10.1186/s12885-022-09723-w
NR 26
TC 1
Z9 1
U1 0
U2 0
PU ELSEVIER SCI LTD
PI London
PA 125 London Wall, London, ENGLAND
SN 2352-3026
J9 LANCET HAEMATOL
JI Lancet Haematol.
PD MAR
PY 2024
VL 11
IS 3
BP e196
EP e205
DI 10.1016/S2352-3026(23)00371-X
EA FEB 2024
PG 10
WC Hematology
WE Science Citation Index Expanded (SCI-EXPANDED)
SC Hematology
GA NO5B3
UT WOS:001201393500001
PM 38301670
DA 2024-06-03
ER

PT J
AU Rozenblum, L
   Houillier, C
   Baptiste, A
   Soussain, C
   Edeline, V
   Naggara, P
   Soret, M
   Causse-Lemercier, V
   Willems, L
   Choquet, S
   Ursu, R
   Galanaud, D
   Belin, L
   Hoang-Xuan, K
   Kas, A
AF Rozenblum, Laura
   Houillier, Caroline
   Baptiste, Amandine
   Soussain, Carole
   Edeline, Veronique
   Naggara, Philippe
   Soret, Marine
   Causse-Lemercier, Valerie
   Willems, Lise
   Choquet, Sylvain
   Ursu, Renata
   Galanaud, Damien
   Belin, Lisa
   Hoang-Xuan, Khe
   Kas, Aurelie
TI Interim FDG-PET improves treatment failure prediction in primary central
   nervous system lymphoma: An LOC network prospective multicentric study
SO NEURO-ONCOLOGY
LA English
DT Article; Early Access
DE early assessment; FDG; PET/MRI; primary central nervous system lymphoma;
   prognosis
ID PRIMARY CNS LYMPHOMA; B-CELL LYMPHOMA; TUMOR IMAGING PROTOCOL; CONSENSUS
   RECOMMENDATIONS; IMMUNOCOMPETENT PATIENTS; RESPONSE ASSESSMENT; INITIAL
   EVALUATION; MRI; METHOTREXATE; PROGNOSIS
AB Background. The purpose of our study was to assess the predictive and prognostic role of 2-18F-fluoro-2-deoxy-d-glucose (FDG) positron emission tomography (PET)/MRI during high-dose methotrexate-based chemotherapy (HD-MBC) in de novo primary central nervous system lymphoma (PCNSL) patients aged 60 and above. Methods. This prospective multicentric ancillary study included 65 immunocompetent patients who received induction HD-MBC as part of the BLOCAGE01 phase III trial. FDG-PET/MRI were acquired at baseline, post 2 cycles (PET/MRI2), and posttreatment (PET/MRI3). FDG-PET response was dichotomized with "positive" indicating persistent tumor uptake higher than the contralateral mirroring brain region. Performances of FDG-PET and International PCNSL Collaborative Group criteria in predicting induction response, progression-free survival (PFS), and overall survival (OS) were compared. Results. Of the 48 PET2 scans performed, 9 were positive and aligned with a partial response (PR) on MRI2. Among these, 8 (89%) progressed by the end of the induction phase. In contrast, 35/39 (90%) of PET2-negative patients achieved complete response (CR). Among the 18 discordant responses at interim (PETCR/MRIPR), 83% ultimately achieved CR. Eighty-seven percent of the PET2-negative patients were disease free at 6 months versus 11% of the PET2-positive patients (P < .001). The MRI2 response did not significantly differentiate patients based on their PFS, regardless of whether they were in CR or PR. Both PET2 and MRI2 independently predicted OS in multivariate analysis, with PET2 showing a stronger association. Conclusions. Our study highlights the potential of interim FDG-PET for early management of PCNSL patients. Response-driven treatment based on PET2 may guide future clinical trials.
C1 [Rozenblum, Laura; Naggara, Philippe; Soret, Marine; Causse-Lemercier, Valerie; Kas, Aurelie] Sorbonne Univ, Grp Hosp Pitie Salpetriere, AP HP, Dept Nucl Med, Paris, France.
   [Rozenblum, Laura; Soret, Marine; Galanaud, Damien; Kas, Aurelie] Sorbonne Univ, Lab Imagerie Biomed, CNRS, INSERM, Paris, France.
   [Houillier, Caroline; Hoang-Xuan, Khe] Sorbonne Univ, Hop Univ La Pitie Salpetriere Charles Foix, AP HP, Inst Cerveau,Dept Neurol Mazarin 2,Inserm,CNRS, Paris, France.
   [Baptiste, Amandine; Belin, Lisa] Sorbonne Univ, Hop Univ Pitie Salpetriere Charles Foix, AP HP, INSERM,Inst Pierre Louis Epidemiol & Sante Publ,De, Paris, France.
   [Soussain, Carole] Univ PSL, Inst Curie, Dept Haematol, Site St Cloud, Paris, France.
   [Soussain, Carole] Univ PSL, INSERM U932 Inst Curie, Paris, France.
   [Edeline, Veronique] Inst Curie, Dept Nucl Med, St Cloud, France.
   [Willems, Lise] Cochin Hosp, AP HP, Dept Haematol, Paris, France.
   [Choquet, Sylvain] Sorbonne Univ, Grp Hosp Pitie Salpetriere, AP HP, Dept Haematol, Paris, France.
   [Ursu, Renata] Hop St Louis, AP HP, Dept Neurol, Paris, France.
   [Galanaud, Damien] Sorbonne Univ, Grp Hosp Pitie Salpetriere, AP HP, Dept Neuroradiol, Paris, France.
   [Rozenblum, Laura] Grp Hosp Pitie Salpetriere, AP HP, Dept Nucl Med, 47-83 Blvd Hop, F-75013 Paris, France.
C3 Assistance Publique Hopitaux Paris (APHP); Hopital Universitaire
   Pitie-Salpetriere - APHP; Sorbonne Universite; Centre National de la
   Recherche Scientifique (CNRS); Institut National de la Sante et de la
   Recherche Medicale (Inserm); Sorbonne Universite; Assistance Publique
   Hopitaux Paris (APHP); Hopital Universitaire Pitie-Salpetriere - APHP;
   Centre National de la Recherche Scientifique (CNRS); Institut National
   de la Sante et de la Recherche Medicale (Inserm); Sorbonne Universite;
   Hopital Universitaire Charles-Foix - APHP; Assistance Publique Hopitaux
   Paris (APHP); Hopital Universitaire Pitie-Salpetriere - APHP; Sorbonne
   Universite; Institut National de la Sante et de la Recherche Medicale
   (Inserm); Hopital Universitaire Charles-Foix - APHP; UNICANCER;
   Universite PSL; Institut Curie; Universite PSL; UNICANCER; Universite
   PSL; Institut Curie; Assistance Publique Hopitaux Paris (APHP);
   Universite Paris Cite; Hopital Universitaire Cochin - APHP; Sorbonne
   Universite; Assistance Publique Hopitaux Paris (APHP); Hopital
   Universitaire Pitie-Salpetriere - APHP; Universite Paris Cite;
   Assistance Publique Hopitaux Paris (APHP); Hopital Universitaire
   Saint-Louis - APHP; Assistance Publique Hopitaux Paris (APHP); Hopital
   Universitaire Pitie-Salpetriere - APHP; Sorbonne Universite; Assistance
   Publique Hopitaux Paris (APHP); Hopital Universitaire Pitie-Salpetriere
   - APHP; Sorbonne Universite
RP Rozenblum, L (corresponding author), Grp Hosp Pitie Salpetriere, AP HP, Dept Nucl Med, 47-83 Blvd Hop, F-75013 Paris, France.
EM
OI Rozenblum, Laura/0000-0002-6306-8008
FU Assistance Publique-Hppitaux de Paris: PHRC LOCALYZE
FX This work was funded by Assistance Publique-Hppitaux de Paris: PHRC
   LOCALYZE.
CR Abrey LE, 2006, J CLIN ONCOL, V24, P5711, DOI 10.1200/JCO.2006.08.2941
   Ahn SY, 2018, CLIN NUCL MED, V43, pE259, DOI 10.1097/RLU.0000000000002154
   Albano D, 2018, J NEURO-ONCOL, V136, P577, DOI 10.1007/s11060-017-2686-3
   Ambady Prakash, 2021, Ann Lymphoma, V5, DOI 10.21037/aol-20-53
   Ambady P, 2017, FLUIDS BARRIERS CNS, V14, DOI 10.1186/s12987-017-0064-3
   Barajas RF, 2021, NEURO-ONCOLOGY, V23, P1056, DOI 10.1093/neuonc/noab020
   Birsen Rudy, 2018, Oncotarget, V9, P16822, DOI 10.18632/oncotarget.24706
   Boellaard R, 2015, EUR J NUCL MED MOL I, V42, P328, DOI 10.1007/s00259-014-2961-x
   Cheson BD, 2014, J CLIN ONCOL, V32, P3059, DOI 10.1200/JCO.2013.54.8800
   de-Bonilla-Damiá A, 2017, REV ESP MED NUCL IMA, V36, P298, DOI 10.1016/j.remn.2017.03.006
   Ellingson BM, 2015, NEURO-ONCOLOGY, V17, P1188, DOI 10.1093/neuonc/nov095
   Furutani K, 2008, J COMPUT ASSIST TOMO, V32, P313, DOI 10.1097/RCT.0b013e318074fd9d
   Hernández-Verdin I, 2023, ANN ONCOL, V34, P186, DOI 10.1016/j.annonc.2022.11.002
   Hoang-Xuan K, 2023, NEURO-ONCOLOGY, V25, P37, DOI 10.1093/neuonc/noac196
   Houillier C, 2020, NEUROLOGY, V94, pE1027, DOI 10.1212/WNL.0000000000008900
   Jang SJ, 2012, CLIN NUCL MED, V37, pE241, DOI 10.1097/RLU.0b013e318252d1ea
   Jo JC, 2017, ANN HEMATOL, V96, P1509, DOI 10.1007/s00277-017-3068-9
   Kaufmann TJ, 2020, NEURO-ONCOLOGY, V22, P757, DOI 10.1093/neuonc/noaa030
   Lauw MIS, 2020, DIAGNOSTICS, V10, DOI 10.3390/diagnostics10121076
   Lin C, 2007, J NUCL MED, V48, P1626, DOI 10.2967/jnumed.107.042093
   Maza S, 2013, RADIOL ONCOL, V47, P103, DOI 10.2478/raon-2013-0016
   Meignan M, 2009, LEUKEMIA LYMPHOMA, V50, P1257, DOI 10.1080/10428190903040048
   Mercadal S, 2015, MED CLIN-BARCELONA, V144, P503, DOI 10.1016/j.medcli.2014.09.010
   Morales-Martinez A, 2022, CURR OPIN ONCOL, V34, P676, DOI 10.1097/CCO.0000000000000896
   Morales-Martinez A, 2021, CANCERS, V13, DOI 10.3390/cancers13143479
   Ostergaard L, 1996, MAGN RESON MED, V36, P715, DOI 10.1002/mrm.1910360510
   Palmedo H, 2006, EUR J NUCL MED MOL I, V33, P164, DOI 10.1007/s00259-005-1917-6
   Postnov A, 2022, CLIN NUCL MED, V47, P699, DOI 10.1097/RLU.0000000000004238
   Pyatigorskaya N, 2020, CURR OPIN NEUROL, V33, P430, DOI 10.1097/WCO.0000000000000841
   Rachdi A, 2023, REV NEUROL-FRANCE, V179, P481, DOI 10.1016/j.neurol.2023.03.012
   Rekowski J, 2021, J NUCL MED, V62, P37, DOI 10.2967/jnumed.120.244145
   ROSEN BR, 1990, MAGNET RESON MED, V14, P249, DOI 10.1002/mrm.1910140211
   Rozenblum L, 2023, EUR J NUCL MED MOL I, V50, P3684, DOI 10.1007/s00259-023-06334-w
   Rozenblum L, 2022, CANCERS, V14, DOI 10.3390/cancers14174071
   Rozenblum L, 2020, CLIN NUCL MED, V45, pE206, DOI 10.1097/RLU.0000000000002959
   Schulte-Altedorneburg G, 2012, EUR J RADIOL, V81, P2380, DOI 10.1016/j.ejrad.2011.05.028
   Siddiqi T, 2021, BLOOD ADV, V5, P4059, DOI 10.1182/bloodadvances.2020004106
   Tabouret E, 2017, NEURO-ONCOLOGY, V19, P422, DOI 10.1093/neuonc/now238
   Thiel E, 2010, LANCET ONCOL, V11, P1036, DOI 10.1016/S1470-2045(10)70229-1
   Trotman J, 2021, LANCET HAEMATOL, V8, pE67, DOI 10.1016/S2352-3026(20)30357-4
   Valles FE, 2013, AM J NEURORADIOL, V34, P35, DOI 10.3174/ajnr.A3165
   van der Meulen M, 2021, NEURO-ONCOL ADV, V3, DOI 10.1093/noajnl/vdab007
   Zhai YJ, 2022, THER ADV MED ONCOL, V14, DOI 10.1177/17588359221093745
NR 43
TC 1
Z9 1
U1 0
U2 0
PU OXFORD UNIV PRESS INC
PI CARY
PA JOURNALS DEPT, 2001 EVANS RD, CARY, NC 27513 USA
SN 1522-8517
EI 1523-5866
J9 NEURO-ONCOLOGY
JI Neuro-Oncology
PD 2024 FEB 16
PY 2024
DI 10.1093/neuonc/noae029
EA FEB 2024
PG 10
WC Oncology; Clinical Neurology
WE Science Citation Index Expanded (SCI-EXPANDED)
SC Oncology; Neurosciences & Neurology
GA MA1B4
UT WOS:001190798800001
DA 2024-06-03
ER

PT J
AU Zuo, JY
   Lei, T
   Zhong, S
   Zhou, JJ
   Liu, R
   Wu, CX
   Li, SW
AF Zuo, Jinyi
   Lei, Ting
   Zhong, Shuai
   Zhou, Jiajun
   Liu, Rui
   Wu, Chenxing
   Li, Shouwei
TI C-reactive protein levels, the prognostic nutritional index, and the
   lactate dehydrogenase-to-lymphocyte ratio are important prognostic
   factors in primary central nervous system lymphoma: a single-center
   study of 223 patients
SO NEUROSURGICAL REVIEW
LA English
DT Article
DE Primary central nervous system lymphoma; C-reactive protein; Prognostic
   nutritional index; Lactate dehydrogenase-to-lymphocyte ratio; Prognostic
   factor; Elderly patients
ID CELL LYMPHOMA; INCREASING INCIDENCE; SURVIVAL; MODEL; IPI; INFLAMMATION;
   NEUTROPHIL; INDICATOR; COUNT; LDH
AB Primary central nervous system lymphoma (PCNSL) is a rare and highly aggressive type of extranodal non-Hodgkin lymphoma (NHL), and the prognosis is poor. Currently, the most used prognostic models are the Memorial Sloan-Kettering Cancer Center (MSKCC) and International Extranodal Lymphoma Study Group (IELSG) scores; however, their predictive effects are changing with increasing incidence and changing treatment regimens. A growing body of evidence has demonstrated that inflammatory and nutritional markers are factors that can determine tumor prognosis. Therefore, the aim of this study was to identify and validate novel prognostic factors for PCNSL. Clinical information was collected from 223 patients with PCNSL. Patients younger than 18 years of age were excluded. Progression-free survival (PFS) and overall survival (OS) were used as endpoints, and receiver operating characteristic (ROC) curve analyses were conducted to determine the cutoff values for the inflammatory indicators. Correlations between variables and PFS or OS were assessed using univariate and multivariate analyses, and positive indicators were selected for survival analysis. A prognostic nutritional index (PNI) < 49.38 was associated with worse PFS (p = 0.003), and outcomes significantly differed between patients with a PNI >= 49.38 and < 49.38 (p < 0.001). Age < 60 years (p < 0.001) and C-reactive protein (CRP) levels < 3.14 (p = 0.001) were associated with better OS. In elderly patients (>= 60 years), a lactate dehydrogenase-to-lymphocyte ratio (LLR) < 95.69 (p = 0.021) was associated with better OS, and the outcome significantly differed between patients with an LLR >= 95.69 and LLR < 95.69 (p = 0.015). The PNI and CRP levels are prognostic factors for PCNSL, and CRP was the first time shown to be a prognosis factor of PCNSL. In elderly patients with PCNSL, the LLR can predict prognosis.
C1 [Zuo, Jinyi; Lei, Ting; Zhong, Shuai; Zhou, Jiajun; Liu, Rui; Wu, Chenxing; Li, Shouwei] Capital Med Univ, Sanbo Brain Hosp, Dept Neurooncol, Beijing, Peoples R China.
C3 Capital Medical University
RP Li, SW (corresponding author), Capital Med Univ, Sanbo Brain Hosp, Dept Neurooncol, Beijing, Peoples R China.
EM
CR Allison SJ, 2014, ONCOGENESIS, V3, DOI 10.1038/oncsis.2014.16
   [Anonymous], 2021, World Health Organization Classification of Tumours of the Central Nervous System
   Campo E, 2022, BLOOD, V140, P1229, DOI 10.1182/blood.2022015851
   Cheson BD, 1999, J CLIN ONCOL, V17, P1244, DOI 10.1200/JCO.1999.17.4.1244
   Corry J, 1998, INT J RADIAT ONCOL, V41, P615, DOI 10.1016/S0360-3016(97)00571-3
   Coussens LM, 2002, NATURE, V420, P860, DOI 10.1038/nature01322
   David KA, 2023, AM J HEMATOL, V98, P900, DOI 10.1002/ajh.26919
   Deng XY, 2020, FRONT ONCOL, V10, DOI 10.3389/fonc.2020.00131
   Ding J, 2017, CANCER BIOMARK, V19, P353, DOI 10.3233/CBM-160336
   Eloranta S, 2018, EUR J HAEMATOL, V100, P61, DOI 10.1111/ejh.12980
   Farrall AL, 2021, CANCERS, V13, DOI 10.3390/cancers13030403
   Ferreri AJM, 2023, NAT REV DIS PRIMERS, V9, DOI 10.1038/s41572-023-00439-0
   Gao YT, 2021, FRONT ONCOL, V11, DOI 10.3389/fonc.2021.696147
   Go SI, 2018, TUMORI J, V104, P292, DOI 10.5301/tj.5000694
   Gupta D, 2010, NUTR J, V9, DOI 10.1186/1475-2891-9-69
   Gupta Tejpal, 2022, CNS Oncol, V11, pCNS83, DOI 10.2217/cns-2022-0004
   Haldorsen IS, 2007, CANCER, V110, P1803, DOI 10.1002/cncr.22989
   Hayabuchi N, 1999, INT J RADIAT ONCOL, V44, P265, DOI 10.1016/S0360-3016(98)00564-1
   Huang JJ, 2011, ANN ONCOL, V22, P149, DOI 10.1093/annonc/mdq314
   Jang JE, 2016, EUR J CANCER, V57, P127, DOI 10.1016/j.ejca.2016.01.016
   Ji HF, 2018, CELL PHYSIOL BIOCHEM, V45, P951, DOI 10.1159/000487288
   Larsen Maja Thim, 2016, Mol Cell Ther, V4, P3
   Li SJ, 2022, FRONT IMMUNOL, V13, DOI 10.3389/fimmu.2022.1039862
   Liu CJ, 2020, CANCER MED-US, V9, P2134, DOI 10.1002/cam4.2872
   Moujaess E, 2017, CRIT REV ONCOL HEMAT, V120, P203, DOI 10.1016/j.critrevonc.2017.11.008
   Ni LW, 2022, FRONT NUTR, V9, DOI 10.3389/fnut.2022.823087
   Onodera T, 1984, Nihon Geka Gakkai Zasshi, V85, P1001
   Park JH, 2014, ANN HEMATOL, V93, P1755, DOI 10.1007/s00277-014-2115-z
   Passardi A, 2015, PLOS ONE, V10, DOI 10.1371/journal.pone.0134732
   Pavlidis A. N., 1993, International Journal of Biological Markers, V8, P14
   Ren HR, 2017, ONCOTARGET, V8, P61365, DOI 10.18632/oncotarget.18135
   Schorb E, 2013, HAEMATOLOGICA, V98, P765, DOI 10.3324/haematol.2012.076075
   Schuetz P, 2019, LANCET, V393, P2312, DOI 10.1016/S0140-6736(18)32776-4
   Troppan K, 2014, BRIT J CANCER, V110, P369, DOI 10.1038/bjc.2013.763
   Troppan KT, 2014, BRIT J CANCER, V111, P55, DOI 10.1038/bjc.2014.277
   Tucci A, 2009, CANCER, V115, P4547, DOI 10.1002/cncr.24490
   Turner DC, 2018, CLIN CANCER RES, V24, P5841, DOI 10.1158/1078-0432.CCR-18-0415
   Velasco R, 2020, J NEURO-ONCOL, V148, P545, DOI 10.1007/s11060-020-03547-z
   Wei L, 2023, J NEURO-ONCOL, V163, P301, DOI 10.1007/s11060-023-04337-z
   Wieduwilt MJ, 2012, CLIN CANCER RES, V18, P1146, DOI 10.1158/1078-0432.CCR-11-0625
   Wong CE, 2023, CLIN EXP MED, V23, P3799, DOI 10.1007/s10238-023-01130-0
   Wu ZT, 2023, FRONT ONCOL, V13, DOI 10.3389/fonc.2023.1104425
NR 42
TC 1
Z9 1
U1 2
U2 2
PU SPRINGER
PI NEW YORK
PA ONE NEW YORK PLAZA, SUITE 4600, NEW YORK, NY, UNITED STATES
SN 0344-5607
EI 1437-2320
J9 NEUROSURG REV
JI Neurosurg. Rev.
PD DEC 19
PY 2023
VL 47
IS 1
AR 17
DI 10.1007/s10143-023-02248-1
PG 11
WC Clinical Neurology; Surgery
WE Science Citation Index Expanded (SCI-EXPANDED)
SC Neurosciences & Neurology; Surgery
GA CS9T9
UT WOS:001127355000001
PM 38112846
OA Green Submitted, Green Published, hybrid
DA 2024-06-03
ER

PT J
AU Karschnia, P
   Arrillaga-Romany, IC
   Eichler, A
   Forst, DA
   Gerstner, E
   Jordan, JT
   Ly, I
   Plotkin, SR
   Wang, NY
   Martinez-Lage, M
   Winter, SF
   Tonn, JC
   Rejeski, K
   von Baumgarten, L
   Cahill, DP
   Nahed, B
   Shankar, GM
   Abramson, JS
   Barnes, JA
   El-Jawahri, A
   Hochberg, EP
   Johnson, PC
   Soumerai, JD
   Takvorian, RW
   Chen, YB
   Frigault, MJ
   Dietrich, J
AF Karschnia, Philipp
   Arrillaga-Romany, Isabel C.
   Eichler, April
   Forst, Deborah A.
   Gerstner, Elizabeth
   Jordan, Justin T.
   Ly, Ina
   Plotkin, Scott R.
   Wang, Nancy
   Martinez-Lage, Maria
   Winter, Sebastian F.
   Tonn, Joerg-Christian
   Rejeski, Kai
   von Baumgarten, Louisa
   Cahill, Daniel P.
   Nahed, Brian, V
   Shankar, Ganesh M.
   Abramson, Jeremy S.
   Barnes, Jeffrey A.
   El-Jawahri, Areej
   Hochberg, Ephraim P.
   Johnson, P. Connor
   Soumerai, Jacob D.
   Takvorian, Ronald W.
   Chen, Yi-Bin
   Frigault, Matthew J.
   Dietrich, Jorg
TI Neurotoxicity and management of primary and secondary central nervous
   system lymphoma after adoptive immunotherapy with CD19-directed chimeric
   antigen receptor T-cells
SO NEURO-ONCOLOGY
LA English
DT Article
DE CNS lymphoma; chimeric antigen receptor; CAR T-cells; neurotoxicity;
   response
ID PRIMARY CNS LYMPHOMA; EFFICACY; THERAPY
AB Background Chimeric antigen receptor (CAR) T-cells targeting CD19 have been established as a leading engineered T-cell therapy for B-cell lymphomas; however, data for patients with central nervous system (CNS) involvement are limited.Methods We retrospectively report on CNS-specific toxicities, management, and CNS response of 45 consecutive CAR T-cell transfusions for patients with active CNS lymphoma at the Massachusetts General Hospital over a 5-year period.Results Our cohort includes 17 patients with primary CNS lymphoma (PCNSL; 1 patient with 2 CAR T-cell transfusions) and 27 patients with secondary CNS lymphoma (SCNSL). Mild ICANS (grade 1-2) was observed after 19/45 transfusions (42.2%) and severe immune effector cell-associated neurotoxicity syndrome (ICANS) (grade 3-4) after 7/45 transfusions (15.6%). A larger increase in C-reactive protein (CRP) levels and higher rates of ICANS were detected in SCNSL. Early fever and baseline C-reactive protein levels were associated with ICANS occurrence. CNS response was seen in 31 cases (68.9%), including a complete response of CNS disease in 18 cases (40.0%) which lasted for a median of 11.4 +/- 4.5 months. Dexamethasone dose at time of lymphodepletion (but not at or after CAR T-cell transfusion) was associated with an increased risk for CNS progression (hazard ratios [HR] per mg/d: 1.16, P = .031). If bridging therapy was warranted, the use of ibrutinib translated into favorable CNS-progression-free survival (5 vs. 1 month, HR 0.28, CI 0.1-0.7; P = .010).Conclusions CAR T-cells exhibit promising antitumor effects and a favorable safety profile in CNS lymphoma. Further evaluation of the role of bridging regimens and corticosteroids is warranted.
C1 [Dietrich, Jorg] Harvard Med Sch, Massachusetts Gen Hosp, Canc Ctr, Div Neurooncol,Dept Neurol, 55 Fruit St,Yawkey 9E, Boston, MA 02114 USA.
   [Karschnia, Philipp; Arrillaga-Romany, Isabel C.; Eichler, April; Forst, Deborah A.; Gerstner, Elizabeth; Jordan, Justin T.; Ly, Ina; Plotkin, Scott R.; Wang, Nancy; Winter, Sebastian F.; Dietrich, Jorg] Harvard Med Sch, Massachusetts Gen Hosp, Canc Ctr, Div Neurooncol,Dept Neurol, Boston, MA USA.
   [Karschnia, Philipp; Tonn, Joerg-Christian; von Baumgarten, Louisa] Ludwig Maximilians Univ Munchen, Dept Neurosurg, Sect Neurooncol, Munich, Germany.
   [Karschnia, Philipp; Tonn, Joerg-Christian; Rejeski, Kai] German Canc Consortium DKTK, Partner Site Munich, Munich, Germany.
   [Martinez-Lage, Maria] Harvard Med Sch, Massachusetts Gen Hosp, Dept Pathol, Div Neuropathol, Boston, MA USA.
   [Rejeski, Kai] Ludwig Maximilians Univ Munchen, Dept Med 3, Sect Cellular Immunotherapy, Munich, Germany.
   [Cahill, Daniel P.; Nahed, Brian, V; Shankar, Ganesh M.] Harvard Med Sch, Massachusetts Gen Hosp, Dept Neurosurg, Boston, MA USA.
   [Abramson, Jeremy S.; Barnes, Jeffrey A.; El-Jawahri, Areej; Hochberg, Ephraim P.; Johnson, P. Connor; Soumerai, Jacob D.; Takvorian, Ronald W.] Harvard Med Sch, Massachusetts Gen Hosp, Dept Med, Oncol Div,Canc Ctr, Boston, MA USA.
   [Chen, Yi-Bin; Frigault, Matthew J.] Harvard Med Sch, Massachusetts Gen Hosp, Canc Ctr, Cellular Immunotherapy Program,Dept Med, Boston, MA USA.
C3 Harvard University; Massachusetts General Hospital; Harvard Medical
   School; Harvard University; Harvard Medical School; Massachusetts
   General Hospital; University of Munich; Helmholtz Association; German
   Cancer Research Center (DKFZ); Harvard University; Massachusetts General
   Hospital; Harvard Medical School; University of Munich; Harvard
   University; Massachusetts General Hospital; Harvard Medical School;
   Harvard University; Massachusetts General Hospital; Harvard Medical
   School; Harvard University; Massachusetts General Hospital; Harvard
   Medical School
RP Dietrich, J (corresponding author), Harvard Med Sch, Massachusetts Gen Hosp, Canc Ctr, Div Neurooncol,Dept Neurol, 55 Fruit St,Yawkey 9E, Boston, MA 02114 USA.
EM
RI Rejeski, Kai/AIE-6115-2022; Winter, Sebastian Friedrich/ABF-8919-2020
OI Rejeski, Kai/0000-0003-3905-0251; Winter, Sebastian
   Friedrich/0000-0001-8836-0235; Plotkin, Scott/0000-0002-6109-6419;
   Karschnia, Philipp/0000-0002-1254-5310
FU Massachusetts General Hospital Cancer Center
FX The authors thank all patients who contributed to the results of the
   study. We appreciate the support of the excellent nurses, staff, and
   providers of the Cellular Immunotherapy Program and the Massachusetts
   General Hospital Cancer Center. Figure 3 has been created with the help
   of BioRender.com.
CR Abramson JS, 2020, LANCET, V396, P839, DOI 10.1016/S0140-6736(20)31366-0
   Abrey LE, 2005, J CLIN ONCOL, V23, P5034, DOI 10.1200/JCO.2005.13.524
   Ahmed G, 2021, BLOOD ADV, V5, P5626, DOI 10.1182/bloodadvances.2021005292
   Alcantara M, 2022, BLOOD, V139, P792, DOI 10.1182/blood.2021012932
   Bachy E, 2022, NAT MED, V28, P2145, DOI 10.1038/s41591-022-01969-y
   Cook MR, 2023, BLOOD ADV, V7, P32, DOI 10.1182/bloodadvances.2022008525
   Ferreri AJM, 2022, LEUKEMIA, V36, P1870, DOI 10.1038/s41375-022-01582-5
   Ferreri AJM, 2021, LANCET HAEMATOL, V8, pe110, DOI 10.1016/S2352-3026(20)30366-5
   Ferreri AJM, 2016, LANCET HAEMATOL, V3, pE217, DOI 10.1016/S2352-3026(16)00036-3
   Fraietta JA, 2016, BLOOD, V127, P1117, DOI 10.1182/blood-2015-11-679134
   Frigault MJ, 2022, BLOOD, V139, P2306, DOI 10.1182/blood.2021014738
   Frigault MJ, 2019, BLOOD, V134, P860, DOI 10.1182/blood.2019001694
   Gauthier J, 2020, BLOOD, V135, P1650, DOI 10.1182/blood.2019002936
   Hirayama AV, 2019, BLOOD, V133, P1876, DOI 10.1182/blood-2018-11-887067
   Karschnia P, 2022, NEUROLOGY, V98, P884, DOI 10.1212/WNL.0000000000200608
   Karschnia P, 2021, NEUROLOGY, V97, P218, DOI 10.1212/WNL.0000000000012193
   Karschnia P, 2019, BLOOD, V133, P2212, DOI 10.1182/blood-2018-12-893396
   Langner-Lemercier S, 2016, NEURO-ONCOLOGY, V18, P1297, DOI 10.1093/neuonc/now033
   Lee DW, 2019, BIOL BLOOD MARROW TR, V25, P625, DOI 10.1016/j.bbmt.2018.12.758
   Liu SY, 2020, BLOOD CANCER J, V10, DOI 10.1038/s41408-020-0280-y
   Long MX, 2017, J CLIN INVEST, V127, P3052, DOI 10.1172/JCI89756
   Mahdi J, 2023, NAT MED, V29, P803, DOI 10.1038/s41591-023-02276-w
   Neelapu SS, 2017, NEW ENGL J MED, V377, P2531, DOI 10.1056/NEJMoa1707447
   Rejeski K, 2022, J IMMUNOTHER CANCER, V10, DOI 10.1136/jitc-2021-004475
   Schuster SJ, 2019, NEW ENGL J MED, V380, P45, DOI 10.1056/NEJMoa1804980
   Siddiqi T, 2021, BLOOD ADV, V5, P4059, DOI 10.1182/bloodadvances.2020004106
   Strati P, 2021, BLOOD, V137, P3272, DOI 10.1182/blood.2020008865
   Xu T, 2023, ONCOIMMUNOLOGY, V12, DOI 10.1080/2162402X.2022.2163781
NR 28
TC 1
Z9 1
U1 0
U2 0
PU OXFORD UNIV PRESS INC
PI CARY
PA JOURNALS DEPT, 2001 EVANS RD, CARY, NC 27513 USA
SN 1522-8517
EI 1523-5866
J9 NEURO-ONCOLOGY
JI Neuro-Oncology
PD DEC 8
PY 2023
VL 25
IS 12
BP 2239
EP 2249
DI 10.1093/neuonc/noad118
PG 11
WC Oncology; Clinical Neurology
WE Science Citation Index Expanded (SCI-EXPANDED)
SC Oncology; Neurosciences & Neurology
GA CJ2A7
UT WOS:001124808300003
PM 37402650
DA 2024-06-03
ER

PT J
AU Hosoi, H
   Okamura, T
   Fukai, J
   Hori, Y
   Murata, S
   Mushino, T
   Nakao, N
   Sonoki, T
AF Hosoi, Hiroki
   Okamura, Tadashi
   Fukai, Junya
   Hori, Yoshikazu
   Murata, Shogo
   Mushino, Toshiki
   Nakao, Naoyuki
   Sonoki, Takashi
TI Comparison of the outcomes of patients with primary central nervous
   system lymphoma between the neurosurgery and hematology/oncology
   departments based on the relative dose intensity of methotrexate
SO ONCOLOGY LETTERS
LA English
DT Article
DE primary central nervous system lymphoma; treating department; dose
   intensity; methotrexate
ID PRIMARY CNS LYMPHOMA; INTERNATIONAL EXTRANODAL LYMPHOMA; ACUTE
   LYMPHOBLASTIC-LEUKEMIA; WHOLE-BRAIN RADIOTHERAPY; B-CELL LYMPHOMA;
   YOUNG-ADULTS; ADOLESCENTS; RITUXIMAB; CHEMOIMMUNOTHERAPY; RANDOMIZATION
AB Primary central nervous system lymphoma (PCNSL) is an extranodal type of lymphoma, which is treated with methotrexate (MTX)-based induction therapy. Although PCNSL is a hematological malignancy, patients with PCNSL may be treated at neurosurgery or hematology/oncology departments; however, the outcomes of PCNSL treatment have not been compared between these two departments. The present study compared the outcomes of 26 patients with newly diagnosed PCNSL that were treated at the Department of Neurological Surgery or Department of Hematology/Oncology (Wakayama Medical University Hospital, Wakayama, Japan) between January 2011 and December 2021. The relative dose intensity (RDI) and relative treatment intensity of MTX were assessed as indicators of the intensity of chemotherapy. The median RDI of MTX was 67 and 93% in the neurosurgery and hematology/oncology groups, respectively (P<0.001). The proportion of patients that achieved a complete response after high-dose MTX-based therapy was significantly higher in the hematology/oncology group than in the neurosurgery group (P=0.038). The estimated 2-year overall survival was 72 and 100% in the neurosurgery and hematology/oncology groups, respectively (P=0.046). As with the difference in the outcomes observed between pediatrics and hematology departments for adolescents with acute lymphoblastic leukemia, the outcomes of patients with PCNSL may differ between neurosurgery and hematology/oncology departments.
C1 [Hosoi, Hiroki; Okamura, Tadashi; Hori, Yoshikazu; Murata, Shogo; Mushino, Toshiki; Sonoki, Takashi] Wakayama Med Univ, Dept Hematol Oncol, 811-1 Kimiidera, Wakayama 6418509, Japan.
   [Fukai, Junya; Nakao, Naoyuki] Wakayama Med Univ, Dept Neurol Surg, Wakayama 6418509, Japan.
C3 Wakayama Medical University; Wakayama Medical University
RP Hosoi, H (corresponding author), Wakayama Med Univ, Dept Hematol Oncol, 811-1 Kimiidera, Wakayama 6418509, Japan.
EM
RI Sonoki, Takashi/KGM-2990-2024
FU Not applicable.
FX Not applicable.
CR Abrey LE, 2006, J CLIN ONCOL, V24, P5711, DOI 10.1200/JCO.2006.08.2941
   Abrey LE, 2005, J CLIN ONCOL, V23, P5034, DOI 10.1200/JCO.2005.13.524
   Bataillard EJ, 2021, BLOOD ADV, V5, P2426, DOI 10.1182/bloodadvances.2021004665
   Boissel N, 2003, J CLIN ONCOL, V21, P774, DOI 10.1200/JCO.2003.02.053
   Calimeriy T, 2021, ESMO OPEN, V6, DOI 10.1016/j.esmoop.2021.100213
   Farhi J, 2018, ANN HEMATOL, V97, P2391, DOI 10.1007/s00277-018-3468-5
   Ferreri AJM, 2003, J CLIN ONCOL, V21, P266, DOI 10.1200/JCO.2003.09.139
   Ferreri AJM, 2017, LANCET HAEMATOL, V4, pE510, DOI 10.1016/S2352-3026(17)30174-6
   Ferreri AJM, 2016, LANCET HAEMATOL, V3, pE217, DOI 10.1016/S2352-3026(16)00036-3
   Hirakawa T, 2010, ANN HEMATOL, V89, P897, DOI 10.1007/s00277-010-0956-7
   Hiroi T, 2023, J GERIATR ONCOL, V14, DOI 10.1016/j.jgo.2022.10.011
   Hoang-Xuan K, 2015, LANCET ONCOL, V16, pE322, DOI 10.1016/S1470-2045(15)00076-5
   Houillier C, 2019, J CLIN ONCOL, V37, P823, DOI 10.1200/JCO.18.00306
   Kanda Y, 2013, BONE MARROW TRANSPL, V48, P452, DOI 10.1038/bmt.2012.244
   Kasenda B, 2015, ANN ONCOL, V26, P1305, DOI 10.1093/annonc/mdv076
   Kondo E, 2019, CANCER CHEMOTH PHARM, V84, P849, DOI 10.1007/s00280-019-03914-2
   LEPAGE E, 1993, ANN ONCOL, V4, P651, DOI 10.1093/oxfordjournals.annonc.a058619
   Morris PG, 2013, J CLIN ONCOL, V31, P3971, DOI 10.1200/JCO.2013.50.4910
   Okamura T, 2022, ANN HEMATOL, V101, P1379, DOI 10.1007/s00277-021-04744-0
   Patel AM, 2022, NEURO-ONCOL PRACT, V9, P183, DOI 10.1093/nop/npab066
   Sasaki N, 2020, JPN J CLIN ONCOL, V50, P999, DOI 10.1093/jjco/hyaa073
   Schaff LR, 2022, BLOOD, V140, P971, DOI 10.1182/blood.2020008377
   Schiffer CA, 2003, J CLIN ONCOL, V21, P760, DOI 10.1200/JCO.2003.11.116
   Stock W, 2008, BLOOD, V112, P1646, DOI 10.1182/blood-2008-01-130237
   Stock W, 2010, HEMATOL-AM SOC HEMAT, P21, DOI 10.1182/asheducation-2010.1.21
NR 25
TC 1
Z9 1
U1 0
U2 0
PU SPANDIDOS PUBL LTD
PI ATHENS
PA POB 18179, ATHENS, 116 10, GREECE
SN 1792-1074
EI 1792-1082
J9 ONCOL LETT
JI Oncol. Lett.
PD NOV
PY 2023
VL 26
IS 5
AR 469
DI 10.3892/ol.2023.14056
PG 9
WC Oncology
WE Science Citation Index Expanded (SCI-EXPANDED)
SC Oncology
GA U0AW1
UT WOS:001081529700001
PM 37809042
OA gold, Green Published
DA 2024-06-03
ER

PT J
AU Liu, X
   Yao, C
   Chen, HY
   Xiang, R
   Wu, H
   Du, P
   Yu, ZK
   Liu, WF
   Liu, J
   Geng, DY
AF Liu, Xiao
   Yao, Chong
   Chen, Hongyi
   Xiang, Rui
   Wu, Hao
   Du, Peng
   Yu, Zekuan
   Liu, Weifan
   Liu, Jie
   Geng, Daoying
TI BTSC-TNAS: A neural architecture search-based transformer for brain
   tumor segmentation and classification
SO COMPUTERIZED MEDICAL IMAGING AND GRAPHICS
LA English
DT Article
DE Transformer; Segmentation; Classification; Neural architecture search;
   Brain tumors multi-modal MR images
ID INFORMATION
AB Glioblastoma (GBM), isolated brain metastasis (SBM), and primary central nervous system lymphoma (PCNSL) possess a high level of similarity in histomorphology and clinical manifestations on multimodal MRI. Such similarities have led to challenges in the clinical diagnosis of these three malignant tumors. However, many existing models solely focus on either the task of segmentation or classification, which limits the application of computer-aided diagnosis in clinical diagnosis and treatment. To solve this problem, we propose a multi-task learning transformer with neural architecture search (NAS) for brain tumor segmentation and classification (BTSC-TNAS). In the segmentation stage, we use a nested transformer U-shape network (NTU-NAS) with NAS to directly predict brain tumor masks from multi-modal MRI images. In the tumor classification stage, we use the multiscale features obtained from the encoder of NTU-NAS as the input features of the classification network (MSC-NET), which are integrated and corrected by the classification feature correction enhancement (CFCE) block to improve the accuracy of classification. The proposed BTSC-TNAS achieves an average Dice coefficient of 80.86% and 87.12% for the segmentation of tumor region and the maximum abnormal region in clinical data respectively. The model achieves a classification accuracy of 0.941. The experiments performed on the BraTS 2019 dataset show that the proposed BTSC-TNAS has excellent generalizability and can provide support for some challenging tasks in the diagnosis and treatment of brain tumors.
C1 [Liu, Xiao; Liu, Jie] Beijing Jiaotong Univ, Sch Comp & Informat Technol, Beijing 100044, Peoples R China.
   [Yao, Chong] China Agr Univ, Coll Informat & Elect Engn, Beijing 100083, Peoples R China.
   [Chen, Hongyi; Xiang, Rui; Yu, Zekuan] Fudan Univ, Acad Engn & Technol, Shanghai 200433, Peoples R China.
   [Wu, Hao] Fudan Univ, Huashan Hosp, Dept Dermatol, Shanghai 200040, Peoples R China.
   [Du, Peng; Geng, Daoying] Fudan Univ, Huashan Hosp, Dept Radiol, Shanghai 200040, Peoples R China.
   [Liu, Weifan] Beijing Forestry Univ, Coll Sci, Beijing 100083, Peoples R China.
C3 Beijing Jiaotong University; China Agricultural University; Fudan
   University; Fudan University; Fudan University; Beijing Forestry
   University
RP Liu, J (corresponding author), Beijing Jiaotong Univ, Sch Comp & Informat Technol, Beijing 100044, Peoples R China.
EM
RI Chen, Yang/KHD-8849-2024; li, cheng/KCZ-0615-2024; guo,
   yi/KHC-4669-2024; li, feiyang/KHW-5210-2024; Sun, Yue/KHU-8159-2024; li,
   li/KHE-5750-2024; zhang, hao/KEJ-2291-2024; wang, nan/KHW-4897-2024;
   Lin, Fan/JZT-1441-2024; li, qing/KHU-6871-2024; yang,
   xiao/KHT-9445-2024; Liu, Yu/KFS-0769-2024; zhu, hao/KHW-3813-2024; su,
   lin/KHC-5034-2024; li, jing/KHC-8303-2024; zhang, yan/KHC-3163-2024;
   Zhang, Lu/KHE-5879-2024; Zhang, Yi/KHW-2039-2024; liu, qi/KFA-4047-2024;
   wang, jin/KHD-7243-2024; liu, qi/KHC-7509-2024; ren, jun/KHG-7717-2024
OI Lin, Fan/0000-0002-7330-3833; Liu, Weifan/0000-0002-0710-9311
CR Balamurugan T, 2023, NEURAL COMPUT APPL, V35, P4739, DOI 10.1007/s00521-022-07934-7
   Cai H, 2019, Arxiv, DOI arXiv:1812.00332
   Cao Hu, 2023, Computer Vision - ECCV 2022 Workshops: Proceedings. Lecture Notes in Computer Science (13803), P205, DOI 10.1007/978-3-031-25066-8_9
   Chen J., 2021, arXiv
   Dushatskiy A., 2022, SPIE, V12032, P193, DOI 10.48550/arXiv.2202.11401
   Elsken T, 2019, J MACH LEARN RES, V20
   Farajzadeh N, 2023, EXPERT SYST APPL, V224, DOI 10.1016/j.eswa.2023.119963
   Gao YH, 2021, LECT NOTES COMPUT SC, V12903, P61, DOI 10.1007/978-3-030-87199-4_6
   Ghosal P., 2019, P 2019 2 INT C ADV C, P1, DOI [10.1109/ICACCP.2019.8882973, DOI 10.1109/ICACCP.2019.8882973]
   Habib H, 2021, J AMB INTEL HUM COMP, DOI 10.1007/s12652-021-03544-8
   Hammerla N.Y., 2018, arXiv, DOI [DOI 10.48550/ARXIV.1804, 10.48550/arXiv.1804]
   Han K, 2023, IEEE T PATTERN ANAL, V45, P87, DOI 10.1109/TPAMI.2022.3152247
   Hatamizadeh A, 2022, IEEE WINT CONF APPL, P1748, DOI 10.1109/WACV51458.2022.00181
   Huang G, 2017, PROC CVPR IEEE, P2261, DOI 10.1109/CVPR.2017.243
   Hussain T., A comparative analysis of efficient CNN-based brain tumor classification models, P259, DOI [10.1142/9789811218842_0011, DOI 10.1142/9789811218842_0011]
   Jin X, 2022, PATTERN RECOGN, V121, DOI 10.1016/j.patcog.2021.108159
   Karlsson RA, 2022, COMPUT METH PROG BIO, V216, DOI 10.1016/j.cmpb.2022.106650
   Khan S, 2022, ACM COMPUT SURV, V54, DOI 10.1145/3505244
   Lambin P, 2012, EUR J CANCER, V48, P441, DOI 10.1016/j.ejca.2011.11.036
   Lin AL, 2022, IEEE T INSTRUM MEAS, V71, DOI 10.1109/TIM.2022.3178991
   Liu CX, 2019, PROC CVPR IEEE, P82, DOI 10.1109/CVPR.2019.00017
   Liu YQ, 2023, IEEE T NEUR NET LEAR, V34, P550, DOI 10.1109/TNNLS.2021.3100554
   Lyu Q, 2020, IEEE T MED IMAGING, V39, P2738, DOI 10.1109/TMI.2020.2974858
   Milletari F, 2016, INT CONF 3D VISION, P565, DOI 10.1109/3DV.2016.79
   Neelima G, 2022, BIOMED SIGNAL PROCES, V74, DOI 10.1016/j.bspc.2022.103537
   Neupane B, 2023, Arxiv, DOI arXiv:2303.09064
   Okuchi S, 2020, RADIOL-IMAG CANCER, V2, DOI 10.1148/rycan.2020190036
   Pinaya WHL, 2022, MED IMAGE ANAL, V79, DOI 10.1016/j.media.2022.102475
   Qin XB, 2020, PATTERN RECOGN, V106, DOI 10.1016/j.patcog.2020.107404
   Qin XB, 2019, PROC CVPR IEEE, P7471, DOI 10.1109/CVPR.2019.00766
   Rehman A, 2020, CIRC SYST SIGNAL PR, V39, P757, DOI 10.1007/s00034-019-01246-3
   Ronneberger O, 2015, LECT NOTES COMPUT SC, V9351, P234, DOI 10.1007/978-3-319-24574-4_28
   Sadad T, 2021, MICROSC RES TECHNIQ, V84, P1296, DOI 10.1002/jemt.23688
   Sehgal A, 2016, 2016 CONFERENCE ON ADVANCES IN SIGNAL PROCESSING (CASP), P104
   Shuying Xu, 2021, 2021 IEEE International Conference on Bioinformatics and Biomedicine (BIBM), P1601, DOI 10.1109/BIBM52615.2021.9669734
   Sobhaninia Z, 2023, BIOMED SIGNAL PROCES, V85, DOI 10.1016/j.bspc.2023.104834
   Sudre CH, 2017, LECT NOTES COMPUT SC, V10553, P240, DOI 10.1007/978-3-319-67558-9_28
   Tan MX, 2021, PR MACH LEARN RES, V139, P7102
   Wang SQ, 2022, TRANSL CANCER RES, V11, P63, DOI 10.21037/tcr-21-1957
   Wang WX, 2021, LECT NOTES COMPUT SC, V12901, P109, DOI 10.1007/978-3-030-87193-2_11
   Wei J., 2020, arXiv, DOI [10.48550/arXiv.2010.15560, DOI 10.48550/ARXIV.2010.15560]
   Weng Y, 2019, IEEE ACCESS, V7, P44247, DOI 10.1109/ACCESS.2019.2908991
   Xingang Yan, 2020, Medical Image Computing and Computer Assisted Intervention - MICCAI 2020. 23rd International Conference. Proceedings. Lecture Notes in Computer Science (LNCS 12261), P388, DOI 10.1007/978-3-030-59710-8_38
   Yang Y, 2018, FRONT NEUROSCI-SWITZ, V12, DOI 10.3389/fnins.2018.00804
   Yuan L, 2021, 2021 IEEE/CVF INTERNATIONAL CONFERENCE ON COMPUTER VISION (ICCV 2021), P538, DOI 10.1109/ICCV48922.2021.00060
   Zhang L, 2023, EUR J RADIOL, V158, DOI 10.1016/j.ejrad.2022.110639
   Zhou Zongwei, 2018, Deep Learn Med Image Anal Multimodal Learn Clin Decis Support (2018), V11045, P3, DOI [10.1007/978-3-030-00889-5_1, 10.1007/978-3-030-00689-1_1]
   Zhu ZQ, 2023, INFORM FUSION, V91, P376, DOI 10.1016/j.inffus.2022.10.022
   Zhuge YZ, 2019, AAAI CONF ARTIF INTE, P9340
NR 49
TC 1
Z9 1
U1 10
U2 10
PU PERGAMON-ELSEVIER SCIENCE LTD
PI OXFORD
PA THE BOULEVARD, LANGFORD LANE, KIDLINGTON, OXFORD OX5 1GB, ENGLAND
SN 0895-6111
EI 1879-0771
J9 COMPUT MED IMAG GRAP
JI Comput. Med. Imaging Graph.
PD DEC
PY 2023
VL 110
AR 102307
DI 10.1016/j.compmedimag.2023.102307
EA OCT 2023
PG 13
WC Engineering, Biomedical; Radiology, Nuclear Medicine & Medical Imaging
WE Science Citation Index Expanded (SCI-EXPANDED)
SC Engineering; Radiology, Nuclear Medicine & Medical Imaging
GA Y4VC3
UT WOS:001105243500001
PM 37913635
DA 2024-06-03
ER

PT J
AU Wang, N
   Mo, ZX
   Pan, L
   Zhou, MH
   Ye, XL
   Liu, XJ
   Cai, X
   Qian, CG
   Chen, FL
   Xiong, Y
   Fan, FS
   Li, WY
AF Wang, Ning
   Mo, Zhenxian
   Pan, Lu
   Zhou, Minhua
   Ye, Xiaolan
   Liu, Xinjian
   Cai, Xiong
   Qian, Changgeng
   Chen, Feili
   Xiong, Yan
   Fan, Fushun
   Li, Wenyu
TI Dual PI3K/HDAC Inhibitor BEBT-908 Exhibits Potent Efficacy as
   Monotherapy for Primary Central Nervous System Lymphoma
SO TARGETED ONCOLOGY
LA English
DT Article
ID PRIMARY CNS LYMPHOMA
AB Background The efficacy of systemic treatment for primary central nervous system lymphoma (PCNSL) is limited because of the blood-brain barrier (BBB) and the ineffectiveness of chemotherapy. The dual PI3K/HDAC inhibitor BEBT-908 has exhibited favorable in vivo distribution and activity in various cancers.Objectives The aims of this study were to assess the efficacy of BEBT-908 in brain orthotopic mouse models of hematological malignancies, to investigate its pharmacologic properties, and to elucidate the underlying mechanism of action.Methods We evaluated the anticancer activity of BEBT-908 in various hematological malignancies through cell viability assays. The impact of BEBT-908 on c-Myc expression and ferroptosis signaling pathways was assessed using Western blotting, qPCR, ROS detection, GSH/GSSG detection, and IHC. Pharmacokinetic and pharmacodynamic profiles were assessed through LC-MS/MS and Western blotting. The effects of BEBT-908 in vivo were examined using xenografts and brain orthotopic mouse models.Results Our findings demonstrate that BEBT-908 exhibits promising anti-tumor activity in vitro and in vivo across multiple subtypes of hematological malignancies. Furthermore, BEBT-908 exhibits excellent BBB penetration and inhibits tumor growth in a brain orthotopic lymphoma model with prolonged survival of host mice. Mechanistically, BEBT-908 downregulated c-Myc expression, which contributed to ferroptosis, ultimately leading to tumor shrinkage.Conclusion Our study provides robust evidence for the dual PI3K/HDAC inhibitor BEBT-908 as an effective anti-cancer agent for PCNSL.
C1 [Wang, Ning; Pan, Lu; Chen, Feili; Li, Wenyu] Southern Med Univ, Guangdong Acad Med Sci, Guangdong Prov Peoples Hosp, 123 Huifu West Rd, Guangzhou 510080, Guangdong, Peoples R China.
   [Fan, Fushun] Jinan Univ, Coll Pharm, 601 Huangpu Ave West, Guangzhou, Peoples R China.
   [Mo, Zhenxian; Zhou, Minhua; Ye, Xiaolan; Liu, Xinjian; Cai, Xiong; Qian, Changgeng; Xiong, Yan; Fan, Fushun] Guangzhou BeBetter Med Inc, 25 Yayingshi Rd, Guangzhou 510000, Guangdong, Peoples R China.
   [Mo, Zhenxian] Jinan Univ, Coll Life Sci & Technol, 601 Huangpu Ave West, Guangzhou, Peoples R China.
   [Wang, Ning; Pan, Lu; Li, Wenyu] South China Univ Technol, Sch Med, Guangzhou, Peoples R China.
   [Cai, Xiong; Qian, Changgeng] Curis Inc, Lexington, MA USA.
C3 Southern Medical University - China; Guangdong Academy of Medical
   Sciences & Guangdong General Hospital; Jinan University; Jinan
   University; South China University of Technology; Curis, Inc.
RP Li, WY (corresponding author), Southern Med Univ, Guangdong Acad Med Sci, Guangdong Prov Peoples Hosp, 123 Huifu West Rd, Guangzhou 510080, Guangdong, Peoples R China.; Fan, FS (corresponding author), Jinan Univ, Coll Pharm, 601 Huangpu Ave West, Guangzhou, Peoples R China.; Fan, FS (corresponding author), Guangzhou BeBetter Med Inc, 25 Yayingshi Rd, Guangzhou 510000, Guangdong, Peoples R China.; Li, WY (corresponding author), South China Univ Technol, Sch Med, Guangzhou, Peoples R China.
EM
RI Zhu, Mengzhao/KFT-2345-2024; han, yang/KHX-8947-2024; li,
   guo/KHU-1749-2024; Wang, YuHan/KGY-2933-2024; WANG,
   SHIHAO/KHC-8263-2024; li, feiyang/KHW-5210-2024; Liu,
   Donghua/KEJ-1974-2024; liu, qi/KFA-4047-2024; Lin, Wei/KFQ-5381-2024;
   wang, yifang/KEI-3766-2024; Wen, Jing/KCL-6614-2024; Zhang,
   jin/KFT-0762-2024; liu, miao/KGL-7043-2024; liu, yang/KFA-8402-2024;
   zhang, xiaoyu/KEJ-0657-2024; Chen, YiJun/KFS-9282-2024; Liu,
   Yu/KFS-0769-2024; Liu, Zhen/KFS-2748-2024; li, qing/KHU-6871-2024; wang,
   jiaqi/KHC-5900-2024; Shen, Yan/KEJ-4617-2024; Liu, Yining/KHC-6217-2024;
   Wang, Jiawei/KHC-8971-2024; liu, qi/KHC-7509-2024; LI,
   yue/KHC-6771-2024; Li, Yuanyuan/KEH-6935-2024; yan, xu/KCY-8174-2024;
   Chen, Yang/KHD-8849-2024; liu, zhen/KFS-0275-2024; Zhang,
   Yulin/KEI-1610-2024; Sun, Yue/KHU-8159-2024; ZHENG, YI/KAM-6536-2024;
   zhang, jinlu/KEE-9374-2024; Liu, Xinru/KEH-2341-2024; WEI,
   ZHEN/KHU-7176-2024; Wang, Xinyi/KHV-4909-2024; yu, xiao/KFT-1725-2024;
   Wang, Zichao/KHX-4954-2024; Yang, Ning/KHD-1133-2024
OI Liu, Donghua/0000-0002-5830-9540; Liu, Yining/0000-0002-2218-2349; Li,
   Yuanyuan/0000-0002-4955-1159; 
FU National Science and Technology Major Project of China [2016ZX09101002];
   Guangdong Pearl River Talents Plan [2014ZT05Y232]; Guangzhou Municipal
   Science and Technology Project [201909020004]
FX This study is funded by National Science and Technology Major Project of
   China (2016ZX09101002), Guangdong Pearl River Talents Plan
   (2014ZT05Y232), Guangzhou Municipal Science and Technology Project
   (201909020004).
CR Alborzinia H, 2022, NAT CANCER, V3, P471, DOI 10.1038/s43018-022-00355-4
   Anderson GR, 2016, SCI TRANSL MED, V8, DOI 10.1126/scitranslmed.aae0348
   Chen T, 2022, J HEMATOL ONCOL, V15, DOI 10.1186/s13045-022-01356-7
   Deckert M, 2011, LEUKEMIA, V25, P1797, DOI 10.1038/leu.2011.169
   Dong Y, 2020, SIGNAL TRANSDUCT TAR, V5, DOI 10.1038/s41392-020-00235-2
   Eyre TA, 2019, BRIT J HAEMATOL, V187, P185, DOI 10.1111/bjh.16070
   Fan FS, 2021, CANCER RES, V81, P6233, DOI 10.1158/0008-5472.CAN-21-1547
   Ferreri AJM, 2016, LANCET HAEMATOL, V3, pE217, DOI 10.1016/S2352-3026(16)00036-3
   Fujimoto K, 2020, NEURO-ONCOL ADV, V2, DOI 10.1093/noajnl/vdaa084
   Ghesquieres H, 2019, ANN ONCOL, V30, P621, DOI 10.1093/annonc/mdz032
   Gleeson M, 2017, ANN ONCOL, V28, P2511, DOI 10.1093/annonc/mdx353
   Grommes C, 2019, BLOOD, V134, DOI 10.1182/blood-2019-126214
   Grommes C, 2017, J CLIN ONCOL, V35, P2410, DOI 10.1200/JCO.2017.72.7602
   Houillier C, 2020, NEUROLOGY, V94, pE1027, DOI 10.1212/WNL.0000000000008900
   Jain N, 2020, BLOOD ADV, V4, P4382, DOI 10.1182/bloodadvances.2020001685
   Kapadia B, 2018, NAT COMMUN, V9, DOI 10.1038/s41467-018-03028-y
   Korfel A, 2016, J CLIN ONCOL, V34, P1757, DOI 10.1200/JCO.2015.64.9897
   Kurland JF, 2008, CANCER RES, V68, P3624, DOI 10.1158/0008-5472.CAN-07-6552
   Langner-Lemercier S, 2016, NEURO-ONCOLOGY, V18, P1297, DOI 10.1093/neuonc/now033
   Li L, 2018, SCI REP-UK, V8, DOI 10.1038/s41598-018-24631-5
   Martz CA, 2014, SCI SIGNAL, V7, DOI 10.1126/scisignal.aaa1877
   Mendez Joe S, 2018, Am Soc Clin Oncol Educ Book, V38, P604, DOI 10.1200/EDBK_200829
   Rahmani M, 2014, CLIN CANCER RES, V20, P4849, DOI 10.1158/1078-0432.CCR-14-0034
   Roberts AW, 2016, NEW ENGL J MED, V374, P311, DOI 10.1056/NEJMoa1513257
   Schabet M, 1999, J NEURO-ONCOL, V43, P199, DOI 10.1023/A:1006290032052
   Schultz C, 1996, J CLIN ONCOL, V14, P556, DOI 10.1200/JCO.1996.14.2.556
   Shao L, 2021, FRONT ONCOL, V11, DOI 10.3389/fonc.2021.689843
   Tarantelli C, 2019, CANCERS, V11, DOI 10.3390/cancers11060775
   von Baumgarten L, 2018, DTSCH ARZTEBL INT, V115, P419, DOI 10.3238/arztebl.2018.0419
   Wilson MR, 2022, BLOOD, V139, P2499, DOI 10.1182/blood.2021014506
   Wu WJ, 2013, CLIN CANCER RES, V19, P5699, DOI 10.1158/1078-0432.CCR-13-1758
   Yu HF, 2021, TRANSL CANCER RES, V10, P1975, DOI 10.21037/tcr-21-50
   Zhang XW, 2022, BMC CANCER, V22, DOI 10.1186/s12885-022-09275-z
   Zhang XW, 2020, CNS NEUROL DISORD-DR, V19, P165, DOI 10.2174/1871527319666200517112252
NR 34
TC 1
Z9 1
U1 2
U2 3
PU SPRINGER
PI DORDRECHT
PA VAN GODEWIJCKSTRAAT 30, 3311 GZ DORDRECHT, NETHERLANDS
SN 1776-2596
EI 1776-260X
J9 TARGET ONCOL
JI Target. Oncol.
PD NOV
PY 2023
VL 18
IS 6
BP 941
EP 952
DI 10.1007/s11523-023-01006-z
EA OCT 2023
PG 12
WC Oncology
WE Science Citation Index Expanded (SCI-EXPANDED)
SC Oncology
GA CN2Q0
UT WOS:001085945300003
PM 37855991
DA 2024-06-03
ER

PT J
AU Calimeri, T
   Steidl, C
   Fiore, P
   Ferreri, AJM
AF Calimeri, Teresa
   Steidl, Carolina
   Fiore, Paolo
   Ferreri, Andres J. M.
TI New hopes in relapsed refractory primary central nervous system lymphoma
SO CURRENT OPINION IN ONCOLOGY
LA English
DT Review
DE blood brain barrier; BTK-I; CAR-T; central nervous system lymphoma;
   relapse; refractory
ID PRIMARY CNS LYMPHOMA; LARGE-B-CELL; RECURRENT PRIMARY CNS;
   RELAPSED/REFRACTORY PRIMARY; SALVAGE THERAPY; PHASE-II; INTRAOCULAR
   LYMPHOMA; R-CHOP; IBRUTINIB; RITUXIMAB
AB Purpose of reviewPatients with relapsed/refractory primary central nervous system lymphoma (rrPCNSL) have poor prognosis, with a median survival after relapse of 6.8 months. In this review, we discuss the evolving landscape and the possible future directions related to this important unmet clinical need.Recent findingsThe modern two-phase approach for newly diagnosed PCNSL based on an induction using high-dose methotrexate (HD-MTX) combinations and a subsequent consolidation, has significantly improved the outcome in this setting. However, this strategy is able to cure more or less 50% of patients. rrPCNSL patients have a very poor prognosis with a reported 5-year overall survival of 18%. Late relapses (after third year) and use of high-dose chemotherapy and autologous stem cell transplantation (HDT-ASCT) represent important factors associated with a better outcome in this setting. On the basis of the growing acquisition of knowledge on the molecular characteristics of PCNSL, the use of non-chemotherapeutic drugs such as bruton tyrosine kinase inhibitors (BTK-is), immunomodulatory drugs (IMiDs) and immune checkpoint blockers (ICBs) is increasing in the last years along with the introduction of novel approaches (CAR-T cells and blood--brain barrier disruption). However, despite high responses in some cases, durations are often short, translating in outcome results still unsatisfactory.Treatment of rrPCNSL patients is challenging. As no standard of care exist in this setting, it is of paramount importance to acquire new knowledge related to this condition and start multidisciplinary collaboration in order to improve pts outcome.
C1 [Calimeri, Teresa; Steidl, Carolina; Fiore, Paolo; Ferreri, Andres J. M.] IRCCS San Raffaele Sci Inst, Lymphoma Unit, Milan, Italy.
   [Fiore, Paolo] Univ Vita Salute San Raffaele, Milan, Italy.
   [Calimeri, Teresa] IRCCS San Raffaele Sci Inst, Lymphoma Unit, Via Olgettina 60, I-20132 Milan, Italy.
C3 Vita-Salute San Raffaele University; IRCCS Ospedale San Raffaele;
   Vita-Salute San Raffaele University; Vita-Salute San Raffaele
   University; IRCCS Ospedale San Raffaele
RP Calimeri, T (corresponding author), IRCCS San Raffaele Sci Inst, Lymphoma Unit, Via Olgettina 60, I-20132 Milan, Italy.
EM
CR Abramson JS, 2017, NEW ENGL J MED, V377, P783, DOI 10.1056/NEJMc1704610
   Aizawa M, 2011, NEUROSCI RES, V69, P343, DOI 10.1016/j.neures.2010.12.019
   Alcantara M, 2022, BLOOD, V139, P792, DOI 10.1182/blood.2021012932
   Ambady P, 2019, LEUKEMIA LYMPHOMA, V60, P515, DOI 10.1080/10428194.2018.1480771
   Arvanitis CD, 2020, NAT REV CANCER, V20, P26, DOI 10.1038/s41568-019-0205-x
   Batchelor TT, 2011, NEUROLOGY, V76, P929, DOI 10.1212/WNL.0b013e31820f2d94
   Bishop MR, 2019, BLOOD ADV, V3, P2230, DOI 10.1182/bloodadvances.2019000151
   Bödör C, 2020, J NEUROPATH EXP NEUR, V79, P176, DOI 10.1093/jnen/nlz125
   Calimeri T, 2022, BLOOD, V140, P6678, DOI 10.1182/blood-2022-170143
   Calimeriy T, 2021, ESMO OPEN, V6, DOI 10.1016/j.esmoop.2021.100213
   Camilleri-Broët S, 2006, BLOOD, V107, P190, DOI 10.1182/blood-2005-03-1024
   Chamberlain MC, 2016, J NEURO-ONCOL, V126, P545, DOI 10.1007/s11060-015-1994-8
   Chapuy B, 2016, BLOOD, V127, P869, DOI 10.1182/blood-2015-10-673236
   Choquet S., 2015, BLOOD, V126, P1524, DOI DOI 10.1182/BLOOD.V126.23.1524.1524
   Cook MR, 2023, BLOOD ADV, V7, P32, DOI 10.1182/bloodadvances.2022008525
   Davis RE, 2010, NATURE, V463, P88, DOI 10.1038/nature08638
   de Groen RAL, 2019, HAEMATOLOGICA, V104, P2337, DOI 10.3324/haematol.2019.227272
   Destito M, 2023, BIOENGINEERING-BASEL, V10, DOI 10.3390/bioengineering10030285
   Dufour J, 2023, ANN HEMATOL, V102, P1159, DOI 10.1007/s00277-023-05108-6
   Enting RH, 2004, NEUROLOGY, V63, P901, DOI 10.1212/01.WNL.0000137050.43114.42
   Ferreri AJM, 2022, LEUKEMIA, V36, P1870, DOI 10.1038/s41375-022-01582-5
   Ferreri AJM, 2020, BLOOD ADV, V4, P3648, DOI 10.1182/bloodadvances.2020002270
   Ferreri AJM, 2019, BLOOD, V134, P252, DOI 10.1182/blood.2019000633
   Ferreri AJM, 2016, LANCET HAEMATOL, V3, pE217, DOI 10.1016/S2352-3026(16)00036-3
   Fischer L, 2006, ANN ONCOL, V17, P1141, DOI 10.1093/annonc/mdl070
   Fox CP, 2021, BLOOD ADV, V5, P4073, DOI 10.1182/bloodadvances.2021004779
   Frigault MJ, 2022, BLOOD, V139, P2306, DOI 10.1182/blood.2021014738
   Ghesquieres H, 2019, ANN ONCOL, V30, P621, DOI 10.1093/annonc/mdz032
   Ghez D, 2018, BLOOD, V131, P1955, DOI 10.1182/blood-2017-11-818286
   Gill S, 2022, BLOOD ADV, V6, P5774, DOI 10.1182/bloodadvances.2022007317
   Grommes C, 2019, BLOOD, V133, P436, DOI 10.1182/blood-2018-09-875732
   Grommes C, 2017, CANCER DISCOV, V7, P1018, DOI 10.1158/2159-8290.CD-17-0613
   Grommes C, 2017, CANCER CELL, V31, P731, DOI 10.1016/j.ccell.2017.05.004
   Heming M, 2022, GENOME MED, V14, DOI 10.1186/s13073-022-01110-1
   Hernández-Verdin I, 2023, ANN ONCOL, V34, P186, DOI 10.1016/j.annonc.2022.11.002
   Hoang-Xuan K, 2020, BLOOD, V136, DOI 10.1182/blood-2020-141773
   Houillier C, 2022, J CLIN ONCOL, V40, P3692, DOI 10.1200/JCO.22.00491
   Houillier C, 2021, NEUROLOGY, V97, P628, DOI 10.1212/WNL.0000000000012515
   Houillier C, 2020, NEUROLOGY, V94, pE1027, DOI 10.1212/WNL.0000000000008900
   Houillier C, 2019, J CLIN ONCOL, V37, P823, DOI 10.1200/JCO.18.00306
   Houillier C, 2015, NEUROLOGY, V84, P325, DOI 10.1212/WNL.0000000000001158
   Ioannou N, 2021, INT J MOL SCI, V22, DOI 10.3390/ijms22168572
   Jacobson CA, 2022, BLOOD, V140, P1060, DOI 10.1182/blood-2022-165256
   Jahnke K, 2006, J NEURO-ONCOL, V80, P159, DOI 10.1007/s11060-006-9165-6
   Kasenda B, 2017, LEUKEMIA, V31, P2623, DOI 10.1038/leu.2017.170
   Vu K, 2019, BRIT J HAEMATOL, V186, P180, DOI 10.1111/bjh.15787
   Korfel A, 2016, J CLIN ONCOL, V34, P1757, DOI 10.1200/JCO.2015.64.9897
   Langner-Lemercier S, 2016, NEURO-ONCOLOGY, V18, P1297, DOI 10.1093/neuonc/now033
   Lionakis MS, 2017, CANCER CELL, V31, P833, DOI 10.1016/j.ccell.2017.04.012
   Locke FL, 2019, LANCET ONCOL, V20, P31, DOI 10.1016/S1470-2045(18)30864-7
   Los-de Vries GT, 2023, CANCER RES, V83, P1917, DOI 10.1158/0008-5472.CAN-22-3814
   Ma JJ, 2022, FRONT ONCOL, V12, DOI 10.3389/fonc.2022.938421
   Mappa S, 2013, HEMATOL ONCOL, V31, P143, DOI 10.1002/hon.2037
   Marcelis L, 2020, CANCER IMMUNOL IMMUN, V69, P1751, DOI 10.1007/s00262-020-02575-y
   Meeks C, 2019, NEURO-ONCOLOGY, V21, P86
   Mendez JS, 2018, NEURO-ONCOLOGY, V20, P687, DOI 10.1093/neuonc/nox187
   Minderman M, 2021, BLOOD, V138, P1194, DOI 10.1182/blood.2021011366
   Mutter JA, 2023, J CLIN ONCOL, V41, P1684, DOI 10.1200/JCO.22.00826
   Nagel D, 2014, ONCOGENE, V33, P5655, DOI 10.1038/onc.2013.565
   Narita Y, 2021, NEURO-ONCOLOGY, V23, P122, DOI 10.1093/neuonc/noaa145
   Nayak L, 2017, BLOOD, V129, P3071, DOI 10.1182/blood-2017-01-764209
   Nayak L, 2013, LEUKEMIA LYMPHOMA, V54, P58, DOI 10.3109/10428194.2012.698736
   Nayak L, 2011, NEURO-ONCOLOGY, V13, P525, DOI 10.1093/neuonc/nor014
   Neelapu SS, 2017, NEW ENGL J MED, V377, P2531, DOI 10.1056/NEJMoa1707447
   Nguyen PL, 2005, J CLIN ONCOL, V23, P1507, DOI 10.1200/JCO.2005.01.161
   Ostrom QT, 2015, NEURO-ONCOLOGY, V16, P1, DOI [10.1093/neuonc/nou327, 10.1093/neuonc/nox158, 10.1093/neuonc/nou223, 10.1093/neuonc/nov189]
   Patel UH, 2015, HEMATOL REP, V7, P12, DOI 10.4081/hr.2015.5704
   Pentsova E, 2014, J NEURO-ONCOL, V117, P161, DOI 10.1007/s11060-014-1370-0
   Plotkin SR, 2004, CLIN CANCER RES, V10, P5643, DOI 10.1158/1078-0432.CCR-04-0159
   Ponzoni M, 2007, BRIT J HAEMATOL, V138, P316, DOI 10.1111/j.1365-2141.2007.06661.x
   Radke J, 2022, NAT COMMUN, V13, DOI 10.1038/s41467-022-30050-y
   Raizer JJ, 2012, CANCER-AM CANCER SOC, V118, P3743, DOI 10.1002/cncr.26709
   Rauschecker AM, 2023, BRIT J HAEMATOL, V201, P154, DOI 10.1111/bjh.18660
   Reinwald M, 2018, CLIN MICROBIOL INFEC, V24, pS53, DOI 10.1016/j.cmi.2018.02.009
   Reni M, 2004, EUR J CANCER, V40, P1682, DOI 10.1016/j.ejca.2004.03.008
   Roddie C., 2022, HEMASPHERE, V6, P1
   Roschewski M, 2020, BLOOD, V136, DOI 10.1182/blood-2020-137637
   Rubenstein JL, 2018, BLOOD ADV, V2, P1595, DOI 10.1182/bloodadvances.2017014845
   Rubenstein JL, 2011, J CLIN ONCOL, V29, pE595, DOI 10.1200/JCO.2011.34.7252
   Rubenstein JL, 2006, BLOOD, V107, P3716, DOI 10.1182/blood-2005-03-0897
   Schenone L, 2022, BONE MARROW TRANSPL, V57, P966, DOI 10.1038/s41409-022-01648-z
   Schorb E, 2022, BLOOD, V140, P1773, DOI 10.1182/blood-2022-162157
   Schuster SJ, 2021, LANCET ONCOL, V22, P1403, DOI 10.1016/S1470-2045(21)00375-2
   Schuster SJ, 2019, NEW ENGL J MED, V380, P45, DOI 10.1056/NEJMoa1804980
   Severson EA, 2023, ONCOLOGIST, V28, pE26, DOI 10.1093/oncolo/oyac190
   Siddiqi T, 2021, BLOOD ADV, V5, P4059, DOI 10.1182/bloodadvances.2020004106
   Soussain C, 2019, EUR J CANCER, V117, P121, DOI 10.1016/j.ejca.2019.05.024
   Soussain C, 2008, J CLIN ONCOL, V26, P2512, DOI 10.1200/JCO.2007.13.5533
   Tun HW, 2018, BLOOD, V132, P2240, DOI 10.1182/blood-2018-02-835496
   Voloschin AD, 2008, J NEURO-ONCOL, V86, P211, DOI 10.1007/s11060-007-9464-6
   Von Roemeling CA, 2023, CLIN CANCER RES, V29, P1751, DOI 10.1158/1078-0432.CCR-22-1682
   Wang XL, 2021, CANCER IMMUNOL RES, V9, P75, DOI 10.1158/2326-6066.CIR-20-0236
   Yang CW, 2022, FRONT ONCOL, V12, DOI 10.3389/fonc.2022.901797
   Zhang Y., 2022, HEMASPHERE, V6, P125, DOI [DOI 10.1097/01.HS9.0000843788.10370.07, 10.1097/01.HS9.0000843788.10370.07]
   Zhang Y, 2021, FRONT ONCOL, V11, DOI 10.3389/fonc.2021.760405
   Zhu Q, 2022, FRONT GENET, V13, DOI 10.3389/fgene.2022.878618
NR 96
TC 1
Z9 1
U1 1
U2 3
PU LIPPINCOTT WILLIAMS & WILKINS
PI PHILADELPHIA
PA TWO COMMERCE SQ, 2001 MARKET ST, PHILADELPHIA, PA 19103 USA
SN 1040-8746
EI 1531-703X
J9 CURR OPIN ONCOL
JI Curr. Opin. Oncol.
PD SEP
PY 2023
VL 35
IS 5
BP 364
EP 372
DI 10.1097/CCO.0000000000000980
PG 9
WC Oncology
WE Science Citation Index Expanded (SCI-EXPANDED)
SC Oncology
GA O5UR9
UT WOS:001044463500003
PM 37551946
DA 2024-06-03
ER

PT J
AU Isbell, LK
   Uibeleisen, R
   Friedl, A
   Burger, E
   Dopatka, T
   Scherer, F
   Orban, A
   Lauer, E
   Malenica, N
   Semenova, I
   Vreden, A
   Valk, E
   Wendler, J
   Neumaier, S
   Fricker, H
   El Rabih, AA
   Gloggengiesser, C
   Hilbig, D
   Bleul, S
   Weis, J
   Gmehlin, D
   Backenstrass, M
   Wirtz, S
   Ihorst, G
   Finke, J
   Illerhaus, G
   Schorb, E
AF Isbell, Lisa K.
   Uibeleisen, Roswitha
   Friedl, Alexander
   Burger, Elvira
   Dopatka, Tatja
   Scherer, Florian
   Orban, Andras
   Lauer, Eliza
   Malenica, Natalie
   Semenova, Inna
   Vreden, Annika
   Valk, Elke
   Wendler, Julia
   Neumaier, Simone
   Fricker, Heidi
   El Rabih, Abed Al Hadi
   Gloggengiesser, Cora
   Hilbig, Daniela
   Bleul, Sabine
   Weis, Joachim
   Gmehlin, Dennis
   Backenstrass, Matthias
   Wirtz, Sebastian
   Ihorst, Gabriele
   Finke, Juergen
   Illerhaus, Gerald
   Schorb, Elisabeth
TI Age-adjusted high-dose chemotherapy followed by autologous stem cell
   transplantation or conventional chemotherapy with R-MP as first-line
   treatment in elderly primary CNS lymphoma patients - the randomized
   phase III PRIMA-CNS trial
SO BMC CANCER
LA English
DT Article
DE Primary central nervous system lymphoma (PCNSL); High-dose chemotherapy
   (HCT); Autologous stem cell transplantation (ASCT); Elderly patients;
   Transplant eligibility
ID NERVOUS-SYSTEM LYMPHOMA; INTERNATIONAL EXTRANODAL LYMPHOMA;
   QUALITY-OF-LIFE; GERIATRIC ASSESSMENT; OLDER; METHOTREXATE;
   CHEMOIMMUNOTHERAPY; MULTICENTER; TOOL; INSTRUMENT
AB BackgroundOlder primary central nervous system lymphoma (PCNSL) patients have an inferior prognosis compared to younger patients because available evidence on best treatment is scarce and treatment delivery is challenging due to comorbidities and reduced performance status. High-dose chemotherapy and autologous stem cell transplantation (HCT-ASCT) after high-dose methotrexate (MTX)-based immuno-chemotherapy has become an increasingly used treatment approach in eligible elderly PCNSL patients with promising feasibility and efficacy, but has not been compared with conventional chemotherapy approaches. In addition, eligibility for HCT-ASCT in elderly PCNSL is not well defined. Geriatric assessment (GA) may be helpful in selecting patients for the best individual treatment choice, but no standardized GA exists to date. A randomized controlled trial, incorporating a GA and comparing age-adapted HCT-ASCT treatment with conventional chemotherapy is needed.MethodsThis open-label, multicenter, randomized phase III trial with two parallel arms will recruit 310 patients with newly diagnosed PCNSL > 65 years of age in 40 centers in Germany and Austria. The primary objective is to demonstrate that intensified chemotherapy followed by consolidating HCT-ASCT is superior to conventional chemotherapy with rituximab, MTX, procarbazine (R-MP) followed by maintenance with procarbazine in terms of progression free survival (PFS). Secondary endpoints include overall survival (OS), event free survival (EFS), (neuro-)toxicity and quality of life (QoL). GA will be conducted at specific time points during the course of the study. All patients will be treated with a pre-phase rituximab-MTX (R-MTX) cycle followed by re-assessment of transplant eligibility. Patients judged transplant eligible will be randomized (1:1). Patients in arm A will be treated with 3 cycles of R-MP followed by maintenance therapy with procarbazine for 6 months. Patients in arm B will be treated with 2 cycles of MARTA (R-MTX/AraC) followed by busulfan- and thiotepa-based HCT-ASCT.DiscussionThe best treatment strategy for elderly PCNSL patients remains unknown. Treatments range from palliative to curative but more toxic therapies, and there is no standardized measure to select patients for the right treatment. This randomized controlled trial will create evidence for the best treatment strategy with the focus on developing a standardized GA to help define eligibility for an intensive treatment approach.
C1 [Isbell, Lisa K.; Burger, Elvira; Dopatka, Tatja; Scherer, Florian; Orban, Andras; Lauer, Eliza; Malenica, Natalie; Semenova, Inna; Vreden, Annika; Fricker, Heidi; El Rabih, Abed Al Hadi; Bleul, Sabine; Finke, Juergen; Schorb, Elisabeth] Univ Freiburg, Dept Med 1, Fac Med, Med Ctr, Hugstetter Str 55, D-79106 Freiburg, Germany.
   [Uibeleisen, Roswitha; Wendler, Julia; Neumaier, Simone; Illerhaus, Gerald] Klinikum Stuttgart, Clin Hematol Oncol & Palliat Care, Kriegsbergstr 60, D-70174 Stuttgart, Germany.
   [Friedl, Alexander] Klinikum Stuttgart, Dept Endocrinol Diabetol & Geriatr, Priessnitzweg 24, D-70374 Bad Cannstatt, Germany.
   [Valk, Elke; Gloggengiesser, Cora; Hilbig, Daniela] Klinikum Stuttgart, Stuttgart Canc Ctr Tumorzentrum Eva Mayer Stihl, Kriegsbergstr 60, D-70174 Stuttgart, Germany.
   [Weis, Joachim] Univ Freiburg, Fac Med, Comprehens Canc Ctr, Endowed Professorship Self Help Res, Freiburg, Germany.
   [Weis, Joachim; Wirtz, Sebastian; Ihorst, Gabriele] Univ Freiburg, Med Ctr, Freiburg, Germany.
   [Gmehlin, Dennis; Backenstrass, Matthias] Klinikum Stuttgart, Inst Clin Psychol, Priessnitzweg 24, D-70374 Bad Cannstatt, Germany.
   [Wirtz, Sebastian; Ihorst, Gabriele] Univ Freiburg, Fac Med, Clin Trials Unit, Elsasser Str 2, D-79110 Freiburg, Germany.
C3 University of Freiburg; Klinikum Stuttgart; Klinikum Stuttgart;
   University of Freiburg; University of Freiburg; University of Freiburg
RP Schorb, E (corresponding author), Univ Freiburg, Dept Med 1, Fac Med, Med Ctr, Hugstetter Str 55, D-79106 Freiburg, Germany.
EM
RI Scherer, Florian/JTT-6048-2023; Ihorst, Gabriele/AAA-7445-2021; Wendler,
   Julia S/IYT-3112-2023
OI Neumaier, Simone/0009-0005-0215-8105
CR AARONSON NK, 1993, J NATL CANCER I, V85, P365, DOI 10.1093/jnci/85.5.365
   Abrey LE, 2005, J CLIN ONCOL, V23, P5034, DOI 10.1200/JCO.2005.13.524
   Bellera CA, 2012, ANN ONCOL, V23, P2166, DOI 10.1093/annonc/mdr587
   Berro M, 2017, BIOL BLOOD MARROW TR, V23, P1646, DOI 10.1016/j.bbmt.2017.06.014
   Brem EA, 2022, J GERIATR ONCOL, V13, P258, DOI 10.1016/j.jgo.2021.10.003
   CHARLSON ME, 1987, J CHRON DIS, V40, P373, DOI 10.1016/0021-9681(87)90171-8
   Correa DD, 2007, ANN ONCOL, V18, P1145, DOI 10.1093/annonc/mdl464
   Farhi J, 2018, ANN HEMATOL, V97, P2391, DOI 10.1007/s00277-018-3468-5
   Ferreri AJM, 2017, LANCET HAEMATOL, V4, pE510, DOI 10.1016/S2352-3026(17)30174-6
   Ferreri AJM, 2016, LANCET HAEMATOL, V3, pE217, DOI 10.1016/S2352-3026(16)00036-3
   Fried LP, 2001, J GERONTOL A-BIOL, V56, pM146, DOI 10.1093/gerona/56.3.M146
   Fritsch K, 2017, LEUKEMIA, V31, P846, DOI 10.1038/leu.2016.334
   Houillier C, 2020, NEUROLOGY, V94, pE1027, DOI 10.1212/WNL.0000000000008900
   Hurria A, 2011, J CLIN ONCOL, V29, P1290, DOI 10.1200/JCO.2010.30.6985
   Illerhaus G, 2022, BLOOD, V140, P10, DOI 10.1182/blood-2022-171733
   Kassam S, 2017, BONE MARROW TRANSPL, V52, P1268, DOI 10.1038/bmt.2017.101
   LACHS MS, 1990, ANN INTERN MED, V112, P699, DOI 10.7326/0003-4819-112-9-699
   Lakshmaiah KC, 2018, EUR J HAEMATOL, V100, P644, DOI 10.1111/ejh.13068
   LAWTON MP, 1969, GERONTOLOGIST, V9, P179, DOI 10.1093/geront/9.3_Part_1.179
   LINN BS, 1968, J AM GERIATR SOC, V16, P622, DOI 10.1111/j.1532-5415.1968.tb02103.x
   Löwe B, 2004, J AFFECT DISORDERS, V81, P61, DOI 10.1016/S0165-0327(03)00198-8
   MAHONEY F I, 1965, Md State Med J, V14, P61
   Martinez-Calle N, 2022, BRIT J HAEMATOL, V196, P473, DOI 10.1111/bjh.17799
   Mendez JS, 2018, NEURO-ONCOLOGY, V20, P687, DOI 10.1093/neuonc/nox187
   Merli F, 2021, J CLIN ONCOL, V39, P1214, DOI 10.1200/JCO.20.02465
   Mutter JA, 2023, J CLIN ONCOL, V41, P1684, DOI 10.1200/JCO.22.00826
   Nasreddine ZS, 2005, J AM GERIATR SOC, V53, P695, DOI 10.1111/j.1532-5415.2005.53221.x
   Nayak L, 2017, NEURO-ONCOLOGY, V19, P625, DOI 10.1093/neuonc/nox029
   Olivier G, 2014, AM J HEMATOL, V89, P1024, DOI 10.1002/ajh.23812
   Omuro A, 2015, LANCET HAEMATOL, V2, pE251, DOI 10.1016/S2352-3026(15)00074-5
   Peyrade F, 2017, LANCET HAEMATOL, V4, pE46, DOI 10.1016/S2352-3026(16)30171-5
   Pfreundschuh M, 2004, BLOOD, V104, P634, DOI 10.1182/blood-2003-06-2095
   Philipp R, 2016, SUPPORT CARE CANCER, V24, P2751, DOI 10.1007/s00520-016-3090-9
   Roth P, 2014, CURR OPIN NEUROL, V27, P697, DOI 10.1097/WCO.0000000000000145
   Roth P, 2012, NEUROLOGY, V79, P890, DOI 10.1212/WNL.0b013e318266fcb2
   Schorb E, 2017, BONE MARROW TRANSPL, V52, P1113, DOI 10.1038/bmt.2017.23
   Schorb E, 2022, HIGH DOSE CHEMOTHERA
   Schorb E., 2021, CANCERS, V13, P17, DOI [10.3390/cancers13174268, DOI 10.3390/CANCERS13174268]
   Schorb E, 2020, BLOOD ADV, V4, P3378, DOI 10.1182/bloodadvances.2020002064
   Schorb E, 2020, BRIT J HAEMATOL, V189, P879, DOI 10.1111/bjh.16451
   Schorb E, 2019, BMC CANCER, V19, DOI 10.1186/s12885-019-5473-z
   Siegal T, 2019, ACTA HAEMATOL-BASEL, V141, P138, DOI 10.1159/000495284
   Spitzer RL, 2006, ARCH INTERN MED, V166, P1092, DOI 10.1001/archinte.166.10.1092
   Strauss E., 2006, COMPENDIUM NEUROPSYC
   Taphoorn MJB, 2010, EUR J CANCER, V46, P1033, DOI 10.1016/j.ejca.2010.01.012
   Tucci A, 2015, LEUKEMIA LYMPHOMA, V56, P921, DOI 10.3109/10428194.2014.953142
   Vreden A, 2021, ONCOL RES TREAT, V44
   Wendler J, 2022, BMC CANCER, V22, DOI 10.1186/s12885-022-09723-w
NR 48
TC 1
Z9 1
U1 0
U2 1
PU BMC
PI LONDON
PA CAMPUS, 4 CRINAN ST, LONDON N1 9XW, ENGLAND
EI 1471-2407
J9 BMC CANCER
JI BMC Cancer
PD AUG 18
PY 2023
VL 23
IS 1
AR 767
DI 10.1186/s12885-023-11193-7
PG 10
WC Oncology
WE Science Citation Index Expanded (SCI-EXPANDED)
SC Oncology
GA Q0EE6
UT WOS:001054317700002
PM 37596517
OA Green Submitted, gold, Green Published
DA 2024-06-03
ER

PT J
AU Therkelsen, KE
   Schaff, LR
   Nandakumar, S
   Omuro, AMP
   DeAngelis, LM
   Grommes, C
AF Therkelsen, Kate Elizabeth
   Schaff, Lauren R.
   Nandakumar, Subhiksha
   Omuro, Antonio M. P.
   DeAngelis, Lisa M.
   Grommes, Christian
TI Long-term Outcomes in Primary CNS Lymphoma After R-MVP and High-Dose
   Chemotherapy With Autologous Hematopoietic Stem Cell Transplant
SO NEUROLOGY
LA English
DT Article
AB Background and ObjectivesPrimary CNS lymphoma (PCNSL), a rare CNS malignancy, is usually treated with high-dose methotrexate in the first-line setting, typically followed by consolidation therapy. Due to the broad range of currently available treatments for PCNSL, comparability in long-term follow-up studies is limited, and data are scattered across small studies.MethodsIn this study, we report the long-term survival of patients with newly diagnosed immunocompetent PCNSL, enrolled in a phase II trial from June 2005 to September 2011. Patients were treated using rituximab, methotrexate, vincristine, and procarbazine (R-MVP) chemotherapy followed by high-dose chemotherapy (HDC) and autologous stem cell transplant (ASCT) in those with partial or complete response to R-MVP. In a post hoc analysis, clinical and imaging features were evaluated in those still alive.Results26 of 32 patients underwent HDC-ASCT consolidation. Of them, 3 patients died of treatment-related toxicity and 2 due to disease progression within 1 year of ASCT. None of the remaining 21 patients had disease progression with a median follow-up of 12.1 years and were included in the analysis. Compared with the post-HDC-ASCT assessment, at the last follow-up, there was no significant difference in the median Karnofsky Performance Status (80 [range: 60-100] vs 90 [range: 70-100]), the median Neurologic Assessment in Neuro-Oncology score (1 [range: 0-4] vs 1 [range: 0-5]), and leukoencephalopathy score (1 [range: 0-3] vs 1 [range: 1-4]).DiscussionLong-term follow-up demonstrated that treatment was well tolerated in most patients enrolled in this study, with stable leukoencephalopathy on imaging and stable clinical performance status. Disease recurrence was not observed beyond 2 years after HDC-ASCT consolidation.
C1 [Therkelsen, Kate Elizabeth] Stanford Univ, Dept Neurol & Neurol Sci, Sch Med, Stanford, CA USA.
   [Schaff, Lauren R.; DeAngelis, Lisa M.; Grommes, Christian] Mem Sloan Kettering Canc Ctr, Dept Neurol, New York, NY 10065 USA.
   [Schaff, Lauren R.; DeAngelis, Lisa M.; Grommes, Christian] Weill Cornell Med Coll, Dept Neurol, New York, NY 10065 USA.
   [Nandakumar, Subhiksha] Mem Sloan Kettering Canc Ctr, Human Oncol & Pathogenesis Program, New York, NY USA.
   [Omuro, Antonio M. P.] Yale Sch Med, Dept Neurol, New Haven, CT USA.
C3 Stanford University; Memorial Sloan Kettering Cancer Center; Cornell
   University; Weill Cornell Medicine; Memorial Sloan Kettering Cancer
   Center; Yale University
RP Grommes, C (corresponding author), Mem Sloan Kettering Canc Ctr, Dept Neurol, New York, NY 10065 USA.; Grommes, C (corresponding author), Weill Cornell Med Coll, Dept Neurol, New York, NY 10065 USA.
EM
OI Schaff, Lauren/0000-0002-4664-670X
FU NIH [1R01NS08094401, P30-CA008748]; Lymphoma Research Foundation Career
   Development Award; Cycle for Survival Equinox Innovation Award
FX This research was supported by grants from the NIH (1R01NS080944-01 to
   I.K.M., P30-CA008748), the Lymphoma Research Foundation Career
   Development Award (C.G), and Cycle for Survival Equinox Innovation Award
   (C.G).
CR Correa DD, 2019, J NEURO-ONCOL, V144, P553, DOI 10.1007/s11060-019-03257-1
   FAZEKAS F, 1987, AM J ROENTGENOL, V149, P351, DOI 10.2214/ajr.149.2.351
   Ferreri AJM, 2017, LANCET HAEMATOL, V4, pE510, DOI 10.1016/S2352-3026(17)30174-6
   Gavrilovic IT, 2006, J CLIN ONCOL, V24, P4570, DOI 10.1200/JCO.2006.06.6910
   Grommes C, 2019, NEURO-ONCOLOGY, V21, P296, DOI 10.1093/neuonc/noy192
   Grommes C, 2017, J CLIN ONCOL, V35, P2410, DOI 10.1200/JCO.2017.72.7602
   Houillier C, 2019, J CLIN ONCOL, V37, P823, DOI 10.1200/JCO.18.00306
   Jahnke K, 2006, J NEURO-ONCOL, V80, P159, DOI 10.1007/s11060-006-9165-6
   Nayak L, 2017, NEURO-ONCOLOGY, V19, P625, DOI 10.1093/neuonc/nox029
   Nayak L, 2011, NEURO-ONCOLOGY, V13, P525, DOI 10.1093/neuonc/nor014
   Omuro A, 2015, BLOOD, V125, P1403, DOI 10.1182/blood-2014-10-604561
   Yamanaka R, 2017, JPN J CLIN ONCOL, V47, P101, DOI 10.1093/jjco/hyw171
NR 12
TC 1
Z9 1
U1 0
U2 0
PU LIPPINCOTT WILLIAMS & WILKINS
PI PHILADELPHIA
PA TWO COMMERCE SQ, 2001 MARKET ST, PHILADELPHIA, PA 19103 USA
SN 0028-3878
EI 1526-632X
J9 NEUROLOGY
JI Neurology
PD AUG 15
PY 2023
VL 101
IS 7
BP E710
EP E716
DI 10.1212/WNL.0000000000207490
PG 7
WC Clinical Neurology
WE Science Citation Index Expanded (SCI-EXPANDED)
SC Neurosciences & Neurology
GA O9SP4
UT WOS:001047146600014
PM 37344228
DA 2024-06-03
ER

PT J
AU Arshad, S
   Fang, X
   Ahn, KW
   Kaur, M
   Scordo, M
   Sauter, CS
   Furqan, F
   Awan, FT
   Hamadani, M
AF Arshad, Shanze
   Fang, Xi
   Ahn, Kwang W.
   Kaur, Manmeet
   Scordo, Michael
   Sauter, Craig S.
   Furqan, Fateeha
   Awan, Farrukh T.
   Hamadani, Mehdi
TI Impact of thiotepa dose-intensity in primary diffuse large B-cell
   lymphoma of the central nervous system undergoing autologous
   hematopoietic cell transplant with thiotepa/carmustine conditioning
SO BONE MARROW TRANSPLANTATION
LA English
DT Article
ID PRIMARY CNS LYMPHOMA; INTERNATIONAL EXTRANODAL LYMPHOMA;
   CHEMOIMMUNOTHERAPY; CONSOLIDATION; RANDOMIZATION; CHEMOTHERAPY;
   METHOTREXATE; RADIOTHERAPY; SURVIVAL; OUTCOMES
AB Thiotepa/carmustine (TT-BCNU) is a commonly used autologous transplant (ASCT) conditioning regimen for primary DLBCL of the CNS (PCNSL). The total thiotepa dose varies among TT-BCNU recipients, with some centers administering a total dose of 20 mg/kg, while others using 10 mg/kg. We retrospectively assessed the impact of thiotepa dose intensity on ASCT outcomes in 218 adult PCNSL patients who underwent a first ASCT with TT-BCNU conditioning and received either a total thiotepa dose of 10 mg/kg (TT-10 group; N = 90), or 20 mg/kg (TT-20 group; N = 128). The median follow-up of survivors was 22 months. The cumulative incidence of 1-year non-relapse mortality (NRM) for TT-10 and TT-20 cohorts were 6% (95%CI = 2-12%) vs. 4% (95%CI = 1-8%), respectively (p = 0.66). The 3-year cumulative incidence of relapse (15% vs. 13%; p = 0.67), progression-free survival (PFS) (71% vs. 80%; p = 0.25) and overall survival (OS) (79% vs. 83%; p = 0.56) were similar in the TT-10 and TT-20 groups, respectively. On multivariate analysis compared to TT-10, the TT-20 cohort was not associated with significantly different risk of NRM (Hazard ration [HR] = 0.77; p = 0.64), relapse/progression (HR = 0.87; p = 0.74), PFS (HR = 0.80; p = 0.48) or OS (HR = 1.10; p = 0.80). In conclusion thiotepa dose-intensity in TT-BCNU conditioning does not impact ASCT outcomes of PCNSL patients.
C1 [Arshad, Shanze] Univ Kentucky, Sch Med, Lexington, KY USA.
   [Fang, Xi; Ahn, Kwang W.] Med Coll Wisconsin, Inst Hlth & Equ, Div Biostat, Milwaukee, WI USA.
   [Kaur, Manmeet] Med Coll Wisconsin, Ctr Int Blood & Marrow Transplant Res, Dept Med, Milwaukee, WI USA.
   [Scordo, Michael] Mem Sloan Kettering Canc Ctr, New York, NY USA.
   [Sauter, Craig S.] Cleveland Clin Fdn, Cleveland, OH USA.
   [Furqan, Fateeha; Hamadani, Mehdi] Med Coll Wisconsin, Dept Med, Milwaukee, WI 53226 USA.
   [Awan, Farrukh T.] UT Southwestern, Div Hematol & Oncol, Dallas, TX USA.
C3 University of Kentucky; Medical College of Wisconsin; Medical College of
   Wisconsin; Center for International Blood & Marrow Transplant Research;
   Memorial Sloan Kettering Cancer Center; Cleveland Clinic Foundation;
   Medical College of Wisconsin; University of Texas System; University of
   Texas Southwestern Medical Center Dallas
RP Hamadani, M (corresponding author), Med Coll Wisconsin, Dept Med, Milwaukee, WI 53226 USA.
EM
OI Hamadani, Mehdi/0000-0001-5372-510X
FU Public Health Service from the National Cancer Institute (NCI)
   [U24CA076518]; National Cancer Institute (NCI); National Heart, Lung and
   Blood Institute (NHLBI); National Institute of Allergy and Infectious
   Diseases (NIAID); National Institute of Allergy and Infectious Diseases
   (NIAID); Health Resources and Services Administration (HRSA)
   [N00014-21-1-2954]; Office of Naval Research [N00014-23-1-2057]; Medical
   College of Wisconsin; National Marrow Donor Program;  [75R60222C00011]
FX This dataset was collected by the Center for International Blood and
   Marrow Transplant Research which is supported primarily by the Public
   Health Service U24CA076518 from the National Cancer Institute (NCI), the
   National Heart, Lung and Blood Institute (NHLBI) and the National
   Institute of Allergy and Infectious Diseases (NIAID); 75R60222C00011
   from the Health Resources and Services Administration (HRSA);
   N00014-21-1-2954 and N00014-23-1-2057 from the Office of Naval Research;
   the Medical College of Wisconsin and the National Marrow Donor Program.
CR Batchelor T, 2021, J CLIN ONCOL, V39, DOI 10.1200/JCO.2021.39.15_suppl.7506
   Cheson BD, 2014, J CLIN ONCOL, V32, P3059, DOI 10.1200/JCO.2013.54.8800
   cibmtr, US
   Commenges D, 1995, Lifetime Data Anal, V1, P145, DOI 10.1007/BF00985764
   Curry LD, 2023, ANN HEMATOL, V102, P141, DOI 10.1007/s00277-022-05018-z
   Epperla N, 2023, HEMATOL ONCOL, V41, P88, DOI 10.1002/hon.3083
   Ferreri AJM, 2022, LEUKEMIA, V36, P1870, DOI 10.1038/s41375-022-01582-5
   Ferreri AJM, 2017, LANCET HAEMATOL, V4, pE510, DOI 10.1016/S2352-3026(17)30174-6
   Ferreri AJM, 2016, LANCET HAEMATOL, V3, pE217, DOI 10.1016/S2352-3026(16)00036-3
   Glass J, 2016, J CLIN ONCOL, V34, P1620, DOI 10.1200/JCO.2015.64.8634
   Houillier C, 2019, J CLIN ONCOL, V37, P823, DOI 10.1200/JCO.18.00306
   Illerhaus G, 2022, BLOOD, V140, P10, DOI 10.1182/blood-2022-171733
   Illerhaus G, 2006, J CLIN ONCOL, V24, P3865, DOI 10.1200/JCO.2006.06.2117
   Khurana A, 2020, BIOL BLOOD MARROW TR, V26, P2217, DOI 10.1016/j.bbmt.2020.08.012
   Phillips Adrienne A, 2017, Am Soc Clin Oncol Educ Book, V37, P526, DOI 10.14694/EDBK_175444
   Roetzer T, 2020, CANCERS, V12, DOI 10.3390/cancers12061593
   Scordo M, 2021, JAMA ONCOL, V7, P993, DOI 10.1001/jamaoncol.2021.1074
   Shenoy PJ, 2011, CANCER-AM CANCER SOC, V117, P2530, DOI 10.1002/cncr.25765
   Steffanoni S, 2023, CANCERS, V15, DOI 10.3390/cancers15020526
   Tao L, 2014, BLOOD, V123, P3553, DOI 10.1182/blood-2013-07-517110
   Zhang X, 2007, COMPUT METH PROG BIO, V88, P95, DOI 10.1016/j.cmpb.2007.07.010
   Zhang X, 2011, COMPUT METH PROG BIO, V101, P87, DOI 10.1016/j.cmpb.2010.07.005
NR 22
TC 1
Z9 1
U1 0
U2 3
PU SPRINGERNATURE
PI LONDON
PA CAMPUS, 4 CRINAN ST, LONDON, N1 9XW, ENGLAND
SN 0268-3369
EI 1476-5365
J9 BONE MARROW TRANSPL
JI Bone Marrow Transplant.
PD NOV
PY 2023
VL 58
IS 11
BP 1203
EP 1208
DI 10.1038/s41409-023-02071-8
EA AUG 2023
PG 6
WC Biophysics; Oncology; Hematology; Immunology; Transplantation
WE Science Citation Index Expanded (SCI-EXPANDED)
SC Biophysics; Oncology; Hematology; Immunology; Transplantation
GA Y0PJ3
UT WOS:001045582400001
PM 37563283
DA 2024-06-03
ER

PT J
AU Usmani, S
   Jain, A
   Al Riyami, K
   Naumani, SP
   Al-Sukaiti, R
   Shamim, MS
AF Usmani, Sharjeel
   Jain, Anjali
   Al Riyami, Khulood
   Naumani, Shah P.
   Al-Sukaiti, Rashid
   Shamim, Muhammad Shahzad
TI The role of <SUP>18</SUP>F-FDG PET/CT in evaluation of Primary CNS
   lymphoma: The path less travelled
SO JOURNAL OF THE PAKISTAN MEDICAL ASSOCIATION
LA English
DT Article
DE F-18-FDG PET/CT; CNS lymphoma; brain tumors
ID NERVOUS-SYSTEM LYMPHOMAS
AB Primary central nervous system lymphoma (PCNSL) is a rare but highly aggressive lymphoma with increasing incidence in immunocompromised patients. MRI is the modality of choice in evaluating brain lesions. However, MRI is often challenging in the detection of early recurrence, assessing residual disease and response evaluation in PCNSL. F-18-FDG PET/ CT has superior diagnostic performance compared with body CT in the evaluation of lymphoma. F-18-FDG PET-CT is helpful in evaluating evaluation of disease extent and differentiating primary CNS lymphoma from systemic lymphoma. Besides diagnostic and prognostic value in primary CNS lymphoma, it might also be helpful in response assessment. The role of FDG-PET in PCNSL is not fully defined. In this article we have reviewed the potential role of F-18-FDG PET/CT in initial diagnosis, baseline staging, restaging, evaluation of treatment response, prognostication, and survival analysis of PCNSL.
C1 [Usmani, Sharjeel; Jain, Anjali; Al Riyami, Khulood; Al-Sukaiti, Rashid] Sultan Qaboos Comprehens Canc Care & Res Ctr, Dept Radiol & Nucl Med, Muscat, Oman.
   [Naumani, Shah P.] SUNY Stony Brook, Dept Radiol, Stony Brook, NY USA.
   [Shamim, Muhammad Shahzad] Aga Khan Univ Hosp, Karachi, Pakistan.
C3 State University of New York (SUNY) System; State University of New York
   (SUNY) Stony Brook; Aga Khan University
RP Shamim, MS (corresponding author), Aga Khan Univ Hosp, Karachi, Pakistan.
EM
RI Usmani, Sharjeel/AAQ-5043-2020
OI Usmani, Sharjeel/0000-0001-9274-7571
CR Bhagavathi S, 2008, ARCH PATHOL LAB MED, V132, P1830, DOI 10.1043/1543-2165-132.11.1830
   de-Bonilla-Damiá A, 2017, REV ESP MED NUCL IMA, V36, P298, DOI 10.1016/j.remn.2017.03.006
   del Rio MS, 2009, ONCOLOGIST, V14, P526, DOI 10.1634/theoncologist.2008-0236
   Fletcher JW, 2008, J NUCL MED, V49, P480, DOI 10.2967/jnumed.107.047787
   HOFFMAN JM, 1993, J NUCL MED, V34, P567
   Kawai N, 2010, J NEURO-ONCOL, V100, P225, DOI 10.1007/s11060-010-0182-0
   Krebs S, 2019, EUR J NUCL MED MOL I, V46, pS260
   Krebs Simone, 2021, Ann Lymphoma, V5, DOI 10.21037/aol-20-52
   Küker W, 2005, J NEURO-ONCOL, V72, P169, DOI 10.1007/s11060-004-3390-7
   Lu SS, 2014, AM J NEURORADIOL, V35, P270, DOI 10.3174/ajnr.A3677
   Makino K, 2011, ANN NUCL MED, V25, P432, DOI 10.1007/s12149-011-0483-1
   Malani R, 2019, LEUKEMIA LYMPHOMA, V60, P2278, DOI 10.1080/10428194.2018.1563294
   Maza S, 2013, RADIOL ONCOL, V47, P103, DOI 10.2478/raon-2013-0016
   Ömür Ö, 2014, DIAGN INTERV RADIOL, V20, P185, DOI 10.5152/dir.2013.13174
   Purandare NC, 2017, NUCL MED COMMUN, V38, P1109, DOI 10.1097/MNM.0000000000000753
   Siegel RL, 2022, CA-CANCER J CLIN, V72, P7, DOI 10.3322/caac.21708
   Thiel E, 2010, LANCET ONCOL, V11, P1036, DOI 10.1016/S1470-2045(10)70229-1
   Usmani Sharjeel, 2021, J Pak Med Assoc, V71, P388
   Zou YR, 2017, ONCOTARGET, V8, P41518, DOI 10.18632/oncotarget.17456
NR 19
TC 1
Z9 1
U1 0
U2 0
PU PAKISTAN MEDICAL ASSOC
PI KARACHI
PA PMA HOUSE, AGA KHAN III RD, KARACHI, 00000, PAKISTAN
SN 0030-9982
J9 J PAK MED ASSOC
JI J. Pak. Med. Assoc.
PD AUG
PY 2023
VL 73
IS 8
BP 1744
EP 1746
DI 10.47391/JPMA.23-59
PG 3
WC Medicine, General & Internal; Medicine, Research & Experimental
WE Science Citation Index Expanded (SCI-EXPANDED)
SC General & Internal Medicine; Research & Experimental Medicine
GA U2VO3
UT WOS:001083432300035
PM 37697779
OA gold
DA 2024-06-03
ER

PT J
AU Jiang, QS
   Zhan, GY
   Jiang, WH
   Xu, YF
   Zheng, GF
   Jiang, C
   Lin, DD
   Wang, K
   Zhu, HW
AF Jiang, Qingsong
   Zhan, Guoyong
   Jiang, Wenhong
   Xu, Yafeng
   Zheng, Guofu
   Jiang, Cai
   Lin, Dongdong
   Wang, Kang
   Zhu, Huiwu
TI Prognostic model and treatment choices for patients with primary
   intracranial central nervous system lymphoma: A population-based study
SO CLINICAL NEUROLOGY AND NEUROSURGERY
LA English
DT Article
DE Primary central nervous system lymphoma; SEER; Prognostic model
ID PRIMARY CNS LYMPHOMA; WHOLE-BRAIN RADIOTHERAPY; INTERNATIONAL EXTRANODAL
   LYMPHOMA; INTRACEREBRAL MALIGNANT-LYMPHOMA; HIGH-DOSE METHOTREXATE;
   RACIAL-DIFFERENCES; RADIATION-THERAPY; RITUXIMAB; SURVIVAL;
   CHEMOIMMUNOTHERAPY
AB Background: Primary central nervous system lymphoma (PCNSL) is a non-Hodgkin lymphoma that occurs in the CNS. With the advancement of medical care, its prognosis and treatment have also undergone tremendous changes. This study aimed to construct a prognostic model and compare the effects of different treatments for intracranial PCNSL. Methods: Cases diagnosed as PCNSL between 2004 and 2015 were obtained from the Surveillance, Epidemiology, and End Results (SEER) database. Data were analyzed using Kaplan-Meier method and Cox regression analysis. Nomogram was built and validated using the R program. Results: A total of 2861 PCNSL patients were included in the analysis. Age, year of diagnosis, surgery and chemotherapy were independent predictors for both overall survival (OS) and cancer-specific survival (CSS). A nomogram was established to predict 3-, 5- and 10-year OS and CSS for patients. Receiver operating characteristic (ROC) curves and decision curve analysis (DCA) showed the nomogram had good predictive performance and clinical application value. We also revealed that gross total resection had significantly better OS and CSS than biopsy alone (P < 0.001). Patients who received only chemotherapy had the best prognosis and did not benefit from additional radiotherapy. Conclusion: We developed a nomogram to predict patient survival rates based on independent predictors. It is an effective tool to help clinicians make survival predictions. Our results showed that patients can benefit from gross total resection of tumor, if it is feasible, and chemotherapy. The role of radiotherapy remained to be further assessed.
C1 [Jiang, Qingsong; Zhan, Guoyong; Zheng, Guofu; Jiang, Cai; Wang, Kang] KeCheng Peoples Hosp, Dept Neurosurg, Quzhou, Zhejiang, Peoples R China.
   [Jiang, Wenhong] KeCheng Peoples Hosp, Dept Med Records Informat Technol, Quzhou, Zhejiang, Peoples R China.
   [Xu, Yafeng] KeCheng Peoples Hosp, Dept Nursing, Quzhou, Zhejiang, Peoples R China.
   [Lin, Dongdong] Zhejiang Univ, Affiliated Zhejiang Hosp, Brain Ctr, Sch Med, Hangzhou, Peoples R China.
   [Zhu, Huiwu] KeCheng Peoples Hosp, Brain Ctr, Quzhou, Zhejiang, Peoples R China.
   [Zhu, Huiwu] 172 Shuanggang Rd, Quzhou, Zhejiang, Peoples R China.
C3 Zhejiang University
RP Zhu, HW (corresponding author), 172 Shuanggang Rd, Quzhou, Zhejiang, Peoples R China.
EM
FU Quzhou Kecheng District Project, China [2021C23]
FX Acknowledgements The study was supported by funds from the Quzhou
   Kecheng District Project, China (Grant Number: 2021C23) .
CR Abrey LE, 2006, J CLIN ONCOL, V24, P5711, DOI 10.1200/JCO.2006.08.2941
   Abrey LE, 2005, J CLIN ONCOL, V23, P5034, DOI 10.1200/JCO.2005.13.524
   Bataille B, 2000, J NEUROSURG, V92, P261, DOI 10.3171/jns.2000.92.2.0261
   Batchelor T, 2003, J CLIN ONCOL, V21, P1044, DOI 10.1200/JCO.2003.03.036
   BERRY MP, 1981, INT J RADIAT ONCOL, V7, P55, DOI 10.1016/0360-3016(81)90060-2
   Coiffier B, 2002, NEW ENGL J MED, V346, P235, DOI 10.1056/NEJMoa011795
   DeAngelis LM, 2002, J CLIN ONCOL, V20, P4643, DOI 10.1200/JCO.2002.11.013
   Ferreri AJM, 2003, J CLIN ONCOL, V21, P266, DOI 10.1200/JCO.2003.09.139
   Ferreri AJM, 2017, LANCET HAEMATOL, V4, pE510, DOI 10.1016/S2352-3026(17)30174-6
   Ferreri AJM, 2016, LANCET HAEMATOL, V3, pE217, DOI 10.1016/S2352-3026(16)00036-3
   Haldorsen IS, 2011, AM J NEURORADIOL, V32, P984, DOI 10.3174/ajnr.A2171
   Morris PG, 2013, J CLIN ONCOL, V31, P3971, DOI 10.1200/JCO.2013.50.4910
   Nguyen PL, 2005, J CLIN ONCOL, V23, P1507, DOI 10.1200/JCO.2005.01.161
   Norden AD, 2011, J NEURO-ONCOL, V101, P487, DOI 10.1007/s11060-010-0269-7
   O'Neill BP, 2013, AM J HEMATOL, V88, P997, DOI 10.1002/ajh.23551
   Olson JE, 2002, CANCER, V95, P1504, DOI 10.1002/cncr.10851
   Ostrom QT, 2018, NEURO-ONCOLOGY, V20, P1, DOI 10.1093/neuonc/noy131
   POLLACK IF, 1989, CANCER-AM CANCER SOC, V63, P939, DOI 10.1002/1097-0142(19890301)63:5<939::AID-CNCR2820630526>3.0.CO;2-V
   Pulido JS, 2009, NEURO-ONCOLOGY, V11, P318, DOI 10.1215/15228517-2008-103
   Schlegel U, 2018, CURR OPIN NEUROL, V31, P733, DOI 10.1097/WCO.0000000000000619
   Shah GD, 2007, J CLIN ONCOL, V25, P4730, DOI 10.1200/JCO.2007.12.5062
   Thiel E, 2010, LANCET ONCOL, V11, P1036, DOI 10.1016/S1470-2045(10)70229-1
   TOMLINSON FH, 1995, J NEUROSURG, V82, P558, DOI 10.3171/jns.1995.82.4.0558
   Villano JL, 2011, BRIT J CANCER, V105, P1414, DOI 10.1038/bjc.2011.357
   Weller M, 2012, NEURO-ONCOLOGY, V14, P1481, DOI 10.1093/neuonc/nos159
NR 25
TC 1
Z9 1
U1 0
U2 0
PU ELSEVIER
PI AMSTERDAM
PA RADARWEG 29, 1043 NX AMSTERDAM, NETHERLANDS
SN 0303-8467
EI 1872-6968
J9 CLIN NEUROL NEUROSUR
JI Clin. Neurol. Neurosurg.
PD OCT
PY 2023
VL 233
AR 107912
DI 10.1016/j.clineuro.2023.107912
EA JUL 2023
PG 8
WC Clinical Neurology; Surgery
WE Science Citation Index Expanded (SCI-EXPANDED)
SC Neurosciences & Neurology; Surgery
GA P5VP1
UT WOS:001051354300001
PM 37531751
DA 2024-06-03
ER

PT J
AU Cohen, YI
   Lebel, E
   Zimran, E
   Shaulov, A
   Stepensky, P
   Grisariu, S
   Avni, B
AF Cohen, Yarden Irit
   Lebel, Eyal
   Zimran, Eran
   Shaulov, Adir
   Stepensky, Polina
   Grisariu, Sigal
   Avni, Batia
TI Long-Term Results with Thiotepa-Containing Conditioning Regimens for
   Autologous Stem Cell Transplantation
SO TRANSPLANTATION AND CELLULAR THERAPY
LA English
DT Article
DE TECAM; Autologous stem cell; transplantation; Lymphoma; Conditioning;
   Thiotepa
ID HIGH-DOSE THERAPY; RESPONSE CRITERIA; HODGKIN-LYMPHOMA; CHEMOTHERAPY
AB Autologous stem cell transplantation (ASCT) remains a cornerstone in the treatment of both Hodgkin lymphoma (HL) and various non-Hodgkin lymphoma (NHL) subtypes. BEAM (carmustine, etoposide, cytarabine, and melphalan) is the most frequently used conditioning regimen; however, owing due to limited availability and toxicity of carmustine, thiotepa-containing regimens have been suggested. We previously reported encouraging results in ASCT with a TECAM (thiotepa, etoposide, cyclophosphamide, cytarabine, and melphalan) conditioning regimen from 2000 to 2013. We aimed to update our experience with the TECAM regimen by adding our experience from 2013 to 2020 to the previously reported cohort. Moreover, we aimed to use the detailed data for the 2 transplant cohorts to identify improvements in ASCT outcomes in the recent era. We retrospectively analyzed all lymphoma patients who underwent ASCT at our center between January 2000 and December 2020. A total of 353 lymphoma patients were included (142 in the newer cohort added to 211 previously reported patients), all of whom were treated with our standard TECAM conditioning regimen. The cohort included 127 patients with HL, 107 with DLBCL, and 119 with other NHL subtypes. The newer cohort was characterized by significantly poorer Eastern Cooperative Oncology Group Performance Status (ECOG-PS) prior to ASCT (45.7% versus 19.3% with ECOG-PS >= 1; P < .01), whereas a higher proportion of patients entered transplantation in complete response (CR) (71.9% versus 47.8%; P < .01). The median follow-up after ASCT was 136.4 months (95% confidence interval [CI], 91.4 to 181.4 months). The 3-year progression-free survival (PFS) and overall survival (OS) rates post-ASCT for the entire cohort were 59.8% and 79.3%, respectively. Evaluating the 303 of 353 patients (86.4%) who entered ASCT with a responsive disease-a population that represents today's approach to the selection of patients for ASCT-the 3-year PFS and OS rates were 61.5% and 81.9%, respectively. In this population, the 3-year PFS rate was 62.2% for HL, 62.6% for DLBCL, 64.3% for primary central nervous system lymphoma (PCNSL), and the 3-year OS rate were 90.1%, 75.2%, and 78.6%, respectively. OS was significantly better in the newer cohort (P < .01), but not when evaluating only patients who entered ASCT with responsive disease. Dose reductions, poor disease status, and poor ECOG-PS at ASCT entry were associated with worse outcomes across all lymphoma subtypes. In accordance with our previous report, patients entering transplantation for DLBCL with a partial response achieved similar outcomes as those with a CR. Eighteen patients died within the first 100 days, 8 due to disease progression and 10 due to transplantation-related complications (2.8%). There were no cases of interstitial pneumonitis syndrome. Twenty-two cases (6.2%) of secondary malignancies were documented. Our results confirm that TECAM is an effective and safe conditioning regimen for ASCT in patients with HL and various NHLs, including favorable results in PCNSL. Despite a higher proportion of frail patients, the newer cohort's outcomes were favorable, driven by better lymphoma control pretransplantation. In the DLBCL cohort, ECOG-PS had more prognostic value than achieving a CR pre-ASCT, a finding relevant to the optimal allocation of patients to different treatment options in the era of chimeric antigen receptor T cell availability.(C) 2023 The American Society for Transplantation and Cellular Therapy. Published by Elsevier Inc. All rights reserved.
C1 [Cohen, Yarden Irit; Lebel, Eyal; Zimran, Eran; Shaulov, Adir; Stepensky, Polina; Grisariu, Sigal; Avni, Batia] Hebrew Univ Jerusalem, Fac Med, Jerusalem, Israel.
   [Cohen, Yarden Irit] Hebrew Univ Jerusalem, Fac Med, Dept Mil Med & Tzameret, Jerusalem, Israel.
   [Cohen, Yarden Irit] Israel Def Forces, Med Corps, Jerusalem, Israel.
   [Lebel, Eyal; Zimran, Eran; Shaulov, Adir; Stepensky, Polina; Grisariu, Sigal; Avni, Batia] Hadassah Hebrew Univ, Bone Marrow Transplantat & Canc Immunotherapy Dept, Med Ctr, Jerusalem, Israel.
   [Lebel, Eyal] Hadassah Med Ctr, Bone Marrow Transplantat & Cellular Therapy Dept, Kalman Mann 1 St, IL-9574869 Jerusalem, Israel.
C3 Hebrew University of Jerusalem; Hebrew University of Jerusalem; Hebrew
   University of Jerusalem; Hadassah University Medical Center; Hebrew
   University of Jerusalem
RP Lebel, E (corresponding author), Hadassah Med Ctr, Bone Marrow Transplantat & Cellular Therapy Dept, Kalman Mann 1 St, IL-9574869 Jerusalem, Israel.
EM
RI Shaulov, Adir/K-7375-2019
OI grisariu, sigal/0000-0001-7190-6326; Zimran, Eran/0000-0002-5483-5180;
   Lebel, Eyal/0000-0001-5215-7278
CR Abdul-Hai A, 2007, BONE MARROW TRANSPL, V40, P891, DOI 10.1038/sj.bmt.1705838
   Ahya VN, 2017, CLIN CHEST MED, V38, P595, DOI 10.1016/j.ccm.2017.07.002
   Becnel Melody, 2017, Ann Lymphoma, V1, DOI 10.21037/aol.2017.11.01
   Caballero MD, 1997, BONE MARROW TRANSPL, V20, P451, DOI 10.1038/sj.bmt.1700913
   Chen YB, 2014, TRANSPLANT CELL THER, V20, P104
   Chen YB, 2015, BIOL BLOOD MARROW TR, V21, P1046, DOI 10.1016/j.bbmt.2015.02.005
   Cheson BD, 1999, J CLIN ONCOL, V17, P1244, DOI 10.1200/JCO.1999.17.4.1244
   Cheson BD, 2007, J CLIN ONCOL, V25, P579, DOI 10.1200/JCO.2006.09.2403
   Cheson BD, 2014, J CLIN ONCOL, V32, P3059, DOI 10.1200/JCO.2013.54.8800
   Chukwueke U, 2022, HEMATOL ONCOL CLIN N, V36, P147, DOI 10.1016/j.hoc.2021.09.004
   Du J, 2021, JAMA NETW OPEN, V4, DOI 10.1001/jamanetworkopen.2021.9807
   Ferreri AJM, 2022, LEUKEMIA, V36, P1870, DOI 10.1038/s41375-022-01582-5
   Goodman KA, 2008, J CLIN ONCOL, V26, P5240, DOI 10.1200/JCO.2007.15.5507
   Grisariu S, 2018, CL LYMPH MYELOM LEUK, V18, P272, DOI 10.1016/j.clml.2018.02.008
   Gunes Ahmet Kursad, 2022, Hematol Oncol Stem Cell Ther, V15, P201, DOI 10.1016/j.hemonc.2021.03.005
   Joffe E, 2018, BONE MARROW TRANSPL, V53, P29, DOI 10.1038/bmt.2017.205
   Kamdar M, 2022, LANCET, V399, P2294, DOI 10.1016/S0140-6736(22)00662-6
   LaCasce AS, 2019, HEMATOL ONCOL, V37, P87, DOI 10.1002/hon.2589
   Locke FL, 2022, NEW ENGL J MED, V386, P640, DOI 10.1056/NEJMoa2116133
   Marchesi F, 2020, LEUKEMIA LYMPHOMA, V61, P2238, DOI 10.1080/10428194.2020.1765238
   Schaaf M, 2012, COCHRANE DB SYST REV, DOI 10.1002/14651858.CD007678.pub2
   Scordo M, 2021, JAMA ONCOL, V7, P993, DOI 10.1001/jamaoncol.2021.1074
   Sellner L, 2016, BONE MARROW TRANSPL, V51, P212, DOI 10.1038/bmt.2015.273
   Shadman M, 2022, BLOOD, V139, P1330, DOI 10.1182/blood.2021013289
   Shah NA, 2014, TRANSPLANT CELL THER, V20, P104
   Stiff P, 2015, HEMATOL-AM SOC HEMAT, P74, DOI 10.1182/asheducation-2015.1.74
   Zahid U, 2017, CURR HEMATOL MALIG R, V12, P217, DOI 10.1007/s11899-017-0382-1
NR 27
TC 1
Z9 1
U1 2
U2 2
PU ELSEVIER SCIENCE INC
PI NEW YORK
PA STE 800, 230 PARK AVE, NEW YORK, NY 10169 USA
SN 2666-6375
EI 2666-6367
J9 TRANSPL CELL THER
JI Transpl. Cell. Ther.
PD AUG
PY 2023
VL 29
IS 8
DI 10.1016/j.jtct.2023.04.022
EA JUL 2023
PG 8
WC Hematology; Immunology; Transplantation
WE Science Citation Index Expanded (SCI-EXPANDED)
SC Hematology; Immunology; Transplantation
GA Q9NR1
UT WOS:001060716300001
PM 37160229
DA 2024-06-03
ER

PT J
AU Wong, CE
   Liao, WA
   Chang, Y
   Lee, PH
   Huang, CC
   Chang, KC
   Lee, JS
AF Wong, Chia-En
   Liao, Wei-An
   Chang, Yu
   Lee, Po-Hsuan
   Huang, Chi-Chen
   Chang, Kung-Chao
   Lee, Jung-Shun
TI The role of comorbidity indices and histochemical markers in surgically
   resected and non-resected primary central nervous system lymphoma
SO CLINICAL AND EXPERIMENTAL MEDICINE
LA English
DT Article
DE Primary central nervous system lymphoma; Surgical resection;
   Stereotactic biopsy; Prognostic nutritional index; PD-L1; Comorbidity
   indices
ID PRIMARY CNS LYMPHOMA; B-CELL LYMPHOMA; PROGNOSTIC-FACTORS; SURVIVAL;
   EXPRESSION; ALBUMIN; IMMUNOHISTOCHEMISTRY; PREDICTOR; OUTCOMES; RISK
AB The role of surgical resection in primary central nervous system lymphoma (PCNSL) was not recognized until recently. However, prognostic factors for surgically treated PCNSL remain unclear. In the present study, we aimed to identify and compare the prognostic value of comorbidity indices and immunohistochemical markers in patients with surgically and non-surgically treated PCNSL. This retrospective single-center study analyzed patients who underwent either surgical resection or stereotactic biopsy for newly diagnosed PCNSL between January 2012 and December 2021. Clinical demographics, comorbidity indices, and immunohistochemical markers were analyzed. We included 23 and 18 patients who underwent stereotactic biopsy and surgical resection, respectively. The median overall survival (OS) was 11.05 months. Using multivariate Cox regression, we identified pretreatment prognostic nutritional index (PNI) (p = 0.009), positive BCL2 staining (p = 0.026), and infratentorial involvement (p = 0.004) as independent prognostic factors of OS. Predictors of progression-free survival (PFS) included PNI (p = 0.040), infratentorial involvement (p = 0.021), and surgical resection for PCNSL (p = 0.048). Subgroup analyses revealed that positive BCL2 (p = 0.048) and PD-L1 (p = 0.037) staining were associated with worse OS in the biopsy group. PNI and infratentorial involvement could significantly impact both OS and PFS in patients with PCNSL. Surgical resection could predict favorable PFS but not OS. Moreover, BCL2 and PD-L1 expression can be employed as prognostic markers in these patients.
C1 [Wong, Chia-En; Chang, Yu; Lee, Po-Hsuan; Huang, Chi-Chen; Lee, Jung-Shun] Natl Cheng Kung Univ, Natl Cheng Kung Univ Hosp, Coll Med, Dept Surg,Sect Neurosurg, Tainan, Taiwan.
   [Liao, Wei-An; Chang, Kung-Chao] Natl Cheng Kung Univ, Natl Cheng Kung Univ Hosp, Coll Med, Dept Pathol, Tainan, Taiwan.
   [Lee, Jung-Shun] Natl Cheng Kung Univ, Coll Med, Dept Cell Biol & Anat, Tainan, Taiwan.
   [Lee, Jung-Shun] Natl Cheng Kung Univ, Inst Basic Med Sci, Coll Med, Tainan, Taiwan.
C3 National Cheng Kung University; National Cheng Kung University Hospital;
   National Cheng Kung University; National Cheng Kung University Hospital;
   National Cheng Kung University; National Cheng Kung University
RP Lee, JS (corresponding author), Natl Cheng Kung Univ, Natl Cheng Kung Univ Hosp, Coll Med, Dept Surg,Sect Neurosurg, Tainan, Taiwan.; Lee, JS (corresponding author), Natl Cheng Kung Univ, Coll Med, Dept Cell Biol & Anat, Tainan, Taiwan.; Lee, JS (corresponding author), Natl Cheng Kung Univ, Inst Basic Med Sci, Coll Med, Tainan, Taiwan.
EM
CR Abdulla M, 2021, ACTA ONCOL, V60, P531, DOI 10.1080/0284186X.2021.1881161
   Aryal M, 2014, ADV DRUG DELIVER REV, V72, P94, DOI 10.1016/j.addr.2014.01.008
   Atas U, 2023, NUTR CANCER, V75, P591, DOI 10.1080/01635581.2022.2142248
   Bobillo S, 2023, HAEMATOLOGICA, V108, P673, DOI 10.3324/haematol.2022.281457
   Boshrabadi Ahmad Pourrashidi, 2017, Surg Neurol Int, V8, P165, DOI 10.4103/sni.sni_446_16
   Correia C, 2015, BLOOD, V125, P658, DOI 10.1182/blood-2014-04-571786
   Cozzolino I, 2016, CANCER CYTOPATHOL, V124, P135, DOI 10.1002/cncy.21626
   Das S, 2022, MEDICINA-LITHUANIA, V58, DOI 10.3390/medicina58020281
   Deng XY, 2020, FRONT ONCOL, V10, DOI 10.3389/fonc.2020.00131
   El-Galaly TC, 2018, EUR J CANCER, V93, P57, DOI 10.1016/j.ejca.2018.01.073
   Farhi J, 2018, ANN HEMATOL, V97, P2391, DOI 10.1007/s00277-018-3468-5
   Ferreri AJM, 2003, J CLIN ONCOL, V21, P266, DOI 10.1200/JCO.2003.09.139
   Ferroli P, 2015, NEUROSURG FOCUS, V39, DOI 10.3171/2015.9.FOCUS15339
   González-Bonet LG, 2017, WORLD NEUROSURG, V99, DOI 10.1016/j.wneu.2016.12.122
   Gibbs J, 1999, ARCH SURG-CHICAGO, V134, P36, DOI 10.1001/archsurg.134.1.36
   Grommes C, 2019, NEURO-ONCOLOGY, V21, P296, DOI 10.1093/neuonc/noy192
   Grommes C, 2017, J CLIN ONCOL, V35, P2410, DOI 10.1200/JCO.2017.72.7602
   Gupta D, 2010, NUTR J, V9, DOI 10.1186/1475-2891-9-69
   Houillier C, 2020, NEUROLOGY, V94, pE1027, DOI 10.1212/WNL.0000000000008900
   Huang JJ, 2011, ANN ONCOL, V22, P149, DOI 10.1093/annonc/mdq314
   Hübner M, 2016, GASTROENT RES PRACT, V2016, DOI 10.1155/2016/8743187
   Jeswani S, 2013, NEUROSURGERY, V73, P240, DOI 10.1227/01.neu.0000430288.85680.37
   Junger ST, 2021, CANCERS, V13, DOI 10.3390/cancers13061435
   Karbhari N, 2022, CASE REP ONCOL, V15, P345, DOI 10.1159/000523858
   Kim DH, 2007, LEUKEMIA, V21, P2227, DOI 10.1038/sj.leu.2404780
   Kim SY, 2016, BMC EVOL BIOL, V16, DOI [10.1186/s12862-016-0613-4, 10.1186/s12911-016-0249-8]
   Kiyasu J, 2015, BLOOD, V126, P2193, DOI 10.1182/blood-2015-02-629600
   Kreher S, 2015, NEURO-ONCOLOGY, V17, P1016, DOI 10.1093/neuonc/nov046
   KROGHJENSEN M, 1995, LEUKEMIA LYMPHOMA, V19, P223
   Kudsk KA, 2003, JPEN-PARENTER ENTER, V27, P1, DOI 10.1177/014860710302700101
   Larsen M.T., 2016, Mol Cell Ther
   Nguyen L, 2017, GENES-BASEL, V8, DOI 10.3390/genes8040116
   Makino K, 2018, J NEURO-ONCOL, V140, P115, DOI 10.1007/s11060-018-2940-3
   Meng XR, 2018, INT J CLIN ONCOL, V23, P783, DOI 10.1007/s10147-018-1268-5
   Moujaess E, 2017, CRIT REV ONCOL HEMAT, V120, P203, DOI 10.1016/j.critrevonc.2017.11.008
   Ni LW, 2022, FRONT NUTR, V9, DOI 10.3389/fnut.2022.823087
   Nosrati A, 2019, ANN HEMATOL, V98, P169, DOI 10.1007/s00277-018-3498-z
   Ou A, 2020, NEURO-ONCOL ADV, V2, DOI 10.1093/noajnl/vdaa018
   Rae AI, 2019, NEUROSURGERY, V84, P935, DOI 10.1093/neuros/nyy096
   Rao IS, 2010, INDIAN J MED PAEDIAT, V31, P145, DOI 10.4103/0971-5851.76201
   Rivera Perla KM, 2021, SURG NEUROL INT, V12
   Roh J, 2022, MODERN PATHOL, V35, P480, DOI 10.1038/s41379-021-00962-z
   Rozet Irene, 2007, Anesthesiol Clin, V25, P631, DOI 10.1016/j.anclin.2007.05.009
   Samhouri Y, 2022, ANTICANCER RES, V42, P1867, DOI 10.21873/anticanres.15663
   Schellekes N, 2021, J NEUROSURG, V135, P1016, DOI 10.3171/2020.9.JNS201980
   Shi QY, 2017, J NEUROPATH EXP NEUR, V76, P942, DOI 10.1093/jnen/nlx083
   Shivakumar L, 2006, CLIN LYMPHOMA MYELOM, V6, P455, DOI 10.3816/CLM.2006.n.025
   Siegal T, 2019, ACTA HAEMATOL-BASEL, V141, P138, DOI 10.1159/000495284
   Soedjana H, 2021, Ann Burns Fire Disasters, V34, P157
   Tsuyama N, 2017, BLOOD, V130, P489, DOI 10.1182/blood-2016-12-759621
   Turner DC, 2018, CLIN CANCER RES, V24, P5841, DOI 10.1158/1078-0432.CCR-18-0415
   Weller M, 2012, NEURO-ONCOLOGY, V14, P1481, DOI 10.1093/neuonc/nos159
   Wu S, 2021, BMC MED IMAGING, V21, DOI 10.1186/s12880-021-00546-y
   Zhao S, 2018, CANCER BIOL MED, V15, P290, DOI 10.20892/j.issn.2095-3941.2018.0047
NR 54
TC 1
Z9 1
U1 0
U2 3
PU SPRINGER-VERLAG ITALIA SRL
PI MILAN
PA VIA DECEMBRIO, 28, MILAN, 20137, ITALY
SN 1591-8890
EI 1591-9528
J9 CLIN EXP MED
JI Clin. Exper. Med.
PD NOV
PY 2023
VL 23
IS 7
BP 3799
EP 3807
DI 10.1007/s10238-023-01130-0
EA JUL 2023
PG 9
WC Medicine, Research & Experimental
WE Science Citation Index Expanded (SCI-EXPANDED)
SC Research & Experimental Medicine
GA AR7H8
UT WOS:001035508200001
PM 37491648
DA 2024-06-03
ER

PT J
AU Wang, F
   Zhou, XF
   Chen, RQ
   Kang, J
   Yang, XY
   Lin, JZ
   Liu, F
   Cao, DR
   Xing, Z
AF Wang, Feng
   Zhou, Xiaofang
   Chen, Ruiquan
   Kang, Jie
   Yang, Xinyi
   Lin, Jinzhu
   Liu, Fang
   Cao, Dairong
   Xing, Zhen
TI Improved performance of non-preloaded and high flip-angle dynamic
   susceptibility contrast perfusion-weighted imaging sequences in the
   presurgical differentiation of brain lymphoma and glioblastoma
SO EUROPEAN RADIOLOGY
LA English
DT Article
DE Glioblastoma; Lymphoma; Perfusion; Cerebral blood volume; Contrast media
ID CEREBRAL BLOOD-VOLUME; PERCENTAGE SIGNAL RECOVERY; NERVOUS-SYSTEM
   LYMPHOMAS; GRADIENT-ECHO; TUMOR GRADE; MRI; RECOMMENDATIONS; PARAMETERS;
   LESIONS; MAPS
AB ObjectiveThis study aimed to compare the accuracy of relative cerebral blood volume (rCBV) and percentage signal recovery (PSR) obtained from high flip-angle dynamic susceptibility contrast perfusion-weighted imaging (DSC-PWI) sequences with and without contrast agent (CA) preload for presurgical discrimination of brain glioblastoma and lymphoma.MethodsConsecutive 336 patients (glioblastoma, 236; PCNSL, 100) were included. All the patients underwent DSC-PWI on 3.0-T magnetic resonance units before surgery. The rCBV and PSR with preloaded and non-preloaded CA were measured. The means of the continuous variables were compared using Welch's t-test. The diagnostic accuracies of the individual parameters were compared using the receiver operating characteristic curve analysis.ResultsThe rCBV was higher with preloaded CA than with non-preloaded CA (glioblastoma, 10.20 vs. 8.90, p = 0.020; PCNSL, 3.88 vs. 3.27, p = 0.020). The PSR was lower with preloaded CA than with non-preloaded CA (glioblastoma, 0.59 vs. 0.90; PCNSL, 0.70 vs. 1.63; all p < 0.001). Regarding the differentiation of glioblastoma and PCNSL, the AUC of rCBV with preloaded CA was indistinguishable from that of non-preloaded CA (0.940 vs. 0.949, p = 0.703), whereas the area under the curve of PSR with preloaded CA was lower than non-preloaded CA (0.529 vs. 0.884, p < 0.001).ConclusionWith preloaded CA, diagnostic performance in differentiating glioblastoma and PCNSL did not improve for rCBV and it was decreased for PSR. Therefore, high flip-angle non-preload DSC-PWI sequences offer excellent accuracy and may be of choice sequence for presurgical discrimination of brain lymphoma and glioblastoma.
C1 [Wang, Feng; Zhou, Xiaofang; Chen, Ruiquan; Kang, Jie; Yang, Xinyi; Lin, Jinzhu; Cao, Dairong; Xing, Zhen] Fujian Med Univ, Affiliated Hosp 1, Dept Radiol, 20 Cha Zhong Rd, Fuzhou 350005, Fujian, Peoples R China.
   [Wang, Feng; Zhou, Xiaofang; Chen, Ruiquan; Kang, Jie; Yang, Xinyi; Lin, Jinzhu; Cao, Dairong; Xing, Zhen] Fujian Med Univ, Affiliated Hosp 1, Natl Reg Med Ctr, Dept Radiol, Binhai Campus, Fuzhou, Peoples R China.
   [Liu, Fang] Fujian Med Univ, Affiliated Hosp 1, Dept Hyperbar Oxygen, Fuzhou, Peoples R China.
   [Cao, Dairong] Fujian Med Univ, Affiliated Hosp 1, Dept Radiol, Fujian Key Lab Precis Med Canc, Fuzhou, Peoples R China.
   [Cao, Dairong] Fujian Med Univ, Affiliated Hosp 1, Fujian Higher Educ Inst, Key Lab Radiat Biol, Fuzhou, Peoples R China.
C3 Fujian Medical University; Fujian Medical University; Fujian Medical
   University; Fujian Medical University; Fujian Medical University
RP Cao, DR; Xing, Z (corresponding author), Fujian Med Univ, Affiliated Hosp 1, Dept Radiol, 20 Cha Zhong Rd, Fuzhou 350005, Fujian, Peoples R China.; Cao, DR; Xing, Z (corresponding author), Fujian Med Univ, Affiliated Hosp 1, Natl Reg Med Ctr, Dept Radiol, Binhai Campus, Fuzhou, Peoples R China.; Cao, DR (corresponding author), Fujian Med Univ, Affiliated Hosp 1, Dept Radiol, Fujian Key Lab Precis Med Canc, Fuzhou, Peoples R China.; Cao, DR (corresponding author), Fujian Med Univ, Affiliated Hosp 1, Fujian Higher Educ Inst, Key Lab Radiat Biol, Fuzhou, Peoples R China.
EM
RI Yang, Xinyi/IVH-6916-2023; zhang, lm/JWP-8874-2024; Zhang,
   Xiaoyu/JXR-6386-2024; Zhou, Xiangfeng/KDO-8724-2024; liu,
   zhao/KGM-5884-2024; CHEN, BING/KHX-6659-2024; Shi, Yaolin/JXN-8322-2024
OI zhen, xing/0000-0002-2104-4681
CR Bell LC, 2017, TOMOGRAPHY, V3, P89, DOI 10.18383/j.tom.2017.00004
   Boxerman JL, 2013, AM J NEURORADIOL, V34, P1364, DOI 10.3174/ajnr.A3477
   Boxerman JL, 2012, AM J NEURORADIOL, V33, P1081, DOI 10.3174/ajnr.A2934
   Boxerman JL, 2020, NEURO-ONCOLOGY, V22, P1262, DOI 10.1093/neuonc/noaa141
   Boxerman JL, 2006, AM J NEURORADIOL, V27, P859
   Cha S, 2002, RADIOLOGY, V223, P11, DOI 10.1148/radiol.2231010594
   Choi KS, 2020, RADIOLOGY, V297, P178, DOI 10.1148/radiol.2020192763
   Cindil E, 2022, ACAD RADIOL, V29, pS52, DOI 10.1016/j.acra.2021.02.002
   Cindil E, 2021, NEURORADIOLOGY, V63, P331, DOI 10.1007/s00234-020-02522-9
   Donahue KM, 2000, MAGNET RESON MED, V43, P845, DOI 10.1002/1522-2594(200006)43:6<845::AID-MRM10>3.0.CO;2-J
   Gulani V, 2017, LANCET NEUROL, V16, P564, DOI 10.1016/S1474-4422(17)30158-8
   Heming M, 2022, GENOME MED, V14, DOI 10.1186/s13073-022-01110-1
   Kickingereder P, 2015, SCI REP-UK, V5, DOI 10.1038/srep16238
   Kim HS, 2020, EUR RADIOL, V30, P2171, DOI 10.1007/s00330-019-06598-7
   Lee JY, 2019, EUR RADIOL, V29, P5539, DOI 10.1007/s00330-019-06097-9
   Lee MD, 2019, AM J NEURORADIOL, V40, P1445, DOI 10.3174/ajnr.A6153
   Lim M, 2018, NAT REV CLIN ONCOL, V15, P422, DOI 10.1038/s41571-018-0003-5
   Mangla R, 2011, AM J NEURORADIOL, V32, P1004, DOI 10.3174/ajnr.A2441
   Paulson ES, 2008, RADIOLOGY, V249, P601, DOI 10.1148/radiol.2492071659
   Paulson ES, 2016, TOMOGRAPHY, V2, P295, DOI 10.18383/j.tom.2016.00217
   Perez-Rodriguez J, 2009, RADIOLOGY, V250, P371, DOI 10.1148/radiol.2502080498
   Pons-Escoda A, 2022, AM J NEURORADIOL, V43, P1567, DOI 10.3174/ajnr.A7668
   Pons-Escoda A, 2020, AM J NEURORADIOL, V41, P1816, DOI 10.3174/ajnr.A6761
   Pons-Escoda A, 2022, EUR RADIOL, V32, P3705, DOI 10.1007/s00330-021-08498-1
   Quarles CC, 2019, NEUROIMAGE, V187, P32, DOI 10.1016/j.neuroimage.2018.04.069
   Schmainda KM, 2004, AM J NEURORADIOL, V25, P1524
   Scordo M, 2021, JAMA ONCOL, V7, P993, DOI 10.1001/jamaoncol.2021.1074
   Song SS, 2021, EUR RADIOL, V31, P4087, DOI 10.1007/s00330-020-07470-9
   Sourbron S, 2010, EUR J RADIOL, V76, P304, DOI 10.1016/j.ejrad.2010.02.017
   Steidl E, 2021, EUR J NUCL MED MOL I, V48, P1956, DOI 10.1007/s00259-020-05114-0
   Welker K, 2015, AM J NEURORADIOL, V36, pE41, DOI 10.3174/ajnr.A4341
   Xing Z, 2014, CLIN NEURORADIOL, V24, P329, DOI 10.1007/s00062-013-0255-5
   Xing Z, 2020, BMC MED IMAGING, V20, DOI 10.1186/s12880-020-00462-7
NR 33
TC 1
Z9 1
U1 4
U2 5
PU SPRINGER
PI NEW YORK
PA ONE NEW YORK PLAZA, SUITE 4600, NEW YORK, NY, UNITED STATES
SN 0938-7994
EI 1432-1084
J9 EUR RADIOL
JI Eur. Radiol.
PD DEC
PY 2023
VL 33
IS 12
BP 8800
EP 8808
DI 10.1007/s00330-023-09917-1
EA JUL 2023
PG 9
WC Radiology, Nuclear Medicine & Medical Imaging
WE Science Citation Index Expanded (SCI-EXPANDED)
SC Radiology, Nuclear Medicine & Medical Imaging
GA CF2U2
UT WOS:001029652100006
PM 37439934
DA 2024-06-03
ER

PT J
AU Cai, MT
   Lai, QL
   Tang, JL
   Du, BQ
   Shen, CH
   Zhang, YX
   Guo, Y
AF Cai, Meng-Ting
   Lai, Qi-Lun
   Tang, Jin-Long
   Du, Bing-Qing
   Shen, Chun-Hong
   Zhang, Yin-Xi
   Guo, Yi
TI Myelin oligodendrocyte glycoprotein antibody-associated disease
   preceding primary central nervous system lymphoma: causality or
   coincidence?
SO NEUROLOGICAL SCIENCES
LA English
DT Article
DE Myelin oligodendrocyte glycoprotein antibody-associated disease; Primary
   central nervous system lymphoma; Sentinel lesions; Biopsy
AB IntroductionPrimarycentral nervous system lymphoma (PCNSL) is a rare extranodal lymphomatous malignancy that affects the brain, spinal cord, leptomeninges, or eyes, in the absence of systemic diffusion. Myelin oligodendrocyte glycoprotein (MOG) antibody-associated disease (MOGAD) is a newly identified benign immune-mediated CNS inflammatory disorder with specific anti-MOG antibody seropositivity. These two seemingly unrelated nosological entities both have abundant clinical and radiological manifestations, and whether there is a potential link between them is unclear.Case reportWe describe a 49-year-old man who presented progressive headache, dizziness, and unsteady gait with multifocal scattered T2 hyperintensities with contrast enhancement. The serum anti-MOG antibody test was positive, and a brain biopsy showed inflammatory infiltration. Initially, he was diagnosed with MOGAD and his condition improved after corticosteroid therapy. The patient relapsed with exacerbation of symptoms and neuroimaging showed new mass-forming lesions four months later. A second brain biopsy confirmed PCNSL.DiscussionThis is the first report of histologically confirmed successive MOGAD and PCNSL. Our case broadens the phenotypic spectrum of sentinel lesions in PCNSL. Though rare, PCNSL should be considered in patients diagnosed with benign CNS inflammatory disorder and responding well to steroid treatment when their clinical symptoms worsen and the imaging deteriorates. A timely biopsy is critical for accurate diagnosis and appropriate therapy.
C1 [Cai, Meng-Ting; Du, Bing-Qing; Shen, Chun-Hong; Zhang, Yin-Xi; Guo, Yi] Zhejiang Univ, Affiliated Hosp 2, Sch Med, Dept Neurol, Hangzhou 310009, Peoples R China.
   [Lai, Qi-Lun] Zhejiang Hosp, Dept Neurol, Hangzhou 310013, Peoples R China.
   [Tang, Jin-Long] Zhejiang Univ, Affiliated Hosp 2, Sch Med, Dept Pathol, Hangzhou 310009, Peoples R China.
   [Guo, Yi] Zhejiang Univ, Affiliated Hosp 2, Sch Med, Dept Gen Practice & Int Med, Hangzhou 310009, Peoples R China.
C3 Zhejiang University; Zhejiang University; Zhejiang University
RP Zhang, YX; Guo, Y (corresponding author), Zhejiang Univ, Affiliated Hosp 2, Sch Med, Dept Neurol, Hangzhou 310009, Peoples R China.; Guo, Y (corresponding author), Zhejiang Univ, Affiliated Hosp 2, Sch Med, Dept Gen Practice & Int Med, Hangzhou 310009, Peoples R China.
EM
RI zhou, yuwei/KHD-4127-2024; li, yuanfang/KHV-1697-2024; guo,
   yi/KHC-4669-2024; feng, yue/KHV-4687-2024; Tang, jinlong/C-4203-2012;
   Zhang, xiaohui/KEE-5747-2024; XIN, WANG/KGK-5385-2024; Zhang,
   yuxuan/JXM-9935-2024; lin, lin/KFB-9548-2024; liu, feng/KCL-0778-2024
OI Guo, Yi/0000-0002-9128-4779
CR Banks SA, 2021, J NEUROL NEUROSUR PS, V92, P384, DOI 10.1136/jnnp-2020-325121
   Banwell B, 2023, LANCET NEUROL, V22, P268, DOI 10.1016/S1474-4422(22)00431-8
   Bataille B, 2000, J NEUROSURG, V92, P261, DOI 10.3171/jns.2000.92.2.0261
   Deckert-Schlüter M, 1998, ACTA NEUROPATHOL, V96, P157, DOI 10.1007/s004010050876
   Grommes C, 2017, J CLIN ONCOL, V35, P2410, DOI 10.1200/JCO.2017.72.7602
   Javier R, 2018, DIAGN PATHOL, V13, DOI 10.1186/s13000-018-0717-9
   Kalus S, 2016, J CLIN NEUROSCI, V24, P146, DOI 10.1016/j.jocn.2015.07.013
   Kvarta MD, 2016, BMC NEUROL, V16, DOI 10.1186/s12883-016-0596-1
   Lu JQ, 2016, WORLD NEUROSURG, V88, DOI 10.1016/j.wneu.2015.11.099
   Nabavizadeh SA, 2016, HEMATOL ONCOL CLIN N, V30, P799, DOI 10.1016/j.hoc.2016.03.005
   Qiu TM, 2021, WORLD NEUROSURG, V145, P172, DOI 10.1016/j.wneu.2020.09.064
   Uzura Y, 2022, J NEUROIMMUNOL, V365, DOI 10.1016/j.jneuroim.2022.577823
   Yamamoto J, 2014, ONCOL LETT, V7, P1835, DOI 10.3892/ol.2014.2033
   Zhang YX, 2022, FRONT NEUROL, V13, DOI 10.3389/fneur.2022.844155
NR 14
TC 1
Z9 1
U1 0
U2 1
PU SPRINGER-VERLAG ITALIA SRL
PI MILAN
PA VIA DECEMBRIO, 28, MILAN, 20137, ITALY
SN 1590-1874
EI 1590-3478
J9 NEUROL SCI
JI Neurol. Sci.
PD OCT
PY 2023
VL 44
IS 10
BP 3711
EP 3715
DI 10.1007/s10072-023-06919-1
EA JUN 2023
PG 5
WC Clinical Neurology; Neurosciences
WE Science Citation Index Expanded (SCI-EXPANDED)
SC Neurosciences & Neurology
GA U5IS9
UT WOS:001020243100001
PM 37389732
DA 2024-06-03
ER

PT J
AU Schrum, DP
   Moorman, MT
   Li, ZG
   Dillon, M
   Peters, KB
   McKinney, M
   Patel, MP
AF Schrum, Daniel P.
   Moorman, Meredith T.
   Li, Zhiguo
   Dillon, Mairead
   Peters, Katherine B.
   McKinney, Matthew
   Patel, Mallika P.
TI Comparison of differing dose levels of methotrexate for patients with
   primary central nervous system lymphoma
SO JOURNAL OF ONCOLOGY PHARMACY PRACTICE
LA English
DT Article
DE Methotrexate; lymphoma; CNS lymphoma; MTR; MATRix
ID CHEMOTHERAPY; CYCLOPHOSPHAMIDE; VINCRISTINE; DOXORUBICIN; RITUXIMAB;
   REGIMEN; TRIAL
AB Introduction It has long been established that high-dose methotrexate is an essential part of therapy for primary central nervous system lymphoma. When regimens utilizing high-dose methotrexate were first studied, a dose of 8 g/m(2) was used. More recently, reduced dosing strategies have been studied and adopted in attempts to reduce rates of adverse events. Studies utilizing 3.5 g/m(2) of methotrexate have shown promising outcomes and improved rates of adverse events but there have never been any randomized head-to-head studies of differing dose levels of high-dose methotrexate. The purpose of this study was to compare efficacy and safety of different dosing strategies of high-dose methotrexate (HD-MTX) for primary central nervous system lymphoma (PCNSL). Methods This single center retrospective review was conducted between 07/01/2013 to 6/3/2020. The patient population was separated into two arms based upon dose of methotrexate. The high intensity (HiHD) arm was defined as patients who received doses > 3.5 g/m(2), while the low intensity (LiHD) arm received <= 3.5 g/m(2). The primary endpoint was overall response rate (ORR) and secondary endpoints include efficacy via 2-year overall survival (OS), progression to transplant, and utilization of consolidation or salvage therapy. Safety was assessed through monitoring of relevant laboratory studies. Results A total of 92 patients were included in this analysis. Baseline demographics were similar between groups, with the LiHD group trending toward older age. There were 78 patients eligible for assessment for ORR; there was no significant difference between the two groups (42.0% LiHD vs. 44.4% HiHD; p = 1.0). Rates of OS, progression to transplant and progression to consolidation chemotherapy were not different between groups. There were statistically significantly higher rates of renal and/or hepatic dysfunction with the first dose in the HiHD group compared with the LiHD group (11.5% LiHD vs. 64.3% HiHD; p <= 0.01). Conclusions In this PCNSL patient cohort, there is no difference in terms of efficacy between HiHD LiHD methotrexate, but patients in the HiHD group had higher rates of renal and hepatic dysfunction. Limitations include small sample size and disparity between group sizes.
C1 [Schrum, Daniel P.; Moorman, Meredith T.; Patel, Mallika P.] Duke Univ, Med Ctr, Dept Pharm, Durham, NC USA.
   [Li, Zhiguo; Dillon, Mairead] Duke Univ, Sch Med, Dept Biostat & Informat, Durham, NC USA.
   [Peters, Katherine B.] Duke Univ, Med Ctr, Dept Neurol, Durham, NC USA.
   [McKinney, Matthew] Duke Univ Med Ctr, Dept Med, Div Hematol Malignancies & Cellular Therapies, Durham, NC USA.
   [Schrum, Daniel P.] Duke Univ Hosp, Dept Pharm, 2301 Erwin Rd, Durham, NC 27710 USA.
C3 Duke University; Duke University; Duke University; Duke University; Duke
   University
RP Schrum, DP (corresponding author), Duke Univ Hosp, Dept Pharm, 2301 Erwin Rd, Durham, NC 27710 USA.
EM
RI McKinney, Matthew/JPK-0745-2023
OI Schrum, Daniel/0000-0002-9779-2723
CR BORSI JD, 1987, CANCER-AM CANCER SOC, V60, P5, DOI 10.1002/1097-0142(19870701)60:1<5::AID-CNCR2820600103>3.0.CO;2-D
   Camilleri-Broet S, 1998, AM J CLIN PATHOL, V110, P607, DOI 10.1093/ajcp/110.5.607
   Dalia S, 2013, BLOOD, V122, DOI 10.1182/blood.V122.21.3040.3040
   Ferreri AJM, 2016, LANCET HAEMATOL, V3, pE217, DOI 10.1016/S2352-3026(16)00036-3
   Glass J, 2016, J CLIN ONCOL, V34, P1620, DOI 10.1200/JCO.2015.64.8634
   Grommes C, 2017, J CLIN ONCOL, V35, P2410, DOI 10.1200/JCO.2017.72.7602
   Harris PA, 2009, J BIOMED INFORM, V42, P377, DOI 10.1016/j.jbi.2008.08.010
   Li Q, 2021, CANCER MANAG RES, V13, P6115, DOI 10.2147/CMAR.S322467
   Mead GM, 2000, CANCER, V89, P1359, DOI 10.1002/1097-0142(20000915)89:6<1359::AID-CNCR21>3.0.CO;2-9
   NELSON DF, 1992, INT J RADIAT ONCOL, V23, P9, DOI 10.1016/0360-3016(92)90538-S
   Pels H, 2003, J CLIN ONCOL, V21, P4489, DOI 10.1200/JCO.2003.04.056
   Rubenstein JL, 2013, J CLIN ONCOL, V31, P3061, DOI 10.1200/JCO.2012.46.9957
   Ruhstaller TW, 2000, ANN ONCOL, V11, P374, DOI 10.1023/A:1008371602708
   Schultz C, 1996, J CLIN ONCOL, V14, P556, DOI 10.1200/JCO.1996.14.2.556
   SHAPIRO WR, 1975, NEW ENGL J MED, V293, P161, DOI 10.1056/NEJM197507242930402
   Shibamoto Y, 2005, INT J RADIAT ONCOL, V62, P809, DOI 10.1016/j.ijrobp.2004.12.043
NR 16
TC 1
Z9 1
U1 0
U2 0
PU SAGE PUBLICATIONS LTD
PI LONDON
PA 1 OLIVERS YARD, 55 CITY ROAD, LONDON EC1Y 1SP, ENGLAND
SN 1078-1552
EI 1477-092X
J9 J ONCOL PHARM PRACT
JI J. Oncol. Pharm. Pract.
PD APR
PY 2024
VL 30
IS 3
BP 513
EP 518
DI 10.1177/10781552231176754
EA MAY 2023
PG 6
WC Oncology; Pharmacology & Pharmacy
WE Science Citation Index Expanded (SCI-EXPANDED)
SC Oncology; Pharmacology & Pharmacy
GA OU0L4
UT WOS:001001171300001
PM 37198894
DA 2024-06-03
ER

PT J
AU Baskaran, AB
   Dixit, KS
   Sonabend, AW
   Horbinski, CM
   Abutalib, SA
   Lukas, RV
AF Baskaran, Archit B.
   Dixit, Karan S.
   Sonabend, Adam W.
   Horbinski, Craig M.
   Abutalib, Syed Ali
   Lukas, Rimas V.
TI From the ER to the OR: Initial Evaluation of Primary Central Nervous
   System Lymphoma
SO NEUROLOGIST
LA English
DT Review
DE primary CNS lymphoma; review; neuro-oncology; neurosurgery
ID PRIMARY CNS LYMPHOMA; SPINAL-CORD INVOLVEMENT; BARR-VIRUS DNA;
   IMMUNOCOMPETENT PATIENTS; CEREBROSPINAL-FLUID; MENINGEAL DISSEMINATION;
   RADIOGRAPHIC FINDINGS; MALIGNANT-LYMPHOMA; DIAGNOSIS; AIDS
AB Background:Primary central nervous system lymphoma (PCNSL) is a rare, often curable neoplasm, often initially presenting in acute care settings by nonneuroscience specialized physicians. Delays in the recognition of specific imaging findings, lack of appropriate specialist consultation, and urgent incorrect medication administration can delay necessary diagnosis and treatment. Review Summary:In this paper, the reader is moved quickly from the initial presentation to the diagnostic surgical intervention for PCNSL in a manner analogous to the experience of clinicians in the frontline setting. We review the clinical presentation of PCNSL, its radiographic features, the effect of prebiopsy steroids, and the role of a biopsy in the diagnosis. In addition, this paper revisits the role of surgical resection for PCNSL and investigational diagnostic studies for PCNSL. Conclusion:PCNSL is a rare tumor that is associated with high morbidity and mortality. However, with appropriate identification of clinical signs, symptoms, and key radiographic findings, the early suspicion of PCNSL can lead to steroid avoidance and timely biopsy for rapid administration of the potentially curative chemoimmunotherapy. Surgical resection presents the potential for improving outcomes for patients with PCNSL, however, this remains controversial. Further research into PCNSL presents the opportunity for better outcomes and longer livelihoods for patients.
C1 [Baskaran, Archit B.] Univ Chicago, Feinberg Sch Med, Chicago, IL USA.
   [Dixit, Karan S.] Univ Chicago, Dept Neurol, Chicago, IL USA.
   [Sonabend, Adam W.] Univ Chicago, Dept Neurol Surg, Chicago, IL USA.
   [Horbinski, Craig M.] Univ Chicago, Dept Pathol, Neuropathol Div, Chicago, IL USA.
   [Horbinski, Craig M.] Univ Chicago, Dept Neurosurg, Neuropathol Div, Chicago, IL USA.
   [Lukas, Rimas V.] Univ Chicago, Dept Neurol, Neurooncol Div, Chicago, IL USA.
   [Baskaran, Archit B.] Univ Chicago, Univ Chicago Med, Chicago, IL USA.
   [Abutalib, Syed Ali] Canc Treatment Ctr Amer, Cellular Therapy & Clin Apheresis Programs, Hematol, BMT, Zion, IL USA.
   [Abutalib, Syed Ali] Rosalind Franklin Univ Med & Sci, N Chicago, IL USA.
   [Lukas, Rimas V.] Northwestern Univ, Lou & Jean Malnati Brain Tumor Inst, Dept Neurol, Chicago, IL 60208 USA.
C3 Northwestern University; Feinberg School of Medicine; University of
   Chicago; University of Chicago; University of Chicago; University of
   Chicago; University of Chicago; University of Chicago; University of
   Chicago; Rosalind Franklin University of Medicine & Science;
   Northwestern University
RP Lukas, RV (corresponding author), Northwestern Univ, Lou & Jean Malnati Brain Tumor Inst, Dept Neurol, Chicago, IL 60208 USA.
EM;
  ;
  
RI Lukas, Rimas/JZD-3282-2024
OI Baskaran, Archit/0000-0002-1729-0142
CR Antinori A, 1999, J CLIN ONCOL, V17, P554, DOI 10.1200/JCO.1999.17.2.554
   Baraniskin Alexander, 2014, CNS Oncol, V3, P77, DOI 10.2217/cns.13.63
   Baraniskin A, 2011, BLOOD, V117, P3140, DOI 10.1182/blood-2010-09-308684
   Bataille B, 2000, J NEUROSURG, V92, P261, DOI 10.3171/jns.2000.92.2.0261
   Bellinzona M, 2005, EJSO, V31, P100, DOI 10.1016/j.ejso.2004.10.002
   Berger JR, 2003, AM J NEURORADIOL, V24, P554
   Bobillo S, 2021, HAEMATOLOGICA, V106, P513, DOI 10.3324/haematol.2019.241208
   Bromberg JEC, 2002, NEUROLOGY, V59, P762, DOI 10.1212/WNL.59.5.762
   Brück W, 2013, PATHOLOGE, V34, P186, DOI 10.1007/s00292-013-1742-9
   Bruno A, 2014, ONCOTARGET, V5, P5065, DOI 10.18632/oncotarget.2080
   Bullis CL, 2020, J CLIN NEUROSCI, V72, P287, DOI 10.1016/j.jocn.2019.10.010
   Camacho DLA, 2003, AM J NEURORADIOL, V24, P633
   Chiavazza C, 2018, BIOMED RES INT, V2018, DOI 10.1155/2018/3606970
   Choi YL, 2006, CLIN NEUROPATHOL, V25, P29
   CINQUE P, 1993, LANCET, V342, P398, DOI 10.1016/0140-6736(93)92814-A
   Cote TR, 1996, J NATL CANCER I, V88, P675, DOI 10.1093/jnci/88.10.675
   Coulon A, 2002, EUR RADIOL, V12, P329, DOI 10.1007/s003300101037
   DeAngelis LM, 2005, CANCER INVEST, V23, P145, DOI 10.1081/CNV-200050458
   Elavarasi A, 2018, J CLIN NEUROSCI, V47, P145, DOI 10.1016/j.jocn.2017.10.027
   Ferreri AJM, 2002, NEUROLOGY, V58, P1513, DOI 10.1212/WNL.58.10.1513
   Ferreri AJM, 2021, BRIT J HAEMATOL, V193, P497, DOI 10.1111/bjh.17357
   FINE HA, 1993, ANN INTERN MED, V119, P1093, DOI 10.7326/0003-4819-119-11-199312010-00007
   Fontanilles M, 2017, ONCOTARGET, V8, P48157, DOI 10.18632/oncotarget.18325
   Fox J, 2019, J NEUROL SCI, V400, P34, DOI 10.1016/j.jns.2019.03.011
   Fukumura K, 2016, ACTA NEUROPATHOL, V131, P865, DOI 10.1007/s00401-016-1536-2
   GEPPERT M, 1990, ACTA NEUROPATHOL, V80, P629, DOI 10.1007/BF00307631
   Goyal P, 2017, QUANT IMAG MED SURG, V7, P511, DOI 10.21037/qims.2017.09.03
   Gray BR, 2020, SEMIN ULTRASOUND CT, V41, P541, DOI 10.1053/j.sult.2020.08.008
   Grommes C, 2019, NEURO-ONCOLOGY, V21, P296, DOI 10.1093/neuonc/noy192
   Grommes C, 2017, J CLIN ONCOL, V35, P2410, DOI 10.1200/JCO.2017.72.7602
   Grossman RI., 1994, NEURORADIOLOGY REQUI, P186
   Gulley ML, 2008, J MOL DIAGN, V10, P279, DOI 10.2353/jmoldx.2008.080023
   Haldorsen IS, 2011, AM J NEURORADIOL, V32, P984, DOI 10.3174/ajnr.A2171
   Haldorsen IS, 2009, AM J NEURORADIOL, V30, P744, DOI 10.3174/ajnr.A1447
   Haldorsen IS, 2008, BMC CANCER, V8, DOI 10.1186/1471-2407-8-225
   Harting I, 2003, NEUROSCI LETT, V342, P163, DOI 10.1016/S0304-3940(03)00272-6
   HENRY JM, 1974, CANCER, V34, P1293, DOI 10.1002/1097-0142(197410)34:4<1293::AID-CNCR2820340441>3.0.CO;2-P
   Holdhoff M, 2020, J NATL COMPR CANC NE, V18, P1571, DOI 10.6004/jnccn.2020.7667
   Houillier C, 2020, NEUROLOGY, V94, pE1027, DOI 10.1212/WNL.0000000000008900
   Huang WY, 2016, AM J NEURORADIOL, V37, P2010, DOI 10.3174/ajnr.A4867
   JACK CR, 1986, AM J ROENTGENOL, V146, P271, DOI 10.2214/ajr.146.2.271
   Jahnke K, 2005, AM J NEURORADIOL, V26, P2446
   Kawasaki K, 2002, NEUROPATHOLOGY, V22, P13, DOI 10.1046/j.0919-6544.2002.00421.x
   Kiewe P, 2010, NEURO-ONCOLOGY, V12, P409, DOI 10.1093/neuonc/nop053
   Kluin PM, 2017, WHO CLASSIFICATION T
   Koeller KK, 1997, RADIOGRAPHICS, V17, P1497, DOI 10.1148/radiographics.17.6.9397461
   Korfel A, 2012, ANN ONCOL, V23, P2374, DOI 10.1093/annonc/mdr627
   Korfel A, 2013, NAT REV NEUROL, V9, P317, DOI 10.1038/nrneurol.2013.83
   Krebs Simone, 2021, Ann Lymphoma, V5, DOI 10.21037/aol-20-52
   Labak CM, 2019, WORLD NEUROSURG, V126, pE1436, DOI 10.1016/j.wneu.2019.02.252
   Lu SS, 2014, AM J NEURORADIOL, V35, P270, DOI 10.3174/ajnr.A3677
   Lu SS, 2015, AM J ROENTGENOL, V204, P827, DOI 10.2214/AJR.14.12677
   Lukas RV, 2018, ONCOLOGY-NY, V32, P17
   Maeyama M, 2020, CANCER MED-US, V9, P4114, DOI 10.1002/cam4.3048
   Malone H, 2015, WORLD NEUROSURG, V84, P1084, DOI 10.1016/j.wneu.2015.05.025
   Manoj N, 2014, NEUROL INDIA, V62, P19, DOI 10.4103/0028-3886.128272
   Mansour A, 2014, CANCER IMAGING, V14, DOI 10.1186/1470-7330-14-22
   Mao XH, 2014, NEUROL SCI, V35, P233, DOI 10.1007/s10072-013-1491-9
   Masouris I, 2021, FRONT NEUROL, V12, DOI 10.3389/fneur.2021.654543
   Mathew BS, 2006, CANCER-AM CANCER SOC, V106, P383, DOI 10.1002/cncr.21583
   Mendelow AD, 2005, LANCET, V365, P387
   MILLER DC, 1994, CANCER-AM CANCER SOC, V74, P1383, DOI 10.1002/1097-0142(19940815)74:4<1383::AID-CNCR2820740432>3.0.CO;2-1
   Mohler AC., 2021, EXPERT REV GASTROENT, P417
   Moini J., 2021, Epidemiology of Brain and Spinal Tumors, DOI DOI 10.1016/C2019-0-03764-8
   Morell AA, 2019, NEURO-ONCOL PRACT, V6, P415, DOI 10.1093/nop/npz015
   Mulazzani M, 2019, J HEMATOL ONCOL, V12, DOI 10.1186/s13045-019-0796-4
   Murase S, 2000, NEUROL RES, V22, P434
   O'Neill BP, 2013, AM J HEMATOL, V88, P997, DOI 10.1002/ajh.23551
   Patrick LB, 2015, CURR ONCOL REP, V17, DOI 10.1007/s11912-015-0483-8
   Pfefferkorn T, 2009, STROKE, V40, P3045, DOI 10.1161/STROKEAHA.109.550871
   Pirotte B, 1997, J NEURO-ONCOL, V32, P63, DOI 10.1023/A:1005733416571
   Porter AB, 2008, ANN NEUROL, V63, P662, DOI 10.1002/ana.21366
   Raco A, 2003, NEUROSURGERY, V53, P1061, DOI 10.1227/01.NEU.0000088766.34559.3E
   Rae AI, 2019, NEUROSURGERY, V84, P935, DOI 10.1093/neuros/nyy096
   Raizer JJ, 2005, J NEURO-ONCOL, V71, P173, DOI 10.1007/s11060-004-1360-8
   Ramachandran Rajoo, 2014, Radiol Case Rep, V9, pe00031, DOI 10.2484/rcr.v9i1.908
   Reni M, 1997, ANN ONCOL, V8, P227, DOI 10.1023/A:1008201717089
   Rubenstein JL, 2013, BLOOD, V121, P4740, DOI 10.1182/blood-2013-01-476333
   Sasayama T, 2012, NEURO-ONCOLOGY, V14, P368, DOI 10.1093/neuonc/nor203
   Saylor Deanna, 2018, Continuum (Minneap Minn), V24, P1397, DOI 10.1212/CON.0000000000000647
   Schabet M, 1999, J NEURO-ONCOL, V43, P199, DOI 10.1023/A:1006290032052
   Scheichel F, 2022, FRONT ONCOL, V12, DOI 10.3389/fonc.2022.884724
   Schob S, 2016, PLOS ONE, V11, DOI 10.1371/journal.pone.0161386
   Scott BJ, 2013, JAMA NEUROL, V70, P311, DOI 10.1001/jamaneurol.2013.606
   Sharma A, 2018, BLOOD, V132, DOI 10.1182/blood-2018-99-113690
   Song YC, 2016, SCI REP-UK, V6, DOI 10.1038/srep21389
   Strehlow F, 2016, J NEURO-ONCOL, V129, P165, DOI 10.1007/s11060-016-2162-5
   Sutherland T, 2012, J MED IMAG RADIAT ON, V56, P295, DOI 10.1111/j.1754-9485.2012.02366.x
   Thurnher MM, 2001, NEURORADIOLOGY, V43, P29, DOI 10.1007/s002340000480
   van Westrhenen A, 2018, BRIT J HAEMATOL, V182, P384, DOI 10.1111/bjh.15410
   Viaccoz A, 2015, NEURO-ONCOLOGY, V17, P1497, DOI 10.1093/neuonc/nov092
   Visco C, 2020, CANCERS, V12, DOI 10.3390/cancers12102913
   Weller M, 2012, NEURO-ONCOLOGY, V14, P1481, DOI 10.1093/neuonc/nos159
   Yanagihara Theodore K., 2014, Applied radiation oncology, V12, P5, DOI DOI 10.37549/ARO1055
   Yim E, 2020, NEURO-ONCOLOGY, V22, P84
   Yousef AF, 2014, EGYPT J RADIOL NUC M, V45, P869, DOI 10.1016/j.ejrnm.2014.04.011
   Yun J., 2016, ARCH CANC RES, V4, P71, DOI 10.21767/2254-6081.100071
   Zhang D, 2010, KOREAN J RADIOL, V11, P269, DOI 10.3348/kjr.2010.11.3.269
   Zhou YY, 2018, NEOPLASIA, V20, P1059, DOI 10.1016/j.neo.2018.08.012
NR 99
TC 1
Z9 1
U1 0
U2 0
PU LIPPINCOTT WILLIAMS & WILKINS
PI PHILADELPHIA
PA TWO COMMERCE SQ, 2001 MARKET ST, PHILADELPHIA, PA 19103 USA
SN 1074-7931
EI 2331-2637
J9 NEUROLOGIST
JI Neurologist
PD MAY
PY 2023
VL 28
IS 3
BP 135
EP 142
DI 10.1097/NRL.0000000000000475
PG 8
WC Clinical Neurology
WE Science Citation Index Expanded (SCI-EXPANDED)
SC Neurosciences & Neurology
GA F3YW5
UT WOS:000981746000001
PM 37027168
DA 2024-06-03
ER

PT J
AU Velicu, MA
   Lavrador, JP
   Sibtain, N
   Vergani, F
   Bhangoo, R
   Gullan, R
   Ashkan, K
AF Velicu, Maria Alexandra
   Lavrador, Jose Pedro
   Sibtain, Naomi
   Vergani, Francesco
   Bhangoo, Ranjeev
   Gullan, Richard
   Ashkan, Keyoumars
TI Neurosurgical Management of Central Nervous System Lymphoma: Lessons
   Learnt from a Neuro-Oncology Multidisciplinary Team Approach
SO JOURNAL OF PERSONALIZED MEDICINE
LA English
DT Article
DE CNS lymphoma; DLBCL; PCNSL; lymphoma CNS relapse; extranodal lymphoma
ID B-CELL LYMPHOMA; SECONDARY CNS INVOLVEMENT; PROPHYLAXIS; OUTCOMES;
   STRATEGIES; THERAPY
AB Central nervous system lymphoma (CNSL) represents one of the most aggressive forms of extranodal lymphoma. The gold standard for CNSL diagnosis remains the stereotactic biopsy, with a limited role for cytoreductive surgery that has not been supported by historical data. Our study aims to provide a comprehensive overview of neurosurgery's role in the diagnosis of systemic relapsed and primary CNSL, with an emphasis on the impact on management and survival. This is a single center retrospective cohort study with data collected between August 2012 and August 2020, including patients referred with a potential diagnosis of CNSL to the local Neuro-oncology Multidisciplinary Team (MDT). The concordance between the MDT outcome and histopathological confirmation was assessed using diagnostic statistics. A Cox regression is used for overall survival (OS) risk factor analysis, and Kaplan-Meier statistics are performed for three prognostic models. The diagnosis of lymphoma is confirmed in all cases of relapsed CNSL, and in all but two patients who underwent neurosurgery. For the relapsed CNSL group, the highest positive predictive value (PPV) is found for an MDT outcome when lymphoma had been considered as single or topmost probable diagnosis. Neuro-oncology MDT has an important role in establishing the diagnosis in CNSL, not only to plan tissue diagnosis but also to stratify the surgical candidates. The MDT outcome based on history and imaging has good predictive value for cases where lymphoma is considered the most probable diagnosis, with the best prediction for cases of relapsed CNSL, questioning the need for invasive tissue diagnosis in the latter group.
C1 [Velicu, Maria Alexandra; Lavrador, Jose Pedro; Vergani, Francesco; Bhangoo, Ranjeev; Gullan, Richard; Ashkan, Keyoumars] Kings Coll Hosp NHS Fdn Trust, Dept Neurosurg, London SE5 9RS, England.
   [Sibtain, Naomi] Kings Coll Hosp NHS Fdn Trust, Dept Neuroradiol, London SE5 9RS, England.
C3 King's College Hospital NHS Foundation Trust; King's College Hospital
   NHS Foundation Trust
RP Velicu, MA (corresponding author), Kings Coll Hosp NHS Fdn Trust, Dept Neurosurg, London SE5 9RS, England.
EM
OI Lavrador, Jose Pedro/0000-0002-9518-111X
CR Armand M, 2019, AM J HEMATOL, V94, P1123, DOI 10.1002/ajh.25578
   Baek DW, 2020, BLOOD RES, V55, P99, DOI 10.5045/br.2020.2020032
   Baraniskin A, 2021, CANCERS, V13, DOI 10.3390/cancers13112665
   Bette S, 2016, ROFO-FORTSCHR RONTG, V188, P1134, DOI 10.1055/s-0042-115572
   Cai QQ, 2016, CHIN J CANCER, V35, DOI 10.1186/s40880-016-0150-y
   Cheng G, 2019, NEUROL SCI, V40, P535, DOI 10.1007/s10072-018-3669-7
   Cirillo M, 2020, CANCER TREAT REV, V91, DOI 10.1016/j.ctrv.2020.102106
   Cloney MB, 2017, J NEURO-ONCOL, V132, P189, DOI 10.1007/s11060-016-2358-8
   El-Galaly TC, 2018, EUR J CANCER, V93, P57, DOI 10.1016/j.ejca.2018.01.073
   Faqah A, 2021, JCO GLOB ONCOL, V7, P486, DOI 10.1200/GO.20.00422
   Ferreri AJM, 2003, J CLIN ONCOL, V21, P266, DOI 10.1200/JCO.2003.09.139
   Ferreri AJM, 2021, LANCET HAEMATOL, V8, pe110, DOI 10.1016/S2352-3026(20)30366-5
   Ferreri AJM, 2015, BRIT J HAEMATOL, V168, P654, DOI 10.1111/bjh.13194
   Fox CP, 2019, BRIT J HAEMATOL, V184, P348, DOI 10.1111/bjh.15661
   Giamouriadis A, 2019, ACTA NEUROCHIR, V161, P2587, DOI 10.1007/s00701-019-04093-y
   Haldorsen IS, 2011, AM J NEURORADIOL, V32, P984, DOI 10.3174/ajnr.A2171
   Hill QA, 2006, BLOOD REV, V20, P319, DOI 10.1016/j.blre.2006.02.001
   Kuitunen H, 2020, ANN HEMATOL, V99, P1823, DOI 10.1007/s00277-020-04140-0
   Lee IH, 2010, EUR J RADIOL, V76, P48, DOI 10.1016/j.ejrad.2009.05.013
   Liu CJ, 2020, CANCER MED-US, V9, P2134, DOI 10.1002/cam4.2872
   Mansour A, 2014, CANCER IMAGING, V14, DOI 10.1186/1470-7330-14-22
   Marenco-Hillembrand L, 2018, FRONT ONCOL, V8, DOI 10.3389/fonc.2018.00558
   Mohammed R, 2019, J BLOOD MED, V10, P71, DOI 10.2147/JBM.S190784
   Narita Y, 2021, NEURO-ONCOLOGY, V23, P122, DOI 10.1093/neuonc/noaa145
   Nder E., 2015, INT J CLIN EXP PATHO, V8, P7798
   Ollila TA, 2018, CURR TREAT OPTION ON, V19, DOI 10.1007/s11864-018-0555-8
   Partovi S, 2014, BRIT J RADIOL, V87, DOI 10.1259/bjr.20130684
   Poynton Edward, 2021, EJHaem, V2, P616, DOI 10.1002/jha2.212
   Qualls D, 2019, HAEMATOLOGICA, V104, P25, DOI 10.3324/haematol.2018.195834
   Rae AI, 2019, NEUROSURGERY, V84, P935, DOI 10.1093/neuros/nyy096
   Rossi D, 2019, HAEMATOLOGICA, V104, P648, DOI 10.3324/haematol.2018.206177
   Tang LJ, 2017, INT J OPHTHALMOL-CHI, V10, P1301, DOI 10.18240/ijo.2017.08.19
   Yamagishi Y, 2021, CANCER SCI, V112, P4702, DOI 10.1111/cas.15133
   Yun J., 2016, ARCH CANC RES, V4, P71, DOI [10.21767/2254-6081.100071, DOI 10.21767/2254-6081.100071]
   Yun J, 2017, FRONT NEUROL, V8, DOI 10.3389/fneur.2017.00478
   Zacharia TT, 2008, J NEUROIMAGING, V18, P411, DOI 10.1111/j.1552-6569.2007.00231.x
   Zhang Y, 2020, CHINESE MED J-PEKING, V133, P1462, DOI 10.1097/CM9.0000000000000844
NR 37
TC 1
Z9 1
U1 1
U2 1
PU MDPI
PI BASEL
PA ST ALBAN-ANLAGE 66, CH-4052 BASEL, SWITZERLAND
EI 2075-4426
J9 J PERS MED
JI J. Pers. Med.
PD APR 30
PY 2023
VL 13
IS 5
AR 783
DI 10.3390/jpm13050783
PG 14
WC Health Care Sciences & Services; Medicine, General & Internal
WE Science Citation Index Expanded (SCI-EXPANDED)
SC Health Care Sciences & Services; General & Internal Medicine
GA L0FX9
UT WOS:001020109800001
PM 37240953
OA gold, Green Published
DA 2024-06-03
ER

PT J
AU Ma, L
   Gong, Q
AF Ma, Le
   Gong, Qiang
TI Recent advances and challenges in primary central nervous system
   lymphoma: a narrative review
SO TRANSLATIONAL CANCER RESEARCH
LA English
DT Review
DE Primary central nervous system lymphoma (PCNSL); diagnosis; treatment
ID PRIMARY CNS LYMPHOMA; HIGH-DOSE METHOTREXATE; B-CELL LYMPHOMA;
   WHOLE-BRAIN RADIOTHERAPY; NON-HODGKINS-LYMPHOMA; INTERNATIONAL
   EXTRANODAL LYMPHOMA; PRIMARY VITREORETINAL LYMPHOMA; PHASE-II;
   ELDERLY-PATIENTS; LEPTOMENINGEAL DISEASE
AB Background and Objective: Primary central nervous system lymphoma (PCNSL) is a rare and highly invasive non-Hodgkin lymphoma that is challenging to diagnose and treat. It is typically confined to the brain, spinal cord, and eyes. The diagnosis of PCNSL lacks specificity, and the misdiagnosis and missed diagnosis rates of PCNSL are high. Traditional treatments for PCNSL, such as surgery, whole-brain radiation therapy, high-dose methotrexate-based chemotherapy, and rituximab (RTX), have been associated with higher initial remission rates. However, the duration of any remission is short, the recurrence rate is high, and treatment-related neurotoxicity is strong, which are challenges for medical researchers. This review provides an overview of and perspectives on the diagnosis, treatment, and evaluation of patients with PCNSL.
   Methods: The PubMed database was searched to retrieve articles published from January 1, 1991, to June 2, 2022 using the following Medical Subject Headings (MeSH) terms: "Primary central nervous system lymphoma" and "clinical trial". The American Society of Clinical Oncology and the National Comprehensive Cancer Network guidelines were also reviewed to obtain additional information. The search was limited to articles published in English, German, and French. In total, 126 articles were deemed eligible for inclusion in this study.
   Key Content and Findings: In terms of the diagnosis of PCNSL, a combination of flow cytometry and cytology has been shown to improve the diagnostic accuracy of PCNSL. Additionally, interleukin 10 and chemokine C-X-C motif ligand 13 are promising biomarkers. In terms of the treatment of PCNSL, programmed death-1 (PD-1) blockage and chimeric antigen receptor T cell (CAR-T) therapy treatments have shown prospective efficacy, but more clinical trials need to be conducted to gather further evidence. We also reviewed and summarized prospective clinical trials on PCNSL.
   Conclusions: PCNSL is a rare and highly aggressive lymphoma. The treatment of PCNSL has progressed significantly, and while the survival of patients has improved, relapse and low long-term survival remain huge challenges. Continuous in-depth research is being conducted on new drug therapies and combination therapies for PCNSL. A combination of targeted drugs (e.g., ibrutinib, lenalidomide, and PD-1 monoclonal antibody) and traditional therapy represents the main research direction for future PCNSL treatments. CAR-T has also shown great potential in the treatment of PCNSL. With the development of these new diagnostic and therapeutic methods and further research into the molecular biology of PCNSL, patients with PCNSL should achieve a better prognosis.
C1 [Ma, Le; Gong, Qiang] Army Med Univ, Affiliated Hosp 1, Southwest Hosp, Dept Hematol, Gaotanyan Rd St, Chongqing 400038, Peoples R China.
C3 Army Medical University
RP Gong, Q (corresponding author), Army Med Univ, Affiliated Hosp 1, Southwest Hosp, Dept Hematol, Gaotanyan Rd St, Chongqing 400038, Peoples R China.
EM
CR Abramson JS, 2017, NEW ENGL J MED, V377, P783, DOI 10.1056/NEJMc1704610
   Abrey LE, 2006, J CLIN ONCOL, V24, P5711, DOI 10.1200/JCO.2006.08.2941
   Abrey LE, 2003, J CLIN ONCOL, V21, P4151, DOI 10.1200/JCO.2003.05.024
   Abrey LE, 2005, J CLIN ONCOL, V23, P5034, DOI 10.1200/JCO.2005.13.524
   Abrey LE, 2000, J CLIN ONCOL, V18, P3144, DOI 10.1200/JCO.2000.18.17.3144
   Adhikari N, 2018, J NEURO-ONCOL, V139, P153, DOI 10.1007/s11060-018-2856-y
   Ahmed G, 2021, BLOOD ADV, V5, P5626, DOI 10.1182/bloodadvances.2021005292
   Akhter A, 2015, J NEURO-ONCOL, V121, P289, DOI 10.1007/s11060-014-1655-3
   Algazi AP, 2009, NEUROTHERAPEUTICS, V6, P587, DOI 10.1016/j.nurt.2009.04.013
   Alnahhas I, 2019, CL LYMPH MYELOM LEUK, V19, pE129, DOI 10.1016/j.clml.2018.11.018
   Baraniskin A, 2012, BRIT J HAEMATOL, V156, P421, DOI 10.1111/j.1365-2141.2011.08928.x
   Baraniskin A, 2011, BLOOD, V117, P3140, DOI 10.1182/blood-2010-09-308684
   Bataille B, 2000, J NEUROSURG, V92, P261, DOI 10.3171/jns.2000.92.2.0261
   Batchelor T, 2003, J CLIN ONCOL, V21, P1044, DOI 10.1200/JCO.2003.03.036
   Batchelor TT, 2011, NEUROLOGY, V76, P929, DOI 10.1212/WNL.0b013e31820f2d94
   Bellinzona M, 2005, EJSO, V31, P100, DOI 10.1016/j.ejso.2004.10.002
   Braggio E, 2015, CLIN CANCER RES, V21, P3986, DOI 10.1158/1078-0432.CCR-14-2116
   Bromberg JEC, 2007, NEUROLOGY, V68, P1674, DOI 10.1212/01.wnl.0000261909.28915.83
   Bromberg JEC, 2019, LANCET ONCOL, V20, P216, DOI 10.1016/S1470-2045(18)30747-2
   Bruno A, 2014, ONCOTARGET, V5, P5065, DOI 10.18632/oncotarget.2080
   BUTLER RW, 1994, J CLIN ONCOL, V12, P2621, DOI 10.1200/JCO.1994.12.12.2621
   Camilleri-Broet S, 1998, AM J CLIN PATHOL, V110, P607, DOI 10.1093/ajcp/110.5.607
   Cazzola M, 2016, BLOOD, V127, P2361, DOI 10.1182/blood-2016-03-657379
   Chamberlain MC, 2010, NEURO-ONCOLOGY, V12, P736, DOI 10.1093/neuonc/noq011
   Chan CC, 2011, ONCOLOGIST, V16, P1589, DOI 10.1634/theoncologist.2011-0210
   Chapuy B, 2016, BLOOD, V127, P869, DOI 10.1182/blood-2015-10-673236
   Chen YS, 2019, ONCOL LETT, V18, P593, DOI 10.3892/ol.2019.10352
   Chiesa S, 2020, ANN HEMATOL, V99, P2367, DOI 10.1007/s00277-020-04220-1
   Coiffier B, 2002, NEW ENGL J MED, V346, P235, DOI 10.1056/NEJMoa011795
   Colombat P, 2006, BONE MARROW TRANSPL, V38, P417, DOI 10.1038/sj.bmt.1705452
   DEANGELIS LM, 1992, J CLIN ONCOL, V10, P635, DOI 10.1200/JCO.1992.10.4.635
   DeAngelis LM, 2002, J CLIN ONCOL, V20, P4643, DOI 10.1200/JCO.2002.11.013
   Dietrich J, 2020, ONCOLOGIST, V25, P747, DOI 10.1634/theoncologist.2020-0489
   Enting RH, 2004, NEUROLOGY, V63, P901, DOI 10.1212/01.WNL.0000137050.43114.42
   Ferreri AJM, 2001, ONCOLOGY-BASEL, V60, P134, DOI 10.1159/000055310
   Ferreri AJM, 2003, J CLIN ONCOL, V21, P266, DOI 10.1200/JCO.2003.09.139
   Ferreri AJM, 2022, LEUKEMIA, V36, P1870, DOI 10.1038/s41375-022-01582-5
   Ferreri AJM, 2021, BRIT J HAEMATOL, V193, P497, DOI 10.1111/bjh.17357
   Ferreri AJM, 2020, BLOOD ADV, V4, P3648, DOI 10.1182/bloodadvances.2020002270
   Ferreri AJM, 2019, BLOOD, V134, P252, DOI 10.1182/blood.2019000633
   Ferreri AJM, 2016, LANCET HAEMATOL, V3, pE217, DOI 10.1016/S2352-3026(16)00036-3
   Ferreri AJM, 2012, BEST PRACT RES CL HA, V25, P119, DOI 10.1016/j.beha.2011.12.001
   Ferreri AJM, 2009, LANCET, V374, P1512, DOI 10.1016/S0140-6736(09)61416-1
   Feugier P, 2004, ANN ONCOL, V15, P129, DOI 10.1093/annonc/mdh013
   Fox CP, 2021, BLOOD ADV, V5, P4073, DOI 10.1182/bloodadvances.2021004779
   Friedman HS, 2000, CLIN CANCER RES, V6, P2585
   Frigault MJ, 2022, BLOOD, V139, P2306, DOI 10.1182/blood.2021014738
   Frigault MJ, 2019, BLOOD, V134, P860, DOI 10.1182/blood.2019001694
   Fritsch K, 2017, LEUKEMIA, V31, P846, DOI 10.1038/leu.2016.334
   Gavrilovic IT, 2006, J CLIN ONCOL, V24, P4570, DOI 10.1200/JCO.2006.06.6910
   Ghesquieres H, 2019, ANN ONCOL, V30, P621, DOI 10.1093/annonc/mdz032
   GLASS J, 1994, J NEUROSURG, V81, P188, DOI 10.3171/jns.1994.81.2.0188
   Glass J, 2016, J CLIN ONCOL, V34, P1620, DOI 10.1200/JCO.2015.64.8634
   Grommes C, 2019, BLOOD, V133, P436, DOI 10.1182/blood-2018-09-875732
   Grommes C, 2017, J CLIN ONCOL, V35, P2410, DOI 10.1200/JCO.2017.72.7602
   Hattab EM, 2010, MODERN PATHOL, V23, P235, DOI 10.1038/modpathol.2009.164
   Hegde U, 2005, BLOOD, V105, P496, DOI 10.1182/blood-2004-05-1982
   Herrlinger U, 2005, ANN NEUROL, V57, P843, DOI 10.1002/ana.20495
   Hoang-Xuan K, 2020, BLOOD, V136, DOI 10.1182/blood-2020-141773
   Holdhoff M, 2014, NEUROLOGY, V83, P235, DOI 10.1212/WNL.0000000000000593
   Hottinger AF, 2007, NEUROLOGY, V69, P1178, DOI 10.1212/01.wnl.0000276986.19602.c1
   Houillier C, 2019, J CLIN ONCOL, V37, P823, DOI 10.1200/JCO.18.00306
   Ikeguchi R, 2018, MULT SCLER J, V24, P1212, DOI 10.1177/1352458517717804
   Illerhaus G, 2006, J CLIN ONCOL, V24, P3865, DOI 10.1200/JCO.2006.06.2117
   Illerhaus G, 2016, LANCET HAEMATOL, V3, pE388, DOI 10.1016/S2352-3026(16)30050-3
   Kasenda B, 2017, LEUKEMIA, V31, P2623, DOI 10.1038/leu.2017.170
   Vu K, 2019, BRIT J HAEMATOL, V186, P180, DOI 10.1111/bjh.15787
   Kobayashi H, 2019, J CHEMOTHERAPY, V31, P35, DOI 10.1080/1120009X.2018.1546984
   Kurzwelly D, 2010, J NEURO-ONCOL, V97, P389, DOI 10.1007/s11060-009-0032-0
   Labak CM, 2019, WORLD NEUROSURG, V126, pE1436, DOI 10.1016/j.wneu.2019.02.252
   Law SC, 2021, AM J TRANSPLANT, V21, P3465, DOI 10.1111/ajt.16628
   Lee DW, 2019, BIOL BLOOD MARROW TR, V25, P625, DOI 10.1016/j.bbmt.2018.12.758
   Lee JY, 2021, BLOOD RES, V56, P285, DOI 10.5045/br.2021.2021073
   Louis DN, 2007, ACTA NEUROPATHOL, V114, P97, DOI 10.1007/s00401-007-0243-4
   Makino K, 2012, J NEURO-ONCOL, V106, P155, DOI 10.1007/s11060-011-0652-z
   Mead GM, 2000, CANCER, V89, P1359, DOI 10.1002/1097-0142(20000915)89:6<1359::AID-CNCR21>3.0.CO;2-9
   Montesinos-Rongen M, 2011, ACTA NEUROPATHOL, V122, P791, DOI 10.1007/s00401-011-0891-2
   Morell AA, 2019, NEURO-ONCOL PRACT, V6, P415, DOI 10.1093/nop/npz015
   Morris PG, 2013, J CLIN ONCOL, V31, P3971, DOI 10.1200/JCO.2013.50.4910
   Nabavizadeh SA, 2016, HEMATOL ONCOL CLIN N, V30, P799, DOI 10.1016/j.hoc.2016.03.005
   Nakamura M, 2004, HISTOL HISTOPATHOL, V19, P211, DOI 10.14670/HH-19.211
   Nakamura T, 2016, NEUROPATH APPL NEURO, V42, P279, DOI 10.1111/nan.12259
   Narita Y, 2021, NEURO-ONCOLOGY, V23, P122, DOI 10.1093/neuonc/noaa145
   Nayak L, 2017, BLOOD, V129, P3071, DOI 10.1182/blood-2017-01-764209
   NELSON DF, 1992, INT J RADIAT ONCOL, V23, P9, DOI 10.1016/0360-3016(92)90538-S
   Neuwelt E, 2008, LANCET NEUROL, V7, P84, DOI 10.1016/S1474-4422(07)70326-5
   Nguyen-Them L, 2016, EUR J CANCER, V61, P69, DOI 10.1016/j.ejca.2016.03.080
   O'Brien P, 2000, J CLIN ONCOL, V18, P519, DOI 10.1200/JCO.2000.18.3.519
   O'Neill BP, 1999, INT J RADIAT ONCOL, V43, P559, DOI 10.1016/S0360-3016(98)00450-7
   Omuro A, 2015, LANCET HAEMATOL, V2, pE251, DOI 10.1016/S2352-3026(15)00074-5
   Omuro A, 2015, BLOOD, V125, P1403, DOI 10.1182/blood-2014-10-604561
   Önder E, 2015, INT J CLIN EXP PATHO, V8, P7798
   Onishi S, 2018, WORLD NEUROSURG, V119, pE890, DOI 10.1016/j.wneu.2018.07.291
   Ostrom QT, 2015, NEURO-ONCOLOGY, V16, P1, DOI [10.1093/neuonc/nou327, 10.1093/neuonc/nox158, 10.1093/neuonc/nou223, 10.1093/neuonc/nov189]
   Otani R, 2019, J CLIN NEUROSCI, V69, P26, DOI 10.1016/j.jocn.2019.08.083
   Palumbo A, 2008, LEUKEMIA, V22, P414, DOI 10.1038/sj.leu.2405062
   Pels H, 2003, J CLIN ONCOL, V21, P4489, DOI 10.1200/JCO.2003.04.056
   Poortmans PMP, 2003, J CLIN ONCOL, V21, P4483, DOI 10.1200/JCO.2003.03.108
   Prasad V, 2017, SEMIN ONCOL, V44, P132, DOI 10.1053/j.seminoncol.2017.06.007
   Qian LR, 2017, BLOOD RES, V52, P159, DOI 10.5045/br.2017.52.3.159
   Quijano S, 2009, J CLIN ONCOL, V27, P1462, DOI 10.1200/JCO.2008.17.7089
   Reni M, 1997, ANN ONCOL, V8, P227, DOI 10.1023/A:1008201717089
   Rubenstein JL, 2018, BLOOD ADV, V2, P1595, DOI 10.1182/bloodadvances.2017014845
   Rubenstein JL, 2013, J CLIN ONCOL, V31, P3061, DOI 10.1200/JCO.2012.46.9957
   Rubenstein JL, 2013, BLOOD, V121, P4740, DOI 10.1182/blood-2013-01-476333
   Sasagawa Y, 2015, J NEURO-ONCOL, V121, P177, DOI 10.1007/s11060-014-1622-z
   Schmitt AM, 2019, HEMATOL ONCOL, V37, P548, DOI 10.1002/hon.2666
   Schorb E, 2017, BONE MARROW TRANSPL, V52, P1113, DOI 10.1038/bmt.2017.23
   Schultz C, 1996, J CLIN ONCOL, V14, P556, DOI 10.1200/JCO.1996.14.2.556
   Scordo M, 2021, JAMA ONCOL, V7, P993, DOI 10.1001/jamaoncol.2021.1074
   Seidel S, 2020, NEUROLOGY, V95, pE3138, DOI 10.1212/WNL.0000000000010949
   Shah NN, 2021, BLOOD, V137, P2854
   Song Y, 2016, SCI REP-UK, V6, DOI 10.1038/srep38671
   Soussain C, 2019, EUR J CANCER, V117, P121, DOI 10.1016/j.ejca.2019.05.024
   Suwinski R, 2021, J THORAC DIS, V13, P3315, DOI 10.21037/jtd-2019-rbmlc-08
   Terziev D, 2018, EUR J HAEMATOL, V101, P115, DOI 10.1111/ejh.13072
   Thiel E, 2010, LANCET ONCOL, V11, P1036, DOI 10.1016/S1470-2045(10)70229-1
   Tun HW, 2018, BLOOD, V132, P2240, DOI 10.1182/blood-2018-02-835496
   van der Meulen M, 2021, J NEURO-ONCOL, V152, P357, DOI 10.1007/s11060-021-03708-8
   Vater I, 2015, LEUKEMIA, V29, P677, DOI 10.1038/leu.2014.264
   Wang A, 2022, LEUKEMIA LYMPHOMA, V63, P1348, DOI 10.1080/10428194.2021.2024818
   Wu JJ, 2018, J NEURO-ONCOL, V140, P427, DOI 10.1007/s11060-018-2970-x
   Yi C, 2019, NUKLEARMED-NUCL MED, V58, P272, DOI 10.1055/a-0835-5746
   Yuan XG, 2020, ANN HEMATOL, V99, P93, DOI 10.1007/s00277-019-03821-9
   Zhou D, 2017, ONCOTARGET, V8, P112384, DOI 10.18632/oncotarget.19736
NR 125
TC 1
Z9 1
U1 1
U2 9
PU AME PUBLISHING COMPANY
PI SHATIN
PA FLAT-RM C 16F, KINGS WING PLAZA 1, NO 3 KWAN ST, SHATIN, HONG KONG
   00000, PEOPLES R CHINA
SN 2218-676X
EI 2219-6803
J9 TRANSL CANCER RES
JI Transl. Cancer Res.
PD MAY
PY 2023
VL 12
IS 5
BP 1335
EP 1352
DI 10.21037/tcr-22-2341
EA APR 2023
PG 18
WC Oncology
WE Science Citation Index Expanded (SCI-EXPANDED)
SC Oncology
GA X4FJ4
UT WOS:000982465300001
PM 37304530
OA Green Published, gold
DA 2024-06-03
ER

PT J
AU Yang, Y
   Li, Q
   Ma, JJ
   Kang, H
   Lin, ZG
   Wang, Y
   Ma, Y
   Chen, BB
AF Yang, Yu
   Li, Qing
   Ma, Jingjing
   Kang, Hui
   Lin, Zhiguang
   Wang, Yang
   Ma, Yan
   Chen, Bobin
TI Radiotherapy or chemotherapy: a real-world study of the first-time
   relapsed and refractory primary central nervous system lymphoma
SO FRONTIERS IN ONCOLOGY
LA English
DT Article
DE relapsed or refractory primary central nervous system lymphoma;
   radiotherapy; chemotherapy; prognosis; salvage therapy
ID PRIMARY CNS LYMPHOMA; WHOLE-BRAIN RADIOTHERAPY; HIGH-DOSE METHOTREXATE;
   LONG-TERM SURVIVAL; B-CELL LYMPHOMA; PHASE-II; LENALIDOMIDE;
   COMBINATION; RECURRENT; RADIATION
AB BackgroundPrimary central nervous system lymphoma (PCNSL) is an uncommon variant of non-Hodgkin lymphoma (NHL) with high aggressiveness and poor prognosis. Although complete remission (CR) could be achieved with therapy, some patients remain refractory or recurrently with a worse response to salvage treatment and poor prognosis. No consensus on rescue therapy has been established currently. This study is aimed to evaluate the efficacy of radiotherapy or chemotherapy in first-time relapsed or refractory progressed PCNSL (R/R PCNSL) and analysis the prognostic factors, to explore differences between relapsed and refractory PCNSL. MethodsTotally 105 R/R PCNSL patients from Huashan Hospital between 1 January 2016 and 31 December 2020 were enrolled, underwent salvage radiotherapy or chemotherapy and received response assessments after each course. PFS1 was defined as the time from diagnosis to the first time of recurrence or refractory progression. Statistical analysis was performed with SPSS version 26.0. ResultsResponse and survival were analyzed over a 17.5months (median) follow-up. Compared to relapsed PCNSL (n = 42), refractory PCNSL (n = 63) had a shorter median PFS1 related to deep lesions. 82.4% of cases were discovered as the second relapse or progression. ORR and PFS were both higher in relapsed PCNSL than those in refractory PCNSL. ORR of radiotherapy in both relapsed and refractory PCNSL was higher than that of chemotherapy. Elevated CSF protein and ocular involvement were related to PFS and OS after recurrence respectively in relapsed PCNSL. Age >= 60y was unfavorable to OS-R (OS after recurrence or progression) in refractory PCNSL. ConclusionsOur results indicate that relapsed PCNSL responds well to inducing and salvage therapy and has a better prognosis compared to refractory PCNSL. Radiotherapy is effective for PCNSL after the first relapse or progression. Age, CSF protein level, and ocular involvement could be potential factors to predict prognosis.
C1 [Yang, Yu; Li, Qing; Ma, Jingjing; Kang, Hui; Lin, Zhiguang; Ma, Yan; Chen, Bobin] Fudan Univ, Huashan Hosp, Shanghai Med Coll, Dept Hematol, Shanghai, Peoples R China.
   [Wang, Yang] Fudan Univ, Huashan Hosp, Shanghai Med Coll, Dept Radiol, Shanghai, Peoples R China.
C3 Fudan University; Fudan University
RP Ma, Y; Chen, BB (corresponding author), Fudan Univ, Huashan Hosp, Shanghai Med Coll, Dept Hematol, Shanghai, Peoples R China.
EM
RI LI, LI/KCJ-5600-2024; Liu, yujing/JQI-7225-2023; li,
   wenjing/JMP-7498-2023; xu, chen/JNE-5010-2023; wang, xi/JNT-5162-2023;
   yan, yan/JVN-1800-2024; wang, KiKi/JFZ-3334-2023; chen,
   xu/JNT-3068-2023; Li, Wei/JLL-4365-2023; wu, p/JDW-5015-2023; li,
   rui/JVM-8999-2024; Yan, Miaochen/JLL-5061-2023; Li, Yao/JJC-2927-2023;
   Zhou, Hong/JKJ-1067-2023; Wang, Jinlong/KHC-3829-2024; Zhang,
   Wei/JKI-3565-2023; Chen, Yu/JLL-0171-2023; Zhang, Shiwei/JIY-4344-2023;
   yuan, lin/JDW-7387-2023; Liu, Jie/JCP-1070-2023; LI, YUN/JTV-7108-2023;
   Yang, Fan/JMA-9594-2023; zhang, xinyu/JKI-8403-2023; liu,
   xiao/JLL-2119-2023; Chen, Xiao/KBD-1464-2024; MA, YAN/HHN-2912-2022;
   yang, peng/JEZ-8452-2023; li, mengyang/JWO-9551-2024; WANG,
   YING/JLM-9219-2023; Yang, Jing/JFK-4046-2023; zhang, yan/JGL-8022-2023;
   qi, li/JFE-7167-2023; Li, Lei/JPE-6543-2023; Wang, Zejun/KBB-8454-2024;
   zhang, chen/JES-0371-2023; Zhao, YiAn/JFK-5881-2023; Wang,
   lingyu/JLM-2013-2023; liu, peng/JSL-1931-2023; zhou, yang/JED-3951-2023;
   li, Li/JPA-0218-2023; zhang, hao/JOJ-7093-2023; Liu, qi/JZT-5038-2024;
   wang, xiaoqiang/JMT-2783-2023; feng, feng/KBR-1814-2024; WANG,
   YANG/JFA-8821-2023; xiao, wei/KCK-6954-2024; liu, yang/JMB-9083-2023;
   Lin, Lin/JTU-1595-2023; chen, Chen/JKJ-2122-2023; liu,
   lin/JFK-3401-2023; Wang, zijun/JNS-5435-2023; chen, gang/JRX-1197-2023;
   Wang, Chao/JHT-6081-2023; li, xiang/JCN-9316-2023; Yang,
   Jie/JDM-6213-2023; yang, li/JGM-1009-2023; wang, wang/JQW-3034-2023
OI Yang, Jing/0009-0004-8274-9863; Yang, Jie/0000-0002-3941-0053; 
FU Clinical Research Plan of Shenkang Hospital Development Center of
   Shanghai [SHDC12020112, SHDC2020CR6005-002]
FX Funding This work was supported by the Clinical Research Plan of
   Shenkang Hospital Development Center of Shanghai (SHDC12020112,
   SHDC2020CR6005-002).
CR Abrey LE, 2006, J CLIN ONCOL, V24, P5711, DOI 10.1200/JCO.2006.08.2941
   Abrey LE, 1998, J CLIN ONCOL, V16, P859, DOI 10.1200/JCO.1998.16.3.859
   Ahn Y, 2017, BLOOD RES, V52, P285, DOI 10.5045/br.2017.52.4.285
   Batchelor TT, 2019, HEMATOL ONCOL, V37, P15, DOI 10.1002/hon.2598
   Bessell EM, 2004, INT J RADIAT ONCOL, V59, P501, DOI 10.1016/j.ijrobp.2003.11.001
   Bhagavathi S, 2008, ARCH PATHOL LAB MED, V132, P1830, DOI 10.1043/1543-2165-132.11.1830
   Chen BB, 2013, CHINESE MED J-PEKING, V126, P482, DOI 10.3760/cma.j.issn.0366-6999.20120844
   Cheson BD, 2014, J CLIN ONCOL, V32, P3059, DOI 10.1200/JCO.2013.54.8800
   Collignon A, 2019, ANN HEMATOL, V98, P915, DOI 10.1007/s00277-018-3564-6
   Crump M, 2017, BLOOD, V130, P1800, DOI 10.1182/blood-2017-03-769620
   Dahiya S, 2013, EXPERT REV ANTICANC, V13, P1109, DOI 10.1586/14737140.2013.829634
   DeAngelis LM, 2002, J CLIN ONCOL, V20, P4643, DOI 10.1200/JCO.2002.11.013
   Ferreri AJM, 2003, J CLIN ONCOL, V21, P266, DOI 10.1200/JCO.2003.09.139
   Ferreri AJM, 2021, LANCET HAEMATOL, V8, pe110, DOI 10.1016/S2352-3026(20)30366-5
   Ferreri AJM, 2011, INT J RADIAT ONCOL, V80, P169, DOI 10.1016/j.ijrobp.2010.01.066
   Ghesquieres H, 2019, ANN ONCOL, V30, P621, DOI 10.1093/annonc/mdz032
   Grommes C, 2017, J CLIN ONCOL, V35, P2410, DOI 10.1200/JCO.2017.72.7602
   Hottinger AF, 2007, NEUROLOGY, V69, P1178, DOI 10.1212/01.wnl.0000276986.19602.c1
   Houillier C, 2021, NEUROLOGY, V97, P628, DOI 10.1212/WNL.0000000000012515
   Li TJ, 2020, FRONT ONCOL, V10, DOI 10.3389/fonc.2020.01306
   Nguyen PL, 2005, J CLIN ONCOL, V23, P1507, DOI 10.1200/JCO.2005.01.161
   O'Neill BP, 2013, AM J HEMATOL, V88, P997, DOI 10.1002/ajh.23551
   Poortmans PMP, 2003, J CLIN ONCOL, V21, P4483, DOI 10.1200/JCO.2003.03.108
   Renaud L, 2021, EUR J HAEMATOL, V107, P370, DOI 10.1111/ejh.13667
   Rubenstein JL, 2018, BLOOD ADV, V2, P1595, DOI 10.1182/bloodadvances.2017014845
   Soussain C, 2019, EUR J CANCER, V117, P121, DOI 10.1016/j.ejca.2019.05.024
   Thiel E, 2010, LANCET ONCOL, V11, P1036, DOI 10.1016/S1470-2045(10)70229-1
   Tun HW, 2018, BLOOD, V132, P2240, DOI 10.1182/blood-2018-02-835496
   Uy GL, 2021, BLOOD, V137, P751, DOI 10.1182/blood.2020007732
   Vallejo A, 2021, CUREUS J MED SCIENCE, V13, DOI 10.7759/cureus.15735
   Zhuang L, 2019, ONCOL REP, V41, P397, DOI 10.3892/or.2018.6781
NR 31
TC 1
Z9 1
U1 0
U2 1
PU FRONTIERS MEDIA SA
PI LAUSANNE
PA AVENUE DU TRIBUNAL FEDERAL 34, LAUSANNE, CH-1015, SWITZERLAND
SN 2234-943X
J9 FRONT ONCOL
JI Front. Oncol.
PD APR 27
PY 2023
VL 13
AR 1098785
DI 10.3389/fonc.2023.1098785
PG 10
WC Oncology
WE Science Citation Index Expanded (SCI-EXPANDED)
SC Oncology
GA F9FU7
UT WOS:000985340800001
PM 37182159
OA gold, Green Published
DA 2024-06-03
ER

PT J
AU Yamasaki, F
   Fudaba, H
   Asano, K
   Sasayama, T
   Natsumeda, M
   Shimabukuro, T
   Taguchi, K
   Koizumi, S
   Nakayama, N
   Fujii, K
   Nishibuchi, I
   Sugiyama, K
   Yoshida, K
   Yonezawa, U
   Yasutomo, M
   Kawasaki, Y
   Kakuta, K
   Katayama, K
   Tanaka, K
   Nagashima, H
   Tsukamoto, Y
   Ideguchi, M
   Nishizaki, T
   Kurozumi, K
   Hosoya, T
   Akita, T
   Kambe, A
AF Yamasaki, Fumiyuki
   Fudaba, Hirotaka
   Asano, Kenichiro
   Sasayama, Takashi
   Natsumeda, Manabu
   Shimabukuro, Taichi
   Taguchi, Kotaro
   Koizumi, Shinichiro
   Nakayama, Noriyuki
   Fujii, Kentaro
   Nishibuchi, Ikuno
   Sugiyama, Kazuhiko
   Yoshida, Kenji
   Yonezawa, Ushio
   Yasutomo, Momii
   Kawasaki, Yukari
   Kakuta, Kiyohide
   Katayama, Kosuke
   Tanaka, Kazuhiro
   Nagashima, Hiroaki
   Tsukamoto, Yoshihiro
   Ideguchi, Makoto
   Nishizaki, Takafumi
   Kurozumi, Kazuhiko
   Hosoya, Tomohiro
   Akita, Tomoyuki
   Kambe, Atsushi
TI Multidrug chemotherapy, whole-brain radiation and cytarabine therapy for
   primary central nervous system lymphoma in elderly patients with dose
   modification based on geriatric assessment: study protocol for a phase
   II, multicentre, non-randomised study
SO BMJ OPEN
LA English
DT Article
DE Clinical trials; Lymphoma; Neurological oncology; GERIATRIC MEDICINE
ID OLDER CANCER-PATIENTS; PRIMARY CNS LYMPHOMA; BREAST-CANCER; SURVIVAL;
   METHOTREXATE; RADIOTHERAPY; PROCARBAZINE; VINCRISTINE; POPULATION;
   TRENDS
AB Introduction Multidrug chemoimmunotherapy with rituximab, high-dose methotrexate, procarbazine and vincristine (R-MPV) is a standard therapy for younger patients with primary central nervous system lymphoma (PCNSL); however, prospective data regarding its use in elderly patients are lacking. This multi-institutional, non-randomised, phase II trial will assess the efficacy and safety of R-MPV and high-dose cytarabine (HD-AraC) for geriatric patients with newly diagnosed PCNSL.
   Methods and analysis Forty-five elderly patients will be included. If R-MPV does not achieve complete response, the patients will undergo reduced-dose, whole-brain radiotherapy comprising 23.4Gy/13 fractions, followed by local boost radiotherapy comprising 21.6Gy/12 fractions. After achieving complete response using R-MPV with or without radiotherapy, the patients will undergo two courses of HD-AraC. All patients will undergo baseline geriatric 8 (G8) assessment before HD-AraC and after three, five and seven R-MPV courses. Patients with screening scores of >= 14 points that decrease to <14 points during subsequent treatment, or those with screening scores <14 points that decrease from the baseline during subsequent treatment are considered unfit for R-MPV/HD-AraC. The primary endpoint is overall survival, and the secondary endpoints are progression-free survival, treatment failure-free survival and frequency of adverse events. The results will guide a later phase III trial and provide information about the utility of a geriatric assessment for defining chemotherapy ineligibility.
   Ethics and dissemination This study complies with the latest Declaration of Helsinki. Written informed consent will be obtained. All participants can quit the study without penalty or impact on treatment. The protocol for the study, statistical analysis plan and informed consent form have been approved by the Certified Review Board at Hiroshima University (CRB6180006) (approval number: CRB2018-0011). The study is ongoing within nine tertiary and two secondary hospitals in Japan. The findings of this trial will be disseminated through national and international presentations and peer-reviewed publications.
   Trial registration jRCTs061180093.
C1 [Yamasaki, Fumiyuki; Yonezawa, Ushio] Hiroshima Univ, Grad Sch Biomed & Hlth Sci, Dept Neurosurg, Hiroshima, Japan.
   [Fudaba, Hirotaka; Yasutomo, Momii; Kawasaki, Yukari] Oita Univ, Fac Med, Dept Neurosurg, Yufu, Japan.
   [Asano, Kenichiro; Kakuta, Kiyohide; Katayama, Kosuke] Hirosaki Univ, Grad Sch Med, Dept Neurosurg, Hirosaki, Aomori, Japan.
   [Sasayama, Takashi; Tanaka, Kazuhiro; Nagashima, Hiroaki] Kobe Univ, Grad Sch Med, Dept Neurosurg, Kobe, Hyogo, Japan.
   [Natsumeda, Manabu; Tsukamoto, Yoshihiro] Niigata Univ, Brain Res Inst, Dept Neurosurg, Niigata, Japan.
   [Shimabukuro, Taichi; Ideguchi, Makoto; Nishizaki, Takafumi] Ube Ind Cent Hosp, Dept Neurosurg, Ube, Yamaguchi, Japan.
   [Taguchi, Kotaro] Natl Hosp Org Yamaguchi Ube Med Ctr, Dept Radiol, Ube, Yamaguchi, Japan.
   [Koizumi, Shinichiro; Kurozumi, Kazuhiko] Hamamatsu Univ Sch Med, Dept Neurosurg, Hamamatsu, Shizuoka, Japan.
   [Nakayama, Noriyuki] Gifu Univ, Dept Neurosurg, Grad Sch Med, Gifu, Japan.
   [Fujii, Kentaro] Okayama Univ, Fac Med Dent & Pharmaceut Sci, Dept Neurol Surg, Okayama, Japan.
   [Nishibuchi, Ikuno] Hiroshima Univ, Grad Sch Biomed & Hlth Sci, Dept Radiat Oncol, Hiroshima, Japan.
   [Sugiyama, Kazuhiko] Hiroshima Univ Hosp, Dept Clin Oncol, Hiroshima, Japan.
   [Sugiyama, Kazuhiko] Hiroshima Univ Hosp, Neurooncol Program, Hiroshima, Japan.
   [Yoshida, Kenji] Tottori Univ Hosp, Dept Radiat Oncol, Yonago, Tottori, Japan.
   [Hosoya, Tomohiro; Kambe, Atsushi] Tottori Univ, Div Neurosurg, Dept Brain & Neurosci, Yonago, Tottori, Japan.
   [Akita, Tomoyuki] Hiroshima Univ Hosp, Clin Res Ctr Hiroshima, Dept Biostat, Hiroshima, Japan.
C3 Hiroshima University; Oita University; Hirosaki University; Kobe
   University; Niigata University; Hamamatsu University School of Medicine;
   Gifu University; Okayama University; Hiroshima University; Hiroshima
   University; Hiroshima University; Tottori University; Tottori
   University; Hiroshima University
RP Yamasaki, F (corresponding author), Hiroshima Univ, Grad Sch Biomed & Hlth Sci, Dept Neurosurg, Hiroshima, Japan.; Kambe, A (corresponding author), Tottori Univ, Div Neurosurg, Dept Brain & Neurosci, Yonago, Tottori, Japan.
EM
RI Natsumeda, Manabu/Z-2072-2019
OI Natsumeda, Manabu/0000-0003-3098-8323; Yamasaki,
   Fumiyuki/0000-0003-0164-6295
FU Tsuchiya Memorial Medical Foundation [J17KF00167]
FX This study is supported by a grant from Tsuchiya Memorial Medical
   Foundation (grant number J17KF00167 to FY).
CR Abrey LE, 2005, J CLIN ONCOL, V23, P5034, DOI 10.1200/JCO.2005.13.524
   Alaggio R, 2022, LEUKEMIA, V36, P1720, DOI 10.1038/s41375-022-01620-2
   [Anonymous], 2017, Neurol Med Chir (Tokyo), V57, P9, DOI 10.2176/nmc.sup.2017-0001
   Bellera CA, 2012, ANN ONCOL, V23, P2166, DOI 10.1093/annonc/mdr587
   Bouchardy C, 2003, J CLIN ONCOL, V21, P3580, DOI 10.1200/JCO.2003.02.046
   Campo E, 2011, BLOOD, V117, P5019, DOI 10.1182/blood-2011-01-293050
   Decoster L, 2015, ANN ONCOL, V26, P288, DOI 10.1093/annonc/mdu210
   Eloranta S, 2018, EUR J HAEMATOL, V100, P61, DOI 10.1111/ejh.12980
   Extermann M, 2005, CRIT REV ONCOL HEMAT, V55, P241, DOI 10.1016/j.critrevonc.2005.06.003
   Fallah J, 2016, BLOOD ADV, V1, P112, DOI 10.1182/bloodadvances.2016000927
   Farhi J, 2018, ANN HEMATOL, V97, P2391, DOI 10.1007/s00277-018-3468-5
   Freeman Tracelyn, 2020, Neurooncol Adv, V2, pvdaa077, DOI 10.1093/noajnl/vdaa077
   Garcia MV, 2021, JAMA ONCOL, V7, P616, DOI 10.1001/jamaoncol.2020.6736
   Giordano SH, 2005, J CLIN ONCOL, V23, P783, DOI 10.1200/JCO.2005.04.175
   Hamaker ME, 2014, LEUKEMIA RES, V38, P275, DOI 10.1016/j.leukres.2013.12.018
   Hamaker ME, 2018, J GERIATR ONCOL, V9, P430, DOI 10.1016/j.jgo.2018.03.014
   Kasenda B, 2015, ANN ONCOL, V26, P1305, DOI 10.1093/annonc/mdv076
   Kerbauy MN, 2017, RADIOTHER ONCOL, V122, P352, DOI 10.1016/j.radonc.2016.12.033
   Lee JY, 2021, BLOOD RES, V56, P285, DOI 10.5045/br.2021.2021073
   Mendez JS, 2018, NEURO-ONCOLOGY, V20, P687, DOI 10.1093/neuonc/nox187
   Monfardini S, 2021, J GERIATR ONCOL, V12, P991, DOI 10.1016/j.jgo.2021.02.015
   Morris PG, 2013, J CLIN ONCOL, V31, P3971, DOI 10.1200/JCO.2013.50.4910
   Omuro A, 2015, LANCET HAEMATOL, V2, pE251, DOI 10.1016/S2352-3026(15)00074-5
   Omuro A, 2015, BLOOD, V125, P1403, DOI 10.1182/blood-2014-10-604561
   Puts MTE, 2012, JNCI-J NATL CANCER I, V104, P1133, DOI 10.1093/jnci/djs285
   RUBENSTEIN L Z, 1991, Journal of the American Geriatrics Society, V39, p8S
   Shiels MS, 2016, BRIT J HAEMATOL, V174, P417, DOI 10.1111/bjh.14073
   Siegal T, 2019, ACTA HAEMATOL-BASEL, V141, P138, DOI 10.1159/000495284
   Suzuki Y, 2022, INTERNAL MED, V61, P1345, DOI 10.2169/internalmedicine.7805-21
   Tang DS, 2022, FRONT ONCOL, V12, DOI 10.3389/fonc.2022.817043
   van der Meulen M, 2017, LEUKEMIA, V31, P1822, DOI 10.1038/leu.2017.128
NR 31
TC 1
Z9 1
U1 0
U2 0
PU BMJ PUBLISHING GROUP
PI LONDON
PA BRITISH MED ASSOC HOUSE, TAVISTOCK SQUARE, LONDON WC1H 9JR, ENGLAND
SN 2044-6055
J9 BMJ OPEN
JI BMJ Open
PD APR
PY 2023
VL 13
IS 4
AR e071350
DI 10.1136/bmjopen-2022-071350
PG 8
WC Medicine, General & Internal
WE Science Citation Index Expanded (SCI-EXPANDED)
SC General & Internal Medicine
GA Q5MG1
UT WOS:001057956000155
PM 37094899
OA gold, Green Published
DA 2024-06-03
ER

PT J
AU Dufour, J
   Choquet, S
   Hoang-Xuan, K
   Schmitt, A
   Ahle, G
   Houot, R
   Taillandier, L
   Gressin, R
   Casasnovas, O
   Marolleau, JP
   Tamburini, J
   Serrier, C
   Perez, E
   Paillassa, J
   Gyan, E
   Chauchet, A
   Ursu, R
   Kas, A
   Soussain, C
   Houillier, C
AF Dufour, J.
   Choquet, S.
   Hoang-Xuan, K.
   Schmitt, A.
   Ahle, G.
   Houot, R.
   Taillandier, L.
   Gressin, R.
   Casasnovas, O.
   Marolleau, J. P.
   Tamburini, J.
   Serrier, C.
   Perez, E.
   Paillassa, J.
   Gyan, E.
   Chauchet, A.
   Ursu, R.
   Kas, A.
   Soussain, C.
   Houillier, C.
TI Systemic relapses of primary CNS lymphomas (PCNSL): a LOC network study
SO ANNALS OF HEMATOLOGY
LA English
DT Article
DE Primary CNS lymphoma; Systemic relapses; PNS relapses; Aggressive
   non-Hodgkin's lymphoma
ID B-CELL LYMPHOMA; CHEMOTHERAPY; INVOLVEMENT; FEATURES; PET
AB Primary central nervous system lymphomas (PCNSLs) classically remain confined within the CNS throughout their evolution for unknown reasons. Our objective was to analyse the rare extracerebral relapses of PCNSL in a nationwide population-based study. We retrospectively selected PCNSL patients who experienced extracerebral relapse during their follow-up from the French LOC database. Of the 1968 PCNSL included in the database from 2011, 30 (1.5%, median age 71 years, median KPS 70) presented an extracerebral relapse, either pure (n = 20) or mixed (both extracerebral and in the CNS) (n = 10), with a histological confirmation in 20 cases. The median delay between initial diagnosis and systemic relapse was 15.5 months [2-121 months]. We found visceral (n = 23, 77%), including testis in 5 (28%) men and breast in 3 (27%) women, lymph node (n = 12, 40%), and peripheral nervous system (PNS) (n = 7, 23%) involvement. Twenty-seven patients were treated with chemotherapy, either with only systemic targets (n = 7) or mixed systemic and CNS targets (n = 20), 4 were consolidated by HCT-ASCT. After systemic relapse, the median progression-free survival and overall survival (OS) were 7 and 12 months, respectively. KPS > 70 and pure systemic relapses were significantly associated with higher OS. Extracerebral PCNSL relapses are rare, mainly extranodal, and frequently involve the testis, breast, and PNS. The prognosis was worse in mixed relapses. Early relapses raise the question of misdiagnosed occult extracerebral lymphoma at diagnostic workup that should systematically include a PET-CT. Paired tumour analysis at diagnosis/relapse would provide a better understanding of the underlying molecular mechanisms.
C1 [Dufour, J.; Hoang-Xuan, K.; Houillier, C.] Sorbonne Univ, Hop Pitie Salpetriere, Serv Neurol 2 Mazarin, AP HP,IHU,ICM, Paris, France.
   [Choquet, S.] Hop La Pitie Salpetriere, Serv Hematol Clin, Paris, France.
   [Schmitt, A.] Inst Bergonie, Serv Hematol, Bordeaux, France.
   [Ahle, G.] Hop Civils Colmar, Serv Neurol, Colmar, France.
   [Houot, R.] Hop Univ Rennes, Serv Hematol, Rennes, France.
   [Taillandier, L.] Hop Univ Nancy, Serv Neurol, Nancy, France.
   [Gressin, R.] Hop Univ Grenoble, Serv Hematol, Grenoble, France.
   [Casasnovas, O.] Hop Univ Dijon, Serv Hematol Clin, Dijon, France.
   [Marolleau, J. P.] Hop Univ Amiens, Serv Hematol Clin, Amiens, France.
   [Tamburini, J.] Hop Cochin, Serv Hematol, Paris, France.
   [Serrier, C.] Ctr Hosp Perpignan, Serv Hematol, Perpignan, France.
   [Perez, E.] Hop Univ Reunion, Serv Oncol Hematol, Paris, France.
   [Paillassa, J.] Hop Univ Angers, Serv Hematol, Angers, France.
   [Gyan, E.] Hop Univ Tours, Serv Hematol, Tours, France.
   [Chauchet, A.] Hop Univ Besancon, Serv Hematol, Besancon, France.
   [Ursu, R.] Hop St Louis, Serv Neurol Orientat Oncol, Paris, France.
   [Kas, A.] Hop La Pitie Salpetriere, Serv Med Nucl, Paris, France.
   [Soussain, C.] Inst Curie, Serv Hematol, St Cloud, France.
   [Soussain, C.] PSL Res Univ, Inst Curie, INSERM, U932, Paris, France.
C3 Assistance Publique Hopitaux Paris (APHP); Hopital Universitaire
   Pitie-Salpetriere - APHP; Sorbonne Universite; Assistance Publique
   Hopitaux Paris (APHP); Hopital Universitaire Pitie-Salpetriere - APHP;
   Sorbonne Universite; UNICANCER; Institut Bergonie; CHU Rennes;
   Universite de Rennes; CHU de Nancy; Communaute Universite Grenoble
   Alpes; Universite Grenoble Alpes (UGA); CHU Grenoble Alpes; CHU Dijon
   Bourgogne; Universite de Picardie Jules Verne (UPJV); CHU Amiens;
   Assistance Publique Hopitaux Paris (APHP); Universite Paris Cite;
   Hopital Universitaire Cochin - APHP; Universite d'Angers; Centre
   Hospitalier Universitaire d'Angers; CHU Tours; Universite de
   Franche-Comte; CHU Besancon; Assistance Publique Hopitaux Paris (APHP);
   Universite Paris Cite; Hopital Universitaire Saint-Louis - APHP;
   Sorbonne Universite; Assistance Publique Hopitaux Paris (APHP); Hopital
   Universitaire Pitie-Salpetriere - APHP; UNICANCER; Universite PSL;
   Institut Curie; Universite PSL; UNICANCER; Institut Curie; Institut
   National de la Sante et de la Recherche Medicale (Inserm)
RP Houillier, C (corresponding author), Sorbonne Univ, Hop Pitie Salpetriere, Serv Neurol 2 Mazarin, AP HP,IHU,ICM, Paris, France.
EM
OI Tamburini, Jerome/0000-0002-0167-5996
CR Abrey LE, 2006, J CLIN ONCOL, V24, P5711, DOI 10.1200/JCO.2006.08.2941
   Abrey LE, 2005, J CLIN ONCOL, V23, P5034, DOI 10.1200/JCO.2005.13.524
   Barajas RF, 2021, NEURO-ONCOLOGY, V23, P1056, DOI 10.1093/neuonc/noab020
   Bertaux M, 2020, J NEURO-ONCOL, V148, P343, DOI 10.1007/s11060-020-03525-5
   Castillo JJ, 2014, AM J HEMATOL, V89, P310, DOI 10.1002/ajh.23638
   Chapuy B, 2016, BLOOD, V127, P869, DOI 10.1182/blood-2015-10-673236
   Cheson BD, 2015, CHIN CLIN ONCOL, V4, DOI 10.3978/j.issn.2304-3865.2014.11.03
   DeAngelis LM, 2002, J CLIN ONCOL, V20, P4643, DOI 10.1200/JCO.2002.11.013
   Ferreri AJM, 2003, J CLIN ONCOL, V21, P266, DOI 10.1200/JCO.2003.09.139
   Gholam D, 2003, LEUKEMIA LYMPHOMA, V44, P1173, DOI 10.1080/1042819031000079195
   Gogia A, 2018, SOUTH ASIAN J CANCER, V7, P200, DOI 10.4103/sajc.sajc_65_18
   Grisariu S, 2010, BLOOD, V115, P5005, DOI 10.1182/blood-2009-12-258210
   Guedj E, 2022, EUR J NUCL MED MOL I, V49, P632, DOI 10.1007/s00259-021-05603-w
   Gupta T, 2021, CL LYMPH MYELOM LEUK, V21, P497, DOI 10.1016/j.clml.2021.03.011
   Hoang-Xuan K, 2015, LANCET ONCOL, V16, pE322, DOI 10.1016/S1470-2045(15)00076-5
   Houillier C, 2020, NEUROLOGY, V94, pE1027, DOI 10.1212/WNL.0000000000008900
   Illerhaus G, 2016, LANCET HAEMATOL, V3, pE388, DOI 10.1016/S2352-3026(16)30050-3
   Jahnke K, 2006, J CLIN ONCOL, V24, P4754, DOI 10.1200/JCO.2006.06.7165
   Jahnke K, 2006, J NEURO-ONCOL, V80, P159, DOI 10.1007/s11060-006-9165-6
   Kridel R, 2017, BRIT J HAEMATOL, V176, P210, DOI 10.1111/bjh.14392
   Langner-Lemercier S, 2016, NEURO-ONCOLOGY, V18, P1297, DOI 10.1093/neuonc/now033
   Mohile NA, 2008, NEURO-ONCOLOGY, V10, P223, DOI 10.1215/15228517-2007-061
   Moller MB, 2004, BRIT J HAEMATOL, V124, P151, DOI 10.1046/j.1365-2141.2003.04749.x
   Ostrom QT, 2018, NEURO-ONCOLOGY, V20, P1, DOI 10.1093/neuonc/noy131
   Park HY, 2021, AM J ROENTGENOL, V216, P1172, DOI 10.2214/AJR.20.24036
   Provencher S, 2011, HEMATOL ONCOL, V29, P10, DOI 10.1002/hon.944
   ROSENBERG SA, 1977, CANCER TREAT REP, V61, P1023
   Shan YD, 2018, DIS MARKERS, V2018, DOI 10.1155/2018/7860494
   SHIPP MA, 1993, NEW ENGL J MED, V329, P987, DOI 10.1056/nejm199309303291402
NR 29
TC 1
Z9 1
U1 0
U2 0
PU SPRINGER
PI NEW YORK
PA ONE NEW YORK PLAZA, SUITE 4600, NEW YORK, NY, UNITED STATES
SN 0939-5555
EI 1432-0584
J9 ANN HEMATOL
JI Ann. Hematol.
PD MAY
PY 2023
VL 102
IS 5
BP 1159
EP 1169
DI 10.1007/s00277-023-05108-6
EA MAR 2023
PG 11
WC Hematology
WE Science Citation Index Expanded (SCI-EXPANDED)
SC Hematology
GA AG6X6
UT WOS:000960793200001
PM 36991231
DA 2024-06-03
ER

PT J
AU Wu, Z
   Li, ZR
   Qiu, XY
   Zhong, MK
   Ding, TL
AF Wu, Zhuo
   Li, Ziran
   Qiu, Xiaoyan
   Zhong, Mingkang
   Ding, Tianling
TI Germline genetic variations in methotrexate pathway are associated with
   pharmacokinetics, outcome, and toxicity in patients with primary central
   nervous system lymphoma
SO EXPERT REVIEW OF CLINICAL PHARMACOLOGY
LA English
DT Article
DE Clinical outcome; genetic variations; methotrexate; PCNSL;
   pharmacokinetics; toxicity
ID HIGH-DOSE METHOTREXATE; SINGLE-NUCLEOTIDE POLYMORPHISMS; ACUTE
   LYMPHOBLASTIC-LEUKEMIA; PLASMA-LEVELS; IMPACT; HYDROLASE; SURVIVAL;
   ADULTS; SNPS
AB BackgroundHigh-dose methotrexate (HD-MTX)-based regimens are the standard treatment for patients with primary central nervous system lymphoma (PCNSL); however, MTX has extensive interpatient variability in pharmacokinetics and clinical outcomes, with genetic variation an important factor involved in the variability in drug response.Methods123 PCNSL patients who received 524 courses of chemotherapy were genotyped for 42 single nucleotide polymorphisms in MTX pathway. The relationship between these polymorphisms and the pharmacokinetics, clinical outcomes, and toxicity of MTX was explored using both univariate and multivariate analyses.ResultsWe found ABCB1 rs2032582 and GGH rs2305558 were substantially associated with the pharmacokinetics of MTX. Patients with GGH rs2305558 T and ABCB1 rs2032582 non-G allele had a higher C-max increased by 20.5%, area under the concentration-time curve (AUC(0-48h)) increased by 19.6% and lower clearance decreased by 19.6%. ABCB1 rs1045642 and rs2032582 might be independent predictors of progression-free survival. Patients with ABCB1 rs1045642 non-A and rs2032582 G allele correlated with higher progression risk of the disease. Furthermore, ATIC rs3821353 was associated with MTX-induced hepatotoxicity (Grade >= 2).ConclusionThese variants may serve as biomarkers to predict the pharmacokinetics, clinical outcomes, and hepatotoxicity of MTX and contribute to personalized therapy for PCNSL patients.
C1 [Wu, Zhuo; Li, Ziran; Qiu, Xiaoyan; Zhong, Mingkang] Fudan Univ, Huashan Hosp, Dept Pharm, Shanghai, Peoples R China.
   [Ding, Tianling] Fudan Univ, Huashan Hosp, Dept Hematol, 12 Middle Urumqi Rd, Shanghai 200040, Peoples R China.
C3 Fudan University; Fudan University
RP Ding, TL (corresponding author), Fudan Univ, Huashan Hosp, Dept Hematol, 12 Middle Urumqi Rd, Shanghai 200040, Peoples R China.
EM
RI ZIRAN, LI/KDN-8374-2024
OI ZIRAN, LI/0000-0001-6740-6003; Wu, Zhuo/0000-0003-1720-0829
CR Abrey LE, 2005, J CLIN ONCOL, V23, P5034, DOI 10.1200/JCO.2005.13.524
   Anglicheau D, 2003, J AM SOC NEPHROL, V14, P1889, DOI 10.1097/01.ASN.0000073901.94759.36
   Avivi I, 2014, LEUKEMIA LYMPHOMA, V55, P565, DOI 10.3109/10428194.2013.789506
   Cole PD, 2001, CANCER RES, V61, P4599
   Crump M, 2017, BLOOD, V130, P1800, DOI 10.1182/blood-2017-03-769620
   de Beaumais TA, 2012, CURR DRUG METAB, V13, P822
   de Deus Dayse Maria Vasconcelos, 2012, Leuk Res Treatment, V2012, P292043, DOI 10.1155/2012/292043
   de Rotte MCFJ, 2015, ANN RHEUM DIS, V74, P408, DOI 10.1136/annrheumdis-2013-203725
   Deckert M, 2011, LEUKEMIA, V25, P1797, DOI 10.1038/leu.2011.169
   Dolecek TA, 2012, NEURO-ONCOLOGY, V14, pv1, DOI 10.1093/neuonc/nos218
   Ferreri AJM, 2003, J CLIN ONCOL, V21, P266, DOI 10.1200/JCO.2003.09.139
   Gregers J, 2015, PHARMACOGENOMICS J, V15, P372, DOI 10.1038/tpj.2014.81
   Jamroziak K, 2004, EUR J HAEMATOL, V72, P314, DOI 10.1111/j.1600-0609.2004.00228.x
   Joerger M, 2012, LEUKEMIA LYMPHOMA, V53, P1867, DOI 10.3109/10428194.2012.676177
   Kato T, 2012, DRUG METAB PHARMACOK, V27, P192, DOI 10.2133/dmpk.DMPK-11-RG-066
   König J, 1999, BBA-BIOMEMBRANES, V1461, P377, DOI 10.1016/S0005-2736(99)00169-8
   Kotnik BF, 2017, RADIOL ONCOL, V51, P455, DOI 10.1515/raon-2017-0040
   Laverdière C, 2002, BLOOD, V100, P3832, DOI 10.1182/blood.V100.10.3832
   Leil TA, 2010, CLIN PHARMACOL THER, V88, P375, DOI 10.1038/clpt.2010.106
   Lima A, 2016, PHARMACOGENOMICS, V17, P1649, DOI 10.2217/pgs-2016-0067
   Liu Y, 2014, PLOS ONE, V9, DOI 10.1371/journal.pone.0082681
   Lui G, 2018, J CLIN PHARMACOL, V58, P1541, DOI 10.1002/jcph.1252
   Mao JJ, 2022, FRONT PHARMACOL, V13, DOI 10.3389/fphar.2022.817673
   Molnar P P, 1999, Neuro Oncol, V1, P89, DOI 10.1093/neuonc/1.2.89
   Pauli-Magnus C, 2004, PHARM RES-DORDR, V21, P904, DOI 10.1023/B:PHAM.0000029276.21063.0b
   Polso AK, 2014, J CLIN PHARM THER, V39, P584, DOI 10.1111/jcpt.12200
   Radtke S, 2013, BLOOD, V121, P5145, DOI 10.1182/blood-2013-01-480335
   Ranganathan P, 2008, J RHEUMATOL, V35, P572
   Rau T, 2006, CLIN PHARMACOL THER, V80, P468, DOI 10.1016/j.clpt.2006.08.012
   Razali RH, 2020, PEDIATR HEMAT ONCOL, V37, P185, DOI 10.1080/08880018.2019.1705949
   Robey RW, 2018, NAT REV CANCER, V18, P452, DOI 10.1038/s41568-018-0005-8
   Rots Marianne G., 1999, Blood, V93, P1677
   Sandhu A, 2018, CLIN RHEUMATOL, V37, P3221, DOI 10.1007/s10067-018-4206-z
   Sandhu A, 2017, THER DRUG MONIT, V39, P157, DOI 10.1097/FTD.0000000000000375
   Shiels MS, 2016, BRIT J HAEMATOL, V174, P417, DOI 10.1111/bjh.14073
   Staatz CE, 2010, CLIN PHARMACOKINET, V49, P141, DOI 10.2165/11317350-000000000-00000
   Suh Y, 2005, MUTAT RES-FUND MOL M, V573, P41, DOI 10.1016/j.mrfmmm.2005.01.005
   Sun FJ, 2021, FRONT PHARMACOL, V12, DOI 10.3389/fphar.2021.641075
   Suthandiram S, 2014, PHARMACOGENOMICS, V15, P1479, DOI [10.2217/PGS.14.97, 10.2217/pgs.14.97]
   Swerdlow SH, 2016, BLOOD, V127, P2375, DOI 10.1182/blood-2016-01-643569
   van de Meeberg MM, 2023, BRIT J CLIN PHARMACO, V89, P61, DOI 10.1111/bcp.15579
   Villano JL, 2011, BRIT J CANCER, V105, P1414, DOI 10.1038/bjc.2011.357
   Wang SM, 2020, PHARMACOTHERAPY, V40, P614, DOI 10.1002/phar.2430
   Wu T, 2019, ANN HEMATOL, V98, P923, DOI 10.1007/s00277-019-03629-7
   Wu Z, 2019, EUR J CLIN PHARMACOL, V75, P33, DOI 10.1007/s00228-018-2546-x
   Xie QF, 2018, CURR PHARM DESIGN, V24, P3558, DOI 10.2174/1381612824666181018153641
   Xu LL, 2018, J BONE ONCOL, V13, P143, DOI 10.1016/j.jbo.2018.10.002
   Yang L, 2017, PHARMACOGENOMICS, V18, P1557, DOI 10.2217/pgs-2017-0110
   Yuan Y, 2021, BIOMARK RES, V9, DOI 10.1186/s40364-021-00282-z
   Zhao Q, 2021, J PHARM PHARMACOL, V73, P1480, DOI 10.1093/jpp/rgab099
NR 50
TC 1
Z9 1
U1 0
U2 0
PU TAYLOR & FRANCIS LTD
PI ABINGDON
PA 2-4 PARK SQUARE, MILTON PARK, ABINGDON OR14 4RN, OXON, ENGLAND
SN 1751-2433
EI 1751-2441
J9 EXPERT REV CLIN PHAR
JI Expert Rev. Clin. Pharmacol.
PD APR 3
PY 2023
VL 16
IS 4
BP 371
EP 381
DI 10.1080/17512433.2023.2194630
EA MAR 2023
PG 11
WC Pharmacology & Pharmacy
WE Science Citation Index Expanded (SCI-EXPANDED)
SC Pharmacology & Pharmacy
GA D6MZ1
UT WOS:000954258500001
PM 36946320
DA 2024-06-03
ER

PT J
AU Zhang, R
   Wei, BY
   Hu, YY
   Lv, WY
   Adilai, A
   Yang, F
   Zhang, JN
   Cheng, G
AF Zhang, Rui
   Wei, Boyuan
   Hu, Yiyang
   Lv, Wenying
   Adilai, Abudurexiti
   Yang, Fan
   Zhang, Jianning
   Cheng, Gang
TI Whole-Exome Sequencing Revealed the Mutational Profiles of Primary
   Central Nervous System Lymphoma
SO CLINICAL LYMPHOMA MYELOMA & LEUKEMIA
LA English
DT Article
DE High-dose methotrexate; MYD88; Genetic alteration; Pathway analysis;
   Interaction network
ID MYD88; EXPRESSION; TRENDS; EPHB1
AB We analyzed 12 cases of Chinese PCNSL using whole-exome sequencing to detect genetic alterations. We identified 448 potentially somatic single nucleotide variants (SNVs) with a median of 12 SNVs per PCNSL sample and 35 small indels with potentially protein-changing features in 9 PCNSL samples. MYD88 with the most mutant frequency affected the activity of NF -KB pathway. PCNSL samples with LRP1B mutations had higher mutation rate than samples with LRP1B wildtype. In conclusion, whole-exome sequencing revealed significantly mutated genes underlying the potential molecular mechanisms of PCNSL, which could serve as therapeutic targets.Background: Primary central nervous system lymphoma (PCNSL) is a highly aggressive type of extranodal non -Hodgkin lymphoma, of which approximately 90% of the cases are diffuse large B-cell lymphoma (DLBCL). In recent years, the incidence of PCNSL has significantly increased in women and older men. Although advanced treatments such as high-dose methotrexate (HD-MTX) and targeted agents have been introduced, the prognosis of these patients remains poorer than those with other forms of non-Hodgkin's lymphoma.Methods: Twelve cases of Chinese PCNSL were analyzed to detect their genetic alterations using whole-exome sequencing (WES). We identified 448 potential somatic single nucleotide variants (SNVs) with a median of 12 SNVs per PCNSL sample and 35 small indels with potentially protein-changing features in 9 PCNSL samples.Results: We found that myeloid differentiation factor 88 (MYD88) had the highest mutation frequency, which affected the activity of the nuclear factor -KB (NF -KB) pathway. PCNSL samples with low-density lipoprotein receptor-related protein 1B (LRP1B) mutations had a higher mutation rate than samples with wild-type LRP1B. Polycystic kidney and hepatic disease 1 (PKHD1), the causal gene of autosomal recessive polycystic kidney disease (ARPKD), was identified in 2 PCNSL cases and exhibited missense mutations. Pathway analysis revealed enrichment in pathways associated with central carbon metabolism in cancer, renal cell carcinoma, nicotine addiction, bladder cancer, and long-term depression.Conclusions: WES revealed significantly mutated genes associated with the molecular mechanisms of PCNSL, which could serve as therapeutic targets to improve patient outcomes.
C1 [Zhang, Rui; Wei, Boyuan; Hu, Yiyang; Adilai, Abudurexiti; Yang, Fan; Zhang, Jianning; Cheng, Gang] Chinese Peoples Liberat Army Gen Hosp, Med Ctr 1, Dept Neurosurg, Beijing, Peoples R China.
   [Hu, Yiyang] Fourth Mil Med Univ, Dept Med Genet & Dev Biol, Xian, Peoples R China.
   [Lv, Wenying] Chinese Peoples Liberat Army Gen Hosp, Med Ctr 6, Dept Neurosurg, Beijing, Peoples R China.
   [Zhang, Jianning; Cheng, Gang] 28 Fuxing Rd, Beijing, Peoples R China.
C3 Chinese People's Liberation Army General Hospital; Air Force Military
   Medical University; Chinese People's Liberation Army General Hospital
RP Zhang, JN; Cheng, G (corresponding author), 28 Fuxing Rd, Beijing, Peoples R China.
EM
CR Alexandrov LB, 2013, NATURE, V500, P415, DOI 10.1038/nature12477
   Bergmann C, 2005, KIDNEY INT, V67, P829, DOI 10.1111/j.1523-1755.2005.00148.x
   Bergmann C, 2003, J AM SOC NEPHROL, V14, P76, DOI 10.1097/01.ASN.0000039578.55705.6E
   Bhatia S, 2015, ONCOTARGET, V6, P8929, DOI 10.18632/oncotarget.3369
   Braggio E, 2011, CLIN CANCER RES, V17, P4245, DOI 10.1158/1078-0432.CCR-11-0395
   Camilleri-Broet S, 1998, AM J CLIN PATHOL, V110, P607, DOI 10.1093/ajcp/110.5.607
   Chen H, 2019, FRONT IMMUNOL, V10, DOI 10.3389/fimmu.2019.01113
   Cibulskis K, 2013, NAT BIOTECHNOL, V31, P213, DOI 10.1038/nbt.2514
   Cote TR, 1996, J NATL CANCER I, V88, P675, DOI 10.1093/jnci/88.10.675
   Davis RE, 2010, NATURE, V463, P88, DOI 10.1038/nature08638
   Du X, 2018, THORAC CANCER, V9, P423, DOI 10.1111/1759-7714.12613
   Gonzalez-Aguilar A, 2012, CLIN CANCER RES, V18, P5203, DOI 10.1158/1078-0432.CCR-12-0845
   Grommes C, 2017, J CLIN ONCOL, V35, P2410, DOI 10.1200/JCO.2017.72.7602
   Gunay-Aygun M, 2010, MOL GENET METAB, V99, P160, DOI 10.1016/j.ymgme.2009.10.010
   Hiemcke-Jiwa LS, 2019, BRIT J HAEMATOL, V185, P974, DOI 10.1111/bjh.15674
   Hoffman S, 2006, NEURO-ONCOLOGY, V8, P27, DOI 10.1215/S1522851705000323
   Jahnke K, 2006, J NEURO-ONCOL, V80, P159, DOI 10.1007/s11060-006-9165-6
   Katoh M, 2012, INT J ONCOL, V41, P1913, DOI 10.3892/ijo.2012.1669
   Lam LT, 2017, MOL CANCER THER, V16, P1511, DOI 10.1158/1535-7163.MCT-16-0459
   Lan SW, 2019, LUNG CANCER, V132, P154, DOI 10.1016/j.lungcan.2019.04.025
   Langner-Lemercier S, 2016, NEURO-ONCOLOGY, V18, P1297, DOI 10.1093/neuonc/now033
   Li BJ, 2019, J CANCER, V10, P577, DOI 10.7150/jca.28905
   Li L, 2017, INT J CLIN EXP PATHO, V10, P10092
   Nakamura T, 2016, NEUROPATH APPL NEURO, V42, P279, DOI 10.1111/nan.12259
   Nosrati A, 2019, ANN HEMATOL, V98, P169, DOI 10.1007/s00277-018-3498-z
   O'Neill BP, 2013, AM J HEMATOL, V88, P997, DOI 10.1002/ajh.23551
   Oh JH, 2020, NPJ GENOM MED, V5, DOI 10.1038/s41525-019-0107-6
   Qian YJ, 2017, MOL MED REP, V16, P6034, DOI 10.3892/mmr.2017.7319
   Saunders CT, 2012, BIOINFORMATICS, V28, P1811, DOI 10.1093/bioinformatics/bts271
   Schwindt H, 2009, LEUKEMIA, V23, P1875, DOI 10.1038/leu.2009.120
   Tabouret E, 2015, J NEUROL SCI, V358, P440, DOI 10.1016/j.jns.2015.09.345
   Turco MC, 2004, LEUKEMIA, V18, P11, DOI 10.1038/sj.leu.2403171
   Villano JL, 2011, BRIT J CANCER, V105, P1414, DOI 10.1038/bjc.2011.357
   Wang K, 2010, NUCLEIC ACIDS RES, V38, DOI 10.1093/nar/gkq603
   Xie C, 2011, NUCLEIC ACIDS RES, V39, pW316, DOI 10.1093/nar/gkr483
   Yamada S, 2015, LEUKEMIA LYMPHOMA, V56, P2141, DOI 10.3109/10428194.2014.979413
   Young RM, 2015, SEMIN HEMATOL, V52, P77, DOI 10.1053/j.seminhematol.2015.01.008
   Yuan Y, 2014, ASIAN PAC J CANCER P, V15, P1441, DOI 10.7314/APJCP.2014.15.3.1441
   Zheng M, 2017, NEUROPATHOLOGY, V37, P509, DOI 10.1111/neup.12405
   Zhou YY, 2018, NEOPLASIA, V20, P1059, DOI 10.1016/j.neo.2018.08.012
   Zhu Q, 2022, FRONT GENET, V13
NR 41
TC 1
Z9 1
U1 0
U2 3
PU CIG MEDIA GROUP, LP
PI DALLAS
PA 3500 MAPLE AVENUE, STE 750, DALLAS, TX 75219-3931 USA
SN 2152-2650
EI 2152-2669
J9 CL LYMPH MYELOM LEUK
JI Clin. Lymphoma Myeloma Leuk.
PD APR
PY 2023
VL 23
IS 4
BP 291
EP 302
DI 10.1016/j.clml.2023.01.003
EA MAR 2023
PG 12
WC Oncology; Hematology
WE Science Citation Index Expanded (SCI-EXPANDED)
SC Oncology; Hematology
GA H2XZ1
UT WOS:000994660600001
PM 36725383
DA 2024-06-03
ER

PT J
AU Ostrowska, B
   Domanska-Czyz, K
   Romejko-Jarosinska, J
   Osowiecki, M
   Targonski, L
   Poplawska, L
   Konecki, R
   Kotarska, M
   Szymanski, M
   Borawska, A
   Swierkowska, M
   Dabrowska-Iwanicka, A
   Druzd-Sitek, A
   Paszkiewicz-Kozik, E
   Mroz-Zycinska, E
   Tajer, J
   Wojciechowska-Lampka, E
   Osiadacz, W
   Rymkiewicz, G
   Lapinska, G
   Wojewodzka-Mirocha, M
   Michalski, W
   Walewski, J
AF Ostrowska, Beata
   Domanska-Czyz, Katarzyna
   Romejko-Jarosinska, Joanna
   Osowiecki, Michal
   Targonski, Lukasz
   Poplawska, Lidia
   Konecki, Robert
   Kotarska, Martyna
   Szymanski, Marcin
   Borawska, Anna
   Swierkowska, Monika
   Dabrowska-Iwanicka, Anna
   Druzd-Sitek, Agnieszka
   Paszkiewicz-Kozik, Ewa
   Mroz-Zycinska, Ewa
   Tajer, Joanna
   Wojciechowska-Lampka, Elzbieta
   Osiadacz, Wlodzimierz
   Rymkiewicz, Grzegorz
   Lapinska, Grazyna
   Wojewodzka-Mirocha, Marta
   Michalski, Wojciech
   Walewski, Jan
TI Safety and efficacy of induction immunochemotherapy with rituximab,
   methotrexate, ifosfamide, and vincristine (R-MIV) in patients with
   primary CNS lymphoma including recent COVID-19 pandemic experience
SO BRITISH JOURNAL OF HAEMATOLOGY
LA English
DT Article
DE COVID-19; HD-MTX; PCNSL; primary CNS lymphoma; remission induction
ID NERVOUS-SYSTEM LYMPHOMA; CHEMOTHERAPY; CYTARABINE
AB Clinical data on primary central nervous system (CNS) lymphoma (PCNSL) patients is mostly generated from prospective studies, and many frail real-world patients are not included. Recently,the diagnosis and treatment of PCNSL patients was confounded by the COVID-19 pandemic. In particular, treatment with high-dose cytarabine was linked to increased risk of pneumonia and virus persistence. We report on outcome of the induction regimen R-MIV (rituximab, methotrexate, ifosfamide, and vincristine) involving intensive administration of high-dose methotrexate (3.5 g/m(2)) with ifosfamide, every 2 weeks and rituximab once per week for six doses. The median age and performance status (PS) for 64 patients was 58 years and 2 (PS 3; 22%) respectively. The overall response rate by magnetic resonance imaging/computed tomography (MRI/CT) was 73% (n = 46/63), with an additional 17.5% (n = 11/63) patients without measurable disease at baseline. Grade 3-4 haematological toxicity was low for R-MIV (neutropenia: 25% and thrombocytopenia: 1%). Three patients (4.7%) died from treatment-related toxicity. Co-existence of SARS-CoV-2 infection with cytomegalovirus reactivation and the varicella-zoster virus in two patients was fatal. Fifty patients (78%) were eligible for consolidation. Median progression-free and overall survival were not reached (median follow-up: 44 months). In conclusion, the R-MIV regimen is feasible in routine practice, effective and safe, even during the COVID-19 pandemic.
C1 [Ostrowska, Beata; Domanska-Czyz, Katarzyna; Romejko-Jarosinska, Joanna; Osowiecki, Michal; Targonski, Lukasz; Poplawska, Lidia; Konecki, Robert; Kotarska, Martyna; Szymanski, Marcin; Borawska, Anna; Swierkowska, Monika; Dabrowska-Iwanicka, Anna; Druzd-Sitek, Agnieszka; Paszkiewicz-Kozik, Ewa; Mroz-Zycinska, Ewa; Tajer, Joanna; Wojciechowska-Lampka, Elzbieta; Osiadacz, Wlodzimierz; Walewski, Jan] Maria Sklodowska Curie Natl Res Inst Oncol, Dept Lymphoid Malignancies, Warsaw, Poland.
   [Rymkiewicz, Grzegorz] Maria Sklodowska Curie Natl Res Inst Oncol, Dept Canc Pathomorphol, Flow Cytometry Lab, Warsaw, Poland.
   [Lapinska, Grazyna; Wojewodzka-Mirocha, Marta] Maria Sklodowska Curie Natl Res Inst Oncol, Dept Oncol Endocrinol & Nucl Med, Warsaw, Poland.
   [Michalski, Wojciech] Maria Sklodowska Curie Natl Res Inst Oncol, Biostat Unit, Warsaw, Poland.
   [Ostrowska, Beata] Maria Sklodowska Curie Natl Res Inst Oncol, Dept Lymphoid Malignancies, WK Roentgena 5, PL-02034 Warsaw, Poland.
RP Ostrowska, B (corresponding author), Maria Sklodowska Curie Natl Res Inst Oncol, Dept Lymphoid Malignancies, WK Roentgena 5, PL-02034 Warsaw, Poland.
EM
RI Rymkiewicz, Grzegorz/AAB-7301-2019
OI Rymkiewicz, Grzegorz/0000-0002-3478-8014; Dabrowska-Iwanicka,
   Anna/0000-0001-8986-0397; Ostrowska, Beata/0000-0001-7318-1031
CR Abrey LE, 2005, J CLIN ONCOL, V23, P5034, DOI 10.1200/JCO.2005.13.524
   Bromberg JEC, 2019, LANCET ONCOL, V20, P216, DOI 10.1016/S1470-2045(18)30747-2
   Calimeriy T, 2021, ESMO OPEN, V6, DOI 10.1016/j.esmoop.2021.100213
   Ferreri AJM, 2022, LEUKEMIA, V36, P1870, DOI 10.1038/s41375-022-01582-5
   Ferreri AJM, 2017, LANCET HAEMATOL, V4, pE510, DOI 10.1016/S2352-3026(17)30174-6
   Ferreri AJM, 2016, LANCET HAEMATOL, V3, pE217, DOI 10.1016/S2352-3026(16)00036-3
   Graham MS, 2018, BEST PRACT RES CL HA, V31, P262, DOI 10.1016/j.beha.2018.07.006
   Gupta Meetakshi, 2019, CNS Oncol, V8, pCNS46, DOI 10.2217/cns-2019-0016
   Gupta T, 2021, CL LYMPH MYELOM LEUK, V21, P497, DOI 10.1016/j.clml.2021.03.011
   Houillier C, 2019, J CLIN ONCOL, V37, P823, DOI 10.1200/JCO.18.00306
   Morris PG, 2013, J CLIN ONCOL, V31, P3971, DOI 10.1200/JCO.2013.50.4910
   Omuro A, 2015, BLOOD, V125, P1403, DOI 10.1182/blood-2014-10-604561
   Ostrowska B, 2013, BLOOD, V122, DOI 10.1182/blood.V122.21.3051.3051
   Pels H, 2003, J CLIN ONCOL, V21, P4489, DOI 10.1200/JCO.2003.04.056
   Rubenstein JL, 2013, J CLIN ONCOL, V31, P3061, DOI 10.1200/JCO.2012.46.9957
   Schmitt AM, 2019, HEMATOL ONCOL, V37, P548, DOI 10.1002/hon.2666
   Schorb E, 2020, BRIT J HAEMATOL, V189, P879, DOI 10.1111/bjh.16451
   Steffanoni S., 2022, NAT MICROBIOL, V00, P1
   Zajdel M, 2019, CANCERS, V11, DOI 10.3390/cancers11111647
NR 19
TC 1
Z9 1
U1 0
U2 2
PU WILEY
PI HOBOKEN
PA 111 RIVER ST, HOBOKEN 07030-5774, NJ USA
SN 0007-1048
EI 1365-2141
J9 BRIT J HAEMATOL
JI Br. J. Haematol.
PD MAY
PY 2023
VL 201
IS 4
BP 663
EP 672
DI 10.1111/bjh.18687
EA FEB 2023
PG 10
WC Hematology
WE Science Citation Index Expanded (SCI-EXPANDED)
SC Hematology
GA F2FO7
UT WOS:000932319600001
PM 36762710
OA hybrid
DA 2024-06-03
ER

PT J
AU Jin, QQ
   Jiang, HY
   Han, Y
   Li, CC
   Zhang, LT
   Zhang, YR
   Chai, Y
   Zeng, PY
   Yue, LL
   Wu, CY
AF Jin, Qiqi
   Jiang, Haoyun
   Han, Ye
   Li, Cuicui
   Zhang, Litian
   Zhang, Yurong
   Chai, Ye
   Zeng, Pengyun
   Yue, Lingling
   Wu, Chongyang
TI Frequent Gene Mutations and Their Possible Roles in the Pathogenesis,
   Treatment and Prognosis of Primary Central Nervous System Lymphoma
SO WORLD NEUROSURGERY
LA English
DT Review
DE CD79B; Gene mutations; Pathogenesis; PCNSL; MYD88
ID PRIMARY CNS LYMPHOMA; NF-KAPPA-B; T-CELL LYMPHOMAS; PHASE-II; IBRUTINIB;
   FEATURES; MYD88; EXPRESSION; RELAPSE; KINASE
AB Primary central nervous system lymphoma (PCNSL) is a rare extranodal non-Hodgkin lymphoma with poor prognosis. In recent years, the emergence of genetic subtypes of systematic diffuse large B-cell lymphoma has highlighted the importance of molecular genetics, but large-scale research on the molecular genetics of PCNSL is lacking. Herein, we summarize the frequent gene mutations and discuss the possible pathogenesis of PCNSL. Myeloid differentiation primary response gene 88 (MYD88) and CD79B mutations, which cause abnormal activation of noncanonical nuclear factor-kB, are prominent genetic abnormalities in PCNSL. They are considered to play a major role in the pathogenesis of PCNSL. Other genes, such as caspase recruitment domain family member 11 (CARD11), tumor necrosis factor alpha induced protein 3 (TNFAIP3), transducin (b)-like 1 X-linked receptor 1, cyclin dependent kinase inhibitor 2A, PR domain zinc finger protein 1, and proviral insertion in murine malignancies 1, are also frequently mutated. Notably, the pathogenesis of immune insufficiencyassociated PCNSL is related to Epstein-Barr virus infection, and its progression may be affected by different signaling pathways. The different mutational patterns in different studies highlight the heterogeneity of PCNSL. However, existing research on the molecular genetics of PCNSL is still limited, and further research into PCNSL is required to clarify the genetic characteristics of PCNSL.
C1 [Jin, Qiqi; Jiang, Haoyun; Li, Cuicui; Zhang, Litian; Zhang, Yurong; Chai, Ye; Zeng, Pengyun; Yue, Lingling; Wu, Chongyang] Lanzhou Univ Second Hosp, Dept Hematol, Lanzhou, Peoples R China.
   [Han, Ye] Xian Cent Hosp, Dept Hematol, Xian, Peoples R China.
RP Wu, CY (corresponding author), Lanzhou Univ Second Hosp, Dept Hematol, Lanzhou, Peoples R China.
EM
FU National Natural Science Foundation of China [8226010026]; Horizontal
   scientific research project of Lanzhou University [(18)0210]; CuiYing
   Science and Technology Innovation" Program of Lanzhou University Second
   Hospital [CY2018-MS07]
FX This work was supported by the National Natural Science Foundation of
   China [grant number 8226010026]; the Horizontal scientific research
   project of Lanzhou University [grant number (18)0210]; and the "CuiYing
   Science and Technology Innovation" Program of Lanzhou University Second
   Hospital [grant number CY2018-MS07].
CR Ambady P, 2019, LEUKEMIA LYMPHOMA, V60, P515, DOI 10.1080/10428194.2018.1480771
   Balint MT, 2016, INT J MOL SCI, V17, DOI 10.3390/ijms17050683
   Barakat M, 2021, CANC TREAT RES COMMU, V27
   Bonzheim I, 2022, BLOOD ADV, V6, P1598, DOI 10.1182/bloodadvances.2021004212
   Braggio E, 2015, CLIN CANCER RES, V21, P3986, DOI 10.1158/1078-0432.CCR-14-2116
   Brennan CW, 2013, CELL, V155, P462, DOI 10.1016/j.cell.2013.09.034
   Bruno A, 2015, ACTA NEUROPATHOL, V130, P439, DOI 10.1007/s00401-015-1461-9
   Bruno A, 2014, ONCOTARGET, V5, P5065, DOI 10.18632/oncotarget.2080
   Chapuy B, 2016, BLOOD, V127, P869, DOI 10.1182/blood-2015-10-673236
   Compagno M, 2009, NATURE, V459, P717, DOI 10.1038/nature07968
   Curran OE, 2021, NEURO-ONCOL ADV, V3, DOI 10.1093/noajnl/vdab090
   Davis RE, 2010, NATURE, V463, P88, DOI 10.1038/nature08638
   Dickson MA, 2014, CLIN CANCER RES, V20, P3379, DOI 10.1158/1078-0432.CCR-13-1551
   Fukumura K, 2016, ACTA NEUROPATHOL, V131, P865, DOI 10.1007/s00401-016-1536-2
   Gandhi MK, 2021, BLOOD, V137, P1468, DOI 10.1182/blood.2020008520
   Ghesquieres H, 2019, ANN ONCOL, V30, P621, DOI 10.1093/annonc/mdz032
   Gonzalez-Aguilar A, 2012, CLIN CANCER RES, V18, P5203, DOI 10.1158/1078-0432.CCR-12-0845
   Grommes C, 2019, NEURO-ONCOLOGY, V21, P306, DOI 10.1093/neuonc/noy193
   Grommes C, 2017, CANCER DISCOV, V7, P1018, DOI 10.1158/2159-8290.CD-17-0613
   Guney E, 2022, BLOOD ADV, V6, P3189, DOI 10.1182/bloodadvances.2021006018
   Hattori K, 2018, CANCER SCI, V109, P225, DOI 10.1111/cas.13450
   Hattori K, 2017, BRIT J HAEMATOL, V177, P492, DOI 10.1111/bjh.14080
   Hiemcke-Jiwa LS, 2018, CRIT REV ONCOL HEMAT, V127, P56, DOI 10.1016/j.critrevonc.2018.05.010
   Honma K, 2009, BLOOD, V114, P2467, DOI 10.1182/blood-2008-12-194852
   Jahnke K, 2006, J NEURO-ONCOL, V80, P159, DOI 10.1007/s11060-006-9165-6
   Kaulen LD, 2021, BRIT J HAEMATOL, V193, P375, DOI 10.1111/bjh.17319
   Kuo HP, 2016, AM J CANCER RES, V6, P2489
   Lenz G, 2008, SCIENCE, V319, P1676, DOI 10.1126/science.1153629
   Lim KH, 2012, IMMUNOL REV, V246, P359, DOI 10.1111/j.1600-065X.2012.01105.x
   Lionakis MS, 2017, CANCER CELL, V31, P833, DOI 10.1016/j.ccell.2017.04.012
   Magnes T, 2021, ESMO OPEN, V6, DOI 10.1016/j.esmoop.2020.100012
   Menon MP, 2015, AM J SURG PATHOL, V39, P1719, DOI 10.1097/PAS.0000000000000503
   Monabati A, 2020, BASIC CLIN NEUROSCI, V11, P491, DOI 10.32598/bcn.11.4.2542.1
   Montesinos-Rongen M, 2004, BLOOD, V103, P1869, DOI 10.1182/blood-2003-05-1465
   Montesinos-Rongen M, 2021, CANCERS, V13, DOI 10.3390/cancers13246334
   Montesinos-Rongen M, 2021, HAEMATOLOGICA, V106, P708, DOI 10.3324/haematol.2019.242701
   Montesinos-Rongen M, 2015, J IMMUNOL, V195, P1312, DOI 10.4049/jimmunol.1402341
   Munch-Petersen HD, 2016, ACTA NEUROPATHOL COM, V4, DOI 10.1186/s40478-016-0307-6
   Nakamura T, 2016, NEUROPATH APPL NEURO, V42, P279, DOI 10.1111/nan.12259
   Narita Y, 2021, NEURO-ONCOLOGY, V23, P122, DOI 10.1093/neuonc/noaa145
   Narlik-Grassow M, 2014, MED RES REV, V34, P136, DOI 10.1002/med.21284
   Nayak L, 2017, BLOOD, V129, P3071, DOI 10.1182/blood-2017-01-764209
   Nayyar N, 2019, BLOOD ADV, V3, P375, DOI 10.1182/bloodadvances.2018027672
   O'Neill BP, 2013, AM J HEMATOL, V88, P997, DOI 10.1002/ajh.23551
   Ondrisova L, 2020, FRONT ONCOL, V10, DOI 10.3389/fonc.2020.591577
   Pedrosa L, 2021, SCI REP-UK, V11, DOI 10.1038/s41598-020-80376-0
   Pels H, 2005, J NEUROL SCI, V228, P143, DOI 10.1016/j.jns.2004.11.038
   Radke J, 2022, NAT COMMUN, V13, DOI 10.1038/s41467-022-30050-y
   Riemersma SA, 2005, J PATHOL, V206, P328, DOI 10.1002/path.1783
   Schaff LR, 2022, BLOOD, V140, P971, DOI 10.1182/blood.2020008377
   Schmitz R, 2018, NEW ENGL J MED, V378, P1396, DOI 10.1056/NEJMoa1801445
   Sethi TK, 2019, LEUKEMIA LYMPHOMA, V60, P2880, DOI 10.1080/10428194.2019.1620942
   Soussain C, 2019, EUR J CANCER, V117, P121, DOI 10.1016/j.ejca.2019.05.024
   Takano S, 2018, WORLD NEUROSURG, V112, pE69, DOI 10.1016/j.wneu.2017.12.028
   Takashima Y, 2019, SCI REP-UK, V9, DOI 10.1038/s41598-019-46473-5
   Takashima Yasuo, 2018, Oncotarget, V9, P27471, DOI 10.18632/oncotarget.25463
   Tun HW, 2018, BLOOD, V132, P2240, DOI 10.1182/blood-2018-02-835496
   Vater I, 2015, LEUKEMIA, V29, P677, DOI 10.1038/leu.2014.264
   Warfel NA, 2015, PHARMACOL THERAPEUT, V151, P41, DOI 10.1016/j.pharmthera.2015.03.001
   Wilson WH, 2015, NAT MED, V21, P922, DOI 10.1038/nm.3884
   Wray-Dutra MN, 2018, J EXP MED, V215, P2445, DOI 10.1084/jem.20180230
   Wright GW, 2020, CANCER CELL, V37, P551, DOI 10.1016/j.ccell.2020.03.015
   Yim J, 2022, AM J SURG PATHOL, V46, P486, DOI 10.1097/PAS.0000000000001859
   Yu L, 2017, BLOOD REV, V31, P77, DOI 10.1016/j.blre.2016.10.001
   Zhang Y, 2020, CHINESE MED J-PEKING, V133, P1462, DOI 10.1097/CM9.0000000000000844
   Zhao Danqing, 2016, Zhonghua Xue Ye Xue Za Zhi, V37, P30, DOI 10.3760/cma.j.issn.0253-2727.2016.01.006
   Zhou JH, 2022, FRONT ONCOL, V12, DOI 10.3389/fonc.2022.824632
   Zhou YY, 2018, NEOPLASIA, V20, P1059, DOI 10.1016/j.neo.2018.08.012
   Zorofchian S, 2018, J NEURO-ONCOL, V140, P509, DOI 10.1007/s11060-018-2990-6
NR 69
TC 1
Z9 1
U1 2
U2 6
PU ELSEVIER SCIENCE INC
PI NEW YORK
PA STE 800, 230 PARK AVE, NEW YORK, NY 10169 USA
SN 1878-8750
EI 1878-8769
J9 WORLD NEUROSURG
JI World Neurosurg.
PD FEB
PY 2023
VL 170
BP 99
EP 106
DI 10.1016/J.wNEu.2022.11.056
EA FEB 2023
PG 8
WC Clinical Neurology; Surgery
WE Science Citation Index Expanded (SCI-EXPANDED)
SC Neurosciences & Neurology; Surgery
GA H0HX1
UT WOS:000992870800001
PM 36396049
DA 2024-06-03
ER

PT J
AU Miao, XL
   Shao, TY
   Wang, YM
   Wang, QJ
   Han, J
   Li, XN
   Li, YX
   Sun, CJ
   Wen, JH
   Liu, JG
AF Miao, Xiuling
   Shao, Tianyu
   Wang, Yaming
   Wang, Qingjun
   Han, Jing
   Li, Xinnan
   Li, Yuxin
   Sun, Chenjing
   Wen, Junhai
   Liu, Jianguo
TI The value of convolutional neural networks-based deep learning model in
   differential diagnosis of space-occupying brain diseases
SO FRONTIERS IN NEUROLOGY
LA English
DT Article
DE convolutional neural network; space-occupying brain lesions; diagnosis;
   differential; magnetic resonance imaging; tumefactive demyelinating
   lesions
ID PRIMARY ANGIITIS; LYMPHOMA; CLASSIFICATION; DEMYELINATION; GUIDELINES
AB ObjectivesIt is still a challenge to differentiate space-occupying brain lesions such as tumefactive demyelinating lesions (TDLs), tumefactive primary angiitis of the central nervous system (TPACNS), primary central nervous system lymphoma (PCNSL), and brain gliomas. Convolutional neural networks (CNNs) have been used to analyze complex medical data and have proven transformative for image-based applications. It can quickly acquire diseases' radiographic features and correct doctors' diagnostic bias to improve diagnostic efficiency and accuracy. The study aimed to assess the value of CNN-based deep learning model in the differential diagnosis of space-occupying brain diseases on MRI. MethodsWe retrospectively analyzed clinical and MRI data from 480 patients with TDLs (n = 116), TPACNS (n = 64), PCNSL (n = 150), and brain gliomas (n = 150). The patients were randomly assigned to training (n = 240), testing (n = 73), calibration (n = 96), and validation (n = 71) groups. And a CNN-implemented deep learning model guided by clinical experts was developed to identify the contrast-enhanced T-1-weighted sequence lesions of these four diseases. We utilized accuracy, sensitivity, specificity, and area under the curve (AUC) to evaluate the performance of the CNN model. The model's performance was then compared to the neuroradiologists' diagnosis. ResultsThe CNN model had a total accuracy of 87% which was higher than senior neuroradiologists (74%), and the AUC of TDLs, PCNSL, TPACNS and gliomas were 0.92, 0.92, 0.89 and 0.88, respectively. ConclusionThe CNN model can accurately identify specific radiographic features of TDLs, TPACNS, PCNSL, and gliomas. It has the potential to be an effective auxiliary diagnostic tool in the clinic, assisting inexperienced clinicians in reducing diagnostic bias and improving diagnostic efficiency.
C1 [Miao, Xiuling; Han, Jing; Liu, Jianguo] South China Univ Technol, Sch Med, Dept Neurol, Guangzhou, Peoples R China.
   [Miao, Xiuling; Li, Xinnan; Li, Yuxin; Sun, Chenjing; Liu, Jianguo] PLA Gen Hosp Beijing, Med Ctr 6, Dept Neurol, Beijing, Peoples R China.
   [Shao, Tianyu; Wen, Junhai] Beijing Inst Technol, Sch Life Sci, Beijing, Peoples R China.
   [Wang, Yaming] Capital Med Univ, Xuanwu Hosp, Dept Neurosurg, Beijing, Peoples R China.
   [Wang, Qingjun] PLA Gen Hosp Beijing, Med Ctr 6, Dept Radiol, Beijing, Peoples R China.
C3 South China University of Technology; Beijing Institute of Technology;
   Capital Medical University
RP Liu, JG (corresponding author), South China Univ Technol, Sch Med, Dept Neurol, Guangzhou, Peoples R China.; Sun, CJ; Liu, JG (corresponding author), PLA Gen Hosp Beijing, Med Ctr 6, Dept Neurol, Beijing, Peoples R China.; Wen, JH (corresponding author), Beijing Inst Technol, Sch Life Sci, Beijing, Peoples R China.
EM
RI Chen, Chao/JHS-6563-2023; Wang, Hao/ABB-8923-2020; wang,
   qingjun/AAB-7599-2020; Wang, Siyi/JNT-2690-2023; Zhang,
   Chi/JSK-0744-2023; Zhao, Xuan/JMR-2135-2023; zhang, jt/JVE-1333-2024;
   Lu, Lu/JPE-5187-2023
OI Wang, Hao/0000-0001-9109-6017; wang, qingjun/0000-0001-8976-3124; 
FU PLA General Hospital Big Data Project [2019MBD-047]
FX Funding This work was supported by the PLA General Hospital Big Data
   Project (2019MBD-047).
CR Akkus Z, 2017, J DIGIT IMAGING, V30, P449, DOI 10.1007/s10278-017-9983-4
   Aneja S, 2019, CURR OPIN NEUROL, V32, P850, DOI 10.1097/WCO.0000000000000761
   [Anonymous], 2007, ACTA NEUROPATHOL, DOI DOI 10.1007/s00401-007-0243-4
   Bajagain Madan, 2020, Surg Neurol Int, V11, P342, DOI 10.25259/SNI_531_2020
   Birnbaum J, 2009, ARCH NEUROL-CHICAGO, V66, P704, DOI 10.1001/archneurol.2009.76
   Chang P, 2018, AM J NEURORADIOL, V39, P1201, DOI 10.3174/ajnr.A5667
   de Boysson H, 2016, STROKE, V47, P2401, DOI 10.1161/STROKEAHA.116.013917
   Faehndrich J, 2011, CLIN NEURORADIOL, V21, P123, DOI 10.1007/s00062-011-0073-6
   Grabowski MM, 2014, J NEUROSURG, V121, P1115, DOI 10.3171/2014.7.JNS132449
   Grommes C, 2017, J CLIN ONCOL, V35, P2410, DOI 10.1200/JCO.2017.72.7602
   Hardy TA, 2019, CURR OPIN NEUROL, V32, P467, DOI 10.1097/WCO.0000000000000683
   He KM, 2016, PROC CVPR IEEE, P770, DOI 10.1109/CVPR.2016.90
   Hoang-Xuan K, 2015, LANCET ONCOL, V16, pE322, DOI 10.1016/S1470-2045(15)00076-5
   Image classification of fine-grained fashion image based on style using pre-trained convolutional neural network, 2018, 2018 IEEE 3 INT C BI, P387
   Jeon HK, 2020, ELECTRONICS-SWITZ, V9, DOI 10.3390/electronics9020311
   Jiang T, 2016, CANCER LETT, V375, P263, DOI 10.1016/j.canlet.2016.01.024
   Krizhevsky A., 2009, Learning multiple layers of features from tiny images
   Krizhevsky A, 2017, COMMUN ACM, V60, P84, DOI 10.1145/3065386
   Kunimatsu A, 2019, MAGN RESON MED SCI, V18, P44, DOI 10.2463/mrms.mp.2017-0178
   Kvarta MD, 2016, BMC NEUROL, V16, DOI 10.1186/s12883-016-0596-1
   LeCun Y, 2015, NATURE, V521, P436, DOI 10.1038/nature14539
   Magadza T, 2021, J IMAGING, V7, DOI 10.3390/jimaging7020019
   Nakayama M, 2021, AM J ROENTGENOL, V217, P186, DOI 10.2214/AJR.20.23226
   Nazir M, 2021, COMPUT MED IMAG GRAP, V91, DOI 10.1016/j.compmedimag.2021.101940
   Ng S, 2007, J CLIN NEUROSCI, V14, P1126, DOI 10.1016/j.jocn.2006.05.003
   Nguyen AV, 2018, NEUROSURG FOCUS, V45, DOI 10.3171/2018.8.FOCUS18325
   Ostrom QT, 2020, NEURO-ONCOLOGY, V22, P1, DOI 10.1093/neuonc/noaa200
   Paoletti M, 2021, RADIOL MED, V126, P827, DOI 10.1007/s11547-021-01334-y
   Qi XK, 2017, CHINESE MED J-PEKING, V130, P1838, DOI 10.4103/0366-6999.211547
   Ryu YJ, 2014, PLOS ONE, V9, DOI 10.1371/journal.pone.0108335
   Suh CH, 2018, AM J NEURORADIOL, V39, P1643, DOI 10.3174/ajnr.A5775
   Suh CH, 2019, J MAGN RESON IMAGING, V50, P560, DOI 10.1002/jmri.26602
   Xia W, 2021, J MAGN RESON IMAGING, V54, P880, DOI 10.1002/jmri.27592
   Yang ZJ, 2017, CNS NEUROL DISORD-DR, V16, P160, DOI 10.2174/1871527315666161018122909
   Yaqub M, 2020, BRAIN SCI, V10, DOI 10.3390/brainsci10070427
   Zaharchuk G, 2018, AM J NEURORADIOL, V39, P1776, DOI 10.3174/ajnr.A5543
   Zhang M, 2020, J MAGN RESON IMAGING, V52, P1227, DOI 10.1002/jmri.27129
   Zhang Y, 2021, FRONT ONCOL, V11, DOI 10.3389/fonc.2021.665891
   Zia R, 2018, INT J IMAG SYST TECH, V28, P153, DOI 10.1002/ima.22266
NR 39
TC 1
Z9 1
U1 2
U2 5
PU FRONTIERS MEDIA SA
PI LAUSANNE
PA AVENUE DU TRIBUNAL FEDERAL 34, LAUSANNE, CH-1015, SWITZERLAND
SN 1664-2295
J9 FRONT NEUROL
JI Front. Neurol.
PD FEB 2
PY 2023
VL 14
AR 1107957
DI 10.3389/fneur.2023.1107957
PG 8
WC Clinical Neurology; Neurosciences
WE Science Citation Index Expanded (SCI-EXPANDED)
SC Neurosciences & Neurology
GA 9P8OA
UT WOS:000944536600001
PM 36816568
OA gold, Green Published
DA 2024-06-03
ER

PT J
AU Feng, L
   Gao, XH
   Jiao, ZY
   Wang, Z
   Min, FL
AF Feng, Lan
   Gao, Xiaohui
   Jiao, Zhiyun
   Wang, Zheng
   Min, Fenglin
TI BTK inhibitor combined with anti-PD-1 monoclonal antibody for the
   treatment of CD20-negative primary central nervous system lymphoma: A
   case report
SO ONCOLOGY LETTERS
LA English
DT Article
DE CD20-negative diffuse large B cell lymphoma; primary central nervous
   system lymphoma; Bruton tyrosine kinase inhibitor; programmed cell death
   protein 1-1 inhibitor
ID BRAIN METASTASES; CD20 EXPRESSION; IBRUTINIB; MECHANISMS; NIVOLUMAB;
   BLOCKADE
AB CD20-negative diffuse large B-cell lymphoma (DLBCL) is a rare type of lymphoproliferative disorder characterized by a high degree of aggressiveness, a tendency for extranodal invasion and chemotherapeutic resistance. CD20-negative DLBCL originating from the nervous system is rarer. In primary central nervous system lymphoma (PCNSL), >90% of cases are histologically classified as DLBCL. The present study reports the case of a 65-year-old female with CD20-negative PCNSL, whose primary clinical symptom was a persistent headache. Serum tests for human immunodeficiency virus, Epstein-Barr virus-DNA, human herpesvirus 8, hepatitis B and hepatitis C were negative. Cranial magnetic resonance imaging suggested multiple intracranial occupancies. The neoplastic cells were found to be positive for CD19, CD79 alpha, Bcl-2 (similar to 92%) and c-Myc (similar to 50%), while showing negative results for CD20, CD138, programmed cell death protein 1 (PD-1) and programmed cell death receptor 1 ligand 1 (PD-L1). The Ki-67 proliferation index was >80%. In the tumor microenvironment, <10% of the tumor-associated macrophages expressed PD-L1. The number of PD-1-positive tumor-infiltrating lymphocytes was 30-40 cells according to high-power field microscopy. The patient's disease progressed during methotrexate-based treatment, leading to a change in the treatment regimen to the Bruton tyrosine kinase inhibitor, zanubrutinib, combined with the anti-PD-1 monoclonal antibody tislelizumab. After two courses of the combined treatment, the patient achieved complete remission (CR) and continued to receive consolidation treatment. In the 20 months of follow-up since CR was achieved, the patient's general condition was good and the disease was in continuous remission. The present case report and literature review show that a combination of drugs targeting different mechanisms may be used to treat PCNSL to prolong patient survival time. The mechanism of the enhanced efficacy of a combination of the two drugs may be related to the enhancement of antitumor T-cell immune responses and reversal of T-cell immune metabolic dysfunctions by the inhibition of glycolysis.
C1 [Feng, Lan; Gao, Xiaohui] Yangzhou Univ, Affiliated Hosp, Dept Hematol, Yangzhou 225009, Jiangsu, Peoples R China.
   [Jiao, Zhiyun] Yangzhou Univ, Affiliated Hosp, Dept Radiol, Yangzhou 225009, Jiangsu, Peoples R China.
   [Wang, Zheng] Yangzhou Univ, Affiliated Hosp, Dept Pathol, Yangzhou 225009, Jiangsu, Peoples R China.
   [Min, Fenglin] Yangzhou Hosp Tradit Chinese Med, Dept Hematol, Yangzhou 225012, Jiangsu, Peoples R China.
   [Min, Fenglin] Yangzhou Hosp Tradit Chinese Med, Dept Hematol, 575 Wengchang Middle Rd, Yangzhou 225012, Jiangsu, Peoples R China.
C3 Yangzhou University; Yangzhou University; Yangzhou University
RP Min, FL (corresponding author), Yangzhou Hosp Tradit Chinese Med, Dept Hematol, 575 Wengchang Middle Rd, Yangzhou 225012, Jiangsu, Peoples R China.
EM
RI GAO, XIAO/JED-3257-2023
CR Ansell SM, 2019, J CLIN ONCOL, V37, P481, DOI 10.1200/JCO.18.00766
   Chang CH, 2015, CELL, V162, P1229, DOI 10.1016/j.cell.2015.08.016
   Chihara D, 2021, CL LYMPH MYELOM LEUK, V21, P73, DOI 10.1016/j.clml.2020.10.015
   Devi K, 2021, OXF MED CASE REP, P447, DOI 10.1093/omcr/omab114
   Eguren-Santamaria I, 2020, CLIN CANCER RES, V26, P4186, DOI 10.1158/1078-0432.CCR-20-0798
   Gamonet C, 2016, EXP HEMATOL ONCOL, V5, DOI 10.1186/s40164-016-0036-3
   Goldwirt L, 2018, CANCER CHEMOTH PHARM, V81, P783, DOI 10.1007/s00280-018-3546-3
   Grommes C, 2017, CANCER DISCOV, V7, P1018, DOI 10.1158/2159-8290.CD-17-0613
   Hanna BS, 2021, HAEMATOLOGICA, V106, P968, DOI 10.3324/haematol.2019.238154
   Katchi T, 2017, BIOMARK RES, V5, DOI 10.1186/s40364-017-0088-5
   Kim KH, 2019, CLIN CANCER RES, V25, P2144, DOI 10.1158/1078-0432.CCR-18-1449
   Lesokhin AM, 2016, J CLIN ONCOL, V34, P2698, DOI 10.1200/JCO.2015.65.9789
   Lionakis MS, 2017, CANCER CELL, V31, P833, DOI 10.1016/j.ccell.2017.04.012
   Nayak L, 2017, BLOOD, V129, P3071, DOI 10.1182/blood-2017-01-764209
   Overman M, 2020, J IMMUNOTHER CANCER, V8, DOI 10.1136/jitc-2020-000587
   Pavlasova G, 2020, HAEMATOLOGICA, V105, P1494, DOI 10.3324/haematol.2019.243543
   Qorraj M, 2017, LEUKEMIA, V31, P470, DOI 10.1038/leu.2016.214
   Qunaj L, 2016, BLOOD, V128, DOI 10.1182/blood.V128.22.4226.4226
   Rubenstein JL, 2017, HEMATOL-AM SOC HEMAT, P556, DOI 10.1182/asheducation-2017.1.556
   Rushton CK, 2020, BLOOD ADV, V4, P2886, DOI 10.1182/bloodadvances.2020001696
   Sagiv-Barfi I, 2015, P NATL ACAD SCI USA, V112, pE966, DOI 10.1073/pnas.1500712112
   Soussain C, 2019, EUR J CANCER, V117, P121, DOI 10.1016/j.ejca.2019.05.024
   Taggart D, 2018, P NATL ACAD SCI USA, V115, pE1540, DOI 10.1073/pnas.1714089115
   Taylor MH, 2022, CLIN CANCER RES, V28, P903, DOI 10.1158/1078-0432.CCR-21-2547
   Terui Y, 2009, CLIN CANCER RES, V15, P2523, DOI 10.1158/1078-0432.CCR-08-1403
   Tomita A, 2016, J CLIN EXP HEMATOP, V56, P89
   Wu JJ, 2022, INVEST NEW DRUG, V40, P650, DOI 10.1007/s10637-022-01219-5
   Zhang T, 2020, CANCER-AM CANCER SOC, V126, P4485, DOI 10.1002/cncr.33067
   Zhang Y., 2022, P EHA2022, pS224
   Zhang Y, 2021, FRONT ONCOL, V11, DOI 10.3389/fonc.2021.760405
   Zhou SJ, 2021, CANCER LETT, V502, P166, DOI 10.1016/j.canlet.2020.12.043
NR 31
TC 1
Z9 1
U1 0
U2 9
PU SPANDIDOS PUBL LTD
PI ATHENS
PA POB 18179, ATHENS, 116 10, GREECE
SN 1792-1074
EI 1792-1082
J9 ONCOL LETT
JI Oncol. Lett.
PD FEB
PY 2023
VL 25
IS 2
AR 48
DI 10.3892/ol.2022.13634
PG 6
WC Oncology
WE Science Citation Index Expanded (SCI-EXPANDED)
SC Oncology
GA 7N4AH
UT WOS:000907283200001
PM 36644138
OA Green Published, gold
DA 2024-06-03
ER

PT J
AU Min, GJ
   Kim, TY
   Jeon, YW
   Hyun, JO
   Choi, BO
   Park, G
   Park, YH
   Cho, SG
AF Min, Gi-June
   Kim, Tong Yoon
   Jeon, Young-Woo
   Hyun, Joo O.
   Choi, Byung-Ock
   Park, Gyeongsin
   Park, Young-Hoon
   Cho, Seok-Goo
TI Diagnosis, treatment, and prognosis of primary intraocular lymphoma:
   Single-center real-world clinical experience
SO CANCER MEDICINE
LA English
DT Article
DE CNS relapse; local intravitreal methotrexate; primary intraocular
   lymphoma; systemic high-dose methotrexate
ID NERVOUS-SYSTEM LYMPHOMA; FEATURES; INTERLEUKIN-10; METHOTREXATE;
   RITUXIMAB; FLUID
AB Background: The diagnosis and management of primary intraocular lymphoma (PIOL) remain challenging. This study identified factors indicative of PIOL, described treatment outcomes, and determined modalities to prevent relapse.Methods: We included 21 PIOL-diagnosed patients, seven via cytology, 12 via genetic evaluation, and two via interleukin (IL) level measurements, who underwent vitrectomy and received local intravitreal methotrexate (IV- MTX) injection. Clinical outcomes, including treatment response and relapse, were compared between patients receiving IV- MTX alone (n = 13) or IV- MTX with systemic high dose methotrexate (HD- MTX) as prophylaxis (n = 8).Results: Twelve ophthalmologic and eight central nervous system (CNS) relapse cases within a median of 20.3 and 11.6 months were shown, regardless of the treatment modalities, with a median progression free survival of 21.3 (95% confidence interval, 9.5- 36.7) months. There was no difference in demographic characteristics between the two groups, except with the poorer performance status in patients in the HD- MTX prophylaxis group. Furthermore, patients demonstrated rapid elevations in the vitreous fluid IL-10/IL- 6 cytokine ratio before ophthalmologic and CNS relapse. Therefore, diagnosis should be based on clinical signs and assisted by vitrectomy, cytologic, molecular, and cytokine studies.Conclusion: For PIOL, aggressive systemic treatment equivalent to that of primary CNS lymphoma (PCNSL) is recommended because solely HD- MTX did not prevent or delay CNS relapse. To prevent PIOL relapse in the CNS efficiently, prospective trials with large numbers of patients and advanced therapeutic regimens are necessary. Furthermore, regular clinical follow up is crucial, and the IL 10/ IL 6 ratio can help evaluate relapse promptly.
C1 [Min, Gi-June; Kim, Tong Yoon; Cho, Seok-Goo] Catholic Univ Korea, Catholic Univ Lymphoma Grp, Seoul St Marys Hosp, Coll Med,Dept Hematol, Seoul, South Korea.
   [Jeon, Young-Woo] Catholic Univ Korea, Catholic Univ Lymphoma Grp, Yeouido St Marys Hosp, Coll Med,Dept Hematol, Seoul, South Korea.
   [Hyun, Joo O.] Catholic Univ Korea, Catholic Univ Lymphoma Grp, Seoul St Marys Hosp, Coll Med,Dept Nucl Med, Seoul, South Korea.
   [Choi, Byung-Ock] Catholic Univ Korea, Catholic Univ Lymphoma Grp, Seoul St Marys Hosp, Coll Med,Dept Radiat Oncol, Seoul, South Korea.
   [Park, Gyeongsin] Catholic Univ Korea, Catholic Univ Lymphoma Grp, Dept Hosp Pathol, Seoul St Marys Hosp,Coll Med, Seoul, South Korea.
   [Park, Young-Hoon] Catholic Univ Korea, Catholic Univ Lymphoma Grp, Seoul St Marys Hosp, Coll Med,Dept Ophthalmol & Visual Sci, Seoul, South Korea.
   [Park, Young-Hoon] Catholic Univ Korea, Catholic Univ Lymphoma Grp, Seoul St Marys Hosp, Coll Med,Dept Ophthalmol & Visual Sci, Banpo Daero 222, Seoul, South Korea.
   [Cho, Seok-Goo] Catholic Univ Korea, Catholic Univ Lymphoma Grp, Seoul St Marys Hosp, Coll Med,Dept Hematol, Banpo Daero 222, Seoul, South Korea.
C3 Seoul St. Mary's Hospital; Catholic University of Korea; Catholic
   University of Korea; Seoul St. Mary's Hospital; Catholic University of
   Korea; Seoul St. Mary's Hospital; Catholic University of Korea; Catholic
   University of Korea; Seoul St. Mary's Hospital; Catholic University of
   Korea; Seoul St. Mary's Hospital; Seoul St. Mary's Hospital; Catholic
   University of Korea; Seoul St. Mary's Hospital; Catholic University of
   Korea
RP Park, YH (corresponding author), Catholic Univ Korea, Catholic Univ Lymphoma Grp, Seoul St Marys Hosp, Coll Med,Dept Ophthalmol & Visual Sci, Banpo Daero 222, Seoul, South Korea.; Cho, SG (corresponding author), Catholic Univ Korea, Catholic Univ Lymphoma Grp, Seoul St Marys Hosp, Coll Med,Dept Hematol, Banpo Daero 222, Seoul, South Korea.
EM
OI Jeon, Youngwoo/0000-0003-3362-8200; Min, Gi June/0000-0001-7636-4693;
   Cho, Seok-Goo/0000-0002-5429-4839
CR Abrey LE, 2005, J CLIN ONCOL, V23, P5034, DOI 10.1200/JCO.2005.13.524
   Augsburger J J, 1989, Trans Pa Acad Ophthalmol Otolaryngol, V41, P796
   Calimeriy T, 2021, ESMO OPEN, V6, DOI 10.1016/j.esmoop.2021.100213
   Cassoux N, 2000, OCUL IMMUNOL INFLAMM, V8, P243, DOI 10.1076/ocii.8.4.243.6463
   CHAN CC, 1995, AM J OPHTHALMOL, V120, P671, DOI 10.1016/S0002-9394(14)72217-2
   Chan Chi-Chao, 2002, Curr Opin Ophthalmol, V13, P411, DOI 10.1097/00055735-200212000-00012
   Coupland SE, 2004, GRAEF ARCH CLIN EXP, V242, P901, DOI 10.1007/s00417-004-0973-0
   Coupland SE, 2003, GRAEF ARCH CLIN EXP, V241, P860, DOI 10.1007/s00417-003-0749-y
   FREEMAN LN, 1987, OPHTHALMOLOGY, V94, P1631
   Frenkel S, 2008, BRIT J OPHTHALMOL, V92, P383, DOI 10.1136/bjo.2007.127928
   Gill MK, 2001, SURV OPHTHALMOL, V45, P463, DOI 10.1016/S0039-6257(01)00217-X
   HERSHEY JM, 1994, OPHTHALMOLOGY, V101, P596
   HOCHBERG FH, 1988, J NEUROSURG, V68, P835, DOI 10.3171/jns.1988.68.6.0835
   Hormigo A, 2004, BRIT J HAEMATOL, V126, P202, DOI 10.1111/j.1365-2141.2004.05028.x
   Kim H, 2006, EXP EYE RES, V82, P760, DOI 10.1016/j.exer.2005.09.018
   Kimura K, 2012, JPN J OPHTHALMOL, V56, P383, DOI 10.1007/s10384-012-0150-7
   Kitzmann AS, 2007, EYE, V21, P1524, DOI 10.1038/sj.eye.6702804
   Lam M, 2021, AM J HEMATOL, V96, P823, DOI 10.1002/ajh.26199
   Matsuo T, 1998, RETINA-J RET VIT DIS, V18, P301
   Matsuo T, 2019, J CLIN EXP HEMATOP, V59, P168, DOI 10.3960/jslrt.19019
   Mendez Joe S, 2018, Am Soc Clin Oncol Educ Book, V38, P604, DOI 10.1200/EDBK_200829
   Omuro A, 2015, BLOOD, V125, P1403, DOI 10.1182/blood-2014-10-604561
   PETERSON K, 1993, CANCER, V72, P843, DOI 10.1002/1097-0142(19930801)72:3<843::AID-CNCR2820720333>3.0.CO;2-#
   Sagoo MS, 2014, SURV OPHTHALMOL, V59, P503, DOI 10.1016/j.survophthal.2013.12.001
   Sasayama T, 2012, NEURO-ONCOLOGY, V14, P368, DOI 10.1093/neuonc/nor203
   WHITCUP SM, 1993, OPHTHALMOLOGY, V100, P1399
   Wolf LA, 2003, OPHTHALMOLOGY, V110, P1671, DOI 10.1016/S0161-6420(03)00811-X
   Yeh S, 2010, EYE, V24, P1625, DOI 10.1038/eye.2010.111
NR 28
TC 1
Z9 2
U1 0
U2 1
PU WILEY
PI HOBOKEN
PA 111 RIVER ST, HOBOKEN 07030-5774, NJ USA
SN 2045-7634
J9 CANCER MED-US
JI Cancer Med.
PD APR
PY 2023
VL 12
IS 7
BP 7911
EP 7922
DI 10.1002/cam4.5567
EA JAN 2023
PG 12
WC Oncology
WE Science Citation Index Expanded (SCI-EXPANDED)
SC Oncology
GA E3CZ7
UT WOS:000924035200001
PM 36721307
OA Green Accepted, gold
DA 2024-06-03
ER

PT J
AU Stifano, V
   Della Pepa, GM
   Offi, M
   Montano, N
   Carcagni, A
   Pallini, R
   Lauretti, L
   Olivi, A
   D'Alessandris, QG
AF Stifano, Vito
   Della Pepa, Giuseppe M.
   Offi, Martina
   Montano, Nicola
   Carcagni, Antonella
   Pallini, Roberto
   Lauretti, Liverana
   Olivi, Alessandro
   D'Alessandris, Quintino Giorgio
TI Resection versus biopsy for management of primary central nervous system
   lymphoma: a meta-analysis
SO NEUROSURGICAL REVIEW
LA English
DT Review
DE primary central nervous system lymphoma; PCNSL; resection; surgery;
   biopsy
ID PRIMARY CNS LYMPHOMA; SURGERY; GUIDELINES; DIAGNOSIS
AB The role of surgery in the management of primary central nervous system lymphomas (PCNSL) is currently confined to diagnosis. However, over recent years, an increasing number of papers have suggested a possible positive prognostic impact of surgery in selected cases. The present work aims to perform a meta-analysis of the available literature evidence. A meta-analysis with meta-regression on the role of surgical resection compared to biopsy in the management of PCNSL was conducted according to the PRISMA statement, searching MEDLINE via PubMed and Embase. The random effect model was used. The quality of evidence was assessed using the GRADE framework. After screening 1395 records, we included 11 papers in our analysis. Patients who underwent surgical resection harbored superficial and single-lesion tumors. At 1-, 2-, and 5-year follow-up, progression-free survival did not differ between the two groups, while overall survival favored resection, even if in a non-significant fashion. Meta-regression analysis showed that the overall survival rate at 2 years, but not at 1 or 5 years, was significantly influenced by tumor location. There were no differences in terms of age, sex, Karnofsky performance status, adjuvant therapy, or procedure-related complications. Overall, the quality of evidence is low. The results of the present meta-analysis do not change the current standard of care for PCNSL. However, surgery could be non-inferior to biopsy with an acceptable risk profile in selected patients harboring single and superficial lesions. The low quality of evidence prompts future randomized studies.
C1 [Stifano, Vito; Della Pepa, Giuseppe M.; Offi, Martina; Montano, Nicola; Pallini, Roberto; Lauretti, Liverana; Olivi, Alessandro; D'Alessandris, Quintino Giorgio] Fdn Policlin Univ A Gemelli IRCCS, Dept Neurosurg, Largo A Gemelli 8, I-00168 Rome, Italy.
   [Carcagni, Antonella] Fdn Policlin Univ A Gemelli IRCCS, Facil Epidemiol & Biostat Gemelli Generator, Largo A Gemelli 8, I-00168 Rome, Italy.
C3 Catholic University of the Sacred Heart; IRCCS Policlinico Gemelli;
   Catholic University of the Sacred Heart; IRCCS Policlinico Gemelli
RP Lauretti, L (corresponding author), Fdn Policlin Univ A Gemelli IRCCS, Dept Neurosurg, Largo A Gemelli 8, I-00168 Rome, Italy.
EM
RI Stifano, Vito/I-9032-2019; Montano, Nicola/M-4565-2016
OI Stifano, Vito/0000-0002-5509-9592; 
CR Abrey LE, 2006, J CLIN ONCOL, V24, P5711, DOI 10.1200/JCO.2006.08.2941
   Balduzzi S, 2019, EVID-BASED MENT HEAL, V22, P153, DOI 10.1136/ebmental-2019-300117
   Balshem H, 2011, J CLIN EPIDEMIOL, V64, P401, DOI 10.1016/j.jclinepi.2010.07.015
   Bataille B, 2000, J NEUROSURG, V92, P261, DOI 10.3171/jns.2000.92.2.0261
   Batchelor TT, 2019, HEMATOL ONCOL, V37, P15, DOI 10.1002/hon.2598
   Chamberlain MC, 2010, NEURO-ONCOLOGY, V12, P736, DOI 10.1093/neuonc/noq011
   Correia CE, 2020, CANCER J, V26, P241, DOI 10.1097/PPO.0000000000000449
   Di Bonaventura R, 2021, J PERS MED, V11, DOI 10.3390/jpm11090909
   Ferreri AJM, 2003, J CLIN ONCOL, V21, P266, DOI 10.1200/JCO.2003.09.139
   Villalonga JF, 2018, ARQ NEURO-PSIQUIAT, V76, P139, DOI [10.1590/0004-282X20180002, 10.1590/0004-282x20180002]
   Gelabert-González M, 2013, NEUROLOGIA, V28, P283, DOI 10.1016/j.nrl.2012.04.005
   Guyatt GH, 2011, J CLIN EPIDEMIOL, V64, P1283, DOI 10.1016/j.jclinepi.2011.01.012
   Hoang-Xuan K, 2015, LANCET ONCOL, V16, pE322, DOI 10.1016/S1470-2045(15)00076-5
   Jahr G, 2018, NEUROSURG REV, V41, P1037, DOI 10.1007/s10143-018-0946-0
   Kim DG, 1996, ACTA NEUROCHIR, V138, P280, DOI 10.1007/BF01411738
   Liu CJ, 2020, CANCER MED-US, V9, P2134, DOI 10.1002/cam4.2872
   Moher D, 2009, BMJ-BRIT MED J, V339, DOI [10.1136/bmj.b2535, 10.1186/2046-4053-4-1, 10.1136/bmj.b2700, 10.1016/j.ijsu.2010.07.299, 10.1371/journal.pmed.1000097, 10.1016/j.ijsu.2010.02.007, 10.1136/bmj.i4086]
   Morris PG, 2009, LANCET NEUROL, V8, P581, DOI 10.1016/S1474-4422(09)70091-2
   National Comprehensive Cancer Network, 2022, NCCN Clinical Practice Guidelines in Oncology, Myeloproliferative Neoplasms
   Olson JE, 2002, CANCER, V95, P1504, DOI 10.1002/cncr.10851
   Ouyang TH, 2020, WORLD NEUROSURG, V139, pE508, DOI 10.1016/j.wneu.2020.04.049
   Rae AI, 2019, NEUROSURGERY, V84, P935, DOI 10.1093/neuros/nyy096
   Schellekes N, 2021, J NEUROSURG, V135, P1016, DOI 10.3171/2020.9.JNS201980
   Singhal D, 2002, CAN J NEUROL SCI, V29, P147, DOI 10.1017/S0317167100120918
   Tseng MY, 1998, J CLIN NEUROSCI, V5, P409, DOI 10.1016/S0967-5868(98)90273-9
   Weller M, 2012, NEURO-ONCOLOGY, V14, P1481, DOI 10.1093/neuonc/nos159
   WHO Classification of Tumours Editorial Board, WHO Classification of Tumours Series
   Wu SQ, 2021, BMC NEUROL, V21, DOI 10.1186/s12883-021-02227-3
NR 28
TC 1
Z9 1
U1 1
U2 2
PU SPRINGER
PI NEW YORK
PA ONE NEW YORK PLAZA, SUITE 4600, NEW YORK, NY, UNITED STATES
SN 0344-5607
EI 1437-2320
J9 NEUROSURG REV
JI Neurosurg. Rev.
PD JAN 16
PY 2023
VL 46
IS 1
AR 37
DI 10.1007/s10143-022-01931-z
PG 9
WC Clinical Neurology; Surgery
WE Science Citation Index Expanded (SCI-EXPANDED)
SC Neurosciences & Neurology; Surgery
GA 7X7LX
UT WOS:000914380300001
PM 36645525
DA 2024-06-03
ER

PT J
AU Chen, JM
   Sun, L
   Dai, YY
   Zhang, L
   Yang, K
   Han, XY
   Ding, XH
   Gao, HL
   Zhou, XG
   Wang, P
AF Chen, Jiamin
   Sun, Lei
   Dai, Yuyang
   Zhang, Liang
   Yang, Kun
   Han, Xiaoyi
   Ding, Xinghuan
   Gao, Haili
   Zhou, Xingang
   Wang, Peng
TI Clinical pathology of primary central nervous system lymphoma in
   HIV-positive patients-a 41 Chinese patients retrospective study
SO ANNALS OF DIAGNOSTIC PATHOLOGY
LA English
DT Article
DE Primary central nervous system lymphoma (PCNSL); HIV; Clinical pathology
ID EPSTEIN-BARR-VIRUS; NO EVIDENCE; PATHOGENESIS; POLYMORPHISM; SURVIVAL;
   EBV
AB Objectives: The purpose of this study was to investigate the clinicopathological characteristics of primary central nervous system lymphoma (PCNSL). Methods: We collected 41 PCNSL formalin-fixed, paraffin-embedded (FFPE) samples from human immunodefi-ciency virus (HIV)-positive patients and performed HE (haematoxylin-eosin) staining, immunohistochemistry (IHC) staining, in situ hybridization, fluorescence in situ hybridization (FISH). Real-time quantitative polymerase chain reaction (RT-qPCR) was performed in 9 cases of FFPE samples. Meanwhile, we analysed the clinical pathological significance of the results. Results: Seven patients had diffuse large B-cell lymphoma (DLBCL) with germinal centre B-cell (GCB)-like DLBCL, 32 had activated B-cell (ABC)-like DLBCL, and 2 had Burkitt lymphoma (BL). GCB-like DLBCL patients were older at onset (P = 0.040).A lower CD4+ T-cell count and a decrease in cerebrospinal fluid (CSF) glucose content were more frequent in ABC-like DLBCL (P = 0.012, P = 0.006). Overexpression of P53 was more in ABC-like DLBCL (P = 0.041). 73.2 % cases were Epstein-Barr encoding region (EBER) positive, which was more likely in ABC-like DLBCL patients (P = 0.037). EBV DNA were detected in 5/7 EBER-negative DLBCL cases and none (0/2) of the BL cases. All the cases were negative for HHV8 staining. None of the 7 Double expressor lymphoma (DEL) cases had BCL2, BCL6, or c-MYC genetic rearrangements. Conclusions: HIV-related PCNSL showed unique clinical pathological significance. None of EBV detected in HIV -related BL and without HHV8 infectious are new sights in our single-center study of Chinese HIV-related PCNSL patients.
C1 [Chen, Jiamin; Sun, Lei; Zhang, Liang; Yang, Kun; Han, Xiaoyi; Gao, Haili; Zhou, Xingang; Wang, Peng] Capital Med Univ, Beijing Ditan Hosp, Dept Pathol, Beijing 100015, Peoples R China.
   [Dai, Yuyang] Capital Med Univ, Beijing Tongren Hosp, Natl Inst Drug Clin Trial, Beijing 100730, Peoples R China.
   [Ding, Xinghuan] Capital Med Univ, Beijing Ditan Hosp, Dept Neurosurg, Beijing 100015, Peoples R China.
C3 Capital Medical University; Capital Medical University; Capital Medical
   University
RP Wang, P (corresponding author), Capital Med Univ, Beijing Ditan Hosp, Dept Pathol, Beijing 100015, Peoples R China.
EM
RI chen, jia/JDW-7660-2023; chen, jiayi/KHV-5520-2024
FU Beijing Ditan Hospital "Yu Miao" Program [DTYM 202103]
FX Funding This work is supported by grant from Beijing Ditan Hospital "Yu
   Miao" Program (DTYM 202103) .
CR Alaggio R, 2022, LEUKEMIA, V36, P1720, DOI 10.1038/s41375-022-01620-2
   Batchelor TT, 2019, HEMATOL ONCOL, V37, P15, DOI 10.1002/hon.2598
   Bayraktar S, 2011, J NEURO-ONCOL, V101, P257, DOI 10.1007/s11060-010-0252-3
   Brandsma Dieta, 2018, Handb Clin Neurol, V152, P177, DOI 10.1016/B978-0-444-63849-6.00014-1
   Cote TR, 1996, J NATL CANCER I, V88, P675, DOI 10.1093/jnci/88.10.675
   Dolcetti R, 2013, SEMIN CANCER BIOL, V23, P441, DOI 10.1016/j.semcancer.2013.07.005
   Franca RA, 2020, PATHOL RES PRACT, V216, DOI 10.1016/j.prp.2020.153192
   Gallagher A, 2003, INT J CANCER, V104, P624, DOI 10.1002/ijc.10979
   Gloghini A, 2013, SEMIN CANCER BIOL, V23, P457, DOI 10.1016/j.semcancer.2013.08.004
   Grommes C, 2017, J CLIN ONCOL, V35, P2410, DOI 10.1200/JCO.2017.72.7602
   Gupta NK, 2017, NEURO-ONCOLOGY, V19, P99, DOI 10.1093/neuonc/now155
   Haldorsen IS, 2008, BMC CANCER, V8, DOI 10.1186/1471-2407-8-225
   He Xiao-feng, 2011, Nan Fang Yi Ke Da Xue Xue Bao, V31, P791
   Kaulen LD, 2019, J NEURO-ONCOL, V144, P107, DOI 10.1007/s11060-019-03208-w
   Li H, 2013, INFECT GENET EVOL, V20, P163, DOI 10.1016/j.meegid.2013.09.010
   Lin YJ, 2013, PLOS ONE, V8, DOI 10.1371/journal.pone.0067572
   Lu Y, 2015, AIDS RES HUM RETROV, V31, P525, DOI [10.1089/aid.2014.0303, 10.1089/AID.2014.0303]
   MACMAHON EME, 1991, LANCET, V338, P969, DOI 10.1016/0140-6736(91)91837-K
   Montesinos-Rongen M, 2001, ACTA NEUROPATHOL, V102, P489
   Mundo L, 2020, MODERN PATHOL, V33, P2407, DOI 10.1038/s41379-020-0575-3
   Mundo L, 2017, FRONT MICROBIOL, V8, DOI 10.3389/fmicb.2017.00229
   Newell ME, 2004, CANCER-AM CANCER SOC, V100, P2627, DOI 10.1002/cncr.20300
   Niller HH, 2017, METHODS MOL BIOL, V1532, P33, DOI 10.1007/978-1-4939-6655-4_2
   Phillips EH, 2014, CURR HEMATOL MALIG R, V9, P243, DOI 10.1007/s11899-014-0217-2
   Qiao YC, 2019, INT J INFECT DIS, V81, P215, DOI 10.1016/j.ijid.2019.02.016
   Roe CJ, 2019, APPL IMMUNOHISTO M M, V27, P155, DOI 10.1097/PAI.0000000000000568
   Schaff LR, 2022, BLOOD, V140, P971, DOI 10.1182/blood.2020008377
   Staratschek-Jox A, 2000, AM J PATHOL, V156, P209, DOI 10.1016/S0002-9440(10)64721-9
   Villano JL, 2011, BRIT J CANCER, V105, P1414, DOI 10.1038/bjc.2011.357
   Wang Hui, 2006, Zhonghua Yixue Yichuanxue Zazhi, V23, P466
NR 30
TC 1
Z9 1
U1 0
U2 2
PU ELSEVIER SCIENCE INC
PI NEW YORK
PA STE 800, 230 PARK AVE, NEW YORK, NY 10169 USA
SN 1092-9134
EI 1532-8198
J9 ANN DIAGN PATHOL
JI Ann. Diagn. Pathol.
PD APR
PY 2023
VL 63
AR 152108
DI 10.1016/j.anndiagpath.2023.152108
EA JAN 2023
PG 7
WC Pathology
WE Science Citation Index Expanded (SCI-EXPANDED)
SC Pathology
GA H1NP3
UT WOS:000993701800001
PM 36638601
DA 2024-06-03
ER

PT J
AU Miyao, K
   Yokota, H
   Sakemura, RL
AF Miyao, Kotaro
   Yokota, Hirofumi
   Sakemura, R. Leo
TI Is CD19-directed chimeric antigen receptor T cell therapy a smart
   strategy to combat central nervous system lymphoma?
SO FRONTIERS IN ONCOLOGY
LA English
DT Review
DE CAR-T cell; PCNSL; CNS; CNS penetration; local administration
ID PRIMARY CNS LYMPHOMA; WHOLE-BRAIN RADIOTHERAPY; HIGH-DOSE METHOTREXATE;
   TERM-FOLLOW-UP; B-CELL; INTENSIVE CHEMOTHERAPY; PD-1 BLOCKADE; PHASE-II;
   TRANSPLANTATION; MULTICENTER
AB Primary central nervous system lymphoma (PCNSL) is a rare form and aggressive type of diffuse large B-cell lymphoma (DLBCL) that occurs in both immunocompetent and immunocompromised adults. While adding rituximab to chemotherapeutic regimens resulted in dramatic improvement in both progression-free survival and overall survival in patients with non-central nervous system (CNS) DLBCL, the outcomes of PCNSL are generally poor due to the immune-privileged tumor microenvironment or suboptimal delivery of systemic agents into tumor tissues. Therefore, more effective therapy for PCNSL generally requires systemic therapy with sufficient CNS penetration, including high-dose intravenous methotrexate with rituximab or high-dose chemotherapy followed by autologous stem cell transplantation. However, overall survival is usually inferior in comparison to non-CNS lymphomas, and treatment options are limited for elderly patients or patients with relapsed/refractory disease. Chimeric antigen receptor T (CAR-T) cell therapy has emerged as a cutting-edge cancer therapy, which led to recent FDA approvals for patients with B-cell malignancies and multiple myeloma. Although CAR-T cell therapy in patients with PCNSL demonstrated promising results without significant toxicities in some small cohorts, most cases of PCNSL are excluded from the pivotal CAR-T cell trials due to the concerns of neurotoxicity after CAR-T cell infusion. In this review, we will provide an overview of PCNSL and highlight current approaches, resistance mechanisms, and future perspectives of CAR-T cell therapy in patients with PCNSL.
C1 [Miyao, Kotaro] Anjo Kosei Hosp, Dept Hematol & Oncol, Anjo, Japan.
   [Yokota, Hirofumi] Nagoya Univ, Dept Hematol & Oncol, Grad Sch Med, Nagoya, Japan.
   [Sakemura, R. Leo] Mayo Clin, T Cell Engn, Rochester, MN 55902 USA.
   [Sakemura, R. Leo] Mayo Clin, Div Hematol, Rochester, MN 55902 USA.
C3 Nagoya University; Mayo Clinic; Mayo Clinic
RP Sakemura, RL (corresponding author), Mayo Clin, T Cell Engn, Rochester, MN 55902 USA.; Sakemura, RL (corresponding author), Mayo Clin, Div Hematol, Rochester, MN 55902 USA.
EM
CR Abramson JS, 2020, LANCET, V396, P839, DOI 10.1016/S0140-6736(20)31366-0
   Abramson JS, 2019, CANCER-AM CANCER SOC, V125, P3692, DOI 10.1002/cncr.32411
   Abramson JS, 2017, NEW ENGL J MED, V377, P783, DOI 10.1056/NEJMc1704610
   Abrey LE, 2005, J CLIN ONCOL, V23, P5034, DOI 10.1200/JCO.2005.13.524
   Alcantara M, 2022, BLOOD, V139, P792, DOI 10.1182/blood.2021012932
   Alimohamed N, 2012, LEUKEMIA LYMPHOMA, V53, P862, DOI 10.3109/10428194.2011.633250
   Better M, 2016, MOL THER, V24, pS115, DOI 10.1016/S1525-0016(16)33096-9
   Bishop MR, 2022, NEW ENGL J MED, V386, P629, DOI 10.1056/NEJMoa2116596
   Bouchkouj N, 2019, CLIN CANCER RES, V25, P1702, DOI 10.1158/1078-0432.CCR-18-2743
   Brandsma Dieta, 2018, Handb Clin Neurol, V152, P177, DOI 10.1016/B978-0-444-63849-6.00014-1
   Chapuy B, 2016, BLOOD, V127, P869, DOI 10.1182/blood-2015-10-673236
   Cheng G, 2003, BONE MARROW TRANSPL, V31, P679, DOI 10.1038/sj.bmt.1703917
   Chow VA, 2018, BLOOD, V132, P777, DOI 10.1182/blood-2018-04-839217
   Colombat P, 2006, BONE MARROW TRANSPL, V38, P417, DOI 10.1038/sj.bmt.1705452
   Cook MR, 2023, BLOOD ADV, V7, P32, DOI 10.1182/bloodadvances.2022008525
   Cote GM, 2012, BIOL BLOOD MARROW TR, V18, P76, DOI 10.1016/j.bbmt.2011.07.006
   Dai HR, 2020, J HEMATOL ONCOL, V13, DOI 10.1186/s13045-020-00856-8
   Dandachi D, 2019, CANCER CAUSE CONTROL, V30, P477, DOI 10.1007/s10552-019-01144-8
   de Larrea CF, 2020, BLOOD CANCER DISCOV, V1, P146, DOI 10.1158/2643-3230.BCD-20-0020
   Deeks SG, 2002, MOL THER, V5, P788, DOI 10.1006/mthe.2002.0611
   Ferreri AJM, 2017, LANCET HAEMATOL, V4, pE510, DOI 10.1016/S2352-3026(17)30174-6
   Fousek K, 2021, LEUKEMIA, V35, P75, DOI 10.1038/s41375-020-0792-2
   Frigault MJ, 2022, BLOOD, V139, P2306, DOI 10.1182/blood.2021014738
   Frigault MJ, 2019, BLOOD, V134, P860, DOI 10.1182/blood.2019001694
   Fry TJ, 2018, NAT MED, V24, P20, DOI 10.1038/nm.4441
   Furuse M, 2017, NEUROL MED-CHIR, V57, P191, DOI 10.2176/nmc.cr.2016-0330
   Garfall AL, 2016, BLOOD, V128, DOI 10.1182/blood.V128.22.5702.5702
   Grommes C, 2017, CANCER DISCOV, V7, P1018, DOI 10.1158/2159-8290.CD-17-0613
   Grommes C, 2017, J CLIN ONCOL, V35, P2410, DOI 10.1200/JCO.2017.72.7602
   Grupp SA, 2013, NEW ENGL J MED, V368, P1509, DOI 10.1056/NEJMoa1215134
   Gust J, 2017, CANCER DISCOV, V7, P1404, DOI 10.1158/2159-8290.CD-17-0698
   Hale M, 2017, MOL THER, V25, P570, DOI 10.1016/j.ymthe.2016.12.023
   Han CH, 2017, CANCER-AM CANCER SOC, V123, P4314, DOI 10.1002/cncr.30965
   Houillier C, 2019, J CLIN ONCOL, V37, P823, DOI 10.1200/JCO.18.00306
   Hunter BD, 2019, JNCI-J NATL CANCER I, V111, P646, DOI 10.1093/jnci/djz017
   Jacobson C, 2020, BLOOD, V136, DOI 10.1182/blood-2020-136834
   Jiang ZT, 2021, FRONT MICROBIOL, V12, DOI 10.3389/fmicb.2021.684016
   June CH, 2018, SCIENCE, V359, P1361, DOI 10.1126/science.aar6711
   Karschnia P, 2022, NEUROLOGY, V98, P884, DOI 10.1212/WNL.0000000000200608
   Kenderian SS, 2017, BIOL BLOOD MARROW TR, V23, P235, DOI 10.1016/j.bbmt.2016.09.002
   Kiefer T, 2012, ANN ONCOL, V23, P1809, DOI 10.1093/annonc/mdr553
   Lee DW, 2019, BIOL BLOOD MARROW TR, V25, P625, DOI 10.1016/j.bbmt.2018.12.758
   Lee DW, 2015, LANCET, V385, P517, DOI 10.1016/S0140-6736(14)61403-3
   Lee DW, 2014, BLOOD, V124, P188, DOI 10.1182/blood-2014-05-552729
   Leibman RS, 2017, PLOS PATHOG, V13, DOI 10.1371/journal.ppat.1006613
   Li TJ, 2020, FRONT ONCOL, V10, DOI 10.3389/fonc.2020.01306
   Liu BF, 2021, J CLIN INVEST, V131, DOI 10.1172/JCI150211
   Liu BF, 2016, J VIROL, V90, P9712, DOI 10.1128/JVI.00852-16
   Locke FL, 2022, NEW ENGL J MED, V386, P640, DOI 10.1056/NEJMoa2116133
   Locke FL, 2019, LANCET ONCOL, V20, P31, DOI 10.1016/S1470-2045(18)30864-7
   Locke FL, 2017, MOL THER, V25, P285, DOI 10.1016/j.ymthe.2016.10.020
   Louis DN, 2016, ACTA NEUROPATHOL, V131, P803, DOI 10.1007/s00401-016-1545-1
   Maude SL, 2018, NEW ENGL J MED, V378, P439, DOI 10.1056/NEJMoa1709866
   Maude SL, 2014, NEW ENGL J MED, V371, P1507, DOI 10.1056/NEJMoa1407222
   Miyao K, 2014, INT J HEMATOL, V100, P152, DOI 10.1007/s12185-014-1608-9
   Montemurro M, 2007, ANN ONCOL, V18, P665, DOI 10.1093/annonc/mdl458
   Munshi NC, 2021, NEW ENGL J MED, V384, P705, DOI 10.1056/NEJMoa2024850
   Narita Y, 2021, NEURO-ONCOLOGY, V23, P122, DOI 10.1093/neuonc/noaa145
   Nayak L, 2017, BLOOD, V129, P3071, DOI 10.1182/blood-2017-01-764209
   Neelapu SS, 2017, NEW ENGL J MED, V377, P2531, DOI 10.1056/NEJMoa1707447
   Neelapu SS, 2017, BLOOD, V130
   NELSON DF, 1992, INT J RADIAT ONCOL, V23, P9, DOI 10.1016/0360-3016(92)90538-S
   O'Neill BP, 2013, AM J HEMATOL, V88, P997, DOI 10.1002/ajh.23551
   Omuro A, 2015, LANCET HAEMATOL, V2, pE251, DOI 10.1016/S2352-3026(15)00074-5
   Omuro A, 2015, BLOOD, V125, P1403, DOI 10.1182/blood-2014-10-604561
   Ou A, 2020, NEURO-ONCOL ADV, V2, DOI 10.1093/noajnl/vdaa018
   Park JH, 2018, NEW ENGL J MED, V378, P449, DOI 10.1056/NEJMoa1709919
   Park JR, 2007, MOL THER, V15, P825, DOI 10.1038/sj.mt.6300104
   Parker KR, 2020, CELL, V183, P126, DOI 10.1016/j.cell.2020.08.022
   Patsoukis N, 2015, NAT COMMUN, V6, DOI 10.1038/ncomms7692
   Pentsova E, 2014, J NEURO-ONCOL, V117, P161, DOI 10.1007/s11060-014-1370-0
   Pettitt D, 2018, MOL THER, V26, P342, DOI 10.1016/j.ymthe.2017.10.019
   Plotkin SR, 2004, CLIN CANCER RES, V10, P5643, DOI 10.1158/1078-0432.CCR-04-0159
   Porter DL, 2015, SCI TRANSL MED, V7, DOI 10.1126/scitranslmed.aac5415
   Qin HY, 2018, MOL THER-ONCOLYTICS, V11, P127, DOI 10.1016/j.omto.2018.10.006
   Qiu YS, 2018, BRIT J HAEMATOL, V183, P674, DOI 10.1111/bjh.15009
   Reni M, 1999, BRIT J CANCER, V79, P530, DOI 10.1038/sj.bjc.6690083
   ROBERTS MR, 1994, BLOOD, V84, P2878
   Rubenstein JL, 2018, BLOOD ADV, V2, P1595, DOI 10.1182/bloodadvances.2017014845
   Rubenstein JL, 2013, J CLIN ONCOL, V31, P3061, DOI 10.1200/JCO.2012.46.9957
   Ruella M, 2018, NAT MED, V24, P1499, DOI 10.1038/s41591-018-0201-9
   Sakemura R, 2022, BLOOD, V139, P3708, DOI 10.1182/blood.2021012811
   Schneider D, 2017, J IMMUNOTHER CANCER, V5, DOI 10.1186/s40425-017-0246-1
   Schuster SJ, 2019, NEW ENGL J MED, V380, P45, DOI 10.1056/NEJMoa1804980
   Shah GD, 2007, J CLIN ONCOL, V25, P4730, DOI 10.1200/JCO.2007.12.5062
   Shah NN, 2020, NAT MED, V26, P1569, DOI 10.1038/s41591-020-1081-3
   Shiels MS, 2016, BRIT J HAEMATOL, V174, P417, DOI 10.1111/bjh.14073
   Siddiqi T, 2021, BLOOD ADV, V5, P4059, DOI 10.1182/bloodadvances.2020004106
   Soussain C, 2008, J CLIN ONCOL, V26, P2512, DOI 10.1200/JCO.2007.13.5533
   Spiegel JY, 2021, NAT MED, V27, P1419, DOI 10.1038/s41591-021-01436-0
   Subklewe M, 2021, BLOOD ADV, V5, P607, DOI 10.1182/bloodadvances.2020001792
   Sukumar M, 2013, J CLIN INVEST, V123, P4479, DOI 10.1172/JCI69589
   Takashima Y, 2019, SCI REP-UK, V9, DOI 10.1038/s41598-019-46473-5
   Theruvath J, 2020, NAT MED, V26, P712, DOI 10.1038/s41591-020-0821-8
   Thiel E, 2010, LANCET ONCOL, V11, P1036, DOI 10.1016/S1470-2045(10)70229-1
   Tong C, 2020, BLOOD, V136, P1632, DOI 10.1182/blood.2020005278
   Tu SF, 2019, FRONT ONCOL, V9, DOI 10.3389/fonc.2019.01350
   Turtle CJ, 2016, J CLIN INVEST, V126, P2123, DOI 10.1172/JCI85309
   Wang MH, 2020, NEW ENGL J MED, V382, P1331, DOI 10.1056/NEJMoa1914347
   Wang XL, 2021, CANCER IMMUNOL RES, V9, P75, DOI 10.1158/2326-6066.CIR-20-0236
   Wang YC, 2019, BLOOD CANCER J, V9, DOI 10.1038/s41408-019-0233-5
   Welch MR, 2015, LEUKEMIA LYMPHOMA, V56, P361, DOI 10.3109/10428194.2014.916800
   Wu JY, 2021, BLOOD CANCER J, V11, DOI 10.1038/s41408-021-00523-2
   Xie BL, 2022, CANCERS, V14, DOI 10.3390/cancers14133230
   Xu-Monette ZY, 2018, BLOOD, V131, P68, DOI 10.1182/blood-2017-07-740993
   Zah E, 2016, CANCER IMMUNOL RES, V4, P498, DOI 10.1158/2326-6066.CIR-15-0231
NR 106
TC 1
Z9 1
U1 4
U2 4
PU FRONTIERS MEDIA SA
PI LAUSANNE
PA AVENUE DU TRIBUNAL FEDERAL 34, LAUSANNE, CH-1015, SWITZERLAND
SN 2234-943X
J9 FRONT ONCOL
JI Front. Oncol.
PD JAN 5
PY 2023
VL 12
AR 1082235
DI 10.3389/fonc.2022.1082235
PG 11
WC Oncology
WE Science Citation Index Expanded (SCI-EXPANDED)
SC Oncology
GA 7W3WO
UT WOS:000913445900001
PM 36686821
OA gold, Green Published
DA 2024-06-03
ER

PT J
AU Chen, X
   Huang, M
   Zhang, ZY
   Jing, HL
   Zou, YL
   Bu, H
AF Chen, Xue
   Huang, Min
   Zhang, Zhenyuan
   Jing, Huilan
   Zou, Yueli
   Bu, Hui
TI Primary meningeal central nervous system lymphoma: A case report and
   literature review
SO MEDICINE
LA English
DT Review
DE cerebrospinal fluid; cytopathology; flow cytometry; meninges; primary
   central nervous system lymphoma
ID B-CELL LYMPHOMAS; FLOW-CYTOMETRY; LEPTOMENINGEAL DISEASE; DIAGNOSIS;
   SURVIVAL; INVOLVEMENT; MANAGEMENT; TRENDS
AB Rationale:Primary central nervous system lymphoma (PCNSL) is a rare extranodal non-Hodgkin lymphoma, and isolated meningeal PCNSL, without evidence of parenchymal involvement, is even less common, occurring in only 10% to 15% of cases. Patient concerns:A 65-years-old female presented to our hospital with progressive lower extremity motor dysfunction and blurred vision. The initial neurological examination revealed decreased muscle strength in both lower extremities and sensory dysfunction of lower extremities, saddle area, and buttocks. Brain magnetic resonance imaging showed no abnormalities. Lumbar enhanced magnetic resonance imaging showed T11 to L3 horizontal meningeal enhancement. Cerebrospinal fluid (CSF) cytology revealed lymphoma cells. Immunohistochemistry and flow cytometry of the CSF were performed as auxiliary methods to establish the diagnosis of lymphoma. Diagnoses:The patient was diagnosed primary meningeal central nervous system lymphoma. Interventions:During hospitalization, the patient was treated with 2 courses of high-dose intrathecal methotrexate and rituximab combined with intrathecal chemotherapy and supportive treatment. Outcomes:After 2 years of follow-up, the patient was able to walk and take care of herself. Lessons:Cases of PCNSL involving only the meninges are rare. Multimodal analysis of the CSF comprises an important component of the diagnostic work-up for patients with primary meningeal central nervous system lymphoma.
C1 [Chen, Xue; Zhang, Zhenyuan; Jing, Huilan; Zou, Yueli; Bu, Hui] Hebei Med Univ, Hosp 2, Dept Neurol, Shijiazhuang, Peoples R China.
   [Huang, Min] Yuncheng Cent Hosp Shanxi Prov, Dept Neurol, Yuncheng, Shanxi, Peoples R China.
   [Bu, Hui] Hebei Med Univ, Hosp 2, Dept Neurol, 215 Heping West Rd, Shijiazhuang 050000, Hebei, Peoples R China.
C3 Hebei Medical University; Shanxi Medical University; Hebei Medical
   University
RP Bu, H (corresponding author), Hebei Med Univ, Hosp 2, Dept Neurol, 215 Heping West Rd, Shijiazhuang 050000, Hebei, Peoples R China.
EM;
  
FU Project of National Key R & D Program of China [2016YFC0904503]
FX This article was supported by the grants from the Project of National
   Key R & D Program of China (No.: 2016YFC0904503).
CR Baraniskin A, 2012, BRIT J HAEMATOL, V156, P421, DOI 10.1111/j.1365-2141.2011.08928.x
   Canovi S, 2016, DIAGN CYTOPATHOL, V44, P841, DOI 10.1002/dc.23539
   Cencini E, 2018, BLOOD RES, V53, P174, DOI 10.5045/br.2018.53.2.174
   Chen T, 2022, J HEMATOL ONCOL, V15, DOI 10.1186/s13045-022-01356-7
   Eloranta S, 2018, EUR J HAEMATOL, V100, P61, DOI 10.1111/ejh.12980
   Han CH, 2017, CANCER-AM CANCER SOC, V123, P4314, DOI 10.1002/cncr.30965
   Hegde U, 2005, BLOOD, V105, P496, DOI 10.1182/blood-2004-05-1982
   Joshi A, 2022, J RADIOL CASE REP, V16, DOI 10.3941/jrcr.v16i7.4562
   Liu J, 2017, ONCOL LETT, V14, P5505, DOI 10.3892/ol.2017.6893
   Liu YX, 2021, INT J ONCOL, V58, P371, DOI 10.3892/ijo.2021.5180
   Mao CL, 2019, WORLD NEUROSURG, V123, pE15, DOI 10.1016/j.wneu.2018.10.034
   Mendez JS, 2018, NEURO-ONCOLOGY, V20, P687, DOI 10.1093/neuonc/nox187
   Rozenblum L, 2022, CANCERS, V14, DOI 10.3390/cancers14174071
   Schinstine M, 2006, CANCER CYTOPATHOL, V108, P157, DOI 10.1002/cncr.21915
   Schroers R, 2010, EUR J HAEMATOL, V85, P520, DOI 10.1111/j.1600-0609.2010.01516.x
   Schroers R, 2010, EUR J HAEMATOL, V85, P236, DOI 10.1111/j.1600-0609.2010.01475.x
   Shiels MS, 2016, BRIT J HAEMATOL, V174, P417, DOI 10.1111/bjh.14073
   Sinicrope K, 2018, NEUROL CLIN, V36, P517, DOI 10.1016/j.ncl.2018.04.008
   Villano JL, 2011, BRIT J CANCER, V105, P1414, DOI 10.1038/bjc.2011.357
NR 19
TC 1
Z9 1
U1 0
U2 3
PU LIPPINCOTT WILLIAMS & WILKINS
PI PHILADELPHIA
PA TWO COMMERCE SQ, 2001 MARKET ST, PHILADELPHIA, PA 19103 USA
SN 0025-7974
EI 1536-5964
J9 MEDICINE
JI Medicine (Baltimore)
PD DEC 30
PY 2022
VL 101
IS 52
DI 10.1097/MD.0000000000032567
PG 5
WC Medicine, General & Internal
WE Science Citation Index Expanded (SCI-EXPANDED)
SC General & Internal Medicine
GA 7M9FZ
UT WOS:000906956700047
PM 36596043
OA gold, Green Published
DA 2024-06-03
ER

PT J
AU Koike, H
   Morikawa, M
   Ishimaru, H
   Ideguchi, R
   Uetani, M
   Hiu, T
   Matsuo, T
   Miyoshi, M
AF Koike, Hirofumi
   Morikawa, Minoru
   Ishimaru, Hideki
   Ideguchi, Reiko
   Uetani, Masataka
   Hiu, Takeshi
   Matsuo, Takayuki
   Miyoshi, Mitsuharu
TI Amide proton transfer MRI differentiates between progressive multifocal
   leukoencephalopathy and malignant brain tumors: a pilot study
SO BMC MEDICAL IMAGING
LA English
DT Article
DE Magnetic resonance imaging; Amide proton transfer imaging; Progressive
   multifocal leukoencephalopathy; Primary central nervous system lymphoma;
   Glioblastoma multiforme
ID CENTRAL-NERVOUS-SYSTEM; MAGNETIZATION-TRANSFER; ENDOGENOUS PROTEIN; CELL
   LYMPHOMA; PATIENT; GLIOMA; RESONANCE; CONTRAST; GLIOBLASTOMA; DISEASE
AB Background: Progressive multifocal leukoencephalopathy (PML) is a demyelinating disease of the central nerve system caused by the John Cunningham virus. On MRI, PML may sometimes appear similar to primary central nervous system lymphoma (PCNSL) and glioblastoma multiforme (GBM). The purpose of this pilot study was to evaluate the potential of amide proton transfer (APT) imaging for differentiating PML from PCNSL and GBM.
   Methods: Patients with PML (n=4; two men; mean age 52.36.1 years), PCNSL (n=7; four women; mean age 74.45.8 years), or GBM (n=11; 6 men; mean age 65.0 +/- 15.2 years) who underwent APT-CEST MRI between January 2021 and September 2022 were retrospectively evaluated. Magnetization transfer ratio asymmetry (MTRasym) values were measured on APT imaging using a region of interest within the lesion. Receiver operating characteristics curve analysis was used to determine diagnostic cutoffs for MTRasym.
   Results: The mean MTRasym values were 0.005 +/- 0.005 in the PML group, 0.025 +/- 0.005 in the PCNSL group, and 0.025 +/- 0.009 in the GBM group. There were significant differences in MTRasym between PML and PCNSL (P=0.023), and between PML and GBM (P=0.015). For differentiating PML from PCNSL, an MTRasym threshold of 0.0165 gave diagnostic sensitivity, specificity, positive predictive value, and negative predictive value of 100% (all). For differentiating PML from GBM, an MTRasym threshold of 0.015 gave diagnostic sensitivity, specificity, positive predictive value, and negative predictive value of 100%, 90.9%, 80.0%, and 100%, respectively.
   Conclusion: MTRasym values obtained from APT imaging allowed patients with PML to be clearly discriminated from patients with PCNSL or GBM.
C1 [Koike, Hirofumi; Uetani, Masataka] Nagasaki Univ, Dept Radiol, Grad Sch Biomed Sci, 1-7-1 Sakamoto, Nagasaki 8528501, Japan.
   [Morikawa, Minoru; Ishimaru, Hideki] Nagasaki Univ Hosp, Dept Radiol, 1-7-1 Sakamoto, Nagasaki 8528501, Japan.
   [Ideguchi, Reiko] Nagasaki Univ, Dept Radioisotope Med, Grad Sch Biomed Sci, 1-7-1 Sakamoto, Nagasaki 8528588, Japan.
   [Hiu, Takeshi; Matsuo, Takayuki] Nagasaki Univ, Dept Neurosurg, Grad Sch Biomed Sci, 1-7-1 Sakamoto, Nagasaki 8528501, Japan.
   [Miyoshi, Mitsuharu] GE Healthcare Japan, MR Applicat & Workflow, 4-7-127 Asahigaoka, Hino, Tokyo 1918503, Japan.
C3 Nagasaki University; Nagasaki University; Nagasaki University; Nagasaki
   University; General Electric
RP Koike, H (corresponding author), Nagasaki Univ, Dept Radiol, Grad Sch Biomed Sci, 1-7-1 Sakamoto, Nagasaki 8528501, Japan.
EM
FU Grants-in-Aid for Scientific Research [20K09328] Funding Source: KAKEN
CR Bathla G, 2016, CLIN RADIOL, V71, P602, DOI 10.1016/j.crad.2016.02.006
   Belden CJ, 2011, RADIOGRAPHICS, V31, P1717, DOI 10.1148/rg.316115512
   Carson KR, 2009, BLOOD, V113, P4834, DOI 10.1182/blood-2008-10-186999
   CBTRUS, 2002, STAT REP PRIM BRIAN
   Commins Deborah L, 2006, Neurosurg Focus, V21, pE2
   Cortese I, 2021, NAT REV NEUROL, V17, P37, DOI 10.1038/s41582-020-00427-y
   DEANGELIS LM, 1993, ANN NEUROL, V33, P308, DOI 10.1002/ana.410330313
   Evens AM, 2010, J CLIN ONCOL, V28, P1038, DOI 10.1200/JCO.2009.25.4961
   FLEISS JL, 1973, EDUC PSYCHOL MEAS, V33, P613, DOI 10.1177/001316447303300309
   Gheuens S, 2010, J NEUROL NEUROSUR PS, V81, P247, DOI 10.1136/jnnp.2009.187666
   Grewal J, 2016, J NEUROVIROL, V22, P683, DOI 10.1007/s13365-016-0459-y
   Gupta HV, 2016, ACTA NEUROL BELG, V116, P391, DOI 10.1007/s13760-015-0532-y
   Henkelman RM, 2001, NMR BIOMED, V14, P57, DOI 10.1002/nbm.683
   Heo HY, 2017, MAGN RESON MED, V77, P1853, DOI 10.1002/mrm.26264
   HO K, 1980, ACTA NEUROPATHOL, V52, P81, DOI 10.1007/BF00687233
   Hong XH, 2014, NEURO-ONCOLOGY, V16, P856, DOI 10.1093/neuonc/not245
   Imai H, 2009, PATHOL INT, V59, P431, DOI 10.1111/j.1440-1827.2009.02390.x
   Jiang SS, 2016, EUR RADIOL, V26, P64, DOI 10.1007/s00330-015-3805-1
   Jones CK, 2006, MAGN RESON MED, V56, P585, DOI 10.1002/mrm.20989
   Kalakunja Aroon, 2003, Clin Adv Hematol Oncol, V1, P554
   Kleinschmidt-DeMasters BK, 2005, NEW ENGL J MED, V353, P369, DOI 10.1056/NEJMoa051782
   Koeller KK, 2016, RADIOL CLIN N AM, V54, P649, DOI 10.1016/j.rcl.2016.03.003
   Korman BD, 2009, ARCH DERMATOL, V145, P937, DOI 10.1001/archdermatol.2009.175
   Kurita K, 1999, INTERNAL MED, V38, P507, DOI 10.2169/internalmedicine.38.507
   Langer-Gould A, 2005, NEW ENGL J MED, V353, P375, DOI 10.1056/NEJMoa051847
   Li CM, 2014, EUR RADIOL, V24, P2631, DOI 10.1007/s00330-014-3241-7
   Lingl JP, 2022, DIAGNOSTICS, V12, DOI 10.3390/diagnostics12020490
   Liu Z, 2015, BRAIN, V138, P1239, DOI 10.1093/brain/awv065
   Ng MC, 2009, J MAGN RESON IMAGING, V29, P523, DOI 10.1002/jmri.21610
   NG S, 1995, J COMPUT ASSIST TOMO, V19, P302, DOI 10.1097/00004728-199503000-00025
   Preusser M, 2010, PATHOLOGY, V42, P547, DOI 10.3109/00313025.2010.508786
   Rees JH, 1996, RADIOGRAPHICS, V16, P1413, DOI 10.1148/radiographics.16.6.8946545
   Rubenstein J, 2008, LEUKEMIA LYMPHOMA, V49, P43, DOI 10.1080/10428190802311441
   Sagiyama K, 2014, P NATL ACAD SCI USA, V111, P4542, DOI 10.1073/pnas.1323855111
   Scheidegger R, 2014, NEUROIMAGE, V99, P256, DOI 10.1016/j.neuroimage.2014.05.036
   Shah R, 2010, CLIN RADIOL, V65, P431, DOI 10.1016/j.crad.2010.03.001
   Tee YK, 2014, NMR BIOMED, V27, P1019, DOI 10.1002/nbm.3147
   Terae S, 1996, NEURORADIOLOGY, V38, P34, DOI 10.1007/BF00593213
   Tietze A, 2014, NMR BIOMED, V27, P163, DOI 10.1002/nbm.3048
   Togao O, 2014, NEURO-ONCOLOGY, V16, P441, DOI 10.1093/neuonc/not158
   Tyler KL, 2009, ARCH NEUROL-CHICAGO, V66, P1065, DOI 10.1001/archneurol.2009.189
   Van Assche G, 2005, NEW ENGL J MED, V353, P362, DOI 10.1056/NEJMoa051586
   van Zijl PCM, 2011, MAGN RESON MED, V65, P927, DOI 10.1002/mrm.22761
   Ward KM, 2000, J MAGN RESON, V143, P79, DOI 10.1006/jmre.1999.1956
   Wen ZB, 2010, NEUROIMAGE, V51, P616, DOI 10.1016/j.neuroimage.2010.02.050
   WHITEMAN MLH, 1993, RADIOLOGY, V187, P233, DOI 10.1148/radiology.187.1.8451420
   WHO Classification of Tumours Editorial Board, 2021, WHO CLASSIFICATION T
   Zhao XN, 2011, MAGN RESON MED, V66, P1033, DOI 10.1002/mrm.22891
   Zhou JY, 2022, MAGN RESON MED, V88, P546, DOI 10.1002/mrm.29241
   Zhou JY, 2013, J MAGN RESON IMAGING, V38, P1119, DOI 10.1002/jmri.24067
   Zhou JY, 2011, NAT MED, V17, P130, DOI 10.1038/nm.2268
   Zhou JY, 2003, MAGNET RESON MED, V50, P1120, DOI 10.1002/mrm.10651
   Zhou JY, 2003, NAT MED, V9, P1085, DOI 10.1038/nm907
   Zu ZL, 2018, NMR BIOMED, V31, DOI 10.1002/nbm.3934
NR 54
TC 1
Z9 2
U1 0
U2 1
PU BMC
PI LONDON
PA CAMPUS, 4 CRINAN ST, LONDON N1 9XW, ENGLAND
SN 1471-2342
J9 BMC MED IMAGING
JI BMC Med. Imag.
PD DEC 26
PY 2022
VL 22
IS 1
AR 227
DI 10.1186/s12880-022-00959-3
PG 10
WC Radiology, Nuclear Medicine & Medical Imaging
WE Science Citation Index Expanded (SCI-EXPANDED)
SC Radiology, Nuclear Medicine & Medical Imaging
GA 7I7DS
UT WOS:000904045200001
PM 36572873
OA Green Published, Green Submitted, gold
DA 2024-06-03
ER

PT J
AU Cheng, CL
   Yao, CY
   Huang, PH
   Yu, CW
   Fang, WQ
   Chuang, WH
   Wu, SJ
   Lin, YJ
   Hung, YC
   Tsai, CH
   Yu, SC
   Chou, WC
   Tien, HF
AF Cheng, Chieh-Lung
   Yao, Chi-Yuan
   Huang, Po-Hao
   Yu, Chih-Wei
   Fang, Wei-Quan
   Chuang, Wen-Hui
   Wu, Shang-Ju
   Lin, Yu-Jen
   Hung, Yu-Chin
   Tsai, Cheng-Hong
   Yu, Shan-Chi
   Chou, Wen-Chien
   Tien, Hwei-Fang
TI Cerebrospinal fluid soluble programmed death-ligand 1 is a useful
   prognostic biomarker in primary central nervous system lymphoma
SO BRITISH JOURNAL OF HAEMATOLOGY
LA English
DT Article
DE biomarker; cerebrospinal fluid; primary central nervous system lymphoma;
   soluble PD-L1; soluble PD-L2
ID PRIMARY CNS LYMPHOMA; B-CELL LYMPHOMA; PHASE-II; EXPRESSION; SURVIVAL;
   INTERLEUKIN-10; LENALIDOMIDE; RITUXIMAB; ANTIBODY; CANCERS
AB The increased expression of programmed death-ligands 1 and 2 (PD-L1 and PD-L2, respectively) on tumour cells contributes to immune evasion, suggesting that these proteins are attractive therapeutic targets. This study aimed to evaluate the validity of cerebrospinal fluid (CSF) soluble PD-L1 (sPD-L1) and soluble PD-L2 (sPD-L2) as biomarkers for primary central nervous system lymphoma (PCNSL). We determined the CSF concentrations of sPD-L1 and sPD-L2 in 46 patients with PCNSL using enzyme-linked immunosorbent assays (ELISAs). A control group comprised 153 patients with other brain tumours, inflammatory/infectious status, or neurodegenerative diseases. Only CSF sPD-L1 levels were significantly higher in patients with PCNSL relative to the controls. CSF sPD-L1 also exhibited superior overall discrimination performance compared to CSF sPD-L2 in diagnosing PCNSL. Compared with patients with PCNSL with low CSF sPD-L1 levels, more patients with high levels had high serum lactate dehydrogenase levels, leptomeningeal involvement, and deep-brain involvement. Furthermore, CSF sPD-L1 could predict poor survival in PCNSL but CSF sPD-L2 could not. Intriguingly, CSF sPD-L1 levels were correlated with disease status and their dynamic changes post treatment could predict time to relapse. In conclusion, this study identified CSF sPD-L1 as a promising prognostic biomarker, indicating a therapeutic potential of PD-L1 blockade in PCNSL.
C1 [Cheng, Chieh-Lung; Chuang, Wen-Hui; Wu, Shang-Ju; Tsai, Cheng-Hong; Chou, Wen-Chien; Tien, Hwei-Fang] Natl Taiwan Univ, Natl Taiwan Univ Hosp, Coll Med, Dept Internal Med,Div Hematol, Taipei, Taiwan.
   [Yao, Chi-Yuan] Natl Taiwan Univ, Natl Taiwan Univ Hosp, Coll Med, Dept Lab Med, Taipei, Taiwan.
   [Huang, Po-Hao; Chou, Wen-Chien] Natl Taiwan Univ, Natl Taiwan Univ Hosp, Coll Med, Dept Surg,Div Neurosurg, Taipei, Taiwan.
   [Yu, Chih-Wei] Natl Taiwan Univ, Natl Taiwan Univ Hosp, Coll Med, Dept Radiol & Med Imaging, Taipei, Taiwan.
   [Fang, Wei-Quan] Ctr Drug Evaluat, Div New Drug, Taipei, Taiwan.
   [Lin, Yu-Jen] Natl Taiwan Univ, Canc Ctr, Dept Hematol Oncol, Taipei, Taiwan.
   [Lin, Yu-Jen] Natl Taiwan Univ Hosp, Dept Internal Med, Div Hematol & Oncol, Yunlin, Taiwan.
   [Wu, Shang-Ju] Natl Taiwan Univ, Natl Taiwan Univ Hosp, Coll Med, Dept Pathol, Taipei, Taiwan.
   [Cheng, Chieh-Lung] Natl Taiwan Univ Hosp, Dept Internal Med, 7 Chung Shan South Rd, Taipei 10002, Taiwan.
C3 National Taiwan University; National Taiwan University Hospital;
   National Taiwan University; National Taiwan University Hospital;
   National Taiwan University; National Taiwan University Hospital;
   National Taiwan University; National Taiwan University Hospital;
   National Taiwan University; National Taiwan University; National Taiwan
   University Hospital; National Taiwan University; National Taiwan
   University Hospital; National Taiwan University; National Taiwan
   University Hospital
RP Cheng, CL (corresponding author), Natl Taiwan Univ Hosp, Dept Internal Med, 7 Chung Shan South Rd, Taipei 10002, Taiwan.
EM
RI Yu, Chih-Wei/GRX-2332-2022; Yu, shanchi/GXZ-6055-2022; Tsai,
   Cheng-Hong/Q-4855-2016
OI Yu, Chih-Wei/0000-0002-8648-8419; Tsai, Cheng-Hong/0000-0002-4174-8766;
   Cheng, Chieh-Lung/0000-0001-5442-1079; CHOU,
   WEN-CHIEN/0000-0003-2967-698X; Yao, Chi-Yuan/0000-0001-5052-9445
FU  [NA-CL-OTHER-PI-008662]
FX ACKNOWLEDGEMENTS Funding for this work was supported by an
   investigator-initiated research grant (NA-CL-OTHER-PI-008662 to
   Chieh-Lung Cheng) from Bristol-Myers Squibb, USA. We also thank the
   relevant medical staff and patients who contributed to this study.
CR Abrey LE, 2005, J CLIN ONCOL, V23, P5034, DOI 10.1200/JCO.2005.13.524
   Alaggio R, 2022, LEUKEMIA, V36, P1720, DOI 10.1038/s41375-022-01620-2
   Ambady P, 2019, LEUKEMIA LYMPHOMA, V60, P515, DOI 10.1080/10428194.2018.1480771
   Baraniskin A, 2011, BLOOD, V117, P3140, DOI 10.1182/blood-2010-09-308684
   Brody R, 2017, LUNG CANCER, V112, P200, DOI 10.1016/j.lungcan.2017.08.005
   Chamberlain MC, 2010, NEURO-ONCOLOGY, V12, P736, DOI 10.1093/neuonc/noq011
   Chapuy B, 2016, BLOOD, V127, P869, DOI 10.1182/blood-2015-10-673236
   Chen YJ, 2011, CYTOKINE, V56, P231, DOI 10.1016/j.cyto.2011.06.004
   Cho H, 2017, ONCOTARGET, V8, P87317, DOI 10.18632/oncotarget.20264
   Cho I, 2020, BMC CANCER, V20, DOI 10.1186/s12885-020-6612-2
   Ding Y, 2017, SCAND J IMMUNOL, V86, P361, DOI 10.1111/sji.12596
   Droeser RA, 2013, EUR J CANCER, V49, P2233, DOI 10.1016/j.ejca.2013.02.015
   Ferreri AJM, 2021, BRIT J HAEMATOL, V193, P497, DOI 10.1111/bjh.17357
   Ferreri Andres J M, 2019, Am Soc Clin Oncol Educ Book, V39, P454, DOI 10.1200/EDBK_242547
   Finkelmeier F, 2016, EUR J CANCER, V59, P152, DOI 10.1016/j.ejca.2016.03.002
   Four M, 2017, HEMATOL ONCOL, V35, P487, DOI 10.1002/hon.2375
   Frigola X, 2012, IMMUNOL LETT, V142, P78, DOI 10.1016/j.imlet.2011.11.001
   Frigola X, 2011, CLIN CANCER RES, V17, P1915, DOI 10.1158/1078-0432.CCR-10-0250
   Furuse M, 2020, BMC CANCER, V20, DOI 10.1186/s12885-020-06755-y
   Gavrilenko AN, 2020, BLOOD, V136, DOI 10.1182/blood-2020-138924
   Ghesquieres H, 2019, ANN ONCOL, V30, P621, DOI 10.1093/annonc/mdz032
   Graber JJ, 2020, LEUKEMIA LYMPHOMA, V61, P1766, DOI 10.1080/10428194.2020.1742903
   Gu DQ, 2018, J IMMUNOTHER CANCER, V6, DOI 10.1186/s40425-018-0449-0
   Hayano A, 2017, ANTICANCER RES, V37, P5655, DOI 10.21873/anticanres.12001
   Herbst RS, 2014, NATURE, V515, P563, DOI 10.1038/nature14011
   Hilal T, 2020, ADV CELL GENE THER, V3, pe82
   Hoang-Xuan K, 2020, BLOOD, V136, DOI 10.1182/blood-2020-141773
   Lauer EM, 2020, BRIT J HAEMATOL, V190, pE110, DOI 10.1111/bjh.16759
   Matikas A, 2019, CLIN CANCER RES, V25, P5717, DOI 10.1158/1078-0432.CCR-19-1131
   Nayak L, 2017, BLOOD, V129, P3071, DOI 10.1182/blood-2017-01-764209
   Okadome K, 2020, BRIT J CANCER, V122, P1535, DOI 10.1038/s41416-020-0811-0
   Okuma Y, 2017, LUNG CANCER, V104, P1, DOI 10.1016/j.lungcan.2016.11.023
   Ostrom QT, 2021, NEURO-ONCOLOGY, V23, P1, DOI 10.1093/neuonc/noab200
   Rossille D, 2014, LEUKEMIA, V28, P2367, DOI 10.1038/leu.2014.137
   Rubenstein JL, 2018, BLOOD ADV, V2, P1595, DOI 10.1182/bloodadvances.2017014845
   Rubenstein JL, 2013, BLOOD, V121, P4740, DOI 10.1182/blood-2013-01-476333
   Sasayama T, 2012, NEURO-ONCOLOGY, V14, P368, DOI 10.1093/neuonc/nor203
   Sethi TK, 2019, LEUKEMIA LYMPHOMA, V60, P2880, DOI 10.1080/10428194.2019.1620942
   Soussain C, 2019, EUR J CANCER, V117, P121, DOI 10.1016/j.ejca.2019.05.024
   Squibb B-M., 2016, STUDY NIVOLUMAB RELA
   Taube JM, 2014, CLIN CANCER RES, V20, P5064, DOI 10.1158/1078-0432.CCR-13-3271
   Topalian SL, 2012, NEW ENGL J MED, V366, P2443, DOI 10.1056/NEJMoa1200690
   Villano JL, 2011, BRIT J CANCER, V105, P1414, DOI 10.1038/bjc.2011.357
   Wang L, 2015, ONCOTARGET, V6, P41228, DOI 10.18632/oncotarget.5682
   Wei D, 2015, MOL NEUROBIOL, V51, P1452, DOI 10.1007/s12035-014-8822-6
   Wei W, 2018, MEDICINE, V97, DOI 10.1097/MD.0000000000009617
   Zhu XX, 2017, ONCOTARGET, V8, P97671, DOI 10.18632/oncotarget.18311
NR 47
TC 1
Z9 1
U1 0
U2 2
PU WILEY
PI HOBOKEN
PA 111 RIVER ST, HOBOKEN 07030-5774, NJ USA
SN 0007-1048
EI 1365-2141
J9 BRIT J HAEMATOL
JI Br. J. Haematol.
PD APR
PY 2023
VL 201
IS 1
BP 75
EP 85
DI 10.1111/bjh.18598
EA DEC 2022
PG 11
WC Hematology
WE Science Citation Index Expanded (SCI-EXPANDED)
SC Hematology
GA C1CM4
UT WOS:000895136100001
PM 36480431
DA 2024-06-03
ER

PT J
AU Ding, XH
   Liang, TY
   Liang, B
   Gao, HL
   Wang, JB
   Liu, HX
   Wang, F
   Zheng, XM
   Li, PL
   Chen, SC
   Feng, ES
AF Ding, Xinghuan
   Liang, Tingyu
   Liang, Bo
   Gao, Haili
   Wang, Jianbo
   Liu, Hongxing
   Wang, Fang
   Zheng, Xinmei
   Li, Peiliang
   Chen, Shichao
   Feng, Enshan
TI Diagnostic value of EBV-DNA in CSF for PCNSL in AIDS patients with focal
   brain lesions: A meta-analysis of diagnostic test
SO MEDICINE
LA English
DT Review
DE diagnosis; Epstein-Barr virus; HIV; AIDS; meta-analysis; PCNSL; PCR
ID CENTRAL-NERVOUS-SYSTEM; EPSTEIN-BARR-VIRUS; POLYMERASE-CHAIN-REACTION;
   CEREBROSPINAL-FLUID; PREDICTIVE-VALUE; VIRAL LOAD; HIV; LYMPHOMA; MARKER
AB Background:Epstein-Barr virus (EBV) infection plays a crucial role in the progression of acquired immunodeficiency syndrome related primary central nervous system lymphoma (AR-PCNSL). This study aimed at evaluating the diagnostic value of cerebrospinal spinal fluid (CSF) EBV-deoxyribonucleic acid (DNA) for PCNSL in patients with infection of human immunodeficiency (HIV) virus through a meta-analysis of diagnostic test. Methods:A systematic search in PubMed, Embase, Web of Science, Wanfang, Chinese Biomedical Database and Chinese National Knowledge Infrastructure was conducted before May 10, 2022. Heterogeneity among the studies was assessed using Q test and I-2 statistics. Publication bias was assessed using the Deek's funnel plot asymmetry test. Statistical analyses were performed using Stata 16.0 software. The pooled sensitivity, specificity, positive and negative likelihood ratios (PLR and NLR), diagnostic odds ratios (DOR) and 95% confidence intervals (CI) were caculated to evaluate the diagnostic value. A symmetric receiver operating characteristic (SROC) curve and the area under the SROC curve (AUC) were constructed to evaluate the test-performance. Results:Twelve studies were included in the final analyses, with a total of 141 patients with AR-PCNSL and 590 controls. The pooled diagnostic values were sensitivity of 0.83 (95% CI: 0.73-0.90), specificity of 0.95 (95%CI: 0.89-0.98), PLR of 17.8 (95%CI: 6.8-46.1), NLR of 0.17 (95%CI: 0.10-0.30), DOR of 102 (95%CI: 28-379), and AUC of 0.94 (95%CI: 0.91-0.96). Conclusion:In summary the overall diagnostic value of CSF EBV-DNA is very high and it can be a reliable diagnostic biomarker for AR-PCNSL.
C1 [Ding, Xinghuan; Liang, Tingyu; Liang, Bo; Wang, Jianbo; Liu, Hongxing; Wang, Fang; Zheng, Xinmei; Li, Peiliang; Chen, Shichao; Feng, Enshan] Capital Med Univ, Beijing Ditan Hosp, Dept Neurosurg, Beijing, Peoples R China.
   [Ding, Xinghuan; Liang, Tingyu; Liang, Bo; Gao, Haili; Wang, Jianbo; Liu, Hongxing; Wang, Fang; Zheng, Xinmei; Li, Peiliang; Chen, Shichao; Feng, Enshan] Capital Med Univ, Beijing Ditan Hosp, Natl Ctr Infect Dis, Beijing, Peoples R China.
   [Gao, Haili] Capital Med Univ, Beijing Ditan Hosp, Dept Pathol, Beijing, Peoples R China.
   [Feng, Enshan] Capital Med Univ, Beijing Ditan Hosp, Dept Neurosurg, Beijing 100015, Peoples R China.
C3 Capital Medical University; Capital Medical University; Capital Medical
   University; Capital Medical University
RP Feng, ES (corresponding author), Capital Med Univ, Beijing Ditan Hosp, Dept Neurosurg, Beijing 100015, Peoples R China.
EM;
  ;
  ;
  
RI LIANG, Tingyu/KIK-8131-2024
FU Seedling Plan of Beijing Ditan Hospital Research Fund [DTYM-202106]
FX This study was funded by Seedling Plan of Beijing Ditan Hospital
   Research Fund (DTYM-202106). The authors would like to acknowledge all
   the authors for their contributions to this study. DX and FE were
   responsible for data analysis. DX, LT, and CS helped design the studies.
   LB, GH, WJ, LH, ZX, and WF participated in the collection of the
   literature. DX, LT, and FE assessed the methodological quality of each
   included study. FE supervised this study. DX is the primary author of
   the manuscript. All authors contributed to the review and development of
   this manuscript.
CR Acosta MC, 2018, HIV MED, V19, P673, DOI 10.1111/hiv.12646
   Antinori A, 1999, J CLIN ONCOL, V17, P554, DOI 10.1200/JCO.1999.17.2.554
   Antinori A, 1999, BRAIN PATHOL, V9, P199
   ARRIBAS JR, 1995, J CLIN MICROBIOL, V33, P1580, DOI 10.1128/JCM.33.6.1580-1583.1995
   Bossolasco S, 2006, CLIN INFECT DIS, V42, pE21, DOI 10.1086/499956
   Bossolasco S, 2002, J NEUROVIROL, V8, P432, DOI 10.1080/13550280260422730
   Brandsma Dieta, 2018, Handb Clin Neurol, V152, P177, DOI 10.1016/B978-0-444-63849-6.00014-1
   Brink NS, 1998, J NEUROL NEUROSUR PS, V65, P191, DOI 10.1136/jnnp.65.2.191
   Cingolani A, 1998, JNCI-J NATL CANCER I, V90, P364, DOI 10.1093/jnci/90.5.364
   Cinque P, 2004, CLIN INFECT DIS, V39, P1396, DOI 10.1086/425132
   CINQUE P, 1993, LANCET, V342, P398, DOI 10.1016/0140-6736(93)92814-A
   Cinque P, 1996, AIDS, V10, P951, DOI 10.1097/00002030-199610090-00004
   Corcoran C, 2008, J CLIN VIROL, V42, P433, DOI 10.1016/j.jcv.2008.03.017
   DELUCA A, 1995, BRIT J HAEMATOL, V90, P844, DOI 10.1111/j.1365-2141.1995.tb05205.x
   Goedert JJ, 2000, SEMIN ONCOL, V27, P390
   Grommes C, 2017, J CLIN ONCOL, V35, P2410, DOI 10.1200/JCO.2017.72.7602
   Higgins JPT, 2002, STAT MED, V21, P1539, DOI 10.1002/sim.1186
   Hirsch Hans H., 1998, Clin Microbiol Infect, V4, P577, DOI 10.1111/j.1469-0691.1998.tb00040.x
   Ivers LC, 2004, CLIN INFECT DIS, V38, P1629, DOI 10.1086/420934
   Skolasky RL, 1999, J NEUROL SCI, V163, P32, DOI 10.1016/S0022-510X(98)00315-3
   Tachikawa N, 1999, INTERNAL MED, V38, P556, DOI 10.2169/internalmedicine.38.556
   van Houwelingen HC, 2002, STAT MED, V21, P589, DOI 10.1002/sim.1040
   Wang J, 2007, HIV MED, V8, P306, DOI 10.1111/j.1468-1293.2007.00475.x
   Wang YL, 2022, BIOMED PHARMACOTHER, V145, DOI 10.1016/j.biopha.2021.112392
   Whiting PF, 2011, ANN INTERN MED, V155, P529, DOI 10.7326/0003-4819-155-8-201110180-00009
   Yanagisawa K, 2013, INTERNAL MED, V52, P955, DOI 10.2169/internalmedicine.52.9088
   Yu GH, 1996, DIAGN CYTOPATHOL, V14, P114, DOI 10.1002/(SICI)1097-0339(199603)14:2<114::AID-DC3>3.0.CO;2-J
NR 27
TC 1
Z9 1
U1 1
U2 2
PU LIPPINCOTT WILLIAMS & WILKINS
PI PHILADELPHIA
PA TWO COMMERCE SQ, 2001 MARKET ST, PHILADELPHIA, PA 19103 USA
SN 0025-7974
EI 1536-5964
J9 MEDICINE
JI Medicine (Baltimore)
PD DEC 2
PY 2022
VL 101
IS 48
DI 10.1097/MD.0000000000031793
PG 8
WC Medicine, General & Internal
WE Science Citation Index Expanded (SCI-EXPANDED)
SC General & Internal Medicine
GA 6W1NU
UT WOS:000895502600110
PM 36482588
OA gold, Green Published
DA 2024-06-03
ER

PT J
AU Chojak, R
   Kozba-Gosztyla, M
   Polanska, K
   Rojek, M
   Chojko, A
   Bogacz, R
   Skorupa, N
   Wieclaw, J
   Czapiga, B
AF Chojak, Rafal
   Kozba-Gosztyla, Marta
   Polanska, Karolina
   Rojek, Marta
   Chojko, Aleksandra
   Bogacz, Rafal
   Skorupa, Natalia
   Wieclaw, Jakub
   Czapiga, Bogdan
TI Surgical resection versus biopsy in the treatment of primary central
   nervous system lymphoma: a systematic review and meta-analysis
SO JOURNAL OF NEURO-ONCOLOGY
LA English
DT Review
DE PCNSL; Surgery; Biopsy; Hazard ratio; Overall survival; Progression-free
   survival
ID PRIMARY CNS LYMPHOMA; HIGH-DOSE METHOTREXATE; INTERNATIONAL EXTRANODAL
   LYMPHOMA; MRI FEATURES; SURVIVAL; SURGERY; RADIOTHERAPY; RITUXIMAB;
   OUTCOMES; REGIMEN
AB Purpose Despite the improvement in treatment and prognosis of primary central nervous system lymphoma (PCNSL) over the last decades, the 5-year survival rate is approximately 30%; thus, new therapeutic approaches are needed to improve patient survival. The study's aim was to evaluate the role of surgical resection of PCNSL. Methods Primary outcomes were the overall survival (OS) and progression-free survival (PFS) of patients with PCNSL who underwent surgical resection versus biopsy alone. The meta-analysis was conducted to calculate pooled hazard ratios (HRs) under a random-effects model for the time-to-event variables. The odds ratios (ORs) were calculated for binary, secondary outcome parameters. Results Seven studies (n = 1046) were included. We found that surgical resection was associated with significantly better OS (HR 0.63 [95% CI 0.51-0.77]) when compared with biopsy. PFS was also significantly improved (HR 0.64 [95% CI 0.49-0.85]) in patients who underwent resection compared with those who underwent biopsy. The heterogeneity for OS and PFS was low (I2 = 7% and 24%, respectively). We also found that patients who underwent biopsy more often had multiple (OR 0.38 [95% CI 0.19-0.79]) or deep-seated (OR 0.20 [95% CI 0.12-0.34]) lesions compared with those who underwent surgical resection. There were no significant differences in chemotherapy or radiotherapy use or the occurrence of postoperative complications between the two groups. Conclusion In selected patients, surgical resection of PCNSL is associated with significantly better overall survival and progression-free survival compared with biopsy alone.
C1 [Chojak, Rafal; Polanska, Karolina; Rojek, Marta; Chojko, Aleksandra; Bogacz, Rafal; Skorupa, Natalia; Wieclaw, Jakub] Wroclaw Med Univ, Fac Med, Ludw Pasteura 1, PL-50367 Wroclaw, Poland.
   [Chojak, Rafal; Kozba-Gosztyla, Marta; Czapiga, Bogdan] 4th Mil Hosp Wroclaw, Dept Neurosurg, Wroclaw, Poland.
   [Czapiga, Bogdan] Wroclaw Med Univ, Fac Hlth Sci, Dept Nervous Syst Dis, Wroclaw, Poland.
C3 Wroclaw Medical University; Wroclaw Medical University
RP Chojak, R (corresponding author), Wroclaw Med Univ, Fac Med, Ludw Pasteura 1, PL-50367 Wroclaw, Poland.; Chojak, R (corresponding author), 4th Mil Hosp Wroclaw, Dept Neurosurg, Wroclaw, Poland.
EM
OI Chojak, Rafal/0000-0002-7644-3444; Czapiga, Bogdan/0000-0002-9819-2455
CR Abrey LE, 2006, J CLIN ONCOL, V24, P5711, DOI 10.1200/JCO.2006.08.2941
   Ahn Y, 2017, BLOOD RES, V52, P285, DOI 10.5045/br.2017.52.4.285
   Bataille B, 2000, J NEUROSURG, V92, P261, DOI 10.3171/jns.2000.92.2.0261
   Bühring U, 2001, NEUROLOGY, V57, P393, DOI 10.1212/WNL.57.3.393
   Chamberlain MC, 2010, NEURO-ONCOLOGY, V12, P736, DOI 10.1093/neuonc/noq011
   Citterio G, 2017, CRIT REV ONCOL HEMAT, V113, P97, DOI 10.1016/j.critrevonc.2017.03.019
   Cloney MB, 2017, J NEURO-ONCOL, V132, P189, DOI 10.1007/s11060-016-2358-8
   Deng XY, 2020, FRONT ONCOL, V10, DOI 10.3389/fonc.2020.00131
   Ferreri AJM, 2003, J CLIN ONCOL, V21, P266, DOI 10.1200/JCO.2003.09.139
   Ferreri AJM, 2022, LEUKEMIA, V36, P1870, DOI 10.1038/s41375-022-01582-5
   Ferreri AJM, 2016, LANCET HAEMATOL, V3, pE217, DOI 10.1016/S2352-3026(16)00036-3
   Ferreri AJM, 2009, LANCET, V374, P1512, DOI 10.1016/S0140-6736(09)61416-1
   Fox CP, 2019, BRIT J HAEMATOL, V184, P348, DOI 10.1111/bjh.15661
   Villalonga JF, 2018, ARQ NEURO-PSIQUIAT, V76, P139, DOI [10.1590/0004-282X20180002, 10.1590/0004-282x20180002]
   Giannini C, 2014, J NEUROPATH EXP NEUR, V73, P478, DOI 10.1097/NEN.0000000000000076
   Green K., 2022, STATPEARLS
   Haldorsen IS, 2011, AM J NEURORADIOL, V32, P984, DOI 10.3174/ajnr.A2171
   Higgins JPT, 2003, BMJ-BRIT MED J, V327, P557, DOI 10.1136/bmj.327.7414.557
   Hiraga S, 1999, J NEUROSURG, V91, P221, DOI 10.3171/jns.1999.91.2.0221
   Hoang-Xuan K, 2023, NEURO-ONCOLOGY, V25, P37, DOI 10.1093/neuonc/noac196
   Hoang-Xuan K, 2015, LANCET ONCOL, V16, pE322, DOI 10.1016/S1470-2045(15)00076-5
   Houillier C, 2020, NEUROLOGY, V94, pE1027, DOI 10.1212/WNL.0000000000008900
   Jahr G, 2018, NEUROSURG REV, V41, P1037, DOI 10.1007/s10143-018-0946-0
   Jahr G, 2018, BRAIN BEHAV, V8, DOI 10.1002/brb3.928
   Kasenda B, 2015, ANN ONCOL, V26, P1305, DOI 10.1093/annonc/mdv076
   Kinslow CJ, 2020, BRIT J NEUROSURG, V34, P690, DOI 10.1080/02688697.2019.1710820
   Küker W, 2005, J NEURO-ONCOL, V72, P169, DOI 10.1007/s11060-004-3390-7
   Labak CM, 2019, WORLD NEUROSURG, V126, pE1436, DOI 10.1016/j.wneu.2019.02.252
   Langner-Lemercier S, 2016, NEURO-ONCOLOGY, V18, P1297, DOI 10.1093/neuonc/now033
   Li Q, 2021, CANCER MANAG RES, V13, P6115, DOI 10.2147/CMAR.S322467
   McGrath S, 2020, BIOMETRICAL J, V62, P69, DOI 10.1002/bimj.201900036
   Mendez JS, 2018, NEURO-ONCOLOGY, V20, P687, DOI 10.1093/neuonc/nox187
   Ostrom QT, 2021, NEURO-ONCOLOGY, V23, P1, DOI 10.1093/neuonc/noab200
   Ottawa Hospital Research Institute, About us
   Ouyang TH, 2020, WORLD NEUROSURG, V139, pE508, DOI 10.1016/j.wneu.2020.04.049
   Page MJ, 2021, BMJ-BRIT MED J, V372, DOI [10.1136/bmj.n71, 10.1016/j.ijsu.2021.105906, 10.1136/bmj.n160]
   Rae AI, 2019, NEUROSURGERY, V84, P935, DOI 10.1093/neuros/nyy096
   Rubenstein JL, 2013, J CLIN ONCOL, V31, P3061, DOI 10.1200/JCO.2012.46.9957
   Schellekes N, 2021, J NEUROSURG, V135, P1016, DOI 10.3171/2020.9.JNS201980
   Schlegel Uwe, 2009, Ther Adv Neurol Disord, V2, P93, DOI 10.1177/1756285608101222
   Shiels MS, 2016, BRIT J HAEMATOL, V174, P417, DOI 10.1111/bjh.14073
   Shin SM, 2016, J RADIOSURGERY SBRT, V4, P247
   Song K-W, 2021, Ann Lymphoma, V5, P16, DOI [DOI 10.21037/AOL-20-50, 10.21037/aol-20-50]
   Sterne JAC, 2011, BMJ-BRIT MED J, V343, DOI 10.1136/bmj.d4002
   Thiel E, 2010, LANCET ONCOL, V11, P1036, DOI 10.1016/S1470-2045(10)70229-1
   Tierney JF, 2007, TRIALS, V8, DOI 10.1186/1745-6215-8-16
   Villano JL, 2011, BRIT J CANCER, V105, P1414, DOI 10.1038/bjc.2011.357
   von Baumgarten L, 2018, DTSCH ARZTEBL INT, V115, P419, DOI 10.3238/arztebl.2018.0419
   Weller M, 2012, NEURO-ONCOLOGY, V14, P1481, DOI 10.1093/neuonc/nos159
   Wu SQ, 2021, BMC NEUROL, V21, DOI 10.1186/s12883-021-02227-3
   Yu JY, 2021, SCI REP-UK, V11, DOI 10.1038/s41598-020-80724-0
   Yun J, 2017, FRONT NEUROL, V8, DOI 10.3389/fneur.2017.00478
   Zhang QJ, 2022, TURK NEUROSURG, V32, P1, DOI 10.5137/1019-5149.JTN.24054-19.2
NR 53
TC 1
Z9 1
U1 0
U2 2
PU SPRINGER
PI NEW YORK
PA ONE NEW YORK PLAZA, SUITE 4600, NEW YORK, NY, UNITED STATES
SN 0167-594X
EI 1573-7373
J9 J NEURO-ONCOL
JI J. Neuro-Oncol.
PD DEC
PY 2022
VL 160
IS 3
BP 753
EP 761
DI 10.1007/s11060-022-04200-7
EA NOV 2022
PG 9
WC Oncology; Clinical Neurology
WE Science Citation Index Expanded (SCI-EXPANDED)
SC Oncology; Neurosciences & Neurology
GA 7F2YV
UT WOS:000892894600001
PM 36449256
OA Green Published, hybrid
DA 2024-06-03
ER

PT J
AU Mo, SS
   Cleveland, J
   Rubenstein, JL
AF Mo, Shirley S.
   Cleveland, Joseph
   Rubenstein, James L.
TI Primary CNS lymphoma: update on molecular pathogenesis and therapy
SO LEUKEMIA & LYMPHOMA
LA English
DT Review
DE Primary CNS lymphoma; brain tumor; NF-kappa B; IMiDs; Bruton's tyrosine
   kinase
ID CENTRAL-NERVOUS-SYSTEM; WHOLE-BRAIN RADIOTHERAPY; INTERNATIONAL
   EXTRANODAL LYMPHOMA; STEM-CELL TRANSPLANTATION; HIGH-DOSE METHOTREXATE;
   NON-HODGKIN-LYMPHOMA; PHASE-II; GENE-EXPRESSION; CYTARABINE; RITUXIMAB
AB Primary central nervous system lymphoma (PCNSL) is a rare and aggressive form of extra-nodal non-Hodgkin lymphoma that as a brain tumor poses a unique set of challenges in diagnosis and management. With the advent of next-generation sequencing, we review updates in the understanding of its molecular and genomic pathogenesis. We also highlight key issues in management, with a focus on emerging technologies and new biological therapies including monoclonal antibodies, IMiDs, BTK inhibitors, PD-1 inhibitors, and CAR-T therapy. Integration of these approaches will likely enhance induction and consolidation strategies to suppress NF-kappa B activation and the anti-tumor immune response, while minimizing the often noxious effects of genotoxic approaches.
C1 [Mo, Shirley S.; Cleveland, Joseph; Rubenstein, James L.] Univ Calif San Francisco, Dept Med, San Francisco, CA 94143 USA.
   [Rubenstein, James L.] Univ Calif San Francisco, Hematol Oncol, San Francisco, CA 94143 USA.
   [Rubenstein, James L.] Univ Calif San Francisco, Helen Diller Family Comprehens Canc Ctr, Dept Med, San Francisco, CA 94143 USA.
C3 University of California System; University of California San Francisco;
   University of California System; University of California San Francisco;
   University of California System; University of California San Francisco;
   UCSF Medical Center; UCSF Helen Diller Family Comprehensive Cancer
   Center
RP Rubenstein, JL (corresponding author), Univ Calif San Francisco, Div Hematol Oncol, M1282 Box 1270, San Francisco, CA 94143 USA.
EM
FU NIH [R01CA139-83-01A1]
FX Supported by NIH R01CA139-83-01A1.
CR Abrey LE, 1998, J CLIN ONCOL, V16, P859, DOI 10.1200/JCO.1998.16.3.859
   Ambady P, 2019, LEUKEMIA LYMPHOMA, V60, P515, DOI 10.1080/10428194.2018.1480771
   Barajas RF, 2021, NEURO-ONCOLOGY, V23, P1056, DOI 10.1093/neuonc/noab020
   Batchelor T, 2021, J CLIN ONCOL, V39, DOI 10.1200/JCO.2021.39.15_suppl.7506
   Birsen R, 2018, HAEMATOLOGICA, V103, pE296, DOI 10.3324/haematol.2017.185843
   Bobillo S, 2021, HAEMATOLOGICA, V106, P513, DOI 10.3324/haematol.2019.241208
   Braaten KM, 2003, CLIN CANCER RES, V9, P1063
   Braggio E, 2015, CLIN CANCER RES, V21, P3986, DOI 10.1158/1078-0432.CCR-14-2116
   Bromberg JEC, 2019, LANCET ONCOL, V20, P216, DOI 10.1016/S1470-2045(18)30747-2
   Carpio C, 2020, BLOOD, V135, P996, DOI 10.1182/blood.2019002395
   Chapuy B, 2016, BLOOD, V127, P869, DOI 10.1182/blood-2015-10-673236
   DEANGELIS LM, 1992, J CLIN ONCOL, V10, P635, DOI 10.1200/JCO.1992.10.4.635
   DeAngelis LM, 2002, J CLIN ONCOL, V20, P4643, DOI 10.1200/JCO.2002.11.013
   ERVIN T, 1980, CANCER, V45, P1556, DOI 10.1002/1097-0142(19800401)45:7<1556::AID-CNCR2820450707>3.0.CO;2-B
   Ferreri AJM, 2017, LANCET HAEMATOL, V4, pE510, DOI 10.1016/S2352-3026(17)30174-6
   Ferreri AJM, 2016, LANCET HAEMATOL, V3, pE217, DOI 10.1016/S2352-3026(16)00036-3
   Frigault MJ, 2022, BLOOD, V139, P2306, DOI 10.1182/blood.2021014738
   Fukumura K, 2016, ACTA NEUROPATHOL, V131, P865, DOI 10.1007/s00401-016-1536-2
   Gandhi MK, 2021, BLOOD, V137, P1468, DOI 10.1182/blood.2020008520
   Geng HM, 2021, NEURO-ONCOLOGY, V23, P1668, DOI 10.1093/neuonc/noab045
   Ghesquieres H, 2019, ANN ONCOL, V30, P621, DOI 10.1093/annonc/mdz032
   Gonzalez-Aguilar A, 2012, CLIN CANCER RES, V18, P5203, DOI 10.1158/1078-0432.CCR-12-0845
   Grommes C, 2019, BLOOD, V133, P436, DOI 10.1182/blood-2018-09-875732
   Grommes C, 2017, CANCER DISCOV, V7, P1018, DOI 10.1158/2159-8290.CD-17-0613
   Guney E, 2022, BLOOD ADV, V6, P3189, DOI 10.1182/bloodadvances.2021006018
   Hoang-Xuan K, 2020, BLOOD, V136, DOI 10.1182/blood-2020-141773
   Houillier C, 2019, J CLIN ONCOL, V37, P823, DOI 10.1200/JCO.18.00306
   Illerhaus G, 2016, LANCET HAEMATOL, V3, pE388, DOI 10.1016/S2352-3026(16)30050-3
   Vu K, 2019, BRIT J HAEMATOL, V186, P180, DOI 10.1111/bjh.15787
   Kim S, 2016, BMC CANCER, V16, DOI 10.1186/s12885-016-2397-8
   Kurtz DM, 2021, NAT BIOTECHNOL, V39, P1537, DOI 10.1038/s41587-021-00981-w
   Lai R, 2002, NEUROLOGY, V59, P1557, DOI 10.1212/01.WNL.0000034256.20173.EA
   Lionakis MS, 2017, CANCER CELL, V31, P833, DOI 10.1016/j.ccell.2017.04.012
   Lv CL, 2022, THER ADV HEMATOL, V13, DOI 10.1177/20406207211066166
   Mahale P, 2018, AM J TRANSPLANT, V18, P453, DOI 10.1111/ajt.14465
   Majumdar S, 2001, J IMMUNOL, V167, P2911, DOI 10.4049/jimmunol.167.5.2911
   Margold M, 2021, NEURO-ONCOLOGY, V23, P2076, DOI 10.1093/neuonc/noab109
   Mondello P, 2020, HEMATOL ONCOL, V38, DOI 10.1002/hon.2679
   Montesinos-Rongen M, 2008, LEUKEMIA, V22, P400, DOI 10.1038/sj.leu.2405019
   Morris PG, 2013, J CLIN ONCOL, V31, P3971, DOI 10.1200/JCO.2013.50.4910
   Mutter JA., 2021, BLOOD, V138, P6, DOI [10.1182/blood-2021-149644, DOI 10.1182/BLOOD-2021-149644]
   Nakamura T, 2016, NEUROPATH APPL NEURO, V42, P279, DOI 10.1111/nan.12259
   Narita Y, 2021, NEURO-ONCOLOGY, V23, P122, DOI 10.1093/neuonc/noaa145
   National Cancer Institute (NCI), 2022, PHASE TRIAL METHOTRE
   Nayak L., 2017, Hematol Oncol, V35, P420, DOI [10.1002/hon.2440_2, DOI 10.1002/HON.2440_2]
   Nayak L, 2017, BLOOD, V129, P3071, DOI 10.1182/blood-2017-01-764209
   Nayyar N, 2019, BLOOD ADV, V3, P375, DOI 10.1182/bloodadvances.2018027672
   NELSON DF, 1992, INT J RADIAT ONCOL, V23, P9, DOI 10.1016/0360-3016(92)90538-S
   Olson JE, 2002, CANCER, V95, P1504, DOI 10.1002/cncr.10851
   Omuro A, 2015, LANCET HAEMATOL, V2, pE251, DOI 10.1016/S2352-3026(15)00074-5
   Omuro A, 2015, BLOOD, V125, P1403, DOI 10.1182/blood-2014-10-604561
   Omuro AMP, 2020, J CLIN ONCOL, V38
   Radke J, 2022, NAT COMMUN, V13, DOI 10.1038/s41467-022-30050-y
   Rubenstein JL, 2018, BLOOD ADV, V2, P1595, DOI 10.1182/bloodadvances.2017014845
   Rubenstein JL, 2013, J CLIN ONCOL, V31, P3061, DOI 10.1200/JCO.2012.46.9957
   Rubenstein JL, 2013, BLOOD, V121, P4740, DOI 10.1182/blood-2013-01-476333
   Rubenstein JL, 2006, BLOOD, V107, P3716, DOI 10.1182/blood-2005-03-0897
   Sasayama T, 2012, NEURO-ONCOLOGY, V14, P368, DOI 10.1093/neuonc/nor203
   Schabet M, 1999, J NEURO-ONCOL, V43, P199, DOI 10.1023/A:1006290032052
   Schmitt AM, 2019, HEMATOL ONCOL, V37, P548, DOI 10.1002/hon.2666
   Schmitz R, 2018, NEW ENGL J MED, V378, P1396, DOI 10.1056/NEJMoa1801445
   Scordo M, 2021, JAMA ONCOL, V7, P993, DOI 10.1001/jamaoncol.2021.1074
   Siddiqi T, 2021, BLOOD ADV, V5, P4059, DOI 10.1182/bloodadvances.2020004106
   SKARIN AT, 1977, BLOOD, V50, P1039
   Soussain C, 2019, EUR J CANCER, V117, P121, DOI 10.1016/j.ejca.2019.05.024
   Tan WJ, 2019, BLOOD, V134, P709, DOI 10.1182/blood.2019000022
   Thomas S, 2015, PLOS ONE, V10, DOI 10.1371/journal.pone.0130078
   Tun HW, 2018, BLOOD, V132, P2240, DOI 10.1182/blood-2018-02-835496
   van der Meulen M, 2021, J NEURO-ONCOL, V152, P357, DOI 10.1007/s11060-021-03708-8
   Vater I, 2015, LEUKEMIA, V29, P677, DOI 10.1038/leu.2014.264
   Villa D, 2019, BLOOD ADV, V3, P3953, DOI 10.1182/bloodadvances.2019000989
   Wieduwilt MJ, 2012, CLIN CANCER RES, V18, P1146, DOI 10.1158/1078-0432.CCR-11-0625
   Zinzani PL, 2021, CLIN CANCER RES, V27, P6124, DOI 10.1158/1078-0432.CCR-21-1471
NR 73
TC 1
Z9 1
U1 1
U2 4
PU TAYLOR & FRANCIS LTD
PI ABINGDON
PA 2-4 PARK SQUARE, MILTON PARK, ABINGDON OR14 4RN, OXON, ENGLAND
SN 1042-8194
EI 1029-2403
J9 LEUKEMIA LYMPHOMA
JI Leuk. Lymphoma
PD JAN 2
PY 2023
VL 64
IS 1
BP 57
EP 65
DI 10.1080/10428194.2022.2133541
EA OCT 2022
PG 9
WC Oncology; Hematology
WE Science Citation Index Expanded (SCI-EXPANDED)
SC Oncology; Hematology
GA 9E4XE
UT WOS:000873532900001
PM 36286546
DA 2024-06-03
ER

PT J
AU Li, XT
   Xiong, H
AF Li, Xintong
   Xiong, Hua
TI Case report: Ventricular primary central nervous system lymphoma with
   partial hypointensity on diffusion-weighted imaging
SO FRONTIERS IN NEUROLOGY
LA English
DT Article
DE primary central nervous system lymphoma; third ventricle; diffusion
   weighted imaging; T2 blackout effect; case report; ventricular PCNSL
ID BIOLOGY; MRI
AB IntroductionPrimary central nervous system lymphoma (PCNSL) is infrequent and represents 3. 1% of primary brain tumors. And the lesions that are restricted to the ventricular system, particularly the third ventricle, are even rarer. There are few pieces of literature or case reports to date. We report a case of PCNSL with partial hypointense on diffusion-weighted imaging (DWI) located in the lateral and third ventricles. Then we reviewed almost all case reports of ventricular PCNSLs in the last 20 years, discuss the imaging presentation, other ventricular tumors with similar imaging findings, and primary treatment measures. Case presentationA 78-year-old man presented with memory loss and poor responsiveness for one week without obvious precipitating factors. Magnetic resonance imaging (MRI) showed lesions in the third ventricle and left lateral ventricles, which were slightly hypointense on T1-weighted imaging (T1WI), and isointense to slightly hypointense on T2-weighted imaging (T2WI). On DWI, the left lateral ventricular lesion was hyperintense, while the third ventricular lesion was hypointense. After the surgical procedure, the pathology and immunohistochemistry revealed diffuse large B-cell lymphoma (DLBCL). ConclusionsVentricular PCNSL is quite rare, and may be confused with other tumors in the same position. However, PCNSL differs from other central nervous system tumors in that it is primarily treated with chemotherapy and/or radiation therapy. So, it is important to recognize PCNSL and differentiate it from other tumors, considering its implications for management planning.
C1 [Li, Xintong] Chongqing Gen Hosp, Dept Radiol, Chongqing, Peoples R China.
   [Xiong, Hua] Peoples Hosp Shapingba Dist, Dept Radiol, Chongqing, Peoples R China.
RP Li, XT (corresponding author), Chongqing Gen Hosp, Dept Radiol, Chongqing, Peoples R China.
EM
CR Algazi AP, 2009, NEUROTHERAPEUTICS, V6, P587, DOI 10.1016/j.nurt.2009.04.013
   Ball MK, 2020, BRAIN TUMOR PATHOL, V37, P22, DOI 10.1007/s10014-019-00354-x
   Bataille B, 2000, J NEUROSURG, V92, P261, DOI 10.3171/jns.2000.92.2.0261
   Brastianos PK, 2012, HEMATOL ONCOL CLIN N, V26, P897, DOI 10.1016/j.hoc.2012.05.003
   Brostigen CS, 2017, ACTA NEUROL SCAND, V135, P484, DOI 10.1111/ane.12632
   Chen X, 2020, FRONT ONCOL, V10, DOI 10.3389/fonc.2020.00502
   Choudhari KA, 2009, BRIT J NEUROSURG, V23, P585, DOI 10.3109/02688690903254350
   Deckert M, 2011, LEUKEMIA, V25, P1797, DOI 10.1038/leu.2011.169
   Erbay MF, 2020, BRIT J RADIOL, V93, DOI 10.1259/bjr.20200552
   Funaro K, 2014, J RADIOL CASE REP, V8, P1, DOI 10.3941/jrcr.v8i3.1361
   Guo RJ, 2019, BMC NEUROL, V19, DOI 10.1186/s12883-019-1430-3
   Haddad R, 2019, WORLD NEUROSURG, V123, P286, DOI 10.1016/j.wneu.2018.12.026
   Haegelen C, 2001, J NEURO-ONCOL, V51, P129, DOI 10.1023/A:1010790325692
   Hiwatashi A, 2003, AM J ROENTGENOL, V181, P1705, DOI 10.2214/ajr.181.6.1811705
   Hsu HI, 2015, INT J SURG CASE REP, V14, P23, DOI 10.1016/j.ijscr.2015.07.006
   Koeller KK, 2002, RADIOGRAPHICS, V22, P1473, DOI 10.1148/rg.226025118
   Liao CH, 2014, J CLIN NEUROSCI, V21, P180, DOI 10.1016/j.jocn.2013.02.036
   Maldjian JA, 2001, AM J NEURORADIOL, V22, P112
   Nowak J, 2014, AM J NEURORADIOL, V35, P1996, DOI 10.3174/ajnr.A4002
   Philippart M, 2019, ACTA NEUROL BELG, V119, P119, DOI 10.1007/s13760-018-1017-6
   Sasani Mehdi, 2011, J Med Case Rep, V5, P213, DOI 10.1186/1752-1947-5-213
   Suri V, 2015, J PEDIATR NEUROSCI, V10, P393, DOI 10.4103/1817-1745.174433
   Taschner CA, 2018, CLIN NEURORADIOL, V28, P139, DOI 10.1007/s00062-018-0668-2
   von Baumgarten L, 2018, DTSCH ARZTEBL INT, V115, P419, DOI 10.3238/arztebl.2018.0419
   Yang DX, 2021, J CRANIOFAC SURG, V32, pE311, DOI 10.1097/SCS.0000000000002514
   Zhu Y, 2015, TURK NEUROSURG, V25, P493, DOI 10.5137/1019-5149.JTN.10496-14.1
NR 26
TC 1
Z9 1
U1 0
U2 8
PU FRONTIERS MEDIA SA
PI LAUSANNE
PA AVENUE DU TRIBUNAL FEDERAL 34, LAUSANNE, CH-1015, SWITZERLAND
SN 1664-2295
J9 FRONT NEUROL
JI Front. Neurol.
PD OCT 21
PY 2022
VL 13
AR 923206
DI 10.3389/fneur.2022.923206
PG 6
WC Clinical Neurology; Neurosciences
WE Science Citation Index Expanded (SCI-EXPANDED)
SC Neurosciences & Neurology
GA 5Y4FG
UT WOS:000879238400001
PM 36341101
OA Green Published, gold
DA 2024-06-03
ER

PT J
AU Ruan, HY
   Wang, Z
   Chen, K
   Tang, XM
   Ding, TL
   Zhang, C
   Guan, M
AF Ruan, Haoyu
   Wang, Zhe
   Chen, Kun
   Tang, Xuemei
   Ding, Tianling
   Zhang, Chao
   Guan, Ming
TI The characteristics of cerebrospinal fluid anaplastic large cells in a
   patient with primary leptomeningeal anaplastic large cell lymphoma
SO CLINICA CHIMICA ACTA
LA English
DT Article
DE Cerebrospinal fluid; Primary leptomeningeal anaplastic large cell
   lymphoma; Single cell transcriptome
ID EXPRESSION
AB Background: Primary central nervous system lymphomas (PCNSL) anaplastic large cell lymphoma (ALCL) are very rare non-Hodgkin's lymphomas, especially in the leptomeninges. Until now, the diagnostics and therapeutics of PCNSL-ALCL is a challenge and urgent need. A 26-y Chinese male presented altered awareness and severe headache.
   Methods: A 26-y Chinese male presented altered awareness and severe headache. Brain magnetic resonance imaging (MRI) delineated no intracerebral lesions and focal leptomeningeal enhancement. Diagnosis was based on cerebrospinal fluid (CSF) examination discovering anaplastic large cells (ALCs) with positive expression for anaplastic lymphoma kinase (ALK) and CD30, and no evidence of systemic involvement. In addition, we firstly performed the single-cell RNA sequencing to identify transcriptome characteristics of CSF-ALCs.
   Results: The case was diagnosed as a rare primary leptomeningeal anaplastic large cell lymphoma (PL-ALCL). Four cycles of systemic chemotherapy with methotrexate and intrathecal cytarabine help achieve complete remission. Compared to normal T cells, 45 genes were specifically upregulated in CSF-ALCs. CSF-ALCs enriched cell proliferation and metabolism pathway and lost features of immune identity. The heterogeneity of CSF-ALCs were manifested in the expression of cancer-testis antigens and cell-cycle signature genes. In addition, the gene regulatory networks (GRNs) revealed the activity of transcription factors EZH2 and NFYC were upregulated in the CSF-ALCs.
   Conclusion: The first analysis of single-cell transcriptome signatures of CSF-ALCs will provide clues for diagnosis and mechanism research of PL-ALCL.
C1 [Ruan, Haoyu; Chen, Kun; Tang, Xuemei; Guan, Ming] Fudan Univ, Huashan Hosp, Dept Lab Med, Shanghai, Peoples R China.
   [Wang, Zhe] Second Mil Med Univ, Dept Physiol, Shanghai 200433, Peoples R China.
   [Zhang, Chao] Tongji Univ, Shanghai YangZhi Rehabil Hosp, Fundamental Res Ctr, Shanghai Sunshine Rehabil Ctr, Shanghai 201619, Peoples R China.
   [Ruan, Haoyu] Nanjing Med Univ, Dept Lab Med, Affiliated Hosp 1, Nanjing, Peoples R China.
   [Ding, Tianling] Fudan Univ, Huashan Hosp, Dept Hematol, Shanghai, Peoples R China.
C3 Fudan University; Naval Medical University; Tongji University; Nanjing
   Medical University; Fudan University
RP Guan, M (corresponding author), Fudan Univ, Huashan Hosp, Dept Lab Med, Shanghai, Peoples R China.; Zhang, C (corresponding author), Tongji Univ, Shanghai YangZhi Rehabil Hosp, Fundamental Res Ctr, Shanghai Sunshine Rehabil Ctr, Shanghai 201619, Peoples R China.
EM
OI Guan, Ming/0000-0002-8796-2653; Ruan, Haoyu/0000-0001-6001-9705
FU National Natural Science Foundation of China [82072367, 82102489,
   32271165]
FX Acknowledgements: This work was funded by the National Natural Science
   Foundation of China,grant number 82072367, 82102489, and 32271165.
CR Aibar S, 2017, NAT METHODS, V14, P1083, DOI [10.1038/NMETH.4463, 10.1038/nmeth.4463]
   Bataille B, 2000, J NEUROSURG, V92, P261, DOI 10.3171/jns.2000.92.2.0261
   Dunbar MJ, 2014, PEDIATR NEUROSURG, V50, P145, DOI 10.1159/000380769
   Duplantier MM, 2006, LEUKEMIA, V20, P1848, DOI 10.1038/sj.leu.2404352
   Felice MS, 2000, MED PEDIATR ONCOL, V34, P361
   Hirano Y, 2021, CANCERS, V13, DOI 10.3390/cancers13174358
   Lamant L, 2007, BLOOD, V109, P2156, DOI 10.1182/blood-2006-06-028969
   Nam SJ, 2016, HISTOL HISTOPATHOL, V31, P213, DOI 10.14670/HH-11-669
   Otasek D, 2019, GENOME BIOL, V20, DOI 10.1186/s13059-019-1758-4
   Pearson JD, 2014, MOL CANCER, V13, DOI 10.1186/1476-4598-13-199
   Ruan HY, 2021, ISCIENCE, V24, DOI 10.1016/j.isci.2021.102972
   Ruan HY, 2020, CLIN TRANSL MED, V10, DOI 10.1002/ctm2.246
   Taga T, 2007, PEDIATR BLOOD CANCER, V48, P477, DOI 10.1002/pbc.20704
   Yoshihara K, 2013, NAT COMMUN, V4, DOI 10.1038/ncomms3612
NR 14
TC 1
Z9 1
U1 0
U2 1
PU ELSEVIER
PI AMSTERDAM
PA RADARWEG 29, 1043 NX AMSTERDAM, NETHERLANDS
SN 0009-8981
EI 1873-3492
J9 CLIN CHIM ACTA
JI Clin. Chim. Acta
PD DEC 1
PY 2022
VL 537
BP 46
EP 50
DI 10.1016/j.cca.2022.09.029
EA OCT 2022
PG 5
WC Medical Laboratory Technology
WE Science Citation Index Expanded (SCI-EXPANDED)
SC Medical Laboratory Technology
GA 6A4KD
UT WOS:000880624600006
PM 36243040
DA 2024-06-03
ER

PT J
AU Kaulen, LD
   Baehring, JM
AF Kaulen, Leon D.
   Baehring, Joachim M.
TI Treatment Options for Recurrent Primary CNS Lymphoma
SO CURRENT TREATMENT OPTIONS IN ONCOLOGY
LA English
DT Review
DE Primary CNS lymphoma; Non-Hodgkin lymphoma; Recurrence; CAR-T cell
   therapy; Immunotherapy; Methotrexate
ID CENTRAL-NERVOUS-SYSTEM; WHOLE-BRAIN RADIOTHERAPY; HIGH-DOSE
   METHOTREXATE; INTERNATIONAL EXTRANODAL LYMPHOMA; STEM-CELL
   TRANSPLANTATION; SALVAGE THERAPY; PHASE-II; INTENSIVE CHEMOTHERAPY;
   INTRAOCULAR LYMPHOMA; PROSPECTIVE TRIAL
