## Supplementary material for "Bibliometric analysis of emerging trend and top 100 most cited articles in primary central nervous system lymphoma": Supple File 1: savedrecs.txt

FN Clarivate Analytics Web of Science
VR 1.0
PT J
AU Abrey, LE
   Batchelor, TT
   Ferreri, AJM
   Gospodarowicz, M
   Pulczynski, EJ
   Zucca, E
   Smith, JR
   Korfel, A
   Soussain, C
   DeAngelis, LM
   Neuwelt, EA
   O'Neill, BP
   Thiel, E
   Shenkier, T
   Graus, F
   van den Bent, M
   Seymour, JF
   Poortmans, P
   Armitage, JO
   Cavalli, F
AF Abrey, LE
   Batchelor, TT
   Ferreri, AJM
   Gospodarowicz, M
   Pulczynski, EJ
   Zucca, E
   Smith, JR
   Korfel, A
   Soussain, C
   DeAngelis, LM
   Neuwelt, EA
   O'Neill, BP
   Thiel, E
   Shenkier, T
   Graus, F
   van den Bent, M
   Seymour, JF
   Poortmans, P
   Armitage, JO
   Cavalli, F
TI Report of an international workshop to standardize baseline evaluation
   and response criteria for primary CNS lymphoma
SO JOURNAL OF CLINICAL ONCOLOGY
LA English
DT Article
ID CENTRAL-NERVOUS-SYSTEM; NON-HODGKINS-LYMPHOMA; CEREBROSPINAL-FLUID
   CYTOLOGY; HIGH-DOSE METHOTREXATE; PHASE-II; DEFERRED RADIOTHERAPY;
   EUROPEAN ORGANIZATION; INTRAOCULAR LYMPHOMA; CHEMOTHERAPY; CANCER
AB Standardized guidelines for the baseline evaluation and response assessment of primary CNS lymphoma (PCNSL) are critical to ensure comparability among clinical trials for newly diagnosed patients. The relative rarity of this tumor precludes rapid completion of large-scale phase III trials and, therefore, our reliance on the results of well-designed phase II trials is critical. To formulate this recommendation, an international group of experts representing hematologic oncology, medical oncology, neuro-oncology, neurology, radiation oncology, neurosurgery, and ophthalmology met to review current standards of reporting and to formulate a consensus opinion regarding minimum baseline evaluation and common standards for assessing response to therapy. The response guidelines were based on the results of neuroimaging, corticosteroid use, ophthalmologic examination, and CSF cytology. A critical issue that requires additional study is the optimal method to assess the neurocognitive impact of therapy and address the quality of life of PCNSL survivors. We hope that these guidelines will improve communication among investigators and comparability among clinical trials in a way that will allow us to develop better therapies for patients.
C1 Mem Sloan Kettering Canc Ctr, Dept Neurol, New York, NY 10021 USA.
C3 Memorial Sloan Kettering Cancer Center
RP Abrey, LE (corresponding author), Mem Sloan Kettering Canc Ctr, Dept Neurol, 1275 York Ave, New York, NY 10021 USA.
EM
RI van den Bent, Martin J/JPK-6126-2023; Poortmans, P.M.P./L-4581-2015;
   Wolf, Max/IRZ-4746-2023; Graus, Francesc/K-9904-2018; Ferreri, Andrés J.
   M./A-6662-2013; Seymour, John/K-7326-2019; Smith, Justine/Y-9044-2019
OI van den Bent, Martin J/0000-0001-5710-5127; Seymour,
   John/0000-0003-2188-6835; Smith, Justine/0000-0002-4756-5493; DeAngelis,
   Lisa/0000-0002-6290-7818; Graus, Francesc/0000-0002-8924-8322; Zucca,
   Emanuele/0000-0002-5522-6109
CR Abrey LE, 2000, J CLIN ONCOL, V18, P3144, DOI 10.1200/JCO.2000.18.17.3144
   Batchelor T, 2003, J CLIN ONCOL, V21, P1044, DOI 10.1200/JCO.2003.03.036
   Bessell EM, 2002, J CLIN ONCOL, V20, P231, DOI 10.1200/JCO.20.1.231
   Bessell EM, 2001, INT J RADIAT ONCOL, V50, P457, DOI 10.1016/S0360-3016(01)01451-1
   Brady L W, 1996, Oncology (Williston Park), V10, P981
   Buccheri G, 1996, EUR J CANCER, V32A, P1135, DOI 10.1016/0959-8049(95)00664-8
   Chamberlain MC, 2001, NEURO-ONCOLOGY, V3, P42, DOI 10.1093/neuonc/3.1.42
   Chan Chi-Chao, 2004, Cancer Control, V11, P285
   Cheson BD, 1999, J CLIN ONCOL, V17, P1244, DOI 10.1200/JCO.1999.17.4.1244
   Correa DD, 2004, NEUROLOGY, V62, P548, DOI 10.1212/01.WNL.0000109673.75316.D8
   DEANGELIS LM, 1992, J CLIN ONCOL, V10, P635, DOI 10.1200/JCO.1992.10.4.635
   DeAngelis LM, 2002, J CLIN ONCOL, V20, P4643, DOI 10.1200/JCO.2002.11.013
   Ferreri AJM, 1996, CANCER, V77, P827, DOI 10.1002/(SICI)1097-0142(19960301)77:5<827::AID-CNCR4>3.0.CO;2-B
   Ferreri AJM, 2003, J CLIN ONCOL, V21, P266, DOI 10.1200/JCO.2003.09.139
   Ferreri AJM, 2004, BRIT J CANCER, V90, P353, DOI 10.1038/sj.bjc.6601472
   Ferreri AJM, 2003, J CLIN ONCOL, V21, P1649, DOI 10.1200/JCO.2003.99.053
   Glantz MJ, 1998, CANCER, V82, P733, DOI 10.1002/(SICI)1097-0142(19980215)82:4<733::AID-CNCR17>3.0.CO;2-Z
   Hegde U, 2005, BLOOD, V105, P496, DOI 10.1182/blood-2004-05-1982
   Herrlinger U, 2002, ANN NEUROL, V51, P247, DOI 10.1002/ana.10102
   Hoang-Xuan K, 2003, J CLIN ONCOL, V21, P2726, DOI 10.1200/JCO.2003.11.036
   Hoffman PM, 2003, EYE, V17, P513, DOI 10.1038/sj.eye.6700378
   LOEFFLER JS, 1985, J CLIN ONCOL, V3, P490, DOI 10.1200/JCO.1985.3.4.490
   MACDONALD DR, 1990, J CLIN ONCOL, V8, P1277, DOI 10.1200/JCO.1990.8.7.1277
   Mead GM, 2000, CANCER, V89, P1359, DOI 10.1002/1097-0142(20000915)89:6<1359::AID-CNCR21>3.0.CO;2-9
   NELSON DF, 1992, INT J RADIAT ONCOL, V23, P9, DOI 10.1016/0360-3016(92)90538-S
   Neuwelt EA, 2005, AM J NEURORADIOL, V26, P258
   O'Brien P, 2000, J CLIN ONCOL, V18, P519, DOI 10.1200/JCO.2000.18.3.519
   Olson JE, 2002, CANCER, V95, P1504, DOI 10.1002/cncr.10851
   ONEILL BP, 1995, J NEURO-ONCOL, V25, P67, DOI 10.1007/BF01054724
   ONEILL BP, 1995, INT J RADIAT ONCOL, V33, P663, DOI 10.1016/0360-3016(95)00207-F
   Pels H, 2003, J CLIN ONCOL, V21, P4489, DOI 10.1200/JCO.2003.04.056
   Poortmans PMP, 2003, J CLIN ONCOL, V21, P4483, DOI 10.1200/JCO.2003.03.108
   Sandor V, 1998, J CLIN ONCOL, V16, P3000, DOI 10.1200/JCO.1998.16.9.3000
   SHAH G, 2004, AM SOC CLIN ONC ANN
   SHIPP MA, 1993, NEW ENGL J MED, V329, P987, DOI 10.1056/nejm199309303291402
   Wu HG, 1999, J KOREAN MED SCI, V14, P565, DOI 10.3346/jkms.1999.14.5.565
NR 36
TC 593
Z9 629
U1 0
U2 19
PU AMER SOC CLINICAL ONCOLOGY
PI ALEXANDRIA
PA 330 JOHN CARLYLE ST, STE 300, ALEXANDRIA, VA 22314 USA
SN 0732-183X
J9 J CLIN ONCOL
JI J. Clin. Oncol.
PD AUG 1
PY 2005
VL 23
IS 22
BP 5034
EP 5043
DI 10.1200/JCO.2005.13.524
PG 10
WC Oncology
WE Science Citation Index Expanded (SCI-EXPANDED)
SC Oncology
GA 952CL
UT WOS:000230979300038
PM 15955902
DA 2024-06-03
ER

PT J
AU Ferreri, AJM
   Blay, JY
   Reni, M
   Pasini, F
   Spina, M
   Ambrosetti, A
   Calderoni, A
   Rossi, A
   Vavassori, V
   Conconi, A
   Devizzi, L
   Berger, F
   Ponzoni, M
   Borisch, B
   Tinguely, M
   Cerati, M
   Milani, M
   Orvieto, E
   Sanchez, J
   Chevreau, C
   Dell'Oro, S
   Zucca, E
   Cavalli, F
AF Ferreri, AJM
   Blay, JY
   Reni, M
   Pasini, F
   Spina, M
   Ambrosetti, A
   Calderoni, A
   Rossi, A
   Vavassori, V
   Conconi, A
   Devizzi, L
   Berger, F
   Ponzoni, M
   Borisch, B
   Tinguely, M
   Cerati, M
   Milani, M
   Orvieto, E
   Sanchez, J
   Chevreau, C
   Dell'Oro, S
   Zucca, E
   Cavalli, F
TI Prognostic scoring system for primary CNS lymphomas: The international
   extranodal lymphoma study group experience
SO JOURNAL OF CLINICAL ONCOLOGY
LA English
DT Article; Proceedings Paper
CT 42nd Annual Meeting of the American-Society-of-Hematology
CY DEC 01-05, 2000
CL SAN FRANCISCO, CALIFORNIA
SP Amer Soc Hematol
ID CENTRAL-NERVOUS-SYSTEM; IMMUNOCOMPETENT PATIENTS; THERAPEUTIC
   MANAGEMENT; RADIOTHERAPY; SURVIVAL; CHEMOTHERAPY; PATTERNS; FAILURE
AB Purpose: To identify survival predictors and to design a prognostic score useful for distinguishing risk groups in immunocompetent patients with primary CNS lymphomas (PCNSL).
   Patients and Methods: The prognostic role of patient-, lymphoma-, and treatment-related variables was analyzed in a multicenter series of 378 PCNSL patients treated at 23 cancer centers from five different countries.
   Results: Age more than 60 years, performance status (PS) more than 1, elevated lactate dehydrogenase (LDH) serum level, high CSF protein concentration, and involvement of deep regions of the brain (periventricular regions, basal ganglia, brainstem, and/or cerebellum) were significantly and independently associated with a worse survival. These five variables were used to design a prognostic score. Each variable was assigned a value of either 0, if favorable, or 1, if unfavorable. The values were then added together to arrive at a final score, which was tested in 105 assessable patients for which complete data of all five variables were available. The 2-year overall survival (OS) +/- SD was 80% +/- 8%, 48% +/- 7%, and 15% +/- 7% (P = .0000 1) for patients with zero to one, two to three, and four to five unfavorable features, respectively. The prognostic role of this score was confirmed by limiting analysis to assessable patients treated with high-dose methotrexate-based chemotherapy (2-year OS +/- SD: 85% +/- 8%, 570% +/- 8%, and 24% +/- 11%; P = .0004).
   Conclusion: Age, PS, LDH serum level, CSF protein concentration, and involvement of deep structures of the brain were independent predictors of survival. A prognostic score including these five parameters seems advisable in distinguishing different risk groups in PCNSL patients. The proposed score and its relevance in therapeutic decision deserve to be validated in further studies. (C) 2003 by American Society of Clinical Oncology.
C1 San Raffaele H Sci Inst, Dept Radiochemotherapy, I-20132 Milan, Italy.
   San Raffaele H Sci Inst, Dept Pathol, I-20132 Milan, Italy.
   Ist Nazl Tumori, Div Med Oncol, I-20133 Milan, Italy.
   Univ Insubria, Osped Circolo, Div Radiotherapia, Varese, Italy.
   Univ Insubria, Fdn Macchi, Varese, Italy.
   Univ Insubria, Osped Circolo Fdn Macchi, Varese, Italy.
   Osped Civile, Div Clinicizzata Oncol Med, I-37126 Verona, Italy.
   Univ Verona, Osped Policlin Borgo Roma, Cattedra Ematol, I-37126 Verona, Italy.
   Azienda Osped Lecco, Div Anat Patol, Lecce, Italy.
   Natl Canc Inst, Ctr Riferimento Oncol, Div Med Oncol A, Aviano, Italy.
   Osped Riuniti Bergamo, Div Ematol, I-24100 Bergamo, Italy.
   Osped Regionale Treviso, Ist Anat & Istol Patol, Treviso, Italy.
   Hop Edouard Herriot, Unite Cytokines & Canc, Lyon, France.
   Ctr Leon Berard, F-69373 Lyon, France.
   Ctr Hosp Lyon Sud, Serv Pathol, Lyon, France.
   Ctr Claudius Regaud, Toulouse, France.
   Osped San Giovannia, Ist Oncol Svizzera Italiana, Bellinzona, Switzerland.
   Inselspital Bern, Inst Med Onkol, Bern, Switzerland.
   Hop Cantonal Univ Geneva, Div Pathol Clin, Geneva, Switzerland.
   Inst Enfermedades Neoplas, Dept Anat Pathol, Lima, Peru.
C3 Vita-Salute San Raffaele University; IRCCS Ospedale San Raffaele;
   Vita-Salute San Raffaele University; IRCCS Ospedale San Raffaele;
   Fondazione IRCCS Istituto Nazionale Tumori Milan; Ospedale Circolo &
   Fondazione Macchi; University of Insubria; University of Insubria;
   Ospedale Circolo & Fondazione Macchi; Ospedale Circolo & Fondazione
   Macchi; University of Insubria; University of Verona; Azienda
   Ospedaliera Universitaria Integrata Verona; IRCCS Aviano (CRO); Ospedali
   Riuniti di Bergamo; ULSS 2 Marca TV; Ospedale Ca' Foncello Treviso; CHU
   Lyon; UNICANCER; Centre Leon Berard; CHU Lyon; UNICANCER; Institut
   Claudius Regaud; Institute of Oncology Research (IOR); Regional Hospital
   of Bellinzona & Valleys, San Giovanni; University of Bern; University
   Hospital of Bern; University of Geneva; Instituto Nacional de
   Enfermedades Neoplasicas
RP Ferreri, AJM (corresponding author), San Raffaele H Sci Inst, Dept Radiochemotherapy, Via Olgettina 60, I-20132 Milan, Italy.
RI Chevreau, Christine/A-5565-2015; Blay, Jean-Yves/N-3966-2016; Ferreri,
   Andrés J. M./A-6662-2013; Spina, Michele/ABE-6014-2020; Borisch,
   Bettina/AAG-8212-2019; Reni, Michele/J-8296-2016; Devizzi,
   Liliana/F-5008-2017
OI Blay, Jean-Yves/0000-0001-7190-120X; Spina, Michele/0000-0001-6576-8182;
   Borisch, Bettina/0000-0002-9902-324X; Reni, Michele/0000-0002-6463-0293;
   Zucca, Emanuele/0000-0002-5522-6109; Sanchez,
   Juvenal/0000-0002-9825-8573; Devizzi, Liliana/0000-0002-1221-0326
CR Abrey LE, 2000, J CLIN ONCOL, V18, P3144, DOI 10.1200/JCO.2000.18.17.3144
   Bataille B, 2000, J NEUROSURG, V92, P261, DOI 10.3171/jns.2000.92.2.0261
   Bessell EM, 2002, J CLIN ONCOL, V20, P231, DOI 10.1200/JCO.20.1.231
   Blay JY, 1998, J CLIN ONCOL, V16, P864, DOI 10.1200/JCO.1998.16.3.864
   BLAY JY, 1993, BRIT J CANCER, V67, P1136, DOI 10.1038/bjc.1993.209
   Corry J, 1998, INT J RADIAT ONCOL, V41, P615, DOI 10.1016/S0360-3016(97)00571-3
   DEANGELIS LM, 1990, NEUROLOGY, V40, P80, DOI 10.1212/WNL.40.1.80
   Ferreri AJM, 1995, CANCER TREAT REV, V21, P415, DOI 10.1016/0305-7372(95)90028-4
   Ferreri AJM, 2000, ANN ONCOL, V11, P927, DOI 10.1023/A:1008376412784
   Ferreri AJM, 2002, NEUROLOGY, V58, P1513, DOI 10.1212/WNL.58.10.1513
   FINE HA, 1993, ANN INTERN MED, V119, P1093, DOI 10.7326/0003-4819-119-11-199312010-00007
   Hayabuchi N, 1999, INT J RADIAT ONCOL, V44, P265, DOI 10.1016/S0360-3016(98)00564-1
   HELLE TL, 1984, J NEUROSURG, V60, P94, DOI 10.3171/jns.1984.60.1.0094
   HOCHBERG FH, 1988, J NEUROSURG, V68, P835, DOI 10.3171/jns.1988.68.6.0835
   JELLINGER KA, 1992, J CANCER RES CLIN, V119, P7, DOI 10.1007/BF01209483
   LEVINSON SA, 1961, CLIN LAB DIAGNOSIS, P916
   LOEFFLER JS, 1985, J CLIN ONCOL, V3, P490, DOI 10.1200/JCO.1985.3.4.490
   Maher EA, 1999, SEMIN ONCOL, V26, P346
   MICHALSKI JM, 1990, RADIOLOGY, V176, P855, DOI 10.1148/radiology.176.3.2389047
   NELSON DF, 1992, INT J RADIAT ONCOL, V23, P9, DOI 10.1016/0360-3016(92)90538-S
   POLLACK IF, 1989, CANCER-AM CANCER SOC, V63, P939, DOI 10.1002/1097-0142(19890301)63:5<939::AID-CNCR2820630526>3.0.CO;2-V
   Reni M, 1997, ANN ONCOL, V8, P227, DOI 10.1023/A:1008201717089
   Shibamoto Y, 1999, J NEURO-ONCOL, V42, P161, DOI 10.1023/A:1006106530795
   SHIPP MA, 1993, NEW ENGL J MED, V329, P987, DOI 10.1056/nejm199309303291402
NR 24
TC 567
Z9 609
U1 0
U2 18
PU AMER SOC CLINICAL ONCOLOGY
PI ALEXANDRIA
PA 330 JOHN CARLYLE ST, STE 300, ALEXANDRIA, VA 22314 USA
SN 0732-183X
J9 J CLIN ONCOL
JI J. Clin. Oncol.
PD JAN 15
PY 2003
VL 21
IS 2
BP 266
EP 272
DI 10.1200/JCO.2003.09.139
PG 7
WC Oncology
WE Conference Proceedings Citation Index - Science (CPCI-S); Science Citation Index Expanded (SCI-EXPANDED)
SC Oncology
GA 685UG
UT WOS:000183281200013
PM 12525518
DA 2024-06-03
ER

PT J
AU DeAngelis, LM
   Seiferheld, W
   Schold, SC
   Fisher, B
   Schultz, CJ
AF DeAngelis, LM
   Seiferheld, W
   Schold, SC
   Fisher, B
   Schultz, CJ
TI Combination chemotherapy and radiotherapy for primary central nervous
   system lymphoma: Radiation Therapy Oncology Group study 93-10
SO JOURNAL OF CLINICAL ONCOLOGY
LA English
DT Article; Proceedings Paper
CT 35th Annual Meeting of the American-Society-of-Clinical-Oncology
CY MAY 13-18, 1999
CL ATLANTA, GA
SP Amer Soc Clin Oncol
ID PRIMARY CNS LYMPHOMA; HIGH-DOSE METHOTREXATE; NON-HODGKINS-LYMPHOMA;
   INTRAOCULAR LYMPHOMA; VINCRISTINE; SURVIVAL; TRIAL; CYCLOPHOSPHAMIDE;
   DOXORUBICIN; INFUSION
AB Purpose: Primary CNS lymphoma (PCNSL) is an aggressive primary brain tumor. Cranial irradiation alone rarely results in long-term disease control or prolonged survival. We prospectively studied the use of combination chemotherapy plus cranial irradiation in newly diagnosed patients with PCNSL.
   Patients and Methods: We enrolled 102 newly diagnosed, immunocompetent patients with PCNSL; 98 were assessable. Patients first received five cycles of methotrexate 2.5 g/m(2), vincristine, procarbazine, and intraventricular methotrexate (12 mg). Whole-brain radiotherapy (RT) was administered to a total dose of 45 Gy and all patients received high-dose cytarabine after RT.
   Results: Fifty-eight percent of patients with measurable disease had a complete response to preirradiation chemotherapy and 36% had a partial (>50%) response, for a 94% response rate. Median progression-free survival was 24.0 months and overall survival was 36.9 months. Age was an important prognostic factor; median survival was 50.4 months in patients younger than 60 and only 21.8 months in those aged 60 or older (P<.001). Fifty-three percent of patients had grade 3 or 4 toxicity during induction chemotherapy, half of which was hematologic. However, 12 patients (15%) experienced severe delayed neurologic toxicity, eight of whom died.
   Conclusion: This is the first multicenter trial demonstrating improved survival with the combination of chemotherapy plus RT compared with previous reports of RT alone. A high-dose methotrexate-based regimen produced a high response rate before RT was administered. High-dose methotrexate combined with cranial irradiation is an effective therapeutic approach to PCNSL, but neurotoxicity is a delayed risk of this approach.
C1 Mem Sloan Kettering Canc Ctr, New York, NY 10021 USA.
   Amer Coll Radiol, Philadelphia, PA USA.
   Univ Pittsburgh, Pittsburgh, PA USA.
   Med Coll Wisconsin, Milwaukee, WI 53226 USA.
   Univ Western Ontario, London, ON, Canada.
C3 Memorial Sloan Kettering Cancer Center; American College of Radiology;
   Pennsylvania Commonwealth System of Higher Education (PCSHE); University
   of Pittsburgh; Medical College of Wisconsin; Western University
   (University of Western Ontario)
RP Mem Sloan Kettering Canc Ctr, 1275 York Ave, New York, NY 10021 USA.
EM
OI DeAngelis, Lisa/0000-0002-6290-7818
FU NCI NIH HHS [U10 CA37422, U10CA21661] Funding Source: Medline; PHS HHS
   [U32115] Funding Source: Medline
CR Abrey LE, 1998, J CLIN ONCOL, V16, P859, DOI 10.1200/JCO.1998.16.3.859
   Abrey LE, 2000, J CLIN ONCOL, V18, P3144, DOI 10.1200/JCO.2000.18.17.3144
   BAUMANN MA, 1986, CANCER-AM CANCER SOC, V57, P1273, DOI 10.1002/1097-0142(19860401)57:7<1273::AID-CNCR2820570702>3.0.CO;2-C
   Central Brain Tumor Registry of the United States, 2000, STAT REP PRIM BRAIN
   DEANGELIS LM, 1992, J CLIN ONCOL, V10, P635, DOI 10.1200/JCO.1992.10.4.635
   DEANGELIS LM, 2001, CANC PRINCIPLES PRAC, P2330
   Ferreri AJM, 2001, ONCOLOGY-BASEL, V60, P134, DOI 10.1159/000055310
   Freilich RJ, 1996, NEUROLOGY, V46, P435, DOI 10.1212/WNL.46.2.435
   FRICK JC, 1986, ARCH INTERN MED, V146, P791, DOI 10.1001/archinte.146.4.791
   GLASS J, 1994, J NEUROSURG, V81, P188, DOI 10.3171/jns.1994.81.2.0188
   Henson JW, 1999, J CLIN ONCOL, V17, P1329
   Herrlinger U, 2002, ANN NEUROL, V51, P247, DOI 10.1002/ana.10102
   Hiraga S, 1999, J NEUROSURG, V91, P221, DOI 10.3171/jns.1999.91.2.0221
   KAPLAN EL, 1958, J AM STAT ASSOC, V53, P457, DOI 10.2307/2281868
   Kellie SJ, 2002, CANCER, V94, P1815, DOI 10.1002/cncr.10397
   Khan RB, 2002, J NEURO-ONCOL, V58, P175, DOI 10.1023/A:1016077907952
   MANTEL NATHAN, 1966, CANCERCHEMOTHERAP REP, V50, P163
   McAllister LD, 2000, NEUROSURGERY, V46, P51, DOI 10.1093/neurosurgery/46.1.51
   Mead GM, 2000, CANCER, V89, P1359, DOI 10.1002/1097-0142(20000915)89:6<1359::AID-CNCR21>3.0.CO;2-9
   NELSON DF, 1992, INT J RADIAT ONCOL, V23, P9, DOI 10.1016/0360-3016(92)90538-S
   O'Brien P, 2000, J CLIN ONCOL, V18, P519, DOI 10.1200/JCO.2000.18.3.519
   Onda K, 1999, Brain Tumor Pathol, V16, P29, DOI 10.1007/BF02478899
   ONEILL BP, 1995, INT J RADIAT ONCOL, V33, P663, DOI 10.1016/0360-3016(95)00207-F
   Sandor V, 1998, J CLIN ONCOL, V16, P3000, DOI 10.1200/JCO.1998.16.9.3000
   Schultz C, 1996, J CLIN ONCOL, V14, P556, DOI 10.1200/JCO.1996.14.2.556
   Tetef ML, 2000, CANCER CHEMOTH PHARM, V46, P19, DOI 10.1007/s002800000118
NR 26
TC 521
Z9 548
U1 1
U2 13
PU AMER SOC CLINICAL ONCOLOGY
PI ALEXANDRIA
PA 2318 MILL ROAD, STE 800, ALEXANDRIA, VA 22314 USA
SN 0732-183X
EI 1527-7755
J9 J CLIN ONCOL
JI J. Clin. Oncol.
PD DEC 15
PY 2002
VL 20
IS 24
BP 4643
EP 4648
DI 10.1200/JCO.2002.11.013
PG 6
WC Oncology
WE Science Citation Index Expanded (SCI-EXPANDED); Conference Proceedings Citation Index - Science (CPCI-S)
SC Oncology
GA 634QK
UT WOS:000180351700007
PM 12488408
DA 2024-06-03
ER

PT J
AU Abrey, LE
   Yahalom, J
   DeAngelis, LM
AF Abrey, LE
   Yahalom, J
   DeAngelis, LM
TI Treatment for primary CNS lymphoma: The next step
SO JOURNAL OF CLINICAL ONCOLOGY
LA English
DT Article
ID NERVOUS-SYSTEM LYMPHOMA; RADIATION-THERAPY; INTRAOCULAR LYMPHOMA;
   CHEMOTHERAPY; SURVIVAL; CYCLOPHOSPHAMIDE; METHOTREXATE; DOXORUBICIN;
   VINCRISTINE; BRAIN
AB Purpose: The use of preradiotherapy (WT) methotrexate (MTX) has improved disease control and survival in patients with primary CNS lymphoma (PCNSL). The reported protocol was designed to optimize and enhance the chemotherapeutic component of treatment.
   Patients and Methods: fifty-two patients were treated with five cycles of high-dose MTX 3.5 g/m(2), procarbazine 100 mg/m(2)/d, and vincristine 1.4 mg/m(2). Thirty patients received whole-brain RT (45 Gy). Twenty-two older patients deferred RT to diminish the risk of delayed neurotoxicity; these patients are compared with 12 older patients who completed the entire treatment regimen. Most patients (n = 35) received high-dose cytarabine after RT.
   Results: Objective response rate to the induction chemotherapy regimen was 90%; overall median survival is 60 months. Grade 3 or 4 myelosuppression was seen in 30 patients, primarily in association with cytarabine; grade 3 nephrotoxicity due to MTX was seen in two patients. Older patients had similar median survival with or without the addition of RT: 32 versus 33 months, respectively. However, late neurotoxicity was significantly more common in those older patients who received RT (P = .00004). Patients younger than 60 years who received the complete treatment regimen have not reached median disease-free or overall survival.
   Conclusion: increasing the dose of MTX and adding procarbazine and vincristine improved disease control and overall survival in patients with newly diagnosed PCNSL. Younger patients in particular fared extremely well with this treatment regimen. In older patients, deferring whole-brain RT did not compromise overall survival but did reduce treatment-related toxicity. (C) 2000 by American Society of Clinical Oncology.
C1 Mem Sloan Kettering Canc Ctr, Dept Neurol, New York, NY 10021 USA.
   Mem Sloan Kettering Canc Ctr, Dept Radiat Oncol, New York, NY 10021 USA.
C3 Memorial Sloan Kettering Cancer Center; Memorial Sloan Kettering Cancer
   Center
RP Abrey, LE (corresponding author), Mem Sloan Kettering Canc Ctr, Dept Neurol, 1275 York Ave, New York, NY 10021 USA.
OI DeAngelis, Lisa/0000-0002-6290-7818
CR Abrey Lauren E., 1997, Neurology, V48, pA18
   Abrey LE, 1998, J CLIN ONCOL, V16, P859, DOI 10.1200/JCO.1998.16.3.859
   Dahlborg S A, 1996, Cancer J Sci Am, V2, P166
   DEANGELIS LM, 1992, J CLIN ONCOL, V10, P635, DOI 10.1200/JCO.1992.10.4.635
   DEANGELIS LM, 1999, P AM SOC CLIN ONCOL, V18, pA140
   Fishburne BC, 1997, ARCH OPHTHALMOL-CHIC, V115, P1152, DOI 10.1001/archopht.1997.01100160322009
   Freilich RJ, 1996, NEUROLOGY, V46, P435, DOI 10.1212/WNL.46.2.435
   GLASS J, 1994, J NEUROSURG, V81, P188, DOI 10.3171/jns.1994.81.2.0188
   Hoang-Xuan Khe, 1997, Neurology, V48, pA17
   KAPLAN EL, 1958, J AM STAT ASSOC, V53, P457, DOI 10.2307/2281868
   LACHANCE DH, 1994, NEUROLOGY, V44, P1721, DOI 10.1212/WNL.44.9.1721
   NELSON DF, 1992, INT J RADIAT ONCOL, V23, P9, DOI 10.1016/0360-3016(92)90538-S
   NEUWELT EA, 1991, J CLIN ONCOL, V9, P1580, DOI 10.1200/JCO.1991.9.9.1580
   PETERSON K, 1993, CANCER, V72, P843, DOI 10.1002/1097-0142(19930801)72:3<843::AID-CNCR2820720333>3.0.CO;2-#
   Sandor V, 1998, J CLIN ONCOL, V16, P3000, DOI 10.1200/JCO.1998.16.9.3000
   Schultz C, 1996, J CLIN ONCOL, V14, P556, DOI 10.1200/JCO.1996.14.2.556
NR 16
TC 458
Z9 474
U1 0
U2 9
PU LIPPINCOTT WILLIAMS & WILKINS
PI PHILADELPHIA
PA 530 WALNUT ST, PHILADELPHIA, PA 19106-3621 USA
SN 0732-183X
J9 J CLIN ONCOL
JI J. Clin. Oncol.
PD SEP
PY 2000
VL 18
IS 17
BP 3144
EP 3150
DI 10.1200/JCO.2000.18.17.3144
PG 7
WC Oncology
WE Science Citation Index Expanded (SCI-EXPANDED)
SC Oncology
GA 349GX
UT WOS:000089038300011
PM 10963643
DA 2024-06-03
ER

PT J
AU Abrey, LE
   DeAngelis, LM
   Yahalom, J
AF Abrey, LE
   DeAngelis, LM
   Yahalom, J
TI Long-term survival in primary CNS lymphoma
SO JOURNAL OF CLINICAL ONCOLOGY
LA English
DT Article
ID NERVOUS-SYSTEM LYMPHOMA; RADIATION-THERAPY; CRANIAL IRRADIATION;
   METHOTREXATE; CHEMOTHERAPY; CYCLOPHOSPHAMIDE; ENCEPHALOPATHY;
   DOXORUBICIN; VINCRISTINE; SEQUELAE
AB Purpose: We have previously reported on 31 patients with primary CNS lymphoma (PCNSL) treated between 1986 and 1992 with methotrexate (MTX), cranial radiotherapy (RT), and high-dose cytarabine who remained free of disease longer than historical controls.
   Patients and Methods: We performed a follow-up analysis of our original cohort and now report their long-term survival and late treatment-related toxicity.
   Results: The median cause-specific survival was 42 months, with a five-year survival of 22.3% compared with 3% to 4% in historical controls treated with RT alone. Age less than 50 years at diagnosis was a significant prognostic factor for survival (P = .01). Median disease-free survival was 40.3 months; 15 patients relapsed, all but one in the CNS. Late treatment-related toxicity was observed in nearly one third of patients and those more than 60 years of age were at substantially higher risk (P < .0001).
   Conclusion: Combined modality therapy for PCNSL has improved survival, but relapse is common and late neurologic toxicity is a significant complication. Although this approach is highly effective for younger patients, efficacious but less neurotoxic regimens need to be developed for older patients. (C) 1998 by American Society of Clinical Oncology.
C1 Mem Sloan Kettering Canc Ctr, Dept Neurol, New York, NY 10021 USA.
   Mem Sloan Kettering Canc Ctr, Dept Radiat Oncol, New York, NY 10021 USA.
C3 Memorial Sloan Kettering Cancer Center; Memorial Sloan Kettering Cancer
   Center
RP DeAngelis, LM (corresponding author), Mem Sloan Kettering Canc Ctr, Dept Neurol, 1275 York Ave, New York, NY 10021 USA.
OI DeAngelis, Lisa/0000-0002-6290-7818
CR BLEYER WA, 1981, CANCER TREAT REP, V65, P89
   CROSSEN JR, 1994, J CLIN ONCOL, V12, P627, DOI 10.1200/JCO.1994.12.3.627
   Dahlborg S A, 1996, Cancer J Sci Am, V2, P166
   DEANGELIS LM, 1989, NEUROLOGY, V39, P789, DOI 10.1212/WNL.39.6.789
   DEANGELIS LM, 1992, J CLIN ONCOL, V10, P635, DOI 10.1200/JCO.1992.10.4.635
   Duffey P, 1996, ARCH NEUROL-CHICAGO, V53, P814, DOI 10.1001/archneur.1996.00550080140022
   FINE HA, 1993, ANN INTERN MED, V119, P1093, DOI 10.7326/0003-4819-119-11-199312010-00007
   Freilich RJ, 1996, NEUROLOGY, V46, P435, DOI 10.1212/WNL.46.2.435
   GABBAI AA, 1989, J NEUROSURG, V70, P190, DOI 10.3171/jns.1989.70.2.0190
   GLASS J, 1994, J NEUROSURG, V81, P188, DOI 10.3171/jns.1994.81.2.0188
   LACHANCE DH, 1994, NEUROLOGY, V44, P1721, DOI 10.1212/WNL.44.9.1721
   MCGUIRT WF, 1992, ANN OTO RHINOL LARYN, V101, P222, DOI 10.1177/000348949210100305
   NELSON DF, 1992, INT J RADIAT ONCOL, V23, P9, DOI 10.1016/0360-3016(92)90538-S
   Schultz C, 1996, J CLIN ONCOL, V14, P556, DOI 10.1200/JCO.1996.14.2.556
   Thiessen Brian, 1997, Neurology, V48, pA29
NR 15
TC 429
Z9 451
U1 0
U2 8
PU W B SAUNDERS CO
PI PHILADELPHIA
PA INDEPENDENCE SQUARE WEST CURTIS CENTER, STE 300, PHILADELPHIA, PA
   19106-3399 USA
SN 0732-183X
J9 J CLIN ONCOL
JI J. Clin. Oncol.
PD MAR
PY 1998
VL 16
IS 3
BP 859
EP 863
DI 10.1200/JCO.1998.16.3.859
PG 5
WC Oncology
WE Science Citation Index Expanded (SCI-EXPANDED)
SC Oncology
GA YZ486
UT WOS:000072258900006
PM 9508166
DA 2024-06-03
ER

PT J
AU Thiel, E
   Korfel, A
   Martus, P
   Kanz, L
   Griesinger, F
   Rauch, M
   Röth, A
   Hertenstein, B
   von Toll, T
   Hundsberger, T
   Mergenthaler, HG
   Leithäuser, M
   Birnbaum, T
   Fischer, L
   Jahnke, K
   Herrlinger, U
   Plasswilm, L
   Nägele, T
   Pietsch, T
   Bamberg, M
   Weller, M
AF Thiel, Eckhard
   Korfel, Agnieszka
   Martus, Peter
   Kanz, Lothar
   Griesinger, Frank
   Rauch, Michael
   Roeth, Alexander
   Hertenstein, Bernd
   von Toll, Theda
   Hundsberger, Thomas
   Mergenthaler, Hans-Guenther
   Leithaeuser, Malte
   Birnbaum, Tobias
   Fischer, Lars
   Jahnke, Kristoph
   Herrlinger, Ulrich
   Plasswilm, Ludwig
   Naegele, Thomas
   Pietsch, Torsten
   Bamberg, Michael
   Weller, Michael
TI High-dose methotrexate with or without whole brain radiotherapy for
   primary CNS lymphoma (G-PCNSL-SG-1): a phase 3, randomised,
   non-inferiority trial
SO LANCET ONCOLOGY
LA English
DT Article
ID NERVOUS-SYSTEM LYMPHOMA; COMBINED-MODALITY THERAPY; CONSOLIDATION
   RADIOTHERAPY; CELL TRANSPLANTATION; 1ST-LINE TREATMENT; INITIAL
   TREATMENT; CHEMOTHERAPY; MULTICENTER; SURVIVAL
AB Background High-dose methotrexate is the standard of care for patients with newly diagnosed primary CNS lymphoma. The role of whole brain radiotherapy is controversial because delayed neurotoxicity limits its acceptance as a standard of care. We aimed to investigate whether first-line chemotherapy based on high-dose methotrexate was non-inferior to the same chemotherapy regimen followed by whole brain radiotherapy for overall survival.
   Methods Immunocompetent patients with newly diagnosed primary CNS lymphoma were enrolled from 75 centres and treated between May, 2000, and May, 2009. Patients were allocated by computer-generated block randomisation to receive first-line chemotherapy based on high-dose methotrexate with or without subsequent whole brain radiotherapy, with stratification by age (<60 vs >= 60 years) and institution (Berlin vs Tubingen vs all other sites). The biostatistics centre assigned patients to treatment groups and informed local centres by fax; physicians and patients were not masked to treatment group after assignment. Patients enrolled between May, 2000, and August, 2006, received high-dose methotrexate (4 g/m(2)) on day 1 of six 14-day cycles; thereafter, patients received high-dose methotrexate plus ifosfamide (1.5 g/m(2)) on days 3-5 of six 14-day cycles. In those assigned to receive first-line chemotherapy followed by radiotherapy, whole brain radiotherapy was given to a total dose of 45 Gy, in 30 fractions of 1.5 Gy given daily on weekdays. Patients allocated to first-line chemotherapy without whole brain radiotherapy who had not achieved complete response were given high-dose cytarabine. The primary endpoint was overall survival, and analysis was per protocol. Our hypothesis was that the omission of whole brain radiotherapy does not compromise overall survival, with a non-inferiority margin of 0.9. This trial is registered with ClinicalTrials.gov, number NCT00153530.
   Findings 551 patients (median age 63 years, IQR 55-69) were enrolled and randomised, of whom 318 were treated per protocol. In the per-protocol population, median overall survival was 32.4 months (95% CI 25.8-39.0) in patients receiving whole brain radiotherapy (n=154), and 37.1 months (27.5-46.7) in those not receiving whole brain radiotherapy (n=164), hazard ratio 1.06 (95% CI 0.80-1.40; p=0.71). Thus our primary hypothesis was not proven. Median progression-free survival was 18.3 months (95% CI 11.6-25.0) in patients receiving whole brain radiotherapy, and 11.9 months (7.3-16.5; p=0.14) in those not receiving whole brain radiotherapy. Treatment-related neurotoxicity in patients with sustained complete response was more common in patients receiving whole brain radiotherapy (22/45, 49% by clinical assessment; 35/49, 71% by neuroradiology) than in those who did not (9/34, 26%; 16/35, 46%).
   Interpretation No significant difference in overall survival was recorded when whole brain radiotherapy was omitted from first-line chemotherapy in patients with newly diagnosed primary CNS lymphoma, but our primary hypothesis was not proven. The progression-free survival benefit afforded by whole brain radiotherapy has to be weighed against the increased risk of neurotoxicity in long-term survivors.
C1 [Weller, Michael] Univ Zurich Hosp, Dept Neurol, CH-8091 Zurich, Switzerland.
   [Thiel, Eckhard; Korfel, Agnieszka; Fischer, Lars; Jahnke, Kristoph] Charite, Dept Hematol & Oncol, Berlin, Germany.
   [Martus, Peter] Charite, Dept Clin Epidemiol & Biostat, Berlin, Germany.
   [Kanz, Lothar] Univ Tubingen Hosp, Dept Hematol & Oncol, Tubingen, Germany.
   [Herrlinger, Ulrich; Weller, Michael] Univ Tubingen Hosp, Dept Neurol, Tubingen, Germany.
   [Plasswilm, Ludwig; Bamberg, Michael] Univ Tubingen Hosp, Dept Radiat Oncol, Tubingen, Germany.
   [Naegele, Thomas] Univ Tubingen Hosp, Dept Neuroradiol, Tubingen, Germany.
   [Griesinger, Frank] Pius Hosp Oldenburg, Dept Radiotherapy & Med Oncol, Oldenburg, Germany.
   [Rauch, Michael] Protestant Hosp Bielefeld, Dept Hematol & Oncol, Bielefeld, Germany.
   [Roeth, Alexander] Univ Hosp Essen, Dept Hematol, Essen, Germany.
   [Hertenstein, Bernd] Hosp Bremen Ctr, Dept Internal Med, Bremen, Germany.
   [von Toll, Theda] Univ Hosp Magdeburg, Dept Hematol & Oncol, Magdeburg, Germany.
   [Hundsberger, Thomas] Univ Hosp Mainz, Dept Neurol, Mainz, Germany.
   [Mergenthaler, Hans-Guenther] Klinikum Stuttgart, Dept Hematol & Oncol, Stuttgart, Germany.
   [Leithaeuser, Malte] Univ Hosp Rostock, Dept Hematol & Oncol, Rostock, Germany.
   [Birnbaum, Tobias] Univ Hosp Munich LMU, Dept Neurol, Munich, Germany.
   [Pietsch, Torsten] Univ Hosp Bonn, Inst Neuropathol, Bonn, Germany.
C3 University of Zurich; University Zurich Hospital; Free University of
   Berlin; Humboldt University of Berlin; Charite Universitatsmedizin
   Berlin; Free University of Berlin; Humboldt University of Berlin;
   Charite Universitatsmedizin Berlin; Eberhard Karls University of
   Tubingen; Eberhard Karls University Hospital; Eberhard Karls University
   of Tubingen; Eberhard Karls University Hospital; Eberhard Karls
   University of Tubingen; Eberhard Karls University Hospital; Eberhard
   Karls University of Tubingen; Eberhard Karls University Hospital;
   Pius-Hospital Oldenburg; University of Duisburg Essen; University
   Hospital Magdeburg; University Hospital Mainz; Klinikum Stuttgart;
   University of Rostock; University of Munich; University of Bonn
RP Weller, M (corresponding author), Univ Zurich Hosp, Dept Neurol, Frauenklin Str 26, CH-8091 Zurich, Switzerland.
EM
RI Hundsberger, Thomas/AAL-6712-2020; McDermott, Michael
   Francis/GZM-5330-2022
OI Hundsberger, Thomas/0000-0002-4419-2767; Weller,
   Michael/0000-0002-1748-174X
FU German Cancer Aid [70-2838-Th 2]
FX The trial received the certificate of quality (Gutesiegel A) of the
   German Cancer Society and was supported by the German Cancer Aid grant
   70-2838-Th 2 to ET and MW. All institutions and investigators
   participating in the trial are listed in webappendix p 6. We thank the
   Competence Network on Malignant Lymphoma (Kompetenznetz Maligne
   Lymphome) for logistical support and the International Primary CNS
   Lymphoma Study Group for fruitful discussions of interim results during
   the study.
CR Abrey LE, 2006, J CLIN ONCOL, V24, P5711, DOI 10.1200/JCO.2006.08.2941
   Abrey LE, 1998, J CLIN ONCOL, V16, P859, DOI 10.1200/JCO.1998.16.3.859
   Abrey LE, 2000, J CLIN ONCOL, V18, P3144, DOI 10.1200/JCO.2000.18.17.3144
   Angelov L, 2009, J CLIN ONCOL, V27, P3503, DOI 10.1200/JCO.2008.19.3789
   Batchelor T, 2003, J CLIN ONCOL, V21, P1044, DOI 10.1200/JCO.2003.03.036
   Bessell EM, 2002, J CLIN ONCOL, V20, P231, DOI 10.1200/JCO.20.1.231
   Bessell EM, 2007, EUR J CANCER, V43, P1141, DOI 10.1016/j.ejca.2006.12.011
   DEANGELIS LM, 1992, J CLIN ONCOL, V10, P635, DOI 10.1200/JCO.1992.10.4.635
   DeAngelis LM, 2002, J CLIN ONCOL, V20, P4643, DOI 10.1200/JCO.2002.11.013
   Ekenel Meltem, 2007, Curr Treat Options Neurol, V9, P271, DOI 10.1007/s11940-007-0013-6
   Ferreri AJM, 2009, LANCET, V374, P1512, DOI 10.1016/S0140-6736(09)61416-1
   Fischer L, 2008, NEUROLOGY, V71, P1102, DOI 10.1212/01.wnl.0000326958.52546.f5
   Fischer L, 2006, HAEMATOLOGICA, V91, P429
   Gavrilovic IT, 2006, J CLIN ONCOL, V24, P4570, DOI 10.1200/JCO.2006.06.6910
   Gerstner E, 2007, EXPERT REV ANTICANC, V7, P689, DOI 10.1586/14737140.7.5.689
   Guha-Thakurta N, 1999, J NEURO-ONCOL, V43, P259, DOI 10.1023/A:1006210703827
   Herrlinger U, 2005, ANN NEUROL, V57, P843, DOI 10.1002/ana.20495
   Herrlinger U, 2002, ANN NEUROL, V51, P247, DOI 10.1002/ana.10102
   Hiraga S, 1999, J NEUROSURG, V91, P221, DOI 10.3171/jns.1999.91.2.0221
   Hoang-Xuan K, 2003, J CLIN ONCOL, V21, P2726, DOI 10.1200/JCO.2003.11.036
   Hottinger AF, 2007, NEUROLOGY, V69, P1178, DOI 10.1212/01.wnl.0000276986.19602.c1
   Illerhaus G, 2007, HAEMATOLOGICA, V93, P147, DOI 10.3324/haematol.11771
   Illerhaus G, 2006, J CLIN ONCOL, V24, P3865, DOI 10.1200/JCO.2006.06.2117
   Jahnke K, 2005, ANN ONCOL, V16, P445, DOI 10.1093/annonc/mdi075
   Jahnke K, 2006, J NEURO-ONCOL, V80, P159, DOI 10.1007/s11060-006-9165-6
   Jahnke K, 2009, EXPERT REV NEUROTHER, V9, P1497, DOI 10.1586/ERN.09.100
   Juergens A, 2010, ANN NEUROL, V67, P182, DOI 10.1002/ana.21824
   Kadan-Lottick NS, 2002, CANCER, V95, P193, DOI 10.1002/cncr.10643
   Korfel A, 2005, BRIT J HAEMATOL, V128, P177, DOI 10.1111/j.1365-2141.2004.05284.x
   Kurzwelly D, 2008, J NEURO-ONCOL, V90, P53, DOI 10.1007/s11060-008-9630-5
   Mead GM, 2000, CANCER, V89, P1359, DOI 10.1002/1097-0142(20000915)89:6<1359::AID-CNCR21>3.0.CO;2-9
   Morris PG, 2009, LANCET NEUROL, V8, P581, DOI 10.1016/S1474-4422(09)70091-2
   NELSON DF, 1992, INT J RADIAT ONCOL, V23, P9, DOI 10.1016/0360-3016(92)90538-S
   O'Brien PC, 2006, INT J RADIAT ONCOL, V64, P408, DOI 10.1016/j.ijrobp.2005.07.958
   Pels H, 2003, J CLIN ONCOL, V21, P4489, DOI 10.1200/JCO.2003.04.056
   Reni M, 2004, J CLIN ONCOL, V22, P1165, DOI 10.1200/JCO.2004.99.259
   Reni M, 2001, INT J RADIAT ONCOL, V51, P419, DOI 10.1016/S0360-3016(01)01639-X
   Shah GD, 2007, J CLIN ONCOL, V25, P4730, DOI 10.1200/JCO.2007.12.5062
NR 38
TC 424
Z9 450
U1 2
U2 35
PU ELSEVIER SCIENCE INC
PI NEW YORK
PA STE 800, 230 PARK AVE, NEW YORK, NY 10169 USA
SN 1470-2045
EI 1474-5488
J9 LANCET ONCOL
JI Lancet Oncol.
PD NOV
PY 2010
VL 11
IS 11
BP 1036
EP 1047
DI 10.1016/S1470-2045(10)70229-1
PG 12
WC Oncology
WE Science Citation Index Expanded (SCI-EXPANDED)
SC Oncology
GA 683AP
UT WOS:000284444400017
PM 20970380
DA 2024-06-03
ER

PT J
AU Abrey, LE
   Ben-Porat, L
   Panageas, KS
   Yahalom, J
   Berkey, B
   Curran, W
   Schultz, C
   Leibel, S
   Nelson, D
   Mehta, M
   DeAngelis, LM
AF Abrey, Lauren E.
   Ben-Porat, Leah
   Panageas, Katherine S.
   Yahalom, Joachim
   Berkey, Brian
   Curran, Walter
   Schultz, Christopher
   Leibel, Steven
   Nelson, Diana
   Mehta, Minesh
   DeAngelis, Lisa M.
TI Primary central nervous system lymphoma: The Memorial Sloan-Kettering
   Cancer Center prognostic model
SO JOURNAL OF CLINICAL ONCOLOGY
LA English
DT Article
ID PRIMARY CNS LYMPHOMA; NON-HODGKINS-LYMPHOMA; ONCOLOGY-GROUP RTOG;
   PHASE-II; CHEMOTHERAPY; RADIOTHERAPY; SURVIVAL; VALIDATION; CHOD/BVAM;
   BRAIN
AB Purpose The purpose of this study was to analyze prognostic factors for patients with newly diagnosed primary CNS lymphoma (PCNSL) in order to establish a predictive model that could be applied to the care of patients and the design of prospective clinical trials.
   Patients and Methods Three hundred thirty-eight consecutive patients with newly diagnosed PCNSL seen at Memorial Sloan-Kettering Cancer Center (MSKCC; New York, NY) between 1983 and 2003 were analyzed. Standard univariate and multivariate analyses were performed. In addition, a formal cut point analysis was used to determine the most statistically significant cut point for age. Recursive partitioning analysis (RPA) was used to create independent prognostic classes. An external validation set obtained from three prospective Radiation Therapy Oncology Group (RTOG) PCNSL clinical trials was used to test the RPA classification.
   Results Age and performance status were the only variables identified on standard multivariate analysis. Cut point analysis of age determined that patients age <= 50 years had significantly improved outcome compared with older patients. RPA of 282 patients identified three distinct prognostic classes: class 1 (patients <50 years), class 2 (patients >= 50; Karnofsky performance score [KPS] >= 70) and class 3 (patients >= 50; KPS <70). These three classes significantly distinguished outcome with regard to both overall and failure-free survival. Analysis of the RTOG data set confirmed the validity of this classification.
   Conclusion The MSKCC prognostic score is a simple, statistically powerful model with universal applicability to patients with newly diagnosed PCNSL. We recommend that it be adopted for the management of newly diagnosed patients and incorporated into the design of prospective clinical trials.
C1 Mem Sloan Kettering Canc Ctr, Dept Neurol, New York, NY 10021 USA.
   Mem Sloan Kettering Canc Ctr, Dept Epidemiol & Biostat, New York, NY 10021 USA.
   Mem Sloan Kettering Canc Ctr, Dept Radiat Oncol, New York, NY 10021 USA.
   Radiat Therapy Oncol Grp, Philadelphia, PA USA.
   Thomas Jefferson Univ, Dept Radiat Oncol, Philadelphia, PA 19107 USA.
   Med Coll Wisconsin, Dept Radiat Oncol, Milwaukee, WI 53226 USA.
   Stanford Comprehens Canc Ctr, Stanford, CA USA.
   Mayo Clin, Div Radiat Oncol, Rochester, MN USA.
C3 Memorial Sloan Kettering Cancer Center; Memorial Sloan Kettering Cancer
   Center; Memorial Sloan Kettering Cancer Center; Radiation Therapy
   Oncology Group (RTOG); Jefferson University; Medical College of
   Wisconsin; Stanford University; Mayo Clinic
RP Abrey, LE (corresponding author), Mem Sloan Kettering Canc Ctr, Dept Neurol, 1275 York Ave, New York, NY 10021 USA.
EM
RI Mehta, Minesh P./AAB-8915-2019
OI Mehta, Minesh P./0000-0002-4812-5713; Price, Leah/0000-0002-5092-4768;
   DeAngelis, Lisa/0000-0002-6290-7818
CR Abrey LE, 2000, J CLIN ONCOL, V18, P3144, DOI 10.1200/JCO.2000.18.17.3144
   Bessell EM, 2004, INT J RADIAT ONCOL, V59, P501, DOI 10.1016/j.ijrobp.2003.11.001
   Bessell EM, 2002, J CLIN ONCOL, V20, P231, DOI 10.1200/JCO.20.1.231
   Breiman L., 1984, Trees, V37, P237, DOI DOI 10.1201/9781315139470
   Corry J, 1998, INT J RADIAT ONCOL, V41, P615, DOI 10.1016/S0360-3016(97)00571-3
   DEANGELIS LM, 1990, NEUROLOGY, V40, P80, DOI 10.1212/WNL.40.1.80
   DeAngelis LM, 2002, J CLIN ONCOL, V20, P4643, DOI 10.1200/JCO.2002.11.013
   Ferreri AJM, 2003, J CLIN ONCOL, V21, P266, DOI 10.1200/JCO.2003.09.139
   Gaspar LE, 2000, INT J RADIAT ONCOL, V47, P1001, DOI 10.1016/S0360-3016(00)00547-2
   Mazumdar M, 2000, STAT MED, V19, P113, DOI 10.1002/(SICI)1097-0258(20000115)19:1<113::AID-SIM245>3.3.CO;2-F
   NELSON DF, 1992, INT J RADIAT ONCOL, V23, P9, DOI 10.1016/0360-3016(92)90538-S
   O'Brien P, 2000, J CLIN ONCOL, V18, P519, DOI 10.1200/JCO.2000.18.3.519
   Panageas KS, 2005, CANCER-AM CANCER SOC, V104, P2466, DOI 10.1002/cncr.21481
   Pels H, 2003, J CLIN ONCOL, V21, P4489, DOI 10.1200/JCO.2003.04.056
   Schultz C, 1996, J CLIN ONCOL, V14, P556, DOI 10.1200/JCO.1996.14.2.556
   Scott CB, 1998, INT J RADIAT ONCOL, V40, P51, DOI 10.1016/S0360-3016(97)00485-9
   Shenkier TN, 2005, CANCER, V103, P1008, DOI 10.1002/cncr.20868
NR 17
TC 411
Z9 429
U1 0
U2 7
PU AMER SOC CLINICAL ONCOLOGY
PI ALEXANDRIA
PA 330 JOHN CARLYLE ST, STE 300, ALEXANDRIA, VA 22314 USA
SN 0732-183X
J9 J CLIN ONCOL
JI J. Clin. Oncol.
PD DEC 20
PY 2006
VL 24
IS 36
BP 5711
EP 5715
DI 10.1200/JCO.2006.08.2941
PG 5
WC Oncology
WE Science Citation Index Expanded (SCI-EXPANDED)
SC Oncology
GA 119EG
UT WOS:000242994400015
PM 17116938
OA Bronze
DA 2024-06-03
ER

PT J
AU Chapuy, B
   Roemer, MGM
   Stewart, C
   Tan, YX
   Abo, RP
   Zhang, LY
   Dunford, AJ
   Meredith, DM
   Thorner, AR
   Jordanova, ES
   Liu, G
   Feuerhake, F
   Ducar, MD
   Illerhaus, G
   Gusenleitner, D
   Linden, EA
   Sun, HH
   Homer, H
   Aono, M
   Pinkus, GS
   Ligon, AH
   Ligon, KL
   Ferry, JA
   Freeman, GJ
   van Hummelen, P
   Golub, TR
   Getz, G
   Rodig, SJ
   de Jong, D
   Monti, S
   Shipp, MA
AF Chapuy, Bjoern
   Roemer, Margaretha G. M.
   Stewart, Chip
   Tan, Yuxiang
   Abo, Ryan P.
   Zhang, Liye
   Dunford, Andrew J.
   Meredith, David M.
   Thorner, Aaron R.
   Jordanova, Ekaterina S.
   Liu, Gang
   Feuerhake, Friedrich
   Ducar, Matthew D.
   Illerhaus, Gerald
   Gusenleitner, Daniel
   Linden, Erica A.
   Sun, Heather H.
   Homer, Heather
   Aono, Miyuki
   Pinkus, Geraldine S.
   Ligon, Azra H.
   Ligon, Keith L.
   Ferry, Judith A.
   Freeman, Gordon J.
   van Hummelen, Paul
   Golub, Todd R.
   Getz, Gad
   Rodig, Scott J.
   de Jong, Daphne
   Monti, Stefano
   Shipp, Margaret A.
TI Targetable genetic features of primary testicular and primary central
   nervous system lymphomas
SO BLOOD
LA English
DT Article
ID B-CELL LYMPHOMA; INTEGRATIVE ANALYSIS REVEALS; HIGH PREVALENCE; CANCER;
   MUTATIONS; EXPRESSION; RECURRENT; GENOME; PROLIFERATION; MYD88
AB Primary central nervous system lymphomas (PCNSLs) and primary testicular lymphomas (PTLs) are extranodal large B-cell lymphomas (LBCLs) with inferior responses to current empiric treatment regimens. To identify targetable genetic features of PCNSL and PTL, we characterized their recurrent somatic mutations, chromosomal rearrangements, copy number alterations (CNAs), and associated driver genes, and compared these comprehensive genetic signatures to those of diffuse LBCL and primary mediastinal large B-cell lymphoma (PMBL). These studies identify unique combinations of genetic alterations in discrete LBCL subtypes and subtype-selective bases for targeted therapy. PCNSLs and PTLs frequently exhibit genomic instability, and near-uniform, often biallelic, CDKN2A loss with rare TP53 mutations. PCNSLs and PTLs also use multiple genetic mechanisms to target key genes and pathways and exhibit near-uniform oncogenic Toll-like receptor signaling as a result of MYD88 mutation and/or NFKBIZ amplification, frequent concurrent B-cell receptor pathway activation, and deregulation of BCL6. Of great interest, PCNSLs and PTLs also have frequent 9p24.1/PD-L1/PD-L2 CNAs and additional translocations of these loci, structural bases of immune evasion that are shared with PMBL.
C1 [Chapuy, Bjoern; Roemer, Margaretha G. M.; Homer, Heather; Aono, Miyuki; Freeman, Gordon J.; Shipp, Margaret A.] Dana Farber Canc Inst, Dept Med Oncol, Boston, MA 02115 USA.
   [Roemer, Margaretha G. M.; de Jong, Daphne] Vrije Univ Amsterdam, Med Ctr, Dept Pathol, Amsterdam, Netherlands.
   [Stewart, Chip; Dunford, Andrew J.; Golub, Todd R.; Getz, Gad] Broad Inst, Cambridge, MA USA.
   [Tan, Yuxiang; Zhang, Liye; Liu, Gang; Gusenleitner, Daniel; Monti, Stefano] Boston Univ, Sch Med, Sect Computat Biomed, Boston, MA 02118 USA.
   [Abo, Ryan P.; Thorner, Aaron R.; Ducar, Matthew D.; van Hummelen, Paul] Dana Farber Canc Inst, Ctr Canc Genome Discovery, Boston, MA 02115 USA.
   [Meredith, David M.; Sun, Heather H.; Pinkus, Geraldine S.; Ligon, Azra H.; Ligon, Keith L.; Rodig, Scott J.] Brigham & Womens Hosp, Dept Pathol, 75 Francis St, Boston, MA 02115 USA.
   [Jordanova, Ekaterina S.] Vrije Univ Amsterdam, Med Ctr, Ctr Gynaecol Oncol, Amsterdam, Netherlands.
   [Feuerhake, Friedrich] Hannover Med Sch, Dept Pathol, Hannover, Germany.
   [Illerhaus, Gerald] Univ Hosp Freiburg, Dept Hematol & Oncol, Freiburg, Germany.
   [Linden, Erica A.] Massachusetts Gen Hosp, North Shore Canc Ctr, Danvers, MA USA.
   [Ferry, Judith A.; Getz, Gad] Massachusetts Gen Hosp, Dept Pathol, Boston, MA 02114 USA.
   [Golub, Todd R.] Dana Farber Canc Inst, Dept Pediat Oncol, Boston, MA 02115 USA.
C3 Harvard University; Dana-Farber Cancer Institute; Vrije Universiteit
   Amsterdam; Harvard University; Massachusetts Institute of Technology
   (MIT); Broad Institute; Boston University; Harvard University;
   Dana-Farber Cancer Institute; Harvard University; Brigham & Women's
   Hospital; Vrije Universiteit Amsterdam; Hannover Medical School;
   University of Freiburg; Harvard University; Massachusetts General
   Hospital; Harvard University; Massachusetts General Hospital; Harvard
   University; Dana-Farber Cancer Institute
RP Shipp, MA (corresponding author), Harvard Univ, Sch Med, Dana Farber Canc Inst, 450 Brookline Ave, Boston, MA 02115 USA.
EM
RI Freeman, Gordon/AAC-5380-2019; Tan, Yuxiang/AFQ-4637-2022; Rodig,
   Scott/Y-3889-2019; Stewart, Chip/IUO-0055-2023; Liu,
   Geoffrey/N-4421-2016
OI Freeman, Gordon/0000-0002-7210-5616; Tan, Yuxiang/0000-0002-1704-1293;
   van hummelen, paul/0000-0002-4801-8600; Jordanova, Ekaterina
   S./0000-0002-8121-1322; Meredith, David/0000-0002-7054-8624; Liu,
   Gang/0000-0001-9270-6796; Monti, Stefano/0000-0002-9376-0660
FU National Institutes of Health, National Cancer Institute [R01 CA161026];
   LLS Translational Research Award; Claudia Adams Barr Program in Basic
   Cancer Research; National Institutes of Health, National Institute of
   Allergy and Infectious Diseases [PO1 AI056299]; Slim Initiative for
   Genomic Medicine; Carlos Slim Foundation in Mexico
FX This work was supported by National Institutes of Health, National
   Cancer Institute grant R01 CA161026 and an LLS Translational Research
   Award (M.A.S.), a Claudia Adams Barr Program in Basic Cancer Research
   (B.C.), National Institutes of Health, National Institute of Allergy and
   Infectious Diseases grant PO1 AI056299 (G.J.F.), and the Slim Initiative
   for Genomic Medicine, a project funded by the Carlos Slim Foundation in
   Mexico (Broad).
CR Abo RP, 2015, NUCLEIC ACIDS RES, V43, DOI 10.1093/nar/gku1211
   Ansell SM, 2015, NEW ENGL J MED, V372, P311, DOI 10.1056/NEJMoa1411087
   Basso K, 2015, NAT REV IMMUNOL, V15, P172, DOI 10.1038/nri3814
   Beroukhim R, 2007, P NATL ACAD SCI USA, V104, P20007, DOI 10.1073/pnas.0710052104
   Booman M, 2008, J PATHOL, V216, P209, DOI 10.1002/path.2399
   Boyd RS, 2003, LEUKEMIA, V17, P1605, DOI 10.1038/sj.leu.2402993
   Braggio E, 2015, CLIN CANCER RES, V21, P3986, DOI 10.1158/1078-0432.CCR-14-2116
   Braggio E, 2011, CLIN CANCER RES, V17, P4245, DOI 10.1158/1078-0432.CCR-11-0395
   Bruno A, 2014, ONCOTARGET, V5, P5065, DOI 10.18632/oncotarget.2080
   Caro P, 2012, CANCER CELL, V22, P547, DOI 10.1016/j.ccr.2012.08.014
   Chang YS, 2013, P NATL ACAD SCI USA, V110, pE3445, DOI 10.1073/pnas.1303002110
   Chapuy B, 2013, CANCER CELL, V24, P777, DOI 10.1016/j.ccr.2013.11.003
   Cheah CY, 2014, BLOOD, V123, P486, DOI 10.1182/blood-2013-10-530659
   Chen BJ, 2013, CLIN CANCER RES, V19, P3462, DOI 10.1158/1078-0432.CCR-13-0855
   Chen LF, 2013, CANCER CELL, V23, P826, DOI 10.1016/j.ccr.2013.05.002
   Davis RE, 2010, NATURE, V463, P88, DOI 10.1038/nature08638
   De Braekeleer E, 2012, LEUKEMIA RES, V36, P945, DOI 10.1016/j.leukres.2012.04.010
   De Silva NS, 2012, IMMUNOL REV, V247, P73, DOI 10.1111/j.1600-065X.2012.01113.x
   Deckert M, 2014, ACTA NEUROPATHOL, V127, P175, DOI 10.1007/s00401-013-1202-x
   Deng L, 2016, LEUKEMIA, V30, P361, DOI 10.1038/leu.2015.237
   Drier Y, 2013, GENOME RES, V23, P228, DOI 10.1101/gr.141382.112
   Gil J, 2006, NAT REV MOL CELL BIO, V7, P667, DOI 10.1038/nrm1987
   Gonzalez-Aguilar A, 2012, CLIN CANCER RES, V18, P5203, DOI 10.1158/1078-0432.CCR-12-0845
   Green MR, 2015, P NATL ACAD SCI USA, V112, pE1116, DOI 10.1073/pnas.1501199112
   Green MR, 2012, CLIN CANCER RES, V18, P1611, DOI 10.1158/1078-0432.CCR-11-1942
   Green MR, 2010, BLOOD, V116, P3268, DOI 10.1182/blood-2010-05-282780
   Hernando E, 2004, NATURE, V430, P797, DOI 10.1038/nature02820
   Horne MJ, 2011, ARCH PATHOL LAB MED, V135, P1363, DOI 10.5858/arpa.2010-0158-RS
   Huang W, 2009, MOL CELL, V35, P48, DOI 10.1016/j.molcel.2009.05.023
   Jabara HH, 2012, NAT IMMUNOL, V13, P612, DOI 10.1038/ni.2305
   Jeelall YS, 2012, J EXP MED, V209, P1907, DOI 10.1084/jem.20112744
   Karras JR, 2014, CANCERS, V6, P1208, DOI 10.3390/cancers6021208
   Keir ME, 2008, ANNU REV IMMUNOL, V26, P677, DOI 10.1146/annurev.immunol.26.021607.090331
   Kraan W, 2014, LEUKEMIA, V28, P719, DOI 10.1038/leu.2013.348
   Krappmann D, 2013, BLOOD, V122, P2146, DOI 10.1182/blood-2013-08-516864
   Lawrence MS, 2013, NATURE, V499, P214, DOI 10.1038/nature12213
   Lenz G, 2008, SCIENCE, V319, P1676, DOI 10.1126/science.1153629
   Lohr JG, 2012, P NATL ACAD SCI USA, V109, P3879, DOI 10.1073/pnas.1121343109
   Malumbres M, 2009, NAT REV CANCER, V9, P153, DOI 10.1038/nrc2602
   Margolin K, 2012, LANCET ONCOL, V13, P459, DOI 10.1016/S1470-2045(12)70090-6
   Monti S, 2005, BLOOD, V105, P1851, DOI 10.1182/blood-2004-07-2947
   Monti S, 2012, CANCER CELL, V22, P359, DOI 10.1016/j.ccr.2012.07.014
   Morin RD, 2013, BLOOD, V122, P1256, DOI 10.1182/blood-2013-02-483727
   Morin RD, 2011, NATURE, V476, P298, DOI 10.1038/nature10351
   Ngo VN, 2011, NATURE, V470, P115, DOI 10.1038/nature09671
   Nogai H, 2013, BLOOD, V122, P2242, DOI 10.1182/blood-2013-06-508028
   Oishi N, 2015, PATHOL INT, V65, P528, DOI 10.1111/pin.12336
   Pardoll DM, 2012, NAT REV CANCER, V12, P252, DOI 10.1038/nrc3239
   Park HY, 2014, J CLIN INVEST, V124, P4976, DOI 10.1172/JCI75667
   Pasqualucci L, 2011, NAT GENET, V43, P830, DOI 10.1038/ng.892
   Rawlings DJ, 2012, NAT REV IMMUNOL, V12, P282, DOI 10.1038/nri3190
   Riemersma SA, 2000, BLOOD, V96, P3569, DOI 10.1182/blood.V96.10.3569.h8003569_3569_3577
   Rubenstein JL, 2013, BLOOD, V122, P2318, DOI 10.1182/blood-2013-06-453084
   Rui LX, 2010, CANCER CELL, V18, P590, DOI 10.1016/j.ccr.2010.11.013
   Schneider C, 2014, P NATL ACAD SCI USA, V111, P8185, DOI 10.1073/pnas.1322466111
   Shlien A, 2008, P NATL ACAD SCI USA, V105, P11264, DOI 10.1073/pnas.0802970105
   Steidl C, 2011, BLOOD, V118, P2659, DOI 10.1182/blood-2011-05-326538
   Steidl C, 2011, NATURE, V471, P377, DOI 10.1038/nature09754
   Tan Y, 2014, INT C INT SYST MOL B
   Thompson SL, 2010, J CELL BIOL, V188, P369, DOI 10.1083/jcb.200905057
   Thurner L, 2014, BLOOD, V124
   Tijchon E, 2013, LEUKEMIA, V27, P541, DOI 10.1038/leu.2012.293
   Trepel M, 2014, BLOOD, V124
   Twa DDW, 2014, BLOOD, V123, P2062, DOI 10.1182/blood-2013-10-535443
   Vater I, 2015, LEUKEMIA, V29, P677, DOI 10.1038/leu.2014.264
   Wang JQ, 2014, J EXP MED, V211, P413, DOI 10.1084/jem.20131424
   Wang L, 2011, MOL BIOL REP, V38, P229, DOI 10.1007/s11033-010-0099-x
   Yamamoto M, 2004, NATURE, V430, P218, DOI 10.1038/nature02738
   Yang YB, 2012, CANCER CELL, V21, P723, DOI 10.1016/j.ccr.2012.05.024
   Ying CY, 2013, NAT IMMUNOL, V14, P1084, DOI 10.1038/ni.2688
   Zhang J, 2013, P NATL ACAD SCI USA, V110, P1398, DOI 10.1073/pnas.1205299110
NR 71
TC 389
Z9 407
U1 2
U2 32
PU AMER SOC HEMATOLOGY
PI WASHINGTON
PA 2021 L ST NW, SUITE 900, WASHINGTON, DC 20036 USA
SN 0006-4971
EI 1528-0020
J9 BLOOD
JI Blood
PD FEB 18
PY 2016
VL 127
IS 7
BP 869
EP 881
DI 10.1182/blood-2015-10-673236
PG 13
WC Hematology
WE Science Citation Index Expanded (SCI-EXPANDED)
SC Hematology
GA DI3JW
UT WOS:000373395900014
PM 26702065
OA Bronze, Green Published
HC Y
HP N
DA 2024-06-03
ER

PT J
AU Villano, JL
   Koshy, M
   Shaikh, H
   Dolecek, TA
   McCarthy, BJ
AF Villano, J. L.
   Koshy, M.
   Shaikh, H.
   Dolecek, T. A.
   McCarthy, B. J.
TI Age, gender, and racial differences in incidence and survival in primary
   CNS lymphoma
SO BRITISH JOURNAL OF CANCER
LA English
DT Article
DE Lymphoma; brain tumour; epidemiology; incidence; trends
ID NERVOUS-SYSTEM LYMPHOMA; HIGH-DOSE METHOTREXATE; NON-HODGKINS-LYMPHOMA;
   UNITED-STATES; BRAIN-TUMORS; TRENDS; EPIDEMIOLOGY; POPULATION;
   RADIOTHERAPY
AB BACKGROUND: Primary central nervous system lymphoma (PCNSL) is a rare subtype of extranodal non-Hodgkin lymphoma that accounts for similar to 4% of newly diagnosed central nervous system (CNS) tumours. The objective of this study was to analyse the epidemiology, incidence, and outcome of these rare tumours.
   METHODS: Primary brain and CNS lymphoma cases were identified from the Surveillance, Epidemiology, and End Results (SEER) research data sets for the years 1980-2008 for analysis of trends in incidence and survival. SEER*Stat v. 7.0.4 software was used to analyse the data.
   RESULTS: The overall incidence rate of PCNSL was 0.47 per 100 000 person-years. The incidence was significantly higher in males compared with females, blacks aged 0-49 years at diagnosis compared with whites, and whites aged 50 years and older at diagnosis compared with blacks. After a significant decline in incidence between 1995 and 1999, incidence rates rose slightly; those aged 75 + years at diagnosis had the most dramatic increase in incidence rates over time. Five-year survival rates were significantly higher in whites compared with blacks aged 0-49 years at diagnosis, but was primarily driven by white women aged 0-49 years.
   CONCLUSION: There is an increase in incidence of PCNSL in the elderly, and elderly blacks have lower incidence compared with white population. Survival remains poor and is negatively dominated by factors associated with HIV infection and advanced age. British Journal of Cancer (2011) 105, 1414-1418. doi:10.1038/bjc.2011.357 www.bjcancer.com Published online 13 September 2011 (C) 2011 Cancer Research UK
C1 [Villano, J. L.; Shaikh, H.] Univ Illinois, Dept Med, Hematol Oncol Sect, Chicago, IL 60680 USA.
   [Koshy, M.] Univ Illinois, Dept Radiat Therapy, Chicago, IL USA.
   [Koshy, M.] Univ Chicago, Dept Radiat & Cellular Oncol, Chicago, IL 60637 USA.
   [Dolecek, T. A.; McCarthy, B. J.] Univ Illinois, Dept Epidemiol Biostat, Chicago, IL USA.
C3 University of Illinois System; University of Illinois Chicago;
   University of Illinois Chicago Hospital; University of Illinois System;
   University of Illinois Chicago; University of Illinois Chicago Hospital;
   University of Chicago; University of Illinois System; University of
   Illinois Chicago; University of Illinois Chicago Hospital
RP Villano, JL (corresponding author), Univ Illinois, Dept Med, Hematol Oncol Sect, 840 S Wood St,M-C 713,820-E,Room 839, Chicago, IL 60680 USA.
EM
OI Koshy, Matthew/0000-0001-6937-7058
FU American Brain Tumor Association; National Brain Tumor Society;
   Pediatric Brain Tumor Foundation; National Cancer Institute, National
   Institutes of Health, Department of Health and Human Services
   [HHSN261000800766P, HHSN261201000576P]
FX All analyses were conducted under contract to the Central Brain Tumor
   Registry of the United States, which receives funding from American
   Brain Tumor Association, National Brain Tumor Society, Pediatric Brain
   Tumor Foundation, and National Cancer Institute, National Institutes of
   Health, Department of Health and Human Services no. HHSN261000800766P
   and no. HHSN261201000576P.
CR Abrey LE, 2009, CURR OPIN NEUROL, V22, P675, DOI 10.1097/WCO.0b013e328332533b
   [Anonymous], 2008, Applied survival analysis: regression modeling of time to event data
   [Anonymous], SEER STAT DAT INC SE
   BROWN CC, 1983, BIOMETRICS, V39, P941, DOI 10.2307/2531329
   Campo E, 2011, BLOOD, V117, P5019, DOI 10.1182/blood-2011-01-293050
   CBTRUS, 2011, CBTRUS STAT REP PRIM
   Chamberlain MC, 2010, NEURO-ONCOLOGY, V12, P736, DOI 10.1093/neuonc/noq011
   Cote TR, 1996, J NATL CANCER I, V88, P675, DOI 10.1093/jnci/88.10.675
   FINE HA, 1993, ANN INTERN MED, V119, P1093, DOI 10.7326/0003-4819-119-11-199312010-00007
   Fritz A.G., 2013, International Classification of Diseases for Oncology (ICD-O)
   Ghafoor A, 2002, CA-CANCER J CLIN, V52, P326, DOI 10.3322/canjclin.52.6.326
   Haldorsen IS, 2007, CANCER, V110, P1803, DOI 10.1002/cncr.22989
   Hoffman S, 2006, NEURO-ONCOLOGY, V8, P27, DOI 10.1215/S1522851705000323
   Kadan-Lottick NS, 2002, CANCER, V95, P193, DOI 10.1002/cncr.10643
   Kim HJ, 2000, STAT MED, V19, P335, DOI 10.1002/(SICI)1097-0258(20000215)19:3<335::AID-SIM336>3.0.CO;2-Z
   KROGHJENSEN M, 1995, LEUKEMIA LYMPHOMA, V19, P223
   MILLER DC, 1994, CANCER-AM CANCER SOC, V74, P1383, DOI 10.1002/1097-0142(19940815)74:4<1383::AID-CNCR2820740432>3.0.CO;2-1
   Montesinos-Rongen M, 2008, LEUKEMIA, V22, P400, DOI 10.1038/sj.leu.2405019
   Morris M, 2006, AM J PUBLIC HEALTH, V96, P1091, DOI 10.2105/AJPH.2004.054759
   Morton LM, 2006, BLOOD, V107, P265, DOI 10.1182/blood-2005-06-2508
   Müller AMS, 2005, ANN HEMATOL, V84, P1, DOI 10.1007/s00277-004-0939-7
   Norden AD, 2011, J NEURO-ONCOL, V101, P487, DOI 10.1007/s11060-010-0269-7
   O'Brien PC, 2009, LANCET, V374, P1477, DOI 10.1016/S0140-6736(09)61488-4
   Olson JE, 2002, CANCER, V95, P1504, DOI 10.1002/cncr.10851
   Panageas KS, 2005, CANCER-AM CANCER SOC, V104, P2466, DOI 10.1002/cncr.21481
   Pulido JS, 2009, NEURO-ONCOLOGY, V11, P318, DOI 10.1215/15228517-2008-103
   Schabet M, 1999, J NEURO-ONCOL, V43, P199, DOI 10.1023/A:1006290032052
   Shiels MS, 2011, J NATL CANCER I, V103, P753, DOI 10.1093/jnci/djr076
   *SURV RES PROGR, 2010, NCI SEER STAT SOFTW
   Thiel E, 2010, LANCET ONCOL, V11, P1036, DOI 10.1016/S1470-2045(10)70229-1
   Westin EH, 2004, SEMIN ONCOL, V31, P198, DOI 10.1053/j.seminoncol.2003.12.030
NR 31
TC 355
Z9 392
U1 1
U2 11
PU NATURE PUBLISHING GROUP
PI LONDON
PA MACMILLAN BUILDING, 4 CRINAN ST, LONDON N1 9XW, ENGLAND
SN 0007-0920
EI 1532-1827
J9 BRIT J CANCER
JI Br. J. Cancer
PD OCT 25
PY 2011
VL 105
IS 9
BP 1414
EP 1418
DI 10.1038/bjc.2011.357
PG 5
WC Oncology
WE Science Citation Index Expanded (SCI-EXPANDED)
SC Oncology
GA 838IR
UT WOS:000296282000024
PM 21915121
OA Green Published, hybrid
DA 2024-06-03
ER

PT J
AU Lionakis, MS
   Dunleavy, K
   Roschewski, M
   Widemann, BC
   Butman, JA
   Schmitz, R
   Yang, Y
   Cole, DE
   Melani, C
   Higham, CS
   Desai, JV
   Ceribelli, M
   Chen, L
   Thomas, CJ
   Little, RF
   Gea-Banacloche, J
   Bhaumik, S
   Stetler-Stevenson, M
   Pittaluga, S
   Jaffe, ES
   Heiss, J
   Lucas, N
   Steinberg, SM
   Staudt, LM
   Wilson, WH
AF Lionakis, Michail S.
   Dunleavy, Kieron
   Roschewski, Mark
   Widemann, Brigitte C.
   Butman, John A.
   Schmitz, Roland
   Yang, Yandan
   Cole, Diane E.
   Melani, Christopher
   Higham, Christine S.
   Desai, Jigar V.
   Ceribelli, Michele
   Chen, Lu
   Thomas, Craig J.
   Little, Richard F.
   Gea-Banacloche, Juan
   Bhaumik, Sucharita
   Stetler-Stevenson, Maryalice
   Pittaluga, Stefania
   Jaffe, Elaine S.
   Heiss, John
   Lucas, Nicole
   Steinberg, Seth M.
   Staudt, Louis M.
   Wilson, Wyndham H.
TI Inhibition of B Cell Receptor Signaling by Ibrutinib in Primary CNS
   Lymphoma
SO CANCER CELL
LA English
DT Article
ID PEGYLATED LIPOSOMAL DOXORUBICIN; BRUTONS TYROSINE KINASE;
   ASPERGILLUS-FUMIGATUS; CD79B MUTATIONS; ALVEOLAR MACROPHAGES;
   IMMUNE-RESPONSES; ONCOGENIC MYD88; HIGH PREVALENCE; SYSTEM; ACTIVATION
AB Primary CNS lymphoma (PCNSL) harbors mutations that reinforce B cell receptor (BCR) signaling. Ibrutinib, a Bruton's tyrosine kinase (BTK) inhibitor, targets BCR signaling and is particularly active in lymphomas with mutations altering the BCR subunit CD79B and MYD88. We performed a proof-of-concept phase Ib study of ibrutinib monotherapy followed by ibrutinib plus chemotherapy (DA-TEDDi-R). In 18 PCNSL patients, 94% showed tumor reductions with ibrutinib alone, including patients having PCNSL with CD79B and/or MYD88 mutations, and 86% of evaluable patients achieved complete remission with DA-TEDDi-R. Increased aspergillosis was observed with ibrutinib monotherapy and DA-TEDDi-R. Aspergillosis was linked to BTK-dependent fungal immunity in a murine model. PCNSL is highly dependent on BCR signaling, and ibrutinib appears to enhance the efficacy of chemotherapy.
C1 [Lionakis, Michail S.] NIAID, Lab Clin Infect Dis, NIH, 9000 Rockville Pike, Bethesda, MD 20892 USA.
   [Dunleavy, Kieron; Roschewski, Mark; Schmitz, Roland; Yang, Yandan; Melani, Christopher; Desai, Jigar V.; Thomas, Craig J.; Lucas, Nicole; Staudt, Louis M.; Wilson, Wyndham H.] NCI, Lymphoid Malignancies Branch, Ctr Canc Res, NIH, Bethesda, MD 20892 USA.
   [Widemann, Brigitte C.; Cole, Diane E.; Higham, Christine S.; Gea-Banacloche, Juan; Bhaumik, Sucharita; Stetler-Stevenson, Maryalice; Pittaluga, Stefania; Jaffe, Elaine S.; Heiss, John; Steinberg, Seth M.] NCI, Pediat Oncol Branch, Ctr Canc Res, NIH, Bethesda, MD 20892 USA.
   [Butman, John A.] NIH, Radiol & Imaging Sci, Ctr Clin, Bethesda, MD 20892 USA.
   [Ceribelli, Michele; Chen, Lu; Thomas, Craig J.] Natl Ctr Adv Translat Sci, Div Preclin Innovat, NIH, Gaithersburg, MD 20850 USA.
   [Little, Richard F.] NCI, Canc Therapy Evaluat Program, NIH, Bethesda, MD 20892 USA.
C3 National Institutes of Health (NIH) - USA; NIH National Institute of
   Allergy & Infectious Diseases (NIAID); National Institutes of Health
   (NIH) - USA; NIH National Cancer Institute (NCI); National Institutes of
   Health (NIH) - USA; NIH National Cancer Institute (NCI); National
   Institutes of Health (NIH) - USA; NIH Clinical Center (CC); National
   Institutes of Health (NIH) - USA; National Institutes of Health (NIH) -
   USA; NIH National Cancer Institute (NCI)
RP Staudt, LM; Wilson, WH (corresponding author), NCI, Lymphoid Malignancies Branch, Ctr Canc Res, NIH, Bethesda, MD 20892 USA.
EM
RI Butman, John A/A-2694-2008; Jaffe, Elaine/G-8981-2014; Desai, Jigar
   V/I-3109-2019; Desai, Jigar/JYQ-1128-2024; Lionakis,
   Michail/J-8840-2017; Heiss, John/AAL-8281-2021; Stetler-Stevenson,
   Maryalice/AAV-7393-2020; Staudt, Louis/AAC-5324-2019; Gea-Banacloche,
   Juan/IQW-8012-2023; Butman, John/J-2780-2013
OI Desai, Jigar/0000-0002-8823-0796; Lionakis, Michail/0000-0003-4994-9500;
   Heiss, John/0000-0002-3890-0165; Butman, John/0000-0002-1547-9195;
   Thomas, Craig/0000-0001-9386-9001; Higham, Christine/0000-0002-4618-3813
FU NIH, National Cancer Institute; National Institute of Allergy and
   Infectious Disease
FX This research was supported by the Intramural Research Program of the
   NIH, National Cancer Institute, and National Institute of Allergy and
   Infectious Disease.
CR Abrey LE, 2005, J CLIN ONCOL, V23, P5034, DOI 10.1200/JCO.2005.13.524
   Ambady Prakash, 2015, CNS Oncol, V4, P393, DOI 10.2217/cns.15.34
   Anders CK, 2013, PLOS ONE, V8, DOI 10.1371/journal.pone.0061359
   Anne PL, 2012, J INVEST DERMATOL, V132, P2118, DOI 10.1038/jid.2012.102
   Baldwin AS, 2001, J CLIN INVEST, V107, P241, DOI 10.1172/JCI11991
   Bliss CI, 1939, ANN APPL BIOL, V26, P585, DOI 10.1111/j.1744-7348.1939.tb06990.x
   Braggio E, 2015, CLIN CANCER RES, V21, P3986, DOI 10.1158/1078-0432.CCR-14-2116
   Bruno A, 2014, ONCOTARGET, V5, P5065, DOI 10.18632/oncotarget.2080
   Byrne JC, 2013, J IMMUNOL, V190, P5207, DOI 10.4049/jimmunol.1300057
   Caraglia M, 2006, CANCER CHEMOTH PHARM, V57, P34, DOI 10.1007/s00280-005-0001-z
   Chapuy B, 2016, BLOOD, V127, P869, DOI 10.1182/blood-2015-10-673236
   Chignard M, 2007, CLIN IMMUNOL, V124, P238, DOI 10.1016/j.clim.2007.05.004
   Dagenais TRT, 2009, CLIN MICROBIOL REV, V22, P447, DOI 10.1128/CMR.00055-08
   Davis RE, 2010, NATURE, V463, P88, DOI 10.1038/nature08638
   de Vries R, 2015, BIOANALYSIS, V7, P2713, DOI 10.4155/bio.15.159
   Dubourdeau M, 2006, J IMMUNOL, V177, P3994, DOI 10.4049/jimmunol.177.6.3994
   Dubovsky JA, 2013, BLOOD, V122, P2539, DOI 10.1182/blood-2013-06-507947
   Ferreri AJM, 2003, J CLIN ONCOL, V21, P266, DOI 10.1200/JCO.2003.09.139
   Fiedler K, 2011, BLOOD, V117, P1329, DOI 10.1182/blood-2010-04-281170
   Fishman JA, 2007, NEW ENGL J MED, V357, P2601, DOI 10.1056/NEJMra064928
   Griner LAM, 2014, P NATL ACAD SCI USA, V111, P2349, DOI 10.1073/pnas.1311846111
   Hattori K, 2017, BRIT J HAEMATOL, V177, P492, DOI 10.1111/bjh.14080
   Herbst S, 2015, EMBO MOL MED, V7, P240, DOI 10.15252/emmm.201404556
   Horwood NJ, 2006, J IMMUNOL, V176, P3635, DOI 10.4049/jimmunol.176.6.3635
   Kasperkovitz PV, 2010, J IMMUNOL, V185, P7614, DOI 10.4049/jimmunol.1002760
   KHAN WN, 1995, IMMUNITY, V3, P283, DOI 10.1016/1074-7613(95)90114-0
   Koukourakis MI, 2000, BRIT J CANCER, V83, P1281, DOI 10.1054/bjoc.2000.1459
   Kraan W, 2014, LEUKEMIA, V28, P719, DOI 10.1038/leu.2013.348
   Kraan W, 2013, BLOOD CANCER J, V3, DOI 10.1038/bcj.2013.28
   Langner-Lemercier S, 2016, NEURO-ONCOLOGY, V18, P1297, DOI 10.1093/neuonc/now033
   Lewis RE, 2013, MYCOSES, V56, P638, DOI 10.1111/myc.12081
   Limper AH, 1997, J CLIN INVEST, V99, P2110, DOI 10.1172/JCI119384
   Lin NU, 2004, J CLIN ONCOL, V22, P3608, DOI 10.1200/JCO.2004.01.175
   Lionakis MS, 2003, LANCET, V362, P1828, DOI 10.1016/S0140-6736(03)14904-5
   Liu XR, 2006, DRUG METAB DISPOS, V34, P1443, DOI 10.1124/dmd.105.008201
   Nakamura T, 2016, NEUROPATH APPL NEURO, V42, P279, DOI 10.1111/nan.12259
   Ngo VN, 2011, NATURE, V470, P115, DOI 10.1038/nature09671
   Ni Gabhann J, 2014, PLOS ONE, V9
   Palmer CD, 2008, BLOOD, V111, P1781, DOI 10.1182/blood-2007-07-102343
   Plebani A, 2002, CLIN IMMUNOL, V104, P221, DOI 10.1006/clim.2002.5241
   Ramaprakash H, 2009, INFECT IMMUN, V77, P108, DOI 10.1128/IAI.00998-08
   Ramirez-Ortiz ZG, 2008, INFECT IMMUN, V76, P2123, DOI 10.1128/IAI.00047-08
   Rao GVS, 2003, J TOXICOL ENV HEAL A, V66, P1441, DOI 10.1080/15287390306417
   Rubenstein JL, 2013, BLOOD, V122, P2318, DOI 10.1182/blood-2013-06-453084
   Rubenstein JL, 2013, J CLIN ONCOL, V31, P3061, DOI 10.1200/JCO.2012.46.9957
   Ruchlemer Rosa, 2016, N Engl J Med, V374, P1593, DOI 10.1056/NEJMc1600328
   Scheers E, 2015, DRUG METAB DISPOS, V43, P289, DOI 10.1124/dmd.114.060061
   SIEGAL T, 1995, J NEUROSURG, V83, P1029, DOI 10.3171/jns.1995.83.6.1029
   Singh N, 2005, CURR OPIN INFECT DIS, V18, P342, DOI 10.1097/01.qco.0000172698.52408.be
   Stadler N., 2017, HAEMATOLOGICA
   Taniguchi K, 2016, AM J SURG PATHOL, V40, P324, DOI 10.1097/PAS.0000000000000592
   Vail DM, 2004, SEMIN ONCOL, V31, P16, DOI 10.1053/j.seminoncol.2004.08.002
   Vater I, 2015, LEUKEMIA, V29, P677, DOI 10.1038/leu.2014.264
   Wager CML, 2014, MUCOSAL IMMUNOL, V7, P1023, DOI 10.1038/mi.2014.65
   Wilson WH, 2015, NAT MED, V21, P922, DOI 10.1038/nm.3884
   Wilson WH, 2013, HEMATOL-AM SOC HEMAT, P584, DOI 10.1182/asheducation-2013.1.584
   Young RM, 2015, P NATL ACAD SCI USA, V112, P13447, DOI 10.1073/pnas.1514944112
NR 57
TC 334
Z9 356
U1 1
U2 45
PU CELL PRESS
PI CAMBRIDGE
PA 50 HAMPSHIRE ST, FLOOR 5, CAMBRIDGE, MA 02139 USA
SN 1535-6108
EI 1878-3686
J9 CANCER CELL
JI Cancer Cell
PD JUN 12
PY 2017
VL 31
IS 6
BP 833
EP +
DI 10.1016/j.ccell.2017.04.012
PG 16
WC Oncology; Cell Biology
WE Science Citation Index Expanded (SCI-EXPANDED)
SC Oncology; Cell Biology
GA EX2RS
UT WOS:000403074800011
PM 28552327
OA Green Accepted, hybrid
DA 2024-06-03
ER

PT J
AU Grommes, C
   DeAngelis, LM
AF Grommes, Christian
   DeAngelis, Lisa M.
TI Primary CNS Lymphoma
SO JOURNAL OF CLINICAL ONCOLOGY
LA English
DT Review
ID NERVOUS-SYSTEM LYMPHOMA; HIGH-DOSE METHOTREXATE; RECURRENT PRIMARY CNS;
   NON-HODGKINS-LYMPHOMA; WHOLE-BRAIN RADIOTHERAPY; STEM-CELL
   TRANSPLANTATION; INTERNATIONAL EXTRANODAL LYMPHOMA; ACUTE
   LYMPHOBLASTIC-LEUKEMIA; COMBINED INITIAL THERAPY; MULTICENTER PHASE-II
AB PrimaryCNSlymphoma(PCNSL) is a rare form of extranodal non-Hodgkin lymphoma that is typically confined to the brain, eyes, and cerebrospinal fluid without evidence of systemic spread. The prognosis of patients with PCNSL has improved during the last decades with the introduction of high-dose methotrexate. However, despite recent progress, results after treatment are durable in half of patients, and therapy can be associated with late neurotoxicity. PCNSL is an uncommon tumor, and only four randomized trials and one phase III trial have been completed so far, all in the first-line setting. To our knowledge, no randomized trial has been conducted for recurrent/refractory disease, leaving many questions unanswered about optimal first-line and salvage treatments. This review will give an overview of the presentation, evaluation, and treatment of immunocompetent patients with PCNSL. (C) 2017 by American Society of Clinical Oncology
C1 [Grommes, Christian; DeAngelis, Lisa M.] Mem Sloan Kettering Canc Ctr, 1275 York Ave, New York, NY 10021 USA.
C3 Memorial Sloan Kettering Cancer Center
RP Grommes, C (corresponding author), Mem Sloan Kettering Canc Ctr, Dept Neurol, 1275 York Ave, New York, NY 10065 USA.
EM
OI DeAngelis, Lisa/0000-0002-6290-7818
FU National Institutes of Health/National Cancer Institute Cancer Center
   Support Grant [P30 CA008748]; Lymphoma Research Foundation Career
   Development Award; Memorial Sloan Kettering Brain Tumor Center; Susan
   and Peter Solomon Divisional Fund; Geoffrey Beene Cancer Research
   Center; Cycle for Survival Equinox Innovation Award
FX Supported by the National Institutes of Health/National Cancer Institute
   Cancer Center Support Grant No. P30 CA008748, Lymphoma Research
   Foundation Career Development Award, Memorial Sloan Kettering Brain
   Tumor Center, Susan and Peter Solomon Divisional Fund, Geoffrey Beene
   Cancer Research Center, and Cycle for Survival Equinox Innovation Award.
CR Abrey LE, 2006, J CLIN ONCOL, V24, P5711, DOI 10.1200/JCO.2006.08.2941
   Abrey LE, 2003, J CLIN ONCOL, V21, P4151, DOI 10.1200/JCO.2003.05.024
   Abrey LE, 2005, J CLIN ONCOL, V23, P5034, DOI 10.1200/JCO.2005.13.524
   Abrey LE, 2000, J CLIN ONCOL, V18, P3144, DOI 10.1200/JCO.2000.18.17.3144
   Alizadeh AA, 2000, NATURE, V403, P503, DOI 10.1038/35000501
   Arellano-Rodrigo E, 2003, EUR J HAEMATOL, V70, P219, DOI 10.1034/j.1600-0609.2003.00045.x
   Bataille B, 2000, J NEUROSURG, V92, P261, DOI 10.3171/jns.2000.92.2.0261
   Batchelor T, 2003, J CLIN ONCOL, V21, P1044, DOI 10.1200/JCO.2003.03.036
   Batchelor TT, 2011, NEUROLOGY, V76, P929, DOI 10.1212/WNL.0b013e31820f2d94
   Bellinzona M, 2005, EJSO, V31, P100, DOI 10.1016/j.ejso.2004.10.002
   BORSI JD, 1987, CANCER-AM CANCER SOC, V60, P5, DOI 10.1002/1097-0142(19870701)60:1<5::AID-CNCR2820600103>3.0.CO;2-D
   Braggio E, 2015, CLIN CANCER RES, V21, P3986, DOI 10.1158/1078-0432.CCR-14-2116
   Brugnoletti F, 2009, PEDIATR BLOOD CANCER, V52, P293, DOI 10.1002/pbc.21764
   Bruno A, 2014, ONCOTARGET, V5, P5065, DOI 10.18632/oncotarget.2080
   Camilleri-Broet S, 1998, AM J CLIN PATHOL, V110, P607, DOI 10.1093/ajcp/110.5.607
   Camilleri-Broët S, 2006, BLOOD, V107, P190, DOI 10.1182/blood-2005-03-1024
   Chamberlain MC, 2016, J NEURO-ONCOL, V126, P545, DOI 10.1007/s11060-015-1994-8
   Chamberlain MC, 2014, J NEURO-ONCOL, V118, P155, DOI 10.1007/s11060-014-1411-8
   Chapuy B, 2016, BLOOD, V127, P869, DOI 10.1182/blood-2015-10-673236
   Choquet C, 2016, AM SOC HEM 58 ANN M
   Coiffier B, 2002, NEW ENGL J MED, V346, P235, DOI 10.1056/NEJMoa011795
   Colombat P, 2006, BONE MARROW TRANSPL, V38, P417, DOI 10.1038/sj.bmt.1705452
   DEANGELIS LM, 1992, J CLIN ONCOL, V10, P635, DOI 10.1200/JCO.1992.10.4.635
   DeAngelis LM, 2002, J CLIN ONCOL, V20, P4643, DOI 10.1200/JCO.2002.11.013
   del Rio MS, 2012, J NEURO-ONCOL, V106, P143, DOI 10.1007/s11060-011-0649-7
   Enting RH, 2004, NEUROLOGY, V63, P901, DOI 10.1212/01.WNL.0000137050.43114.42
   Ferreri AJM, 2001, ONCOLOGY-BASEL, V60, P134, DOI 10.1159/000055310
   Ferreri AJM, 2003, J CLIN ONCOL, V21, P266, DOI 10.1200/JCO.2003.09.139
   Ferreri AJM, 2016, LANCET HAEMATOL, V3, pE217, DOI 10.1016/S2352-3026(16)00036-3
   Ferreri AJM, 2009, LANCET, V374, P1512, DOI 10.1016/S0140-6736(09)61416-1
   Fischer L, 2006, ANN ONCOL, V17, P1141, DOI 10.1093/annonc/mdl070
   Gavrilovic IT, 2006, J CLIN ONCOL, V24, P4570, DOI 10.1200/JCO.2006.06.6910
   Ghesquieres H, 2016, AM SOC HEM 58 ANN M
   GLASS J, 1994, J NEUROSURG, V81, P188, DOI 10.3171/jns.1994.81.2.0188
   Glass J, 2016, J CLIN ONCOL, V34, P1620, DOI 10.1200/JCO.2015.64.8634
   Grimm SA, 2007, ANN ONCOL, V18, P1851, DOI 10.1093/annonc/mdm340
   Grommes C, 2016, AM SOC HEM 58 ANN M
   Hans CP, 2004, BLOOD, V103, P275, DOI 10.1182/blood-2003-05-1545
   Herrlinger U, 2000, NEUROLOGY, V54, P1707, DOI 10.1212/WNL.54.8.1707
   Herrlinger U, 2005, ANN NEUROL, V57, P843, DOI 10.1002/ana.20495
   Holdhoff M, 2014, NEUROLOGY, V83, P235, DOI 10.1212/WNL.0000000000000593
   Hottinger AF, 2007, NEUROLOGY, V69, P1178, DOI 10.1212/01.wnl.0000276986.19602.c1
   Houillier C, 2015, NEUROLOGY, V84, P325, DOI 10.1212/WNL.0000000000001158
   Illerhaus G, 2006, J CLIN ONCOL, V24, P3865, DOI 10.1200/JCO.2006.06.2117
   Illerhaus G, 2016, LANCET HAEMATOL, V3, pE388, DOI 10.1016/S2352-3026(16)30050-3
   Iwamoto FM, 2007, CANCER, V110, P2528, DOI 10.1002/cncr.23077
   Jahnke K, 2006, J NEURO-ONCOL, V80, P159, DOI 10.1007/s11060-006-9165-6
   Khan RB, 2002, J NEURO-ONCOL, V58, P175, DOI 10.1023/A:1016077907952
   Korfel A, 2016, J CLIN ONCOL, V34, P1757, DOI 10.1200/JCO.2015.64.9897
   Makino K, 2012, J NEURO-ONCOL, V106, P155, DOI 10.1007/s11060-011-0652-z
   Mead GM, 2000, CANCER, V89, P1359, DOI 10.1002/1097-0142(20000915)89:6<1359::AID-CNCR21>3.0.CO;2-9
   Montemurro M, 2007, ANN ONCOL, V18, P665, DOI 10.1093/annonc/mdl458
   Morris PG, 2013, J CLIN ONCOL, V31, P3971, DOI 10.1200/JCO.2013.50.4910
   Nakamura T, 2016, NEUROPATH APPL NEURO, V42, P279, DOI 10.1111/nan.12259
   Nayak L, 2013, LEUKEMIA LYMPHOMA, V54, P58, DOI 10.3109/10428194.2012.698736
   Nayak L, 2011, NEURO-ONCOLOGY, V13, P525, DOI 10.1093/neuonc/nor014
   NELSON DF, 1992, INT J RADIAT ONCOL, V23, P9, DOI 10.1016/0360-3016(92)90538-S
   Nguyen PL, 2005, J CLIN ONCOL, V23, P1507, DOI 10.1200/JCO.2005.01.161
   O'Brien P, 2000, J CLIN ONCOL, V18, P519, DOI 10.1200/JCO.2000.18.3.519
   O'Neill BP, 1999, INT J RADIAT ONCOL, V43, P559, DOI 10.1016/S0360-3016(98)00450-7
   O'Neill BP, 2013, AM J HEMATOL, V88, P997, DOI 10.1002/ajh.23551
   Omuro AMP, 2005, ARCH NEUROL-CHICAGO, V62, P1595, DOI 10.1001/archneur.62.10.1595
   Omuro A, 2015, LANCET HAEMATOL, V2, pE251, DOI 10.1016/S2352-3026(15)00074-5
   Omuro A, 2015, BLOOD, V125, P1403, DOI 10.1182/blood-2014-10-604561
   ONEILL BP, 1995, INT J RADIAT ONCOL, V33, P663, DOI 10.1016/0360-3016(95)00207-F
   Pasqualucci L, 2015, SEMIN HEMATOL, V52, P67, DOI 10.1053/j.seminhematol.2015.01.005
   Pels H, 2003, J CLIN ONCOL, V21, P4489, DOI 10.1200/JCO.2003.04.056
   Pentsova E, 2014, J NEURO-ONCOL, V117, P161, DOI 10.1007/s11060-014-1370-0
   Plotkin SR, 2004, CLIN CANCER RES, V10, P5643, DOI 10.1158/1078-0432.CCR-04-0159
   Poortmans PMP, 2003, J CLIN ONCOL, V21, P4483, DOI 10.1200/JCO.2003.03.108
   Raizer JJ, 2012, CANCER-AM CANCER SOC, V118, P3743, DOI 10.1002/cncr.26709
   Reni M, 2007, BRIT J CANCER, V96, P864, DOI 10.1038/sj.bjc.6603660
   Reni M, 1999, BRIT J CANCER, V79, P530, DOI 10.1038/sj.bjc.6690083
   Reni M, 1997, ANN ONCOL, V8, P227, DOI 10.1023/A:1008201717089
   Rubenstein JL, 2007, J CLIN ONCOL, V25, P1350, DOI 10.1200/JCO.2006.09.7311
   Rubenstein JL, 2013, J CLIN ONCOL, V31, P3061, DOI 10.1200/JCO.2012.46.9957
   Rubenstein JL, 2013, BLOOD, V121, P745, DOI 10.1182/blood-2012-07-440974
   Ruhstaller TW, 2000, ANN ONCOL, V11, P374, DOI 10.1023/A:1008371602708
   Schultz C, 1996, J CLIN ONCOL, V14, P556, DOI 10.1200/JCO.1996.14.2.556
   SHAPIRO WR, 1975, NEW ENGL J MED, V293, P161, DOI 10.1056/NEJM197507242930402
   Shibamoto Y, 2005, INT J RADIAT ONCOL, V62, P809, DOI 10.1016/j.ijrobp.2004.12.043
   Shibamoto Y, 2003, CANCER-AM CANCER SOC, V97, P128, DOI 10.1002/cncr.11035
   Soussain C, 2008, J CLIN ONCOL, V26, P2512, DOI 10.1200/JCO.2007.13.5533
   Thiel E, 2010, LANCET ONCOL, V11, P1036, DOI 10.1016/S1470-2045(10)70229-1
   Vater I, 2015, LEUKEMIA, V29, P677, DOI 10.1038/leu.2014.264
   Villano JL, 2011, BRIT J CANCER, V105, P1414, DOI 10.1038/bjc.2011.357
   Weller M, 2012, NEURO-ONCOLOGY, V14, P1481, DOI 10.1093/neuonc/nos159
   Wong ET, 2004, CANCER-AM CANCER SOC, V101, P139, DOI 10.1002/cncr.20339
   Wong SF, 2012, J CLIN NEUROSCI, V19, P1501, DOI 10.1016/j.jocn.2012.04.001
   Zhang JP, 2013, J NEURO-ONCOL, V115, P71, DOI 10.1007/s11060-013-1196-1
NR 90
TC 324
Z9 346
U1 1
U2 48
PU AMER SOC CLINICAL ONCOLOGY
PI ALEXANDRIA
PA 2318 MILL ROAD, STE 800, ALEXANDRIA, VA 22314 USA
SN 0732-183X
EI 1527-7755
J9 J CLIN ONCOL
JI J. Clin. Oncol.
PD JUL 20
PY 2017
VL 35
IS 21
BP 2410
EP +
DI 10.1200/JCO.2017.72.7602
PG 11
WC Oncology
WE Science Citation Index Expanded (SCI-EXPANDED)
SC Oncology
GA FA6TX
UT WOS:000405578300010
PM 28640701
OA Green Published
HC Y
HP N
DA 2024-06-03
ER

PT J
AU Rubenstein, JL
   Hsi, ED
   Johnson, JL
   Jung, SH
   Nakashima, MO
   Grant, B
   Cheson, BD
   Kaplan, LD
AF Rubenstein, James L.
   Hsi, Eric D.
   Johnson, Jeffrey L.
   Jung, Sin-Ho
   Nakashima, Megan O.
   Grant, Barbara
   Cheson, Bruce D.
   Kaplan, Lawrence D.
TI Intensive Chemotherapy and Immunotherapy in Patients With Newly
   Diagnosed Primary CNS Lymphoma: CALGB 50202 (Alliance 50202)
SO JOURNAL OF CLINICAL ONCOLOGY
LA English
DT Article
ID CENTRAL-NERVOUS-SYSTEM; WHOLE-BRAIN RADIOTHERAPY; HIGH-DOSE
   METHOTREXATE; QUALITY-OF-LIFE; CENTER B-CELLS; RESPONSE CRITERIA;
   POOR-PROGNOSIS; TEMOZOLOMIDE; IMMUNOCHEMOTHERAPY; EXPRESSION
AB Purpose
   Concerns regarding neurocognitive toxicity of whole-brain radiotherapy (WBRT) have motivated development of alternative, dose-intensive chemotherapeutic strategies as consolidation in primary CNS lymphoma (PCNSL). We performed a multicenter study of high-dose consolidation, without WBRT, in PCNSL. Objectives were to determine: one, rate of complete response (CR) after remission induction therapy with methotrexate, temozolomide, and rituximab (MT-R); two, feasibility of a two-step approach using high-dose consolidation with etoposide plus cytarabine (EA); three, progression-free survival (PFS); and four, correlation between clinical and molecular prognostic factors and outcome.
   Patients and Methods
   Forty-four patients with newly diagnosed PCNSL were treated with induction MT-R, and patients who achieved CR received EA consolidation. We performed a prospective analysis of molecular prognostic biomarkers in PCNSL in the setting of a clinical trial.
   Results
   The rate of CR to MT-R was 66%. The overall 2-year PFS was 0.57, with median follow-up of 4.9 years. The 2-year time to progression was 0.59, and for patients who completed consolidation, it was 0.77. Patients age >60 years did as well as younger patients, and the most significant clinical prognostic variable was treatment delay. High BCL6 expression correlated with shorter survival.
   Conclusion
   CALGB 50202 demonstrates for the first time to our knowledge that dose-intensive consolidation for PCNSL is feasible in the multicenter setting and yields rates of PFS and OS at least comparable to those of regimens involving WBRT. On the basis of these encouraging results, an intergroup study has been activated comparing EA consolidation with myeloablative chemotherapy in this randomized trial in PCNSL, in which neither arm involves WBRT. (C) 2013 by American Society of Clinical Oncology
C1 [Rubenstein, James L.; Kaplan, Lawrence D.] Univ Calif San Francisco, Helen Diller Comprehens Canc Ctr, San Francisco, CA 94143 USA.
   [Hsi, Eric D.; Nakashima, Megan O.] Cleveland Clin, Cleveland, OH 44106 USA.
   [Johnson, Jeffrey L.; Jung, Sin-Ho] Duke Univ, Alliance Stat & Data Ctr, Duke Comprehens Canc Ctr, Durham, NC USA.
   [Grant, Barbara] Univ Vermont, Burlington, VT USA.
   [Cheson, Bruce D.] Georgetown Univ Hosp, Washington, DC 20007 USA.
C3 University of California System; University of California San Francisco;
   UCSF Medical Center; UCSF Helen Diller Family Comprehensive Cancer
   Center; Cleveland Clinic Foundation; Duke University; University of
   Vermont; Georgetown University
RP Rubenstein, JL (corresponding author), Univ Calif San Francisco, 505 Parnassus Ave,Rm M1282, San Francisco, CA 94143 USA.
EM
RI Nakashima, Megan/AAB-8321-2019
OI Nakashima, Megan/0000-0002-8127-7663
FU National Cancer Institute [RO1CA1398301, CA60138, CA21115, CA33601,
   CA77406, CA77597, CA31946]; Leukemia and Lymphoma Society
FX Supported by National Cancer Institute Grants No. RO1CA1398301 and
   CA60138 (J.L.R., L.D.K.), CA21115 (E.D.H., M.O.N.), CA33601 (J.L.J.,
   S.-H.J.), CA77406 (B. G.), CA77597 (B.D.C.), CA31946 to the Alliance for
   Clinical Trials in Oncology, and CA33601 to the Alliance Statistics and
   Data Center. J.L.R. is also supported by the Leukemia and Lymphoma
   Society.
CR Abrey LE, 2006, J CLIN ONCOL, V24, P5711, DOI 10.1200/JCO.2006.08.2941
   Abrey LE, 2005, J CLIN ONCOL, V23, P5034, DOI 10.1200/JCO.2005.13.524
   Abrey LE, 1998, J CLIN ONCOL, V16, P859, DOI 10.1200/JCO.1998.16.3.859
   Angelov L, 2009, J CLIN ONCOL, V27, P3503, DOI 10.1200/JCO.2008.19.3789
   Batchelor T, 2006, J CLIN ONCOL, V24, P1281, DOI 10.1200/JCO.2005.04.8819
   Batchelor T, 2003, J CLIN ONCOL, V21, P1044, DOI 10.1200/JCO.2003.03.036
   Boehme V, 2007, ANN ONCOL, V18, P149, DOI 10.1093/annonc/mdl327
   Braaten KM, 2003, CLIN CANCER RES, V9, P1063
   Camilleri-Broët S, 2006, BLOOD, V107, P190, DOI 10.1182/blood-2005-03-1024
   Cerchietti LC, 2010, CANCER CELL, V17, P400, DOI 10.1016/j.ccr.2009.12.050
   Chang CC, 2003, ARCH PATHOL LAB MED, V127, P208
   Cheson BD, 2007, J CLIN ONCOL, V25, P579, DOI 10.1200/JCO.2006.09.2403
   Damon LE, 2009, J CLIN ONCOL, V27, P6101, DOI 10.1200/JCO.2009.22.2554
   DeAngelis LM, 2002, J CLIN ONCOL, V20, P4643, DOI 10.1200/JCO.2002.11.013
   Ferreri AJM, 2003, J CLIN ONCOL, V21, P266, DOI 10.1200/JCO.2003.09.139
   Ferreri AJM, 2009, LANCET, V374, P1512, DOI 10.1016/S0140-6736(09)61416-1
   Haldorsen IS, 2005, ACTA ONCOL, V44, P728, DOI 10.1080/02841860500256272
   Herrlinger U, 2002, ANN NEUROL, V51, P247, DOI 10.1002/ana.10102
   Hoang-Xuan K, 2003, J CLIN ONCOL, V21, P2726, DOI 10.1200/JCO.2003.11.036
   Hormigo A, 2004, BRIT J HAEMATOL, V126, P202, DOI 10.1111/j.1365-2141.2004.05028.x
   Illerhaus G, 2007, HAEMATOLOGICA, V93, P147, DOI 10.3324/haematol.11771
   Kluk MJ, 2012, PLOS ONE, V7, DOI 10.1371/journal.pone.0033813
   Levy O, 2008, CANCER, V112, P151, DOI 10.1002/cncr.23149
   Momota H, 2010, J NEURO-ONCOL, V98, P341, DOI 10.1007/s11060-009-0078-z
   Montesinos-Rongen M, 1999, AM J PATHOL, V155, P2077, DOI 10.1016/S0002-9440(10)65526-5
   Osoba D, 2000, J CLIN ONCOL, V18, P1481, DOI 10.1200/JCO.2000.18.7.1481
   Osoba D, 2000, EUR J CANCER, V36, P1788, DOI 10.1016/S0959-8049(00)00165-9
   OTT RJ, 1991, EUR J CANCER, V27, P1356, DOI 10.1016/0277-5379(91)90009-3
   Relling MV, 1996, J CLIN ONCOL, V14, P399, DOI 10.1200/JCO.1996.14.2.399
   Reni M, 2007, BRIT J CANCER, V96, P864, DOI 10.1038/sj.bjc.6603660
   Reni M, 2001, ANN HEMATOL, V80, pB113
   Rubenstein JL, 2006, BLOOD, V107, P3716, DOI 10.1182/blood-2005-03-0897
   Shah GD, 2007, J CLIN ONCOL, V25, P4730, DOI 10.1200/JCO.2007.12.5062
   Soussain C, 2001, J CLIN ONCOL, V19, P742, DOI 10.1200/JCO.2001.19.3.742
   Sun A, 2011, J CLIN ONCOL, V29, P279, DOI 10.1200/JCO.2010.29.6053
   Thiel E, 2010, LANCET ONCOL, V11, P1036, DOI 10.1016/S1470-2045(10)70229-1
   Thompsett AR, 1999, BLOOD, V94, P1738, DOI 10.1182/blood.V94.5.1738.417k10_1738_1746
   Wieduwilt MJ, 2012, CLIN CANCER RES, V18, P1146, DOI 10.1158/1078-0432.CCR-11-0625
   Winter JN, 2006, BLOOD, V107, P4207, DOI 10.1182/blood-2005-10-4222
   Wong ET, 2004, CANCER-AM CANCER SOC, V101, P139, DOI 10.1002/cncr.20339
NR 40
TC 317
Z9 332
U1 0
U2 25
PU AMER SOC CLINICAL ONCOLOGY
PI ALEXANDRIA
PA 2318 MILL ROAD, STE 800, ALEXANDRIA, VA 22314 USA
SN 0732-183X
EI 1527-7755
J9 J CLIN ONCOL
JI J. Clin. Oncol.
PD SEP 1
PY 2013
VL 31
IS 25
BP 3061
EP +
DI 10.1200/JCO.2012.46.9957
PG 13
WC Oncology
WE Science Citation Index Expanded (SCI-EXPANDED)
SC Oncology
GA 301OC
UT WOS:000330540600006
PM 23569323
OA Bronze, Green Published
DA 2024-06-03
ER

PT J
AU Nayak, L
   Iwamoto, FM
   LaCasce, A
   Mukundan, S
   Roemer, MGM
   Chapuy, B
   Armand, P
   Rodig, SJ
   Shipp, MA
AF Nayak, Lakshmi
   Iwamoto, Fabio M.
   LaCasce, Ann
   Mukundan, Srinivasan
   Roemer, Margaretha G. M.
   Chapuy, Bjoern
   Armand, Philippe
   Rodig, Scott J.
   Shipp, Margaret A.
TI PD-1 blockade with nivolumab in relapsed/refractory primary central
   nervous system and testicular lymphoma
SO BLOOD
LA English
DT Article
ID PRIMARY CNS LYMPHOMA; CLASSICAL HODGKIN LYMPHOMA; STEM-CELL
   TRANSPLANTATION; WHOLE-BRAIN RADIATION; IMMUNOCOMPETENT PATIENTS;
   FEATURES; FAILURE; DIAGNOSIS; SINGLE
AB Primary central nervous system (CNS) lymphoma (PCNSL) and primary testicular lymphoma (PTL) are rare extranodal large B-cell lymphomas with similar genetic signatures. There are no standard-of-care treatment options for patients with relapsed and refractory PCNSL and PTL, and the overall prognosis is poor. PCNSLs and PTLs exhibit frequent 9p24.1 copy-number alterations and infrequent translocations of 9p24.1 and associated increased expression of the programmed cell death protein 1 (PD-1) ligands, PD-L1 and PD-L2. The activity of PD-1 blockade in other lymphomas with 9p24.1 alterations prompted us to test the efficacy of the anti-PD1 antibody, nivolumab, in 4 patients with relapsed/refractory PCNSL and 1 patient with CNS relapse of PTL. All 5 patients had clinical and radiographic responses to PD-1 blockade, and 3 patients remain progression-free at 13 1 to 17 1 months. Our data suggest that nivolumab is active in relapsed/refractory PCNSL and PTL and support further investigation of PD-1 blockade in these diseases.
C1 [Nayak, Lakshmi; LaCasce, Ann; Mukundan, Srinivasan; Roemer, Margaretha G. M.; Chapuy, Bjoern; Armand, Philippe; Rodig, Scott J.; Shipp, Margaret A.] Dana Farber Canc Inst, 450 Brookline Ave,DA 2120, Boston, MA 02215 USA.
   [Nayak, Lakshmi; LaCasce, Ann; Mukundan, Srinivasan; Armand, Philippe; Rodig, Scott J.; Shipp, Margaret A.] Brigham & Womens Hosp, 75 Francis St, Boston, MA 02115 USA.
   [Iwamoto, Fabio M.] New York Presbyterian Hosp, New York, NY USA.
C3 Harvard University; Dana-Farber Cancer Institute; Harvard University;
   Brigham & Women's Hospital; NewYork-Presbyterian Hospital
RP Nayak, L (corresponding author), Dana Farber Canc Inst, 450 Brookline Ave,DA 2120, Boston, MA 02215 USA.
EM
RI iwamoto, fabio/KJX-6609-2024; Rodig, Scott/Y-3889-2019
FU Novocure; Bristol-Myers Squibb; Celldex; Northwest Biotherapeutics;
   Stemline; Regeneron; Incyte; Immunocellular Therapeutics; Merck;
   Affimed; Pfizer; Otsuka; Sequenta; Sigma Tau; Bayer
FX F.M.I. is a consultant for Merck, Novocure, Regeneron, Abbvie, and
   Alexion and received research support from Novocure, Bristol-Myers
   Squibb, Celldex, Northwest Biotherapeutics, Stemline, Regeneron, Incyte,
   Immunocellular Therapeutics, and Merck. A.L. is a member of the
   Bristol-Mers Squibb Data & Safety Monitoring Board. P.A. is a consultant
   for Merck, Bristol-Myers Squibb, and Infinity and received research
   support from Bristol-Myers Squibb, Merck, Affimed, Pfizer, Otsuka,
   Sequenta, and Sigma Tau. S.J.R. received research funding from
   Bristol-Myers Squibb. M.A.S. is a consultant for Bristol-Myers Squibb,
   Cell Signaling, Gilead, Takeda, Seattle Genetics, and AstraZeneca and
   received research funding from Bristol-Myers Squibb and Bayer. The
   remaining authors declare no competing financial interests.
CR Ansell SM, 2015, NEW ENGL J MED, V372, P311, DOI 10.1056/NEJMoa1411087
   Armand P, J CLIN ONCOL
   Braggio E, 2015, CLIN CANCER RES, V21, P3986, DOI 10.1158/1078-0432.CCR-14-2116
   Bruno A, 2014, ONCOTARGET, V5, P5065, DOI 10.18632/oncotarget.2080
   Chapuy B, 2016, BLOOD, V127, P869, DOI 10.1182/blood-2015-10-673236
   Cheah CY, 2014, BLOOD, V123
   Correa DD, 2012, NEURO-ONCOLOGY, V14, P101, DOI 10.1093/neuonc/nor186
   Cortazar FB, 2016, KIDNEY INT, V90, P638, DOI 10.1016/j.kint.2016.04.008
   Deng L, 2016, LEUKEMIA, V30, P361, DOI 10.1038/leu.2015.237
   Ferreri AJM, 2016, BLOOD, V127, P1642, DOI 10.1182/blood-2015-10-636340
   Hoang-Xuan K, 2015, LANCET ONCOL, V16, pE322, DOI 10.1016/S1470-2045(15)00076-5
   Kasenda B, 2016, BLOOD, V128, P32, DOI 10.1182/blood-2016-01-650101
   Korfel A, 2013, NAT REV NEUROL, V9, P317, DOI 10.1038/nrneurol.2013.83
   Kraan W, 2014, LEUKEMIA, V28, P719, DOI 10.1038/leu.2013.348
   Langner-Lemercier S, 2016, NEURO-ONCOLOGY, V18, P1297, DOI 10.1093/neuonc/now033
   Moskowitz CH, 2016, BLOOD, V128, DOI 10.1182/blood.V128.22.1107.1107
   Nayak L, 2013, CURR TREAT OPTION ON, V14, P539, DOI 10.1007/s11864-013-0252-6
   Nguyen PL, 2005, J CLIN ONCOL, V23, P1507, DOI 10.1200/JCO.2005.01.161
   Pauken KE, 2015, TRENDS IMMUNOL, V36, P265, DOI 10.1016/j.it.2015.02.008
   Roemer MGM, 2016, J CLIN ONCOL, V34, P2690, DOI 10.1200/JCO.2016.66.4482
   Soussain C, 2008, J CLIN ONCOL, V26, P2512, DOI 10.1200/JCO.2007.13.5533
   Vater I, 2015, LEUKEMIA, V29, P677, DOI 10.1038/leu.2014.264
   Younes A, 2016, LANCET ONCOL, V17, P1283, DOI 10.1016/S1470-2045(16)30167-X
NR 23
TC 307
Z9 325
U1 2
U2 42
PU AMER SOC HEMATOLOGY
PI WASHINGTON
PA 2021 L ST NW, SUITE 900, WASHINGTON, DC 20036 USA
SN 0006-4971
EI 1528-0020
J9 BLOOD
JI Blood
PD JUN 8
PY 2017
VL 129
IS 23
BP 3071
EP 3073
DI 10.1182/blood-2017-01-764209
PG 3
WC Hematology
WE Science Citation Index Expanded (SCI-EXPANDED)
SC Hematology
GA EW9YK
UT WOS:000402875100006
PM 28356247
OA Green Published, Bronze
HC Y
HP N
DA 2024-06-03
ER

PT J
AU Pels, H
   Schmidt-Wolf, IGH
   Glasmacher, A
   Schulz, H
   Engert, A
   Diehl, V
   Zellner, A
   Schackert, G
   Reichmann, H
   Kroschinsky, F
   Vogt-Schaden, M
   Egerer, G
   Bode, U
   Schaller, C
   Deckert, M
   Fimmers, R
   Helmstaedter, C
   Atasoy, A
   Klockgether, T
   Schlegel, U
AF Pels, H
   Schmidt-Wolf, IGH
   Glasmacher, A
   Schulz, H
   Engert, A
   Diehl, V
   Zellner, A
   Schackert, G
   Reichmann, H
   Kroschinsky, F
   Vogt-Schaden, M
   Egerer, G
   Bode, U
   Schaller, C
   Deckert, M
   Fimmers, R
   Helmstaedter, C
   Atasoy, A
   Klockgether, T
   Schlegel, U
TI Primary central nervous system lymphoma: Results of a pilot and phase II
   study of systemic and intraventricular chemotherapy with deferred
   radiotherapy
SO JOURNAL OF CLINICAL ONCOLOGY
LA English
DT Article; Proceedings Paper
CT 44th Annual Meeting of the American-Society-of-Hematology
CY DEC 06-10, 2002
CL PHILADELPHIA, PA
SP Amer Soc Hematol
ID PRIMARY CNS LYMPHOMA; HIGH-DOSE METHOTREXATE; NON-HODGKINS-LYMPHOMA;
   INTRAOCULAR LYMPHOMA; THERAPY; TRIAL; MULTICENTER; NEOPLASMS; SURVIVAL;
   TEMOZOLOMIDE
AB Purpose: To evaluate response rate, response duration, overall survival (OS), and toxicity in primary CNS lymphoma (PCNSL) after systemic and intraventricular chemotherapy with deferred radiotherapy.
   Patients and Methods: From September 1995 to July 2001, 65 consecutive patients with PCNSL (median age, 62 years) were enrolled onto a pilot and phase 11 study evaluating chemotherapy without radiotherapy. A high-dose methotrexate (MTX; cycles 1, 2, 4, and 5) and cytarabine (ARA-C; cycles 3 and 6)-based systemic therapy (including dexamethasone, vinca-alkaloids, ifosfamide, and cyclophosphamide) was combined with intraventricular MTX, prednisolone, and ARA-C.
   Results: Sixty-one of 65 patients were assessable for response. Of these, 37 patients (61%) achieved complete response, six (10%) achieved partial response, and 12 (19%) progressed under therapy. Six (9%) of 65 patients died because of treatment-related complications. Follow-up is 0 to 87 months (median, 26 months). The Kaplan-Meier estimates for median time to treatment failure (TTF) and median OS were 21 months and 50 months, respectively. For patients older than 60 years, median survival was 34 months, and the median TTF was 15 months. In patients younger than 61 years, median survival and median TTF have not been reached yet; the 5-year survival fraction is 75%. Systemic toxicity was mainly hematologic. Ommaya reservoir infection occurred in 12 patients (19%), and permanent cognitive dysfunction possibly as a result of treatment occurred in only two patients (3%).
   Conclusion: Primary chemotherapy based on high-dose MTX and ARA-C is highly efficient in PCNSL. Response rate and response duration in this series are comparable to the response rates and durations reported after combined radiotherapy and chemotherapy. Neurotoxicity was infrequent. (C) 2003 by American Society of Clinical Oncology.
C1 Univ Bonn, Inst Biostat, Dept Neurol, D-5300 Bonn, Germany.
   Univ Bonn, Inst Biostat, Dept Internal Med, D-5300 Bonn, Germany.
   Univ Bonn, Inst Biostat, Dept Pediat Hematooncol, D-5300 Bonn, Germany.
   Univ Bonn, Inst Biostat, Dept Neurosurg, D-5300 Bonn, Germany.
   Univ Bonn, Dept Epileptol, Neuropsychol Unit, D-5300 Bonn, Germany.
   Univ Cologne, Dept Internal Med, D-5000 Cologne 41, Germany.
   Univ Cologne, Dept Neuropathol, D-5000 Cologne 41, Germany.
   Univ Dresden, Dept Neurosurg, Dresden, Germany.
   Univ Dresden, Dept Neurol, Dresden, Germany.
   Univ Dresden, Dept Internal Med, Dresden, Germany.
   Heidelberg Univ, Dept Neurol, D-6900 Heidelberg, Germany.
   Heidelberg Univ, Dept Internal Med, D-6900 Heidelberg, Germany.
C3 University of Bonn; University of Bonn; University of Bonn; University
   of Bonn; University of Bonn; University of Cologne; University of
   Cologne; Technische Universitat Dresden; Technische Universitat Dresden;
   Technische Universitat Dresden; Ruprecht Karls University Heidelberg;
   Ruprecht Karls University Heidelberg
RP Univ Hosp Bonn, Dept Neurol, Sigmund Freud Str 25, D-53105 Bonn, Germany.
EM
RI Deckert, Martina/P-2842-2016; Glasmacher, Axel/J-9634-2019
OI Glasmacher, Axel/0000-0003-2611-7656
CR Abrey LE, 1998, J CLIN ONCOL, V16, P859, DOI 10.1200/JCO.1998.16.3.859
   Abrey LE, 2000, J CLIN ONCOL, V18, P3144, DOI 10.1200/JCO.2000.18.17.3144
   [Anonymous], 2001, PATHOLOGY GENETICS T
   Batchelor T, 2003, J CLIN ONCOL, V21, P1044, DOI 10.1200/JCO.2003.03.036
   Bessell EM, 2001, INT J RADIAT ONCOL, V50, P457, DOI 10.1016/S0360-3016(01)01451-1
   Blay JY, 1998, J CLIN ONCOL, V16, P864, DOI 10.1200/JCO.1998.16.3.864
   BLEYER WA, 1978, BLOOD, V51, P835
   Central Brain Tumor Registry of the United States, 2000, STAT REP PRIM BRAIN
   DeAngelis L M, 2001, Curr Treat Options Oncol, V2, P309, DOI 10.1007/s11864-001-0024-6
   DEANGELIS LM, 1992, J CLIN ONCOL, V10, P635, DOI 10.1200/JCO.1992.10.4.635
   DeAngelis LM, 2002, J CLIN ONCOL, V20, P4643, DOI 10.1200/JCO.2002.11.013
   Ferreri AJM, 2001, ONCOLOGY-BASEL, V60, P134, DOI 10.1159/000055310
   Ferreri AJM, 2002, NEUROLOGY, V58, P1513, DOI 10.1212/WNL.58.10.1513
   Fine HA, 2002, J CLIN ONCOL, V20, P4615, DOI 10.1200/JCO.2002.20.24.4615
   Fliessbach K, 2003, ARCH NEUROL-CHICAGO, V60, P563, DOI 10.1001/archneur.60.4.563
   Guha-Thakurta N, 1999, J NEURO-ONCOL, V43, P259, DOI 10.1023/A:1006210703827
   HARRIS NL, 1994, BLOOD, V84, P1361
   Herrlinger U, 2002, NEUROLOGY, V58, P1573, DOI 10.1212/WNL.58.10.1573
   Herrlinger U, 2002, ANN NEUROL, V51, P247, DOI 10.1002/ana.10102
   Hiraga S, 1999, J NEUROSURG, V91, P221, DOI 10.3171/jns.1999.91.2.0221
   KAPLAN EL, 1958, J AM STAT ASSOC, V53, P457, DOI 10.2307/2281868
   Khan RB, 2002, J NEURO-ONCOL, V58, P175, DOI 10.1023/A:1016077907952
   MACDONALD DR, 1990, J CLIN ONCOL, V8, P1277, DOI 10.1200/JCO.1990.8.7.1277
   Maher EA, 1999, SEMIN ONCOL, V26, P346
   MANTEL NATHAN, 1966, CANCERCHEMOTHERAP REP, V50, P163
   McAllister LD, 2000, NEUROSURGERY, V46, P51, DOI 10.1093/neurosurgery/46.1.51
   Nelson DF, 1999, J NEURO-ONCOL, V43, P241, DOI 10.1023/A:1006206602918
   NELSON DF, 1992, INT J RADIAT ONCOL, V23, P9, DOI 10.1016/0360-3016(92)90538-S
   O'Brien P, 2000, J CLIN ONCOL, V18, P519, DOI 10.1200/JCO.2000.18.3.519
   Reiter A, 1999, BLOOD, V94, P3294
   Reni M, 2000, J NATL CANCER I, V92, P575, DOI 10.1093/jnci/92.7.575
   Sandor V, 1998, J CLIN ONCOL, V16, P3000, DOI 10.1200/JCO.1998.16.9.3000
   Schlegel U, 2001, J NEUROL NEUROSUR PS, V71, P118, DOI 10.1136/jnnp.71.1.118
   Soussain C, 2001, J CLIN ONCOL, V19, P742, DOI 10.1200/JCO.2001.19.3.742
   World Health Organization, 1979, WHO OFFS PUBL, P14
NR 35
TC 296
Z9 307
U1 0
U2 14
PU AMER SOC CLINICAL ONCOLOGY
PI ALEXANDRIA
PA 2318 MILL ROAD, STE 800, ALEXANDRIA, VA 22314 USA
SN 0732-183X
EI 1527-7755
J9 J CLIN ONCOL
JI J. Clin. Oncol.
PD DEC 15
PY 2003
VL 21
IS 24
BP 4489
EP 4495
DI 10.1200/JCO.2003.04.056
PG 7
WC Oncology
WE Science Citation Index Expanded (SCI-EXPANDED); Conference Proceedings Citation Index - Science (CPCI-S)
SC Oncology
GA 755VN
UT WOS:000187432300007
PM 14597744
DA 2024-06-03
ER

PT J
AU Doolittle, ND
   Miner, ME
   Hall, WA
   Siegal, T
   Hanson, EJ
   Osztie, E
   McAllister, LD
   Bubalo, JS
   Kraemer, DF
   Fortin, D
   Nixon, R
   Muldoon, LL
   Neuwelt, EA
AF Doolittle, ND
   Miner, ME
   Hall, WA
   Siegal, T
   Hanson, EJ
   Osztie, E
   McAllister, LD
   Bubalo, JS
   Kraemer, DF
   Fortin, D
   Nixon, R
   Muldoon, LL
   Neuwelt, EA
TI Safety and efficacy of a multicenter study using intraarterial
   chemotherapy in conjunction with osmotic opening of the blood-brain
   barrier for the treatment of patients with malignant brain tumors
SO CANCER
LA English
DT Article
DE neuro-oncology; brain tumors; blood-brain barrier; intraarterial
   chemotherapy
ID INTRACAROTID INFUSION; ANAPLASTIC GLIOMAS; BRADYKININ ANALOG;
   DRUG-DELIVERY; RODENT MODEL; PHASE-II; CISPLATIN; CARBOPLATIN;
   DISRUPTION; ETOPOSIDE
AB BACKGROUND. The aim of this study was to determine the safety and efficacy of intraarterial chemotherapy with osmotic opening of the blood-brain barrier (BBB) for the treatment of malignant brain tumors when administered across multiple centers.
   METHODS. Patients with primary central nervous system lymphoma (PCNSL), primitive neuroectodermal tumor (PNET), germ cell tumor, cancer metastasis to the brain, or low or high grade glioma were eligible. Prior to entry, magnetic resonance imaging or computed tomography brain scan, medical history, neurologic status, and Karnofsky performance status were reviewed at the coordinating center. Standardized anesthesia and intraarterial catheterization guidelines were followed by a multidisciplinary ream at each center. Between March 1994 and November 1997, 5 universities treated 221 adult patients with intraarterial chemotherapy with or without osmotic opening of the BBB (2464 procedures).
   RESULTS. Of evaluable patients with PCNSL, 40 of 53 (75%) achieved complete response (CR). All evaluable patients with PNET (n = 17), metastatic disease (n = 12), or germ cell tumor (n = 4) achieved stable disease (SD) or better. Of 57 evaluable patients with glioblastoma multiforme, 45 (79%) achieved SD or better. Asymptomatic subintimal tear occurred in 11 of 221 patients (5%), pulmonary embolism in 6 of 221 (2.7%), and renal toxicity in 4 of 221 (1.8%). One patient with extensive glioma expired within 48 hours after treatment.
   CONCLUSIONS. Using standard guidelines and protocols, intraarterial chemotherapy with or without osmotic opening of the BBB is feasible across multiple centers with a low incidence of carherer-related complications. Tn patients with chemotherapy-sensitive tumors, such as PCNSL, PNET, germ cell tumor, and cancer metastasis to the central nervous system, enhanced delivery results in a high degree of tumor response, with an efficacy profile that is reproducible across multiple centers. (C) 2000 American Cancer Society.
C1 Oregon Hlth Sci Univ, Dept Neurol, Portland, OR 97201 USA.
   Ohio State Univ, Div Neurosurg, Columbus, OH 43210 USA.
   Univ Minnesota, Dept Neurosurg, Minneapolis, MN USA.
   Sharett Inst Oncol, Neurooncol Clin, Jerusalem, Israel.
   Trinity Lutheran Hosp, Kansas City, MO USA.
   Oregon Hlth Sci Univ, Dept Pharmacol, Portland, OR 97201 USA.
   Oregon Hlth Sci Univ, Div Med Informat & Outcomes Res, Portland, OR 97201 USA.
   Oregon Hlth Sci Univ, Dept Anat Pathol, Portland, OR 97201 USA.
C3 Oregon Health & Science University; University System of Ohio; Ohio
   State University; University of Minnesota System; University of
   Minnesota Twin Cities; Oregon Health & Science University; Oregon Health
   & Science University; Oregon Health & Science University
RP Doolittle, ND (corresponding author), Oregon Hlth Sci Univ, Dept Neurol, 3181 SW Sam Jackson Pk Rd L603, Portland, OR 97201 USA.
FU NCI NIH HHS [R01 CA 31770] Funding Source: Medline; NINDS NIH HHS [R01
   NS 34608, R01 NS 33618] Funding Source: Medline
CR Assietti R, 1996, J NEURO-ONCOL, V27, P251, DOI 10.1007/BF00165482
   Barth RF, 1997, CANCER RES, V57, P1129
   BASHIR R, 1988, J NEUROSURG, V68, P917, DOI 10.3171/jns.1988.68.6.0917
   Black KL, 1997, J NEUROSURG, V86, P603, DOI 10.3171/jns.1997.86.4.0603
   Boiardi A, 1997, J NEURO-ONCOL, V34, P179, DOI 10.1023/A:1005705624858
   CHERUKU R, 1991, CANCER, V68, P2621, DOI 10.1002/1097-0142(19911215)68:12<2621::AID-CNCR2820681218>3.0.CO;2-7
   Cloughesy TF, 1997, J NEURO-ONCOL, V35, P121, DOI 10.1023/A:1005856002264
   CROSSEN JR, 1992, NEUROSURGERY, V30, P23, DOI 10.1227/00006123-199201000-00005
   Dahlborg S A, 1996, Cancer J Sci Am, V2, P166
   Dahlborg SA, 1998, CANCER J, V4, P110
   Doolittle N D, 1998, J Neurosci Nurs, V30, P81
   DROPCHO EJ, 1992, J CLIN ONCOL, V10, P452, DOI 10.1200/JCO.1992.10.3.452
   FENSTERMACHER JD, 1977, CANCER TREAT REP, V61, P519
   FEUN LG, 1984, CANCER-AM CANCER SOC, V54, P794, DOI 10.1002/1097-0142(19840901)54:5<794::AID-CNCR2820540503>3.0.CO;2-F
   Fortin D, 1999, AM J NEURORADIOL, V20, P1794
   GUMERLOCK MK, 1992, J NEURO-ONCOL, V12, P33
   HIESIGER EM, 1995, J NEURO-ONCOL, V25, P143, DOI 10.1007/BF01057758
   Hirano Y, 1998, INT J ONCOL, V13, P537
   HOCHBERG FH, 1985, J NEUROSURG, V63, P876, DOI 10.3171/jns.1985.63.6.0876
   JEREMIC B, 1992, J CLIN ONCOL, V10, P1074, DOI 10.1200/JCO.1992.10.7.1074
   Kroll RA, 1998, NEUROSURGERY, V43, P879, DOI 10.1097/00006123-199810000-00090
   Kroll RA, 1998, NEUROSURGERY, V42, P1083, DOI 10.1097/00006123-199805000-00082
   MADAJEWICZ S, 1991, CANCER, V67, P2844, DOI 10.1002/1097-0142(19910601)67:11<2844::AID-CNCR2820671123>3.0.CO;2-K
   MAHALEY MS, 1989, J NEUROSURG, V70, P371, DOI 10.3171/jns.1989.70.3.0371
   Matsukado K, 1996, NEUROSURGERY, V39, P125, DOI 10.1097/00006123-199607000-00025
   McAllister LD, 2000, NEUROSURGERY, V46, P51, DOI 10.1093/neurosurgery/46.1.51
   MORTIMER JE, 1992, CANCER, V69, P1220
   NAKAGAWA H, 1994, SURG NEUROL, V41, P19, DOI 10.1016/0090-3019(94)90203-8
   Neuwelt E A., 1989, Implications of the Blood Brain Barrier and its Manipulation: Clinical Aspects, P195
   Neuwelt EA, 1998, CLIN CANCER RES, V4, P1549
   NEUWELT EA, 1986, NEUROSURGERY, V19, P573, DOI 10.1227/00006123-198610000-00011
   Neuwelt EA, 1998, J PHARMACOL EXP THER, V286, P77
   QUEVEDO JF, 1994, MAYO CLIN PROC, V69, P329, DOI 10.1016/S0025-6196(12)62216-2
   Remsen LG, 1995, CLIN CANCER RES, V1, P731
   ROMANGOLDSTEIN S, 1994, AM J NEURORADIOL, V15, P581
   SARIS SC, 1991, J NEUROSURG, V74, P763, DOI 10.3171/jns.1991.74.5.0763
   SHAPIRO WR, 1992, J NEUROSURG, V76, P772, DOI 10.3171/jns.1992.76.5.0772
   STEWART DJ, 1992, NEUROSURGERY, V30, P512
   STEWART DJ, 1983, CANCER RES, V43, P917
   WATNE K, 1992, NEUROSURGERY, V30, P223, DOI 10.1227/00006123-199202000-00012
   WILLIAMS PC, 1995, NEUROSURGERY, V37, P17, DOI 10.1227/00006123-199507000-00003
NR 41
TC 285
Z9 316
U1 0
U2 22
PU WILEY-LISS
PI NEW YORK
PA DIV JOHN WILEY & SONS INC, 605 THIRD AVE, NEW YORK, NY 10158-0012 USA
SN 0008-543X
J9 CANCER
JI Cancer
PD FEB 1
PY 2000
VL 88
IS 3
BP 637
EP 647
DI 10.1002/(SICI)1097-0142(20000201)88:3<637::AID-CNCR22>3.0.CO;2-Y
PG 11
WC Oncology
WE Science Citation Index Expanded (SCI-EXPANDED)
SC Oncology
GA 276JQ
UT WOS:000084874900022
PM 10649259
OA Bronze
DA 2024-06-03
ER

PT J
AU Camilleri-Broët, S
   Crinière, E
   Broët, P
   Delwail, V
   Mokhtari, K
   Moreau, A
   Kujas, M
   Raphaël, M
   Iraqi, W
   Sautès-Fridman, C
   Colombat, P
   Hoang-Xuan, K
   Martin, A
AF Camilleri-Broët, S
   Crinière, E
   Broët, P
   Delwail, V
   Mokhtari, K
   Moreau, A
   Kujas, M
   Raphaël, M
   Iraqi, W
   Sautès-Fridman, C
   Colombat, P
   Hoang-Xuan, K
   Martin, A
TI A uniform activated B-cell-like in-immunophenotype might explain the
   poor prognosis of primary central nervous system lymphomas:: analysis of
   83 cases
SO BLOOD
LA English
DT Article
ID BCL-2 PROTEIN EXPRESSION; NON-HODGKINS; GERMINAL-CENTER; CHEMOTHERAPY;
   SURVIVAL; RADIOTHERAPY; PROFILE; CLASSIFICATION; PREDICTION; DERIVATION
AB Most primary central nervous system lymphomas (PCNSLs) in immunocompetent patients are diffuse large B-cell lymphomas (DLBCLs), characterized by poor prognosis, compared with systemic forms. A germinal center B-cell-like (GCB) origin of PCNSL was hypothesized on the basis of BCL-6 expression and ongoing mutational activity. Our goal herein was to determine, for 83 PCNSLs, the percentages of GCB and activated B-cell-like (ABC) phenotypes and their prognostic significance. CD10, BCL-6, MUM1, BCL-2, and CD138 antigens were immunohistochemically labeled on paraffin-embedded sections; the first 4 were positive in 2.4%, 55.5%, 92.6%, and 55.5% of the tumors, respectively. None of the 56 tested samples expressed CD138. Among the 82 patients with complete information, 79 (96.3%) were classified as ABC; 42 (51.2%) expressed BCL-6(+)MUM1(+), suggesting an '' activated GCB '' origin; 33 (40.2%) were exclusively MUM1(+), and the remaining 4 (4.9%) were negative for all markers tested. These findings provide new insights into interpreting the poor PCNSL prognostic, which may, in part, be due to biologic aggressiveness associated with its activated B-cell-like pattern. We postulate assigning PCNSL a histogenetic '' time-slot,'' overlapping late GC and early post-GC, that could explain the predominant ABC phenotype observed.
C1 Hop Hotel Dieu, Serv Anat Pathol, F-75181 Paris 04, France.
   Univ Paris 05, Fac Med, Paris, France.
   Grp Hosp Pitie Salpetriere, Lab Neuropathol R Escourolle, AP HP, Paris, France.
   Grp Hosp Pitie Salpetriere, Serv Neurol Mazarin, AP HP, Paris, France.
   INSERM U255, Ctr Rech Biomed Cordeliers, Paris, France.
   Univ Paris 06, INSERM U711, Grp Hosp Pitie Salpetriere, Paris, France.
   Fac Med Paris Sud, Serv Sante Publ, AP HP, Villejuif, France.
   Hop Bernard, Poitiers, France.
   Hop Hotel Dieu, Nantes, France.
   Univ Paris Sud, CHU Bicetre, Serv Hematol & Immunol Biol Cytogenet, INSERM E109, Le Kremlin Bicetre, France.
   Hop Bretonneau, Tours, France.
   Hop Avicenne, Fac Med Paris 13, AP HP, Bobigny, France.
C3 Assistance Publique Hopitaux Paris (APHP); Universite Paris Cite;
   Hopital Universitaire Hotel-Dieu - APHP; Universite Paris Cite; Sorbonne
   Universite; Assistance Publique Hopitaux Paris (APHP); Hopital
   Universitaire Pitie-Salpetriere - APHP; Sorbonne Universite; Assistance
   Publique Hopitaux Paris (APHP); Hopital Universitaire Pitie-Salpetriere
   - APHP; Institut National de la Sante et de la Recherche Medicale
   (Inserm); Sorbonne Universite; Universite Paris Cite; Assistance
   Publique Hopitaux Paris (APHP); Hopital Universitaire Pitie-Salpetriere
   - APHP; Sorbonne Universite; Institut National de la Sante et de la
   Recherche Medicale (Inserm); Assistance Publique Hopitaux Paris (APHP);
   Hopital Universitaire Paul-Brousse - APHP; Universite Paris Saclay;
   Universite de Poitiers; Nantes Universite; CHU de Nantes; Universite
   Paris Saclay; Assistance Publique Hopitaux Paris (APHP); Hopital
   Universitaire Bicetre - APHP; Institut National de la Sante et de la
   Recherche Medicale (Inserm); Universite Paris Cite; CHU Tours;
   Universite Paris 13; Assistance Publique Hopitaux Paris (APHP); Hopital
   Universitaire Avicenne - APHP
RP Hop Hotel Dieu, Serv Anat Pathol, 1 Pl Parvis Notre Dame, F-75181 Paris 04, France.
EM
RI Hoang-Xuan, Khe/O-7527-2017; sautes-fridman, catherine/JOZ-3968-2023
OI Hoang Xuan, Khe/0000-0003-4072-1999; BROET,
   PHILIPPE/0000-0001-9122-2810; sautes-fridman,
   catherine/0000-0003-1735-8722; Delwail, Vincent/0000-0003-0680-1418
CR Alizadeh AA, 2000, NATURE, V403, P503, DOI 10.1038/35000501
   Bessell EM, 2004, INT J RADIAT ONCOL, V59, P501, DOI 10.1016/j.ijrobp.2003.11.001
   Bessell EM, 2001, INT J RADIAT ONCOL, V50, P457, DOI 10.1016/S0360-3016(01)01451-1
   Braaten KM, 2003, CLIN CANCER RES, V9, P1063
   Camilleri-Broët S, 2000, MODERN PATHOL, V13, P158, DOI 10.1038/modpathol.3880030
   Camilleri-Broet S, 1998, AM J CLIN PATHOL, V110, P607, DOI 10.1093/ajcp/110.5.607
   Carbone A, 2001, BLOOD, V97, P744, DOI 10.1182/blood.V97.3.744
   Chang CC, 2004, AM J SURG PATHOL, V28, P464, DOI 10.1097/00000478-200404000-00005
   Chang CC, 2003, ARCH PATHOL LAB MED, V127, P208
   Coiffier B, 2001, CURR OPIN ONCOL, V13, P325, DOI 10.1097/00001622-200109000-00003
   Colomo L, 2003, BLOOD, V101, P78, DOI 10.1182/blood-2002-04-1286
   COX DR, 1972, J R STAT SOC B, V34, P187
   DeAngelis LM, 2002, J CLIN ONCOL, V20, P4643, DOI 10.1200/JCO.2002.11.013
   Gascoyne RD, 1997, BLOOD, V90, P244
   Hans CP, 2004, BLOOD, V103, P275, DOI 10.1182/blood-2003-05-1545
   Harada K, 2001, CANCER GENET CYTOGEN, V125, P147, DOI 10.1016/S0165-4608(00)00377-0
   Hedvat CV, 2002, HUM PATHOL, V33, P968, DOI 10.1053/hupa.2002.127438
   Hermine O, 1996, BLOOD, V87, P265, DOI 10.1182/blood.V87.1.265.bloodjournal871265
   Hoang-Xuan K, 2004, CURR OPIN ONCOL, V16, P601, DOI 10.1097/01.cco.0000142927.88773.7e
   JAFFE ES, 2002, WHO CLASSIFICATION T, P171
   Kadan-Lottick NS, 2002, CANCER, V95, P193, DOI 10.1002/cncr.10643
   KAPLAN EL, 1958, J AM STAT ASSOC, V53, P457, DOI 10.2307/2281868
   Larocca LM, 1998, BLOOD, V92, P1011
   Lossos IS, 2004, NEW ENGL J MED, V350, P1828, DOI 10.1056/NEJMoa032520
   Montesinos-Rongen M, 1999, AM J PATHOL, V155, P2077, DOI 10.1016/S0002-9440(10)65526-5
   Montesinos-Rongen M, 2004, BLOOD, V103, P1869, DOI 10.1182/blood-2003-05-1465
   Mounier N, 2003, BLOOD, V101, P4279, DOI 10.1182/blood-2002-11-3442
   Niu HF, 2002, HEMATOL ONCOL, V20, P155, DOI 10.1002/hon.689
   Paulus W, 1999, J NEURO-ONCOL, V43, P203, DOI 10.1023/A:1006242116122
   Rickert CH, 1999, AM J PATHOL, V155, P1445, DOI 10.1016/S0002-9440(10)65458-2
   Rosenwald A, 2002, NEW ENGL J MED, V346, P1937, DOI 10.1056/NEJMoa012914
   Sáez AI, 2004, AM J PATHOL, V164, P613, DOI 10.1016/S0002-9440(10)63150-1
   Shipp MA, 2002, NAT MED, V8, P68, DOI 10.1038/nm0102-68
   Thompsett AR, 1999, BLOOD, V94, P1738, DOI 10.1182/blood.V94.5.1738.417k10_1738_1746
NR 34
TC 283
Z9 305
U1 0
U2 3
PU AMER SOC HEMATOLOGY
PI WASHINGTON
PA 2021 L ST NW, SUITE 900, WASHINGTON, DC 20036 USA
SN 0006-4971
EI 1528-0020
J9 BLOOD
JI Blood
PD JAN 1
PY 2006
VL 107
IS 1
BP 190
EP 196
DI 10.1182/blood-2005-03-1024
PG 7
WC Hematology
WE Science Citation Index Expanded (SCI-EXPANDED)
SC Hematology
GA 997HD
UT WOS:000234235200037
PM 16150948
OA Bronze
DA 2024-06-03
ER

PT J
AU Grommes, C
   Pastore, A
   Palaskas, N
   Tang, SS
   Campos, C
   Schartz, D
   Codega, P
   Nichol, D
   Clark, O
   Hsieh, WY
   Rohle, D
   Rosenblum, M
   Viale, A
   Tabar, VS
   Brennan, CW
   Gavrilovic, IT
   Kaley, TJ
   Nolan, CP
   Omuro, A
   Pentsova, E
   Thomas, AA
   Tsyvkin, E
   Noy, A
   Palomba, ML
   Hamlin, P
   Sauter, CS
   Moskowitz, CH
   Wolfe, J
   Dogan, A
   Won, M
   Glass, J
   Peak, S
   Lallana, EC
   Hatzoglou, V
   Reiner, AS
   Gutin, PH
   Huse, JT
   Panageas, KS
   Graeber, TG
   Schultz, N
   DeAngelis, LM
   Mellinghoff, IK
AF Grommes, Christian
   Pastore, Alessandro
   Palaskas, Nicolaos
   Tang, Sarah S.
   Campos, Carl
   Schartz, Derrek
   Codega, Paolo
   Nichol, Donna
   Clark, Owen
   Hsieh, Wan-Ying
   Rohle, Dan
   Rosenblum, Marc
   Viale, Agnes
   Tabar, Viviane S.
   Brennan, Cameron W.
   Gavrilovic, Igor T.
   Kaley, Thomas J.
   Nolan, Craig P.
   Omuro, Antonio
   Pentsova, Elena
   Thomas, Alissa A.
   Tsyvkin, Elina
   Noy, Ariela
   Palomba, M. Lia
   Hamlin, Paul
   Sauter, Craig S.
   Moskowitz, Craig H.
   Wolfe, Julia
   Dogan, Ahmet
   Won, Minhee
   Glass, Jon
   Peak, Scott
   Lallana, Enrico C.
   Hatzoglou, Vaios
   Reiner, Anne S.
   Gutin, Philip H.
   Huse, Jason T.
   Panageas, Katherine S.
   Graeber, Thomas G.
   Schultz, Nikolaus
   DeAngelis, Lisa M.
   Mellinghoff, Ingo K.
TI Ibrutinib Unmasks Critical Role of Bruton Tyrosine Kinase in Primary CNS
   Lymphoma
SO CANCER DISCOVERY
LA English
DT Article
ID B-CELL LYMPHOMA; NF-KAPPA-B; GENE-EXPRESSION; TARGETING BTK; CANCER;
   MUTATIONS; PI3K; GENOME; HETEROGENEITY; HYBRIDIZATION
AB Bruton tyrosine kinase (BTK) links the B-cell antigen receptor (BCR) and Toll-like receptors with NF-kappa B. The role of BTK in primary central nervous system (CNS) lymphoma (PCNSL) is unknown. We performed a phase I clinical trial with ibrutinib, the first-in-class BTK inhibitor, for patients with relapsed or refractory CNS lymphoma. Clinical responses to ibrutinib occurred in 10 of 13 (77%) patients with PCNSL, including five complete responses. The only PCNSL with complete ibrutinib resistance harbored a mutation within the coiled-coil domain of CARD11, a known ibrutinib resistance mechanism. Incomplete tumor responses were associated with mutations in the B-cell antigen receptor-associated protein CD79B. CD79B-mutant PCNSLs showed enrichment of mammalian target of rapamycin (mTOR)-related gene sets and increased staining with PI3K/mTOR activation markers. Inhibition of the PI3K isoforms p110a/p110d or mTOR synergized with ibrutinib to induce cell death in CD79B-mutant PCNSL cells.
   SIGNIFICANCE: Ibrutinib has substantial activity in patients with relapsed or refractory B-cell lymphoma of the CNS. Response rates in PCNSL were considerably higher than reported for diffuse large B-cell lymphoma outside the CNS, suggesting a divergent molecular pathogenesis. Combined inhibition of BTK and PI3K/mTOR may augment the ibrutinib response in CD79B-mutant human PCNSLs. (C) 2017 AACR.
C1 [Grommes, Christian; Gavrilovic, Igor T.; Kaley, Thomas J.; Nolan, Craig P.; Omuro, Antonio; Pentsova, Elena; Thomas, Alissa A.; Wolfe, Julia; DeAngelis, Lisa M.; Mellinghoff, Ingo K.] Mem Sloan Kettering Canc Ctr, Dept Neurol, 1275 York Ave, New York, NY 10021 USA.
   [Grommes, Christian; Palaskas, Nicolaos; Tang, Sarah S.; Campos, Carl; Schartz, Derrek; Codega, Paolo; Nichol, Donna; Clark, Owen; Hsieh, Wan-Ying; Rohle, Dan; Huse, Jason T.; Schultz, Nikolaus; Mellinghoff, Ingo K.] Mem Sloan Kettering Canc Ctr, Human Oncol & Pathogenesis Program, 1275 York Ave, New York, NY 10021 USA.
   [Grommes, Christian; Gavrilovic, Igor T.; Kaley, Thomas J.; Nolan, Craig P.; Omuro, Antonio; Pentsova, Elena; DeAngelis, Lisa M.; Mellinghoff, Ingo K.] Weill Cornell Med Coll, Dept Neurol, New York, NY USA.
   [Pastore, Alessandro] Mem Sloan Kettering Canc Ctr, Dept Computat Biol Program, 1275 York Ave, New York, NY 10021 USA.
   [Rosenblum, Marc; Dogan, Ahmet; Huse, Jason T.] Mem Sloan Kettering Canc Ctr, Dept Pathol, 1275 York Ave, New York, NY 10021 USA.
   [Viale, Agnes; Schultz, Nikolaus] Mem Sloan Kettering Canc Ctr, Marie Josee & Henry R Kravis Ctr Mol Oncol, 1275 York Ave, New York, NY 10021 USA.
   [Tabar, Viviane S.; Brennan, Cameron W.; Gutin, Philip H.] Mem Sloan Kettering Canc Ctr, Dept Neurosurg, 1275 York Ave, New York, NY 10021 USA.
   [Tsyvkin, Elina; Noy, Ariela; Palomba, M. Lia; Hamlin, Paul; Sauter, Craig S.; Moskowitz, Craig H.] Mem Sloan Kettering Canc Ctr, Dept Med, 1275 York Ave, New York, NY 10021 USA.
   [Tsyvkin, Elina; Noy, Ariela; Palomba, M. Lia; Hamlin, Paul; Sauter, Craig S.; Moskowitz, Craig H.] Weill Cornell Med Coll, Dept Med, New York, NY USA.
   [Won, Minhee] NRG Oncol Stat & Data Management Ctr, Philadelphia, PA USA.
   [Glass, Jon] Thomas Jefferson Univ, Dept Neurosurg, Philadelphia, PA 19107 USA.
   [Peak, Scott] Permanente Med Grp Inc, Dept Neurosurg, Sacramento, CA USA.
   [Lallana, Enrico C.] Permanente Med Grp Inc, Dept Neurooncol, Redwood City, CA USA.
   [Hatzoglou, Vaios] Mem Sloan Kettering Canc Ctr, Dept Radiol, 1275 York Ave, New York, NY 10021 USA.
   [Reiner, Anne S.; Panageas, Katherine S.; Schultz, Nikolaus] Mem Sloan Kettering Canc Ctr, Dept Epidemiol & Biostat, New York, NY 10021 USA.
   [Graeber, Thomas G.] Univ Calif Los Angeles, Dept Mol & Med Pharmacol, Crump Inst Mol Imaging, Los Angeles, CA USA.
   [Mellinghoff, Ingo K.] Weill Cornell Med Coll, Dept Pharmacol, New York, NY USA.
   [Palaskas, Nicolaos] Univ Calif Los Angeles, Div Hematol Oncol, Los Angeles, CA USA.
   [Nichol, Donna] Personal Genome Diagnost, Baltimore, MD USA.
   [Rohle, Dan] Roche, Basel, Switzerland.
   [Omuro, Antonio] Univ Miami, Dept Neurol, Miami, FL USA.
   [Thomas, Alissa A.] Univ Vermont, Dept Neurol, Burlington, VT USA.
   [Huse, Jason T.] MD Anderson Canc Ctr, Dept Pathol, Houston, TX USA.
   [Huse, Jason T.] MD Anderson Canc Ctr, Dept Translat Mol Pathol, Houston, TX USA.
C3 Memorial Sloan Kettering Cancer Center; Memorial Sloan Kettering Cancer
   Center; Cornell University; Weill Cornell Medicine; Memorial Sloan
   Kettering Cancer Center; Memorial Sloan Kettering Cancer Center;
   Memorial Sloan Kettering Cancer Center; Memorial Sloan Kettering Cancer
   Center; Memorial Sloan Kettering Cancer Center; Cornell University;
   Weill Cornell Medicine; Jefferson University; Permanente Medical Groups;
   Permanente Medical Groups; Memorial Sloan Kettering Cancer Center;
   Memorial Sloan Kettering Cancer Center; University of California System;
   University of California Los Angeles; Cornell University; Weill Cornell
   Medicine; University of California System; University of California Los
   Angeles; Roche Holding; University of Miami; University of Vermont;
   University of Texas System; UTMD Anderson Cancer Center; University of
   Texas System; UTMD Anderson Cancer Center
RP DeAngelis, LM (corresponding author), Mem Sloan Kettering Canc Ctr, Dept Neurol, 1275 York Ave, New York, NY 10021 USA.; Schultz, N (corresponding author), Mem Sloan Kettering Canc Ctr, Human Oncol & Pathogenesis Program, 1275 York Ave, New York, NY 10021 USA.; DeAngelis, LM (corresponding author), Weill Cornell Med Coll, Dept Neurol, New York, NY USA.; Schultz, N (corresponding author), Mem Sloan Kettering Canc Ctr, Marie Josee & Henry R Kravis Ctr Mol Oncol, 1275 York Ave, New York, NY 10021 USA.; Schultz, N (corresponding author), Mem Sloan Kettering Canc Ctr, Dept Epidemiol & Biostat, New York, NY 10021 USA.; Mellinghoff, IK (corresponding author), Mem Sloan Kettering Canc Ctr, 1275 York Ave, New York, NY 10065 USA.
EM
RI Doğan, Ahmet/JVD-6557-2023; Schultz, Nikolaus/ABE-6347-2020; Brennan,
   Cameron/IWU-9193-2023; Jones, Steven/JTV-4458-2023; Brennan,
   Cameron/R-7082-2019
OI Brennan, Cameron/0000-0003-4064-8891; PALASKAS,
   NICOLAOS/0000-0003-0783-0683; Reiner, Anne/0000-0002-1258-6112;
   DeAngelis, Lisa/0000-0002-6290-7818; Hatzoglou,
   Vaios/0000-0003-0252-0694; Mellinghoff, Ingo/0000-0002-4347-8149;
   Hamlin, Paul/0000-0001-9500-1804
FU NIH [1R01NS080944-01, P30-CA008748]; National Brain Tumor Society;
   Geoffrey Beene Cancer Research Center; Memorial Sloan Kettering Brain
   Tumor Center; Society of Memorial Sloan Kettering Cancer Center;
   American Brain Tumor Association Basic Research Fellowship Award;
   Lymphoma Research Foundation Career Development Award; Susan and Peter
   Solomon Divisional Fund; Cycle for Survival Equinox Innovation Award;
   MSK SPORE in Lymphoma Developmental Research Program (DRP) award [5 P50
   CA 192937-02]; Mildred-Scheel Postdoctoral Research Fellowship of the
   Deutsche Krebshilfe e.V. [111354]; Robertson Foundation; Radiation
   Therapy Oncology Group (RTOG) translational research program grant (TRP)
   [176]
FX This research was supported by grants from the NIH (1R01NS080944-01 to
   I.K. Mellinghoff, P30-CA008748), the National Brain Tumor Society (I.K.
   Mellinghoff), the Geoffrey Beene Cancer Research Center (I. K.
   Mellinghoff), the Memorial Sloan Kettering Brain Tumor Center (C.
   Grommes), the Society of Memorial Sloan Kettering Cancer Center (C.
   Grommes), the American Brain Tumor Association Basic Research Fellowship
   Award (C. Grommes), the Lymphoma Research Foundation Career Development
   Award (C. Grommes), Susan and Peter Solomon Divisional Fund (C.
   Grommes), Cycle for Survival Equinox Innovation Award (C. Grommes), an
   MSK SPORE in Lymphoma (5 P50 CA 192937-02) Developmental Research
   Program (DRP) award (C. Grommes), a Mildred-Scheel Postdoctoral Research
   Fellowship of the Deutsche Krebshilfe e.V. (# 111354; A. Pastore), and
   the Robertson Foundation (N. Schultz). This research includes tissue
   contributed through a Radiation Therapy Oncology Group (RTOG)
   translational research program grant (TRP#176).
CR Abrey LE, 2005, J CLIN ONCOL, V23, P5034, DOI 10.1200/JCO.2005.13.524
   Advani RH, 2013, J CLIN ONCOL, V31, P88, DOI 10.1200/JCO.2012.42.7906
   Alizadeh AA, 2000, NATURE, V403, P503, DOI 10.1038/35000501
   [Anonymous], 2015, AM SOC HEMATOLOGY
   Braggio E, 2011, CLIN CANCER RES, V17, P4245, DOI 10.1158/1078-0432.CCR-11-0395
   Byrd JC, 2013, NEW ENGL J MED, V369, P32, DOI 10.1056/NEJMoa1215637
   Carnevale J, 2016, HEMATOL ONCOL CLIN N, V30, P1293, DOI 10.1016/j.hoc.2016.07.013
   Cheng DT, 2015, J MOL DIAGN, V17, P251, DOI 10.1016/j.jmoldx.2014.12.006
   Chong CR, 2013, NAT MED, V19, P1389, DOI 10.1038/nm.3388
   Cibulskis K, 2013, NAT BIOTECHNOL, V31, P213, DOI 10.1038/nbt.2514
   Compagno M, 2009, NATURE, V459, P717, DOI 10.1038/nature07968
   Davis RE, 2010, NATURE, V463, P88, DOI 10.1038/nature08638
   Ferreri AJM, 2009, HEMATOL ONCOL, V27, P61, DOI 10.1002/hon.881
   Fischer L, 2006, ANN ONCOL, V17, P1141, DOI 10.1093/annonc/mdl070
   Gay NJ, 2014, NAT REV IMMUNOL, V14, P546, DOI 10.1038/nri3713
   Gonzalez-Perez A, 2012, NUCLEIC ACIDS RES, V40, DOI 10.1093/nar/gks743
   Hans CP, 2004, BLOOD, V103, P275, DOI 10.1182/blood-2003-05-1545
   Harrow F, 2005, J IMMUNOL, V175, P6659, DOI 10.4049/jimmunol.175.10.6659
   Hendriks RW, 2014, NAT REV CANCER, V14, P219, DOI 10.1038/nrc3702
   Hoshida Y, 2008, NEW ENGL J MED, V359, P1995, DOI 10.1056/NEJMoa0804525
   Hsieh AC, 2012, NATURE, V485, P55, DOI 10.1038/nature10912
   Kato M, 2009, NATURE, V459, P712, DOI 10.1038/nature07969
   Khodabakhshi AH, 2012, ONCOTARGET, V3, P1308
   Kim JH, 2016, LEUKEMIA LYMPHOMA, V57, P1413, DOI 10.3109/10428194.2015.1113276
   Korfel A, 2016, J CLIN ONCOL, V34, P1757, DOI 10.1200/JCO.2015.64.9897
   Lawrence MS, 2013, NATURE, V499, P214, DOI 10.1038/nature12213
   Lenz G, 2008, SCIENCE, V319, P1676, DOI 10.1126/science.1153629
   Lien EC, 2017, CURR OPIN CELL BIOL, V45, P62, DOI 10.1016/j.ceb.2017.02.007
   Limon JJ, 2010, SCI SIGNAL, V3, DOI 10.1126/scisignal.3134pe25
   Lohr JG, 2012, P NATL ACAD SCI USA, V109, P3879, DOI 10.1073/pnas.1121343109
   Maira SM, 2012, MOL CANCER THER, V11, P317, DOI 10.1158/1535-7163.MCT-11-0474
   Montesinos-Rongen M, 2015, J IMMUNOL, V195, P1312, DOI 10.4049/jimmunol.1402341
   Montesinos-Rongen M, 2012, ACTA NEUROPATHOL, V124, P905, DOI 10.1007/s00401-012-1064-7
   Montesinos-Rongen M, 2011, ACTA NEUROPATHOL, V122, P791, DOI 10.1007/s00401-011-0891-2
   Morin RD, 2013, BLOOD, V122, P1256, DOI 10.1182/blood-2013-02-483727
   Nakamura T, 2016, NEUROPATH APPL NEURO, V42, P279, DOI 10.1111/nan.12259
   Nayak L, 2013, LEUKEMIA LYMPHOMA, V54, P58, DOI 10.3109/10428194.2012.698736
   Ngo VN, 2011, NATURE, V470, P115, DOI 10.1038/nature09671
   Nogai H, 2011, J CLIN ONCOL, V29, P1803, DOI 10.1200/JCO.2010.33.3252
   Okkenhaug K, 2003, NAT REV IMMUNOL, V3, P317, DOI 10.1038/nri1056
   Palaskas N, 2011, CANCER RES, V71, P5164, DOI 10.1158/0008-5472.CAN-10-4633
   Pasqualucci L, 2001, NATURE, V412, P341, DOI 10.1038/35085588
   Pasqualucci L, 2011, NAT GENET, V43, P830, DOI 10.1038/ng.892
   Paul J, 2017, CANCER CELL, V31, P64, DOI 10.1016/j.ccell.2016.12.003
   Pentsova E, 2014, J NEURO-ONCOL, V117, P161, DOI 10.1007/s11060-014-1370-0
   Spies E, 2015, J IMMUNOL, V195, P4549, DOI 10.4049/jimmunol.1501964
   Srinivasan L, 2009, CELL, V139, P573, DOI 10.1016/j.cell.2009.08.041
   Subramanian A, 2005, P NATL ACAD SCI USA, V102, P15545, DOI 10.1073/pnas.0506580102
   Thorpe LM, 2015, NAT REV CANCER, V15, P7, DOI 10.1038/nrc3860
   Treon SP, 2015, NEW ENGL J MED, V372, P1430, DOI 10.1056/NEJMoa1501548
   Vanhaesebroeck B, 2010, NAT REV MOL CELL BIO, V11, P329, DOI 10.1038/nrm2882
   Vater I, 2015, LEUKEMIA, V29, P677, DOI 10.1038/leu.2014.264
   Wang ML, 2013, NEW ENGL J MED, V369, P507, DOI 10.1056/NEJMoa1306220
   Wilson WH, 2015, NAT MED, V21, P922, DOI 10.1038/nm.3884
   Wu CL, 2016, ONCOTARGET, V7, P38180, DOI 10.18632/oncotarget.9500
   Yap TA, 2015, CURR OPIN PHARMACOL, V23, P98, DOI 10.1016/j.coph.2015.05.016
   Young RM, 2013, NAT REV DRUG DISCOV, V12, P229, DOI 10.1038/nrd3937
NR 57
TC 266
Z9 278
U1 4
U2 42
PU AMER ASSOC CANCER RESEARCH
PI PHILADELPHIA
PA 615 CHESTNUT ST, 17TH FLOOR, PHILADELPHIA, PA 19106-4404 USA
SN 2159-8274
EI 2159-8290
J9 CANCER DISCOV
JI Cancer Discov.
PD SEP
PY 2017
VL 7
IS 9
BP 1018
EP 1029
DI 10.1158/2159-8290.CD-17-0613
PG 12
WC Oncology
WE Science Citation Index Expanded (SCI-EXPANDED)
SC Oncology
GA FF5HG
UT WOS:000409013300022
PM 28619981
OA Green Submitted, Bronze, Green Accepted
HC Y
HP N
DA 2024-06-03
ER

PT J
AU Haldorsen, IS
   Espeland, A
   Larsson, EM
AF Haldorsen, I. S.
   Espeland, A.
   Larsson, E. -M.
TI Central Nervous System Lymphoma: Characteristic Findings on Traditional
   and Advanced Imaging
SO AMERICAN JOURNAL OF NEURORADIOLOGY
LA English
DT Article
ID APPARENT DIFFUSION-COEFFICIENT; PRIMARY CEREBRAL LYMPHOMA; PRIMARY CNS
   LYMPHOMA; NON-HODGKIN-LYMPHOMA; BRAIN PERFUSION; MRI FEATURES; AIDS; CT;
   DIAGNOSIS; THERAPY
AB CNS lymphoma consists of 2 major subtypes: secondary CNS involvement by systemic lymphoma and PCNSL. Contrast-enhanced MR imaging is the method of choice for detecting CNS lymphoma. In leptomeningeal CNS lymphoma, representing two-thirds of secondary CNS lymphomas, imaging typically shows leptomeningeal, subependymal, dural, or cranial nerve enhancement. Single or multiple periventricular and/or superficial contrast-enhancing lesions are characteristic of parenchymal CNS lymphoma, representing one-third of secondary CNS lymphomas and almost 100% of PCNSLs. New CT and MR imaging techniques and metabolic imaging have demonstrated characteristic findings in CNS lymphoma, aiding in its differentiation from other CNS lesions. Advanced imaging techniques may, in the future, substantially improve the diagnostic accuracy of imaging, ultimately facilitating a noninvasive method of diagnosis. Furthermore, these imaging techniques may play a pivotal role in planning targeted therapies, prognostication, and monitoring treatment response.
C1 [Haldorsen, I. S.; Espeland, A.] Haukeland Hosp, Dept Radiol, N-5021 Bergen, Norway.
   [Haldorsen, I. S.; Espeland, A.] Univ Bergen, Dept Surg Sci, Sect Radiol, Bergen, Norway.
   [Larsson, E. -M.] Univ Uppsala Hosp, Dept Radiol, Uppsala, Sweden.
C3 University of Bergen; Haukeland University Hospital; University of
   Bergen; Uppsala University; Uppsala University Hospital
RP Haldorsen, IS (corresponding author), Haukeland Hosp, Dept Radiol, Jonas Liesvei 65, N-5021 Bergen, Norway.
EM
OI Haldorsen, Ingfrid S./0000-0001-9313-7564
CR Bakshi R, 2004, FRONT BIOSCI-LANDMRK, V9, P632, DOI 10.2741/1256
   Barajas RF, 2010, AM J NEURORADIOL, V31, P60, DOI 10.3174/ajnr.A1750
   Bataille B, 2000, J NEUROSURG, V92, P261, DOI 10.3171/jns.2000.92.2.0261
   Besson C, 2001, BLOOD, V98, P2339, DOI 10.1182/blood.V98.8.2339
   Bierman P, 2005, HEMATOL ONCOL CLIN N, V19, P597, DOI 10.1016/j.hoc.2005.05.003
   Bühring U, 2001, NEUROLOGY, V57, P393, DOI 10.1212/WNL.57.3.393
   Calli C, 2006, EUR J RADIOL, V58, P394, DOI 10.1016/j.ejrad.2005.12.032
   Chang L, 1997, NEUROIMAG CLIN N AM, V7, P409
   CHANG L, 1995, RADIOLOGY, V197, P525, DOI 10.1148/radiology.197.2.7480706
   Chen W, 2008, SEMIN NUCL MED, V38, P240, DOI 10.1053/j.semnuclmed.2008.02.005
   Cianfoni A, 2007, RADIOL MED, V112, P1225, DOI 10.1007/s11547-007-0219-4
   Cote TR, 1996, J NATL CANCER I, V88, P675, DOI 10.1093/jnci/88.10.675
   Coulon A, 2002, EUR RADIOL, V12, P329, DOI 10.1007/s003300101037
   DeAngelis LM, 2005, CANCER INVEST, V23, P145, DOI 10.1081/CNV-200050458
   Diamond C, 2006, CANCER, V106, P128, DOI 10.1002/cncr.21562
   Eichler April F, 2006, Neurosurg Focus, V21, pE15
   Erdag N, 2001, AM J ROENTGENOL, V176, P1319, DOI 10.2214/ajr.176.5.1761319
   Fainardi E, 2010, ACT NEUR S, V106, P283, DOI 10.1007/978-3-211-98811-4_53
   FINE HA, 1993, ANN INTERN MED, V119, P1093, DOI 10.7326/0003-4819-119-11-199312010-00007
   Fitzsimmons A, 2005, HEMATOL ONCOL CLIN N, V19, P689, DOI 10.1016/j.hoc.2005.05.009
   FREILICH RJ, 1995, ANN NEUROL, V38, P51, DOI 10.1002/ana.410380111
   Gleissner B, 2007, J NEURO-ONCOL, V84, P107, DOI 10.1007/s11060-007-9353-z
   Gliemroth J, 2003, CLIN NEUROL NEUROSUR, V105, P78, DOI 10.1016/S0303-8467(02)00105-1
   Go John L, 2006, Neurosurg Focus, V21, pE4
   Gunduz Kaan, 2006, Neurosurg Focus, V21, pE9
   Hakyemez B, 2006, J MAGN RESON IMAGING, V24, P817, DOI 10.1002/jmri.20707
   Haldorsen IS, 2009, AM J NEURORADIOL, V30, P744, DOI 10.3174/ajnr.A1447
   Haldorsen IS, 2007, CANCER, V110, P1803, DOI 10.1002/cncr.22989
   Haldorsen IS, 2008, BMC CANCER, V8, DOI 10.1186/1471-2407-8-225
   Haldorsen IS, 2005, ACTA ONCOL, V44, P728, DOI 10.1080/02841860500256272
   Harting I, 2003, NEUROSCI LETT, V342, P163, DOI 10.1016/S0304-3940(03)00272-6
   Hartmann M, 2003, NEUROSCI LETT, V338, P119, DOI 10.1016/S0304-3940(02)01367-8
   HAYAKAWA T, 1994, J NEURO-ONCOL, V19, P197, DOI 10.1007/BF01053274
   Heiss WD, 2009, EUR J NUCL MED MOL I, V36, P105, DOI 10.1007/s00259-008-0962-3
   Hill QA, 2006, BLOOD REV, V20, P319, DOI 10.1016/j.blre.2006.02.001
   Iwamoto Fabio M, 2006, Neurosurg Focus, V21, pE5
   Kasamon YL, 2005, HEMATOL ONCOL CLIN N, V19, P665, DOI 10.1016/j.hoc.2005.05.004
   Kim HS, 2009, AM J NEURORADIOL, V30, P1574, DOI 10.3174/ajnr.A1635
   Koeller KK, 1997, RADIOGRAPHICS, V17, P1497, DOI 10.1148/radiographics.17.6.9397461
   Küker W, 2005, J NEURO-ONCOL, V72, P169, DOI 10.1007/s11060-004-3390-7
   Küker W, 2002, J NEUROL, V249, P1713, DOI 10.1007/s00415-002-0919-6
   Larsson HBW, 2009, MAGN RESON MED, V62, P1270, DOI 10.1002/mrm.22136
   Manninger SP, 2005, AM J NEURORADIOL, V26, P2290
   Mohile NA, 2007, SEMIN RADIAT ONCOL, V17, P223, DOI 10.1016/j.semradonc.2007.02.008
   Montoto S, 2005, HEMATOL ONCOL CLIN N, V19, P751, DOI 10.1016/j.hoc.2005.05.008
   Morris PG, 2009, LANCET NEUROL, V8, P581, DOI 10.1016/S1474-4422(09)70091-2
   OGAWA T, 1994, RADIOGRAPHICS, V14, P101, DOI 10.1148/radiographics.14.1.8128041
   Olson JE, 2002, CANCER, V95, P1504, DOI 10.1002/cncr.10851
   Palmedo H, 2006, EUR J NUCL MED MOL I, V33, P164, DOI 10.1007/s00259-005-1917-6
   Peng XH, 2008, INT J NANOMED, V3, P311
   ROSENFELD SS, 1992, J NUCL MED, V33, P532
   RUIZ A, 1994, AM J NEURORADIOL, V15, P1885
   Schlegel U, 2000, J NEUROL SCI, V181, P1, DOI 10.1016/S0022-510X(00)00385-3
   Schroeder PC, 2006, NEURORADIOLOGY, V48, P715, DOI 10.1007/s00234-006-0123-y
   SENOCAK E, 2010, EUR J RADIOL    0302
   Shinoda J, 2003, J NEURO-ONCOL, V61, P261, DOI 10.1023/A:1022583610812
   Stadnik TW, 2001, AM J NEURORADIOL, V22, P969
   Taillibert S, 2008, J NEURO-ONCOL, V86, P225, DOI 10.1007/s11060-007-9468-2
   Thurnher MM, 2001, NEURORADIOLOGY, V43, P29, DOI 10.1007/s002340000480
   Toh CH, 2008, AM J NEURORADIOL, V29, P471, DOI 10.3174/ajnr.A0872
   van der Sanden GAC, 2002, CANCER, V94, P1548, DOI 10.1002/cncr.10357
   van Oostenbrugge RJ, 1999, NEUROLOGY, V53, P382, DOI 10.1212/WNL.53.2.382
   Weaver JD, 2007, NEUROLOGIST, V13, P150, DOI 10.1097/01.nrl.0000254706.85609.95
   Wong ET, 2005, CLIN CANCER RES, V11, p7151S, DOI 10.1158/1078-0432.CCR-1004-0002
   Wu O, 2005, NEUROIMAG CLIN N AM, V15, P623, DOI 10.1016/j.nic.2005.08.009
   Zacharia TT, 2008, J NEUROIMAGING, V18, P411, DOI 10.1111/j.1552-6569.2007.00231.x
NR 66
TC 265
Z9 293
U1 0
U2 19
PU AMER SOC NEURORADIOLOGY
PI DENVILLE
PA PO BOX 3000, DENVILLE, NJ 07834-9349 USA
SN 0195-6108
EI 1936-959X
J9 AM J NEURORADIOL
JI Am. J. Neuroradiol.
PD JUN-JUL
PY 2011
VL 32
IS 6
BP 984
EP 992
DI 10.3174/ajnr.A2171
PG 9
WC Clinical Neurology; Neuroimaging; Radiology, Nuclear Medicine & Medical
   Imaging
WE Science Citation Index Expanded (SCI-EXPANDED)
SC Neurosciences & Neurology; Radiology, Nuclear Medicine & Medical Imaging
GA 783FJ
UT WOS:000292066600005
PM 20616176
OA Green Published, hybrid
DA 2024-06-03
ER

PT J
AU Batchelor, T
   Loeffler, JS
AF Batchelor, T
   Loeffler, JS
TI Primary CNS lymphoma
SO JOURNAL OF CLINICAL ONCOLOGY
LA English
DT Review
ID CENTRAL-NERVOUS-SYSTEM; NON-HODGKINS-LYMPHOMA; HIGH-DOSE METHOTREXATE;
   STEM-CELL RESCUE; MALIGNANT-LYMPHOMA; PHASE-II; DEFERRED RADIOTHERAPY;
   SALVAGE CHEMOTHERAPY; BRAIN LYMPHOMA; SURVIVAL
AB Primary CNS lymphoma (PCNSL), an uncommon form of extranodal non-Hodgkin's lymphoma (NHL), has increased in incidence during the last three decades and occurs in both immunocompromised and immunocompetent hosts. PCNSL in immunocompetent patients is associated with unique diagnostic, prognostic, and therapeutic issues, and the management of this malignancy is different from that of other forms of extranodal NHL. Characteristic imaging features should be suggestive of the diagnosis, avoidance of corticosteroids, if possible, and early neurosurgical consultation for stereotactic biopsy. Because PCNSL may involve the brain, CSF, and eyes, diagnostic evaluation should include assessment of all of these regions as well as screening for possible occult systemic disease. Resection provides no therapeutic benefit and should be reserved for the rare patient with neurologic deterioration due to brain herniation. Whole-brain radiation therapy (WBRT) alone is insufficient for durable tumor control and is associated with a high risk of neurotoxicity in patients older than age 60. Neurotoxicity typically is associated with significant cognitive, motor, and autonomic dysfunction, and has a negative impact on quality of life. Chemotherapy and WBRT together improve tumor response rates and survival compared with WBRT alone. Methotrexate-based multiagent chemotherapy without WBRT is associated with similar tumor response rates and survival compared with regimens that include WBRT, although controlled trials have not been performed. The risk of neurotoxicity is lower in patients treated with chemotherapy alone.
C1 Massachusetts Gen Hosp, Dept Neurol, Div Neurooncol, Stephen E & Catherine Pappas Ctr Neurooncol, Boston, MA 02114 USA.
   Massachusetts Gen Hosp, Dept Radiat Oncol, Boston, MA 02114 USA.
   Harvard Univ, Sch Med, Boston, MA USA.
C3 Harvard University; Massachusetts General Hospital; Harvard University;
   Massachusetts General Hospital; Harvard University; Harvard Medical
   School
RP Batchelor, T (corresponding author), Massachusetts Gen Hosp, Dept Neurol, Div Neurooncol, Stephen E & Catherine Pappas Ctr Neurooncol, Yawkey 9E,55 Fruit St, Boston, MA 02114 USA.
EM
CR Abrey LE, 2003, J CLIN ONCOL, V21, P4151, DOI 10.1200/JCO.2003.05.024
   Abrey LE, 2005, J CLIN ONCOL, V23, P5034, DOI 10.1200/JCO.2005.13.524
   Abrey LE, 1998, J CLIN ONCOL, V16, P859, DOI 10.1200/JCO.1998.16.3.859
   ATLAS SW, 2002, MAGNETIC RESONANCE I, P565
   BALMACEDA C, 1995, ANN NEUROL, V38, P202, DOI 10.1002/ana.410380212
   Bataille B, 2000, J NEUROSURG, V92, P261, DOI 10.3171/jns.2000.92.2.0261
   Batchelor T, 2003, J CLIN ONCOL, V21, P1044, DOI 10.1200/JCO.2003.03.036
   BATCHELOR TT, 2004, LYMPHOMA NERVOUS SYS, P29
   Bellinzona M, 2005, EJSO, V31, P100, DOI 10.1016/j.ejso.2004.10.002
   Blay JY, 1998, J CLIN ONCOL, V16, P864, DOI 10.1200/JCO.1998.16.3.864
   Braaten KM, 2003, CLIN CANCER RES, V9, P1063
   *CENTR BRAIN TUM R, 2002, PRIM BRAIN TUM US 19
   Chan WC, 1997, BLOOD, V89, P3909
   Correa DD, 2004, NEUROLOGY, V62, P548, DOI 10.1212/01.WNL.0000109673.75316.D8
   Cote TR, 1996, J NATL CANCER I, V88, P675, DOI 10.1093/jnci/88.10.675
   DEANGELIS LM, 1990, NEUROLOGY, V40, P80, DOI 10.1212/WNL.40.1.80
   DeAngelis LM, 2002, J CLIN ONCOL, V20, P4643, DOI 10.1200/JCO.2002.11.013
   EBY NL, 1988, CANCER-AM CANCER SOC, V62, P2461, DOI 10.1002/1097-0142(19881201)62:11<2461::AID-CNCR2820621135>3.0.CO;2-M
   Enting RH, 2004, NEUROLOGY, V63, P901, DOI 10.1212/01.WNL.0000137050.43114.42
   Ferreri AJM, 1996, CANCER, V77, P827, DOI 10.1002/(SICI)1097-0142(19960301)77:5<827::AID-CNCR4>3.0.CO;2-B
   Ferreri AJM, 2003, J CLIN ONCOL, V21, P2407, DOI 10.1200/JCO.2003.01.135
   Ferreri AJM, 2003, J CLIN ONCOL, V21, P266, DOI 10.1200/JCO.2003.09.139
   Ferreri AJM, 2004, BRIT J HAEMATOL, V126, P657, DOI 10.1111/j.1365-2141.2004.05109.x
   Ferreri AJM, 2004, BRIT J CANCER, V90, P353, DOI 10.1038/sj.bjc.6601472
   FINE HA, 1993, ANN INTERN MED, V119, P1093, DOI 10.7326/0003-4819-119-11-199312010-00007
   Fitzsimmons A, 2005, HEMATOL ONCOL CLIN N, V19, P689, DOI 10.1016/j.hoc.2005.05.009
   Fliessbach K, 2005, NEUROLOGY, V64, P1184, DOI 10.1212/01.WNL.0000156350.49336.E2
   HENRY JM, 1974, CANCER, V34, P1293, DOI 10.1002/1097-0142(197410)34:4<1293::AID-CNCR2820340441>3.0.CO;2-P
   Henson JW, 2004, LYMPHOMA NERVOUS SYS, P183
   Herrlinger U, 2000, NEUROLOGY, V54, P1707, DOI 10.1212/WNL.54.8.1707
   Kadan-Lottick NS, 2002, CANCER, V95, P193, DOI 10.1002/cncr.10643
   Küker W, 2005, J NEURO-ONCOL, V72, P169, DOI 10.1007/s11060-004-3390-7
   Lai R, 2004, NEUROLOGY, V62, P451, DOI 10.1212/01.WNL.0000106941.51340.A2
   LOEFFLER JS, 1985, J CLIN ONCOL, V3, P490, DOI 10.1200/JCO.1985.3.4.490
   Lossos IS, 2001, BLOOD, V98, P945, DOI 10.1182/blood.V98.4.945
   MILLER DC, 1994, CANCER-AM CANCER SOC, V74, P1383, DOI 10.1002/1097-0142(19940815)74:4<1383::AID-CNCR2820740432>3.0.CO;2-1
   MURRAY K, 1986, J NEUROSURG, V65, P600, DOI 10.3171/jns.1986.65.5.0600
   NELSON DF, 1992, INT J RADIAT ONCOL, V23, P9, DOI 10.1016/0360-3016(92)90538-S
   Neuwelt EA, 2005, AM J NEURORADIOL, V26, P258
   Nguyen PL, 2005, J CLIN ONCOL, V23, P1507, DOI 10.1200/JCO.2005.01.161
   ONEILL BP, 1995, J NEURO-ONCOL, V25, P67, DOI 10.1007/BF01054724
   Pels H, 2003, J CLIN ONCOL, V21, P4489, DOI 10.1200/JCO.2003.04.056
   Plotkin SR, 2004, CLIN CANCER RES, V10, P5643, DOI 10.1158/1078-0432.CCR-04-0159
   Poortmans PMP, 2003, J CLIN ONCOL, V21, P4483, DOI 10.1200/JCO.2003.03.108
   Reni M, 2004, EUR J CANCER, V40, P1682, DOI 10.1016/j.ejca.2004.03.008
   Schabet M, 1999, J NEURO-ONCOL, V43, P199, DOI 10.1023/A:1006290032052
   SHAPIRO WR, 1975, NEW ENGL J MED, V293, P161, DOI 10.1056/NEJM197507242930402
   Shibamoto Y, 2005, INT J RADIAT ONCOL, V62, P809, DOI 10.1016/j.ijrobp.2004.12.043
   Soussain C, 2001, J CLIN ONCOL, V19, P742, DOI 10.1200/JCO.2001.19.3.742
   Takeshita M, 2000, AM J SURG PATHOL, V24, P1641, DOI 10.1097/00000478-200012000-00008
   Voloschin AD, 2004, NEUROLOGY, V62, pA478
NR 51
TC 252
Z9 286
U1 0
U2 14
PU AMER SOC CLINICAL ONCOLOGY
PI ALEXANDRIA
PA 330 JOHN CARLYLE ST, STE 300, ALEXANDRIA, VA 22314 USA
SN 0732-183X
J9 J CLIN ONCOL
JI J. Clin. Oncol.
PD MAR 10
PY 2006
VL 24
IS 8
BP 1281
EP 1288
DI 10.1200/JCO.2005.04.8819
PG 8
WC Oncology
WE Science Citation Index Expanded (SCI-EXPANDED)
SC Oncology
GA 024ZS
UT WOS:000236235700008
PM 16525183
DA 2024-06-03
ER

PT J
AU Küker, W
   Nägele, T
   Korfel, A
   Heckl, S
   Thiel, E
   Bamberg, M
   Weller, M
   Herrlinger, U
AF Küker, W
   Nägele, T
   Korfel, A
   Heckl, S
   Thiel, E
   Bamberg, M
   Weller, M
   Herrlinger, U
TI Primary central nervous system lymphomas (PCNSL):: MRI features at
   presentation in 100 patients
SO JOURNAL OF NEURO-ONCOLOGY
LA English
DT Article
DE brain neoplasm; DW-MRI; lymphoma; MRI; PCNSL; spectroscopy
ID PRIMARY CNS LYMPHOMA; PRIMARY CEREBRAL LYMPHOMA; INTRAVASCULAR
   LYMPHOMATOSIS; PROTON SPECTROSCOPY; BRAIN; MACROMOLECULES;
   LEUKOENCEPHALOPATHY; RADIOTHERAPY
AB To avoid an unnecessary extend of surgery in primary central nervous system lymphoma (PCNSL), the diagnosis should be suspected after MRI.
   Pre-treatment MRI examinations of 100 immunologically competent patients with biopsy-proven PCNSL were evaluated. All patients had T2- and T1-weighted images with contrast enhancement. Diffusion-weighted MRI (DW-MRI) was available in 15, proton-MR-spectroscopy (H-1-MRS) in four patients.
   The number of lesions ranged from one (n=65 patients) to eight (n=1) with a mean of 1.7. The most frequent locations were the cerebral hemispheres (n=66), the basal ganglia (n=27) and the corpus callosum (n=24). In the 65 patients with a solitary lesion, hemispheric lesions were most frequent (n=23) followed by corpus callosum (n=18). Contrast enhancement was found in all but one patient. H-1-MRS revealed a uniformly pathologic pattern of metabolite concentrations in all patients.
   Characteristic imaging features of PCNSL are contrast-enhancing lesions with a diameter of at least 15 mm in contact with the subarachnoid space. DW-MRI and proton spectroscopy may aid in differential diagnosis.
C1 Univ Tubingen, Sch Med, Dept Neuroradiol, Tubingen, Germany.
   Univ Tubingen, Sch Med, Dept Neurol, Tubingen, Germany.
   Univ Tubingen, Sch Med, Dept Radiat Oncol, Tubingen, Germany.
   Univ Hosp Benjamin Franklin, Dept Hematol & Oncol, Berlin, Germany.
C3 Eberhard Karls University of Tubingen; Eberhard Karls University of
   Tubingen; Eberhard Karls University of Tubingen; Free University of
   Berlin; Humboldt University of Berlin; Charite Universitatsmedizin
   Berlin
RP Küker, W (corresponding author), Radcliffe Infirm, Dept Neuroradiol, Woodstock Rd, Oxford OX2 6HE, England.
EM
RI McDermott, Michael Francis/GZM-5330-2022
OI Kuker, Wilhelm/0000-0002-1739-6115; Weller, Michael/0000-0002-1748-174X
CR Ayuso-Peralta L, 2001, J NEUROL NEUROSUR PS, V71, P243, DOI 10.1136/jnnp.71.2.243
   Bailey P, 1929, ARCH SURG-CHICAGO, V18, P1359, DOI 10.1001/archsurg.1929.01140130449031
   Bühring U, 2001, NEUROLOGY, V57, P393, DOI 10.1212/WNL.57.3.393
   DeAngelis LM, 1999, J NEURO-ONCOL, V43, P249, DOI 10.1023/A:1006258619757
   Hartmann M, 2003, NEUROSCI LETT, V338, P119, DOI 10.1016/S0304-3940(02)01367-8
   Herrlinger U, 2002, ANN NEUROL, V51, P247, DOI 10.1002/ana.10102
   Herrlinger U, 2002, NEURORADIOLOGY, V44, P239, DOI 10.1007/s002340100701
   JELLINGER KA, 1992, J CANCER RES CLIN, V119, P7, DOI 10.1007/BF01209483
   Kadan-Lottick NS, 2002, CANCER, V95, P193, DOI 10.1002/cncr.10643
   Kaminogo M, 2001, NEURORADIOLOGY, V43, P353, DOI 10.1007/s002340000473
   KLOSE U, 1990, MAGN RESON MED, V14, P26, DOI 10.1002/mrm.1910140104
   Küker W, 2002, J NEUROL, V249, P1713, DOI 10.1007/s00415-002-0919-6
   Lanfermann H, 1997, ACTA RADIOL, V38, P259
   Larocca LM, 1998, BLOOD, V92, P1011
   LING SM, 1994, CANCER-AM CANCER SOC, V73, P2570, DOI 10.1002/1097-0142(19940515)73:10<2570::AID-CNCR2820731019>3.0.CO;2-1
   Mader I, 2001, BRAIN, V124, P953, DOI 10.1093/brain/124.5.953
   Moulignier A, 2003, AIDS, V17, P1111, DOI 10.1097/00002030-200305020-00032
   Naegele T, 2000, RADIOLOGY, V216, P683, DOI 10.1148/radiology.216.3.r00se27683
   Nakahara T, 1999, J NEUROL NEUROSUR PS, V67, P403, DOI 10.1136/jnnp.67.3.403
   PETERSON K, 1993, CANCER, V72, P843, DOI 10.1002/1097-0142(19930801)72:3<843::AID-CNCR2820720333>3.0.CO;2-#
   Pramstaller PP, 1999, J NEUROL, V246, P934, DOI 10.1007/s004150050485
   ROMANGOLDSTEIN SM, 1992, AM J NEURORADIOL, V13, P1207
   Seeger U, 2003, MAGNET RESON MED, V49, P19, DOI 10.1002/mrm.10332
   Seeger U, 2001, MAGN RESON MED, V45, P948, DOI 10.1002/mrm.1127
   Song DK, 2002, AM J NEURORADIOL, V23, P239
   Williams RL, 1998, AM J NEURORADIOL, V19, P427
NR 26
TC 248
Z9 278
U1 0
U2 10
PU SPRINGER
PI NEW YORK
PA 233 SPRING STREET, NEW YORK, NY 10013 USA
SN 0167-594X
J9 J NEURO-ONCOL
JI J. Neuro-Oncol.
PD APR
PY 2005
VL 72
IS 2
BP 169
EP 177
DI 10.1007/s11060-004-3390-7
PG 9
WC Oncology; Clinical Neurology
WE Science Citation Index Expanded (SCI-EXPANDED)
SC Oncology; Neurosciences & Neurology
GA 931SG
UT WOS:000229504900013
PM 15925998
DA 2024-06-03
ER

PT J
AU Soussain, C
   Hoang-Xuan, K
   Taillandier, L
   Fourme, E
   Choquet, S
   Witz, F
   Casasnovas, O
   Dupriez, B
   Souleau, B
   Taksin, AL
   Gisselbrecht, C
   Jaccard, A
   Omuro, A
   Sanson, M
   Janvier, M
   Kolb, B
   Zini, JM
   Leblond, V
AF Soussain, Carole
   Hoang-Xuan, Khe
   Taillandier, Luc
   Fourme, Emmanuelle
   Choquet, Sylvain
   Witz, Francis
   Casasnovas, Olivier
   Dupriez, Brigitte
   Souleau, Bertrand
   Taksin, Anne-Laure
   Gisselbrecht, Christian
   Jaccard, Arnaud
   Omuro, Antonio
   Sanson, Marc
   Janvier, Maud
   Kolb, Brigitte
   Zini, Jean-Marc
   Leblond, Veronique
TI Intensive chemotherapy followed by hematopoietic stem-cell rescue for
   refractory and recurrent primary CNS and intraocular lymphoma: Societe
   Francaise de Greffe de Moelle Osseuse-Therapie Cellulaire
SO JOURNAL OF CLINICAL ONCOLOGY
LA English
DT Article
ID NERVOUS-SYSTEM LYMPHOMA; HIGH-DOSE CHEMOTHERAPY; NON-HODGKINS-LYMPHOMA;
   PHASE-II; SALVAGE CHEMOTHERAPY; TRANSPLANTATION; METHOTREXATE;
   TEMOZOLOMIDE; BRAIN; PCNSL
AB Purpose
   The prognosis of relapsing primary CNS lymphoma (PCNSL) is poor. We report the results of a prospective multicenter trial of intensive chemotherapy followed by autologous hematopoietic stem-cell rescue (IC + HCR) in immunocompetent adult patients with PCNSL or intraocular lymphoma (IOL) after failure of high-dose methotrexate-based treatment.
   Patients and Methods
   Salvage treatment consisted of two cycles of high-dose cytarabine and etoposide (CYVE). Intensive chemotherapy combined thiotepa, busulfan, and cyclophosphamide. Forty-three patients (median age, 52 years; range, 23 to 65 years) were included, with relapse (n = 22), refractory disease (n = 17), or a partial response to first-line treatment (n = 4). The response to CYVE was not assessable in three cases because of treatment-related death. Twenty patients (47%) were chemosensitive to CYVE: 15 of them proceeded to IC + HCR. IC + HCR was also administered to 12 patients who did not respond to CYVE. All but one of the 27 patients who underwent IC + HCR entered complete remission.
   Results
   With a median follow-up of 36 months, the median overall survival was 18.3 months in the overall population, and 58.6 months among patients who completed IC + HCR. The respective median progression-free survival (PFS) times after IC + HCR were 11.6 and 41.1 months. The 2-year overall survival probability was 45% in the whole population and 69% among the 27 patients who received IC + HCR. The 2-year PFS probability was 43% among all the patients and 58% in the IC + HCR subpopulation.
   Conclusion IC + HCR is an effective treatment for refractory and recurrent PCNSL.
C1 [Soussain, Carole] Ctr Rene Huguenin, Serv Hematol, Dept Hematol, Biostat Unit, F-92210 St Cloud, France.
   Hop La Pitie Salpetriere, Dept Neurol, Paris, France.
   Hop La Pitie Salpetriere, Dept Hematol, Paris, France.
   Hop Lariboisiere, Dept Hematol, F-75475 Paris, France.
   Ctr Hosp Univ CHU, Dept Neurol, Nancy, France.
   Ctr Hosp Univ CHU, Dept Hematol, Nancy, France.
   CHU Dijon, Dept Hematol, Dijon, France.
   CH Lens, Dept Hematol, Lens, France.
   Hop Mil, Dept Hematol, Percy, France.
   Hop Versailles, Dept Hematol, Versailles, France.
   Hop St Louis, Dept Hematol, Paris, France.
   CHU, Dept Hematol, Limoges, France.
   CHU Reims, Dept Hematol, Reims, France.
C3 Rene Huguenin Hospital; Assistance Publique Hopitaux Paris (APHP);
   Hopital Universitaire Pitie-Salpetriere - APHP; Sorbonne Universite;
   Assistance Publique Hopitaux Paris (APHP); Hopital Universitaire
   Pitie-Salpetriere - APHP; Sorbonne Universite; Universite Paris Cite;
   Assistance Publique Hopitaux Paris (APHP); Hopital Universitaire
   Lariboisiere-Fernand-Widal - APHP; CHU de Nancy; CHU de Nancy; CHU Dijon
   Bourgogne; Centre Hospitalier de Versailles; Universite Paris Cite;
   Assistance Publique Hopitaux Paris (APHP); Hopital Universitaire
   Saint-Louis - APHP; CHU Limoges; Universite de Reims Champagne-Ardenne;
   CHU de Reims
RP Soussain, C (corresponding author), Ctr Rene Huguenin, Serv Hematol, Dept Hematol, Biostat Unit, 35 Rue Dailly, F-92210 St Cloud, France.
EM
RI choquet, sylvain/O-7952-2017; Arnaud, Jaccard/O-8819-2016; Hoang-Xuan,
   Khe/O-7527-2017
OI choquet, sylvain/0000-0002-7791-0470; Arnaud,
   Jaccard/0000-0001-7091-8253; Casasnovas,
   Rene-Olivier/0000-0002-1156-8983; Hoang Xuan, Khe/0000-0003-4072-1999
CR Abrey LE, 2003, J CLIN ONCOL, V21, P4151, DOI 10.1200/JCO.2003.05.024
   Abrey LE, 2005, J CLIN ONCOL, V23, P5034, DOI 10.1200/JCO.2005.13.524
   Arellano-Rodrigo E, 2003, EUR J HAEMATOL, V70, P219, DOI 10.1034/j.1600-0609.2003.00045.x
   Colombat P, 2006, BONE MARROW TRANSPL, V38, P417, DOI 10.1038/sj.bmt.1705452
   Correa DD, 2007, ANN ONCOL, V18, P1145, DOI 10.1093/annonc/mdl464
   Deangelis Lisa M, 2006, Hematology Am Soc Hematol Educ Program, P311, DOI 10.1182/asheducation-2006.1.311
   EGORIN MJ, 1984, CANCER TREAT REP, V68, P1265
   Enting RH, 2004, NEUROLOGY, V63, P901, DOI 10.1212/01.WNL.0000137050.43114.42
   Fischer L, 2004, NEUROLOGY, V62, P1885, DOI 10.1212/01.WNL.0000125334.16420.D4
   HASSAN M, 1992, CANCER CHEMOTH PHARM, V30, P81, DOI 10.1007/BF00686397
   HEIDEMAN RL, 1989, CANCER RES, V49, P736
   Illerhaus G, 2006, J CLIN ONCOL, V24, P3865, DOI 10.1200/JCO.2006.06.2117
   Jahnke K, 2006, J NEURO-ONCOL, V80, P159, DOI 10.1007/s11060-006-9165-6
   Montemurro M, 2007, ANN ONCOL, V18, P665, DOI 10.1093/annonc/mdl458
   Muldoon LL, 2007, J CLIN ONCOL, V25, P2295, DOI 10.1200/JCO.2006.09.9861
   Nguyen PL, 2005, J CLIN ONCOL, V23, P1507, DOI 10.1200/JCO.2005.01.161
   ONEILL BP, 1995, J NEURO-ONCOL, V25, P67, DOI 10.1007/BF01054724
   Patte C., 1990, MED PEDIATR ONCOL, V18, P406
   PHILIP T, 1995, NEW ENGL J MED, V333, P1540, DOI 10.1056/NEJM199512073332305
   Plotkin SR, 2004, CLIN CANCER RES, V10, P5643, DOI 10.1158/1078-0432.CCR-04-0159
   Reni M, 2004, EUR J CANCER, V40, P1682, DOI 10.1016/j.ejca.2004.03.008
   Smith GA, 1997, J CLIN ONCOL, V15, P833, DOI 10.1200/JCO.1997.15.2.833
   Soussain C, 2001, J CLIN ONCOL, V19, P742, DOI 10.1200/JCO.2001.19.3.742
   SOUSSAIN C, 1995, BLOOD, V85, P664, DOI 10.1182/blood.V85.3.664.bloodjournal853664
   Tyson RM, 2003, LEUKEMIA LYMPHOMA, V44, P627, DOI 10.1080/1042819021000055057
   VASSAL G, 1989, CANCER CHEMOTH PHARM, V24, P386
   Wong ET, 2004, CANCER-AM CANCER SOC, V101, P139, DOI 10.1002/cncr.20339
NR 27
TC 238
Z9 242
U1 0
U2 2
PU AMER SOC CLINICAL ONCOLOGY
PI ALEXANDRIA
PA 2318 MILL ROAD, STE 800, ALEXANDRIA, VA 22314 USA
SN 0732-183X
EI 1527-7755
J9 J CLIN ONCOL
JI J. Clin. Oncol.
PD MAY 20
PY 2008
VL 26
IS 15
BP 2512
EP 2518
DI 10.1200/JCO.2007.13.5533
PG 7
WC Oncology
WE Science Citation Index Expanded (SCI-EXPANDED)
SC Oncology
GA 302LR
UT WOS:000255970300019
PM 18413641
OA Bronze
DA 2024-06-03
ER

PT J
AU Baraniskin, A
   Kuhnhenn, J
   Schlegel, U
   Chan, A
   Deckert, M
   Gold, R
   Maghnouj, A
   Zöllner, H
   Reinacher-Schick, A
   Schmiegel, W
   Hahn, SA
   Schroers, R
AF Baraniskin, Alexander
   Kuhnhenn, Jan
   Schlegel, Uwe
   Chan, Andrew
   Deckert, Martina
   Gold, Ralf
   Maghnouj, Abdelouahid
   Zoellner, Hannah
   Reinacher-Schick, Anke
   Schmiegel, Wolff
   Hahn, Stephan A.
   Schroers, Roland
TI Identification of microRNAs in the cerebrospinal fluid as marker for
   primary diffuse large B-cell lymphoma of the central nervous system
SO BLOOD
LA English
DT Article
ID PRIMARY CNS LYMPHOMA; MENINGEAL DISSEMINATION; EXPRESSION; CANCER;
   DIAGNOSIS; BIOMARKER; SURVIVAL; DISEASE; SERUM; CSF
AB The diagnosis of primary central nervous system lymphoma (PCNSL) depends on histopathology of brain biopsies, because disease markers in the cerebrospinal fluid (CSF) with sufficient diagnostic accuracy are not available yet. MicroRNAs (miRNAs) are regulatory RNA molecules that are deregulated in many disease types, including cancer. Recently, miRNAs have shown promise as markers for cancer diagnosis. In this study, we demonstrate that miRNAs are present in the CSF of patients with PCNSL. With a candidate approach and miRNA quantification by reverse transcription polymerase chain reaction, miRNAs with significant levels in the CSF of patients with PCNSL were identified. MiR-21, miR-19, and miR-92a levels in CSF collected from patients with PCNSL and from controls with inflammatory CNS disorders and other neurologic disorders indicated a significant diagnostic value of this method. Receiver-operating characteristic analyses showed area under the curves of 0.94, 0.98, and 0.97, respectively, for miR-21, miR-19, and miR-92a CSF levels in discriminating PCNSL from controls. More importantly, combined miRNA analyses resulted in an increased diagnostic accuracy with 95.7% sensitivity and 96.7% specificity. We also demonstrated a remarkable stability of miRNAs in the CSF. In conclusion, CSF miRNAs are potentially useful tools as novel noninvasive biomarker for the diagnosis of PCNSL. (Blood. 2011; 117(11):3140-3146)
C1 [Baraniskin, Alexander; Reinacher-Schick, Anke; Schmiegel, Wolff; Hahn, Stephan A.; Schroers, Roland] Ruhr Univ Bochum, Dept Med Hematol & Oncol, D-44892 Bochum, Germany.
   [Baraniskin, Alexander; Maghnouj, Abdelouahid; Zoellner, Hannah; Hahn, Stephan A.] Ruhr Univ Bochum, Clin Res Ctr, D-44892 Bochum, Germany.
   [Kuhnhenn, Jan; Schlegel, Uwe; Chan, Andrew; Gold, Ralf] Ruhr Univ Bochum, Dept Neurol, D-44892 Bochum, Germany.
   [Deckert, Martina] Univ Cologne, Dept Neuropathol, D-5000 Cologne 41, Germany.
C3 Ruhr University Bochum; Ruhr University Bochum; Ruhr University Bochum;
   University of Cologne
RP Schroers, R (corresponding author), Ruhr Univ Bochum, Knappschaftskrankenhaus Bochum Langendreer, Schornau 23-25, D-44892 Bochum, Germany.
EM
RI Reinacher-Schick, Anke C/F-2722-2012; Deckert, Martina/P-2842-2016;
   Hahn, Stephan A/E-3880-2010; Hahn, Stephan A./AAV-6062-2021
OI Hahn, Stephan A/0000-0003-0855-9741; Hahn, Stephan
   A./0000-0003-0855-9741; Chan, Andrew/0000-0003-3434-7283
FU Protein Research Department (Ruhr-University); Ministry of Science,
   North Rhine-Westphalia, Germany
FX This work was supported by the Protein Research Department
   (Ruhr-University) and by a grant (PURE) from the Ministry of Science,
   North Rhine-Westphalia, Germany.
CR Abrey LE, 2005, J CLIN ONCOL, V23, P5034, DOI 10.1200/JCO.2005.13.524
   Bataille B, 2000, J NEUROSURG, V92, P261, DOI 10.3171/jns.2000.92.2.0261
   Cogswell JP, 2008, J ALZHEIMERS DIS, V14, P27, DOI 10.3233/jad-2008-14103
   Croce CM, 2009, NAT REV GENET, V10, P704, DOI 10.1038/nrg2634
   Fischer L, 2008, NEUROLOGY, V71, P1102, DOI 10.1212/01.wnl.0000326958.52546.f5
   Hu ZB, 2010, J CLIN ONCOL, V28, P1721, DOI 10.1200/JCO.2009.24.9342
   Juergens A, 2010, ANN NEUROL, V67, P182, DOI 10.1002/ana.21824
   Kersten MJ, 1996, BLOOD, V87, P1985, DOI 10.1182/blood.V87.5.1985.1985
   Kiewe P, 2010, NEURO-ONCOLOGY, V12, P409, DOI 10.1093/neuonc/nop053
   Kosaka N, 2010, CANCER SCI, V101, P2087, DOI 10.1111/j.1349-7006.2010.01650.x
   LANDIS JR, 1977, BIOMETRICS, V33, P159, DOI 10.2307/2529310
   Lawrie CH, 2008, BRIT J HAEMATOL, V141, P672, DOI 10.1111/j.1365-2141.2008.07077.x
   Lawrie CH, 2007, BRIT J HAEMATOL, V137, P503, DOI 10.1111/j.1365-2141.2007.06611.x
   Lawrie CH, 2007, INT J CANCER, V121, P1156, DOI 10.1002/ijc.22800
   Mitchell PS, 2008, P NATL ACAD SCI USA, V105, P10513, DOI 10.1073/pnas.0804549105
   Ney DE., 2010, NONHODGKIN LYMPHOMAS, P527
   Pels H, 2003, J CLIN ONCOL, V21, P4489, DOI 10.1200/JCO.2003.04.056
   Pels Hendrik, 2006, Curr Treat Options Neurol, V8, P346, DOI 10.1007/s11940-006-0024-8
   RABIN EZ, 1977, CAN J NEUROL SCI, V4, P125, DOI 10.1017/S0317167100120529
   Robertus JL, 2009, MODERN PATHOL, V22, P547, DOI 10.1038/modpathol.2009.10
   Roy S, 2008, J CLIN ONCOL, V26, P96, DOI 10.1200/JCO.2007.12.1053
   Schroers R, 2010, EUR J HAEMATOL, V85, P520, DOI 10.1111/j.1600-0609.2010.01516.x
   Schroers R, 2010, EUR J HAEMATOL, V85, P236, DOI 10.1111/j.1600-0609.2010.01475.x
   Szafranska AE, 2007, ONCOGENE, V26, P4442, DOI 10.1038/sj.onc.1210228
NR 24
TC 237
Z9 260
U1 0
U2 29
PU AMER SOC HEMATOLOGY
PI WASHINGTON
PA 1900 M STREET. NW SUITE 200, WASHINGTON, DC 20036 USA
SN 0006-4971
J9 BLOOD
JI Blood
PD MAR 17
PY 2011
VL 117
IS 11
BP 3140
EP 3146
DI 10.1182/blood-2010-09-308684
PG 7
WC Hematology
WE Science Citation Index Expanded (SCI-EXPANDED)
SC Hematology
GA 736LN
UT WOS:000288496300024
PM 21200023
DA 2024-06-03
ER

PT J
AU Omuro, A
   Correa, DD
   DeAngelis, LM
   Moskowitz, CH
   Matasar, MJ
   Kaley, TJ
   Gavrilovic, IT
   Nolan, C
   Pentsova, E
   Grommes, CC
   Panageas, KS
   Baser, RE
   Faivre, G
   Abrey, LE
   Sauter, CS
AF Omuro, Antonio
   Correa, Denise D.
   DeAngelis, Lisa M.
   Moskowitz, Craig H.
   Matasar, Matthew J.
   Kaley, Thomas J.
   Gavrilovic, Igor T.
   Nolan, Craig
   Pentsova, Elena
   Grommes, Christian C.
   Panageas, Katherine S.
   Baser, Raymond E.
   Faivre, Geraldine
   Abrey, Lauren E.
   Sauter, Craig S.
TI R-MPV followed by high-dose chemotherapy with TBC and autologous
   stem-cell transplant for newly diagnosed primary CNS lymphoma
SO BLOOD
LA English
DT Article
ID CENTRAL-NERVOUS-SYSTEM; WHOLE-BRAIN RADIOTHERAPY; SINGLE-CENTER
   EXPERIENCE; RECURRENT PRIMARY CNS; NON-HODGKINS-LYMPHOMA; INTENSIVE
   CHEMOTHERAPY; PHASE-II; INTRAOCULAR LYMPHOMA; CONSOLIDATING
   RADIOTHERAPY; DEFERRED RADIOTHERAPY
AB High-dose methotrexate-based chemotherapy is the mainstay of treatment of primary central nervous system lymphoma (PCNSL), but relapses remain frequent. High-dose chemotherapy (HDC) with autologous stem-cell transplant (ASCT) may provide an alternative to address chemoresistance and overcome the blood-brain barrier. In this single-center phase-2 study, newly diagnosed PCNSL patients received 5 to 7 cycles of chemotherapy with rituximab, methotrexate (3.5 g/m(2)), procarbazine, and vincristine (R-MPV). Those with a complete or partial response proceeded with consolidation HDC with thiotepa, cyclophosphamide, and busulfan, followed by ASCT and no radiotherapy. Primary end point was 1-year progression-free survival (PFS), N532. Median age was 57, and median Karnofsky performance status 80. Following R-MPV, objective response rate was 97%, and 26 (81%) patients proceeded with HDC-ASCT. Among all patients, median PFS and overall survival (OS) were not reached (median follow-up: 45 months). Two-year PFS was 79% (95% confidence interval [CI], 58-90), with no events observed beyond 2 years. Two-year OS was 81% (95% CI, 63-91). In transplanted patients, 2-year PFS and OS were 81%. There were 3 treatment-related deaths. Prospective neuropsychological evaluations suggested relatively stable cognitive functions posttransplant. In conclusion, this treatment was associated with excellent disease control and survival, an acceptable toxicity profile, and no evidence of neurotoxicity thus far. This trial was registered at www.clinicaltrials.gov as NCT00596154.
C1 [Omuro, Antonio; Correa, Denise D.; DeAngelis, Lisa M.; Kaley, Thomas J.; Gavrilovic, Igor T.; Nolan, Craig; Pentsova, Elena; Grommes, Christian C.; Faivre, Geraldine; Abrey, Lauren E.] Mem Sloan Kettering Canc Ctr, Dept Neurol, New York, NY 10065 USA.
   [Moskowitz, Craig H.; Matasar, Matthew J.; Sauter, Craig S.] Mem Sloan Kettering Canc Ctr, Dept Med, New York, NY 10065 USA.
   [Panageas, Katherine S.; Baser, Raymond E.] Mem Sloan Kettering Canc Ctr, Dept Epidemiol & Biostat, New York, NY 10065 USA.
C3 Memorial Sloan Kettering Cancer Center; Memorial Sloan Kettering Cancer
   Center; Memorial Sloan Kettering Cancer Center
RP Omuro, A (corresponding author), Mem Sloan Kettering Canc Ctr, Dept Neurol, 1275 York Ave, New York, NY 10065 USA.
EM
OI Kaley, Thomas/0000-0002-2540-8518; DeAngelis, Lisa/0000-0002-6290-7818;
   Matasar, Matthew/0000-0002-4581-3721; Nolan, Craig/0000-0003-3534-4615;
   Gavrilovic, Igor/0000-0001-9104-6132; Omuro, Antonio/0000-0003-4299-3664
FU Memorial Sloan Kettering Cancer Center Department of Neurology Research
   and Development Funds; National Institutes of Health, National Cancer
   Institute [P30 CA008748]
FX This work was an investigator-initiated clinical trial sponsored by
   Memorial Sloan Kettering Cancer Center Department of Neurology Research
   and Development Funds and was conducted with no industry support. K.S.P.
   received research funding from National Institutes of Health, National
   Cancer Institute (grant P30 CA008748).
CR Abrey LE, 2006, J CLIN ONCOL, V24, P5711, DOI 10.1200/JCO.2006.08.2941
   Abrey LE, 2003, J CLIN ONCOL, V21, P4151, DOI 10.1200/JCO.2003.05.024
   Abrey LE, 2005, J CLIN ONCOL, V23, P5034, DOI 10.1200/JCO.2005.13.524
   Abrey LE, 2000, J CLIN ONCOL, V18, P3144, DOI 10.1200/JCO.2000.18.17.3144
   Alimohamed N, 2012, LEUKEMIA LYMPHOMA, V53, P862, DOI 10.3109/10428194.2011.633250
   Anagnostopoulos A, 2004, CANCER-AM CANCER SOC, V100, P2607, DOI 10.1002/cncr.20294
   Batchelor T, 2003, J CLIN ONCOL, V21, P1044, DOI 10.1200/JCO.2003.03.036
   Brevet M, 2005, EUR J HAEMATOL, V75, P288, DOI 10.1111/j.1600-0609.2005.00508.x
   Bromberg JE, 2013, HAEMATOLOGICA, V98, P808, DOI 10.3324/haematol.2012.070839
   Chen YB, 2015, CANCER-AM CANCER SOC, V121, P226, DOI 10.1002/cncr.29023
   Colombat P, 2006, BONE MARROW TRANSPL, V38, P417, DOI 10.1038/sj.bmt.1705452
   Cote GM, 2012, BIOL BLOOD MARROW TR, V18, P76, DOI 10.1016/j.bbmt.2011.07.006
   DEANGELIS LM, 1992, J CLIN ONCOL, V10, P635, DOI 10.1200/JCO.1992.10.4.635
   FAZEKAS F, 1987, AM J ROENTGENOL, V149, P351, DOI 10.2214/ajr.149.2.351
   Ferreri AJM, 2009, LANCET, V374, P1512, DOI 10.1016/S0140-6736(09)61416-1
   GABBAI AA, 1989, J NEUROSURG, V70, P190, DOI 10.3171/jns.1989.70.2.0190
   GLASS J, 1994, J NEUROSURG, V81, P188, DOI 10.3171/jns.1994.81.2.0188
   Graber JJ, 2011, CURR OPIN NEUROL, V24, P633, DOI 10.1097/WCO.0b013e32834cbdef
   Hottinger AF, 2007, NEUROLOGY, V69, P1178, DOI 10.1212/01.wnl.0000276986.19602.c1
   Illerhaus G, 2012, BLOOD, V120
   Illerhaus G, 2007, HAEMATOLOGICA, V93, P147, DOI 10.3324/haematol.11771
   Illerhaus G, 2006, J CLIN ONCOL, V24, P3865, DOI 10.1200/JCO.2006.06.2117
   Montemurro M, 2007, ANN ONCOL, V18, P665, DOI 10.1093/annonc/mdl458
   Morris PG, 2013, J CLIN ONCOL, V31, P3971, DOI 10.1200/JCO.2013.50.4910
   NELSON DF, 1992, INT J RADIAT ONCOL, V23, P9, DOI 10.1016/0360-3016(92)90538-S
   Nguyen PL, 2005, J CLIN ONCOL, V23, P1507, DOI 10.1200/JCO.2005.01.161
   Omuro A, 2011, J NEURO-ONCOL, V104, P323, DOI 10.1007/s11060-010-0497-x
   Omuro AMP, 2005, ARCH NEUROL-CHICAGO, V62, P1595, DOI 10.1001/archneur.62.10.1595
   ONEILL BP, 1995, INT J RADIAT ONCOL, V33, P663, DOI 10.1016/0360-3016(95)00207-F
   Pels H, 2003, J CLIN ONCOL, V21, P4489, DOI 10.1200/JCO.2003.04.056
   Poortmans PMP, 2003, J CLIN ONCOL, V21, P4483, DOI 10.1200/JCO.2003.03.108
   PRZEPIORKA D, 1995, LEUKEMIA LYMPHOMA, V17, P427, DOI 10.3109/10428199509056853
   Rubenstein JL, 2013, J CLIN ONCOL, V31, P3061, DOI 10.1200/JCO.2012.46.9957
   Schorb E, 2013, HAEMATOLOGICA, V98, P765, DOI 10.3324/haematol.2012.076075
   Schultz C, 1996, J CLIN ONCOL, V14, P556, DOI 10.1200/JCO.1996.14.2.556
   Shimoni A, 2001, BONE MARROW TRANSPL, V27, P821, DOI 10.1038/sj.bmt.1703007
   Soussain C, 2001, J CLIN ONCOL, V19, P742, DOI 10.1200/JCO.2001.19.3.742
   Soussain C, 2008, J CLIN ONCOL, V26, P2512, DOI 10.1200/JCO.2007.13.5533
   Soussain C, 2012, HAEMATOL-HEMATOL J, V97, P1751, DOI 10.3324/haematol.2011.060434
   Thiel E, 2010, LANCET ONCOL, V11, P1036, DOI 10.1016/S1470-2045(10)70229-1
   Welch MR, 2015, LEUKEMIA LYMPHOMA, V56, P361, DOI 10.3109/10428194.2014.916800
   Yoon DH, 2011, BONE MARROW TRANSPL, V46, P105, DOI 10.1038/bmt.2010.71
NR 42
TC 223
Z9 232
U1 0
U2 12
PU AMER SOC HEMATOLOGY
PI WASHINGTON
PA 2021 L ST NW, SUITE 900, WASHINGTON, DC 20036 USA
SN 0006-4971
EI 1528-0020
J9 BLOOD
JI Blood
PD FEB 26
PY 2015
VL 125
IS 9
BP 1403
EP 1410
DI 10.1182/blood-2014-10-604561
PG 8
WC Hematology
WE Science Citation Index Expanded (SCI-EXPANDED)
SC Hematology
GA CD1FT
UT WOS:000350820900012
PM 25568347
OA Green Published, Bronze
DA 2024-06-03
ER

PT J
AU Olson, JE
   Janney, CA
   Rao, RD
   Cerhan, JR
   Kurtin, PJ
   Schiff, D
   Kaplan, RS
   O'Neill, BP
AF Olson, JE
   Janney, CA
   Rao, RD
   Cerhan, JR
   Kurtin, PJ
   Schiff, D
   Kaplan, RS
   O'Neill, BP
TI The continuing increase in the incidence of primary central nervous
   system non-Hodgkin lymphoma - A surveillance, epidemiology, and end
   results analysis
SO CANCER
LA English
DT Article; Proceedings Paper
CT 37th Annual Meeting of the American-Society-of-Clinical-Oncology
CY MAY 12-15, 2001
CL SAN FRANCISCO, CALIFORNIA
SP Amer Soc Clin Oncol
DE primary central nervous system lymphoma (PCNSL); epidemiology; brain
   tumor; Surveillance, Epidemiology, and End Results (SEER) program;
   lymphoma
ID HIGH-DOSE METHOTREXATE; PRIMARY CNS LYMPHOMA; BRAIN LYMPHOMA; SURVIVAL;
   GLIOMAS; TRENDS
AB BACKGROUND. Primary central nervous system lymphoma (PCNSL) is an extranodal form of non-Hodgkin lymphoma arising in the craniospinal axis. The incidence of PCNSL appears to be increasing.
   METHODS. PCNSL incidence data from 1973-1997 were obtained from the nine Surveillance, Epidemiology and End Results (SEER) registries. To limit the influence of the human immunodeficiency virus on incidence rates, data of never-married males and females and persons of unknown marital status were excluded. As a surrogate for new technology, SEER data were reviewed by dates of diagnosis (surrogate for imaging) and compared with glioma incidence (surrogate for stereotactic neurosurgery and improved diagnostic neuropathology). Age-adjusted incidence rates were estimated and compared for the period prior to computed tomography (CT) (1973-1984) and the magnetic resonance imaging (MRI) period (1985-1997). The estimated annual percent change was calculated based on linear regression analyses using SEER*STAT.
   RESULTS. The incidence of PCNSL appears to be increasing in all SEER registries examined. All age groups demonstrated an increase over tune. This increase was observed both in the CT era as well as in the MRI era. PCNSL age-adjusted incidence (0.15 to 0.48, a 3-fold increase) outpaced that of systemic lymphoma (14.1 to 18.5, a 33% increase) for the same registries over the same time periods. The rate of increase has begun to slow since 1985; the estimated annual percent change for PCNSL was three-fold higher during the period 1973-1985 compared with 1986-1997.
   CONCLUSION. The incidence rate of PCNSL continues to rise. The increase is evident in all age groups and in both genders. Data from the current study suggest that improved diagnostic tools, such as CT or MRI, cannot explain this increase.
C1 Mayo Clin & Mayo Fdn, Dept Neurol, Rochester, MN 55905 USA.
   Mayo Clin, Ctr Canc, Rochester, MN USA.
   Mayo Clin & Mayo Fdn, Dept Oncol, Rochester, MN 55905 USA.
   Univ Pittsburgh, Med Ctr, Dept Neurosurg, Pittsburgh, PA USA.
   Pittsburgh Canc Inst, Pittsburgh, PA 15213 USA.
   NCI, Off Trials Evaluat Program, Rockville, MD USA.
   Mayo Clin & Mayo Fdn, Dept Neurol, Rochester, MN 55905 USA.
C3 Mayo Clinic; Mayo Clinic; Mayo Clinic; Pennsylvania Commonwealth System
   of Higher Education (PCSHE); University of Pittsburgh; Pennsylvania
   Commonwealth System of Higher Education (PCSHE); University of
   Pittsburgh; Magee-Womens Research Institute; National Institutes of
   Health (NIH) - USA; NIH National Cancer Institute (NCI); Mayo Clinic
RP O'Neill, BP (corresponding author), Mayo Clin & Mayo Fdn, Dept Neurol, 200 1St St SW, Rochester, MN 55905 USA.
OI Cerhan, James/0000-0002-7482-178X; Kaplan, Richard
   S./0000-0002-0189-8348; Janney, Carol/0000-0002-4821-4679
FU NCI NIH HHS [P30 CA15030, K07 CA64220] Funding Source: Medline
CR Besson C, 2001, BLOOD, V98, P2339, DOI 10.1182/blood.V98.8.2339
   Blay JY, 1998, J CLIN ONCOL, V16, P864, DOI 10.1200/JCO.1998.16.3.864
   Corn BW, 1997, CANCER, V79, P2409
   Cote TR, 1996, J NATL CANCER I, V88, P675, DOI 10.1093/jnci/88.10.675
   EBY NL, 1988, CANCER-AM CANCER SOC, V62, P2461, DOI 10.1002/1097-0142(19881201)62:11<2461::AID-CNCR2820621135>3.0.CO;2-M
   Ferreri AJM, 1996, CANCER, V77, P827, DOI 10.1002/(SICI)1097-0142(19960301)77:5<827::AID-CNCR4>3.0.CO;2-B
   FINE HA, 1993, ANN INTERN MED, V119, P1093, DOI 10.7326/0003-4819-119-11-199312010-00007
   FINE HA, 1998, LYMPHOMAS, P481
   Fleury A, 1997, CANCER, V79, P1195, DOI 10.1002/(SICI)1097-0142(19970315)79:6<1195::AID-CNCR19>3.0.CO;2-V
   GABBAI AA, 1989, J NEUROSURG, V70, P190, DOI 10.3171/jns.1989.70.2.0190
   Garber K, 2001, J NATL CANCER I, V93, P494, DOI 10.1093/jnci/93.7.494
   Hao D, 1999, ANN ONCOL, V10, P65, DOI 10.1023/A:1008308313914
   Legler JM, 1999, JNCI-J NATL CANCER I, V91, P1382, DOI 10.1093/jnci/91.16.1382
   LUTZ JM, 1994, BRIT J CANCER, V70, P716, DOI 10.1038/bjc.1994.381
   *NCI, 2000, SEER ASTERISK STAT 3
   NEUWELT EA, 1991, J CLIN ONCOL, V9, P1580, DOI 10.1200/JCO.1991.9.9.1580
   O'Neill B P, 1999, Neuro Oncol, V1, P196, DOI 10.1093/neuonc/1.3.196
   ONEILL BP, 1995, J NEURO-ONCOL, V25, P67, DOI 10.1007/BF01054724
   Percy C., 1990, WHO INT CLASSIFICATI
   PERCY C, 1990, INT CLASSIFICATION D, P144
   RIES L, 2000, NIH PUB NCI
   WERNER MH, 1995, CANCER, V76, P1634, DOI 10.1002/1097-0142(19951101)76:9<1634::AID-CNCR2820760921>3.0.CO;2-0
NR 22
TC 214
Z9 245
U1 0
U2 6
PU JOHN WILEY & SONS INC
PI HOBOKEN
PA 111 RIVER ST, HOBOKEN, NJ 07030 USA
SN 0008-543X
J9 CANCER
JI Cancer
PD OCT 1
PY 2002
VL 95
IS 7
BP 1504
EP 1510
DI 10.1002/cncr.10851
PG 7
WC Oncology
WE Conference Proceedings Citation Index - Science (CPCI-S); Science Citation Index Expanded (SCI-EXPANDED)
SC Oncology
GA 595VE
UT WOS:000178128600013
PM 12237919
DA 2024-06-03
ER

PT J
AU Soussain, C
   Suzan, F
   Hoang-Xuan, K
   Cassoux, N
   Levy, V
   Azar, N
   Belanger, C
   Achour, E
   Ribrag, V
   Gerber, S
   Delattre, JY
   Leblond, V
AF Soussain, C
   Suzan, F
   Hoang-Xuan, K
   Cassoux, N
   Levy, V
   Azar, N
   Belanger, C
   Achour, E
   Ribrag, V
   Gerber, S
   Delattre, JY
   Leblond, V
TI Results of intensive chemotherapy followed by hematopoietic stem-cell
   rescue in 22 patients with refractory or recurrent primary CNS lymphoma
   or intraocular lymphoma
SO JOURNAL OF CLINICAL ONCOLOGY
LA English
DT Article
ID BONE-MARROW TRANSPLANTATION; HIGH-DOSE CHEMOTHERAPY; NERVOUS-SYSTEM
   LYMPHOMA; THERAPY; METHOTREXATE; RADIOTHERAPY; MALIGNANCIES; RADIATION;
   PROTOCOL; MODALITY
AB Purpose: To assess the feasibility and efficacy of intensive chemotherapy with hematopoietic stem-cell rescue (IC + HCR) in patients with refractory or recurrent primary CNS lymphoma (PCNSL) or intraocular lymphoma (IOL).
   Patients and Methods: IC consisted of thiotepa 250 mg/m(2)/d days -9 through -7, busulfan 10 mg/kg (total dose) days -6 through -4, and cyclophosphamide 60 mg/kg/d days -3 and -2. intravenous clonazepam 2 mg/d was given prophylactically from the day before initiation of busulfan therapy to the day after completion of busulfan therapy. Patients with refractory or recurrent PCNSL underwent IC + HCR only if they were chemosensitive to two cycles of salvage treatment with cytarabine (2 g/m(2)/d days 2 through 5 and 50 mg/m2/d days 1 through 5 in a 15-hour infusion) and etoposide (VP-16; 200 mg/m(2)/d days 2 through 5) (CYVE). Patients with IOL refractory to high-dose methotrexate (MTX) and cytarabine entered the IC + HCR program directly.
   Results: Twenty-two patients (10 with relapses, 12 with refractory disease) were enrolled. Twenty patients entered the IC + HCR program: twelve entered after CYVE treatment, seven entered directly, and one had previously been retreated with high-dose MTX. Before IC, eight patients were in complete remission (CR), four were in partial remission (PR), one had stable disease, and seven had refractory disease. After IC + HCR, 16 patients entered CR, two remained in PR, one had stable disease, and one had disease progression. Fourteen patients remained alive (median follow up time, 41.5 months). The overall probability of survival at 3 years was 63.7%. After IC, that probability was 60% and the 3-year probability of event-free survival was 53%. Seven patients had neurologic adverse events during the entire procedure.
   Conclusion: IC + HCR proved feasible and effective in patients with refractory or recurrent PCNSL or IOL. The entire procedure seemed to be most toxic in patients greater than or equal to 60 years. A prospective multicenter study is ongoing. (C) 2001 by American Society of Clinical Oncology.
C1 Hop Meaux, Hematol Serv, Meaux, France.
   Hop Versailles, Hematol Serv, Versailles, France.
   Hop La Pitie Salpetriere, Serv Neurooncol, Paris, France.
   Hop La Pitie Salpetriere, Serv Ophthalmol, Paris, France.
   Hop La Pitie Salpetriere, Serv Neuroradiol, Paris, France.
   Hop La Pitie Salpetriere, Hematol Serv, Paris, France.
   Hop La Pitie Salpetriere, Ctr Transfus, Paris, France.
   Hop St Louis, Dept Biostat & Informat Med, Paris, France.
   Hop Necker Enfants Malad, Hematol Serv, Paris, France.
   Hop Colmar, Hematol Serv, Colmar, France.
   Inst Gustave Roussy, Hematol Serv, Villejuif, France.
C3 Centre Hospitalier de Versailles; Sorbonne Universite; Assistance
   Publique Hopitaux Paris (APHP); Hopital Universitaire Pitie-Salpetriere
   - APHP; Sorbonne Universite; Assistance Publique Hopitaux Paris (APHP);
   Hopital Universitaire Pitie-Salpetriere - APHP; Assistance Publique
   Hopitaux Paris (APHP); Hopital Universitaire Pitie-Salpetriere - APHP;
   Sorbonne Universite; Sorbonne Universite; Assistance Publique Hopitaux
   Paris (APHP); Hopital Universitaire Pitie-Salpetriere - APHP; Sorbonne
   Universite; Assistance Publique Hopitaux Paris (APHP); Hopital
   Universitaire Pitie-Salpetriere - APHP; Universite Paris Cite;
   Assistance Publique Hopitaux Paris (APHP); Hopital Universitaire
   Saint-Louis - APHP; Assistance Publique Hopitaux Paris (APHP);
   Universite Paris Cite; Hopital Universitaire Necker-Enfants Malades -
   APHP; UNICANCER; Gustave Roussy
RP Soussain, C (corresponding author), CHG Meaux, Hematol Serv, 6-8 Rue St Fiacre,BP 218, F-77104 Meaux, France.
RI Hoang-Xuan, Khe/O-7527-2017; Cassoux, Nathalie/D-6844-2015
OI Cassoux, Nathalie/0000-0002-5535-9649; Hoang Xuan,
   Khe/0000-0003-4072-1999; levy, vincent/0000-0003-0423-1255
CR BALIS FM, 1989, AM J PEDIAT HEMATOL, V11, P74
   Blay JY, 1998, J CLIN ONCOL, V16, P864, DOI 10.1200/JCO.1998.16.3.864
   BLAY JY, 1995, BLOOD, V86, P2922
   COHEN IJ, 1986, CANCER-AM CANCER SOC, V57, P6, DOI 10.1002/1097-0142(19860101)57:1<6::AID-CNCR2820570104>3.0.CO;2-B
   Corry J, 1998, INT J RADIAT ONCOL, V41, P615, DOI 10.1016/S0360-3016(97)00571-3
   DEANGELIS LM, 1990, NEUROLOGY, V40, P80, DOI 10.1212/WNL.40.1.80
   DEANGELIS LM, 1992, J CLIN ONCOL, V10, P635, DOI 10.1200/JCO.1992.10.4.635
   Doolittle ND, 2000, CANCER, V88, P637, DOI 10.1002/(SICI)1097-0142(20000201)88:3<637::AID-CNCR22>3.0.CO;2-Y
   Freilich RJ, 1996, NEUROLOGY, V46, P435, DOI 10.1212/WNL.46.2.435
   KAPLAN EL, 1958, J AM STAT ASSOC, V53, P457, DOI 10.2307/2281868
   Khalfallah S, 1996, BONE MARROW TRANSPL, V18, P1021
   MACDONALD DR, 1990, J CLIN ONCOL, V8, P1277, DOI 10.1200/JCO.1990.8.7.1277
   MARKS P, 1999, ANN ONCOL S3, V10, P15
   ONEIL BP, 1995, P AM SOC CLIN ONCOL, V14, P141
   Patte C., 1990, MED PEDIATR ONCOL, V18, P406
   PHILIP T, 1995, NEW ENGL J MED, V333, P150
   PRZEPIORKA D, 1995, LEUKEMIA LYMPHOMA, V17, P427, DOI 10.3109/10428199509056853
   Sarazin M, 1995, EUR J CANCER, V31A, P2003, DOI 10.1016/0959-8049(95)00345-2
   Soussain C, 1996, LEUKEMIA LYMPHOMA, V23, P339, DOI 10.3109/10428199609054837
   SOUSSAIN C, 1995, BLOOD, V85, P664, DOI 10.1182/blood.V85.3.664.bloodjournal853664
   TALHA MRZ, 1980, BRIT J CANCER, V41, P140, DOI 10.1038/bjc.1980.17
   vanBesien K, 1996, J CLIN ONCOL, V14, P3036, DOI 10.1200/JCO.1996.14.11.3036
   vanBesien K, 1997, ANN ONCOL, V8, P515, DOI 10.1023/A:1008248315859
   WOLFF SN, 1995, HIGH DOSE CANC THERA, P879
NR 24
TC 213
Z9 223
U1 0
U2 1
PU LIPPINCOTT WILLIAMS & WILKINS
PI PHILADELPHIA
PA 530 WALNUT ST, PHILADELPHIA, PA 19106-3621 USA
SN 0732-183X
J9 J CLIN ONCOL
JI J. Clin. Oncol.
PD FEB 1
PY 2001
VL 19
IS 3
BP 742
EP 749
DI 10.1200/JCO.2001.19.3.742
PG 8
WC Oncology
WE Science Citation Index Expanded (SCI-EXPANDED)
SC Oncology
GA 399GK
UT WOS:000166803100019
PM 11157026
DA 2024-06-03
ER

PT J
AU Illerhaus, G
   Marks, R
   Ihorst, G
   Guttenberger, R
   Ostertag, C
   Derigs, G
   Frickhofen, N
   Feuerhake, F
   Volk, B
   Finke, J
AF Illerhaus, Gerald
   Marks, Reinhard
   Ihorst, Gabriele
   Guttenberger, Roland
   Ostertag, Christoph
   Derigs, Guenther
   Frickhofen, Norbert
   Feuerhake, Friedrich
   Volk, Benedikt
   Finke, Juergen
TI High-dose chemotherapy with autologous stern-cell transplantation and
   hyperfractionated radiotherapy as first-line treatment of primary CNS
   lymphoma
SO JOURNAL OF CLINICAL ONCOLOGY
LA English
DT Article; Proceedings Paper
CT 47th Annual Meeting of the American-Society-of-Hematology
CY DEC 10-13, 2005
CL Atlanta, GA
SP Amer Soc Hematol
ID CENTRAL-NERVOUS-SYSTEM; NON-HODGKINS-LYMPHOMA; PHASE-II; DEFERRED
   RADIOTHERAPY; RESPONSE CRITERIA; INITIAL TREATMENT; METHOTREXATE;
   THERAPY; MULTICENTER; SURVIVAL
AB Purpose To improve survival and reduce toxicity in primary CNS lymphoma (PCNSL) treatment, we conducted a multicenter phase II study with early high-dose chemotherapy (HDT) and autologous stem-cell transplantation ASCT) followed by hyperfractionated whole-brain radiotherapy (WBRT) for newly diagnosed PCNSL patients younger than 65 years of age.
   Patients and Methods Chemotherapy included three steps: three cycles of methortrexate (8 g/m(2)); cytarabine (AraC; two doses of 3 g/m(2)) and thiotepa (40 mg/m(2)) followed by stem-cell harvest; HDT with carmustine (400 mg/m(2)) and thiotepa (two doses of 5 mg/kg body weight) followed by ASCT. WBRT (45 Gy, two doses of 1 Gy/d) was administered for consolidation.
   Results Thirty patients with PCNSL younger than 65 years of age (median, 54 years; range, 27 years to 64 years) were enrolled (nine pilot-phase; 21 phase II). Twenty-eight patients responded to methotrexate: six patients with complete remission (CR), 15 patients with partial remission (PR), and seven patients with stable disease (SD) with clinical improvement. Of 26 patients proceeding to AraC and thiotepa, 10 patients achieved CR, 14 patients achieved PR, one patient experienced SD with clinical improvement, and one patient suffered disease progression. Twenty-three patients received HDT plus ASCT, resulting in 15 patients with CRs and eight patients with PRs. After WBRT, 21 of 21 patients had CRs. One patient died from liver failure after methotrexate. HDT was well tolerated apart from WHO grade 3/4 cytopenia. With a median follow-up of (33 months (range, 4 months to 84 months), 5-year overall survival probability is 69% for all patients and 87% for the 23 patients receiving HDT plus ASCT. The 5-year probability of relapse-related death is 21% for all patients (n=30) and 8.7% for patients treated with HDT plus ASCT (n=23).
   Conclusion Sequential systemic methotrexate and AraC and thiotepa followed by HDT plus ASCT and hyperfractionated WBRT is very effective with little toxicity as initial therapy for PCNSL.
C1 Univ Med Ctr Freiburg, Dept Hematol & Oncol, Ctr Clin Trials, D-79106 Freiburg, Germany.
   Univ Med Ctr Freiburg, Dept Stereotact Neurosurg, Ctr Clin Trials, D-79106 Freiburg, Germany.
   Univ Med Ctr Freiburg, Dept Radiotherapy, Ctr Clin Trials, D-79106 Freiburg, Germany.
   Univ Med Ctr Freiburg, Dept Neuropathol, Ctr Clin Trials, D-79106 Freiburg, Germany.
   Univ Hosp Mainz, Mainz, Germany.
   Dr Horst Schmidt Klin, Wiesbaden, Germany.
C3 University of Freiburg; University of Freiburg; University of Freiburg;
   University of Freiburg; University Hospital Mainz
RP Finke, J (corresponding author), Univ Med Ctr Freiburg, Dept Hematol & Oncol, Ctr Clin Trials, Hugstetter Str 55, D-79106 Freiburg, Germany.
EM
RI Frickhofen, Norbert/KDN-5002-2024; Ihorst, Gabriele/AAA-7445-2021
OI Frickhofen, Norbert/0000-0001-9916-2226; 
CR Abrey LE, 2003, J CLIN ONCOL, V21, P4151, DOI 10.1200/JCO.2003.05.024
   Abrey LE, 2005, J CLIN ONCOL, V23, P5034, DOI 10.1200/JCO.2005.13.524
   Abrey LE, 1998, J CLIN ONCOL, V16, P859, DOI 10.1200/JCO.1998.16.3.859
   Abrey LE, 2000, J CLIN ONCOL, V18, P3144, DOI 10.1200/JCO.2000.18.17.3144
   Batchelor T, 2003, J CLIN ONCOL, V21, P1044, DOI 10.1200/JCO.2003.03.036
   Bessell EM, 1996, J CLIN ONCOL, V14, P945, DOI 10.1200/JCO.1996.14.3.945
   Blay JY, 1998, J CLIN ONCOL, V16, P864, DOI 10.1200/JCO.1998.16.3.864
   BLAY JY, 1995, BLOOD, V86, P2922
   COLOMBAT P, 2003, P AN M AM SOC CLIN, V22, P100
   Corn BW, 1997, CANCER, V79, P2409
   DeAngelis LM, 2002, J CLIN ONCOL, V20, P4643, DOI 10.1200/JCO.2002.11.013
   DeAngelis LM, 1999, J NEURO-ONCOL, V43, P249, DOI 10.1023/A:1006258619757
   Ferreri AJM, 2001, ONCOLOGY-BASEL, V60, P134, DOI 10.1159/000055310
   Filley CM, 2001, NEW ENGL J MED, V345, P425, DOI 10.1056/NEJM200108093450606
   Fisher B, 2005, J NEURO-ONCOL, V74, P201, DOI 10.1007/s11060-004-6596-9
   GLASS J, 1994, J NEUROSURG, V81, P188, DOI 10.3171/jns.1994.81.2.0188
   Guha-Thakurta N, 1999, J NEURO-ONCOL, V43, P259, DOI 10.1023/A:1006210703827
   Herrlinger U, 2002, ANN NEUROL, V51, P247, DOI 10.1002/ana.10102
   Korfel A, 1998, LEUKEMIA LYMPHOMA, V30, P609, DOI 10.3109/10428199809057573
   MACDONALD DR, 1990, J CLIN ONCOL, V8, P1277, DOI 10.1200/JCO.1990.8.7.1277
   Marubini E., 1994, Analysing survival data from clinical trials and observational studies
   MILLER DC, 1994, CANCER-AM CANCER SOC, V74, P1383, DOI 10.1002/1097-0142(19940815)74:4<1383::AID-CNCR2820740432>3.0.CO;2-1
   Milpied N, 2004, NEW ENGL J MED, V350, P1287, DOI 10.1056/NEJMoa031770
   Nelson DF, 1999, J NEURO-ONCOL, V43, P241, DOI 10.1023/A:1006206602918
   O'Brien P, 2000, J CLIN ONCOL, V18, P519, DOI 10.1200/JCO.2000.18.3.519
   Pels H, 2003, J CLIN ONCOL, V21, P4489, DOI 10.1200/JCO.2003.04.056
   Soussain C, 2001, J CLIN ONCOL, V19, P742, DOI 10.1200/JCO.2001.19.3.742
   STEWART DA, 2004, BLOOD, V104, P4315
NR 28
TC 210
Z9 221
U1 0
U2 11
PU AMER SOC CLINICAL ONCOLOGY
PI ALEXANDRIA
PA 330 JOHN CARLYLE ST, STE 300, ALEXANDRIA, VA 22314 USA
SN 0732-183X
J9 J CLIN ONCOL
JI J. Clin. Oncol.
PD AUG 20
PY 2006
VL 24
IS 24
BP 3865
EP 3870
DI 10.1200/JCO.2006.06.2117
PG 6
WC Oncology
WE Conference Proceedings Citation Index - Science (CPCI-S); Science Citation Index Expanded (SCI-EXPANDED)
SC Oncology
GA 077TD
UT WOS:000240052300011
PM 16864853
OA Bronze
DA 2024-06-03
ER

PT J
AU Abrey, LE
   Moskowitz, CH
   Mason, WP
   Crump, M
   Stewart, D
   Forsyth, P
   Paleologos, N
   Correa, DD
   Anderson, ND
   Caron, D
   Zelenetz, A
   Nimer, SD
   DeAngelis, LM
AF Abrey, LE
   Moskowitz, CH
   Mason, WP
   Crump, M
   Stewart, D
   Forsyth, P
   Paleologos, N
   Correa, DD
   Anderson, ND
   Caron, D
   Zelenetz, A
   Nimer, SD
   DeAngelis, LM
TI Intensive methotrexate and cytarabine followed by high-dose chemotherapy
   with autologous stem-cell rescue in patients with newly diagnosed
   primary CNS lymphoma: An intent-to-treat analysis
SO JOURNAL OF CLINICAL ONCOLOGY
LA English
DT Article; Proceedings Paper
CT 37th Annual Meeting of the American-Society-of-Clinical-Oncology
CY MAY 12-15, 2001
CL SAN FRANCISCO, CA
SP Amer Soc Clin Oncol
ID NON-HODGKINS-LYMPHOMA; NERVOUS-SYSTEM LYMPHOMA; THERAPY; MULTICENTER;
   TRANSPLANTATION; RADIOTHERAPY; REGIMENS; SURVIVAL; TRIAL; RISK
AB Purpose: To assess the safety and efficacy of intensive methotrexate-based chemotherapy followed by high-dose chemotherapy (HDT) with autologous stem-cell rescue in patients with newly diagnosed primary CNS lymphoma (PCNSL).
   Patients and Methods: Twenty-eight patients received induction chemotherapy using high-dose systemic methotrexate (3.5 g/m(2)) and cytarabine (3 g/m(2) daily for 2 days). Fourteen patients with chemosensitive disease evident on neuroimaging then received high-dose therapy using carmustine, etoposide, cytarabine, and melphalan with autologous stem-cell rescue.
   Results: The objective response rate to the induction-phase chemotherapy was 57%, and median overall survival is not yet assessable, with a median follow-up time of 28 months. The overall median event-free survival time is 5.6 months for all patients and 9.3 months for 14 patients who underwent transplantation. Six of these 14 patients (43%) remained disease-free at last follow-up. Treatment was well tolerated; there was one transplantation-related death. Prospective neuropsychologic evaluations have revealed no evidence of treatment-related neurotoxicity.
   Conclusion: This treatment approach is feasible in patients with newly diagnosed PCNSL without evidence of significant related neurotoxicity. Although the transplantation results are similar to those achieved in patients with aggressive or poor-prognosis systemic lymphoma, the low response rate to induction chemotherapy and the significant number of patients who experienced relapse soon after HDT suggest that more aggressive induction chemotherapy may be warranted. (C) 2003 by American Society of Clinical Oncology.
C1 Mem Sloan Kettering Canc Ctr, Dept Neurol, New York, NY 10021 USA.
   Mem Sloan Kettering Canc Ctr, Dept Med, New York, NY 10021 USA.
   Mem Sloan Kettering Canc Ctr, Off Clin Res, New York, NY 10021 USA.
   Univ Toronto, Princess Margaret Hosp, Toronto, ON, Canada.
   Univ Calgary, Tom Baker Canc Ctr, Calgary, AB, Canada.
   Northwestern Univ, Evanston Hosp, Dept Neurol, Chicago, IL 60611 USA.
C3 Memorial Sloan Kettering Cancer Center; Memorial Sloan Kettering Cancer
   Center; Memorial Sloan Kettering Cancer Center; University of Toronto;
   University Health Network Toronto; Princess Margaret Cancer Centre; Tom
   Baker Cancer Clinic; University of Calgary; Northwestern University;
   NorthShore University Health System
RP Mem Sloan Kettering Canc Ctr, Dept Neurol, 1275 York Ave, New York, NY 10021 USA.
EM
RI Anderson, Nicole D/E-9101-2011
OI Anderson, Nicole/0000-0002-5979-2220; DeAngelis,
   Lisa/0000-0002-6290-7818; Zelenetz, Andrew/0000-0003-1403-6883
CR Abrey LE, 1998, J CLIN ONCOL, V16, P859, DOI 10.1200/JCO.1998.16.3.859
   Abrey LE, 2000, J CLIN ONCOL, V18, P3144, DOI 10.1200/JCO.2000.18.17.3144
   Abrey LE, 1997, NEUROLOGY, V48, pV1003
   Batchelor T, 2003, J CLIN ONCOL, V21, P1044, DOI 10.1200/JCO.2003.03.036
   CADMAN E, 1979, J CLIN INVEST, V64, P788, DOI 10.1172/JCI109525
   Cairncross G, 2000, NEURO-ONCOLOGY, V2, P114, DOI 10.1093/neuonc/2.2.114
   Cheng G, 2003, BONE MARROW TRANSPL, V31, P679, DOI 10.1038/sj.bmt.1703917
   COLTMAN AC, 1986, UPDATE TREATMENT DIF, P71
   Deangelis L M, 1995, Oncology (Williston Park), V9, P63
   DeAngelis LM, 2002, J CLIN ONCOL, V20, P4643, DOI 10.1200/JCO.2002.11.013
   Doolittle ND, 2000, CANCER, V88, P637, DOI 10.1002/(SICI)1097-0142(20000201)88:3<637::AID-CNCR22>3.0.CO;2-Y
   EDELSTEIN M, 1975, CANCER RES, V35, P1555
   FISHER RI, 1993, NEW ENGL J MED, V328, P1002, DOI 10.1056/NEJM199304083281404
   GLASS J, 1994, J NEUROSURG, V81, P188, DOI 10.3171/jns.1994.81.2.0188
   Haioun C, 2000, J CLIN ONCOL, V18, P3025, DOI 10.1200/JCO.2000.18.16.3025
   Herrlinger U, 2002, ANN NEUROL, V51, P247, DOI 10.1002/ana.10102
   KAPLAN EL, 1958, J AM STAT ASSOC, V53, P457, DOI 10.2307/2281868
   Kewalramani T, 2001, Curr Oncol Rep, V3, P271, DOI 10.1007/s11912-001-0061-0
   KLUINNELEMANS HC, 2001, J NATL CANCER I, V93, P4
   Martelli M, 2003, J CLIN ONCOL, V21, P1255, DOI 10.1200/JCO.2003.01.117
   PHILIP T, 1995, NEW ENGL J MED, V333, P1540, DOI 10.1056/NEJM199512073332305
   Santini G, 2000, HAEMATOLOGICA, V85, P160
   Soussain C, 2001, J CLIN ONCOL, V19, P742, DOI 10.1200/JCO.2001.19.3.742
   WIEBE VJ, 1992, CRIT REV ONCOL HEMAT, V13, P241, DOI 10.1016/1040-8428(92)90092-5
NR 24
TC 210
Z9 216
U1 0
U2 1
PU AMER SOC CLINICAL ONCOLOGY
PI ALEXANDRIA
PA 2318 MILL ROAD, STE 800, ALEXANDRIA, VA 22314 USA
SN 0732-183X
EI 1527-7755
J9 J CLIN ONCOL
JI J. Clin. Oncol.
PD NOV 15
PY 2003
VL 21
IS 22
BP 4151
EP 4156
DI 10.1200/JCO.2003.05.024
PG 6
WC Oncology
WE Science Citation Index Expanded (SCI-EXPANDED); Conference Proceedings Citation Index - Science (CPCI-S)
SC Oncology
GA 745BW
UT WOS:000186669300015
PM 14615443
DA 2024-06-03
ER

PT J
AU Schultz, C
   Scott, C
   Sherman, W
   Donahue, B
   Fields, J
   Murray, K
   Fisher, B
   Abrams, R
   MeisKindblom, J
AF Schultz, C
   Scott, C
   Sherman, W
   Donahue, B
   Fields, J
   Murray, K
   Fisher, B
   Abrams, R
   MeisKindblom, J
TI Preirradiation chemotherapy with cyclophosphamide, doxorubicin,
   vincristine, and dexamethasone for primary CNS lymphomas: Initial report
   of Radiation Therapy Oncology Group protocol 88-06
SO JOURNAL OF CLINICAL ONCOLOGY
LA English
DT Article
ID CENTRAL-NERVOUS-SYSTEM; PRIMARY MALIGNANT-LYMPHOMA; RENAL-TRANSPLANT
   RECIPIENTS; IMMUNE-DEFICIENCY SYNDROME; NON-HODGKINS-LYMPHOMA;
   BRAIN-TUMORS; SURVIVAL; METHOTREXATE; PATTERNS; FAILURE
AB Purpose: This study was a prospective phase I/II trial performed by the Radiation Therapy Oncology Group (RTOG) to test the tolerance and efficacy of preirradiation cyclophosphamide, doxorubicin, vincristine, and dexamethasone (CHOD) chemotherapy followed by large-volume, high-dose brain radiation therapy (Ri) for patients with primary CNS lymphoma (PCNSL).
   Patients and Methods: Fifty-four (52 assessable) human immunodeficiency virus (HIV)-negative patients with PCNSL were entered on study and received two (n = 20) or three (n = 32) cycles of CHOD (six patients with positive CSF cytology received intrathecal methotrexate in addition to CHOD). Whole-brain RT to 41.4 Gy and tumor boost to 18 Gy (total dose, 59.4 Gy) followed chemotherapy.
   Results: As of July 1994, with a minimum potential follow-up time of 20 months, 12 of 52 assessable patients remain alive without evidence of progression. The median survival time for the entire group is 16.1 months, with a 2-year survival rate of 42%. By univariate analysis, patient age was found to be a significant prognostic factor with respect to survival (P =.005) in favor of age less than 60 years, Karnofsky performance status (KPS) was of borderline significance (P =.057). Survival for patients treated on RTOG 88-06 was compared with that of patients treated on RTOG 83-15, which tested RT alone. No difference in overall survival was found (P =.53). Grade 4 neutropenia developed in 29 of 51 patients during chemotherapy, There were two deaths during chemotherapy: one as a result of sepsis and one of a pulmonary embolus. The worst toxicity during RT was less than or equal to grade 2 in 50 of 52 patients.
   Conclusion: preirradiation CHOD chemotherapy does not significantly improve survival over RT alone for patients with PCNSL. Age remains a powerful prognostic factor independent of therapy and must be considered in testing alternative combined approaches. (C) 1996 by American Society of Clinical Oncology.
C1 RADIAT THERAPY ONCOL GRP,PHILADELPHIA,PA.
   COLUMBIA PRESBYTERIAN MED CTR,DEPT RADIAT ONCOL,NEW YORK,NY 10032.
   NYU,MED CTR,DEPT RADIAT ONCOL,NEW YORK,NY.
   UNIV WASHINGTON,DEPT RADIAT ONCOL,ST LOUIS,MO.
   LONDON REG CANC CTR,DEPT RADIAT ONCOL,LONDON,ON N6A 4L6,CANADA.
   JOHNS HOPKINS ONCOL CTR,DEPT RADIAT ONCOL,BALTIMORE,MD.
   LOUISIANA STATE UNIV,MED CTR,DEPT PATHOL,NEW ORLEANS,LA 70112.
C3 Radiation Therapy Oncology Group (RTOG); NewYork-Presbyterian Hospital;
   Columbia University; New York University; Washington University (WUSTL);
   Western University (University of Western Ontario); Johns Hopkins
   University; Johns Hopkins Medicine; Louisiana State University System
RP Schultz, C (corresponding author), MED COLL WISCONSIN,DEPT RADIAT ONCOL,8700 W WISCONSIN AVE,DH-165,MILWAUKEE,WI 53226, USA.
FU NCI NIH HHS [CA-21661, CA-32115] Funding Source: Medline
CR BERRY MP, 1981, INT J RADIAT ONCOL, V7, P55, DOI 10.1016/0360-3016(81)90060-2
   CHAMBERLAIN MC, 1992, J NEURO-ONCOL, V14, P271
   DEANGELIS LM, 1992, J CLIN ONCOL, V10, P635, DOI 10.1200/JCO.1992.10.4.635
   EBY NL, 1988, CANCER-AM CANCER SOC, V62, P2461, DOI 10.1002/1097-0142(19881201)62:11<2461::AID-CNCR2820621135>3.0.CO;2-M
   EDGAR R, 1961, NEUROLOGY, V11, P239, DOI 10.1212/WNL.11.3.239
   ERVIN T, 1980, CANCER, V45, P1556, DOI 10.1002/1097-0142(19800401)45:7<1556::AID-CNCR2820450707>3.0.CO;2-B
   FREEMAN CR, 1986, CANCER-AM CANCER SOC, V58, P1106, DOI 10.1002/1097-0142(19860901)58:5<1106::AID-CNCR2820580521>3.0.CO;2-3
   GLASS J, 1994, J NEUROSURG, V81, P188, DOI 10.3171/jns.1994.81.2.0188
   GREIG NH, 1990, JNCI-J NATL CANCER I, V82, P1621, DOI 10.1093/jnci/82.20.1621
   GUINAN ME, 1987, JAMA-J AM MED ASSOC, V257, P2039
   GUNDERSON CH, 1971, J NEUROSURG, V35, P406, DOI 10.3171/jns.1971.35.4.0406
   HENRY JM, 1974, CANCER, V34, P1293, DOI 10.1002/1097-0142(197410)34:4<1293::AID-CNCR2820340441>3.0.CO;2-P
   HERBST KD, 1976, CANCER-AM CANCER SOC, V38, P1476, DOI 10.1002/1097-0142(197610)38:4<1476::AID-CNCR2820380405>3.0.CO;2-J
   HOOVER R, 1973, LANCET, V2, P55
   JACK CR, 1988, RADIOLOGY, V167, P211, DOI 10.1148/radiology.167.1.3279454
   Jellinger K, 1975, Acta Neuropathol Suppl, VSuppl 6, P95
   KAWAKAMI Y, 1985, J NEUROSURG, V62, P522, DOI 10.3171/jns.1985.62.4.0522
   LACHANCE D H, 1992, Neurology, V42, P343
   LOEFFLER JS, 1985, J CLIN ONCOL, V3, P490, DOI 10.1200/JCO.1985.3.4.490
   MCLAUGHLIN P, 1988, JNCI-J NATL CANCER I, V80, P1408, DOI 10.1093/jnci/80.17.1408
   MENDENHALL NP, 1983, CANCER-AM CANCER SOC, V52, P1993, DOI 10.1002/1097-0142(19831201)52:11<1993::AID-CNCR2820521104>3.0.CO;2-C
   MICHALSKI JM, 1990, RADIOLOGY, V176, P855, DOI 10.1148/radiology.176.3.2389047
   MURRAY K, 1986, J NEUROSURG, V65, P600, DOI 10.3171/jns.1986.65.5.0600
   NELSON DF, 1992, INT J RADIAT ONCOL, V23, P9, DOI 10.1016/0360-3016(92)90538-S
   NEUWELT EA, 1991, J CLIN ONCOL, V9, P1580, DOI 10.1200/JCO.1991.9.9.1580
   OTT RJ, 1991, EUR J CANCER, V27, P1356, DOI 10.1016/0277-5379(91)90009-3
   SCHAUMBURG HH, 1972, BRAIN, V95, P199, DOI 10.1093/brain/95.2.199
   SCHNECK SA, 1971, LANCET, V1, P983
   SHIBAMOTO Y, 1990, CANCER-AM CANCER SOC, V65, P1907, DOI 10.1002/1097-0142(19900501)65:9<1907::AID-CNCR2820650906>3.0.CO;2-L
   SNIDER WD, 1983, NEW ENGL J MED, V308, P45
   STEWART DJ, 1984, J NEURO-ONCOL, V2, P289
   WELCH K, 1984, JAMA-J AM MED ASSOC, V252, P1152, DOI 10.1001/jama.252.9.1152
   Yahalom Joachim, 1994, International Journal of Radiation Oncology Biology Physics, V30, P215, DOI 10.1016/0360-3016(94)90732-3
   YAMASAKI T, 1993, SURG NEUROL, V40, P383, DOI 10.1016/0090-3019(93)90217-O
   ZIEGLER JL, 1982, LANCET, V2, P631
NR 35
TC 203
Z9 220
U1 0
U2 0
PU W B SAUNDERS CO
PI PHILADELPHIA
PA INDEPENDENCE SQUARE WEST CURTIS CENTER, STE 300, PHILADELPHIA, PA
   19106-3399
SN 0732-183X
J9 J CLIN ONCOL
JI J. Clin. Oncol.
PD FEB
PY 1996
VL 14
IS 2
BP 556
EP 564
DI 10.1200/JCO.1996.14.2.556
PG 9
WC Oncology
WE Science Citation Index Expanded (SCI-EXPANDED)
SC Oncology
GA TU998
UT WOS:A1996TU99800034
PM 8636771
DA 2024-06-03
ER

PT J
AU Smith, JR
   Rosenbaum, JT
   Wilson, DJ
   Doolittle, ND
   Siegal, T
   Neuwelt, EA
   Pe'er, J
AF Smith, JR
   Rosenbaum, JT
   Wilson, DJ
   Doolittle, ND
   Siegal, T
   Neuwelt, EA
   Pe'er, J
TI Role of intravitreal methotrexate in the management of primary central
   nervous system lymphoma with ocular involvement
SO OPHTHALMOLOGY
LA English
DT Article
ID POLYMERASE-CHAIN-REACTION; PRIMARY CNS; INTRAOCULAR LYMPHOMA;
   CHEMOTHERAPY; EPIDEMIOLOGY; DIAGNOSIS; THERAPY
AB Purpose: To evaluate the safety and efficacy of intravitreal methotrexate in the management of primary central nervous system lymphoma (PCNSL) involving the eye.
   Design: Retrospective noncomparative interventional case series.
   Participants: Sixteen human immunodeficiency virus-negative white patients (5 males and 11 females, aged 30-76 years) with intraocular B cell lymphoma treated with intravitreal methotrexate at Oregon Health & Science University or Hadassah University Hospital between August 1995 and September 2000.
   Intervention: Patients were treated with intravitreal methotrexate (400 mug/0.1 ml) according to a standard induction-consolidation-maintenance regimen and monitored by serial examinations, including measurement of visual acuity, slit-lamp biomicroscopy, and dilated funduscopy.
   Main Outcome Measures: Clinical response to intravitreal chemotherapy, number of injections for clinical remission, visual acuity, complications during study period, length of survival from diagnosis, and cause of death.
   Results. Time of follow-up from commencement of the methotrexate injections was 6 to 35 months (median, 18.5 months). Twenty-six of 26 eyes (100%) were cleared clinically of malignant cells after a maximum of 12 methotrexate injections. A second remission was induced in three patients, who were treated with a further course of intravitreal chemotherapy after their tumor recurred within the eye. Complications that occurred during the period of treatment and follow-up included cataract (73% of 26 eyes), corneal epitheliopathy (58% of 26 eyes), maculopathy (42% of 26 eyes), vitreous hemorrhage (8% of 26 eyes), optic atrophy (4% of 26 eyes), and sterile endophthalmitis (4% of 26 eyes). No patient had irreversible loss of vision that could be definitely attributed to the intravitreal injection of methotrexate.
   Conclusions: Intravitreal chemotherapy with methotrexate is effective in inducing clinical remission of intraocular tumor in PCNSL with acceptable morbidity. Further study is indicated to determine whether this approach extends life expectancy.
C1 Oregon Hlth & Sci Univ, Casey Eye Inst, Portland, OR 97201 USA.
   Oregon Hlth & Sci Univ, Dept Med, Portland, OR 97201 USA.
   Oregon Hlth & Sci Univ, Dept Cell Biol, Portland, OR 97201 USA.
   Oregon Hlth & Sci Univ, Dept Neurol, Portland, OR 97201 USA.
   Hadassah Univ Hosp, Neurooncol Ctr, IL-91120 Jerusalem, Israel.
   Hebrew Univ Jerusalem, Hadassah Fac Med, Jerusalem, Israel.
   Hadassah Univ Hosp, Dept Ophthalmol, IL-91120 Jerusalem, Israel.
C3 Oregon Health & Science University; Oregon Health & Science University;
   Oregon Health & Science University; Oregon Health & Science University;
   Hebrew University of Jerusalem; Hebrew University of Jerusalem; Hebrew
   University of Jerusalem
RP Oregon Hlth & Sci Univ, Casey Eye Inst, 3375 SW Terwilliger Blvd, Portland, OR 97201 USA.
RI Peer, Jacob/HSE-3187-2023; Smith, Justine/Y-9044-2019
OI Smith, Justine/0000-0002-4756-5493
FU NCI NIH HHS [R13-CA86959, CA31770] Funding Source: Medline; NEI NIH HHS
   [EY06484] Funding Source: Medline; NINDS NIH HHS [NS33618, NS34608]
   Funding Source: Medline
CR Abrey LE, 1998, J CLIN ONCOL, V16, P859, DOI 10.1200/JCO.1998.16.3.859
   Abrey LE, 2000, J CLIN ONCOL, V18, P3144, DOI 10.1200/JCO.2000.18.17.3144
   Buggage RR, 1999, INVEST OPHTH VIS SCI, V40, P2462
   CHAR DH, 1988, OPHTHALMOLOGY, V95, P625
   Cote TR, 1996, J NATL CANCER I, V88, P675, DOI 10.1093/jnci/88.10.675
   CUNHAVAZ J, 1979, SURV OPHTHALMOL, V23, P279, DOI 10.1016/0039-6257(79)90158-9
   de Smet MD, 1999, BRIT J OPHTHALMOL, V83, P448, DOI 10.1136/bjo.83.4.448
   deSmet MD, 1996, AM J OPHTHALMOL, V121, P442, DOI 10.1016/S0002-9394(14)70444-1
   Doolittle ND, 2000, CANCER, V88, P637, DOI 10.1002/(SICI)1097-0142(20000201)88:3<637::AID-CNCR22>3.0.CO;2-Y
   Fishburne BC, 1997, ARCH OPHTHALMOL-CHIC, V115, P1152, DOI 10.1001/archopht.1997.01100160322009
   Henson JW, 1999, J CLIN ONCOL, V17, P1329
   Hoang-Xuan K, 1999, J NEURO-ONCOL, V43, P287, DOI 10.1023/A:1006266821574
   HOCHBERG FH, 1988, J NEUROSURG, V68, P835, DOI 10.3171/jns.1988.68.6.0835
   Kraemer DF, 2001, NEUROSURGERY, V48, P1033, DOI 10.1097/00006123-200105000-00013
   MARGOLIS L, 1980, CANCER-AM CANCER SOC, V45, P688, DOI 10.1002/1097-0142(19800215)45:4<688::AID-CNCR2820450412>3.0.CO;2-F
   Martin DF, 1999, AM J OPHTHALMOL, V127, P329
   MILLAY RH, 1986, AM J OPHTHALMOL, V102, P626, DOI 10.1016/0002-9394(86)90536-2
   PETERSON K, 1993, CANCER, V72, P843, DOI 10.1002/1097-0142(19930801)72:3<843::AID-CNCR2820720333>3.0.CO;2-#
   Rhodes CH, 1996, CANCER-AM CANCER SOC, V77, P543
   Sandor V, 1998, J CLIN ONCOL, V16, P3000, DOI 10.1200/JCO.1998.16.9.3000
   Schabet M, 1999, J NEURO-ONCOL, V43, P199, DOI 10.1023/A:1006290032052
   Schlaepfer R, 2000, MANAG FOR ECOSYST, V1, P1
   Shen DF, 1998, OPHTHALMOLOGY, V105, P1664, DOI 10.1016/S0161-6420(98)99036-4
   SIEGEL MJ, 1989, BRIT J OPHTHALMOL, V73, P342, DOI 10.1136/bjo.73.5.342
   Slowik F, 1975, ACTA NEUROPATH BER S, V6, P95, DOI DOI 10.1007/978-3-662-08456-4_16
   Spaeth, 1982, OPHTHALMIC SURG PRIN, P410
   White VA, 1999, ARCH OPHTHALMOL-CHIC, V117, P761
   ZAMBER RW, 1992, WESTERN J MED, V157, P530
NR 28
TC 200
Z9 215
U1 1
U2 8
PU ELSEVIER SCIENCE INC
PI NEW YORK
PA STE 800, 230 PARK AVE, NEW YORK, NY 10169 USA
SN 0161-6420
EI 1549-4713
J9 OPHTHALMOLOGY
JI Ophthalmology
PD SEP
PY 2002
VL 109
IS 9
BP 1709
EP 1716
AR PII S0161-6420(02)01125-9
DI 10.1016/S0161-6420(02)01125-9
PG 8
WC Ophthalmology
WE Science Citation Index Expanded (SCI-EXPANDED)
SC Ophthalmology
GA 592CT
UT WOS:000177919700032
PM 12208721
DA 2024-06-03
ER

PT J
AU Bühring, U
   Herrlinger, U
   Krings, T
   Thiex, R
   Weller, M
   Küker, W
AF Bühring, U
   Herrlinger, U
   Krings, T
   Thiex, R
   Weller, M
   Küker, W
TI MRI features of primary central nervous system lymphomas at presentation
SO NEUROLOGY
LA English
DT Review
ID PRIMARY CNS LYMPHOMA; CHEMOTHERAPY
AB Pretreatment MRI examinations of 40 immunologically competent patients with primary CNS lymphoma (PCNSL) were evaluated (24 men, 16 Women, median age 63 years). Seventy lesions were found (mean size: 19.9 mm). The number of lesions ranged from one (n = 25) to six (n = 1). The most frequent locations were the cerebral hemispheres (n = 22), the corpus callosum (n = 11), and the basal ganglia (n = 11). Cerebellar manifestations were found in 10 patients. Ocular (n = 2) and medullary cord (n = 1) manifestations were rare. Contrast enhancement was encountered in all lesions. Although 39 patients had lesions adjacent to the CSF space, leptomeningeal spread was only present in five patients. Necrosis was seen in two lesions only. Edema was extensive in 24 patients, moderate in 11 patients, and absent in five patients. Contrast-enhancing lesions in contact with the subarachnoid space and without necrosis are characteristic of PCNSL.
C1 Univ Tubingen, Sch Med, Dept Neuroradiol, D-72076 Tubingen, Germany.
   Univ Tubingen, Sch Med, Dept Neurol, D-72076 Tubingen, Germany.
   Rhein Westfal TH Aachen, Sch Med, Dept Neuroradiol, Aachen, Germany.
   Rhein Westfal TH Aachen, Sch Med, Dept Neurosurg, Aachen, Germany.
C3 Eberhard Karls University of Tubingen; Eberhard Karls University of
   Tubingen; RWTH Aachen University; RWTH Aachen University
EM
RI McDermott, Michael Francis/GZM-5330-2022
OI Weller, Michael/0000-0002-1748-174X; Kuker, Wilhelm/0000-0002-1739-6115
CR Abrey LE, 2000, J CLIN ONCOL, V18, P3144, DOI 10.1200/JCO.2000.18.17.3144
   Bailey P, 1929, ARCH SURG-CHICAGO, V18, P1359, DOI 10.1001/archsurg.1929.01140130449031
   EBY NL, 1988, CANCER-AM CANCER SOC, V62, P2461, DOI 10.1002/1097-0142(19881201)62:11<2461::AID-CNCR2820621135>3.0.CO;2-M
   GOLDSTEIN JD, 1991, J COMPUT ASSIST TOMO, V15, P39, DOI 10.1097/00004728-199101000-00004
   Guha-Thakurta N, 1999, J NEURO-ONCOL, V43, P259, DOI 10.1023/A:1006210703827
   Herrlinger U, 2000, NEUROLOGY, V54, P1707, DOI 10.1212/WNL.54.8.1707
   Herrlinger U, 2001, CANCER, V91, P130, DOI 10.1002/1097-0142(20010101)91:1<130::AID-CNCR17>3.0.CO;2-8
   JELLINGER KA, 1992, J CANCER RES CLIN, V119, P7, DOI 10.1007/BF01209483
   Korfel A, 1998, LEUKEMIA LYMPHOMA, V30, P609, DOI 10.3109/10428199809057573
   LING SM, 1994, CANCER-AM CANCER SOC, V73, P2570, DOI 10.1002/1097-0142(19940515)73:10<2570::AID-CNCR2820731019>3.0.CO;2-1
   PETERSON K, 1993, CANCER, V72, P843, DOI 10.1002/1097-0142(19930801)72:3<843::AID-CNCR2820720333>3.0.CO;2-#
   Pramstaller PP, 1999, J NEUROL, V246, P934, DOI 10.1007/s004150050485
   ROMANGOLDSTEIN SM, 1992, AM J NEURORADIOL, V13, P1207
   Surawicz T S, 1999, Neuro Oncol, V1, P14, DOI 10.1093/neuonc/1.1.14
NR 14
TC 195
Z9 215
U1 0
U2 3
PU LIPPINCOTT WILLIAMS & WILKINS
PI PHILADELPHIA
PA TWO COMMERCE SQ, 2001 MARKET ST, PHILADELPHIA, PA 19103 USA
SN 0028-3878
EI 1526-632X
J9 NEUROLOGY
JI Neurology
PD AUG 14
PY 2001
VL 57
IS 3
BP 393
EP 396
DI 10.1212/WNL.57.3.393
PG 4
WC Clinical Neurology
WE Science Citation Index Expanded (SCI-EXPANDED)
SC Neurosciences & Neurology
GA 462FR
UT WOS:000170410500007
PM 11515505
DA 2024-06-03
ER

PT J
AU Omuro, AMP
   Ben-Porat, LS
   Panageas, KS
   Kim, AK
   Correa, DD
   Yahalom, J
   DeAngelis, LM
   Abrey, LE
AF Omuro, AMP
   Ben-Porat, LS
   Panageas, KS
   Kim, AK
   Correa, DD
   Yahalom, J
   DeAngelis, LM
   Abrey, LE
TI Delayed neurotoxicity in primary central nervous system lymphoma
SO ARCHIVES OF NEUROLOGY
LA English
DT Article
ID PRIMARY CNS LYMPHOMA; HIGH-DOSE METHOTREXATE; WHITE-MATTER LESIONS;
   PHASE-II; DEFERRED RADIOTHERAPY; EUROPEAN ORGANIZATION;
   IONIZING-RADIATION; PROGENITOR CELLS; COMPETING RISK; CHEMOTHERAPY
AB Background: Treatment for primary central nervous lymphoma (PCNSL) with chemotherapy and radiotherapy has resulted in improved survival, but some patients develop neurologic deterioration that represents a treatment-related toxic effect. This delayed neurotoxicity has been poorly defined in the literature, and the underlying mechanisms are unknown.
   Objective: To describe the clinical findings, time course, and pathophysiologic mechanisms associated with neurotoxicity in an attempt to generate hypotheses for future studies that address prevention and treatment of this complication of successful PCNSL therapy.
   Design: Retrospective review
   Setting: Department of Neurology, Memorial Sloan-Kettering Cancer Center.
   Patients: One hundred eighty-five patients treated for PCNSL, including 43 who developed neurotoxicity.
   Main Outcome Measures: Potential risk factors, clinical course, and neuropsychological, neuroimaging, and histologic findings.
   Results: The 5-year cumulative incidence of neurotoxicity was 24%; this incidence increases over time. Neurotoxicity presented as a rapidly progressive subcortical dementia characterized by psychomotor slowing, executive and memory dysfunction, behavioral changes, gait ataxia, and incontinence. Imaging findings revealed diffuse white matter disease and cortical-subcortical atrophy. Available autopsy data showed white matter damage with gliosis, thickening of small vessels, and demyelination. Statistical analyses were performed, accounting for death as a competing risk. Older age (P=.01), mental status changes at diagnosis (P=.04), female sex (P=.05), and radiotherapy (P <.001) predicted neurotoxicity on univariate analysis, but only radiotherapy remained significant in the multivariate setting.
   Conclusion: These findings suggest that the core pathophysiologic mechanism is the interruption of frontal subcortical circuits mediated by radiation damage, possibly caused by progressive microvascular alterations, loss of oligodendrocyte progenitors, or oxidative stress.
C1 Mem Sloan Kettering Canc Ctr, Dept Neurol, New York, NY 10021 USA.
   Mem Sloan Kettering Canc Ctr, Dept Epidemiol & Biostat, New York, NY 10021 USA.
   Mem Sloan Kettering Canc Ctr, Dept Radiat Oncol, New York, NY 10021 USA.
C3 Memorial Sloan Kettering Cancer Center; Memorial Sloan Kettering Cancer
   Center; Memorial Sloan Kettering Cancer Center
RP Mem Sloan Kettering Canc Ctr, Dept Neurol, 1275 York Ave, New York, NY 10021 USA.
EM
OI Price, Leah/0000-0002-5092-4768; Omuro, Antonio/0000-0003-4299-3664;
   DeAngelis, Lisa/0000-0002-6290-7818
CR Abrey LE, 1998, J CLIN ONCOL, V16, P859, DOI 10.1200/JCO.1998.16.3.859
   Abrey LE, 2000, J CLIN ONCOL, V18, P3144, DOI 10.1200/JCO.2000.18.17.3144
   ALLEN JC, 1980, CANCER TREAT REP, V64, P1261
   Batchelor T, 2003, J CLIN ONCOL, V21, P1044, DOI 10.1200/JCO.2003.03.036
   Blay JY, 1998, J CLIN ONCOL, V16, P864, DOI 10.1200/JCO.1998.16.3.864
   BLEYER WA, 1981, CANCER TREAT REP, V65, P89
   Chan AS, 2004, CANCER, V100, P398, DOI 10.1002/cncr.11885
   Correa DD, 2004, NEUROLOGY, V62, P548, DOI 10.1212/01.WNL.0000109673.75316.D8
   CUMMINGS JL, 1993, ARCH NEUROL-CHICAGO, V50, P873, DOI 10.1001/archneur.1993.00540080076020
   FAZEKAS F, 1987, AM J ROENTGENOL, V149, P351, DOI 10.2214/ajr.149.2.351
   Fine JP, 1999, J AM STAT ASSOC, V94, P496, DOI 10.2307/2670170
   GLANTZ MJ, 1994, NEUROLOGY, V44, P2020, DOI 10.1212/WNL.44.11.2020
   Goldman S, 2003, TRENDS NEUROSCI, V26, P590, DOI 10.1016/j.tins.2003.09.011
   GRAY RJ, 1988, ANN STAT, V16, P1141, DOI 10.1214/aos/1176350951
   Harder H, 2004, NEUROLOGY, V62, P544, DOI 10.1212/WNL.62.4.544
   Hebb AO, 2001, NEUROSURGERY, V49, P1166, DOI 10.1097/00006123-200111000-00028
   Herrlinger U, 2002, ANN NEUROL, V51, P247, DOI 10.1002/ana.10102
   Hoang-Xuan K, 2003, J CLIN ONCOL, V21, P2726, DOI 10.1200/JCO.2003.11.036
   Hulshof MCCM, 2002, STRAHLENTHER ONKOL, V178, P192, DOI 10.1007/s00066-002-0916-9
   Lai R, 2002, NEUROLOGY, V59, P1557, DOI 10.1212/01.WNL.0000034256.20173.EA
   Lai R, 2004, NEUROLOGY, V62, P451, DOI 10.1212/01.WNL.0000106941.51340.A2
   Mulhern RK, 1999, ANN NEUROL, V46, P834, DOI 10.1002/1531-8249(199912)46:6<834::AID-ANA5>3.0.CO;2-M
   MULLENIX PJ, 1994, PEDIATR RES, V35, P171, DOI 10.1203/00006450-199402000-00009
   Nieder C, 2004, ANTICANCER RES, V24, P3803
   O'Neill BP, 2004, NEUROLOGY, V62, P532, DOI 10.1212/WNL.62.4.532
   Pels H, 2003, J CLIN ONCOL, V21, P4489, DOI 10.1200/JCO.2003.04.056
   Poortmans PMP, 2003, J CLIN ONCOL, V21, P4483, DOI 10.1200/JCO.2003.03.108
   Román GC, 2002, LANCET NEUROL, V1, P426, DOI 10.1016/S1474-4422(02)00190-4
   Steen RG, 2001, ANN NEUROL, V50, P787, DOI 10.1002/ana.10029
   Tofilon PJ, 2000, RADIAT RES, V153, P357, DOI 10.1667/0033-7587(2000)153[0357:TROTCN]2.0.CO;2
   Tullberg M, 2004, NEUROLOGY, V63, P246, DOI 10.1212/01.WNL.0000130530.55104.B5
   VANDERKOGEL AJ, 1985, RADIOTHER ONCOL, V4, P239, DOI 10.1016/S0167-8140(85)80089-X
   VANDERMAAZEN RWM, 1993, INT J RADIAT BIOL, V63, P661, DOI 10.1080/09553009314450861
   Wassenberg MWM, 2001, J NEURO-ONCOL, V52, P73, DOI 10.1023/A:1010676807228
NR 34
TC 194
Z9 206
U1 0
U2 6
PU AMER MEDICAL ASSOC
PI CHICAGO
PA 330 N WABASH AVE, STE 39300, CHICAGO, IL 60611-5885 USA
SN 0003-9942
EI 1538-3687
J9 ARCH NEUROL-CHICAGO
JI Arch. Neurol.
PD OCT
PY 2005
VL 62
IS 10
BP 1595
EP 1600
DI 10.1001/archneur.62.10.1595
PG 6
WC Clinical Neurology
WE Science Citation Index Expanded (SCI-EXPANDED)
SC Neurosciences & Neurology
GA 973DK
UT WOS:000232502900015
PM 16216945
DA 2024-06-03
ER

PT J
AU PETERSON, K
   GORDON, KB
   HEINEMANN, MH
   DEANGELIS, LM
AF PETERSON, K
   GORDON, KB
   HEINEMANN, MH
   DEANGELIS, LM
TI THE CLINICAL SPECTRUM OF OCULAR LYMPHOMA
SO CANCER
LA English
DT Article
DE NON-HODGKIN LYMPHOMA; BRAIN NEOPLASM; OCULAR NEOPLASM; INTRAOCULAR
   LYMPHOMA; UVEITIS; VITREITIS; CENTRAL NERVOUS SYSTEM LYMPHOMA
ID RETICULUM-CELL SARCOMA; INTRAOCULAR LYMPHOMA; MALIGNANT-LYMPHOMA;
   CHEMOTHERAPY; MANAGEMENT; DIAGNOSIS; THERAPY; BRAIN; FEATURES; EYE
AB Background. Ocular lymphoma is an uncommon cause of chronic vitreitis or uveitis, often refractory to steroid treatment, and frequently representing another site of multifocal primary central nervous system lymphoma (PCNSL).
   Methods. The authors reviewed the medical and ophthalmologic records of 24 patients with ocular lymphoma; 23 had associated PCNSL.
   Results. In half, the eyes were the initial site of disease; in the others central nervous system (CNS) lymphoma developed before, or concurrent with, ocular lymphoma. Most patients had bilateral ocular symptoms; slit-lamp examination revealed asymptomatic disease in four. Vitrectomy was not always diagnostic, particularly in patients who had received steroids. Ocular ultrasound, performed on seven patients, provided an objective measure of disease and treatment response. Patients received a variety of therapeutic combinations of steroids, radiation, and chemotherapy, including high-dose cytosine arabinoside. Despite therapy, eventual ocular or CNS relapse or both was common. Thirteen patients have died, 12 with known recurrent CNS disease.
   Conclusions. Ocular lymphoma frequently is associated with PCNSL. The diagnosis should be considered in patients with steroid-resistant chronic vitreitis or uveitis. Patients with PCNSL should be carefully evaluated for ocular involvement, regardless of symptoms. Treatment can contribute to prolonged remission, but eventual ocular or CNS relapse is the rule.
C1 MEM SLOAN KETTERING CANC CTR,DEPT NEUROL,1275 YORK AVE,NEW YORK,NY 10021.
   MEM SLOAN KETTERING CANC CTR,DEPT OPHTHALMOL,NEW YORK,NY 10021.
   CORNELL UNIV,MED CTR,COLL MED,DEPT NEUROL & NEUROSCI,NEW YORK,NY 10021.
   CORNELL UNIV,MED CTR,COLL MED,DEPT OPHTHALMOL,NEW YORK,NY 10021.
C3 Memorial Sloan Kettering Cancer Center; Memorial Sloan Kettering Cancer
   Center; Cornell University; Cornell University
CR APPEN RE, 1975, ARCH OPHTHALMOL-CHIC, V93, P123
   BAUMANN MA, 1986, CANCER-AM CANCER SOC, V57, P1273, DOI 10.1002/1097-0142(19860401)57:7<1273::AID-CNCR2820570702>3.0.CO;2-C
   BOSLEY TM, 1987, BRIT J OPHTHALMOL, V71, P526, DOI 10.1136/bjo.71.7.526
   CHAR DH, 1988, OPHTHALMOLOGY, V95, P625
   CHAR DH, 1981, AM J OPHTHALMOL, V91, P480, DOI 10.1016/0002-9394(81)90236-1
   COOPER EL, 1951, AM J OPHTHALMOL, V34, P1153, DOI 10.1016/0002-9394(51)90691-5
   COX DR, 1972, J R STAT SOC B, V34, P187
   Deangelis L M, 1987, Oncology (Williston Park), V1, P52
   DEANGELIS LM, 1990, NEUROLOGY, V40, P80, DOI 10.1212/WNL.40.1.80
   EBY NL, 1988, CANCER-AM CANCER SOC, V62, P2461, DOI 10.1002/1097-0142(19881201)62:11<2461::AID-CNCR2820621135>3.0.CO;2-M
   FREEMAN LN, 1987, OPHTHALMOLOGY, V94, P1631
   GASS JDM, 1991, ARCH OPHTHALMOL-CHIC, V109, P1134, DOI 10.1001/archopht.1991.01080080094039
   HOCHBERG FH, 1988, J NEUROSURG, V68, P835, DOI 10.3171/jns.1988.68.6.0835
   KAPLAN EL, 1958, J AM STAT ASSOC, V53, P457, DOI 10.2307/2281868
   KIRMANI MH, 1987, BRIT J OPHTHALMOL, V71, P748, DOI 10.1136/bjo.71.10.748
   KLINGELE TG, 1975, AM J OPHTHALMOL, V79, P39, DOI 10.1016/0002-9394(75)90453-5
   MARGOLIS L, 1980, CANCER-AM CANCER SOC, V45, P688, DOI 10.1002/1097-0142(19800215)45:4<688::AID-CNCR2820450412>3.0.CO;2-F
   MARSTAD MK, 1973, ACTA OPHTHALMOL, V52, P211
   MICHELSON JB, 1981, ARCH OPHTHALMOL-CHIC, V99, P1409, DOI 10.1001/archopht.1981.03930020283017
   NEAULT RW, 1972, AM J OPHTHALMOL, V73, P431, DOI 10.1016/0002-9394(72)90072-4
   PARVER LM, 1979, ARCH OPHTHALMOL-CHIC, V97, P1505
   QUALMAN SJ, 1983, CANCER-AM CANCER SOC, V52, P878, DOI 10.1002/1097-0142(19830901)52:5<878::AID-CNCR2820520523>3.0.CO;2-D
   ROCKWOOD EJ, 1984, J NEUROSURG, V61, P369, DOI 10.3171/jns.1984.61.2.0369
   ROSENBAUM TJ, 1979, J NEUROSURG, V50, P660, DOI 10.3171/jns.1979.50.5.0660
   SCHAUMBURG HH, 1972, BRAIN, V95, P199, DOI 10.1093/brain/95.2.199
   SIEGEL MJ, 1989, BRIT J OPHTHALMOL, V73, P342, DOI 10.1136/bjo.73.5.342
   SIMON JW, 1980, BRIT J OPHTHALMOL, V64, P793, DOI 10.1136/bjo.64.10.793
   SLOAS HA, 1981, ARCH OPHTHALMOL-CHIC, V99, P1048
   STRAUCHEN JA, 1989, CANCER-AM CANCER SOC, V63, P1918, DOI 10.1002/1097-0142(19890515)63:10<1918::AID-CNCR2820631008>3.0.CO;2-D
   SULLIVAN SF, 1977, ANN OPHTHALMOL, V9, P401
   VOGEL MH, 1968, AM J OPHTHALMOL, V66, P205, DOI 10.1016/0002-9394(68)92065-5
   WEITER JJ, 1985, NEW ENGL J MED, V313, P436
NR 32
TC 193
Z9 203
U1 0
U2 0
PU WILEY-LISS
PI NEW YORK
PA DIV JOHN WILEY & SONS INC 605 THIRD AVE, NEW YORK, NY 10158-0012
SN 0008-543X
J9 CANCER
JI Cancer
PD AUG 1
PY 1993
VL 72
IS 3
BP 843
EP 849
DI 10.1002/1097-0142(19930801)72:3<843::AID-CNCR2820720333>3.0.CO;2-#
PG 7
WC Oncology
WE Science Citation Index Expanded (SCI-EXPANDED)
SC Oncology
GA LP720
UT WOS:A1993LP72000031
PM 8334638
DA 2024-06-03
ER

PT J
AU Gonzalez-Aguilar, A
   Idbaih, A
   Boisselier, B
   Habbita, N
   Rossetto, M
   Laurenge, A
   Bruno, A
   Jouvet, A
   Polivka, M
   Adam, C
   Figarella-Branger, D
   Miquel, C
   Vital, A
   Ghesquières, H
   Gressin, R
   Delwail, V
   Taillandier, L
   Chinot, O
   Soubeyran, P
   Gyan, E
   Choquet, S
   Houillier, C
   Soussain, C
   Tanguy, ML
   Marie, Y
   Mokhtari, K
   Hoang-Xuan, K
AF Gonzalez-Aguilar, Alberto
   Idbaih, Ahmed
   Boisselier, Blandine
   Habbita, Naima
   Rossetto, Marta
   Laurenge, Alice
   Bruno, Aurelie
   Jouvet, Anne
   Polivka, Marc
   Adam, Clovis
   Figarella-Branger, Dominique
   Miquel, Catherine
   Vital, Anne
   Ghesquieres, Herve
   Gressin, Remy
   Delwail, Vincent
   Taillandier, Luc
   Chinot, Olivier
   Soubeyran, Pierre
   Gyan, Emmanuel
   Choquet, Sylvain
   Houillier, Caroline
   Soussain, Carole
   Tanguy, Marie-Laure
   Marie, Yannick
   Mokhtari, Karima
   Hoang-Xuan, Khe
TI Recurrent Mutations of <i>MYD88</i> and <i>TBL1XR1</i> in Primary
   Central Nervous System Lymphomas
SO CLINICAL CANCER RESEARCH
LA English
DT Article
ID COMPARATIVE GENOMIC HYBRIDIZATION; B-CELL LYMPHOMA; ACUTE
   LYMPHOBLASTIC-LEUKEMIA; COPY NUMBER CHANGES; GENE-EXPRESSION; PRIMARY
   CNS; CHROMOSOMAL IMBALANCES; POOR-PROGNOSIS; IDENTIFICATION;
   INACTIVATION
AB Purpose: Our objective was to identify the genetic changes involved in primary central nervous system lymphoma (PCNSL) oncogenesis and evaluate their clinical relevance.
   Experimental Design: We investigated a series of 29 newly diagnosed, HIV-negative, PCNSL patients using high-resolution single-nucleotide polymorphism (SNP) arrays (n = 29) and whole-exome sequencing (n = 4) approaches. Recurrent homozygous deletions and somatic gene mutations found were validated by quantitative real-time PCR and Sanger sequencing, respectively. Molecular results were correlated with prognosis.
   Results: All PCNSLs were diffuse large B-cell lymphomas, and the patients received chemotherapy without radiotherapy as initial treatment. The SNP analysis revealed recurrent large and focal chromosome imbalances that target candidate genes in PCNSL oncogenesis. The most frequent genomic abnormalities were (i) 6p21.32 loss (HLA locus), (ii) 6q loss, (iii) CDKN2A homozygous deletions, (iv) 12q12-q22, and (v) chromosome 7q21 and 7q31 gains. Homozygous deletions of PRMD1, TOX, and DOCK5 and the amplification of HDAC9 were also detected. Sequencing of matched tumor and blood DNA samples identified novel somatic mutations in MYD88 and TBL1XR1 in 38% and 14% of the cases, respectively. The correlation of genetic abnormalities with clinical outcomes using multivariate analysis showed that 6q22 loss (P = 0.006 and P = 0.01) and CDKN2 Ahomozygous deletion (P = 0.02 and P = 0.01) were significantly associated with shorter progression-free survival and overall survival.
   Conclusions: Our study provides new insights into the molecular tumorigenesis of PCNSL and identifies novel genetic alterations in this disease, especially MYD88 and TBL1XR1 mutations activating the NF-kappa B signaling pathway, which may be promising targets for future therapeutic strategies. Clin Cancer Res; 18(19); 5203-11. (C)2012 AACR.
C1 [Hoang-Xuan, Khe] Grp Hosp Pitie Salpetriere, Assistance Publ Hop Paris, Serv Neurol Mazarin, Ctr Expert Natl Lymphomes Primitifs SNC Reseau LO, F-75013 Paris, France.
   [Gonzalez-Aguilar, Alberto; Idbaih, Ahmed; Boisselier, Blandine; Habbita, Naima; Rossetto, Marta; Laurenge, Alice; Bruno, Aurelie; Houillier, Caroline; Tanguy, Marie-Laure; Marie, Yannick; Mokhtari, Karima; Hoang-Xuan, Khe] Univ Paris 06, Ctr Rech Inst Cerveau & Moelle Epiniere CRICM UMR, INSERM U975, CNRS,UMR 7225, Paris, France.
   [Polivka, Marc] Ctr Hosp Univ Lariboisiere, Serv Anatomopathol, Paris, France.
   [Miquel, Catherine] Hop St Anne, Serv Anatomopathol, F-75674 Paris, France.
   [Jouvet, Anne] Hop Neurol, Hosp Civils Lyon, Bron, France.
   [Jouvet, Anne] Univ Lyon 1, UMR S842, INSERM U842, F-69365 Lyon, France.
   [Ghesquieres, Herve] Ctr Leon Berard, Dept Hematol, F-69373 Lyon, France.
   [Adam, Clovis] CHU Bicetre, Serv Anatomopathol, Le Kremlin Bicetre, France.
   [Figarella-Branger, Dominique; Chinot, Olivier] Ctr Hosp Univ La Timone, Assistance Publ Hop Marseille, Marseille, France.
   [Figarella-Branger, Dominique; Chinot, Olivier] Univ Mediterranee, CRO2, INSERM U911, Marseille, France.
   [Vital, Anne] Ctr Hosp Univ Bordeaux, Serv Pathol, Bordeaux, France.
   [Soubeyran, Pierre] Inst Bergonie, Serv Hematol, Bordeaux, France.
   [Gressin, Remy] Univ Grenoble 1, CHU Grenoble, INSERM, Inst Albert Bonniot,U823, Grenoble, France.
   [Delwail, Vincent] Ctr Hosp Univ Poitiers, Serv Hematol, Poitiers, France.
   [Taillandier, Luc] Ctr Hosp Univ Nancy, Hop Cent, Nancy, France.
   [Gyan, Emmanuel] CHU Tours, Serv Hematol & Therapie Cellulaire, CIC, Inserm U202, Tours, France.
   [Soussain, Carole] Hop Rene Huguenin, Inst Curie, Serv Hematol St Cloud, St Cloud, France.
C3 Assistance Publique Hopitaux Paris (APHP); Hopital Universitaire
   Pitie-Salpetriere - APHP; Universite Paris Cite; Hopital Universitaire
   Saint-Louis - APHP; Aix-Marseille Universite; Assistance
   Publique-Hopitaux de Marseille; Sorbonne Universite; Institut National
   de la Sante et de la Recherche Medicale (Inserm); Centre National de la
   Recherche Scientifique (CNRS); CNRS - National Institute for Biology
   (INSB); Sorbonne Universite; Assistance Publique Hopitaux Paris (APHP);
   Universite Paris Cite; Hopital Universitaire Lariboisiere-Fernand-Widal
   - APHP; Universite Paris Cite; GHU PARIS Psychiatrie Neurosciences; CHU
   Lyon; Institut National de la Sante et de la Recherche Medicale
   (Inserm); Universite Claude Bernard Lyon 1; UNICANCER; Centre Leon
   Berard; Assistance Publique Hopitaux Paris (APHP); Hopital Universitaire
   Bicetre - APHP; Aix-Marseille Universite; Assistance Publique-Hopitaux
   de Marseille; Institut National de la Sante et de la Recherche Medicale
   (Inserm); Aix-Marseille Universite; Universite de Bordeaux; CHU
   Bordeaux; UNICANCER; Institut Bergonie; Communaute Universite Grenoble
   Alpes; Universite Grenoble Alpes (UGA); Institut National de la Sante et
   de la Recherche Medicale (Inserm); CHU Grenoble Alpes; CHU Poitiers;
   Universite de Poitiers; CHU de Nancy; CHU Tours; Institut National de la
   Sante et de la Recherche Medicale (Inserm); UNICANCER; Universite PSL;
   Institut Curie; Rene Huguenin Hospital
RP Hoang-Xuan, K (corresponding author), Grp Hosp Pitie Salpetriere, Assistance Publ Hop Paris, Serv Neurol Mazarin, Ctr Expert Natl Lymphomes Primitifs SNC Reseau LO, 47-83 Blvd Hop, F-75013 Paris, France.
EM
RI Remy, GRESSIN/G-7303-2014; choquet, sylvain/O-7952-2017; González,
   Alberto/IYJ-8593-2023; BOISSELIER, Blandine/K-4377-2015; Hoang-Xuan,
   Khe/O-7527-2017; miquel, catherine/B-5536-2013; Idbaih,
   Ahmed/N-8746-2017; chinot, olivier/M-9196-2018; Soubeyran,
   Pierre-Louis/F-4523-2012
OI Remy, GRESSIN/0000-0002-9188-3501; choquet, sylvain/0000-0002-7791-0470;
   GYAN, Emmanuel/0000-0002-7651-9189; Houillier,
   Caroline/0000-0003-2931-7522; Idbaih, Ahmed/0000-0001-5290-1204; Hoang
   Xuan, Khe/0000-0003-4072-1999; Gonzalez Aguilar,
   Alberto/0000-0001-8916-6258; Laurenge, Alice/0000-0002-0058-3176;
   chinot, olivier/0000-0001-6317-9691; Delwail,
   Vincent/0000-0003-0680-1418; Soubeyran, Pierre-Louis/0000-0003-3244-7299
FU Ligue Nationale Contre le Cancer (France); ARTC (Association Pour la
   Recherche Contre les Tumeurs Cerebrales)
FX This work is part of the National Program Cartes d'Identite des Tumeurs
   (CIT) http://cit.ligue-cancer.net/ funded and developed by the Ligue
   Nationale Contre le Cancer (France). This study benefited from the LOC
   study group network (Reseau National de Centres Experts des Lymphomes
   Primitifs du SNC, INCa).; A. Gonzalez-Aguilar, B. Boisselier, N.
   Habbita, M. Rossetto, and A. Bruno are recipients of grants from the
   ARTC (Association Pour la Recherche Contre les Tumeurs Cerebrales).
CR Aliahmad P, 2008, J EXP MED, V205, P245, DOI 10.1084/jem.20071944
   Aliahmad P, 2010, NAT IMMUNOL, V11, P945, DOI 10.1038/ni.1930
   Alizadeh AA, 2000, NATURE, V403, P503, DOI 10.1038/35000501
   Bardet V, 2002, GENE CHROMOSOME CANC, V33, P178, DOI 10.1002/gcc.10014
   Beà S, 2004, J CLIN ONCOL, V22, P3498, DOI 10.1200/JCO.2004.11.025
   Booman M, 2008, J PATHOL, V216, P209, DOI 10.1002/path.2399
   Boonstra R, 2003, VIRCHOWS ARCH, V443, P164, DOI 10.1007/s00428-003-0836-9
   Braggio E, 2011, CLIN CANCER RES, V17, P4245, DOI 10.1158/1078-0432.CCR-11-0395
   Brazier H, 2006, J BONE MINER RES, V21, P1387, DOI 10.1359/JBMR.060613
   Cady FM, 2008, J CLIN ONCOL, V26, P4814, DOI 10.1200/JCO.2008.16.1455
   Calado DP, 2010, CANCER CELL, V18, P580, DOI 10.1016/j.ccr.2010.11.024
   Camilleri-Broët S, 2006, BLOOD, V107, P190, DOI 10.1182/blood-2005-03-1024
   Chen WY, 2006, BLOOD, V107, P2477, DOI 10.1182/blood-2005-07-2950
   Choi YW, 2007, INT J GYNECOL CANCER, V17, P687, DOI 10.1111/j.1525-1438.2007.00834.x
   Chu LC, 2006, INT J CANCER, V119, P2487, DOI 10.1002/ijc.22124
   Cobbers JMJL, 1998, BRAIN PATHOL, V8, P263
   Courts C, 2008, J NEUROPATH EXP NEUR, V67, P720, DOI 10.1097/NEN.0b013e31817dd02d
   Deckert M, 2011, LEUKEMIA, V25, P1797, DOI 10.1038/leu.2011.169
   Deckert M., 2007, WHO Classification of Tumours of the Central Nervous System, Vfourth, P188
   Fischer L, 2011, NEURO-ONCOLOGY, V13, P1090, DOI 10.1093/neuonc/nor107
   Gnirke A, 2009, NAT BIOTECHNOL, V27, P182, DOI 10.1038/nbt.1523
   Hayashi Y, 2001, J NEURO-ONCOL, V55, P51, DOI 10.1023/A:1012946812930
   Kadota M, 2009, CANCER RES, V69, P7357, DOI 10.1158/0008-5472.CAN-09-0064
   Kreisel F, 2011, CANCER GENET-NY, V204, P129, DOI 10.1016/j.cancergen.2010.12.010
   Lenz G, 2008, P NATL ACAD SCI USA, V105, P13520, DOI 10.1073/pnas.0804295105
   Montesinos-Rongen M, 2008, LEUKEMIA, V22, P400, DOI 10.1038/sj.leu.2405019
   Montesinos-Rongen M, 2004, BLOOD, V103, P1869, DOI 10.1182/blood-2003-05-1465
   Montesinos-Rongen M, 2010, ACTA NEUROPATHOL, V120, P529, DOI 10.1007/s00401-010-0709-7
   Montesinos-Rongen M, 2009, BLOOD, V113, P7, DOI 10.1182/blood-2008-04-149005
   Moreno DA, 2010, BRIT J HAEMATOL, V150, P665, DOI 10.1111/j.1365-2141.2010.08301.x
   Morin RD, 2011, NATURE, V476, P298, DOI 10.1038/nature10351
   Nakamura M, 2003, CANCER RES, V63, P737
   Ngo VN, 2011, NATURE, V470, P115, DOI 10.1038/nature09671
   Parker H, 2008, GENE CHROMOSOME CANC, V47, P1118, DOI 10.1002/gcc.20613
   Pasqualucci L, 2006, J EXP MED, V203, P311, DOI 10.1084/jem.20052204
   Petrie K, 2003, J BIOL CHEM, V278, P16059, DOI 10.1074/jbc.M212935200
   Puente XS, 2011, NATURE, V475, P101, DOI 10.1038/nature10113
   Riemersma SA, 2000, BLOOD, V96, P3569, DOI 10.1182/blood.V96.10.3569.h8003569_3569_3577
   Rimsza LM, 2004, BLOOD, V103, P4251, DOI 10.1182/blood-2003-07-2365
   Robledo C, 2009, CANCER-AM CANCER SOC, V115, P3728, DOI 10.1002/cncr.24430
   Rubenstein JL, 2006, BLOOD, V107, P3716, DOI 10.1182/blood-2005-03-0897
   Sadikovic B, 2009, HUM MOL GENET, V18, P1962, DOI 10.1093/hmg/ddp117
   Schwindt H, 2009, LEUKEMIA, V23, P1875, DOI 10.1038/leu.2009.120
   Shah N, 2008, EUR J CANCER, V44, P2144, DOI 10.1016/j.ejca.2008.06.044
   Sung CO, 2011, BLOOD, V117, P1291, DOI 10.1182/blood-2010-07-297861
   Tam W, 2006, BLOOD, V107, P4090, DOI 10.1182/blood-2005-09-3778
   Tun HW, 2008, BLOOD, V111, P3200, DOI 10.1182/blood-2007-10-119099
   Weber T, 2000, BRAIN PATHOL, V10, P73
   Zhang JH, 2011, BLOOD, V118, P3080, DOI 10.1182/blood-2011-03-341412
   Zhang XM, 2006, BMC CELL BIOL, V7, DOI 10.1186/1471-2121-7-31
NR 50
TC 188
Z9 197
U1 0
U2 17
PU AMER ASSOC CANCER RESEARCH
PI PHILADELPHIA
PA 615 CHESTNUT ST, 17TH FLOOR, PHILADELPHIA, PA 19106-4404 USA
SN 1078-0432
EI 1557-3265
J9 CLIN CANCER RES
JI Clin. Cancer Res.
PD OCT 1
PY 2012
VL 18
IS 19
BP 5203
EP 5211
DI 10.1158/1078-0432.CCR-12-0845
PG 9
WC Oncology
WE Science Citation Index Expanded (SCI-EXPANDED)
SC Oncology
GA 048JF
UT WOS:000311906600008
PM 22837180
OA Green Submitted
DA 2024-06-03
ER

PT J
AU Schabet, M
AF Schabet, M
TI Epidemiology of primary CNS lymphoma
SO JOURNAL OF NEURO-ONCOLOGY
LA English
DT Article; Proceedings Paper
CT UICC International Symposium on Primary CNS Lymphoma
CY OCT   18, 1997
CL TUBINGEN, GERMANY
DE epidemiology; non-Hodgkin's lymphoma; primary CNS lymphoma
ID CENTRAL-NERVOUS-SYSTEM; ACQUIRED-IMMUNODEFICIENCY-SYNDROME; INCREASING
   INCIDENCE; RADIATION-THERAPY; CANCER; AIDS; DISORDERS; VIRUS
AB In the beginning of the nineties the National Cancer Institute Surveillance, Epidemiology, and End Results Program calculated the incidence of primary central nervous system non-Hodgkin's lymphoma (PCNSL) as 1:100 000. The incidence of PCNSL has been increasing since the seventies in immunocompetent patients. The main increase, however, is taking place since the mid-eighties and is due to the increase of immunodeficieny and immunosuppression. The risk is 2-6% in AIDS patients according to clinical data and will probably further increase with the length of survival in these patients. Transplant patients carry a risk of 1-5% to develop a PCNSL. The risk is 1-2% for renal, and 2-7% for cardiac, lung or liver transplant recipients. Patients with congenital immune deficiency have a risk of 4%. PCNSL may also present as a secondary malignancy.
C1 Univ Tubingen, Dept Neurol, D-72076 Tubingen, Germany.
C3 Eberhard Karls University of Tubingen
RP Schabet, M (corresponding author), Univ Tubingen, Dept Neurol, Hoppe Seyler Str 3, D-72076 Tubingen, Germany.
CR Ahsan H, 1995, INT J EPIDEMIOL, V24, P1078, DOI 10.1093/ije/24.6.1078
   BAUMGARTNER JE, 1990, J NEUROSURG, V73, P206, DOI 10.3171/jns.1990.73.2.0206
   Boubenider S, 1997, J NEPHROL, V10, P136
   Confavreux C, 1996, NEUROLOGY, V46, P1607, DOI 10.1212/WNL.46.6.1607
   DEANGELIS LM, 1991, CANCER-AM CANCER SOC, V67, P1431, DOI 10.1002/1097-0142(19910301)67:5<1431::AID-CNCR2820670527>3.0.CO;2-K
   EBY NL, 1988, CANCER-AM CANCER SOC, V62, P2461, DOI 10.1002/1097-0142(19881201)62:11<2461::AID-CNCR2820621135>3.0.CO;2-M
   FERRACINI R, 1993, PATHOL RES PRACT, V189, P249, DOI 10.1016/S0344-0338(11)80507-8
   FILIPOVICH AH, 1987, AM J PEDIAT HEMATOL, V9, P183
   FINE HA, 1993, ANN INTERN MED, V119, P1093, DOI 10.7326/0003-4819-119-11-199312010-00007
   FORMENTI SC, 1989, CANCER-AM CANCER SOC, V63, P1101, DOI 10.1002/1097-0142(19890315)63:6<1101::AID-CNCR2820630611>3.0.CO;2-R
   Goplen AK, 1997, J ACQ IMMUN DEF SYND, V14, P351, DOI 10.1097/00042560-199704010-00007
   HOCHBERG FH, 1988, J NEUROSURG, V68, P835, DOI 10.3171/jns.1988.68.6.0835
   JELLINGER KA, 1995, J NEURO-ONCOL, V24, P33, DOI 10.1007/BF01052655
   KINLEN LJ, 1985, AM J MED, V78, P44, DOI 10.1016/0002-9343(85)90245-1
   LEVY RM, 1988, J ACQ IMMUN DEF SYND, V1, P31
   LING SM, 1994, CANCER-AM CANCER SOC, V73, P2570, DOI 10.1002/1097-0142(19940515)73:10<2570::AID-CNCR2820731019>3.0.CO;2-1
   MACMAHON EME, 1991, LANCET, V338, P969, DOI 10.1016/0140-6736(91)91837-K
   ONEILL BP, 1993, ANN NEUROL, V34, P313
   OSULLIVAN MG, 1991, LANCET, V338, P895, DOI 10.1016/0140-6736(91)91561-8
   PATCHELL RA, 1988, NEUROL CLIN, V6, P297, DOI 10.1016/S0733-8619(18)30871-5
   PENN I, 1995, TRANSPLANTATION, V59, P240, DOI 10.1097/00007890-199501270-00016
   Penn I, 1996, Liver Transpl Surg, V2, P52, DOI 10.1002/lt.500020109
   PENN I, 1993, J HEART LUNG TRANSPL, V12, P328
   PERRY GS, 1980, J PEDIATR-US, V97, P72, DOI 10.1016/S0022-3476(80)80133-8
   Reni M, 1997, ANN ONCOL, V8, P227, DOI 10.1023/A:1008201717089
   ROSENBLUM ML, 1988, ANN NEUROL S, V23, P13
NR 26
TC 170
Z9 185
U1 0
U2 5
PU KLUWER ACADEMIC PUBL
PI DORDRECHT
PA SPUIBOULEVARD 50, PO BOX 17, 3300 AA DORDRECHT, NETHERLANDS
SN 0167-594X
J9 J NEURO-ONCOL
JI J. Neuro-Oncol.
PD JUL
PY 1999
VL 43
IS 3
BP 199
EP 201
DI 10.1023/A:1006290032052
PG 3
WC Oncology; Clinical Neurology
WE Conference Proceedings Citation Index - Science (CPCI-S); Science Citation Index Expanded (SCI-EXPANDED)
SC Oncology; Neurosciences & Neurology
GA 239YQ
UT WOS:000082798500002
PM 10563423
DA 2024-06-03
ER

PT J
AU Shiels, MS
   Pfeiffer, RM
   Besson, C
   Clarke, CA
   Morton, LM
   Nogueira, L
   Pawlish, K
   Yanik, EL
   Suneja, G
   Engels, EA
AF Shiels, Meredith S.
   Pfeiffer, Ruth M.
   Besson, Caroline
   Clarke, Christina A.
   Morton, Lindsay M.
   Nogueira, Leticia
   Pawlish, Karen
   Yanik, Elizabeth L.
   Suneja, Gita
   Engels, Eric A.
TI Trends in primary central nervous system lymphoma incidence and survival
   in the US
SO BRITISH JOURNAL OF HAEMATOLOGY
LA English
DT Article
DE Lymphoma; central nervous system; incidence; survival; human
   immunodeficiency virus; transplant
ID PRIMARY CNS LYMPHOMA; NON-HODGKINS-LYMPHOMA; UNITED-STATES; CANCER-RISK;
   IMMUNOCOMPETENT PATIENTS; HIV; SURVEILLANCE; PEOPLE; AIDS
AB It is suspected that primary central nervous system lymphoma (PCNSL) rates are increasing among immunocompetent people. We estimated PCNSL trends in incidence and survival among immunocompetent persons by excluding cases among human immunodeficiency virus (HIV)-infected persons and transplant recipients. PCNSL data were derived from 10 Surveillance, Epidemiology and End Results (SEER) cancer registries (1992-2011). HIV-infected cases had reported HIV infection or death due to HIV. Transplant recipient cases were estimated from the Transplant Cancer Match Study. We estimated PCNSL trends overall and among immunocompetent individuals, and survival by HIV status. A total of 4158 PCNSLs were diagnosed (36% HIV-infected; 09% transplant recipients). HIV prevalence in PCNSL cases declined from 641% (1992-1996) to 127% (2007-2011), while the prevalence of transplant recipients remained low. General population PCNSL rates were strongly influenced by immunosuppressed cases, particularly in 20-39 year-old men. Among immunocompetent people, PCNSL rates in men and women aged 65+ years increased significantly (17% and 16%/year), but remained stable in other age groups. Five-year survival was poor, particularly among HIV-infected cases (90%). Among HIV-uninfected cases, 5-year survival increased from 191% (1992-1994) to 301% (2004-2006). In summary, PCNSL rates have increased among immunocompetent elderly adults, but not in younger individuals. Survival remains poor for both HIV-infected and HIV-uninfected PCNSL patients.
C1 [Shiels, Meredith S.; Pfeiffer, Ruth M.; Morton, Lindsay M.; Yanik, Elizabeth L.; Engels, Eric A.] NCI, Div Canc Epidemiol & Genet, Rockville, MD USA.
   [Besson, Caroline] Bicetre Univ Hosp, Le Kremlin Bicetre, France.
   [Clarke, Christina A.] Canc Prevent Inst Calif, Fremont, CA USA.
   [Nogueira, Leticia] Texas Dept State Hlth Serv, Austin, TX USA.
   [Pawlish, Karen] New Jersey Dept Hlth, Trenton, NJ USA.
   [Suneja, Gita] Univ Utah, Dept Radiat Oncol, Salt Lake City, UT USA.
C3 National Institutes of Health (NIH) - USA; NIH National Cancer Institute
   (NCI); Assistance Publique Hopitaux Paris (APHP); Hopital Universitaire
   Bicetre - APHP; Cancer Prevention Institute of California; Texas
   Department of State Health Services; New Jersey Department of Health &
   Senior Services; Utah System of Higher Education; University of Utah
RP Shiels, MS (corresponding author), NCI, NIH, 9609 Med Ctr Dr,Room 6E-218 MSC 9767, Bethesda, MD 20892 USA.
EM
RI Nogueira, Leticia/J-2547-2019; Morton, Lindsay/AAE-7495-2022; Yanik,
   Elizabeth L./ABF-7130-2021; Besson, Caroline/H-4648-2018
OI Morton, Lindsay/0000-0001-9767-2310; Yanik, Elizabeth
   L./0000-0002-5835-0201; Besson, Caroline/0000-0003-4364-7173
FU Intramural Research Program of the National Cancer Institute;
   Minneapolis Medical Research Foundation, Minneapolis, MN
   [HHSH250201000018C]; Arbor Research Collaborative for Health in Ann
   Arbor, MI [HHSH234200537009C]; SEER Program of the National Cancer
   Institute: California [HHSN261201000036C, HHSN261201000035C,
   HHSN261201000034C]; SEER Program of the National Cancer Institute:
   Connecticut [HHSN261201000024C]; SEER Program of the National Cancer
   Institute: Hawaii [HHSN261201 000037C, N01-PC-35137, N01-PC-35139]; SEER
   Program of the National Cancer Institute: Iowa [HSN 261201000032C,
   N01-PC-35143]; SEER Program of the National Cancer Institute: New Jersey
   [HHSN261201300021I, N01-PC-2013-00021]; SEER Program of the National
   Cancer Institute: Seattle-Puget Sound [N01-PC-35142]; SEER Program of
   the National Cancer Institute: Utah [HHSN2612013000171]; National
   Program of Cancer Registries of the Centers for Disease Control and
   Prevention [1U58 DP000807-01, U58 DP000848-04, 5U58DP003875-01,
   5U58DP003883-03, U58DP12-1205 3919-03, 5U58DP003921-03,
   5U58/DP003931-02, U58DP 003879, U58DP000832, 5U58DP000824-04];
   Massachusetts Cancer Prevention and Control Cooperative Agreement
   [5458DP003920]; New Jersey (Memorandum of Agreement)
   [5-100-046-4220-496-6140]; University of Utah; Fred Hutchinson Cancer
   Research Center in Seattle, WA
FX This work was supported by the Intramural Research Program of the
   National Cancer Institute.; The SRTR is currently operated under
   contract number HHSH250201000018C (Health Resources and Services
   Administration) by the Minneapolis Medical Research Foundation,
   Minneapolis, MN. During the initial period when registry linkages were
   performed, the SRTR was managed by Arbor Research Collaborative for
   Health in Ann Arbor, MI (contract HHSH234200537009C). The following
   cancer registries were supported by the SEER Program of the National
   Cancer Institute: California (contracts HHSN261201000036C,
   HHSN261201000035C, and HHSN261201000034C), Connecticut
   (HHSN261201000024C), Hawaii (HHSN261201 000037C, N01-PC-35137, and
   N01-PC-35139), Iowa (HSN 261201000032C and N01-PC-35143), New Jersey
   (HHSN261201300021I, N01-PC-2013-00021), Seattle-Puget Sound
   (N01-PC-35142), and Utah (HHSN2612013000171). The following cancer
   registries were supported by the National Program of Cancer Registries
   of the Centers for Disease Control and Prevention: California (agreement
   1U58 DP000807-01), Colorado (U58 DP000848-04), Georgia
   (5U58DP003875-01), Illinois (5U58DP003883-03), Maryland (U58DP12-1205
   3919-03), Michigan (5U58DP003921-03), New Jersey (5U58/ DP003931-02),
   New York (U58DP 003879), North Carolina (U58DP000832) and Texas
   (5U58DP000824-04). Additional support was provided by the states of
   California, Colorado, Connecticut, Illinois, Iowa, Massachusetts
   (Massachusetts Cancer Prevention and Control Cooperative Agreement
   5458DP003920), New Jersey (Memorandum of Agreement
   5-100-046-4220-496-6140), New York (including the Cancer Surveillance
   Initiative), Texas, Utah, and Washington, as well as the University of
   Utah and Fred Hutchinson Cancer Research Center in Seattle, WA.
CR Abrey LE, 2009, CURR OPIN NEUROL, V22, P675, DOI 10.1097/WCO.0b013e328332533b
   [Anonymous], 1992, MMWR Recomm. Rep, V41
   [Anonymous], GUIDELINES USE ANTIR
   [Anonymous], OPTN SRTR 2012 ANN D
   [Anonymous], 2013, HIV SURV REP 2011
   Bayraktar S, 2011, J NEURO-ONCOL, V101, P257, DOI 10.1007/s11060-010-0252-3
   Buell JF, 2005, TRANSPL P, V37, P954, DOI 10.1016/j.transproceed.2004.12.130
   Chimienti E, 2009, CLIN LYMPHOMA MYELOM, V9, P353, DOI 10.3816/CLM.2009.n.070
   Clarke CA, 2013, BRIT J CANCER, V109, P280, DOI 10.1038/bjc.2013.294
   Clarke CA, 2004, JAIDS-J ACQ IMM DEF, V36, P1083, DOI 10.1097/00126334-200408150-00012
   Diamond C, 2007, CANCER CAUSE CONTROL, V18, P135, DOI 10.1007/s10552-006-0096-5
   Engels EA, 2008, INT J CANCER, V123, P187, DOI 10.1002/ijc.23487
   Engels EA, 2006, AIDS, V20, P1645, DOI 10.1097/01.aids.0000238411.75324.59
   Engels EA, 2011, JAMA-J AM MED ASSOC, V306, P1891, DOI 10.1001/jama.2011.1592
   Evens AM, 2013, AM J TRANSPLANT, V13, P1512, DOI 10.1111/ajt.12211
   Ferreri AJM, 2011, BLOOD, V118, P510, DOI 10.1182/blood-2011-03-321349
   Fritz A.G., 2013, International Classification of Diseases for Oncology (ICD-O)
   Gibson Todd M, 2014, AIDS, V28, P2313, DOI 10.1097/QAD.0000000000000428
   Gloghini A, 2013, SEMIN CANCER BIOL, V23, P457, DOI 10.1016/j.semcancer.2013.08.004
   Hoang-Xuan K, 2015, LANCET ONCOL, V16, pE322, DOI 10.1016/S1470-2045(15)00076-5
   Kim HJ, 2000, STAT MED, V19, P335, DOI 10.1002/(SICI)1097-0258(20000215)19:3<335::AID-SIM336>3.0.CO;2-Z
   National Comprehensive Cancer Network I, 2015, NCCN GUID VERS 1 201
   Norden AD, 2011, J NEURO-ONCOL, V101, P487, DOI 10.1007/s11060-010-0269-7
   O'Neill BP, 2013, AM J HEMATOL, V88, P997, DOI 10.1002/ajh.23551
   Olson JE, 2002, CANCER, V95, P1504, DOI 10.1002/cncr.10851
   Shiels MS, 2013, CANCER EPIDEM BIOMAR, V22, P1069, DOI 10.1158/1055-9965.EPI-13-0040
   Shiels MS, 2011, JAMA-J AM MED ASSOC, V305, P1450, DOI 10.1001/jama.2011.396
   Surveillance Epidemiology and End Results (SEER) Program, 2014, INC SEER 13 REGS RES
   Vardiman JW., 2008, WHO CLASSIFICATION T
   Villano JL, 2011, BRIT J CANCER, V105, P1414, DOI 10.1038/bjc.2011.357
NR 30
TC 168
Z9 176
U1 0
U2 6
PU WILEY-BLACKWELL
PI HOBOKEN
PA 111 RIVER ST, HOBOKEN 07030-5774, NJ USA
SN 0007-1048
EI 1365-2141
J9 BRIT J HAEMATOL
JI Br. J. Haematol.
PD AUG
PY 2016
VL 174
IS 3
BP 417
EP 424
DI 10.1111/bjh.14073
PG 8
WC Hematology
WE Science Citation Index Expanded (SCI-EXPANDED)
SC Hematology
GA DS4CO
UT WOS:000380729200008
PM 27018254
OA Green Accepted, Bronze
DA 2024-06-03
ER

PT J
AU Angelov, L
   Doolittle, ND
   Kraemer, DF
   Siegal, T
   Barnett, GH
   Peereboom, DM
   Stevens, G
   McGregor, J
   Jahnke, K
   Lacy, CA
   Hedrick, NA
   Shalom, E
   Ference, S
   Bell, S
   Sorenson, L
   Tyson, RM
   Haluska, M
   Neuwelt, EA
AF Angelov, Lilyana
   Doolittle, Nancy D.
   Kraemer, Dale F.
   Siegal, Tali
   Barnett, Gene H.
   Peereboom, David M.
   Stevens, Glen
   McGregor, John
   Jahnke, Kristoph
   Lacy, Cynthia A.
   Hedrick, Nancy A.
   Shalom, Edna
   Ference, Sandra
   Bell, Susan
   Sorenson, Lisa
   Tyson, Rose Marie
   Haluska, Marianne
   Neuwelt, Edward A.
TI Blood-Brain Barrier Disruption and Intra-Arterial Methotrexate-Based
   Therapy for Newly Diagnosed Primary CNS Lymphoma: A Multi-Institutional
   Experience
SO JOURNAL OF CLINICAL ONCOLOGY
LA English
DT Article; Proceedings Paper
CT 43rd Annual Meeting of the American-Society-of-Clinical-Oncology (ASCO)
CY JUN 01-05, 2007
CL Chicago, IL
SP Amer Soc Clin Oncol (ASCO)
ID NERVOUS-SYSTEM LYMPHOMA; HIGH-DOSE METHOTREXATE; TERM-FOLLOW-UP;
   PHASE-II; CHEMOTHERAPY DELIVERY; EUROPEAN ORGANIZATION; COGNITIVE
   FUNCTIONS; RADIOTHERAPY; MULTICENTER; SURVIVAL
AB Purpose Primary CNS lymphoma (PCNSL) is confined to the CNS and/or the eyes at presentation and is usually initially treated with intravenous methotrexate-based chemotherapy and whole-brain radiotherapy (WBRT). However, the intact blood-brain barrier (BBB) can limit diffusion of methotrexate into brain and tumor. With BBB disruption (BBBD), enhanced drug delivery to the tumor can be achieved.
   Patients and Methods This report summarizes the multi-institutional experience of 149 newly diagnosed (with no prior WBRT) patients with PCNSL treated with osmotic BBBD and intra-arterial (IA) methotrexate at four institutions from 1982 to 2005. In this series, 47.6% of patients were age >= 60 years, and 42.3% had Karnofsky performance score (KPS) less than 70 at diagnosis.
   Results The overall response rate was 81.9% (57.8% complete; 24.2% partial). Median overall survival (OS) was 3.1 years (25% estimated survival at 8.5 years). Median progression-free survival (PFS) was 1.8 years, with 5-year PFS of 31% and 7-year PFS of 25%. In low-risk patients (age >= 60 years and KPS >= 70), median OS was approximately 14 years, with a plateau after approximately 8 years. Procedures were generally well tolerated; focal seizures (9.2%) were the most frequent side effect and lacked long-term sequelae.
   Conclusion This large series of patients treated over a 23-year period demonstrates that BBBD/IA methotrexate-based chemotherapy results in successful and durable tumor control and outcomes that are comparable or superior to other PCNSL treatment regimens.
C1 Cleveland Clin, Brain Tumor & Neuro Oncol Ctr, Cleveland, OH USA.
   Cleveland Clin, Dept Neurosurg, Cleveland, OH USA.
   Cleveland Clin, Dept Hematol, Cleveland, OH USA.
   Cleveland Clin, Dept Med Oncol, Cleveland, OH USA.
   Ohio State Univ, Med Ctr, Dept Neurosurg, Columbus, OH 43210 USA.
   Oregon Hlth & Sci Univ, Dept Neurol, Dept Med Informat & Clin Epidemiol, Portland, OR 97201 USA.
   Oregon Hlth & Sci Univ, Dept Publ Hlth, Dept Med Informat & Clin Epidemiol, Portland, OR 97201 USA.
   Oregon Hlth & Sci Univ, Dept Prevent Med, Dept Med Informat & Clin Epidemiol, Portland, OR 97201 USA.
   Oregon State Univ, Dept Pharm Practice, Corvallis, OR 97331 USA.
   Portland VA Med Ctr, Portland, OR USA.
   Hadassah Hebrew Univ Hosp, Leslie & Michael Gaffin Ctr Neurooncol, Jerusalem, Israel.
C3 Cleveland Clinic Foundation; Cleveland Clinic Foundation; Cleveland
   Clinic Foundation; Cleveland Clinic Foundation; University System of
   Ohio; Ohio State University; Oregon Health & Science University; Oregon
   Health & Science University; Oregon Health & Science University; Oregon
   State University; US Department of Veterans Affairs; Veterans Health
   Administration (VHA); Portland VA Medical Center; Hebrew University of
   Jerusalem; Hadassah University Medical Center
RP Neuwelt, EA (corresponding author), 3181 SW Sam Jackson Pk Rd,Mailcode L603, Portland, OR 97239 USA.
EM
RI McGregor, John Michael/K-6870-2013
FU NCI NIH HHS [CA137488, R01 CA137488-15, R13 CA086959, R01 CA137488, R13
   CA086959-10] Funding Source: Medline; NINDS NIH HHS [NS34608, R01
   NS044687, R01 NS034608, NS44687, R37 NS044687] Funding Source: Medline
CR Abrey LE, 2006, J CLIN ONCOL, V24, P5711, DOI 10.1200/JCO.2006.08.2941
   Abrey LE, 1998, J CLIN ONCOL, V16, P859, DOI 10.1200/JCO.1998.16.3.859
   Abrey LE, 2000, J CLIN ONCOL, V18, P3144, DOI 10.1200/JCO.2000.18.17.3144
   Batchelor T, 2006, J CLIN ONCOL, V24, P1281, DOI 10.1200/JCO.2005.04.8819
   Batchelor T, 2003, J CLIN ONCOL, V21, P1044, DOI 10.1200/JCO.2003.03.036
   Blay JY, 1998, J CLIN ONCOL, V16, P864, DOI 10.1200/JCO.1998.16.3.864
   Cohen IJ, 2004, J PEDIAT HEMATOL ONC, V26, P156, DOI 10.1097/00043426-200403000-00004
   Correa DD, 2007, ANN ONCOL, V18, P1145, DOI 10.1093/annonc/mdl464
   Correa DD, 2004, NEUROLOGY, V62, P548, DOI 10.1212/01.WNL.0000109673.75316.D8
   CROSSEN JR, 1992, NEUROSURGERY, V30, P23, DOI 10.1227/00006123-199201000-00005
   CROSSEN JR, 1994, J CLIN ONCOL, V12, P627, DOI 10.1200/JCO.1994.12.3.627
   Dahlborg S A, 1996, Cancer J Sci Am, V2, P166
   DEANGELIS LM, 1992, J CLIN ONCOL, V10, P635, DOI 10.1200/JCO.1992.10.4.635
   DeAngelis LM, 2002, J CLIN ONCOL, V20, P4643, DOI 10.1200/JCO.2002.11.013
   Doolittle ND, 2008, BLOOD, V111, P1085, DOI 10.1182/blood-2007-07-101402
   Doolittle ND, 2007, LEUKEMIA LYMPHOMA, V48, P1712, DOI 10.1080/10428190701493902
   Doolittle ND, 2000, CANCER, V88, P637, DOI 10.1002/(SICI)1097-0142(20000201)88:3<637::AID-CNCR22>3.0.CO;2-Y
   Ferreri AJM, 2003, J CLIN ONCOL, V21, P2407, DOI 10.1200/JCO.2003.01.135
   Ferreri AJM, 2003, J CLIN ONCOL, V21, P266, DOI 10.1200/JCO.2003.09.139
   Feugier P, 2004, ANN ONCOL, V15, P129, DOI 10.1093/annonc/mdh013
   Gavrilovic IT, 2006, J CLIN ONCOL, V24, P4570, DOI 10.1200/JCO.2006.06.6910
   GLASS J, 1994, J NEUROSURG, V81, P188, DOI 10.3171/jns.1994.81.2.0188
   Harder H, 2004, NEUROLOGY, V62, P544, DOI 10.1212/WNL.62.4.544
   Herrlinger U, 2002, ANN NEUROL, V51, P247, DOI 10.1002/ana.10102
   Hoang-Xuan K, 2003, J CLIN ONCOL, V21, P2726, DOI 10.1200/JCO.2003.11.036
   Jahnke Kristoph, 2006, Neurosurg Focus, V21, pE11
   Lai R, 2004, NEUROLOGY, V62, P451, DOI 10.1212/01.WNL.0000106941.51340.A2
   MACDONALD DR, 1990, J CLIN ONCOL, V8, P1277, DOI 10.1200/JCO.1990.8.7.1277
   Marchi N, 2007, EPILEPSIA, V48, P732, DOI 10.1111/j.1528-1167.2007.00988.x
   McAllister LD, 2000, NEUROSURGERY, V46, P51, DOI 10.1093/neurosurgery/46.1.51
   Muldoon LL, 2007, J CLIN ONCOL, V25, P2295, DOI 10.1200/JCO.2006.09.9861
   Neuwelt EA, 2005, AM J NEURORADIOL, V26, P258
   NEUWELT EA, 1991, J CLIN ONCOL, V9, P1580, DOI 10.1200/JCO.1991.9.9.1580
   NEUWELT EA, 1981, ANN INTERN MED, V94, P449, DOI 10.7326/0003-4819-94-4-449
   Neuwelt E, 2008, LANCET NEUROL, V7, P84, DOI 10.1016/S1474-4422(07)70326-5
   O'Brien P, 2000, J CLIN ONCOL, V18, P519, DOI 10.1200/JCO.2000.18.3.519
   Omuro AMP, 2005, NEUROLOGY, V64, P69, DOI 10.1212/01.WNL.0000148641.98241.5E
   Omuro AMP, 2005, ARCH NEUROL-CHICAGO, V62, P1595, DOI 10.1001/archneur.62.10.1595
   Panageas KS, 2005, CANCER-AM CANCER SOC, V104, P2466, DOI 10.1002/cncr.21481
   Pels H, 2003, J CLIN ONCOL, V21, P4489, DOI 10.1200/JCO.2003.04.056
   Poortmans PMP, 2003, J CLIN ONCOL, V21, P4483, DOI 10.1200/JCO.2003.03.108
   ROMANGOLDSTEIN S, 1995, AM J NEURORADIOL, V16, P543
   Rubenstein JL, 2003, BLOOD, V101, P466, DOI 10.1182/blood-2002-06-1636
   Shah GD, 2007, J CLIN ONCOL, V25, P4730, DOI 10.1200/JCO.2007.12.5062
   Smith JR, 2002, OPHTHALMOLOGY, V109, P1709, DOI 10.1016/S0161-6420(02)01125-9
   Tyson RM, 2003, LEUKEMIA LYMPHOMA, V44, P627, DOI 10.1080/1042819021000055057
NR 46
TC 167
Z9 188
U1 0
U2 5
PU AMER SOC CLINICAL ONCOLOGY
PI ALEXANDRIA
PA 2318 MILL ROAD, STE 800, ALEXANDRIA, VA 22314 USA
SN 0732-183X
EI 1527-7755
J9 J CLIN ONCOL
JI J. Clin. Oncol.
PD JUL 20
PY 2009
VL 27
IS 21
BP 3503
EP 3509
DI 10.1200/JCO.2008.19.3789
PG 7
WC Oncology
WE Science Citation Index Expanded (SCI-EXPANDED); Conference Proceedings Citation Index - Science (CPCI-S)
SC Oncology
GA 471II
UT WOS:000268049100016
PM 19451444
OA Green Published, Bronze
DA 2024-06-03
ER

PT J
AU Freilich, RJ
   Delattre, JY
   Monjour, A
   DeAngelis, LM
AF Freilich, RJ
   Delattre, JY
   Monjour, A
   DeAngelis, LM
TI Chemotherapy without radiation therapy as initial treatment for primary
   CNS lymphoma in older patients
SO NEUROLOGY
LA English
DT Article; Proceedings Paper
CT 47th Annual Meeting of the American-Academy-of-Neurology
CY MAY, 1995
CL SEATTLE, WA
SP Amer Acad Neurol
ID CENTRAL NERVOUS-SYSTEM; PRIMARY BRAIN LYMPHOMA; SURVIVAL; MANAGEMENT
AB Chemotherapy plus radiation therapy (RT) for primary CNS lymphoma (PCNSL) has significantly improved patient survival over RT alone, but there are late neurologic sequelae of RT, particularly in the elderly. We treated 13 patients over age 50 years (mean age 74 years) with chemotherapy alone as initial treatment for PCNSL. All received methotrexate (MTX) and procarbazine; in addition, five received thiotepa, four vincristine, and four vincristine and cytarabine. Ten achieved a complete response (CR), 2 a partial response (PR), and 1 progressed through treatment. Two patients with ocular lymphoma responded to MTX, procarbazine, and vincristine. Four of six patients who relapsed after achieving a CR or PR were treated with additional chemotherapy or RT; three achieved a CR and one a PR. Five patients remain in CR at 7.5 to 30 months, one is alive at 35 months but with progressive disease, six died of PCNSL at 5 to 30.5 months, and one died in CR of sulfur allergy 2 months after diagnosis. The Karnofsky Performance Status improved in 11 of 13 patients with treatment. Cognitive deficits were present in nine patients at diagnosis and improved in eight of these nine after chemotherapy. Only one patient developed new cognitive deficits, due to progressive tumor and possibly MTX leukoencephalopathy. Chemotherapy alone for PCNSL is effective in the elderly and eliminates the risk of RT-related neurotoxicity. RT can salvage those who relapse after chemotherapy.
C1 MEM SLOAN KETTERING CANC CTR,DEPT NEUROL,NEUROL SERV,NEW YORK,NY 10021.
   HOP LA PITIE SALPETRIERE,DEPT NEUROL,PARIS,FRANCE.
   UNIV STRASBOURG 1,DEPT NEUROL,COLMAR,FRANCE.
C3 Memorial Sloan Kettering Cancer Center; Sorbonne Universite; Assistance
   Publique Hopitaux Paris (APHP); Hopital Universitaire Pitie-Salpetriere
   - APHP; Universites de Strasbourg Etablissements Associes; Universite de
   Strasbourg
OI DeAngelis, Lisa/0000-0002-6290-7818
CR BOIARDI A, 1993, J NEUROL, V241, P96, DOI 10.1007/BF00869771
   COHEN IJ, 1986, CANCER-AM CANCER SOC, V57, P6, DOI 10.1002/1097-0142(19860101)57:1<6::AID-CNCR2820570104>3.0.CO;2-B
   Deangelis L M, 1987, Oncology (Williston Park), V1, P52
   DEANGELIS LM, 1989, NEUROLOGY, V39, P789, DOI 10.1212/WNL.39.6.789
   DEANGELIS LM, 1992, J CLIN ONCOL, V10, P635, DOI 10.1200/JCO.1992.10.4.635
   EBY NL, 1988, CANCER-AM CANCER SOC, V62, P2461, DOI 10.1002/1097-0142(19881201)62:11<2461::AID-CNCR2820621135>3.0.CO;2-M
   GLASS J, 1994, J NEUROSURG, V81, P188, DOI 10.3171/jns.1994.81.2.0188
   HENRY JM, 1974, CANCER, V34, P1293, DOI 10.1002/1097-0142(197410)34:4<1293::AID-CNCR2820340441>3.0.CO;2-P
   KAWAKAMI Y, 1985, J NEUROSURG, V62, P522, DOI 10.3171/jns.1985.62.4.0522
   LOEFFLER JS, 1985, J CLIN ONCOL, V3, P490, DOI 10.1200/JCO.1985.3.4.490
   MARGOLIS L, 1980, CANCER-AM CANCER SOC, V45, P688, DOI 10.1002/1097-0142(19800215)45:4<688::AID-CNCR2820450412>3.0.CO;2-F
   MCLAUGHLIN P, 1988, JNCI-J NATL CANCER I, V80, P1408, DOI 10.1093/jnci/80.17.1408
   MILLER DC, 1994, CANCER-AM CANCER SOC, V74, P1383, DOI 10.1002/1097-0142(19940815)74:4<1383::AID-CNCR2820740432>3.0.CO;2-1
   MURRAY K, 1986, J NEUROSURG, V65, P600, DOI 10.3171/jns.1986.65.5.0600
   NELSON DF, 1992, INT J RADIAT ONCOL, V23, P9, DOI 10.1016/0360-3016(92)90538-S
   NEUWELT EA, 1991, J CLIN ONCOL, V9, P1580, DOI 10.1200/JCO.1991.9.9.1580
   PETERSON K, 1993, CANCER, V72, P843, DOI 10.1002/1097-0142(19930801)72:3<843::AID-CNCR2820720333>3.0.CO;2-#
   SCHULTZ C, 1994, P AN M AM SOC CLIN, V13, P174
   STRAUCHEN JA, 1989, CANCER-AM CANCER SOC, V63, P1918, DOI 10.1002/1097-0142(19890515)63:10<1918::AID-CNCR2820631008>3.0.CO;2-D
NR 19
TC 166
Z9 174
U1 0
U2 1
PU LITTLE BROWN CO
PI BOSTON
PA 34 BEACON STREET, BOSTON, MA 02108-1493
SN 0028-3878
J9 NEUROLOGY
JI Neurology
PD FEB
PY 1996
VL 46
IS 2
BP 435
EP 439
DI 10.1212/WNL.46.2.435
PG 5
WC Clinical Neurology
WE Conference Proceedings Citation Index - Science (CPCI-S); Science Citation Index Expanded (SCI-EXPANDED)
SC Neurosciences & Neurology
GA TZ712
UT WOS:A1996TZ71200024
PM 8614508
DA 2024-06-03
ER

PT J
AU Hartmann, M
   Heiland, S
   Harting, I
   Tronnier, VM
   Sommer, C
   Ludwig, R
   Sartor, K
AF Hartmann, M
   Heiland, S
   Harting, I
   Tronnier, VM
   Sommer, C
   Ludwig, R
   Sartor, K
TI Distinguishing of primary cerebral lymphoma from high-grade glioma with
   perfusion-weighted magnetic resonance imaging
SO NEUROSCIENCE LETTERS
LA English
DT Article
DE lymphoma; cerebral glioma; perfusion-weighted magnetic resonance imaging
ID BLOOD-BRAIN-BARRIER; NERVOUS-SYSTEM LYMPHOMAS; PRIMARY CNS LYMPHOMA;
   VOLUME MAPS; MR; LESIONS
AB To assess the usefulness of perfusion-weighted echo-planar magnetic resonance imaging in the differential diagnosis of primary supratentorial lymphoma (PCNSL) and glioblastoma (GBM), 12 patients with a PCNSL and 12 with a GBM were examined using a 1.5 T magnetic resonance (MR) imager. With dynamic-susceptibility contrast MR imaging the intensity-time curves of each tumor were analyzed, and we determined the relative regional cerebral blood volume ratios (rrCBV [tumor/contralateral white matter (WM)]) to find out whether these parameters could be used to separate PCNSL from GBM. The maximum rrCBV ratio in the PCNSL was significantly lower than that of the GBM (P < 0.0001). Comparing the intensity-time curves for the two tumor groups, the PCNSL showed a characteristic type of curve with a significant increase in signal intensity above the baseline due to massive leakage of contrast media into the interstitial space. PCNSL tend to have low maximum CBV ratios and typical intensity-time curves. These two parameters may be useful in distinguishing PCNSL from GBM. (C) 2002 Elsevier Science Ireland Ltd. All rights reserved.
C1 Univ Heidelberg, Med Ctr, Dept Neurol, Div Neuroradiol, D-69120 Heidelberg, Germany.
   Univ Heidelberg, Med Ctr, Dept Neurosurg, D-69120 Heidelberg, Germany.
   Univ Ulm, Dept Pathol, Div Neuropathol, D-89081 Ulm, Germany.
C3 Ruprecht Karls University Heidelberg; Ruprecht Karls University
   Heidelberg; Ulm University
RP Hartmann, M (corresponding author), Univ Heidelberg, Med Ctr, Dept Neurol, Div Neuroradiol, Neuenheimer Feld 400, D-69120 Heidelberg, Germany.
CR Abrey LE, 2000, J CLIN ONCOL, V18, P3144, DOI 10.1200/JCO.2000.18.17.3144
   ARONEN HJ, 1995, ACTA RADIOL, V36, P520
   ARONEN HJ, 1994, RADIOLOGY, V191, P41, DOI 10.1148/radiology.191.1.8134596
   Bühring U, 2001, NEUROLOGY, V57, P393, DOI 10.1212/WNL.57.3.393
   Coulon A, 2002, EUR RADIOL, V12, P329, DOI 10.1007/s003300101037
   GOMORI JM, 1995, INT J NEURORADIOL, V1, P127
   Hartmann M, 1997, RADIOLOGE, V37, P42, DOI 10.1007/s001170050176
   Heiland S, 2000, ROFO-FORTSCHR RONTG, V172, P812, DOI 10.1055/s-2000-7899
   Heiland S, 1999, MAGN RESON IMAGING, V17, P21, DOI 10.1016/S0730-725X(98)00149-0
   Heiland S, 1999, ROFO-FORTSCHR RONTG, V171, P3, DOI 10.1055/s-1999-9889
   JELLINGER KA, 1992, J CANCER RES CLIN, V119, P7, DOI 10.1007/BF01209483
   Kleihues P, 1999, HISTOLOGICAL TYPING
   Knopp EA, 1999, RADIOLOGY, V211, P791, DOI 10.1148/radiology.211.3.r99jn46791
   Ostergaard L, 1999, J NEUROSURG, V90, P300, DOI 10.3171/jns.1999.90.2.0300
   ROMANGOLDSTEIN SM, 1992, AM J NEURORADIOL, V13, P1207
   Sugahara T, 1998, AM J ROENTGENOL, V171, P1479, DOI 10.2214/ajr.171.6.9843274
   Sugahara T, 1999, J COMPUT ASSIST TOMO, V23, P232, DOI 10.1097/00004728-199903000-00011
NR 17
TC 164
Z9 177
U1 0
U2 7
PU ELSEVIER SCI IRELAND LTD
PI CLARE
PA CUSTOMER RELATIONS MANAGER, BAY 15, SHANNON INDUSTRIAL ESTATE CO, CLARE,
   IRELAND
SN 0304-3940
J9 NEUROSCI LETT
JI Neurosci. Lett.
PD FEB 27
PY 2003
VL 338
IS 2
BP 119
EP 122
DI 10.1016/S0304-3940(02)01367-8
PG 4
WC Neurosciences
WE Science Citation Index Expanded (SCI-EXPANDED)
SC Neurosciences & Neurology
GA 646BE
UT WOS:000181014300008
PM 12566167
DA 2024-06-03
ER

PT J
AU Nguyen, PL
   Chakravarti, A
   Finkelstein, DM
   Hochberg, FH
   Batchelor, TT
   Loeffler, JS
AF Nguyen, PL
   Chakravarti, A
   Finkelstein, DM
   Hochberg, FH
   Batchelor, TT
   Loeffler, JS
TI Results of whole-brain radiation as salvage of methotrexate failure for
   immunocompetent patients with primary CNS lymphoma
SO JOURNAL OF CLINICAL ONCOLOGY
LA English
DT Article
ID CENTRAL-NERVOUS-SYSTEM; DEFERRED RADIOTHERAPY; THERAPY; SURVIVAL;
   CHEMOTHERAPY
AB Purpose
   This study evaluates the efficacy and toxicity of whole-brain radiation therapy (WBRT) as salvage therapy for immunocompetent patients who failed initial high-dose methotrexate for primary CNS lymphoma (PCNSL).
   Patients and Methods
   The study cohort included 27 consecutive patients who failed initial high-dose methotrexate and then received salvage WBRT (median dose, 36 Gy). Actuarial survival was measured from the initiation of radiotherapy.
   Results
   Ten patients (37%) achieved a complete radiographic response (CR), and 10 patients (37%) a partial response to WBRT, for a 74% overall radiographic response rate. At the time of maximal response, Karnofsky performance status improved in 12 (44%) of 27 patients and at least stabilized in 67%. Median estimated survival from initiation of WBRT was 10.9 months (range, 0.3 to 63.7 months). The univariate predictor of longer survival was age less than 60 years at the time of WBRT (P = .028). Among patients who survived 4 months, achievement of a CR to WBRT by 4 months (P = .002) predicted longer survival. Late treatment-associated neurotoxicity was diagnosed in four patients (15%) and was significantly associated with total radiation doses greater than 36 Gy (P = .04). No patient treated with daily fractions less than 1.8 Gy developed late neurotoxicity.
   Conclusion
   For patients with PCNSL who experience treatment failure with methotrexate, WBRT provides high response rates (74%) and a median survival of 10.9 months. Age less than 60 years and response to WBRT predict post-WBRT survival. Modest rates of late neurotoxicity (15%) were seen and were associated with a total dose greater than 36 Gy.
C1 Harvard Univ, Massachusetts Gen Hosp, Sch Med, Dept Radiat Oncol, Boston, MA 02114 USA.
   Harvard Univ, Massachusetts Gen Hosp, Sch Med, Dept Neurol, Boston, MA 02114 USA.
   Harvard Univ, Massachusetts Gen Hosp, Sch Med, Dept Med, Boston, MA 02114 USA.
   Harvard Univ, Massachusetts Gen Hosp, Sch Med, Dept Biostat, Boston, MA 02114 USA.
C3 Harvard University; Harvard Medical School; Massachusetts General
   Hospital; Harvard University; Harvard Medical School; Massachusetts
   General Hospital; Harvard University; Harvard Medical School;
   Massachusetts General Hospital; Harvard University; Massachusetts
   General Hospital; Harvard Medical School
RP Harvard Univ, Massachusetts Gen Hosp, Sch Med, Dept Radiat Oncol, 100 Blossom St,Cox 301, Boston, MA 02114 USA.
EM
CR Abrey LE, 1998, J CLIN ONCOL, V16, P859, DOI 10.1200/JCO.1998.16.3.859
   Abrey LE, 2000, J CLIN ONCOL, V18, P3144, DOI 10.1200/JCO.2000.18.17.3144
   Abrey LE, 1997, NEUROLOGY, V48, pV1003
   Ahsan H, 1995, INT J EPIDEMIOL, V24, P1078, DOI 10.1093/ije/24.6.1078
   Batchelor T, 2003, J CLIN ONCOL, V21, P1044, DOI 10.1200/JCO.2003.03.036
   CBTRUS, 2002, STAT REPORT PRIMARY
   Cher L, 1996, NEUROLOGY, V46, P1757, DOI 10.1212/WNL.46.6.1757
   DeAngelis LM, 2002, J CLIN ONCOL, V20, P4643, DOI 10.1200/JCO.2002.11.013
   DESABLENS B, 1999, ANN ONCOL, V10, P14
   Ferreri AJM, 2003, J CLIN ONCOL, V21, P266, DOI 10.1200/JCO.2003.09.139
   Guha-Thakurta N, 1999, J NEURO-ONCOL, V43, P259, DOI 10.1023/A:1006210703827
   KORN EL, 1986, STAT MED, V5, P255, DOI 10.1002/sim.4780050306
   LAWLER BE, 2002, P AN M AM SOC CLIN, V21, pA75
   LIANG BC, 1999, HOSP PHYSICIAN, V34, P54
   LOEFFLER JS, 1985, J CLIN ONCOL, V3, P490, DOI 10.1200/JCO.1985.3.4.490
   MACDONALD DR, 1990, J CLIN ONCOL, V8, P1277, DOI 10.1200/JCO.1990.8.7.1277
   NELSON DF, 1992, INT J RADIAT ONCOL, V23, P9, DOI 10.1016/0360-3016(92)90538-S
   Plotkin SR, 2001, LANCET ONCOL, V2, P455
   Reni M, 2001, INT J RADIAT ONCOL, V51, P419, DOI 10.1016/S0360-3016(01)01639-X
NR 19
TC 162
Z9 176
U1 0
U2 1
PU AMER SOC CLINICAL ONCOLOGY
PI ALEXANDRIA
PA 2318 MILL ROAD, STE 800, ALEXANDRIA, VA 22314 USA
SN 0732-183X
EI 1527-7755
J9 J CLIN ONCOL
JI J. Clin. Oncol.
PD MAR 1
PY 2005
VL 23
IS 7
BP 1507
EP 1513
DI 10.1200/JCO.2005.01.161
PG 7
WC Oncology
WE Science Citation Index Expanded (SCI-EXPANDED)
SC Oncology
GA 902PB
UT WOS:000227367900023
PM 15735126
OA Bronze
DA 2024-06-03
ER

PT J
AU Soussain, C
   Choquet, S
   Blonski, M
   Leclercq, D
   Houillier, C
   Rezai, K
   Bijou, F
   Houot, R
   Boyle, E
   Gressin, R
   Nicolas-Virelizier, E
   Barrie, M
   Moluçon-Chabrot, C
   Lelez, ML
   Clavert, A
   Coisy, S
   Leruez, S
   Touitou, V
   Cassoux, N
   Daniau, M
   de la Bretonnière, ME
   El Yamani, A
   Ghesquieres, H
   Hoang-Xuan, K
AF Soussain, C.
   Choquet, S.
   Blonski, M.
   Leclercq, D.
   Houillier, C.
   Rezai, K.
   Bijou, F.
   Houot, R.
   Boyle, E.
   Gressin, R.
   Nicolas-Virelizier, E.
   Barrie, M.
   Molucon-Chabrot, C.
   Lelez, M. L.
   Clavert, A.
   Coisy, S.
   Leruez, S.
   Touitou, V.
   Cassoux, N.
   Daniau, M.
   de la Bretonniere, M. Ertault
   El Yamani, A.
   Ghesquieres, H.
   Hoang-Xuan, K.
TI Ibrutinib monotherapy for relapse or refractory primary CNS lymphoma and
   primary vitreoretinal lymphoma: Final analysis of the phase II
   'proof-of-concept' <i>i</i>LOC study by the Lymphoma study association
   (LYSA) and the French oculo-cerebral lymphoma (LOC) network
SO EUROPEAN JOURNAL OF CANCER
LA English
DT Article
DE Primary CNS lymphoma; Primary vitreoretinal lymphoma; Relapse; Ibrutinib
ID CENTRAL-NERVOUS-SYSTEM; BRUTON TYROSINE KINASE; INHIBITOR IBRUTINIB
AB Background: Primary central nervous system lymphomas (PCNSLs) are mainly diffuse large B-cell lymphomas (DLBCLs) of the non-germinal centre B-cell subtype, with unmet medical needs. This study aimed to evaluate the efficacy and toxicity of ibrutinib in DLBCL-PCNSL
   Patients and methods: This prospective, multicentre, phase II study involved patients with relapse or refractory(R/R) DLBCL-PCNSL or primary vitreoretinal lymphoma. The treatment consisted of ibrutinib (560 mg/day) until disease progression or unacceptable toxicity occurred. The primary outcome was the disease control (DC) rate after two months of treatment (P0 < 10%; P1 > 30%).
   Results: Fifty-two patients were recruited. Forty-four patients were evaluable for response. After 2 months of treatment, the DC was 70% in evaluable patients and 62% in the intent-to-treat analysis, including 10 complete responses (19%), 17 partial responses (33%) and 5 stable diseases (10%). With a median follow-up of 25.7 months (range, 0.7-30.5), the median progression-free and overall survivals were 4.8 months (95% confidence interval [CI]; 2.8 -12.7) and 19.2 months (95% CI; 7.2-NR), respectively. Thirteen patients received ibrutinib for more than 12 months. Two patients experienced pulmonary aspergillosis with a favourable (n = 1) or fatal outcome (n = 1). Ibrutinib was detectable in the cerebrospinal fluid (CSF). The clinical response to ibrutinib seemed independent of the gene mutations in the BCR pathway.
   Conclusion: Ibrutinib showed clinical activity in the brain, the CSF and the intraocular compartment and was tolerated in R/R PCNSL. The addition of ibrutinib to standard methotrexate-base induction chemotherapy will be further evaluated in the first-line treatment. (C) 2019 Elsevier Ltd. All rights reserved.
C1 [Soussain, C.] Inst Curie, Hematol, Site St Cloud, St Cloud, France.
   [Choquet, S.] Grp Hosp Pitie Salpetriere, AP HP, Hematol, Paris, France.
   [Blonski, M.] CHU Nancy, Neurol, Nancy, France.
   [Leclercq, D.] Grp Hosp Pitie Salpetriere, AP HP, Neuroradiol, Paris, France.
   [Houillier, C.; Hoang-Xuan, K.] Sorbonne Univ, Grp Hosp Pitie Salpetriere, AP HP, IHU,ICM,Neurol, Paris, France.
   [Rezai, K.] Inst Curie, Radiopharmacol, Site St Cloud, St Cloud, France.
   [Bijou, F.] Inst Bergonie, Hematol, Bordeaux, France.
   [Houot, R.] Univ Rennes, CHU Rennes, MICMAC MIcroenvironm Cell Differentiat iMmunol &, INSERM,Dept Hematol,UMR S 1236, F-35000 Rennes, France.
   [Boyle, E.] CHU Lille, Hematol, Lille, France.
   [Gressin, R.] CHU Grenoble, Hematol, Grenoble, France.
   [Nicolas-Virelizier, E.] Ctr Leon Berard, Hematol, Lyon, France.
   [Barrie, M.] CHU Timone, Neurooncol, Marseille, France.
   [Molucon-Chabrot, C.] CHU Clermont Ferrand, Hematol, Clermont Ferrand, France.
   [Lelez, M. L.] CHU Tours, Ophthalmol, Tours, France.
   [Clavert, A.] CHU Angers, Hematol, Angers, France.
   [Coisy, S.; Leruez, S.] CHU Angers, Ophthalmol, Angers, France.
   [Touitou, V.] Grp Hosp Pitie Salpetriere, AP HP, Ophthalmol, Paris, France.
   [Cassoux, N.] Univ Paris Descartes & PSL Paris Sci & Lettre, Inst Curie, Ophthalmol, Site Paris, Paris, France.
   [Daniau, M.] Grp Hosp Pitie Salpetriere, Inst Cerveau & Moelle, Mol Biol, Paris, France.
   [de la Bretonniere, M. Ertault] Univ Tours, CHU Tours, Hematol & Cell Therapy, Tours, France.
   [El Yamani, A.] Ctr Hosp Blois, Hematol, Blois, France.
   [Ghesquieres, H.] Univ Claude Bernard Lyon 1, Ctr Hosp Lyon Sud, Hematol, Pierre Benite, France.
C3 UNICANCER; Universite PSL; Institut Curie; Assistance Publique Hopitaux
   Paris (APHP); Hopital Universitaire Pitie-Salpetriere - APHP; Sorbonne
   Universite; CHU de Nancy; Sorbonne Universite; Assistance Publique
   Hopitaux Paris (APHP); Hopital Universitaire Pitie-Salpetriere - APHP;
   Assistance Publique Hopitaux Paris (APHP); Hopital Universitaire
   Pitie-Salpetriere - APHP; Sorbonne Universite; UNICANCER; Universite
   PSL; Institut Curie; UNICANCER; Institut Bergonie; Universite de Rennes;
   Institut National de la Sante et de la Recherche Medicale (Inserm); CHU
   Rennes; Universite de Lille; CHU Lille; Communaute Universite Grenoble
   Alpes; Universite Grenoble Alpes (UGA); CHU Grenoble Alpes; UNICANCER;
   Centre Leon Berard; Aix-Marseille Universite; Assistance
   Publique-Hopitaux de Marseille; CHU Clermont Ferrand; CHU Tours;
   Universite d'Angers; Centre Hospitalier Universitaire d'Angers;
   Universite d'Angers; Centre Hospitalier Universitaire d'Angers; Sorbonne
   Universite; Assistance Publique Hopitaux Paris (APHP); Hopital
   Universitaire Pitie-Salpetriere - APHP; Universite Paris Cite;
   UNICANCER; Universite PSL; Institut Curie; Sorbonne Universite;
   Assistance Publique Hopitaux Paris (APHP); Hopital Universitaire
   Pitie-Salpetriere - APHP; CHU Tours; Universite de Tours; CHU Lyon;
   Universite Claude Bernard Lyon 1
RP Soussain, C (corresponding author), Inst Curie, Hematol, 35 Rue Dailly, St Cloud, France.
EM
RI Boyle, Eileen M./Y-5179-2019; choquet, sylvain/O-7952-2017; Hoang-Xuan,
   Khe/O-7527-2017
OI Boyle, Eileen M./0000-0001-8791-1744; choquet,
   sylvain/0000-0002-7791-0470; Hoang Xuan, Khe/0000-0003-4072-1999;
   Houillier, Caroline/0000-0003-2931-7522
FU Janssen; LYSARC (The Lymphoma Academic Research Organization)
FX This investigator-initiated study was funded by Janssen and sponsored by
   the LYSARC (The Lymphoma Academic Research Organization). Janssen did
   not participate in the conception, conduction or analysis of the trial
   or the writing of this report.
CR Abrey LE, 2005, J CLIN ONCOL, V23, P5034, DOI 10.1200/JCO.2005.13.524
   Advani RH, 2013, J CLIN ONCOL, V31, P88, DOI 10.1200/JCO.2012.42.7906
   Belhouachi N, 2013, HAEMATOLOGICA, V98, pS1
   Bruno A, 2014, ONCOTARGET, V5, P5065, DOI 10.18632/oncotarget.2080
   Camilleri-Broët S, 2006, BLOOD, V107, P190, DOI 10.1182/blood-2005-03-1024
   Cassoux N, 2007, INVEST OPHTH VIS SCI, V48, P3253, DOI 10.1167/iovs.06-0031
   Chamoun K, 2017, NEUROLOGY, V88, P101, DOI 10.1212/WNL.0000000000003420
   Chapuy B, 2016, BLOOD, V127, P869, DOI 10.1182/blood-2015-10-673236
   Costopoulos M, 2016, OPHTHALMOLOGY, V123, P1626, DOI 10.1016/j.ophtha.2016.01.037
   GHESQUIERES H, 2016, BLOOD, V128
   Ghez D, 2018, BLOOD, V131, P1955, DOI 10.1182/blood-2017-11-818286
   Goldwirt L, 2018, CANCER CHEMOTH PHARM, V81, P783, DOI 10.1007/s00280-018-3546-3
   Gonzalez-Aguilar A, 2012, CLIN CANCER RES, V18, P5203, DOI 10.1158/1078-0432.CCR-12-0845
   Grommes C, 2017, CANCER DISCOV, V7, P1018, DOI 10.1158/2159-8290.CD-17-0613
   Hans CP, 2004, BLOOD, V103, P275, DOI 10.1182/blood-2003-05-1545
   Lionakis MS, 2017, CANCER CELL, V31, P833, DOI 10.1016/j.ccell.2017.04.012
   Montesinos-Rongen M, 2008, LEUKEMIA, V22, P400, DOI 10.1038/sj.leu.2405019
   Montesinos-Rongen M, 2004, BLOOD, V103, P1869, DOI 10.1182/blood-2003-05-1465
   Nayak L., 2017, Hematol Oncol, V35, P420, DOI [10.1002/hon.2440_2, DOI 10.1002/HON.2440_2]
   Pouzoulet F, 2016, BLOOD, V128, DOI 10.1182/blood.V128.22.4170.4170
   Qiu YS, 2018, BRIT J HAEMATOL, V183, P674, DOI 10.1111/bjh.15009
   ROGERS KA, 2017, BLOOD S1, V130
   Thompsett AR, 1999, BLOOD, V94, P1738, DOI 10.1182/blood.V94.5.1738.417k10_1738_1746
   Tillman BF, 2018, EUR J HAEMATOL, V100, P325, DOI 10.1111/ejh.13020
   Tun HW, 2018, BLOOD, V132, P2240, DOI 10.1182/blood-2018-02-835496
   Wilson WH, 2015, NAT MED, V21, P922, DOI 10.1038/nm.3884
   Yang YB, 2012, CANCER CELL, V21, P723, DOI 10.1016/j.ccr.2012.05.024
NR 27
TC 160
Z9 169
U1 1
U2 8
PU ELSEVIER SCI LTD
PI OXFORD
PA THE BOULEVARD, LANGFORD LANE, KIDLINGTON, OXFORD OX5 1GB, OXON, ENGLAND
SN 0959-8049
EI 1879-0852
J9 EUR J CANCER
JI Eur. J. Cancer
PD AUG
PY 2019
VL 117
BP 121
EP 130
DI 10.1016/j.ejca.2019.05.024
PG 10
WC Oncology
WE Science Citation Index Expanded (SCI-EXPANDED)
SC Oncology
GA II8YH
UT WOS:000475479200016
PM 31279304
OA Green Published, hybrid
DA 2024-06-03
ER

PT J
AU Weller, M
   Martus, P
   Roth, P
   Thiel, E
   Korfel, A
AF Weller, Michael
   Martus, Peter
   Roth, Patrick
   Thiel, Eckhard
   Korfel, Agnieszka
CA German PCNSL Study Grp
TI Surgery for primary CNS lymphoma? Challenging a paradigm
SO NEURO-ONCOLOGY
LA English
DT Article
DE CNS lymphoma; resection; surgery
ID CENTRAL-NERVOUS-SYSTEM; RADIOTHERAPY
AB The standard of care for primary central nervous system lymphoma (PCNSL) is systemic chemotherapy with or without whole brain radiotherapy or intrathecal chemotherapy. In contrast to treatment for other brain tumors, efforts at resection are discouraged. This is a secondary analysis of the German PCNSL Study Group1 trial, a large randomized phase III study comprising 526 patients with PCNSL. Progression-free survival (hazard ratio [HR]: 1.39; 95 confidence interval [CI]: 1.101.74; P = .005) and overall survival (HR: 1.33; 95 CI: 1.041.70; P = .024) were significantly shorter in biopsied patients compared with patients with subtotal or gross total resections. This difference in outcome was not due to age or Karnofsky performance status (KPS). When controlled for the number of lesions, the HR of biopsy versus subtotal or gross total resection remained unchanged for progression-free survival (HR = 1.37; P = .009) but was smaller for overall survival (HR = 1.27; P = .085). This analysis of the largest PCNSL trial ever performed challenges the traditional view that the extent of resection has no prognostic impact on this disease. Therefore, we propose to reconsider the statement that efforts at resection should be discouraged, at least if resection seems safe, as is often the case in treatment of single PCNSL lesions.
C1 [Weller, Michael; Roth, Patrick] Univ Zurich Hosp, Dept Neurol, CH-8091 Zurich, Switzerland.
   [Martus, Peter] Univ Tubingen, Dept Med Biometry, Tubingen, Germany.
   [Thiel, Eckhard; Korfel, Agnieszka] Charite, Dept Hematol & Oncol, D-13353 Berlin, Germany.
C3 University of Zurich; University Zurich Hospital; Eberhard Karls
   University of Tubingen; Eberhard Karls University Hospital; Free
   University of Berlin; Humboldt University of Berlin; Charite
   Universitatsmedizin Berlin
RP Weller, M (corresponding author), Univ Zurich Hosp, Dept Neurol, Frauenklin Str 26, CH-8091 Zurich, Switzerland.
EM
RI McDermott, Michael Francis/GZM-5330-2022; Roth, Patrick/R-6671-2018
OI Weller, Michael/0000-0002-1748-174X
FU German Cancer Aid grant [70-2838-Th 2]; Merck Serono; Roche; Pfizer;
   Amgen; Celgene
FX The G-PCNSL-SG-1 trial was supported by the German Cancer Aid grant
   70-2838-Th 2 to E.T. and M.W.; M.W. received research support from Merck
   Serono and Roche and honoraria for advisory board and lecture activities
   from Merck Serono, MSD, Roche, and Magforce. P. M. received honoraria
   from Parexel for serving on a safety board. P. R. reports no
   disclosures. E. T. received honoraria, grants, and congress support from
   Pfizer, Amgen, and Celgene. A.K. reports no disclosures.
CR Bataille B, 2000, J NEUROSURG, V92, P261, DOI 10.3171/jns.2000.92.2.0261
   Bellinzona M, 2005, EJSO, V31, P100, DOI 10.1016/j.ejso.2004.10.002
   DEANGELIS LM, 1990, NEUROLOGY, V40, P80, DOI 10.1212/WNL.40.1.80
   HENRY JM, 1974, CANCER, V34, P1293, DOI 10.1002/1097-0142(197410)34:4<1293::AID-CNCR2820340441>3.0.CO;2-P
   Thiel E, 2010, LANCET ONCOL, V11, P1036, DOI 10.1016/S1470-2045(10)70229-1
NR 5
TC 159
Z9 169
U1 0
U2 2
PU OXFORD UNIV PRESS INC
PI CARY
PA JOURNALS DEPT, 2001 EVANS RD, CARY, NC 27513 USA
SN 1522-8517
J9 NEURO-ONCOLOGY
JI Neuro-Oncology
PD DEC
PY 2012
VL 14
IS 12
BP 1481
EP 1484
DI 10.1093/neuonc/nos159
PG 4
WC Oncology; Clinical Neurology
WE Science Citation Index Expanded (SCI-EXPANDED)
SC Oncology; Neurosciences & Neurology
GA 040HP
UT WOS:000311313600008
PM 22984018
OA Green Published, Bronze
DA 2024-06-03
ER

PT J
AU Antinori, A
   Ammassari, A
   DeLuca, A
   Cingolani, A
   Murri, R
   Scoppettuolo, G
   Fortini, M
   Tartaglione, T
   Larocca, LM
   Zannoni, G
   Cattani, P
   Grillo, R
   Roselli, R
   Iacoangeli, M
   Scerrati, M
   Ortona, L
AF Antinori, A
   Ammassari, A
   DeLuca, A
   Cingolani, A
   Murri, R
   Scoppettuolo, G
   Fortini, M
   Tartaglione, T
   Larocca, LM
   Zannoni, G
   Cattani, P
   Grillo, R
   Roselli, R
   Iacoangeli, M
   Scerrati, M
   Ortona, L
TI Diagnosis of AIDS-related focal brain lesions: A decision-making
   analysis based on clinical and neuroradiologic characteristics combined
   with polymerase chain reaction assays in CSF
SO NEUROLOGY
LA English
DT Article
ID CENTRAL-NERVOUS-SYSTEM; ACQUIRED-IMMUNODEFICIENCY-SYNDROME;
   IMMUNE-DEFICIENCY-SYNDROME; PROGRESSIVE MULTIFOCAL LEUKOENCEPHALOPATHY;
   PNEUMOCYSTIS-CARINII PNEUMONIA; CEREBROSPINAL-FLUID; HIV-INFECTION;
   TOXOPLASMIC ENCEPHALITIS; AEROSOLIZED PENTAMIDINE; CEREBRAL
   TOXOPLASMOSIS
AB Objective: To identify disease patterns in AIDS-related focal brain lesions (FBL) and to design a decision-making strategy for differential diagnosis. Design: Prospective study. Probabilities of CNS disorders were calculated using Bayes' theorem according to clinical variables (mass effect at CT or MRI, Toxoplasma serology, anti-Toxoplasma prophylaxis) and to the results of polymerase chain reaction (PCR) assays. Patients: 136 consecutive HIV-infected patients with a definitive diagnosis of FBL-causing disorder observed from 1991 to 1995 in a single clinical setting, Interventions: Patients underwent empiric anti-Toxoplasma therapy. After 3 weeks, patients with progressive: stable disease underwent brain biopsy. In 66 patients Epstein-Barr virus (EBV)-DNA, JC virus (JCV)-DNA, and T gondii-DNA amplification was performed by PCR in CSF. Diagnostic criteria were histopathologic examination of bioptic or autoptic tissue specimens for all disorders and complete/partial resolution of FBL after empiric therapy for toxoplasmic encephalitis (TE). Results: Neuroradiologic characteristics did not discriminate between TE and primary CNS lymphoma (PCNSL). Probability of TE was 0.87 in Toxoplasma-seropositive patients with mass effect who were not receiving anti-Toxoplasma prophylaxis, but only 0.59 if prophylaxis was performed. In seronegative patients with mass effect, the likelihood of PCNSL was 0.74, If EBV-DNA or T gondii-DNA tests were positive, the probability of PCNSL or TE increased to more than 0.96, The absence of T gondii-DNA did not exclude the possibility of a TE diagnosis, Among FBL without mass effect, the probability of progressive multifocal leukoencephalopathy (PML) was 0.81; this increased to 0.99 if JCV-DNA testing was positive. Sensitivity of brain biopsy was 93%, with a perioperative morbidity of 12% and a mortality of 2%. Conclusions: Due to the low diagnostic capability of clinical variables, PCR amplifications in CSF, especially for EBV-DNA and for JCV-DNA, represent, in most cases, an essential step in the differential diagnosis of AIDS-related FBL. This is particularly true in patients with FBL without mass effect or with mass effect and who are either seronegative or undergoing anti-Toxoplasma prophylaxis Brain biopsy remains a necessary procedure in EBV-DNA-positive cases and in seronegative patients with FBL displaying a mass effect. Positive JCV-DNA testing may obviate the need for brain biopsy in patients with FBL without mass effect, An advanced diagnostic strategy based on combined clinical criteria and PCR tests may allow rapid and accurate identification of patients for prompt brain biopsy or specific therapy.
C1 UNIV CATTOLICA SACRO CUORE, DEPT RADIOL, I-00168 ROME, ITALY.
   UNIV CATTOLICA SACRO CUORE, DEPT PATHOL, I-00168 ROME, ITALY.
   UNIV CATTOLICA SACRO CUORE, DEPT MICROBIOL, I-00168 ROME, ITALY.
   UNIV CATTOLICA SACRO CUORE, DEPT NEUROSURG, I-00168 ROME, ITALY.
   NATL STAT INST, DEPT METHODOL STUDIES, ROME, ITALY.
C3 Catholic University of the Sacred Heart; IRCCS Policlinico Gemelli;
   Catholic University of the Sacred Heart; IRCCS Policlinico Gemelli;
   Catholic University of the Sacred Heart; IRCCS Policlinico Gemelli;
   Catholic University of the Sacred Heart; IRCCS Policlinico Gemelli
RP Antinori, A (corresponding author), UNIV CATTOLICA SACRO CUORE, DEPT INFECT DIS, LGO A GEMELLI 8, I-00168 ROME, ITALY.
RI Antinori, Andrea/K-7125-2016; Larocca, Luigi Maria/M-9590-2019; Cattani,
   Paola/AAC-8218-2019; Scoppettuolo, Giancarlo/AAR-6970-2020; Zannoni,
   Gian Franco/AAC-4754-2020; Larocca, Luigi M/A-1577-2008; TARTAGLIONE,
   TOMMASO/K-5390-2017; De Luca, Andrea/G-8810-2011; cingolani,
   antonella/J-8921-2018
OI Antinori, Andrea/0000-0003-2121-4684; Larocca, Luigi
   Maria/0000-0003-1739-4758; Scoppettuolo, Giancarlo/0000-0003-0893-7044;
   Zannoni, Gian Franco/0000-0002-4473-7560; TARTAGLIONE,
   TOMMASO/0000-0003-3896-4078; IACOANGELI, Maurizio/0000-0002-3629-5301;
   De Luca, Andrea/0000-0002-9561-9193; Cattani, Paola/0000-0001-5231-4229;
   Murri, Rita/0000-0001-9468-5458; cingolani,
   antonella/0000-0002-3793-2755; Ammassari, Adriana/0000-0002-0284-0340
CR AMMASSARI A, 1996, CURR TOP MICROBIOL I, V219, P201
   ANTINORI A, 1993, LANCET, V341, P1411, DOI 10.1016/0140-6736(93)90977-O
   ANTINORI A, 1995, AIDS, V9, P1343, DOI 10.1097/00002030-199512000-00007
   ANTINORI A, 1994, AIDS, V8, P1022, DOI 10.1097/00002030-199407000-00030
   Armitage P, 1990, STAT METHODS MED RES
   BACELLAR H, 1994, NEUROLOGY, V44, P1892, DOI 10.1212/WNL.44.10.1892
   BAUMGARTNER JE, 1990, J NEUROSURG, V73, P206, DOI 10.3171/jns.1990.73.2.0206
   BERGER JR, 1987, ANN INTERN MED, V107, P78, DOI 10.7326/0003-4819-107-1-78
   [Anonymous], 1992, MMWR Recomm Rep, V41, P1
   CHAMBERLAIN MC, 1994, CANCER, V73, P1728, DOI 10.1002/1097-0142(19940315)73:6<1728::AID-CNCR2820730629>3.0.CO;2-8
   CHAPPELL ET, 1992, NEUROSURGERY, V30, P825, DOI 10.1227/00006123-199206000-00001
   CIMINO C, 1991, ARCH INTERN MED, V151, P1381, DOI 10.1001/archinte.151.7.1381
   Cingolani A, 1996, J MED MICROBIOL, V45, P472, DOI 10.1099/00222615-45-6-472
   CINQUE P, 1993, LANCET, V342, P398, DOI 10.1016/0140-6736(93)92814-A
   CIRILLO SF, 1991, J NEUROSURG, V73, P720
   COHN JA, 1989, AM J MED, V86, P521, DOI 10.1016/0002-9343(89)90378-1
   CORN BW, 1995, CANCER, V76, P163, DOI 10.1002/1097-0142(19950715)76:2<163::AID-CNCR2820760202>3.0.CO;2-Z
   DELUCA A, 1995, BRIT J HAEMATOL, V90, P844, DOI 10.1111/j.1365-2141.1995.tb05205.x
   DeLuca A, 1996, J CLIN MICROBIOL, V34, P1343, DOI 10.1128/JCM.34.5.1343-1346.1996
   DONAHUE BR, 1995, CANCER, V76, P328, DOI 10.1002/1097-0142(19950715)76:2<328::AID-CNCR2820760225>3.0.CO;2-7
   DUPON M, 1995, J CLIN MICROBIOL, V33, P2421, DOI 10.1128/JCM.33.9.2421-2426.1995
   FORMENTI SC, 1989, CANCER-AM CANCER SOC, V63, P1101, DOI 10.1002/1097-0142(19890315)63:6<1101::AID-CNCR2820630611>3.0.CO;2-R
   FORSYTH PA, 1994, NEUROLOGY, V44, P1473, DOI 10.1212/WNL.44.8.1473
   GALLETO G, 1993, JAMA-J AM MED ASSOC, V269, P92
   GIRARD PM, 1993, NEW ENGL J MED, V328, P1514, DOI 10.1056/NEJM199305273282102
   GOLDSTEIN JD, 1991, CANCER, V67, P2756, DOI 10.1002/1097-0142(19910601)67:11<2756::AID-CNCR2820671108>3.0.CO;2-3
   HAVERKOS HW, 1987, AM J MED, V82, P907, DOI 10.1016/0002-9343(87)90151-3
   HOFFMAN JM, 1993, J NUCL MED, V34, P567
   IACOANGELI M, 1994, BRIT J SURG, V81, P1508, DOI 10.1002/bjs.1800811036
   LEVY RM, 1992, NEUROSURGERY, V30, P186, DOI 10.1227/00006123-199202000-00006
   LING SM, 1994, CANCER-AM CANCER SOC, V73, P2570, DOI 10.1002/1097-0142(19940515)73:10<2570::AID-CNCR2820731019>3.0.CO;2-1
   LUFT BJ, 1993, NEW ENGL J MED, V329, P995, DOI 10.1056/NEJM199309303291403
   Luzzati R, 1996, ARCH INTERN MED, V156, P565, DOI 10.1001/archinte.156.5.565
   MATHEWS C, 1995, AIDS, V9, P1243, DOI 10.1097/00002030-199511000-00005
   MOOD AM, 1974, INTRO THEORY STATIST
   NAVIA BA, 1986, ANN NEUROL, V19, P224, DOI 10.1002/ana.410190303
   OKSENHENDLER E, 1994, AIDS, V8, P483, DOI 10.1097/00002030-199404000-00010
   OMALLEY JP, 1994, AM J ROENTGENOL, V163, P417, DOI 10.2214/ajr.163.2.8037042
   PLUDA JM, 1990, ANN INTERN MED, V113, P276, DOI 10.7326/0003-4819-113-4-276
   PORTER SB, 1992, NEW ENGL J MED, V327, P1643, DOI 10.1056/NEJM199212033272306
   SNIDER WD, 1983, ANN NEUROL, V14, P403, DOI 10.1002/ana.410140404
   Vach W., 1994, Logistic regression with missing values in the covariates
   1992, LANCET, V340, P1135
NR 43
TC 159
Z9 185
U1 0
U2 6
PU LIPPINCOTT-RAVEN PUBL
PI PHILADELPHIA
PA 227 EAST WASHINGTON SQ, PHILADELPHIA, PA 19106
SN 0028-3878
J9 NEUROLOGY
JI Neurology
PD MAR
PY 1997
VL 48
IS 3
BP 687
EP 694
DI 10.1212/WNL.48.3.687
PG 8
WC Clinical Neurology
WE Science Citation Index Expanded (SCI-EXPANDED)
SC Neurosciences & Neurology
GA XC475
UT WOS:A1997XC47500025
PM 9065549
DA 2024-06-03
ER

PT J
AU Braggio, E
   Van Wier, S
   Ojha, J
   McPhail, E
   Asmann, YW
   Egan, J
   da Silva, JA
   Schiff, D
   Lopes, MB
   Decker, PA
   Valdez, R
   Tibes, R
   Eckloff, B
   Witzig, TE
   Stewart, AK
   Fonseca, R
   O'Neill, BP
AF Braggio, Esteban
   Van Wier, Scott
   Ojha, Juhi
   McPhail, Ellen
   Asmann, Yan W.
   Egan, Jan
   da Silva, Jackline Ayres
   Schiff, David
   Lopes, M. Beatriz
   Decker, Paul A.
   Valdez, Riccardo
   Tibes, Raoul
   Eckloff, Bruce
   Witzig, Thomas E.
   Stewart, A. Keith
   Fonseca, Rafael
   O'Neill, Brian Patrick
TI Genome-Wide Analysis Uncovers Novel Recurrent Alterations in Primary
   Central Nervous System Lymphomas
SO CLINICAL CANCER RESEARCH
LA English
DT Article
ID B-CELL LYMPHOMAS; KINASE-C-DELTA; L265P SOMATIC MUTATION;
   GENE-EXPRESSION; MYD88 L265P; PKC-DELTA; TOX; MACROGLOBULINEMIA;
   DIFFERENTIATION; IDENTIFICATION
AB Purpose: Primary central nervous system lymphoma (PCNSL) is an aggressive non-Hodgkin lymphoma confined to the central nervous system. Whether there is a PCNSL-specific genomic signature and, if so, how it differs from systemic diffuse large B-cell lymphoma (DLBCL) is uncertain.
   Experimental Design: We performed a comprehensive genomic study of tumor samples from 19 immunocompetent PCNSL patients. Testing comprised array-comparative genomic hybridization and whole exome sequencing.
   Results: Biallelic inactivation of TOX and PRKCD was recurrently found in PCNSL but not in systemic DLBCL, suggesting a specific role in PCNSL pathogenesis. In addition, we found a high prevalence of MYD88 mutations (79%) and CDKN2A biallelic loss (60%). Several genes recurrently affected in PCNSL were common with systemic DLBCL, including loss of TNFAIP3, PRDM1, GNA13, TMEM30A, TBL1XR1, B2M, CD58, activating mutations of CD79B, CARD11, and translocations IgH-BCL6. Overall, B-cell receptor/Toll-like receptor/NF-kappa B pathways were altered in >90% of PNCSL, highlighting its value for targeted therapeutic approaches. Furthermore, integrated analysis showed enrichment of pathways associated with immune response, proliferation, apoptosis, and lymphocyte differentiation.
   Conclusions: In summary, genome-wide analysis uncovered novel recurrent alterations, including TOX and PRKCD, helping to differentiate PCNSL from systemic DLBCL and related lymphomas. (C)2015 AACR.
C1 [Braggio, Esteban; Van Wier, Scott; Ojha, Juhi; Egan, Jan; Valdez, Riccardo; Tibes, Raoul; Stewart, A. Keith; Fonseca, Rafael] Mayo Clin, Scottsdale, AZ 85259 USA.
   [McPhail, Ellen; Decker, Paul A.; Eckloff, Bruce; Witzig, Thomas E.; O'Neill, Brian Patrick] Mayo Clin, Rochester, MN USA.
   [Asmann, Yan W.] Mayo Clin, Jacksonville, FL 32224 USA.
   [da Silva, Jackline Ayres] Natl Inst Canc, Sao Paulo, Brazil.
   [Schiff, David; Lopes, M. Beatriz] Univ Virginia, Charlottesville, VA USA.
C3 Mayo Clinic; Mayo Clinic Phoenix; Mayo Clinic; Mayo Clinic; University
   of Virginia
RP Braggio, E (corresponding author), Mayo Clin, 13400 Shea Blvd,MCCRB Bldg,Room 3-029, Scottsdale, AZ 85259 USA.
EM
RI Lopes, Maria/GXF-7507-2022; McPhail, Ellen D. Remstein/AAF-3474-2020;
   Tibes, Raoul/AAV-1683-2020; Ayres-Silva, Jackline/GLS-6204-2022;
   Ayres-Silva, Jackline/I-7733-2014
OI McPhail, Ellen D. Remstein/0000-0001-6918-3376; Ayres-Silva,
   Jackline/0000-0003-2225-7723; Ayres-Silva, Jackline/0000-0003-2225-7723;
   Tibes, Raoul/0000-0002-1372-8733; Braggio, Esteban/0000-0003-3860-4830
FU Mayo Clinic SPORE in Brain Cancer [P50 CA108961]; Iowa/Mayo Lymphoma
   SPORE [P50 CA97274]; George Haub Family Career Development Award Fund in
   Cancer Research; Steve's Run grant; Henry Predolin Foundation
FX This work was supported by the Mayo Clinic SPORE in Brain Cancer (P50
   CA108961; Principal Investigator: B.P. O'Neill), the Iowa/Mayo Lymphoma
   SPORE (P50 CA97274; Principal Investigators: George Weiner and Thomas
   Witzig), the George Haub Family Career Development Award Fund in Cancer
   Research, Steve's Run grant and the Henry Predolin Foundation.
CR Adzhubei IA, 2010, NAT METHODS, V7, P248, DOI 10.1038/nmeth0410-248
   Aliahmad P, 2004, J EXP MED, V199, P1089, DOI 10.1084/jem.20040051
   Aliahmad P, 2008, J EXP MED, V205, P245, DOI 10.1084/jem.20071944
   Aliahmad P, 2010, NAT IMMUNOL, V11, P945, DOI 10.1038/ni.1930
   [Anonymous], 2008, WHO CLASSIFICATION T
   Belot A, 2013, ARTHRITIS RHEUM-US, V65, P2161, DOI 10.1002/art.38008
   Booman M, 2008, J PATHOL, V216, P209, DOI 10.1002/path.2399
   Braggio E, 2011, CLIN CANCER RES, V17, P4245, DOI 10.1158/1078-0432.CCR-11-0395
   Cady FM, 2008, J CLIN ONCOL, V26, P4814, DOI 10.1200/JCO.2008.16.1455
   Cataldo KA, 1999, AM J SURG PATHOL, V23, P1386, DOI 10.1097/00000478-199911000-00009
   Courts C, 2008, J NEUROPATH EXP NEUR, V67, P720, DOI 10.1097/NEN.0b013e31817dd02d
   Descatoire M, 2014, J EXP MED, V211, P986, DOI 10.1084/jem.20132203
   Ferreri AJ, 2011, CANCER RES, V118, P510
   Ghayur T, 1996, J EXP MED, V184, P2399, DOI 10.1084/jem.184.6.2399
   Gonzalez-Aguilar A, 2012, CLIN CANCER RES, V18, P5203, DOI 10.1158/1078-0432.CCR-12-0845
   Kay NE, 2010, CANCER GENET CYTOGEN, V203, P161, DOI 10.1016/j.cancergencyto.2010.09.003
   Ke G, 2013, ONCOGENE, V32, P3019, DOI 10.1038/onc.2012.323
   Kraan W, 2013, BLOOD CANCER J, V3, DOI 10.1038/bcj.2013.28
   Kuehn HS, 2013, BLOOD, V121, P3117, DOI 10.1182/blood-2012-12-469544
   Larson DE, 2012, BIOINFORMATICS, V28, P311, DOI 10.1093/bioinformatics/btr665
   Limnander A, 2011, NAT IMMUNOL, V12, P425, DOI 10.1038/ni.2016
   Lu ZM, 1997, MOL CELL BIOL, V17, P3418, DOI 10.1128/MCB.17.6.3418
   McKenna A, 2010, GENOME RES, V20, P1297, DOI 10.1101/gr.107524.110
   MISCHAK H, 1993, J BIOL CHEM, V268, P6090
   Miyamoto A, 2002, NATURE, V416, P865, DOI 10.1038/416865a
   Montesinos-Rongen M, 2008, LEUKEMIA, V22, P400, DOI 10.1038/sj.leu.2405019
   Montesinos-Rongen M, 2002, J NEUROPATH EXP NEUR, V61, P926, DOI 10.1093/jnen/61.10.926
   Montesinos-Rongen M, 2012, ACTA NEUROPATHOL, V124, P905, DOI 10.1007/s00401-012-1064-7
   Montesinos-Rongen M, 2011, ACTA NEUROPATHOL, V122, P791, DOI 10.1007/s00401-011-0891-2
   Montesinos-Rongen M, 2010, ACTA NEUROPATHOL, V120, P529, DOI 10.1007/s00401-010-0709-7
   Ngo VN, 2011, NATURE, V470, P115, DOI 10.1038/nature09671
   Pek JW, 2011, P NATL ACAD SCI USA, V108, P12007, DOI 10.1073/pnas.1106245108
   PESSINO A, 1995, BIOCHEM J, V312, P549, DOI 10.1042/bj3120549
   Puente XS, 2011, NATURE, V475, P101, DOI 10.1038/nature10113
   Quesada V, 2012, NAT GENET, V44, P47, DOI 10.1038/ng.1032
   Remstein ED, 2006, AM J SURG PATHOL, V30, P1546, DOI 10.1097/01.pas.0000213275.60962.2a
   Richter J, 2009, BMC CANCER, V9, DOI 10.1186/1471-2407-9-455
   Robinson JT, 2011, NAT BIOTECHNOL, V29, P24, DOI 10.1038/nbt.1754
   Rothberg JM, 2011, NATURE, V475, P348, DOI 10.1038/nature10242
   Rubenstein JL, 2006, BLOOD, V107, P3716, DOI 10.1182/blood-2005-03-0897
   Schwindt H, 2009, LEUKEMIA, V23, P1875, DOI 10.1038/leu.2009.120
   Sung CO, 2011, BLOOD, V117, P1291, DOI 10.1182/blood-2010-07-297861
   Treon SP, 2012, NEW ENGL J MED, V367, P826, DOI 10.1056/NEJMoa1200710
   Tun HW, 2008, BLOOD, V111, P3200, DOI 10.1182/blood-2007-10-119099
   Varettoni M, 2013, BLOOD, V121, P2522, DOI 10.1182/blood-2012-09-457101
   Wang LL, 2011, NEW ENGL J MED, V365, P2497, DOI 10.1056/NEJMoa1109016
   Wilkinson B, 2002, NAT IMMUNOL, V3, P272, DOI 10.1038/ni767
   Xu L, 2013, BLOOD, V121, P2051, DOI 10.1182/blood-2012-09-454355
NR 48
TC 157
Z9 159
U1 0
U2 8
PU AMER ASSOC CANCER RESEARCH
PI PHILADELPHIA
PA 615 CHESTNUT ST, 17TH FLOOR, PHILADELPHIA, PA 19106-4404 USA
SN 1078-0432
EI 1557-3265
J9 CLIN CANCER RES
JI Clin. Cancer Res.
PD SEP 1
PY 2015
VL 21
IS 17
BP 3986
EP 3994
DI 10.1158/1078-0432.CCR-14-2116
PG 9
WC Oncology
WE Science Citation Index Expanded (SCI-EXPANDED)
SC Oncology
GA CS2OF
UT WOS:000361910000022
PM 25991819
OA Green Accepted, Bronze
DA 2024-06-03
ER

PT J
AU Coupland, SE
   Heimann, H
   Bechrakis, NE
AF Coupland, SE
   Heimann, H
   Bechrakis, NE
TI Primary intraocular lymphoma: a review of the clinical,
   histopathological and molecular biological features
SO GRAEFES ARCHIVE FOR CLINICAL AND EXPERIMENTAL OPHTHALMOLOGY
LA English
DT Review
ID NERVOUS-SYSTEM LYMPHOMA; RETICULUM-CELL SARCOMA; NON-HODGKINS-LYMPHOMA;
   PRIMARY CNS LYMPHOMA; POLYMERASE-CHAIN-REACTION; T-CELL; B-CELL; OCULAR
   INVOLVEMENT; RADIATION-THERAPY; PIGMENT-EPITHELIUM
AB Introduction: Primary intraocular lymphoma (PIOL) is a rare non-Hodgkin lymphoma which arises in the retina or the vitreous. It can occur either together with or independently of primary cerebral nervous system lymphoma (PCNSL); the incidence of the latter has significantly increased over the past three decades. PIOL remains one of the most difficult diagnoses to establish, particularly due to its ability to mimic other diseases in the eye and to the limited material which is often available for examination. Methods: The article reviews the clinical, histopathological, molecular biological and biochemical approaches to the diagnosis of PIOL. The differential diagnoses, including other lymphomatous manifestations in the eye, e.g. primary uveal lymphoma, as well as non-neoplastic uveal diseases are addressed. Furthermore, the treatment strategies for PIOL are summarised. Results: Diagnostic progress has been made in various fields, including flow cytometry and immunocytology, cytokine analysis, and as well as molecular biological analysis of the immunoglobulin heavy and light chains using polymerase chain reaction on both fixed and non-fixed material. The optimal therapy of PIOL remains to be determined: the current trends suggest that combined radiotherapy and chemotherapy, as well as intravitreal chemotherapy, are of value. Novel therapies which may have a role in the future include oral trofosfamide. Conclusion: Our understanding of the pathogenesis of PIOL/PCNSL remains far from complete. Intensified efforts must be made to determine the cell of origin of PIOL, as well as to establish "molecular signatures", which could be used to decrease diagnostic delay. Further studies, possibly prospective ones, are required to establish the optimal therapy for initial and recurrent disease.
C1 Charite, Dept Pathol, D-12200 Berlin, Germany.
   Charite, Dept Ophthalmol, D-12200 Berlin, Germany.
C3 Free University of Berlin; Humboldt University of Berlin; Charite
   Universitatsmedizin Berlin; Free University of Berlin; Humboldt
   University of Berlin; Charite Universitatsmedizin Berlin
RP Charite, Dept Pathol, Campus Benjamin Franklin,Hindenburgdamm 30, D-12200 Berlin, Germany.
EM
RI Heimann, Heinrich/AAP-8747-2020
OI Heimann, Heinrich/0000-0002-3298-4644; Coupland,
   Sarah/0000-0002-1464-2069
CR Abrey LE, 1998, J CLIN ONCOL, V16, P859, DOI 10.1200/JCO.1998.16.3.859
   Akpek EK, 1999, OPHTHALMOLOGY, V106, P1805, DOI 10.1016/S0161-6420(99)90341-X
   Anagnostopoulos I, 2000, PATHOLOGE, V21, P178, DOI 10.1007/s002920050384
   APPEN RE, 1975, ARCH OPHTHALMOL-CHIC, V93, P123
   Barr C C, 1975, Surv Ophthalmol, V19, P224
   Basso U, 2002, EUR J CANCER, V38, P1298, DOI 10.1016/S0959-8049(02)00031-X
   Batchelor TT, 2003, CLIN CANCER RES, V9, P711
   BAUMANN MA, 1986, CANCER-AM CANCER SOC, V57, P1273, DOI 10.1002/1097-0142(19860401)57:7<1273::AID-CNCR2820570702>3.0.CO;2-C
   BEASLEY H, 1961, AM J OPHTHALMOL, V51, P1294, DOI 10.1016/0002-9394(61)92476-X
   Bechrakis NE, 2002, OPHTHALMOLOGY, V109, P235, DOI 10.1016/S0161-6420(01)00931-9
   BENEZRA D, 1989, BRIT J OPHTHALMOL, V73, P846, DOI 10.1136/bjo.73.10.846
   BROWN SM, 1994, SURV OPHTHALMOL, V39, P133, DOI 10.1016/0039-6257(94)90158-9
   Buggage RR, 2001, CURR OPIN ONCOL, V13, P137, DOI 10.1097/00001622-200105000-00001
   Cassoux N, 2000, OCUL IMMUNOL INFLAMM, V8, P243, DOI 10.1076/ocii.8.4.243.6463
   CHAN CC, 1995, AM J OPHTHALMOL, V120, P671, DOI 10.1016/S0002-9394(14)72217-2
   Chan Chi-Chao, 2002, Curr Opin Ophthalmol, V13, P411, DOI 10.1097/00055735-200212000-00012
   Chang TS, 1996, ARCH OPHTHALMOL-CHIC, V114, P669, DOI 10.1001/archopht.1996.01100130661005
   CHAR DH, 1988, BRIT J OPHTHALMOL, V72, P905, DOI 10.1136/bjo.72.12.905
   CHAR DH, 1988, OPHTHALMOLOGY, V95, P625
   CHAR DH, 1981, AM J OPHTHALMOL, V91, P480, DOI 10.1016/0002-9394(81)90236-1
   CHEUNG MK, 1994, AM J OPHTHALMOL, V118, P457, DOI 10.1016/S0002-9394(14)75796-4
   CIRICILLO SF, 1990, J NEUROSURG, V73, P720, DOI 10.3171/jns.1990.73.5.0720
   Ciulla TA, 1997, AM J OPHTHALMOL, V123, P420, DOI 10.1016/S0002-9394(14)70150-3
   Ciulla TA, 1997, SURV OPHTHALMOL, V41, P467, DOI 10.1016/S0039-6257(97)00015-5
   Cochereau I, 1996, AM J OPHTHALMOL, V121, P322, DOI 10.1016/S0002-9394(14)70284-3
   Cockerham GC, 2000, OPHTHALMOLOGY, V107, P151, DOI 10.1016/S0161-6420(99)00025-1
   COOK C, 1954, BRIT J OPHTHALMOL, V38, P182, DOI 10.1136/bjo.38.3.182
   COOPER EL, 1951, AM J OPHTHALMOL, V34, P1153, DOI 10.1016/0002-9394(51)90691-5
   Corn BW, 1997, CANCER J SCI AM, V3, P52
   CORRIVEAU C, 1986, CAN J OPHTHALMOL, V21, P144
   Coupland SE, 2005, GRAEF ARCH CLIN EXP, V243, P189, DOI 10.1007/s00417-004-0890-2
   Coupland SE, 2003, GRAEF ARCH CLIN EXP, V241, P860, DOI 10.1007/s00417-003-0749-y
   Coupland SE, 2002, J PATHOL, V197, P333, DOI 10.1002/path.1130
   Coupland SE, 2001, OPHTHALMOLOGY, V108, P1289, DOI 10.1016/S0161-6420(01)00594-2
   Coupland SE, 1999, OPHTHALMOLOGY, V106, P2109, DOI 10.1016/S0161-6420(99)90492-X
   COUPLAND SE, 2004, IN PRESS J OPHTHALMO
   CROOKES GP, 1967, AM J OPHTHALMOL, V63, P962, DOI 10.1016/0002-9394(67)90040-2
   Davis JL, 1997, AM J OPHTHALMOL, V124, P362, DOI 10.1016/S0002-9394(14)70828-1
   DAVIS JL, 1992, OPHTHALMOLOGY, V99, P250
   Dean JM, 1996, RETINA-J RET VIT DIS, V16, P47, DOI 10.1097/00006982-199616010-00009
   DeAngelis L M, 2001, Curr Treat Options Oncol, V2, P309, DOI 10.1007/s11864-001-0024-6
   DeAngelis L M, 1994, Recent Results Cancer Res, V135, P155
   DEANGELIS LM, 1992, J CLIN ONCOL, V10, P635, DOI 10.1200/JCO.1992.10.4.635
   DeAngelis LM, 2002, J CLIN ONCOL, V20, P4643, DOI 10.1200/JCO.2002.11.013
   DeAngelis LM, 1999, CURR OPIN NEUROL, V12, P687, DOI 10.1097/00019052-199912000-00005
   DESMET MD, 1990, INT OPHTHALMOL, V14, P413, DOI 10.1007/BF00163568
   DESROCHES G, 1983, ARCH OPHTHALMOL-CHIC, V101, P725, DOI 10.1001/archopht.1983.01040010725004
   DUKER JS, 1987, RETINA-J RET VIT DIS, V7, P41, DOI 10.1097/00006982-198707010-00010
   DUKER JS, 1989, ARCH OPHTHALMOL-CHIC, V107, P446, DOI 10.1001/archopht.1989.01070010456044
   EBY NL, 1988, CANCER-AM CANCER SOC, V62, P2461, DOI 10.1002/1097-0142(19881201)62:11<2461::AID-CNCR2820621135>3.0.CO;2-M
   Engel H M, 1981, Retina, V1, P121
   ERNY BC, 1991, BRIT J OPHTHALMOL, V75, P698, DOI 10.1136/bjo.75.11.698
   ERNY BC, BR J OPHTHALMOL, V76, P128
   ESCOFFERY RF, 1985, RETINA-J RET VIT DIS, V5, P91, DOI 10.1097/00006982-198500520-00005
   Falini B, 2000, BLOOD, V95, P2084, DOI 10.1182/blood.V95.6.2084
   FAULBORN J, 1983, KLIN MONATSBL AUGENH, V183, P120, DOI 10.1055/s-2008-1054886
   Feiden W, 2002, PATHOLOGE, V23, P284, DOI 10.1007/s00292-002-0539-z
   Ferreri AJM, 2002, ANN ONCOL, V13, P531, DOI 10.1093/annonc/mdf080
   Feugier P, 2004, ANN ONCOL, V15, P129, DOI 10.1093/annonc/mdh013
   FINE HA, 1993, ANN INTERN MED, V119, P1093, DOI 10.7326/0003-4819-119-11-199312010-00007
   Fishburne BC, 1997, ARCH OPHTHALMOL-CHIC, V115, P1152, DOI 10.1001/archopht.1997.01100160322009
   FOERSTER HC, 1960, OPHTHALMOLOGY, V64, P308
   FREDRICK DR, 1989, ARCH OPHTHALMOL-CHIC, V107, P395, DOI 10.1001/archopht.1989.01070010405034
   FREEMAN LN, 1987, OPHTHALMOLOGY, V94, P1631
   Freilich RJ, 1996, NEUROLOGY, V46, P435, DOI 10.1212/WNL.46.2.435
   FREILICH RJ, 1995, NEUROL CLIN, V13, P901, DOI 10.1016/S0733-8619(18)30024-0
   GARTNER J, 1989, KLIN MONATSBL AUGENH, V131, P61
   GASS JDM, 1991, ARCH OPHTHALMOL-CHIC, V109, P1134, DOI 10.1001/archopht.1991.01080080094039
   GASS JDM, 1984, RETINA-J RET VIT DIS, V4, P135, DOI 10.1097/00006982-198400430-00001
   GASS JDM, 1967, AM J OPHTHALMOL, V64, P612
   Gill MK, 2001, SURV OPHTHALMOL, V45, P463, DOI 10.1016/S0039-6257(01)00217-X
   Goeminne JC, 1999, OPHTHALMOLOGICA, V213, P323, DOI 10.1159/000027447
   GOLDEY SH, 1989, ARCH OPHTHALMOL-CHIC, V107, P1349, DOI 10.1001/archopht.1989.01070020419047
   Gorochov G, 2003, INVEST OPHTH VIS SCI, V44, P5235, DOI 10.1167/iovs.03-0035
   Grossniklaus HE, 1998, OPHTHALMOLOGY, V105, P1265, DOI 10.1016/S0161-6420(98)97032-4
   Helbig H, 2003, OPHTHALMOLOGE, V100, P145, DOI 10.1007/s00347-002-0692-z
   HENRY JM, 1974, CANCER, V34, P1293, DOI 10.1002/1097-0142(197410)34:4<1293::AID-CNCR2820340441>3.0.CO;2-P
   Herrlinger U, 1999, J NEURO-ONCOL, V43, P219, DOI 10.1023/A:1006298201101
   HOCHBERG FH, 1988, J NEUROSURG, V68, P835, DOI 10.3171/jns.1988.68.6.0835
   Hoffman PM, 2003, EYE, V17, P513, DOI 10.1038/sj.eye.6700378
   Holz FG, 1999, RETINA-J RET VIT DIS, V19, P343, DOI 10.1097/00006982-199919040-00015
   Jaffe E.S., 2001, PATHOLOGY GENETICS T, V3
   Jahnke K, 2004, GRAEF ARCH CLIN EXP, V242, P771, DOI 10.1007/s00417-004-0947-2
   JAKOBIEC FA, 1987, OPHTHALMOLOGY, V94, P397
   JENSEN OA, 1994, GRAEF ARCH CLIN EXP, V232, P148, DOI 10.1007/BF00176784
   KAPLAN HJ, 1980, ARCH OPHTHALMOL-CHIC, V98, P707, DOI 10.1001/archopht.1980.01020030701010
   Katai N, 1997, GRAEF ARCH CLIN EXP, V235, P431, DOI 10.1007/BF00947062
   Kawai Y, 1999, INT J HEMATOL, V69, P256
   KELTNER JL, 1977, ARCH OPHTHALMOL-CHIC, V95, P645, DOI 10.1001/archopht.1977.04450040111017
   KINCAID MC, 1983, SURV OPHTHALMOL, V27, P211, DOI 10.1016/0039-6257(83)90123-6
   KIRMANI MH, 1987, BRIT J OPHTHALMOL, V71, P748, DOI 10.1136/bjo.71.10.748
   KLINGELE TG, 1975, AM J OPHTHALMOL, V79, P39, DOI 10.1016/0002-9394(75)90453-5
   KOHNO T, 1993, OPHTHALMOLOGY, V100, P1794
   Küker W, 2002, J NEUROL, V249, P1713, DOI 10.1007/s00415-002-0919-6
   KUMAR SR, 1994, ARCH OPHTHALMOL-CHIC, V112, P954, DOI 10.1001/archopht.1994.01090190102028
   Lafaut B A, 1994, Bull Soc Belge Ophtalmol, V252, P67
   LEITCH RJ, 1993, BRIT J OPHTHALMOL, V77, P126, DOI 10.1136/bjo.77.2.126
   Levy-Clarke GA, 2001, RETINA-J RET VIT DIS, V21, P281, DOI 10.1097/00006982-200106000-00023
   LJUNG BM, 1988, ACTA CYTOL, V32, P840
   Lobo A, 2003, CLIN EXP OPHTHALMOL, V31, P155, DOI 10.1046/j.1442-9071.2003.00624.x
   Lobo A, 2003, OPHTHALMOLOGY, V110, P595, DOI 10.1016/S0161-6420(02)01895-X
   Lois N, 2000, ARCH OPHTHALMOL-CHIC, V118, P1692, DOI 10.1001/archopht.118.12.1692
   MARGOLIS L, 1980, CANCER-AM CANCER SOC, V45, P688, DOI 10.1002/1097-0142(19800215)45:4<688::AID-CNCR2820450412>3.0.CO;2-F
   MARTIN DF, 1993, OPHTHALMOLOGY, V100, P705
   Mason JO, 2003, OPHTHALMOLOGY, V110, P1241, DOI 10.1016/S0161-6420(03)00268-9
   McGavic JS, 1943, ARCH OPHTHALMOL-CHIC, V30, P179
   Merchant A, 1997, INT OPHTHALMOL CLIN, V37, P101, DOI 10.1097/00004397-199703740-00009
   Merle-Béral H, 2004, BRIT J HAEMATOL, V124, P469, DOI 10.1046/j.1365-2141.2003.04800.x
   MICHELS RG, 1975, ARCH OPHTHALMOL-CHIC, V93, P1331, DOI 10.1001/archopht.1975.01010020961005
   NATHWANI BN, 1978, CANCER-AM CANCER SOC, V41, P303, DOI 10.1002/1097-0142(197801)41:1<303::AID-CNCR2820410142>3.0.CO;2-G
   NELSON DF, 1992, INT J RADIAT ONCOL, V23, P9, DOI 10.1016/0360-3016(92)90538-S
   Palexas G N, 1995, Trans Am Ophthalmol Soc, V93, P281
   PARVER LM, 1979, ARCH OPHTHALMOL-CHIC, V97, P1505
   PAVAN PR, 1995, ARCH OPHTHALMOL-CHIC, V113, P1233, DOI 10.1001/archopht.1995.01100100021014
   Pels H, 2003, J CLIN ONCOL, V21, P4489, DOI 10.1200/JCO.2003.04.056
   PETERSON K, 1993, CANCER, V72, P843, DOI 10.1002/1097-0142(19930801)72:3<843::AID-CNCR2820720333>3.0.CO;2-#
   Plowman P N, 1993, Clin Oncol (R Coll Radiol), V5, P315, DOI 10.1016/S0936-6555(05)80909-4
   QUALMAN SJ, 1983, CANCER-AM CANCER SOC, V52, P878, DOI 10.1002/1097-0142(19830901)52:5<878::AID-CNCR2820520523>3.0.CO;2-D
   Read RW, 2002, SURV OPHTHALMOL, V47, P81, DOI 10.1016/S0039-6257(01)00305-8
   Reim H, 1990, Fortschr Ophthalmol, V87, P557
   RIDLEY ME, 1992, OPHTHALMOLOGY, V99, P1153
   ROSENTHAL AR, 1983, OPHTHALMOLOGY, V90, P899
   Rothova A, 2001, OPHTHALMOLOGY, V108, P386, DOI 10.1016/S0161-6420(00)00499-1
   Rubenstein JL, 2003, BLOOD, V101, P466, DOI 10.1182/blood-2002-06-1636
   RYAN S J, 1972, Transactions of the American Academy of Ophthalmology and Oto-Laryngology, V76, P652
   RYAN SJ, 1971, AM J OPHTHALMOL, V72, P586, DOI 10.1016/0002-9394(71)90855-5
   SAGA T, 1984, ARCH OPHTHALMOL-CHIC, V102, P399
   SCHACHAT AP, 1989, ARCH OPHTHALMOL-CHIC, V107, P697, DOI 10.1001/archopht.1989.01070010715033
   SCROGGS MW, 1990, ACTA CYTOL, V34, P401
   Shen DF, 1998, OPHTHALMOLOGY, V105, P1664, DOI 10.1016/S0161-6420(98)99036-4
   SHIELDS JA, 1974, ARCH OPHTHALMOL-CHIC, V91, P259
   SHIELDS JA, 1981, ARCH OPHTHALMOL-CHIC, V99, P2147
   Soussain C, 1996, LEUKEMIA LYMPHOMA, V23, P339, DOI 10.3109/10428199609054837
   Soussain C, 2001, J CLIN ONCOL, V19, P742, DOI 10.1200/JCO.2001.19.3.742
   STENSON S, 1981, ARCH OPHTHALMOL-CHIC, V99, P272
   STRAUCHEN JA, 1989, CANCER-AM CANCER SOC, V63, P1918, DOI 10.1002/1097-0142(19890515)63:10<1918::AID-CNCR2820631008>3.0.CO;2-D
   Triebenstein O., 1920, KLIN MONATSBL AUGENH, V64, P825
   Umland O, 2003, J HISTOCHEM CYTOCHEM, V51, P977, DOI 10.1177/002215540305100714
   Ursea R, 1997, RETINA-J RET VIT DIS, V17, P118, DOI 10.1097/00006982-199703000-00006
   VALLURI S, 1995, RETINA-J RET VIT DIS, V15, P125, DOI 10.1097/00006982-199515020-00006
   van Dongen JJM, 2003, LEUKEMIA, V17, P2257, DOI 10.1038/sj.leu.2403202
   Velez G, 2000, SURV OPHTHALMOL, V44, P518, DOI 10.1016/S0039-6257(00)00118-1
   Velez G, 2002, RETINA-J RET VIT DIS, V22, P37, DOI 10.1097/00006982-200202000-00007
   Verbraeken HE, 1997, BRIT J OPHTHALMOL, V81, P31, DOI 10.1136/bjo.81.1.31
   WENDER A, 1994, ARCH OPHTHALMOL-CHIC, V112, P450, DOI 10.1001/archopht.1994.01090160024009
   Whitcup SM, 1997, ARCH OPHTHALMOL-CHIC, V115, P1157, DOI 10.1001/archopht.1997.01100160327010
   WHITCUP SM, 1993, OPHTHALMOLOGY, V100, P1399
   White VA, 1999, ARCH OPHTHALMOL-CHIC, V117, P761
   Williams GC, 2000, ARCH OPHTHALMOL-CHIC, V118, P278
   WOLTER JR, 1963, AM J OPHTHALMOL, V55, P317, DOI 10.1016/0002-9394(63)92689-8
   Wylen EL, 1998, NEUROL RES, V20, P307, DOI 10.1080/01616412.1998.11740523
   Yahalom C, 2002, ARCH OPHTHALMOL-CHIC, V120, P204
   Yamada K, 2003, AM J OPHTHALMOL, V136, P380, DOI 10.1016/S0002-9394(03)00324-6
   Young TL, 1996, J PEDIATR OPHTHALMOL, V33, P268
   YUNIS JJ, 1982, NEW ENGL J MED, V307, P1231, DOI 10.1056/NEJM198211113072002
   ZAUBERMAN H, 1971, OPHTHALMOLOGICA, V163, P65, DOI 10.1159/000306325
NR 156
TC 157
Z9 166
U1 0
U2 7
PU SPRINGER
PI NEW YORK
PA ONE NEW YORK PLAZA, SUITE 4600, NEW YORK, NY, UNITED STATES
SN 0721-832X
EI 1435-702X
J9 GRAEF ARCH CLIN EXP
JI Graefes Arch. Clin. Exp. Ophthalmol.
PD NOV
PY 2004
VL 242
IS 11
BP 901
EP 913
DI 10.1007/s00417-004-0973-0
PG 13
WC Ophthalmology
WE Science Citation Index Expanded (SCI-EXPANDED)
SC Ophthalmology
GA 882XI
UT WOS:000225973300001
PM 15565454
DA 2024-06-03
ER

PT J
AU Sandor, V
   Stark-Vancs, V
   Pearson, D
   Nussenblat, R
   Whitcup, SM
   Brouwers, P
   Patronas, N
   Heiss, J
   Jaffe, E
   deSmet, M
   Kohler, D
   Simon, R
   Wittes, R
AF Sandor, V
   Stark-Vancs, V
   Pearson, D
   Nussenblat, R
   Whitcup, SM
   Brouwers, P
   Patronas, N
   Heiss, J
   Jaffe, E
   deSmet, M
   Kohler, D
   Simon, R
   Wittes, R
TI Phase II trial of chemotherapy alone for primary CNS and intraocular
   lymphoma
SO JOURNAL OF CLINICAL ONCOLOGY
LA English
DT Article
ID CENTRAL-NERVOUS-SYSTEM; NON-HODGKINS-LYMPHOMA; PRIMARY BRAIN LYMPHOMA;
   HIGH-DOSE METHOTREXATE; SURVIVAL; THERAPY; PHARMACOLOGY
AB Purpose: Primary CNS lymphoma (PCNSL) and primary intraocular lymphoma (IOL) are usually treated with radiation therapy alone or in combination with chemotherapy. The neurotoxicity of these treatments can be substantial. This study attempts to define the toxicity and efficacy of the treatment of this disease with chemotherapy alone.
   Patients and Methods: Fourteen nonimmunocompromised patients were accrued to a chemotherapy regimen that incorporated a 24-hour infusion of high dose methotrexate total dose of 8.4 g/m(2) with leucovorin rescue; thiotepa 35 mg/m(2); vincristine 1.4 mg/m(2); dexa methasone; and intrathecal cytarabine (Ara-C) and methotrexate (MN) administered in 21-day cycles. Seven patients were prospectively followed up with formal neuropsychologic assessments for evidence of CNS toxicity.
   Results: The response rate was 100% with 11 (79%) complete responses and three (21%) partial responses. Cumulative survival and progression-free survival rates at more than 4.5 years were 68.8% and 34.3%, respectively. Median survival has not been reached, and median progression free survival was 16.5 months. Toxicity included severe leukoencephalopathy that was clearly attributable to chemotherapy (two patients), grade 3 or 4 neutropenia in 50% of the cycles administered, ileus (one patient), and seizures (two patients). Mucositis and renal and hepatic toxicity were mild and not therapy limiting,
   Conclusion: The MN regimen is generally well tolerated and produces a high complete response rate. Chemotherapy alone should be investigated further in this disease to assess the necessity of initial radiation therapy, either alone or in combined modality regimens, for the achievement of optimal response and survival. J Clin Oncol 16:3000-3006. (C) 1998 by American Society of Clinical Oncology.
C1 NCI, Dept Neurosurg, Div Canc Treatment & Diagnosis, NIH, Bethesda, MD 20892 USA.
   NEI, NIH, Bethesda, MD 20892 USA.
   NIH, Dept Neuroradiol, Bethesda, MD 20892 USA.
   NIH, Dept Pharm, Ctr Clin, Bethesda, MD 20892 USA.
   NIH, Dept Pathol, Bethesda, MD 20892 USA.
   NIH, Div Biostat, Bethesda, MD 20892 USA.
C3 National Institutes of Health (NIH) - USA; NIH National Cancer Institute
   (NCI); National Institutes of Health (NIH) - USA; NIH National Eye
   Institute (NEI); National Institutes of Health (NIH) - USA; National
   Institutes of Health (NIH) - USA; NIH Clinical Center (CC); National
   Institutes of Health (NIH) - USA; National Institutes of Health (NIH) -
   USA
RP Wittes, R (corresponding author), NCI, Dept Neurosurg, Div Canc Treatment & Diagnosis, NIH, Bld 31,Rm 3A44,31 Ctr Dr, Bethesda, MD 20892 USA.
EM
RI Heiss, John/AAL-8281-2021; Jaffe, Elaine S/G-8984-2014
OI Heiss, John/0000-0002-3890-0165; 
CR BALIS FM, 1989, AM J PEDIAT HEMATOL, V11, P74
   BLAY JY, 1995, BLOOD, V86, P2922
   BLEYER WA, 1977, CANCER TREAT REV, V4, P87, DOI 10.1016/S0305-7372(77)80007-8
   DEANGELIS LM, 1992, J CLIN ONCOL, V10, P635, DOI 10.1200/JCO.1992.10.4.635
   EBY NL, 1988, CANCER-AM CANCER SOC, V62, P2461, DOI 10.1002/1097-0142(19881201)62:11<2461::AID-CNCR2820621135>3.0.CO;2-M
   FINE HA, 1993, ANN INTERN MED, V119, P1093, DOI 10.7326/0003-4819-119-11-199312010-00007
   GABBAI AA, 1989, J NEUROSURG, V70, P190, DOI 10.3171/jns.1989.70.2.0190
   GILL PS, 1985, AM J MED, V78, P742, DOI 10.1016/0002-9343(85)90277-3
   GLANTZ MJ, 1995, CANCER, V75, P2919, DOI 10.1002/1097-0142(19950615)75:12<2919::AID-CNCR2820751220>3.0.CO;2-9
   HENRY JM, 1974, CANCER, V34, P1293, DOI 10.1002/1097-0142(197410)34:4<1293::AID-CNCR2820340441>3.0.CO;2-P
   HOCHBERG FH, 1991, J NEURO-ONCOL, V10, P191
   MACMAHON EME, 1991, LANCET, V338, P969, DOI 10.1016/0140-6736(91)91837-K
   MILLER DC, 1994, CANCER-AM CANCER SOC, V74, P1383, DOI 10.1002/1097-0142(19940815)74:4<1383::AID-CNCR2820740432>3.0.CO;2-1
   MURRAY K, 1986, J NEUROSURG, V65, P600, DOI 10.3171/jns.1986.65.5.0600
   NELSON DF, 1992, INT J RADIAT ONCOL, V23, P9, DOI 10.1016/0360-3016(92)90538-S
   NEUWELT EA, 1991, J CLIN ONCOL, V9, P1580, DOI 10.1200/JCO.1991.9.9.1580
   SAGERMAN RH, 1967, RADIOLOGY, V88, P552, DOI 10.1148/88.3.542
   Schultz C, 1996, J CLIN ONCOL, V14, P556, DOI 10.1200/JCO.1996.14.2.556
   SKARIN AT, 1977, BLOOD, V50, P1039
   Thall PF, 1996, J CLIN ONCOL, V14, P296, DOI 10.1200/JCO.1996.14.1.296
   TRUDEAU M, 1988, AM J CLIN ONCOL-CANC, V11, P126, DOI 10.1097/00000421-198804000-00008
NR 21
TC 157
Z9 163
U1 0
U2 2
PU W B SAUNDERS CO
PI PHILADELPHIA
PA INDEPENDENCE SQUARE WEST CURTIS CENTER, STE 300, PHILADELPHIA, PA
   19106-3399 USA
SN 0732-183X
J9 J CLIN ONCOL
JI J. Clin. Oncol.
PD SEP
PY 1998
VL 16
IS 9
BP 3000
EP 3006
DI 10.1200/JCO.1998.16.9.3000
PG 7
WC Oncology
WE Science Citation Index Expanded (SCI-EXPANDED)
SC Oncology
GA 116PY
UT WOS:000075734800014
PM 9738568
DA 2024-06-03
ER

PT J
AU Houillier, C
   Taillandier, L
   Dureau, S
   Lamy, T
   Laadhari, M
   Chinot, O
   Moluçon-Chabrot, C
   Soubeyran, P
   Gressin, R
   Choquet, S
   Damaj, G
   Thyss, A
   Abraham, J
   Delwail, V
   Gyan, E
   Sanhes, L
   Cornillon, J
   Garidi, R
   Delmer, A
   Tanguy, ML
   Al Jijakli, A
   Morel, P
   Bourquard, P
   Moles, MP
   Chauchet, A
   Gastinne, T
   Constans, JM
   Langer, A
   Martin, A
   Moisson, P
   Lacomblez, L
   Martin-Duverneuil, N
   Delgadillo, D
   Turbiez, I
   Feuvret, L
   Cassoux, N
   Touitou, V
   Ricard, D
   Hoang-Xuan, K
   Soussain, C
AF Houillier, Caroline
   Taillandier, Luc
   Dureau, Sylvain
   Lamy, Thierry
   Laadhari, Mouna
   Chinot, Olivier
   Molucon-Chabrot, Cecile
   Soubeyran, Pierre
   Gressin, Remy
   Choquet, Sylvain
   Damaj, Gandhi
   Thyss, Antoine
   Abraham, Julie
   Delwail, Vincent
   Gyan, Emmanuel
   Sanhes, Laurence
   Cornillon, Jerome
   Garidi, Reda
   Delmer, Alain
   Tanguy, Marie-Laure
   Al Jijakli, Ahmad
   Morel, Pierre
   Bourquard, Pascal
   Moles, Marie-Pierre
   Chauchet, Adrien
   Gastinne, Thomas
   Constans, Jean-Marc
   Langer, Adriana
   Martin, Antoine
   Moisson, Patricia
   Lacomblez, Lucette
   Martin-Duverneuil, Nadine
   Delgadillo, Daniel
   Turbiez, Isabelle
   Feuvret, Loic
   Cassoux, Nathalie
   Touitou, Valerie
   Ricard, Damien
   Hoang-Xuan, Khe
   Soussain, Carole
CA Intergrp GOELAMS-ANOCEF
   LOC Network CNS Lymphoma
TI Radiotherapy or Autologous Stem-Cell Transplantation for Primary CNS
   Lymphoma in Patients 60 Years of Age and Younger: Results o the
   Intergroup ANOCEF-GOELAMS Randomized Phase II PRECIS Study
SO JOURNAL OF CLINICAL ONCOLOGY
LA English
DT Article
ID HIGH-DOSE METHOTREXATE; NERVOUS-SYSTEM LYMPHOMA; WHOLE-BRAIN
   RADIOTHERAPY; INTERNATIONAL EXTRANODAL LYMPHOMA; LONG-TERM SURVIVAL;
   INTENSIVE CHEMOTHERAPY; INTRAOCULAR LYMPHOMA; REFRACTORY PRIMARY;
   FOLLOW-UP; THIOTEPA
AB PURPOSE To determine the efficacy and toxicity of chemoimmunotherapy followed by either whole-brain radiotherapy (WBRT) or intensive chemotherapy and autologous stem-cell transplantation (ASCT) as a first-line treatment of primary CNS lymphoma (PCNSL).
   PATIENTS AND METHOL Immunocompetent patients (18 to 60 years of age) with untreated PCNSL were randomly assigned to receive WBRT or ASCT as consolidation treatment after induction chemotherapy consisting of two cycles of R-MBVP (rituximab 375 mg/m(2) day (D) 1, methotrexate 3 g/m(2) Dl; D15, VP16 100 mg/m(2) D2, BCNU 100 mg/m(2) D3, prednisone 60 mg/kg/d D1-D5) followed by two cycles of R-AraC (rituximab 375 mg/m(2) D1, cytarabine 3 g/m(2) D1 to D2). Intensive chemotherapy consisted of thiotepa (250 mg/m(2) /d D9; D8; D7), busulfan (8 mg/kg D6 through D4), and cyclophosphamide (60 mg/kg/d D3; D2). WBRT delivered 40 Gy (2 Gy/ fraction). The primary end point was 2-year progression-free survival. Cognitive outcome was the main secondary end point. Analysis was intention to treat in a noncomparative phase II trial.
   RESULTS Between October 2008 and February 2014, 140 patients were recruited from 23 French centers. Both WBRT and ASCT met the predetermined threshold (among the first 38 patients in each group, at least 24 patients were alive and disease free at 2 years). The 2-year progression-free survival rates were 63% (95% CI, 49% to 81%) and 87% (95% CI, 77% to 98%) in the WBRT and ASCT arms, respectively. Toxicity deaths were recorded in one and five patients after WBRT and ASCT, respectively. Cognitive impairment was observed after WBRT, whereas cognitive functions were preserved or improved after ASCT.
   CONCLUSION WBRT and ASCT are effective consolidation treatments for patients with PCNSL who are 60 years of age and younger. The efficacy end points tended to favor the ASCT arm. The specific risk of each procedure should be considered. (C) 2019 by American Society of Clinical Oncology
C1 [Houillier, Caroline; Choquet, Sylvain; Lacomblez, Lucette; Martin-Duverneuil, Nadine; Delgadillo, Daniel; Feuvret, Loic; Touitou, Valerie; Hoang-Xuan, Khe] Grp Hosp Univ Pitie Salpetriere, Paris, France.
   [Taillandier, Luc] CHU Nancy, Nancy, France.
   [Dureau, Sylvain; Laadhari, Mouna; Langer, Adriana; Moisson, Patricia; Turbiez, Isabelle; Cassoux, Nathalie; Soussain, Carole] Inst Curie, Site St Cloud,35 Rue Dailly, F-92210 St Cloud, France.
   [Lamy, Thierry] CHU Rennes, Rennes, France.
   [Chinot, Olivier] CHU Timone, Marseille, France.
   [Molucon-Chabrot, Cecile] CHU Clermont Ferrand, Clermont Ferrand, France.
   [Soubeyran, Pierre] Inst Bergonie, Bordeaux, France.
   [Gressin, Remy] CHU Grenoble, La Tronche, France.
   [Damaj, Gandhi; Constans, Jean-Marc] CHU Amiens, Salouel, France.
   [Thyss, Antoine] Ctr Antoine Lacassagne, Nice, France.
   [Abraham, Julie] CHU Limoges, Limoges, France.
   [Delwail, Vincent] CHU Poitiers, Poitiers, France.
   [Gyan, Emmanuel; Tanguy, Marie-Laure] Ctr Hosp Reg Univ Tours, Tours, France.
   [Sanhes, Laurence] Ctr Hosp Perpignan, Perpignan, France.
   [Cornillon, Jerome] Inst Cancerol Loire, St Priest En Jarez, France.
   [Garidi, Reda] Ctr Hosp St Quentin, St Quentin en Yvelines, France.
   [Delmer, Alain] CHU Reims, Reims, France.
   [Al Jijakli, Ahmad] Ctr Hosp Argenteuil, Argenteuil, France.
   [Morel, Pierre] Ctr Hosp Lens, Lens, France.
   [Bourquard, Pascal] CHU Nimes, Nimes, France.
   [Moles, Marie-Pierre] CHU Angers, Angers, France.
   [Chauchet, Adrien] CHU Besancon, Besancon, France.
   [Gastinne, Thomas] CHU Nantes, Nantes, France.
   [Martin, Antoine] Hop Avicenne, Bobigny, France.
   [Ricard, Damien] Hop Instruct Armees Percy, Clamart, France.
C3 Sorbonne Universite; Assistance Publique Hopitaux Paris (APHP); Hopital
   Universitaire Pitie-Salpetriere - APHP; CHU de Nancy; UNICANCER;
   Universite PSL; Institut Curie; CHU Rennes; Universite de Rennes;
   Aix-Marseille Universite; Assistance Publique-Hopitaux de Marseille; CHU
   Clermont Ferrand; UNICANCER; Institut Bergonie; CHU Grenoble Alpes;
   Communaute Universite Grenoble Alpes; Universite Grenoble Alpes (UGA);
   Universite de Picardie Jules Verne (UPJV); CHU Amiens; UNICANCER; Centre
   Antoine Lacassagne; CHU Limoges; CHU Poitiers; Universite de Poitiers;
   CHU Tours; Institut de Cancerologie de la Loire; Universite Paris
   Saclay; CHU de Reims; Universite de Reims Champagne-Ardenne; Universite
   de Montpellier; CHU de Nimes; Universite d'Angers; Centre Hospitalier
   Universitaire d'Angers; Universite de Franche-Comte; CHU Besancon;
   Nantes Universite; CHU de Nantes; Universite Paris 13; Assistance
   Publique Hopitaux Paris (APHP); Hopital Universitaire Avicenne - APHP
RP Soussain, C (corresponding author), Inst Curie, Site St Cloud,35 Rue Dailly, F-92210 St Cloud, France.
EM
RI Hoang-Xuan, Khe/O-7527-2017; Langer, Adriana/ABF-4247-2021; choquet,
   sylvain/O-7952-2017; CHINOT, Olivier/M-9196-2018; Soubeyran,
   Pierre-Louis/F-4523-2012
OI choquet, sylvain/0000-0002-7791-0470; Hoang Xuan,
   Khe/0000-0003-4072-1999; Delmer, Alain/0000-0002-1430-2574; Dureau,
   Sylvain/0000-0002-7104-185X; Delwail, Vincent/0000-0003-0680-1418;
   Soubeyran, Pierre-Louis/0000-0003-3244-7299; Houillier,
   Caroline/0000-0003-2931-7522
FU French Government [PHRC-K 2007 AOM 07183]; Roche; Amgen; Pierre Fabre
FX Supported by Grant No. PHRC-K 2007 AOM 07183 from the French Government
   and by Roche, Amgen, and Pierre Fabre.
CR Abrey LE, 2005, J CLIN ONCOL, V23, P5034, DOI 10.1200/JCO.2005.13.524
   Abrey LE, 1998, J CLIN ONCOL, V16, P859, DOI 10.1200/JCO.1998.16.3.859
   Batchelor T, 2003, J CLIN ONCOL, V21, P1044, DOI 10.1200/JCO.2003.03.036
   Bessell EM, 2004, INT J RADIAT ONCOL, V59, P501, DOI 10.1016/j.ijrobp.2003.11.001
   Chen YB, 2015, CANCER-AM CANCER SOC, V121, P226, DOI 10.1002/cncr.29023
   Correa DD, 2012, NEURO-ONCOLOGY, V14, P101, DOI 10.1093/neuonc/nor186
   DEANGELIS LM, 1992, J CLIN ONCOL, V10, P635, DOI 10.1200/JCO.1992.10.4.635
   Doolittle ND, 2013, NEUROLOGY, V81, P84, DOI 10.1212/WNL.0b013e318297eeba
   Ekenel M, 2008, CANCER-AM CANCER SOC, V113, P1025, DOI 10.1002/cncr.23670
   Ferreri AJM, 2017, LANCET HAEMATOL, V4, pE510, DOI 10.1016/S2352-3026(17)30174-6
   Ferreri AJM, 2016, LANCET HAEMATOL, V3, pE217, DOI 10.1016/S2352-3026(16)00036-3
   Gavrilovic IT, 2006, J CLIN ONCOL, V24, P4570, DOI 10.1200/JCO.2006.06.6910
   Gerard LM, 2011, LEUKEMIA LYMPHOMA, V52, P1882, DOI 10.3109/10428194.2011.584004
   Ghesquières H, 2010, ANN ONCOL, V21, P842, DOI 10.1093/annonc/mdp529
   Illerhaus G, 2016, LANCET HAEMATOL, V3, pE388, DOI 10.1016/S2352-3026(16)30050-3
   Kasenda B, 2017, LEUKEMIA, V31, P2623, DOI 10.1038/leu.2017.170
   Kasenda B, 2015, ANN ONCOL, V26, P608, DOI 10.1093/annonc/mdv002
   Morris PG, 2013, J CLIN ONCOL, V31, P3971, DOI 10.1200/JCO.2013.50.4910
   O'Brien P, 2000, J CLIN ONCOL, V18, P519, DOI 10.1200/JCO.2000.18.3.519
   Omuro A, 2011, J NEURO-ONCOL, V104, P323, DOI 10.1007/s11060-010-0497-x
   Omuro AMP, 2005, ARCH NEUROL-CHICAGO, V62, P1595, DOI 10.1001/archneur.62.10.1595
   Omuro A, 2015, BLOOD, V125, P1403, DOI 10.1182/blood-2014-10-604561
   Poortmans PMP, 2003, J CLIN ONCOL, V21, P4483, DOI 10.1200/JCO.2003.03.108
   Scordo M, 2017, BIOL BLOOD MARROW TR, V23, P38, DOI 10.1016/j.bbmt.2016.09.024
   Soussain C, 2008, J CLIN ONCOL, V26, P2512, DOI 10.1200/JCO.2007.13.5533
   Soussain C, 2012, HAEMATOL-HEMATOL J, V97, P1751, DOI 10.3324/haematol.2011.060434
   Thiel E, 2010, LANCET ONCOL, V11, P1036, DOI 10.1016/S1470-2045(10)70229-1
   Welch MR, 2015, LEUKEMIA LYMPHOMA, V56, P361, DOI 10.3109/10428194.2014.916800
NR 28
TC 156
Z9 163
U1 1
U2 17
PU AMER SOC CLINICAL ONCOLOGY
PI ALEXANDRIA
PA 2318 MILL ROAD, STE 800, ALEXANDRIA, VA 22314 USA
SN 0732-183X
EI 1527-7755
J9 J CLIN ONCOL
JI J. Clin. Oncol.
PD APR 1
PY 2019
VL 37
IS 10
BP 823
EP +
DI 10.1200/JCO.18.00306
PG 12
WC Oncology
WE Science Citation Index Expanded (SCI-EXPANDED)
SC Oncology
GA HS5SS
UT WOS:000463932000008
PM 30785830
HC Y
HP N
DA 2024-06-03
ER

PT J
AU Barajas, RF
   Rubenstein, JL
   Chang, JS
   Hwang, J
   Cha, S
AF Barajas, R. F., Jr.
   Rubenstein, J. L.
   Chang, J. S.
   Hwang, J.
   Cha, S.
TI Diffusion-Weighted MR Imaging Derived Apparent Diffusion Coefficient Is
   Predictive of Clinical Outcome in Primary Central Nervous System
   Lymphoma
SO AMERICAN JOURNAL OF NEURORADIOLOGY
LA English
DT Article
ID PRIMARY CNS; GENE-EXPRESSION; CHEMOTHERAPY; BIOMARKER; SURVIVAL;
   IDENTIFICATION; METHOTREXATE; RADIOSURGERY; DIAGNOSIS; MAP
AB MATERIALS AND METHODS: PCNSL tumors from 18 immunocompetent patients, treated uniformly with methotrexate-based chemotherapy, were studied with pretherapeutic DWI. Enhancing lesions were diagnosed by pathologic analysis as high-grade B-cell lymphomas, Regions of interest were placed around all enhancing lesions allowing calculation of mean, 25th percentile (ADC(25%)), and minimum ADC values. Histopathologic tumor cellularity was quantitatively measured in all patients. High and low ADC groups were stratified by the median ADC value of the cohort. The Welch t test assessed differences between groups. The Pearson correlation examined relationships between ADC measurements and tumor cellular density. Single and multivanable survival analysis was performed.
   BACKGROUND AND PURPOSE: There is evidence that increased tumor cellular density within diagnostic specimens of primary central nervous system lymphoma (PCNSL) may have significant prognostic implications. Because cellular density may influence measurements of apparent diffusion coefficient (ADC) by using diffusion-weighted MR imaging (DWI), we hypothesized that ADC measured from contrast-enhancing regions might correlate with clinical outcome in patients with PCNSL.
   RESULTS: We detected significant intra- and intertumor heterogeneity in ADC measurements. An inverse correlation between cellular density and ADC measurements was observed (P < .05). ADC(25%) measurements less than the median value of 692 (low ADC group) were associated with significantly shorter progression-free and overall survival. Patients with improved clinical outcome were noted to exhibit a significant decrease in ADC measurements following high-dose methotrexate chemotherapy.
   CONCLUSIONS: Our study provides evidence that ADC measurements within contrast-enhancing regions of PCNSL tumors may provide noninvasive insight into clinical outcome.
C1 [Barajas, R. F., Jr.; Chang, J. S.; Cha, S.] Univ Calif San Francisco, Dept Radiol, Neuroradiol Sect, San Francisco, CA 94143 USA.
   [Rubenstein, J. L.] Univ Calif San Francisco, Div Hematol Oncol, San Francisco, CA 94143 USA.
   [Hwang, J.] Univ Calif San Francisco, Helen Diller Family Comprehens Canc Ctr, San Francisco, CA 94143 USA.
C3 University of California System; University of California San Francisco;
   University of California System; University of California San Francisco;
   University of California System; University of California San Francisco;
   UCSF Medical Center; UCSF Helen Diller Family Comprehensive Cancer
   Center
RP Cha, S (corresponding author), UCSF Dept Radiol, 350 Parnassus Ave,Ste 307,Room 307H,Box 0336, San Francisco, CA 94117 USA.
EM
OI Barajas jr, Ramon/0000-0001-5093-4168
FU National Center for Research Resources, a component of the National
   Institutes of Health (NIH) [NS045013, TL1 RR024129-01]; NIH Roadmap for
   Medical Research; National Cancer Institute; NIH [P50 CA097257];
   Gabrielle's Angel Foundation for Cancer Research; American Cancer
   Society
FX This work was supported by NS045013 (S C) and TL1 RR024129-01 (R.F.B.)
   from the, National Center for Research Resources, a component of the
   National Institutes of Health (NIH) and NIH Roadmap for Medical
   Research; by a National Cancer Institute Research Career Award, NIH
   Brain Tumor Specialized Programs of Research Excellence (SPORE) grant
   P50 CA097257, and by grants from Gabrielle's Angel Foundation for Cancer
   Research and the American Cancer Society (J.L.R.).
CR Abrey LE, 2005, J CLIN ONCOL, V23, P5034, DOI 10.1200/JCO.2005.13.524
   Abrey LE, 1998, J CLIN ONCOL, V16, P859, DOI 10.1200/JCO.1998.16.3.859
   Blay JY, 1998, J CLIN ONCOL, V16, P864, DOI 10.1200/JCO.1998.16.3.864
   BURGER PC, 1986, AM J SURG PATHOL, V10, P611, DOI 10.1097/00000478-198609000-00003
   *CENTR BRAIN TUM R, 2005, STAT REP PRIM BRAIN
   Chenevert TL, 2000, JNCI-J NATL CANCER I, V92, P2029, DOI 10.1093/jnci/92.24.2029
   Ducreux D, 2003, NEURORADIOLOGY, V45, P865, DOI 10.1007/s00234-003-1107-9
   Gavrilovic IT, 2006, J CLIN ONCOL, V24, P4570, DOI 10.1200/JCO.2006.06.6910
   GLASS J, 1994, J NEUROSURG, V81, P188, DOI 10.3171/jns.1994.81.2.0188
   Guo AC, 2002, RADIOLOGY, V224, P177, DOI 10.1148/radiol.2241010637
   Hamstra DA, 2005, P NATL ACAD SCI USA, V102, P16759, DOI 10.1073/pnas.0508347102
   Hochberg FH, 2007, NAT CLIN PRACT NEURO, V3, P24, DOI 10.1038/ncpneuro0395
   Kitis O, 2005, EUR J RADIOL, V55, P393, DOI 10.1016/j.ejrad.2005.02.004
   LEBIHAN D, 1986, RADIOLOGY, V161, P401, DOI 10.1148/radiology.161.2.3763909
   LEBIHAN D, 1992, AM J ROENTGENOL, V159, P591, DOI 10.2214/ajr.159.3.1503032
   Mardor Y, 2003, J CLIN ONCOL, V21, P1094, DOI 10.1200/JCO.2003.05.069
   Matsumoto Y, 2007, INT J HEMATOL, V85, P333, DOI 10.1532/IJH97.06205
   McAllister LD, 2000, NEUROSURGERY, V46, P51, DOI 10.1093/neurosurgery/46.1.51
   Moffat BA, 2005, P NATL ACAD SCI USA, V102, P5524, DOI 10.1073/pnas.0501532102
   MURRAY K, 1986, J NEUROSURG, V65, P600, DOI 10.3171/jns.1986.65.5.0600
   Olson JE, 2002, CANCER, V95, P1504, DOI 10.1002/cncr.10851
   ONEILL BP, 1987, NEUROLOGY, V37, P1160, DOI 10.1212/WNL.37.7.1160
   ONEILL BP, 1989, MAYO CLIN PROC, V64, P1005, DOI 10.1016/S0025-6196(12)61228-2
   Reiche W, 2007, CLIN NEUROL NEUROSUR, V109, P92, DOI 10.1016/j.clineuro.2006.03.007
   Rowley HA, 1999, NEUROIMAG CLIN N AM, V9, P343
   Roy S, 2008, J CLIN ONCOL, V26, P96, DOI 10.1200/JCO.2007.12.1053
   Rubenstein JL, 2007, J CLIN ONCOL, V25, P1350, DOI 10.1200/JCO.2006.09.7311
   Rubenstein JL, 2009, BLOOD, V113, P266, DOI 10.1182/blood-2008-04-152835
   Rubenstein JL, 2006, BLOOD, V107, P3716, DOI 10.1182/blood-2005-03-0897
   Ryken TC, 2001, INT J RADIAT ONCOL, V51, P1152, DOI 10.1016/S0360-3016(01)01756-4
   Soussain C, 2008, J CLIN ONCOL, V26, P2512, DOI 10.1200/JCO.2007.13.5533
   Stadnik TW, 2001, AM J NEURORADIOL, V22, P969
   Surawicz T S, 1999, Neuro Oncol, V1, P14, DOI 10.1093/neuonc/1.1.14
   Toh CH, 2008, AM J NEURORADIOL, V29, P471, DOI 10.3174/ajnr.A0872
NR 34
TC 155
Z9 173
U1 0
U2 6
PU AMER SOC NEURORADIOLOGY
PI DENVILLE
PA PO BOX 3000, DENVILLE, NJ 07834-9349 USA
SN 0195-6108
EI 1936-959X
J9 AM J NEURORADIOL
JI Am. J. Neuroradiol.
PD JAN
PY 2010
VL 31
IS 1
BP 60
EP 66
DI 10.3174/ajnr.A1750
PG 7
WC Clinical Neurology; Neuroimaging; Radiology, Nuclear Medicine & Medical
   Imaging
WE Science Citation Index Expanded (SCI-EXPANDED)
SC Neurosciences & Neurology; Radiology, Nuclear Medicine & Medical Imaging
GA 548EY
UT WOS:000273943700011
PM 19729544
OA Green Accepted, hybrid, Green Published
DA 2024-06-03
ER

PT J
AU Enting, RH
   Demopoulos, A
   DeAngelis, LM
   Abrey, LE
AF Enting, RH
   Demopoulos, A
   DeAngelis, LM
   Abrey, LE
TI Salvage therapy for primary CNS lymphoma with a combination of rituximab
   and temozolomide
SO NEUROLOGY
LA English
DT Article
ID NERVOUS-SYSTEM LYMPHOMA; CHEMOTHERAPY
AB The authors evaluated the efficacy of a combination of rituximab and temozolomide for recurrent or refractory primary CNS lymphoma (PCNSL). Fifteen patients with a median age of 69 years had a 53% objective response rate with acceptable toxicity. Median overall survival is 14 months and median progression free survival of responding patients is 7.7 months. This combination merits further study and provides a reasonable therapeutic alternative for older patients with progressive PCNSL.
C1 Mem Sloan Kettering Canc Ctr, Dept Neurol, New York, NY 10021 USA.
   Univ Groningen Hosp, Dept Neurol, Groningen, Netherlands.
C3 Memorial Sloan Kettering Cancer Center; University of Groningen
RP Mem Sloan Kettering Canc Ctr, Dept Neurol, 1275 York Ave, New York, NY 10021 USA.
EM
OI DeAngelis, Lisa/0000-0002-6290-7818
CR Abrey LE, 2000, J CLIN ONCOL, V18, P3144, DOI 10.1200/JCO.2000.18.17.3144
   Arellano-Rodrigo E, 2003, EUR J HAEMATOL, V70, P219, DOI 10.1034/j.1600-0609.2003.00045.x
   CIORDIA R, 2000, 36 ANN M AM SOC CLIN
   FIGUEROA F, 2000, 36 ANN M AM SOC CLIN
   Guha-Thakurta N, 1999, J NEURO-ONCOL, V43, P259, DOI 10.1023/A:1006210703827
   Herrlinger U, 2000, NEUROLOGY, V54, P1707, DOI 10.1212/WNL.54.8.1707
   O'Brien SM, 2001, J CLIN ONCOL, V19, P2165, DOI 10.1200/JCO.2001.19.8.2165
   Pels H, 2003, ONKOLOGIE, V26, P351, DOI 10.1159/000072095
   RENI M, 2003, P AN M AM SOC CLIN, V22, P571
   Soussain C, 2001, J CLIN ONCOL, V19, P742, DOI 10.1200/JCO.2001.19.3.742
NR 10
TC 155
Z9 161
U1 0
U2 1
PU LIPPINCOTT WILLIAMS & WILKINS
PI PHILADELPHIA
PA TWO COMMERCE SQ, 2001 MARKET ST, PHILADELPHIA, PA 19103 USA
SN 0028-3878
EI 1526-632X
J9 NEUROLOGY
JI Neurology
PD SEP 14
PY 2004
VL 63
IS 5
BP 901
EP 903
DI 10.1212/01.WNL.0000137050.43114.42
PG 3
WC Clinical Neurology
WE Science Citation Index Expanded (SCI-EXPANDED)
SC Neurosciences & Neurology
GA 853ME
UT WOS:000223830900027
PM 15365145
DA 2024-06-03
ER

PT J
AU Nakamura, T
   Tateishi, K
   Niwa, T
   Matsushita, Y
   Tamura, K
   Kinoshita, M
   Tanaka, K
   Fukushima, S
   Takami, H
   Arita, H
   Kubo, A
   Shuto, T
   Ohno, M
   Miyakita, Y
   Kocialkowski, S
   Sasayama, T
   Hashimoto, N
   Maehara, T
   Shibui, S
   Ushijima, T
   Kawahara, N
   Narita, Y
   Ichimura, K
AF Nakamura, T.
   Tateishi, K.
   Niwa, T.
   Matsushita, Y.
   Tamura, K.
   Kinoshita, M.
   Tanaka, K.
   Fukushima, S.
   Takami, H.
   Arita, H.
   Kubo, A.
   Shuto, T.
   Ohno, M.
   Miyakita, Y.
   Kocialkowski, S.
   Sasayama, T.
   Hashimoto, N.
   Maehara, T.
   Shibui, S.
   Ushijima, T.
   Kawahara, N.
   Narita, Y.
   Ichimura, K.
TI Recurrent mutations of <i>CD79B</i> and <i>MYD88</i> are the hallmark of
   primary central nervous system lymphomas
SO NEUROPATHOLOGY AND APPLIED NEUROBIOLOGY
LA English
DT Article
DE B-cell receptor signalling; CD79B; MYD88; NF-B; PCNSL
ID B-CELL LYMPHOMA; ONCOLOGY-GROUP; INACTIVATION; GENOME
AB AimsPrimary central nervous system lymphoma (PCNSL) manifest aggressive clinical behaviour and have poor prognosis. Although constitutive activation of the nuclear factor-B (NF-B) pathway has been documented, knowledge about the genetic alterations leading to the impairment of the NF-B pathway in PCNSLs is still limited. This study was aimed to unravel the underlying genetic profiles of PCNSL.
   MethodsWe conducted the systematic sequencing of 21 genes relevant to the NF-B signalling network for 71 PCNSLs as well as the pyrosequencing of CD79B and MYD88 mutation hotspots in a further 35 PCNSLs and 46 glioblastomas (GBMs) for validation.
   ResultsThe results showed that 68 out of 71 PCNSLs had mutations in the NF-B gene network, most commonly affecting CD79B (83%), MYD88 (76%), TBL1XR1 (23%), PRDM1 (20%) and CREBBP1 (20%). These mutations, particularly CD79B and MYD88, frequently coincided within each tumour in various combinations, simultaneously affecting diverse pathways within the network. No GBMs had hotspot mutation of CD79B Y196 and MYD88 L265.
   ConclusionsThe prevalence of CD79B and MYD88 mutations in PCNSLs was considerably higher than reported in systemic diffuse large B-cell lymphomas. This observation could reflect the paucity of antigen stimuli from the immune system in the central nervous system (CNS) and the necessity to substitute them by the constitutive activation of CD79B and MYD88 that would initiate the signalling cascades. These hotspot mutations may serve as a genetic hallmark for PCNSL serving as a genetic marker for diagnose and potential targets for molecular therapy.
C1 [Nakamura, T.; Tateishi, K.; Fukushima, S.; Takami, H.; Ichimura, K.] Tokyo Med & Dent Univ, Div Brain Tumor Translat Res, Natl Canc Ctr, Res Inst, Tokyo, Japan.
   [Niwa, T.; Ushijima, T.] Tokyo Med & Dent Univ, Natl Canc Ctr, Res Inst, Div Epigen, Tokyo, Japan.
   [Arita, H.; Ohno, M.; Miyakita, Y.; Shibui, S.; Narita, Y.] Tokyo Med & Dent Univ, Natl Canc Ctr, Dept Neurosurg & Neurooncol, Tokyo, Japan.
   [Tamura, K.; Maehara, T.] Tokyo Med & Dent Univ, Dept Neurosurg, Tokyo, Japan.
   [Nakamura, T.; Tateishi, K.; Kawahara, N.] Yokohama City Univ, Grad Sch Med, Dept Neurosurg, Yokohama, Japan.
   [Shuto, T.] Yokohama Rosai Hosp, Dept Neurosurg, Yokohama, Kanagawa, Japan.
   [Kinoshita, M.; Arita, H.; Hashimoto, N.] Osaka Univ, Grad Sch Med, Dept Neurosurg, Osaka, Japan.
   [Tanaka, K.] Kobe Univ, Grad Sch Med, Dept Neurosurg, Kobe, Hyogo 657, Japan.
   [Kubo, A.] Yokosuka Kyosai Hosp, Dept Neurosurg, Yokosuka, Kanagawa, Japan.
   [Kocialkowski, S.] Univ Cambridge, Dept Pathol, Tennis Court Rd, Cambridge CB2 1QP, England.
C3 Tokyo Medical & Dental University (TMDU); National Cancer Center -
   Japan; Tokyo Medical & Dental University (TMDU); National Cancer Center
   - Japan; National Cancer Center - Japan; Tokyo Medical & Dental
   University (TMDU); Tokyo Medical & Dental University (TMDU); Yokohama
   City University; Yokohama Rosai Hospital; Osaka University; Kobe
   University; University of Cambridge
RP Ichimura, K (corresponding author), Natl Canc Ctr, Res Inst, Div Brain Tumor Translat Res, Chuo Ku, 5-1-1 Tsukiji, Tokyo 1040045, Japan.
EM
RI Arita, Hideyuki/AAH-2301-2019; Takami, Hirokazu/H-6903-2013; Ushijima,
   Toshikazu/AAP-5742-2021
OI Arita, Hideyuki/0000-0002-1126-8155; Ushijima,
   Toshikazu/0000-0003-3405-7817; Kinoshita, Manabu/0000-0001-7923-6902;
   Sasayama, Takashi/0000-0002-5593-9045
FU National Cancer Center Research and Development Fund [26-A-9];
   Grants-in-Aid for Scientific Research [25462256] Funding Source: KAKEN
FX We are grateful to Dr. Hiromi Sakamoto, Ms. Mineko Ushiama and Dr. Kai
   Yamasaki for their generous technical help with this project. This work
   was supported by the National Cancer Center Research and Development
   Fund (26-A-9 (K. I.)).
CR Abrey LE, 2006, J CLIN ONCOL, V24, P5711, DOI 10.1200/JCO.2006.08.2941
   [Anonymous], 2008, WHO Classification of Tumors of Haematopoietic and Lymphoid Tissues
   [Anonymous], 2014, NEUROLOGIA MEDICOCHI
   Brennan CW, 2013, CELL, V155, P462, DOI 10.1016/j.cell.2013.09.034
   Bruno A, 2014, ONCOTARGET, V5, P5065, DOI 10.18632/oncotarget.2080
   Courts C, 2008, J NEUROPATH EXP NEUR, V67, P720, DOI 10.1097/NEN.0b013e31817dd02d
   Davis RE, 2010, NATURE, V463, P88, DOI 10.1038/nature08638
   Ferreri AJM, 2009, LANCET, V374, P1512, DOI 10.1016/S0140-6736(09)61416-1
   Gaspar L, 1997, INT J RADIAT ONCOL, V37, P745, DOI 10.1016/S0360-3016(96)00619-0
   Gonzalez-Aguilar A, 2012, CLIN CANCER RES, V18, P5203, DOI 10.1158/1078-0432.CCR-12-0845
   Kato M, 2009, NATURE, V459, P712, DOI 10.1038/nature07969
   Kraan W, 2013, BLOOD CANCER J, V3, DOI 10.1038/bcj.2013.28
   Lenz G, 2008, SCIENCE, V319, P1676, DOI 10.1126/science.1153629
   Lohr JG, 2012, P NATL ACAD SCI USA, V109, P3879, DOI 10.1073/pnas.1121343109
   Michallet AS, 2009, BLOOD REV, V23, P11, DOI 10.1016/j.blre.2008.05.002
   Montesinos-Rongen M, 2012, ACTA NEUROPATHOL, V124, P905, DOI 10.1007/s00401-012-1064-7
   Montesinos-Rongen M, 2010, ACTA NEUROPATHOL, V120, P529, DOI 10.1007/s00401-010-0709-7
   Morin RD, 2013, BLOOD, V122, P1256, DOI 10.1182/blood-2013-02-483727
   Muldoon LL, 2013, J CEREBR BLOOD F MET, V33, P13, DOI 10.1038/jcbfm.2012.153
   Ngo VN, 2011, NATURE, V470, P115, DOI 10.1038/nature09671
   Odegard VH, 2006, NAT REV IMMUNOL, V6, P573, DOI 10.1038/nri1896
   OKEN MM, 1982, AM J CLIN ONCOL-CANC, V5, P649, DOI 10.1097/00000421-198212000-00014
   Pasqualucci L, 2011, NAT GENET, V43, P830, DOI 10.1038/ng.892
   Poulain S, 2014, BRIT J HAEMATOL, V167, P506, DOI 10.1111/bjh.13078
   Takami H, 2015, BRAIN PATHOL, V25, P256, DOI 10.1111/bpa.12173
   Vater I, 2015, LEUKEMIA, V29, P677, DOI 10.1038/leu.2014.264
   Zhang J, 2013, P NATL ACAD SCI USA, V110, P1398, DOI 10.1073/pnas.1205299110
   Zhou HJ, 2007, BLOOD, V110, P2121, DOI 10.1182/blood-2007-02-073080
NR 28
TC 152
Z9 157
U1 0
U2 13
PU WILEY-BLACKWELL
PI HOBOKEN
PA 111 RIVER ST, HOBOKEN 07030-5774, NJ USA
SN 0305-1846
EI 1365-2990
J9 NEUROPATH APPL NEURO
JI Neuropathol. Appl. Neurobiol.
PD APR
PY 2016
VL 42
IS 3
BP 279
EP 290
DI 10.1111/nan.12259
PG 12
WC Clinical Neurology; Neurosciences; Pathology
WE Science Citation Index Expanded (SCI-EXPANDED)
SC Neurosciences & Neurology; Pathology
GA DI4TP
UT WOS:000373492500006
PM 26111727
DA 2024-06-03
ER

PT J
AU Smith, JR
   Braziel, RM
   Paoletti, S
   Lipp, M
   Uguccioni, M
   Rosenbaum, JT
AF Smith, JR
   Braziel, RM
   Paoletti, S
   Lipp, M
   Uguccioni, M
   Rosenbaum, JT
TI Expression of B-cell-attracting chemokine 1 (CXCL13) by malignant
   lymphocytes and vascular endothelium in primary central nervous system
   lymphoma
SO BLOOD
LA English
DT Article; Proceedings Paper
CT 42nd Annual Meeting of the American-Society-of-Hematology
CY DEC 01-05, 2000
CL SAN FRANCISCO, CA
SP Amer Soc Hematol
ID GERMINAL CENTER; T-LYMPHOCYTES; EPIDEMIOLOGY; ACTIVATION; RECEPTORS;
   FOLLICLES; MIGRATION; TISSUE
AB Primary central nervous system lymphoma (PCNSL) is a rare but often rapidly fatal form of non-Hodgkin B-cell lymphoma that arises within the central nervous system (CNS) and has a low propensity to metastasize. We performed immunohistochemistry on formalin-fixed, paraffin-embedded brain biopsy specimens from 24 patients with PCNSL to investigate the expression of B cell-attracting chemokine 1 (BCA-1, CXCL13), a lymphoid chemokine involved in B-cell compartmental homing within secondary lymphoid organs and recently implicated in the pathogenesis of inflammatory and malignant lymphocyte-mediated diseases. Whereas BCA-1 was not detected in normal human brain, all 24 brain biopsy specimens containing PCNSL were positive for BCA-1. Double immunostaining on selected specimens localized BCA-1 to malignant B lymphocytes and vascular endothelium. In contrast, 2 chemokines implicated particularly in T-cell movement, secondary lymphoid tissue chemokine (SLC, CCL21) and Epstein-Barr virus-induced molecule 1 ligand chemokine (ELC, CCL19), were expressed only by occasional stromal cells in 2 and 4 of the 24 specimens, respectively. Tumor cells stained positively for CXCR5, the primary receptor for BCA-1. In situ hybridization verified the expression of BCA-1 mRNA by malignant B cells, but not vascular endothelium, within the tumor mass, suggesting that vascular endothelial BCA-1 expression may be consequent to transcytosis. In PCNSL, expression of BCA-1 by malignant lymphocytes and vascular endothelium may influence tumor development and localization to CNS. (C) 2003 by The American Society of Hematology.
C1 Oregon Hlth & Sci Univ, Casey Eye Inst, Portland, OR 97201 USA.
   Oregon Hlth & Sci Univ, Dept Pathol, Portland, OR 97201 USA.
   Max Delbruck Ctr Mol Med, Berlin, Germany.
   Inst Res Biomed, Bellinzona, Switzerland.
C3 Oregon Health & Science University; Oregon Health & Science University;
   Helmholtz Association; Max Delbruck Center for Molecular Medicine;
   Universita della Svizzera Italiana
RP Oregon Hlth & Sci Univ, Casey Eye Inst, 3375 Sw Terwilliger Blvd, Portland, OR 97201 USA.
EM
RI Paoletti, Samantha/AAG-8152-2021; Lipp, Martin/G-2235-2010; Smith,
   Justine/Y-9044-2019
OI Paoletti, Samantha/0000-0001-7982-6478; Smith,
   Justine/0000-0002-4756-5493; Lipp, Martin/0000-0002-0087-2672;
   Uguccioni, Mariagrazia/0000-0002-9570-7011
FU NEI NIH HHS [EYO6484] Funding Source: Medline
CR Alderson L, 1996, J NEUROL NEUROSUR PS, V60, P102, DOI 10.1136/jnnp.60.1.102
   Alizadeh AA, 2000, NATURE, V403, P503, DOI 10.1038/35000501
   Amft N, 2001, ARTHRITIS RHEUM-US, V44, P2633, DOI 10.1002/1529-0131(200111)44:11<2633::AID-ART443>3.0.CO;2-9
   Ansel KM, 2000, NATURE, V406, P309, DOI 10.1038/35018581
   Ansell SM, 2001, J CLIN ONCOL, V19, P720, DOI 10.1200/JCO.2001.19.3.720
   Baekkevold ES, 2001, J EXP MED, V193, P1105, DOI 10.1084/jem.193.9.1105
   Browning JL, 1997, J IMMUNOL, V159, P3288
   Cote TR, 1996, J NATL CANCER I, V88, P675, DOI 10.1093/jnci/88.10.675
   Cyster JG, 1999, SCIENCE, V286, P2098, DOI 10.1126/science.286.5447.2098
   Drillenburg P, 2000, BLOOD, V95, P1900, DOI 10.1182/blood.V95.6.1900
   FORSTER R, 1994, BLOOD, V84, P830
   GRANT JW, 1992, BRAIN PATHOL, V2, P97, DOI 10.1111/j.1750-3639.1992.tb00677.x
   HOCHBERG FH, 1988, J NEUROSURG, V68, P835, DOI 10.3171/jns.1988.68.6.0835
   Hostager BS, 2002, J IMMUNOL, V168, P3318, DOI 10.4049/jimmunol.168.7.3318
   Ishikawa S, 2001, J EXP MED, V193, P1393, DOI 10.1084/jem.193.12.1393
   JAFFE ES, 2001, TUMOURS HAEMOPOIETIC
   Koulis A, 1997, AM J PATHOL, V151, P1353
   Lossos IS, 2000, BLOOD, V95, P1797, DOI 10.1182/blood.V95.5.1797.005k30_1797_1803
   Luther SA, 2000, IMMUNITY, V12, P471, DOI 10.1016/S1074-7613(00)80199-5
   Mazzucchelli L, 1999, J CLIN INVEST, V104, pR49, DOI 10.1172/JCI7830
   Michel CC, 1996, CARDIOVASC RES, V32, P644, DOI 10.1016/S0008-6363(96)00064-8
   Müller A, 2001, NATURE, V410, P50, DOI 10.1038/35065016
   Müller G, 2001, BIOL CHEM, V382, P1387, DOI 10.1515/BC.2001.171
   MURRAY K, 1986, J NEUROSURG, V65, P600, DOI 10.3171/jns.1986.65.5.0600
   NAKHLEH RE, 1989, ARCH PATHOL LAB MED, V113, P1050
   Paulus W, 1999, J NEURO-ONCOL, V43, P203, DOI 10.1023/A:1006242116122
   Pileri SA, 2001, ANN ONCOL, V12, P1445, DOI 10.1023/A:1012559725243
   Robertson MJ, 2000, CELL IMMUNOL, V199, P8, DOI 10.1006/cimm.1999.1601
   Salomonsson S, 2002, SCAND J IMMUNOL, V55, P336, DOI 10.1046/j.1365-3083.2002.01058.x
   Schabet M, 1999, J NEURO-ONCOL, V43, P199, DOI 10.1023/A:1006290032052
   Schaerli P, 2001, J IMMUNOL, V167, P6082, DOI 10.4049/jimmunol.167.11.6082
   SCHAERLI P, 2000, J EXP MED, V164, P3955
   Shi KR, 2001, J IMMUNOL, V166, P650, DOI 10.4049/jimmunol.166.1.650
   Slowik F, 1975, ACTA NEUROPATH BER S, V6, P95, DOI DOI 10.1007/978-3-662-08456-4_16
   Stein JV, 2000, J EXP MED, V191, P61, DOI 10.1084/jem.191.1.61
   Takemura S, 2001, J IMMUNOL, V167, P1072, DOI 10.4049/jimmunol.167.2.1072
   Xanthou G, 2001, ARTHRITIS RHEUM-US, V44, P408
NR 37
TC 152
Z9 166
U1 0
U2 2
PU AMER SOC HEMATOLOGY
PI WASHINGTON
PA 2021 L ST NW, SUITE 900, WASHINGTON, DC 20036 USA
SN 0006-4971
EI 1528-0020
J9 BLOOD
JI Blood
PD FEB 1
PY 2003
VL 101
IS 3
BP 815
EP 821
DI 10.1182/blood-2002-05-1576
PG 7
WC Hematology
WE Science Citation Index Expanded (SCI-EXPANDED); Conference Proceedings Citation Index - Science (CPCI-S)
SC Hematology
GA 640GG
UT WOS:000180679700004
PM 12393412
OA Bronze
DA 2024-06-03
ER

PT J
AU Sagoo, MS
   Mehta, H
   Swampillai, AJ
   Cohen, VML
   Amin, SZ
   Plowman, PN
   Lightman, S
AF Sagoo, Mandeep S.
   Mehta, Hemal
   Swampillai, Andrew J.
   Cohen, Victoria M. L.
   Amin, Sepideh Z.
   Plowman, P. Nicholas
   Lightman, Sue
TI Primary intraocular lymphoma
SO SURVEY OF OPHTHALMOLOGY
LA English
DT Review
DE lymphoma; ocular; masquerade; vitreoretinal
ID CENTRAL-NERVOUS-SYSTEM; POLYMERASE-CHAIN-REACTION; B-CELL LYMPHOMA;
   NON-HODGKINS-LYMPHOMA; PRIMARY CNS LYMPHOMA;
   BONE-MARROW-TRANSPLANTATION; PRIMARY OCULAR LYMPHOMA; HIGH-DOSE
   METHOTREXATE; RECURRENT PRIMARY CNS; CLINICAL-FEATURES
AB Primary intraocular lymphoma (PIOL) is an ocular malignancy that is a subset of primary central system lymphoma (PCNSL). Approximately one-third of PIOL patients will have concurrent PCNSL at presentation, and 42-92% will develop PCNSL within a mean of 8-29 months. Although rare, the incidence has been rising in both immunocompromised and immunocompetent populations. The majority of PIOL is diffuse large B-cell lymphoma, though rare T-cell variants are described. Recently, PIOL has been classified by main site of involvement in the eye, with vitreoretinal lymphoma as the most common type of ocular lymphoma related to PCNSL. Diagnosis remains challenging for ophthalmologists and pathologists. PIOL can masquerade as noninfectious or infectious uveitis, white dot syndromes, or occasionally as other neoplasms such as metastatic cancers. Laboratory diagnosis by cytology has been much aided by the use of immunocytochemistry, flow cytometry, biochemical finding of interleukin changes (IL10:IL6 ratio > 1), and cellular microdissection with polymerase chain reaction amplification for clonality. Use of several tests improves the diagnostic yield. Approaches to treatment have centered on systemic methotrexate-based chemotherapy, often with cytarabine (Ara-C) and radiotherapy. Use of intravitreal chemotherapy with methotrexate (0.4 mg/0.1 mL) is promising in controlling ocular disease, and intravitreal rituximab (anti-CD20 monoclonal antibody) has also been tried. Despite these advances, prognosis remains poor. (C) 2014 The Authors. Published by Elsevier Inc. All rights reserved.
C1 [Sagoo, Mandeep S.; Amin, Sepideh Z.; Lightman, Sue] UCL Inst Ophthalmol, London, England.
   [Sagoo, Mandeep S.; Mehta, Hemal; Cohen, Victoria M. L.; Lightman, Sue] Moorfields Eye Hosp, London EC1V 2PD, England.
   [Sagoo, Mandeep S.; Swampillai, Andrew J.; Cohen, Victoria M. L.] St Bartholomews Hosp, Dept Ophthalmol, London, England.
   [Plowman, P. Nicholas] St Bartholomews Hosp, Dept Oncol, London, England.
C3 University of London; University College London; University of London;
   University College London; Moorfields Eye Hospital NHS Foundation Trust;
   University of London; Queen Mary University London; University of
   London; Queen Mary University London
RP Sagoo, MS (corresponding author), Moorfields Eye Hosp, UCL Inst Ophthalmol, City Rd, London EC1V 2PD, England.
EM
RI Mehta, Hemal/HSB-3343-2023
OI Mehta, Hemal/0000-0002-0606-9004; Sagoo, Mandeep/0000-0003-1530-3824;
   Cohen, Victoria/0000-0002-7793-160X
CR AbdelReheim FA, 1996, ARCH PATHOL LAB MED, V120, P357
   Akpek EK, 1999, OPHTHALMOLOGY, V106, P2291
   Akpek EK, 1999, OPHTHALMOLOGY, V106, P1805, DOI 10.1016/S0161-6420(99)90341-X
   Akpek EK, 2000, ARCH OPHTHALMOL-CHIC, V118, P731
   Androudi S, 2009, OCUL IMMUNOL INFLAMM, V17, P73, DOI 10.1080/09273940802596542
   Augsburger J J, 1989, Trans Pa Acad Ophthalmol Otolaryngol, V41, P796
   Baehring JM, 2005, CANCER-AM CANCER SOC, V104, P591, DOI 10.1002/cncr.21191
   Balansard B, 2005, BRIT J OPHTHALMOL, V89, P96, DOI 10.1136/bjo.2004.042226
   BANCHEREAU J, 1994, STEM CELLS, V12, P278, DOI 10.1002/stem.5530120304
   Bardenstein, 1998, Cancer Control, V5, P317
   Barr C C, 1975, Surv Ophthalmol, V19, P224
   BARTON K, 1994, EYE, V8, P288, DOI 10.1038/eye.1994.59
   Batchelor TT, 2003, CLIN CANCER RES, V9, P711
   BAUMANN MA, 1986, CANCER-AM CANCER SOC, V57, P1273, DOI 10.1002/1097-0142(19860401)57:7<1273::AID-CNCR2820570702>3.0.CO;2-C
   Berenbom A, 2007, EYE, V21, P1198, DOI 10.1038/sj.eye.6702437
   Bessell EM, 2007, EUR J CANCER, V43, P1141, DOI 10.1016/j.ejca.2006.12.011
   Bhat PV, 2009, AM J OPHTHALMOL, V148, P350, DOI 10.1016/j.ajo.2009.04.005
   BLUMENKRANZ MS, 1992, RETINA-J RET VIT DIS, V12, pS64, DOI 10.1097/00006982-199212031-00014
   BROWN SM, 1994, SURV OPHTHALMOL, V39, P133, DOI 10.1016/0039-6257(94)90158-9
   Browning DJ, 2007, EYE, V21, P880, DOI 10.1038/sj.eye.6702756
   BUETTNER H, 1993, MAYO CLIN PROC, V68, P1011, DOI 10.1016/S0025-6196(12)62276-9
   Buggage RR, 1999, ARCH OPHTHALMOL-CHIC, V117, P1239
   Buggage RR, 1999, INVEST OPHTH VIS SCI, V40, P2462
   Buggage RR, 2001, CURR OPIN ONCOL, V13, P137, DOI 10.1097/00001622-200105000-00001
   Calfa CJ, 2007, AM J OPHTHALMOL, V144, P326, DOI 10.1016/j.ajo.2007.03.031
   Cassoux N, 2000, OCUL IMMUNOL INFLAMM, V8, P243, DOI 10.1076/ocii.8.4.243.6463
   Cassoux N, 2007, INVEST OPHTH VIS SCI, V48, P3253, DOI 10.1167/iovs.06-0031
   Caya J G, 1985, Diagn Cytopathol, V1, P267, DOI 10.1002/dc.2840010404
   CBTRUS, 1998, STAT REP PRIM BRAIN
   CHAN CC, 1995, AM J OPHTHALMOL, V120, P671, DOI 10.1016/S0002-9394(14)72217-2
   Chan CC, 2003, OPHTHALMOLOGY, V110, P421, DOI 10.1016/S0161-6420(02)01737-2
   Chan CC, 2003, CLIN LYMPHOMA, V4, P30, DOI 10.1016/S1526-9655(11)70005-7
   Chan CC, 1998, DIAGN MOL PATHOL, V7, P63, DOI 10.1097/00019606-199802000-00011
   Chan Chi-Chao, 2004, Cancer Control, V11, P285
   Chan Chi-Chao, 2003, Trans Am Ophthalmol Soc, V101, P275
   Chan CC, 2011, ONCOLOGIST, V16, P1589, DOI 10.1634/theoncologist.2011-0210
   Chan Chi-Chao, 2002, Curr Opin Ophthalmol, V13, P411, DOI 10.1097/00055735-200212000-00012
   Chan CC, 2009, OCUL IMMUNOL INFLAMM, V17, P375, DOI 10.3109/09273940903434804
   Chan SM, 2000, OPHTHALMOLOGY, V107, P1479, DOI 10.1016/S0161-6420(00)00172-X
   Char Devron H, 2005, Ophthalmol Clin North Am, V18, P177, DOI 10.1016/j.ohc.2004.08.007
   CHAR DH, 1988, BRIT J OPHTHALMOL, V72, P905, DOI 10.1136/bjo.72.12.905
   CHAR DH, 1988, OPHTHALMOLOGY, V95, P625
   Chatzistefanou K, 1998, OPHTHALMOLOGY, V105, P347, DOI 10.1016/S0161-6420(98)93523-0
   Chawla R, 2005, EUR J OPHTHALMOL, V15, P153, DOI 10.1177/112067210501500127
   Choi John Y, 2006, Semin Ophthalmol, V21, P125, DOI 10.1080/08820530500350498
   Cockerham GC, 2000, OPHTHALMOLOGY, V107, P151, DOI 10.1016/S0161-6420(99)00025-1
   COOPER EL, 1951, AM J OPHTHALMOL, V34, P1153, DOI 10.1016/0002-9394(51)90691-5
   CORRIVEAU C, 1986, CAN J OPHTHALMOL, V21, P144
   COSKUNCAN NM, 1994, ARCH OPHTHALMOL-CHIC, V112, P372, DOI 10.1001/archopht.1994.01090150102031
   Coupland SE, 2008, CLIN EXP OPHTHALMOL, V36, P564, DOI 10.1111/j.1442-9071.2008.01843.x
   Coupland SE, 2005, INVEST OPHTH VIS SCI, V46, P3957, DOI 10.1167/iovs.05-0318
   Coupland SE, 2005, INVEST OPHTH VIS SCI, V46, P3507, DOI 10.1167/iovs.05-0401
   Coupland SE, 2005, GRAEF ARCH CLIN EXP, V243, P847, DOI 10.1007/s00417-005-1166-1
   Coupland SE, 2005, GRAEF ARCH CLIN EXP, V243, P482, DOI 10.1007/s00417-004-1050-4
   Coupland SE, 2005, GRAEF ARCH CLIN EXP, V243, P189, DOI 10.1007/s00417-004-0890-2
   Coupland SE, 2004, GRAEF ARCH CLIN EXP, V242, P901, DOI 10.1007/s00417-004-0973-0
   Coupland SE, 2003, GRAEF ARCH CLIN EXP, V241, P860, DOI 10.1007/s00417-003-0749-y
   Coupland SE, 2002, J PATHOL, V197, P333, DOI 10.1002/path.1130
   Coutinho AB, 2005, CAN J OPHTHALMOL, V40, P623, DOI 10.1016/S0008-4182(05)80058-4
   Cursiefen C, 2000, ARCH OPHTHALMOL-CHIC, V118, P1437
   Dark F, 1996, PSYCHOSOMATICS, V37, P396, DOI 10.1016/S0033-3182(96)71557-8
   Davis JL, 2005, AM J OPHTHALMOL, V140, P822, DOI 10.1016/j.ajo.2005.05.032
   Davis JL, 2004, OCUL IMMUNOL INFLAMM, V12, P7, DOI 10.1076/ocii.12.1.7.28072
   Davis JL, 1997, AM J OPHTHALMOL, V124, P362, DOI 10.1016/S0002-9394(14)70828-1
   DAVIS JL, 1992, OPHTHALMOLOGY, V99, P250
   de Smet MD, 1999, BRIT J OPHTHALMOL, V83, P448, DOI 10.1136/bjo.83.4.448
   Dean JM, 1996, RETINA-J RET VIT DIS, V16, P47, DOI 10.1097/00006982-199616010-00009
   DeAngelis L M, 2001, Curr Treat Options Oncol, V2, P309, DOI 10.1007/s11864-001-0024-6
   Derzko-Dzulynsky LA, 1998, AM J HEMATOL, V57, P228, DOI 10.1002/(SICI)1096-8652(199803)57:3<228::AID-AJH9>3.0.CO;2-X
   deSmet MD, 1996, AM J OPHTHALMOL, V121, P442, DOI 10.1016/S0002-9394(14)70444-1
   Dunleavy K, 2006, LEUKEMIA LYMPHOMA, V47, P1726, DOI 10.1080/10428190600674339
   EBY NL, 1988, CANCER-AM CANCER SOC, V62, P2461, DOI 10.1002/1097-0142(19881201)62:11<2461::AID-CNCR2820621135>3.0.CO;2-M
   El-Shabrawi Y, 1998, OPHTHALMOLOGY, V105, P1659, DOI 10.1016/S0161-6420(98)99035-2
   Eljarrat-Binstock E, 2007, CURR EYE RES, V32, P639, DOI 10.1080/02713680701528674
   Fahim DK, 2005, J NEURO-OPHTHALMOL, V25, P33, DOI 10.1097/00041327-200503000-00009
   Faia LJ, 2009, ARCH PATHOL LAB MED, V133, P1228, DOI 10.1043/1543-2165-133.8.1228
   Fardeau C, 2009, AM J OPHTHALMOL, V147, P886, DOI 10.1016/j.ajo.2008.12.025
   Farkas T, 2004, ACTA CYTOL, V48, P487, DOI 10.1159/000326409
   Feilmeier MR, 2008, LEUKEMIA LYMPHOMA, V49, P1
   Ferreri AJM, 2002, ANN ONCOL, V13, P531, DOI 10.1093/annonc/mdf080
   Finger PT, 2006, BRIT J OPHTHALMOL, V90, P690, DOI 10.1136/bjo.2005.087346
   Fishburne BC, 1997, ARCH OPHTHALMOL-CHIC, V115, P1152, DOI 10.1001/archopht.1997.01100160322009
   Forooghian Farzin, 2011, Ophthalmic Surg Lasers Imaging, V42 Online, pe97, DOI 10.3928/15428877-20110922-02
   FREDRICK DR, 1989, ARCH OPHTHALMOL-CHIC, V107, P395, DOI 10.1001/archopht.1989.01070010405034
   FREEMAN LN, 1987, OPHTHALMOLOGY, V94, P1631
   Frenkel S, 2008, BRIT J OPHTHALMOL, V92, P383, DOI 10.1136/bjo.2007.127928
   Gallagher MJ, 2008, EUR J OPHTHALMOL, V18, P567, DOI 10.1177/112067210801800411
   GASS JDM, 1984, RETINA-J RET VIT DIS, V4, P135, DOI 10.1097/00006982-198400430-00001
   Gerber S, 1998, RIV NEURORADIOL, V11, P161, DOI 10.1177/197140099801100203
   Gill MK, 2001, SURV OPHTHALMOL, V45, P463, DOI 10.1016/S0039-6257(01)00217-X
   GIVNER I, 1955, AM J OPHTHALMOL, V39, P29, DOI 10.1016/0002-9394(55)92650-7
   Goeminne JC, 1999, OPHTHALMOLOGICA, V213, P323, DOI 10.1159/000027447
   Goldberg DE, 2005, RETINA-J RET VIT DIS, V25, P633, DOI 10.1097/00006982-200507000-00015
   Gombos DS, 1999, OPHTHALMOL CLIN N AM, V12, P225
   Gonzales John A., 2007, International Ophthalmology, V27, P241, DOI 10.1007/s10792-007-9065-6
   Gooi P, 2008, CLIN OPHTHALMOL, V2, P969
   Gorochov G, 2003, INVEST OPHTH VIS SCI, V44, P5235, DOI 10.1167/iovs.03-0035
   Grimm SA, 2008, NEUROLOGY, V71, P1355, DOI 10.1212/01.wnl.0000327672.04729.8c
   Grimm SA, 2007, ANN ONCOL, V18, P1851, DOI 10.1093/annonc/mdm340
   Gunduz Kaan, 2006, Neurosurg Focus, V21, pE9
   Harris NL, 1999, J CLIN ONCOL, V17, P3835, DOI 10.1200/JCO.1999.17.12.3835
   HARRIS NL, 1994, BLOOD, V84, P1361
   Hedayatfar A, 2012, J OPHTHALMIC INFLAMM, V2, P49, DOI 10.1007/s12348-011-0045-7
   Herrlinger U, 1999, J NEURO-ONCOL, V43, P227, DOI 10.1023/A:1006250317940
   Higashide T, 2004, AM J OPHTHALMOL, V138, P306, DOI 10.1016/j.ajo.2004.03.003
   HOCHBERG FH, 1988, J NEUROSURG, V68, P835, DOI 10.3171/jns.1988.68.6.0835
   Hoffman PM, 2003, EYE, V17, P513, DOI 10.1038/sj.eye.6700378
   Hon C, 2005, ANN HEMATOL, V84, P203, DOI 10.1007/s00277-004-0975-3
   Hormigo A, 2004, BRIT J HAEMATOL, V126, P202, DOI 10.1111/j.1365-2141.2004.05028.x
   Hormigo A, 2003, CLIN LYMPHOMA, V4, P22, DOI 10.3816/CLM.2003.n.010
   HYKIN PG, 1990, J ROY SOC MED, V83, P51, DOI 10.1177/014107689008300122
   Intzedy L, 2008, EYE, V22, P289, DOI 10.1038/sj.eye.6702965
   Isobe K, 2006, LEUKEMIA LYMPHOMA, V47, P1800, DOI 10.1080/10428190600632881
   Itty S, 2009, RETINA-J RET VIT DIS, V29, P415, DOI 10.1097/IAE.0b013e318196b1f6
   JACK CR, 1988, RADIOLOGY, V167, P211, DOI 10.1148/radiology.167.1.3279454
   Jahnke K, 2006, GRAEF ARCH CLIN EXP, V244, P663, DOI 10.1007/s00417-005-0138-9
   Jahnke K, 2005, ANN ONCOL, V16, P1974, DOI 10.1093/annonc/mdi409
   Jahnke K, 2004, GRAEF ARCH CLIN EXP, V242, P771, DOI 10.1007/s00417-004-0947-2
   Jahnke K, 2009, J NEURO-ONCOL, V93, P213, DOI 10.1007/s11060-008-9761-8
   JOHNSON BL, 1992, OPHTHALMOLOGY, V99, P987
   Johnston RL, 2004, OPHTHALMOLOGY, V111, P522, DOI 10.1016/j.ophtha.2002.10.002
   Karikehalli S, 2004, ACTA CYTOL, V48, P207, DOI 10.1159/000326317
   Karma A, 2007, OPHTHALMOLOGY, V114, P1372, DOI 10.1016/j.ophtha.2006.11.009
   Katai N, 1997, GRAEF ARCH CLIN EXP, V235, P431, DOI 10.1007/BF00947062
   Khatri VP, 1998, CANCER IMMUNOL IMMUN, V46, P239, DOI 10.1007/s002620050483
   KHETERPAL S, 1993, AM J OPHTHALMOL, V116, P507, DOI 10.1016/S0002-9394(14)71416-3
   Kim Eunah, 2009, Korean J Ophthalmol, V23, P210, DOI 10.3341/kjo.2009.23.3.210
   Kim H, 2006, EXP EYE RES, V82, P760, DOI 10.1016/j.exer.2005.09.018
   Kitzmann AS, 2007, EYE, V21, P1524, DOI 10.1038/sj.eye.6702804
   KLINGELE TG, 1975, AM J OPHTHALMOL, V79, P39, DOI 10.1016/0002-9394(75)90453-5
   Küker W, 2002, J NEUROL, V249, P1713, DOI 10.1007/s00415-002-0919-6
   KUMAR SR, 1994, ARCH OPHTHALMOL-CHIC, V112, P954, DOI 10.1001/archopht.1994.01090190102028
   Levy-Clarke GA, 2005, HEMATOL ONCOL CLIN N, V19, P739, DOI 10.1016/j.hoc.2005.05.011
   Levy-Clarke GA, 2001, RETINA-J RET VIT DIS, V21, P281, DOI 10.1097/00006982-200106000-00023
   Levy-Clarke GA, 2008, SURV OPHTHALMOL, V53, P285, DOI 10.1016/j.survophthal.2008.02.004
   Liang F, 2011, GRAEF ARCH CLIN EXP, V249, P1425, DOI 10.1007/s00417-011-1669-x
   LJUNG BM, 1988, ACTA CYTOL, V32, P840
   Lobo A, 2003, CLIN EXP OPHTHALMOL, V31, P155, DOI 10.1046/j.1442-9071.2003.00624.x
   Lobo A, 2003, OPHTHALMOLOGY, V110, P595, DOI 10.1016/S0161-6420(02)01895-X
   Lobo A, 2007, J MOL DIAGN, V9, P113, DOI 10.2353/jmoldx.2007.050121
   Malumbres R, 2007, BRIT J HAEMATOL, V138, P749, DOI 10.1111/j.1365-2141.2007.06744.x
   MARCUS HC, 1963, ARCH OPHTHALMOL-CHIC, V69, P251
   Marcus R, GUIDELINES DIAGNOSIS
   MARGOLIS L, 1980, CANCER-AM CANCER SOC, V45, P688, DOI 10.1002/1097-0142(19800215)45:4<688::AID-CNCR2820450412>3.0.CO;2-F
   Margolis R, 2008, CURR OPIN OPHTHALMOL, V19, P218, DOI 10.1097/ICU.0b013e3282fc261d
   Mason JO, 2003, OPHTHALMOLOGY, V110, P1241, DOI 10.1016/S0161-6420(03)00268-9
   MATSUO K, 1995, OPHTHALMOLOGICA, V209, P331, DOI 10.1159/000310653
   Matsuo T, 2009, J CLIN EXP HEMATOP, V49, P77, DOI 10.3960/jslrt.49.77
   MATZKIN DC, 1994, OPHTHALMOLOGY, V101, P850
   Merle-Béral H, 2004, BRIT J HAEMATOL, V124, P469, DOI 10.1046/j.1365-2141.2003.04800.x
   Mochizuki M, 2009, OCUL IMMUNOL INFLAMM, V17, P69, DOI 10.1080/09273940902957305
   Mruthyunjaya P, 2002, OPHTHALMOLOGY, V109, P1123, DOI 10.1016/S0161-6420(02)01033-3
   Muhaya M, 1999, CLIN EXP IMMUNOL, V116, P410
   Nasir MA, 1996, RETINA-J RET VIT DIS, V16, P324, DOI 10.1097/00006982-199616040-00008
   Nasir S, 2000, ONCOLOGY-NY, V14, P228
   Neudorfer M, 2004, ACTA OPHTHALMOL SCAN, V82, P754, DOI 10.1111/j.1600-0420.2004.00354.x
   NODA S, 1993, ANN OPHTHALMOL, V25, P30
   Nussenblatt RB, 2006, OCUL IMMUNOL INFLAMM, V14, P139, DOI 10.1080/09273940600630170
   Ohta K, 2009, OCUL IMMUNOL INFLAMM, V17, P142, DOI 10.1080/09273940802702553
   Ongkosuwito JV, 1998, BRIT J OPHTHALMOL, V82, P245, DOI 10.1136/bjo.82.3.245
   Ormerod LD, 1997, OCUL IMMUNOL INFLAMM, V5, P271, DOI 10.3109/09273949709085068
   Parikh AH, 2005, AM J OPHTHALMOL, V139, P573, DOI 10.1016/j.ajo.2004.09.047
   PETERSON K, 1993, CANCER, V72, P843, DOI 10.1002/1097-0142(19930801)72:3<843::AID-CNCR2820720333>3.0.CO;2-#
   Peyman GA, 1996, OPHTHALMIC SURG LAS, V27, P246
   Plotkin SR, 2001, LANCET ONCOL, V2, P455
   Plowman P N, 1993, Clin Oncol (R Coll Radiol), V5, P315, DOI 10.1016/S0936-6555(05)80909-4
   Plowman PN, 2002, CLIN ONCOL-UK, V14, P521, DOI 10.1053/clon.2002.0154
   QUALMAN SJ, 1983, CANCER-AM CANCER SOC, V52, P878, DOI 10.1002/1097-0142(19830901)52:5<878::AID-CNCR2820520523>3.0.CO;2-D
   Raparia K, 2009, ARCH PATHOL LAB MED, V133, P1233, DOI 10.1043/1543-2165-133.8.1233
   Rattray KM, 2000, EYE, V14, P706, DOI 10.1038/eye.2000.188
   Read RW, 2002, SURV OPHTHALMOL, V47, P81, DOI 10.1016/S0039-6257(01)00305-8
   Rivero ME, 1999, ARCH OPHTHALMOL-CHIC, V117, P616
   Rizkalla K, 2005, CAN J OPHTHALMOL, V40, P585, DOI 10.1016/S0008-4182(05)80050-X
   Rothova A, 2001, OPHTHALMOLOGY, V108, P386, DOI 10.1016/S0161-6420(00)00499-1
   Sandor V, 1998, J CLIN ONCOL, V16, P3000, DOI 10.1200/JCO.1998.16.9.3000
   Sarafzadeh S, 2010, ACTA OPHTHALMOL, V88, P705, DOI 10.1111/j.1755-3768.2009.01531.x
   Schabet M, 1999, J NEURO-ONCOL, V43, P199, DOI 10.1023/A:1006290032052
   Sen HN, 2008, OCUL IMMUNOL INFLAMM, V16, P29, DOI 10.1080/09273940801899764
   Sen HN, 2009, OCUL IMMUNOL INFLAMM, V17, P133, DOI 10.1080/09273940903108544
   Shah GK, 2001, ARCH OPHTHALMOL-CHIC, V119, P617
   Shen DF, 1998, OPHTHALMOLOGY, V105, P1664, DOI 10.1016/S0161-6420(98)99036-4
   Shen DF, 2001, MODERN PATHOL, V14, P995, DOI 10.1038/modpathol.3880424
   Sobrin L, 2005, RETINA-J RET VIT DIS, V25, P370, DOI 10.1097/00006982-200504000-00021
   Sou R, 2008, JPN J OPHTHALMOL, V52, P167, DOI 10.1007/s10384-008-0519-9
   Soussain C, 1996, LEUKEMIA LYMPHOMA, V23, P339, DOI 10.3109/10428199609054837
   Soussain C, 2001, J CLIN ONCOL, V19, P742, DOI 10.1200/JCO.2001.19.3.742
   Soussain C, 2008, J CLIN ONCOL, V26, P2512, DOI 10.1200/JCO.2007.13.5533
   Soussain C, 2012, HAEMATOL-HEMATOL J, V97, P1751, DOI 10.3324/haematol.2011.060434
   STANTON CA, 1992, ARCH OPHTHALMOL-CHIC, V110, P1614, DOI 10.1001/archopht.1992.01080230114034
   STRAUCHEN JA, 1989, CANCER-AM CANCER SOC, V63, P1918, DOI 10.1002/1097-0142(19890515)63:10<1918::AID-CNCR2820631008>3.0.CO;2-D
   Sugita S, 2009, JPN J OPHTHALMOL, V53, P209, DOI 10.1007/s10384-009-0662-y
   Taoka K, 2012, BRIT J HAEMATOL, V157, P252, DOI 10.1111/j.1365-2141.2011.08938.x
   Tempescul A, 2011, ANN HEMATOL, V90, P1117, DOI 10.1007/s00277-010-1139-2
   TRUDEAU M, 1988, AM J CLIN ONCOL-CANC, V11, P126, DOI 10.1097/00000421-198804000-00008
   Tuaillon N., 2001, Current Molecular Medicine (Hilversum), V1, P259, DOI 10.2174/1566524013363915
   Ursea R, 1997, RETINA-J RET VIT DIS, V17, P118, DOI 10.1097/00006982-199703000-00006
   VALLURI S, 1995, RETINA-J RET VIT DIS, V15, P125, DOI 10.1097/00006982-199515020-00006
   van Dongen JJM, 2003, LEUKEMIA, V17, P2257, DOI 10.1038/sj.leu.2403202
   Vasconcelos-Santos DV, 2011, J OPHTHALMIC INFLAMM, V1, P171, DOI 10.1007/s12348-011-0034-x
   Velez G, 2000, SURV OPHTHALMOL, V44, P518, DOI 10.1016/S0039-6257(00)00118-1
   Velez G, 2002, OPHTHALMIC SURG LAS, V33, P329
   Velez G, 2002, RETINA-J RET VIT DIS, V22, P37, DOI 10.1097/00006982-200202000-00007
   Vicuna-Kojchen J, 2008, BRIT J OPHTHALMOL, V92, P231, DOI 10.1136/bjo.2007.130062
   VOGEL MH, 1968, AM J OPHTHALMOL, V66, P205, DOI 10.1016/0002-9394(68)92065-5
   Wakefield D, 2009, OCUL IMMUNOL INFLAMM, V17, P6, DOI 10.1080/09273940902834413
   WALKER J, 1993, INT OPHTHALMOL, V17, P331
   Wallace DJ, 2006, INVEST OPHTH VIS SCI, V47, P2750, DOI 10.1167/iovs.05-1312
   Wallace DJ, 2006, SURV OPHTHALMOL, V51, P41, DOI 10.1016/j.survophthal.2005.11.002
   Wang JK, 2006, JPN J OPHTHALMOL, V50, P474, DOI 10.1007/s10384-005-0327-4
   Wang YJ, 2011, INT J MOL SCI, V12, P5684, DOI 10.3390/ijms12095684
   WENDER A, 1994, ARCH OPHTHALMOL-CHIC, V112, P450, DOI 10.1001/archopht.1994.01090160024009
   Whitcup SM, 1997, ARCH OPHTHALMOL-CHIC, V115, P1157, DOI 10.1001/archopht.1997.01100160327010
   WHITCUP SM, 1993, OPHTHALMOLOGY, V100, P1399
   Whitcup SM, 2000, ARCH OPHTHALMOL-CHIC, V118, P446
   White VA, 1999, ARCH OPHTHALMOL-CHIC, V117, P761
   WILSON DJ, 1992, ARCH OPHTHALMOL-CHIC, V110, P1455, DOI 10.1001/archopht.1992.01080220117032
   Winiarska M, 2008, PLOS MED, V5, DOI 10.1371/journal.pmed.0050064
   Woei-A-Jin FJSH, 2012, ANN HEMATOL, V91, P1821, DOI 10.1007/s00277-012-1479-1
   Wolf LA, 2003, OPHTHALMOLOGY, V110, P1671, DOI 10.1016/S0161-6420(03)00811-X
   Woolf DK, 2012, CLIN ONCOL-UK, V24, P339, DOI 10.1016/j.clon.2012.03.001
   Zaldivar RA, 2004, OPHTHALMOLOGY, V111, P1762, DOI 10.1016/j.ophtha.2004.03.021
NR 221
TC 149
Z9 160
U1 0
U2 9
PU ELSEVIER SCIENCE INC
PI NEW YORK
PA STE 800, 230 PARK AVE, NEW YORK, NY 10169 USA
SN 0039-6257
EI 1879-3304
J9 SURV OPHTHALMOL
JI Surv. Ophthalmol.
PD SEP-OCT
PY 2014
VL 59
IS 5
BP 503
EP 516
DI 10.1016/j.survophthal.2013.12.001
PG 14
WC Ophthalmology
WE Science Citation Index Expanded (SCI-EXPANDED)
SC Ophthalmology
GA AN6WO
UT WOS:000340739300003
PM 24560125
OA hybrid
DA 2024-06-03
ER

PT J
AU Reni, M
   Ferreri, AJM
   Garancini, P
   Villa, E
AF Reni, M
   Ferreri, AJM
   Garancini, P
   Villa, E
TI Therapeutic management of primary central nervous system lymphoma in
   immunocompetent patients: Results of a critical review of the literature
SO ANNALS OF ONCOLOGY
LA English
DT Review
DE brain neoplasms; chemotherapy; combined treatment; extranodal lymphomas;
   primary central nervous system lymphoma; radiotherapy
ID NON-HODGKINS-LYMPHOMA; PRIMARY MALIGNANT-LYMPHOMA; PRIMARY CEREBRAL
   LYMPHOMA; PRIMARY CNS LYMPHOMA; UNIVERSITY-OF-CALIFORNIA; RETICULUM-CELL
   SARCOMA; RADIATION-THERAPY; PROGNOSTIC FACTORS; BRAIN LYMPHOMA;
   RADIOTHERAPY
AB Background: The optimal treatment for primary central nervous system lymphomas (PCNSL) has not yet been defined.
   Patients and methods: Therapeutic results of 1180 immunocompetent patients (pts) with PCNSL reported in 50 series published in English between 1980 and 1995 were analysed. The impact on survival of age, treatment strategy, radiation field and doses, systemic and intrathecal chemotherapy (CHT) and treatment sequence was evaluated.
   Results: Univariate analyses showed a longer survival in pts of less than or equal to 60 years (P < 0.00001); pts treated with > 40 Gy to whole brain (WB) (P = 0.02); pts receiving > 50 Gy to the tumor bed after a WB dose > 40 Gy (P = 0.02). pts submitted to a combined treatment as opposed to CHT alone (P = 0.007)
   or radiotherapy alone (P < 0.00001); pts receiving CHT followed by radiotherapy rather than in the reverse sequence (P = 0.05); pts treated with high-dose methotrexate (HD-MTX) (P = 0.04) and pts receiving intrathecal CHT (P = 0.03). Multivariate analysis confirmed the independent prognostic value of age, WE dose, HD-MTX and intrathecal CHT.
   Conclusions: Current data confirm the prognostic value of age and appear to support the use of systemic CHT consisting of HD-MTX and intrathecal drug administration followed by 41-50 Gy to WB and > 50 Gy to the tumor bed in the treatment of PCNSL in immunocompetent pts.
C1 SAN RAFFAELE SCI INST,DEPT RADIOCHEMOTHERAPY,I-20132 MILAN,ITALY.
   SAN RAFFAELE SCI INST,UNIT EPIDEMIOL & MED STAT,I-20132 MILAN,ITALY.
C3 Vita-Salute San Raffaele University; IRCCS Ospedale San Raffaele;
   Vita-Salute San Raffaele University; IRCCS Ospedale San Raffaele
RI Ferreri, Andrés J. M./A-6662-2013; Reni, Michele/J-8296-2016
OI Reni, Michele/0000-0002-6463-0293
CR ASHBY MA, 1988, CLIN RADIOL, V39, P173
   BERRY MP, 1981, INT J RADIAT ONCOL, V7, P55, DOI 10.1016/0360-3016(81)90060-2
   BLAY JY, 1993, BRIT J CANCER, V67, P1136, DOI 10.1038/bjc.1993.209
   BOGDAHN U, 1986, ACTA NEUROL SCAND, V73, P602, DOI 10.1111/j.1600-0404.1986.tb04607.x
   BOIARDI A, 1993, J NEUROL, V241, P96, DOI 10.1007/BF00869771
   BRADA M, 1990, INT J RADIAT ONCOL, V18, P787, DOI 10.1016/0360-3016(90)90398-4
   BRAUS DF, 1992, J NEUROL, V239, P117, DOI 10.1007/BF00833908
   CHAMBERLAIN MC, 1990, ARCH NEUROL-CHICAGO, V47, P1113, DOI 10.1001/archneur.1990.00530100081017
   COX DR, 1972, J R STAT SOC B, V34, P187
   COX JD, 1974, RADIOLOGY, V112, P179, DOI 10.1148/112.1.179
   DEANGELIS LM, 1990, NEUROLOGY, V40, P80, DOI 10.1212/WNL.40.1.80
   DEANGELIS LM, 1992, J CLIN ONCOL, V10, P635, DOI 10.1200/JCO.1992.10.4.635
   DEANGELIS LM, 1994, PRINCIPLES PRACTICE, V6, P1
   DIMARCO A, 1986, TUMORI, V72, P565, DOI 10.1177/030089168607200605
   DORREEN MS, 1988, Q J MED, V67, P387
   EBY NL, 1988, CANCER-AM CANCER SOC, V62, P2461, DOI 10.1002/1097-0142(19881201)62:11<2461::AID-CNCR2820621135>3.0.CO;2-M
   Ferreri AJM, 1995, EUR J CANCER, V31A, P2008, DOI 10.1016/0959-8049(95)00346-0
   FINE HA, 1993, ANN INTERN MED, V119, P1093, DOI 10.7326/0003-4819-119-11-199312010-00007
   FINE HA, 1995, BLOOD, V86, P2873, DOI 10.1182/blood.V86.8.2873.bloodjournal8682873
   FRANK G, 1985, SURG NEUROL, V23, P3, DOI 10.1016/0090-3019(85)90150-8
   FREEMAN CR, 1986, CANCER-AM CANCER SOC, V58, P1106, DOI 10.1002/1097-0142(19860901)58:5<1106::AID-CNCR2820580521>3.0.CO;2-3
   GEYER JR, 1988, INT J RADIAT ONCOL, V15, P373
   GLASS J, 1989, J NEUROSURG, V70, P190
   GONZALEZ DG, 1983, CANCER-AM CANCER SOC, V51, P2048, DOI 10.1002/1097-0142(19830601)51:11<2048::AID-CNCR2820511115>3.0.CO;2-T
   HARDWIDGE C, 1990, CLIN NEUROPATHOL, V9, P217
   HAYAKAWA T, 1994, J NEURO-ONCOL, V19, P197, DOI 10.1007/BF01053274
   HELLE TL, 1984, J NEUROSURG, V60, P94, DOI 10.3171/jns.1984.60.1.0094
   HENRY JM, 1974, CANCER, V34, P1293, DOI 10.1002/1097-0142(197410)34:4<1293::AID-CNCR2820340441>3.0.CO;2-P
   HILL BT, 1990, INT J RADIAT ONCOL, V19, P75, DOI 10.1016/0360-3016(90)90137-9
   HOCHBERG FH, 1988, J NEUROSURG, V68, P835, DOI 10.3171/jns.1988.68.6.0835
   JELLINGER K, 1979, CLIN NEUROL NEUROSUR, V81, P173, DOI 10.1016/0303-8467(79)90005-2
   JELLINGER KA, 1992, J CANCER RES CLIN, V119, P7, DOI 10.1007/BF01209483
   KAPLAN EL, 1958, J AM STAT ASSOC, V53, P457, DOI 10.2307/2281868
   KAWAKAMI Y, 1985, J NEUROSURG, V62, P522, DOI 10.3171/jns.1985.62.4.0522
   KROGHJENSEN M, 1994, ANN ONCOL, V5, P349, DOI 10.1093/oxfordjournals.annonc.a058839
   LACHANCE DH, 1994, NEUROLOGY, V44, P1721, DOI 10.1212/WNL.44.9.1721
   LETENDRE L, 1982, CANCER-AM CANCER SOC, V49, P939, DOI 10.1002/1097-0142(19820301)49:5<939::AID-CNCR2820490518>3.0.CO;2-1
   LING SM, 1994, CANCER-AM CANCER SOC, V73, P2570, DOI 10.1002/1097-0142(19940515)73:10<2570::AID-CNCR2820731019>3.0.CO;2-1
   LITTMAN P, 1975, CANCER, V35, P1412, DOI 10.1002/1097-0142(197505)35:5<1412::AID-CNCR2820350526>3.0.CO;2-T
   LOEFFLER JS, 1985, J CLIN ONCOL, V3, P490, DOI 10.1200/JCO.1985.3.4.490
   MACKINTOSH FR, 1982, CANCER-AM CANCER SOC, V49, P586, DOI 10.1002/1097-0142(19820201)49:3<586::AID-CNCR2820490331>3.0.CO;2-C
   MCLAUGHLIN P, 1988, JNCI-J NATL CANCER I, V80, P1408, DOI 10.1093/jnci/80.17.1408
   MENDENHALL NP, 1983, CANCER-AM CANCER SOC, V52, P1993, DOI 10.1002/1097-0142(19831201)52:11<1993::AID-CNCR2820521104>3.0.CO;2-C
   MICHALSKI JM, 1990, RADIOLOGY, V176, P855, DOI 10.1148/radiology.176.3.2389047
   MILLER DC, 1994, CANCER-AM CANCER SOC, V74, P1383, DOI 10.1002/1097-0142(19940815)74:4<1383::AID-CNCR2820740432>3.0.CO;2-1
   MORGELLO S, 1989, NEUROLOGY, V39, P1190, DOI 10.1212/WNL.39.9.1190
   MURRAY K, 1986, J NEUROSURG, V65, P600, DOI 10.3171/jns.1986.65.5.0600
   NELSON DF, 1992, INT J RADIAT ONCOL, V23, P9, DOI 10.1016/0360-3016(92)90538-S
   NEUWELT EA, 1991, J CLIN ONCOL, V9, P1580, DOI 10.1200/JCO.1991.9.9.1580
   ONEILL BP, 1995, INT J RADIAT ONCOL, V33, P663, DOI 10.1016/0360-3016(95)00207-F
   OTT RJ, 1991, EUR J CANCER, V27, P1356, DOI 10.1016/0277-5379(91)90009-3
   PETO R, 1973, BIOMETRICS, V29, P579, DOI 10.2307/2529177
   PITMAN SW, 1977, CANCER TREAT REP, V61, P695
   PITTMAN KB, 1991, ONCOLOGY, V48, P184
   POLLACK IF, 1989, CANCER-AM CANCER SOC, V63, P939, DOI 10.1002/1097-0142(19890301)63:5<939::AID-CNCR2820630526>3.0.CO;2-V
   RAMPEN FHJ, 1980, EUR J CANCER, V16, P177, DOI 10.1016/0014-2964(80)90149-8
   REMICK SC, 1990, MEDICINE, V69, P345, DOI 10.1097/00005792-199011000-00003
   SAGERMAN RH, 1983, RADIOLOGY, V149, P567, DOI 10.1148/radiology.149.2.6353480
   SAGERMAN RH, 1967, RADIOLOGY, V88, P552, DOI 10.1148/88.3.542
   Sarazin M, 1995, EUR J CANCER, V31A, P2003, DOI 10.1016/0959-8049(95)00345-2
   SCHAUMBURG HH, 1972, BRAIN, V95, P199, DOI 10.1093/brain/95.2.199
   Schultz C, 1996, J CLIN ONCOL, V14, P556, DOI 10.1200/JCO.1996.14.2.556
   SELCH MT, 1994, AM J CLIN ONCOL-CANC, V17, P286, DOI 10.1097/00000421-199408000-00002
   SHIBAMOTO Y, 1990, CANCER-AM CANCER SOC, V65, P1907, DOI 10.1002/1097-0142(19900501)65:9<1907::AID-CNCR2820650906>3.0.CO;2-L
   SLEVIN ML, 1983, J CLIN ONCOL, V1, P546, DOI 10.1200/JCO.1983.1.9.546
   SOCIE G, 1990, CANCER-AM CANCER SOC, V65, P322, DOI 10.1002/1097-0142(19900115)65:2<322::AID-CNCR2820650223>3.0.CO;2-D
   SPAUN E, 1985, SURG NEUROL, V24, P646, DOI 10.1016/0090-3019(85)90123-5
   STEWART DJ, 1984, J NEURO-ONCOL, V2, P289
   TOMLINSON FH, 1995, J NEUROSURG, V82, P558, DOI 10.3171/jns.1995.82.4.0558
   UEMATSU M, 1992, ACTA ONCOL, V31, P551, DOI 10.3109/02841869209088305
   VAKILI ST, 1986, J SURG ONCOL, V33, P95, DOI 10.1002/jso.2930330208
   WATNE K, 1992, ACTA ONCOL, V31, P545, DOI 10.3109/02841869209088304
   WOODMAN R, 1985, MEDICINE, V64, P425, DOI 10.1097/00005792-198511000-00006
   YASUNAGA T, 1986, ACTA RADIOL ONCOL, V25, P23, DOI 10.3109/02841868609136372
NR 74
TC 149
Z9 159
U1 0
U2 1
PU KLUWER ACADEMIC PUBL
PI DORDRECHT
PA SPUIBOULEVARD 50, PO BOX 17, 3300 AA DORDRECHT, NETHERLANDS
SN 0923-7534
J9 ANN ONCOL
JI Ann. Oncol.
PD MAR
PY 1997
VL 8
IS 3
BP 227
EP 234
DI 10.1023/A:1008201717089
PG 8
WC Oncology
WE Science Citation Index Expanded (SCI-EXPANDED)
SC Oncology
GA WV106
UT WOS:A1997WV10600009
PM 9137790
OA hybrid
DA 2024-06-03
ER

PT J
AU Larocca, LM
   Capello, D
   Rinelli, A
   Nori, S
   Antinori, A
   Gloghini, A
   Cingolani, A
   Migliazza, A
   Saglio, G
   Cammilleri-Broet, S
   Raphael, M
   Carbone, A
   Gaidano, G
AF Larocca, LM
   Capello, D
   Rinelli, A
   Nori, S
   Antinori, A
   Gloghini, A
   Cingolani, A
   Migliazza, A
   Saglio, G
   Cammilleri-Broet, S
   Raphael, M
   Carbone, A
   Gaidano, G
TI The molecular and phenotypic profile of primary central nervous system
   lymphoma identifies distinct categories of the disease and is consistent
   with histogenetic derivation from germinal center-related B cells
SO BLOOD
LA English
DT Article
ID ACQUIRED-IMMUNODEFICIENCY-SYNDROME; EPSTEIN-BARR-VIRUS;
   NON-HODGKINS-LYMPHOMA; LATENT GENE-EXPRESSION; FINGER ENCODING GENE;
   BCL-6 PROTEIN; HMSH2; TRANSLOCATIONS; ASSOCIATION; MSH2
AB Primary central nervous system lymphoma (PCNSL) is a major cause of morbidity and mortality among human immunodeficiency virus (HIV)-infected individuals. The precise histogenetic derivation and the molecular pathogenesis of PCNSL is poorly understood. In an attempt to clarify the histogenesis and pathogenesis of these lymphomas, 49 PCNSL (26 acquired immunodeficiency syndrome [AIDS]related and 23 AIDS-unrelated) were analyzed for multiple biologic markers, which are known to bear histogenetic and pathogenetic significance for mature B-cell neoplasms. PCNSL associated frequently (50.0%) with mutations of BCL-6 5' noncoding regions, which are regarded as a marker of B-cell transition through the germinal center (GC), Expression of BCL-6 protein, which is restricted to GC B cells throughout physiologic B-cell maturation, was detected in 100% AIDS-unrelated PCNSL and in 56.2% AIDS-related cases, Notably, among AIDS-related PCNSL, expression of BCL-6 was mutually exclusive with expression of Epstein-Barr virus (EBV)-encoded latent membrane protein (LMP)-1 and, with few exceptions, also of BCL-2. All but one PCNSL expressed hMSH2, which among mature B cells selectively stains GC B cells. These data suggest that PCNSL may be frequently related to GC B cells and may be segregated into two major biologic categories based on the expression pattern of BCL-6, LMP-1, and BCL-2. BCL-6(+)/LMP-1(-)/BCL-2(-) PCNSL occur both in the presence and in the absence of HIV infection and consistently display a large noncleaved cell morphology. Conversely, BCL-6(-)/LMP-1(+)/BCL-2(+) PCNSL are restricted to HIV-infected hosts and are represented by lymphomas with immunoblastic features. These data are relevant for the pathogenesis and histogenesis of PCNSL and may be helpful to segregate distinct biologic and prognostic categories of these lymphomas, (C) 1998 by The American Society of Hematology.
C1 Univ Cattolica Sacro Cuore, Div Pathol, Inst Pathol, I-00168 Rome, Italy.
   Univ Turin, Dept Med Sci, Div Internal Med, Novara, Italy.
   Univ Cattolica Sacro Cuore, Inst Infect Dis, I-00168 Rome, Italy.
   INRCCS, Ctr Riferimento Oncol, Div Pathol, Aviano, Italy.
   Univ Turin, Dept Clin & Biol Sci, Orbassano, Italy.
   Columbia Univ, Coll Phys & Surg, Dept Pathol, Div Oncol, New York, NY USA.
   Neuropathol Lab, Paris, France.
   Univ Paris 13, Hop Avicenne, Serv Hematol Biol, Paris, France.
C3 Catholic University of the Sacred Heart; IRCCS Policlinico Gemelli;
   University of Turin; Catholic University of the Sacred Heart; IRCCS
   Policlinico Gemelli; IRCCS Aviano (CRO); University of Turin; Columbia
   University; Assistance Publique Hopitaux Paris (APHP); Hopital
   Universitaire Avicenne - APHP; Universite Paris 13
RP Univ Cattolica Sacro Cuore, Div Pathol, Inst Pathol, Largo F Vito 1, I-00168 Rome, Italy.
EM
RI Gaidano, Gianluca/AAW-2773-2021; Carbone, Antonino/AAA-2217-2019;
   Antinori, Andrea/K-7125-2016; Larocca, Luigi Maria/M-9590-2019; Capello,
   Daniela/J-4110-2012; Larocca, Luigi M/A-1577-2008; Gloghini,
   Annunziata/C-1992-2017; Gloghini, Annunziata/AAB-4809-2019; cingolani,
   antonella/J-8921-2018
OI Gaidano, Gianluca/0000-0002-4681-0151; Carbone,
   Antonino/0000-0003-2211-639X; Antinori, Andrea/0000-0003-2121-4684;
   Larocca, Luigi Maria/0000-0003-1739-4758; Capello,
   Daniela/0000-0001-9157-8753; Gloghini, Annunziata/0000-0002-7226-1942;
   Gloghini, Annunziata/0000-0002-7226-1942; cingolani,
   antonella/0000-0002-3793-2755; Saglio, Giuseppe/0000-0002-1046-3514
CR Antinori A, 1997, NEUROLOGY, V48, P687, DOI 10.1212/WNL.48.3.687
   BALLERINI P, 1993, BLOOD, V81, P166
   BARON BW, 1993, P NATL ACAD SCI USA, V90, P5262, DOI 10.1073/pnas.90.11.5262
   BERAL V, 1991, LANCET, V337, P805, DOI 10.1016/0140-6736(91)92513-2
   CAMILLERIBROET S, 1995, BLOOD, V86, P432, DOI 10.1182/blood.V86.2.432.bloodjournal862432
   CamilleriBroet S, 1997, HUM PATHOL, V28, P367, DOI 10.1016/S0046-8177(97)90137-4
   Capello D, 1997, BRIT J HAEMATOL, V99, P168, DOI 10.1046/j.1365-2141.1997.3573171.x
   CARBONE A, 1993, J CLIN ONCOL, V11, P1674, DOI 10.1200/JCO.1993.11.9.1674
   Carbone A, 1997, AM J PATHOL, V150, P155
   Carbone A, 1998, BLOOD, V91, P747
   Cattoretti G., 1997, Blood, V90, p175A
   CATTORETTI G, 1995, BLOOD, V86, P45, DOI 10.1182/blood.V86.1.45.bloodjournal86145
   Cote TR, 1996, J NATL CANCER I, V88, P675, DOI 10.1093/jnci/88.10.675
   Deangelis Lisa M., 1997, P2233
   Dent AL, 1997, SCIENCE, V276, P589, DOI 10.1126/science.276.5312.589
   FINE HA, 1998, LYMPHOMAS, P481
   FISHEL R, 1993, CELL, V75, P1027, DOI 10.1016/0092-8674(93)90546-3
   FLENGHI L, 1995, AM J PATHOL, V147, P405
   Gaidano G., 1997, Annals of Oncology, V8, pS105, DOI 10.1023/A:1008254921497
   Gaidano G, 1997, BLOOD, V89, P3755
   GAIDANO G, 1994, BLOOD, V84, P397
   Gaidano G, 1996, AIDS, V10, P941, DOI 10.1097/00002030-199610090-00003
   GEDDES JF, 1992, J CLIN PATHOL, V45, P587, DOI 10.1136/jcp.45.7.587
   Goplen AK, 1997, J ACQ IMMUN DEF SYND, V14, P351, DOI 10.1097/00042560-199704010-00007
   HAMILTONDUTOIT SJ, 1993, AM J PATHOL, V143, P1072
   HARRIS NL, 1994, BLOOD, V84, P1361
   ITOYAMA T, 1994, CANCER-AM CANCER SOC, V73, P455, DOI 10.1002/1097-0142(19940115)73:2<455::AID-CNCR2820730234>3.0.CO;2-0
   JELLINGER KA, 1992, J CANCER RES CLIN, V119, P7, DOI 10.1007/BF01209483
   KERCKAERT JP, 1993, NAT GENET, V5, P66, DOI 10.1038/ng0993-66
   Leach FS, 1996, CANCER RES, V56, P235
   Lennert K., 1992, HISTOPATHOLOGY NONHO
   LEVINE AM, 1993, JNCI-J NATL CANCER I, V85, P1382, DOI 10.1093/jnci/85.17.1382
   MACMAHON EME, 1991, LANCET, V338, P969, DOI 10.1016/0140-6736(91)91837-K
   Marra G, 1996, ONCOGENE, V13, P2189
   Migliazza A., 1997, Blood, V90, p177A
   Migliazza A, 1995, P NATL ACAD SCI USA, V92, P12520, DOI 10.1073/pnas.92.26.12520
   MILLER DC, 1992, J NEUROONCOL, V12, P264
   MILLER SA, 1988, NUCLEIC ACIDS RES, V16, P1215, DOI 10.1093/nar/16.3.1215
   MORGELLO S, 1992, AM J PATHOL, V141, P441
   ROSENBERG SA, 1982, CANCER, V49, P2112
   Thibodeau SN, 1996, CANCER RES, V56, P4836
   Volpe G, 1996, GENE CHROMOSOME CANC, V16, P21, DOI 10.1002/(SICI)1098-2264(199605)16:1<21::AID-GCC3>3.0.CO;2-5
   WILSON TM, 1995, CANCER RES, V55, P5146
   Ye BH, 1997, NAT GENET, V16, P161, DOI 10.1038/ng0697-161
   YE BH, 1993, SCIENCE, V262, P747, DOI 10.1126/science.8235596
   YE BHH, 1993, CANCER RES, V53, P2732
   BLOOD
NR 47
TC 147
Z9 151
U1 0
U2 3
PU AMER SOC HEMATOLOGY
PI WASHINGTON
PA 2021 L ST NW, SUITE 900, WASHINGTON, DC 20036 USA
SN 0006-4971
EI 1528-0020
J9 BLOOD
JI Blood
PD AUG 1
PY 1998
VL 92
IS 3
BP 1011
EP 1019
PG 9
WC Hematology
WE Science Citation Index Expanded (SCI-EXPANDED)
SC Hematology
GA 104UY
UT WOS:000075035300035
PM 9680371
DA 2024-06-03
ER

PT J
AU Correa, DD
   DeAngelis, LM
   Shi, W
   Thaler, H
   Glass, A
   Abrey, LE
AF Correa, DD
   DeAngelis, LM
   Shi, W
   Thaler, H
   Glass, A
   Abrey, LE
TI Cognitive functions in survivors of primary central nervous system
   lymphoma
SO NEUROLOGY
LA English
DT Article
ID PRIMARY CNS LYMPHOMA; RADIATION-THERAPY; CRANIAL IRRADIATION;
   CHEMOTHERAPY; RADIOTHERAPY; ABNORMALITIES; PERFORMANCE; DEMENTIA;
   CANCER; MR
AB Background: The standard treatment for primary CNS lymphoma (PCNSL) involves high-dose methotrexate-based (MTX) chemotherapy and whole brain radiotherapy (WBRT). This combined regimen prolongs patient survival, but also carries a substantial risk for delayed neurotoxicity particularly in the elderly. However, cognitive outcome evaluations have not been included in most clinical trials. Objective: To assess cognitive functioning and quality of life in PCNSL survivors treated either with WBRT MTX-based chemotherapy or chemotherapy alone. Methods: Twenty-eight PCNSL patients in disease remission received a post-treatment baseline neuropsychological evaluation, and a subset of patients were available for an 8-month follow-up evaluation. Assessment of quality of life and extent of white matter disease on MRI were also performed. Results: Patients displayed mild to moderate impairments across several cognitive domains. These were of sufficient severity to reduce quality of life in half of the patient sample. Comparisons according to treatment type revealed more pronounced cognitive impairment, particularly in the memory and attention/executive domains, among patients treated with WBRT +/- chemotherapy. Extent of white matter disease correlated with attention/executive, memory, and language impairment. Conclusions: PCNSL survivors treated with WBRT chemotherapy displayed more pronounced cognitive dysfunction than patients treated with MTX-based chemotherapy alone.
C1 Mem Sloan Kettering Canc Ctr, Dept Neurol, ABPPCN, New York, NY 10021 USA.
   Mem Sloan Kettering Canc Ctr, Dept Epidemiol & Biostat, New York, NY 10021 USA.
   Cornell Univ, Weill Med Coll, Dept Neurol & Neurosci, New York, NY USA.
C3 Memorial Sloan Kettering Cancer Center; Memorial Sloan Kettering Cancer
   Center; Cornell University; Weill Cornell Medicine
RP Mem Sloan Kettering Canc Ctr, Dept Neurol, ABPPCN, 1275 York Ave, New York, NY 10021 USA.
EM
OI DeAngelis, Lisa/0000-0002-6290-7818
CR Abrey LE, 1998, J CLIN ONCOL, V16, P859, DOI 10.1200/JCO.1998.16.3.859
   Ahles TA, 2002, J CLIN ONCOL, V20, P485, DOI 10.1200/JCO.2002.20.2.485
   [Anonymous], 1989, CLIN NEUROPSYCHOL, DOI DOI 10.1080/13854048908403285
   Armstrong CL, 2002, NEUROLOGY, V59, P40, DOI 10.1212/WNL.59.1.40
   BARONA A, 1984, J CONSULT CLIN PSYCH, V52, P885, DOI 10.1037/0022-006X.52.5.885
   Beck AT., 1987, Beck depression inventory
   Benedict RHB, 1998, CLIN NEUROPSYCHOL, V12, P43, DOI 10.1076/clin.12.1.43.1726
   Benton A. L., 1989, Multilingual aphasia examination manual
   CONSTINE LS, 1988, INT J RADIAT ONCOL, V15, P319, DOI 10.1016/S0360-3016(98)90011-6
   Corry J, 1998, INT J RADIAT ONCOL, V41, P615, DOI 10.1016/S0360-3016(97)00571-3
   CROSSEN JR, 1994, J CLIN ONCOL, V12, P627, DOI 10.1200/JCO.1994.12.3.627
   Deangelis L M, 1995, Oncology (Williston Park), V9, P63
   DEANGELIS LM, 1992, J CLIN ONCOL, V10, P635, DOI 10.1200/JCO.1992.10.4.635
   DeAngelis LM, 2002, J CLIN ONCOL, V20, P4643, DOI 10.1200/JCO.2002.11.013
   DEANGELIS LM, 1991, RAD INJURY NERVOUS S, P361
   Duffey P, 1996, ARCH NEUROL-CHICAGO, V53, P814, DOI 10.1001/archneur.1996.00550080140022
   FAZEKAS F, 1987, AM J NEURORADIOL, V8, P421
   Ferreri AJM, 2003, J CLIN ONCOL, V21, P2407, DOI 10.1200/JCO.2003.01.135
   Fliessbach K, 2003, ARCH NEUROL-CHICAGO, V60, P563, DOI 10.1001/archneur.60.4.563
   FOLSTEIN MF, 1975, J PSYCHIAT RES, V12, P189, DOI 10.1016/0022-3956(75)90026-6
   Freilich RJ, 1996, NEUROLOGY, V46, P435, DOI 10.1212/WNL.46.2.435
   Golden C.J., 1978, Stroop color and word test
   Herrlinger U, 1998, ACTA NEUROL SCAND, V97, P257
   Kaplan E.F., 1983, The Boston Naming test, V2nd
   KLOVE H, 1963, MED CLIN N AM, V47, P1647, DOI 10.1016/S0025-7125(16)33515-5
   Korfel A, 2001, ANN HEMATOL, V80, pB20
   LEIBEL SA, 1987, J NEUROSURG, V66, P1, DOI 10.3171/jns.1987.66.1.0001
   McAllister LD, 2000, NEUROSURGERY, V46, P51, DOI 10.1093/neurosurgery/46.1.51
   NEUWELT EA, 1991, J CLIN ONCOL, V9, P1580, DOI 10.1200/JCO.1991.9.9.1580
   Olson JE, 2002, CANCER, V95, P1504, DOI 10.1002/cncr.10851
   ONEILL BP, 1989, MAYO CLIN PROC, V64, P1005, DOI 10.1016/S0025-6196(12)61228-2
   PETERSON K, 1997, HAND CLINIC, V68, P257
   Postma TJ, 2002, NEUROLOGY, V59, P121, DOI 10.1212/WNL.59.1.121
   REITAN R. M., 1958, PERCEPT MOT SKILLS, V8, P271
   Schretlen D., 1997, Brief test of attention
   SHELINE GE, 1980, INT J RADIAT ONCOL, V6, P1215, DOI 10.1016/0360-3016(80)90175-3
   SUNDERLAND T, 1989, J AM GERIATR SOC, V37, P725, DOI 10.1111/j.1532-5415.1989.tb02233.x
   Surma-aho O, 2001, NEUROLOGY, V56, P1285, DOI 10.1212/WNL.56.10.1285
   Wechsler D., 2009, WMS-IV: Wechsler Memory Scale-Fourth Edition
   WEITZNER MA, 1995, CANCER, V75, P1151, DOI 10.1002/1097-0142(19950301)75:5<1151::AID-CNCR2820750515>3.0.CO;2-Q
   Weitzner MA, 1997, PSYCHO-ONCOL, V6, P169, DOI 10.1002/(SICI)1099-1611(199709)6:3<169::AID-PON269>3.3.CO;2-R
NR 41
TC 146
Z9 154
U1 0
U2 9
PU LIPPINCOTT WILLIAMS & WILKINS
PI PHILADELPHIA
PA TWO COMMERCE SQ, 2001 MARKET ST, PHILADELPHIA, PA 19103 USA
SN 0028-3878
EI 1526-632X
J9 NEUROLOGY
JI Neurology
PD FEB 24
PY 2004
VL 62
IS 4
BP 548
EP 555
DI 10.1212/01.WNL.0000109673.75316.D8
PG 8
WC Clinical Neurology
WE Science Citation Index Expanded (SCI-EXPANDED)
SC Neurosciences & Neurology
GA 778JR
UT WOS:000189238900006
PM 14981169
DA 2024-06-03
ER

PT J
AU Ferreri, AJM
   Abrey, LE
   Blay, JY
   Borisch, B
   Hochman, J
   Neuwelt, EA
   Yahalom, J
   Zucca, E
   Cavalli, F
   Armitage, J
   Batchelor, T
AF Ferreri, AJM
   Abrey, LE
   Blay, JY
   Borisch, B
   Hochman, J
   Neuwelt, EA
   Yahalom, J
   Zucca, E
   Cavalli, F
   Armitage, J
   Batchelor, T
TI Summary statement on primary central nervous system lymphomas from the
   Eighth International Conference on Malignant Lymphoma, Lugano,
   Switzerland, June 12 to 15, 2002
SO JOURNAL OF CLINICAL ONCOLOGY
LA English
DT Article
ID PRIMARY CNS LYMPHOMA; HIGH-DOSE METHOTREXATE; NON-HODGKINS-LYMPHOMA;
   CENTER B-CELLS; RADIATION-THERAPY; BLOOD-BRAIN; CONSOLIDATION
   RADIOTHERAPY; INTRAVITREAL METHOTREXATE; INTRATHECAL CHEMOTHERAPY;
   CEREBROSPINAL-FLUID
AB Under the sponsorship of the International Extranodal Lymphoma Study Group, a Multidisciplinary Workshop on primary CNS lymphoma (PCNSL) with over 50 participants from Europe, North America, Israel, and Australia was held as part of the Eighth International Conference on Malignant Lymphoma in Lugano, Switzerland (June 12 to 15, 2002). The main purposes of the Workshop were to exchange the latest scientific information, to analyze methodologic issues in the design of clinical trials, to reach a consensus on treatment recommendations and prognostic factors, to discuss clinical and molecular targets for future studies, and to establish an international collaborative group to conduct laboratory and clinical investigations in PCNSL. This article summarizes the contents of the Workshop, analyzes the current knowledge on the most relevant biologic and clinical issues in PCNSL, and focuses on fundamental challenges to be addressed in future studies.
C1 Ist Sci San Raffaele, Dept Radiochemotherapy, I-20132 Milan, Italy.
   Mem Sloan Kettering Canc Ctr, Dept Neurol, New York, NY 10021 USA.
   Mem Sloan Kettering Canc Ctr, Dept Radiat Oncol, New York, NY 10021 USA.
   Oregon Hlth & Sci Univ, Dept Neurol & Neurosurg, Portland, OR 97201 USA.
   Univ Nebraska, Med Ctr, Coll Med, Omaha, NE USA.
   Harvard Univ, Massachusetts Gen Hosp, Sch Med, Dept Neurol, Boston, MA USA.
   Hop Edouard Herriot, Unite Cytokines & Canc, Lyon, France.
   Ctr Leon Berard, F-69373 Lyon, France.
   Univ Geneva, Dept Pathol, CH-1211 Geneva, Switzerland.
   Osped San Giovanni Bellinzona, Div Med Oncol, Oncol Inst So Switzerland, Bellinzona, Switzerland.
   Hebrew Univ Jerusalem, Inst Life Sci, Dept Cell & Anim Biol, IL-91904 Jerusalem, Israel.
C3 Vita-Salute San Raffaele University; IRCCS Ospedale San Raffaele;
   Memorial Sloan Kettering Cancer Center; Memorial Sloan Kettering Cancer
   Center; Oregon Health & Science University; University of Nebraska
   System; University of Nebraska Medical Center; Harvard University;
   Massachusetts General Hospital; Harvard Medical School; CHU Lyon;
   UNICANCER; Centre Leon Berard; University of Geneva; Institute of
   Oncology Research (IOR); Regional Hospital of Bellinzona & Valleys, San
   Giovanni; Hebrew University of Jerusalem
RP Ist Sci San Raffaele, Dept Radiochemotherapy, Via Olgettina 60, I-20132 Milan, Italy.
EM
RI Ferreri, Andrés J. M./A-6662-2013; Blay, Jean-Yves/N-3966-2016; Borisch,
   Bettina/AAG-8212-2019
OI Blay, Jean-Yves/0000-0001-7190-120X; Borisch,
   Bettina/0000-0002-9902-324X; Zucca, Emanuele/0000-0002-5522-6109
CR Abrey LE, 1998, J CLIN ONCOL, V16, P859, DOI 10.1200/JCO.1998.16.3.859
   Abrey LE, 2000, J CLIN ONCOL, V18, P3144, DOI 10.1200/JCO.2000.18.17.3144
   ABREY LE, 2001, P AN M AM SOC CLIN, V20, pA53
   AHO R, 1994, NEUROPATH APPL NEURO, V20, P551, DOI 10.1111/j.1365-2990.1994.tb01009.x
   Alizadeh AA, 2000, NATURE, V403, P503, DOI 10.1038/35000501
   Assaf N, 1997, VIRCHOWS ARCH, V431, P459, DOI 10.1007/s004280050124
   BALMACEDA C, 1995, ANN NEUROL, V38, P202, DOI 10.1002/ana.410380212
   Batchelor TT, 2003, CLIN CANCER RES, V9, P711
   BENKE R, 1988, INVAS METAST, V8, P159
   Bessell EM, 2002, J CLIN ONCOL, V20, P231, DOI 10.1200/JCO.20.1.231
   Bessell EM, 1996, J CLIN ONCOL, V14, P945, DOI 10.1200/JCO.1996.14.3.945
   BIONDI A, 1993, LEUKEMIA, V7, P281
   Blay JY, 1998, J CLIN ONCOL, V16, P864, DOI 10.1200/JCO.1998.16.3.864
   BLAY JY, 1993, BRIT J CANCER, V67, P1136, DOI 10.1038/bjc.1993.209
   BLAY JY, 1995, BLOOD, V86, P2922
   BORSI JD, 1987, CANCER-AM CANCER SOC, V60, P5, DOI 10.1002/1097-0142(19870701)60:1<5::AID-CNCR2820600103>3.0.CO;2-D
   Braaten KM, 2003, CLIN CANCER RES, V9, P1063
   Brada M, 1998, INT J RADIAT ONCOL, V40, P1157, DOI 10.1016/S0360-3016(98)00002-9
   Camilleri-Broet S, 1998, AM J CLIN PATHOL, V110, P607, DOI 10.1093/ajcp/110.5.607
   *CBTRUS, 2000, STAT REP PRIM BRAIN
   Cheng AL, 1998, CANCER, V82, P1946, DOI 10.1002/(SICI)1097-0142(19980515)82:10<1946::AID-CNCR19>3.0.CO;2-T
   Cher L, 1996, NEUROLOGY, V46, P1757, DOI 10.1212/WNL.46.6.1757
   DEANGELIS LM, 1990, J NEURO-ONCOL, V9, P177, DOI 10.1007/BF02427840
   DeAngelis LM, 2002, J CLIN ONCOL, V20, P4643, DOI 10.1200/JCO.2002.11.013
   Dent S, 1996, J NEURO-ONCOL, V28, P25, DOI 10.1007/BF00300443
   Doolittle ND, 2000, CANCER, V88, P637, DOI 10.1002/(SICI)1097-0142(20000201)88:3<637::AID-CNCR22>3.0.CO;2-Y
   Ferreri AJM, 1995, CANCER TREAT REV, V21, P415, DOI 10.1016/0305-7372(95)90028-4
   Ferreri AJM, 2000, ANN ONCOL, V11, P927, DOI 10.1023/A:1008376412784
   Ferreri AJM, 2002, ANN ONCOL, V13, P531, DOI 10.1093/annonc/mdf080
   Ferreri AJM, 2001, ONCOLOGY-BASEL, V60, P134, DOI 10.1159/000055310
   Ferreri AJM, 2003, J CLIN ONCOL, V21, P266, DOI 10.1200/JCO.2003.09.139
   Ferreri AJM, 2002, NEUROLOGY, V58, P1513, DOI 10.1212/WNL.58.10.1513
   Fishburne BC, 1997, ARCH OPHTHALMOL-CHIC, V115, P1152, DOI 10.1001/archopht.1997.01100160322009
   Freilich RJ, 1996, NEUROLOGY, V46, P435, DOI 10.1212/WNL.46.2.435
   Glass J, 1996, J NEURO-ONCOL, V30, P257
   GLASS J, 1994, J NEUROSURG, V81, P188, DOI 10.3171/jns.1994.81.2.0188
   Guha-Thakurta N, 1999, J NEURO-ONCOL, V43, P259, DOI 10.1023/A:1006210703827
   Herrlinger U, 2000, NEUROLOGY, V54, P1707, DOI 10.1212/WNL.54.8.1707
   Herrlinger U, 2002, ANN NEUROL, V51, P247, DOI 10.1002/ana.10102
   Hiraga S, 1999, J NEUROSURG, V91, P221, DOI 10.3171/jns.1999.91.2.0221
   Hochman J, 2001, CANCER RES, V61, P5242
   ILLERHAUS G, 2001, ONKOLOGIE, V54, P14
   Jaeckle KA, 2001, BRIT J CANCER, V84, P157, DOI 10.1054/bjoc.2000.1574
   Khan RB, 2002, J NEURO-ONCOL, V58, P175, DOI 10.1023/A:1016077907952
   Korfel A, 1998, LEUKEMIA LYMPHOMA, V30, P609, DOI 10.3109/10428199809057573
   Kraemer Dale F, 2002, Curr Neurol Neurosci Rep, V2, P216, DOI 10.1007/s11910-002-0080-8
   Larocca LM, 1998, BLOOD, V92, P1011
   LAWLER BE, 2002, P AN M AM SOC CLIN, V21, pA75
   Mead GM, 2000, CANCER, V89, P1359, DOI 10.1002/1097-0142(20000915)89:6<1359::AID-CNCR21>3.0.CO;2-9
   Migliazza A, 1995, P NATL ACAD SCI USA, V92, P12520, DOI 10.1073/pnas.92.26.12520
